# Genomic Surveillance of SARS-CoV-2 in Erie County, New York

**DOI:** 10.1101/2021.07.01.21259869

**Authors:** Natalie A. Lamb, Jonathan E. Bard, Alyssa Pohlman, Amanda Boccolucci, Donald A. Yergeau, Brandon J. Marzullo, Carleen Pope, Gale Burstein, John Tomaszewski, Norma J. Nowak, Jennifer A. Surtees

**Author notes:** Corresponding author: Jennifer A. Surtees, Department of Biochemistry, Jacobs School of Medicine and Biomedical Sciences, University at Buffalo, State University of New York, Rm 4215 - 955 Main Street, Buffalo, NY, 14203, USA. Denotes that these authors contributed equally to this manuscript (Natalie A. Lamb, Jonathan E. Bard).

## Abstract

Early in the SAR-CoV-2 pandemic, we established a whole genome sequencing pipeline to assess lineages circulating in Western New York. Initial sequences revealed entry into the region via Europe, similar to observations in New York City. However, as the pandemic progressed and variants of concern emerged, we observed distinct patterns in lineages relative to NYC. Notably, B.1.427 became dominant in Western New York, before it was displaced by B.1.1.7. Our hierarchical cluster analysis of B.1.1.7 lineages, which by May 2021 made up ∼ 80% of all cases, indicated both multiple introductions and community spread. Our work highlights the importance of widespread, regional surveillance of SARS-CoV-2 across the United States.

## Introduction

As SARS-CoV-2 moves through populations around the world, viral genomes accumulate genetic variants as a result of endogenous, low level of replication errors. Genomic information generated *during* the ongoing pandemic provides important insights into the mode and rate of viral evolution and can assist epidemiological efforts. It can also help support vaccine and diagnostic test development and can be a tool in assessing treatments. Faster and broader generation of genomic sequence can thus contribute to improved public health and can help to prepare for future outbreaks of this or other viruses.

As the SARS-CoV-2 virus spreads from person-to-person, it undergoes multiple rounds of replication, accumulating more mutations, which are specific to that lineage. Therefore, the pattern and combination of mutations provide information about 1) the molecular evolution of the virus and 2) its relatedness to other versions of the virus. Both allow us to develop phylogenetic trees, with which to infer the global path the virus has taken to infect an individual in a particular community. In this way, it is possible to predict that the virus that infected one person in the United States is derived from a lineage that came from China and is distinct from a virus that arrived via Europe. Additional mutations will accumulate within a community, making it possible to monitor community spread via viral genome sequencing.^1^ However, we do not yet have a clear picture of the mutation profile of SARS-CoV-2 or rate of genome instability.

The mutations that have been observed through global genome sequencing efforts are typically single nucleotide variants (SNVs) or nucleotide insertions or deletions (in/dels) that lead to frame shift mutations; there is some evidence for mutation clusters (nextstrain.org).^2^ There is little information about structural rearrangements that may be occurring as the virus spreads, although it is known that this type of virus does undergo recombination with some frequency. The presence of rearrangements would have significant implications for understanding the phylogeny and spread of the virus, as well as having potential functional consequences. The prediction is that beneficial (for the virus) mutations will become more prevalent in a population. However, their presence may simply be a result of genetic drift.

In coding regions, SNVs may result in synonymous changes (no change in coding sequence) or non-synonymous changes (change in amino acid residue), which may have functional, biological relevance for viral entry into the cell, infection and disease, and ultimately transmission to a new host.^3^ For example, there is some evidence that the increasingly prevalent D614G change in the Spike (S) protein, which arose in European populations,^4^ affects the stability of the protein and thus infectivity, using both pseudovirus and live, infectious virus.^4-7^ Non-coding mutations in 5’ or 3’ UTRs may also play a role in viral function by altering gene expression of genome replication signals and has not been assessed. Furthermore, *combinations* of mutations could have additive or synergistic effects on viral biological function. Similarly, structural rearrangements may impact viral activities.

SARS-CoV-2 accumulates mutations as it infects new individuals – each round of infection (replication) has the potential to introduce novel mutations. Those mutations that result in viable virus will be dispersed and allow the virus to continue to spread. The rate of mutation and the variant types have functional consequences for the virus, as well as for development of vaccines against the virus. Regional variations in mutation profiles will functionally distinguish viral lineages and are important for developing public health policy and clinical decisions based on the presence and prevalence of different variants of concern and variants of interest.

## Methods and Materials

All SARS-CoV-2 samples were de-identified. This study was reviewed by the University at Buffalo Institutional Review Board and determined to be “Not Human Research” (IRB ID: STUDY00004515).

### SARS-CoV-2 Amplicon Tiling, Library preparation and Sequencing

Patient nasopharyngeal swabs are collected from various locations throughout Erie County, New York and processed at the Erie County Public Health Lab (ECPHL) following standard pipelines for COVID PCR testing. Positive samples are de-identified, re-numbered and the positive RNA samples are delivered to the University at Buffalo Genomics and Bioinformatics Core (UB GBC) for library preparation and sequencing. To ensure efficient sequencing library preparation, we presort positive samples with Ct values between 15-30 and proceed with cDNA manufacture. Samples with these Ct values provide enough template for optimal multiplex PCR whereas Ct values above 30 typically do not.

#### RNA extraction from ECPHL and Kaleida Health samples

Nasopharyngeal swabs were placed into viral storage media (KHL samples) or in viral lysis buffer (ECPHL) and delivered to the UB Genomics Core either fresh or frozen. All viral RNA extractions were performed in a BL2.1 level Biosafety cabinet. An aliquot (200 μl) of the viral media was removed and spun for 10 minutes at 1500 *x g* to pellet contaminating cells. Following spindown, 140 ul of the viral supernatant was transferred to a new tube with lysis buffer for RNA extraction using the Qiagen Viral RNA extraction kit following the manufacturer’s protocol. Carrier RNA was added to facilitate efficient viral RNA isolation. Extracted RNA was eluted from the final column using EB buffer (60 μl) and RNA was stored at -80 °C until used for cDNA synthesis and library prep.

The overall amplicon tiling and library prep is based on the ARTIC v2 protocol developed at the Wadsworth Center, New York State Department of Health with minimal modifications for Illumina MiSeq sequencing.^8, 9^ Additional samples were processed using the New England Biolabs SARS-CoV-2 FS DNA library prep kit and Illumina NextSeq500 sequencing. Final libraries were cleaned up twice using AMPURE XP Beads and then quantified using Invitrogen Quant-iT fluorescence and NGS Agilent Fragment Analyzer. The final libraries were pooled (up to 192 per sequencing run) to 10 nM and further quantified in triplicate using the Kapa Biosystems Universal qPCR kit.

Once quantified by qPCR, pooled libraries (4 nM) were denatured using 0.2 N sodium hydroxide. Denatured libraries were diluted to a final concentration in Illumina HT1 buffer(12.5 pM for MiSeq and 1.5 pM for NextSeq). The diluted libraries were loaded onto an Illumina MiSeq sequencer PE300 V3 sequencing or the NextSeq for PE75 V1.5 sequencing in mid-output mode. Control PhiX was added at 1% for loading alignment control. Upon completion of the sequencing run, data were transferred to the high-performance computing facility (Center for Computational Research) located in the Center of Excellence building at the University at Buffalo.

### UB GBC SARS-COV-2 Bioinformatics Analysis

The GBC SARS-COV-2 analysis pipeline (https://github.com/UBGBC/fastq-to-consensus) is modelled off of the recommendations provided by the CDC SARS-COV-2 spheres working group (https://github.com/CDCgov/SARS-CoV-2_Sequencing), and is written in the python pipeline framework Snakemake. This rule-based framework establishes a complete chain of custody of all commands issued with version control, while integrating directly into CCR’s SLURM workload manager. To briefly describe the pipeline, sequencer results are first demultiplexed using illumina’s bcl2fastq utility, producing FASTQ format files. Then, reads are checked for initial quality using fastqc, fastq_screen, and multiqc, prior to adapter removal analysis via the tool Cutadapt. Once samples are fully evaluated for sequencing run quality, each sample is aligned to the SARS-COV-2 genome build MN908947.2 using the bwa alignment tool. Next, IVAR filters ARTIC primer locations, followed by bcftools variant calling, normalization, and filtering. The GBC pipeline produces highly accurate variant call set in VCF format, capable of detecting SNPS, insertions, and deletions. In addition to calling consensus variants, low frequency variants are also captured in separate VCF format files to facilitate more in-depth downstream analysis.

Following consensus sequence generation, variant calls are inspected using Broad’s Integrated Genomics Viewer (IGV) browser, files are added to a local instance of Nextstrain,^2^ evaluated for quality and clade variants in Nextclade, and uploaded to the GISAID^10, 11^ repository for public access. In addition to Nextclade, every sample is also analyzed using the online tool Pangolin COVID-19 Lineage Assigner in order to resolve lineage placement.^12^

### Hierarchical clustering

To characterize variants that are unique within distinct lineages, hierarchical cluster analysis was performed in the R statistical programming language. Variant call files for every sample belonging to the B.1.1.7 lineage were subset, and clustered with other members of the corresponding lineage. To detect inter-lineage variation, we compared each sample’s spread of variants utilizing the Bedtools jaccard function, which generates a Jaccard index score between every sample. The Jaccard index is a measure of similarity and diversity between finite sets. The resulting similarity matrix was then used as input for hierarchical cluster analysis in RStudio.^13^

### Phylogenetic analysis

Consensus genomes were aligned using the command line version of the MAFFT multiple sequencing alignment algorithm.^14^ The resulting alignment was then used as input into the FastTree algorithm [price, 2009; price, 2010}, inferring maximum-likelihood phylogeny using the jukes-cantor distance model of nucleotide evolution, generating a newick formatted phylogenetic tree. For data visualizations, the R packages TreeIO,^15^ and ggtree,^16^ using the pangolin lineage calls^12^ as data overlays to highlight lineage specific branchpoints of the phylogenetic tree. In addition to R based phylogenetic trees, screen captures using our local instance of Nextstrain was used to highlight different variant spreads.

## Results and Discussion

### Early infections derived from Europe

In April 2020, through a partnership with Erie County Public Health Laboratories (ECPHL), 50 de-identified COVID-19+ patient samples from Erie County were prepared for sequencing. From the initial set of 50 samples, we obtained robust sequence data from 32 genomes. These data were integrated in to the Nextstrain database^2^ to predict phylogenetic relationships with viral genomes worldwide. The majority of samples are predicted to have arrived in the United States via Europe (**Fig. 1**), while a handful arrived via China or Singapore. This was consistent with European origins of SARS-CoV-2 cases in New York City^17^ and was distinct from migration patterns on the west coast of the United States, where early infections were predicted to have arrived via Asia.^18^ Notably, all of the viruses derived from Europe contained the D614G Spike protein mutation (**Fig. 2**).^19^

**Fig. 1.**
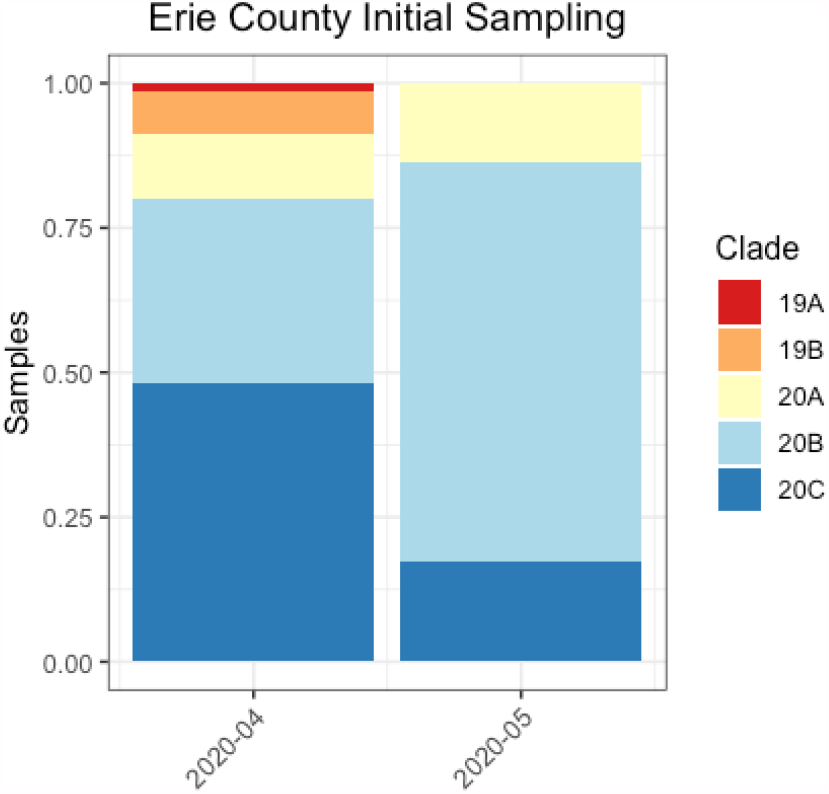
Early Samples were largely from the 20A/B/C lineages. Nextstrain clade assignments of initial sampling suggest European transmission into Erie County.

**Fig. 2.**
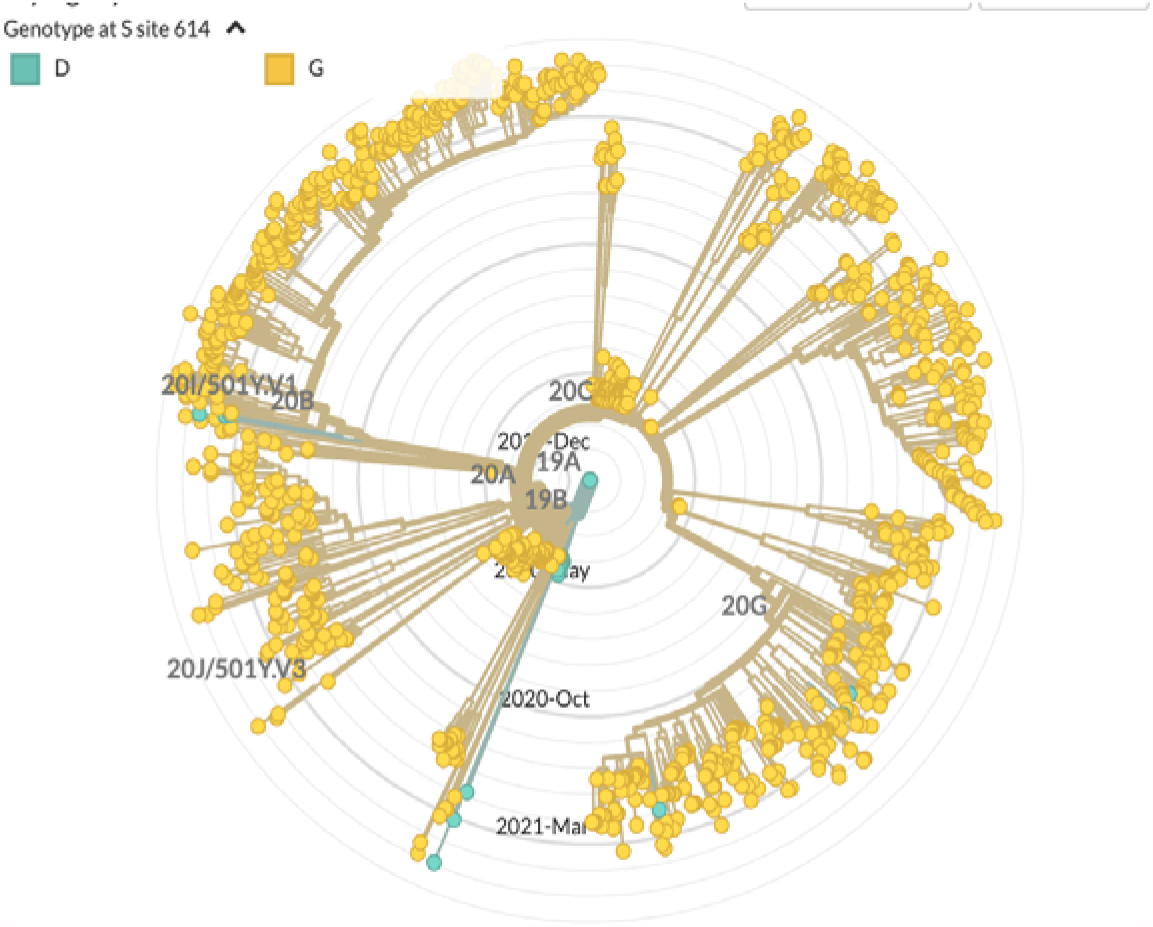
Samples predicted to have arrived from Europe (beige clades) all contain the D614G change (yellow circle) in the Spike protein. None of the samples from Asia (teal clades) contain this change (teal circles).

We received an additional ∼ 200 samples from April through early July 2020. The majority of these samples were placed in Nextstrain clades 20A, 20B, 20C, with the largest proportion in 20C. The 20A clade was descended from the original 19A clade, distinguished by signature C3037T, C14408T and A23403G mutations; it emerged and dominated the major European outbreak in early 2020. 20B emerged as a second European clade distinguished by G28881A, G28882A, and G28883C mutations. 20C was primarily North American and was characterized by C1059T and G25563T mutations.

Notably, positive SARS-CoV-2 cases were very low in Erie County and WNY during the summer of 2020. The Wadsworth Center, which was doing spot sequencing from around New York State, similarly saw a significant drop-off in samples from Erie County to sequence over the summer. As a result, there is a significant gap in genomic sequence information.

Unfortunately, cases began to increase again in the fall of 2020, with significant spikes in late November and December, providing samples for genomic surveillance. Sequencing from late 2020 and into 2021 revealed that the majority of samples in Erie County could be assigned to the 20G clade, which is derived from 20C and is concentrated in the United States (**Figure 3**). 20G is characterized by mutations resulting in the following residues changes: ORF1b N1653D, ORF3a G172V, N67S and N199L.

**Fig. 3.**
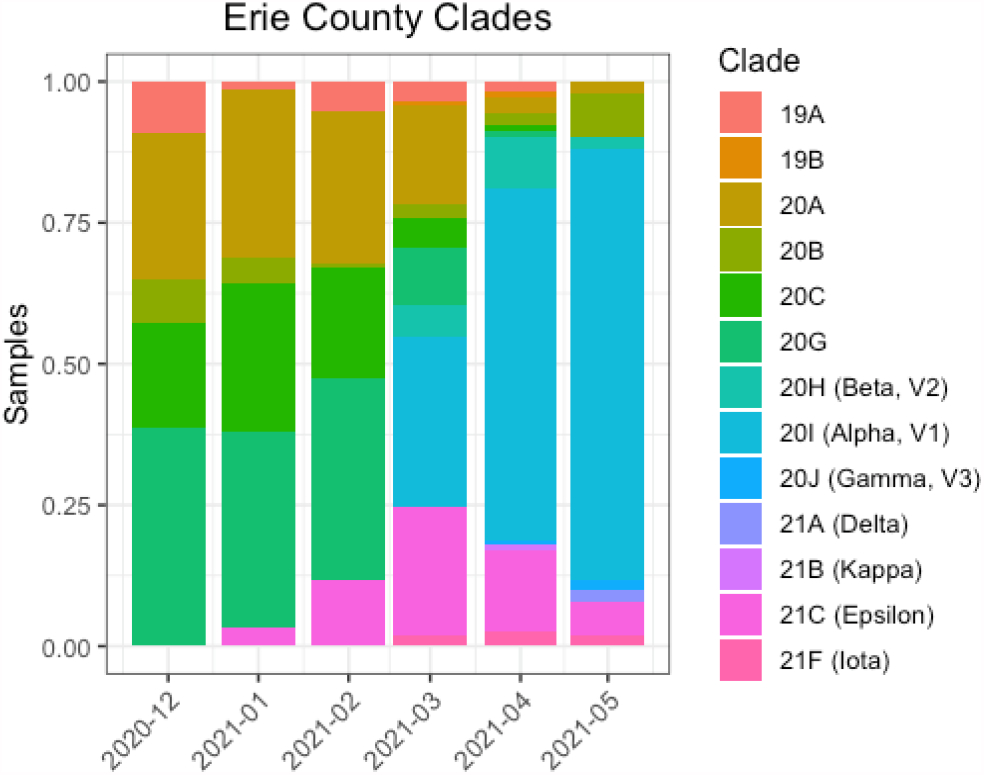
Nextstrain clade breakdown of Erie County samples. Transitions from 20C/20G to variants of concern have occurred rapidly over the span of five months.

### Detection of variants of concern and variants of interest in early 2021

In 2021, we adopted the pangolin nomenclature in our bioinformatic pipeline, which provided more granularity as SARS-CoV-2 lineages have continued to emerge. A subset of those lineages, e.g. B.1.1.7, B.1.351, P.1, and so forth, are so-called variants of concern (VOC), as defined by WHO and the CDC. These lineages have been associated with increased transmissibility and resistance to some treatment options ^20^ and have become increasingly prevalent in many regions since late 2020. In January, 2021, out of ninety-one samples sequenced, we identified five positive samples assigned to the B.1.427/429 lineages (5.5%), the so-called California variants **(Figure 4)**^21, 22^ These lineages are classified as variants of concern by the CDC and variants of interest by the WHO. Three were B.1.427 samples collected on Jan. 15; these three samples had identical genomic sequences, consistent with a common source of infection. Two B.1.429 samples were collected on Jan. 20; the genomic sequences were identical. We also identified one P.2 variant, currently designated a variant of interest by WHO and the CDC, originally identified in Brazil.

**Fig. 4.**
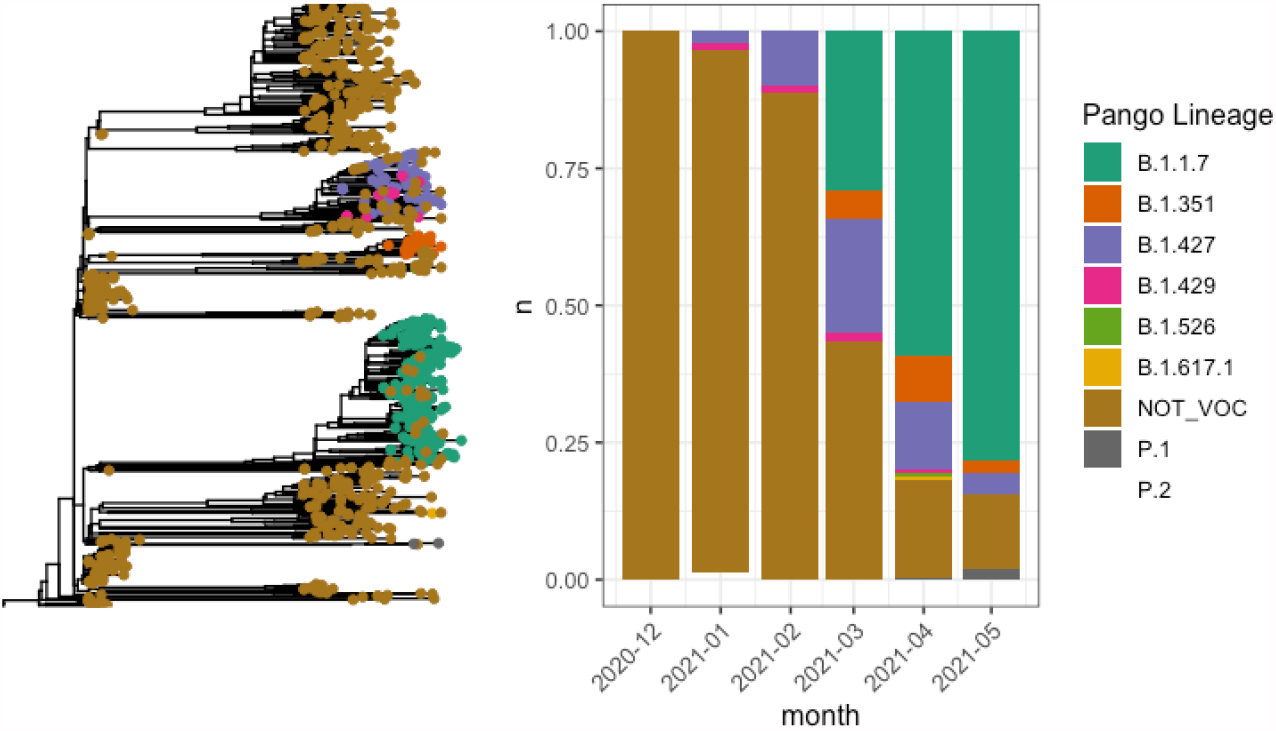
Detection of VOCs in Erie County clade breakdown of Erie County samples. Rapid transitions to variants of interest/concern occurred, with B.1.1.7 becoming the dominant variant as of April 2021.

Through February and March, the number of variants continued to increase in WNY. We also began to sequence SARS-CoV-2 samples from Kaleida Health. In February, an additional thirteen B.1.427 samples and one B.1.429 samples were collected out of 99 samples (14%) from Erie County Public Health Laboratory (ECPHL); no other variants of concern were observed. From Kaleida Health Laboratory (KHL), three of the 40 samples sequenced (7.5%) were B.427, for an overall frequency of 12.2% variants of concern – all B.1.427/429. In March, the proportion of variants of concern increased significantly and additional variants emerged in the samples. From ECPHL, 59/85 (69%) were variants of concern (24 x B.1.427, 34 x B.1.1.7, 1 x B.1.351). Two samples were variant of interest, B.1.526.1. We observed similar trends in the KHL samples; 29/55 (52%) were variants of concern (13 x B.1.427, 11 x B.1.1.7, 5 x B.351). One sample was B.1.526.1; two were B.1.526.2. These data indicate that the proportions of variants observed from ECPHL is representative of WNY. Combined, our WNY data for March showed a marked shift in the variant profile. The frequency of VOC was 62.9%, a dramatic increase from the 12.2% in the previous month. There was also a broader distribution, with the appearance of B.1.1.7 (45/140), B.1.351 (6/140) in addition to B.1.427 (37/140). We also noted 5 so-called NY variants of interest (3 x B.1.526.1, 2 x B.1.526.2). In April, the variants, and particularly B.1.1.7, had taken over. 35/38 (92.1%) samples sequenced were variants (8 x B.1.427, 2 x B.1.351, 25 x B.1.1.7). This transition to B.1.1.7 suggests that this variant is able to outcompete B.1.427, as has been seen in other places, including California.^9, 23^ Indeed, of the 51 samples sequencing May 2021, 80% were called B.1.1.7, and an additional 8% corresponded to other VOC lineages.

Although increases in VOCs and VOIs were observed across the country, there were clearly regional differences in the types of variants observed. For example, in WNY, primarily B.1.1.7 and B.1.427 variants were observed. This is in direct contrast to the variant profiles seen in NYC, where cases were dominated by B.1.526 lineages in addition to B.1.1.7. Smaller numbers of B.1.351 were observed in both NYC and WNY. This highlights the importance of regional surveillance to provide the most relevant data to clinicians in an area. (Figure 5).

**Fig. 5.**
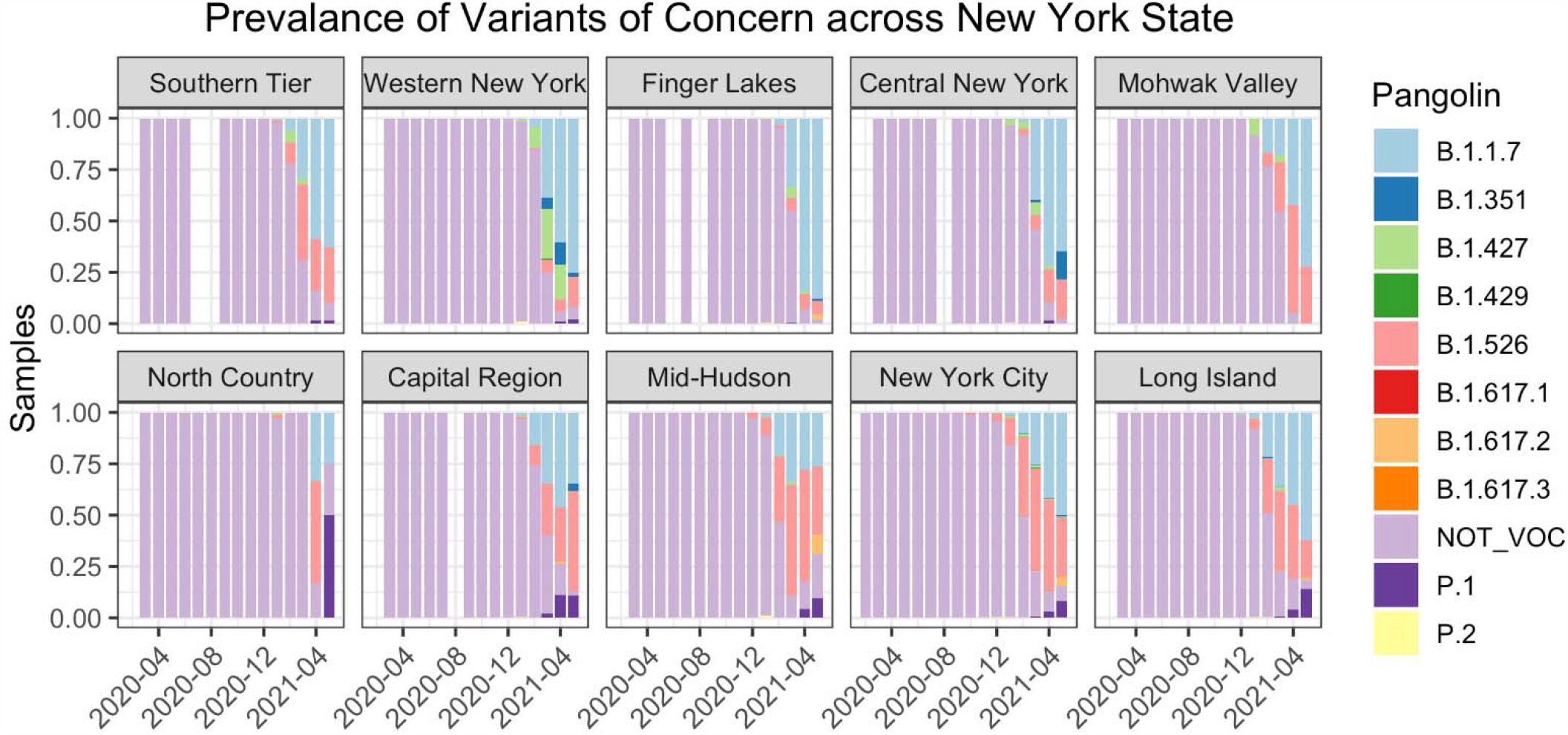
Spread of VOCs across NYS 8 regions. Distinct differences exist in the strains of COVID-19 circulating at opposite ends of New York State. N = 44,648 samples retrieved from GISAID.

We note that the samples sequenced are essentially “samples of convenience” and are therefore not necessarily representative of the absolute percentages of variants of concern and variants of interest in WNY. We were able to sequence samples to which we had access, which was only a fraction of positive cases in the region in this time period. Indeed, out of the nearly 90,000 positive diagnoses in Erie County, we have profiled just over 1% of these cases. Nonetheless, this data does provide insight into COVID-19 strain dynamics in Erie County, and makes clear that there was a sudden increase in VOCs and VOIs in early 2021, similar to other regions of the United States.

### Predicting relationships -Hierarchical cluster and maximum likelihood phylogenetic analyses

In addition to standard phylogenetic analysis, to gain additional insights into the heterogeneity within COVID-19 in Erie County, we performed hierarchical cluster analysis as an alternative mode for data visualization. Cluster analysis allowed for the identification of groups of similar variants, which combined with collection date can inform how viral evolution occurred on a local scale. We evaluated the B.1.1.7 lineage, prominent in our samples (234 samples). Variant profiles between samples were compared and a similarity matrix was generated. The similarity matrix then underwent hierarchical clustering to determine variant group sub structures. In the B.1.1.7 samples, several distinct variant patterns emerged (Figure 6). Cluster label I show most similarity to the reported B.1.1.7 variant spread and is the most dominant subgroup. The cluster labeled II has accumulated additional mutations-notably C445T (Figure 6C), C4754T and T10612A. These three mutations in combination are not observed in any strains on Nextstrain, representing a B.1.1.7 sub lineage specific to WNY. Cluster III contains a spread of variants that are observed in other B.1.1.7 lineages, found across the United States, as well as in Ghana. While the C745T mutation in cluster IV is found in other B.1.1.7 lineages on Nextstrain, the other three mutations (C6651T, A10042G, and C22388T) are unique. Each sub group has unique signatures indicating multiple B.1.1.7 strains are circulating within Erie County.

**Figure 6.**
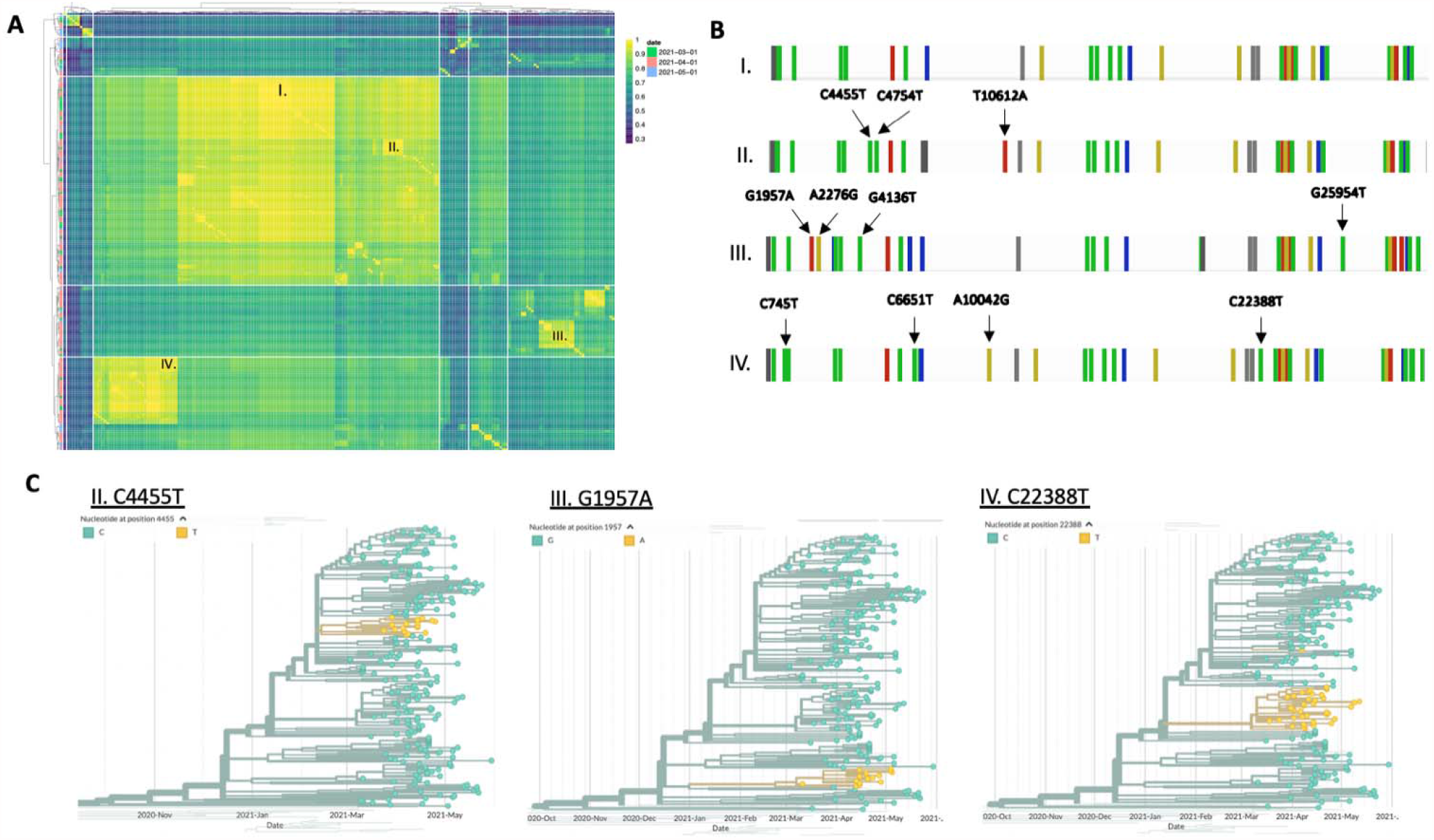
Sub lineages of B.1.1.7 in WNY. **(A)** Cluster analysis of the B.1.1.7 samples based on similarity scores of consensus variants. Unique B.1.1.7 sub-lineages are labeled I, II, III and IV. **(B)** Representative variant spreads from the four distinct B.1.1.7 sub-groups. Unique mutations are labeled with arrows. **(C)** Phylogenetic trees of three distinct subgroups labeled II, III, and IV. Samples with the designated variant are highlighted in yellow.

## Conclusion

Regional sequencing of the SARS-CoV-2 in Erie County has revealed differences in the strains circulating as compared to other NYS designated Regional Councils. These distinctions serve as evidence for the importance of region-specific surveillance as the pandemic and variants of concern continue to evolve.

Although our analysis is limited to just over 1% of all SARS-CoV-2 infections in Erie County, inspection of lineage specific mutations, such as those seen in Erie County B.1.1.7 reveal mutational patterns indicating community spread and inter-lineage competition of fitness. Thus, it is our opinion that continued surveillance of the viral genomes present at the local level will provide valuable information to health-care policy makers.

## Supporting information

Supplemental ackowledgment table#1

Supplemental acknowledgement table #2

Supplemental acknowledgement table #3

Supplemental acknowledgement table #4

Supplemental acknowledgement table #5

## Data Availability

All sequences have been uploaded to GISAID.

## Acknowledgments

This work required collaboration among several entities in the Western New York region, including Erie County Medical Center, Kaleida Health Networks, and Erie County Public Health Laboratories. In addition, we graciously thank Wadsworth Laboratories for their initial advisement of our protocol development. The immense work of all laboratories and scientists contributing to the GISAID repository (supplemental tables provided) across New York State proved and invaluable resource for out studies. All work was carried out using University at Buffalo’s Center for Computational Research supercomputing facility, located at the Center of Excellence in Bioinformatics and Life Sciences. This work was initially funded by a State University of New York Research Foundation pilot project award (COVID202044) to J.A.S. Subsequent work was funded by UB’s Genome, Environment and Microbiome Community of Excellence and Erie County Department of Health (J.A.S.).

