## Supplemental ackowledgment table#1 for "Genomic Surveillance of SARS-CoV-2 in Erie County, New York"

We gratefully acknowledge the following Authors from the Originating laboratories responsible for obtaining the specimens, as well as the Submitting laboratories where the genome data were generated and shared via GISAID, on which this research is based.

All Submitters of data may be contacted directly via www.gisaid.org

Authors are sorted alphabetically.

| Accession ID | Originating Laboratory | Submitting Laboratory | Authors |
| --- | --- | --- | --- |
| EPI_ISL_1001460 | Clinical Microbiology Laboratory, NewYork Presbyterian Hospital/Columbia University Irving Medical Center | Uhlemann Laboratory, Columbia University Irving Medical Center | Medini K. Annavajhala, Anne-Catrin Uhlemann, Hiroshi Mori, David Ho |
| EPI_ISL_1233899 | Respiratory Viruses Branch, Centers for Disease Control and Prevention | Respiratory Viruses Branch, Centers for Disease Control and Prevention | Cook,P.W., Howard,D.T., Batra,D., Rambo-Martin,B.L., de Feo,E., Antico,J., Tran,C., Tolentino,M., Wickline,S., Gietzen,K., Sickler,B., Liu,J., Allen,E., Febbo,P., Galloway,S., Washington,N.L., White,S., Levan,G., Barrett,K.S., Cirulli,E., Bolze,A., Ascencio,A., Rivera-Garcia,C., Cho,R., Nguyen,J., Wang,S., Ramirez,J., Cassens,T., Sandoval,E., Isaksson,M., Lee,W., Becker,D., Laurent,M., Lu,J., Paden,C.R., Tong,S., MacCannell,D. |
| EPI_ISL_1233937 | Division of Viral Diseases, CDC Pathogen Discovery Team | Division of Viral Diseases, CDC Pathogen Discovery Team | Queen,K., Li,Y., Tao,Y., Zhang,J., Uehara,A., Montmayeur,A., Paden,C.R., Cook,P.W., Marine,R., Sheth,M., Padilla,J., Nobles,S., Burroughs,M., Rowe,L., Wang,H., Rambo-Martin,B.L., Batra,D., Knipe,K., Lee,J.S., Tong,S. |
| EPI_ISL_1300448 | Epidemiology of Microbial Diseases, Yale School of Public Health | Yale School of Public Health | Fauver,J.R. |
| EPI_ISL_414476 | MSHS Clinical Microbiology Laboratories | MSHS Pathogen Surveillance Program | Gopi Patel, Emilia Sordillo, Melissa Gitman, Alberto Paniz-mondolfi, Matthew Hernandez, Shelcie Fabre, Jose Polanco, Ana Sylvia Gonzalez-Reiche, Zenab Khan, Nancy Francoeur, Melissa Smith, Robert Sebra, Lisa Miorin, Wen-chun Liu, Randy Albrecht, Judith Aberg, Florian Krammer, Adolfo Garcia-Sarstre, Viviana Simon, Harm van Bakel |
| EPI_ISL_414639 | NYU Langone Health | Departments of Pathology and Medicine, New York University School of Medicine | John Chen, Dacia Dimartino, Xiaojun Feng, Adriana Heguy, Megan Hogan, Emily Huang, George Jour, Christian Marier, Matthew T. Maurano, Mark J. Mulligan, Peter Meyn, Marie Samanovic-Golden, Amy Rapkiewicz, Guomiao Shen, Matija Snuderl, Gael Westby, Paul Zappile |
| EPI_ISL_415151 | MSHS Clinical Microbiology Laboratories | MSHS Pathogen Surveillance Program | Gopi Patel, Emilia Sordillo, Melissa Gitman, Alberto Paniz-mondolfi, Matthew Hernandez, Shelcie Fabre, Jose Polanco, Ana Silvia Gonzalez-Reiche, Zenab Khan, Nancy Francoeur, Melissa Smith, Robert Sebra, Lisa Miorin, Wen-chun Liu, Randy Albrecht, Judith Aberg, Florian Krammer, Adolfo Garcia-Sarstre, Viviana Simon, Harm van Bakel |
| EPI_ISL_416830, EPI_ISL_416831, EPI_ISL_416832 | NYU Langone Health | Department of Pathology and Medicine, New York University School of Medicine | John Chen, Dacia Dimartino, Xiaojun Feng, Adriana Heguy, Megan Hogan, Emily Huang, George Jour, Christian Marier, Matt Maurano, Mark Mulligan, Peter Meyn, Marie Samanovic-Golden, Amy Rapkiewicz, Guomiao Shen, Matija Snuderl, Gael Westby, Paul Zappile |
| EPI_ISL_418190, EPI_ISL_418191, EPI_ISL_418192, EPI_ISL_418193, EPI_ISL_418194, EPI_ISL_418195, EPI_ISL_418196, EPI_ISL_418197, EPI_ISL_418198, EPI_ISL_418199, EPI_ISL_418200, EPI_ISL_418201, EPI_ISL_418202, EPI_ISL_418203, EPI_ISL_418204, EPI_ISL_418205, EPI_ISL_418254 | | | |
| see above | NYU Langone Health | Department of Pathology and Medicine, New York University School of Medicine | Margaret Black, John Cadley, Paolo Cotzia, John Chen, Dacia Dimartino, Xiaojun Feng, Adriana Heguy, Megan Hogan, Emily Huang, George Jour, Christian Marier, Matthew T. Maurano, Mark J. Mulligan, Peter Meyn, Jared Pinnell, Amy Rapkiewicz, Marie Samanovic-Golden, Antonio Serrano, Guomiao Shen, Matija Snuderl, Nick Vulpescu, Gael Westby, Paul Zappile |
| EPI_ISL_418968, EPI_ISL_418969, EPI_ISL_418970, EPI_ISL_418971, EPI_ISL_418972, EPI_ISL_418973, EPI_ISL_418974, EPI_ISL_418975, EPI_ISL_418976, EPI_ISL_418977, EPI_ISL_418978, EPI_ISL_418979, EPI_ISL_418980 | | | |
| see above | NYU Langone Health | Department of Pathology and Medicine, New York University School of Medicine | Maria Aguero-Rosenfeld, Margaret Black, John Cadley, Paolo Cotzia, John Chen, Dacia Dimartino, Xiaojun Feng, Adriana Heguy, Megan Hogan, Emily Huang, George Jour, Christian Marier, Matthew T. Maurano, Mark J. Mulligan, Peter Meyn, Jared Pinnell, Sitharam Ramaswami, Amy Rapkiewicz, Marie Samanovic-Golden, Antonio Serrano, Guomiao Shen, Matija Snuderl, Nick Vulpescu, Gael Westby, Paul Zappile, Yutong Zhang |
| EPI_ISL_419523 | Yale Clinical Virology Laboratory | Grubaugh Lab - Yale School of Public Health | Joseph Fauver, Anderson Brito, Tara Alpert, Chantal Vogels, Ellen Foxman, Albert Ko, Marie Landry, Nathan Grubaugh |
| EPI_ISL_419696, EPI_ISL_419697, EPI_ISL_419698, EPI_ISL_419699, EPI_ISL_419700, EPI_ISL_419701, EPI_ISL_419702, EPI_ISL_419703, EPI_ISL_419704, EPI_ISL_419705 | NYU Langone Health | Departments of Pathology and Medicine, New York University School of Medicine | Maria Aguero-Rosenfeld, Margaret Black, John Cadley, Paolo Cotzia, John Chen, Dacia Dimartino, Xiaojun Feng, Adriana Heguy, Megan Hogan, Emily Huang, George Jour, Christian Marier, Matthew T. Maurano, Mark J. Mulligan, Peter Meyn, Jared Pinnell, Sitharam Ramaswami, Amy Rapkiewicz, Marie Samanovic-Golden, Antonio Serrano, Guomiao Shen, Matija Snuderl, Nick Vulpescu, Gael Westby, Paul Zappile, Yutong Zhang |
| EPI_ISL_420090, EPI_ISL_420093, EPI_ISL_420094 | Yale Clinical Virology Laboratory | Grubaugh Lab - Yale School of Public Health | Joseph Fauver, Anderson Brito, Tara Alpert, Chantal Vogels, Ellen Foxman, Albert Ko, Marie Landry, Nathan Grubaugh |
| EPI_ISL_420293 | Wildlife Conservation Society, Bronx Zoo | Diagnostic Virology Laboratory, United States Department of Agriculture, National Veterinary Services Laboratories | Patrick K. Mitchell, Renee R. Anderson, Brittany Chilson, Roopa Venugopalan, D. G. Diel, Laura B. Goodman, L. Wang, F. Yuan, Y. Fang, Mary Lea Killian, Kerrie Franzen, Nichole Hines Bergeson, Ivan Kuzmin, Melinda Jenkins-Moore, Tod P. Stuber |
| EPI_ISL_420296, EPI_ISL_420297, EPI_ISL_420298, EPI_ISL_420299, EPI_ISL_420300, EPI_ISL_420301, EPI_ISL_420302, EPI_ISL_420307, EPI_ISL_420308, EPI_ISL_420309 | NYU Langone Health | Departments of Pathology and Medicine, New York University School of Medicine | Maria Aguero-Rosenfeld, Margaret Black, John Cadley, Paolo Cotzia, John Chen, Dacia Dimartino, Xiaojun Feng, Adriana Heguy, Megan Hogan, Emily Huang, George Jour, Christian Marier, Matthew T. Maurano, Mark J. Mulligan, Peter Meyn, Jared Pinnell, Sitharam Ramaswami, Amy Rapkiewicz, Marie Samanovic-Golden, Antonio Serrano, Guomiao Shen, Matija Snuderl, Nick Vulpescu, Gael Westby, Paul Zappile, Yutong Zhang |
| EPI_ISL_420570, EPI_ISL_420571, EPI_ISL_420572, EPI_ISL_420573, EPI_ISL_420574, EPI_ISL_420575, EPI_ISL_420576, EPI_ISL_420577, EPI_ISL_420578, EPI_ISL_420579, EPI_ISL_420580, EPI_ISL_420581, EPI_ISL_420582, EPI_ISL_420584, EPI_ISL_420585, EPI_ISL_420586, EPI_ISL_420587, EPI_ISL_420588, EPI_ISL_420589, EPI_ISL_420590, EPI_ISL_420591 | | | |
| see above | NYU Langone Health | Departments of Pathology and Medicine, New York University School of Medicine | Maria Aguero-Rosenfeld, Brendan Belovarac, Margaret Black, Ludovic Boytard, John Cadley, Paolo Cotzia, John Chen, Dacia Dimartino, Xiaojun Feng, Tatyana Gindin, Adriana Heguy, Megan Hogan, Emily Huang, George Jour, Andrew Lytle, Christian Marier, Matthew T. Maurano, Mark J. Mulligan, Peter Meyn, Iman Osman, Jared Pinnell, Sitharam Ramaswami, Amy Rapkiewicz, Marie Samanovic-Golden, Antonio Serrano, Guomiao Shen, Matija Snuderl, Theodore Vougiouklakis, Nick Vulpescu, Gael Westby, Paul Zappile, Yutong Zhang |
| EPI_ISL_420793 | NYC Department of Health and Mental Hygiene | Pathogen Discovery, Respiratory Viruses Branch, Division of Viral Diseases, Centers for Disease Control and Prevention | Krista Queen, Yan Li, Ying Tao, Jing Zhang, Anne Uehara, Clinton R. Paden, Haibin Wang, Rachel Marine, Mary S. Keckler, Alison S. Laufer Halpin, Jasmine Padilla, Justin Lee, Christopher A. Elkins, Suxiang Tong |
| EPI_ISL_421348, EPI_ISL_421349, EPI_ISL_421350, EPI_ISL_421351, EPI_ISL_421352, EPI_ISL_421353, EPI_ISL_421354, EPI_ISL_421355, EPI_ISL_421356, EPI_ISL_421357, EPI_ISL_421358, EPI_ISL_421359, EPI_ISL_421360, EPI_ISL_421361, EPI_ISL_421362, EPI_ISL_421363, EPI_ISL_421364, EPI_ISL_421365, EPI_ISL_421366, EPI_ISL_421367, EPI_ISL_421368, EPI_ISL_421369, EPI_ISL_421370, EPI_ISL_421371, EPI_ISL_421372, EPI_ISL_421373, EPI_ISL_421374, EPI_ISL_421375, EPI_ISL_421376, EPI_ISL_421377, EPI_ISL_421378, EPI_ISL_421379, EPI_ISL_421380, EPI_ISL_421381, EPI_ISL_421382, EPI_ISL_421383, EPI_ISL_421384, EPI_ISL_421385, EPI_ISL_421386, EPI_ISL_421387, EPI_ISL_421388, EPI_ISL_421389, EPI_ISL_421390, EPI_ISL_421391, EPI_ISL_421392, EPI_ISL_421393, EPI_ISL_421394, EPI_ISL_421395, EPI_ISL_421396, EPI_ISL_421397, EPI_ISL_421398, EPI_ISL_421399, EPI_ISL_421400, EPI_ISL_421401, EPI_ISL_421402, EPI_ISL_421403, EPI_ISL_421404, EPI_ISL_421405, EPI_ISL_421406, EPI_ISL_421407, EPI_ISL_421408, EPI_ISL_421409, EPI_ISL_421410, EPI_ISL_421411, EPI_ISL_421412, EPI_ISL_421413, EPI_ISL_421414, EPI_ISL_421415, EPI_ISL_421416, EPI_ISL_421417, EPI_ISL_421418, EPI_ISL_421419, EPI_ISL_421420, EPI_ISL_421421, EPI_ISL_421422, EPI_ISL_421423, EPI_ISL_421424, EPI_ISL_421425, EPI_ISL_421426, EPI_ISL_421427, EPI_ISL_421428, EPI_ISL_421429, EPI_ISL_421430, EPI_ISL_421431, EPI_ISL_421432, EPI_ISL_421433, EPI_ISL_421434, EPI_ISL_421435 | | | |
| see above | MSHS Clinical Microbiology Laboratories | MSHS Pathogen Surveillance Program | Ana S. Gonzalez-Reiche, Mitchell Sullivan, Ajay Obla, Gopi Patel, Emilia Sordillo, Melissa Gitman, Alberto Paniz-mondolfi, Matthew Hernandez, Shelcie Fabre, Jose Polanco, Zenab Khan, Bremy Albuquerque, Jayeeta Dutta, Juan Soto, Shwetha Sridhar Hara, Ying-Chih Wang, Melissa Smith, Robert Sebra, Lisa Miorin, Wen-chun Liu, Randy Albrecht, Judith Aberg, Florian Krammer, Adolfo Garcia-Sarstre, Viviana Simon, Harm van Bakel |
| EPI_ISL_421577, EPI_ISL_421578, EPI_ISL_421579, EPI_ISL_421580, EPI_ISL_421581, EPI_ISL_421582, EPI_ISL_421583, EPI_ISL_421586, EPI_ISL_421587, EPI_ISL_421588, EPI_ISL_421589, EPI_ISL_421590, EPI_ISL_421591, EPI_ISL_421592 | | | |
| see above | NYU Langone Health | Departments of Pathology and Medicine, New York University School of Medicine | Maria Aguero-Rosenfeld, Brendan Belovarac, Margaret Black, Ludovic Boytard, John Cadley, Paolo Cotzia, John Chen, Dacia Dimartino, Xiaojun Feng, Tatyana Gindin, Adriana Heguy, Megan Hogan, Emily Huang, George Jour, Andrew Lytle, Christian Marier, Matthew T. Maurano, Mark J. Mulligan, Peter Meyn, Iman Osman, Jared Pinnell, Sitharam Ramaswami, Amy Rapkiewicz, Marie Samanovic-Golden, Antonio Serrano, Guomiao Shen, Matija Snuderl, Theodore Vougiouklakis, Nick Vulpescu, Gael Westby, Paul Zappile, Yutong Zhang |

| | | | |
|---|---|---|---|
| EPI_ISL_421593, EPI_ISL_421594, EPI_ISL_421595, EPI_ISL_421596, EPI_ISL_421597, EPI_ISL_421598, EPI_ISL_421599, EPI_ISL_421600, EPI_ISL_421601, EPI_ISL_421602, EPI_ISL_421603, EPI_ISL_421604, EPI_ISL_421605, EPI_ISL_421606, EPI_ISL_421607, EPI_ISL_421608, EPI_ISL_421609, EPI_ISL_421610, EPI_ISL_421611, EPI_ISL_421612, EPI_ISL_421613, EPI_ISL_421614, EPI_ISL_421615, EPI_ISL_421616, EPI_ISL_421617, EPI_ISL_421618, EPI_ISL_421620, EPI_ISL_421621, EPI_ISL_421622, EPI_ISL_421623, EPI_ISL_421624, EPI_ISL_421625, EPI_ISL_421626, EPI_ISL_421627, EPI_ISL_421628, EPI_ISL_421629, EPI_ISL_421630, EPI_ISL_421631, EPI_ISL_421632, EPI_ISL_421633, EPI_ISL_421634, EPI_ISL_421635 | | | |
| see above | MSHS Clinical Microbiology Laboratories | MSHS Pathogen Surveillance Program | Ana S. Gonzalez-Reiche, Mitchell Sullivan, Ajay Obla, Gopi Patel, Emilia Sordillo, Melissa Gitman, Alberto Paniz-mondolfi, Matthew Hernandez, Shelcie Fabre, Jose Polanco, Zenab Khan, Bremy Albuquerque, Jayeeta Dutta, Juan Soto, Shwetha Sridhar Hara, Ying-Chih Wang, Melissa Smith, Robert Sebra, Lisa Miorin, Wen-chun Liu, Randy Albrecht, Judith Aberg, Florian Krammer, Adolfo Garcia-Sarstre, Viviana Simon, Harm van Bakel |
| EPI_ISL_421704, EPI_ISL_421705, EPI_ISL_421706, EPI_ISL_421707, EPI_ISL_421708, EPI_ISL_421709, EPI_ISL_421710, EPI_ISL_421711, EPI_ISL_421712, EPI_ISL_421713, EPI_ISL_421714, EPI_ISL_421715, EPI_ISL_421716, EPI_ISL_421717, EPI_ISL_421719, EPI_ISL_421720, EPI_ISL_421721, EPI_ISL_421722, EPI_ISL_421724, EPI_ISL_421725, EPI_ISL_421726, EPI_ISL_421727, EPI_ISL_421728, EPI_ISL_421729, EPI_ISL_421730, EPI_ISL_421731, EPI_ISL_421732, EPI_ISL_421733 | | | |
| see above | NYU Langone Health | Departments of Pathology and Medicine, New York University School of Medicine | Maria Aguero-Rosenfeld, Brendan Belovarac, Margaret Black, Ludovic Boytard, John Cadley, Paolo Cotzia, John Chen, Dacia Dimartino, Xiaojun Feng, Tatyana Gindin, Adriana Heguy, Megan Hogan, Emily Huang, George Jour, Andrew Lytle, Christian Marier, Matthew T. Maurano, Mark J. Mulligan, Peter Meyn, Iman Osman, Jared Pinnell, Sitharam Ramaswami, Amy Rapkiewicz, Marie Samanovic-Golden, Antonio Serrano, Guomiao Shen, Matija Snuderl, Theodore Vougiouklakis, Nick Vulpescu, Gael Westby, Paul Zappile, Yutong Zhang |
| EPI_ISL_422488, EPI_ISL_422489, EPI_ISL_422490, EPI_ISL_422491, EPI_ISL_422492, EPI_ISL_422493, EPI_ISL_422494, EPI_ISL_422495, EPI_ISL_422496, EPI_ISL_422497, EPI_ISL_422498, EPI_ISL_422499, EPI_ISL_422500, EPI_ISL_422501, EPI_ISL_422502, EPI_ISL_422503, EPI_ISL_422504, EPI_ISL_422505, EPI_ISL_422506, EPI_ISL_422507, EPI_ISL_422508, EPI_ISL_422509, EPI_ISL_422510, EPI_ISL_422511, EPI_ISL_422512, EPI_ISL_422513, EPI_ISL_422514, EPI_ISL_422515, EPI_ISL_422516, EPI_ISL_422517, EPI_ISL_422518, EPI_ISL_422519, EPI_ISL_422520, EPI_ISL_422521, EPI_ISL_422522, EPI_ISL_422523, EPI_ISL_422524, EPI_ISL_422525, EPI_ISL_422526, EPI_ISL_422527, EPI_ISL_422528, EPI_ISL_422529, EPI_ISL_422530, EPI_ISL_422531, EPI_ISL_422532, EPI_ISL_422533, EPI_ISL_422534, EPI_ISL_422535, EPI_ISL_422536, EPI_ISL_422537, EPI_ISL_422538, EPI_ISL_422539, EPI_ISL_422540, EPI_ISL_422541, EPI_ISL_422542, EPI_ISL_422543, EPI_ISL_422544, EPI_ISL_422545, EPI_ISL_422546, EPI_ISL_422547, EPI_ISL_422548, EPI_ISL_422549, EPI_ISL_422550, EPI_ISL_422551, EPI_ISL_422552, EPI_ISL_422553, EPI_ISL_422554, EPI_ISL_422555, EPI_ISL_422556, EPI_ISL_422557, EPI_ISL_422558, EPI_ISL_422559, EPI_ISL_422560, EPI_ISL_422561, EPI_ISL_422562 | | | |
| see above | MSHS Clinical Microbiology Laboratories | MSHS Pathogen Surveillance Program | Ana S. Gonzalez-Reiche, Mitchell Sullivan, Ajay Obla, Gopi Patel, Emilia Sordillo, Melissa Gitman, Alberto Paniz-mondolfi, Matthew Hernandez, Shelcie Fabre, Jose Polanco, Zenab Khan, Bremy Albuquerque, Jayeeta Dutta, Juan Soto, Shwetha Sridhar Hara, Ying-Chih Wang, Melissa Smith, Robert Sebra, Lisa Miorin, Wen-chun Liu, Randy Albrecht, Judith Aberg, Florian Krammer, Adolfo Garcia-Sarstre, Viviana Simon, Harm van Bakel |
| EPI_ISL_424906 | NYC Department of Health and Mental Hygiene | Pathogen Discovery, Respiratory Viruses Branch, Division of Viral Diseases, Centers for Disease Control and Prevention | Ying Tao, Clinton R. Paden, Jing Zhang, Krista Queen, Anna Uehara, Yan Li, Haibin Wang, Mary S. Keckler, Alison S. Laufer Halpin, Christopher A. Elkins, Suxiang Tong |
| EPI_ISL_424929, EPI_ISL_424930, EPI_ISL_424931, EPI_ISL_424932, EPI_ISL_424933, EPI_ISL_424934, EPI_ISL_424935, EPI_ISL_424936, EPI_ISL_424937, EPI_ISL_424938, EPI_ISL_424939, EPI_ISL_424940, EPI_ISL_424941, EPI_ISL_424942, EPI_ISL_424943, EPI_ISL_424944, EPI_ISL_424945, EPI_ISL_424946, EPI_ISL_424947, EPI_ISL_424948, EPI_ISL_424949, EPI_ISL_424950, EPI_ISL_424951, EPI_ISL_424952, EPI_ISL_424953, EPI_ISL_424954, EPI_ISL_424955, EPI_ISL_424956, EPI_ISL_424957, EPI_ISL_424958, EPI_ISL_424959, EPI_ISL_424960, EPI_ISL_424961, EPI_ISL_424962, EPI_ISL_424963, EPI_ISL_424964, EPI_ISL_424965, EPI_ISL_424966, EPI_ISL_424967, EPI_ISL_424968 | | | |
| see above | NYU Langone Health | Departments of Pathology and Medicine, New York University School of Medicine | Maria Aguero-Rosenfeld, Brendan Belovarac, Margaret Black, Ludovic Boytard, John Cadley, Paolo Cotzia, John Chen, Dacia Dimartino, Xiaojun Feng, Tatyana Gindin, Adriana Heguy, Megan Hogan, Emily Huang, George Jour, Andrew Lytle, Christian Marier, Matthew T. Maurano, Mark J. Mulligan, Peter Meyn, Iman Osman, Jared Pinnell, Sitharam Ramaswami, Amy Rapkiewicz, Marie Samanovic-Golden, Antonio Serrano, Guomiao Shen, Matija Snuderl, Theodore Vougiouklakis, Nick Vulpescu, Gael Westby, Paul Zappile, Yutong Zhang |
| EPI_ISL_425224 | Wadsworth Center, New York State Department of Health | Wadsworth Center, New York State Department of Health | Kirsten St. George, Daryl M. Lamson, Sara Griesemer, Jonathan Plitnick, Navjot Singh, Matthew D. Shudt, Erica Lasek-Nesselquist |
| EPI_ISL_426025, EPI_ISL_426026, EPI_ISL_426027, EPI_ISL_426028, EPI_ISL_426029, EPI_ISL_426030, EPI_ISL_426031, EPI_ISL_426032, EPI_ISL_426033, EPI_ISL_426034, EPI_ISL_426035, EPI_ISL_426036, EPI_ISL_426037, EPI_ISL_426038, EPI_ISL_426039, EPI_ISL_426040, EPI_ISL_426041, EPI_ISL_426042, EPI_ISL_426043, EPI_ISL_426044, EPI_ISL_426045, EPI_ISL_426046, EPI_ISL_426047, EPI_ISL_426048, EPI_ISL_426049, EPI_ISL_426050 | | | |
| see above | Wadsworth Center, New York State Department.of Health | Wadsworth Center, New York State Department.of Health | Kirsten St. George, Daryl M. Lamson, Sara Griesemer, Jonathan Plitnick, Navjot Singh, Matthew D. Shudt, Erica Lasek-Nesselquist |
| EPI_ISL_426290 | Wadsworth Center, New York State Department.of Health | Wadsworth Center, New York State Department.of Health | Kirsten St. George, Daryl M. Lamson, Sara Griesemer, Jonathan Plitnick, Navjot Singh, Matthew D. Shudt, Erica Lasek-Nesselquist |
| EPI_ISL_426291, EPI_ISL_426292, EPI_ISL_426293, EPI_ISL_426294, EPI_ISL_426295, EPI_ISL_426296, EPI_ISL_426297, EPI_ISL_426298, EPI_ISL_426299, EPI_ISL_426300, EPI_ISL_426301, EPI_ISL_426302, EPI_ISL_426303, EPI_ISL_426304, EPI_ISL_426305, EPI_ISL_426306, EPI_ISL_426307, EPI_ISL_426308, EPI_ISL_426309, EPI_ISL_426310, EPI_ISL_426311, EPI_ISL_426312, EPI_ISL_426313, EPI_ISL_426314, EPI_ISL_426315, EPI_ISL_426316, EPI_ISL_426317, EPI_ISL_426318, EPI_ISL_426319, EPI_ISL_426320, EPI_ISL_426321, EPI_ISL_426322, EPI_ISL_426323, EPI_ISL_426324, EPI_ISL_426325, EPI_ISL_426326, EPI_ISL_426327, EPI_ISL_426328 | | | |
| see above | Wadsworth Center, New York State Department.of Health | Wadsworth Center, New York State Department.of Health | Kirsten St. George, Daryl M. Lamson, Sara Griesemer, Jonathan Plitnick, Navjot Singh, Matthew D. Shudt, Erica Lasek-Nesselquist |
| EPI_ISL_426617, EPI_ISL_426618, EPI_ISL_426619, EPI_ISL_426620, EPI_ISL_426621, EPI_ISL_426622, EPI_ISL_426623, EPI_ISL_426624, EPI_ISL_426625, EPI_ISL_426626, EPI_ISL_427469, EPI_ISL_427470, EPI_ISL_427471, EPI_ISL_427472, EPI_ISL_427473, EPI_ISL_427474, EPI_ISL_427475, EPI_ISL_427476, EPI_ISL_427477, EPI_ISL_427478, EPI_ISL_427479, EPI_ISL_427480, EPI_ISL_427481, EPI_ISL_427482, EPI_ISL_427483, EPI_ISL_427484, EPI_ISL_427485, EPI_ISL_427486, EPI_ISL_427487, EPI_ISL_427488, EPI_ISL_427489, EPI_ISL_427490, EPI_ISL_427491, EPI_ISL_427492, EPI_ISL_427493, EPI_ISL_427494, EPI_ISL_427495, EPI_ISL_427496, EPI_ISL_427497, EPI_ISL_427498, EPI_ISL_427499, EPI_ISL_427500, EPI_ISL_427501, EPI_ISL_427502, EPI_ISL_427503, EPI_ISL_427504, EPI_ISL_427505, EPI_ISL_427506, EPI_ISL_427507, EPI_ISL_427508, EPI_ISL_427509, EPI_ISL_427510, EPI_ISL_427511, EPI_ISL_427512, EPI_ISL_427513, EPI_ISL_427514, EPI_ISL_427515, EPI_ISL_427516, EPI_ISL_427517, EPI_ISL_427518, EPI_ISL_427519, EPI_ISL_427520, EPI_ISL_427521, EPI_ISL_427524, EPI_ISL_427525 | | | |
| see above | NYU Langone Health | Departments of Pathology and Medicine, New York University School of Medicine | Maria Aguero-Rosenfeld, Brendan Belovarac, Margaret Black, Ludovic Boytard, John Cadley, Paolo Cotzia, John Chen, Dacia Dimartino, Xiaojun Feng, Tatyana Gindin, Emily Guzman, Adriana Heguy, Megan Hogan, Emily Huang, George Jour, Andrew Lytle, Christian Marier, Matthew T. Maurano, Mark J. Mulligan, Peter Meyn, Iman Osman, Jared Pinnell, Vanessa Raabe, Sitharam Ramaswami, Amy Rapkiewicz, Marie Samanovic-Golden, Antonio Serrano, Guomiao Shen, Matija Snuderl, Theodore Vougiouklakis, Nick Vulpescu, Gael Westby, Paul Zappile, Yutong Zhang |
| EPI_ISL_427526, EPI_ISL_427527, EPI_ISL_427528, EPI_ISL_427529, EPI_ISL_427530, EPI_ISL_427531, EPI_ISL_427532, EPI_ISL_427533, EPI_ISL_427534, EPI_ISL_427535, EPI_ISL_427536, EPI_ISL_427537, EPI_ISL_427538, EPI_ISL_427539, EPI_ISL_427540, EPI_ISL_427541, EPI_ISL_427542, EPI_ISL_427543, EPI_ISL_427544, EPI_ISL_427545, EPI_ISL_427546, EPI_ISL_427547, EPI_ISL_427548, EPI_ISL_427549, EPI_ISL_427550, EPI_ISL_427551, EPI_ISL_427552, EPI_ISL_427553, EPI_ISL_427554, EPI_ISL_427555, EPI_ISL_427556, EPI_ISL_427557, EPI_ISL_427558, EPI_ISL_427559, EPI_ISL_427560, EPI_ISL_427561, EPI_ISL_427562, EPI_ISL_427563, EPI_ISL_427564, EPI_ISL_427565, EPI_ISL_427566, EPI_ISL_427567, EPI_ISL_427568, EPI_ISL_427569, EPI_ISL_427570, EPI_ISL_427571, EPI_ISL_427572, EPI_ISL_427573, EPI_ISL_427574, EPI_ISL_427575, EPI_ISL_427576, EPI_ISL_427577, EPI_ISL_427578, EPI_ISL_427579, EPI_ISL_427580, EPI_ISL_427581, EPI_ISL_427582, EPI_ISL_427583, EPI_ISL_427584, EPI_ISL_427585, EPI_ISL_427586, EPI_ISL_427587, EPI_ISL_427588, EPI_ISL_427589, EPI_ISL_427590, EPI_ISL_427591, EPI_ISL_427592, EPI_ISL_427593, EPI_ISL_427594, EPI_ISL_427595, EPI_ISL_427596, EPI_ISL_427597, EPI_ISL_427598, EPI_ISL_427599, EPI_ISL_427600, EPI_ISL_427601, EPI_ISL_427602, EPI_ISL_427603, EPI_ISL_427604, EPI_ISL_427605, EPI_ISL_427606, EPI_ISL_427607, EPI_ISL_427608, EPI_ISL_427609, EPI_ISL_427610, EPI_ISL_427611, EPI_ISL_427612, EPI_ISL_427613, EPI_ISL_427614, EPI_ISL_427615, EPI_ISL_427616, EPI_ISL_427617, EPI_ISL_427618 | | | |
| see above | NewYork-Presbyterian & Mason Lab | Mason Lab | Daniel J. Butler, Christopher Mozsary, Cem Meydan, David Danko, Jonathan Foox, Joel Rosiene, Alon Shaiber, Matthew MacKay, Ebrahim Afshinnekoo, Fritz J. Sedlazeck, Nikolay A. Ivanov, Maria Sierra, Craig D. Westover, Krista Ryon, Benjamin Young, Chandrima Bhattacharya, Phyllis Ruggiero, Justyna Gawrys, Iman Hajirasouliha, Dmitry Meleshko, Mirella Salvatore, Dong Xu, Jenny Xiang, John Sipley, Lin Cong, Arryn Craney, Priya Velu, Lars F. Westblade, Massimo Loda, Shawn Levy, Melissa Cushing, Marcin Imielinski, Hanna Rennert, Christopher E. Mason |
| EPI_ISL_427627, EPI_ISL_427628, EPI_ISL_427629, EPI_ISL_427630, EPI_ISL_427631, EPI_ISL_427632, EPI_ISL_427633, EPI_ISL_427634, EPI_ISL_427635, EPI_ISL_427636, EPI_ISL_427637, EPI_ISL_427638, EPI_ISL_427639, EPI_ISL_427640, EPI_ISL_427641, EPI_ISL_427642, EPI_ISL_428757, EPI_ISL_428758, EPI_ISL_428759, EPI_ISL_428760, EPI_ISL_428761, EPI_ISL_428762, EPI_ISL_428763, EPI_ISL_428764, EPI_ISL_428765, EPI_ISL_428766, EPI_ISL_428767, EPI_ISL_428768, EPI_ISL_428769, EPI_ISL_428770, EPI_ISL_428771, EPI_ISL_428772, EPI_ISL_428773, EPI_ISL_428774, EPI_ISL_428775, EPI_ISL_428776, EPI_ISL_428777, EPI_ISL_428778, EPI_ISL_428779, EPI_ISL_428780, EPI_ISL_428781, EPI_ISL_428782, EPI_ISL_428783, EPI_ISL_428784, EPI_ISL_428785, EPI_ISL_428786, EPI_ISL_428787, EPI_ISL_428788, EPI_ISL_428789, EPI_ISL_428790, EPI_ISL_428791, EPI_ISL_428792, EPI_ISL_428793, EPI_ISL_428794, EPI_ISL_428795, EPI_ISL_428796, EPI_ISL_428797, EPI_ISL_428798, EPI_ISL_428799, EPI_ISL_428800, EPI_ISL_428801, EPI_ISL_428802, EPI_ISL_428803, EPI_ISL_428804, EPI_ISL_428805 | | | |
| see above | NYU Langone Health | Departments of Pathology and Medicine, New York University School of Medicine | Maria Aguero-Rosenfeld, Brendan Belovarac, Margaret Black, Ludovic Boytard, John Cadley, Paolo Cotzia, John Chen, Dacia Dimartino, Xiaojun Feng, Tatyana Gindin, Emily Guzman, Adriana Heguy, Megan Hogan, Emily Huang, George Jour, Andrew Lytle, Christian Marier, Matthew T. Maurano, Mark J. Mulligan, Peter Meyn, Iman Osman, Jared Pinnell, Vanessa Raabe, Sitharam Ramaswami, Amy Rapkiewicz, Marie Samanovic-Golden, Antonio Serrano, Guomiao Shen, Matija Snuderl, Theodore Vougiouklakis, Nick Vulpescu, Gael Westby, Paul Zappile, Yutong Zhang |
| EPI_ISL_430319, EPI_ISL_430320, EPI_ISL_430321, EPI_ISL_430322, EPI_ISL_430323, EPI_ISL_430324, EPI_ISL_430325, EPI_ISL_430326, EPI_ISL_430327, EPI_ISL_430328, EPI_ISL_430329, EPI_ISL_430330, EPI_ISL_430331, EPI_ISL_430332, EPI_ISL_430333, EPI_ISL_430334, EPI_ISL_430335, EPI_ISL_430336, EPI_ISL_430337, EPI_ISL_430338, EPI_ISL_430339, EPI_ISL_430340, EPI_ISL_430341, EPI_ISL_430342, EPI_ISL_430343, EPI_ISL_430344, EPI_ISL_430345, EPI_ISL_430346, EPI_ISL_430347, EPI_ISL_430348, EPI_ISL_430349, EPI_ISL_430350, EPI_ISL_430351, EPI_ISL_430352, EPI_ISL_430353, EPI_ISL_430354, EPI_ISL_430355, EPI_ISL_430357, EPI_ISL_430358, EPI_ISL_430359, EPI_ISL_430360, EPI_ISL_430361, EPI_ISL_430362, EPI_ISL_430363, EPI_ISL_430364, EPI_ISL_430365, EPI_ISL_430366, EPI_ISL_430367, EPI_ISL_430368, EPI_ISL_430369, EPI_ISL_430370, EPI_ISL_430372, EPI_ISL_430373, EPI_ISL_430374, EPI_ISL_430375, EPI_ISL_430376, EPI_ISL_430377, EPI_ISL_430378, EPI_ISL_430379, EPI_ISL_430380, EPI_ISL_430381, EPI_ISL_430382, EPI_ISL_430383, EPI_ISL_430384, EPI_ISL_430385, EPI_ISL_430386, EPI_ISL_430387, EPI_ISL_430388, EPI_ISL_430389, EPI_ISL_430390, EPI_ISL_430391, EPI_ISL_430392, EPI_ISL_430393, EPI_ISL_430395, EPI_ISL_430396, EPI_ISL_430397, EPI_ISL_430398, EPI_ISL_430399, EPI_ISL_430400, EPI_ISL_430402, EPI_ISL_430403, EPI_ISL_430404, EPI_ISL_430406, EPI_ISL_430407, EPI_ISL_430408, EPI_ISL_430409, EPI_ISL_430410, EPI_ISL_430411, EPI_ISL_430412, EPI_ISL_430413, EPI_ISL_430414, EPI_ISL_430415, EPI_ISL_430416, EPI_ISL_430417, EPI_ISL_430418, EPI_ISL_430419, EPI_ISL_430420, EPI_ISL_430421, EPI_ISL_430422, EPI_ISL_430423, EPI_ISL_430424, EPI_ISL_430425, EPI_ISL_430426, EPI_ISL_430427, EPI_ISL_430429, EPI_ISL_430430, EPI_ISL_430431, EPI_ISL_430432, EPI_ISL_430433, EPI_ISL_430434, EPI_ISL_435475, EPI_ISL_435476, EPI_ISL_435477, EPI_ISL_435478, EPI_ISL_435479, EPI_ISL_435480, EPI_ISL_435481, EPI_ISL_435482, EPI_ISL_435484, EPI_ISL_435485, EPI_ISL_435486, EPI_ISL_435487, EPI_ISL_435489, EPI_ISL_435490, EPI_ISL_435491, EPI_ISL_435492, EPI_ISL_435493, EPI_ISL_435494, EPI_ISL_435495, EPI_ISL_435496, EPI_ISL_435497, EPI_ISL_435498, EPI_ISL_435499, EPI_ISL_435500, EPI_ISL_435501, EPI_ISL_435502, EPI_ISL_435503, EPI_ISL_435504, EPI_ISL_435505, EPI_ISL_435506, EPI_ISL_435507, EPI_ISL_435508, EPI_ISL_435509, EPI_ISL_435510, EPI_ISL_435511, EPI_ISL_435512, EPI_ISL_435513, EPI_ISL_435514, EPI_ISL_435515, EPI_ISL_435516, EPI_ISL_435517, EPI_ISL_435518, EPI_ISL_435520, EPI_ISL_435521, EPI_ISL_435522, EPI_ISL_435523, EPI_ISL_435524, EPI_ISL_435525, EPI_ISL_435526, EPI_ISL_435527, EPI_ISL_435528, EPI_ISL_435529, EPI_ISL_435530, EPI_ISL_435531, EPI_ISL_435532, EPI_ISL_435533, EPI_ISL_435534, EPI_ISL_435535, EPI_ISL_435536, EPI_ISL_435537, EPI_ISL_435538, EPI_ISL_435539, EPI_ISL_435540, EPI_ISL_435541, EPI_ISL_435542, EPI_ISL_435543, EPI_ISL_435544, EPI_ISL_435545, EPI_ISL_435546, EPI_ISL_435547, EPI_ISL_435548, EPI_ISL_435549, EPI_ISL_435721, EPI_ISL_435722 | | | |
| see above | NYU Langone Health | Departments of Pathology and Medicine, New York University | Maria Aguero-Rosenfeld, Brendan Belovarac, Margaret Black, Ludovic Boytard, John Cadley, Paolo Cotzia, John Chen, Dacia Dimartino, Xiaojun Feng, |

| | | | |
|---|---|---|---|
| | | School of Medicine | Tatyana Gindin, Emily Guzman, Adriana Heguy, Megan Hogan, Emily Huang, George Jour, Lawrence H. Lin, Raven Luther, Andrew Lytle, Christian Marier, Matthew T. Maurano, Mark J. Mulligan, Peter Meyn, Raquel Ordonez Ciriza, Iman Osman, Jared Pinnell, Vanessa Raabe, Sitharam Ramaswami, Amy Rapkiewicz, Andre M. Ribeiro-dos-Santos, Marie Samanovic-Golden, Antonio Serrano, Guomiao Shen, Matija Snuderl, Theodore Vougiouklakis, Nick Vulpescu, Gael Westby, Paul Zappile, Yutong Zhang |
| EPI_ISL_436047, EPI_ISL_436048, EPI_ISL_436049, EPI_ISL_436050, EPI_ISL_436051, EPI_ISL_436052, EPI_ISL_436053, EPI_ISL_436054, EPI_ISL_436055, EPI_ISL_436056, EPI_ISL_436057, EPI_ISL_436058, EPI_ISL_436059, EPI_ISL_436060, EPI_ISL_436061, EPI_ISL_436062, EPI_ISL_436063, EPI_ISL_436064, EPI_ISL_436065, EPI_ISL_436066, EPI_ISL_436067, EPI_ISL_436068, EPI_ISL_436069, EPI_ISL_436070, EPI_ISL_436071, EPI_ISL_436072, EPI_ISL_436073, EPI_ISL_436074, EPI_ISL_436075, EPI_ISL_436076, EPI_ISL_436077, EPI_ISL_436078, EPI_ISL_436079, EPI_ISL_436080, EPI_ISL_436081, EPI_ISL_436082 | | | |
| see above | NYC Department of Health and Mental Hygiene | Pathogen Discovery, Respiratory Viruses Branch, Division of Viral Diseases, Centers for Disease Control and Prevention | Ying Tao, Krista Queen, Christy Harrison, Jennifer Rakeman, Clinton R. Paden, Jing Zhang, Anna Uehara, Yan Li, Haibin Wang, Jasmine Padilla, Justin Lee, Bettina Bankamp, Zachary Weiner, Suxiang Tong |
| EPI_ISL_436733, EPI_ISL_436734, EPI_ISL_436735, EPI_ISL_436736, EPI_ISL_436737, EPI_ISL_436738, EPI_ISL_436739, EPI_ISL_436740, EPI_ISL_436741, EPI_ISL_436742, EPI_ISL_436743 | | | |
| see above | NYU Langone Health | Departments of Pathology and Medicine, New York University School of Medicine | Maria Aguero-Rosenfeld, Brendan Belovarac, Margaret Black, Ludovic Boytard, John Cadley, Paolo Cotzia, John Chen, Dacia Dimartino, Xiaojun Feng, Tatyana Gindin, Emily Guzman, Adriana Heguy, Megan Hogan, Emily Huang, George Jour, Alireza Khodadadi-Jamayran, Lawrence H. Lin, Raven Luther, Andrew Lytle, Christian Marier, Matthew T. Maurano, Mark J. Mulligan, Peter Meyn, Raquel Ordonez Ciriza, Iman Osman, Jared Pinnell, Vanessa Raabe, Sitharam Ramaswami, Amy Rapkiewicz, Andre M. Ribeiro-dos-Santos, Marie Samanovic-Golden, Antonio Serrano, Guomiao Shen, Matija Snuderl, Theodore Vougiouklakis, Nick Vulpescu, Gael Westby, Paul Zappile, Yutong Zhang |
| EPI_ISL_444519 | Molecular Infectious Disease | Molecular Infectious Disease | Anderson,B.P., Rosenthal,S.H., Gerasimova,A., Kagan,R.M. and Owen,R. |
| EPI_ISL_444613, EPI_ISL_444614, EPI_ISL_444615, EPI_ISL_444616, EPI_ISL_444617, EPI_ISL_444618, EPI_ISL_444619, EPI_ISL_444620, EPI_ISL_444621, EPI_ISL_444622, EPI_ISL_444623, EPI_ISL_444624, EPI_ISL_444625, EPI_ISL_444626, EPI_ISL_444627, EPI_ISL_444628, EPI_ISL_444629, EPI_ISL_444630, EPI_ISL_444631, EPI_ISL_444632, EPI_ISL_444633, EPI_ISL_444634, EPI_ISL_444635, EPI_ISL_444636, EPI_ISL_444637, EPI_ISL_444638, EPI_ISL_444639, EPI_ISL_444640, EPI_ISL_444641, EPI_ISL_444642, EPI_ISL_444643, EPI_ISL_444644, EPI_ISL_444645, EPI_ISL_444646, EPI_ISL_444647, EPI_ISL_444648, EPI_ISL_444649, EPI_ISL_444650, EPI_ISL_444651, EPI_ISL_444652, EPI_ISL_444653, EPI_ISL_444654, EPI_ISL_444656, EPI_ISL_444657, EPI_ISL_444658, EPI_ISL_444659, EPI_ISL_444660, EPI_ISL_444661, EPI_ISL_444662, EPI_ISL_444663, EPI_ISL_444664, EPI_ISL_444665, EPI_ISL_444666, EPI_ISL_444667, EPI_ISL_444668, EPI_ISL_444669, EPI_ISL_444671, EPI_ISL_444672, EPI_ISL_444673, EPI_ISL_444674, EPI_ISL_444675, EPI_ISL_444676, EPI_ISL_444677, EPI_ISL_444678, EPI_ISL_444679, EPI_ISL_444680, EPI_ISL_444681, EPI_ISL_444682, EPI_ISL_444683, EPI_ISL_444684, EPI_ISL_444685, EPI_ISL_444686, EPI_ISL_444687, EPI_ISL_444688, EPI_ISL_444689, EPI_ISL_444690, EPI_ISL_444691, EPI_ISL_444692, EPI_ISL_444693, EPI_ISL_444694, EPI_ISL_444695, EPI_ISL_444696, EPI_ISL_444697, EPI_ISL_444698, EPI_ISL_444699, EPI_ISL_444700, EPI_ISL_444701, EPI_ISL_444702, EPI_ISL_444703, EPI_ISL_444704, EPI_ISL_444705, EPI_ISL_444706, EPI_ISL_444707, EPI_ISL_444708, EPI_ISL_444709, EPI_ISL_444710, EPI_ISL_444711, EPI_ISL_444712, EPI_ISL_444713, EPI_ISL_444714, EPI_ISL_444715, EPI_ISL_444716, EPI_ISL_444717, EPI_ISL_444718, EPI_ISL_444719, EPI_ISL_444720, EPI_ISL_444721, EPI_ISL_444722, EPI_ISL_444723, EPI_ISL_444724, EPI_ISL_444725, EPI_ISL_444726, EPI_ISL_444727, EPI_ISL_444728, EPI_ISL_444729, EPI_ISL_444730, EPI_ISL_444731, EPI_ISL_444732, EPI_ISL_444733, EPI_ISL_444734, EPI_ISL_444735, EPI_ISL_444736, EPI_ISL_444737, EPI_ISL_444738, EPI_ISL_444739, EPI_ISL_444740, EPI_ISL_444741, EPI_ISL_444743, EPI_ISL_444745, EPI_ISL_444746, EPI_ISL_444747, EPI_ISL_444748, EPI_ISL_444749, EPI_ISL_444750, EPI_ISL_444751, EPI_ISL_444752, EPI_ISL_444753, EPI_ISL_444754, EPI_ISL_444756, EPI_ISL_444757, EPI_ISL_444758, EPI_ISL_444759, EPI_ISL_444760, EPI_ISL_444761, EPI_ISL_444762, EPI_ISL_444763, EPI_ISL_444764, EPI_ISL_444765, EPI_ISL_444766, EPI_ISL_444767, EPI_ISL_444768, EPI_ISL_444769, EPI_ISL_444770, EPI_ISL_444771, EPI_ISL_444772, EPI_ISL_444773, EPI_ISL_444774, EPI_ISL_444775, EPI_ISL_444776, EPI_ISL_444777, EPI_ISL_444778, EPI_ISL_444779, EPI_ISL_444781, EPI_ISL_444782, EPI_ISL_444783, EPI_ISL_444784, EPI_ISL_444785, EPI_ISL_444786, EPI_ISL_444787, EPI_ISL_444788, EPI_ISL_444789, EPI_ISL_444790, EPI_ISL_444791, EPI_ISL_444792 | | | |
| see above | NYU Langone Health | Departments of Pathology and Medicine, New York University School of Medicine | Maria Aguero-Rosenfeld, Brendan Belovarac, Margaret Black, Ludovic Boytard, John Cadley, Paolo Cotzia, John Chen, Dacia Dimartino, Xiaojun Feng, Tatyana Gindin, Emily Guzman, Adriana Heguy, Megan Hogan, Emily Huang, George Jour, Alireza Khodadadi-Jamayran, Lawrence H. Lin, Raven Luther, Andrew Lytle, Christian Marier, Matthew T. Maurano, Mark J. Mulligan, Peter Meyn, Raquel Ordonez Ciriza, Iman Osman, Jared Pinnell, Vanessa Raabe, Sitharam Ramaswami, Amy Rapkiewicz, Andre M. Ribeiro-dos-Santos, Marie Samanovic-Golden, Antonio Serrano, Guomiao Shen, Matija Snuderl, Theodore Vougiouklakis, Nick Vulpescu, Gael Westby, Paul Zappile, Yutong Zhang |
| EPI_ISL_450004, EPI_ISL_450005, EPI_ISL_450006, EPI_ISL_450007, EPI_ISL_450008, EPI_ISL_450009, EPI_ISL_450010, EPI_ISL_450011, EPI_ISL_450012, EPI_ISL_450013, EPI_ISL_450014, EPI_ISL_450015, EPI_ISL_450016, EPI_ISL_450017, EPI_ISL_450018, EPI_ISL_450019, EPI_ISL_450020, EPI_ISL_450021, EPI_ISL_450022, EPI_ISL_450023, EPI_ISL_450024, EPI_ISL_450025, EPI_ISL_450026, EPI_ISL_450027, EPI_ISL_450028, EPI_ISL_450029, EPI_ISL_450030, EPI_ISL_450031, EPI_ISL_450032, EPI_ISL_450033, EPI_ISL_450034, EPI_ISL_450035, EPI_ISL_450036, EPI_ISL_450037, EPI_ISL_450038, EPI_ISL_450039, EPI_ISL_450040, EPI_ISL_450041, EPI_ISL_450042, EPI_ISL_450043, EPI_ISL_450044, EPI_ISL_450045, EPI_ISL_450046, EPI_ISL_450047, EPI_ISL_450048, EPI_ISL_450050, EPI_ISL_450051, EPI_ISL_450052, EPI_ISL_450053, EPI_ISL_450054, EPI_ISL_450055, EPI_ISL_450056, EPI_ISL_450057, EPI_ISL_450058, EPI_ISL_450059, EPI_ISL_450060, EPI_ISL_450061, EPI_ISL_450062, EPI_ISL_450063, EPI_ISL_450064, EPI_ISL_450065, EPI_ISL_450066, EPI_ISL_450067, EPI_ISL_450068, EPI_ISL_450069, EPI_ISL_450070, EPI_ISL_450071, EPI_ISL_450072, EPI_ISL_450073, EPI_ISL_450074, EPI_ISL_450075, EPI_ISL_450076, EPI_ISL_450077, EPI_ISL_450078, EPI_ISL_450079, EPI_ISL_450080, EPI_ISL_450081, EPI_ISL_450082, EPI_ISL_450083, EPI_ISL_450084, EPI_ISL_450085, EPI_ISL_450086, EPI_ISL_450087, EPI_ISL_450088, EPI_ISL_450089, EPI_ISL_450090, EPI_ISL_450091, EPI_ISL_450092, EPI_ISL_450093, EPI_ISL_450094, EPI_ISL_450095, EPI_ISL_450096, EPI_ISL_450097, EPI_ISL_450098, EPI_ISL_450099, EPI_ISL_450100, EPI_ISL_450101, EPI_ISL_450102, EPI_ISL_450103, EPI_ISL_450104, EPI_ISL_450105, EPI_ISL_450106, EPI_ISL_450107, EPI_ISL_450108, EPI_ISL_450109, EPI_ISL_450110, EPI_ISL_450111, EPI_ISL_450112, EPI_ISL_450113, EPI_ISL_450114, EPI_ISL_450115, EPI_ISL_450116, EPI_ISL_450117, EPI_ISL_450118, EPI_ISL_450119, EPI_ISL_450120, EPI_ISL_450121, EPI_ISL_450122, EPI_ISL_450123, EPI_ISL_450124, EPI_ISL_450125, EPI_ISL_450126, EPI_ISL_450127, EPI_ISL_450128, EPI_ISL_450129, EPI_ISL_450130, EPI_ISL_450131, EPI_ISL_450132, EPI_ISL_450133, EPI_ISL_450134, EPI_ISL_450135, EPI_ISL_450136, EPI_ISL_450137, EPI_ISL_450139, EPI_ISL_450140, EPI_ISL_450141, EPI_ISL_450142, EPI_ISL_450143, EPI_ISL_450144, EPI_ISL_450146, EPI_ISL_450147, EPI_ISL_450148, EPI_ISL_450149, EPI_ISL_450150, EPI_ISL_450151, EPI_ISL_450152, EPI_ISL_450153, EPI_ISL_450154, EPI_ISL_450155, EPI_ISL_450156, EPI_ISL_450157, EPI_ISL_450158, EPI_ISL_450159, EPI_ISL_450160, EPI_ISL_450161, EPI_ISL_450162, EPI_ISL_450163, EPI_ISL_450164, EPI_ISL_450165, EPI_ISL_450166, EPI_ISL_450167, EPI_ISL_450168, EPI_ISL_450169, EPI_ISL_450170 | | | |
| see above | MSHS Clinical Microbiology Laboratories | MSHS Pathogen Surveillance Program | Ana S. Gonzalez-Reiche, Mitchell Sullivan, Ajay Obla, Gopi Patel, Emilia Sordillo, Melissa Gitman, Alberto Paniz-mondolfi, Matthew Hernandez, Shelcie Fabre, Jose Polanco, Zenab Khan, Bremy Alburquerque, Jayeeta Dutta, Juan Soto, Shwetha Sridhar Hara, Ying-Chih Wang, Melissa Smith, Robert Sebra, Lisa Miorin, Wen-chun Liu, Randy Albrecht, Judith Aberg, Florian Krammer, Adolfo Garcia-Sastre, Viviana Simon, Harm van Bakel |
| EPI_ISL_450393, EPI_ISL_450394, EPI_ISL_450395, EPI_ISL_450396, EPI_ISL_450397, EPI_ISL_450398, EPI_ISL_450400, EPI_ISL_450401, EPI_ISL_450402, EPI_ISL_451401, EPI_ISL_451402, EPI_ISL_451403, EPI_ISL_451404, EPI_ISL_451405, EPI_ISL_451406, EPI_ISL_451407, EPI_ISL_451408, EPI_ISL_451409, EPI_ISL_451410, EPI_ISL_451411, EPI_ISL_451412, EPI_ISL_451413, EPI_ISL_451414, EPI_ISL_451415, EPI_ISL_451416, EPI_ISL_451417, EPI_ISL_451418, EPI_ISL_451419, EPI_ISL_451420, EPI_ISL_451421, EPI_ISL_451422, EPI_ISL_451423, EPI_ISL_451424, EPI_ISL_451425, EPI_ISL_451426, EPI_ISL_451427, EPI_ISL_451428, EPI_ISL_451429, EPI_ISL_451430, EPI_ISL_451431, EPI_ISL_451432, EPI_ISL_451433, EPI_ISL_451434, EPI_ISL_451435, EPI_ISL_451436, EPI_ISL_451437, EPI_ISL_451438, EPI_ISL_451439, EPI_ISL_451440, EPI_ISL_451441, EPI_ISL_451442, EPI_ISL_451443, EPI_ISL_451445, EPI_ISL_451446, EPI_ISL_451447, EPI_ISL_451448, EPI_ISL_451449, EPI_ISL_451450, EPI_ISL_451451, EPI_ISL_451452, EPI_ISL_451453, EPI_ISL_451454, EPI_ISL_451455, EPI_ISL_451456, EPI_ISL_451457, EPI_ISL_451458, EPI_ISL_451459, EPI_ISL_451460, EPI_ISL_451461, EPI_ISL_451462, EPI_ISL_451463, EPI_ISL_451464, EPI_ISL_451465, EPI_ISL_451466, EPI_ISL_451467, EPI_ISL_451468, EPI_ISL_451469, EPI_ISL_451470, EPI_ISL_451471, EPI_ISL_451472, EPI_ISL_451473, EPI_ISL_451475, EPI_ISL_451476, EPI_ISL_451477, EPI_ISL_451478, EPI_ISL_451479, EPI_ISL_451665 | | | |
| see above | NYU Langone Health | Departments of Pathology and Medicine, New York University School of Medicine | Maria Aguero-Rosenfeld, Brendan Belovarac, Margaret Black, Ludovic Boytard, John Cadley, Paolo Cotzia, John Chen, Dacia Dimartino, Xiaojun Feng, Tatyana Gindin, Emily Guzman, Adriana Heguy, Megan Hogan, Emily Huang, George Jour, Alireza Khodadadi-Jamayran, Lawrence H. Lin, Raven Luther, Andrew Lytle, Christian Marier, Matthew T. Maurano, Mark J. Mulligan, Peter Meyn, Raquel Ordonez Ciriza, Iman Osman, Jared Pinnell, Vanessa Raabe, Sitharam Ramaswami, Amy Rapkiewicz, Andre M. Ribeiro-dos-Santos, Marie Samanovic-Golden, Antonio Serrano, Guomiao Shen, Matija Snuderl, Theodore Vougiouklakis, Nick Vulpescu, Gael Westby, Paul Zappile, Yutong Zhang |
| EPI_ISL_454691 | Quest Diagnostics | Quest Diagnostics | Anderson,B.P., Rosenthal,S.H., Gerasimova,A., Kagan,R.M. and Owen, R. |
| EPI_ISL_456003, EPI_ISL_456004, EPI_ISL_456005, EPI_ISL_456006, EPI_ISL_456007, EPI_ISL_456008, EPI_ISL_456009, EPI_ISL_456010, EPI_ISL_456011, EPI_ISL_456012, EPI_ISL_456013, EPI_ISL_456014, EPI_ISL_456015, EPI_ISL_456016, EPI_ISL_456017, EPI_ISL_456018, EPI_ISL_456019, EPI_ISL_456020, EPI_ISL_456021, EPI_ISL_456022, EPI_ISL_456023, EPI_ISL_456024, EPI_ISL_456025, EPI_ISL_456026, EPI_ISL_456027, EPI_ISL_456028, EPI_ISL_456029, EPI_ISL_456030, EPI_ISL_456031, EPI_ISL_456032, EPI_ISL_456033, EPI_ISL_456034, EPI_ISL_456035, EPI_ISL_456036, EPI_ISL_456037, EPI_ISL_456038, EPI_ISL_456039, EPI_ISL_456040, EPI_ISL_456041, EPI_ISL_456042, EPI_ISL_456043, EPI_ISL_456044, EPI_ISL_456045, EPI_ISL_456046, EPI_ISL_456047, EPI_ISL_456048, EPI_ISL_456049, EPI_ISL_456050, EPI_ISL_456052, EPI_ISL_456053, EPI_ISL_456054, EPI_ISL_456055, EPI_ISL_456056, EPI_ISL_456057, EPI_ISL_456058, EPI_ISL_456059, EPI_ISL_456060, EPI_ISL_456061, EPI_ISL_456062, EPI_ISL_456063, EPI_ISL_456064, EPI_ISL_456065, EPI_ISL_456066, EPI_ISL_456067, EPI_ISL_456069, EPI_ISL_456070, EPI_ISL_456107, EPI_ISL_456108, EPI_ISL_456109, EPI_ISL_456110, EPI_ISL_456111, EPI_ISL_456112, EPI_ISL_456114, EPI_ISL_456115, EPI_ISL_458017, EPI_ISL_458018, EPI_ISL_458019, EPI_ISL_458020, EPI_ISL_458021, EPI_ISL_458022, EPI_ISL_460603, EPI_ISL_467377, EPI_ISL_467378, EPI_ISL_467379, EPI_ISL_467380, EPI_ISL_467381, EPI_ISL_467382, EPI_ISL_467383, EPI_ISL_467384, EPI_ISL_467385, EPI_ISL_467386, EPI_ISL_467387, EPI_ISL_467388, EPI_ISL_467389, EPI_ISL_467390, EPI_ISL_467391, EPI_ISL_467392, EPI_ISL_467393, EPI_ISL_467394, EPI_ISL_467395, EPI_ISL_467396, EPI_ISL_467397, EPI_ISL_467398, EPI_ISL_467399, EPI_ISL_467400, EPI_ISL_467401, EPI_ISL_467402, EPI_ISL_467403, EPI_ISL_467404, EPI_ISL_467405, EPI_ISL_467406, EPI_ISL_467407, EPI_ISL_467408, EPI_ISL_467409, EPI_ISL_467410, EPI_ISL_467411, EPI_ISL_467412, EPI_ISL_467413, EPI_ISL_467414, EPI_ISL_467415, EPI_ISL_467416, EPI_ISL_467417, EPI_ISL_467418, EPI_ISL_467419, EPI_ISL_467420, EPI_ISL_467421, EPI_ISL_467422 | | | |
| see above | NYU Langone Health | Departments of Pathology and Medicine, New York University School of Medicine | Maria Aguero-Rosenfeld, Brendan Belovarac, Margaret Black, Ludovic Boytard, John Cadley, Paolo Cotzia, John Chen, Dacia Dimartino, Xiaojun Feng, Tatyana Gindin, Emily Guzman, Adriana Heguy, Megan Hogan, Emily Huang, George Jour, Alireza Khodadadi-Jamayran, Lawrence H. Lin, Raven Luther, Andrew Lytle, Christian Marier, Matthew T. Maurano, Mark J. Mulligan, Peter Meyn, Raquel Ordonez Ciriza, Iman Osman, Jared Pinnell, Vanessa Raabe, Sitharam Ramaswami, Amy Rapkiewicz, Andre M. Ribeiro-dos-Santos, Marie Samanovic-Golden, Antonio Serrano, Guomiao Shen, Matija Snuderl, Theodore Vougiouklakis, Nick Vulpescu, Gael Westby, Paul Zappile, Yutong Zhang |
| EPI_ISL_467884, EPI_ISL_467885, EPI_ISL_467886, EPI_ISL_467887, EPI_ISL_467888, EPI_ISL_467889, EPI_ISL_467890, EPI_ISL_467891, EPI_ISL_467892, EPI_ISL_467893, EPI_ISL_467894, EPI_ISL_467895, EPI_ISL_467896, EPI_ISL_467897, EPI_ISL_467898, EPI_ISL_467899, EPI_ISL_467900, EPI_ISL_467901, EPI_ISL_467902, EPI_ISL_467903 | | | |
| see above | Quest Diagnostics | Quest Diagnostics | Anderson,B.P., Rosenthal,S.H., Gerasimova,A., Kagan,R.M. and Owen, R. |
| EPI_ISL_467945 | Montefiore Medical Center, Dept. of Pathology, Clinical Virology | Albert Einstein College of Medicine, Dept. of Microbiology & Immunology, Chandran lab | J. Maximilian Fels, Saad Khan, Robert Forster, Karin A. Skalina, Ariel S. Wirchnianski, Denise Haslwanter, Catalina Florez, Robert H. Bortz III, M. Eugenia Dieterle, Ethan Laudermilch, Rohit K. Jangra, Amanda Mengotto, Duncan Kimmel, Shahina B. Maqbool, John M. Greally, Wendy A. Szymczak, Amy S. Fox, Michael B. Prystowsky, D. Yitzchak Goldstein, Johanna P. Daily, Libusha Kelly, Kartik Chandran |
| EPI_ISL_468576, EPI_ISL_468577, EPI_ISL_468578, EPI_ISL_468579, EPI_ISL_468580, EPI_ISL_468581, EPI_ISL_468582, EPI_ISL_468583, EPI_ISL_468584, EPI_ISL_468585, EPI_ISL_468586, EPI_ISL_468587, EPI_ISL_468588 | | | |
| see above | Quest Diagnostics | Quest Diagnostics | Anderson,B.P., Rosenthal,S.H., Gerasimova,A., Kagan,R.M. and Owen, R. |
| EPI_ISL_476524 | Yale Clinical Virology Laboratory | Grubaugh Lab - Yale School of Public Health | Joseph Fauver, Tara Alpert, Anderson Brito, Anne Wyllie, Chantal Vogels, Mary Petrone, Cole Jensen, Chaney Kalinich, Isabel Ott, Arnau Casanovas, Catherine Muenker, Adam Moore, Alice Lu, Maria Tokuyama, Patrick Wong, Peiwen Lu, Saad Omer, Richard Martinello, Allison Nelson, Shelli Farhadian, |

| | | | |
|---|---|---|---|
| | | | Akiko Iwasaki, Charlese Dela Cruz, Albert Ko, Nathan Grubaugh |
| EPI_ISL_479663, EPI_ISL_479664, EPI_ISL_479665, EPI_ISL_479666, EPI_ISL_479667, EPI_ISL_479668, EPI_ISL_479669, EPI_ISL_479670, EPI_ISL_479671, EPI_ISL_479672, EPI_ISL_479673, EPI_ISL_479674, EPI_ISL_479675 | | | |
| see above | Center for Genomics and System Biology, New York University | Center for Genomics and System Biology, New York University | Roder,A., Banakis,S., Johnson,K., Khalfan,M., Borenstein,E.S., Samanovic,M., Cornelius,A., Herati,R., Ulrich,R., Fleming,A., Kottkamp,A., Raabe,V., Mulligan,M.J., Gresham,D., Ghedin,E. |
| EPI_ISL_480784, EPI_ISL_480785 | NYC Department of Health and Mental Hygiene | Pathogen Discovery, Respiratory Viruses Branch, Division of Viral Diseases, Centers for Disease Control and Prevention | Krista Queen, Christine Mahl, Jennifer Rakeman, Anna Uehara, Ying Tao, Jing Zhang, Yan Li, Clinton R. Paden, Haibin Wang, Jasmine Padilla, Justin Lee, Sally Slavinski, Suxiang Tong |
| EPI_ISL_481283 | Center for Genomics and System Biology, New York University | Center for Genomics and System Biology, New York University | Roder,A., Banakis,S., Johnson,K., Khalfan,M., Borenstein,E.S., Samanovic,M., Cornelius,A., Herati,R., Ulrich,R., Fleming,A., Kottkamp,A., Raabe,V., Mulligan,M.J., Gresham,D. and Ghedin,E. |
| EPI_ISL_484959, EPI_ISL_484972, EPI_ISL_491320, EPI_ISL_491321, EPI_ISL_495497 | University of Wisconsin-Madison AIDS Vaccine Research Laboratories | University of Wisconsin-Madison AIDS Vaccine Research Laboratories | Gage Moreno, Katarina Braun, et al. AIDS Vaccine Research Laboratories |
| EPI_ISL_500503, EPI_ISL_500504, EPI_ISL_500505, EPI_ISL_500506, EPI_ISL_500507, EPI_ISL_500525, EPI_ISL_501082 | Mayo Clinic Laboratories | University of Washington Virology Lab | Pavitra Roychoudhury, Hong Xie, Lasata Shrestha, Amin Addetia, Truong Nguyen, Victoria M Rachleff, Meei-Li Huang, Keith R Jerome, Alexander Greninger |
| EPI_ISL_525575, EPI_ISL_525576, EPI_ISL_525577, EPI_ISL_525578, EPI_ISL_525579, EPI_ISL_525580, EPI_ISL_525581, EPI_ISL_525582, EPI_ISL_525583, EPI_ISL_525584, EPI_ISL_525585, EPI_ISL_525586, EPI_ISL_525587, EPI_ISL_525588, EPI_ISL_525589, EPI_ISL_525590, EPI_ISL_525591, EPI_ISL_525592, EPI_ISL_525593, EPI_ISL_525594, EPI_ISL_525595, EPI_ISL_525596, EPI_ISL_525597, EPI_ISL_525598, EPI_ISL_525599, EPI_ISL_525600, EPI_ISL_525601, EPI_ISL_525602, EPI_ISL_525603, EPI_ISL_525604, EPI_ISL_525605, EPI_ISL_525606, EPI_ISL_525607, EPI_ISL_525608, EPI_ISL_525609, EPI_ISL_525610, EPI_ISL_525611, EPI_ISL_525612, EPI_ISL_525613, EPI_ISL_525614, EPI_ISL_525615, EPI_ISL_525616, EPI_ISL_525617, EPI_ISL_525618, EPI_ISL_525619, EPI_ISL_525620, EPI_ISL_525621, EPI_ISL_525622, EPI_ISL_525623, EPI_ISL_525624, EPI_ISL_525625, EPI_ISL_525626, EPI_ISL_525627, EPI_ISL_525628, EPI_ISL_525629, EPI_ISL_525630, EPI_ISL_525631, EPI_ISL_525632, EPI_ISL_525633, EPI_ISL_525634, EPI_ISL_525635, EPI_ISL_525636, EPI_ISL_525637, EPI_ISL_525638, EPI_ISL_525639, EPI_ISL_525640, EPI_ISL_525641, EPI_ISL_525642, EPI_ISL_525643, EPI_ISL_525644, EPI_ISL_525645, EPI_ISL_525646, EPI_ISL_525647, EPI_ISL_525648, EPI_ISL_525649, EPI_ISL_525650, EPI_ISL_525651, EPI_ISL_525652, EPI_ISL_525653, EPI_ISL_525654, EPI_ISL_525655, EPI_ISL_525656, EPI_ISL_525657, EPI_ISL_525658, EPI_ISL_525659, EPI_ISL_525660, EPI_ISL_525661, EPI_ISL_525662, EPI_ISL_525663, EPI_ISL_525664, EPI_ISL_525665, EPI_ISL_525666, EPI_ISL_525667, EPI_ISL_525668, EPI_ISL_525669, EPI_ISL_525670, EPI_ISL_525671, EPI_ISL_525672, EPI_ISL_525673, EPI_ISL_525674, EPI_ISL_525675, EPI_ISL_525676, EPI_ISL_525677, EPI_ISL_525678, EPI_ISL_525679, EPI_ISL_525680, EPI_ISL_525681, EPI_ISL_525682, EPI_ISL_525683, EPI_ISL_525684, EPI_ISL_525685, EPI_ISL_525686 | | | |
| see above | Wadsworth Center, New York State Department of Health | Wadsworth Center, New York State Department of Health | Kirsten St. George, Daryl M. Lamson, Sara Griesemer, Jonathan Plitnick, Navjot Singh, Matthew D. Shudt, Erica Lasek-Nesselquist |
| EPI_ISL_526686, EPI_ISL_526687, EPI_ISL_526688 | Faith Laboratory, Immunology Institute, Icahn School of Medicine at Mount Sinai | van Bakel Laboratory, Genetics and Genomics Sciences, Icahn School of Medicine at Mount Sinai | Graham J. Britton, Alice Chen-Liaw, Francesca Cossarini, Alexandra Livanos, Matthew P. Spindler, Tamar Plitt, Joseph Eggers, Ilaria Mogno, Ana S. Gonzalez-Reiche, Sophia Sui, Michael Tankelevich, Lauren Tal Grinspan, Rebekah E. Dixon, Divya Jha, Gustavo Martinez-Delgado, Fatima Amanat, Daisy Hoagland, Benjamin R. tenOever, Marla C. Dubinsky, Miriam Merad, Harm Van Bakel, Florian Krammer, Gerold Bongers, Saurabh Mehandru and Jeremiah J. Faith |
| EPI_ISL_529033, EPI_ISL_529034, EPI_ISL_529035, EPI_ISL_529036, EPI_ISL_534731 | Wadsworth Center, New York State Department of Health | Wadsworth Center, New York State Department of Health | Kirsten St. George, Daryl M. Lamson, Sara Griesemer, Jonathan Plitnick, Navjot Singh, Matthew D. Shudt, Erica Lasek-Nesselquist |
| EPI_ISL_536661, EPI_ISL_536702, EPI_ISL_536714, EPI_ISL_536722, EPI_ISL_536752, EPI_ISL_536760, EPI_ISL_536773 | University of Wisconsin-Madison AIDS Vaccine Research Laboratories | University of Wisconsin-Madison AIDS Vaccine Research Laboratories | Gage Moreno, Katarina Braun, et al. AIDS Vaccine Research Laboratories |
| EPI_ISL_548243, EPI_ISL_548244 | Faith Laboratory, Immunology Institute, Icahn School of Medicine at Mount Sinai | van Bakel Laboratory, Genetics and Genomics Sciences, Icahn School of Medicine at Mount Sinai | Graham J. Britton, Alice Chen-Liaw, Francesca Cossarini, Alexandra Livanos, Matthew P. Spindler, Tamar Plitt, Joseph Eggers, Ilaria Mogno, Ana S. Gonzalez-Reiche, Sophia Sui, Michael Tankelevich, Lauren Tal Grinspan, Rebekah E. Dixon, Divya Jha, Gustavo Martinez-Delgado, Fatima Amanat, Daisy Hoagland, Benjamin R. tenOever, Marla C. Dubinsky, Miriam Merad, Harm Van Bakel, Florian Krammer, Gerold Bongers, Saurabh Mehandru and Jeremiah J. Faith |
| EPI_ISL_566036, EPI_ISL_566037, EPI_ISL_566038, EPI_ISL_566039, EPI_ISL_566040, EPI_ISL_566041, EPI_ISL_566042, EPI_ISL_566043 | Wildlife Conservation Society, Bronx Zoo | Diagnostic Virology Laboratory, United States Department of Agriculture, National Veterinary Services Laboratories, Population Medicine and Diagnostic Sciences, Cornell University | McAloose,D., Laverack,M., Wang,L., Killian,M.L., Caserta,L.C., Yuan,F., Mitchell,P.K., Queen,K., Mauldin,M.R., Cronk,B.D., Bartlett,S.L., Sykes,J.M., Zec,S., Stokol,T., Ingerman,K., Delaney,M.A., Frederikson,R., Ivancic,M., Jenkins-Moore,M., Mozingo,K., Farnzen,K., Kuzmin,I., Hines Bergeson,N., Goodman,L., Wang,H., Tong,S., Fang,Y., Olmstead,C., McCann,C., Thomas,P., Goodrich,E., Elvinger,F., Smith,D.C., Slavinski,S., Calle,P.P., Terio,K., Torchetti,M.K., Diel,D.G., Cardia Caserta,L. |
| EPI_ISL_566044 | Wildlife Conservation Society, Bronx Zoo | Diagnostic Virology Laboratory, United States Department of Agriculture, National Veterinary Services Laboratories, Population Medicine and Diagnostic Sciences, Cornell University | McAloose,D., Laverack,M., Wang,L., Killian,M.L., Caserta,L.C., Yuan,F., Mitchell,P.K., Queen,K., Mauldin,M.R., Cronk,B.D., Bartlett,S.L., Sykes,J.M., Zec,S., Stokol,T., Ingerman,K., Delaney,M.A., Frederikson,R., Ivancic,M., Jenkins-Moore,M., Mozingo,K., Farnzen,K., Kuzmin,I., Hines Bergeson,N., Goodman,L., Wang,H., Tong,S., Fang,Y., Olmstead,C., McCann,C., Thomas,P., Goodrich,E., Elvinger,F., Smith,D.C., Slavinski,S., Calle,P.P., Terio,K., Torchetti,M.K., Diel,D.G., Cardia Caserta,L., Mauldin,M. |
| EPI_ISL_571550, EPI_ISL_571647, EPI_ISL_571648, EPI_ISL_571649, EPI_ISL_571650, EPI_ISL_571651, EPI_ISL_571652, EPI_ISL_571653, EPI_ISL_571654, EPI_ISL_571655, EPI_ISL_571656, EPI_ISL_571657, EPI_ISL_571658, EPI_ISL_571659, EPI_ISL_571660, EPI_ISL_571661, EPI_ISL_571662, EPI_ISL_571663, EPI_ISL_571664, EPI_ISL_571665, EPI_ISL_571666, EPI_ISL_571667, EPI_ISL_571668, EPI_ISL_571669, EPI_ISL_571670, EPI_ISL_571671, EPI_ISL_571672, EPI_ISL_571673, EPI_ISL_571674, EPI_ISL_571675, EPI_ISL_571676, EPI_ISL_571677, EPI_ISL_571678, EPI_ISL_571679, EPI_ISL_571680, EPI_ISL_571681, EPI_ISL_571682, EPI_ISL_571683, EPI_ISL_571684, EPI_ISL_571799, EPI_ISL_571984, EPI_ISL_571985, EPI_ISL_571986, EPI_ISL_571987, EPI_ISL_571988, EPI_ISL_571989, EPI_ISL_571990, EPI_ISL_571991, EPI_ISL_571992, EPI_ISL_571993, EPI_ISL_571994, EPI_ISL_571995, EPI_ISL_571996, EPI_ISL_571997, EPI_ISL_571998, EPI_ISL_571999, EPI_ISL_572000, EPI_ISL_572001, EPI_ISL_572002, EPI_ISL_572003, EPI_ISL_572004, EPI_ISL_572005, EPI_ISL_572006, EPI_ISL_572007, EPI_ISL_572008, EPI_ISL_572009, EPI_ISL_572010, EPI_ISL_572011, EPI_ISL_572012, EPI_ISL_572013, EPI_ISL_572014, EPI_ISL_572015, EPI_ISL_572016, EPI_ISL_572017, EPI_ISL_572018, EPI_ISL_572019, EPI_ISL_572020, EPI_ISL_572021, EPI_ISL_572022, EPI_ISL_572023, EPI_ISL_572024, EPI_ISL_572025, EPI_ISL_572026, EPI_ISL_572027, EPI_ISL_572028, EPI_ISL_572029, EPI_ISL_572030, EPI_ISL_572031 | | | |
| see above | Quest Diagnostics | Quest Diagnostics | Rosenthal,S.H., Gerasimova,A., Kagan,R.M., Anderson, B., Grover, D., Livingston, K.E., Hua, M., Liu Y., Shalhout, D.F., Owen, R., Lacbawan, F. |
| EPI_ISL_583426 | Garcia-Sastre Laboratory, Department of Microbiology, Icahn School of Medicine at Mount Sinai | van Bakel Laboratory, Genetics and Genomics Sciences, Icahn School of Medicine at Mount Sinai | Teresa Aydillo, Ana S. Gonzalez-Reiche, Sadaf Aslam, Adriana van de Guchte, Zenab Khan, Ajay Obla, Jayeeta Dutta, Harm van Bakel, Judith Aberg, Adolfo García-Sastre, Gunjan Shah, Tobias Hohl, Genovefa Papanicolaou, Miguel-Angel Perales, Kent Sepkowitz, Ngolela Esther Babady, and Mini Kamboj |
| EPI_ISL_583427, EPI_ISL_583428 | Memorial Sloan Kettering Cancer Center | van Bakel Laboratory, Genetics and Genomics Sciences, Icahn School of Medicine at Mount Sinai | Teresa Aydillo, Ana S. Gonzalez-Reiche, Sadaf Aslam, Adriana van de Guchte, Zenab Khan, Ajay Obla, Jayeeta Dutta, Harm van Bakel, Judith Aberg, Adolfo García-Sastre, Gunjan Shah, Tobias Hohl, Genovefa Papanicolaou, Miguel-Angel Perales, Kent Sepkowitz, Ngolela Esther Babady, and Mini Kamboj |
| EPI_ISL_583429 | Garcia-Sastre Laboratory, Department of Microbiology, Icahn School of Medicine at Mount Sinai | van Bakel Laboratory, Genetics and Genomics Sciences, Icahn School of Medicine at Mount Sinai | Teresa Aydillo, Ana S. Gonzalez-Reiche, Sadaf Aslam, Adriana van de Guchte, Zenab Khan, Ajay Obla, Jayeeta Dutta, Harm van Bakel, Judith Aberg, Adolfo García-Sastre, Gunjan Shah, Tobias Hohl, Genovefa Papanicolaou, Miguel-Angel Perales, Kent Sepkowitz, Ngolela Esther Babady, and Mini Kamboj |
| EPI_ISL_583430, EPI_ISL_583431, EPI_ISL_583432, EPI_ISL_583433 | Memorial Sloan Kettering Cancer Center | van Bakel Laboratory, Genetics and Genomics Sciences, Icahn School of Medicine at Mount Sinai | Teresa Aydillo, Ana S. Gonzalez-Reiche, Sadaf Aslam, Adriana van de Guchte, Zenab Khan, Ajay Obla, Jayeeta Dutta, Harm van Bakel, Judith Aberg, Adolfo García-Sastre, Gunjan Shah, Tobias Hohl, Genovefa Papanicolaou, Miguel-Angel Perales, Kent Sepkowitz, Ngolela Esther Babady, and Mini Kamboj |
| EPI_ISL_583434, EPI_ISL_583435, EPI_ISL_583436 | Garcia-Sastre Laboratory, Department of Microbiology, Icahn School of Medicine at Mount Sinai | van Bakel Laboratory, Genetics and Genomics Sciences, Icahn School of Medicine at Mount Sinai | Teresa Aydillo, Ana S. Gonzalez-Reiche, Sadaf Aslam, Adriana van de Guchte, Zenab Khan, Ajay Obla, Jayeeta Dutta, Harm van Bakel, Judith Aberg, Adolfo García-Sastre, Gunjan Shah, Tobias Hohl, Genovefa Papanicolaou, Miguel-Angel Perales, Kent Sepkowitz, Ngolela Esther Babady, and Mini Kamboj |
| EPI_ISL_583437, EPI_ISL_583438, EPI_ISL_583439, EPI_ISL_583440 | Memorial Sloan Kettering Cancer Center | van Bakel Laboratory, Genetics and Genomics Sciences, Icahn School of Medicine at Mount Sinai | Teresa Aydillo, Ana S. Gonzalez-Reiche, Sadaf Aslam, Adriana van de Guchte, Zenab Khan, Ajay Obla, Jayeeta Dutta, Harm van Bakel, Judith Aberg, Adolfo García-Sastre, Gunjan Shah, Tobias Hohl, Genovefa Papanicolaou, Miguel-Angel Perales, Kent Sepkowitz, Ngolela Esther Babady, and Mini Kamboj |
| EPI_ISL_583441, EPI_ISL_583442 | Garcia-Sastre Laboratory, Department of Microbiology, Icahn School of Medicine at Mount Sinai | van Bakel Laboratory, Genetics and Genomics Sciences, Icahn School of Medicine at Mount Sinai | Teresa Aydillo, Ana S. Gonzalez-Reiche, Sadaf Aslam, Adriana van de Guchte, Zenab Khan, Ajay Obla, Jayeeta Dutta, Harm van Bakel, Judith Aberg, Adolfo García-Sastre, Gunjan Shah, Tobias Hohl, Genovefa Papanicolaou, Miguel-Angel Perales, Kent Sepkowitz, Ngolela Esther Babady, and Mini Kamboj |
| EPI_ISL_583443, EPI_ISL_583444, EPI_ISL_583445, EPI_ISL_583446, | Memorial Sloan Kettering Cancer Center | van Bakel Laboratory, Genetics and Genomics Sciences, Icahn School of Medicine at Mount Sinai | Teresa Aydillo, Ana S. Gonzalez-Reiche, Sadaf Aslam, Adriana van de Guchte, Zenab Khan, Ajay Obla, Jayeeta Dutta, Harm van Bakel, Judith Aberg, Adolfo García-Sastre, Gunjan Shah, Tobias Hohl, Genovefa Papanicolaou, Miguel-Angel Perales, Kent Sepkowitz, Ngolela Esther Babady, and Mini |

| | | | |
|---|---|---|---|
| EPI_ISL_583447, EPI_ISL_583448, EPI_ISL_583449, EPI_ISL_583450, EPI_ISL_583451 | | | Kamboj |
| EPI_ISL_583452, EPI_ISL_583453, EPI_ISL_583454 | Garcia-Sastre Laboratory, Department of Microbiology, Icahn School of Medicine at Mount Sinai | van Bakel Laboratory, Genetics and Genomics Sciences, Icahn School of Medicine at Mount Sinai | Teresa Aydillo, Ana S. Gonzalez-Reiche, Sadaf Aslam, Adriana van de Guchte, Zenab Khan, Ajay Obla, Jayeeta Dutta, Harm van Bakel, Judith Aberg, Adolfo García-Sastre, Gunjan Shah, Tobias Hohl, Genovefa Papanicolaou, Miguel-Angel Perales, Kent Sepkowitz, Ngolela Esther Babady, and Mini Kamboj |
| EPI_ISL_583455, EPI_ISL_583456, EPI_ISL_583457 | Memorial Sloan Kettering Cancer Center | van Bakel Laboratory, Genetics and Genomics Sciences, Icahn School of Medicine at Mount Sinai | Teresa Aydillo, Ana S. Gonzalez-Reiche, Sadaf Aslam, Adriana van de Guchte, Zenab Khan, Ajay Obla, Jayeeta Dutta, Harm van Bakel, Judith Aberg, Adolfo García-Sastre, Gunjan Shah, Tobias Hohl, Genovefa Papanicolaou, Miguel-Angel Perales, Kent Sepkowitz, Ngolela Esther Babady, and Mini Kamboj |
| EPI_ISL_583458, EPI_ISL_583459 | Garcia-Sastre Laboratory, Department of Microbiology, Icahn School of Medicine at Mount Sinai | van Bakel Laboratory, Genetics and Genomics Sciences, Icahn School of Medicine at Mount Sinai | Teresa Aydillo, Ana S. Gonzalez-Reiche, Sadaf Aslam, Adriana van de Guchte, Zenab Khan, Ajay Obla, Jayeeta Dutta, Harm van Bakel, Judith Aberg, Adolfo García-Sastre, Gunjan Shah, Tobias Hohl, Genovefa Papanicolaou, Miguel-Angel Perales, Kent Sepkowitz, Ngolela Esther Babady, and Mini Kamboj |
| EPI_ISL_583460, EPI_ISL_583461, EPI_ISL_583462 | Memorial Sloan Kettering Cancer Center | van Bakel Laboratory, Genetics and Genomics Sciences, Icahn School of Medicine at Mount Sinai | Teresa Aydillo, Ana S. Gonzalez-Reiche, Sadaf Aslam, Adriana van de Guchte, Zenab Khan, Ajay Obla, Jayeeta Dutta, Harm van Bakel, Judith Aberg, Adolfo García-Sastre, Gunjan Shah, Tobias Hohl, Genovefa Papanicolaou, Miguel-Angel Perales, Kent Sepkowitz, Ngolela Esther Babady, and Mini Kamboj |
| EPI_ISL_583463, EPI_ISL_583464 | Garcia-Sastre Laboratory, Department of Microbiology, Icahn School of Medicine at Mount Sinai | van Bakel Laboratory, Genetics and Genomics Sciences, Icahn School of Medicine at Mount Sinai | Teresa Aydillo, Ana S. Gonzalez-Reiche, Sadaf Aslam, Adriana van de Guchte, Zenab Khan, Ajay Obla, Jayeeta Dutta, Harm van Bakel, Judith Aberg, Adolfo García-Sastre, Gunjan Shah, Tobias Hohl, Genovefa Papanicolaou, Miguel-Angel Perales, Kent Sepkowitz, Ngolela Esther Babady, and Mini Kamboj |
| EPI_ISL_583465, EPI_ISL_583466 | Memorial Sloan Kettering Cancer Center | van Bakel Laboratory, Genetics and Genomics Sciences, Icahn School of Medicine at Mount Sinai | Teresa Aydillo, Ana S. Gonzalez-Reiche, Sadaf Aslam, Adriana van de Guchte, Zenab Khan, Ajay Obla, Jayeeta Dutta, Harm van Bakel, Judith Aberg, Adolfo García-Sastre, Gunjan Shah, Tobias Hohl, Genovefa Papanicolaou, Miguel-Angel Perales, Kent Sepkowitz, Ngolela Esther Babady, and Mini Kamboj |
| EPI_ISL_583467 | Garcia-Sastre Laboratory, Department of Microbiology, Icahn School of Medicine at Mount Sinai | van Bakel Laboratory, Genetics and Genomics Sciences, Icahn School of Medicine at Mount Sinai | Teresa Aydillo, Ana S. Gonzalez-Reiche, Sadaf Aslam, Adriana van de Guchte, Zenab Khan, Ajay Obla, Jayeeta Dutta, Harm van Bakel, Judith Aberg, Adolfo García-Sastre, Gunjan Shah, Tobias Hohl, Genovefa Papanicolaou, Miguel-Angel Perales, Kent Sepkowitz, Ngolela Esther Babady, and Mini Kamboj |
| EPI_ISL_583468, EPI_ISL_583469, EPI_ISL_583470 | Memorial Sloan Kettering Cancer Center | van Bakel Laboratory, Genetics and Genomics Sciences, Icahn School of Medicine at Mount Sinai | Teresa Aydillo, Ana S. Gonzalez-Reiche, Sadaf Aslam, Adriana van de Guchte, Zenab Khan, Ajay Obla, Jayeeta Dutta, Harm van Bakel, Judith Aberg, Adolfo García-Sastre, Gunjan Shah, Tobias Hohl, Genovefa Papanicolaou, Miguel-Angel Perales, Kent Sepkowitz, Ngolela Esther Babady, and Mini Kamboj |
| EPI_ISL_583471 | Garcia-Sastre Laboratory, Department of Microbiology, Icahn School of Medicine at Mount Sinai | van Bakel Laboratory, Genetics and Genomics Sciences, Icahn School of Medicine at Mount Sinai | Teresa Aydillo, Ana S. Gonzalez-Reiche, Sadaf Aslam, Adriana van de Guchte, Zenab Khan, Ajay Obla, Jayeeta Dutta, Harm van Bakel, Judith Aberg, Adolfo García-Sastre, Gunjan Shah, Tobias Hohl, Genovefa Papanicolaou, Miguel-Angel Perales, Kent Sepkowitz, Ngolela Esther Babady, and Mini Kamboj |
| EPI_ISL_583472 | Memorial Sloan Kettering Cancer Center | van Bakel Laboratory, Genetics and Genomics Sciences, Icahn School of Medicine at Mount Sinai | Teresa Aydillo, Ana S. Gonzalez-Reiche, Sadaf Aslam, Adriana van de Guchte, Zenab Khan, Ajay Obla, Jayeeta Dutta, Harm van Bakel, Judith Aberg, Adolfo García-Sastre, Gunjan Shah, Tobias Hohl, Genovefa Papanicolaou, Miguel-Angel Perales, Kent Sepkowitz, Ngolela Esther Babady, and Mini Kamboj |
| EPI_ISL_583473 | Garcia-Sastre Laboratory, Department of Microbiology, Icahn School of Medicine at Mount Sinai | van Bakel Laboratory, Genetics and Genomics Sciences, Icahn School of Medicine at Mount Sinai | Teresa Aydillo, Ana S. Gonzalez-Reiche, Sadaf Aslam, Adriana van de Guchte, Zenab Khan, Ajay Obla, Jayeeta Dutta, Harm van Bakel, Judith Aberg, Adolfo García-Sastre, Gunjan Shah, Tobias Hohl, Genovefa Papanicolaou, Miguel-Angel Perales, Kent Sepkowitz, Ngolela Esther Babady, and Mini Kamboj |
| EPI_ISL_583474, EPI_ISL_583475, EPI_ISL_583476 | Memorial Sloan Kettering Cancer Center | van Bakel Laboratory, Genetics and Genomics Sciences, Icahn School of Medicine at Mount Sinai | Teresa Aydillo, Ana S. Gonzalez-Reiche, Sadaf Aslam, Adriana van de Guchte, Zenab Khan, Ajay Obla, Jayeeta Dutta, Harm van Bakel, Judith Aberg, Adolfo García-Sastre, Gunjan Shah, Tobias Hohl, Genovefa Papanicolaou, Miguel-Angel Perales, Kent Sepkowitz, Ngolela Esther Babady, and Mini Kamboj |
| EPI_ISL_583477 | Garcia-Sastre Laboratory, Department of Microbiology, Icahn School of Medicine at Mount Sinai | van Bakel Laboratory, Genetics and Genomics Sciences, Icahn School of Medicine at Mount Sinai | Teresa Aydillo, Ana S. Gonzalez-Reiche, Sadaf Aslam, Adriana van de Guchte, Zenab Khan, Ajay Obla, Jayeeta Dutta, Harm van Bakel, Judith Aberg, Adolfo García-Sastre, Gunjan Shah, Tobias Hohl, Genovefa Papanicolaou, Miguel-Angel Perales, Kent Sepkowitz, Ngolela Esther Babady, and Mini Kamboj |
| EPI_ISL_583478, EPI_ISL_583479, EPI_ISL_583480 | Memorial Sloan Kettering Cancer Center | van Bakel Laboratory, Genetics and Genomics Sciences, Icahn School of Medicine at Mount Sinai | Teresa Aydillo, Ana S. Gonzalez-Reiche, Sadaf Aslam, Adriana van de Guchte, Zenab Khan, Ajay Obla, Jayeeta Dutta, Harm van Bakel, Judith Aberg, Adolfo García-Sastre, Gunjan Shah, Tobias Hohl, Genovefa Papanicolaou, Miguel-Angel Perales, Kent Sepkowitz, Ngolela Esther Babady, and Mini Kamboj |
| EPI_ISL_594457 | NYS Department of Health | Pathogen Discovery, Respiratory Viruses Branch, Division of Viral Diseases, Centers for Disease Control and Prevention | Ying Tao, Yan Li, Clinton Paden, Jing Zhang, Krista Queen, Anna Uehara, Haibin Wang, Julu Bhatnagar, Suxiang Tong |
| EPI_ISL_602562, EPI_ISL_602563 | Department of Microbiology and Immunology, Stony Brook University | Department of Microbiology and Immunology, Stony Brook University | Nascimento Conde,J., Luperchio,A.M., Gorbunova,E.E., Mackow,E.R. |
| EPI_ISL_603842, EPI_ISL_604235, EPI_ISL_604236, EPI_ISL_604237, EPI_ISL_604238, EPI_ISL_604239, EPI_ISL_604240, EPI_ISL_604241, EPI_ISL_604242, EPI_ISL_604243, EPI_ISL_604244, EPI_ISL_604245, EPI_ISL_604246, EPI_ISL_604247, EPI_ISL_604248, EPI_ISL_604249, EPI_ISL_604250, EPI_ISL_604251, EPI_ISL_604252, EPI_ISL_604253, EPI_ISL_604254, EPI_ISL_604255, EPI_ISL_604256, EPI_ISL_604257, EPI_ISL_604258, EPI_ISL_604259, EPI_ISL_604260, EPI_ISL_604261, EPI_ISL_604262, EPI_ISL_604263, EPI_ISL_604264, EPI_ISL_604265, EPI_ISL_604266, EPI_ISL_604267, EPI_ISL_604268, EPI_ISL_604269, EPI_ISL_604270, EPI_ISL_604271, EPI_ISL_604272, EPI_ISL_604273, EPI_ISL_604274, EPI_ISL_604275, EPI_ISL_604276, EPI_ISL_604277, EPI_ISL_604278, EPI_ISL_604357, EPI_ISL_604358, EPI_ISL_604359, EPI_ISL_604360, EPI_ISL_604361, EPI_ISL_604362, EPI_ISL_604363, EPI_ISL_604364, EPI_ISL_604365, EPI_ISL_604366, EPI_ISL_604367, EPI_ISL_604368, EPI_ISL_604369, EPI_ISL_604370, EPI_ISL_604371, EPI_ISL_604484, EPI_ISL_604565, EPI_ISL_604566, EPI_ISL_604567, EPI_ISL_604568, EPI_ISL_604569, EPI_ISL_604570, EPI_ISL_604571, EPI_ISL_604572, EPI_ISL_604573, EPI_ISL_604574, EPI_ISL_604575, EPI_ISL_604576, EPI_ISL_604577, EPI_ISL_604705, EPI_ISL_604706, EPI_ISL_604707, EPI_ISL_604708, EPI_ISL_604709, EPI_ISL_604710, EPI_ISL_604711, EPI_ISL_604712, EPI_ISL_604713, EPI_ISL_604714, EPI_ISL_604715, EPI_ISL_604716, EPI_ISL_604717, EPI_ISL_604794, EPI_ISL_604795, EPI_ISL_604796, EPI_ISL_604797, EPI_ISL_604798, EPI_ISL_604799, EPI_ISL_604800, EPI_ISL_604801, EPI_ISL_604802, EPI_ISL_604803, EPI_ISL_604804, EPI_ISL_604805, EPI_ISL_604806, EPI_ISL_604807, EPI_ISL_604808, EPI_ISL_604809, EPI_ISL_604810, EPI_ISL_604811, EPI_ISL_604812, EPI_ISL_604813, EPI_ISL_604814, EPI_ISL_604815, EPI_ISL_604816, EPI_ISL_604817, EPI_ISL_604818, EPI_ISL_604819, EPI_ISL_604820, EPI_ISL_604821, EPI_ISL_604822, EPI_ISL_604823, EPI_ISL_604824, EPI_ISL_604825, EPI_ISL_604826, EPI_ISL_604827, EPI_ISL_604828, EPI_ISL_604829, EPI_ISL_604830, EPI_ISL_604831, EPI_ISL_604832, EPI_ISL_604833, EPI_ISL_604834, EPI_ISL_604955, EPI_ISL_604956, EPI_ISL_604957, EPI_ISL_604958, EPI_ISL_605038, EPI_ISL_605039, EPI_ISL_605040, EPI_ISL_605041, EPI_ISL_605042, EPI_ISL_605043, EPI_ISL_605044, EPI_ISL_605045, EPI_ISL_605046, EPI_ISL_605047, EPI_ISL_605048, EPI_ISL_605049, EPI_ISL_605050, EPI_ISL_605051, EPI_ISL_605052, EPI_ISL_605053, EPI_ISL_605054, EPI_ISL_605055, EPI_ISL_605056 | | | |
| see above | Quest Diagnostics | Quest Diagnostics | Rosenthal,S.H., Gerasimova,A., Kagan,R.M., Anderson, B., Grover, D., Livingston, K.E., Hua, M., Liu Y., Shalhout, D.F., Owen, R., Lacbawan, F. |
| EPI_ISL_614294 | Maimonides Medical Center | New York City Public Health Laboratory | Jade Wang, et al. |
| EPI_ISL_631502, EPI_ISL_631503 | New York-Presbyterian-Columbia University Medical Center | New York City Public Health Laboratory | Jade Wang, et al. |
| EPI_ISL_631504 | Wyckoff Heights Medical Center | New York City Public Health Laboratory | Jade Wang, et al. |
| EPI_ISL_631505 | Northwell Health-GoHealth Urgent Care 10025 | New York City Public Health Laboratory | Jade Wang, et al. |
| EPI_ISL_631506 | NYC Department Of Health And Mental Hygiene | New York City Public Health Laboratory | Jade Wang, et al. |
| EPI_ISL_631507 | Mount Sinai West | New York City Public Health Laboratory | Jade Wang, et al. |
| EPI_ISL_631508 | Brooklyn Hospital Center | New York City Public Health Laboratory | Jade Wang, et al. |
| EPI_ISL_631509 | New York Presbyterian Lower Manhattan | New York City Public Health Laboratory | Jade Wang, et al. |
| EPI_ISL_631510, EPI_ISL_631511 | New York Presbyterian/ Weill Cornell Medical Center | New York City Public Health Laboratory | Jade Wang, et al. |
| EPI_ISL_631512 | Mount Sinai Hospital | New York City Public Health Laboratory | Jade Wang, et al. |
| EPI_ISL_631513 | Montefiore Medical Center | New York City Public Health Laboratory | Jade Wang, et al. |
| EPI_ISL_631514 | New York Community Hospital | New York City Public Health Laboratory | Jade Wang, et al. |
| EPI_ISL_631515 | NYU Langone Health | New York City Public Health Laboratory | Jade Wang, et al. |

| | | | |
|---|---|---|---|
| EPI_ISL_631516 | Jamaica Hospital Medical Center | New York City Public Health Laboratory | Jade Wang, et al. |
| EPI_ISL_631517, EPI_ISL_631518, EPI_ISL_631519 | New York Presbyterian-Brooklyn Methodist Hospital | New York City Public Health Laboratory | Jade Wang, et al. |
| EPI_ISL_631520 | NYC HH Lincoln Medical And Mental Health Center | New York City Public Health Laboratory | Jade Wang, et al. |
| EPI_ISL_631521 | Mount Sinai Hospital | New York City Public Health Laboratory | Jade Wang, et al. |
| EPI_ISL_631522 | Robert Kutnick | New York City Public Health Laboratory | Jade Wang, et al. |
| EPI_ISL_631523 | NYU Langone Health | New York City Public Health Laboratory | Jade Wang, et al. |
| EPI_ISL_631524 | Jamaica Hospital Medical Center | New York City Public Health Laboratory | Jade Wang, et al. |
| EPI_ISL_631525 | Mount Sinai Hospital | New York City Public Health Laboratory | Jade Wang, et al. |
| EPI_ISL_631526, EPI_ISL_631527 | NYU Langone Health | New York City Public Health Laboratory | Jade Wang, et al. |
| EPI_ISL_631528 | Maimonides Medical Center | New York City Public Health Laboratory | Jade Wang, et al. |
| EPI_ISL_631529 | St. Johns Episcopal Hospital | New York City Public Health Laboratory | Jade Wang, et al. |
| EPI_ISL_631530 | New York Presbyterian Queens | New York City Public Health Laboratory | Jade Wang, et al. |
| EPI_ISL_631531 | NYU Langone Health | New York City Public Health Laboratory | Jade Wang, et al. |
| EPI_ISL_631532 | Mount Sinai Hospital | New York City Public Health Laboratory | Jade Wang, et al. |
| EPI_ISL_631533 | New York Presbyterian Lower Manhattan | New York City Public Health Laboratory | Jade Wang, et al. |
| EPI_ISL_631534 | Mount Sinai Hospital | New York City Public Health Laboratory | Jade Wang, et al. |
| EPI_ISL_631535 | NYC Department Of Health And Mental Hygiene | New York City Public Health Laboratory | Jade Wang, et al. |
| EPI_ISL_631536 | New York Presbyterian Lower Manhattan | New York City Public Health Laboratory | Jade Wang, et al. |
| EPI_ISL_631537 | Mount Sinai Hospital | New York City Public Health Laboratory | Jade Wang, et al. |
| EPI_ISL_631538 | Maimonides Medical Center | New York City Public Health Laboratory | Jade Wang, et al. |
| EPI_ISL_631539, EPI_ISL_631540 | New York Presbyterian Lower Manhattan | New York City Public Health Laboratory | Jade Wang, et al. |
| EPI_ISL_631541 | NYC Department Of Health And Mental Hygiene | New York City Public Health Laboratory | Jade Wang, et al. |
| EPI_ISL_631542 | New York Presbyterian Lower Manhattan | New York City Public Health Laboratory | Jade Wang, et al. |
| EPI_ISL_631543 | Robert Kutnick | New York City Public Health Laboratory | Jade Wang, et al. |
| EPI_ISL_631544, EPI_ISL_631545 | New York Presbyterian-Brooklyn Methodist Hospital | New York City Public Health Laboratory | Jade Wang, et al. |
| EPI_ISL_631546 | Maimonides Medical Center | New York City Public Health Laboratory | Jade Wang, et al. |
| EPI_ISL_631547 | New York Presbyterian Queens | New York City Public Health Laboratory | Jade Wang, et al. |
| EPI_ISL_631548 | New York Presbyterian-Brooklyn Methodist Hospital | New York City Public Health Laboratory | Jade Wang, et al. |
| EPI_ISL_631549 | Mount Sinai Hospital | New York City Public Health Laboratory | Jade Wang, et al. |
| EPI_ISL_631550, EPI_ISL_631551 | Jamaica Hospital Medical Center | New York City Public Health Laboratory | Jade Wang, et al. |
| EPI_ISL_631552, EPI_ISL_631553 | Brookdale University Hospital Medical Center | New York City Public Health Laboratory | Jade Wang, et al. |
| EPI_ISL_631554 | Jamaica Hospital Medical Center | New York City Public Health Laboratory | Jade Wang, et al. |
| EPI_ISL_631555 | Richmond University Medical Center | New York City Public Health Laboratory | Jade Wang, et al. |
| EPI_ISL_631556 | NYC HH Jacobi Medical Center | New York City Public Health Laboratory | Jade Wang, et al. |
| EPI_ISL_631557 | NYC HH Lincoln Medical And Mental Health Center | New York City Public Health Laboratory | Jade Wang, et al. |
| EPI_ISL_631558, EPI_ISL_631559 | NYU Langone Health | New York City Public Health Laboratory | Jade Wang, et al. |
| EPI_ISL_631560, EPI_ISL_631561 | Montefiore Medical Center | New York City Public Health Laboratory | Jade Wang, et al. |
| EPI_ISL_631562 | Brookdale University Hospital Medical Center | New York City Public Health Laboratory | Jade Wang, et al. |
| EPI_ISL_631563 | Flushing Hospital Medical Center | New York City Public Health Laboratory | Jade Wang, et al. |
| EPI_ISL_631564 | NYC HH Elmhurst Hospital Medical Center | New York City Public Health Laboratory | Jade Wang, et al. |
| EPI_ISL_631565 | Brookdale University Hospital Medical Center | New York City Public Health Laboratory | Jade Wang, et al. |
| EPI_ISL_631566, EPI_ISL_631567 | New York Presbyterian-Brooklyn Methodist Hospital | New York City Public Health Laboratory | Jade Wang, et al. |
| EPI_ISL_631568, EPI_ISL_631569 | New York Presbyterian Queens | New York City Public Health Laboratory | Jade Wang, et al. |
| EPI_ISL_631570 | OCME Office Of Chief Medical Examiner | New York City Public Health Laboratory | Jade Wang, et al. |
| EPI_ISL_631571 | Jamaica Hospital Medical Center | New York City Public Health Laboratory | Jade Wang, et al. |
| EPI_ISL_631572 | NYC Department Of Health And Mental Hygiene | New York City Public Health Laboratory | Jade Wang, et al. |
| EPI_ISL_631573 | Brookdale University Hospital Medical Center | New York City Public Health Laboratory | Jade Wang, et al. |
| EPI_ISL_631574, EPI_ISL_631575 | Jamaica Hospital Medical Center | New York City Public Health Laboratory | Jade Wang, et al. |
| EPI_ISL_631576 | Richmond University Medical Center | New York City Public Health Laboratory | Jade Wang, et al. |
| EPI_ISL_631577 | NYC HH Lincoln Medical And Mental Health Center | New York City Public Health Laboratory | Jade Wang, et al. |
| EPI_ISL_631578 | Jamaica Hospital Medical Center | New York City Public Health Laboratory | Jade Wang, et al. |
| EPI_ISL_631579 | Flushing Hospital Medical Center | New York City Public Health Laboratory | Jade Wang, et al. |
| EPI_ISL_631580 | Bellevue Hospital Center | New York City Public Health Laboratory | Jade Wang, et al. |
| EPI_ISL_631581 | SUNY Downstate Medical Center | New York City Public Health Laboratory | Jade Wang, et al. |
| EPI_ISL_631582 | Jamaica Hospital Medical Center | New York City Public Health Laboratory | Jade Wang, et al. |
| EPI_ISL_631583 | Richmond University Medical Center | New York City Public Health Laboratory | Jade Wang, et al. |
| EPI_ISL_631584 | NYC HH Lincoln Medical And Mental Health Center | New York City Public Health Laboratory | Jade Wang, et al. |
| EPI_ISL_631585 | Jamaica Hospital Medical Center | New York City Public Health Laboratory | Jade Wang, et al. |

| | | | |
|---|---|---|---|
| EPI_ISL_631586 | Flushing Hospital Medical Center | New York City Public Health Laboratory | Jade Wang, et al. |
| EPI_ISL_631587 | NYC HH Lincoln Medical And Mental Health Center | New York City Public Health Laboratory | Jade Wang, et al. |
| EPI_ISL_631588 | SUNY Downstate Medical Center | New York City Public Health Laboratory | Jade Wang, et al. |
| EPI_ISL_631589 | Flushing Hospital Medical Center | New York City Public Health Laboratory | Jade Wang, et al. |
| EPI_ISL_631590 | Jamaica Hospital Medical Center | New York City Public Health Laboratory | Jade Wang, et al. |
| EPI_ISL_631591 | OCME Office Of Chief Medical Examiner | New York City Public Health Laboratory | Jade Wang, et al. |
| EPI_ISL_631592, EPI_ISL_631593 | Jamaica Hospital Medical Center | New York City Public Health Laboratory | Jade Wang, et al. |
| EPI_ISL_631594 | OCME Office Of Chief Medical Examiner | New York City Public Health Laboratory | Jade Wang, et al. |
| EPI_ISL_631595 | Jamaica Hospital Medical Center | New York City Public Health Laboratory | Jade Wang, et al. |
| EPI_ISL_631596 | James J. Peters VA Medical Center | New York City Public Health Laboratory | Jade Wang, et al. |
| EPI_ISL_631597 | Jamaica Hospital Medical Center | New York City Public Health Laboratory | Jade Wang, et al. |
| EPI_ISL_631598 | OCME Office Of Chief Medical Examiner | New York City Public Health Laboratory | Jade Wang, et al. |
| EPI_ISL_631599 | Jamaica Hospital Medical Center | New York City Public Health Laboratory | Jade Wang, et al. |
| EPI_ISL_631600 | James J. Peters VA Medical Center | New York City Public Health Laboratory | Jade Wang, et al. |
| EPI_ISL_631601 | Jamaica Hospital Medical Center | New York City Public Health Laboratory | Jade Wang, et al. |
| EPI_ISL_631602 | Flushing Hospital Medical Center | New York City Public Health Laboratory | Jade Wang, et al. |
| EPI_ISL_631603 | NYC HH Lincoln Medical And Mental Health Center | New York City Public Health Laboratory | Jade Wang, et al. |
| EPI_ISL_631604 | Jamaica Hospital Medical Center | New York City Public Health Laboratory | Jade Wang, et al. |
| EPI_ISL_631605 | OCME Office Of Chief Medical Examiner | New York City Public Health Laboratory | Jade Wang, et al. |
| EPI_ISL_631606 | Flushing Hospital Medical Center | New York City Public Health Laboratory | Jade Wang, et al. |
| EPI_ISL_631607 | NYC HH Lincoln Medical And Mental Health Center | New York City Public Health Laboratory | Jade Wang, et al. |
| EPI_ISL_631608, EPI_ISL_631609, EPI_ISL_631610 | Jamaica Hospital Medical Center | New York City Public Health Laboratory | Jade Wang, et al. |
| EPI_ISL_631611 | OCME Office Of Chief Medical Examiner | New York City Public Health Laboratory | Jade Wang, et al. |
| EPI_ISL_631612, EPI_ISL_631613 | Jamaica Hospital Medical Center | New York City Public Health Laboratory | Jade Wang, et al. |
| EPI_ISL_631614 | Flushing Hospital Medical Center | New York City Public Health Laboratory | Jade Wang, et al. |
| EPI_ISL_631615 | Jamaica Hospital Medical Center | New York City Public Health Laboratory | Jade Wang, et al. |
| EPI_ISL_631616 | Flushing Hospital Medical Center | New York City Public Health Laboratory | Jade Wang, et al. |
| EPI_ISL_631617, EPI_ISL_631618, EPI_ISL_631619 | OCME Office Of Chief Medical Examiner | New York City Public Health Laboratory | Jade Wang, et al. |
| EPI_ISL_631620 | Flushing Hospital Medical Center | New York City Public Health Laboratory | Jade Wang, et al. |
| EPI_ISL_631621 | Jamaica Hospital Medical Center | New York City Public Health Laboratory | Jade Wang, et al. |
| EPI_ISL_631622, EPI_ISL_631623, EPI_ISL_631624, EPI_ISL_631625, EPI_ISL_631626, EPI_ISL_631627, EPI_ISL_631628, EPI_ISL_631629, EPI_ISL_631630, EPI_ISL_631631, EPI_ISL_631632, EPI_ISL_631633 | | | |
| see above | Richmond University Medical Center | New York City Public Health Laboratory | Jade Wang, et al. |
| EPI_ISL_631634 | Jamaica Hospital Medical Center | New York City Public Health Laboratory | Jade Wang, et al. |
| EPI_ISL_631635 | OCME Office Of Chief Medical Examiner | New York City Public Health Laboratory | Jade Wang, et al. |
| EPI_ISL_631636, EPI_ISL_631637 | Jamaica Hospital Medical Center | New York City Public Health Laboratory | Jade Wang, et al. |
| EPI_ISL_631638 | Flushing Hospital Medical Center | New York City Public Health Laboratory | Jade Wang, et al. |
| EPI_ISL_631639, EPI_ISL_631685 | OCME Office Of Chief Medical Examiner | New York City Public Health Laboratory | Jade Wang, et al. |
| EPI_ISL_631686 | NYC HH Lincoln Medical And Mental Health Center | New York City Public Health Laboratory | Jade Wang, et al. |
| EPI_ISL_631687 | OCME Office Of Chief Medical Examiner | New York City Public Health Laboratory | Jade Wang, et al. |
| EPI_ISL_631688 | Wildlife Conservation Society - Bronx Zoo | New York City Public Health Laboratory | Jade Wang, et al. |
| EPI_ISL_631689, EPI_ISL_631690 | Lincoln Medical Center | New York City Public Health Laboratory | Jade Wang, et al. |
| EPI_ISL_631691 | NYC HH Lincoln Medical And Mental Health Center | New York City Public Health Laboratory | Jade Wang, et al. |
| EPI_ISL_631692 | Flushing Hospital Medical Center | New York City Public Health Laboratory | Jade Wang, et al. |
| EPI_ISL_631693 | NYC HH Lincoln Medical And Mental Health Center | New York City Public Health Laboratory | Jade Wang, et al. |
| EPI_ISL_631694, EPI_ISL_631695 | OCME Office Of Chief Medical Examiner | New York City Public Health Laboratory | Jade Wang, et al. |
| EPI_ISL_631696 | Flushing Hospital Medical Center | New York City Public Health Laboratory | Jade Wang, et al. |
| EPI_ISL_631697, EPI_ISL_631698 | Jamaica Hospital Medical Center | New York City Public Health Laboratory | Jade Wang, et al. |
| EPI_ISL_631699, EPI_ISL_631700, EPI_ISL_631701 | Flushing Hospital Medical Center | New York City Public Health Laboratory | Jade Wang, et al. |
| EPI_ISL_631702, EPI_ISL_631703, EPI_ISL_631704, EPI_ISL_631705, EPI_ISL_631706 | Jamaica Hospital Medical Center | New York City Public Health Laboratory | Jade Wang, et al. |
| EPI_ISL_631707, EPI_ISL_631708 | Flushing Hospital Medical Center | New York City Public Health Laboratory | Jade Wang, et al. |
| EPI_ISL_631709 | Jamaica Hospital Medical Center | New York City Public Health Laboratory | Jade Wang, et al. |
| EPI_ISL_631710 | Flushing Hospital Medical Center | New York City Public Health Laboratory | Jade Wang, et al. |
| EPI_ISL_631711, EPI_ISL_631712, EPI_ISL_631713 | Jamaica Hospital Medical Center | New York City Public Health Laboratory | Jade Wang, et al. |
| EPI_ISL_631714, EPI_ISL_631715 | Flushing Hospital Medical Center | New York City Public Health Laboratory | Jade Wang, et al. |
| EPI_ISL_631716, EPI_ISL_631717, | Jamaica Hospital Medical Center | New York City Public Health Laboratory | Jade Wang, et al. |

| | | | |
|---|---|---|---|
| EPI_ISL_631718, EPI_ISL_631719, EPI_ISL_631720, EPI_ISL_631721, EPI_ISL_631722, EPI_ISL_631723 | | | |
| EPI_ISL_631724, EPI_ISL_631725 | Flushing Hospital Medical Center | New York City Public Health Laboratory | Jade Wang, et al. |
| EPI_ISL_631726 | Jamaica Hospital Medical Center | New York City Public Health Laboratory | Jade Wang, et al. |
| EPI_ISL_631727 | Flushing Hospital Medical Center | New York City Public Health Laboratory | Jade Wang, et al. |
| EPI_ISL_631728, EPI_ISL_631729, EPI_ISL_631730, EPI_ISL_631731 | Jamaica Hospital Medical Center | New York City Public Health Laboratory | Jade Wang, et al. |
| EPI_ISL_631732 | Flushing Hospital Medical Center | New York City Public Health Laboratory | Jade Wang, et al. |
| EPI_ISL_631733, EPI_ISL_631734, EPI_ISL_631735, EPI_ISL_631736, EPI_ISL_631737, EPI_ISL_631738, EPI_ISL_631739, EPI_ISL_631740, EPI_ISL_631741, EPI_ISL_631742, EPI_ISL_631743, EPI_ISL_631744, EPI_ISL_631745, EPI_ISL_631746, EPI_ISL_631747 | | | |
| see above | Jamaica Hospital Medical Center | New York City Public Health Laboratory | Jade Wang, et al. |
| EPI_ISL_631748, EPI_ISL_631749, EPI_ISL_631750, EPI_ISL_631751 | St Barnabas Hospital | New York City Public Health Laboratory | Jade Wang, et al. |
| EPI_ISL_631752, EPI_ISL_631753, EPI_ISL_631754 | NYC HH Lincoln Medical And Mental Health Center | New York City Public Health Laboratory | Jade Wang, et al. |
| EPI_ISL_631755 | Richmond University Medical Center | New York City Public Health Laboratory | Jade Wang, et al. |
| EPI_ISL_631756 | Flushing Hospital Medical Center | New York City Public Health Laboratory | Jade Wang, et al. |
| EPI_ISL_631757 | Richmond University Medical Center | New York City Public Health Laboratory | Jade Wang, et al. |
| EPI_ISL_631758, EPI_ISL_631759, EPI_ISL_631760, EPI_ISL_631761, EPI_ISL_631762, EPI_ISL_631763 | OCME Office Of Chief Medical Examiner | New York City Public Health Laboratory | Jade Wang, et al. |
| EPI_ISL_631764 | Mount Sinai Hospital | New York City Public Health Laboratory | Jade Wang, et al. |
| EPI_ISL_631765 | Bellevue Hospital Center | New York City Public Health Laboratory | Jade Wang, et al. |
| EPI_ISL_631766 | Flushing Hospital Medical Center | New York City Public Health Laboratory | Jade Wang, et al. |
| EPI_ISL_631767, EPI_ISL_631768 | Jamaica Hospital Medical Center | New York City Public Health Laboratory | Jade Wang, et al. |
| EPI_ISL_631769, EPI_ISL_631770, EPI_ISL_631771 | Flushing Hospital Medical Center | New York City Public Health Laboratory | Jade Wang, et al. |
| EPI_ISL_631772 | Jamaica Hospital Medical Center | New York City Public Health Laboratory | Jade Wang, et al. |
| EPI_ISL_631773, EPI_ISL_631774 | Flushing Hospital Medical Center | New York City Public Health Laboratory | Jade Wang, et al. |
| EPI_ISL_631775, EPI_ISL_631776, EPI_ISL_631777 | Jamaica Hospital Medical Center | New York City Public Health Laboratory | Jade Wang, et al. |
| EPI_ISL_631778, EPI_ISL_631779, EPI_ISL_631780, EPI_ISL_631781, EPI_ISL_631782, EPI_ISL_631783, EPI_ISL_631784 | St Barnabas Hospital | New York City Public Health Laboratory | Jade Wang, et al. |
| EPI_ISL_631785, EPI_ISL_631786 | Mount Sinai Hospital | New York City Public Health Laboratory | Jade Wang, et al. |
| EPI_ISL_631787 | Flushing Hospital Medical Center | New York City Public Health Laboratory | Jade Wang, et al. |
| EPI_ISL_631788, EPI_ISL_631789, EPI_ISL_631790, EPI_ISL_631791 | Jamaica Hospital Medical Center | New York City Public Health Laboratory | Jade Wang, et al. |
| EPI_ISL_631792 | Richmond University Medical Center | New York City Public Health Laboratory | Jade Wang, et al. |
| EPI_ISL_631793, EPI_ISL_631794 | SUNY Downstate Medical Center | New York City Public Health Laboratory | Jade Wang, et al. |
| EPI_ISL_631795 | Jamaica Hospital Medical Center | New York City Public Health Laboratory | Jade Wang, et al. |
| EPI_ISL_631796 | Maimonides Medical Center | New York City Public Health Laboratory | Jade Wang, et al. |
| EPI_ISL_631797 | OCME Office Of Chief Medical Examiner | New York City Public Health Laboratory | Jade Wang, et al. |
| EPI_ISL_631798 | Jamaica Hospital Medical Center | New York City Public Health Laboratory | Jade Wang, et al. |
| EPI_ISL_631799 | NYC HH Lincoln Medical And Mental Health Center | New York City Public Health Laboratory | Jade Wang, et al. |
| EPI_ISL_631800 | Brookdale University Hospital Medical Center | New York City Public Health Laboratory | Jade Wang, et al. |
| EPI_ISL_631801 | Richmond University Medical Center | New York City Public Health Laboratory | Jade Wang, et al. |
| EPI_ISL_631802 | SUNY Downstate Medical Center | New York City Public Health Laboratory | Jade Wang, et al. |
| EPI_ISL_631803 | Brookdale University Hospital Medical Center | New York City Public Health Laboratory | Jade Wang, et al. |
| EPI_ISL_631804 | Bellevue Hospital Center | New York City Public Health Laboratory | Jade Wang, et al. |
| EPI_ISL_631805, EPI_ISL_631806, EPI_ISL_631807 | Brookdale University Hospital Medical Center | New York City Public Health Laboratory | Jade Wang, et al. |
| EPI_ISL_631808 | Wyckoff Heights Medical Center | New York City Public Health Laboratory | Jade Wang, et al. |
| EPI_ISL_631809, EPI_ISL_631810 | Jamaica Hospital Medical Center | New York City Public Health Laboratory | Jade Wang, et al. |
| EPI_ISL_631811 | Wyckoff Heights Medical Center | New York City Public Health Laboratory | Jade Wang, et al. |
| EPI_ISL_631812, EPI_ISL_631813 | Jamaica Hospital Medical Center | New York City Public Health Laboratory | Jade Wang, et al. |
| EPI_ISL_631814, EPI_ISL_631815, EPI_ISL_631816, EPI_ISL_631817, EPI_ISL_631818 | Richmond University Medical Center | New York City Public Health Laboratory | Jade Wang, et al. |
| EPI_ISL_631819 | Brookdale University Hospital Medical Center | New York City Public Health Laboratory | Jade Wang, et al. |
| EPI_ISL_631820 | Jamaica Hospital Medical Center | New York City Public Health Laboratory | Jade Wang, et al. |
| EPI_ISL_631821 | Richmond University Medical Center | New York City Public Health Laboratory | Jade Wang, et al. |
| EPI_ISL_631822, EPI_ISL_631823, | Jamaica Hospital Medical Center | New York City Public Health Laboratory | Jade Wang, et al. |

| | | | |
|---|---|---|---|
| EPI_ISL_631825 | | | |
| EPI_ISL_631826 | Flushing Hospital Medical Center | New York City Public Health Laboratory | Jade Wang, et al. |
| EPI_ISL_631827 | NYC HH Lincoln Medical And Mental Health Center | New York City Public Health Laboratory | Jade Wang, et al. |
| EPI_ISL_631828 | Richmond University Medical Center | New York City Public Health Laboratory | Jade Wang, et al. |
| EPI_ISL_631829 | Brookdale University Hospital Medical Center | New York City Public Health Laboratory | Jade Wang, et al. |
| EPI_ISL_631830 | Mount Sinai Hospital | New York City Public Health Laboratory | Jade Wang, et al. |
| EPI_ISL_631831 | VA NY Harbor Healthcare System | New York City Public Health Laboratory | Jade Wang, et al. |
| EPI_ISL_631832 | Metro Urgicare | New York City Public Health Laboratory | Jade Wang, et al. |
| EPI_ISL_631833 | Jamaica Hospital Medical Center | New York City Public Health Laboratory | Jade Wang, et al. |
| EPI_ISL_631834 | Mount Sinai Hospital | New York City Public Health Laboratory | Jade Wang, et al. |
| EPI_ISL_631835, EPI_ISL_631836 | Jamaica Hospital Medical Center | New York City Public Health Laboratory | Jade Wang, et al. |
| EPI_ISL_631837 | Richmond University Medical Center | New York City Public Health Laboratory | Jade Wang, et al. |
| EPI_ISL_631838 | Bellevue Hospital Center | New York City Public Health Laboratory | Jade Wang, et al. |
| EPI_ISL_631839 | Mount Sinai Hospital | New York City Public Health Laboratory | Jade Wang, et al. |
| EPI_ISL_631840, EPI_ISL_631841, EPI_ISL_631842, EPI_ISL_631843, EPI_ISL_631844, EPI_ISL_631845, EPI_ISL_631846 | OCME Office Of Chief Medical Examiner | New York City Public Health Laboratory | Jade Wang, et al. |
| EPI_ISL_631847 | Jamaica Hospital Medical Center | New York City Public Health Laboratory | Jade Wang, et al. |
| EPI_ISL_631848 | Flushing Hospital Medical Center | New York City Public Health Laboratory | Jade Wang, et al. |
| EPI_ISL_631849, EPI_ISL_631850 | Jamaica Hospital Medical Center | New York City Public Health Laboratory | Jade Wang, et al. |
| EPI_ISL_631851 | Flushing Hospital Medical Center | New York City Public Health Laboratory | Jade Wang, et al. |
| EPI_ISL_631852, EPI_ISL_631853, EPI_ISL_631854 | Jamaica Hospital Medical Center | New York City Public Health Laboratory | Jade Wang, et al. |
| EPI_ISL_631855 | Flushing Hospital Medical Center | New York City Public Health Laboratory | Jade Wang, et al. |
| EPI_ISL_631856, EPI_ISL_631857, EPI_ISL_631858, EPI_ISL_631859 | St Barnabas Hospital | New York City Public Health Laboratory | Jade Wang, et al. |
| EPI_ISL_631860 | New York Presbyterian/ Weill Cornell Medical Center | New York City Public Health Laboratory | Jade Wang, et al. |
| EPI_ISL_631861 | New York Presbyterian Queens | New York City Public Health Laboratory | Jade Wang, et al. |
| EPI_ISL_631862 | NYC Department Of Health And Mental Hygiene | New York City Public Health Laboratory | Jade Wang, et al. |
| EPI_ISL_631863 | Maimonides Medical Center | New York City Public Health Laboratory | Jade Wang, et al. |
| EPI_ISL_631864 | Brookdale University Hospital Medical Center | New York City Public Health Laboratory | Jade Wang, et al. |
| EPI_ISL_631865 | NYC HH Elmhurst Hospital Medical Center | New York City Public Health Laboratory | Jade Wang, et al. |
| EPI_ISL_631866, EPI_ISL_631867 | Jamaica Hospital Medical Center | New York City Public Health Laboratory | Jade Wang, et al. |
| EPI_ISL_631868, EPI_ISL_631869 | Richmond University Medical Center | New York City Public Health Laboratory | Jade Wang, et al. |
| EPI_ISL_631870, EPI_ISL_631871, EPI_ISL_631872 | Jamaica Hospital Medical Center | New York City Public Health Laboratory | Jade Wang, et al. |
| EPI_ISL_631873 | Richmond University Medical Center | New York City Public Health Laboratory | Jade Wang, et al. |
| EPI_ISL_631874 | James J. Peters VA Medical Center | New York City Public Health Laboratory | Jade Wang, et al. |
| EPI_ISL_631875 | Richmond University Medical Center | New York City Public Health Laboratory | Jade Wang, et al. |
| EPI_ISL_631876, EPI_ISL_631877, EPI_ISL_631878, EPI_ISL_631879 | Jamaica Hospital Medical Center | New York City Public Health Laboratory | Jade Wang, et al. |
| EPI_ISL_631880 | Mount Sinai West | New York City Public Health Laboratory | Jade Wang, et al. |
| EPI_ISL_631881 | Maimonides Medical Center | New York City Public Health Laboratory | Jade Wang, et al. |
| EPI_ISL_631882, EPI_ISL_631883 | Jamaica Hospital Medical Center | New York City Public Health Laboratory | Jade Wang, et al. |
| EPI_ISL_631884 | Brookdale University Hospital Medical Center | New York City Public Health Laboratory | Jade Wang, et al. |
| EPI_ISL_631885 | Flushing Hospital Medical Center | New York City Public Health Laboratory | Jade Wang, et al. |
| EPI_ISL_631886 | NYC HH Lincoln Medical And Mental Health Center | New York City Public Health Laboratory | Jade Wang, et al. |
| EPI_ISL_631887 | Jamaica Hospital Medical Center | New York City Public Health Laboratory | Jade Wang, et al. |
| EPI_ISL_631888 | St Barnabas Hospital | New York City Public Health Laboratory | Jade Wang, et al. |
| EPI_ISL_631889, EPI_ISL_631890 | Jamaica Hospital Medical Center | New York City Public Health Laboratory | Jade Wang, et al. |
| EPI_ISL_631891 | Flushing Hospital Medical Center | New York City Public Health Laboratory | Jade Wang, et al. |
| EPI_ISL_631892 | Richmond University Medical Center | New York City Public Health Laboratory | Jade Wang, et al. |
| EPI_ISL_631893, EPI_ISL_631894 | Flushing Hospital Medical Center | New York City Public Health Laboratory | Jade Wang, et al. |
| EPI_ISL_631895, EPI_ISL_631896 | Jamaica Hospital Medical Center | New York City Public Health Laboratory | Jade Wang, et al. |
| EPI_ISL_631897 | Richmond University Medical Center | New York City Public Health Laboratory | Jade Wang, et al. |
| EPI_ISL_631898 | OCME Office Of Chief Medical Examiner | New York City Public Health Laboratory | Jade Wang, et al. |
| EPI_ISL_631899 | Richmond University Medical Center | New York City Public Health Laboratory | Jade Wang, et al. |
| EPI_ISL_631900 | St Barnabas Hospital | New York City Public Health Laboratory | Jade Wang, et al. |
| EPI_ISL_631901, EPI_ISL_631902 | Jamaica Hospital Medical Center | New York City Public Health Laboratory | Jade Wang, et al. |
| EPI_ISL_631903 | Richmond University Medical Center | New York City Public Health Laboratory | Jade Wang, et al. |

| | | | |
|---|---|---|---|
| EPI_ISL_631904 | Jamaica Hospital Medical Center | New York City Public Health Laboratory | Jade Wang, et al. |
| EPI_ISL_631905 | Richmond University Medical Center | New York City Public Health Laboratory | Jade Wang, et al. |
| EPI_ISL_631906, EPI_ISL_631907, EPI_ISL_631908 | Flushing Hospital Medical Center | New York City Public Health Laboratory | Jade Wang, et al. |
| EPI_ISL_631909, EPI_ISL_631910 | Jamaica Hospital Medical Center | New York City Public Health Laboratory | Jade Wang, et al. |
| EPI_ISL_631911, EPI_ISL_631912 | St Barnabas Hospital | New York City Public Health Laboratory | Jade Wang, et al. |
| EPI_ISL_631913, EPI_ISL_631914 | Mount Sinai Hospital | New York City Public Health Laboratory | Jade Wang, et al. |
| EPI_ISL_631915 | Maimonides Medical Center | New York City Public Health Laboratory | Jade Wang, et al. |
| EPI_ISL_631916 | Flushing Hospital Medical Center | New York City Public Health Laboratory | Jade Wang, et al. |
| EPI_ISL_631917 | St Barnabas Hospital | New York City Public Health Laboratory | Jade Wang, et al. |
| EPI_ISL_631918 | Flushing Hospital Medical Center | New York City Public Health Laboratory | Jade Wang, et al. |
| EPI_ISL_631919, EPI_ISL_631920 | Richmond University Medical Center | New York City Public Health Laboratory | Jade Wang, et al. |
| EPI_ISL_631921, EPI_ISL_631922 | Jamaica Hospital Medical Center | New York City Public Health Laboratory | Jade Wang, et al. |
| EPI_ISL_631923 | Flushing Hospital Medical Center | New York City Public Health Laboratory | Jade Wang, et al. |
| EPI_ISL_631924 | St Barnabas Hospital | New York City Public Health Laboratory | Jade Wang, et al. |
| EPI_ISL_631925, EPI_ISL_631926, EPI_ISL_631927, EPI_ISL_631928 | Jamaica Hospital Medical Center | New York City Public Health Laboratory | Jade Wang, et al. |
| EPI_ISL_631929 | Flushing Hospital Medical Center | New York City Public Health Laboratory | Jade Wang, et al. |
| EPI_ISL_631930 | Jamaica Hospital Medical Center | New York City Public Health Laboratory | Jade Wang, et al. |
| EPI_ISL_631931 | Flushing Hospital Medical Center | New York City Public Health Laboratory | Jade Wang, et al. |
| EPI_ISL_631932 | OCME Office Of Chief Medical Examiner | New York City Public Health Laboratory | Jade Wang, et al. |
| EPI_ISL_631933 | St Barnabas Hospital | New York City Public Health Laboratory | Jade Wang, et al. |
| EPI_ISL_631934 | OCME Office Of Chief Medical Examiner | New York City Public Health Laboratory | Jade Wang, et al. |
| EPI_ISL_631935 | St Barnabas Hospital | New York City Public Health Laboratory | Jade Wang, et al. |
| EPI_ISL_631936, EPI_ISL_631937, EPI_ISL_631938 | OCME Office Of Chief Medical Examiner | New York City Public Health Laboratory | Jade Wang, et al. |
| EPI_ISL_631939, EPI_ISL_631940, EPI_ISL_631941, EPI_ISL_631942, EPI_ISL_631943, EPI_ISL_631944, EPI_ISL_631945, EPI_ISL_631946, EPI_ISL_631947, EPI_ISL_631948, EPI_ISL_631949 | | | |
| see above | St Barnabas Hospital | New York City Public Health Laboratory | Jade Wang, et al. |
| EPI_ISL_631950, EPI_ISL_631951 | Jamaica Hospital Medical Center | New York City Public Health Laboratory | Jade Wang, et al. |
| EPI_ISL_631952 | St Barnabas Hospital | New York City Public Health Laboratory | Jade Wang, et al. |
| EPI_ISL_631953 | NYC HH Lincoln Medical And Mental Health Center | New York City Public Health Laboratory | Jade Wang, et al. |
| EPI_ISL_631954 | Richmond University Medical Center | New York City Public Health Laboratory | Jade Wang, et al. |
| EPI_ISL_631955 | Flushing Hospital Medical Center | New York City Public Health Laboratory | Jade Wang, et al. |
| EPI_ISL_631956 | OCME Office Of Chief Medical Examiner | New York City Public Health Laboratory | Jade Wang, et al. |
| EPI_ISL_631957, EPI_ISL_631958 | St Barnabas Hospital | New York City Public Health Laboratory | Jade Wang, et al. |
| EPI_ISL_631959, EPI_ISL_631960, EPI_ISL_631961 | Flushing Hospital Medical Center | New York City Public Health Laboratory | Jade Wang, et al. |
| EPI_ISL_631962 | Richmond University Medical Center | New York City Public Health Laboratory | Jade Wang, et al. |
| EPI_ISL_631963 | Jamaica Hospital Medical Center | New York City Public Health Laboratory | Jade Wang, et al. |
| EPI_ISL_631964 | Flushing Hospital Medical Center | New York City Public Health Laboratory | Jade Wang, et al. |
| EPI_ISL_631965 | Richmond University Medical Center | New York City Public Health Laboratory | Jade Wang, et al. |
| EPI_ISL_631966 | Jamaica Hospital Medical Center | New York City Public Health Laboratory | Jade Wang, et al. |
| EPI_ISL_631967 | Richmond University Medical Center | New York City Public Health Laboratory | Jade Wang, et al. |
| EPI_ISL_631968 | Flushing Hospital Medical Center | New York City Public Health Laboratory | Jade Wang, et al. |
| EPI_ISL_631969, EPI_ISL_631970 | Jamaica Hospital Medical Center | New York City Public Health Laboratory | Jade Wang, et al. |
| EPI_ISL_631971 | St Barnabas Hospital | New York City Public Health Laboratory | Jade Wang, et al. |
| EPI_ISL_631972 | Richmond University Medical Center | New York City Public Health Laboratory | Jade Wang, et al. |
| EPI_ISL_631973 | Jamaica Hospital Medical Center | New York City Public Health Laboratory | Jade Wang, et al. |
| EPI_ISL_631974 | NYC HH Coney Island Hospital | New York City Public Health Laboratory | Jade Wang, et al. |
| EPI_ISL_631975 | Richmond University Medical Center | New York City Public Health Laboratory | Jade Wang, et al. |
| EPI_ISL_631976 | Brookdale University Hospital Medical Center | New York City Public Health Laboratory | Jade Wang, et al. |
| EPI_ISL_631977 | SUNY Downstate Medical Center | New York City Public Health Laboratory | Jade Wang, et al. |
| EPI_ISL_631978 | Brookdale University Hospital Medical Center | New York City Public Health Laboratory | Jade Wang, et al. |
| EPI_ISL_631979 | Jamaica Hospital Medical Center | New York City Public Health Laboratory | Jade Wang, et al. |
| EPI_ISL_631980 | St Barnabas Hospital | New York City Public Health Laboratory | Jade Wang, et al. |
| EPI_ISL_631981 | Brookdale University Hospital Medical Center | New York City Public Health Laboratory | Jade Wang, et al. |
| EPI_ISL_631982 | Montefiore Medical Center | New York City Public Health Laboratory | Jade Wang, et al. |
| EPI_ISL_631983 | Mount Sinai Hospital | New York City Public Health Laboratory | Jade Wang, et al. |
| EPI_ISL_631984 | Brookdale University Hospital Medical Center | New York City Public Health Laboratory | Jade Wang, et al. |
| EPI_ISL_631985 | St Barnabas Hospital | New York City Public Health Laboratory | Jade Wang, et al. |

| | | | |
|---|---|---|---|
| EPI_ISL_631986 | Flushing Hospital Medical Center | New York City Public Health Laboratory | Jade Wang, et al. |
| EPI_ISL_631987, EPI_ISL_631988 | St Barnabas Hospital | New York City Public Health Laboratory | Jade Wang, et al. |
| EPI_ISL_631989 | Flushing Hospital Medical Center | New York City Public Health Laboratory | Jade Wang, et al. |
| EPI_ISL_631990 | St Barnabas Hospital | New York City Public Health Laboratory | Jade Wang, et al. |
| EPI_ISL_631991 | Flushing Hospital Medical Center | New York City Public Health Laboratory | Jade Wang, et al. |
| EPI_ISL_631992 | Richmond University Medical Center | New York City Public Health Laboratory | Jade Wang, et al. |
| EPI_ISL_631993 | Maimonides Medical Center | New York City Public Health Laboratory | Jade Wang, et al. |
| EPI_ISL_631994 | Jamaica Hospital Medical Center | New York City Public Health Laboratory | Jade Wang, et al. |
| EPI_ISL_631995 | Wyckoff Heights Medical Center | New York City Public Health Laboratory | Jade Wang, et al. |
| EPI_ISL_631996, EPI_ISL_631997 | Jamaica Hospital Medical Center | New York City Public Health Laboratory | Jade Wang, et al. |
| EPI_ISL_631998, EPI_ISL_631999, EPI_ISL_632000, EPI_ISL_632001, EPI_ISL_632002 | Richmond University Medical Center | New York City Public Health Laboratory | Jade Wang, et al. |
| EPI_ISL_632003, EPI_ISL_632004 | Flushing Hospital Medical Center | New York City Public Health Laboratory | Jade Wang, et al. |
| EPI_ISL_632005 | Jamaica Hospital Medical Center | New York City Public Health Laboratory | Jade Wang, et al. |
| EPI_ISL_632006 | OCME Office Of Chief Medical Examiner | New York City Public Health Laboratory | Jade Wang, et al. |
| EPI_ISL_632007, EPI_ISL_632008, EPI_ISL_632009 | Jamaica Hospital Medical Center | New York City Public Health Laboratory | Jade Wang, et al. |
| EPI_ISL_632010 | Flushing Hospital Medical Center | New York City Public Health Laboratory | Jade Wang, et al. |
| EPI_ISL_632011, EPI_ISL_632012 | Jamaica Hospital Medical Center | New York City Public Health Laboratory | Jade Wang, et al. |
| EPI_ISL_632013, EPI_ISL_632014, EPI_ISL_632015, EPI_ISL_632016 | Richmond University Medical Center | New York City Public Health Laboratory | Jade Wang, et al. |
| EPI_ISL_632017 | Jamaica Hospital Medical Center | New York City Public Health Laboratory | Jade Wang, et al. |
| EPI_ISL_632018, EPI_ISL_632019, EPI_ISL_632020 | Flushing Hospital Medical Center | New York City Public Health Laboratory | Jade Wang, et al. |
| EPI_ISL_632021 | Jamaica Hospital Medical Center | New York City Public Health Laboratory | Jade Wang, et al. |
| EPI_ISL_632022 | Flushing Hospital Medical Center | New York City Public Health Laboratory | Jade Wang, et al. |
| EPI_ISL_632023 | Jamaica Hospital Medical Center | New York City Public Health Laboratory | Jade Wang, et al. |
| EPI_ISL_632024 | Flushing Hospital Medical Center | New York City Public Health Laboratory | Jade Wang, et al. |
| EPI_ISL_632025 | Jamaica Hospital Medical Center | New York City Public Health Laboratory | Jade Wang, et al. |
| EPI_ISL_632026 | NYC HH Lincoln Medical And Mental Health Center | New York City Public Health Laboratory | Jade Wang, et al. |
| EPI_ISL_632027 | Flushing Hospital Medical Center | New York City Public Health Laboratory | Jade Wang, et al. |
| EPI_ISL_632028, EPI_ISL_632029 | Jamaica Hospital Medical Center | New York City Public Health Laboratory | Jade Wang, et al. |
| EPI_ISL_632030, EPI_ISL_632031, EPI_ISL_632032, EPI_ISL_632033 | Richmond University Medical Center | New York City Public Health Laboratory | Jade Wang, et al. |
| EPI_ISL_632034 | Flushing Hospital Medical Center | New York City Public Health Laboratory | Jade Wang, et al. |
| EPI_ISL_632035 | Jamaica Hospital Medical Center | New York City Public Health Laboratory | Jade Wang, et al. |
| EPI_ISL_632036 | Richmond University Medical Center | New York City Public Health Laboratory | Jade Wang, et al. |
| EPI_ISL_632037 | Jamaica Hospital Medical Center | New York City Public Health Laboratory | Jade Wang, et al. |
| EPI_ISL_632038 | New York Presbyterian Lower Manhattan | New York City Public Health Laboratory | Jade Wang, et al. |
| EPI_ISL_632039 | NYC Department Of Health And Mental Hygiene | New York City Public Health Laboratory | Jade Wang, et al. |
| EPI_ISL_632040 | New York Presbyterian Queens | New York City Public Health Laboratory | Jade Wang, et al. |
| EPI_ISL_632041, EPI_ISL_632042 | New York Presbyterian Lower Manhattan | New York City Public Health Laboratory | Jade Wang, et al. |
| EPI_ISL_632043, EPI_ISL_632044 | NYU Langone Health | New York City Public Health Laboratory | Jade Wang, et al. |
| EPI_ISL_632045 | New York Presbyterian Lower Manhattan | New York City Public Health Laboratory | Jade Wang, et al. |
| EPI_ISL_632046 | New York Presbyterian-Brooklyn Methodist Hospital | New York City Public Health Laboratory | Jade Wang, et al. |
| EPI_ISL_632047 | New York Presbyterian Lower Manhattan | New York City Public Health Laboratory | Jade Wang, et al. |
| EPI_ISL_632048 | New York Presbyterian-Brooklyn Methodist Hospital | New York City Public Health Laboratory | Jade Wang, et al. |
| EPI_ISL_632049 | New York Presbyterian Queens | New York City Public Health Laboratory | Jade Wang, et al. |
| EPI_ISL_632050 | Long Island Jewish Medical Center | New York City Public Health Laboratory | Jade Wang, et al. |
| EPI_ISL_632051 | Jamaica Hospital Medical Center | New York City Public Health Laboratory | Jade Wang, et al. |
| EPI_ISL_632052, EPI_ISL_632053 | New York Presbyterian Queens | New York City Public Health Laboratory | Jade Wang, et al. |
| EPI_ISL_632054 | New York Presbyterian-Brooklyn Methodist Hospital | New York City Public Health Laboratory | Jade Wang, et al. |
| EPI_ISL_632055, EPI_ISL_632056 | New York Community Hospital | New York City Public Health Laboratory | Jade Wang, et al. |
| EPI_ISL_632057 | New York-Presbyterian-Columbia University Medical Center | New York City Public Health Laboratory | Jade Wang, et al. |
| EPI_ISL_632058, EPI_ISL_632059 | Proximedical Urgent Care | New York City Public Health Laboratory | Jade Wang, et al. |
| EPI_ISL_632060 | NYC HH Queens Hospital Center | New York City Public Health Laboratory | Jade Wang, et al. |
| EPI_ISL_632061 | Jamaica Hospital Medical Center | New York City Public Health Laboratory | Jade Wang, et al. |
| EPI_ISL_632062 | NYC Department Of Health And Mental Hygiene | New York City Public Health Laboratory | Jade Wang, et al. |
| EPI_ISL_632063, EPI_ISL_632064 | New York Presbyterian Queens | New York City Public Health Laboratory | Jade Wang, et al. |
| EPI_ISL_632065, EPI_ISL_632066 | Jamaica Hospital Medical Center | New York City Public Health Laboratory | Jade Wang, et al. |

| | | | |
|---|---|---|---|
| EPI_ISL_632067 | NYU Langone Health | New York City Public Health Laboratory | Jade Wang, et al. |
| EPI_ISL_632068, EPI_ISL_632069 | Bellevue Hospital Center | New York City Public Health Laboratory | Jade Wang, et al. |
| EPI_ISL_632070, EPI_ISL_632071 | New York Presbyterian Queens | New York City Public Health Laboratory | Jade Wang, et al. |
| EPI_ISL_632072 | NYC HH Lincoln Medical And Mental Health Center | New York City Public Health Laboratory | Jade Wang, et al. |
| EPI_ISL_632073 | OCME Office Of Chief Medical Examiner | New York City Public Health Laboratory | Jade Wang, et al. |
| EPI_ISL_632074 | NYC Department Of Health And Mental Hygiene | New York City Public Health Laboratory | Jade Wang, et al. |
| EPI_ISL_632075 | Jamaica Hospital Medical Center | New York City Public Health Laboratory | Jade Wang, et al. |
| EPI_ISL_632076 | NYU Langone Health | New York City Public Health Laboratory | Jade Wang, et al. |
| EPI_ISL_632077, EPI_ISL_632078 | Brookdale University Hospital Medical Center | New York City Public Health Laboratory | Jade Wang, et al. |
| EPI_ISL_632079 | NYU Langone Health | New York City Public Health Laboratory | Jade Wang, et al. |
| EPI_ISL_632080 | SUNY Downstate Medical Center | New York City Public Health Laboratory | Jade Wang, et al. |
| EPI_ISL_632081 | New York Presbyterian-Brooklyn Methodist Hospital | New York City Public Health Laboratory | Jade Wang, et al. |
| EPI_ISL_632082 | New York Presbyterian/ Weill Cornell Medical Center | New York City Public Health Laboratory | Jade Wang, et al. |
| EPI_ISL_632083, EPI_ISL_632084 | NYC Department Of Health And Mental Hygiene | New York City Public Health Laboratory | Jade Wang, et al. |
| EPI_ISL_632085 | Richmond University Medical Center | New York City Public Health Laboratory | Jade Wang, et al. |
| EPI_ISL_632086 | Jamaica Hospital Medical Center | New York City Public Health Laboratory | Jade Wang, et al. |
| EPI_ISL_632087 | Flushing Hospital Medical Center | New York City Public Health Laboratory | Jade Wang, et al. |
| EPI_ISL_632088 | Jamaica Hospital Medical Center | New York City Public Health Laboratory | Jade Wang, et al. |
| EPI_ISL_632089 | Brookdale University Hospital Medical Center | New York City Public Health Laboratory | Jade Wang, et al. |
| EPI_ISL_632090 | Jamaica Hospital Medical Center | New York City Public Health Laboratory | Jade Wang, et al. |
| EPI_ISL_632091, EPI_ISL_632092 | SUNY Downstate Medical Center | New York City Public Health Laboratory | Jade Wang, et al. |
| EPI_ISL_632093, EPI_ISL_632094, EPI_ISL_632095, EPI_ISL_632096 | Jamaica Hospital Medical Center | New York City Public Health Laboratory | Jade Wang, et al. |
| EPI_ISL_632097 | Flushing Hospital Medical Center | New York City Public Health Laboratory | Jade Wang, et al. |
| EPI_ISL_632098 | Bellevue Hospital Center | New York City Public Health Laboratory | Jade Wang, et al. |
| EPI_ISL_632099 | VA NY Harbor Healthcare System | New York City Public Health Laboratory | Jade Wang, et al. |
| EPI_ISL_632100 | Brookdale University Hospital Medical Center | New York City Public Health Laboratory | Jade Wang, et al. |
| EPI_ISL_632101, EPI_ISL_632102 | Mount Sinai Queens | New York City Public Health Laboratory | Jade Wang, et al. |
| EPI_ISL_632103, EPI_ISL_632104, EPI_ISL_632105 | Flushing Hospital Medical Center | New York City Public Health Laboratory | Jade Wang, et al. |
| EPI_ISL_632106, EPI_ISL_632107, EPI_ISL_632108, EPI_ISL_632109, EPI_ISL_632110 | Jamaica Hospital Medical Center | New York City Public Health Laboratory | Jade Wang, et al. |
| EPI_ISL_632111 | Montefiore Medical Center | New York City Public Health Laboratory | Jade Wang, et al. |
| EPI_ISL_632112, EPI_ISL_632113 | Richmond University Medical Center | New York City Public Health Laboratory | Jade Wang, et al. |
| EPI_ISL_632114 | Brookdale University Hospital Medical Center | New York City Public Health Laboratory | Jade Wang, et al. |
| EPI_ISL_632115 | Richmond University Medical Center | New York City Public Health Laboratory | Jade Wang, et al. |
| EPI_ISL_632116 | NYC HH Lincoln Medical And Mental Health Center | New York City Public Health Laboratory | Jade Wang, et al. |
| EPI_ISL_632117, EPI_ISL_632118 | Richmond University Medical Center | New York City Public Health Laboratory | Jade Wang, et al. |
| EPI_ISL_632119, EPI_ISL_632120 | Flushing Hospital Medical Center | New York City Public Health Laboratory | Jade Wang, et al. |
| EPI_ISL_632121 | Bellevue Hospital Center | New York City Public Health Laboratory | Jade Wang, et al. |
| EPI_ISL_632122 | Flushing Hospital Medical Center | New York City Public Health Laboratory | Jade Wang, et al. |
| EPI_ISL_632123 | Brookdale University Hospital Medical Center | New York City Public Health Laboratory | Jade Wang, et al. |
| EPI_ISL_632124, EPI_ISL_632125, EPI_ISL_632126, EPI_ISL_632127 | Flushing Hospital Medical Center | New York City Public Health Laboratory | Jade Wang, et al. |
| EPI_ISL_632128 | Jamaica Hospital Medical Center | New York City Public Health Laboratory | Jade Wang, et al. |
| EPI_ISL_632129 | Wyckoff Heights Medical Center | New York City Public Health Laboratory | Jade Wang, et al. |
| EPI_ISL_632130 | Jamaica Hospital Medical Center | New York City Public Health Laboratory | Jade Wang, et al. |
| EPI_ISL_632131 | NYC HH Lincoln Medical And Mental Health Center | New York City Public Health Laboratory | Jade Wang, et al. |
| EPI_ISL_632132, EPI_ISL_632133, EPI_ISL_632134 | Flushing Hospital Medical Center | New York City Public Health Laboratory | Jade Wang, et al. |
| EPI_ISL_632135 | SUNY Downstate Medical Center | New York City Public Health Laboratory | Jade Wang, et al. |
| EPI_ISL_632136 | Bellevue Hospital Center | New York City Public Health Laboratory | Jade Wang, et al. |
| EPI_ISL_632137, EPI_ISL_632138, EPI_ISL_632139, EPI_ISL_632140 | Flushing Hospital Medical Center | New York City Public Health Laboratory | Jade Wang, et al. |
| EPI_ISL_632141 | Wyckoff Heights Medical Center | New York City Public Health Laboratory | Jade Wang, et al. |
| EPI_ISL_632142 | Brookdale University Hospital Medical Center | New York City Public Health Laboratory | Jade Wang, et al. |
| EPI_ISL_632143 | Jamaica Hospital Medical Center | New York City Public Health Laboratory | Jade Wang, et al. |
| EPI_ISL_632144 | Flushing Hospital Medical Center | New York City Public Health Laboratory | Jade Wang, et al. |
| EPI_ISL_632145 | Jamaica Hospital Medical Center | New York City Public Health Laboratory | Jade Wang, et al. |
| EPI_ISL_632146, EPI_ISL_632147 | NYC HH Lincoln Medical And Mental Health Center | New York City Public Health Laboratory | Jade Wang, et al. |

| | | | |
|---|---|---|---|
| EPI_ISL_632148 | SUNY Downstate Medical Center | New York City Public Health Laboratory | Jade Wang, et al. |
| EPI_ISL_632149, EPI_ISL_632150, EPI_ISL_632151, EPI_ISL_632152 | Jamaica Hospital Medical Center | New York City Public Health Laboratory | Jade Wang, et al. |
| EPI_ISL_632153 | SUNY Downstate Medical Center | New York City Public Health Laboratory | Jade Wang, et al. |
| EPI_ISL_632154 | Richmond University Medical Center | New York City Public Health Laboratory | Jade Wang, et al. |
| EPI_ISL_632155, EPI_ISL_632156 | Brookdale University Hospital Medical Center | New York City Public Health Laboratory | Jade Wang, et al. |
| EPI_ISL_632157 | Wyckoff Heights Medical Center | New York City Public Health Laboratory | Jade Wang, et al. |
| EPI_ISL_632158 | SUNY Downstate Medical Center | New York City Public Health Laboratory | Jade Wang, et al. |
| EPI_ISL_632159 | Wyckoff Heights Medical Center | New York City Public Health Laboratory | Jade Wang, et al. |
| EPI_ISL_632160, EPI_ISL_632161 | SUNY Downstate Medical Center | New York City Public Health Laboratory | Jade Wang, et al. |
| EPI_ISL_632162 | Jamaica Hospital Medical Center | New York City Public Health Laboratory | Jade Wang, et al. |
| EPI_ISL_632163, EPI_ISL_632164, EPI_ISL_632165 | Richmond University Medical Center | New York City Public Health Laboratory | Jade Wang, et al. |
| EPI_ISL_632166, EPI_ISL_632167, EPI_ISL_632168, EPI_ISL_632169, EPI_ISL_632170, EPI_ISL_632171, EPI_ISL_632172 | Jamaica Hospital Medical Center | New York City Public Health Laboratory | Jade Wang, et al. |
| EPI_ISL_632173 | Richmond University Medical Center | New York City Public Health Laboratory | Jade Wang, et al. |
| EPI_ISL_632174 | Flushing Hospital Medical Center | New York City Public Health Laboratory | Jade Wang, et al. |
| EPI_ISL_632175, EPI_ISL_632176 | OCME Office Of Chief Medical Examiner | New York City Public Health Laboratory | Jade Wang, et al. |
| EPI_ISL_632177 | NYC HH Lincoln Medical And Mental Health Center | New York City Public Health Laboratory | Jade Wang, et al. |
| EPI_ISL_632178 | OCME Office Of Chief Medical Examiner | New York City Public Health Laboratory | Jade Wang, et al. |
| EPI_ISL_632179 | James J. Peters VA Medical Center | New York City Public Health Laboratory | Jade Wang, et al. |
| EPI_ISL_632180 | Jamaica Hospital Medical Center | New York City Public Health Laboratory | Jade Wang, et al. |
| EPI_ISL_632181 | Flushing Hospital Medical Center | New York City Public Health Laboratory | Jade Wang, et al. |
| EPI_ISL_632182 | Jamaica Hospital Medical Center | New York City Public Health Laboratory | Jade Wang, et al. |
| EPI_ISL_632183 | OCME Office Of Chief Medical Examiner | New York City Public Health Laboratory | Jade Wang, et al. |
| EPI_ISL_632184 | Jamaica Hospital Medical Center | New York City Public Health Laboratory | Jade Wang, et al. |
| EPI_ISL_632185 | Flushing Hospital Medical Center | New York City Public Health Laboratory | Jade Wang, et al. |
| EPI_ISL_632186, EPI_ISL_632187 | NYC HH Lincoln Medical And Mental Health Center | New York City Public Health Laboratory | Jade Wang, et al. |
| EPI_ISL_632188, EPI_ISL_632189 | Jamaica Hospital Medical Center | New York City Public Health Laboratory | Jade Wang, et al. |
| EPI_ISL_632190 | Flushing Hospital Medical Center | New York City Public Health Laboratory | Jade Wang, et al. |
| EPI_ISL_632191 | Jamaica Hospital Medical Center | New York City Public Health Laboratory | Jade Wang, et al. |
| EPI_ISL_632192, EPI_ISL_632193 | NYC HH Lincoln Medical And Mental Health Center | New York City Public Health Laboratory | Jade Wang, et al. |
| EPI_ISL_632194 | Flushing Hospital Medical Center | New York City Public Health Laboratory | Jade Wang, et al. |
| EPI_ISL_632195 | Jamaica Hospital Medical Center | New York City Public Health Laboratory | Jade Wang, et al. |
| EPI_ISL_632196, EPI_ISL_632197 | Flushing Hospital Medical Center | New York City Public Health Laboratory | Jade Wang, et al. |
| EPI_ISL_632198 | Jamaica Hospital Medical Center | New York City Public Health Laboratory | Jade Wang, et al. |
| EPI_ISL_632199 | Richmond University Medical Center | New York City Public Health Laboratory | Jade Wang, et al. |
| EPI_ISL_632200 | James J. Peters VA Medical Center | New York City Public Health Laboratory | Jade Wang, et al. |
| EPI_ISL_632201 | Richmond University Medical Center | New York City Public Health Laboratory | Jade Wang, et al. |
| EPI_ISL_632202 | OCME Office Of Chief Medical Examiner | New York City Public Health Laboratory | Jade Wang, et al. |
| EPI_ISL_632203 | Richmond University Medical Center | New York City Public Health Laboratory | Jade Wang, et al. |
| EPI_ISL_632204 | Jamaica Hospital Medical Center | New York City Public Health Laboratory | Jade Wang, et al. |
| EPI_ISL_632205, EPI_ISL_632206, EPI_ISL_632207, EPI_ISL_632208 | Richmond University Medical Center | New York City Public Health Laboratory | Jade Wang, et al. |
| EPI_ISL_632209, EPI_ISL_632210 | Jamaica Hospital Medical Center | New York City Public Health Laboratory | Jade Wang, et al. |
| EPI_ISL_632211, EPI_ISL_632212, EPI_ISL_632213 | Flushing Hospital Medical Center | New York City Public Health Laboratory | Jade Wang, et al. |
| EPI_ISL_632214 | Jamaica Hospital Medical Center | New York City Public Health Laboratory | Jade Wang, et al. |
| EPI_ISL_632215, EPI_ISL_632216, EPI_ISL_632217, EPI_ISL_632218 | Richmond University Medical Center | New York City Public Health Laboratory | Jade Wang, et al. |
| EPI_ISL_632219 | Jamaica Hospital Medical Center | New York City Public Health Laboratory | Jade Wang, et al. |
| EPI_ISL_632220 | Flushing Hospital Medical Center | New York City Public Health Laboratory | Jade Wang, et al. |
| EPI_ISL_632221 | Jamaica Hospital Medical Center | New York City Public Health Laboratory | Jade Wang, et al. |
| EPI_ISL_632222, EPI_ISL_632223, EPI_ISL_632224, EPI_ISL_632225, EPI_ISL_632226 | Flushing Hospital Medical Center | New York City Public Health Laboratory | Jade Wang, et al. |
| EPI_ISL_632227, EPI_ISL_632228, EPI_ISL_632229 | Jamaica Hospital Medical Center | New York City Public Health Laboratory | Jade Wang, et al. |
| EPI_ISL_632230, EPI_ISL_632231, EPI_ISL_632232, EPI_ISL_632233, EPI_ISL_632234 | Richmond University Medical Center | New York City Public Health Laboratory | Jade Wang, et al. |

| | | | |
|---|---|---|---|
| EPI_ISL_632235 | NYC HH Lincoln Medical And Mental Health Center | New York City Public Health Laboratory | Jade Wang, et al. |
| EPI_ISL_632236, EPI_ISL_632237 | Flushing Hospital Medical Center | New York City Public Health Laboratory | Jade Wang, et al. |
| EPI_ISL_632238 | OCME Office Of Chief Medical Examiner | New York City Public Health Laboratory | Jade Wang, et al. |
| EPI_ISL_632239, EPI_ISL_632240, EPI_ISL_632241, EPI_ISL_632939 | Jamaica Hospital Medical Center | New York City Public Health Laboratory | Jade Wang, et al. |
| EPI_ISL_632940 | Richmond University Medical Center | New York City Public Health Laboratory | Jade Wang, et al. |
| EPI_ISL_632941 | Jamaica Hospital Medical Center | New York City Public Health Laboratory | Jade Wang, et al. |
| EPI_ISL_632942, EPI_ISL_632943 | Richmond University Medical Center | New York City Public Health Laboratory | Jade Wang, et al. |
| EPI_ISL_632944 | Jamaica Hospital Medical Center | New York City Public Health Laboratory | Jade Wang, et al. |
| EPI_ISL_632945, EPI_ISL_632946 | OCME Office Of Chief Medical Examiner | New York City Public Health Laboratory | Jade Wang, et al. |
| EPI_ISL_632947 | Flushing Hospital Medical Center | New York City Public Health Laboratory | Jade Wang, et al. |
| EPI_ISL_632948 | Richmond University Medical Center | New York City Public Health Laboratory | Jade Wang, et al. |
| EPI_ISL_632949, EPI_ISL_632950 | Jamaica Hospital Medical Center | New York City Public Health Laboratory | Jade Wang, et al. |
| EPI_ISL_632951, EPI_ISL_632952 | Flushing Hospital Medical Center | New York City Public Health Laboratory | Jade Wang, et al. |
| EPI_ISL_632953 | Jamaica Hospital Medical Center | New York City Public Health Laboratory | Jade Wang, et al. |
| EPI_ISL_632954, EPI_ISL_632955, EPI_ISL_632956 | Flushing Hospital Medical Center | New York City Public Health Laboratory | Jade Wang, et al. |
| EPI_ISL_632957, EPI_ISL_632958 | Jamaica Hospital Medical Center | New York City Public Health Laboratory | Jade Wang, et al. |
| EPI_ISL_632959 | Richmond University Medical Center | New York City Public Health Laboratory | Jade Wang, et al. |
| EPI_ISL_632960 | Flushing Hospital Medical Center | New York City Public Health Laboratory | Jade Wang, et al. |
| EPI_ISL_632961 | Jamaica Hospital Medical Center | New York City Public Health Laboratory | Jade Wang, et al. |
| EPI_ISL_632962 | Richmond University Medical Center | New York City Public Health Laboratory | Jade Wang, et al. |
| EPI_ISL_632963 | NYC Department Of Health And Mental Hygiene | New York City Public Health Laboratory | Jade Wang, et al. |
| EPI_ISL_632964 | Department of Homeless Services | New York City Public Health Laboratory | Jade Wang, et al. |
| EPI_ISL_632965 | Richmond University Medical Center | New York City Public Health Laboratory | Jade Wang, et al. |
| EPI_ISL_632966, EPI_ISL_632967, EPI_ISL_632968 | Flushing Hospital Medical Center | New York City Public Health Laboratory | Jade Wang, et al. |
| EPI_ISL_632969, EPI_ISL_632970, EPI_ISL_632971, EPI_ISL_632972 | Jamaica Hospital Medical Center | New York City Public Health Laboratory | Jade Wang, et al. |
| EPI_ISL_632973 | Flushing Hospital Medical Center | New York City Public Health Laboratory | Jade Wang, et al. |
| EPI_ISL_632974 | Jamaica Hospital Medical Center | New York City Public Health Laboratory | Jade Wang, et al. |
| EPI_ISL_632975 | Richmond University Medical Center | New York City Public Health Laboratory | Jade Wang, et al. |
| EPI_ISL_632976, EPI_ISL_632977, EPI_ISL_632978, EPI_ISL_632979 | Empire City Laboratories | New York City Public Health Laboratory | Jade Wang, et al. |
| EPI_ISL_632980, EPI_ISL_632981, EPI_ISL_632982, EPI_ISL_632983, EPI_ISL_632984, EPI_ISL_632985, EPI_ISL_632986, EPI_ISL_632987, EPI_ISL_632988, EPI_ISL_632989, EPI_ISL_632990, EPI_ISL_632991, EPI_ISL_632992, EPI_ISL_632993 | | | |
| see above | DOHMH Corona | New York City Public Health Laboratory | Jade Wang, et al. |
| EPI_ISL_632994, EPI_ISL_632995, EPI_ISL_632996, EPI_ISL_632997, EPI_ISL_632998 | DOHMH Crown Heights | New York City Public Health Laboratory | Jade Wang, et al. |
| EPI_ISL_632999, EPI_ISL_633000, EPI_ISL_633001, EPI_ISL_633002, EPI_ISL_633003, EPI_ISL_633004, EPI_ISL_633005, EPI_ISL_633006, EPI_ISL_633007, EPI_ISL_633008 | DOHMH Jamaica | New York City Public Health Laboratory | Jade Wang, et al. |
| EPI_ISL_633009, EPI_ISL_633010 | DOHMH Central Harlem | New York City Public Health Laboratory | Jade Wang, et al. |
| EPI_ISL_633011 | DOHMH PHL | New York City Public Health Laboratory | Jade Wang, et al. |
| EPI_ISL_633012 | DOHMH Riverside | New York City Public Health Laboratory | Jade Wang, et al. |
| EPI_ISL_633013 | DOHMH PHL | New York City Public Health Laboratory | Jade Wang, et al. |
| EPI_ISL_633014 | DOHMH Chelsea | New York City Public Health Laboratory | Jade Wang, et al. |
| EPI_ISL_633015 | DOHMH Morrisania | New York City Public Health Laboratory | Jade Wang, et al. |
| EPI_ISL_633016, EPI_ISL_633017 | DOHMH Chelsea | New York City Public Health Laboratory | Jade Wang, et al. |
| EPI_ISL_633018 | DOHMH Central Harlem | New York City Public Health Laboratory | Jade Wang, et al. |
| EPI_ISL_633019, EPI_ISL_633020, EPI_ISL_633021, EPI_ISL_633022 | DOHMH Morrisania | New York City Public Health Laboratory | Jade Wang, et al. |
| EPI_ISL_633023 | DOHMH Chelsea | New York City Public Health Laboratory | Jade Wang, et al. |
| EPI_ISL_633024 | DOHMH Morrisania | New York City Public Health Laboratory | Jade Wang, et al. |
| EPI_ISL_633025 | DOHMH Chelsea | New York City Public Health Laboratory | Jade Wang, et al. |
| EPI_ISL_633026 | DOHMH PHL | New York City Public Health Laboratory | Jade Wang, et al. |
| EPI_ISL_633027, EPI_ISL_633028, EPI_ISL_633029, EPI_ISL_633030 | DOHMH Morrisania | New York City Public Health Laboratory | Jade Wang, et al. |
| EPI_ISL_633031 | DOHMH Chelsea | New York City Public Health Laboratory | Jade Wang, et al. |
| EPI_ISL_633032 | DOHMH Morrisania | New York City Public Health Laboratory | Jade Wang, et al. |
| EPI_ISL_633033 | DOHMH Chelsea | New York City Public Health Laboratory | Jade Wang, et al. |

| | | | |
|---|---|---|---|
| EPI_ISL_633034 | DOHMH Riverside | New York City Public Health Laboratory | Jade Wang, et al. |
| EPI_ISL_633035 | DOHMH PHL | New York City Public Health Laboratory | Jade Wang, et al. |
| EPI_ISL_633036 | DOHMH Morrisania | New York City Public Health Laboratory | Jade Wang, et al. |
| EPI_ISL_633037 | DOHMH Central Harlem | New York City Public Health Laboratory | Jade Wang, et al. |
| EPI_ISL_633038 | DOHMH PHL | New York City Public Health Laboratory | Jade Wang, et al. |
| EPI_ISL_633039, EPI_ISL_633040 | DOHMH Chelsea | New York City Public Health Laboratory | Jade Wang, et al. |
| EPI_ISL_633041, EPI_ISL_633042, EPI_ISL_633043 | DOHMH Riverside | New York City Public Health Laboratory | Jade Wang, et al. |
| EPI_ISL_633044 | DOHMH Morrisania | New York City Public Health Laboratory | Jade Wang, et al. |
| EPI_ISL_633045 | DOHMH PHL | New York City Public Health Laboratory | Jade Wang, et al. |
| EPI_ISL_633046, EPI_ISL_633047 | DOHMH Chelsea | New York City Public Health Laboratory | Jade Wang, et al. |
| EPI_ISL_633048, EPI_ISL_633049 | DOHMH Morrisania | New York City Public Health Laboratory | Jade Wang, et al. |
| EPI_ISL_633050, EPI_ISL_633051 | DOHMH PHL | New York City Public Health Laboratory | Jade Wang, et al. |
| EPI_ISL_633052 | DOHMH Chelsea | New York City Public Health Laboratory | Jade Wang, et al. |
| EPI_ISL_633053 | DOHMH Morrisania | New York City Public Health Laboratory | Jade Wang, et al. |
| EPI_ISL_633054, EPI_ISL_633055 | DOHMH PHL | New York City Public Health Laboratory | Jade Wang, et al. |
| EPI_ISL_633056 | DOHMH Central Harlem | New York City Public Health Laboratory | Jade Wang, et al. |
| EPI_ISL_633057, EPI_ISL_633058 | DOHMH Morrisania | New York City Public Health Laboratory | Jade Wang, et al. |
| EPI_ISL_633059 | DOHMH Corona | New York City Public Health Laboratory | Jade Wang, et al. |
| EPI_ISL_633060, EPI_ISL_633061, EPI_ISL_633062 | DOHMH Jamaica | New York City Public Health Laboratory | Jade Wang, et al. |
| EPI_ISL_633063 | DOHMH Crown Heights | New York City Public Health Laboratory | Jade Wang, et al. |
| EPI_ISL_633064 | DOHMH Jamaica | New York City Public Health Laboratory | Jade Wang, et al. |
| EPI_ISL_633065 | DOHMH Morrisania | New York City Public Health Laboratory | Jade Wang, et al. |
| EPI_ISL_633066 | DOHMH Chelsea | New York City Public Health Laboratory | Jade Wang, et al. |
| EPI_ISL_633067, EPI_ISL_633068 | DOHMH Morrisania | New York City Public Health Laboratory | Jade Wang, et al. |
| EPI_ISL_633069 | DOHMH Chelsea | New York City Public Health Laboratory | Jade Wang, et al. |
| EPI_ISL_633070 | Flushing Hospital Medical Center | New York City Public Health Laboratory | Jade Wang, et al. |
| EPI_ISL_633071 | DOHMH PHL | New York City Public Health Laboratory | Jade Wang, et al. |
| EPI_ISL_633072 | DOHMH Riverside | New York City Public Health Laboratory | Jade Wang, et al. |
| EPI_ISL_633073 | DOHMH Fort Greene | New York City Public Health Laboratory | Jade Wang, et al. |
| EPI_ISL_633074 | DOHMH Corona | New York City Public Health Laboratory | Jade Wang, et al. |
| EPI_ISL_633075 | DOHMH Morrisania | New York City Public Health Laboratory | Jade Wang, et al. |
| EPI_ISL_633076, EPI_ISL_633077, EPI_ISL_633078 | DOHMH Chelsea | New York City Public Health Laboratory | Jade Wang, et al. |
| EPI_ISL_633079 | DOHMH PHL | New York City Public Health Laboratory | Jade Wang, et al. |
| EPI_ISL_633080, EPI_ISL_633081, EPI_ISL_633082 | DOHMH Morrisania | New York City Public Health Laboratory | Jade Wang, et al. |
| EPI_ISL_641311 | Respiratory virus Laboratory, Chinese Academy of Medical Science | Respiratory virus Laboratory, Chinese Academy of Medical Science | Li,J., Zhen,H., Chen,Y. and Liu,L. |
| EPI_ISL_661254, EPI_ISL_666964 | Quest Diagnostics | Quest Diagnostics | Rosenthal,S.H., Gerasimova,A., Kagan,R.M., Anderson, B., Owen, R., Lacbawan, F. |
| EPI_ISL_671674 | DOHMH Corona | New York City Public Health Laboratory | Jade Wang, et al. |
| EPI_ISL_671675 | DOHMH Jamaica | New York City Public Health Laboratory | Jade Wang, et al. |
| EPI_ISL_671676 | DOHMH PHL | New York City Public Health Laboratory | Jade Wang, et al. |
| EPI_ISL_671677 | DOHMH Corona | New York City Public Health Laboratory | Jade Wang, et al. |
| EPI_ISL_671678 | DOHMH Fort Greene | New York City Public Health Laboratory | Jade Wang, et al. |
| EPI_ISL_671679, EPI_ISL_671680 | DOHMH Morrisania | New York City Public Health Laboratory | Jade Wang, et al. |
| EPI_ISL_671681, EPI_ISL_671682, EPI_ISL_671683 | DOHMH Corona | New York City Public Health Laboratory | Jade Wang, et al. |
| EPI_ISL_671684 | DOHMH Chelsea | New York City Public Health Laboratory | Jade Wang, et al. |
| EPI_ISL_671685 | DOHMH Corona | New York City Public Health Laboratory | Jade Wang, et al. |
| EPI_ISL_671686, EPI_ISL_671687, EPI_ISL_671688 | DOHMH Morrisania | New York City Public Health Laboratory | Jade Wang, et al. |
| EPI_ISL_671689 | DOHMH Central Harlem | New York City Public Health Laboratory | Jade Wang, et al. |
| EPI_ISL_671690 | DOHMH Crown Heights | New York City Public Health Laboratory | Jade Wang, et al. |
| EPI_ISL_671691 | DOHMH Corona | New York City Public Health Laboratory | Jade Wang, et al. |
| EPI_ISL_671692 | DOHMH Jamaica | New York City Public Health Laboratory | Jade Wang, et al. |
| EPI_ISL_671693 | DOHMH Chelsea | New York City Public Health Laboratory | Jade Wang, et al. |
| EPI_ISL_671694 | DOHMH Riverside | New York City Public Health Laboratory | Jade Wang, et al. |
| EPI_ISL_671695 | DOHMH Corona | New York City Public Health Laboratory | Jade Wang, et al. |
| EPI_ISL_671696 | DOHMH PHL | New York City Public Health Laboratory | Jade Wang, et al. |

| | | | |
|---|---|---|---|
| EPI_ISL_671697 | DOHMH Corona | New York City Public Health Laboratory | Jade Wang, et al. |
| EPI_ISL_671698 | DOHMH Jamaica | New York City Public Health Laboratory | Jade Wang, et al. |
| EPI_ISL_671699, EPI_ISL_671700 | DOHMH Corona | New York City Public Health Laboratory | Jade Wang, et al. |
| EPI_ISL_671701 | DOHMH Riverside | New York City Public Health Laboratory | Jade Wang, et al. |
| EPI_ISL_671702 | DOHMH Chelsea | New York City Public Health Laboratory | Jade Wang, et al. |
| EPI_ISL_671703 | DOHMH Corona | New York City Public Health Laboratory | Jade Wang, et al. |
| EPI_ISL_671704 | DOHMH Central Harlem | New York City Public Health Laboratory | Jade Wang, et al. |
| EPI_ISL_671705 | DOHMH Corona | New York City Public Health Laboratory | Jade Wang, et al. |
| EPI_ISL_671706, EPI_ISL_671707 | DOHMH Jamaica | New York City Public Health Laboratory | Jade Wang, et al. |
| EPI_ISL_671708 | DOHMH Morrisania | New York City Public Health Laboratory | Jade Wang, et al. |
| EPI_ISL_671709 | DOHMH Riverside | New York City Public Health Laboratory | Jade Wang, et al. |
| EPI_ISL_671710, EPI_ISL_671711 | DOHMH Corona | New York City Public Health Laboratory | Jade Wang, et al. |
| EPI_ISL_671712 | DOHMH Jamaica | New York City Public Health Laboratory | Jade Wang, et al. |
| EPI_ISL_671713 | DOHMH Chelsea | New York City Public Health Laboratory | Jade Wang, et al. |
| EPI_ISL_671714 | DOHMH Jamaica | New York City Public Health Laboratory | Jade Wang, et al. |
| EPI_ISL_671715, EPI_ISL_671716 | DOHMH PHL | New York City Public Health Laboratory | Jade Wang, et al. |
| EPI_ISL_671717, EPI_ISL_671718, EPI_ISL_671719, EPI_ISL_671720, EPI_ISL_671721 | DOHMH Corona | New York City Public Health Laboratory | Jade Wang, et al. |
| EPI_ISL_671722 | DOHMH Chelsea | New York City Public Health Laboratory | Jade Wang, et al. |
| EPI_ISL_671723, EPI_ISL_671724 | DOHMH Jamaica | New York City Public Health Laboratory | Jade Wang, et al. |
| EPI_ISL_671725 | DOHMH Riverside | New York City Public Health Laboratory | Jade Wang, et al. |
| EPI_ISL_671726, EPI_ISL_671727, EPI_ISL_671728, EPI_ISL_671729 | DOHMH Morrisania | New York City Public Health Laboratory | Jade Wang, et al. |
| EPI_ISL_671730 | DOHMH Jamaica | New York City Public Health Laboratory | Jade Wang, et al. |
| EPI_ISL_671731, EPI_ISL_671732 | DOHMH Corona | New York City Public Health Laboratory | Jade Wang, et al. |
| EPI_ISL_671733 | DOHMH Riverside | New York City Public Health Laboratory | Jade Wang, et al. |
| EPI_ISL_671734 | DOHMH PHL | New York City Public Health Laboratory | Jade Wang, et al. |
| EPI_ISL_671735 | DOHMH Jamaica | New York City Public Health Laboratory | Jade Wang, et al. |
| EPI_ISL_671736 | DOHMH Morrisania | New York City Public Health Laboratory | Jade Wang, et al. |
| EPI_ISL_671737 | DOHMH PHL | New York City Public Health Laboratory | Jade Wang, et al. |
| EPI_ISL_671738 | DOHMH Fort Greene | New York City Public Health Laboratory | Jade Wang, et al. |
| EPI_ISL_671739 | DOHMH Jamaica | New York City Public Health Laboratory | Jade Wang, et al. |
| EPI_ISL_671740, EPI_ISL_671741, EPI_ISL_671742 | DOHMH Morrisania | New York City Public Health Laboratory | Jade Wang, et al. |
| EPI_ISL_671743 | DOHMH Jamaica | New York City Public Health Laboratory | Jade Wang, et al. |
| EPI_ISL_671744 | DOHMH PHL | New York City Public Health Laboratory | Jade Wang, et al. |
| EPI_ISL_671745 | DOHMH Riverside | New York City Public Health Laboratory | Jade Wang, et al. |
| EPI_ISL_671746, EPI_ISL_671747 | DOHMH Corona | New York City Public Health Laboratory | Jade Wang, et al. |
| EPI_ISL_671748, EPI_ISL_671749, EPI_ISL_671750 | DOHMH Riverside | New York City Public Health Laboratory | Jade Wang, et al. |
| EPI_ISL_671751, EPI_ISL_671752, EPI_ISL_671753, EPI_ISL_671754, EPI_ISL_671755, EPI_ISL_671756, EPI_ISL_671757, EPI_ISL_671758 | DOHMH Morrisania | New York City Public Health Laboratory | Jade Wang, et al. |
| EPI_ISL_671759 | DOHMH Fort Greene | New York City Public Health Laboratory | Jade Wang, et al. |
| EPI_ISL_671760 | DOHMH Crown Heights | New York City Public Health Laboratory | Jade Wang, et al. |
| EPI_ISL_671761 | DOHMH Corona | New York City Public Health Laboratory | Jade Wang, et al. |
| EPI_ISL_671762 | DOHMH Riverside | New York City Public Health Laboratory | Jade Wang, et al. |
| EPI_ISL_671763 | DOHMH Chelsea | New York City Public Health Laboratory | Jade Wang, et al. |
| EPI_ISL_676648, EPI_ISL_676649, EPI_ISL_676650 | Wadsworth Center, New York State Department of Health | Wadsworth Center, New York State Department of Health | Kirsten St. George, Daryl M. Lamson, Sara Griesemer, Jonathan Plitnick, Navjot Singh, Matthew D. Shudt, Erica Lasek-Nesselquist |
| EPI_ISL_676651 | Wadsworth Center, New York State Department.of Health | Wadsworth Center, New York State Department.of Health | Kirsten St. George, Daryl M. Lamson, Alexis Russel, Jonathan Plitnick, Navjot Singh, John Kelly, Sara Griesemer, Erasmus Schneider, Erica Lasek-Nesselquist |
| EPI_ISL_676652 | Masonic Medical Research Institute | Wadsworth Center, New York State Department.of Health | Nathan Tucker, Kirsten St. George, Daryl M. Lamson, Alexis Russel, Jonathan Plitnick, Navjot Singh, John Kelly, Sara Griesemer, Erasmus Schneider, Erica Lasek-Nesselquist |
| EPI_ISL_676653, EPI_ISL_676654, EPI_ISL_676655, EPI_ISL_676656, EPI_ISL_676657, EPI_ISL_676658 | Wadsworth Center, New York State Department.of Health | Wadsworth Center, New York State Department.of Health | Kirsten St. George, Daryl M. Lamson, Alexis Russel, Jonathan Plitnick, Navjot Singh, John Kelly, Sara Griesemer, Erasmus Schneider, Erica Lasek-Nesselquist |
| EPI_ISL_676659 | Masonic Medical Research Institute | Wadsworth Center, New York State Department.of Health | Nathan Tucker, Kirsten St. George, Daryl M. Lamson, Alexis Russel, Jonathan Plitnick, Navjot Singh, John Kelly, Sara Griesemer, Erasmus Schneider, Erica Lasek-Nesselquist |
| EPI_ISL_676660 | Wadsworth Center, New York State Department.of Health | Wadsworth Center, New York State Department.of Health | Kirsten St. George, Daryl M. Lamson, Alexis Russel, Jonathan Plitnick, Navjot Singh, John Kelly, Sara Griesemer, Erasmus Schneider, Erica Lasek-Nesselquist |
| EPI_ISL_676661 | Masonic Medical Research Institute | Wadsworth Center, New York State Department.of Health | Nathan Tucker, Kirsten St. George, Daryl M. Lamson, Alexis Russel, Jonathan Plitnick, Navjot Singh, John Kelly, Sara Griesemer, Erasmus Schneider, |

| | | | |
|---|---|---|---|
| | | | Erica Lasek-Nesselquist |
| EPI_ISL_676662 | Wadsworth Center, New York State Department.of Health | Wadsworth Center, New York State Department.of Health | Kirsten St. George, Daryl M. Lamson, Alexis Russel, Jonathan Plitnick, Navjot Singh, John Kelly, Sara Griesemer, Erasmus Schneider, Erica Lasek-Nesselquist |
| EPI_ISL_676663 | Masonic Medical Research Institute | Wadsworth Center, New York State Department.of Health | Nathan Tucker, Kirsten St. George, Daryl M. Lamson, Alexis Russel, Jonathan Plitnick, Navjot Singh, John Kelly, Sara Griesemer, Erasmus Schneider, Erica Lasek-Nesselquist |
| EPI_ISL_676664 | Wadsworth Center, New York State Department.of Health | Wadsworth Center, New York State Department.of Health | Kirsten St. George, Daryl M. Lamson, Alexis Russel, Jonathan Plitnick, Navjot Singh, John Kelly, Sara Griesemer, Erasmus Schneider, Erica Lasek-Nesselquist |
| EPI_ISL_676665 | Masonic Medical Research Institute | Wadsworth Center, New York State Department.of Health | Nathan Tucker, Kirsten St. George, Daryl M. Lamson, Alexis Russel, Jonathan Plitnick, Navjot Singh, John Kelly, Sara Griesemer, Erasmus Schneider, Erica Lasek-Nesselquist |
| EPI_ISL_676666, EPI_ISL_676667 | Wadsworth Center, New York State Department.of Health | Wadsworth Center, New York State Department.of Health | Kirsten St. George, Daryl M. Lamson, Alexis Russel, Jonathan Plitnick, Navjot Singh, John Kelly, Sara Griesemer, Erasmus Schneider, Erica Lasek-Nesselquist |
| EPI_ISL_676668 | Masonic Medical Research Institute | Wadsworth Center, New York State Department.of Health | Nathan Tucker, Kirsten St. George, Daryl M. Lamson, Alexis Russel, Jonathan Plitnick, Navjot Singh, John Kelly, Sara Griesemer, Erasmus Schneider, Erica Lasek-Nesselquist |
| EPI_ISL_676669, EPI_ISL_676670, EPI_ISL_676671, EPI_ISL_676672 | Wadsworth Center, New York State Department.of Health | Wadsworth Center, New York State Department.of Health | Kirsten St. George, Daryl M. Lamson, Alexis Russel, Jonathan Plitnick, Navjot Singh, John Kelly, Sara Griesemer, Erasmus Schneider, Erica Lasek-Nesselquist |
| EPI_ISL_676673 | Masonic Medical Research Institute | Wadsworth Center, New York State Department.of Health | Nathan Tucker, Kirsten St. George, Daryl M. Lamson, Alexis Russel, Jonathan Plitnick, Navjot Singh, John Kelly, Sara Griesemer, Erasmus Schneider, Erica Lasek-Nesselquist |
| EPI_ISL_676674, EPI_ISL_676675, EPI_ISL_676676, EPI_ISL_676677, EPI_ISL_676678, EPI_ISL_676679 | Wadsworth Center, New York State Department.of Health | Wadsworth Center, New York State Department.of Health | Kirsten St. George, Daryl M. Lamson, Alexis Russel, Jonathan Plitnick, Navjot Singh, John Kelly, Sara Griesemer, Erasmus Schneider, Erica Lasek-Nesselquist |
| EPI_ISL_676680 | Masonic Medical Research Institute | Wadsworth Center, New York State Department.of Health | Nathan Tucker, Kirsten St. George, Daryl M. Lamson, Alexis Russel, Jonathan Plitnick, Navjot Singh, John Kelly, Sara Griesemer, Erasmus Schneider, Erica Lasek-Nesselquist |
| EPI_ISL_676681, EPI_ISL_676682, EPI_ISL_676683, EPI_ISL_676684, EPI_ISL_676685, EPI_ISL_676686, EPI_ISL_676687, EPI_ISL_676688, EPI_ISL_676689, EPI_ISL_676690, EPI_ISL_676691, EPI_ISL_676692, EPI_ISL_676693, EPI_ISL_676694, EPI_ISL_676695, EPI_ISL_676696, EPI_ISL_676697, EPI_ISL_676698, EPI_ISL_676699, EPI_ISL_676700, EPI_ISL_676701, EPI_ISL_676702, EPI_ISL_676703, EPI_ISL_676704, EPI_ISL_676705, EPI_ISL_676706, EPI_ISL_676707, EPI_ISL_676708 | | | |
| see above | Wadsworth Center, New York State Department.of Health | Wadsworth Center, New York State Department.of Health | Kirsten St. George, Daryl M. Lamson, Alexis Russel, Jonathan Plitnick, Navjot Singh, John Kelly, Sara Griesemer, Erasmus Schneider, Erica Lasek-Nesselquist |
| EPI_ISL_676709 | Masonic Medical Research Institute | Wadsworth Center, New York State Department.of Health | Nathan Tucker, Kirsten St. George, Daryl M. Lamson, Alexis Russel, Jonathan Plitnick, Navjot Singh, John Kelly, Sara Griesemer, Erasmus Schneider, Erica Lasek-Nesselquist |
| EPI_ISL_676710, EPI_ISL_676711, EPI_ISL_676712 | Wadsworth Center, New York State Department.of Health | Wadsworth Center, New York State Department.of Health | Kirsten St. George, Daryl M. Lamson, Alexis Russel, Jonathan Plitnick, Navjot Singh, John Kelly, Sara Griesemer, Erasmus Schneider, Erica Lasek-Nesselquist |
| EPI_ISL_676713 | Masonic Medical Research Institute | Wadsworth Center, New York State Department.of Health | Nathan Tucker, Kirsten St. George, Daryl M. Lamson, Alexis Russel, Jonathan Plitnick, Navjot Singh, John Kelly, Sara Griesemer, Erasmus Schneider, Erica Lasek-Nesselquist |
| EPI_ISL_676714, EPI_ISL_676715, EPI_ISL_676716, EPI_ISL_676717, EPI_ISL_676718, EPI_ISL_676719, EPI_ISL_676720, EPI_ISL_676721, EPI_ISL_676722, EPI_ISL_676723, EPI_ISL_676724, EPI_ISL_676725 | | | |
| see above | Wadsworth Center, New York State Department.of Health | Wadsworth Center, New York State Department.of Health | Kirsten St. George, Daryl M. Lamson, Alexis Russel, Jonathan Plitnick, Navjot Singh, John Kelly, Sara Griesemer, Erasmus Schneider, Erica Lasek-Nesselquist |
| EPI_ISL_676726 | Masonic Medical Research Institute | Wadsworth Center, New York State Department.of Health | Nathan Tucker, Kirsten St. George, Daryl M. Lamson, Alexis Russel, Jonathan Plitnick, Navjot Singh, John Kelly, Sara Griesemer, Erasmus Schneider, Erica Lasek-Nesselquist |
| EPI_ISL_676727, EPI_ISL_676728, EPI_ISL_676729, EPI_ISL_676730, EPI_ISL_676731, EPI_ISL_676732, EPI_ISL_676733, EPI_ISL_676734, EPI_ISL_676735, EPI_ISL_676736, EPI_ISL_676737, EPI_ISL_676738, EPI_ISL_676739, EPI_ISL_676740, EPI_ISL_676741, EPI_ISL_676742, EPI_ISL_676743, EPI_ISL_676744, EPI_ISL_676745, EPI_ISL_676746, EPI_ISL_676747, EPI_ISL_676748, EPI_ISL_676749, EPI_ISL_676750, EPI_ISL_676751, EPI_ISL_676752, EPI_ISL_676753, EPI_ISL_676754, EPI_ISL_676755, EPI_ISL_676756, EPI_ISL_676757, EPI_ISL_676758, EPI_ISL_676759, EPI_ISL_676760, EPI_ISL_676761, EPI_ISL_676762, EPI_ISL_676763, EPI_ISL_676764, EPI_ISL_676765, EPI_ISL_676766, EPI_ISL_676767, EPI_ISL_676768, EPI_ISL_676769, EPI_ISL_676770, EPI_ISL_676771, EPI_ISL_676772, EPI_ISL_676773, EPI_ISL_676774, EPI_ISL_676775, EPI_ISL_676776, EPI_ISL_676777, EPI_ISL_676778, EPI_ISL_676779, EPI_ISL_676780, EPI_ISL_676781, EPI_ISL_676782, EPI_ISL_676783, EPI_ISL_676784, EPI_ISL_676785, EPI_ISL_676786, EPI_ISL_676787, EPI_ISL_676788, EPI_ISL_676789, EPI_ISL_676790, EPI_ISL_676791, EPI_ISL_676792, EPI_ISL_676793, EPI_ISL_676794, EPI_ISL_676795, EPI_ISL_676796, EPI_ISL_676797, EPI_ISL_676798, EPI_ISL_676799, EPI_ISL_676800, EPI_ISL_676801, EPI_ISL_676802, EPI_ISL_676803, EPI_ISL_676804, EPI_ISL_676805, EPI_ISL_676806, EPI_ISL_676807, EPI_ISL_676808, EPI_ISL_676809, EPI_ISL_676810, EPI_ISL_676811, EPI_ISL_676812, EPI_ISL_676813, EPI_ISL_676814, EPI_ISL_676815, EPI_ISL_676816, EPI_ISL_676817, EPI_ISL_676818, EPI_ISL_676819, EPI_ISL_676820, EPI_ISL_676821, EPI_ISL_676822, EPI_ISL_676823, EPI_ISL_676824, EPI_ISL_676825, EPI_ISL_676826, EPI_ISL_676827, EPI_ISL_676828, EPI_ISL_676829, EPI_ISL_676830, EPI_ISL_676831, EPI_ISL_676832, EPI_ISL_676833, EPI_ISL_676834, EPI_ISL_676835, EPI_ISL_676836, EPI_ISL_676837, EPI_ISL_676838, EPI_ISL_676839, EPI_ISL_676840, EPI_ISL_676841, EPI_ISL_676842, EPI_ISL_676843, EPI_ISL_676844, EPI_ISL_676845, EPI_ISL_676846, EPI_ISL_676847, EPI_ISL_676848, EPI_ISL_676849, EPI_ISL_676850, EPI_ISL_676851, EPI_ISL_676852, EPI_ISL_676853, EPI_ISL_676854, EPI_ISL_676855, EPI_ISL_676856, EPI_ISL_676857, EPI_ISL_676858, EPI_ISL_676859, EPI_ISL_676860, EPI_ISL_676861, EPI_ISL_676862, EPI_ISL_676863, EPI_ISL_676864, EPI_ISL_676865, EPI_ISL_676866, EPI_ISL_676867, EPI_ISL_676868, EPI_ISL_676869, EPI_ISL_676870, EPI_ISL_676871, EPI_ISL_676872, EPI_ISL_676873, EPI_ISL_676874, EPI_ISL_676875, EPI_ISL_676876, EPI_ISL_676877, EPI_ISL_676878, EPI_ISL_676879, EPI_ISL_676880, EPI_ISL_676881, EPI_ISL_676882, EPI_ISL_676883, EPI_ISL_676884, EPI_ISL_676885, EPI_ISL_676886, EPI_ISL_676887, EPI_ISL_676888, EPI_ISL_676889, EPI_ISL_676890, EPI_ISL_676891, EPI_ISL_676892, EPI_ISL_676893, EPI_ISL_676894, EPI_ISL_676895, EPI_ISL_676896, EPI_ISL_676897, EPI_ISL_676898, EPI_ISL_676899, EPI_ISL_676900, EPI_ISL_676901, EPI_ISL_676902, EPI_ISL_676903, EPI_ISL_676904, EPI_ISL_676905, EPI_ISL_676906, EPI_ISL_676907, EPI_ISL_676908, EPI_ISL_676909, EPI_ISL_676910, EPI_ISL_676911, EPI_ISL_676912, EPI_ISL_676913, EPI_ISL_676914, EPI_ISL_676915, EPI_ISL_676916, EPI_ISL_676917, EPI_ISL_676918, EPI_ISL_676919, EPI_ISL_676920, EPI_ISL_676921, EPI_ISL_676922, EPI_ISL_676923, EPI_ISL_676924, EPI_ISL_676925, EPI_ISL_676926, EPI_ISL_676927, EPI_ISL_676928, EPI_ISL_676929, EPI_ISL_676930, EPI_ISL_676931, EPI_ISL_676932, EPI_ISL_676933, EPI_ISL_676934, EPI_ISL_676935, EPI_ISL_676936, EPI_ISL_676937, EPI_ISL_676938, EPI_ISL_676939, EPI_ISL_676940, EPI_ISL_676941, EPI_ISL_676942, EPI_ISL_676943, EPI_ISL_676944, EPI_ISL_676945, EPI_ISL_676946, EPI_ISL_676947, EPI_ISL_676948, EPI_ISL_676949, EPI_ISL_676950, EPI_ISL_676951, EPI_ISL_676952, EPI_ISL_676953, EPI_ISL_676954, EPI_ISL_676955, EPI_ISL_676956, EPI_ISL_676957, EPI_ISL_676958, EPI_ISL_676959, EPI_ISL_676960, EPI_ISL_676961, EPI_ISL_676962, EPI_ISL_676963, EPI_ISL_676964, EPI_ISL_676965, EPI_ISL_676966, EPI_ISL_676967, EPI_ISL_676968, EPI_ISL_676969, EPI_ISL_676970, EPI_ISL_676971, EPI_ISL_676972, EPI_ISL_676973, EPI_ISL_676974, EPI_ISL_676975, EPI_ISL_676976, EPI_ISL_676977, EPI_ISL_676978, EPI_ISL_676979, EPI_ISL_676980, EPI_ISL_676981, EPI_ISL_676982, EPI_ISL_676983, EPI_ISL_676984, EPI_ISL_676985, EPI_ISL_676986, EPI_ISL_676987, EPI_ISL_676988, EPI_ISL_676989, EPI_ISL_676990, EPI_ISL_676991, EPI_ISL_676992, EPI_ISL_676993, EPI_ISL_676994 | | | |
| see above | Wadsworth Center, New York State Department.of Health | Wadsworth Center, New York State Department.of Health | Kirsten St. George, Daryl M. Lamson, Alexis Russel, Jonathan Plitnick, Navjot Singh, John Kelly, Sara Griesemer, Erasmus Schneider, Erica Lasek-Nesselquist |
| EPI_ISL_676995, EPI_ISL_676996, EPI_ISL_676997, EPI_ISL_676998, EPI_ISL_676999, EPI_ISL_677000, EPI_ISL_677001, EPI_ISL_677002, EPI_ISL_677003, EPI_ISL_677004, EPI_ISL_677005, EPI_ISL_677006, EPI_ISL_677007, EPI_ISL_677008, EPI_ISL_677009, EPI_ISL_677010, EPI_ISL_677011, EPI_ISL_677012, EPI_ISL_677013, EPI_ISL_677014, EPI_ISL_677015, EPI_ISL_677016, EPI_ISL_677017, EPI_ISL_677018, EPI_ISL_677019, EPI_ISL_677020, EPI_ISL_677021, EPI_ISL_677022, EPI_ISL_677023, EPI_ISL_677024, EPI_ISL_677025 | | | |
| see above | Masonic Medical Research Institute | Wadsworth Center, New York State Department.of Health | Nathan Tucker, Kirsten St. George, Daryl M. Lamson, Alexis Russel, Jonathan Plitnick, Navjot Singh, John Kelly, Sara Griesemer, Erasmus Schneider, Erica Lasek-Nesselquist |
| EPI_ISL_677026, EPI_ISL_677027, EPI_ISL_677028, EPI_ISL_677029, EPI_ISL_677030, EPI_ISL_677031, EPI_ISL_677032, EPI_ISL_677033, EPI_ISL_677034, EPI_ISL_677035, EPI_ISL_677036, EPI_ISL_677037, EPI_ISL_677038, EPI_ISL_677039, EPI_ISL_677040, EPI_ISL_677041, EPI_ISL_677042, EPI_ISL_677043, EPI_ISL_677044, EPI_ISL_677045, EPI_ISL_677046, EPI_ISL_677047, EPI_ISL_677048 | | | |
| see above | Wadsworth Center, New York State Department.of Health | Wadsworth Center, New York State Department.of Health | Kirsten St. George, Daryl M. Lamson, Alexis Russel, Jonathan Plitnick, Navjot Singh, John Kelly, Sara Griesemer, Erasmus Schneider, Erica Lasek-Nesselquist |
| EPI_ISL_677049, EPI_ISL_677050, EPI_ISL_677051, EPI_ISL_677052, EPI_ISL_677053, EPI_ISL_677054, EPI_ISL_677055, EPI_ISL_677056, EPI_ISL_677057, EPI_ISL_677058, EPI_ISL_677059, EPI_ISL_677060, EPI_ISL_677061, EPI_ISL_677062, EPI_ISL_677063, EPI_ISL_677064, EPI_ISL_677065, EPI_ISL_677066, EPI_ISL_677067, EPI_ISL_677068, EPI_ISL_677069, EPI_ISL_677070, EPI_ISL_677071, EPI_ISL_677072, EPI_ISL_677073, EPI_ISL_677074, EPI_ISL_677075, EPI_ISL_677076 | | | |
| see above | Masonic Medical Research Institute | Wadsworth Center, New York State Department.of Health | Nathan Tucker, Kirsten St. George, Daryl M. Lamson, Alexis Russel, Jonathan Plitnick, Navjot Singh, John Kelly, Sara Griesemer, Erasmus Schneider, Erica Lasek-Nesselquist |
| EPI_ISL_677077, EPI_ISL_677078, EPI_ISL_677079, EPI_ISL_677080, EPI_ISL_677081, EPI_ISL_677082, EPI_ISL_677083, EPI_ISL_677084, EPI_ISL_677085, EPI_ISL_677086, EPI_ISL_677087, EPI_ISL_677088, EPI_ISL_677089, EPI_ISL_677090, EPI_ISL_677091, EPI_ISL_677092, EPI_ISL_677093, EPI_ISL_677094, EPI_ISL_677095, EPI_ISL_677096, EPI_ISL_677097, EPI_ISL_677098, EPI_ISL_677099, EPI_ISL_677100, EPI_ISL_677101, EPI_ISL_677102, EPI_ISL_677103, EPI_ISL_677104, EPI_ISL_677105, EPI_ISL_677106, EPI_ISL_677107, EPI_ISL_677108, EPI_ISL_677109, EPI_ISL_677110 | | | |
| see above | Wadsworth Center, New York State Department.of Health | Wadsworth Center, New York State Department.of Health | Kirsten St. George, Daryl M. Lamson, Alexis Russel, Jonathan Plitnick, Navjot Singh, John Kelly, Sara Griesemer, Erasmus Schneider, Erica Lasek-Nesselquist |
| EPI_ISL_677111, EPI_ISL_677112, EPI_ISL_677113, EPI_ISL_677114, EPI_ISL_677115, EPI_ISL_677116, EPI_ISL_677117, EPI_ISL_677118, EPI_ISL_677119, EPI_ISL_677120, EPI_ISL_677121, EPI_ISL_677122, EPI_ISL_677123, EPI_ISL_677124, EPI_ISL_677125, EPI_ISL_677126, EPI_ISL_677127, EPI_ISL_677128, EPI_ISL_677129 | | | |
| see above | Masonic Medical Research Institute | Wadsworth Center, New York State Department.of Health | Nathan Tucker, Kirsten St. George, Daryl M. Lamson, Alexis Russel, Jonathan Plitnick, Navjot Singh, John Kelly, Sara Griesemer, Erasmus Schneider, |

| | | | |
|---|---|---|---|
| | | | Erica Lasek-Nesselquist |
| EPI_ISL_677130, EPI_ISL_677131, EPI_ISL_677132, EPI_ISL_677133, EPI_ISL_677664, EPI_ISL_677665, EPI_ISL_677666, EPI_ISL_677667, EPI_ISL_677668 | Wadsworth Center, New York State Department.of Health | Wadsworth Center, New York State Department.of Health | Kirsten St. George, Daryl M. Lamson, Alexis Russel, Jonathan Plitnick, Navjot Singh, John Kelly, Sara Griesemer, Erasmus Schneider, Erica Lasek-Nesselquist |
| EPI_ISL_677669 | Masonic Medical Research Institute | Wadsworth Center, New York State Department.of Health | Nathan Tucker, Kirsten St. George, Daryl M. Lamson, Alexis Russel, Jonathan Plitnick, Navjot Singh, John Kelly, Sara Griesemer, Erasmus Schneider, Erica Lasek-Nesselquist |
| EPI_ISL_677670, EPI_ISL_677671 | Wadsworth Center, New York State Department.of Health | Wadsworth Center, New York State Department.of Health | Kirsten St. George, Daryl M. Lamson, Alexis Russel, Jonathan Plitnick, Navjot Singh, John Kelly, Sara Griesemer, Erasmus Schneider, Erica Lasek-Nesselquist |
| EPI_ISL_677672 | Masonic Medical Research Institute | Wadsworth Center, New York State Department.of Health | Nathan Tucker, Kirsten St. George, Daryl M. Lamson, Alexis Russel, Jonathan Plitnick, Navjot Singh, John Kelly, Sara Griesemer, Erasmus Schneider, Erica Lasek-Nesselquist |
| EPI_ISL_677673 | Wadsworth Center, New York State Department.of Health | Wadsworth Center, New York State Department.of Health | Kirsten St. George, Daryl M. Lamson, Alexis Russel, Jonathan Plitnick, Navjot Singh, John Kelly, Sara Griesemer, Erasmus Schneider, Erica Lasek-Nesselquist |
| EPI_ISL_677678, EPI_ISL_677679, EPI_ISL_677680, EPI_ISL_677681, EPI_ISL_677682, EPI_ISL_677683, EPI_ISL_677684, EPI_ISL_677685, EPI_ISL_677686, EPI_ISL_677687, EPI_ISL_677688, EPI_ISL_677689, EPI_ISL_677692, EPI_ISL_677695, EPI_ISL_677696, EPI_ISL_677699 | | | |
| see above | Vanda Pharmaceuticals Clinical Site | Vanda Pharmaceuticals | Vanda Pharmaceuticals |
| EPI_ISL_683753 | DOHMH Corona | New York City Public Health Laboratory | Jade Wang, et al. |
| EPI_ISL_683754 | DOHMH Morrisania | New York City Public Health Laboratory | Jade Wang, et al. |
| EPI_ISL_683755 | DOHMH PHL | New York City Public Health Laboratory | Jade Wang, et al. |
| EPI_ISL_683756 | DOHMH Central Harlem | New York City Public Health Laboratory | Jade Wang, et al. |
| EPI_ISL_683757, EPI_ISL_683758 | DOHMH Corona | New York City Public Health Laboratory | Jade Wang, et al. |
| EPI_ISL_683759 | DOHMH PHL | New York City Public Health Laboratory | Jade Wang, et al. |
| EPI_ISL_683760 | DOHMH Morrisania | New York City Public Health Laboratory | Jade Wang, et al. |
| EPI_ISL_683761 | DOHMH Corona | New York City Public Health Laboratory | Jade Wang, et al. |
| EPI_ISL_683762, EPI_ISL_683763 | DOHMH Morrisania | New York City Public Health Laboratory | Jade Wang, et al. |
| EPI_ISL_683764 | DOHMH Corona | New York City Public Health Laboratory | Jade Wang, et al. |
| EPI_ISL_683765 | DOHMH Chelsea | New York City Public Health Laboratory | Jade Wang, et al. |
| EPI_ISL_683766 | DOHMH Corona | New York City Public Health Laboratory | Jade Wang, et al. |
| EPI_ISL_683767, EPI_ISL_683768, EPI_ISL_683769 | DOHMH Riverside | New York City Public Health Laboratory | Jade Wang, et al. |
| EPI_ISL_683770 | DOHMH Corona | New York City Public Health Laboratory | Jade Wang, et al. |
| EPI_ISL_683771 | DOHMH Fort Greene | New York City Public Health Laboratory | Jade Wang, et al. |
| EPI_ISL_683772 | DOHMH Morrisania | New York City Public Health Laboratory | Jade Wang, et al. |
| EPI_ISL_683773 | DOHMH Corona | New York City Public Health Laboratory | Jade Wang, et al. |
| EPI_ISL_683774 | DOHMH PHL | New York City Public Health Laboratory | Jade Wang, et al. |
| EPI_ISL_683775, EPI_ISL_683776 | DOHMH Jamaica | New York City Public Health Laboratory | Jade Wang, et al. |
| EPI_ISL_683777 | DOHMH Corona | New York City Public Health Laboratory | Jade Wang, et al. |
| EPI_ISL_683778 | DOHMH Morrisania | New York City Public Health Laboratory | Jade Wang, et al. |
| EPI_ISL_683779 | DOHMH Corona | New York City Public Health Laboratory | Jade Wang, et al. |
| EPI_ISL_683780 | DOHMH Chelsea | New York City Public Health Laboratory | Jade Wang, et al. |
| EPI_ISL_683781, EPI_ISL_683782 | DOHMH Jamaica | New York City Public Health Laboratory | Jade Wang, et al. |
| EPI_ISL_683783 | DOHMH PHL | New York City Public Health Laboratory | Jade Wang, et al. |
| EPI_ISL_683784 | DOHMH Riverside | New York City Public Health Laboratory | Jade Wang, et al. |
| EPI_ISL_683785 | DOHMH Corona | New York City Public Health Laboratory | Jade Wang, et al. |
| EPI_ISL_683786 | DOHMH Jamaica | New York City Public Health Laboratory | Jade Wang, et al. |
| EPI_ISL_683787 | DOHMH PHL | New York City Public Health Laboratory | Jade Wang, et al. |
| EPI_ISL_683788 | DOHMH Chelsea | New York City Public Health Laboratory | Jade Wang, et al. |
| EPI_ISL_683789 | DOHMH PHL | New York City Public Health Laboratory | Jade Wang, et al. |
| EPI_ISL_683790 | DOHMH Morrisania | New York City Public Health Laboratory | Jade Wang, et al. |
| EPI_ISL_683791 | DOHMH Jamaica | New York City Public Health Laboratory | Jade Wang, et al. |
| EPI_ISL_683792 | DOHMH Chelsea | New York City Public Health Laboratory | Jade Wang, et al. |
| EPI_ISL_683793, EPI_ISL_683794 | DOHMH Corona | New York City Public Health Laboratory | Jade Wang, et al. |
| EPI_ISL_683795 | DOHMH Central Harlem | New York City Public Health Laboratory | Jade Wang, et al. |
| EPI_ISL_683796 | DOHMH Corona | New York City Public Health Laboratory | Jade Wang, et al. |
| EPI_ISL_683797 | DOHMH PHL | New York City Public Health Laboratory | Jade Wang, et al. |
| EPI_ISL_683798, EPI_ISL_683799, EPI_ISL_683800 | DOHMH Corona | New York City Public Health Laboratory | Jade Wang, et al. |
| EPI_ISL_683801 | DOHMH Morrisania | New York City Public Health Laboratory | Jade Wang, et al. |
| EPI_ISL_683802 | DOHMH Corona | New York City Public Health Laboratory | Jade Wang, et al. |
| EPI_ISL_683803, EPI_ISL_683804, EPI_ISL_683805, EPI_ISL_683806 | DOHMH Morrisania | New York City Public Health Laboratory | Jade Wang, et al. |
| EPI_ISL_683807, EPI_ISL_683808 | DOHMH Chelsea | New York City Public Health Laboratory | Jade Wang, et al. |

| | | | |
|---|---|---|---|
| EPI_ISL_683809 | DOHMH Corona | New York City Public Health Laboratory | Jade Wang, et al. |
| EPI_ISL_683810 | DOHMH PHL | New York City Public Health Laboratory | Jade Wang, et al. |
| EPI_ISL_683811, EPI_ISL_683812 | DOHMH Corona | New York City Public Health Laboratory | Jade Wang, et al. |
| EPI_ISL_683813 | DOHMH Central Harlem | New York City Public Health Laboratory | Jade Wang, et al. |
| EPI_ISL_683814 | DOHMH Morrisania | New York City Public Health Laboratory | Jade Wang, et al. |
| EPI_ISL_683815 | DOHMH Corona | New York City Public Health Laboratory | Jade Wang, et al. |
| EPI_ISL_683816 | DOHMH Fort Greene | New York City Public Health Laboratory | Jade Wang, et al. |
| EPI_ISL_683817, EPI_ISL_683818 | DOHMH Corona | New York City Public Health Laboratory | Jade Wang, et al. |
| EPI_ISL_683819, EPI_ISL_683820 | DOHMH Riverside | New York City Public Health Laboratory | Jade Wang, et al. |
| EPI_ISL_683821 | DOHMH Crown Heights | New York City Public Health Laboratory | Jade Wang, et al. |
| EPI_ISL_683822 | DOHMH Jamaica | New York City Public Health Laboratory | Jade Wang, et al. |
| EPI_ISL_683823 | DOHMH Morrisania | New York City Public Health Laboratory | Jade Wang, et al. |
| EPI_ISL_683824 | DOHMH Corona | New York City Public Health Laboratory | Jade Wang, et al. |
| EPI_ISL_683825, EPI_ISL_683826 | DOHMH Morrisania | New York City Public Health Laboratory | Jade Wang, et al. |
| EPI_ISL_683827 | DOHMH Crown Heights | New York City Public Health Laboratory | Jade Wang, et al. |
| EPI_ISL_683828 | DOHMH Chelsea | New York City Public Health Laboratory | Jade Wang, et al. |
| EPI_ISL_683829 | DOHMH Central Harlem | New York City Public Health Laboratory | Jade Wang, et al. |
| EPI_ISL_683830 | DOHMH Chelsea | New York City Public Health Laboratory | Jade Wang, et al. |
| EPI_ISL_683831 | DOHMH Corona | New York City Public Health Laboratory | Jade Wang, et al. |
| EPI_ISL_683832 | DOHMH Fort Greene | New York City Public Health Laboratory | Jade Wang, et al. |
| EPI_ISL_683833 | DOHMH Corona | New York City Public Health Laboratory | Jade Wang, et al. |
| EPI_ISL_683834 | DOHMH Central Harlem | New York City Public Health Laboratory | Jade Wang, et al. |
| EPI_ISL_683836 | DOHMH PHL | New York City Public Health Laboratory | Jade Wang, et al. |
| EPI_ISL_683837, EPI_ISL_683838 | DOHMH Jamaica | New York City Public Health Laboratory | Jade Wang, et al. |
| EPI_ISL_683839, EPI_ISL_683840 | DOHMH Morrisania | New York City Public Health Laboratory | Jade Wang, et al. |
| EPI_ISL_683841 | DOHMH Corona | New York City Public Health Laboratory | Jade Wang, et al. |
| EPI_ISL_683842 | DOHMH Central Harlem | New York City Public Health Laboratory | Jade Wang, et al. |
| EPI_ISL_683843, EPI_ISL_683844, EPI_ISL_683845 | DOHMH Morrisania | New York City Public Health Laboratory | Jade Wang, et al. |
| EPI_ISL_683846 | DOHMH PHL | New York City Public Health Laboratory | Jade Wang, et al. |
| EPI_ISL_683847 | DOHMH Corona | New York City Public Health Laboratory | Jade Wang, et al. |
| EPI_ISL_683848 | DOHMH Central Harlem | New York City Public Health Laboratory | Jade Wang, et al. |
| EPI_ISL_683849 | DOHMH Corona | New York City Public Health Laboratory | Jade Wang, et al. |
| EPI_ISL_683850 | DOHMH Jamaica | New York City Public Health Laboratory | Jade Wang, et al. |
| EPI_ISL_683851 | DOHMH Corona | New York City Public Health Laboratory | Jade Wang, et al. |
| EPI_ISL_683852 | DOHMH PHL | New York City Public Health Laboratory | Jade Wang, et al. |
| EPI_ISL_683853, EPI_ISL_683854 | DOHMH Morrisania | New York City Public Health Laboratory | Jade Wang, et al. |
| EPI_ISL_683855 | DOHMH Corona | New York City Public Health Laboratory | Jade Wang, et al. |
| EPI_ISL_683856 | DOHMH Crown Heights | New York City Public Health Laboratory | Jade Wang, et al. |
| EPI_ISL_683857 | DOHMH Corona | New York City Public Health Laboratory | Jade Wang, et al. |
| EPI_ISL_683858 | DOHMH Jamaica | New York City Public Health Laboratory | Jade Wang, et al. |
| EPI_ISL_683859 | DOHMH Central Harlem | New York City Public Health Laboratory | Jade Wang, et al. |
| EPI_ISL_683860 | DOHMH Corona | New York City Public Health Laboratory | Jade Wang, et al. |
| EPI_ISL_683861, EPI_ISL_683862 | DOHMH Jamaica | New York City Public Health Laboratory | Jade Wang, et al. |
| EPI_ISL_683863 | DOHMH Morrisania | New York City Public Health Laboratory | Jade Wang, et al. |
| EPI_ISL_683864 | DOHMH Jamaica | New York City Public Health Laboratory | Jade Wang, et al. |
| EPI_ISL_683865 | DOHMH Chelsea | New York City Public Health Laboratory | Jade Wang, et al. |
| EPI_ISL_683866 | DOHMH Riverside | New York City Public Health Laboratory | Jade Wang, et al. |
| EPI_ISL_683867 | DOHMH Corona | New York City Public Health Laboratory | Jade Wang, et al. |
| EPI_ISL_683868 | DOHMH Fort Greene | New York City Public Health Laboratory | Jade Wang, et al. |
| EPI_ISL_683869 | DOHMH Morrisania | New York City Public Health Laboratory | Jade Wang, et al. |
| EPI_ISL_683870 | DOHMH Corona | New York City Public Health Laboratory | Jade Wang, et al. |
| EPI_ISL_683871 | DOHMH PHL | New York City Public Health Laboratory | Jade Wang, et al. |
| EPI_ISL_683872, EPI_ISL_683873 | DOHMH Corona | New York City Public Health Laboratory | Jade Wang, et al. |
| EPI_ISL_683874 | DOHMH Jamaica | New York City Public Health Laboratory | Jade Wang, et al. |
| EPI_ISL_683875 | DOHMH PHL | New York City Public Health Laboratory | Jade Wang, et al. |
| EPI_ISL_683876 | DOHMH Corona | New York City Public Health Laboratory | Jade Wang, et al. |
| EPI_ISL_683877 | DOHMH Central Harlem | New York City Public Health Laboratory | Jade Wang, et al. |
| EPI_ISL_683878 | DOHMH Jamaica | New York City Public Health Laboratory | Jade Wang, et al. |

| | | | |
|---|---|---|---|
| EPI_ISL_683879, EPI_ISL_683880 | DOHMH Morrisania | New York City Public Health Laboratory | Jade Wang, et al. |
| EPI_ISL_683881 | DOHMH Corona | New York City Public Health Laboratory | Jade Wang, et al. |
| EPI_ISL_683882, EPI_ISL_683883 | DOHMH Central Harlem | New York City Public Health Laboratory | Jade Wang, et al. |
| EPI_ISL_683884 | DOHMH Morrisania | New York City Public Health Laboratory | Jade Wang, et al. |
| EPI_ISL_683885 | DOHMH Jamaica | New York City Public Health Laboratory | Jade Wang, et al. |
| EPI_ISL_683886 | DOHMH Morrisania | New York City Public Health Laboratory | Jade Wang, et al. |
| EPI_ISL_683887, EPI_ISL_683888 | DOHMH Fort Greene | New York City Public Health Laboratory | Jade Wang, et al. |
| EPI_ISL_683889 | DOHMH Morrisania | New York City Public Health Laboratory | Jade Wang, et al. |
| EPI_ISL_683890 | DOHMH PHL | New York City Public Health Laboratory | Jade Wang, et al. |
| EPI_ISL_683891 | DOHMH Crown Heights | New York City Public Health Laboratory | Jade Wang, et al. |
| EPI_ISL_683892 | DOHMH Jamaica | New York City Public Health Laboratory | Jade Wang, et al. |
| EPI_ISL_683893 | DOHMH Corona | New York City Public Health Laboratory | Jade Wang, et al. |
| EPI_ISL_683894, EPI_ISL_683895 | DOHMH Riverside | New York City Public Health Laboratory | Jade Wang, et al. |
| EPI_ISL_683896, EPI_ISL_683897, EPI_ISL_683898, EPI_ISL_683899, EPI_ISL_683900 | DOHMH Corona | New York City Public Health Laboratory | Jade Wang, et al. |
| EPI_ISL_683901 | DOHMH Jamaica | New York City Public Health Laboratory | Jade Wang, et al. |
| EPI_ISL_683902 | DOHMH Riverside | New York City Public Health Laboratory | Jade Wang, et al. |
| EPI_ISL_683903 | DOHMH Jamaica | New York City Public Health Laboratory | Jade Wang, et al. |
| EPI_ISL_683904 | DOHMH Crown Heights | New York City Public Health Laboratory | Jade Wang, et al. |
| EPI_ISL_683905 | DOHMH Morrisania | New York City Public Health Laboratory | Jade Wang, et al. |
| EPI_ISL_683906 | DOHMH Fort Greene | New York City Public Health Laboratory | Jade Wang, et al. |
| EPI_ISL_683907 | DOHMH Riverside | New York City Public Health Laboratory | Jade Wang, et al. |
| EPI_ISL_683908, EPI_ISL_683909 | DOHMH Corona | New York City Public Health Laboratory | Jade Wang, et al. |
| EPI_ISL_683910 | DOHMH Crown Heights | New York City Public Health Laboratory | Jade Wang, et al. |
| EPI_ISL_683911, EPI_ISL_683912 | DOHMH Corona | New York City Public Health Laboratory | Jade Wang, et al. |
| EPI_ISL_683913 | DOHMH Fort Greene | New York City Public Health Laboratory | Jade Wang, et al. |
| EPI_ISL_683914, EPI_ISL_683915, EPI_ISL_683916 | DOHMH Jamaica | New York City Public Health Laboratory | Jade Wang, et al. |
| EPI_ISL_683917, EPI_ISL_683918, EPI_ISL_683919 | DOHMH Morrisania | New York City Public Health Laboratory | Jade Wang, et al. |
| EPI_ISL_683920, EPI_ISL_683921 | DOHMH Jamaica | New York City Public Health Laboratory | Jade Wang, et al. |
| EPI_ISL_683922 | DOHMH Fort Greene | New York City Public Health Laboratory | Jade Wang, et al. |
| EPI_ISL_683923 | DOHMH Chelsea | New York City Public Health Laboratory | Jade Wang, et al. |
| EPI_ISL_683924 | DOHMH Central Harlem | New York City Public Health Laboratory | Jade Wang, et al. |
| EPI_ISL_683925 | DOHMH Jamaica | New York City Public Health Laboratory | Jade Wang, et al. |
| EPI_ISL_683926 | DOHMH Corona | New York City Public Health Laboratory | Jade Wang, et al. |
| EPI_ISL_683927, EPI_ISL_683928 | DOHMH Central Harlem | New York City Public Health Laboratory | Jade Wang, et al. |
| EPI_ISL_683929 | DOHMH Corona | New York City Public Health Laboratory | Jade Wang, et al. |
| EPI_ISL_683930 | DOHMH Jamaica | New York City Public Health Laboratory | Jade Wang, et al. |
| EPI_ISL_683931 | DOHMH Chelsea | New York City Public Health Laboratory | Jade Wang, et al. |
| EPI_ISL_683932 | DOHMH Corona | New York City Public Health Laboratory | Jade Wang, et al. |
| EPI_ISL_683933 | DOHMH Central Harlem | New York City Public Health Laboratory | Jade Wang, et al. |
| EPI_ISL_683934 | DOHMH Corona | New York City Public Health Laboratory | Jade Wang, et al. |
| EPI_ISL_683935 | DOHMH Crown Heights | New York City Public Health Laboratory | Jade Wang, et al. |
| EPI_ISL_683936 | DOHMH Chelsea | New York City Public Health Laboratory | Jade Wang, et al. |
| EPI_ISL_683937 | DOHMH Central Harlem | New York City Public Health Laboratory | Jade Wang, et al. |
| EPI_ISL_683938 | DOHMH Morrisania | New York City Public Health Laboratory | Jade Wang, et al. |
| EPI_ISL_683939 | DOHMH Jamaica | New York City Public Health Laboratory | Jade Wang, et al. |
| EPI_ISL_683940 | DOHMH Morrisania | New York City Public Health Laboratory | Jade Wang, et al. |
| EPI_ISL_683941 | DOHMH Chelsea | New York City Public Health Laboratory | Jade Wang, et al. |
| EPI_ISL_683942 | DOHMH Riverside | New York City Public Health Laboratory | Jade Wang, et al. |
| EPI_ISL_683943 | DOHMH PHL | New York City Public Health Laboratory | Jade Wang, et al. |
| EPI_ISL_683944, EPI_ISL_683945 | DOHMH Jamaica | New York City Public Health Laboratory | Jade Wang, et al. |
| EPI_ISL_683946, EPI_ISL_683947 | DOHMH Morrisania | New York City Public Health Laboratory | Jade Wang, et al. |
| EPI_ISL_683948 | DOHMH Jamaica | New York City Public Health Laboratory | Jade Wang, et al. |
| EPI_ISL_683949, EPI_ISL_683950 | DOHMH Riverside | New York City Public Health Laboratory | Jade Wang, et al. |
| EPI_ISL_683951 | DOHMH Jamaica | New York City Public Health Laboratory | Jade Wang, et al. |
| EPI_ISL_683952, EPI_ISL_683953 | DOHMH Corona | New York City Public Health Laboratory | Jade Wang, et al. |
| EPI_ISL_683954 | DOHMH PHL | New York City Public Health Laboratory | Jade Wang, et al. |

| | | | |
|---|---|---|---|
| EPI_ISL_683955 | DOHMH Riverside | New York City Public Health Laboratory | Jade Wang, et al. |
| EPI_ISL_683956 | DOHMH Jamaica | New York City Public Health Laboratory | Jade Wang, et al. |
| EPI_ISL_683957 | DOHMH Chelsea | New York City Public Health Laboratory | Jade Wang, et al. |
| EPI_ISL_683958 | DOHMH Jamaica | New York City Public Health Laboratory | Jade Wang, et al. |
| EPI_ISL_683959 | DOHMH Morrisania | New York City Public Health Laboratory | Jade Wang, et al. |
| EPI_ISL_683960 | DOHMH Corona | New York City Public Health Laboratory | Jade Wang, et al. |
| EPI_ISL_683961 | DOHMH Chelsea | New York City Public Health Laboratory | Jade Wang, et al. |
| EPI_ISL_683962 | DOHMH Central Harlem | New York City Public Health Laboratory | Jade Wang, et al. |
| EPI_ISL_683963 | DOHMH Fort Greene | New York City Public Health Laboratory | Jade Wang, et al. |
| EPI_ISL_683964 | DOHMH Central Harlem | New York City Public Health Laboratory | Jade Wang, et al. |
| EPI_ISL_683965, EPI_ISL_683966 | DOHMH Morrisania | New York City Public Health Laboratory | Jade Wang, et al. |
| EPI_ISL_683967 | DOHMH Chelsea | New York City Public Health Laboratory | Jade Wang, et al. |
| EPI_ISL_683968 | DOHMH Central Harlem | New York City Public Health Laboratory | Jade Wang, et al. |
| EPI_ISL_683969 | DOHMH Morrisania | New York City Public Health Laboratory | Jade Wang, et al. |
| EPI_ISL_683970 | DOHMH Crown Heights | New York City Public Health Laboratory | Jade Wang, et al. |
| EPI_ISL_683971, EPI_ISL_683972 | DOHMH Chelsea | New York City Public Health Laboratory | Jade Wang, et al. |
| EPI_ISL_683973 | DOHMH Fort Greene | New York City Public Health Laboratory | Jade Wang, et al. |
| EPI_ISL_683974 | DOHMH Central Harlem | New York City Public Health Laboratory | Jade Wang, et al. |
| EPI_ISL_683975 | DOHMH Jamaica | New York City Public Health Laboratory | Jade Wang, et al. |
| EPI_ISL_683976, EPI_ISL_683977 | DOHMH Central Harlem | New York City Public Health Laboratory | Jade Wang, et al. |
| EPI_ISL_683978, EPI_ISL_683979 | DOHMH Corona | New York City Public Health Laboratory | Jade Wang, et al. |
| EPI_ISL_683980, EPI_ISL_683981 | DOHMH Chelsea | New York City Public Health Laboratory | Jade Wang, et al. |
| EPI_ISL_683982, EPI_ISL_683983, EPI_ISL_683984, EPI_ISL_683985 | DOHMH Central Harlem | New York City Public Health Laboratory | Jade Wang, et al. |
| EPI_ISL_683986 | DOHMH Jamaica | New York City Public Health Laboratory | Jade Wang, et al. |
| EPI_ISL_683987, EPI_ISL_683988 | DOHMH PHL | New York City Public Health Laboratory | Jade Wang, et al. |
| EPI_ISL_683989 | DOHMH Chelsea | New York City Public Health Laboratory | Jade Wang, et al. |
| EPI_ISL_683990 | DOHMH Jamaica | New York City Public Health Laboratory | Jade Wang, et al. |
| EPI_ISL_683991 | DOHMH Central Harlem | New York City Public Health Laboratory | Jade Wang, et al. |
| EPI_ISL_683992 | DOHMH Morrisania | New York City Public Health Laboratory | Jade Wang, et al. |
| EPI_ISL_683993 | DOHMH PHL | New York City Public Health Laboratory | Jade Wang, et al. |
| EPI_ISL_683994 | DOHMH Riverside | New York City Public Health Laboratory | Jade Wang, et al. |
| EPI_ISL_683995 | DOHMH PHL | New York City Public Health Laboratory | Jade Wang, et al. |
| EPI_ISL_683996 | DOHMH Fort Greene | New York City Public Health Laboratory | Jade Wang, et al. |
| EPI_ISL_734287, EPI_ISL_734288, EPI_ISL_734289, EPI_ISL_734290, EPI_ISL_734291, EPI_ISL_734292, EPI_ISL_734293, EPI_ISL_734294, EPI_ISL_734295, EPI_ISL_734296, EPI_ISL_734297, EPI_ISL_734298, EPI_ISL_734299, EPI_ISL_734300, EPI_ISL_734301, EPI_ISL_734302, EPI_ISL_734303, EPI_ISL_734304, EPI_ISL_734305, EPI_ISL_734306, EPI_ISL_734307, EPI_ISL_734308, EPI_ISL_734309, EPI_ISL_734310, EPI_ISL_734311, EPI_ISL_734312, EPI_ISL_734313, EPI_ISL_734314, EPI_ISL_734315, EPI_ISL_734316, EPI_ISL_734317, EPI_ISL_734318, EPI_ISL_734319, EPI_ISL_734320, EPI_ISL_734321, EPI_ISL_734322, EPI_ISL_734323, EPI_ISL_734324, EPI_ISL_734325, EPI_ISL_734326, EPI_ISL_734327, EPI_ISL_734328, EPI_ISL_734329, EPI_ISL_734330, EPI_ISL_734331, EPI_ISL_734332, EPI_ISL_734333, EPI_ISL_734334, EPI_ISL_734335, EPI_ISL_734336, EPI_ISL_734337, EPI_ISL_734338, EPI_ISL_734339, EPI_ISL_734340, EPI_ISL_734341, EPI_ISL_734342, EPI_ISL_734343, EPI_ISL_734344, EPI_ISL_734345, EPI_ISL_734346, EPI_ISL_734347, EPI_ISL_734348, EPI_ISL_734349, EPI_ISL_734350, EPI_ISL_734351, EPI_ISL_734352, EPI_ISL_734353, EPI_ISL_734354, EPI_ISL_734355, EPI_ISL_734356, EPI_ISL_734357, EPI_ISL_734358, EPI_ISL_734359, EPI_ISL_734360, EPI_ISL_734361, EPI_ISL_734362, EPI_ISL_734363, EPI_ISL_734364, EPI_ISL_734365, EPI_ISL_734366, EPI_ISL_734367, EPI_ISL_734368, EPI_ISL_734369, EPI_ISL_734370, EPI_ISL_734371, EPI_ISL_734372, EPI_ISL_734373, EPI_ISL_734374, EPI_ISL_734375, EPI_ISL_734376, EPI_ISL_734377, EPI_ISL_734378, EPI_ISL_734379, EPI_ISL_734380, EPI_ISL_734381, EPI_ISL_734382, EPI_ISL_734383, EPI_ISL_734384, EPI_ISL_734385, EPI_ISL_734386, EPI_ISL_734387, EPI_ISL_734388, EPI_ISL_734389, EPI_ISL_734390, EPI_ISL_734391, EPI_ISL_734392, EPI_ISL_734393, EPI_ISL_734394, EPI_ISL_734395, EPI_ISL_734396, EPI_ISL_734397, EPI_ISL_734398, EPI_ISL_734399, EPI_ISL_734400, EPI_ISL_734401, EPI_ISL_734402, EPI_ISL_734403, EPI_ISL_734404, EPI_ISL_734405, EPI_ISL_734406, EPI_ISL_734407, EPI_ISL_734408, EPI_ISL_734409, EPI_ISL_734410, EPI_ISL_734411, EPI_ISL_734412, EPI_ISL_734413, EPI_ISL_734414, EPI_ISL_734415, EPI_ISL_734416, EPI_ISL_734417, EPI_ISL_734418, EPI_ISL_734419, EPI_ISL_734420, EPI_ISL_734421, EPI_ISL_734422, EPI_ISL_734423, EPI_ISL_734424, EPI_ISL_734425, EPI_ISL_734426, EPI_ISL_734427, EPI_ISL_734428, EPI_ISL_734429, EPI_ISL_734430, EPI_ISL_734431, EPI_ISL_734432, EPI_ISL_734433, EPI_ISL_734434, EPI_ISL_734435, EPI_ISL_734436, EPI_ISL_734437, EPI_ISL_734438, EPI_ISL_734439, EPI_ISL_734440, EPI_ISL_734441, EPI_ISL_734442, EPI_ISL_734443, EPI_ISL_734444, EPI_ISL_734445, EPI_ISL_734446 | | | |
| see above | Wadsworth Center, New York State Department.of Health | Wadsworth Center, New York State Department.of Health | Kirsten St. George, Daryl M. Lamson, Alexis Russel, Jonathan Plitnick, Navjot Singh, John Kelly, Sara Griesemer, Erasmus Schneider, Erica Lasek-Nesselquist |
| EPI_ISL_734447, EPI_ISL_734448, EPI_ISL_734449, EPI_ISL_734450, EPI_ISL_734451, EPI_ISL_734452, EPI_ISL_734453, EPI_ISL_734454, EPI_ISL_734455, EPI_ISL_734456, EPI_ISL_734457, EPI_ISL_734458, EPI_ISL_734459, EPI_ISL_734460, EPI_ISL_734461, EPI_ISL_734462, EPI_ISL_734463, EPI_ISL_734464, EPI_ISL_734465, EPI_ISL_734466, EPI_ISL_734467, EPI_ISL_734468, EPI_ISL_734469, EPI_ISL_734470, EPI_ISL_734471, EPI_ISL_734472, EPI_ISL_734473, EPI_ISL_734474, EPI_ISL_734475, EPI_ISL_734476, EPI_ISL_734477, EPI_ISL_734478, EPI_ISL_734479, EPI_ISL_734480, EPI_ISL_734481 | | | |
| see above | Masonic Medical Research Institute | Wadsworth Center, New York State Department.of Health | Kirsten St. George, Nathan Tucker, Ryan D. Pfeiffer, Daryl M. Lamson, Alexis Russel, Jonathan Plitnick, Navjot Singh, John Kelly, Sara Griesemer, Erasmus Schneider, Erica Lasek-Nesselquist |
| EPI_ISL_745398 | DOHMH Jamaica | New York City Public Health Laboratory | Jade Wang, et al. |
| EPI_ISL_745399 | DOHMH Morrisania | New York City Public Health Laboratory | Jade Wang, et al. |
| EPI_ISL_745400 | DOHMH Central Harlem | New York City Public Health Laboratory | Jade Wang, et al. |
| EPI_ISL_745401 | DOHMH Corona | New York City Public Health Laboratory | Jade Wang, et al. |
| EPI_ISL_745402 | DOHMH Jamaica | New York City Public Health Laboratory | Jade Wang, et al. |
| EPI_ISL_745403 | DOHMH Corona | New York City Public Health Laboratory | Jade Wang, et al. |
| EPI_ISL_745404 | DOHMH Central Harlem | New York City Public Health Laboratory | Jade Wang, et al. |
| EPI_ISL_745405 | DOHMH Jamaica | New York City Public Health Laboratory | Jade Wang, et al. |
| EPI_ISL_745406 | DOHMH Morrisania | New York City Public Health Laboratory | Jade Wang, et al. |
| EPI_ISL_745407 | DOHMH Jamaica | New York City Public Health Laboratory | Jade Wang, et al. |
| EPI_ISL_745408 | DOHMH Riverside | New York City Public Health Laboratory | Jade Wang, et al. |
| EPI_ISL_745409 | DOHMH Corona | New York City Public Health Laboratory | Jade Wang, et al. |
| EPI_ISL_745410 | DOHMH Chelsea | New York City Public Health Laboratory | Jade Wang, et al. |
| EPI_ISL_745411 | DOHMH Corona | New York City Public Health Laboratory | Jade Wang, et al. |

| | | | |
|---|---|---|---|
| EPI_ISL_745412 | DOHMH Morrisania | New York City Public Health Laboratory | Jade Wang, et al. |
| EPI_ISL_745413 | DOHMH Central Harlem | New York City Public Health Laboratory | Jade Wang, et al. |
| EPI_ISL_745414, EPI_ISL_745415, EPI_ISL_745416, EPI_ISL_745417, EPI_ISL_745418 | DOHMH Jamaica | New York City Public Health Laboratory | Jade Wang, et al. |
| EPI_ISL_745419, EPI_ISL_745420 | DOHMH Crown Heights | New York City Public Health Laboratory | Jade Wang, et al. |
| EPI_ISL_745421, EPI_ISL_745422 | DOHMH Chelsea | New York City Public Health Laboratory | Jade Wang, et al. |
| EPI_ISL_745423, EPI_ISL_745424 | DOHMH Corona | New York City Public Health Laboratory | Jade Wang, et al. |
| EPI_ISL_745425 | DOHMH Central Harlem | New York City Public Health Laboratory | Jade Wang, et al. |
| EPI_ISL_745426, EPI_ISL_745427 | DOHMH Corona | New York City Public Health Laboratory | Jade Wang, et al. |
| EPI_ISL_745428, EPI_ISL_745429 | DOHMH Riverside | New York City Public Health Laboratory | Jade Wang, et al. |
| EPI_ISL_745430, EPI_ISL_745431, EPI_ISL_745432, EPI_ISL_745433 | DOHMH Morrisania | New York City Public Health Laboratory | Jade Wang, et al. |
| EPI_ISL_745434, EPI_ISL_745435 | DOHMH Corona | New York City Public Health Laboratory | Jade Wang, et al. |
| EPI_ISL_745436 | DOHMH Chelsea | New York City Public Health Laboratory | Jade Wang, et al. |
| EPI_ISL_745437, EPI_ISL_745438 | DOHMH PHL | New York City Public Health Laboratory | Jade Wang, et al. |
| EPI_ISL_745439 | DOHMH Riverside | New York City Public Health Laboratory | Jade Wang, et al. |
| EPI_ISL_745440 | DOHMH Jamaica | New York City Public Health Laboratory | Jade Wang, et al. |
| EPI_ISL_745441, EPI_ISL_745442, EPI_ISL_745443 | DOHMH Morrisania | New York City Public Health Laboratory | Jade Wang, et al. |
| EPI_ISL_745444 | DOHMH Jamaica | New York City Public Health Laboratory | Jade Wang, et al. |
| EPI_ISL_745445 | DOHMH Crown Heights | New York City Public Health Laboratory | Jade Wang, et al. |
| EPI_ISL_745446, EPI_ISL_745447 | DOHMH Corona | New York City Public Health Laboratory | Jade Wang, et al. |
| EPI_ISL_745448 | DOHMH Crown Heights | New York City Public Health Laboratory | Jade Wang, et al. |
| EPI_ISL_745449, EPI_ISL_745450 | DOHMH Morrisania | New York City Public Health Laboratory | Jade Wang, et al. |
| EPI_ISL_745451 | DOHMH Central Harlem | New York City Public Health Laboratory | Jade Wang, et al. |
| EPI_ISL_745452 | DOHMH Crown Heights | New York City Public Health Laboratory | Jade Wang, et al. |
| EPI_ISL_745453 | DOHMH Chelsea | New York City Public Health Laboratory | Jade Wang, et al. |
| EPI_ISL_745454 | DOHMH Crown Heights | New York City Public Health Laboratory | Jade Wang, et al. |
| EPI_ISL_745455 | DOHMH Corona | New York City Public Health Laboratory | Jade Wang, et al. |
| EPI_ISL_745456 | DOHMH PHL | New York City Public Health Laboratory | Jade Wang, et al. |
| EPI_ISL_745457 | DOHMH Chelsea | New York City Public Health Laboratory | Jade Wang, et al. |
| EPI_ISL_745458, EPI_ISL_745459, EPI_ISL_745460, EPI_ISL_745461, EPI_ISL_745462 | DOHMH Corona | New York City Public Health Laboratory | Jade Wang, et al. |
| EPI_ISL_745463 | DOHMH Fort Greene | New York City Public Health Laboratory | Jade Wang, et al. |
| EPI_ISL_745464 | DOHMH Corona | New York City Public Health Laboratory | Jade Wang, et al. |
| EPI_ISL_745465 | DOHMH PHL | New York City Public Health Laboratory | Jade Wang, et al. |
| EPI_ISL_745466, EPI_ISL_745467 | DOHMH Corona | New York City Public Health Laboratory | Jade Wang, et al. |
| EPI_ISL_745468 | DOHMH Riverside | New York City Public Health Laboratory | Jade Wang, et al. |
| EPI_ISL_745469 | DOHMH Central Harlem | New York City Public Health Laboratory | Jade Wang, et al. |
| EPI_ISL_745470 | DOHMH Morrisania | New York City Public Health Laboratory | Jade Wang, et al. |
| EPI_ISL_747039, EPI_ISL_747040, EPI_ISL_747041, EPI_ISL_747042, EPI_ISL_747043 | Respiratory Viruses Branch, Centers for Disease Control and Prevention | Respiratory Viruses Branch, Centers for Disease Control and Prevention | Queen,K., Li,Y., Tao,Y., Uehara,A., Montmayeur,A., Paden,C.R., Cook,P.W., Marine,R., Sheth,M., Wang,H., Lee,J., Tong,S. |
| EPI_ISL_751545, EPI_ISL_751548, EPI_ISL_751560, EPI_ISL_751562, EPI_ISL_751580, EPI_ISL_751581, EPI_ISL_751658, EPI_ISL_751682, EPI_ISL_751717 | NYSDOH Wadsworth Center, Virology Lab | Genomics and Discovery, Respiratory Viruses Branch, Division of Viral Diseases, Centers for Disease Control and Prevention | Krista Queen, Yan Li, Ying Tao, Jing Zhang, Anna Uehara, Anna Montmayeur, Clinton R. Paden, Peter W. Cook,Rachel Marine, Mili Sheth, Haibin Wang, Justin Lee, Suxiang Tong |

EPI_ISL_754394, EPI_ISL_754395, EPI_ISL_754396, EPI_ISL_754397, EPI_ISL_754398, EPI_ISL_754399, EPI_ISL_754400, EPI_ISL_754401, EPI_ISL_754402, EPI_ISL_754403, EPI_ISL_754404, EPI_ISL_754405, EPI_ISL_754406, EPI_ISL_754407, EPI_ISL_754408, EPI_ISL_754409, EPI_ISL_754410, EPI_ISL_754411, EPI_ISL_754412, EPI_ISL_754413, EPI_ISL_754414, EPI_ISL_754415, EPI_ISL_754416, EPI_ISL_754417, EPI_ISL_754418, EPI_ISL_754419, EPI_ISL_754420, EPI_ISL_754421, EPI_ISL_754422, EPI_ISL_754423, EPI_ISL_754424, EPI_ISL_754425, EPI_ISL_754426, EPI_ISL_754427, EPI_ISL_754428, EPI_ISL_754429, EPI_ISL_754430, EPI_ISL_754431, EPI_ISL_754432, EPI_ISL_754433, EPI_ISL_754434, EPI_ISL_754435, EPI_ISL_754436, EPI_ISL_754437, EPI_ISL_754438, EPI_ISL_754439, EPI_ISL_754440, EPI_ISL_754441, EPI_ISL_754442, EPI_ISL_754443, EPI_ISL_754444, EPI_ISL_754445, EPI_ISL_754446, EPI_ISL_754447, EPI_ISL_754448, EPI_ISL_754449, EPI_ISL_754450, EPI_ISL_754451, EPI_ISL_754452, EPI_ISL_754453, EPI_ISL_754454, EPI_ISL_754455, EPI_ISL_754456, EPI_ISL_754457, EPI_ISL_754458, EPI_ISL_754459, EPI_ISL_754460, EPI_ISL_754461, EPI_ISL_754462, EPI_ISL_754463, EPI_ISL_754464, EPI_ISL_754465, EPI_ISL_754466, EPI_ISL_754467, EPI_ISL_754468, EPI_ISL_754469, EPI_ISL_754470, EPI_ISL_754471, EPI_ISL_754472, EPI_ISL_754473, EPI_ISL_754474, EPI_ISL_754475, EPI_ISL_754476, EPI_ISL_754477, EPI_ISL_754478, EPI_ISL_754479, EPI_ISL_754480, EPI_ISL_754481, EPI_ISL_754482, EPI_ISL_754483, EPI_ISL_754484, EPI_ISL_754485, EPI_ISL_754486, EPI_ISL_754487, EPI_ISL_754488, EPI_ISL_754489, EPI_ISL_754490, EPI_ISL_754491, EPI_ISL_754492, EPI_ISL_754493, EPI_ISL_754494, EPI_ISL_754495, EPI_ISL_754496, EPI_ISL_754497, EPI_ISL_754498, EPI_ISL_754499, EPI_ISL_754500, EPI_ISL_754501, EPI_ISL_754502, EPI_ISL_754503, EPI_ISL_754504, EPI_ISL_754505, EPI_ISL_754506, EPI_ISL_754507, EPI_ISL_754508, EPI_ISL_754509, EPI_ISL_754510, EPI_ISL_754511, EPI_ISL_754512, EPI_ISL_754513, EPI_ISL_754514, EPI_ISL_754515, EPI_ISL_754516, EPI_ISL_754517, EPI_ISL_754518, EPI_ISL_754519, EPI_ISL_754520, EPI_ISL_754521, EPI_ISL_754522, EPI_ISL_754523, EPI_ISL_754524, EPI_ISL_754525, EPI_ISL_754526, EPI_ISL_754527, EPI_ISL_754528, EPI_ISL_754529, EPI_ISL_754530, EPI_ISL_754531, EPI_ISL_754532, EPI_ISL_754533, EPI_ISL_754534, EPI_ISL_754535, EPI_ISL_754536, EPI_ISL_754537, EPI_ISL_754538, EPI_ISL_754539, EPI_ISL_754540, EPI_ISL_754541, EPI_ISL_754542, EPI_ISL_754543, EPI_ISL_754544, EPI_ISL_754545, EPI_ISL_754546, EPI_ISL_754547, EPI_ISL_754548, EPI_ISL_754549, EPI_ISL_754550, EPI_ISL_754551, EPI_ISL_754552, EPI_ISL_754553, EPI_ISL_754554, EPI_ISL_754555, EPI_ISL_754556, EPI_ISL_754557, EPI_ISL_754558, EPI_ISL_754559, EPI_ISL_754560, EPI_ISL_754561, EPI_ISL_754562, EPI_ISL_754563, EPI_ISL_754564, EPI_ISL_754565, EPI_ISL_754566, EPI_ISL_754567, EPI_ISL_754568, EPI_ISL_754569, EPI_ISL_754570, EPI_ISL_754571, EPI_ISL_754572, EPI_ISL_754573, EPI_ISL_754574, EPI_ISL_754575, EPI_ISL_754576, EPI_ISL_754577, EPI_ISL_754578, EPI_ISL_754579, EPI_ISL_754580, EPI_ISL_754581, EPI_ISL_754582, EPI_ISL_754583, EPI_ISL_754584, EPI_ISL_754585, EPI_ISL_754586, EPI_ISL_754587, EPI_ISL_754588, EPI_ISL_754589, EPI_ISL_754590, EPI_ISL_754591, EPI_ISL_754592, EPI_ISL_754593, EPI_ISL_754594, EPI_ISL_754595, EPI_ISL_754596, EPI_ISL_754597, EPI_ISL_754598, EPI_ISL_754599, EPI_ISL_754600, EPI_ISL_754601, EPI_ISL_754602, EPI_ISL_754603, EPI_ISL_754604, EPI_ISL_754605, EPI_ISL_754606, EPI_ISL_754607, EPI_ISL_754608, EPI_ISL_754609, EPI_ISL_754610, EPI_ISL_754611, EPI_ISL_754612, EPI_ISL_754613, EPI_ISL_754614, EPI_ISL_754615, EPI_ISL_754616, EPI_ISL_754617, EPI_ISL_754618, EPI_ISL_754677, EPI_ISL_754678, EPI_ISL_754679, EPI_ISL_754680, EPI_ISL_754681, EPI_ISL_754682, EPI_ISL_754683, EPI_ISL_754684, EPI_ISL_754685, EPI_ISL_754686, EPI_ISL_754687, EPI_ISL_754688, EPI_ISL_754689, EPI_ISL_754690, EPI_ISL_754691, EPI_ISL_754692, EPI_ISL_754693, EPI_ISL_754694, EPI_ISL_754695, EPI_ISL_754696, EPI_ISL_754697, EPI_ISL_754698, EPI_ISL_754699, EPI_ISL_754700, EPI_ISL_754701, EPI_ISL_754702, EPI_ISL_754703, EPI_ISL_754704, EPI_ISL_754705, EPI_ISL_754706, EPI_ISL_754707, EPI_ISL_754708, EPI_ISL_754709, EPI_ISL_754710, EPI_ISL_754711, EPI_ISL_754712, EPI_ISL_754713, EPI_ISL_754714, EPI_ISL_754715, EPI_ISL_754716, EPI_ISL_754717, EPI_ISL_754718, EPI_ISL_754719, EPI_ISL_754720, EPI_ISL_754721, EPI_ISL_754722, EPI_ISL_754723, EPI_ISL_754724, EPI_ISL_754725, EPI_ISL_754726, EPI_ISL_754727, EPI_ISL_754728, EPI_ISL_754729, EPI_ISL_754730, EPI_ISL_754731, EPI_ISL_754732, EPI_ISL_754733, EPI_ISL_754734, EPI_ISL_754735, EPI_ISL_754736, EPI_ISL_754737, EPI_ISL_754738, EPI_ISL_754739, EPI_ISL_754740, EPI_ISL_754741, EPI_ISL_754742, EPI_ISL_754743, EPI_ISL_754744, EPI_ISL_754745, EPI_ISL_754779, EPI_ISL_754780, EPI_ISL_754781, EPI_ISL_754782, EPI_ISL_754783, EPI_ISL_754784, EPI_ISL_754785, EPI_ISL_754786, EPI_ISL_754787, EPI_ISL_754788, EPI_ISL_754789, EPI_ISL_754790, EPI_ISL_754791, EPI_ISL_754792, EPI_ISL_754793, EPI_ISL_754794, EPI_ISL_754795, EPI_ISL_754796, EPI_ISL_754797, EPI_ISL_754798, EPI_ISL_754799, EPI_ISL_754800, EPI_ISL_754801, EPI_ISL_754802, EPI_ISL_754803, EPI_ISL_754804, EPI_ISL_754805, EPI_ISL_754806, EPI_ISL_754807, EPI_ISL_754808,

| | | | |
|---|---|---|---|
| EPI_ISL_754809, EPI_ISL_754810, EPI_ISL_754811, EPI_ISL_754812, EPI_ISL_754813, EPI_ISL_754814, EPI_ISL_754815, EPI_ISL_754816, EPI_ISL_754817, EPI_ISL_754818, EPI_ISL_754819, EPI_ISL_754820, EPI_ISL_754821, EPI_ISL_754822, EPI_ISL_754823, EPI_ISL_754824, EPI_ISL_754825, EPI_ISL_754826, EPI_ISL_754827, EPI_ISL_754828, EPI_ISL_754829, EPI_ISL_754830, EPI_ISL_754831, EPI_ISL_754832, EPI_ISL_754833, EPI_ISL_754834, EPI_ISL_754835, EPI_ISL_754836, EPI_ISL_754837, EPI_ISL_754838, EPI_ISL_754839, EPI_ISL_754840, EPI_ISL_754841, EPI_ISL_765480 | | | |
| see above | Wadsworth Center, New York State Department.of Health | Wadsworth Center, New York State Department.of Health | Kirsten St. George, Daryl M. Lamson, Alexis Russel, Matthew Shudt, Melissa A Leisner, Jonathan Plitnick, Navjot Singh, John Kelly, Sara Griesemer, Erasmus Schneider, Erica Lasek-Nesselquist |
| EPI_ISL_765481 | SARATOGA HOSPITAL LABORATORY | Wadsworth Center, New York State Department.of Health | Kirsten St. George, Daryl M. Lamson, Alexis Russel, Matthew Shudt, Melissa A Leisner, Jonathan Plitnick, Navjot Singh, John Kelly, Sara Griesemer, Erasmus Schneider, Erica Lasek-Nesselquist |
| EPI_ISL_765482 | Wadsworth Center, New York State Department.of Health | Wadsworth Center, New York State Department.of Health | Kirsten St. George, Daryl M. Lamson, Alexis Russel, Matthew Shudt, Melissa A Leisner, Jonathan Plitnick, Navjot Singh, John Kelly, Sara Griesemer, Erasmus Schneider, Erica Lasek-Nesselquist |
| EPI_ISL_765483 | SARATOGA HOSPITAL LABORATORY | Wadsworth Center, New York State Department.of Health | Kirsten St. George, Daryl M. Lamson, Alexis Russel, Matthew Shudt, Melissa A Leisner, Jonathan Plitnick, Navjot Singh, John Kelly, Sara Griesemer, Erasmus Schneider, Erica Lasek-Nesselquist |
| EPI_ISL_765484, EPI_ISL_765485, EPI_ISL_765486, EPI_ISL_765487, EPI_ISL_765488 | Wadsworth Center, New York State Department.of Health | Wadsworth Center, New York State Department.of Health | Kirsten St. George, Daryl M. Lamson, Alexis Russel, Matthew Shudt, Melissa A Leisner, Jonathan Plitnick, Navjot Singh, John Kelly, Sara Griesemer, Erasmus Schneider, Erica Lasek-Nesselquist |
| EPI_ISL_765489, EPI_ISL_765490, EPI_ISL_765491, EPI_ISL_765492, EPI_ISL_765493, EPI_ISL_765494, EPI_ISL_765495 | MONTEFIORE MEDICAL CENTER LABORATORIES | Wadsworth Center, New York State Department.of Health | Kirsten St. George, Daryl M. Lamson, Alexis Russel, Matthew Shudt, Melissa A Leisner, Jonathan Plitnick, Navjot Singh, John Kelly, Sara Griesemer, Erasmus Schneider, Erica Lasek-Nesselquist |
| EPI_ISL_765496, EPI_ISL_765497, EPI_ISL_765498, EPI_ISL_765499, EPI_ISL_765500, EPI_ISL_765501, EPI_ISL_765502, EPI_ISL_765503, EPI_ISL_765504, EPI_ISL_765505, EPI_ISL_765506, EPI_ISL_765507, EPI_ISL_765508, EPI_ISL_765509, EPI_ISL_765510, EPI_ISL_765511, EPI_ISL_765512, EPI_ISL_765513, EPI_ISL_765514, EPI_ISL_765515, EPI_ISL_765516 | | | |
| see above | Wadsworth Center, New York State Department.of Health | Wadsworth Center, New York State Department.of Health | Kirsten St. George, Daryl M. Lamson, Alexis Russel, Matthew Shudt, Melissa A Leisner, Jonathan Plitnick, Navjot Singh, John Kelly, Sara Griesemer, Erasmus Schneider, Erica Lasek-Nesselquist |
| EPI_ISL_765517, EPI_ISL_765518, EPI_ISL_765519, EPI_ISL_765520, EPI_ISL_765521, EPI_ISL_765522, EPI_ISL_765523, EPI_ISL_765524, EPI_ISL_765525, EPI_ISL_765526, EPI_ISL_765527, EPI_ISL_765528, EPI_ISL_765529 | | | |
| see above | SARATOGA HOSPITAL LABORATORY | Wadsworth Center, New York State Department.of Health | Kirsten St. George, Daryl M. Lamson, Alexis Russel, Matthew Shudt, Melissa A Leisner, Jonathan Plitnick, Navjot Singh, John Kelly, Sara Griesemer, Erasmus Schneider, Erica Lasek-Nesselquist |
| EPI_ISL_765530, EPI_ISL_765531, EPI_ISL_765532, EPI_ISL_765533, EPI_ISL_765534, EPI_ISL_765535, EPI_ISL_765536, EPI_ISL_765537, EPI_ISL_765538, EPI_ISL_765539, EPI_ISL_765540, EPI_ISL_765541, EPI_ISL_765542, EPI_ISL_765543, EPI_ISL_765544, EPI_ISL_765545, EPI_ISL_765546, EPI_ISL_765547, EPI_ISL_765548, EPI_ISL_765549, EPI_ISL_765550, EPI_ISL_765551, EPI_ISL_765552, EPI_ISL_765553, EPI_ISL_765554, EPI_ISL_765555, EPI_ISL_765556, EPI_ISL_765557, EPI_ISL_765558, EPI_ISL_765559, EPI_ISL_765560, EPI_ISL_765561, EPI_ISL_765562, EPI_ISL_765563, EPI_ISL_765564, EPI_ISL_765565, EPI_ISL_765566 | | | |
| see above | MONTEFIORE MEDICAL CENTER LABORATORIES | Wadsworth Center, New York State Department.of Health | Kirsten St. George, Daryl M. Lamson, Alexis Russel, Matthew Shudt, Melissa A Leisner, Jonathan Plitnick, Navjot Singh, John Kelly, Sara Griesemer, Erasmus Schneider, Erica Lasek-Nesselquist |
| EPI_ISL_767417 | URMC LABS | Wadsworth Center, New York State Department.of Health | Kirsten St. George, Daryl M. Lamson, Alexis Russel, Matthew Shudt, Melissa A Leisner, Jonathan Plitnick, Navjot Singh, John Kelly, Sara Griesemer, Erasmus Schneider, Erica Lasek-Nesselquist |
| EPI_ISL_767418 | Wadsworth Center, New York State Department.of Health | Wadsworth Center, New York State Department.of Health | Kirsten St. George, Daryl M. Lamson, Alexis Russel, Matthew Shudt, Melissa A Leisner, Jonathan Plitnick, Navjot Singh, John Kelly, Sara Griesemer, Erasmus Schneider, Erica Lasek-Nesselquist |
| EPI_ISL_767419 | URMC LABS | Wadsworth Center, New York State Department.of Health | Kirsten St. George, Daryl M. Lamson, Alexis Russel, Matthew Shudt, Melissa A Leisner, Jonathan Plitnick, Navjot Singh, John Kelly, Sara Griesemer, Erasmus Schneider, Erica Lasek-Nesselquist |
| EPI_ISL_767420, EPI_ISL_767421, EPI_ISL_767422 | Wadsworth Center, New York State Department.of Health | Wadsworth Center, New York State Department.of Health | Kirsten St. George, Daryl M. Lamson, Alexis Russel, Matthew Shudt, Melissa A Leisner, Jonathan Plitnick, Navjot Singh, John Kelly, Sara Griesemer, Erasmus Schneider, Erica Lasek-Nesselquist |
| EPI_ISL_767423, EPI_ISL_767424 | URMC LABS | Wadsworth Center, New York State Department.of Health | Kirsten St. George, Daryl M. Lamson, Alexis Russel, Matthew Shudt, Melissa A Leisner, Jonathan Plitnick, Navjot Singh, John Kelly, Sara Griesemer, Erasmus Schneider, Erica Lasek-Nesselquist |
| EPI_ISL_767425, EPI_ISL_767426, EPI_ISL_767427, EPI_ISL_767428, EPI_ISL_767429, EPI_ISL_767430, EPI_ISL_767431, EPI_ISL_767432, EPI_ISL_767433, EPI_ISL_767434, EPI_ISL_767435, EPI_ISL_767436, EPI_ISL_767437, EPI_ISL_767438, EPI_ISL_767439, EPI_ISL_767440, EPI_ISL_767441, EPI_ISL_767442, EPI_ISL_767443, EPI_ISL_767444, EPI_ISL_767445, EPI_ISL_767446, EPI_ISL_767447, EPI_ISL_767448, EPI_ISL_767449, EPI_ISL_767450, EPI_ISL_767451, EPI_ISL_767452, EPI_ISL_767453, EPI_ISL_767454, EPI_ISL_767455, EPI_ISL_767456, EPI_ISL_767457, EPI_ISL_767458, EPI_ISL_767459, EPI_ISL_767460, EPI_ISL_767461, EPI_ISL_767462, EPI_ISL_767463, EPI_ISL_767464, EPI_ISL_767465, EPI_ISL_767466, EPI_ISL_767467, EPI_ISL_767468, EPI_ISL_767469, EPI_ISL_767470, EPI_ISL_767471, EPI_ISL_767472, EPI_ISL_767473, EPI_ISL_767474, EPI_ISL_767475, EPI_ISL_767476, EPI_ISL_767477, EPI_ISL_767478, EPI_ISL_767479, EPI_ISL_767480, EPI_ISL_767481, EPI_ISL_767482, EPI_ISL_767483, EPI_ISL_767484, EPI_ISL_767485, EPI_ISL_767486, EPI_ISL_767487, EPI_ISL_767488, EPI_ISL_767489, EPI_ISL_767490, EPI_ISL_767491, EPI_ISL_767492, EPI_ISL_767493, EPI_ISL_767494, EPI_ISL_767495, EPI_ISL_767496, EPI_ISL_767497, EPI_ISL_767498, EPI_ISL_767499, EPI_ISL_767500, EPI_ISL_767501, EPI_ISL_767502, EPI_ISL_767503, EPI_ISL_767504, EPI_ISL_767505, EPI_ISL_767506, EPI_ISL_767507, EPI_ISL_767508, EPI_ISL_767509, EPI_ISL_767510, EPI_ISL_767511, EPI_ISL_767512, EPI_ISL_767513, EPI_ISL_767514, EPI_ISL_767515, EPI_ISL_767516, EPI_ISL_767517, EPI_ISL_767518, EPI_ISL_767519, EPI_ISL_767520, EPI_ISL_767521, EPI_ISL_767522, EPI_ISL_767523, EPI_ISL_767524, EPI_ISL_767525, EPI_ISL_767526, EPI_ISL_767527, EPI_ISL_767528, EPI_ISL_767529, EPI_ISL_767530, EPI_ISL_767531, EPI_ISL_767532, EPI_ISL_767533, EPI_ISL_767534, EPI_ISL_767535, EPI_ISL_767536, EPI_ISL_767537 | | | |
| see above | Wadsworth Center, New York State Department.of Health | Wadsworth Center, New York State Department.of Health | Kirsten St. George, Daryl M. Lamson, Alexis Russel, Matthew Shudt, Melissa A Leisner, Jonathan Plitnick, Navjot Singh, John Kelly, Sara Griesemer, Erasmus Schneider, Erica Lasek-Nesselquist |
| EPI_ISL_767538, EPI_ISL_767539, EPI_ISL_767540, EPI_ISL_767541, EPI_ISL_767542, EPI_ISL_767543, EPI_ISL_767544, EPI_ISL_767545, EPI_ISL_767546, EPI_ISL_767547, EPI_ISL_767548, EPI_ISL_767549, EPI_ISL_767550, EPI_ISL_767551, EPI_ISL_767552, EPI_ISL_767553, EPI_ISL_767554, EPI_ISL_767555, EPI_ISL_767556, EPI_ISL_767557, EPI_ISL_767558, EPI_ISL_767559, EPI_ISL_767560, EPI_ISL_767561, EPI_ISL_767562, EPI_ISL_767563, EPI_ISL_767564, EPI_ISL_767565, EPI_ISL_767566, EPI_ISL_767567, EPI_ISL_767568, EPI_ISL_767569, EPI_ISL_767570, EPI_ISL_767571, EPI_ISL_767572, EPI_ISL_767573, EPI_ISL_767574, EPI_ISL_767575, EPI_ISL_767576, EPI_ISL_767577 | | | |
| see above | URMC LABS | Wadsworth Center, New York State Department.of Health | Kirsten St. George, Daryl M. Lamson, Alexis Russel, Matthew Shudt, Melissa A Leisner, Jonathan Plitnick, Navjot Singh, John Kelly, Sara Griesemer, Erasmus Schneider, Erica Lasek-Nesselquist |
| EPI_ISL_767578 | Wadsworth Center, New York State Department.of Health | Wadsworth Center, New York State Department.of Health | Kirsten St. George, Daryl M. Lamson, Alexis Russel, Matthew Shudt, Melissa A Leisner, Jonathan Plitnick, Navjot Singh, John Kelly, Sara Griesemer, Erasmus Schneider, Erica Lasek-Nesselquist |
| EPI_ISL_767579 | URMC LABS | Wadsworth Center, New York State Department.of Health | Kirsten St. George, Daryl M. Lamson, Alexis Russel, Matthew Shudt, Melissa A Leisner, Jonathan Plitnick, Navjot Singh, John Kelly, Sara Griesemer, Erasmus Schneider, Erica Lasek-Nesselquist |
| EPI_ISL_767580, EPI_ISL_767581, EPI_ISL_767582 | WHITE PLAINS HOSPITAL CENTER LABORATORY | Wadsworth Center, New York State Department.of Health | Kirsten St. George, Daryl M. Lamson, Alexis Russel, Matthew Shudt, Melissa A Leisner, Jonathan Plitnick, Navjot Singh, John Kelly, Sara Griesemer, Erasmus Schneider, Erica Lasek-Nesselquist |
| EPI_ISL_767583 | MEMORIAL SLOAN KETTERING CANCER CENTER | Wadsworth Center, New York State Department.of Health | Kirsten St. George, Daryl M. Lamson, Alexis Russel, Matthew Shudt, Melissa A Leisner, Jonathan Plitnick, Navjot Singh, John Kelly, Sara Griesemer, Erasmus Schneider, Erica Lasek-Nesselquist |
| EPI_ISL_767584, EPI_ISL_767585, EPI_ISL_767586, EPI_ISL_767587 | WHITE PLAINS HOSPITAL CENTER LABORATORY | Wadsworth Center, New York State Department.of Health | Kirsten St. George, Daryl M. Lamson, Alexis Russel, Matthew Shudt, Melissa A Leisner, Jonathan Plitnick, Navjot Singh, John Kelly, Sara Griesemer, Erasmus Schneider, Erica Lasek-Nesselquist |
| EPI_ISL_767588 | MEMORIAL SLOAN KETTERING CANCER CENTER | Wadsworth Center, New York State Department.of Health | Kirsten St. George, Daryl M. Lamson, Alexis Russel, Matthew Shudt, Melissa A Leisner, Jonathan Plitnick, Navjot Singh, John Kelly, Sara Griesemer, Erasmus Schneider, Erica Lasek-Nesselquist |
| EPI_ISL_767589 | SARATOGA HOSPITAL LABORATORY | Wadsworth Center, New York State Department.of Health | Kirsten St. George, Daryl M. Lamson, Alexis Russel, Matthew Shudt, Melissa A Leisner, Jonathan Plitnick, Navjot Singh, John Kelly, Sara Griesemer, Erasmus Schneider, Erica Lasek-Nesselquist |
| EPI_ISL_767590 | WHITE PLAINS HOSPITAL CENTER LABORATORY | Wadsworth Center, New York State Department.of Health | Kirsten St. George, Daryl M. Lamson, Alexis Russel, Matthew Shudt, Melissa A Leisner, Jonathan Plitnick, Navjot Singh, John Kelly, Sara Griesemer, Erasmus Schneider, Erica Lasek-Nesselquist |
| EPI_ISL_767591, EPI_ISL_767592, EPI_ISL_767593 | MEMORIAL SLOAN KETTERING CANCER CENTER | Wadsworth Center, New York State Department.of Health | Kirsten St. George, Daryl M. Lamson, Alexis Russel, Matthew Shudt, Melissa A Leisner, Jonathan Plitnick, Navjot Singh, John Kelly, Sara Griesemer, Erasmus Schneider, Erica Lasek-Nesselquist |
| EPI_ISL_767594 | SARATOGA HOSPITAL LABORATORY | Wadsworth Center, New York State Department.of Health | Kirsten St. George, Daryl M. Lamson, Alexis Russel, Matthew Shudt, Melissa A Leisner, Jonathan Plitnick, Navjot Singh, John Kelly, Sara Griesemer, |

| | | | |
|---|---|---|---|
| | | | Erasmus Schneider, Erica Lasek-Nesselquist |
| EPI_ISL_767595 | WHITE PLAINS HOSPITAL CENTER LABORATORY | Wadsworth Center, New York State Department.of Health | Kirsten St. George, Daryl M. Lamson, Alexis Russel, Matthew Shudt, Melissa A Leisner, Jonathan Plitnick, Navjot Singh, John Kelly, Sara Griesemer, Erasmus Schneider, Erica Lasek-Nesselquist |
| EPI_ISL_767596, EPI_ISL_767597 | MEMORIAL SLOAN KETTERING CANCER CENTER | Wadsworth Center, New York State Department.of Health | Kirsten St. George, Daryl M. Lamson, Alexis Russel, Matthew Shudt, Melissa A Leisner, Jonathan Plitnick, Navjot Singh, John Kelly, Sara Griesemer, Erasmus Schneider, Erica Lasek-Nesselquist |
| EPI_ISL_767598 | WHITE PLAINS HOSPITAL CENTER LABORATORY | Wadsworth Center, New York State Department.of Health | Kirsten St. George, Daryl M. Lamson, Alexis Russel, Matthew Shudt, Melissa A Leisner, Jonathan Plitnick, Navjot Singh, John Kelly, Sara Griesemer, Erasmus Schneider, Erica Lasek-Nesselquist |
| EPI_ISL_767599, EPI_ISL_767600, EPI_ISL_767601, EPI_ISL_767602, EPI_ISL_767603, EPI_ISL_767604, EPI_ISL_767605, EPI_ISL_767606, EPI_ISL_767607, EPI_ISL_767608, EPI_ISL_767609, EPI_ISL_767610, EPI_ISL_767611, EPI_ISL_767612, EPI_ISL_767613, EPI_ISL_767614 | | | |
| see above | SARATOGA HOSPITAL LABORATORY | Wadsworth Center, New York State Department.of Health | Kirsten St. George, Daryl M. Lamson, Alexis Russel, Matthew Shudt, Melissa A Leisner, Jonathan Plitnick, Navjot Singh, John Kelly, Sara Griesemer, Erasmus Schneider, Erica Lasek-Nesselquist |
| EPI_ISL_767615, EPI_ISL_767616, EPI_ISL_767617, EPI_ISL_767618, EPI_ISL_767619, EPI_ISL_767620, EPI_ISL_767621, EPI_ISL_767622, EPI_ISL_767623, EPI_ISL_767624, EPI_ISL_767625, EPI_ISL_767626, EPI_ISL_767627, EPI_ISL_767628 | | | |
| see above | WHITE PLAINS HOSPITAL CENTER LABORATORY | Wadsworth Center, New York State Department.of Health | Kirsten St. George, Daryl M. Lamson, Alexis Russel, Matthew Shudt, Melissa A Leisner, Jonathan Plitnick, Navjot Singh, John Kelly, Sara Griesemer, Erasmus Schneider, Erica Lasek-Nesselquist |
| EPI_ISL_767629, EPI_ISL_767630, EPI_ISL_767631, EPI_ISL_767632, EPI_ISL_767633, EPI_ISL_767634, EPI_ISL_767635, EPI_ISL_767636, EPI_ISL_767637, EPI_ISL_767638, EPI_ISL_767639, EPI_ISL_767640, EPI_ISL_767641, EPI_ISL_767642, EPI_ISL_767643, EPI_ISL_767644, EPI_ISL_767645, EPI_ISL_767646, EPI_ISL_767647 | | | |
| see above | MEMORIAL SLOAN KETTERING CANCER CENTER | Wadsworth Center, New York State Department.of Health | Kirsten St. George, Daryl M. Lamson, Alexis Russel, Matthew Shudt, Melissa A Leisner, Jonathan Plitnick, Navjot Singh, John Kelly, Sara Griesemer, Erasmus Schneider, Erica Lasek-Nesselquist |
| EPI_ISL_767648 | BIO-REFERENCE LABORATORIES | Wadsworth Center, New York State Department.of Health | Kirsten St. George, Daryl M. Lamson, Alexis Russel, Matthew Shudt, Melissa A Leisner, Jonathan Plitnick, Navjot Singh, John Kelly, Sara Griesemer, Erasmus Schneider, Erica Lasek-Nesselquist |
| EPI_ISL_767649 | MEMORIAL SLOAN KETTERING CANCER CENTER | Wadsworth Center, New York State Department.of Health | Kirsten St. George, Daryl M. Lamson, Alexis Russel, Matthew Shudt, Melissa A Leisner, Jonathan Plitnick, Navjot Singh, John Kelly, Sara Griesemer, Erasmus Schneider, Erica Lasek-Nesselquist |
| EPI_ISL_767650, EPI_ISL_767651, EPI_ISL_767652, EPI_ISL_767653 | BIO-REFERENCE LABORATORIES | Wadsworth Center, New York State Department.of Health | Kirsten St. George, Daryl M. Lamson, Alexis Russel, Matthew Shudt, Melissa A Leisner, Jonathan Plitnick, Navjot Singh, John Kelly, Sara Griesemer, Erasmus Schneider, Erica Lasek-Nesselquist |
| EPI_ISL_767654, EPI_ISL_767655, EPI_ISL_767656, EPI_ISL_767657, EPI_ISL_767658, EPI_ISL_767659, EPI_ISL_767660, EPI_ISL_767661, EPI_ISL_767662, EPI_ISL_767663, EPI_ISL_767664 | | | |
| see above | MEMORIAL SLOAN KETTERING CANCER CENTER | Wadsworth Center, New York State Department.of Health | Kirsten St. George, Daryl M. Lamson, Alexis Russel, Matthew Shudt, Melissa A Leisner, Jonathan Plitnick, Navjot Singh, John Kelly, Sara Griesemer, Erasmus Schneider, Erica Lasek-Nesselquist |
| EPI_ISL_767665, EPI_ISL_767666, EPI_ISL_767667, EPI_ISL_767668, EPI_ISL_767669, EPI_ISL_767670, EPI_ISL_767671, EPI_ISL_767672, EPI_ISL_767673, EPI_ISL_767674, EPI_ISL_767675, EPI_ISL_767676, EPI_ISL_767677, EPI_ISL_767678, EPI_ISL_767679, EPI_ISL_767680 | | | |
| see above | BIO-REFERENCE LABORATORIES | Wadsworth Center, New York State Department.of Health | Kirsten St. George, Daryl M. Lamson, Alexis Russel, Matthew Shudt, Melissa A Leisner, Jonathan Plitnick, Navjot Singh, John Kelly, Sara Griesemer, Erasmus Schneider, Erica Lasek-Nesselquist |
| EPI_ISL_767681, EPI_ISL_767682, EPI_ISL_767683, EPI_ISL_767684, EPI_ISL_767685, EPI_ISL_767686, EPI_ISL_767687, EPI_ISL_767688, EPI_ISL_767689, EPI_ISL_767690, EPI_ISL_767691, EPI_ISL_767692, EPI_ISL_767693, EPI_ISL_767694, EPI_ISL_767695, EPI_ISL_767696, EPI_ISL_767697, EPI_ISL_767698, EPI_ISL_767699, EPI_ISL_767700, EPI_ISL_767701, EPI_ISL_767702, EPI_ISL_767703, EPI_ISL_767704, EPI_ISL_767705, EPI_ISL_767706, EPI_ISL_767707, EPI_ISL_767708, EPI_ISL_767709, EPI_ISL_767710, EPI_ISL_767711, EPI_ISL_767712, EPI_ISL_767713, EPI_ISL_767714, EPI_ISL_767715 | | | |
| see above | MEMORIAL SLOAN KETTERING CANCER CENTER | Wadsworth Center, New York State Department.of Health | Kirsten St. George, Daryl M. Lamson, Alexis Russel, Matthew Shudt, Melissa A Leisner, Jonathan Plitnick, Navjot Singh, John Kelly, Sara Griesemer, Erasmus Schneider, Erica Lasek-Nesselquist |
| EPI_ISL_769916, EPI_ISL_769917, EPI_ISL_769918, EPI_ISL_769919, EPI_ISL_769920, EPI_ISL_769921, EPI_ISL_769922, EPI_ISL_769923, EPI_ISL_769924, EPI_ISL_769925, EPI_ISL_769926, EPI_ISL_769927, EPI_ISL_769928, EPI_ISL_769929, EPI_ISL_769930, EPI_ISL_769931, EPI_ISL_769932, EPI_ISL_769933, EPI_ISL_769934, EPI_ISL_769935, EPI_ISL_769936, EPI_ISL_769937, EPI_ISL_769938, EPI_ISL_769939, EPI_ISL_769940, EPI_ISL_769941, EPI_ISL_769942, EPI_ISL_769943, EPI_ISL_769944, EPI_ISL_769945, EPI_ISL_769946, EPI_ISL_769947, EPI_ISL_769948, EPI_ISL_769949, EPI_ISL_769950, EPI_ISL_769951, EPI_ISL_769952, EPI_ISL_769953, EPI_ISL_769954, EPI_ISL_769955, EPI_ISL_769956, EPI_ISL_769957, EPI_ISL_769958, EPI_ISL_769959, EPI_ISL_769960, EPI_ISL_769961, EPI_ISL_769962, EPI_ISL_769963, EPI_ISL_769964, EPI_ISL_769965, EPI_ISL_769966, EPI_ISL_769967, EPI_ISL_769968, EPI_ISL_769969, EPI_ISL_769970, EPI_ISL_769971, EPI_ISL_769972, EPI_ISL_769973, EPI_ISL_769974, EPI_ISL_769975, EPI_ISL_769976, EPI_ISL_769977, EPI_ISL_769978, EPI_ISL_769979, EPI_ISL_769980, EPI_ISL_769981, EPI_ISL_769982, EPI_ISL_769983, EPI_ISL_769984, EPI_ISL_769985 | | | |
| see above | Albany Medical Center Hospital Clinical Laboratories | Wadsworth Center, New York State Department.of Health | Kirsten St. George, Daryl M. Lamson, Alexis Russel, Matthew Shudt, Melissa A Leisner, Jonathan Plitnick, Navjot Singh, John Kelly, Sara Griesemer, Erasmus Schneider, Erica Lasek-Nesselquist |
| EPI_ISL_794014, EPI_ISL_794015, EPI_ISL_794016, EPI_ISL_794017, EPI_ISL_794018, EPI_ISL_794019, EPI_ISL_794020, EPI_ISL_794021, EPI_ISL_794022, EPI_ISL_794023, EPI_ISL_794024, EPI_ISL_794025, EPI_ISL_794026, EPI_ISL_794027, EPI_ISL_794028, EPI_ISL_794029, EPI_ISL_794030, EPI_ISL_794031, EPI_ISL_794032, EPI_ISL_794033, EPI_ISL_794034, EPI_ISL_794035, EPI_ISL_794036, EPI_ISL_794037, EPI_ISL_794038, EPI_ISL_794039, EPI_ISL_794040, EPI_ISL_794041, EPI_ISL_794042, EPI_ISL_794043, EPI_ISL_794044, EPI_ISL_794045, EPI_ISL_794046, EPI_ISL_794047, EPI_ISL_794048, EPI_ISL_794049, EPI_ISL_794050, EPI_ISL_794051, EPI_ISL_794052, EPI_ISL_794053, EPI_ISL_794054, EPI_ISL_794055, EPI_ISL_794056, EPI_ISL_794057, EPI_ISL_794058, EPI_ISL_794059, EPI_ISL_794060, EPI_ISL_794061, EPI_ISL_794062, EPI_ISL_794063, EPI_ISL_794064, EPI_ISL_794065, EPI_ISL_794066, EPI_ISL_794067, EPI_ISL_794068, EPI_ISL_794069, EPI_ISL_794070, EPI_ISL_794071, EPI_ISL_794072, EPI_ISL_794073, EPI_ISL_794074, EPI_ISL_794075, EPI_ISL_794076, EPI_ISL_794077, EPI_ISL_794078, EPI_ISL_794079, EPI_ISL_794080, EPI_ISL_794081, EPI_ISL_794082 | | | |
| see above | Wadsworth Center, New York State Department.of Health | Wadsworth Center, New York State Department.of Health | Kirsten St. George, Daryl M. Lamson, Alexis Russel, Matthew Shudt, Melissa A Leisner, Jonathan Plitnick, Navjot Singh, John Kelly, Sara Griesemer, Erasmus Schneider, Erica Lasek-Nesselquist |
| EPI_ISL_794083, EPI_ISL_794084, EPI_ISL_794085, EPI_ISL_794086, EPI_ISL_794087, EPI_ISL_794088, EPI_ISL_794089, EPI_ISL_794090, EPI_ISL_794091, EPI_ISL_794092, EPI_ISL_794093, EPI_ISL_794094, EPI_ISL_794095, EPI_ISL_794096, EPI_ISL_794097, EPI_ISL_794098, EPI_ISL_794099, EPI_ISL_794100, EPI_ISL_794101, EPI_ISL_794102, EPI_ISL_794103, EPI_ISL_794104, EPI_ISL_794105, EPI_ISL_794106 | | | |
| see above | URMC LABS | Wadsworth Center, New York State Department.of Health | Kirsten St. George, Daryl M. Lamson, Alexis Russel, Matthew Shudt, Melissa A Leisner, Jonathan Plitnick, Navjot Singh, John Kelly, Sara Griesemer, Erasmus Schneider, Erica Lasek-Nesselquist |
| EPI_ISL_794107, EPI_ISL_794108, EPI_ISL_794109, EPI_ISL_794110, EPI_ISL_794111, EPI_ISL_794112, EPI_ISL_794113, EPI_ISL_794114, EPI_ISL_794115, EPI_ISL_794116, EPI_ISL_794117, EPI_ISL_794118 | | | |
| see above | GLENS FALLS HOSPITAL LABORATORY | Wadsworth Center, New York State Department.of Health | Kirsten St. George, Daryl M. Lamson, Alexis Russel, Matthew Shudt, Melissa A Leisner, Jonathan Plitnick, Navjot Singh, John Kelly, Sara Griesemer, Erasmus Schneider, Erica Lasek-Nesselquist |
| EPI_ISL_794119, EPI_ISL_794120, EPI_ISL_794121, EPI_ISL_794122, EPI_ISL_794123, EPI_ISL_794124, EPI_ISL_794125, EPI_ISL_794126, EPI_ISL_794127, EPI_ISL_794128, EPI_ISL_794129, EPI_ISL_794130, EPI_ISL_794131, EPI_ISL_794132, EPI_ISL_794133, EPI_ISL_794134, EPI_ISL_794135, EPI_ISL_794136, EPI_ISL_794137, EPI_ISL_794138, EPI_ISL_794139, EPI_ISL_794140, EPI_ISL_794141, EPI_ISL_794142, EPI_ISL_794143, EPI_ISL_794144, EPI_ISL_794145, EPI_ISL_794146, EPI_ISL_794147, EPI_ISL_794148, EPI_ISL_794149, EPI_ISL_794150, EPI_ISL_794151, EPI_ISL_794152, EPI_ISL_794153, EPI_ISL_794154, EPI_ISL_794155, EPI_ISL_794156, EPI_ISL_794157, EPI_ISL_794158 | | | |
| see above | NORTHWELL HEALTH LABORATORIES | Wadsworth Center, New York State Department.of Health | Kirsten St. George, Daryl M. Lamson, Alexis Russel, Matthew Shudt, Melissa A Leisner, Jonathan Plitnick, Navjot Singh, John Kelly, Sara Griesemer, Erasmus Schneider, Erica Lasek-Nesselquist |
| EPI_ISL_794159, EPI_ISL_794160, EPI_ISL_794161, EPI_ISL_794162, EPI_ISL_794163, EPI_ISL_794164, EPI_ISL_794165, EPI_ISL_794166, EPI_ISL_794167, EPI_ISL_794168, EPI_ISL_794169, EPI_ISL_794170, EPI_ISL_794171, EPI_ISL_794172, EPI_ISL_794173, EPI_ISL_794174, EPI_ISL_794175, EPI_ISL_794176, EPI_ISL_794177, EPI_ISL_794178, EPI_ISL_794179, EPI_ISL_794180, EPI_ISL_794181 | | | |
| see above | Wadsworth Center, New York State Department.of Health | Wadsworth Center, New York State Department.of Health | Kirsten St. George, Daryl M. Lamson, Alexis Russel, Matthew Shudt, Melissa A Leisner, Jonathan Plitnick, Navjot Singh, John Kelly, Sara Griesemer, Erasmus Schneider, Erica Lasek-Nesselquist |
| EPI_ISL_794182, EPI_ISL_794183, EPI_ISL_794184, EPI_ISL_794185, EPI_ISL_794186, EPI_ISL_794187, EPI_ISL_794188, EPI_ISL_794189, EPI_ISL_794190, EPI_ISL_794191, EPI_ISL_794192, EPI_ISL_794193, EPI_ISL_794194, EPI_ISL_794195, EPI_ISL_794196, EPI_ISL_794197, EPI_ISL_794198, EPI_ISL_794199, EPI_ISL_794200, EPI_ISL_794201, EPI_ISL_794202, EPI_ISL_794203, EPI_ISL_794204, EPI_ISL_794205 | | | |
| see above | URMC LABS | Wadsworth Center, New York State Department.of Health | Kirsten St. George, Daryl M. Lamson, Alexis Russel, Matthew Shudt, Melissa A Leisner, Jonathan Plitnick, Navjot Singh, John Kelly, Sara Griesemer, Erasmus Schneider, Erica Lasek-Nesselquist |
| EPI_ISL_794206, EPI_ISL_794207 | Wadsworth Center, New York State Department.of Health | Wadsworth Center, New York State Department.of Health | Kirsten St. George, Daryl M. Lamson, Alexis Russel, Matthew Shudt, Melissa A Leisner, Jonathan Plitnick, Navjot Singh, John Kelly, Sara Griesemer, Erasmus Schneider, Erica Lasek-Nesselquist |
| EPI_ISL_794208, EPI_ISL_794209, EPI_ISL_794210, EPI_ISL_794211, EPI_ISL_794212, EPI_ISL_794213, EPI_ISL_794214, EPI_ISL_794215, EPI_ISL_794216, EPI_ISL_794217, EPI_ISL_794218, EPI_ISL_794219, EPI_ISL_794220, EPI_ISL_794221, EPI_ISL_794222, EPI_ISL_794223, EPI_ISL_794224, EPI_ISL_794225, EPI_ISL_794226, EPI_ISL_794227, EPI_ISL_794228, EPI_ISL_794229, EPI_ISL_794230, EPI_ISL_794231, EPI_ISL_794232, EPI_ISL_794233, EPI_ISL_794234, EPI_ISL_794235, EPI_ISL_794236, EPI_ISL_794237, EPI_ISL_794238, EPI_ISL_794239, EPI_ISL_794240, EPI_ISL_794241, EPI_ISL_794242, EPI_ISL_794243, | | | |

| | | | |
|---|---|---|---|
| EPI_ISL_794244, EPI_ISL_794245, EPI_ISL_794246, EPI_ISL_794247, EPI_ISL_794248, EPI_ISL_794249, EPI_ISL_794250, EPI_ISL_794251, EPI_ISL_794252, EPI_ISL_794253, EPI_ISL_794254, EPI_ISL_794255, EPI_ISL_794256, EPI_ISL_794257, EPI_ISL_794258, EPI_ISL_794259, EPI_ISL_794260, EPI_ISL_794261, EPI_ISL_794262, EPI_ISL_794263, EPI_ISL_794264, EPI_ISL_794265, EPI_ISL_794266, EPI_ISL_794267, EPI_ISL_794268, EPI_ISL_794269, EPI_ISL_794270, EPI_ISL_794271, EPI_ISL_794272, EPI_ISL_794273, EPI_ISL_794274, EPI_ISL_794275, EPI_ISL_794276, EPI_ISL_794277, EPI_ISL_794278, EPI_ISL_794279, EPI_ISL_794280, EPI_ISL_794281, EPI_ISL_794282 | | | |
| see above | WESTCHESTER MEDICAL CENTER | Wadsworth Center, New York State Department.of Health | Kirsten St. George, Daryl M. Lamson, Alexis Russel, Matthew Shudt, Melissa A Leisner, Jonathan Plitnick, Navjot Singh, John Kelly, Sara Griesemer, Erasmus Schneider, Erica Lasek-Nesselquist |
| EPI_ISL_794283, EPI_ISL_794284, EPI_ISL_794285, EPI_ISL_794286, EPI_ISL_794287, EPI_ISL_794288, EPI_ISL_794289, EPI_ISL_794290, EPI_ISL_794291, EPI_ISL_794292, EPI_ISL_794293, EPI_ISL_794294, EPI_ISL_794295, EPI_ISL_794296, EPI_ISL_794297, EPI_ISL_794298, EPI_ISL_794299, EPI_ISL_794300, EPI_ISL_794301, EPI_ISL_794302, EPI_ISL_794303, EPI_ISL_794304, EPI_ISL_794305, EPI_ISL_794306, EPI_ISL_794307 | | | |
| see above | Columbia University Irving Medical Center | Wadsworth Center, New York State Department.of Health | Kirsten St. George, Daryl M. Lamson, Alexis Russel, Matthew Shudt, Melissa A Leisner, Jonathan Plitnick, Navjot Singh, John Kelly, Sara Griesemer, Erasmus Schneider, Erica Lasek-Nesselquist |
| EPI_ISL_794308, EPI_ISL_794309, EPI_ISL_794310, EPI_ISL_794311, EPI_ISL_794312, EPI_ISL_794313, EPI_ISL_794314, EPI_ISL_794315, EPI_ISL_794316, EPI_ISL_794317, EPI_ISL_794318, EPI_ISL_794319, EPI_ISL_794320 | | | |
| see above | Wadsworth Center, New York State Department.of Health | Wadsworth Center, New York State Department.of Health | Kirsten St. George, Daryl M. Lamson, Alexis Russel, Matthew Shudt, Melissa A Leisner, Jonathan Plitnick, Navjot Singh, John Kelly, Sara Griesemer, Erasmus Schneider, Erica Lasek-Nesselquist |
| EPI_ISL_801861, EPI_ISL_801862, EPI_ISL_801863, EPI_ISL_801864, EPI_ISL_801865, EPI_ISL_801866, EPI_ISL_801867, EPI_ISL_801868, EPI_ISL_801869, EPI_ISL_801870, EPI_ISL_801871, EPI_ISL_801872, EPI_ISL_801873, EPI_ISL_801874, EPI_ISL_801875, EPI_ISL_801876, EPI_ISL_801877, EPI_ISL_801878, EPI_ISL_801879, EPI_ISL_801880, EPI_ISL_801881, EPI_ISL_801882, EPI_ISL_801883, EPI_ISL_801884, EPI_ISL_801885, EPI_ISL_801886, EPI_ISL_801887, EPI_ISL_801888, EPI_ISL_801889, EPI_ISL_801890, EPI_ISL_801891, EPI_ISL_801892, EPI_ISL_801893, EPI_ISL_801894, EPI_ISL_801895, EPI_ISL_801896, EPI_ISL_801897, EPI_ISL_801898, EPI_ISL_801899, EPI_ISL_801900, EPI_ISL_801901, EPI_ISL_801902, EPI_ISL_801903, EPI_ISL_801904, EPI_ISL_801905, EPI_ISL_801906, EPI_ISL_801907, EPI_ISL_801908, EPI_ISL_801909, EPI_ISL_801910, EPI_ISL_801911, EPI_ISL_801912, EPI_ISL_801913, EPI_ISL_801914, EPI_ISL_801915, EPI_ISL_801916, EPI_ISL_801917, EPI_ISL_801918, EPI_ISL_801919, EPI_ISL_801920, EPI_ISL_801921, EPI_ISL_801922, EPI_ISL_801923, EPI_ISL_801924, EPI_ISL_801925, EPI_ISL_801926, EPI_ISL_801927, EPI_ISL_801928, EPI_ISL_801929, EPI_ISL_801930, EPI_ISL_801931, EPI_ISL_801932, EPI_ISL_801933, EPI_ISL_801934, EPI_ISL_801935, EPI_ISL_801936, EPI_ISL_801937, EPI_ISL_801938, EPI_ISL_801939, EPI_ISL_801940, EPI_ISL_801941, EPI_ISL_801942, EPI_ISL_801943, EPI_ISL_801944, EPI_ISL_801945, EPI_ISL_801946, EPI_ISL_801947, EPI_ISL_801948, EPI_ISL_801949, EPI_ISL_801950, EPI_ISL_801951, EPI_ISL_801952, EPI_ISL_801953, EPI_ISL_801954, EPI_ISL_801955, EPI_ISL_801956, EPI_ISL_801957, EPI_ISL_801958, EPI_ISL_801959, EPI_ISL_801960, EPI_ISL_801961, EPI_ISL_801962, EPI_ISL_801963, EPI_ISL_801964, EPI_ISL_801965, EPI_ISL_801966, EPI_ISL_801967, EPI_ISL_801968, EPI_ISL_801969, EPI_ISL_801970, EPI_ISL_801971, EPI_ISL_801972, EPI_ISL_801973, EPI_ISL_801974, EPI_ISL_801975, EPI_ISL_801976, EPI_ISL_801977, EPI_ISL_801978, EPI_ISL_801979, EPI_ISL_801980, EPI_ISL_801981, EPI_ISL_801982, EPI_ISL_801983, EPI_ISL_801984, EPI_ISL_801985, EPI_ISL_801986, EPI_ISL_801987, EPI_ISL_801988, EPI_ISL_801989, EPI_ISL_801990, EPI_ISL_801991, EPI_ISL_801992, EPI_ISL_801993, EPI_ISL_801994, EPI_ISL_801995, EPI_ISL_801996, EPI_ISL_801997, EPI_ISL_801998, EPI_ISL_801999, EPI_ISL_802000, EPI_ISL_802001, EPI_ISL_802002, EPI_ISL_802003, EPI_ISL_802004, EPI_ISL_802005, EPI_ISL_802006, EPI_ISL_802007, EPI_ISL_802008, EPI_ISL_802009, EPI_ISL_802010, EPI_ISL_802011, EPI_ISL_802012, EPI_ISL_802013, EPI_ISL_802014, EPI_ISL_802015, EPI_ISL_802016, EPI_ISL_802017, EPI_ISL_802018, EPI_ISL_802019, EPI_ISL_802020, EPI_ISL_802021, EPI_ISL_802022, EPI_ISL_802023, EPI_ISL_802024, EPI_ISL_802025, EPI_ISL_802026, EPI_ISL_802027, EPI_ISL_802028, EPI_ISL_802029, EPI_ISL_802030, EPI_ISL_802031, EPI_ISL_802032, EPI_ISL_802033, EPI_ISL_802034, EPI_ISL_802035, EPI_ISL_802036, EPI_ISL_802037, EPI_ISL_802038, EPI_ISL_802039, EPI_ISL_802040, EPI_ISL_802041, EPI_ISL_802042, EPI_ISL_802043, EPI_ISL_802044, EPI_ISL_802045, EPI_ISL_802046, EPI_ISL_802047, EPI_ISL_802048, EPI_ISL_802049, EPI_ISL_802050, EPI_ISL_802051, EPI_ISL_802052, EPI_ISL_802053, EPI_ISL_802054, EPI_ISL_802055, EPI_ISL_802056, EPI_ISL_802057, EPI_ISL_802058, EPI_ISL_802059, EPI_ISL_802060, EPI_ISL_802061, EPI_ISL_802062, EPI_ISL_802063, EPI_ISL_802064, EPI_ISL_802065, EPI_ISL_802066, EPI_ISL_802067, EPI_ISL_802068, EPI_ISL_802069, EPI_ISL_802070, EPI_ISL_802071, EPI_ISL_802072, EPI_ISL_802073, EPI_ISL_802074, EPI_ISL_802075, EPI_ISL_802076, EPI_ISL_802077, EPI_ISL_802078, EPI_ISL_802080, EPI_ISL_802081, EPI_ISL_802082, EPI_ISL_802083, EPI_ISL_802084, EPI_ISL_802085, EPI_ISL_802086, EPI_ISL_802087, EPI_ISL_802088, EPI_ISL_802089, EPI_ISL_802090, EPI_ISL_802091, EPI_ISL_802092, EPI_ISL_802093, EPI_ISL_802094, EPI_ISL_802095, EPI_ISL_802096, EPI_ISL_802097, EPI_ISL_802098, EPI_ISL_802099, EPI_ISL_802100, EPI_ISL_802101, EPI_ISL_802102, EPI_ISL_802104, EPI_ISL_802105, EPI_ISL_802106, EPI_ISL_802107, EPI_ISL_802109, EPI_ISL_802110, EPI_ISL_802111, EPI_ISL_802112, EPI_ISL_802113, EPI_ISL_802114, EPI_ISL_802115, EPI_ISL_802116, EPI_ISL_802117, EPI_ISL_802118, EPI_ISL_802119, EPI_ISL_802120, EPI_ISL_802121, EPI_ISL_802122, EPI_ISL_802123, EPI_ISL_802124, EPI_ISL_802125, EPI_ISL_802126, EPI_ISL_802127, EPI_ISL_802128, EPI_ISL_802129, EPI_ISL_802130, EPI_ISL_802131, EPI_ISL_802132, EPI_ISL_802133, EPI_ISL_802134, EPI_ISL_802135, EPI_ISL_802137, EPI_ISL_802138, EPI_ISL_802139, EPI_ISL_802140, EPI_ISL_802141, EPI_ISL_802142, EPI_ISL_802143, EPI_ISL_802144, EPI_ISL_802145, EPI_ISL_802146, EPI_ISL_802147, EPI_ISL_802149, EPI_ISL_802150, EPI_ISL_802151, EPI_ISL_802152, EPI_ISL_802153, EPI_ISL_802154, EPI_ISL_802155, EPI_ISL_802156, EPI_ISL_802157, EPI_ISL_802158, EPI_ISL_802159, EPI_ISL_802160, EPI_ISL_802161, EPI_ISL_802162, EPI_ISL_802163, EPI_ISL_802164, EPI_ISL_802165, EPI_ISL_802166, EPI_ISL_802167, EPI_ISL_802168, EPI_ISL_802169, EPI_ISL_802170, EPI_ISL_802171, EPI_ISL_802172, EPI_ISL_802173, EPI_ISL_802174, EPI_ISL_802175, EPI_ISL_802176, EPI_ISL_802177, EPI_ISL_802178, EPI_ISL_802179, EPI_ISL_802180, EPI_ISL_802181, EPI_ISL_802182, EPI_ISL_802183, EPI_ISL_802184, EPI_ISL_802185, EPI_ISL_802186, EPI_ISL_802187, EPI_ISL_802189, EPI_ISL_802190, EPI_ISL_802191, EPI_ISL_802192, EPI_ISL_802193, EPI_ISL_802194, EPI_ISL_802195, EPI_ISL_802196, EPI_ISL_802197, EPI_ISL_802198, EPI_ISL_802199, EPI_ISL_802200, EPI_ISL_802201, EPI_ISL_802203, EPI_ISL_802204, EPI_ISL_802205, EPI_ISL_802206, EPI_ISL_802207, EPI_ISL_802208, EPI_ISL_802209, EPI_ISL_802210, EPI_ISL_802211, EPI_ISL_802212, EPI_ISL_802213, EPI_ISL_802214, EPI_ISL_802215, EPI_ISL_802216, EPI_ISL_802217, EPI_ISL_802218, EPI_ISL_802219, EPI_ISL_802220, EPI_ISL_802221, EPI_ISL_802222, EPI_ISL_802223, EPI_ISL_802224, EPI_ISL_802225, EPI_ISL_802226, EPI_ISL_802227, EPI_ISL_802228, EPI_ISL_802229, EPI_ISL_802230, EPI_ISL_802231, EPI_ISL_802232, EPI_ISL_802234, EPI_ISL_802235, EPI_ISL_802236, EPI_ISL_802237, EPI_ISL_802238, EPI_ISL_802239, EPI_ISL_802240, EPI_ISL_802241, EPI_ISL_802242, EPI_ISL_802243, EPI_ISL_802245, EPI_ISL_802246, EPI_ISL_802247, EPI_ISL_802248, EPI_ISL_802249, EPI_ISL_802250, EPI_ISL_802251, EPI_ISL_802252, EPI_ISL_802253, EPI_ISL_802254, EPI_ISL_802255, EPI_ISL_802256, EPI_ISL_802257, EPI_ISL_802259, EPI_ISL_802260, EPI_ISL_802261, EPI_ISL_802262, EPI_ISL_802263, EPI_ISL_802264, EPI_ISL_802265, EPI_ISL_802266, EPI_ISL_802267, EPI_ISL_802268, EPI_ISL_802269, EPI_ISL_802270, EPI_ISL_802271, EPI_ISL_802272, EPI_ISL_802273, EPI_ISL_802274, EPI_ISL_802276, EPI_ISL_802277, EPI_ISL_802278, EPI_ISL_802279, EPI_ISL_802280, EPI_ISL_802281, EPI_ISL_802282, EPI_ISL_802283, EPI_ISL_802284, EPI_ISL_802285, EPI_ISL_802286, EPI_ISL_802287, EPI_ISL_802288, EPI_ISL_802289, EPI_ISL_802290, EPI_ISL_802291, EPI_ISL_802292, EPI_ISL_802293, EPI_ISL_802294, EPI_ISL_802295, EPI_ISL_802296, EPI_ISL_802297, EPI_ISL_802298, EPI_ISL_802299, EPI_ISL_802300, EPI_ISL_802301, EPI_ISL_802302, EPI_ISL_802303, EPI_ISL_802304, EPI_ISL_802305, EPI_ISL_802306, EPI_ISL_802307, EPI_ISL_802308, EPI_ISL_802309, EPI_ISL_802310, EPI_ISL_802311, EPI_ISL_802312, EPI_ISL_802313, EPI_ISL_802314, EPI_ISL_802315, EPI_ISL_802316, EPI_ISL_802317, EPI_ISL_802318, EPI_ISL_802319, EPI_ISL_802320, EPI_ISL_802321, EPI_ISL_802322, EPI_ISL_802323, EPI_ISL_802324, EPI_ISL_802325, EPI_ISL_802326, EPI_ISL_802327, EPI_ISL_802328, EPI_ISL_802329, EPI_ISL_802330, EPI_ISL_802331, EPI_ISL_802332, EPI_ISL_802333, EPI_ISL_802334, EPI_ISL_802335, EPI_ISL_802336, EPI_ISL_802337, EPI_ISL_802338, EPI_ISL_802339, EPI_ISL_802340, EPI_ISL_802342, EPI_ISL_802343, EPI_ISL_802344, EPI_ISL_802345, EPI_ISL_802346, EPI_ISL_802347, EPI_ISL_802348, EPI_ISL_802349, EPI_ISL_802350, EPI_ISL_802351, EPI_ISL_802352, EPI_ISL_802353, EPI_ISL_802354, EPI_ISL_802355, EPI_ISL_802356, EPI_ISL_802358, EPI_ISL_802359, EPI_ISL_802360, EPI_ISL_802361, EPI_ISL_802362, EPI_ISL_802363, EPI_ISL_802364, EPI_ISL_802365, EPI_ISL_802366, EPI_ISL_802367, EPI_ISL_802368, EPI_ISL_802369, EPI_ISL_802370, EPI_ISL_802371, EPI_ISL_802372, EPI_ISL_802373, EPI_ISL_802374, EPI_ISL_802375, EPI_ISL_802376, EPI_ISL_802377, EPI_ISL_802378, EPI_ISL_802379, EPI_ISL_802380, EPI_ISL_802381, EPI_ISL_802382, EPI_ISL_802383, EPI_ISL_802384, EPI_ISL_802385, EPI_ISL_802386, EPI_ISL_802387, EPI_ISL_802388, EPI_ISL_802389, EPI_ISL_802391, EPI_ISL_802394, EPI_ISL_802395, EPI_ISL_802396, EPI_ISL_802397, EPI_ISL_802398, EPI_ISL_802399, EPI_ISL_802400, EPI_ISL_802401, EPI_ISL_802402, EPI_ISL_802403, EPI_ISL_802404, EPI_ISL_802405, EPI_ISL_802406, EPI_ISL_802407, EPI_ISL_802408, EPI_ISL_802409, EPI_ISL_802410, EPI_ISL_802411, EPI_ISL_802412, EPI_ISL_802413, EPI_ISL_802414, EPI_ISL_802415, EPI_ISL_802416, EPI_ISL_802418, EPI_ISL_802419, EPI_ISL_802420, EPI_ISL_802421, EPI_ISL_802422, EPI_ISL_802423, EPI_ISL_802426, EPI_ISL_802427 | | | |
| see above | MSHS Clinical Microbiology Laboratories | MSHS Pathogen Surveillance Program | Ana S. Gonzalez-Reiche, Hala Alshammary, Mitchell J. Sullivan, Brianne Ciferri, Ajay Obla, Angela Amoako, Mahmoud Awawda, Elena Hirsch, Ashley S. Salimbangon, Levy Sominsky, Katherine Beach, Kayla Russo, Charles Gleason, Shelcie Fabre, Giulio Kleiner, Zenab Khan, Bremy Alburquerque, Adriana van de Guchte, Komal Srivastava, Matthew M. Hernandez, Jayeeta Dutta, Denise Jurczyszak, Emily Ferreri, Rachel Chernet, Nancy Francoeur, Betsaida Salom Melo, Irina Oussenko, Gintaras Deikus, Juan Soto, Shwetha Hara Sridhar, Ying-Chih Wang, Kathryn Twyman, Andrew Kasarskis, Deena R. Altman, Robert Sebra, Adolfo García-Sastre, Marta Luksza, Gopi Patel, Sarah Schaefer, Melissa Gitman, Michael D. Nowak, Alberto Paniz-Mondolfi, Emilia Mia Sordillo, Viviana Simon, Harm van Bakel |
| EPI_ISL_802428 | SARATOGA HOSPITAL LABORATORY | Wadsworth Center, New York State Department.of Health | Kirsten St. George, Daryl M. Lamson, Alexis Russel, Matthew Shudt, Melissa A Leisner, Jonathan Plitnick, Navjot Singh, John Kelly, Sara Griesemer, Erasmus Schneider, Erica Lasek-Nesselquist |
| EPI_ISL_802429 | Wadsworth Center, New York State Department.of Health | Wadsworth Center, New York State Department.of Health | Kirsten St. George, Daryl M. Lamson, Alexis Russel, Matthew Shudt, Melissa A Leisner, Jonathan Plitnick, Navjot Singh, John Kelly, Sara Griesemer, Erasmus Schneider, Erica Lasek-Nesselquist |
| EPI_ISL_802430 | BIO-REFERENCE LABORATORIES | Wadsworth Center, New York State Department.of Health | Kirsten St. George, Daryl M. Lamson, Alexis Russel, Matthew Shudt, Melissa A Leisner, Jonathan Plitnick, Navjot Singh, John Kelly, Sara Griesemer, Erasmus Schneider, Erica Lasek-Nesselquist |
| EPI_ISL_802431 | Wadsworth Center, New York State Department.of Health | Wadsworth Center, New York State Department.of Health | Kirsten St. George, Daryl M. Lamson, Alexis Russel, Matthew Shudt, Melissa A Leisner, Jonathan Plitnick, Navjot Singh, John Kelly, Sara Griesemer, Erasmus Schneider, Erica Lasek-Nesselquist |
| EPI_ISL_802432 | SARATOGA HOSPITAL LABORATORY | Wadsworth Center, New York State Department.of Health | Kirsten St. George, Daryl M. Lamson, Alexis Russel, Matthew Shudt, Melissa A Leisner, Jonathan Plitnick, Navjot Singh, John Kelly, Sara Griesemer, Erasmus Schneider, Erica Lasek-Nesselquist |
| EPI_ISL_802433 | Wadsworth Center, New York State Department.of Health | Wadsworth Center, New York State Department.of Health | Kirsten St. George, Daryl M. Lamson, Alexis Russel, Matthew Shudt, Melissa A Leisner, Jonathan Plitnick, Navjot Singh, John Kelly, Sara Griesemer, Erasmus Schneider, Erica Lasek-Nesselquist |
| EPI_ISL_802434, EPI_ISL_802435 | WESTCHESTER MEDICAL CENTER | Wadsworth Center, New York State Department.of Health | Kirsten St. George, Daryl M. Lamson, Alexis Russel, Matthew Shudt, Melissa A Leisner, Jonathan Plitnick, Navjot Singh, John Kelly, Sara Griesemer, Erasmus Schneider, Erica Lasek-Nesselquist |
| EPI_ISL_802436 | TEMPUS LABS INC | Wadsworth Center, New York State Department.of Health | Kirsten St. George, Daryl M. Lamson, Alexis Russel, Matthew Shudt, Melissa A Leisner, Jonathan Plitnick, Navjot Singh, John Kelly, Sara Griesemer, Erasmus Schneider, Erica Lasek-Nesselquist |
| EPI_ISL_802437 | SARATOGA HOSPITAL LABORATORY | Wadsworth Center, New York State Department.of Health | Kirsten St. George, Daryl M. Lamson, Alexis Russel, Matthew Shudt, Melissa A Leisner, Jonathan Plitnick, Navjot Singh, John Kelly, Sara Griesemer, Erasmus Schneider, Erica Lasek-Nesselquist |
| EPI_ISL_802438 | Wadsworth Center, New York State Department.of Health | Wadsworth Center, New York State Department.of Health | Kirsten St. George, Daryl M. Lamson, Alexis Russel, Matthew Shudt, Melissa A Leisner, Jonathan Plitnick, Navjot Singh, John Kelly, Sara Griesemer, Erasmus Schneider, Erica Lasek-Nesselquist |
| EPI_ISL_802439 | SARATOGA HOSPITAL LABORATORY | Wadsworth Center, New York State Department.of Health | Kirsten St. George, Daryl M. Lamson, Alexis Russel, Matthew Shudt, Melissa A Leisner, Jonathan Plitnick, Navjot Singh, John Kelly, Sara Griesemer, Erasmus Schneider, Erica Lasek-Nesselquist |
| EPI_ISL_802440, EPI_ISL_802441 | WESTCHESTER MEDICAL CENTER | Wadsworth Center, New York State Department.of Health | Kirsten St. George, Daryl M. Lamson, Alexis Russel, Matthew Shudt, Melissa A Leisner, Jonathan Plitnick, Navjot Singh, John Kelly, Sara Griesemer, Erasmus Schneider, Erica Lasek-Nesselquist |
| EPI_ISL_802442 | Wadsworth Center, New York State Department.of Health | Wadsworth Center, New York State Department.of Health | Kirsten St. George, Daryl M. Lamson, Alexis Russel, Matthew Shudt, Melissa A Leisner, Jonathan Plitnick, Navjot Singh, John Kelly, Sara Griesemer, Erasmus Schneider, Erica Lasek-Nesselquist |
| EPI_ISL_802443 | WESTCHESTER MEDICAL CENTER | Wadsworth Center, New York State Department.of Health | Kirsten St. George, Daryl M. Lamson, Alexis Russel, Matthew Shudt, Melissa A Leisner, Jonathan Plitnick, Navjot Singh, John Kelly, Sara Griesemer, Erasmus Schneider, Erica Lasek-Nesselquist |

| | | | |
|---|---|---|---|
| EPI_ISL_802444 | SUNRISE MEDICAL LABORATORIES | Wadsworth Center, New York State Department.of Health | Kirsten St. George, Daryl M. Lamson, Alexis Russel, Matthew Shudt, Melissa A Leisner, Jonathan Plitnick, Navjot Singh, John Kelly, Sara Griesemer, Erasmus Schneider, Erica Lasek-Nesselquist |
| EPI_ISL_802445, EPI_ISL_802446, EPI_ISL_802447 | BIO-REFERENCE LABORATORIES | Wadsworth Center, New York State Department.of Health | Kirsten St. George, Daryl M. Lamson, Alexis Russel, Matthew Shudt, Melissa A Leisner, Jonathan Plitnick, Navjot Singh, John Kelly, Sara Griesemer, Erasmus Schneider, Erica Lasek-Nesselquist |
| EPI_ISL_802448 | WESTCHESTER MEDICAL CENTER | Wadsworth Center, New York State Department.of Health | Kirsten St. George, Daryl M. Lamson, Alexis Russel, Matthew Shudt, Melissa A Leisner, Jonathan Plitnick, Navjot Singh, John Kelly, Sara Griesemer, Erasmus Schneider, Erica Lasek-Nesselquist |
| EPI_ISL_802449, EPI_ISL_802450, EPI_ISL_802451, EPI_ISL_802452, EPI_ISL_802453, EPI_ISL_802454, EPI_ISL_802455, EPI_ISL_802456, EPI_ISL_802457, EPI_ISL_802458, EPI_ISL_802459, EPI_ISL_802460, EPI_ISL_802461, EPI_ISL_802462, EPI_ISL_802463, EPI_ISL_802464, EPI_ISL_802465, EPI_ISL_802466 | | | |
| see above | Wadsworth Center, New York State Department.of Health | Wadsworth Center, New York State Department.of Health | Kirsten St. George, Daryl M. Lamson, Alexis Russel, Matthew Shudt, Melissa A Leisner, Jonathan Plitnick, Navjot Singh, John Kelly, Sara Griesemer, Erasmus Schneider, Erica Lasek-Nesselquist |
| EPI_ISL_802467, EPI_ISL_802468 | SARATOGA HOSPITAL LABORATORY | Wadsworth Center, New York State Department.of Health | Kirsten St. George, Daryl M. Lamson, Alexis Russel, Matthew Shudt, Melissa A Leisner, Jonathan Plitnick, Navjot Singh, John Kelly, Sara Griesemer, Erasmus Schneider, Erica Lasek-Nesselquist |
| EPI_ISL_802469, EPI_ISL_802470, EPI_ISL_802471, EPI_ISL_802472, EPI_ISL_802473, EPI_ISL_802474, EPI_ISL_802475, EPI_ISL_802476, EPI_ISL_802477, EPI_ISL_802478, EPI_ISL_802479, EPI_ISL_802480, EPI_ISL_802481, EPI_ISL_802482, EPI_ISL_802483, EPI_ISL_802484, EPI_ISL_802485, EPI_ISL_802486, EPI_ISL_802487, EPI_ISL_802488, EPI_ISL_802489, EPI_ISL_802490 | | | |
| see above | WESTCHESTER MEDICAL CENTER | Wadsworth Center, New York State Department.of Health | Kirsten St. George, Daryl M. Lamson, Alexis Russel, Matthew Shudt, Melissa A Leisner, Jonathan Plitnick, Navjot Singh, John Kelly, Sara Griesemer, Erasmus Schneider, Erica Lasek-Nesselquist |
| EPI_ISL_802491, EPI_ISL_802492, EPI_ISL_802493, EPI_ISL_802494, EPI_ISL_802495, EPI_ISL_802496, EPI_ISL_802497 | SARATOGA HOSPITAL LABORATORY | Wadsworth Center, New York State Department.of Health | Kirsten St. George, Daryl M. Lamson, Alexis Russel, Matthew Shudt, Melissa A Leisner, Jonathan Plitnick, Navjot Singh, John Kelly, Sara Griesemer, Erasmus Schneider, Erica Lasek-Nesselquist |
| EPI_ISL_802498, EPI_ISL_802499, EPI_ISL_802500 | Wadsworth Center, New York State Department.of Health | Wadsworth Center, New York State Department.of Health | Kirsten St. George, Daryl M. Lamson, Alexis Russel, Matthew Shudt, Melissa A Leisner, Jonathan Plitnick, Navjot Singh, John Kelly, Sara Griesemer, Erasmus Schneider, Erica Lasek-Nesselquist |
| EPI_ISL_802777, EPI_ISL_802778, EPI_ISL_802779, EPI_ISL_802780, EPI_ISL_802781, EPI_ISL_802782, EPI_ISL_802783, EPI_ISL_802784, EPI_ISL_802785, EPI_ISL_802786, EPI_ISL_802787, EPI_ISL_802788, EPI_ISL_802789 | | | |
| see above | Columbia University Irving Medical Center | Wadsworth Center, New York State Department.of Health | Kirsten St. George, Daryl M. Lamson, Alexis Russel, Matthew Shudt, Melissa A Leisner, Jonathan Plitnick, Navjot Singh, John Kelly, Sara Griesemer, Erasmus Schneider, Erica Lasek-Nesselquist |
| EPI_ISL_802790, EPI_ISL_802791, EPI_ISL_802792, EPI_ISL_802793 | BIO-REFERENCE LABORATORIES | Wadsworth Center, New York State Department.of Health | Kirsten St. George, Daryl M. Lamson, Alexis Russel, Matthew Shudt, Melissa A Leisner, Jonathan Plitnick, Navjot Singh, John Kelly, Sara Griesemer, Erasmus Schneider, Erica Lasek-Nesselquist |
| EPI_ISL_804945 | Wadsworth Center, New York State Department.of Health | Wadsworth Center, New York State Department.of Health | Kirsten St. George, Daryl M. Lamson, Alexis Russel, Matthew Shudt, Melissa A Leisner, Jonathan Plitnick, Navjot Singh, John Kelly, Sara Griesemer, Erasmus Schneider, Erica Lasek-Nesselquist |
| EPI_ISL_804946 | BIO-REFERENCE LABORATORIES | Wadsworth Center, New York State Department.of Health | Kirsten St. George, Daryl M. Lamson, Alexis Russel, Matthew Shudt, Melissa A Leisner, Jonathan Plitnick, Navjot Singh, John Kelly, Sara Griesemer, Erasmus Schneider, Erica Lasek-Nesselquist |
| EPI_ISL_804947, EPI_ISL_804948 | Wadsworth Center, New York State Department.of Health | Wadsworth Center, New York State Department.of Health | Kirsten St. George, Daryl M. Lamson, Alexis Russel, Matthew Shudt, Melissa A Leisner, Jonathan Plitnick, Navjot Singh, John Kelly, Sara Griesemer, Erasmus Schneider, Erica Lasek-Nesselquist |
| EPI_ISL_804980 | Columbia University Irving Medical Center | Wadsworth Center, New York State Department of Health | Kirsten St. George, Daryl M. Lamson, Alexis Russel, Matthew Shudt, Melissa A Leisner, Jonathan Plitnick, Navjot Singh, John Kelly, Erasmus Schneider, Erica Lasek-Nesselquist |
| EPI_ISL_804981, EPI_ISL_804982, EPI_ISL_804983, EPI_ISL_804984 | BIO-REFERENCE LABORATORIES | Wadsworth Center, New York State Department of Health | Kirsten St. George, Daryl M. Lamson, Alexis Russel, Matthew Shudt, Melissa A Leisner, Jonathan Plitnick, Navjot Singh, John Kelly, Erasmus Schneider, Erica Lasek-Nesselquist |
| EPI_ISL_812201 | NYC Department of Health and Mental Hygiene | Pathogen Discovery, Respiratory Viruses Branch, Division of Viral Diseases, Centers for Disease Control and Prevention | Yan Li, Ying Tao, Anna Montmayeur, Jing Zhang, Brian Lynch, Krista Queen, Anna Uehara, Rachel Marine, Peter Cook, Clinton R. Paden, Haibin Wang, Suxiang Tong |
| EPI_ISL_812722, EPI_ISL_812723, EPI_ISL_812724 | DOHMH PHL | New York City Public Health Laboratory | Jade Wang, et al. |
| EPI_ISL_812725 | DOHMH Crown Heights | New York City Public Health Laboratory | Jade Wang, et al. |
| EPI_ISL_812726 | DOHMH Corona | New York City Public Health Laboratory | Jade Wang, et al. |
| EPI_ISL_812727 | DOHMH Jamaica | New York City Public Health Laboratory | Jade Wang, et al. |
| EPI_ISL_812728 | DOHMH Riverside | New York City Public Health Laboratory | Jade Wang, et al. |
| EPI_ISL_812729 | DOHMH Central Harlem | New York City Public Health Laboratory | Jade Wang, et al. |
| EPI_ISL_812730 | DOHMH Crown Heights | New York City Public Health Laboratory | Jade Wang, et al. |
| EPI_ISL_812731 | DOHMH Jamaica | New York City Public Health Laboratory | Jade Wang, et al. |
| EPI_ISL_812732, EPI_ISL_812733 | DOHMH Corona | New York City Public Health Laboratory | Jade Wang, et al. |
| EPI_ISL_812734 | DOHMH Crown Heights | New York City Public Health Laboratory | Jade Wang, et al. |
| EPI_ISL_812735, EPI_ISL_812736, EPI_ISL_812737 | DOHMH Morrisania | New York City Public Health Laboratory | Jade Wang, et al. |
| EPI_ISL_812738 | DOHMH Crown Heights | New York City Public Health Laboratory | Jade Wang, et al. |
| EPI_ISL_812739 | DOHMH Central Harlem | New York City Public Health Laboratory | Jade Wang, et al. |
| EPI_ISL_812740 | DOHMH Corona | New York City Public Health Laboratory | Jade Wang, et al. |
| EPI_ISL_812741 | DOHMH Central Harlem | New York City Public Health Laboratory | Jade Wang, et al. |
| EPI_ISL_812742, EPI_ISL_812743, EPI_ISL_812744 | DOHMH Corona | New York City Public Health Laboratory | Jade Wang, et al. |
| EPI_ISL_812745 | DOHMH Riverside | New York City Public Health Laboratory | Jade Wang, et al. |
| EPI_ISL_812746, EPI_ISL_812747, EPI_ISL_812748 | DOHMH Corona | New York City Public Health Laboratory | Jade Wang, et al. |
| EPI_ISL_812749 | DOHMH Central Harlem | New York City Public Health Laboratory | Jade Wang, et al. |
| EPI_ISL_812750, EPI_ISL_812751 | DOHMH Corona | New York City Public Health Laboratory | Jade Wang, et al. |
| EPI_ISL_812752 | DOHMH Crown Heights | New York City Public Health Laboratory | Jade Wang, et al. |
| EPI_ISL_812753, EPI_ISL_812754, EPI_ISL_812755, EPI_ISL_812756, EPI_ISL_812757 | DOHMH Morrisania | New York City Public Health Laboratory | Jade Wang, et al. |
| EPI_ISL_812758 | DOHMH Crown Heights | New York City Public Health Laboratory | Jade Wang, et al. |

| | | | |
|---|---|---|---|
| EPI_ISL_812759 | DOHMH Riverside | New York City Public Health Laboratory | Jade Wang, et al. |
| EPI_ISL_812760 | DOHMH Corona | New York City Public Health Laboratory | Jade Wang, et al. |
| EPI_ISL_823813 | DOHMH Jamaica | New York City Public Health Laboratory | Jade Wang, et al. |
| EPI_ISL_823814 | DOHMH Riverside | New York City Public Health Laboratory | Jade Wang, et al. |
| EPI_ISL_823815, EPI_ISL_823816 | DOHMH Chelsea | New York City Public Health Laboratory | Jade Wang, et al. |
| EPI_ISL_823817, EPI_ISL_823818, EPI_ISL_823819 | DOHMH Corona | New York City Public Health Laboratory | Jade Wang, et al. |
| EPI_ISL_823820 | DOHMH PHL | New York City Public Health Laboratory | Jade Wang, et al. |
| EPI_ISL_823821, EPI_ISL_823822 | DOHMH Jamaica | New York City Public Health Laboratory | Jade Wang, et al. |
| EPI_ISL_823823, EPI_ISL_823824, EPI_ISL_823825 | DOHMH Riverside | New York City Public Health Laboratory | Jade Wang, et al. |
| EPI_ISL_823826, EPI_ISL_823827, EPI_ISL_823828 | DOHMH Corona | New York City Public Health Laboratory | Jade Wang, et al. |
| EPI_ISL_823829, EPI_ISL_823830, EPI_ISL_823831 | DOHMH Morrisania | New York City Public Health Laboratory | Jade Wang, et al. |
| EPI_ISL_823832, EPI_ISL_823833, EPI_ISL_823834 | DOHMH Central Harlem | New York City Public Health Laboratory | Jade Wang, et al. |
| EPI_ISL_823835, EPI_ISL_823836, EPI_ISL_823837 | DOHMH Corona | New York City Public Health Laboratory | Jade Wang, et al. |
| EPI_ISL_823838, EPI_ISL_823839, EPI_ISL_823840 | DOHMH Central Harlem | New York City Public Health Laboratory | Jade Wang, et al. |
| EPI_ISL_823841 | DOHMH Riverside | New York City Public Health Laboratory | Jade Wang, et al. |
| EPI_ISL_823842 | DOHMH PHL | New York City Public Health Laboratory | Jade Wang, et al. |
| EPI_ISL_823843, EPI_ISL_823844 | DOHMH Central Harlem | New York City Public Health Laboratory | Jade Wang, et al. |
| EPI_ISL_823845 | DOHMH Morrisania | New York City Public Health Laboratory | Jade Wang, et al. |
| EPI_ISL_823846, EPI_ISL_823847, EPI_ISL_823848 | DOHMH Jamaica | New York City Public Health Laboratory | Jade Wang, et al. |
| EPI_ISL_823849 | DOHMH Fort Greene | New York City Public Health Laboratory | Jade Wang, et al. |
| EPI_ISL_823850 | DOHMH Corona | New York City Public Health Laboratory | Jade Wang, et al. |
| EPI_ISL_823851 | DOHMH Fort Greene | New York City Public Health Laboratory | Jade Wang, et al. |
| EPI_ISL_823852, EPI_ISL_823853 | DOHMH Crown Heights | New York City Public Health Laboratory | Jade Wang, et al. |
| EPI_ISL_823854, EPI_ISL_823855, EPI_ISL_823856, EPI_ISL_823857, EPI_ISL_823858, EPI_ISL_823859, EPI_ISL_823860 | DOHMH Morrisania | New York City Public Health Laboratory | Jade Wang, et al. |
| EPI_ISL_823861, EPI_ISL_823862, EPI_ISL_823863 | DOHMH Corona | New York City Public Health Laboratory | Jade Wang, et al. |
| EPI_ISL_823864 | DOHMH Chelsea | New York City Public Health Laboratory | Jade Wang, et al. |
| EPI_ISL_823865 | DOHMH Central Harlem | New York City Public Health Laboratory | Jade Wang, et al. |
| EPI_ISL_823866, EPI_ISL_823867 | DOHMH Fort Greene | New York City Public Health Laboratory | Jade Wang, et al. |
| EPI_ISL_823868 | DOHMH Corona | New York City Public Health Laboratory | Jade Wang, et al. |
| EPI_ISL_823869 | DOHMH Central Harlem | New York City Public Health Laboratory | Jade Wang, et al. |
| EPI_ISL_823870, EPI_ISL_823871 | DOHMH Jamaica | New York City Public Health Laboratory | Jade Wang, et al. |
| EPI_ISL_823872, EPI_ISL_823873, EPI_ISL_823874 | DOHMH Morrisania | New York City Public Health Laboratory | Jade Wang, et al. |
| EPI_ISL_823875 | DOHMH PHL | New York City Public Health Laboratory | Jade Wang, et al. |
| EPI_ISL_823876, EPI_ISL_823877 | DOHMH Jamaica | New York City Public Health Laboratory | Jade Wang, et al. |
| EPI_ISL_823878, EPI_ISL_823879, EPI_ISL_823880, EPI_ISL_823881, EPI_ISL_823882 | DOHMH Corona | New York City Public Health Laboratory | Jade Wang, et al. |
| EPI_ISL_823883 | DOHMH Riverside | New York City Public Health Laboratory | Jade Wang, et al. |
| EPI_ISL_823884, EPI_ISL_823885, EPI_ISL_823886, EPI_ISL_823887 | DOHMH Central Harlem | New York City Public Health Laboratory | Jade Wang, et al. |
| EPI_ISL_823888 | DOHMH Riverside | New York City Public Health Laboratory | Jade Wang, et al. |
| EPI_ISL_823889, EPI_ISL_823890, EPI_ISL_823891 | DOHMH PHL | New York City Public Health Laboratory | Jade Wang, et al. |
| EPI_ISL_823892, EPI_ISL_823893 | DOHMH Morrisania | New York City Public Health Laboratory | Jade Wang, et al. |
| EPI_ISL_823894, EPI_ISL_823895 | DOHMH Jamaica | New York City Public Health Laboratory | Jade Wang, et al. |
| EPI_ISL_823896, EPI_ISL_823897 | DOHMH PHL | New York City Public Health Laboratory | Jade Wang, et al. |
| EPI_ISL_823898, EPI_ISL_823899 | DOHMH Corona | New York City Public Health Laboratory | Jade Wang, et al. |
| EPI_ISL_823900 | DOHMH Riverside | New York City Public Health Laboratory | Jade Wang, et al. |
| EPI_ISL_823901, EPI_ISL_823902 | DOHMH Central Harlem | New York City Public Health Laboratory | Jade Wang, et al. |
| EPI_ISL_823903 | DOHMH Morrisania | New York City Public Health Laboratory | Jade Wang, et al. |
| EPI_ISL_823904 | DOHMH Fort Greene | New York City Public Health Laboratory | Jade Wang, et al. |

| | | | |
|---|---|---|---|
| EPI_ISL_823905, EPI_ISL_823906 | DOHMH Corona | New York City Public Health Laboratory | Jade Wang, et al. |
| EPI_ISL_823907, EPI_ISL_823908 | DOHMH Jamaica | New York City Public Health Laboratory | Jade Wang, et al. |
| EPI_ISL_823909 | DOHMH PHL | New York City Public Health Laboratory | Jade Wang, et al. |
| EPI_ISL_823910, EPI_ISL_823911 | DOHMH Fort Greene | New York City Public Health Laboratory | Jade Wang, et al. |
| EPI_ISL_823912 | DOHMH Central Harlem | New York City Public Health Laboratory | Jade Wang, et al. |
| EPI_ISL_823913, EPI_ISL_823914 | DOHMH PHL | New York City Public Health Laboratory | Jade Wang, et al. |
| EPI_ISL_823915, EPI_ISL_823916, EPI_ISL_823917 | DOHMH Jamaica | New York City Public Health Laboratory | Jade Wang, et al. |
| EPI_ISL_823918, EPI_ISL_823919, EPI_ISL_823920, EPI_ISL_823921, EPI_ISL_823922 | DOHMH Corona | New York City Public Health Laboratory | Jade Wang, et al. |
| EPI_ISL_823923 | DOHMH Fort Greene | New York City Public Health Laboratory | Jade Wang, et al. |
| EPI_ISL_823924 | DOHMH Chelsea | New York City Public Health Laboratory | Jade Wang, et al. |
| EPI_ISL_823925 | DOHMH Crown Heights | New York City Public Health Laboratory | Jade Wang, et al. |
| EPI_ISL_823926 | DOHMH Jamaica | New York City Public Health Laboratory | Jade Wang, et al. |
| EPI_ISL_823927 | DOHMH Central Harlem | New York City Public Health Laboratory | Jade Wang, et al. |
| EPI_ISL_823928 | DOHMH Jamaica | New York City Public Health Laboratory | Jade Wang, et al. |
| EPI_ISL_823929 | DOHMH Morrisania | New York City Public Health Laboratory | Jade Wang, et al. |
| EPI_ISL_823930 | DOHMH Chelsea | New York City Public Health Laboratory | Jade Wang, et al. |
| EPI_ISL_823931 | DOHMH Corona | New York City Public Health Laboratory | Jade Wang, et al. |
| EPI_ISL_823932 | DOHMH Morrisania | New York City Public Health Laboratory | Jade Wang, et al. |
| EPI_ISL_823933 | DOHMH Corona | New York City Public Health Laboratory | Jade Wang, et al. |
| EPI_ISL_823934 | DOHMH Chelsea | New York City Public Health Laboratory | Jade Wang, et al. |
| EPI_ISL_823935, EPI_ISL_823936 | DOHMH Crown Heights | New York City Public Health Laboratory | Jade Wang, et al. |
| EPI_ISL_823937 | DOHMH PHL | New York City Public Health Laboratory | Jade Wang, et al. |
| EPI_ISL_823938 | DOHMH Fort Greene | New York City Public Health Laboratory | Jade Wang, et al. |
| EPI_ISL_823939 | DOHMH Riverside | New York City Public Health Laboratory | Jade Wang, et al. |
| EPI_ISL_823940, EPI_ISL_823941 | DOHMH Jamaica | New York City Public Health Laboratory | Jade Wang, et al. |
| EPI_ISL_823942 | DOHMH Crown Heights | New York City Public Health Laboratory | Jade Wang, et al. |
| EPI_ISL_823943, EPI_ISL_823944, EPI_ISL_823945 | DOHMH Corona | New York City Public Health Laboratory | Jade Wang, et al. |
| EPI_ISL_823946 | DOHMH Jamaica | New York City Public Health Laboratory | Jade Wang, et al. |
| EPI_ISL_823947 | DOHMH Morrisania | New York City Public Health Laboratory | Jade Wang, et al. |
| EPI_ISL_823948 | DOHMH Chelsea | New York City Public Health Laboratory | Jade Wang, et al. |
| EPI_ISL_823949 | DOHMH Jamaica | New York City Public Health Laboratory | Jade Wang, et al. |
| EPI_ISL_823950 | DOHMH Central Harlem | New York City Public Health Laboratory | Jade Wang, et al. |
| EPI_ISL_823951, EPI_ISL_823952 | DOHMH Riverside | New York City Public Health Laboratory | Jade Wang, et al. |
| EPI_ISL_823953 | DOHMH Corona | New York City Public Health Laboratory | Jade Wang, et al. |
| EPI_ISL_823954 | DOHMH Chelsea | New York City Public Health Laboratory | Jade Wang, et al. |
| EPI_ISL_823955, EPI_ISL_823956 | DOHMH Jamaica | New York City Public Health Laboratory | Jade Wang, et al. |
| EPI_ISL_823957 | DOHMH Corona | New York City Public Health Laboratory | Jade Wang, et al. |
| EPI_ISL_823958 | DOHMH PHL | New York City Public Health Laboratory | Jade Wang, et al. |
| EPI_ISL_823959, EPI_ISL_823960 | DOHMH Morrisania | New York City Public Health Laboratory | Jade Wang, et al. |
| EPI_ISL_823961 | DOHMH Jamaica | New York City Public Health Laboratory | Jade Wang, et al. |
| EPI_ISL_823962 | DOHMH Chelsea | New York City Public Health Laboratory | Jade Wang, et al. |
| EPI_ISL_823963, EPI_ISL_823964, EPI_ISL_823965, EPI_ISL_823966, EPI_ISL_823967 | DOHMH Corona | New York City Public Health Laboratory | Jade Wang, et al. |
| EPI_ISL_823968, EPI_ISL_823969 | DOHMH Jamaica | New York City Public Health Laboratory | Jade Wang, et al. |
| EPI_ISL_823970 | Department of Homeless Services | New York City Public Health Laboratory | Jade Wang, et al. |
| EPI_ISL_823971, EPI_ISL_823972 | OCME Office Of Chief Medical Examiner | New York City Public Health Laboratory | Jade Wang, et al. |
| EPI_ISL_823973 | Department of Homeless Services | New York City Public Health Laboratory | Jade Wang, et al. |
| EPI_ISL_823974 | OCME Office Of Chief Medical Examiner | New York City Public Health Laboratory | Jade Wang, et al. |
| EPI_ISL_823975 | Department of Homeless Services | New York City Public Health Laboratory | Jade Wang, et al. |
| EPI_ISL_824295 | DOHMH Morrisania | New York City Public Health Laboratory | Jade Wang, et al. |
| EPI_ISL_824296, EPI_ISL_824297 | DOHMH Jamaica | New York City Public Health Laboratory | Jade Wang, et al. |
| EPI_ISL_824298 | DOHMH Central Harlem | New York City Public Health Laboratory | Jade Wang, et al. |
| EPI_ISL_824299 | DOHMH Morrisania | New York City Public Health Laboratory | Jade Wang, et al. |
| EPI_ISL_824300 | DOHMH Chelsea | New York City Public Health Laboratory | Jade Wang, et al. |
| EPI_ISL_824301 | DOHMH Crown Heights | New York City Public Health Laboratory | Jade Wang, et al. |
| EPI_ISL_824302 | DOHMH Jamaica | New York City Public Health Laboratory | Jade Wang, et al. |

| | | | |
|---|---|---|---|
| EPI_ISL_824303 | DOHMH Morrisania | New York City Public Health Laboratory | Jade Wang, et al. |
| EPI_ISL_824304, EPI_ISL_824305 | DOHMH Jamaica | New York City Public Health Laboratory | Jade Wang, et al. |
| EPI_ISL_824306, EPI_ISL_824307 | DOHMH PHL | New York City Public Health Laboratory | Jade Wang, et al. |
| EPI_ISL_824308, EPI_ISL_824309 | DOHMH Jamaica | New York City Public Health Laboratory | Jade Wang, et al. |
| EPI_ISL_824310 | DOHMH Crown Heights | New York City Public Health Laboratory | Jade Wang, et al. |
| EPI_ISL_824311, EPI_ISL_824312 | DOHMH Fort Greene | New York City Public Health Laboratory | Jade Wang, et al. |
| EPI_ISL_824313 | DOHMH Jamaica | New York City Public Health Laboratory | Jade Wang, et al. |
| EPI_ISL_824314, EPI_ISL_824315 | DOHMH Corona | New York City Public Health Laboratory | Jade Wang, et al. |
| EPI_ISL_824316 | DOHMH Crown Heights | New York City Public Health Laboratory | Jade Wang, et al. |
| EPI_ISL_824317, EPI_ISL_824318 | DOHMH Central Harlem | New York City Public Health Laboratory | Jade Wang, et al. |
| EPI_ISL_824319 | DOHMH Chelsea | New York City Public Health Laboratory | Jade Wang, et al. |
| EPI_ISL_824320 | DOHMH Corona | New York City Public Health Laboratory | Jade Wang, et al. |
| EPI_ISL_824321, EPI_ISL_824322 | DOHMH Fort Greene | New York City Public Health Laboratory | Jade Wang, et al. |
| EPI_ISL_824323, EPI_ISL_824324, EPI_ISL_824325, EPI_ISL_824326, EPI_ISL_824327 | DOHMH Morrisania | New York City Public Health Laboratory | Jade Wang, et al. |
| EPI_ISL_824328 | DOHMH Corona | New York City Public Health Laboratory | Jade Wang, et al. |
| EPI_ISL_824329, EPI_ISL_824330 | DOHMH Jamaica | New York City Public Health Laboratory | Jade Wang, et al. |
| EPI_ISL_824331 | DOHMH Corona | New York City Public Health Laboratory | Jade Wang, et al. |
| EPI_ISL_826578, EPI_ISL_826579, EPI_ISL_826580, EPI_ISL_826581, EPI_ISL_826582, EPI_ISL_826583, EPI_ISL_826584, EPI_ISL_826585, EPI_ISL_826586, EPI_ISL_826587, EPI_ISL_826588, EPI_ISL_826589, EPI_ISL_826590, EPI_ISL_826591, EPI_ISL_826592, EPI_ISL_826593, EPI_ISL_826594, EPI_ISL_826595, EPI_ISL_826596, EPI_ISL_826597, EPI_ISL_826598, EPI_ISL_826599, EPI_ISL_826600, EPI_ISL_826601, EPI_ISL_826602, EPI_ISL_826603, EPI_ISL_826604, EPI_ISL_826605, EPI_ISL_826606, EPI_ISL_826607, EPI_ISL_826608, EPI_ISL_826609, EPI_ISL_826610, EPI_ISL_826611, EPI_ISL_826612, EPI_ISL_826613, EPI_ISL_826614, EPI_ISL_826615, EPI_ISL_826616, EPI_ISL_826617, EPI_ISL_826618, EPI_ISL_826619, EPI_ISL_826620, EPI_ISL_826621, EPI_ISL_826622, EPI_ISL_826623, EPI_ISL_826624, EPI_ISL_826625, EPI_ISL_826626, EPI_ISL_826627, EPI_ISL_826628, EPI_ISL_826629, EPI_ISL_826630, EPI_ISL_826631, EPI_ISL_826632, EPI_ISL_826633, EPI_ISL_826634, EPI_ISL_826635, EPI_ISL_826636, EPI_ISL_826637, EPI_ISL_826638, EPI_ISL_826639, EPI_ISL_826640, EPI_ISL_826641, EPI_ISL_826642, EPI_ISL_826643, EPI_ISL_826644, EPI_ISL_826645, EPI_ISL_826646, EPI_ISL_826647, EPI_ISL_826648, EPI_ISL_826649, EPI_ISL_826650, EPI_ISL_826651, EPI_ISL_826652, EPI_ISL_826653, EPI_ISL_826654, EPI_ISL_826655, EPI_ISL_826656, EPI_ISL_826657, EPI_ISL_826658, EPI_ISL_826659, EPI_ISL_826660, EPI_ISL_826661, EPI_ISL_826662, EPI_ISL_826663, EPI_ISL_826664, EPI_ISL_826665, EPI_ISL_826666, EPI_ISL_826667, EPI_ISL_826668, EPI_ISL_826669, EPI_ISL_826670 | | | |
| see above | Montefiore Medical Center | Albert Einstein College of Medicine, Dept. of Microbiology & Immunology, Chandran lab | J. Maximilian Fels, Saad Khan, Ryan Forster, Karin A. Skalina, Surksha Sirichand, Amy S. Fox, Aviv Bergman, William B. Mitchell, Lucia R. Wolgast, Wendy Szymczak, Robert H. Bortz III, M. Eugenia Dieterle, Catalina Florez, Denise Haslwanter, Rohit K. Jangra, Ethan Laudermilch, Ariel S. Wirchnianski, Jason Barnhill, David L. Goldman, Hnin Khine, D. Yitzchak Goldstein, Johanna P. Daily, Kartik Chandran, Libusha Kelly |
| EPI_ISL_830208, EPI_ISL_830210, EPI_ISL_830212 | KALEIDA CENTER FOR LABORATORY MEDICINE | Wadsworth Center, New York State Department of Health | Kirsten St. George, Daryl M. Lamson, Alexis Russel, Matthew Shudt, Melissa A Leisner, Jonathan Plitnick, Navjot Singh, John Kelly, Erasmus Schneider, Erica Lasek-Nesselquist |
| EPI_ISL_830214 | BOSTON HEART DIAGNOSTICS CORP | Wadsworth Center, New York State Department of Health | Kirsten St. George, Daryl M. Lamson, Alexis Russel, Matthew Shudt, Melissa A Leisner, Jonathan Plitnick, Navjot Singh, John Kelly, Erasmus Schneider, Erica Lasek-Nesselquist |
| EPI_ISL_830216, EPI_ISL_830218, EPI_ISL_830220, EPI_ISL_830221 | Wadsworth Center, New York State Department of Health | Wadsworth Center, New York State Department of Health | Kirsten St. George, Daryl M. Lamson, Alexis Russel, Matthew Shudt, Melissa A Leisner, Jonathan Plitnick, Navjot Singh, John Kelly, Erasmus Schneider, Erica Lasek-Nesselquist |
| EPI_ISL_830223, EPI_ISL_830225, EPI_ISL_830227, EPI_ISL_830229, EPI_ISL_830230, EPI_ISL_830232, EPI_ISL_830234, EPI_ISL_830236, EPI_ISL_830238, EPI_ISL_830239, EPI_ISL_830241, EPI_ISL_830243 | | | |
| see above | KALEIDA CENTER FOR LABORATORY MEDICINE | Wadsworth Center, New York State Department of Health | Kirsten St. George, Daryl M. Lamson, Alexis Russel, Matthew Shudt, Melissa A Leisner, Jonathan Plitnick, Navjot Singh, John Kelly, Erasmus Schneider, Erica Lasek-Nesselquist |
| EPI_ISL_830244, EPI_ISL_830246, EPI_ISL_830248, EPI_ISL_830250, EPI_ISL_830252, EPI_ISL_830253, EPI_ISL_830255, EPI_ISL_830257, EPI_ISL_830259, EPI_ISL_830261, EPI_ISL_830263, EPI_ISL_830265, EPI_ISL_830267, EPI_ISL_830269, EPI_ISL_830270, EPI_ISL_830272, EPI_ISL_830274, EPI_ISL_830276, EPI_ISL_830278, EPI_ISL_830280, EPI_ISL_830282, EPI_ISL_830284, EPI_ISL_830286, EPI_ISL_830288, EPI_ISL_830289, EPI_ISL_830291, EPI_ISL_830293, EPI_ISL_830294, EPI_ISL_830296, EPI_ISL_830298, EPI_ISL_830300, EPI_ISL_830302, EPI_ISL_830304, EPI_ISL_830306, EPI_ISL_830308, EPI_ISL_830310, EPI_ISL_830312, EPI_ISL_830313, EPI_ISL_830315, EPI_ISL_830317, EPI_ISL_830319, EPI_ISL_830321, EPI_ISL_830323, EPI_ISL_830325, EPI_ISL_830326, EPI_ISL_830329, EPI_ISL_830331, EPI_ISL_830333, EPI_ISL_830334, EPI_ISL_830335, EPI_ISL_830337, EPI_ISL_830338, EPI_ISL_830340, EPI_ISL_830342, EPI_ISL_830344, EPI_ISL_830346, EPI_ISL_830348, EPI_ISL_830349 | | | |
| see above | Wadsworth Center, New York State Department of Health | Wadsworth Center, New York State Department of Health | Kirsten St. George, Daryl M. Lamson, Alexis Russel, Matthew Shudt, Melissa A Leisner, Jonathan Plitnick, Navjot Singh, John Kelly, Erasmus Schneider, Erica Lasek-Nesselquist |
| EPI_ISL_830571 | KALEIDA CENTER FOR LABORATORY MEDICINE | Wadsworth Center, New York State Department of Health | Kirsten St. George, Daryl M. Lamson, Alexis Russel, Matthew Shudt, Melissa A Leisner, Jonathan Plitnick, Navjot Singh, John Kelly, Erasmus Schneider, Erica Lasek-Nesselquist |
| EPI_ISL_830572 | NORTHWELL HEALTH LABORATORIES | Wadsworth Center, New York State Department of Health | Kirsten St. George, Daryl M. Lamson, Alexis Russel, Matthew Shudt, Melissa A Leisner, Jonathan Plitnick, Navjot Singh, John Kelly, Erasmus Schneider, Erica Lasek-Nesselquist |
| EPI_ISL_830573 | KALEIDA CENTER FOR LABORATORY MEDICINE | Wadsworth Center, New York State Department of Health | Kirsten St. George, Daryl M. Lamson, Alexis Russel, Matthew Shudt, Melissa A Leisner, Jonathan Plitnick, Navjot Singh, John Kelly, Erasmus Schneider, Erica Lasek-Nesselquist |
| EPI_ISL_830574 | NORTHWELL HEALTH LABORATORIES | Wadsworth Center, New York State Department of Health | Kirsten St. George, Daryl M. Lamson, Alexis Russel, Matthew Shudt, Melissa A Leisner, Jonathan Plitnick, Navjot Singh, John Kelly, Erasmus Schneider, Erica Lasek-Nesselquist |
| EPI_ISL_830575 | KALEIDA CENTER FOR LABORATORY MEDICINE | Wadsworth Center, New York State Department of Health | Kirsten St. George, Daryl M. Lamson, Alexis Russel, Matthew Shudt, Melissa A Leisner, Jonathan Plitnick, Navjot Singh, John Kelly, Erasmus Schneider, Erica Lasek-Nesselquist |
| EPI_ISL_830576, EPI_ISL_830577, EPI_ISL_830578 | NORTHWELL HEALTH LABORATORIES | Wadsworth Center, New York State Department of Health | Kirsten St. George, Daryl M. Lamson, Alexis Russel, Matthew Shudt, Melissa A Leisner, Jonathan Plitnick, Navjot Singh, John Kelly, Erasmus Schneider, Erica Lasek-Nesselquist |
| EPI_ISL_830579, EPI_ISL_830580, EPI_ISL_830581, EPI_ISL_830582, EPI_ISL_830583, EPI_ISL_830584, EPI_ISL_830585, EPI_ISL_830586, EPI_ISL_830587, EPI_ISL_830588, EPI_ISL_830589, EPI_ISL_830590, EPI_ISL_830591, EPI_ISL_830592, EPI_ISL_830593, EPI_ISL_830594, EPI_ISL_830595, EPI_ISL_830596, EPI_ISL_830597, EPI_ISL_830598, EPI_ISL_830599, EPI_ISL_830600, EPI_ISL_830601, EPI_ISL_830602, EPI_ISL_830603, EPI_ISL_830604, EPI_ISL_830605, EPI_ISL_830606, EPI_ISL_830607, EPI_ISL_830608, EPI_ISL_830609, EPI_ISL_830610, EPI_ISL_830611, EPI_ISL_830612, EPI_ISL_830613, EPI_ISL_830614, EPI_ISL_830615, EPI_ISL_830616, EPI_ISL_830617, EPI_ISL_830618, EPI_ISL_830619, EPI_ISL_830620, EPI_ISL_830621, EPI_ISL_830622, EPI_ISL_830623, EPI_ISL_830624, EPI_ISL_830625, EPI_ISL_830626, EPI_ISL_830627 | | | |
| see above | KALEIDA CENTER FOR LABORATORY MEDICINE | Wadsworth Center, New York State Department of Health | Kirsten St. George, Daryl M. Lamson, Alexis Russel, Matthew Shudt, Melissa A Leisner, Jonathan Plitnick, Navjot Singh, John Kelly, Erasmus Schneider, Erica Lasek-Nesselquist |
| EPI_ISL_830628, EPI_ISL_830629, EPI_ISL_830630, EPI_ISL_830631, EPI_ISL_830632, EPI_ISL_830633 | NORTHWELL HEALTH LABORATORIES | Wadsworth Center, New York State Department of Health | Kirsten St. George, Daryl M. Lamson, Alexis Russel, Matthew Shudt, Melissa A Leisner, Jonathan Plitnick, Navjot Singh, John Kelly, Erasmus Schneider, Erica Lasek-Nesselquist |
| EPI_ISL_830634 | MARY IMOGENE BASSETT HOSPITAL | Wadsworth Center, New York State Department of Health | Kirsten St. George, Daryl M. Lamson, Alexis Russel, Matthew Shudt, Melissa A Leisner, Jonathan Plitnick, Navjot Singh, John Kelly, Erasmus Schneider, Erica Lasek-Nesselquist |
| EPI_ISL_830635, EPI_ISL_830636, EPI_ISL_830637, EPI_ISL_830638, EPI_ISL_830639, EPI_ISL_830640, EPI_ISL_830641, EPI_ISL_830642 | NORTHWELL HEALTH LABORATORIES | Wadsworth Center, New York State Department of Health | Kirsten St. George, Daryl M. Lamson, Alexis Russel, Matthew Shudt, Melissa A Leisner, Jonathan Plitnick, Navjot Singh, John Kelly, Erasmus Schneider, Erica Lasek-Nesselquist |

| | | | |
|---|---|---|---|
| EPI_ISL_830643, EPI_ISL_830644, EPI_ISL_830645, EPI_ISL_830646, EPI_ISL_830647 | Wadsworth Center, New York State Department of Health | Wadsworth Center, New York State Department of Health | Kirsten St. George, Daryl M. Lamson, Alexis Russel, Matthew Shudt, Melissa A Leisner, Jonathan Plitnick, Navjot Singh, John Kelly, Erasmus Schneider, Erica Lasek-Nesselquist |
| EPI_ISL_830650, EPI_ISL_830651, EPI_ISL_830652, EPI_ISL_830653, EPI_ISL_830654, EPI_ISL_830655, EPI_ISL_830656, EPI_ISL_830657, EPI_ISL_830658, EPI_ISL_830659, EPI_ISL_830660, EPI_ISL_830661, EPI_ISL_830662, EPI_ISL_830663, EPI_ISL_830664, EPI_ISL_830665, EPI_ISL_830666, EPI_ISL_830667, EPI_ISL_830668, EPI_ISL_830669, EPI_ISL_830670, EPI_ISL_830671, EPI_ISL_830672, EPI_ISL_830673, EPI_ISL_830674, EPI_ISL_830675, EPI_ISL_830676, EPI_ISL_830677, EPI_ISL_830678, EPI_ISL_830679, EPI_ISL_830680, EPI_ISL_830681, EPI_ISL_830682 | | | |
| see above | SUNY UPSTATE MEDICAL UNIVERSITY | Wadsworth Center, New York State Department of Health | Kirsten St. George, Daryl M. Lamson, Alexis Russel, Matthew Shudt, Melissa A Leisner, Jonathan Plitnick, Navjot Singh, John Kelly, Erasmus Schneider, Erica Lasek-Nesselquist |
| EPI_ISL_830683, EPI_ISL_830684, EPI_ISL_830685, EPI_ISL_830686, EPI_ISL_830687, EPI_ISL_830688, EPI_ISL_830689, EPI_ISL_830690, EPI_ISL_830691, EPI_ISL_830692, EPI_ISL_830693, EPI_ISL_830694, EPI_ISL_830695, EPI_ISL_830696, EPI_ISL_830697, EPI_ISL_830698, EPI_ISL_830699, EPI_ISL_830700, EPI_ISL_830701, EPI_ISL_830702, EPI_ISL_830703, EPI_ISL_830704, EPI_ISL_830705, EPI_ISL_830706, EPI_ISL_830707, EPI_ISL_830708, EPI_ISL_830709, EPI_ISL_830710, EPI_ISL_830711, EPI_ISL_830712, EPI_ISL_830713, EPI_ISL_830714, EPI_ISL_830715, EPI_ISL_830716, EPI_ISL_830717, EPI_ISL_830718, EPI_ISL_830719 | | | |
| see above | ALBANY MEDICAL CENTER HOSPITAL CLINICAL LABORATORIES | Wadsworth Center, New York State Department of Health | Kirsten St. George, Daryl M. Lamson, Alexis Russel, Matthew Shudt, Melissa A Leisner, Jonathan Plitnick, Navjot Singh, John Kelly, Erasmus Schneider, Erica Lasek-Nesselquist |
| EPI_ISL_830720, EPI_ISL_830721, EPI_ISL_830722, EPI_ISL_830723 | BIO-REFERENCE LABORATORIES | Wadsworth Center, New York State Department of Health | Kirsten St. George, Daryl M. Lamson, Alexis Russel, Matthew Shudt, Melissa A Leisner, Jonathan Plitnick, Navjot Singh, John Kelly, Erasmus Schneider, Erica Lasek-Nesselquist |
| EPI_ISL_830724, EPI_ISL_830725 | ACUTIS DIAGNOSTICS | Wadsworth Center, New York State Department of Health | Kirsten St. George, Daryl M. Lamson, Alexis Russel, Matthew Shudt, Melissa A Leisner, Jonathan Plitnick, Navjot Singh, John Kelly, Erasmus Schneider, Erica Lasek-Nesselquist |
| EPI_ISL_830726 | Wadsworth Center, New York State Department of Health | Wadsworth Center, New York State Department of Health | Kirsten St. George, Daryl M. Lamson, Alexis Russel, Matthew Shudt, Melissa A Leisner, Jonathan Plitnick, Navjot Singh, John Kelly, Erasmus Schneider, Erica Lasek-Nesselquist |
| EPI_ISL_832243 | DOHMH Corona | New York City Public Health Laboratory | Jade Wang, et al. |
| EPI_ISL_832244 | DOHMH Crown Heights | New York City Public Health Laboratory | Jade Wang, et al. |
| EPI_ISL_832245 | DOHMH Corona | New York City Public Health Laboratory | Jade Wang, et al. |
| EPI_ISL_832246 | DOHMH Jamaica | New York City Public Health Laboratory | Jade Wang, et al. |
| EPI_ISL_832247, EPI_ISL_832248 | DOHMH Morrisania | New York City Public Health Laboratory | Jade Wang, et al. |
| EPI_ISL_832249 | DOHMH PHL | New York City Public Health Laboratory | Jade Wang, et al. |
| EPI_ISL_832250 | DOHMH Jamaica | New York City Public Health Laboratory | Jade Wang, et al. |
| EPI_ISL_832251, EPI_ISL_832252, EPI_ISL_832253, EPI_ISL_832254 | DOHMH Corona | New York City Public Health Laboratory | Jade Wang, et al. |
| EPI_ISL_832255 | DOHMH Morrisania | New York City Public Health Laboratory | Jade Wang, et al. |
| EPI_ISL_832256 | DOHMH Central Harlem | New York City Public Health Laboratory | Jade Wang, et al. |
| EPI_ISL_832257 | DOHMH Morrisania | New York City Public Health Laboratory | Jade Wang, et al. |
| EPI_ISL_832258, EPI_ISL_832259 | DOHMH PHL | New York City Public Health Laboratory | Jade Wang, et al. |
| EPI_ISL_832260, EPI_ISL_832261, EPI_ISL_832262 | DOHMH Corona | New York City Public Health Laboratory | Jade Wang, et al. |
| EPI_ISL_832263 | DOHMH Jamaica | New York City Public Health Laboratory | Jade Wang, et al. |
| EPI_ISL_832264 | DOHMH Morrisania | New York City Public Health Laboratory | Jade Wang, et al. |
| EPI_ISL_832265 | DOHMH Jamaica | New York City Public Health Laboratory | Jade Wang, et al. |
| EPI_ISL_832266, EPI_ISL_832267 | DOHMH Central Harlem | New York City Public Health Laboratory | Jade Wang, et al. |
| EPI_ISL_832268, EPI_ISL_832269 | DOHMH Fort Greene | New York City Public Health Laboratory | Jade Wang, et al. |
| EPI_ISL_832270, EPI_ISL_832271 | DOHMH Riverside | New York City Public Health Laboratory | Jade Wang, et al. |
| EPI_ISL_832272, EPI_ISL_832273, EPI_ISL_832274 | DOHMH Jamaica | New York City Public Health Laboratory | Jade Wang, et al. |
| EPI_ISL_832275 | DOHMH Crown Heights | New York City Public Health Laboratory | Jade Wang, et al. |
| EPI_ISL_832276 | DOHMH Jamaica | New York City Public Health Laboratory | Jade Wang, et al. |
| EPI_ISL_832277, EPI_ISL_832278, EPI_ISL_832279 | DOHMH Corona | New York City Public Health Laboratory | Jade Wang, et al. |
| EPI_ISL_832280, EPI_ISL_832281 | DOHMH Morrisania | New York City Public Health Laboratory | Jade Wang, et al. |
| EPI_ISL_832282 | DOHMH Chelsea | New York City Public Health Laboratory | Jade Wang, et al. |
| EPI_ISL_832283 | DOHMH Jamaica | New York City Public Health Laboratory | Jade Wang, et al. |
| EPI_ISL_832284 | DOHMH Central Harlem | New York City Public Health Laboratory | Jade Wang, et al. |
| EPI_ISL_832285 | DOHMH Morrisania | New York City Public Health Laboratory | Jade Wang, et al. |
| EPI_ISL_832286 | DOHMH Fort Greene | New York City Public Health Laboratory | Jade Wang, et al. |
| EPI_ISL_832287, EPI_ISL_832288 | DOHMH Crown Heights | New York City Public Health Laboratory | Jade Wang, et al. |
| EPI_ISL_832289 | DOHMH Morrisania | New York City Public Health Laboratory | Jade Wang, et al. |
| EPI_ISL_832290, EPI_ISL_832291 | DOHMH Corona | New York City Public Health Laboratory | Jade Wang, et al. |
| EPI_ISL_832292, EPI_ISL_832293 | DOHMH Crown Heights | New York City Public Health Laboratory | Jade Wang, et al. |
| EPI_ISL_832294 | DOHMH Jamaica | New York City Public Health Laboratory | Jade Wang, et al. |
| EPI_ISL_832295 | DOHMH Crown Heights | New York City Public Health Laboratory | Jade Wang, et al. |
| EPI_ISL_832296 | DOHMH Riverside | New York City Public Health Laboratory | Jade Wang, et al. |
| EPI_ISL_832297, EPI_ISL_832298 | DOHMH Jamaica | New York City Public Health Laboratory | Jade Wang, et al. |
| EPI_ISL_832299 | DOHMH Morrisania | New York City Public Health Laboratory | Jade Wang, et al. |
| EPI_ISL_832300 | DOHMH Central Harlem | New York City Public Health Laboratory | Jade Wang, et al. |
| EPI_ISL_832301, EPI_ISL_832302 | DOHMH Chelsea | New York City Public Health Laboratory | Jade Wang, et al. |
| EPI_ISL_832303, EPI_ISL_832304 | DOHMH Corona | New York City Public Health Laboratory | Jade Wang, et al. |

| | | | |
|---|---|---|---|
| EPI_ISL_832305 | DOHMH Riverside | New York City Public Health Laboratory | Jade Wang, et al. |
| EPI_ISL_832306, EPI_ISL_832307, EPI_ISL_832308, EPI_ISL_832309, EPI_ISL_832310, EPI_ISL_832311 | DOHMH Jamaica | New York City Public Health Laboratory | Jade Wang, et al. |
| EPI_ISL_832312 | DOHMH Central Harlem | New York City Public Health Laboratory | Jade Wang, et al. |
| EPI_ISL_832313, EPI_ISL_832314 | DOHMH Chelsea | New York City Public Health Laboratory | Jade Wang, et al. |
| EPI_ISL_832315, EPI_ISL_832316 | DOHMH Morrisania | New York City Public Health Laboratory | Jade Wang, et al. |
| EPI_ISL_832317, EPI_ISL_832318, EPI_ISL_832319 | DOHMH Jamaica | New York City Public Health Laboratory | Jade Wang, et al. |
| EPI_ISL_832320, EPI_ISL_832321, EPI_ISL_832322 | DOHMH Corona | New York City Public Health Laboratory | Jade Wang, et al. |
| EPI_ISL_832323, EPI_ISL_832324 | DOHMH Jamaica | New York City Public Health Laboratory | Jade Wang, et al. |
| EPI_ISL_832325 | DOHMH Corona | New York City Public Health Laboratory | Jade Wang, et al. |
| EPI_ISL_832326 | DOHMH Riverside | New York City Public Health Laboratory | Jade Wang, et al. |
| EPI_ISL_832327 | DOHMH Morrisania | New York City Public Health Laboratory | Jade Wang, et al. |
| EPI_ISL_832328, EPI_ISL_832329, EPI_ISL_832330 | DOHMH Jamaica | New York City Public Health Laboratory | Jade Wang, et al. |
| EPI_ISL_832331, EPI_ISL_832332, EPI_ISL_832333, EPI_ISL_832334, EPI_ISL_832335 | DOHMH Corona | New York City Public Health Laboratory | Jade Wang, et al. |
| EPI_ISL_832336, EPI_ISL_832337 | DOHMH Central Harlem | New York City Public Health Laboratory | Jade Wang, et al. |
| EPI_ISL_832338 | DOHMH Crown Heights | New York City Public Health Laboratory | Jade Wang, et al. |
| EPI_ISL_832339 | DOHMH Riverside | New York City Public Health Laboratory | Jade Wang, et al. |
| EPI_ISL_832340 | DOHMH Crown Heights | New York City Public Health Laboratory | Jade Wang, et al. |
| EPI_ISL_832341 | DOHMH Chelsea | New York City Public Health Laboratory | Jade Wang, et al. |
| EPI_ISL_832342, EPI_ISL_832343, EPI_ISL_832344, EPI_ISL_832345 | DOHMH Morrisania | New York City Public Health Laboratory | Jade Wang, et al. |
| EPI_ISL_832346, EPI_ISL_832347, EPI_ISL_832348, EPI_ISL_832349, EPI_ISL_832350 | DOHMH Central Harlem | New York City Public Health Laboratory | Jade Wang, et al. |
| EPI_ISL_832351 | DOHMH Corona | New York City Public Health Laboratory | Jade Wang, et al. |
| EPI_ISL_832352, EPI_ISL_832353 | DOHMH PHL | New York City Public Health Laboratory | Jade Wang, et al. |
| EPI_ISL_832354 | DOHMH Jamaica | New York City Public Health Laboratory | Jade Wang, et al. |
| EPI_ISL_832355, EPI_ISL_832356 | DOHMH Corona | New York City Public Health Laboratory | Jade Wang, et al. |
| EPI_ISL_832357, EPI_ISL_832358 | DOHMH Jamaica | New York City Public Health Laboratory | Jade Wang, et al. |
| EPI_ISL_832359, EPI_ISL_832360 | DOHMH Corona | New York City Public Health Laboratory | Jade Wang, et al. |
| EPI_ISL_832361 | DOHMH Jamaica | New York City Public Health Laboratory | Jade Wang, et al. |
| EPI_ISL_832362, EPI_ISL_832363 | DOHMH Chelsea | New York City Public Health Laboratory | Jade Wang, et al. |
| EPI_ISL_847741 | Chiu Laboratory, University of California, San Francisco | Chiu Laboratory, University of California, San Francisco | Charles Chiu, Xianding (Wayne) Deng, Candace Wang, Brian Bushnell, Scot Federman, Jill Hacker, Debra Wadford |
| EPI_ISL_847830 | Tempus | Grubaugh Lab - Yale School of Public Health | Tara Alpert, Joseph Fauver, Anderson Brito, Mallery Breban, Anne Wyllie, Chantal Vogels, Mary Petrone, Chaney Kalinich, Isabel Ott, Nathan Grubaugh |
| EPI_ISL_850558, EPI_ISL_850565 | Helix/Illumina | Genomics and Discovery, Respiratory Viruses Branch, Division of Viral Diseases, Centers for Disease Control and Prevention | Peter W. Cook, Dhwani Batra, Ben L. Rambo-Martin Eileen de Feo, Jan Antico, Christine Tran, Matthew Tolentino, Shannon Wickline, Kim Gietzen, Brad Sickler, Jingtao Liu, Eric Allen, Phil Febbo, Summer Galloway, Nicole L. Washington, Simon White, Geraint Levan, Kelly Schiabor Barrett, Elizabeth Cirulli, Alexandre Bolze, Ary Ascencio, Charlotte Rivera-Garcia, Ryan Cho, Jason Nguyen, Sherry Wang, Jimmy Ramirez, Tyler Cassens, Efren Sandoval, Magnus Isaksson, William Lee, David Becker, Marc Laurent, James Lu, Clinton R. Paden, Suxiang Tong, Duncan MacCannell |
| EPI_ISL_850770, EPI_ISL_850874, EPI_ISL_850925, EPI_ISL_850934 | Helix / Illumina | Genomics and Discovery, Respiratory Viruses Branch, Division of Viral Diseases, Centers for Disease Control and Prevention | Peter W. Cook, Dhwani Batra, Ben L. Rambo-Martin Eileen de Feo, Jan Antico, Christine Tran, Matthew Tolentino, Shannon Wickline, Kim Gietzen, Brad Sickler, Jingtao Liu, Eric Allen, Phil Febbo, Summer Galloway, Nicole L. Washington, Simon White, Geraint Levan, Kelly Schiabor Barrett, Elizabeth Cirulli, Alexandre Bolze, Ary Ascencio, Charlotte Rivera-Garcia, Ryan Cho, Jason Nguyen, Sherry Wang, Jimmy Ramirez, Tyler Cassens, Efren Sandoval, Magnus Isaksson, William Lee, David Becker, Marc Laurent, James Lu, Clinton R. Paden, Suxiang Tong, Duncan MacCannell |
| EPI_ISL_850975 | Helix/Illumina | Genomics and Discovery, Respiratory Viruses Branch, Division of Viral Diseases, Centers for Disease Control and Prevention | Peter W. Cook, Dhwani Batra, Ben L. Rambo-Martin Eileen de Feo, Jan Antico, Christine Tran, Matthew Tolentino, Shannon Wickline, Kim Gietzen, Brad Sickler, Jingtao Liu, Eric Allen, Phil Febbo, Summer Galloway, Nicole L. Washington, Simon White, Geraint Levan, Kelly Schiabor Barrett, Elizabeth Cirulli, Alexandre Bolze, Ary Ascencio, Charlotte Rivera-Garcia, Ryan Cho, Jason Nguyen, Sherry Wang, Jimmy Ramirez, Tyler Cassens, Efren Sandoval, Magnus Isaksson, William Lee, David Becker, Marc Laurent, James Lu, Clinton R. Paden, Suxiang Tong, Duncan MacCannell |
| EPI_ISL_853648, EPI_ISL_853649 | THE MARY IMOGENE BASSETT HOSPITAL | Wadsworth Center, New York State Department of Health | Kirsten St. George, Daryl M. Lamson, Alexis Russel, Matthew Shudt, Melissa A Leisner, Jonathan Plitnick, Navjot Singh, John Kelly, Erasmus Schneider, Erica Lasek-Nesselquist |
| EPI_ISL_853650, EPI_ISL_853651 | NORTHWELL HEALTH LABORATORIES | Wadsworth Center, New York State Department of Health | Kirsten St. George, Daryl M. Lamson, Alexis Russel, Matthew Shudt, Melissa A Leisner, Jonathan Plitnick, Navjot Singh, John Kelly, Erasmus Schneider, Erica Lasek-Nesselquist |
| EPI_ISL_853652 | THE MARY IMOGENE BASSETT HOSPITAL | Wadsworth Center, New York State Department of Health | Kirsten St. George, Daryl M. Lamson, Alexis Russel, Matthew Shudt, Melissa A Leisner, Jonathan Plitnick, Navjot Singh, John Kelly, Erasmus Schneider, Erica Lasek-Nesselquist |
| EPI_ISL_853653, EPI_ISL_853654, EPI_ISL_853655, EPI_ISL_853656, EPI_ISL_853657, EPI_ISL_853658, EPI_ISL_853659, EPI_ISL_853660, EPI_ISL_853661, EPI_ISL_853662, EPI_ISL_853663, EPI_ISL_853664, EPI_ISL_853665, EPI_ISL_853666 | | | |
| see above | NORTHWELL HEALTH LABORATORIES | Wadsworth Center, New York State Department of Health | Kirsten St. George, Daryl M. Lamson, Alexis Russel, Matthew Shudt, Melissa A Leisner, Jonathan Plitnick, Navjot Singh, John Kelly, Erasmus Schneider, Erica Lasek-Nesselquist |
| EPI_ISL_853667, EPI_ISL_853668, EPI_ISL_853669 | THE MARY IMOGENE BASSETT HOSPITAL | Wadsworth Center, New York State Department of Health | Kirsten St. George, Daryl M. Lamson, Alexis Russel, Matthew Shudt, Melissa A Leisner, Jonathan Plitnick, Navjot Singh, John Kelly, Erasmus Schneider, Erica Lasek-Nesselquist |
| EPI_ISL_853670, EPI_ISL_853671, EPI_ISL_853672, EPI_ISL_853673, EPI_ISL_853674, EPI_ISL_853675 | NORTHWELL HEALTH LABORATORIES | Wadsworth Center, New York State Department of Health | Kirsten St. George, Daryl M. Lamson, Alexis Russel, Matthew Shudt, Melissa A Leisner, Jonathan Plitnick, Navjot Singh, John Kelly, Erasmus Schneider, Erica Lasek-Nesselquist |
| EPI_ISL_853676, EPI_ISL_853677, EPI_ISL_853678, EPI_ISL_853679, | ACUTIS DIAGNOSTICS | Wadsworth Center, New York State Department of Health | Kirsten St. George, Daryl M. Lamson, Alexis Russel, Matthew Shudt, Melissa A Leisner, Jonathan Plitnick, Navjot Singh, John Kelly, Erasmus Schneider, Erica Lasek-Nesselquist |

| | | | |
|---|---|---|---|
| EPI_ISL_853680, EPI_ISL_853681, EPI_ISL_853682 | | | |
| EPI_ISL_853683 | NORTHWELL HEALTH LABORATORIES | Wadsworth Center, New York State Department of Health | Kirsten St. George, Daryl M. Lamson, Alexis Russel, Matthew Shudt, Melissa A Leisner, Jonathan Plitnick, Navjot Singh, John Kelly, Erasmus Schneider, Erica Lasek-Nesselquist |
| EPI_ISL_853684, EPI_ISL_853685, EPI_ISL_853686, EPI_ISL_853687 | ALBANY MEDICAL CENTER HOSPITAL CLINICAL LABORATORIES | Wadsworth Center, New York State Department of Health | Kirsten St. George, Daryl M. Lamson, Alexis Russel, Matthew Shudt, Melissa A Leisner, Jonathan Plitnick, Navjot Singh, John Kelly, Erasmus Schneider, Erica Lasek-Nesselquist |
| EPI_ISL_853688 | THE MARY IMOGENE BASSETT HOSPITAL | Wadsworth Center, New York State Department of Health | Kirsten St. George, Daryl M. Lamson, Alexis Russel, Matthew Shudt, Melissa A Leisner, Jonathan Plitnick, Navjot Singh, John Kelly, Erasmus Schneider, Erica Lasek-Nesselquist |
| EPI_ISL_853689 | SUNY UPSTATE MEDICAL UNIVERSITY | Wadsworth Center, New York State Department of Health | Kirsten St. George, Daryl M. Lamson, Alexis Russel, Matthew Shudt, Melissa A Leisner, Jonathan Plitnick, Navjot Singh, John Kelly, Erasmus Schneider, Erica Lasek-Nesselquist |
| EPI_ISL_853690, EPI_ISL_853691, EPI_ISL_853692, EPI_ISL_853693, EPI_ISL_853694, EPI_ISL_853695, EPI_ISL_853696, EPI_ISL_853697, EPI_ISL_853698, EPI_ISL_853699, EPI_ISL_853700, EPI_ISL_853701, EPI_ISL_853702, EPI_ISL_853703, EPI_ISL_853704 | | | |
| see above | ALBANY MEDICAL CENTER HOSPITAL CLINICAL LABORATORIES | Wadsworth Center, New York State Department of Health | Kirsten St. George, Daryl M. Lamson, Alexis Russel, Matthew Shudt, Melissa A Leisner, Jonathan Plitnick, Navjot Singh, John Kelly, Erasmus Schneider, Erica Lasek-Nesselquist |
| EPI_ISL_853705 | NORTHWELL HEALTH LABORATORIES | Wadsworth Center, New York State Department of Health | Kirsten St. George, Daryl M. Lamson, Alexis Russel, Matthew Shudt, Melissa A Leisner, Jonathan Plitnick, Navjot Singh, John Kelly, Erasmus Schneider, Erica Lasek-Nesselquist |
| EPI_ISL_853706, EPI_ISL_853707, EPI_ISL_853708 | SUNY UPSTATE MEDICAL UNIVERSITY | Wadsworth Center, New York State Department of Health | Kirsten St. George, Daryl M. Lamson, Alexis Russel, Matthew Shudt, Melissa A Leisner, Jonathan Plitnick, Navjot Singh, John Kelly, Erasmus Schneider, Erica Lasek-Nesselquist |
| EPI_ISL_853709, EPI_ISL_853710, EPI_ISL_853711 | ALBANY MEDICAL CENTER HOSPITAL CLINICAL LABORATORIES | Wadsworth Center, New York State Department of Health | Kirsten St. George, Daryl M. Lamson, Alexis Russel, Matthew Shudt, Melissa A Leisner, Jonathan Plitnick, Navjot Singh, John Kelly, Erasmus Schneider, Erica Lasek-Nesselquist |
| EPI_ISL_853712, EPI_ISL_853713, EPI_ISL_853714 | NORTHWELL HEALTH LABORATORIES | Wadsworth Center, New York State Department of Health | Kirsten St. George, Daryl M. Lamson, Alexis Russel, Matthew Shudt, Melissa A Leisner, Jonathan Plitnick, Navjot Singh, John Kelly, Erasmus Schneider, Erica Lasek-Nesselquist |
| EPI_ISL_853715 | SUNY UPSTATE MEDICAL UNIVERSITY | Wadsworth Center, New York State Department of Health | Kirsten St. George, Daryl M. Lamson, Alexis Russel, Matthew Shudt, Melissa A Leisner, Jonathan Plitnick, Navjot Singh, John Kelly, Erasmus Schneider, Erica Lasek-Nesselquist |
| EPI_ISL_853716, EPI_ISL_853717 | NORTHWELL HEALTH LABORATORIES | Wadsworth Center, New York State Department of Health | Kirsten St. George, Daryl M. Lamson, Alexis Russel, Matthew Shudt, Melissa A Leisner, Jonathan Plitnick, Navjot Singh, John Kelly, Erasmus Schneider, Erica Lasek-Nesselquist |
| EPI_ISL_854305 | ESOTERIX GENETIC LABORATORIES LLC | Wadsworth Center, New York State Department of Health | Kirsten St. George, Daryl M. Lamson, Alexis Russel, Matthew Shudt, Melissa A Leisner, Jonathan Plitnick, Navjot Singh, John Kelly, Erasmus Schneider, Erica Lasek-Nesselquist |
| EPI_ISL_854306, EPI_ISL_854307, EPI_ISL_854308, EPI_ISL_854309, EPI_ISL_854310, EPI_ISL_854311, EPI_ISL_854312, EPI_ISL_854313, EPI_ISL_854314, EPI_ISL_854315, EPI_ISL_854316, EPI_ISL_854317, EPI_ISL_854318, EPI_ISL_854319, EPI_ISL_854320, EPI_ISL_854321, EPI_ISL_854322, EPI_ISL_854323, EPI_ISL_854324, EPI_ISL_854325, EPI_ISL_854326, EPI_ISL_854327, EPI_ISL_854328, EPI_ISL_854329, EPI_ISL_854330, EPI_ISL_854331, EPI_ISL_854332, EPI_ISL_854333, EPI_ISL_854334, EPI_ISL_854335, EPI_ISL_854336, EPI_ISL_854337, EPI_ISL_854338, EPI_ISL_854339, EPI_ISL_854340, EPI_ISL_854341, EPI_ISL_854342, EPI_ISL_854343, EPI_ISL_854344, EPI_ISL_854345, EPI_ISL_854346, EPI_ISL_854347, EPI_ISL_854348, EPI_ISL_854349 | | | |
| see above | URMC LABS | Wadsworth Center, New York State Department of Health | Kirsten St. George, Daryl M. Lamson, Alexis Russel, Matthew Shudt, Melissa A Leisner, Jonathan Plitnick, Navjot Singh, John Kelly, Erasmus Schneider, Erica Lasek-Nesselquist |
| EPI_ISL_854350, EPI_ISL_854351, EPI_ISL_854352, EPI_ISL_854353, EPI_ISL_854354, EPI_ISL_854355, EPI_ISL_854356, EPI_ISL_854357, EPI_ISL_854358, EPI_ISL_854359, EPI_ISL_854360, EPI_ISL_854361, EPI_ISL_854362, EPI_ISL_854363, EPI_ISL_854364, EPI_ISL_854365, EPI_ISL_854366, EPI_ISL_854367 | | | |
| see above | BIO-REFERENCE LABORATORIES | Wadsworth Center, New York State Department of Health | Kirsten St. George, Daryl M. Lamson, Alexis Russel, Matthew Shudt, Melissa A Leisner, Jonathan Plitnick, Navjot Singh, John Kelly, Erasmus Schneider, Erica Lasek-Nesselquist |
| EPI_ISL_854368 | Wadsworth Center, New York State Department of Health | Wadsworth Center, New York State Department of Health | Kirsten St. George, Daryl M. Lamson, Alexis Russel, Matthew Shudt, Melissa A Leisner, Jonathan Plitnick, Navjot Singh, John Kelly, Erasmus Schneider, Erica Lasek-Nesselquist |
| EPI_ISL_854369 | BIO-REFERENCE LABORATORIES | Wadsworth Center, New York State Department of Health | Kirsten St. George, Daryl M. Lamson, Alexis Russel, Matthew Shudt, Melissa A Leisner, Jonathan Plitnick, Navjot Singh, John Kelly, Erasmus Schneider, Erica Lasek-Nesselquist |
| EPI_ISL_854370, EPI_ISL_854371 | URMC LABS | Wadsworth Center, New York State Department of Health | Kirsten St. George, Daryl M. Lamson, Alexis Russel, Matthew Shudt, Melissa A Leisner, Jonathan Plitnick, Navjot Singh, John Kelly, Erasmus Schneider, Erica Lasek-Nesselquist |
| EPI_ISL_854372 | BIO-REFERENCE LABORATORIES | Wadsworth Center, New York State Department of Health | Kirsten St. George, Daryl M. Lamson, Alexis Russel, Matthew Shudt, Melissa A Leisner, Jonathan Plitnick, Navjot Singh, John Kelly, Erasmus Schneider, Erica Lasek-Nesselquist |
| EPI_ISL_854373 | MONTEFIORE MEDICAL CENTER LABORATORIES | Wadsworth Center, New York State Department of Health | Kirsten St. George, Daryl M. Lamson, Alexis Russel, Matthew Shudt, Melissa A Leisner, Jonathan Plitnick, Navjot Singh, John Kelly, Erasmus Schneider, Erica Lasek-Nesselquist |
| EPI_ISL_854374, EPI_ISL_854375, EPI_ISL_854376, EPI_ISL_854377, EPI_ISL_854378, EPI_ISL_854379, EPI_ISL_854380, EPI_ISL_854381, EPI_ISL_854382, EPI_ISL_854383, EPI_ISL_854384, EPI_ISL_854385, EPI_ISL_854386, EPI_ISL_854387, EPI_ISL_854388, EPI_ISL_854389, EPI_ISL_854390, EPI_ISL_854391, EPI_ISL_854392, EPI_ISL_854393, EPI_ISL_854394, EPI_ISL_854395, EPI_ISL_854396, EPI_ISL_854397, EPI_ISL_854398 | | | |
| see above | WESTCHESTER MEDICAL CENTER | Wadsworth Center, New York State Department of Health | Kirsten St. George, Daryl M. Lamson, Alexis Russel, Matthew Shudt, Melissa A Leisner, Jonathan Plitnick, Navjot Singh, John Kelly, Erasmus Schneider, Erica Lasek-Nesselquist |
| EPI_ISL_854399 | URMC LABS | Wadsworth Center, New York State Department of Health | Kirsten St. George, Daryl M. Lamson, Alexis Russel, Matthew Shudt, Melissa A Leisner, Jonathan Plitnick, Navjot Singh, John Kelly, Erasmus Schneider, Erica Lasek-Nesselquist |
| EPI_ISL_854400, EPI_ISL_854401, EPI_ISL_854402, EPI_ISL_854403 | WESTCHESTER MEDICAL CENTER | Wadsworth Center, New York State Department of Health | Kirsten St. George, Daryl M. Lamson, Alexis Russel, Matthew Shudt, Melissa A Leisner, Jonathan Plitnick, Navjot Singh, John Kelly, Erasmus Schneider, Erica Lasek-Nesselquist |
| EPI_ISL_854404 | URMC LABS | Wadsworth Center, New York State Department of Health | Kirsten St. George, Daryl M. Lamson, Alexis Russel, Matthew Shudt, Melissa A Leisner, Jonathan Plitnick, Navjot Singh, John Kelly, Erasmus Schneider, Erica Lasek-Nesselquist |
| EPI_ISL_854405, EPI_ISL_854406, EPI_ISL_854407, EPI_ISL_854408, EPI_ISL_854409, EPI_ISL_854410, EPI_ISL_854411, EPI_ISL_854412, EPI_ISL_854413, EPI_ISL_854414, EPI_ISL_854415, EPI_ISL_854416, EPI_ISL_854417, EPI_ISL_854418, EPI_ISL_854419, EPI_ISL_854420 | | | |
| see above | MONTEFIORE MEDICAL CENTER LABORATORIES | Wadsworth Center, New York State Department of Health | Kirsten St. George, Daryl M. Lamson, Alexis Russel, Matthew Shudt, Melissa A Leisner, Jonathan Plitnick, Navjot Singh, John Kelly, Erasmus Schneider, Erica Lasek-Nesselquist |
| EPI_ISL_854421 | SARATOGA HOSPITAL LABORATORY | Wadsworth Center, New York State Department of Health | Kirsten St. George, Daryl M. Lamson, Alexis Russel, Matthew Shudt, Melissa A Leisner, Jonathan Plitnick, Navjot Singh, John Kelly, Erasmus Schneider, Erica Lasek-Nesselquist |
| EPI_ISL_854422, EPI_ISL_854423, EPI_ISL_854424, EPI_ISL_854425, EPI_ISL_854426, EPI_ISL_854427, EPI_ISL_854428, EPI_ISL_854429, EPI_ISL_854430, EPI_ISL_854431, EPI_ISL_854432, EPI_ISL_854433, EPI_ISL_854434, EPI_ISL_854435 | | | |
| see above | MONTEFIORE MEDICAL CENTER LABORATORIES | Wadsworth Center, New York State Department of Health | Kirsten St. George, Daryl M. Lamson, Alexis Russel, Matthew Shudt, Melissa A Leisner, Jonathan Plitnick, Navjot Singh, John Kelly, Erasmus Schneider, Erica Lasek-Nesselquist |
| EPI_ISL_854436, EPI_ISL_854437, EPI_ISL_854438, EPI_ISL_854439, EPI_ISL_854440, EPI_ISL_854441, EPI_ISL_854442, EPI_ISL_854443, EPI_ISL_854444, EPI_ISL_854445 | SARATOGA HOSPITAL LABORATORY | Wadsworth Center, New York State Department of Health | Kirsten St. George, Daryl M. Lamson, Alexis Russel, Matthew Shudt, Melissa A Leisner, Jonathan Plitnick, Navjot Singh, John Kelly, Erasmus Schneider, Erica Lasek-Nesselquist |
| EPI_ISL_854446 | BOSTON HEART DIAGNOSTICS CORP | Wadsworth Center, New York State Department of Health | Kirsten St. George, Daryl M. Lamson, Alexis Russel, Matthew Shudt, Melissa A Leisner, Jonathan Plitnick, Navjot Singh, John Kelly, Erasmus Schneider, Erica Lasek-Nesselquist |

| | | | |
|---|---|---|---|
| EPI_ISL_854447, EPI_ISL_854448 | TEMPUS LABS INC | Wadsworth Center, New York State Department of Health | Kirsten St. George, Daryl M. Lamson, Alexis Russel, Matthew Shudt, Melissa A Leisner, Jonathan Plitnick, Navjot Singh, John Kelly, Erasmus Schneider, Erica Lasek-Nesselquist |
| EPI_ISL_854449 | WESTCHESTER MEDICAL CENTER | Wadsworth Center, New York State Department of Health | Kirsten St. George, Daryl M. Lamson, Alexis Russel, Matthew Shudt, Melissa A Leisner, Jonathan Plitnick, Navjot Singh, John Kelly, Erasmus Schneider, Erica Lasek-Nesselquist |
| EPI_ISL_854450 | MONTEFIORE MEDICAL CENTER LABORATORIES | Wadsworth Center, New York State Department of Health | Kirsten St. George, Daryl M. Lamson, Alexis Russel, Matthew Shudt, Melissa A Leisner, Jonathan Plitnick, Navjot Singh, John Kelly, Erasmus Schneider, Erica Lasek-Nesselquist |
| EPI_ISL_854451 | WESTCHESTER MEDICAL CENTER | Wadsworth Center, New York State Department of Health | Kirsten St. George, Daryl M. Lamson, Alexis Russel, Matthew Shudt, Melissa A Leisner, Jonathan Plitnick, Navjot Singh, John Kelly, Erasmus Schneider, Erica Lasek-Nesselquist |
| EPI_ISL_854452, EPI_ISL_854453, EPI_ISL_854454, EPI_ISL_854455, EPI_ISL_854456, EPI_ISL_854457, EPI_ISL_854458, EPI_ISL_854459 | MONTEFIORE MEDICAL CENTER LABORATORIES | Wadsworth Center, New York State Department of Health | Kirsten St. George, Daryl M. Lamson, Alexis Russel, Matthew Shudt, Melissa A Leisner, Jonathan Plitnick, Navjot Singh, John Kelly, Erasmus Schneider, Erica Lasek-Nesselquist |
| EPI_ISL_855165 | Quest Diagnostics | Quest Diagnostics | Rosenthal,S.H., Gerasimova,A., Kagan,R.M., Anderson, B., Hua, M., Liu Y., Bernstein, L.E., Livingston, K.E., Perez, A., Shalhout, D.F., Shlyakhter, I.A., Owen, R., Tanpaiboon, P., Lacbawan, F. |
| EPI_ISL_855186, EPI_ISL_855187, EPI_ISL_855188, EPI_ISL_855189, EPI_ISL_855190, EPI_ISL_855191, EPI_ISL_855192, EPI_ISL_855193, EPI_ISL_855194, EPI_ISL_855195, EPI_ISL_855196, EPI_ISL_855197, EPI_ISL_855198, EPI_ISL_855199, EPI_ISL_855200, EPI_ISL_855201, EPI_ISL_855202, EPI_ISL_855203, EPI_ISL_855204, EPI_ISL_855205, EPI_ISL_855206, EPI_ISL_855207, EPI_ISL_855208, EPI_ISL_855209, EPI_ISL_855210, EPI_ISL_855211, EPI_ISL_855212, EPI_ISL_855213, EPI_ISL_855214, EPI_ISL_855215, EPI_ISL_855216, EPI_ISL_855217, EPI_ISL_855218, EPI_ISL_855219, EPI_ISL_855220, EPI_ISL_855221, EPI_ISL_855222, EPI_ISL_855223, EPI_ISL_855224, EPI_ISL_855225, EPI_ISL_855226, EPI_ISL_855227, EPI_ISL_855228, EPI_ISL_855229, EPI_ISL_855230, EPI_ISL_855231, EPI_ISL_855232, EPI_ISL_855233, EPI_ISL_855234, EPI_ISL_855235, EPI_ISL_855236, EPI_ISL_855237, EPI_ISL_855238, EPI_ISL_855239, EPI_ISL_855240, EPI_ISL_855241, EPI_ISL_855242, EPI_ISL_855243, EPI_ISL_855244, EPI_ISL_855245, EPI_ISL_855246, EPI_ISL_855247, EPI_ISL_855248, EPI_ISL_855249, EPI_ISL_855250, EPI_ISL_855251, EPI_ISL_855252, EPI_ISL_855253, EPI_ISL_855254, EPI_ISL_855255, EPI_ISL_855256, EPI_ISL_855257, EPI_ISL_855258, EPI_ISL_855259, EPI_ISL_855260, EPI_ISL_855261, EPI_ISL_855262, EPI_ISL_855263, EPI_ISL_855264, EPI_ISL_855265, EPI_ISL_855266, EPI_ISL_855267, EPI_ISL_855268, EPI_ISL_855269, EPI_ISL_855270, EPI_ISL_855271, EPI_ISL_855272, EPI_ISL_855273, EPI_ISL_855274, EPI_ISL_855275, EPI_ISL_855276, EPI_ISL_855277, EPI_ISL_855278, EPI_ISL_855279, EPI_ISL_855280, EPI_ISL_855281, EPI_ISL_855282, EPI_ISL_855283, EPI_ISL_855284, EPI_ISL_855285, EPI_ISL_855286, EPI_ISL_855287, EPI_ISL_855288, EPI_ISL_855289, EPI_ISL_855290, EPI_ISL_855291, EPI_ISL_855292, EPI_ISL_855293, EPI_ISL_855294, EPI_ISL_855295, EPI_ISL_855296, EPI_ISL_855297, EPI_ISL_855298, EPI_ISL_855299, EPI_ISL_855300, EPI_ISL_855301, EPI_ISL_855302, EPI_ISL_855303, EPI_ISL_855304, EPI_ISL_855305, EPI_ISL_855306, EPI_ISL_855307, EPI_ISL_855308, EPI_ISL_855309, EPI_ISL_855310, EPI_ISL_855311, EPI_ISL_855312, EPI_ISL_855313, EPI_ISL_855314, EPI_ISL_855315, EPI_ISL_855316, EPI_ISL_855317, EPI_ISL_855318, EPI_ISL_855319, EPI_ISL_855320, EPI_ISL_855321, EPI_ISL_855322, EPI_ISL_855323, EPI_ISL_855324, EPI_ISL_855325, EPI_ISL_855326, EPI_ISL_855327, EPI_ISL_855328, EPI_ISL_855329, EPI_ISL_855330, EPI_ISL_855331, EPI_ISL_855332, EPI_ISL_855333, EPI_ISL_855334, EPI_ISL_855335, EPI_ISL_855336, EPI_ISL_855337, EPI_ISL_855338, EPI_ISL_855339 | | | |
| see above | Institute for Computational Biomedicine, Weill Cornell Medicine | Institute for Computational Biomedicine, Weill Cornell Medicine | Butler,D.J., Mozsary,C., Meydan,C., Danko,D., Foox,J., Rosiene,J., Shaiber,A., Afshinnekoo,E., MacKay,M., Sedlazeck,F.J., Ivanov,N.A., Sierra,M., Pohle,D., Zietz,M., Gisladottir,U., Ramlall,V., Westover,C.D., Ryon,K., Young,B., Bhattacharya,C., Ruggiero,P., Langhorst,B.W., Tanner,N., Gawrys,J., Meleshko,D., Xu,D., Steel,P.A.D., Shemesh,A.J., Xiang,J., Thierry-Mieg,J., Thierry-Mieg,D., Schwartz,R.E., Iftner,A., Bezdan,D., Sipley,J., Cong,L., Craney,A., Velu,P., Melnick,A.M., Hajirasouliha,I., Horner,S.M., Iftner,T., Salvatore,M., Loda,M., Westblade,L.F., Cushing,M., Levy,S., Wu,S., Tatonetti,N., Imielinski,M., Rennert,H., Mason,C.E. |
| EPI_ISL_857056, EPI_ISL_857057, EPI_ISL_857058, EPI_ISL_857059, EPI_ISL_857060, EPI_ISL_857061, EPI_ISL_857062 | OCME Office Of Chief Medical Examiner | New York City Public Health Laboratory | Jade Wang, et al. |
| EPI_ISL_857063, EPI_ISL_857064, EPI_ISL_857065 | DOHMH PHL | New York City Public Health Laboratory | Jade Wang, et al. |
| EPI_ISL_857066 | OCME Office Of Chief Medical Examiner | New York City Public Health Laboratory | Jade Wang, et al. |
| EPI_ISL_857067 | DOHMH Corona | New York City Public Health Laboratory | Jade Wang, et al. |
| EPI_ISL_857068 | DOHMH PHL | New York City Public Health Laboratory | Jade Wang, et al. |
| EPI_ISL_857069 | DOHMH Jamaica | New York City Public Health Laboratory | Jade Wang, et al. |
| EPI_ISL_857070 | DOHMH PHL | New York City Public Health Laboratory | Jade Wang, et al. |
| EPI_ISL_857071 | DOHMH Morrisania | New York City Public Health Laboratory | Jade Wang, et al. |
| EPI_ISL_857072 | DOHMH PHL | New York City Public Health Laboratory | Jade Wang, et al. |
| EPI_ISL_857073 | DOHMH Corona | New York City Public Health Laboratory | Jade Wang, et al. |
| EPI_ISL_857074 | DOHMH Morrisania | New York City Public Health Laboratory | Jade Wang, et al. |
| EPI_ISL_857075 | DOHMH Jamaica | New York City Public Health Laboratory | Jade Wang, et al. |
| EPI_ISL_857076 | DOHMH Chelsea | New York City Public Health Laboratory | Jade Wang, et al. |
| EPI_ISL_857077, EPI_ISL_857078, EPI_ISL_857079, EPI_ISL_857080 | DOHMH Central Harlem | New York City Public Health Laboratory | Jade Wang, et al. |
| EPI_ISL_857081 | DOHMH Morrisania | New York City Public Health Laboratory | Jade Wang, et al. |
| EPI_ISL_857082, EPI_ISL_857083, EPI_ISL_857084 | DOHMH Corona | New York City Public Health Laboratory | Jade Wang, et al. |
| EPI_ISL_857085, EPI_ISL_857086 | DOHMH Central Harlem | New York City Public Health Laboratory | Jade Wang, et al. |
| EPI_ISL_857087, EPI_ISL_857088 | DOHMH PHL | New York City Public Health Laboratory | Jade Wang, et al. |
| EPI_ISL_857089 | DOHMH Crown Heights | New York City Public Health Laboratory | Jade Wang, et al. |
| EPI_ISL_857090 | DOHMH Riverside | New York City Public Health Laboratory | Jade Wang, et al. |
| EPI_ISL_857091 | DOHMH Jamaica | New York City Public Health Laboratory | Jade Wang, et al. |
| EPI_ISL_857092 | DOHMH Riverside | New York City Public Health Laboratory | Jade Wang, et al. |
| EPI_ISL_857093 | DOHMH Morrisania | New York City Public Health Laboratory | Jade Wang, et al. |
| EPI_ISL_857094 | DOHMH Central Harlem | New York City Public Health Laboratory | Jade Wang, et al. |
| EPI_ISL_857095 | DOHMH Fort Greene | New York City Public Health Laboratory | Jade Wang, et al. |
| EPI_ISL_857096 | DOHMH Jamaica | New York City Public Health Laboratory | Jade Wang, et al. |
| EPI_ISL_857097 | DOHMH Crown Heights | New York City Public Health Laboratory | Jade Wang, et al. |
| EPI_ISL_857098, EPI_ISL_857099 | DOHMH Morrisania | New York City Public Health Laboratory | Jade Wang, et al. |
| EPI_ISL_857100 | DOHMH Central Harlem | New York City Public Health Laboratory | Jade Wang, et al. |
| EPI_ISL_857101, EPI_ISL_857102, EPI_ISL_857103, EPI_ISL_857104, EPI_ISL_857105 | DOHMH Corona | New York City Public Health Laboratory | Jade Wang, et al. |
| EPI_ISL_857106, EPI_ISL_857107, EPI_ISL_857108, EPI_ISL_857109, EPI_ISL_857110 | DOHMH Chelsea | New York City Public Health Laboratory | Jade Wang, et al. |

| | | | |
|---|---|---|---|
| EPI_ISL_857111 | DOHMH Riverside | New York City Public Health Laboratory | Jade Wang, et al. |
| EPI_ISL_857112, EPI_ISL_857113, EPI_ISL_857114, EPI_ISL_857115 | DOHMH Fort Greene | New York City Public Health Laboratory | Jade Wang, et al. |
| EPI_ISL_857116 | DOHMH Central Harlem | New York City Public Health Laboratory | Jade Wang, et al. |
| EPI_ISL_857117 | DOHMH Chelsea | New York City Public Health Laboratory | Jade Wang, et al. |
| EPI_ISL_857118 | DOHMH Central Harlem | New York City Public Health Laboratory | Jade Wang, et al. |
| EPI_ISL_857119 | DOHMH Morrisania | New York City Public Health Laboratory | Jade Wang, et al. |
| EPI_ISL_857120 | DOHMH Central Harlem | New York City Public Health Laboratory | Jade Wang, et al. |
| EPI_ISL_857121 | DOHMH PHL | New York City Public Health Laboratory | Jade Wang, et al. |
| EPI_ISL_857122, EPI_ISL_857123, EPI_ISL_857124, EPI_ISL_857125 | DOHMH Morrisania | New York City Public Health Laboratory | Jade Wang, et al. |
| EPI_ISL_857126 | DOHMH Central Harlem | New York City Public Health Laboratory | Jade Wang, et al. |
| EPI_ISL_857127, EPI_ISL_857128, EPI_ISL_857129 | DOHMH PHL | New York City Public Health Laboratory | Jade Wang, et al. |
| EPI_ISL_857130, EPI_ISL_857131 | DOHMH Chelsea | New York City Public Health Laboratory | Jade Wang, et al. |
| EPI_ISL_857132, EPI_ISL_857133 | DOHMH Central Harlem | New York City Public Health Laboratory | Jade Wang, et al. |
| EPI_ISL_857134 | DOHMH PHL | New York City Public Health Laboratory | Jade Wang, et al. |
| EPI_ISL_857135 | DOHMH Corona | New York City Public Health Laboratory | Jade Wang, et al. |
| EPI_ISL_857136, EPI_ISL_857137, EPI_ISL_857138, EPI_ISL_857139 | DOHMH Chelsea | New York City Public Health Laboratory | Jade Wang, et al. |
| EPI_ISL_857140, EPI_ISL_857141 | DOHMH Morrisania | New York City Public Health Laboratory | Jade Wang, et al. |
| EPI_ISL_857142 | DOHMH Central Harlem | New York City Public Health Laboratory | Jade Wang, et al. |
| EPI_ISL_857143, EPI_ISL_857144 | DOHMH Riverside | New York City Public Health Laboratory | Jade Wang, et al. |
| EPI_ISL_857145, EPI_ISL_857146, EPI_ISL_857147 | DOHMH Corona | New York City Public Health Laboratory | Jade Wang, et al. |
| EPI_ISL_857148 | DOHMH Fort Greene | New York City Public Health Laboratory | Jade Wang, et al. |
| EPI_ISL_857149, EPI_ISL_857150, EPI_ISL_857151, EPI_ISL_857152 | DOHMH Jamaica | New York City Public Health Laboratory | Jade Wang, et al. |
| EPI_ISL_857153 | DOHMH Corona | New York City Public Health Laboratory | Jade Wang, et al. |
| EPI_ISL_857154, EPI_ISL_857155 | DOHMH Jamaica | New York City Public Health Laboratory | Jade Wang, et al. |
| EPI_ISL_857156 | DOHMH Corona | New York City Public Health Laboratory | Jade Wang, et al. |
| EPI_ISL_857157 | DOHMH PHL | New York City Public Health Laboratory | Jade Wang, et al. |
| EPI_ISL_857158 | DOHMH Corona | New York City Public Health Laboratory | Jade Wang, et al. |
| EPI_ISL_857159, EPI_ISL_857160 | DOHMH Morrisania | New York City Public Health Laboratory | Jade Wang, et al. |
| EPI_ISL_857161, EPI_ISL_857162 | DOHMH PHL | New York City Public Health Laboratory | Jade Wang, et al. |
| EPI_ISL_857163 | DOHMH Central Harlem | New York City Public Health Laboratory | Jade Wang, et al. |
| EPI_ISL_857164 | DOHMH Morrisania | New York City Public Health Laboratory | Jade Wang, et al. |
| EPI_ISL_857165 | DOHMH Corona | New York City Public Health Laboratory | Jade Wang, et al. |
| EPI_ISL_857166, EPI_ISL_857167 | DOHMH Fort Greene | New York City Public Health Laboratory | Jade Wang, et al. |
| EPI_ISL_857168 | DOHMH Corona | New York City Public Health Laboratory | Jade Wang, et al. |
| EPI_ISL_857169, EPI_ISL_857170, EPI_ISL_857171 | DOHMH Morrisania | New York City Public Health Laboratory | Jade Wang, et al. |
| EPI_ISL_857172, EPI_ISL_857173 | DOHMH Crown Heights | New York City Public Health Laboratory | Jade Wang, et al. |
| EPI_ISL_857174, EPI_ISL_857175, EPI_ISL_857176, EPI_ISL_857177 | DOHMH Jamaica | New York City Public Health Laboratory | Jade Wang, et al. |
| EPI_ISL_857178 | DOHMH Riverside | New York City Public Health Laboratory | Jade Wang, et al. |
| EPI_ISL_857179 | DOHMH Jamaica | New York City Public Health Laboratory | Jade Wang, et al. |
| EPI_ISL_857180, EPI_ISL_857181, EPI_ISL_857182, EPI_ISL_857183 | DOHMH Central Harlem | New York City Public Health Laboratory | Jade Wang, et al. |
| EPI_ISL_857184 | DOHMH Riverside | New York City Public Health Laboratory | Jade Wang, et al. |
| EPI_ISL_857185 | DOHMH PHL | New York City Public Health Laboratory | Jade Wang, et al. |
| EPI_ISL_857186 | OCME Office Of Chief Medical Examiner | New York City Public Health Laboratory | Jade Wang, et al. |
| EPI_ISL_857187 | DOHMH PHL | New York City Public Health Laboratory | Jade Wang, et al. |
| EPI_ISL_857188 | Department of Homeless Services | New York City Public Health Laboratory | Jade Wang, et al. |
| EPI_ISL_857189 | DOHMH Corona | New York City Public Health Laboratory | Jade Wang, et al. |
| EPI_ISL_857190 | DOHMH PHL | New York City Public Health Laboratory | Jade Wang, et al. |
| EPI_ISL_857191 | DOHMH Morrisania | New York City Public Health Laboratory | Jade Wang, et al. |
| EPI_ISL_857192 | DOHMH PHL | New York City Public Health Laboratory | Jade Wang, et al. |
| EPI_ISL_857193 | OCME Office Of Chief Medical Examiner | New York City Public Health Laboratory | Jade Wang, et al. |
| EPI_ISL_857194 | DOHMH Central Harlem | New York City Public Health Laboratory | Jade Wang, et al. |
| EPI_ISL_857195 | DOHMH Riverside | New York City Public Health Laboratory | Jade Wang, et al. |

| | | | |
|---|---|---|---|
| EPI_ISL_857196 | DOHMH Crown Heights | New York City Public Health Laboratory | Jade Wang, et al. |
| EPI_ISL_857197 | DOHMH Jamaica | New York City Public Health Laboratory | Jade Wang, et al. |
| EPI_ISL_857198 | Department of Homeless Services | New York City Public Health Laboratory | Jade Wang, et al. |
| EPI_ISL_857199 | OCME Office Of Chief Medical Examiner | New York City Public Health Laboratory | Jade Wang, et al. |
| EPI_ISL_857200 | DOHMH Central Harlem | New York City Public Health Laboratory | Jade Wang, et al. |
| EPI_ISL_857201 | DOHMH Jamaica | New York City Public Health Laboratory | Jade Wang, et al. |
| EPI_ISL_857202 | DOHMH PHL | New York City Public Health Laboratory | Jade Wang, et al. |
| EPI_ISL_857203 | DOHMH Riverside | New York City Public Health Laboratory | Jade Wang, et al. |
| EPI_ISL_857204, EPI_ISL_857205 | DOHMH Central Harlem | New York City Public Health Laboratory | Jade Wang, et al. |
| EPI_ISL_857206 | DOHMH PHL | New York City Public Health Laboratory | Jade Wang, et al. |
| EPI_ISL_857207, EPI_ISL_857208 | DOHMH Morrisania | New York City Public Health Laboratory | Jade Wang, et al. |
| EPI_ISL_857209 | DOHMH Central Harlem | New York City Public Health Laboratory | Jade Wang, et al. |
| EPI_ISL_857210, EPI_ISL_857211, EPI_ISL_857212, EPI_ISL_857213, EPI_ISL_857214, EPI_ISL_857215, EPI_ISL_857216, EPI_ISL_857217 | DOHMH Jamaica | New York City Public Health Laboratory | Jade Wang, et al. |
| EPI_ISL_857218, EPI_ISL_857219, EPI_ISL_857220 | DOHMH Corona | New York City Public Health Laboratory | Jade Wang, et al. |
| EPI_ISL_857221 | DOHMH Fort Greene | New York City Public Health Laboratory | Jade Wang, et al. |
| EPI_ISL_857222, EPI_ISL_857223, EPI_ISL_857224 | DOHMH Jamaica | New York City Public Health Laboratory | Jade Wang, et al. |
| EPI_ISL_857225 | DOHMH Corona | New York City Public Health Laboratory | Jade Wang, et al. |
| EPI_ISL_857226 | DOHMH Jamaica | New York City Public Health Laboratory | Jade Wang, et al. |
| EPI_ISL_857227, EPI_ISL_857228 | DOHMH Riverside | New York City Public Health Laboratory | Jade Wang, et al. |
| EPI_ISL_857229 | DOHMH Morrisania | New York City Public Health Laboratory | Jade Wang, et al. |
| EPI_ISL_857230 | DOHMH Chelsea | New York City Public Health Laboratory | Jade Wang, et al. |
| EPI_ISL_857231, EPI_ISL_857232, EPI_ISL_857233 | DOHMH Central Harlem | New York City Public Health Laboratory | Jade Wang, et al. |
| EPI_ISL_857234, EPI_ISL_857235, EPI_ISL_857236, EPI_ISL_857237 | DOHMH Jamaica | New York City Public Health Laboratory | Jade Wang, et al. |
| EPI_ISL_857238, EPI_ISL_857239, EPI_ISL_857240 | DOHMH Corona | New York City Public Health Laboratory | Jade Wang, et al. |
| EPI_ISL_857241, EPI_ISL_857242, EPI_ISL_857243, EPI_ISL_857244, EPI_ISL_857245, EPI_ISL_857246, EPI_ISL_857247, EPI_ISL_857248, EPI_ISL_857249, EPI_ISL_857250 | DOHMH Jamaica | New York City Public Health Laboratory | Jade Wang, et al. |
| EPI_ISL_857251, EPI_ISL_857252 | DOHMH Morrisania | New York City Public Health Laboratory | Jade Wang, et al. |
| EPI_ISL_857253, EPI_ISL_857254 | DOHMH Chelsea | New York City Public Health Laboratory | Jade Wang, et al. |
| EPI_ISL_857255 | DOHMH Central Harlem | New York City Public Health Laboratory | Jade Wang, et al. |
| EPI_ISL_857256 | DOHMH Fort Greene | New York City Public Health Laboratory | Jade Wang, et al. |
| EPI_ISL_857257 | DOHMH PHL | New York City Public Health Laboratory | Jade Wang, et al. |
| EPI_ISL_857258, EPI_ISL_857259 | DOHMH Central Harlem | New York City Public Health Laboratory | Jade Wang, et al. |
| EPI_ISL_857260, EPI_ISL_857261, EPI_ISL_857262 | DOHMH Chelsea | New York City Public Health Laboratory | Jade Wang, et al. |
| EPI_ISL_857263, EPI_ISL_857264 | DOHMH Corona | New York City Public Health Laboratory | Jade Wang, et al. |
| EPI_ISL_857265 | DOHMH Fort Greene | New York City Public Health Laboratory | Jade Wang, et al. |
| EPI_ISL_857266 | DOHMH PHL | New York City Public Health Laboratory | Jade Wang, et al. |
| EPI_ISL_857267, EPI_ISL_857268, EPI_ISL_857269, EPI_ISL_857270 | DOHMH Jamaica | New York City Public Health Laboratory | Jade Wang, et al. |
| EPI_ISL_857271, EPI_ISL_857272, EPI_ISL_857273 | DOHMH Morrisania | New York City Public Health Laboratory | Jade Wang, et al. |
| EPI_ISL_857274, EPI_ISL_857275, EPI_ISL_857276 | DOHMH Jamaica | New York City Public Health Laboratory | Jade Wang, et al. |
| EPI_ISL_857277 | DOHMH Corona | New York City Public Health Laboratory | Jade Wang, et al. |
| EPI_ISL_857278 | DOHMH PHL | New York City Public Health Laboratory | Jade Wang, et al. |
| EPI_ISL_857279, EPI_ISL_857280 | DOHMH Fort Greene | New York City Public Health Laboratory | Jade Wang, et al. |
| EPI_ISL_857281 | DOHMH Riverside | New York City Public Health Laboratory | Jade Wang, et al. |
| EPI_ISL_857282, EPI_ISL_857283 | DOHMH Morrisania | New York City Public Health Laboratory | Jade Wang, et al. |
| EPI_ISL_857284 | DOHMH PHL | New York City Public Health Laboratory | Jade Wang, et al. |
| EPI_ISL_857285, EPI_ISL_857286 | DOHMH Central Harlem | New York City Public Health Laboratory | Jade Wang, et al. |
| EPI_ISL_857287, EPI_ISL_857288 | Department of Homeless Services | New York City Public Health Laboratory | Jade Wang, et al. |
| EPI_ISL_857289 | OCME Office Of Chief Medical Examiner | New York City Public Health Laboratory | Jade Wang, et al. |
| EPI_ISL_857290 | Department of Homeless Services | New York City Public Health Laboratory | Jade Wang, et al. |

| | | | |
|---|---|---|---|
| EPI_ISL_857291, EPI_ISL_857292 | OCME Office Of Chief Medical Examiner | New York City Public Health Laboratory | Jade Wang, et al. |
| EPI_ISL_857293 | Department of Homeless Services | New York City Public Health Laboratory | Jade Wang, et al. |
| EPI_ISL_857294, EPI_ISL_857295, EPI_ISL_857296, EPI_ISL_857297 | OCME Office Of Chief Medical Examiner | New York City Public Health Laboratory | Jade Wang, et al. |
| EPI_ISL_857298 | Department of Homeless Services | New York City Public Health Laboratory | Jade Wang, et al. |
| EPI_ISL_857299, EPI_ISL_857300 | OCME Office Of Chief Medical Examiner | New York City Public Health Laboratory | Jade Wang, et al. |
| EPI_ISL_857301 | Department of Homeless Services | New York City Public Health Laboratory | Jade Wang, et al. |
| EPI_ISL_857302, EPI_ISL_857303, EPI_ISL_857304, EPI_ISL_857305 | OCME Office Of Chief Medical Examiner | New York City Public Health Laboratory | Jade Wang, et al. |
| EPI_ISL_857306 | Department of Homeless Services | New York City Public Health Laboratory | Jade Wang, et al. |
| EPI_ISL_857307 | OCME Office Of Chief Medical Examiner | New York City Public Health Laboratory | Jade Wang, et al. |
| EPI_ISL_861106, EPI_ISL_861107, EPI_ISL_861108 | KALEIDA CENTER FOR LABORATORY MEDICINE | Wadsworth Center, New York State Department of Health | Kirsten St. George, Daryl M. Lamson, Alexis Russel, Matthew Shudt, Melissa A Leisner, Jonathan Plitnick, Navjot Singh, John Kelly, Erasmus Schneider, Erica Lasek-Nesselquist |
| EPI_ISL_861109, EPI_ISL_861110, EPI_ISL_861111, EPI_ISL_861112, EPI_ISL_861113, EPI_ISL_861114 | New York Presbyterian Hospital | Wadsworth Center, New York State Department of Health | Kirsten St. George, Daryl M. Lamson, Alexis Russel, Matthew Shudt, Melissa A Leisner, Jonathan Plitnick, Navjot Singh, John Kelly, Erasmus Schneider, Erica Lasek-Nesselquist |
| EPI_ISL_861115, EPI_ISL_861116, EPI_ISL_861117 | Wadsworth Center, New York State Department of Health | Wadsworth Center, New York State Department of Health | Kirsten St. George, Daryl M. Lamson, Alexis Russel, Matthew Shudt, Melissa A Leisner, Jonathan Plitnick, Navjot Singh, John Kelly, Erasmus Schneider, Erica Lasek-Nesselquist |
| EPI_ISL_861118, EPI_ISL_861119, EPI_ISL_861120, EPI_ISL_861121, EPI_ISL_861122, EPI_ISL_861123, EPI_ISL_861124, EPI_ISL_861125, EPI_ISL_861126, EPI_ISL_861127, EPI_ISL_861128, EPI_ISL_861129, EPI_ISL_861130, EPI_ISL_861131, EPI_ISL_861132, EPI_ISL_861133 | | | |
| see above | New York Presbyterian Hospital | Wadsworth Center, New York State Department of Health | Kirsten St. George, Daryl M. Lamson, Alexis Russel, Matthew Shudt, Melissa A Leisner, Jonathan Plitnick, Navjot Singh, John Kelly, Erasmus Schneider, Erica Lasek-Nesselquist |
| EPI_ISL_861134 | KALEIDA CENTER FOR LABORATORY MEDICINE | Wadsworth Center, New York State Department of Health | Kirsten St. George, Daryl M. Lamson, Alexis Russel, Matthew Shudt, Melissa A Leisner, Jonathan Plitnick, Navjot Singh, John Kelly, Erasmus Schneider, Erica Lasek-Nesselquist |
| EPI_ISL_861135, EPI_ISL_861136, EPI_ISL_861137, EPI_ISL_861138, EPI_ISL_861139, EPI_ISL_861140, EPI_ISL_861141, EPI_ISL_861142, EPI_ISL_861143 | ADIRONDACK MEDICAL CENTER | Wadsworth Center, New York State Department of Health | Kirsten St. George, Daryl M. Lamson, Alexis Russel, Matthew Shudt, Melissa A Leisner, Jonathan Plitnick, Navjot Singh, John Kelly, Erasmus Schneider, Erica Lasek-Nesselquist |
| EPI_ISL_861144, EPI_ISL_861145, EPI_ISL_861146, EPI_ISL_861147, EPI_ISL_861148, EPI_ISL_861149, EPI_ISL_861150, EPI_ISL_861151, EPI_ISL_861152, EPI_ISL_861153, EPI_ISL_861154, EPI_ISL_861155, EPI_ISL_861156, EPI_ISL_861157, EPI_ISL_861158, EPI_ISL_861159, EPI_ISL_861160, EPI_ISL_861161, EPI_ISL_861162, EPI_ISL_861163, EPI_ISL_861164, EPI_ISL_861165, EPI_ISL_861166, EPI_ISL_861167, EPI_ISL_861168, EPI_ISL_861169, EPI_ISL_861170, EPI_ISL_861171, EPI_ISL_861172, EPI_ISL_861173, EPI_ISL_861174 | | | |
| see above | KALEIDA CENTER FOR LABORATORY MEDICINE | Wadsworth Center, New York State Department of Health | Kirsten St. George, Daryl M. Lamson, Alexis Russel, Matthew Shudt, Melissa A Leisner, Jonathan Plitnick, Navjot Singh, John Kelly, Erasmus Schneider, Erica Lasek-Nesselquist |
| EPI_ISL_861175, EPI_ISL_861176, EPI_ISL_861177, EPI_ISL_861178, EPI_ISL_861179, EPI_ISL_861180, EPI_ISL_861181, EPI_ISL_861182 | BIO-REFERENCE LABORATORIES | Wadsworth Center, New York State Department of Health | Kirsten St. George, Daryl M. Lamson, Alexis Russel, Matthew Shudt, Melissa A Leisner, Jonathan Plitnick, Navjot Singh, John Kelly, Erasmus Schneider, Erica Lasek-Nesselquist |
| EPI_ISL_861183, EPI_ISL_861184 | URMC LABS | Wadsworth Center, New York State Department of Health | Kirsten St. George, Daryl M. Lamson, Alexis Russel, Matthew Shudt, Melissa A Leisner, Jonathan Plitnick, Navjot Singh, John Kelly, Erasmus Schneider, Erica Lasek-Nesselquist |
| EPI_ISL_861185 | BIO-REFERENCE LABORATORIES | Wadsworth Center, New York State Department of Health | Kirsten St. George, Daryl M. Lamson, Alexis Russel, Matthew Shudt, Melissa A Leisner, Jonathan Plitnick, Navjot Singh, John Kelly, Erasmus Schneider, Erica Lasek-Nesselquist |
| EPI_ISL_861186 | NORTH SHORE UNIVERSITY HOSPITAL | Wadsworth Center, New York State Department of Health | Kirsten St. George, Daryl M. Lamson, Alexis Russel, Matthew Shudt, Melissa A Leisner, Jonathan Plitnick, Navjot Singh, John Kelly, Erasmus Schneider, Erica Lasek-Nesselquist |
| EPI_ISL_861187 | BIO-REFERENCE LABORATORIES | Wadsworth Center, New York State Department of Health | Kirsten St. George, Daryl M. Lamson, Alexis Russel, Matthew Shudt, Melissa A Leisner, Jonathan Plitnick, Navjot Singh, John Kelly, Erasmus Schneider, Erica Lasek-Nesselquist |
| EPI_ISL_861188, EPI_ISL_861189, EPI_ISL_861190, EPI_ISL_861191, EPI_ISL_861192, EPI_ISL_861193, EPI_ISL_861194, EPI_ISL_861195, EPI_ISL_861196, EPI_ISL_861197, EPI_ISL_861198 | | | |
| see above | NORTH SHORE UNIVERSITY HOSPITAL | Wadsworth Center, New York State Department of Health | Kirsten St. George, Daryl M. Lamson, Alexis Russel, Matthew Shudt, Melissa A Leisner, Jonathan Plitnick, Navjot Singh, John Kelly, Erasmus Schneider, Erica Lasek-Nesselquist |
| EPI_ISL_861199, EPI_ISL_861200, EPI_ISL_861201, EPI_ISL_861202, EPI_ISL_861203, EPI_ISL_861204, EPI_ISL_861205, EPI_ISL_861206, EPI_ISL_861207, EPI_ISL_861208, EPI_ISL_861209 | | | |
| see above | URMC LABS | Wadsworth Center, New York State Department of Health | Kirsten St. George, Daryl M. Lamson, Alexis Russel, Matthew Shudt, Melissa A Leisner, Jonathan Plitnick, Navjot Singh, John Kelly, Erasmus Schneider, Erica Lasek-Nesselquist |
| EPI_ISL_861210, EPI_ISL_861211, EPI_ISL_861212, EPI_ISL_861213, EPI_ISL_861214, EPI_ISL_861215, EPI_ISL_861216, EPI_ISL_861217, EPI_ISL_861218, EPI_ISL_861219, EPI_ISL_861220, EPI_ISL_861221, EPI_ISL_861222, EPI_ISL_861223, EPI_ISL_861224, EPI_ISL_861225, EPI_ISL_861226, EPI_ISL_861227, EPI_ISL_861228, EPI_ISL_861229, EPI_ISL_861230, EPI_ISL_861231, EPI_ISL_861232 | | | |
| see above | BIO-REFERENCE LABORATORIES | Wadsworth Center, New York State Department of Health | Kirsten St. George, Daryl M. Lamson, Alexis Russel, Matthew Shudt, Melissa A Leisner, Jonathan Plitnick, Navjot Singh, John Kelly, Erasmus Schneider, Erica Lasek-Nesselquist |
| EPI_ISL_861244, EPI_ISL_861245, EPI_ISL_861246, EPI_ISL_861247, EPI_ISL_861248, EPI_ISL_861249, EPI_ISL_861250, EPI_ISL_861251, EPI_ISL_861252, EPI_ISL_861253, EPI_ISL_861254, EPI_ISL_861255, EPI_ISL_861256, EPI_ISL_861257, EPI_ISL_861258, EPI_ISL_861259, EPI_ISL_861260, EPI_ISL_861261, EPI_ISL_861262, EPI_ISL_861263, EPI_ISL_861264, EPI_ISL_861265, EPI_ISL_861266, EPI_ISL_861267, EPI_ISL_861268, EPI_ISL_861269, EPI_ISL_861270, EPI_ISL_861271, EPI_ISL_861272, EPI_ISL_861273, EPI_ISL_861274, EPI_ISL_861275, EPI_ISL_861276, EPI_ISL_861277, EPI_ISL_861278, EPI_ISL_861279, EPI_ISL_861280, EPI_ISL_861281, EPI_ISL_861282, EPI_ISL_861283, EPI_ISL_861284, EPI_ISL_861285, EPI_ISL_861286, EPI_ISL_861287, EPI_ISL_861288, EPI_ISL_861289, EPI_ISL_861290, EPI_ISL_861291, EPI_ISL_861292, EPI_ISL_861293, EPI_ISL_861294, EPI_ISL_861295, EPI_ISL_861296, EPI_ISL_861297, EPI_ISL_861298, EPI_ISL_861299, EPI_ISL_861300, EPI_ISL_861301, EPI_ISL_861302, EPI_ISL_861303, EPI_ISL_861304, EPI_ISL_861305, EPI_ISL_861306, EPI_ISL_861307, EPI_ISL_861308, EPI_ISL_861309, EPI_ISL_861310, EPI_ISL_861311, EPI_ISL_861312, EPI_ISL_861313, EPI_ISL_861314, EPI_ISL_861315, EPI_ISL_861316, EPI_ISL_861317, EPI_ISL_861318, EPI_ISL_861319, EPI_ISL_861320, EPI_ISL_861321, EPI_ISL_861322, EPI_ISL_861323, EPI_ISL_861324, EPI_ISL_861325, EPI_ISL_861326, EPI_ISL_861327, EPI_ISL_861328, EPI_ISL_861329, EPI_ISL_861330, EPI_ISL_861331 | | | |
| see above | MONTEFIORE MEDICAL CENTER LABORATORIES | Wadsworth Center, New York State Department of Health | Kirsten St. George, Daryl M. Lamson, Alexis Russel, Matthew Shudt, Melissa A Leisner, Jonathan Plitnick, Navjot Singh, John Kelly, Erasmus Schneider, Erica Lasek-Nesselquist |
| EPI_ISL_861332 | DOHMH Central Harlem | New York City Public Health Laboratory | Jade Wang, et al. |
| EPI_ISL_861333, EPI_ISL_861334, EPI_ISL_861335, EPI_ISL_861336, EPI_ISL_861337, EPI_ISL_861338, EPI_ISL_861339, EPI_ISL_861340, EPI_ISL_861341, EPI_ISL_861342, EPI_ISL_861343, EPI_ISL_861344, EPI_ISL_861345, EPI_ISL_861346, EPI_ISL_861347, EPI_ISL_861348, EPI_ISL_861349, EPI_ISL_861350, EPI_ISL_861351, EPI_ISL_861352, EPI_ISL_861353, EPI_ISL_861354, EPI_ISL_861355, EPI_ISL_861356, EPI_ISL_861357, EPI_ISL_861358, EPI_ISL_861359, EPI_ISL_861360, EPI_ISL_861361, EPI_ISL_861362, EPI_ISL_861363, EPI_ISL_861364, EPI_ISL_861365, EPI_ISL_861366, EPI_ISL_861367, EPI_ISL_861368, EPI_ISL_861369, EPI_ISL_861370, EPI_ISL_861371, EPI_ISL_861372, EPI_ISL_861373, EPI_ISL_861374, EPI_ISL_861375, EPI_ISL_861376, EPI_ISL_861377, EPI_ISL_861378, EPI_ISL_861379, EPI_ISL_861380, EPI_ISL_861381, EPI_ISL_861382, EPI_ISL_861383, EPI_ISL_861384, EPI_ISL_861385, EPI_ISL_861386, EPI_ISL_861387, EPI_ISL_861388, EPI_ISL_861389, EPI_ISL_861390, EPI_ISL_861391, EPI_ISL_861392, EPI_ISL_861393, EPI_ISL_861394, EPI_ISL_861395, EPI_ISL_861396, EPI_ISL_861397, EPI_ISL_861398, EPI_ISL_861399, EPI_ISL_861400, EPI_ISL_861401, EPI_ISL_861402, EPI_ISL_861403, EPI_ISL_861404, EPI_ISL_861405, EPI_ISL_861406, EPI_ISL_861407, EPI_ISL_861408, EPI_ISL_861409, EPI_ISL_861410, EPI_ISL_861411, EPI_ISL_861412 | | | |
| see above | WESTCHESTER MEDICAL CENTER | Wadsworth Center, New York State Department of Health | Kirsten St. George, Daryl M. Lamson, Alexis Russel, Matthew Shudt, Melissa A Leisner, Jonathan Plitnick, Navjot Singh, John Kelly, Erasmus Schneider, Erica Lasek-Nesselquist |
| EPI_ISL_861413, EPI_ISL_861414, EPI_ISL_861415, EPI_ISL_861416, | NORTH SHORE UNIVERSITY HOSPITAL | Wadsworth Center, New York State Department of Health | Kirsten St. George, Daryl M. Lamson, Alexis Russel, Matthew Shudt, Melissa A Leisner, Jonathan Plitnick, Navjot Singh, John Kelly, Erasmus Schneider, Erica Lasek-Nesselquist |

| | | | |
|---|---|---|---|
| EPI_ISL_861417, EPI_ISL_861418 | | | |
| EPI_ISL_861758 | Murphy Medical Associates | Grubaugh Lab - Yale School of Public Health | Tara Alpert, Joseph Fauver, Anderson Brito, Mallery Breban, Anne Wyllie, Chantal Vogels, Mary Petrone, Annie Watkins, Chaney Kalinich, Isabel Ott, Nathan Grubaugh |
| EPI_ISL_876032, EPI_ISL_876033, EPI_ISL_876034, EPI_ISL_876035, EPI_ISL_876036, EPI_ISL_876037, EPI_ISL_876038, EPI_ISL_876039, EPI_ISL_876040, EPI_ISL_876041 | Montefiore Medical Center | Albert Einstein College of Medicine, Dept. of Microbiology & Immunology, Chandran lab | J. Maximilian Fels, Saad Khan, Ryan Forster, Karin A. Skalina, Surksha Sirichand, Amy S. Fox, Aviv Bergman, William B. Mitchell, Lucia R. Wolgast, Wendy Szymczak, Robert H. Bortz III, M. Eugenia Dieterle, Catalina Florez, Denise Haslwanter, Rohit K. Jangra, Ethan Laudermilch, Ariel S. Wirchnianski, Jason Barnhill, David L. Goldman, Hnin Khine, D. Yitzchak Goldstein, Johanna P. Daily, Kartik Chandran, Libusha Kelly |
| EPI_ISL_876519 | DOHMH Jamaica | New York City Public Health Laboratory | Jade Wang, et al. |
| EPI_ISL_876711, EPI_ISL_876712 | Helix/Illumina | Genomics and Discovery, Respiratory Viruses Branch, Division of Viral Diseases, Centers for Disease Control and Prevention | Peter W. Cook,Dhwani Batra,Ben L. Rambo-Martin,Eileen de Feo,Jan Antico,Christine Tran,Matthew Tolentino,Shannon Wickline,Kim Gietzen,Brad Sickler,Jingtao Liu,Eric Allen,Phil Febbo,Summer Galloway,Nicole L. Washington,Simon White,Geraint Levan,Kelly Schiabor Barrett,Elizabeth Cirulli,Alexandre Bolze,Ary Ascencio,Charlotte Rivera-Garcia,Ryan Cho,Jason Nguyen,Sherry Wang,Jimmy Ramirez,Tyler Cassens,Efren Sandoval,Magnus Isaksson,William Lee,David Becker,Marc Laurent,James Lu,Clinton R. Paden,Suxiang Tong,Duncan MacCannell, |
| EPI_ISL_876925, EPI_ISL_876951, EPI_ISL_877121 | Quest Diagnostics | Quest Diagnostics | Rosenthal,S.H., Gerasimova,A., Kagan,R.M., Anderson, B., Hua, M., Liu Y., Bernstein, L.E., Livingston, K.E., Perez, A., Shalhout, D.F., Shlyakhter, I.A., Owen, R., Tanpaiboon, P., Lacbawan, F. |
| EPI_ISL_883307, EPI_ISL_883308 | Pandemic Response Laboratory | New York City Public Health Laboratory | Jade Wang, et al. |
| EPI_ISL_883309 | DOHMH Central Harlem | New York City Public Health Laboratory | Jade Wang, et al. |
| EPI_ISL_883310 | DOHMH Jamaica | New York City Public Health Laboratory | Jade Wang, et al. |
| EPI_ISL_883311 | Pandemic Response Laboratory | New York City Public Health Laboratory | Jade Wang, et al. |
| EPI_ISL_883312, EPI_ISL_883313 | DOHMH Corona | New York City Public Health Laboratory | Jade Wang, et al. |
| EPI_ISL_883314 | DOHMH Jamaica | New York City Public Health Laboratory | Jade Wang, et al. |
| EPI_ISL_883315, EPI_ISL_883316 | OCME Office Of Chief Medical Examiner | New York City Public Health Laboratory | Jade Wang, et al. |
| EPI_ISL_883317 | Pandemic Response Laboratory | New York City Public Health Laboratory | Jade Wang, et al. |
| EPI_ISL_883318, EPI_ISL_883319, EPI_ISL_883320, EPI_ISL_883321, EPI_ISL_883322, EPI_ISL_883323 | DOHMH Jamaica | New York City Public Health Laboratory | Jade Wang, et al. |
| EPI_ISL_883324 | DOHMH Central Harlem | New York City Public Health Laboratory | Jade Wang, et al. |
| EPI_ISL_883325 | OCME Office Of Chief Medical Examiner | New York City Public Health Laboratory | Jade Wang, et al. |
| EPI_ISL_883326 | Department of Homeless Services | New York City Public Health Laboratory | Jade Wang, et al. |
| EPI_ISL_883327 | DOHMH Central Harlem | New York City Public Health Laboratory | Jade Wang, et al. |
| EPI_ISL_883328 | OCME Office Of Chief Medical Examiner | New York City Public Health Laboratory | Jade Wang, et al. |
| EPI_ISL_883329 | DOHMH Central Harlem | New York City Public Health Laboratory | Jade Wang, et al. |
| EPI_ISL_883330 | OCME Office Of Chief Medical Examiner | New York City Public Health Laboratory | Jade Wang, et al. |
| EPI_ISL_883331 | DOHMH Morrisania | New York City Public Health Laboratory | Jade Wang, et al. |
| EPI_ISL_883332 | Department of Homeless Services | New York City Public Health Laboratory | Jade Wang, et al. |
| EPI_ISL_883333, EPI_ISL_883334 | DOHMH Riverside | New York City Public Health Laboratory | Jade Wang, et al. |
| EPI_ISL_883335 | DOHMH Chelsea | New York City Public Health Laboratory | Jade Wang, et al. |
| EPI_ISL_883336 | OCME Office Of Chief Medical Examiner | New York City Public Health Laboratory | Jade Wang, et al. |
| EPI_ISL_883337 | DOHMH Crown Heights | New York City Public Health Laboratory | Jade Wang, et al. |
| EPI_ISL_883338 | DOHMH Morrisania | New York City Public Health Laboratory | Jade Wang, et al. |
| EPI_ISL_883339 | DOHMH Chelsea | New York City Public Health Laboratory | Jade Wang, et al. |
| EPI_ISL_883340, EPI_ISL_883341, EPI_ISL_883342, EPI_ISL_883343 | DOHMH PHL | New York City Public Health Laboratory | Jade Wang, et al. |
| EPI_ISL_883344, EPI_ISL_883345 | OCME Office Of Chief Medical Examiner | New York City Public Health Laboratory | Jade Wang, et al. |
| EPI_ISL_883346 | DOHMH PHL | New York City Public Health Laboratory | Jade Wang, et al. |
| EPI_ISL_883347 | DOHMH Morrisania | New York City Public Health Laboratory | Jade Wang, et al. |
| EPI_ISL_883348 | DOHMH Corona | New York City Public Health Laboratory | Jade Wang, et al. |
| EPI_ISL_883349 | DOHMH Jamaica | New York City Public Health Laboratory | Jade Wang, et al. |
| EPI_ISL_883350, EPI_ISL_883351, EPI_ISL_883352, EPI_ISL_883353 | OCME Office Of Chief Medical Examiner | New York City Public Health Laboratory | Jade Wang, et al. |
| EPI_ISL_883354, EPI_ISL_883355, EPI_ISL_883356 | DOHMH Corona | New York City Public Health Laboratory | Jade Wang, et al. |
| EPI_ISL_883357, EPI_ISL_883358, EPI_ISL_883359, EPI_ISL_883360 | DOHMH Crown Heights | New York City Public Health Laboratory | Jade Wang, et al. |
| EPI_ISL_883361, EPI_ISL_883362, EPI_ISL_883363 | DOHMH PHL | New York City Public Health Laboratory | Jade Wang, et al. |
| EPI_ISL_883364 | DOHMH Crown Heights | New York City Public Health Laboratory | Jade Wang, et al. |
| EPI_ISL_883365, EPI_ISL_883366, EPI_ISL_883367 | DOHMH Morrisania | New York City Public Health Laboratory | Jade Wang, et al. |
| EPI_ISL_883368, EPI_ISL_883369 | DOHMH Chelsea | New York City Public Health Laboratory | Jade Wang, et al. |
| EPI_ISL_883370 | DOHMH Crown Heights | New York City Public Health Laboratory | Jade Wang, et al. |
| EPI_ISL_883371, EPI_ISL_883372 | DOHMH Central Harlem | New York City Public Health Laboratory | Jade Wang, et al. |
| EPI_ISL_883373, EPI_ISL_883374, EPI_ISL_883375 | DOHMH Jamaica | New York City Public Health Laboratory | Jade Wang, et al. |
| EPI_ISL_883376, EPI_ISL_883377, | DOHMH Central Harlem | New York City Public Health Laboratory | Jade Wang, et al. |

| | | | |
|---|---|---|---|
| EPI_ISL_883378, EPI_ISL_883379 | | | |
| EPI_ISL_883380, EPI_ISL_883381, EPI_ISL_883382 | DOHMH PHL | New York City Public Health Laboratory | Jade Wang, et al. |
| EPI_ISL_883383, EPI_ISL_883384, EPI_ISL_883385, EPI_ISL_883386, EPI_ISL_883387, EPI_ISL_883388, EPI_ISL_883389 | Pandemic Response Laboratory | New York City Public Health Laboratory | Jade Wang, et al. |
| EPI_ISL_883390, EPI_ISL_883391, EPI_ISL_883392, EPI_ISL_883393, EPI_ISL_883394 | OCME Office Of Chief Medical Examiner | New York City Public Health Laboratory | Jade Wang, et al. |
| EPI_ISL_883395, EPI_ISL_883396 | Department of Homeless Services | New York City Public Health Laboratory | Jade Wang, et al. |
| EPI_ISL_883397, EPI_ISL_883398, EPI_ISL_883399, EPI_ISL_883400, EPI_ISL_883401, EPI_ISL_883402 | OCME Office Of Chief Medical Examiner | New York City Public Health Laboratory | Jade Wang, et al. |
| EPI_ISL_883403 | Department of Homeless Services | New York City Public Health Laboratory | Jade Wang, et al. |
| EPI_ISL_883404, EPI_ISL_883405, EPI_ISL_883406, EPI_ISL_883407, EPI_ISL_883408 | OCME Office Of Chief Medical Examiner | New York City Public Health Laboratory | Jade Wang, et al. |
| EPI_ISL_883409 | DOHMH Central Harlem | New York City Public Health Laboratory | Jade Wang, et al. |
| EPI_ISL_883410, EPI_ISL_883411 | DOHMH Corona | New York City Public Health Laboratory | Jade Wang, et al. |
| EPI_ISL_883412 | DOHMH Crown Heights | New York City Public Health Laboratory | Jade Wang, et al. |
| EPI_ISL_883413, EPI_ISL_883414, EPI_ISL_883415 | DOHMH Central Harlem | New York City Public Health Laboratory | Jade Wang, et al. |
| EPI_ISL_883416, EPI_ISL_883417 | DOHMH Fort Greene | New York City Public Health Laboratory | Jade Wang, et al. |
| EPI_ISL_883418, EPI_ISL_883419, EPI_ISL_883420, EPI_ISL_883421, EPI_ISL_883422, EPI_ISL_883423 | DOHMH Jamaica | New York City Public Health Laboratory | Jade Wang, et al. |
| EPI_ISL_883424 | DOHMH PHL | New York City Public Health Laboratory | Jade Wang, et al. |
| EPI_ISL_883425 | BOSTON HEART DIAGNOSTICS CORP | Wadsworth Center, New York State Department of Health | Kirsten St. George, Daryl M. Lamson, Alexis Russel, Matthew Shudt, Melissa A Leisner, Jonathan Plitnick, Navjot Singh, John Kelly, Erasmus Schneider, Erica Lasek-Nesselquist |
| EPI_ISL_883426, EPI_ISL_883427, EPI_ISL_883428 | NORTHWELL HEALTH LABORATORIES | Wadsworth Center, New York State Department of Health | Kirsten St. George, Daryl M. Lamson, Alexis Russel, Matthew Shudt, Melissa A Leisner, Jonathan Plitnick, Navjot Singh, John Kelly, Erasmus Schneider, Erica Lasek-Nesselquist |
| EPI_ISL_883429, EPI_ISL_883430 | BOSTON HEART DIAGNOSTICS CORP | Wadsworth Center, New York State Department of Health | Kirsten St. George, Daryl M. Lamson, Alexis Russel, Matthew Shudt, Melissa A Leisner, Jonathan Plitnick, Navjot Singh, John Kelly, Erasmus Schneider, Erica Lasek-Nesselquist |
| EPI_ISL_883431, EPI_ISL_883432, EPI_ISL_883433, EPI_ISL_883434, EPI_ISL_883435 | NORTHWELL HEALTH LABORATORIES | Wadsworth Center, New York State Department of Health | Kirsten St. George, Daryl M. Lamson, Alexis Russel, Matthew Shudt, Melissa A Leisner, Jonathan Plitnick, Navjot Singh, John Kelly, Erasmus Schneider, Erica Lasek-Nesselquist |
| EPI_ISL_883436, EPI_ISL_883437, EPI_ISL_883438, EPI_ISL_883439, EPI_ISL_883440, EPI_ISL_883441, EPI_ISL_883442, EPI_ISL_883443, EPI_ISL_883444 | ADIRONDACK MEDICAL CENTER | Wadsworth Center, New York State Department of Health | Kirsten St. George, Daryl M. Lamson, Alexis Russel, Matthew Shudt, Melissa A Leisner, Jonathan Plitnick, Navjot Singh, John Kelly, Erasmus Schneider, Erica Lasek-Nesselquist |
| EPI_ISL_883445, EPI_ISL_883446, EPI_ISL_883447, EPI_ISL_883448, EPI_ISL_883449, EPI_ISL_883450, EPI_ISL_883451, EPI_ISL_883452, EPI_ISL_883453, EPI_ISL_883454, EPI_ISL_883455, EPI_ISL_883456, EPI_ISL_883457, EPI_ISL_883458, EPI_ISL_883459, EPI_ISL_883460, EPI_ISL_883461, EPI_ISL_883462, EPI_ISL_883463, EPI_ISL_883464, EPI_ISL_883465, EPI_ISL_883466, EPI_ISL_883467, EPI_ISL_883468, EPI_ISL_883469, EPI_ISL_883470, EPI_ISL_883471, EPI_ISL_883472 | | | |
| see above | NORTHWELL HEALTH LABORATORIES | Wadsworth Center, New York State Department of Health | Kirsten St. George, Daryl M. Lamson, Alexis Russel, Matthew Shudt, Melissa A Leisner, Jonathan Plitnick, Navjot Singh, John Kelly, Erasmus Schneider, Erica Lasek-Nesselquist |
| EPI_ISL_883473 | ADIRONDACK MEDICAL CENTER | Wadsworth Center, New York State Department of Health | Kirsten St. George, Daryl M. Lamson, Alexis Russel, Matthew Shudt, Melissa A Leisner, Jonathan Plitnick, Navjot Singh, John Kelly, Erasmus Schneider, Erica Lasek-Nesselquist |
| EPI_ISL_883474, EPI_ISL_883475, EPI_ISL_883476, EPI_ISL_883477, EPI_ISL_883478, EPI_ISL_883479, EPI_ISL_883480, EPI_ISL_883481, EPI_ISL_883482, EPI_ISL_883483, EPI_ISL_883484, EPI_ISL_883485, EPI_ISL_883486, EPI_ISL_883487, EPI_ISL_883488, EPI_ISL_883489, EPI_ISL_883490, EPI_ISL_883491, EPI_ISL_883492, EPI_ISL_883493, EPI_ISL_883494, EPI_ISL_883495, EPI_ISL_883496, EPI_ISL_883497, EPI_ISL_883498 | | | |
| see above | NORTHWELL HEALTH LABORATORIES | Wadsworth Center, New York State Department of Health | Kirsten St. George, Daryl M. Lamson, Alexis Russel, Matthew Shudt, Melissa A Leisner, Jonathan Plitnick, Navjot Singh, John Kelly, Erasmus Schneider, Erica Lasek-Nesselquist |
| EPI_ISL_884008, EPI_ISL_884009, EPI_ISL_884010, EPI_ISL_884011, EPI_ISL_884012 | ALBANY MEDICAL CENTER HOSPITAL CLINICAL LABORATORIES | Wadsworth Center, New York State Department of Health | Kirsten St. George, Daryl M. Lamson, Alexis Russel, Matthew Shudt, Melissa A Leisner, Jonathan Plitnick, Navjot Singh, John Kelly, Erasmus Schneider, Erica Lasek-Nesselquist |
| EPI_ISL_884013, EPI_ISL_884014, EPI_ISL_884015, EPI_ISL_884016 | Wadsworth Center, New York State Department of Health | Wadsworth Center, New York State Department of Health | Kirsten St. George, Daryl M. Lamson, Alexis Russel, Matthew Shudt, Melissa A Leisner, Jonathan Plitnick, Navjot Singh, John Kelly, Erasmus Schneider, Erica Lasek-Nesselquist |
| EPI_ISL_884017, EPI_ISL_884018, EPI_ISL_884019, EPI_ISL_884020, EPI_ISL_884021, EPI_ISL_884022, EPI_ISL_884023, EPI_ISL_884024, EPI_ISL_884025, EPI_ISL_884026, EPI_ISL_884027, EPI_ISL_884028, EPI_ISL_884029, EPI_ISL_884030, EPI_ISL_884031 | | | |
| see above | ALBANY MEDICAL CENTER HOSPITAL CLINICAL LABORATORIES | Wadsworth Center, New York State Department of Health | Kirsten St. George, Daryl M. Lamson, Alexis Russel, Matthew Shudt, Melissa A Leisner, Jonathan Plitnick, Navjot Singh, John Kelly, Erasmus Schneider, Erica Lasek-Nesselquist |
| EPI_ISL_884032, EPI_ISL_884033, EPI_ISL_884034, EPI_ISL_884035, EPI_ISL_884036, EPI_ISL_884037, EPI_ISL_884038, EPI_ISL_884039, EPI_ISL_884040, EPI_ISL_884041 | Wadsworth Center, New York State Department of Health | Wadsworth Center, New York State Department of Health | Kirsten St. George, Daryl M. Lamson, Alexis Russel, Matthew Shudt, Melissa A Leisner, Jonathan Plitnick, Navjot Singh, John Kelly, Erasmus Schneider, Erica Lasek-Nesselquist |
| EPI_ISL_884042, EPI_ISL_884043, EPI_ISL_884044, EPI_ISL_884045, EPI_ISL_884046 | ALBANY MEDICAL CENTER HOSPITAL CLINICAL LABORATORIES | Wadsworth Center, New York State Department of Health | Kirsten St. George, Daryl M. Lamson, Alexis Russel, Matthew Shudt, Melissa A Leisner, Jonathan Plitnick, Navjot Singh, John Kelly, Erasmus Schneider, Erica Lasek-Nesselquist |
| EPI_ISL_884047 | Wadsworth Center, New York State Department of Health | Wadsworth Center, New York State Department of Health | Kirsten St. George, Daryl M. Lamson, Alexis Russel, Matthew Shudt, Melissa A Leisner, Jonathan Plitnick, Navjot Singh, John Kelly, Erasmus Schneider, Erica Lasek-Nesselquist |
| EPI_ISL_884048, EPI_ISL_884049, EPI_ISL_884050, EPI_ISL_884051, | ALBANY MEDICAL CENTER HOSPITAL CLINICAL LABORATORIES | Wadsworth Center, New York State Department of Health | Kirsten St. George, Daryl M. Lamson, Alexis Russel, Matthew Shudt, Melissa A Leisner, Jonathan Plitnick, Navjot Singh, John Kelly, Erasmus Schneider, Erica Lasek-Nesselquist |

| | | | |
|---|---|---|---|
| EPI_ISL_884052, EPI_ISL_884053 | | | |
| EPI_ISL_884054 | TEMPUS LABS INC | Wadsworth Center, New York State Department of Health | Kirsten St. George, Daryl M. Lamson, Alexis Russel, Matthew Shudt, Melissa A Leisner, Jonathan Plitnick, Navjot Singh, John Kelly, Erasmus Schneider, Erica Lasek-Nesselquist |
| EPI_ISL_884055, EPI_ISL_884056, EPI_ISL_884057, EPI_ISL_884058 | Wadsworth Center, New York State Department of Health | Wadsworth Center, New York State Department of Health | Kirsten St. George, Daryl M. Lamson, Alexis Russel, Matthew Shudt, Melissa A Leisner, Jonathan Plitnick, Navjot Singh, John Kelly, Erasmus Schneider, Erica Lasek-Nesselquist |
| EPI_ISL_884059, EPI_ISL_884060, EPI_ISL_884061, EPI_ISL_884062, EPI_ISL_884063, EPI_ISL_884064, EPI_ISL_884065, EPI_ISL_884066, EPI_ISL_884067, EPI_ISL_884068, EPI_ISL_884069, EPI_ISL_884070, EPI_ISL_884071, EPI_ISL_884072, EPI_ISL_884073, EPI_ISL_884074, EPI_ISL_884075, EPI_ISL_884076, EPI_ISL_884077, EPI_ISL_884078, EPI_ISL_884079 | | | |
| see above | ALBANY MEDICAL CENTER HOSPITAL CLINICAL LABORATORIES | Wadsworth Center, New York State Department of Health | Kirsten St. George, Daryl M. Lamson, Alexis Russel, Matthew Shudt, Melissa A Leisner, Jonathan Plitnick, Navjot Singh, John Kelly, Erasmus Schneider, Erica Lasek-Nesselquist |
| EPI_ISL_884080 | Wadsworth Center, New York State Department of Health | Wadsworth Center, New York State Department of Health | Kirsten St. George, Daryl M. Lamson, Alexis Russel, Matthew Shudt, Melissa A Leisner, Jonathan Plitnick, Navjot Singh, John Kelly, Erasmus Schneider, Erica Lasek-Nesselquist |
| EPI_ISL_884081, EPI_ISL_884082, EPI_ISL_884083, EPI_ISL_884084, EPI_ISL_884085 | ALBANY MEDICAL CENTER HOSPITAL CLINICAL LABORATORIES | Wadsworth Center, New York State Department of Health | Kirsten St. George, Daryl M. Lamson, Alexis Russel, Matthew Shudt, Melissa A Leisner, Jonathan Plitnick, Navjot Singh, John Kelly, Erasmus Schneider, Erica Lasek-Nesselquist |
| EPI_ISL_884086 | Wadsworth Center, New York State Department of Health | Wadsworth Center, New York State Department of Health | Kirsten St. George, Daryl M. Lamson, Alexis Russel, Matthew Shudt, Melissa A Leisner, Jonathan Plitnick, Navjot Singh, John Kelly, Erasmus Schneider, Erica Lasek-Nesselquist |
| EPI_ISL_884390, EPI_ISL_884391, EPI_ISL_884392, EPI_ISL_884393 | Infectious Diseases, Quest Diagnostics | Infectious Diseases, Quest Diagnostics | Rosenthal,S.H., Gerasimova,A., Kagan,R.M., Anderson,B., Bernstein,L.E., Livingston,K.E., Hua,M., Liu,Y., Shalhout,D.F., Owen,R., Lacbawan,F. |
| EPI_ISL_884713, EPI_ISL_884795 | Respiratory Viruses Branch, Centers for Disease Control and Prevention | Respiratory Viruses Branch, Centers for Disease Control and Prevention | Cook,P.W., Batra,D., Rambo-Martin,B.L., de Feo,E., Antico,J., Tran,C., Tolentino,M., Wickline,S., Gietzen,K., Sickler,B., Liu,J., Allen,E., Febbo,P., Galloway,S., Washington,N.L., White,S., Levan,G., Barret,K.S., Cirulli,E., Bolze,A., Ascencio,A., Rivera-Garcia,C., Cho,R., Nguyen,J., Wang,S., Ramirez,J., Cassens,T., Sandoval,E., Isaksson,M., Lee,W., Becker,D., Laurent,M., Lu,J., Paden,C.R., Tong,S., MacCannell,D. |
| EPI_ISL_886231, EPI_ISL_886365, EPI_ISL_886366, EPI_ISL_886405, EPI_ISL_886508, EPI_ISL_886563, EPI_ISL_886666, EPI_ISL_886772, EPI_ISL_886830, EPI_ISL_886837, EPI_ISL_886838, EPI_ISL_886892, EPI_ISL_886918, EPI_ISL_886919, EPI_ISL_886920, EPI_ISL_886927, EPI_ISL_886991, EPI_ISL_886998, EPI_ISL_887004, EPI_ISL_887034, EPI_ISL_887036, EPI_ISL_887046, EPI_ISL_887075, EPI_ISL_887087, EPI_ISL_887102, EPI_ISL_887601, EPI_ISL_887612, EPI_ISL_887631, EPI_ISL_887660, EPI_ISL_887662, EPI_ISL_887665, EPI_ISL_887711, EPI_ISL_887770, EPI_ISL_887792, EPI_ISL_887814, EPI_ISL_887894, EPI_ISL_888114, EPI_ISL_888141, EPI_ISL_888144, EPI_ISL_888146, EPI_ISL_888172, EPI_ISL_888229, EPI_ISL_888271, EPI_ISL_888349, EPI_ISL_888367, EPI_ISL_888405, EPI_ISL_888416, EPI_ISL_888452, EPI_ISL_888456, EPI_ISL_888481, EPI_ISL_888513, EPI_ISL_888562 | | | |
| see above | Labcorp | Genomics and Discovery, Respiratory Viruses Branch, Division of Viral Diseases, Centers for Disease Control and Prevention | Peter W. Cook,Dhwani Batra,Ben L. Rambo-Martin,Summer Galloway,Brian Krueger,Minoo Agarwal,Eyad Almasri,Debbie Boles,Ayla Burns,Nuthawin Charoensri,Oren Cohen,Susan Countryman,Mary Ann Cristobal,Bobbi Croy,Suzanne Dale,Hrushikesh Deshmukh,Amanda Douglas,Vincent Drouillon,Marcia Eisenberg,Howard Engler,Rama Ghatti,Prashant Gupta,Susan Hicks,Jake Humphrey,Lax Iyer,Manoj Jain,Mohan Kolli,Tim Kuphal,Stanley Letovsky,Michael Levandoski,Craig Lukasik,Jonathan Meltzer,Brian Norvell,Mindy Nye,Scott Parker,Christos Petropoulos,John Pruitt,Steven Ragan,Scott Ryan,Mike Sapeta,Jana Schroth,Suresh Babu Selvaraju,Goran Stevovic,Amanda Suchanek,Andrea Throop,Lyndon Tilson,Thomas Urban,Joe Voshell,Kimberly Wagner,Jonathan Williams,Mary Williamson,Qian Zeng,Tricia Zwiefelhofer,Clinton R. Paden,Suxiang Tong,Duncan MacCannell, |
| EPI_ISL_891212 | Population Medicine and Diagnostic Sciences, Cornell University | Population Medicine and Diagnostic Sciences, Cornell University | Cardia Caserta,L., Mitchel,P.K., Plachorzyk,E. and Diel,D.G. |
| EPI_ISL_896216, EPI_ISL_896217 | Columbia University Irving Medical Center | Wadsworth Center, New York State Department of Health | Kirsten St. George, Daryl M. Lamson, Alexis Russel, Matthew Shudt, Melissa A Leisner, Jonathan Plitnick, Navjot Singh, John Kelly, Erasmus Schneider, Erica Lasek-Nesselquist |
| EPI_ISL_896218, EPI_ISL_896219, EPI_ISL_896220, EPI_ISL_896221, EPI_ISL_896222, EPI_ISL_896223, EPI_ISL_896224, EPI_ISL_896225 | SUNY UPSTATE MEDICAL UNIVERSITY | Wadsworth Center, New York State Department of Health | Kirsten St. George, Daryl M. Lamson, Alexis Russel, Matthew Shudt, Melissa A Leisner, Jonathan Plitnick, Navjot Singh, John Kelly, Erasmus Schneider, Erica Lasek-Nesselquist |
| EPI_ISL_896226 | TEMPUS LABS INC | Wadsworth Center, New York State Department of Health | Kirsten St. George, Daryl M. Lamson, Alexis Russel, Matthew Shudt, Melissa A Leisner, Jonathan Plitnick, Navjot Singh, John Kelly, Erasmus Schneider, Erica Lasek-Nesselquist |
| EPI_ISL_896227 | Columbia University Irving Medical Center | Wadsworth Center, New York State Department of Health | Kirsten St. George, Daryl M. Lamson, Alexis Russel, Matthew Shudt, Melissa A Leisner, Jonathan Plitnick, Navjot Singh, John Kelly, Erasmus Schneider, Erica Lasek-Nesselquist |
| EPI_ISL_896228, EPI_ISL_896229, EPI_ISL_896230 | SUNY UPSTATE MEDICAL UNIVERSITY | Wadsworth Center, New York State Department of Health | Kirsten St. George, Daryl M. Lamson, Alexis Russel, Matthew Shudt, Melissa A Leisner, Jonathan Plitnick, Navjot Singh, John Kelly, Erasmus Schneider, Erica Lasek-Nesselquist |
| EPI_ISL_896231, EPI_ISL_896232, EPI_ISL_896233, EPI_ISL_896234 | Columbia University Irving Medical Center | Wadsworth Center, New York State Department of Health | Kirsten St. George, Daryl M. Lamson, Alexis Russel, Matthew Shudt, Melissa A Leisner, Jonathan Plitnick, Navjot Singh, John Kelly, Erasmus Schneider, Erica Lasek-Nesselquist |
| EPI_ISL_896235 | SUNY UPSTATE MEDICAL UNIVERSITY | Wadsworth Center, New York State Department of Health | Kirsten St. George, Daryl M. Lamson, Alexis Russel, Matthew Shudt, Melissa A Leisner, Jonathan Plitnick, Navjot Singh, John Kelly, Erasmus Schneider, Erica Lasek-Nesselquist |
| EPI_ISL_896236, EPI_ISL_896237, EPI_ISL_896238, EPI_ISL_896239, EPI_ISL_896240, EPI_ISL_896241, EPI_ISL_896242, EPI_ISL_896243, EPI_ISL_896244, EPI_ISL_896245, EPI_ISL_896246 | | | |
| see above | Columbia University Irving Medical Center | Wadsworth Center, New York State Department of Health | Kirsten St. George, Daryl M. Lamson, Alexis Russel, Matthew Shudt, Melissa A Leisner, Jonathan Plitnick, Navjot Singh, John Kelly, Erasmus Schneider, Erica Lasek-Nesselquist |
| EPI_ISL_896247, EPI_ISL_896248, EPI_ISL_896249, EPI_ISL_896250, EPI_ISL_896251, EPI_ISL_896252, EPI_ISL_896253, EPI_ISL_896254, EPI_ISL_896255, EPI_ISL_896256, EPI_ISL_896257, EPI_ISL_896258, EPI_ISL_896259, EPI_ISL_896260, EPI_ISL_896261, EPI_ISL_896262, EPI_ISL_896263, EPI_ISL_896264, EPI_ISL_896265, EPI_ISL_896266, EPI_ISL_896267, EPI_ISL_896268, EPI_ISL_896269, EPI_ISL_896270, EPI_ISL_896271, EPI_ISL_896272, EPI_ISL_896273, EPI_ISL_896274, EPI_ISL_896275, EPI_ISL_896276, EPI_ISL_896277, EPI_ISL_896278, EPI_ISL_896279, EPI_ISL_896280, EPI_ISL_896281, EPI_ISL_896282, EPI_ISL_896283, EPI_ISL_896284, EPI_ISL_896285, EPI_ISL_896286, EPI_ISL_896287, EPI_ISL_896288, EPI_ISL_896289, EPI_ISL_896290, EPI_ISL_896291, EPI_ISL_896292, EPI_ISL_896293 | | | |
| see above | SUNY UPSTATE MEDICAL UNIVERSITY | Wadsworth Center, New York State Department of Health | Kirsten St. George, Daryl M. Lamson, Alexis Russel, Matthew Shudt, Melissa A Leisner, Jonathan Plitnick, Navjot Singh, John Kelly, Erasmus Schneider, Erica Lasek-Nesselquist |
| EPI_ISL_896294 | New York Presbyterian Hospital | Wadsworth Center, New York State Department of Health | Kirsten St. George, Daryl M. Lamson, Alexis Russel, Matthew Shudt, Melissa A Leisner, Jonathan Plitnick, Navjot Singh, John Kelly, Erasmus Schneider, Erica Lasek-Nesselquist |
| EPI_ISL_896295 | MEMORIAL SLOAN KETTERING CANCER CENTER | Wadsworth Center, New York State Department of Health | Kirsten St. George, Daryl M. Lamson, Alexis Russel, Matthew Shudt, Melissa A Leisner, Jonathan Plitnick, Navjot Singh, John Kelly, Erasmus Schneider, Erica Lasek-Nesselquist |
| EPI_ISL_896296 | New York Presbyterian Hospital | Wadsworth Center, New York State Department of Health | Kirsten St. George, Daryl M. Lamson, Alexis Russel, Matthew Shudt, Melissa A Leisner, Jonathan Plitnick, Navjot Singh, John Kelly, Erasmus Schneider, Erica Lasek-Nesselquist |
| EPI_ISL_896297 | BIO-REFERENCE LABORATORIES | Wadsworth Center, New York State Department of Health | Kirsten St. George, Daryl M. Lamson, Alexis Russel, Matthew Shudt, Melissa A Leisner, Jonathan Plitnick, Navjot Singh, John Kelly, Erasmus Schneider, Erica Lasek-Nesselquist |
| EPI_ISL_896298 | New York Presbyterian Hospital | Wadsworth Center, New York State Department of Health | Kirsten St. George, Daryl M. Lamson, Alexis Russel, Matthew Shudt, Melissa A Leisner, Jonathan Plitnick, Navjot Singh, John Kelly, Erasmus Schneider, Erica Lasek-Nesselquist |
| EPI_ISL_896299 | MEMORIAL SLOAN KETTERING CANCER CENTER | Wadsworth Center, New York State Department of Health | Kirsten St. George, Daryl M. Lamson, Alexis Russel, Matthew Shudt, Melissa A Leisner, Jonathan Plitnick, Navjot Singh, John Kelly, Erasmus Schneider, Erica Lasek-Nesselquist |
| EPI_ISL_896300 | BIO-REFERENCE LABORATORIES | Wadsworth Center, New York State Department of Health | Kirsten St. George, Daryl M. Lamson, Alexis Russel, Matthew Shudt, Melissa A Leisner, Jonathan Plitnick, Navjot Singh, John Kelly, Erasmus Schneider, Erica Lasek-Nesselquist |
| EPI_ISL_896301 | STONY BROOK UNIVERSITY HOSPITAL | Wadsworth Center, New York State Department of Health | Kirsten St. George, Daryl M. Lamson, Alexis Russel, Matthew Shudt, Melissa A Leisner, Jonathan Plitnick, Navjot Singh, John Kelly, Erasmus Schneider, Erica Lasek-Nesselquist |

| | | | |
|---|---|---|---|
| EPI_ISL_896302, EPI_ISL_896303, EPI_ISL_896304, EPI_ISL_896305, EPI_ISL_896306 | New York Presbyterian Hospital | Wadsworth Center, New York State Department of Health | Kirsten St. George, Daryl M. Lamson, Alexis Russel, Matthew Shudt, Melissa A Leisner, Jonathan Plitnick, Navjot Singh, John Kelly, Erasmus Schneider, Erica Lasek-Nesselquist |
| EPI_ISL_896307, EPI_ISL_896308, EPI_ISL_896309 | Wadsworth Center, New York State Department of Health | Wadsworth Center, New York State Department of Health | Kirsten St. George, Daryl M. Lamson, Alexis Russel, Matthew Shudt, Melissa A Leisner, Jonathan Plitnick, Navjot Singh, John Kelly, Erasmus Schneider, Erica Lasek-Nesselquist |
| EPI_ISL_896310, EPI_ISL_896311, EPI_ISL_896312, EPI_ISL_896313, EPI_ISL_896314, EPI_ISL_896315, EPI_ISL_896316, EPI_ISL_896317, EPI_ISL_896318, EPI_ISL_896319, EPI_ISL_896320, EPI_ISL_896321, EPI_ISL_896322, EPI_ISL_896323, EPI_ISL_896324, EPI_ISL_896325, EPI_ISL_896326, EPI_ISL_896327, EPI_ISL_896328, EPI_ISL_896329, EPI_ISL_896330, EPI_ISL_896331, EPI_ISL_896332, EPI_ISL_896333, EPI_ISL_896334, EPI_ISL_896335, EPI_ISL_896336, EPI_ISL_896337 | | | |
| see above | MEMORIAL SLOAN KETTERING CANCER CENTER | Wadsworth Center, New York State Department of Health | Kirsten St. George, Daryl M. Lamson, Alexis Russel, Matthew Shudt, Melissa A Leisner, Jonathan Plitnick, Navjot Singh, John Kelly, Erasmus Schneider, Erica Lasek-Nesselquist |
| EPI_ISL_896338, EPI_ISL_896339, EPI_ISL_896340, EPI_ISL_896341, EPI_ISL_896342 | New York Presbyterian Hospital | Wadsworth Center, New York State Department of Health | Kirsten St. George, Daryl M. Lamson, Alexis Russel, Matthew Shudt, Melissa A Leisner, Jonathan Plitnick, Navjot Singh, John Kelly, Erasmus Schneider, Erica Lasek-Nesselquist |
| EPI_ISL_896343, EPI_ISL_896344, EPI_ISL_896345, EPI_ISL_896346 | MEMORIAL SLOAN KETTERING CANCER CENTER | Wadsworth Center, New York State Department of Health | Kirsten St. George, Daryl M. Lamson, Alexis Russel, Matthew Shudt, Melissa A Leisner, Jonathan Plitnick, Navjot Singh, John Kelly, Erasmus Schneider, Erica Lasek-Nesselquist |
| EPI_ISL_896347, EPI_ISL_896348 | New York Presbyterian Hospital | Wadsworth Center, New York State Department of Health | Kirsten St. George, Daryl M. Lamson, Alexis Russel, Matthew Shudt, Melissa A Leisner, Jonathan Plitnick, Navjot Singh, John Kelly, Erasmus Schneider, Erica Lasek-Nesselquist |
| EPI_ISL_896349 | MEMORIAL SLOAN KETTERING CANCER CENTER | Wadsworth Center, New York State Department of Health | Kirsten St. George, Daryl M. Lamson, Alexis Russel, Matthew Shudt, Melissa A Leisner, Jonathan Plitnick, Navjot Singh, John Kelly, Erasmus Schneider, Erica Lasek-Nesselquist |
| EPI_ISL_896350, EPI_ISL_896351, EPI_ISL_896352, EPI_ISL_896353, EPI_ISL_896354, EPI_ISL_896355, EPI_ISL_896356, EPI_ISL_896357, EPI_ISL_896358, EPI_ISL_896359, EPI_ISL_896360, EPI_ISL_896361, EPI_ISL_896362, EPI_ISL_896363, EPI_ISL_896364, EPI_ISL_896365, EPI_ISL_896366, EPI_ISL_896367, EPI_ISL_896368, EPI_ISL_896369, EPI_ISL_896370, EPI_ISL_896371, EPI_ISL_896372 | | | |
| see above | New York Presbyterian Hospital | Wadsworth Center, New York State Department of Health | Kirsten St. George, Daryl M. Lamson, Alexis Russel, Matthew Shudt, Melissa A Leisner, Jonathan Plitnick, Navjot Singh, John Kelly, Erasmus Schneider, Erica Lasek-Nesselquist |
| EPI_ISL_896382, EPI_ISL_896383, EPI_ISL_896384 | Columbia University Irving Medical Center | Wadsworth Center, New York State Department of Health | Kirsten St. George, Daryl M. Lamson, Alexis Russel, Matthew Shudt, Melissa A Leisner, Jonathan Plitnick, Navjot Singh, John Kelly, Erasmus Schneider, Erica Lasek-Nesselquist |
| EPI_ISL_896385 | URMC LABS | Wadsworth Center, New York State Department of Health | Kirsten St. George, Daryl M. Lamson, Alexis Russel, Matthew Shudt, Melissa A Leisner, Jonathan Plitnick, Navjot Singh, John Kelly, Erasmus Schneider, Erica Lasek-Nesselquist |
| EPI_ISL_896386 | Columbia University Irving Medical Center | Wadsworth Center, New York State Department of Health | Kirsten St. George, Daryl M. Lamson, Alexis Russel, Matthew Shudt, Melissa A Leisner, Jonathan Plitnick, Navjot Singh, John Kelly, Erasmus Schneider, Erica Lasek-Nesselquist |
| EPI_ISL_896387, EPI_ISL_896388 | URMC LABS | Wadsworth Center, New York State Department of Health | Kirsten St. George, Daryl M. Lamson, Alexis Russel, Matthew Shudt, Melissa A Leisner, Jonathan Plitnick, Navjot Singh, John Kelly, Erasmus Schneider, Erica Lasek-Nesselquist |
| EPI_ISL_896389 | Columbia University Irving Medical Center | Wadsworth Center, New York State Department of Health | Kirsten St. George, Daryl M. Lamson, Alexis Russel, Matthew Shudt, Melissa A Leisner, Jonathan Plitnick, Navjot Singh, John Kelly, Erasmus Schneider, Erica Lasek-Nesselquist |
| EPI_ISL_896390 | URMC LABS | Wadsworth Center, New York State Department of Health | Kirsten St. George, Daryl M. Lamson, Alexis Russel, Matthew Shudt, Melissa A Leisner, Jonathan Plitnick, Navjot Singh, John Kelly, Erasmus Schneider, Erica Lasek-Nesselquist |
| EPI_ISL_896391, EPI_ISL_896392, EPI_ISL_896393, EPI_ISL_896394, EPI_ISL_896395 | Columbia University Irving Medical Center | Wadsworth Center, New York State Department of Health | Kirsten St. George, Daryl M. Lamson, Alexis Russel, Matthew Shudt, Melissa A Leisner, Jonathan Plitnick, Navjot Singh, John Kelly, Erasmus Schneider, Erica Lasek-Nesselquist |
| EPI_ISL_896396, EPI_ISL_896397, EPI_ISL_896398, EPI_ISL_896399, EPI_ISL_896400, EPI_ISL_896401, EPI_ISL_896402, EPI_ISL_896403, EPI_ISL_896404, EPI_ISL_896405, EPI_ISL_896406, EPI_ISL_896407, EPI_ISL_896408, EPI_ISL_896409, EPI_ISL_896410, EPI_ISL_896411, EPI_ISL_896412, EPI_ISL_896413, EPI_ISL_896414, EPI_ISL_896415, EPI_ISL_896416, EPI_ISL_896417, EPI_ISL_896418, EPI_ISL_896419, EPI_ISL_896420, EPI_ISL_896421 | | | |
| see above | URMC LABS | Wadsworth Center, New York State Department of Health | Kirsten St. George, Daryl M. Lamson, Alexis Russel, Matthew Shudt, Melissa A Leisner, Jonathan Plitnick, Navjot Singh, John Kelly, Erasmus Schneider, Erica Lasek-Nesselquist |
| EPI_ISL_896422, EPI_ISL_896423, EPI_ISL_896424, EPI_ISL_896425, EPI_ISL_896426, EPI_ISL_896427, EPI_ISL_896428, EPI_ISL_896429, EPI_ISL_896430, EPI_ISL_896431, EPI_ISL_896432, EPI_ISL_896433, EPI_ISL_896434, EPI_ISL_896435, EPI_ISL_896436, EPI_ISL_896437, EPI_ISL_896438, EPI_ISL_896439, EPI_ISL_896440, EPI_ISL_896441, EPI_ISL_896442, EPI_ISL_896443, EPI_ISL_896444, EPI_ISL_896445, EPI_ISL_896446 | | | |
| see above | Columbia University Irving Medical Center | Wadsworth Center, New York State Department of Health | Kirsten St. George, Daryl M. Lamson, Alexis Russel, Matthew Shudt, Melissa A Leisner, Jonathan Plitnick, Navjot Singh, John Kelly, Erasmus Schneider, Erica Lasek-Nesselquist |
| EPI_ISL_896495, EPI_ISL_896496, EPI_ISL_896497 | URMC LABS | Wadsworth Center, New York State Department of Health | Kirsten St. George, Daryl M. Lamson, Alexis Russel, Matthew Shudt, Melissa A Leisner, Jonathan Plitnick, Navjot Singh, John Kelly, Erasmus Schneider, Erica Lasek-Nesselquist |
| EPI_ISL_896498, EPI_ISL_896499, EPI_ISL_896500, EPI_ISL_896501 | MEMORIAL SLOAN KETTERING CANCER CENTER | Wadsworth Center, New York State Department of Health | Kirsten St. George, Daryl M. Lamson, Alexis Russel, Matthew Shudt, Melissa A Leisner, Jonathan Plitnick, Navjot Singh, John Kelly, Erasmus Schneider, Erica Lasek-Nesselquist |
| EPI_ISL_896502, EPI_ISL_896503 | URMC LABS | Wadsworth Center, New York State Department of Health | Kirsten St. George, Daryl M. Lamson, Alexis Russel, Matthew Shudt, Melissa A Leisner, Jonathan Plitnick, Navjot Singh, John Kelly, Erasmus Schneider, Erica Lasek-Nesselquist |
| EPI_ISL_896504, EPI_ISL_896505, EPI_ISL_896506, EPI_ISL_896507 | New York Presbyterian Hospital | Wadsworth Center, New York State Department of Health | Kirsten St. George, Daryl M. Lamson, Alexis Russel, Matthew Shudt, Melissa A Leisner, Jonathan Plitnick, Navjot Singh, John Kelly, Erasmus Schneider, Erica Lasek-Nesselquist |
| EPI_ISL_896508, EPI_ISL_896509, EPI_ISL_896510, EPI_ISL_896511, EPI_ISL_896512, EPI_ISL_896513, EPI_ISL_896514, EPI_ISL_896515, EPI_ISL_896516, EPI_ISL_896517, EPI_ISL_896518, EPI_ISL_896519 | | | |
| see above | URMC LABS | Wadsworth Center, New York State Department of Health | Kirsten St. George, Daryl M. Lamson, Alexis Russel, Matthew Shudt, Melissa A Leisner, Jonathan Plitnick, Navjot Singh, John Kelly, Erasmus Schneider, Erica Lasek-Nesselquist |
| EPI_ISL_896520, EPI_ISL_896521, EPI_ISL_896522, EPI_ISL_896523, EPI_ISL_896524 | MEMORIAL SLOAN KETTERING CANCER CENTER | Wadsworth Center, New York State Department of Health | Kirsten St. George, Daryl M. Lamson, Alexis Russel, Matthew Shudt, Melissa A Leisner, Jonathan Plitnick, Navjot Singh, John Kelly, Erasmus Schneider, Erica Lasek-Nesselquist |
| EPI_ISL_896525, EPI_ISL_896526, EPI_ISL_896527, EPI_ISL_896528, EPI_ISL_896529 | New York Presbyterian Hospital | Wadsworth Center, New York State Department of Health | Kirsten St. George, Daryl M. Lamson, Alexis Russel, Matthew Shudt, Melissa A Leisner, Jonathan Plitnick, Navjot Singh, John Kelly, Erasmus Schneider, Erica Lasek-Nesselquist |
| EPI_ISL_896530 | MEMORIAL SLOAN KETTERING CANCER CENTER | Wadsworth Center, New York State Department of Health | Kirsten St. George, Daryl M. Lamson, Alexis Russel, Matthew Shudt, Melissa A Leisner, Jonathan Plitnick, Navjot Singh, John Kelly, Erasmus Schneider, Erica Lasek-Nesselquist |
| EPI_ISL_896531, EPI_ISL_896532 | New York Presbyterian Hospital | Wadsworth Center, New York State Department of Health | Kirsten St. George, Daryl M. Lamson, Alexis Russel, Matthew Shudt, Melissa A Leisner, Jonathan Plitnick, Navjot Singh, John Kelly, Erasmus Schneider, Erica Lasek-Nesselquist |
| EPI_ISL_896533 | MEMORIAL SLOAN KETTERING CANCER CENTER | Wadsworth Center, New York State Department of Health | Kirsten St. George, Daryl M. Lamson, Alexis Russel, Matthew Shudt, Melissa A Leisner, Jonathan Plitnick, Navjot Singh, John Kelly, Erasmus Schneider, Erica Lasek-Nesselquist |
| EPI_ISL_896534 | New York Presbyterian Hospital | Wadsworth Center, New York State Department of Health | Kirsten St. George, Daryl M. Lamson, Alexis Russel, Matthew Shudt, Melissa A Leisner, Jonathan Plitnick, Navjot Singh, John Kelly, Erasmus Schneider, Erica Lasek-Nesselquist |
| EPI_ISL_896535, EPI_ISL_896536 | MEMORIAL SLOAN KETTERING CANCER CENTER | Wadsworth Center, New York State Department of Health | Kirsten St. George, Daryl M. Lamson, Alexis Russel, Matthew Shudt, Melissa A Leisner, Jonathan Plitnick, Navjot Singh, John Kelly, Erasmus Schneider, |

| | | | |
|---|---|---|---|
| | | | Erica Lasek-Nesselquist |
| EPI_ISL_896537, EPI_ISL_896538, EPI_ISL_896539, EPI_ISL_896540 | New York Presbyterian Hospital | Wadsworth Center, New York State Department of Health | Kirsten St. George, Daryl M. Lamson, Alexis Russel, Matthew Shudt, Melissa A Leisner, Jonathan Plitnick, Navjot Singh, John Kelly, Erasmus Schneider, Erica Lasek-Nesselquist |
| EPI_ISL_896541, EPI_ISL_896542, EPI_ISL_896543, EPI_ISL_896544, EPI_ISL_896545, EPI_ISL_896546, EPI_ISL_896547, EPI_ISL_896548 | MEMORIAL SLOAN KETTERING CANCER CENTER | Wadsworth Center, New York State Department of Health | Kirsten St. George, Daryl M. Lamson, Alexis Russel, Matthew Shudt, Melissa A Leisner, Jonathan Plitnick, Navjot Singh, John Kelly, Erasmus Schneider, Erica Lasek-Nesselquist |
| EPI_ISL_896549, EPI_ISL_896550, EPI_ISL_896551 | New York Presbyterian Hospital | Wadsworth Center, New York State Department of Health | Kirsten St. George, Daryl M. Lamson, Alexis Russel, Matthew Shudt, Melissa A Leisner, Jonathan Plitnick, Navjot Singh, John Kelly, Erasmus Schneider, Erica Lasek-Nesselquist |
| EPI_ISL_896552 | MEMORIAL SLOAN KETTERING CANCER CENTER | Wadsworth Center, New York State Department of Health | Kirsten St. George, Daryl M. Lamson, Alexis Russel, Matthew Shudt, Melissa A Leisner, Jonathan Plitnick, Navjot Singh, John Kelly, Erasmus Schneider, Erica Lasek-Nesselquist |
| EPI_ISL_896553, EPI_ISL_896554 | New York Presbyterian Hospital | Wadsworth Center, New York State Department of Health | Kirsten St. George, Daryl M. Lamson, Alexis Russel, Matthew Shudt, Melissa A Leisner, Jonathan Plitnick, Navjot Singh, John Kelly, Erasmus Schneider, Erica Lasek-Nesselquist |
| EPI_ISL_896555 | MEMORIAL SLOAN KETTERING CANCER CENTER | Wadsworth Center, New York State Department of Health | Kirsten St. George, Daryl M. Lamson, Alexis Russel, Matthew Shudt, Melissa A Leisner, Jonathan Plitnick, Navjot Singh, John Kelly, Erasmus Schneider, Erica Lasek-Nesselquist |
| EPI_ISL_896556, EPI_ISL_896557, EPI_ISL_896558, EPI_ISL_896559, EPI_ISL_896560, EPI_ISL_896561 | New York Presbyterian Hospital | Wadsworth Center, New York State Department of Health | Kirsten St. George, Daryl M. Lamson, Alexis Russel, Matthew Shudt, Melissa A Leisner, Jonathan Plitnick, Navjot Singh, John Kelly, Erasmus Schneider, Erica Lasek-Nesselquist |
| EPI_ISL_896562 | MEMORIAL SLOAN KETTERING CANCER CENTER | Wadsworth Center, New York State Department of Health | Kirsten St. George, Daryl M. Lamson, Alexis Russel, Matthew Shudt, Melissa A Leisner, Jonathan Plitnick, Navjot Singh, John Kelly, Erasmus Schneider, Erica Lasek-Nesselquist |
| EPI_ISL_896563, EPI_ISL_896564, EPI_ISL_896565, EPI_ISL_896566, EPI_ISL_896567, EPI_ISL_896568, EPI_ISL_896569 | URMC LABS | Wadsworth Center, New York State Department of Health | Kirsten St. George, Daryl M. Lamson, Alexis Russel, Matthew Shudt, Melissa A Leisner, Jonathan Plitnick, Navjot Singh, John Kelly, Erasmus Schneider, Erica Lasek-Nesselquist |
| EPI_ISL_896570, EPI_ISL_896571 | MEMORIAL SLOAN KETTERING CANCER CENTER | Wadsworth Center, New York State Department of Health | Kirsten St. George, Daryl M. Lamson, Alexis Russel, Matthew Shudt, Melissa A Leisner, Jonathan Plitnick, Navjot Singh, John Kelly, Erasmus Schneider, Erica Lasek-Nesselquist |
| EPI_ISL_912169 | East Gene | Grubaugh Lab - Yale School of Public Health | Tara Alpert, Joseph Fauver, Anderson Brito, Mallery Breban, Anne Wyllie, Chantal Vogels, Mary Petrone, Annie Watkins, Chaney Kalinich, Isabel Ott, Nathan Grubaugh |
| EPI_ISL_915340, EPI_ISL_915341, EPI_ISL_915342, EPI_ISL_915343, EPI_ISL_915344, EPI_ISL_915345 | Quest Diagnostics | Quest Diagnostics | Rosenthal,S.H., Gerasimova,A., Kagan,R.M., Anderson, B., Hua, M., Liu Y., Bernstein, L.E., Livingston, K.E., Perez, A., Shalhout, D.F., Shlyakhter, I.A., Owen, R., Tanpaiboon, P., Lacbawan, F. |
| EPI_ISL_935973, EPI_ISL_935974, EPI_ISL_935975, EPI_ISL_935976, EPI_ISL_935977, EPI_ISL_935978, EPI_ISL_935979, EPI_ISL_935980, EPI_ISL_935981, EPI_ISL_935982, EPI_ISL_935983, EPI_ISL_935984, EPI_ISL_935985, EPI_ISL_935986, EPI_ISL_935987, EPI_ISL_935988, EPI_ISL_935989, EPI_ISL_935990, EPI_ISL_935991, EPI_ISL_935992 | | | |
| see above | GLENS FALLS HOSPITAL LABORATORY | Wadsworth Center, New York State Department of Health | Kirsten St. George, Daryl M. Lamson, Alexis Russel, Matthew Shudt, Melissa A Leisner, Jonathan Plitnick, Navjot Singh, John Kelly, Erasmus Schneider, Erica Lasek-Nesselquist |
| EPI_ISL_935993, EPI_ISL_935994, EPI_ISL_935995, EPI_ISL_935996, EPI_ISL_935997, EPI_ISL_935998, EPI_ISL_935999, EPI_ISL_936000, EPI_ISL_936001, EPI_ISL_936002, EPI_ISL_936003, EPI_ISL_936004, EPI_ISL_936005, EPI_ISL_936006, EPI_ISL_936007, EPI_ISL_936008, EPI_ISL_936009, EPI_ISL_936010, EPI_ISL_936011, EPI_ISL_936012, EPI_ISL_936013, EPI_ISL_936014, EPI_ISL_936015, EPI_ISL_936016, EPI_ISL_936017, EPI_ISL_936018, EPI_ISL_936019, EPI_ISL_936020, EPI_ISL_936021, EPI_ISL_936022 | | | |
| see above | SUNY UPSTATE MEDICAL UNIVERSITY | Wadsworth Center, New York State Department of Health | Kirsten St. George, Daryl M. Lamson, Alexis Russel, Matthew Shudt, Melissa A Leisner, Jonathan Plitnick, Navjot Singh, John Kelly, Erasmus Schneider, Erica Lasek-Nesselquist |
| EPI_ISL_936023, EPI_ISL_936024, EPI_ISL_936025, EPI_ISL_936026, EPI_ISL_936027, EPI_ISL_936028, EPI_ISL_936029, EPI_ISL_936030, EPI_ISL_936031, EPI_ISL_936032, EPI_ISL_936033, EPI_ISL_936034, EPI_ISL_936035 | | | |
| see above | THE MARY IMOGENE BASSETT HOSPITAL | Wadsworth Center, New York State Department of Health | Kirsten St. George, Daryl M. Lamson, Alexis Russel, Matthew Shudt, Melissa A Leisner, Jonathan Plitnick, Navjot Singh, John Kelly, Erasmus Schneider, Erica Lasek-Nesselquist |
| EPI_ISL_936036, EPI_ISL_936037, EPI_ISL_936038, EPI_ISL_936039, EPI_ISL_936040, EPI_ISL_936041, EPI_ISL_936042, EPI_ISL_936043, EPI_ISL_936044, EPI_ISL_936045 | Columbia University Irving Medical Center | Wadsworth Center, New York State Department of Health | Kirsten St. George, Daryl M. Lamson, Alexis Russel, Matthew Shudt, Melissa A Leisner, Jonathan Plitnick, Navjot Singh, John Kelly, Erasmus Schneider, Erica Lasek-Nesselquist |
| EPI_ISL_936046, EPI_ISL_936047, EPI_ISL_936048, EPI_ISL_936049, EPI_ISL_936050, EPI_ISL_936051, EPI_ISL_936052, EPI_ISL_936053, EPI_ISL_936054, EPI_ISL_936055, EPI_ISL_936056, EPI_ISL_936057, EPI_ISL_936058, EPI_ISL_936059 | | | |
| see above | ADIRONDACK MEDICAL CENTER | Wadsworth Center, New York State Department of Health | Kirsten St. George, Daryl M. Lamson, Alexis Russel, Matthew Shudt, Melissa A Leisner, Jonathan Plitnick, Navjot Singh, John Kelly, Erasmus Schneider, Erica Lasek-Nesselquist |
| EPI_ISL_936060, EPI_ISL_936061, EPI_ISL_936062 | KALEIDA CENTER FOR LABORATORY MEDICINE | Wadsworth Center, New York State Department of Health | Kirsten St. George, Daryl M. Lamson, Alexis Russel, Matthew Shudt, Melissa A Leisner, Jonathan Plitnick, Navjot Singh, John Kelly, Erasmus Schneider, Erica Lasek-Nesselquist |
| EPI_ISL_936063, EPI_ISL_936064, EPI_ISL_936065, EPI_ISL_936066 | Columbia University Irving Medical Center | Wadsworth Center, New York State Department of Health | Kirsten St. George, Daryl M. Lamson, Alexis Russel, Matthew Shudt, Melissa A Leisner, Jonathan Plitnick, Navjot Singh, John Kelly, Erasmus Schneider, Erica Lasek-Nesselquist |
| EPI_ISL_936067, EPI_ISL_936068 | KALEIDA CENTER FOR LABORATORY MEDICINE | Wadsworth Center, New York State Department of Health | Kirsten St. George, Daryl M. Lamson, Alexis Russel, Matthew Shudt, Melissa A Leisner, Jonathan Plitnick, Navjot Singh, John Kelly, Erasmus Schneider, Erica Lasek-Nesselquist |
| EPI_ISL_936069 | Columbia University Irving Medical Center | Wadsworth Center, New York State Department of Health | Kirsten St. George, Daryl M. Lamson, Alexis Russel, Matthew Shudt, Melissa A Leisner, Jonathan Plitnick, Navjot Singh, John Kelly, Erasmus Schneider, Erica Lasek-Nesselquist |
| EPI_ISL_936070, EPI_ISL_936071, EPI_ISL_936072, EPI_ISL_936073, EPI_ISL_936074, EPI_ISL_936075, EPI_ISL_936076, EPI_ISL_936077, EPI_ISL_936078, EPI_ISL_936079, EPI_ISL_936080, EPI_ISL_936081 | | | |
| see above | KALEIDA CENTER FOR LABORATORY MEDICINE | Wadsworth Center, New York State Department of Health | Kirsten St. George, Daryl M. Lamson, Alexis Russel, Matthew Shudt, Melissa A Leisner, Jonathan Plitnick, Navjot Singh, John Kelly, Erasmus Schneider, Erica Lasek-Nesselquist |
| EPI_ISL_936082 | Columbia University Irving Medical Center | Wadsworth Center, New York State Department of Health | Kirsten St. George, Daryl M. Lamson, Alexis Russel, Matthew Shudt, Melissa A Leisner, Jonathan Plitnick, Navjot Singh, John Kelly, Erasmus Schneider, Erica Lasek-Nesselquist |
| EPI_ISL_936083, EPI_ISL_936084, EPI_ISL_936085 | KALEIDA CENTER FOR LABORATORY MEDICINE | Wadsworth Center, New York State Department of Health | Kirsten St. George, Daryl M. Lamson, Alexis Russel, Matthew Shudt, Melissa A Leisner, Jonathan Plitnick, Navjot Singh, John Kelly, Erasmus Schneider, Erica Lasek-Nesselquist |
| EPI_ISL_936086 | Columbia University Irving Medical Center | Wadsworth Center, New York State Department of Health | Kirsten St. George, Daryl M. Lamson, Alexis Russel, Matthew Shudt, Melissa A Leisner, Jonathan Plitnick, Navjot Singh, John Kelly, Erasmus Schneider, Erica Lasek-Nesselquist |
| EPI_ISL_936087, EPI_ISL_936088 | KALEIDA CENTER FOR LABORATORY MEDICINE | Wadsworth Center, New York State Department of Health | Kirsten St. George, Daryl M. Lamson, Alexis Russel, Matthew Shudt, Melissa A Leisner, Jonathan Plitnick, Navjot Singh, John Kelly, Erasmus Schneider, Erica Lasek-Nesselquist |
| EPI_ISL_936089 | Columbia University Irving Medical Center | Wadsworth Center, New York State Department of Health | Kirsten St. George, Daryl M. Lamson, Alexis Russel, Matthew Shudt, Melissa A Leisner, Jonathan Plitnick, Navjot Singh, John Kelly, Erasmus Schneider, Erica Lasek-Nesselquist |

| | | | |
|---|---|---|---|
| EPI_ISL_936090, EPI_ISL_936091 | KALEIDA CENTER FOR LABORATORY MEDICINE | Wadsworth Center, New York State Department of Health | Kirsten St. George, Daryl M. Lamson, Alexis Russel, Matthew Shudt, Melissa A Leisner, Jonathan Plitnick, Navjot Singh, John Kelly, Erasmus Schneider, Erica Lasek-Nesselquist |
| EPI_ISL_936092, EPI_ISL_936093 | Columbia University Irving Medical Center | Wadsworth Center, New York State Department of Health | Kirsten St. George, Daryl M. Lamson, Alexis Russel, Matthew Shudt, Melissa A Leisner, Jonathan Plitnick, Navjot Singh, John Kelly, Erasmus Schneider, Erica Lasek-Nesselquist |
| EPI_ISL_936094, EPI_ISL_936095, EPI_ISL_936096, EPI_ISL_936097, EPI_ISL_936098, EPI_ISL_936099, EPI_ISL_936100, EPI_ISL_936101, EPI_ISL_936102 | KALEIDA CENTER FOR LABORATORY MEDICINE | Wadsworth Center, New York State Department of Health | Kirsten St. George, Daryl M. Lamson, Alexis Russel, Matthew Shudt, Melissa A Leisner, Jonathan Plitnick, Navjot Singh, John Kelly, Erasmus Schneider, Erica Lasek-Nesselquist |
| EPI_ISL_936103, EPI_ISL_936104, EPI_ISL_936105, EPI_ISL_936106, EPI_ISL_936107, EPI_ISL_936108, EPI_ISL_936109, EPI_ISL_936110, EPI_ISL_936111, EPI_ISL_936112, EPI_ISL_936113, EPI_ISL_936114, EPI_ISL_936115, EPI_ISL_936116, EPI_ISL_936117, EPI_ISL_936118, EPI_ISL_936119, EPI_ISL_936120, EPI_ISL_936121, EPI_ISL_936122, EPI_ISL_936123, EPI_ISL_936124, EPI_ISL_936125, EPI_ISL_936126, EPI_ISL_936127, EPI_ISL_936128, EPI_ISL_936129 | | | |
| see above | WESTCHESTER MEDICAL CENTER | Wadsworth Center, New York State Department of Health | Kirsten St. George, Daryl M. Lamson, Alexis Russel, Matthew Shudt, Melissa A Leisner, Jonathan Plitnick, Navjot Singh, John Kelly, Erasmus Schneider, Erica Lasek-Nesselquist |
| EPI_ISL_936130, EPI_ISL_936131, EPI_ISL_936132, EPI_ISL_936133, EPI_ISL_936134, EPI_ISL_936135, EPI_ISL_936136, EPI_ISL_936137 | New York Presbyterian Hospital | Wadsworth Center, New York State Department of Health | Kirsten St. George, Daryl M. Lamson, Alexis Russel, Matthew Shudt, Melissa A Leisner, Jonathan Plitnick, Navjot Singh, John Kelly, Erasmus Schneider, Erica Lasek-Nesselquist |
| EPI_ISL_936138, EPI_ISL_936139, EPI_ISL_936140, EPI_ISL_936141 | THE MARY IMOGENE BASSETT HOSPITAL | Wadsworth Center, New York State Department of Health | Kirsten St. George, Daryl M. Lamson, Alexis Russel, Matthew Shudt, Melissa A Leisner, Jonathan Plitnick, Navjot Singh, John Kelly, Erasmus Schneider, Erica Lasek-Nesselquist |
| EPI_ISL_936142, EPI_ISL_936143 | SUNY UPSTATE MEDICAL UNIVERSITY | Wadsworth Center, New York State Department of Health | Kirsten St. George, Daryl M. Lamson, Alexis Russel, Matthew Shudt, Melissa A Leisner, Jonathan Plitnick, Navjot Singh, John Kelly, Erasmus Schneider, Erica Lasek-Nesselquist |
| EPI_ISL_936144 | KALEIDA CENTER FOR LABORATORY MEDICINE | Wadsworth Center, New York State Department of Health | Kirsten St. George, Daryl M. Lamson, Alexis Russel, Matthew Shudt, Melissa A Leisner, Jonathan Plitnick, Navjot Singh, John Kelly, Erasmus Schneider, Erica Lasek-Nesselquist |
| EPI_ISL_936145, EPI_ISL_936146, EPI_ISL_936147 | New York Presbyterian Hospital | Wadsworth Center, New York State Department of Health | Kirsten St. George, Daryl M. Lamson, Alexis Russel, Matthew Shudt, Melissa A Leisner, Jonathan Plitnick, Navjot Singh, John Kelly, Erasmus Schneider, Erica Lasek-Nesselquist |
| EPI_ISL_936148 | Wadsworth Center, New York State Department of Health | Wadsworth Center, New York State Department of Health | Kirsten St. George, Daryl M. Lamson, Alexis Russel, Matthew Shudt, Melissa A Leisner, Jonathan Plitnick, Navjot Singh, John Kelly, Erasmus Schneider, Erica Lasek-Nesselquist |
| EPI_ISL_936149, EPI_ISL_936150, EPI_ISL_936151, EPI_ISL_936152 | MEMORIAL SLOAN KETTERING CANCER CENTER | Wadsworth Center, New York State Department of Health | Kirsten St. George, Daryl M. Lamson, Alexis Russel, Matthew Shudt, Melissa A Leisner, Jonathan Plitnick, Navjot Singh, John Kelly, Erasmus Schneider, Erica Lasek-Nesselquist |
| EPI_ISL_936153 | THE MARY IMOGENE BASSETT HOSPITAL | Wadsworth Center, New York State Department of Health | Kirsten St. George, Daryl M. Lamson, Alexis Russel, Matthew Shudt, Melissa A Leisner, Jonathan Plitnick, Navjot Singh, John Kelly, Erasmus Schneider, Erica Lasek-Nesselquist |
| EPI_ISL_936154, EPI_ISL_936155, EPI_ISL_936156, EPI_ISL_936157, EPI_ISL_936158, EPI_ISL_936159, EPI_ISL_936160, EPI_ISL_936161, EPI_ISL_936162, EPI_ISL_936163, EPI_ISL_936164, EPI_ISL_936165, EPI_ISL_936166, EPI_ISL_936167, EPI_ISL_936168, EPI_ISL_936169, EPI_ISL_936170, EPI_ISL_936171, EPI_ISL_936172 | | | |
| see above | MEMORIAL SLOAN KETTERING CANCER CENTER | Wadsworth Center, New York State Department of Health | Kirsten St. George, Daryl M. Lamson, Alexis Russel, Matthew Shudt, Melissa A Leisner, Jonathan Plitnick, Navjot Singh, John Kelly, Erasmus Schneider, Erica Lasek-Nesselquist |
| EPI_ISL_936173 | TEMPUS LABS INC | Wadsworth Center, New York State Department of Health | Kirsten St. George, Daryl M. Lamson, Alexis Russel, Matthew Shudt, Melissa A Leisner, Jonathan Plitnick, Navjot Singh, John Kelly, Erasmus Schneider, Erica Lasek-Nesselquist |
| EPI_ISL_936174, EPI_ISL_936175, EPI_ISL_936176, EPI_ISL_936177, EPI_ISL_936178, EPI_ISL_936179 | MEMORIAL SLOAN KETTERING CANCER CENTER | Wadsworth Center, New York State Department of Health | Kirsten St. George, Daryl M. Lamson, Alexis Russel, Matthew Shudt, Melissa A Leisner, Jonathan Plitnick, Navjot Singh, John Kelly, Erasmus Schneider, Erica Lasek-Nesselquist |
| EPI_ISL_936180, EPI_ISL_936181 | ALBANY MEDICAL CENTER | Wadsworth Center, New York State Department of Health | Kirsten St. George, Daryl M. Lamson, Alexis Russel, Matthew Shudt, Melissa A Leisner, Jonathan Plitnick, Navjot Singh, John Kelly, Erasmus Schneider, Erica Lasek-Nesselquist |
| EPI_ISL_936212, EPI_ISL_936213, EPI_ISL_936214, EPI_ISL_936215, EPI_ISL_936216, EPI_ISL_936217, EPI_ISL_936218, EPI_ISL_936219, EPI_ISL_936220, EPI_ISL_936221, EPI_ISL_936222, EPI_ISL_936223, EPI_ISL_936224, EPI_ISL_936225, EPI_ISL_936226, EPI_ISL_936227, EPI_ISL_936228, EPI_ISL_936229, EPI_ISL_936230, EPI_ISL_936231, EPI_ISL_936232, EPI_ISL_936233, EPI_ISL_936234, EPI_ISL_936235, EPI_ISL_936236, EPI_ISL_936237, EPI_ISL_936238, EPI_ISL_936239, EPI_ISL_936240, EPI_ISL_936241, EPI_ISL_936242, EPI_ISL_936243, EPI_ISL_936244, EPI_ISL_936245, EPI_ISL_936246, EPI_ISL_936247 | | | |
| see above | Wadsworth Center, New York State Department of Health | Wadsworth Center, New York State Department of Health | Kirsten St. George, Daryl M. Lamson, Alexis Russel, Matthew Shudt, Melissa A Leisner, Jonathan Plitnick, Navjot Singh, John Kelly, Erasmus Schneider, Erica Lasek-Nesselquist |
| EPI_ISL_936248, EPI_ISL_936249, EPI_ISL_936250, EPI_ISL_936251, EPI_ISL_936252, EPI_ISL_936253, EPI_ISL_936254, EPI_ISL_936255, EPI_ISL_936256, EPI_ISL_936257, EPI_ISL_936258, EPI_ISL_936259, EPI_ISL_936260, EPI_ISL_936261, EPI_ISL_936262, EPI_ISL_936263, EPI_ISL_936264, EPI_ISL_936265, EPI_ISL_936266, EPI_ISL_936267, EPI_ISL_936268, EPI_ISL_936269, EPI_ISL_936270, EPI_ISL_936271, EPI_ISL_936272, EPI_ISL_936273, EPI_ISL_936274, EPI_ISL_936275, EPI_ISL_936276, EPI_ISL_936277, EPI_ISL_936278, EPI_ISL_936279, EPI_ISL_936280, EPI_ISL_936281, EPI_ISL_936282, EPI_ISL_936283, EPI_ISL_936284, EPI_ISL_936285, EPI_ISL_936286, EPI_ISL_936287, EPI_ISL_936288, EPI_ISL_936289, EPI_ISL_936290, EPI_ISL_936291, EPI_ISL_936292, EPI_ISL_936293, EPI_ISL_936294, EPI_ISL_936295, EPI_ISL_936296, EPI_ISL_936297, EPI_ISL_936298, EPI_ISL_936299 | | | |
| see above | MONTEFIORE MEDICAL CENTER LABORATORIES | Wadsworth Center, New York State Department of Health | Kirsten St. George, Daryl M. Lamson, Alexis Russel, Matthew Shudt, Melissa A Leisner, Jonathan Plitnick, Navjot Singh, John Kelly, Erasmus Schneider, Erica Lasek-Nesselquist |
| EPI_ISL_936300, EPI_ISL_936301, EPI_ISL_936302 | Wadsworth Center, New York State Department of Health | Wadsworth Center, New York State Department of Health | Kirsten St. George, Daryl M. Lamson, Alexis Russel, Matthew Shudt, Melissa A Leisner, Jonathan Plitnick, Navjot Singh, John Kelly, Erasmus Schneider, Erica Lasek-Nesselquist |
| EPI_ISL_937126, EPI_ISL_937127, EPI_ISL_937128 | DOHMH Riverside | New York City Public Health Laboratory | Jade Wang, et al. |
| EPI_ISL_937129, EPI_ISL_937130, EPI_ISL_937131 | DOHMH Morrisania | New York City Public Health Laboratory | Jade Wang, et al. |
| EPI_ISL_937132, EPI_ISL_937133 | DOHMH Central Harlem | New York City Public Health Laboratory | Jade Wang, et al. |
| EPI_ISL_937134, EPI_ISL_937135 | DOHMH Fort Greene | New York City Public Health Laboratory | Jade Wang, et al. |
| EPI_ISL_937136, EPI_ISL_937137 | DOHMH Chelsea | New York City Public Health Laboratory | Jade Wang, et al. |
| EPI_ISL_937138 | DOHMH Crown Heights | New York City Public Health Laboratory | Jade Wang, et al. |
| EPI_ISL_937139, EPI_ISL_937140, EPI_ISL_937141, EPI_ISL_937142, EPI_ISL_937143, EPI_ISL_937144 | DOHMH Corona | New York City Public Health Laboratory | Jade Wang, et al. |
| EPI_ISL_937145, EPI_ISL_937146, EPI_ISL_937147, EPI_ISL_937148, EPI_ISL_937149, EPI_ISL_937150, EPI_ISL_937151 | DOHMH Jamaica | New York City Public Health Laboratory | Jade Wang, et al. |
| EPI_ISL_937152, EPI_ISL_937153, EPI_ISL_937154, EPI_ISL_937155, EPI_ISL_937156 | DOHMH PHL | New York City Public Health Laboratory | Jade Wang, et al. |
| EPI_ISL_937157, EPI_ISL_937158, | DOHMH Jamaica | New York City Public Health Laboratory | Jade Wang, et al. |

| | | | |
|---|---|---|---|
| EPI_ISL_937159, EPI_ISL_937160, EPI_ISL_937161, EPI_ISL_937162, EPI_ISL_937163 | | | |
| EPI_ISL_937164, EPI_ISL_937165 | DOHMH Corona | New York City Public Health Laboratory | Jade Wang, et al. |
| EPI_ISL_937166 | DOHMH Riverside | New York City Public Health Laboratory | Jade Wang, et al. |
| EPI_ISL_937167 | DOHMH Central Harlem | New York City Public Health Laboratory | Jade Wang, et al. |
| EPI_ISL_937168 | DOHMH Crown Heights | New York City Public Health Laboratory | Jade Wang, et al. |
| EPI_ISL_937169, EPI_ISL_937170, EPI_ISL_937171 | DOHMH PHL | New York City Public Health Laboratory | Jade Wang, et al. |
| EPI_ISL_937172 | DOHMH Central Harlem | New York City Public Health Laboratory | Jade Wang, et al. |
| EPI_ISL_937173 | DOHMH Crown Heights | New York City Public Health Laboratory | Jade Wang, et al. |
| EPI_ISL_937174 | DOHMH Chelsea | New York City Public Health Laboratory | Jade Wang, et al. |
| EPI_ISL_937175, EPI_ISL_937176, EPI_ISL_937177 | DOHMH Jamaica | New York City Public Health Laboratory | Jade Wang, et al. |
| EPI_ISL_937178, EPI_ISL_937179, EPI_ISL_937180, EPI_ISL_937181, EPI_ISL_937182, EPI_ISL_937183, EPI_ISL_937184 | DOHMH Corona | New York City Public Health Laboratory | Jade Wang, et al. |
| EPI_ISL_937185 | DOHMH PHL | New York City Public Health Laboratory | Jade Wang, et al. |
| EPI_ISL_937186, EPI_ISL_937187, EPI_ISL_937188, EPI_ISL_937189, EPI_ISL_937190 | OCME Office Of Chief Medical Examiner | New York City Public Health Laboratory | Jade Wang, et al. |
| EPI_ISL_937191, EPI_ISL_937192 | Department of Homeless Services | New York City Public Health Laboratory | Jade Wang, et al. |
| EPI_ISL_937193, EPI_ISL_937194, EPI_ISL_937195, EPI_ISL_937196, EPI_ISL_937197, EPI_ISL_937198, EPI_ISL_937199 | OCME Office Of Chief Medical Examiner | New York City Public Health Laboratory | Jade Wang, et al. |
| EPI_ISL_937200, EPI_ISL_937201 | DOHMH Fort Greene | New York City Public Health Laboratory | Jade Wang, et al. |
| EPI_ISL_937202, EPI_ISL_937203 | DOHMH Chelsea | New York City Public Health Laboratory | Jade Wang, et al. |
| EPI_ISL_937204, EPI_ISL_937205 | DOHMH Central Harlem | New York City Public Health Laboratory | Jade Wang, et al. |
| EPI_ISL_937206, EPI_ISL_937207 | DOHMH Morrisania | New York City Public Health Laboratory | Jade Wang, et al. |
| EPI_ISL_937208 | DOHMH Corona | New York City Public Health Laboratory | Jade Wang, et al. |
| EPI_ISL_937209, EPI_ISL_937210, EPI_ISL_937211, EPI_ISL_937212 | DOHMH PHL | New York City Public Health Laboratory | Jade Wang, et al. |
| EPI_ISL_937213 | DOHMH Jamaica | New York City Public Health Laboratory | Jade Wang, et al. |
| EPI_ISL_937214 | DOHMH Corona | New York City Public Health Laboratory | Jade Wang, et al. |
| EPI_ISL_937215 | DOHMH PHL | New York City Public Health Laboratory | Jade Wang, et al. |
| EPI_ISL_937216 | DOHMH Central Harlem | New York City Public Health Laboratory | Jade Wang, et al. |
| EPI_ISL_937217, EPI_ISL_937218 | DOHMH Chelsea | New York City Public Health Laboratory | Jade Wang, et al. |
| EPI_ISL_937219, EPI_ISL_937220, EPI_ISL_937221, EPI_ISL_937222, EPI_ISL_937223, EPI_ISL_937224, EPI_ISL_937225, EPI_ISL_937226 | DOHMH Jamaica | New York City Public Health Laboratory | Jade Wang, et al. |
| EPI_ISL_937227, EPI_ISL_937228, EPI_ISL_937229, EPI_ISL_937230, EPI_ISL_937231 | DOHMH Corona | New York City Public Health Laboratory | Jade Wang, et al. |
| EPI_ISL_937232, EPI_ISL_937233, EPI_ISL_937234, EPI_ISL_937235, EPI_ISL_937236 | DOHMH Morrisania | New York City Public Health Laboratory | Jade Wang, et al. |
| EPI_ISL_937237 | DOHMH Riverside | New York City Public Health Laboratory | Jade Wang, et al. |
| EPI_ISL_937238, EPI_ISL_937239 | DOHMH Morrisania | New York City Public Health Laboratory | Jade Wang, et al. |
| EPI_ISL_937240, EPI_ISL_937241, EPI_ISL_937242, EPI_ISL_937243, EPI_ISL_937244, EPI_ISL_937245, EPI_ISL_937246, EPI_ISL_937247 | OCME Office Of Chief Medical Examiner | New York City Public Health Laboratory | Jade Wang, et al. |
| EPI_ISL_937248, EPI_ISL_937249, EPI_ISL_937250 | Department of Homeless Services | New York City Public Health Laboratory | Jade Wang, et al. |
| EPI_ISL_937251, EPI_ISL_937252, EPI_ISL_937253, EPI_ISL_937254, EPI_ISL_937255 | OCME Office Of Chief Medical Examiner | New York City Public Health Laboratory | Jade Wang, et al. |
| EPI_ISL_937256, EPI_ISL_937257, EPI_ISL_937258, EPI_ISL_937259, EPI_ISL_937260, EPI_ISL_937261, EPI_ISL_937262, EPI_ISL_937263, EPI_ISL_937264, EPI_ISL_937265, EPI_ISL_937266, EPI_ISL_937267, EPI_ISL_937268, EPI_ISL_937269, EPI_ISL_937270, EPI_ISL_937271, EPI_ISL_937272 | | | |
| see above | NYC HH Elmhurst Hospital Medical Center | New York City Public Health Laboratory | Jade Wang, et al. |
| EPI_ISL_955265 | Sunrise Medical Laboratory | Pathogen Discovery, Respiratory Viruses Branch, Division of Viral Diseases, Centers for Disease Control and Prevention | Ying Tao, Jing Zhang, Yan Li, Krista Queen, Anna Uehara, Peter Cook, Clinton R. Paden, Haibin Wang, Suxiang Tong |
| EPI_ISL_955271 | American Esoteric Laboratory | Pathogen Discovery, Respiratory Viruses Branch, Division of Viral Diseases, Centers for Disease Control and Prevention | Ying Tao, Jing Zhang, Yan Li, Krista Queen, Anna Uehara, Peter Cook, Clinton R. Paden, Haibin Wang, Suxiang Tong |
| EPI_ISL_955300 | Sunrise Medical Laboratory | Pathogen Discovery, Respiratory Viruses Branch, Division of Viral Diseases, Centers for Disease Control and Prevention | Ying Tao, Jing Zhang, Yan Li, Krista Queen, Anna Uehara, Peter Cook, Clinton R. Paden, Haibin Wang, Suxiang Tong |

| | | | |
|---|---|---|---|
| EPI_ISL_961289, EPI_ISL_961290, EPI_ISL_961291, EPI_ISL_961292, EPI_ISL_961293, EPI_ISL_961294, EPI_ISL_961295, EPI_ISL_961296, EPI_ISL_961297, EPI_ISL_961298, EPI_ISL_961299, EPI_ISL_961300, EPI_ISL_961301, EPI_ISL_961302, EPI_ISL_961303, EPI_ISL_961304, EPI_ISL_961305, EPI_ISL_961306, EPI_ISL_961307, EPI_ISL_961308, EPI_ISL_961309, EPI_ISL_961310, EPI_ISL_961311, EPI_ISL_961312, EPI_ISL_961313, EPI_ISL_961314, EPI_ISL_961315, EPI_ISL_961328, EPI_ISL_961329, EPI_ISL_961330, EPI_ISL_961331, EPI_ISL_961332, EPI_ISL_961333, EPI_ISL_961334, EPI_ISL_961335, EPI_ISL_961336, EPI_ISL_961337, EPI_ISL_961338, EPI_ISL_961339, EPI_ISL_961340, EPI_ISL_961341, EPI_ISL_961342, EPI_ISL_961343, EPI_ISL_961344, EPI_ISL_961345, EPI_ISL_961346, EPI_ISL_961347, EPI_ISL_961348, EPI_ISL_961349, EPI_ISL_961350, EPI_ISL_961351 | | | |
| see above | Biotia | Biotia | Dorottya Nagy-Szakal, Mara Couto-Rodriguez, Heather Wells, Joseph Barrows, Marilyne Debieu, Kristin Butcher, Siyuan Chen, Agnes Berki, Courteny Hager, Robert Boorstein, Mariah Taylor, Colleen Jonsson, Christopher Mason, Niamh O'Hara |
| EPI_ISL_965303, EPI_ISL_965528, EPI_ISL_965770, EPI_ISL_965812, EPI_ISL_965814, EPI_ISL_965826, EPI_ISL_965837, EPI_ISL_965838, EPI_ISL_965839, EPI_ISL_965910, EPI_ISL_965948, EPI_ISL_965962, EPI_ISL_965963, EPI_ISL_965964, EPI_ISL_965965, EPI_ISL_965967, EPI_ISL_965968, EPI_ISL_965969, EPI_ISL_965970, EPI_ISL_965971, EPI_ISL_965972, EPI_ISL_965973, EPI_ISL_965977, EPI_ISL_965980, EPI_ISL_965989, EPI_ISL_966020, EPI_ISL_966044, EPI_ISL_966062, EPI_ISL_966084, EPI_ISL_966108, EPI_ISL_966160, EPI_ISL_966184, EPI_ISL_966204, EPI_ISL_966227, EPI_ISL_966245, EPI_ISL_966264, EPI_ISL_966289, EPI_ISL_966308, EPI_ISL_966311, EPI_ISL_966333, EPI_ISL_966357, EPI_ISL_966375, EPI_ISL_966376, EPI_ISL_966377, EPI_ISL_966378 | | | |
| see above | NYU Langone Health | Departments of Pathology and Medicine, New York University School of Medicine | Adriana Heguy, Dacia Dimartino, Emily Guzman, Christian Marier, Peter Meyn, Sitharam Ramaswami, Gael Westby, Paul Zappile, Yutong Zhang, Paolo Cotzia, Guiqing Wang |
| EPI_ISL_966379 | OCME Office Of Chief Medical Examiner | New York City Public Health Laboratory | Jade Wang, et al. |
| EPI_ISL_966380 | NYU Langone Health | Departments of Pathology and Medicine, New York University School of Medicine | Adriana Heguy, Dacia Dimartino, Emily Guzman, Christian Marier, Peter Meyn, Sitharam Ramaswami, Gael Westby, Paul Zappile, Yutong Zhang, Paolo Cotzia, Guiqing Wang |
| EPI_ISL_966381 | OCME Office Of Chief Medical Examiner | New York City Public Health Laboratory | Jade Wang, et al. |
| EPI_ISL_966382 | DOHMH Jamaica | New York City Public Health Laboratory | Jade Wang, et al. |
| EPI_ISL_966383 | OCME Office Of Chief Medical Examiner | New York City Public Health Laboratory | Jade Wang, et al. |
| EPI_ISL_966384 | DOHMH Jamaica | New York City Public Health Laboratory | Jade Wang, et al. |
| EPI_ISL_966385 | Department of Homeless Services | New York City Public Health Laboratory | Jade Wang, et al. |
| EPI_ISL_966386 | DOHMH Corona | New York City Public Health Laboratory | Jade Wang, et al. |
| EPI_ISL_966387, EPI_ISL_966388 | DOHMH Jamaica | New York City Public Health Laboratory | Jade Wang, et al. |
| EPI_ISL_966389 | DOHMH Central Harlem | New York City Public Health Laboratory | Jade Wang, et al. |
| EPI_ISL_966390, EPI_ISL_966391 | OCME Office Of Chief Medical Examiner | New York City Public Health Laboratory | Jade Wang, et al. |
| EPI_ISL_966392 | DOHMH Corona | New York City Public Health Laboratory | Jade Wang, et al. |
| EPI_ISL_966393 | OCME Office Of Chief Medical Examiner | New York City Public Health Laboratory | Jade Wang, et al. |
| EPI_ISL_966394 | DOHMH Corona | New York City Public Health Laboratory | Jade Wang, et al. |
| EPI_ISL_966395 | DOHMH Jamaica | New York City Public Health Laboratory | Jade Wang, et al. |
| EPI_ISL_966396, EPI_ISL_966397 | OCME Office Of Chief Medical Examiner | New York City Public Health Laboratory | Jade Wang, et al. |
| EPI_ISL_966398 | DOHMH Jamaica | New York City Public Health Laboratory | Jade Wang, et al. |
| EPI_ISL_966399 | DOHMH Corona | New York City Public Health Laboratory | Jade Wang, et al. |
| EPI_ISL_966400 | OCME Office Of Chief Medical Examiner | New York City Public Health Laboratory | Jade Wang, et al. |
| EPI_ISL_966401 | Department of Homeless Services | New York City Public Health Laboratory | Jade Wang, et al. |
| EPI_ISL_966402, EPI_ISL_966403 | OCME Office Of Chief Medical Examiner | New York City Public Health Laboratory | Jade Wang, et al. |
| EPI_ISL_966404 | DOHMH PHL | New York City Public Health Laboratory | Jade Wang, et al. |
| EPI_ISL_966405 | DOHMH Jamaica | New York City Public Health Laboratory | Jade Wang, et al. |
| EPI_ISL_966406 | NYU Langone Health | Departments of Pathology and Medicine, New York University School of Medicine | Adriana Heguy, Dacia Dimartino, Emily Guzman, Christian Marier, Peter Meyn, Sitharam Ramaswami, Gael Westby, Paul Zappile, Yutong Zhang, Paolo Cotzia, Guiqing Wang |
| EPI_ISL_966407, EPI_ISL_966408 | DOHMH Morrisania | New York City Public Health Laboratory | Jade Wang, et al. |
| EPI_ISL_966409, EPI_ISL_966410 | DOHMH Corona | New York City Public Health Laboratory | Jade Wang, et al. |
| EPI_ISL_966411 | OCME Office Of Chief Medical Examiner | New York City Public Health Laboratory | Jade Wang, et al. |
| EPI_ISL_966412 | DOHMH Jamaica | New York City Public Health Laboratory | Jade Wang, et al. |
| EPI_ISL_966413 | DOHMH Central Harlem | New York City Public Health Laboratory | Jade Wang, et al. |
| EPI_ISL_966414 | DOHMH Morrisania | New York City Public Health Laboratory | Jade Wang, et al. |
| EPI_ISL_966415 | DOHMH Corona | New York City Public Health Laboratory | Jade Wang, et al. |
| EPI_ISL_966416 | DOHMH Central Harlem | New York City Public Health Laboratory | Jade Wang, et al. |
| EPI_ISL_966417, EPI_ISL_966418, EPI_ISL_966419 | OCME Office Of Chief Medical Examiner | New York City Public Health Laboratory | Jade Wang, et al. |
| EPI_ISL_966420 | DOHMH Morrisania | New York City Public Health Laboratory | Jade Wang, et al. |
| EPI_ISL_966421, EPI_ISL_966422 | DOHMH Jamaica | New York City Public Health Laboratory | Jade Wang, et al. |
| EPI_ISL_966423 | DOHMH Morrisania | New York City Public Health Laboratory | Jade Wang, et al. |
| EPI_ISL_966424 | DOHMH Jamaica | New York City Public Health Laboratory | Jade Wang, et al. |
| EPI_ISL_966425 | DOHMH Corona | New York City Public Health Laboratory | Jade Wang, et al. |
| EPI_ISL_966426 | DOHMH PHL | New York City Public Health Laboratory | Jade Wang, et al. |
| EPI_ISL_966427 | OCME Office Of Chief Medical Examiner | New York City Public Health Laboratory | Jade Wang, et al. |
| EPI_ISL_966428 | NYU Langone Health | Departments of Pathology and Medicine, New York University School of Medicine | Adriana Heguy, Dacia Dimartino, Emily Guzman, Christian Marier, Peter Meyn, Sitharam Ramaswami, Gael Westby, Paul Zappile, Yutong Zhang, Paolo Cotzia, Guiqing Wang |
| EPI_ISL_966429, EPI_ISL_966430, EPI_ISL_966431 | OCME Office Of Chief Medical Examiner | New York City Public Health Laboratory | Jade Wang, et al. |
| EPI_ISL_966432, EPI_ISL_966433 | DOHMH Jamaica | New York City Public Health Laboratory | Jade Wang, et al. |
| EPI_ISL_966434, EPI_ISL_966435, EPI_ISL_966436, EPI_ISL_966437 | DOHMH Morrisania | New York City Public Health Laboratory | Jade Wang, et al. |
| EPI_ISL_966438, EPI_ISL_966439, EPI_ISL_966440, EPI_ISL_966441, EPI_ISL_966442 | OCME Office Of Chief Medical Examiner | New York City Public Health Laboratory | Jade Wang, et al. |

| | | | |
|---|---|---|---|
| EPI_ISL_966443 | DOHMH Fort Greene | New York City Public Health Laboratory | Jade Wang, et al. |
| EPI_ISL_966444 | DOHMH PHL | New York City Public Health Laboratory | Jade Wang, et al. |
| EPI_ISL_966445, EPI_ISL_966446 | DOHMH Corona | New York City Public Health Laboratory | Jade Wang, et al. |
| EPI_ISL_966447, EPI_ISL_966448, EPI_ISL_966449 | DOHMH Jamaica | New York City Public Health Laboratory | Jade Wang, et al. |
| EPI_ISL_966450 | DOHMH Central Harlem | New York City Public Health Laboratory | Jade Wang, et al. |
| EPI_ISL_966451 | DOHMH Morrisania | New York City Public Health Laboratory | Jade Wang, et al. |
| EPI_ISL_966452 | NYU Langone Health | Departments of Pathology and Medicine, New York University School of Medicine | Adriana Heguy, Dacia Dimartino, Emily Guzman, Christian Marier, Peter Meyn, Sitharam Ramaswami, Gael Westby, Paul Zappile, Yutong Zhang, Paolo Cotzia, Guiqing Wang |
| EPI_ISL_966453, EPI_ISL_966454, EPI_ISL_966455, EPI_ISL_966456, EPI_ISL_966457, EPI_ISL_966458 | OCME Office Of Chief Medical Examiner | New York City Public Health Laboratory | Jade Wang, et al. |
| EPI_ISL_966459 | DOHMH Corona | New York City Public Health Laboratory | Jade Wang, et al. |
| EPI_ISL_966460 | DOHMH Morrisania | New York City Public Health Laboratory | Jade Wang, et al. |
| EPI_ISL_966461, EPI_ISL_966462, EPI_ISL_966463 | DOHMH Jamaica | New York City Public Health Laboratory | Jade Wang, et al. |
| EPI_ISL_966464, EPI_ISL_966465, EPI_ISL_966466 | DOHMH PHL | New York City Public Health Laboratory | Jade Wang, et al. |
| EPI_ISL_966467 | DOHMH Corona | New York City Public Health Laboratory | Jade Wang, et al. |
| EPI_ISL_966468 | DOHMH Chelsea | New York City Public Health Laboratory | Jade Wang, et al. |
| EPI_ISL_966469 | Department of Homeless Services | New York City Public Health Laboratory | Jade Wang, et al. |
| EPI_ISL_966470, EPI_ISL_966471 | OCME Office Of Chief Medical Examiner | New York City Public Health Laboratory | Jade Wang, et al. |
| EPI_ISL_966472 | NYU Langone Health | Departments of Pathology and Medicine, New York University School of Medicine | Adriana Heguy, Dacia Dimartino, Emily Guzman, Christian Marier, Peter Meyn, Sitharam Ramaswami, Gael Westby, Paul Zappile, Yutong Zhang, Paolo Cotzia, Guiqing Wang |
| EPI_ISL_966473 | OCME Office Of Chief Medical Examiner | New York City Public Health Laboratory | Jade Wang, et al. |
| EPI_ISL_966474 | DOHMH Corona | New York City Public Health Laboratory | Jade Wang, et al. |
| EPI_ISL_966475 | DOHMH Central Harlem | New York City Public Health Laboratory | Jade Wang, et al. |
| EPI_ISL_966476, EPI_ISL_966477, EPI_ISL_966478 | DOHMH Corona | New York City Public Health Laboratory | Jade Wang, et al. |
| EPI_ISL_966479 | DOHMH Jamaica | New York City Public Health Laboratory | Jade Wang, et al. |
| EPI_ISL_966480 | DOHMH Corona | New York City Public Health Laboratory | Jade Wang, et al. |
| EPI_ISL_966481 | DOHMH Jamaica | New York City Public Health Laboratory | Jade Wang, et al. |
| EPI_ISL_966482 | DOHMH Corona | New York City Public Health Laboratory | Jade Wang, et al. |
| EPI_ISL_966483 | DOHMH Chelsea | New York City Public Health Laboratory | Jade Wang, et al. |
| EPI_ISL_966484 | DOHMH Crown Heights | New York City Public Health Laboratory | Jade Wang, et al. |
| EPI_ISL_966485, EPI_ISL_966486, EPI_ISL_966487, EPI_ISL_966488 | OCME Office Of Chief Medical Examiner | New York City Public Health Laboratory | Jade Wang, et al. |
| EPI_ISL_966489 | NYU Langone Health | Departments of Pathology and Medicine, New York University School of Medicine | Adriana Heguy, Dacia Dimartino, Emily Guzman, Christian Marier, Peter Meyn, Sitharam Ramaswami, Gael Westby, Paul Zappile, Yutong Zhang, Paolo Cotzia, Guiqing Wang |
| EPI_ISL_966490 | OCME Office Of Chief Medical Examiner | New York City Public Health Laboratory | Jade Wang, et al. |
| EPI_ISL_966491 | DOHMH Corona | New York City Public Health Laboratory | Jade Wang, et al. |
| EPI_ISL_966492, EPI_ISL_966493, EPI_ISL_966494, EPI_ISL_966495 | OCME Office Of Chief Medical Examiner | New York City Public Health Laboratory | Jade Wang, et al. |
| EPI_ISL_966496 | NYU Langone Health | Departments of Pathology and Medicine, New York University School of Medicine | Adriana Heguy, Dacia Dimartino, Emily Guzman, Christian Marier, Peter Meyn, Sitharam Ramaswami, Gael Westby, Paul Zappile, Yutong Zhang, Paolo Cotzia, Guiqing Wang |
| EPI_ISL_966525 | Helix/Illumina | Respiratory Viruses Branch, Division of Viral Diseases, Centers for Disease Control and Prevention | Peter W. Cook,Dakota Howard,Dhwani Batra,Ben L. Rambo-Martin,Eileen de Feo,Jan Antico,Christine Tran,Matthew Tolentino,Shannon Wickline,Kim Gietzen,Brad Sickler,Jingtao Liu,Eric Allen,Phil Febbo,Summer Galloway,Nicole L. Washington,Simon White,Geraint Levan,Kelly Schiabor Barrett,Elizabeth Cirulli,Alexandre Bolze,Ary Ascencio,Charlotte Rivera-Garcia,Ryan Cho,Jason Nguyen,Sherry Wang,Jimmy Ramirez,Tyler Cassens,Efren Sandoval,Magnus Isaksson,William Lee,David Becker,Marc Laurent,James Lu,Clinton R. Paden,Suxiang Tong,Duncan MacCannell, |
| EPI_ISL_966549, EPI_ISL_966551, EPI_ISL_966552, EPI_ISL_966565, EPI_ISL_966591, EPI_ISL_966605, EPI_ISL_966623, EPI_ISL_966646, EPI_ISL_966663, EPI_ISL_966682 | NYU Langone Health | Departments of Pathology and Medicine, New York University School of Medicine | Adriana Heguy, Dacia Dimartino, Emily Guzman, Christian Marier, Peter Meyn, Sitharam Ramaswami, Gael Westby, Paul Zappile, Yutong Zhang, Paolo Cotzia, Guiqing Wang |
| EPI_ISL_966706 | Helix/Illumina | Respiratory Viruses Branch, Division of Viral Diseases, Centers for Disease Control and Prevention | Peter W. Cook,Dakota Howard,Dhwani Batra,Ben L. Rambo-Martin,Eileen de Feo,Jan Antico,Christine Tran,Matthew Tolentino,Shannon Wickline,Kim Gietzen,Brad Sickler,Jingtao Liu,Eric Allen,Phil Febbo,Summer Galloway,Nicole L. Washington,Simon White,Geraint Levan,Kelly Schiabor Barrett,Elizabeth Cirulli,Alexandre Bolze,Ary Ascencio,Charlotte Rivera-Garcia,Ryan Cho,Jason Nguyen,Sherry Wang,Jimmy Ramirez,Tyler Cassens,Efren Sandoval,Magnus Isaksson,William Lee,David Becker,Marc Laurent,James Lu,Clinton R. Paden,Suxiang Tong,Duncan MacCannell, |
| EPI_ISL_966707 | NYU Langone Health | Departments of Pathology and Medicine, New York University School of Medicine | Adriana Heguy, Dacia Dimartino, Emily Guzman, Christian Marier, Peter Meyn, Sitharam Ramaswami, Gael Westby, Paul Zappile, Yutong Zhang, Paolo Cotzia, Guiqing Wang |
| EPI_ISL_966952, EPI_ISL_967055, EPI_ISL_967085, EPI_ISL_967137, EPI_ISL_967207, EPI_ISL_967230, EPI_ISL_967296, EPI_ISL_967332, EPI_ISL_967358, EPI_ISL_967678, EPI_ISL_967789, EPI_ISL_967880 | | | |
| see above | Helix/Illumina | Respiratory Viruses Branch, Division of Viral Diseases, Centers for Disease Control and Prevention | Peter W. Cook,Dakota Howard,Dhwani Batra,Ben L. Rambo-Martin,Eileen de Feo,Jan Antico,Christine Tran,Matthew Tolentino,Shannon Wickline,Kim Gietzen,Brad Sickler,Jingtao Liu,Eric Allen,Phil Febbo,Summer Galloway,Nicole L. Washington,Simon White,Geraint Levan,Kelly Schiabor Barrett,Elizabeth Cirulli,Alexandre Bolze,Ary Ascencio,Charlotte Rivera-Garcia,Ryan Cho,Jason Nguyen,Sherry Wang,Jimmy Ramirez,Tyler Cassens,Efren Sandoval,Magnus Isaksson,William Lee,David Becker,Marc Laurent,James Lu,Clinton R. Paden,Suxiang Tong,Duncan MacCannell, |
| EPI_ISL_968080 | Pandemic Response Lab - NYC | Pandemic Response Lab, R&D | Henry Lee, Michael Hammerling, Melissa Hopkins, Cybill del Castillo, William Ward, Pradeep Bugga, Haiping Hao, Jon Laurent |
| EPI_ISL_977666 | NYU Langone Health | Departments of Pathology and Medicine, New York University School of Medicine | Adriana Heguy, Dacia Dimartino, Emily Guzman, Christian Marier, Peter Meyn, Sitharam Ramaswami, Gael Westby, Paul Zappile, Yutong Zhang, Paolo Cotzia, Guiqing Wang |

| | | | |
|---|---|---|---|
| EPI_ISL_978603, EPI_ISL_978726, EPI_ISL_978736, EPI_ISL_978741 | Helix/Illumina | Respiratory Viruses Branch, Division of Viral Diseases, Centers for Disease Control and Prevention | Peter W. Cook,Dakota Howard,Dhwani Batra,Ben L. Rambo-Martin,Eileen de Feo,Jan Antico,Christine Tran,Matthew Tolentino,Shannon Wickline,Kim Gietzen,Brad Sickler,Jingtao Liu,Eric Allen,Phil Febbo,Summer Galloway,Nicole L. Washington,Simon White,Geraint Levan,Kelly Schiabor Barrett,Elizabeth Cirulli,Alexandre Bolze,Ary Ascencio,Charlotte Rivera-Garcia,Ryan Cho,Jason Nguyen,Sherry Wang,Jimmy Ramirez,Tyler Cassens,Efren Sandoval,Magnus Isaksson,William Lee,David Becker,Marc Laurent,James Lu,Clinton R. Paden,Suxiang Tong,Duncan MacCannell, |
| EPI_ISL_982425, EPI_ISL_982426, EPI_ISL_982427, EPI_ISL_982428, EPI_ISL_982429, EPI_ISL_982430, EPI_ISL_982431, EPI_ISL_982432, EPI_ISL_982433, EPI_ISL_982434, EPI_ISL_982435, EPI_ISL_982436, EPI_ISL_982437, EPI_ISL_982438, EPI_ISL_982439, EPI_ISL_982440, EPI_ISL_982441, EPI_ISL_982442, EPI_ISL_982443, EPI_ISL_982444, EPI_ISL_982445, EPI_ISL_982446, EPI_ISL_982447, EPI_ISL_982448, EPI_ISL_982449, EPI_ISL_982450, EPI_ISL_982451, EPI_ISL_982452, EPI_ISL_982453, EPI_ISL_982454, EPI_ISL_982455, EPI_ISL_982456, EPI_ISL_982457, EPI_ISL_982458, EPI_ISL_982459, EPI_ISL_982460, EPI_ISL_982461, EPI_ISL_982462, EPI_ISL_982463, EPI_ISL_982464, EPI_ISL_982465, EPI_ISL_982466, EPI_ISL_982467, EPI_ISL_982468, EPI_ISL_982469, EPI_ISL_982470, EPI_ISL_982471, EPI_ISL_982472, EPI_ISL_982473, EPI_ISL_982474, EPI_ISL_982475, EPI_ISL_982476, EPI_ISL_982477, EPI_ISL_982478, EPI_ISL_982479, EPI_ISL_982480, EPI_ISL_982481, EPI_ISL_982482, EPI_ISL_982483, EPI_ISL_982484, EPI_ISL_982485, EPI_ISL_982486, EPI_ISL_982487, EPI_ISL_982488, EPI_ISL_982489, EPI_ISL_982490, EPI_ISL_982491, EPI_ISL_982492, EPI_ISL_982493, EPI_ISL_982494, EPI_ISL_982495, EPI_ISL_982496, EPI_ISL_982497, EPI_ISL_982498 | | | |
| see above | MONTEFIORE MEDICAL CENTER LABORATORIES | Wadsworth Center, New York State Department of Health | Kirsten St. George, Daryl M. Lamson, Alexis Russel, Matthew Shudt, Melissa A Leisner, Jonathan Plitnick, Navjot Singh, John Kelly, Erasmus Schneider, Erica Lasek-Nesselquist |
| EPI_ISL_982499, EPI_ISL_982500, EPI_ISL_982501, EPI_ISL_982502, EPI_ISL_982503, EPI_ISL_982504 | ADIRONDACK MEDICAL CENTER | Wadsworth Center, New York State Department of Health | Kirsten St. George, Daryl M. Lamson, Alexis Russel, Matthew Shudt, Melissa A Leisner, Jonathan Plitnick, Navjot Singh, John Kelly, Erasmus Schneider, Erica Lasek-Nesselquist |
| EPI_ISL_983099, EPI_ISL_983100, EPI_ISL_983101, EPI_ISL_983102, EPI_ISL_983103, EPI_ISL_983104, EPI_ISL_983105, EPI_ISL_983106, EPI_ISL_983107, EPI_ISL_983108, EPI_ISL_983109, EPI_ISL_983110, EPI_ISL_983111, EPI_ISL_983112, EPI_ISL_983113, EPI_ISL_983114, EPI_ISL_983115 | | | |
| see above | MONTEFIORE MEDICAL CENTER LABORATORIES | Wadsworth Center, New York State Department of Health | Kirsten St. George, Daryl M. Lamson, Alexis Russel, Matthew Shudt, Melissa A Leisner, Jonathan Plitnick, Navjot Singh, John Kelly, Erasmus Schneider, Erica Lasek-Nesselquist |
| EPI_ISL_983116, EPI_ISL_983117, EPI_ISL_983118 | SUNY UPSTATE MEDICAL UNIVERSITY | Wadsworth Center, New York State Department of Health | Kirsten St. George, Daryl M. Lamson, Alexis Russel, Matthew Shudt, Melissa A Leisner, Jonathan Plitnick, Navjot Singh, John Kelly, Erasmus Schneider, Erica Lasek-Nesselquist |
| EPI_ISL_983119, EPI_ISL_983120, EPI_ISL_983121, EPI_ISL_983122, EPI_ISL_983123, EPI_ISL_983124, EPI_ISL_983125, EPI_ISL_983126, EPI_ISL_983127, EPI_ISL_983128, EPI_ISL_983129, EPI_ISL_983130, EPI_ISL_983131 | | | |
| see above | THE MARY IMOGENE BASSETT HOSPITAL | Wadsworth Center, New York State Department of Health | Kirsten St. George, Daryl M. Lamson, Alexis Russel, Matthew Shudt, Melissa A Leisner, Jonathan Plitnick, Navjot Singh, John Kelly, Erasmus Schneider, Erica Lasek-Nesselquist |
| EPI_ISL_983132, EPI_ISL_983133, EPI_ISL_983134, EPI_ISL_983135, EPI_ISL_983136, EPI_ISL_983137, EPI_ISL_983138, EPI_ISL_983139, EPI_ISL_983140, EPI_ISL_983141, EPI_ISL_983142, EPI_ISL_983143, EPI_ISL_983144, EPI_ISL_983145, EPI_ISL_983146, EPI_ISL_983147, EPI_ISL_983148, EPI_ISL_983149, EPI_ISL_983150, EPI_ISL_983151, EPI_ISL_983152, EPI_ISL_983153, EPI_ISL_983154, EPI_ISL_983155, EPI_ISL_983156, EPI_ISL_983157, EPI_ISL_983158, EPI_ISL_983159, EPI_ISL_983160, EPI_ISL_983161, EPI_ISL_983162, EPI_ISL_983163, EPI_ISL_983164, EPI_ISL_983165, EPI_ISL_983166, EPI_ISL_983167, EPI_ISL_983168, EPI_ISL_983169, EPI_ISL_983170 | | | |
| see above | MONTEFIORE MEDICAL CENTER LABORATORIES | Wadsworth Center, New York State Department of Health | Kirsten St. George, Daryl M. Lamson, Alexis Russel, Matthew Shudt, Melissa A Leisner, Jonathan Plitnick, Navjot Singh, John Kelly, Erasmus Schneider, Erica Lasek-Nesselquist |
| EPI_ISL_983240, EPI_ISL_983241, EPI_ISL_983242 | KALEIDA CENTER FOR LABORATORY MEDICINE | Wadsworth Center, New York State Department of Health | Kirsten St. George, Daryl M. Lamson, Alexis Russel, Matthew Shudt, Melissa A Leisner, Jonathan Plitnick, Navjot Singh, John Kelly, Erasmus Schneider, Erica Lasek-Nesselquist |
| EPI_ISL_983243 | SUNY UPSTATE MEDICAL UNIVERSITY | Wadsworth Center, New York State Department of Health | Kirsten St. George, Daryl M. Lamson, Alexis Russel, Matthew Shudt, Melissa A Leisner, Jonathan Plitnick, Navjot Singh, John Kelly, Erasmus Schneider, Erica Lasek-Nesselquist |
| EPI_ISL_983244 | KALEIDA CENTER FOR LABORATORY MEDICINE | Wadsworth Center, New York State Department of Health | Kirsten St. George, Daryl M. Lamson, Alexis Russel, Matthew Shudt, Melissa A Leisner, Jonathan Plitnick, Navjot Singh, John Kelly, Erasmus Schneider, Erica Lasek-Nesselquist |
| EPI_ISL_983245 | SUNY UPSTATE MEDICAL UNIVERSITY | Wadsworth Center, New York State Department of Health | Kirsten St. George, Daryl M. Lamson, Alexis Russel, Matthew Shudt, Melissa A Leisner, Jonathan Plitnick, Navjot Singh, John Kelly, Erasmus Schneider, Erica Lasek-Nesselquist |
| EPI_ISL_983246, EPI_ISL_983247 | KALEIDA CENTER FOR LABORATORY MEDICINE | Wadsworth Center, New York State Department of Health | Kirsten St. George, Daryl M. Lamson, Alexis Russel, Matthew Shudt, Melissa A Leisner, Jonathan Plitnick, Navjot Singh, John Kelly, Erasmus Schneider, Erica Lasek-Nesselquist |
| EPI_ISL_983248, EPI_ISL_983249 | SUNY UPSTATE MEDICAL UNIVERSITY | Wadsworth Center, New York State Department of Health | Kirsten St. George, Daryl M. Lamson, Alexis Russel, Matthew Shudt, Melissa A Leisner, Jonathan Plitnick, Navjot Singh, John Kelly, Erasmus Schneider, Erica Lasek-Nesselquist |
| EPI_ISL_983250, EPI_ISL_983251, EPI_ISL_983252, EPI_ISL_983253, EPI_ISL_983254, EPI_ISL_983255, EPI_ISL_983256, EPI_ISL_983257, EPI_ISL_983258, EPI_ISL_983259, EPI_ISL_983260, EPI_ISL_983261, EPI_ISL_983262, EPI_ISL_983263, EPI_ISL_983264, EPI_ISL_983265, EPI_ISL_983266, EPI_ISL_983267, EPI_ISL_983268, EPI_ISL_983269, EPI_ISL_983270, EPI_ISL_983271, EPI_ISL_983272, EPI_ISL_983273, EPI_ISL_983274, EPI_ISL_983275, EPI_ISL_983276, EPI_ISL_983277, EPI_ISL_983278, EPI_ISL_983279, EPI_ISL_983280, EPI_ISL_983281, EPI_ISL_983282 | | | |
| see above | KALEIDA CENTER FOR LABORATORY MEDICINE | Wadsworth Center, New York State Department of Health | Kirsten St. George, Daryl M. Lamson, Alexis Russel, Matthew Shudt, Melissa A Leisner, Jonathan Plitnick, Navjot Singh, John Kelly, Erasmus Schneider, Erica Lasek-Nesselquist |
| EPI_ISL_983283 | SUNY UPSTATE MEDICAL UNIVERSITY | Wadsworth Center, New York State Department of Health | Kirsten St. George, Daryl M. Lamson, Alexis Russel, Matthew Shudt, Melissa A Leisner, Jonathan Plitnick, Navjot Singh, John Kelly, Erasmus Schneider, Erica Lasek-Nesselquist |
| EPI_ISL_983284, EPI_ISL_983285, EPI_ISL_983286, EPI_ISL_983287, EPI_ISL_983288 | KALEIDA CENTER FOR LABORATORY MEDICINE | Wadsworth Center, New York State Department of Health | Kirsten St. George, Daryl M. Lamson, Alexis Russel, Matthew Shudt, Melissa A Leisner, Jonathan Plitnick, Navjot Singh, John Kelly, Erasmus Schneider, Erica Lasek-Nesselquist |
| EPI_ISL_983289, EPI_ISL_983290, EPI_ISL_983291, EPI_ISL_983292, EPI_ISL_983293, EPI_ISL_983294, EPI_ISL_983295 | SUNY UPSTATE MEDICAL UNIVERSITY | Wadsworth Center, New York State Department of Health | Kirsten St. George, Daryl M. Lamson, Alexis Russel, Matthew Shudt, Melissa A Leisner, Jonathan Plitnick, Navjot Singh, John Kelly, Erasmus Schneider, Erica Lasek-Nesselquist |
| EPI_ISL_983296, EPI_ISL_983297, EPI_ISL_983298, EPI_ISL_983299 | KALEIDA CENTER FOR LABORATORY MEDICINE | Wadsworth Center, New York State Department of Health | Kirsten St. George, Daryl M. Lamson, Alexis Russel, Matthew Shudt, Melissa A Leisner, Jonathan Plitnick, Navjot Singh, John Kelly, Erasmus Schneider, Erica Lasek-Nesselquist |
| EPI_ISL_983300, EPI_ISL_983301, EPI_ISL_983302, EPI_ISL_983303, EPI_ISL_983304, EPI_ISL_983305, EPI_ISL_983306, EPI_ISL_983307, EPI_ISL_983308, EPI_ISL_983309, EPI_ISL_983310, EPI_ISL_983311, EPI_ISL_983312, EPI_ISL_983313, EPI_ISL_983314, EPI_ISL_983315, EPI_ISL_983316, EPI_ISL_983317, EPI_ISL_983318, EPI_ISL_983319 | | | |
| see above | SUNY UPSTATE MEDICAL UNIVERSITY | Wadsworth Center, New York State Department of Health | Kirsten St. George, Daryl M. Lamson, Alexis Russel, Matthew Shudt, Melissa A Leisner, Jonathan Plitnick, Navjot Singh, John Kelly, Erasmus Schneider, Erica Lasek-Nesselquist |
| EPI_ISL_983438, EPI_ISL_983439, EPI_ISL_983440, EPI_ISL_983441, EPI_ISL_983442, EPI_ISL_983443, EPI_ISL_983444 | URMC LABS | Wadsworth Center, New York State Department of Health | Kirsten St. George, Daryl M. Lamson, Alexis Russel, Matthew Shudt, Melissa A Leisner, Jonathan Plitnick, Navjot Singh, John Kelly, Erasmus Schneider, Erica Lasek-Nesselquist |
| EPI_ISL_983445, EPI_ISL_983446 | THE MARY IMOGENE BASSETT HOSPITAL | Wadsworth Center, New York State Department of Health | Kirsten St. George, Daryl M. Lamson, Alexis Russel, Matthew Shudt, Melissa A Leisner, Jonathan Plitnick, Navjot Singh, John Kelly, Erasmus Schneider, Erica Lasek-Nesselquist |
| EPI_ISL_983447, EPI_ISL_983448, EPI_ISL_983449, EPI_ISL_983450, EPI_ISL_983451, EPI_ISL_983452, EPI_ISL_983453, EPI_ISL_983454, EPI_ISL_983455, EPI_ISL_983456, EPI_ISL_983457, EPI_ISL_983458, EPI_ISL_983459, EPI_ISL_983460, EPI_ISL_983461, EPI_ISL_983462, EPI_ISL_983463, EPI_ISL_983464, EPI_ISL_983465, EPI_ISL_983466, EPI_ISL_983467, EPI_ISL_983468, EPI_ISL_983469, EPI_ISL_983470, EPI_ISL_983471, EPI_ISL_983472, EPI_ISL_983473, EPI_ISL_983474, EPI_ISL_983475 | | | |
| see above | URMC LABS | Wadsworth Center, New York State Department of Health | Kirsten St. George, Daryl M. Lamson, Alexis Russel, Matthew Shudt, Melissa A Leisner, Jonathan Plitnick, Navjot Singh, John Kelly, Erasmus Schneider, Erica Lasek-Nesselquist |
| EPI_ISL_983476 | Wadsworth Center, New York State Department of Health | Wadsworth Center, New York State Department of Health | Kirsten St. George, Daryl M. Lamson, Alexis Russel, Matthew Shudt, Melissa A Leisner, Jonathan Plitnick, Navjot Singh, John Kelly, Erasmus Schneider, Erica Lasek-Nesselquist |
| EPI_ISL_983477 | THE MARY IMOGENE BASSETT HOSPITAL | Wadsworth Center, New York State Department of Health | Kirsten St. George, Daryl M. Lamson, Alexis Russel, Matthew Shudt, Melissa A Leisner, Jonathan Plitnick, Navjot Singh, John Kelly, Erasmus Schneider, Erica Lasek-Nesselquist |

| | | | |
|---|---|---|---|
| EPI_ISL_983478, EPI_ISL_983479, EPI_ISL_983480, EPI_ISL_983481, EPI_ISL_983482, EPI_ISL_983483, EPI_ISL_983484 | URMC LABS | Wadsworth Center, New York State Department of Health | Kirsten St. George, Daryl M. Lamson, Alexis Russel, Matthew Shudt, Melissa A Leisner, Jonathan Plitnick, Navjot Singh, John Kelly, Erasmus Schneider, Erica Lasek-Nesselquist |
| EPI_ISL_983485, EPI_ISL_983486, EPI_ISL_983487, EPI_ISL_983488, EPI_ISL_983489, EPI_ISL_983490, EPI_ISL_983491, EPI_ISL_983492 | THE MARY IMOGENE BASSETT HOSPITAL | Wadsworth Center, New York State Department of Health | Kirsten St. George, Daryl M. Lamson, Alexis Russel, Matthew Shudt, Melissa A Leisner, Jonathan Plitnick, Navjot Singh, John Kelly, Erasmus Schneider, Erica Lasek-Nesselquist |
| EPI_ISL_984756, EPI_ISL_984757, EPI_ISL_984758, EPI_ISL_984759, EPI_ISL_984760, EPI_ISL_984761, EPI_ISL_984762, EPI_ISL_984763, EPI_ISL_984764, EPI_ISL_984765, EPI_ISL_984766, EPI_ISL_984767, EPI_ISL_984768, EPI_ISL_984769, EPI_ISL_984770, EPI_ISL_984771, EPI_ISL_984772, EPI_ISL_984773, EPI_ISL_984774, EPI_ISL_984775, EPI_ISL_984776, EPI_ISL_984777, EPI_ISL_984778, EPI_ISL_984779, EPI_ISL_984780, EPI_ISL_984781, EPI_ISL_984782, EPI_ISL_984783, EPI_ISL_984784, EPI_ISL_984785, EPI_ISL_984786, EPI_ISL_984787, EPI_ISL_984788, EPI_ISL_984789, EPI_ISL_984790, EPI_ISL_984791, EPI_ISL_984792, EPI_ISL_984793, EPI_ISL_984794, EPI_ISL_984795, EPI_ISL_984796, EPI_ISL_984797, EPI_ISL_984798, EPI_ISL_984799, EPI_ISL_984800, EPI_ISL_984801, EPI_ISL_984802, EPI_ISL_984803, EPI_ISL_984804, EPI_ISL_984805, EPI_ISL_984806, EPI_ISL_984807, EPI_ISL_984808, EPI_ISL_984809, EPI_ISL_984810, EPI_ISL_984811, EPI_ISL_984812, EPI_ISL_984813, EPI_ISL_984814, EPI_ISL_984815, EPI_ISL_984816, EPI_ISL_984817, EPI_ISL_984818, EPI_ISL_984819, EPI_ISL_984820, EPI_ISL_984821, EPI_ISL_984822, EPI_ISL_984823, EPI_ISL_984824, EPI_ISL_984825, EPI_ISL_984826, EPI_ISL_984827, EPI_ISL_984828, EPI_ISL_984829, EPI_ISL_984830, EPI_ISL_984831, EPI_ISL_984832, EPI_ISL_984833, EPI_ISL_984834, EPI_ISL_984835, EPI_ISL_984836, EPI_ISL_984837, EPI_ISL_984838, EPI_ISL_984839, EPI_ISL_984840, EPI_ISL_984841, EPI_ISL_984842, EPI_ISL_984843, EPI_ISL_984844, EPI_ISL_984845, EPI_ISL_984846, EPI_ISL_984847, EPI_ISL_984848, EPI_ISL_984849, EPI_ISL_984850, EPI_ISL_984851, EPI_ISL_984852, EPI_ISL_984853, EPI_ISL_984854, EPI_ISL_984855, EPI_ISL_984856, EPI_ISL_984857, EPI_ISL_984858, EPI_ISL_984859, EPI_ISL_984860, EPI_ISL_984861, EPI_ISL_984862, EPI_ISL_984863, EPI_ISL_984864, EPI_ISL_984865, EPI_ISL_984866, EPI_ISL_984867, EPI_ISL_984868, EPI_ISL_984869, EPI_ISL_984870, EPI_ISL_984871, EPI_ISL_984872, EPI_ISL_984873, EPI_ISL_984874, EPI_ISL_984875, EPI_ISL_984876, EPI_ISL_984877, EPI_ISL_984878, EPI_ISL_984879, EPI_ISL_984880, EPI_ISL_984881, EPI_ISL_984882, EPI_ISL_984883, EPI_ISL_984884, EPI_ISL_984885, EPI_ISL_984886, EPI_ISL_984887, EPI_ISL_984888, EPI_ISL_984889, EPI_ISL_984890, EPI_ISL_984891, EPI_ISL_984892, EPI_ISL_984893, EPI_ISL_984894, EPI_ISL_984895, EPI_ISL_984896, EPI_ISL_984897, EPI_ISL_984898, EPI_ISL_984899, EPI_ISL_984900, EPI_ISL_984901, EPI_ISL_984902, EPI_ISL_984903, EPI_ISL_984904, EPI_ISL_984905, EPI_ISL_984906, EPI_ISL_984907, EPI_ISL_984908, EPI_ISL_984909, EPI_ISL_984910, EPI_ISL_984911, EPI_ISL_984912, EPI_ISL_984913, EPI_ISL_984914, EPI_ISL_984915, EPI_ISL_984916, EPI_ISL_984917, EPI_ISL_984918, EPI_ISL_984919, EPI_ISL_984920, EPI_ISL_984921, EPI_ISL_984922, EPI_ISL_984923, EPI_ISL_984924, EPI_ISL_984925, EPI_ISL_984926, EPI_ISL_984927, EPI_ISL_984928, EPI_ISL_984929, EPI_ISL_984930, EPI_ISL_984931, EPI_ISL_984932, EPI_ISL_984933, EPI_ISL_984934, EPI_ISL_984935, EPI_ISL_984936, EPI_ISL_984937, EPI_ISL_984938, EPI_ISL_984939, EPI_ISL_984940, EPI_ISL_984941, EPI_ISL_984942, EPI_ISL_984943, EPI_ISL_984944, EPI_ISL_984945, EPI_ISL_984946, EPI_ISL_984947, EPI_ISL_994802, EPI_ISL_994803, EPI_ISL_994804, EPI_ISL_994805, EPI_ISL_994806, EPI_ISL_994807, EPI_ISL_994808, EPI_ISL_994809, EPI_ISL_994810, EPI_ISL_994811, EPI_ISL_994812, EPI_ISL_994813, EPI_ISL_994814, EPI_ISL_994815, EPI_ISL_994816, EPI_ISL_994817, EPI_ISL_994818, EPI_ISL_994819, EPI_ISL_994820, EPI_ISL_994821, EPI_ISL_994822, EPI_ISL_994823, EPI_ISL_994824, EPI_ISL_994825, EPI_ISL_994826, EPI_ISL_994827, EPI_ISL_994828, EPI_ISL_994829, EPI_ISL_994830, EPI_ISL_994831, EPI_ISL_994832, EPI_ISL_994833, EPI_ISL_994834, EPI_ISL_994835, EPI_ISL_994836, EPI_ISL_994837, EPI_ISL_994838, EPI_ISL_994839, EPI_ISL_994840, EPI_ISL_994841, EPI_ISL_994842, EPI_ISL_994843, EPI_ISL_994844, EPI_ISL_994845, EPI_ISL_994846, EPI_ISL_994847, EPI_ISL_994848, EPI_ISL_994849, EPI_ISL_994850, EPI_ISL_994851, EPI_ISL_994852, EPI_ISL_994853, EPI_ISL_994854, EPI_ISL_994855, EPI_ISL_994856, EPI_ISL_994857, EPI_ISL_994858, EPI_ISL_994859, EPI_ISL_994860, EPI_ISL_994861, EPI_ISL_994862, EPI_ISL_994863, EPI_ISL_994864, EPI_ISL_994865, EPI_ISL_994866, EPI_ISL_994867, EPI_ISL_994868, EPI_ISL_994869, EPI_ISL_994870, EPI_ISL_994871, EPI_ISL_994872, EPI_ISL_994873, EPI_ISL_994874, EPI_ISL_994875, EPI_ISL_994876, EPI_ISL_994877, EPI_ISL_994878, EPI_ISL_994879, EPI_ISL_994880, EPI_ISL_994881, EPI_ISL_994882, EPI_ISL_994883, EPI_ISL_994884, EPI_ISL_994885, EPI_ISL_994886, EPI_ISL_994887, EPI_ISL_994888, EPI_ISL_994889, EPI_ISL_994890, EPI_ISL_994891, EPI_ISL_994892, EPI_ISL_994893, EPI_ISL_994894, EPI_ISL_994895, EPI_ISL_994896, EPI_ISL_994897, EPI_ISL_994898, EPI_ISL_994899, EPI_ISL_994900, EPI_ISL_994901, EPI_ISL_994902, EPI_ISL_994903, EPI_ISL_994904, EPI_ISL_994905, EPI_ISL_994906, EPI_ISL_994907, EPI_ISL_994908, EPI_ISL_994909, EPI_ISL_994910, EPI_ISL_994911, EPI_ISL_994912, EPI_ISL_994913, EPI_ISL_994914, EPI_ISL_994915, EPI_ISL_994916, EPI_ISL_994917, EPI_ISL_994918, EPI_ISL_994919, EPI_ISL_994920, EPI_ISL_994921, EPI_ISL_994922, EPI_ISL_994923, EPI_ISL_994924, EPI_ISL_994925, EPI_ISL_994926, EPI_ISL_994927, EPI_ISL_994928, EPI_ISL_994929, EPI_ISL_994930, EPI_ISL_994931, EPI_ISL_994932, EPI_ISL_994933, EPI_ISL_994934, EPI_ISL_994935, EPI_ISL_994936, EPI_ISL_994937, EPI_ISL_994938, EPI_ISL_994939, EPI_ISL_994940, EPI_ISL_994941, EPI_ISL_994942, EPI_ISL_994943, EPI_ISL_994944, EPI_ISL_994945, EPI_ISL_994946, EPI_ISL_994947, EPI_ISL_994948, EPI_ISL_994949, EPI_ISL_994950, EPI_ISL_994951, EPI_ISL_994952, EPI_ISL_994953, EPI_ISL_994954, EPI_ISL_994955, EPI_ISL_994956, EPI_ISL_994957, EPI_ISL_994958, EPI_ISL_994959, EPI_ISL_994960, EPI_ISL_994961, EPI_ISL_994962, EPI_ISL_994963, EPI_ISL_994964, EPI_ISL_994965, EPI_ISL_994966, EPI_ISL_994967, EPI_ISL_994968, EPI_ISL_994969, EPI_ISL_994970, EPI_ISL_994971, EPI_ISL_994972, EPI_ISL_994973, EPI_ISL_994974, EPI_ISL_994975, EPI_ISL_994976, EPI_ISL_994977, EPI_ISL_994978, EPI_ISL_994979, EPI_ISL_994980, EPI_ISL_994981, EPI_ISL_994982, EPI_ISL_994983, EPI_ISL_994984, EPI_ISL_994985, EPI_ISL_994986, EPI_ISL_994987, EPI_ISL_994988, EPI_ISL_994989, EPI_ISL_994990, EPI_ISL_994991, EPI_ISL_994992, EPI_ISL_994993, EPI_ISL_994994, EPI_ISL_994995, EPI_ISL_994996, EPI_ISL_994997, EPI_ISL_994998, EPI_ISL_994999, EPI_ISL_995000, EPI_ISL_995001, EPI_ISL_995002, EPI_ISL_995003, EPI_ISL_995004, EPI_ISL_995005, EPI_ISL_995006, EPI_ISL_995007, EPI_ISL_995008, EPI_ISL_995009, EPI_ISL_995010, EPI_ISL_995011, EPI_ISL_995012, EPI_ISL_995013, EPI_ISL_995014, EPI_ISL_995015, EPI_ISL_995016, EPI_ISL_995017, EPI_ISL_995018, EPI_ISL_995019, EPI_ISL_995020, EPI_ISL_995021, EPI_ISL_995022, EPI_ISL_995023, EPI_ISL_995024, EPI_ISL_995025, EPI_ISL_995026, EPI_ISL_995027, EPI_ISL_995028, EPI_ISL_995029, EPI_ISL_995030, EPI_ISL_995031, EPI_ISL_995032, EPI_ISL_995033, EPI_ISL_995034, EPI_ISL_995035, EPI_ISL_995036, EPI_ISL_995037, EPI_ISL_995038, EPI_ISL_995039, EPI_ISL_995040, EPI_ISL_995041, EPI_ISL_995042, EPI_ISL_995043, EPI_ISL_995044, EPI_ISL_995045, EPI_ISL_995046, EPI_ISL_995047, EPI_ISL_995048, EPI_ISL_995049, EPI_ISL_995050, EPI_ISL_995051, EPI_ISL_995052, EPI_ISL_995053, EPI_ISL_995054, EPI_ISL_995055, EPI_ISL_995056, EPI_ISL_995057, EPI_ISL_995058, EPI_ISL_995059, EPI_ISL_995060, EPI_ISL_995061, EPI_ISL_995062, EPI_ISL_995063, EPI_ISL_995064, EPI_ISL_995065, EPI_ISL_995066, EPI_ISL_995067, EPI_ISL_995068, EPI_ISL_995069, EPI_ISL_995070, EPI_ISL_995071, EPI_ISL_995072, EPI_ISL_995073, EPI_ISL_995074, EPI_ISL_995075, EPI_ISL_995076, EPI_ISL_995077, EPI_ISL_995078, EPI_ISL_995079, EPI_ISL_995080, EPI_ISL_995081, EPI_ISL_995082, EPI_ISL_995083, EPI_ISL_995084, EPI_ISL_995085, EPI_ISL_995086, EPI_ISL_995087, EPI_ISL_995088, EPI_ISL_995089, EPI_ISL_995090, EPI_ISL_995091, EPI_ISL_995092, EPI_ISL_995093, EPI_ISL_995094, EPI_ISL_995095, EPI_ISL_995096, EPI_ISL_995097, EPI_ISL_995098, EPI_ISL_995099, EPI_ISL_995100, EPI_ISL_995101, EPI_ISL_995102, EPI_ISL_995103, EPI_ISL_995104, EPI_ISL_995105, EPI_ISL_995106, EPI_ISL_995107, EPI_ISL_995108, EPI_ISL_995109, EPI_ISL_995110, EPI_ISL_995111, EPI_ISL_995112, EPI_ISL_995113, EPI_ISL_995114, EPI_ISL_995115, EPI_ISL_995116, EPI_ISL_995117, EPI_ISL_995118, EPI_ISL_995119, EPI_ISL_995120, EPI_ISL_995121, EPI_ISL_995122, EPI_ISL_995123, EPI_ISL_995124, EPI_ISL_995125, EPI_ISL_995126, EPI_ISL_995127, EPI_ISL_995128, EPI_ISL_995129, EPI_ISL_995130, EPI_ISL_995131, EPI_ISL_995132, EPI_ISL_995133, EPI_ISL_995134, EPI_ISL_995135, EPI_ISL_995136, EPI_ISL_995137, EPI_ISL_995138, EPI_ISL_995139, EPI_ISL_995140, EPI_ISL_995141, EPI_ISL_995142, EPI_ISL_995143, EPI_ISL_995144, EPI_ISL_995145, EPI_ISL_995146, EPI_ISL_995147, EPI_ISL_995148, EPI_ISL_995149, EPI_ISL_995150, EPI_ISL_995151, EPI_ISL_995152, EPI_ISL_995153, EPI_ISL_995154, EPI_ISL_995155, EPI_ISL_995156, EPI_ISL_995157, EPI_ISL_995158, EPI_ISL_995159, EPI_ISL_995160, EPI_ISL_995161, EPI_ISL_995162, EPI_ISL_995163, EPI_ISL_995164, EPI_ISL_995165, EPI_ISL_995166, EPI_ISL_995167, EPI_ISL_995168, EPI_ISL_995169, EPI_ISL_995170, EPI_ISL_995171 | | | |
| see above | Pandemic Response Lab - NYC | Pandemic Response Lab, R&D | Henry Lee, Michael Hammerling, Melissa Hopkins, Cybill del Castillo, William Ward, Pradeep Bugga, Haiping Hao, Jon Laurent |
