## Supplemental acknowledgement table #2 for "Genomic Surveillance of SARS-CoV-2 in Erie County, New York"

We gratefully acknowledge the following Authors from the Originating laboratories responsible for obtaining the specimens, as well as the Submitting laboratories where the genome data were generated and shared via GISAID, on which this research is based.

All Submitters of data may be contacted directly via www.gisaid.org

Authors are sorted alphabetically.

| Accession ID | Originating Laboratory | Submitting Laboratory | Authors |
|---|---|---|---|
| EPI_ISL_1009188, EPI_ISL_1009189, EPI_ISL_1009190, EPI_ISL_1009192, EPI_ISL_1009193 | American Esoteric Laboratories | Colleen B. Jonsson | Mariah K. Taylor, Walter Reichard, Jyothi Parvathareddy, Colleen B. Jonsson |
| EPI_ISL_1009289, EPI_ISL_1009290, EPI_ISL_1009291, EPI_ISL_1009292, EPI_ISL_1009293, EPI_ISL_1009294, EPI_ISL_1009295, EPI_ISL_1009296, EPI_ISL_1009297, EPI_ISL_1009298, EPI_ISL_1009299, EPI_ISL_1009300, EPI_ISL_1009301, EPI_ISL_1009302, EPI_ISL_1009303, EPI_ISL_1009304, EPI_ISL_1009305, EPI_ISL_1009306, EPI_ISL_1009307, EPI_ISL_1009308, EPI_ISL_1009309, EPI_ISL_1009310, EPI_ISL_1009311, EPI_ISL_1009312, EPI_ISL_1009313, EPI_ISL_1009314, EPI_ISL_1009315, EPI_ISL_1009316, EPI_ISL_1009317, EPI_ISL_1009318, EPI_ISL_1009319, EPI_ISL_1009320, EPI_ISL_1009321, EPI_ISL_1009322, EPI_ISL_1009323, EPI_ISL_1009324, EPI_ISL_1009325, EPI_ISL_1009326, EPI_ISL_1009327, EPI_ISL_1009328, EPI_ISL_1009329, EPI_ISL_1009330, EPI_ISL_1009331, EPI_ISL_1009332, EPI_ISL_1009333, EPI_ISL_1009334, EPI_ISL_1009335, EPI_ISL_1009336, EPI_ISL_1009337, EPI_ISL_1009338, EPI_ISL_1009339, EPI_ISL_1009340, EPI_ISL_1009341, EPI_ISL_1009342, EPI_ISL_1009343, EPI_ISL_1009344, EPI_ISL_1009345, EPI_ISL_1009346, EPI_ISL_1009347, EPI_ISL_1009348, EPI_ISL_1009349, EPI_ISL_1009350, EPI_ISL_1009351, EPI_ISL_1009352, EPI_ISL_1009353, EPI_ISL_1009354, EPI_ISL_1009355, EPI_ISL_1009356, EPI_ISL_1009357, EPI_ISL_1009358, EPI_ISL_1009359, EPI_ISL_1009360, EPI_ISL_1009361, EPI_ISL_1009362, EPI_ISL_1009363, EPI_ISL_1009364, EPI_ISL_1009365, EPI_ISL_1009366, EPI_ISL_1009367, EPI_ISL_1009368, EPI_ISL_1009369, EPI_ISL_1009370, EPI_ISL_1009371, EPI_ISL_1009372, EPI_ISL_1009373, EPI_ISL_1009374, EPI_ISL_1009375, EPI_ISL_1009376, EPI_ISL_1009377, EPI_ISL_1009378, EPI_ISL_1009379, EPI_ISL_1009380, EPI_ISL_1009381, EPI_ISL_1009382, EPI_ISL_1009383, EPI_ISL_1009384, EPI_ISL_1009385, EPI_ISL_1009386, EPI_ISL_1009387, EPI_ISL_1009388, EPI_ISL_1009389, EPI_ISL_1009390, EPI_ISL_1009391, EPI_ISL_1009392, EPI_ISL_1009393, EPI_ISL_1009394, EPI_ISL_1009395, EPI_ISL_1009396, EPI_ISL_1009397, EPI_ISL_1009398, EPI_ISL_1009399, EPI_ISL_1009400, EPI_ISL_1009401, EPI_ISL_1009402, EPI_ISL_1009403, EPI_ISL_1009404, EPI_ISL_1009405, EPI_ISL_1009406, EPI_ISL_1009407, EPI_ISL_1009408, EPI_ISL_1009409, EPI_ISL_1009410, EPI_ISL_1009411, EPI_ISL_1009412, EPI_ISL_1009413, EPI_ISL_1009414, EPI_ISL_1009415, EPI_ISL_1009416, EPI_ISL_1009417, EPI_ISL_1009418, EPI_ISL_1009419, EPI_ISL_1009420, EPI_ISL_1009421, EPI_ISL_1009422, EPI_ISL_1009423, EPI_ISL_1009424, EPI_ISL_1009425, EPI_ISL_1009426, EPI_ISL_1009427, EPI_ISL_1009428, EPI_ISL_1009429, EPI_ISL_1009430, EPI_ISL_1009431, EPI_ISL_1009432, EPI_ISL_1009433, EPI_ISL_1009434, EPI_ISL_1009435, EPI_ISL_1009436, EPI_ISL_1009437, EPI_ISL_1009438, EPI_ISL_1009439, EPI_ISL_1009440, EPI_ISL_1009441, EPI_ISL_1009442, EPI_ISL_1009443, EPI_ISL_1009444, EPI_ISL_1009445, EPI_ISL_1009446, EPI_ISL_1009447, EPI_ISL_1009448, EPI_ISL_1009449, EPI_ISL_1009450, EPI_ISL_1009451, EPI_ISL_1009452, EPI_ISL_1009453, EPI_ISL_1009454, EPI_ISL_1009455, EPI_ISL_1009456, EPI_ISL_1009457, EPI_ISL_1009458, EPI_ISL_1009459, EPI_ISL_1009460, EPI_ISL_1009461, EPI_ISL_1009462, EPI_ISL_1009463, EPI_ISL_1009464, EPI_ISL_1009465, EPI_ISL_1009466, EPI_ISL_1009467, EPI_ISL_1009468, EPI_ISL_1009469, EPI_ISL_1009470, EPI_ISL_1009471, EPI_ISL_1009472, EPI_ISL_1009473, EPI_ISL_1009474, EPI_ISL_1009475, EPI_ISL_1009476, EPI_ISL_1009477, EPI_ISL_1009478, EPI_ISL_1009479, EPI_ISL_1009480, EPI_ISL_1009481, EPI_ISL_1009482, EPI_ISL_1009483, EPI_ISL_1009484, EPI_ISL_1009485, EPI_ISL_1009486, EPI_ISL_1009487, EPI_ISL_1009488, EPI_ISL_1009489, EPI_ISL_1009490, EPI_ISL_1009491, EPI_ISL_1009492, EPI_ISL_1009493, EPI_ISL_1009494, EPI_ISL_1009495, EPI_ISL_1009496, EPI_ISL_1009497, EPI_ISL_1009498, EPI_ISL_1009499, EPI_ISL_1009500, EPI_ISL_1009501, EPI_ISL_1009502, EPI_ISL_1009503, EPI_ISL_1009504, EPI_ISL_1009505, EPI_ISL_1009506, EPI_ISL_1009507, EPI_ISL_1009508, EPI_ISL_1009509, EPI_ISL_1009510, EPI_ISL_1009511, EPI_ISL_1009512, EPI_ISL_1009513, EPI_ISL_1009514, EPI_ISL_1009515, EPI_ISL_1009516, EPI_ISL_1009517, EPI_ISL_1009518, EPI_ISL_1009519, EPI_ISL_1009520, EPI_ISL_1009521, EPI_ISL_1009522, EPI_ISL_1009523, EPI_ISL_1009524, EPI_ISL_1009525, EPI_ISL_1009526, EPI_ISL_1009527, EPI_ISL_1009528, EPI_ISL_1009529, EPI_ISL_1009530, EPI_ISL_1009531, EPI_ISL_1009532, EPI_ISL_1009533, EPI_ISL_1009534, EPI_ISL_1009535, EPI_ISL_1009536, EPI_ISL_1009537, EPI_ISL_1009538, EPI_ISL_1009539, EPI_ISL_1009540, EPI_ISL_1009541, EPI_ISL_1009542, EPI_ISL_1009543, EPI_ISL_1009544, EPI_ISL_1009545, EPI_ISL_1009546, EPI_ISL_1009547, EPI_ISL_1009548, EPI_ISL_1009549, EPI_ISL_1009550, EPI_ISL_1009551, EPI_ISL_1009552, EPI_ISL_1009553, EPI_ISL_1009554, EPI_ISL_1009555, EPI_ISL_1009556, EPI_ISL_1009557, EPI_ISL_1009558, EPI_ISL_1009559, EPI_ISL_1009560, EPI_ISL_1009561, EPI_ISL_1009562, EPI_ISL_1009563, EPI_ISL_1009564, EPI_ISL_1009565, EPI_ISL_1009566, EPI_ISL_1009567, EPI_ISL_1009568, EPI_ISL_1009569, EPI_ISL_1009570, EPI_ISL_1009571, EPI_ISL_1009572, EPI_ISL_1009573, EPI_ISL_1009574, EPI_ISL_1009575, EPI_ISL_1009576, EPI_ISL_1009577, EPI_ISL_1009578, EPI_ISL_1009579, EPI_ISL_1009580, EPI_ISL_1009581, EPI_ISL_1009582, EPI_ISL_1009583, EPI_ISL_1009584, EPI_ISL_1009585, EPI_ISL_1009586, EPI_ISL_1009587, EPI_ISL_1009588, EPI_ISL_1009589, EPI_ISL_1009590, EPI_ISL_1009591, EPI_ISL_1009592, EPI_ISL_1009593, EPI_ISL_1009594, EPI_ISL_1009595, EPI_ISL_1009596, EPI_ISL_1009597, EPI_ISL_1009598, EPI_ISL_1009599, EPI_ISL_1009600, EPI_ISL_1009601, EPI_ISL_1009602, EPI_ISL_1009603, EPI_ISL_1009604, EPI_ISL_1009605, EPI_ISL_1009606, EPI_ISL_1009607, EPI_ISL_1009608, EPI_ISL_1009609, EPI_ISL_1009610, EPI_ISL_1009611, EPI_ISL_1009612, EPI_ISL_1009613, EPI_ISL_1009614, EPI_ISL_1009615, EPI_ISL_1009616 | | | |
| see above | Erie County Public Health (ECPHL) | University at Buffalo Genomics and Bioinformatics Core | Jonathan Bard, Natalie Lamb, Alyssa Pohlman, Brandon Marzullo, Amanda Boccolucci, Norma Nowak, Donald Yergeau, Jennifer Surtees |
| EPI_ISL_1015653, EPI_ISL_1015654, EPI_ISL_1015655, EPI_ISL_1015656, EPI_ISL_1015657, EPI_ISL_1015658, EPI_ISL_1015659, EPI_ISL_1015660, EPI_ISL_1015661, EPI_ISL_1015662, EPI_ISL_1015663, EPI_ISL_1015664, EPI_ISL_1015665, EPI_ISL_1015666, EPI_ISL_1015667, EPI_ISL_1015668, EPI_ISL_1015669, EPI_ISL_1015670, EPI_ISL_1015671, EPI_ISL_1015672, EPI_ISL_1015673, EPI_ISL_1015674, EPI_ISL_1015675, EPI_ISL_1015676, EPI_ISL_1015677, EPI_ISL_1015678, EPI_ISL_1015679, EPI_ISL_1015680, EPI_ISL_1015681, EPI_ISL_1015682, EPI_ISL_1015683, EPI_ISL_1015684, EPI_ISL_1015685, EPI_ISL_1015686, EPI_ISL_1015687, EPI_ISL_1015688, EPI_ISL_1015689, EPI_ISL_1015690, EPI_ISL_1015691, EPI_ISL_1015692, EPI_ISL_1015693, EPI_ISL_1015694, EPI_ISL_1015695, EPI_ISL_1015696, EPI_ISL_1015697, EPI_ISL_1015698, EPI_ISL_1015699, EPI_ISL_1015700, EPI_ISL_1015701, EPI_ISL_1015702, EPI_ISL_1015703, EPI_ISL_1015704, EPI_ISL_1015705, EPI_ISL_1015706, EPI_ISL_1015707, EPI_ISL_1015708, EPI_ISL_1015709, EPI_ISL_1015710, EPI_ISL_1015711, EPI_ISL_1015712 | | | |
| see above | Columbia University Irving Medical Center | Wadsworth Center, New York State Department of Health | Kirsten St. George, Daryl M. Lamson, Alexis Russel, Matthew Shudt, Melissa A Leisner, Jonathan Plitnick, Navjot Singh, John Kelly, Erasmus Schneider, Erica Lasek-Nesselquist |
| EPI_ISL_1016013, EPI_ISL_1016014, EPI_ISL_1016015, EPI_ISL_1016016, EPI_ISL_1016017, EPI_ISL_1016018, EPI_ISL_1016075, EPI_ISL_1016100 | NYU Langone Health | Departments of Pathology and Medicine, New York University School of Medicine | Adriana Heguy, Dacia Dimartino, Emily Guzman, Christian Marier, Peter Meyn, Sitharam Ramaswami, Gael Westby, Paul Zappile, Yutong Zhang, Paolo Cotzia, Guiqing Wang |
| EPI_ISL_1016101, EPI_ISL_1016102, EPI_ISL_1016103, EPI_ISL_1016104, EPI_ISL_1016105, EPI_ISL_1016106, EPI_ISL_1016107, EPI_ISL_1016108, EPI_ISL_1016109, EPI_ISL_1016110 | URMC LABS | Wadsworth Center, New York State Department of Health | Kirsten St. George, Daryl M. Lamson, Alexis Russel, Matthew Shudt, Melissa A Leisner, Jonathan Plitnick, Navjot Singh, John Kelly, Erasmus Schneider, Erica Lasek-Nesselquist |
| EPI_ISL_1016111, EPI_ISL_1016112, EPI_ISL_1016113, EPI_ISL_1016114, EPI_ISL_1016115, EPI_ISL_1016116, EPI_ISL_1016117 | Columbia University Irving Medical Center | Wadsworth Center, New York State Department of Health | Kirsten St. George, Daryl M. Lamson, Alexis Russel, Matthew Shudt, Melissa A Leisner, Jonathan Plitnick, Navjot Singh, John Kelly, Erasmus Schneider, Erica Lasek-Nesselquist |
| EPI_ISL_1016118, EPI_ISL_1016119, EPI_ISL_1016120, EPI_ISL_1016121, EPI_ISL_1016122, EPI_ISL_1016123, EPI_ISL_1016124, EPI_ISL_1016125, EPI_ISL_1016126 | URMC LABS | Wadsworth Center, New York State Department of Health | Kirsten St. George, Daryl M. Lamson, Alexis Russel, Matthew Shudt, Melissa A Leisner, Jonathan Plitnick, Navjot Singh, John Kelly, Erasmus Schneider, Erica Lasek-Nesselquist |
| EPI_ISL_1016127, EPI_ISL_1016128, EPI_ISL_1016129, EPI_ISL_1016130, EPI_ISL_1016131, EPI_ISL_1016132, EPI_ISL_1016133, EPI_ISL_1016134, EPI_ISL_1016135 | THE MARY IMOGENE BASSETT HOSPITAL | Wadsworth Center, New York State Department of Health | Kirsten St. George, Daryl M. Lamson, Alexis Russel, Matthew Shudt, Melissa A Leisner, Jonathan Plitnick, Navjot Singh, John Kelly, Erasmus Schneider, Erica Lasek-Nesselquist |
| EPI_ISL_1016136 | NORTHWELL HEALTH LABORATORIES | Wadsworth Center, New York State Department of Health | Kirsten St. George, Daryl M. Lamson, Alexis Russel, Matthew Shudt, Melissa A Leisner, Jonathan Plitnick, Navjot Singh, John Kelly, Erasmus Schneider, Erica Lasek-Nesselquist |
| EPI_ISL_1016137 | THE MARY IMOGENE BASSETT HOSPITAL | Wadsworth Center, New York State Department of Health | Kirsten St. George, Daryl M. Lamson, Alexis Russel, Matthew Shudt, Melissa A Leisner, Jonathan Plitnick, Navjot Singh, John Kelly, Erasmus Schneider, Erica Lasek-Nesselquist |
| EPI_ISL_1016138 | NORTHWELL HEALTH LABORATORIES | Wadsworth Center, New York State Department of Health | Kirsten St. George, Daryl M. Lamson, Alexis Russel, Matthew Shudt, Melissa A Leisner, Jonathan Plitnick, Navjot Singh, John Kelly, Erasmus Schneider, Erica Lasek-Nesselquist |
| EPI_ISL_1016139 | THE MARY IMOGENE BASSETT HOSPITAL | Wadsworth Center, New York State Department of Health | Kirsten St. George, Daryl M. Lamson, Alexis Russel, Matthew Shudt, Melissa A Leisner, Jonathan Plitnick, Navjot Singh, John Kelly, Erasmus Schneider, Erica Lasek-Nesselquist |
| EPI_ISL_1016140 | NORTHWELL HEALTH LABORATORIES | Wadsworth Center, New York State Department of Health | Kirsten St. George, Daryl M. Lamson, Alexis Russel, Matthew Shudt, Melissa A Leisner, Jonathan Plitnick, Navjot Singh, John Kelly, Erasmus Schneider, Erica Lasek-Nesselquist |
| EPI_ISL_1016141, EPI_ISL_1016142 | THE MARY IMOGENE BASSETT HOSPITAL | Wadsworth Center, New York State Department of Health | Kirsten St. George, Daryl M. Lamson, Alexis Russel, Matthew Shudt, Melissa A Leisner, Jonathan Plitnick, Navjot Singh, John Kelly, Erasmus Schneider, Erica Lasek-Nesselquist |
| EPI_ISL_1016143, EPI_ISL_1016144, EPI_ISL_1016145, EPI_ISL_1016146, EPI_ISL_1016147, EPI_ISL_1016148, EPI_ISL_1016149, EPI_ISL_1016150, EPI_ISL_1016151, EPI_ISL_1016152, EPI_ISL_1016153 | | | |
| see above | NORTHWELL HEALTH LABORATORIES | Wadsworth Center, New York State Department of Health | Kirsten St. George, Daryl M. Lamson, Alexis Russel, Matthew Shudt, Melissa A Leisner, Jonathan Plitnick, Navjot Singh, John Kelly, Erasmus Schneider, |

| | | | |
|---|---|---|---|
| | | | Erica Lasek-Nesselquist |
| EPI_ISL_1016154, EPI_ISL_1016155 | THE MARY IMOGENE BASSETT HOSPITAL | Wadsworth Center, New York State Department of Health | Kirsten St. George, Daryl M. Lamson, Alexis Russel, Matthew Shudt, Melissa A Leisner, Jonathan Plitnick, Navjot Singh, John Kelly, Erasmus Schneider, Erica Lasek-Nesselquist |
| EPI_ISL_1016156, EPI_ISL_1016157, EPI_ISL_1016158, EPI_ISL_1016159, EPI_ISL_1016160, EPI_ISL_1016161, EPI_ISL_1016162, EPI_ISL_1016163, EPI_ISL_1016164, EPI_ISL_1016165, EPI_ISL_1016166, EPI_ISL_1016167, EPI_ISL_1016168 | | | |
| see above | NORTHWELL HEALTH LABORATORIES | Wadsworth Center, New York State Department of Health | Kirsten St. George, Daryl M. Lamson, Alexis Russel, Matthew Shudt, Melissa A Leisner, Jonathan Plitnick, Navjot Singh, John Kelly, Erasmus Schneider, Erica Lasek-Nesselquist |
| EPI_ISL_1016169 | TEMPUS LABS INC | Wadsworth Center, New York State Department of Health | Kirsten St. George, Daryl M. Lamson, Alexis Russel, Matthew Shudt, Melissa A Leisner, Jonathan Plitnick, Navjot Singh, John Kelly, Erasmus Schneider, Erica Lasek-Nesselquist |
| EPI_ISL_1016170 | NYU Langone Health | Departments of Pathology and Medicine, New York University School of Medicine | Adriana Heguy, Dacia Dimartino, Emily Guzman, Christian Marier, Peter Meyn, Sitharam Ramaswami, Gael Westby, Paul Zappile, Yutong Zhang, Paolo Cotzia, Guiqing Wang |
| EPI_ISL_1016171, EPI_ISL_1016172, EPI_ISL_1016173, EPI_ISL_1016174, EPI_ISL_1016175, EPI_ISL_1016176, EPI_ISL_1016177, EPI_ISL_1016178, EPI_ISL_1016179, EPI_ISL_1016180, EPI_ISL_1016181, EPI_ISL_1016182, EPI_ISL_1016183, EPI_ISL_1016184, EPI_ISL_1016185, EPI_ISL_1016186, EPI_ISL_1016187, EPI_ISL_1016188, EPI_ISL_1016189, EPI_ISL_1016190, EPI_ISL_1016191, EPI_ISL_1016192, EPI_ISL_1016193, EPI_ISL_1016194, EPI_ISL_1016195, EPI_ISL_1016196, EPI_ISL_1016197, EPI_ISL_1016198, EPI_ISL_1016199, EPI_ISL_1016200, EPI_ISL_1016201, EPI_ISL_1016202, EPI_ISL_1016203, EPI_ISL_1016204, EPI_ISL_1016205, EPI_ISL_1016206, EPI_ISL_1016207, EPI_ISL_1016208, EPI_ISL_1016209, EPI_ISL_1016210, EPI_ISL_1016211, EPI_ISL_1016212, EPI_ISL_1016213, EPI_ISL_1016214, EPI_ISL_1016215, EPI_ISL_1016216, EPI_ISL_1016217, EPI_ISL_1016218, EPI_ISL_1016219 | | | |
| see above | MONTEFIORE MEDICAL CENTER LABORATORIES | Wadsworth Center, New York State Department of Health | Kirsten St. George, Daryl M. Lamson, Alexis Russel, Matthew Shudt, Melissa A Leisner, Jonathan Plitnick, Navjot Singh, John Kelly, Erasmus Schneider, Erica Lasek-Nesselquist |
| EPI_ISL_1016220, EPI_ISL_1016221, EPI_ISL_1016222, EPI_ISL_1016223, EPI_ISL_1016224, EPI_ISL_1016225, EPI_ISL_1016226, EPI_ISL_1016227, EPI_ISL_1016228, EPI_ISL_1016229, EPI_ISL_1016230, EPI_ISL_1016231, EPI_ISL_1016232, EPI_ISL_1016233, EPI_ISL_1016234, EPI_ISL_1016235, EPI_ISL_1016236, EPI_ISL_1016237, EPI_ISL_1016238, EPI_ISL_1016239, EPI_ISL_1016240, EPI_ISL_1016241, EPI_ISL_1016242, EPI_ISL_1016243, EPI_ISL_1016244, EPI_ISL_1016245, EPI_ISL_1016246, EPI_ISL_1016247, EPI_ISL_1016248 | | | |
| see above | New York Presbyterian Hospital | Wadsworth Center, New York State Department of Health | Kirsten St. George, Daryl M. Lamson, Alexis Russel, Matthew Shudt, Melissa A Leisner, Jonathan Plitnick, Navjot Singh, John Kelly, Erasmus Schneider, Erica Lasek-Nesselquist |
| EPI_ISL_1016249, EPI_ISL_1016250, EPI_ISL_1016251, EPI_ISL_1016252, EPI_ISL_1016253, EPI_ISL_1016254, EPI_ISL_1016255, EPI_ISL_1016256 | TEMPUS LABS INC | Wadsworth Center, New York State Department of Health | Kirsten St. George, Daryl M. Lamson, Alexis Russel, Matthew Shudt, Melissa A Leisner, Jonathan Plitnick, Navjot Singh, John Kelly, Erasmus Schneider, Erica Lasek-Nesselquist |
| EPI_ISL_1016257 | THE MARY IMOGENE BASSETT HOSPITAL | Wadsworth Center, New York State Department of Health | Kirsten St. George, Daryl M. Lamson, Alexis Russel, Matthew Shudt, Melissa A Leisner, Jonathan Plitnick, Navjot Singh, John Kelly, Erasmus Schneider, Erica Lasek-Nesselquist |
| EPI_ISL_1016258, EPI_ISL_1016259 | New York Presbyterian Hospital | Wadsworth Center, New York State Department of Health | Kirsten St. George, Daryl M. Lamson, Alexis Russel, Matthew Shudt, Melissa A Leisner, Jonathan Plitnick, Navjot Singh, John Kelly, Erasmus Schneider, Erica Lasek-Nesselquist |
| EPI_ISL_1016260, EPI_ISL_1016261 | Columbia University Irving Medical Center | Wadsworth Center, New York State Department of Health | Kirsten St. George, Daryl M. Lamson, Alexis Russel, Matthew Shudt, Melissa A Leisner, Jonathan Plitnick, Navjot Singh, John Kelly, Erasmus Schneider, Erica Lasek-Nesselquist |
| EPI_ISL_1016262, EPI_ISL_1016263, EPI_ISL_1016264, EPI_ISL_1016265, EPI_ISL_1016266 | New York Presbyterian Hospital | Wadsworth Center, New York State Department of Health | Kirsten St. George, Daryl M. Lamson, Alexis Russel, Matthew Shudt, Melissa A Leisner, Jonathan Plitnick, Navjot Singh, John Kelly, Erasmus Schneider, Erica Lasek-Nesselquist |
| EPI_ISL_1016267, EPI_ISL_1016268 | Columbia University Irving Medical Center | Wadsworth Center, New York State Department of Health | Kirsten St. George, Daryl M. Lamson, Alexis Russel, Matthew Shudt, Melissa A Leisner, Jonathan Plitnick, Navjot Singh, John Kelly, Erasmus Schneider, Erica Lasek-Nesselquist |
| EPI_ISL_1016269 | New York Presbyterian Hospital | Wadsworth Center, New York State Department of Health | Kirsten St. George, Daryl M. Lamson, Alexis Russel, Matthew Shudt, Melissa A Leisner, Jonathan Plitnick, Navjot Singh, John Kelly, Erasmus Schneider, Erica Lasek-Nesselquist |
| EPI_ISL_1016270 | Columbia University Irving Medical Center | Wadsworth Center, New York State Department of Health | Kirsten St. George, Daryl M. Lamson, Alexis Russel, Matthew Shudt, Melissa A Leisner, Jonathan Plitnick, Navjot Singh, John Kelly, Erasmus Schneider, Erica Lasek-Nesselquist |
| EPI_ISL_1016271, EPI_ISL_1016272, EPI_ISL_1016273, EPI_ISL_1016274, EPI_ISL_1016275, EPI_ISL_1016276, EPI_ISL_1016277, EPI_ISL_1016278, EPI_ISL_1016279, EPI_ISL_1016280 | New York Presbyterian Hospital | Wadsworth Center, New York State Department of Health | Kirsten St. George, Daryl M. Lamson, Alexis Russel, Matthew Shudt, Melissa A Leisner, Jonathan Plitnick, Navjot Singh, John Kelly, Erasmus Schneider, Erica Lasek-Nesselquist |
| EPI_ISL_1016281, EPI_ISL_1016282, EPI_ISL_1016283, EPI_ISL_1016284 | Columbia University Irving Medical Center | Wadsworth Center, New York State Department of Health | Kirsten St. George, Daryl M. Lamson, Alexis Russel, Matthew Shudt, Melissa A Leisner, Jonathan Plitnick, Navjot Singh, John Kelly, Erasmus Schneider, Erica Lasek-Nesselquist |
| EPI_ISL_1016285, EPI_ISL_1016286 | SUNY UPSTATE MEDICAL UNIVERSITY | Wadsworth Center, New York State Department of Health | Kirsten St. George, Daryl M. Lamson, Alexis Russel, Matthew Shudt, Melissa A Leisner, Jonathan Plitnick, Navjot Singh, John Kelly, Erasmus Schneider, Erica Lasek-Nesselquist |
| EPI_ISL_1016287, EPI_ISL_1016288, EPI_ISL_1016289, EPI_ISL_1016290, EPI_ISL_1016291, EPI_ISL_1016292, EPI_ISL_1016293, EPI_ISL_1016294, EPI_ISL_1016295, EPI_ISL_1016296, EPI_ISL_1016297 | | | |
| see above | Columbia University Irving Medical Center | Wadsworth Center, New York State Department of Health | Kirsten St. George, Daryl M. Lamson, Alexis Russel, Matthew Shudt, Melissa A Leisner, Jonathan Plitnick, Navjot Singh, John Kelly, Erasmus Schneider, Erica Lasek-Nesselquist |
| EPI_ISL_1016298, EPI_ISL_1016299, EPI_ISL_1016300, EPI_ISL_1016301, EPI_ISL_1016302, EPI_ISL_1016303 | SUNY UPSTATE MEDICAL UNIVERSITY | Wadsworth Center, New York State Department of Health | Kirsten St. George, Daryl M. Lamson, Alexis Russel, Matthew Shudt, Melissa A Leisner, Jonathan Plitnick, Navjot Singh, John Kelly, Erasmus Schneider, Erica Lasek-Nesselquist |
| EPI_ISL_1016304, EPI_ISL_1016305, EPI_ISL_1016306, EPI_ISL_1016307, EPI_ISL_1016308, EPI_ISL_1016309, EPI_ISL_1016310 | New York Presbyterian Hospital | Wadsworth Center, New York State Department of Health | Kirsten St. George, Daryl M. Lamson, Alexis Russel, Matthew Shudt, Melissa A Leisner, Jonathan Plitnick, Navjot Singh, John Kelly, Erasmus Schneider, Erica Lasek-Nesselquist |
| EPI_ISL_1016311, EPI_ISL_1016312, EPI_ISL_1016313, EPI_ISL_1016314, EPI_ISL_1016315, EPI_ISL_1016316, EPI_ISL_1016317, EPI_ISL_1016318, EPI_ISL_1016319, EPI_ISL_1016320, EPI_ISL_1016321, EPI_ISL_1016322 | | | |
| see above | SUNY UPSTATE MEDICAL UNIVERSITY | Wadsworth Center, New York State Department of Health | Kirsten St. George, Daryl M. Lamson, Alexis Russel, Matthew Shudt, Melissa A Leisner, Jonathan Plitnick, Navjot Singh, John Kelly, Erasmus Schneider, Erica Lasek-Nesselquist |
| EPI_ISL_1016323, EPI_ISL_1016324, EPI_ISL_1016325, EPI_ISL_1016326 | Columbia University Irving Medical Center | Wadsworth Center, New York State Department of Health | Kirsten St. George, Daryl M. Lamson, Alexis Russel, Matthew Shudt, Melissa A Leisner, Jonathan Plitnick, Navjot Singh, John Kelly, Erasmus Schneider, Erica Lasek-Nesselquist |
| EPI_ISL_1016327 | SUNY UPSTATE MEDICAL UNIVERSITY | Wadsworth Center, New York State Department of Health | Kirsten St. George, Daryl M. Lamson, Alexis Russel, Matthew Shudt, Melissa A Leisner, Jonathan Plitnick, Navjot Singh, John Kelly, Erasmus Schneider, Erica Lasek-Nesselquist |
| EPI_ISL_1016328, EPI_ISL_1016329, EPI_ISL_1016330, EPI_ISL_1016331 | Columbia University Irving Medical Center | Wadsworth Center, New York State Department of Health | Kirsten St. George, Daryl M. Lamson, Alexis Russel, Matthew Shudt, Melissa A Leisner, Jonathan Plitnick, Navjot Singh, John Kelly, Erasmus Schneider, Erica Lasek-Nesselquist |
| EPI_ISL_1016332 | WESTCHESTER MEDICAL CENTER | Wadsworth Center, New York State Department of Health | Kirsten St. George, Daryl M. Lamson, Alexis Russel, Matthew Shudt, Melissa A Leisner, Jonathan Plitnick, Navjot Singh, John Kelly, Erasmus Schneider, Erica Lasek-Nesselquist |
| EPI_ISL_1016333, EPI_ISL_1016334 | URMC LABS | Wadsworth Center, New York State Department of Health | Kirsten St. George, Daryl M. Lamson, Alexis Russel, Matthew Shudt, Melissa A Leisner, Jonathan Plitnick, Navjot Singh, John Kelly, Erasmus Schneider, Erica Lasek-Nesselquist |
| EPI_ISL_1016335, EPI_ISL_1016336 | New York Presbyterian Hospital | Wadsworth Center, New York State Department of Health | Kirsten St. George, Daryl M. Lamson, Alexis Russel, Matthew Shudt, Melissa A Leisner, Jonathan Plitnick, Navjot Singh, John Kelly, Erasmus Schneider, Erica Lasek-Nesselquist |

| | | | |
|---|---|---|---|
| EPI_ISL_1016337, EPI_ISL_1016338 | MEMORIAL SLOAN KETTERING CANCER CENTER | Wadsworth Center, New York State Department of Health | Kirsten St. George, Daryl M. Lamson, Alexis Russel, Matthew Shudt, Melissa A Leisner, Jonathan Plitnick, Navjot Singh, John Kelly, Erasmus Schneider, Erica Lasek-Nesselquist |
| EPI_ISL_1016339 | URMC LABS | Wadsworth Center, New York State Department of Health | Kirsten St. George, Daryl M. Lamson, Alexis Russel, Matthew Shudt, Melissa A Leisner, Jonathan Plitnick, Navjot Singh, John Kelly, Erasmus Schneider, Erica Lasek-Nesselquist |
| EPI_ISL_1016340 | WESTCHESTER MEDICAL CENTER | Wadsworth Center, New York State Department of Health | Kirsten St. George, Daryl M. Lamson, Alexis Russel, Matthew Shudt, Melissa A Leisner, Jonathan Plitnick, Navjot Singh, John Kelly, Erasmus Schneider, Erica Lasek-Nesselquist |
| EPI_ISL_1016341 | New York Presbyterian Hospital | Wadsworth Center, New York State Department of Health | Kirsten St. George, Daryl M. Lamson, Alexis Russel, Matthew Shudt, Melissa A Leisner, Jonathan Plitnick, Navjot Singh, John Kelly, Erasmus Schneider, Erica Lasek-Nesselquist |
| EPI_ISL_1016342, EPI_ISL_1016343 | MEMORIAL SLOAN KETTERING CANCER CENTER | Wadsworth Center, New York State Department of Health | Kirsten St. George, Daryl M. Lamson, Alexis Russel, Matthew Shudt, Melissa A Leisner, Jonathan Plitnick, Navjot Singh, John Kelly, Erasmus Schneider, Erica Lasek-Nesselquist |
| EPI_ISL_1016344, EPI_ISL_1016345, EPI_ISL_1016346 | WESTCHESTER MEDICAL CENTER | Wadsworth Center, New York State Department of Health | Kirsten St. George, Daryl M. Lamson, Alexis Russel, Matthew Shudt, Melissa A Leisner, Jonathan Plitnick, Navjot Singh, John Kelly, Erasmus Schneider, Erica Lasek-Nesselquist |
| EPI_ISL_1016347, EPI_ISL_1016348, EPI_ISL_1016349, EPI_ISL_1016350, EPI_ISL_1016351, EPI_ISL_1016352, EPI_ISL_1016353, EPI_ISL_1016354, EPI_ISL_1016355, EPI_ISL_1016356, EPI_ISL_1016357, EPI_ISL_1016358, EPI_ISL_1016359, EPI_ISL_1016360, EPI_ISL_1016361, EPI_ISL_1016362, EPI_ISL_1016363, EPI_ISL_1016364 | | | |
| see above | URMC LABS | Wadsworth Center, New York State Department of Health | Kirsten St. George, Daryl M. Lamson, Alexis Russel, Matthew Shudt, Melissa A Leisner, Jonathan Plitnick, Navjot Singh, John Kelly, Erasmus Schneider, Erica Lasek-Nesselquist |
| EPI_ISL_1016365 | WESTCHESTER MEDICAL CENTER | Wadsworth Center, New York State Department of Health | Kirsten St. George, Daryl M. Lamson, Alexis Russel, Matthew Shudt, Melissa A Leisner, Jonathan Plitnick, Navjot Singh, John Kelly, Erasmus Schneider, Erica Lasek-Nesselquist |
| EPI_ISL_1016366, EPI_ISL_1016367, EPI_ISL_1016368, EPI_ISL_1016369, EPI_ISL_1016370, EPI_ISL_1016371, EPI_ISL_1016372 | New York Presbyterian Hospital | Wadsworth Center, New York State Department of Health | Kirsten St. George, Daryl M. Lamson, Alexis Russel, Matthew Shudt, Melissa A Leisner, Jonathan Plitnick, Navjot Singh, John Kelly, Erasmus Schneider, Erica Lasek-Nesselquist |
| EPI_ISL_1016373, EPI_ISL_1016374, EPI_ISL_1016375, EPI_ISL_1016376, EPI_ISL_1016377, EPI_ISL_1016378, EPI_ISL_1016379, EPI_ISL_1016380 | WESTCHESTER MEDICAL CENTER | Wadsworth Center, New York State Department of Health | Kirsten St. George, Daryl M. Lamson, Alexis Russel, Matthew Shudt, Melissa A Leisner, Jonathan Plitnick, Navjot Singh, John Kelly, Erasmus Schneider, Erica Lasek-Nesselquist |
| EPI_ISL_1016381, EPI_ISL_1016382, EPI_ISL_1016383, EPI_ISL_1016384, EPI_ISL_1016385, EPI_ISL_1016386, EPI_ISL_1016387, EPI_ISL_1016388, EPI_ISL_1016389, EPI_ISL_1016390, EPI_ISL_1016391, EPI_ISL_1016392, EPI_ISL_1016393, EPI_ISL_1016394, EPI_ISL_1016395, EPI_ISL_1016396, EPI_ISL_1016397, EPI_ISL_1016398, EPI_ISL_1016399, EPI_ISL_1016400, EPI_ISL_1016401 | | | |
| see above | New York Presbyterian Hospital | Wadsworth Center, New York State Department of Health | Kirsten St. George, Daryl M. Lamson, Alexis Russel, Matthew Shudt, Melissa A Leisner, Jonathan Plitnick, Navjot Singh, John Kelly, Erasmus Schneider, Erica Lasek-Nesselquist |
| EPI_ISL_1016402, EPI_ISL_1016403, EPI_ISL_1016404, EPI_ISL_1016405, EPI_ISL_1016406, EPI_ISL_1016407, EPI_ISL_1016408, EPI_ISL_1016409, EPI_ISL_1016410, EPI_ISL_1016411 | MEMORIAL SLOAN KETTERING CANCER CENTER | Wadsworth Center, New York State Department of Health | Kirsten St. George, Daryl M. Lamson, Alexis Russel, Matthew Shudt, Melissa A Leisner, Jonathan Plitnick, Navjot Singh, John Kelly, Erasmus Schneider, Erica Lasek-Nesselquist |
| EPI_ISL_1016412 | STONY BROOK UNIVERSITY HOSPITAL | Wadsworth Center, New York State Department of Health | Kirsten St. George, Daryl M. Lamson, Alexis Russel, Matthew Shudt, Melissa A Leisner, Jonathan Plitnick, Navjot Singh, John Kelly, Erasmus Schneider, Erica Lasek-Nesselquist |
| EPI_ISL_1016413 | NORTHWELL HEALTH LABORATORIES | Wadsworth Center, New York State Department of Health | Kirsten St. George, Daryl M. Lamson, Alexis Russel, Matthew Shudt, Melissa A Leisner, Jonathan Plitnick, Navjot Singh, John Kelly, Erasmus Schneider, Erica Lasek-Nesselquist |
| EPI_ISL_1016414, EPI_ISL_1016415 | URMC LABS | Wadsworth Center, New York State Department of Health | Kirsten St. George, Daryl M. Lamson, Alexis Russel, Matthew Shudt, Melissa A Leisner, Jonathan Plitnick, Navjot Singh, John Kelly, Erasmus Schneider, Erica Lasek-Nesselquist |
| EPI_ISL_1016416 | STONY BROOK UNIVERSITY HOSPITAL | Wadsworth Center, New York State Department of Health | Kirsten St. George, Daryl M. Lamson, Alexis Russel, Matthew Shudt, Melissa A Leisner, Jonathan Plitnick, Navjot Singh, John Kelly, Erasmus Schneider, Erica Lasek-Nesselquist |
| EPI_ISL_1016417 | ALBANY MEDICAL CENTER HOSPITAL CLINICAL LABORATORIES | Wadsworth Center, New York State Department of Health | Kirsten St. George, Daryl M. Lamson, Alexis Russel, Matthew Shudt, Melissa A Leisner, Jonathan Plitnick, Navjot Singh, John Kelly, Erasmus Schneider, Erica Lasek-Nesselquist |
| EPI_ISL_1016418, EPI_ISL_1016419, EPI_ISL_1016420, EPI_ISL_1016421, EPI_ISL_1016422, EPI_ISL_1016423, EPI_ISL_1016424, EPI_ISL_1016425, EPI_ISL_1016426, EPI_ISL_1016427, EPI_ISL_1016428, EPI_ISL_1016429, EPI_ISL_1016430, EPI_ISL_1016431, EPI_ISL_1016432, EPI_ISL_1016433, EPI_ISL_1016434, EPI_ISL_1016435, EPI_ISL_1016436, EPI_ISL_1016437, EPI_ISL_1016438, EPI_ISL_1016439, EPI_ISL_1016440, EPI_ISL_1016441, EPI_ISL_1016442, EPI_ISL_1016443, EPI_ISL_1016444, EPI_ISL_1016445, EPI_ISL_1016446, EPI_ISL_1016447, EPI_ISL_1016448, EPI_ISL_1016449, EPI_ISL_1016450, EPI_ISL_1016451, EPI_ISL_1016452, EPI_ISL_1016453, EPI_ISL_1016454, EPI_ISL_1016455, EPI_ISL_1016456, EPI_ISL_1016457 | | | |
| see above | URMC LABS | Wadsworth Center, New York State Department of Health | Kirsten St. George, Daryl M. Lamson, Alexis Russel, Matthew Shudt, Melissa A Leisner, Jonathan Plitnick, Navjot Singh, John Kelly, Erasmus Schneider, Erica Lasek-Nesselquist |
| EPI_ISL_1016458 | NORTHWELL HEALTH LABORATORIES | Wadsworth Center, New York State Department of Health | Kirsten St. George, Daryl M. Lamson, Alexis Russel, Matthew Shudt, Melissa A Leisner, Jonathan Plitnick, Navjot Singh, John Kelly, Erasmus Schneider, Erica Lasek-Nesselquist |
| EPI_ISL_1016459, EPI_ISL_1016460, EPI_ISL_1016461, EPI_ISL_1016462, EPI_ISL_1016463, EPI_ISL_1016464, EPI_ISL_1016465, EPI_ISL_1016466, EPI_ISL_1016467 | STONY BROOK UNIVERSITY HOSPITAL | Wadsworth Center, New York State Department of Health | Kirsten St. George, Daryl M. Lamson, Alexis Russel, Matthew Shudt, Melissa A Leisner, Jonathan Plitnick, Navjot Singh, John Kelly, Erasmus Schneider, Erica Lasek-Nesselquist |
| EPI_ISL_1016468, EPI_ISL_1016469, EPI_ISL_1016470, EPI_ISL_1016471, EPI_ISL_1016472, EPI_ISL_1016473, EPI_ISL_1016474, EPI_ISL_1016475 | ALBANY MEDICAL CENTER HOSPITAL CLINICAL LABORATORIES | Wadsworth Center, New York State Department of Health | Kirsten St. George, Daryl M. Lamson, Alexis Russel, Matthew Shudt, Melissa A Leisner, Jonathan Plitnick, Navjot Singh, John Kelly, Erasmus Schneider, Erica Lasek-Nesselquist |
| EPI_ISL_1016476, EPI_ISL_1016477, EPI_ISL_1016478 | STONY BROOK UNIVERSITY HOSPITAL | Wadsworth Center, New York State Department of Health | Kirsten St. George, Daryl M. Lamson, Alexis Russel, Matthew Shudt, Melissa A Leisner, Jonathan Plitnick, Navjot Singh, John Kelly, Erasmus Schneider, Erica Lasek-Nesselquist |
| EPI_ISL_1016565, EPI_ISL_1016602, EPI_ISL_1016643 | Helix/Illumina | Respiratory Viruses Branch, Division of Viral Diseases, Centers for Disease Control and Prevention | Peter W. Cook, Dakota Howard, Dhwani Batra, Ben L. Rambo-Martin, Eileen de Feo, Jan Antico, Christine Tran, Matthew Tolentino, Shannon Wickline, Kim Gietzen, Brad Sickler, Jingtao Liu, Eric Allen, Phil Febbo, Summer Galloway, Nicole L. Washington, Simon White, Geraint Levan, Kelly Schiabor Barrett, Elizabeth Cirulli, Alexandre Bolze, Ary Ascencio, Charlotte Rivera-Garcia, Ryan Cho, Jason Nguyen, Sherry Wang, Jimmy Ramirez, Tyler Cassens, Efren Sandoval, Magnus Isaksson, William Lee, David Becker, Marc Laurent, James Lu, Clinton R. Paden, Suxiang Tong, Duncan MacCannell |
| EPI_ISL_1017489, EPI_ISL_1017491, EPI_ISL_1017500, EPI_ISL_1017502, EPI_ISL_1017514, EPI_ISL_1017525, EPI_ISL_1017526, EPI_ISL_1017536, EPI_ISL_1017541, EPI_ISL_1017547, EPI_ISL_1017566, EPI_ISL_1017568, EPI_ISL_1017571, EPI_ISL_1017575, EPI_ISL_1017576, EPI_ISL_1017577, EPI_ISL_1017582, EPI_ISL_1017583, EPI_ISL_1017584 | | | |
| see above | Murphy Medical Associates | Grubaugh Lab - Yale School of Public Health | Mary Petrone, Joseph Fauver, Caleb Neal, Steven Murphy, Chantal Vogels, Mallery Breban, Annie Watkins, Tara Alpert, Nathan Grubaugh |
| EPI_ISL_1017969, EPI_ISL_1017970 | DOHMH PHL | New York City Public Health Laboratory | Jade Wang, et al. |
| EPI_ISL_1017971, EPI_ISL_1017972, EPI_ISL_1017973 | DOHMH Riverside | New York City Public Health Laboratory | Jade Wang, et al. |

| | | | |
|---|---|---|---|
| EPI_ISL_1017974 | DOHMH Morrisania | New York City Public Health Laboratory | Jade Wang, et al. |
| EPI_ISL_1017975 | DOHMH Corona | New York City Public Health Laboratory | Jade Wang, et al. |
| EPI_ISL_1017976, EPI_ISL_1017977 | DOHMH Jamaica | New York City Public Health Laboratory | Jade Wang, et al. |
| EPI_ISL_1017978 | DOHMH Chelsea | New York City Public Health Laboratory | Jade Wang, et al. |
| EPI_ISL_1017979 | DOHMH PHL | New York City Public Health Laboratory | Jade Wang, et al. |
| EPI_ISL_1017980, EPI_ISL_1017981 | DOHMH Morrisania | New York City Public Health Laboratory | Jade Wang, et al. |
| EPI_ISL_1017982, EPI_ISL_1017983, EPI_ISL_1017984, EPI_ISL_1017985 | DOHMH Central Harlem | New York City Public Health Laboratory | Jade Wang, et al. |
| EPI_ISL_1017986, EPI_ISL_1017987, EPI_ISL_1017988 | DOHMH Corona | New York City Public Health Laboratory | Jade Wang, et al. |
| EPI_ISL_1017989, EPI_ISL_1017990 | Department of Homeless Services | New York City Public Health Laboratory | Jade Wang, et al. |
| EPI_ISL_1017991 | DOHMH PHL | New York City Public Health Laboratory | Jade Wang, et al. |
| EPI_ISL_1017992, EPI_ISL_1017993, EPI_ISL_1017994 | DOHMH Jamaica | New York City Public Health Laboratory | Jade Wang, et al. |
| EPI_ISL_1017995, EPI_ISL_1017996, EPI_ISL_1017997, EPI_ISL_1017998, EPI_ISL_1017999 | DOHMH Corona | New York City Public Health Laboratory | Jade Wang, et al. |
| EPI_ISL_1018000 | DOHMH Morrisania | New York City Public Health Laboratory | Jade Wang, et al. |
| EPI_ISL_1018001 | DOHMH Central Harlem | New York City Public Health Laboratory | Jade Wang, et al. |
| EPI_ISL_1018002, EPI_ISL_1018003, EPI_ISL_1018004 | DOHMH PHL | New York City Public Health Laboratory | Jade Wang, et al. |
| EPI_ISL_1018005 | DOHMH Corona | New York City Public Health Laboratory | Jade Wang, et al. |
| EPI_ISL_1018006, EPI_ISL_1018007, EPI_ISL_1018008, EPI_ISL_1018009, EPI_ISL_1018010 | DOHMH Morrisania | New York City Public Health Laboratory | Jade Wang, et al. |
| EPI_ISL_1018011 | DOHMH Chelsea | New York City Public Health Laboratory | Jade Wang, et al. |
| EPI_ISL_1018012, EPI_ISL_1018013, EPI_ISL_1018014, EPI_ISL_1018015, EPI_ISL_1018016, EPI_ISL_1018017, EPI_ISL_1018018, EPI_ISL_1018019, EPI_ISL_1018020 | DOHMH Jamaica | New York City Public Health Laboratory | Jade Wang, et al. |
| EPI_ISL_1018021 | DOHMH Crown Heights | New York City Public Health Laboratory | Jade Wang, et al. |
| EPI_ISL_1018022, EPI_ISL_1018023 | DOHMH Jamaica | New York City Public Health Laboratory | Jade Wang, et al. |
| EPI_ISL_1018024 | DOHMH Crown Heights | New York City Public Health Laboratory | Jade Wang, et al. |
| EPI_ISL_1018025, EPI_ISL_1018026 | DOHMH Morrisania | New York City Public Health Laboratory | Jade Wang, et al. |
| EPI_ISL_1018027, EPI_ISL_1018028, EPI_ISL_1018029, EPI_ISL_1018030, EPI_ISL_1018031 | DOHMH Corona | New York City Public Health Laboratory | Jade Wang, et al. |
| EPI_ISL_1018032, EPI_ISL_1018033, EPI_ISL_1018034, EPI_ISL_1018035, EPI_ISL_1018036, EPI_ISL_1018037, EPI_ISL_1018038, EPI_ISL_1018039, EPI_ISL_1018040, EPI_ISL_1018041, EPI_ISL_1018042, EPI_ISL_1018043, EPI_ISL_1018044, EPI_ISL_1018045, EPI_ISL_1018046, EPI_ISL_1018047, EPI_ISL_1018048, EPI_ISL_1018049 | | | |
| see above | OCME Office Of Chief Medical Examiner | New York City Public Health Laboratory | Jade Wang, et al. |
| EPI_ISL_1020487, EPI_ISL_1020488, EPI_ISL_1020489, EPI_ISL_1020490, EPI_ISL_1020491, EPI_ISL_1020492, EPI_ISL_1020493, EPI_ISL_1020494, EPI_ISL_1020495, EPI_ISL_1020496, EPI_ISL_1020497, EPI_ISL_1020498, EPI_ISL_1020499, EPI_ISL_1020500, EPI_ISL_1020501, EPI_ISL_1020502, EPI_ISL_1020503, EPI_ISL_1020504, EPI_ISL_1020505, EPI_ISL_1020506, EPI_ISL_1020507, EPI_ISL_1020508, EPI_ISL_1020509, EPI_ISL_1020510, EPI_ISL_1020511, EPI_ISL_1020512, EPI_ISL_1020513, EPI_ISL_1020514, EPI_ISL_1020515, EPI_ISL_1020516, EPI_ISL_1020517, EPI_ISL_1020518, EPI_ISL_1020519, EPI_ISL_1020520, EPI_ISL_1020521, EPI_ISL_1020522, EPI_ISL_1020523, EPI_ISL_1020524, EPI_ISL_1020525, EPI_ISL_1020526, EPI_ISL_1020527, EPI_ISL_1020528, EPI_ISL_1020529, EPI_ISL_1020530, EPI_ISL_1020531, EPI_ISL_1020532, EPI_ISL_1020533, EPI_ISL_1020534, EPI_ISL_1020535, EPI_ISL_1020536, EPI_ISL_1020537, EPI_ISL_1020538, EPI_ISL_1020539, EPI_ISL_1020540, EPI_ISL_1020541, EPI_ISL_1020542, EPI_ISL_1020543, EPI_ISL_1020544, EPI_ISL_1020545, EPI_ISL_1020546, EPI_ISL_1020547, EPI_ISL_1020548, EPI_ISL_1020549, EPI_ISL_1020550 | | | |
| see above | Columbia University Irving Medical Center | Wadsworth Center, New York State Department of Health | Kirsten St. George, Daryl M. Lamson, Alexis Russel, Matthew Shudt, Melissa A Leisner, Jonathan Plitnick, Navjot Singh, John Kelly, Erasmus Schneider, Erica Lasek-Nesselquist |
| EPI_ISL_1020593, EPI_ISL_1020774, EPI_ISL_1020778, EPI_ISL_1020810, EPI_ISL_1020839, EPI_ISL_1020872, EPI_ISL_1020911, EPI_ISL_1020934, EPI_ISL_1020989, EPI_ISL_1020994, EPI_ISL_1021030, EPI_ISL_1021033, EPI_ISL_1021041, EPI_ISL_1021054, EPI_ISL_1021057, EPI_ISL_1021066, EPI_ISL_1021081, EPI_ISL_1021094, EPI_ISL_1021098, EPI_ISL_1021121, EPI_ISL_1021123, EPI_ISL_1021125, EPI_ISL_1021128, EPI_ISL_1021135, EPI_ISL_1021136, EPI_ISL_1021168, EPI_ISL_1021181, EPI_ISL_1021210, EPI_ISL_1021229, EPI_ISL_1021257, EPI_ISL_1021265, EPI_ISL_1021359, EPI_ISL_1021360, EPI_ISL_1021361, EPI_ISL_1021368, EPI_ISL_1021383, EPI_ISL_1021384, EPI_ISL_1021394, EPI_ISL_1021413, EPI_ISL_1021428, EPI_ISL_1021437, EPI_ISL_1021501, EPI_ISL_1021507, EPI_ISL_1021508, EPI_ISL_1021540, EPI_ISL_1021542, EPI_ISL_1021561, EPI_ISL_1021562, EPI_ISL_1021592, EPI_ISL_1021630, EPI_ISL_1021655, EPI_ISL_1021669, EPI_ISL_1021722, EPI_ISL_1021735, EPI_ISL_1021736, EPI_ISL_1021750, EPI_ISL_1021751, EPI_ISL_1021761, EPI_ISL_1021803, EPI_ISL_1021819, EPI_ISL_1021824, EPI_ISL_1021832, EPI_ISL_1021857, EPI_ISL_1021865, EPI_ISL_1021868, EPI_ISL_1021928, EPI_ISL_1021963, EPI_ISL_1021969, EPI_ISL_1021970, EPI_ISL_1021981, EPI_ISL_1022042, EPI_ISL_1022048, EPI_ISL_1022085, EPI_ISL_1022107, EPI_ISL_1022108, EPI_ISL_1022130, EPI_ISL_1022131, EPI_ISL_1022132, EPI_ISL_1022133, EPI_ISL_1022167, EPI_ISL_1022176, EPI_ISL_1022205, EPI_ISL_1022209, EPI_ISL_1022214, EPI_ISL_1022227, EPI_ISL_1022261, EPI_ISL_1022269, EPI_ISL_1022288, EPI_ISL_1022311, EPI_ISL_1022325, EPI_ISL_1022336, EPI_ISL_1022371, EPI_ISL_1022372, EPI_ISL_1022373, EPI_ISL_1022374, EPI_ISL_1022395, EPI_ISL_1022396, EPI_ISL_1026002, EPI_ISL_1026006, EPI_ISL_1026023, EPI_ISL_1026103, EPI_ISL_1026107, EPI_ISL_1026212, EPI_ISL_1026215, EPI_ISL_1026241, EPI_ISL_1026300, EPI_ISL_1026306, EPI_ISL_1026346, EPI_ISL_1026443, EPI_ISL_1026456, EPI_ISL_1026490, EPI_ISL_1026532, EPI_ISL_1026540, EPI_ISL_1026613, EPI_ISL_1026630, EPI_ISL_1026672, EPI_ISL_1026695, EPI_ISL_1026726, EPI_ISL_1026732, EPI_ISL_1026854, EPI_ISL_1027684, EPI_ISL_1027727, EPI_ISL_1027736, EPI_ISL_1027771, EPI_ISL_1027772, EPI_ISL_1027774, EPI_ISL_1027777, EPI_ISL_1027793, EPI_ISL_1027812, EPI_ISL_1027837, EPI_ISL_1027866, EPI_ISL_1027919, EPI_ISL_1027979, EPI_ISL_1027990, EPI_ISL_1028017, EPI_ISL_1028026, EPI_ISL_1028037, EPI_ISL_1028113, EPI_ISL_1028119, EPI_ISL_1028181, EPI_ISL_1028310, EPI_ISL_1028315, EPI_ISL_1028330, EPI_ISL_1028384, EPI_ISL_1028401, EPI_ISL_1028439, EPI_ISL_1028460, EPI_ISL_1028469, EPI_ISL_1028524, EPI_ISL_1028539, EPI_ISL_1028544, EPI_ISL_1028547, EPI_ISL_1028558, EPI_ISL_1028572, EPI_ISL_1028615, EPI_ISL_1028632, EPI_ISL_1028640, EPI_ISL_1028645, EPI_ISL_1028656, EPI_ISL_1028687, EPI_ISL_1028690, EPI_ISL_1028716, EPI_ISL_1028743, EPI_ISL_1028773, EPI_ISL_1028812, EPI_ISL_1028847, EPI_ISL_1028870, EPI_ISL_1028936, EPI_ISL_1028951, EPI_ISL_1028954, EPI_ISL_1029021, EPI_ISL_1029031, EPI_ISL_1029056, EPI_ISL_1029070, EPI_ISL_1029074, EPI_ISL_1029084, EPI_ISL_1029086, EPI_ISL_1029087, EPI_ISL_1029088, EPI_ISL_1029244, EPI_ISL_1029245, EPI_ISL_1029287, EPI_ISL_1029288, EPI_ISL_1029289, EPI_ISL_1029328, EPI_ISL_1029401, EPI_ISL_1029411, EPI_ISL_1029440, EPI_ISL_1029441, EPI_ISL_1029442, EPI_ISL_1029485, EPI_ISL_1029486, EPI_ISL_1029487, EPI_ISL_1029488, EPI_ISL_1029501, EPI_ISL_1029502, EPI_ISL_1029917, EPI_ISL_1029939, EPI_ISL_1029941, EPI_ISL_1029943, EPI_ISL_1030530, EPI_ISL_1030622, EPI_ISL_1030672, EPI_ISL_1030682, EPI_ISL_1030748, EPI_ISL_1030785, EPI_ISL_1030833, EPI_ISL_1030880, EPI_ISL_1030908, EPI_ISL_1030946, EPI_ISL_1030998, EPI_ISL_1031026, EPI_ISL_1031031, EPI_ISL_1031049, EPI_ISL_1031122, EPI_ISL_1031224, EPI_ISL_1031246, EPI_ISL_1031247, EPI_ISL_1031251, EPI_ISL_1031276, EPI_ISL_1031295, EPI_ISL_1031374, EPI_ISL_1031503, EPI_ISL_1031511, EPI_ISL_1031544, EPI_ISL_1031563, EPI_ISL_1031660, EPI_ISL_1031678, EPI_ISL_1031687, EPI_ISL_1031765, EPI_ISL_1031766, EPI_ISL_1031797, EPI_ISL_1031873, EPI_ISL_1032050, EPI_ISL_1032060, EPI_ISL_1032067, EPI_ISL_1032163, EPI_ISL_1032196, EPI_ISL_1032197, EPI_ISL_1032201, EPI_ISL_1032205, EPI_ISL_1032344, EPI_ISL_1032393, EPI_ISL_1032394, EPI_ISL_1032395, EPI_ISL_1032399, EPI_ISL_1032401, EPI_ISL_1032405, EPI_ISL_1032567, EPI_ISL_1032580, EPI_ISL_1032581 | | | |
| see above | Laboratory Corporation of America | Respiratory Viruses Branch, Division of Viral Diseases, Centers for Disease Control and Prevention | Peter W. Cook, Dakota Howard, Dhwani Batra, Ben L. Rambo-Martin, Clinton R. Paden, Suxiang Tong, Duncan MacCannell |
| EPI_ISL_1034279 | Northwest Pathology | UW Virology Lab | Pavitra Roychoudhury, Lasata Shrestha, Shah A Mohamed Bakhash, Michelle Lin, Hong Xie, Meei-Li Huang, Keith R Jerome, Alexander Greninger |
| EPI_ISL_1036954, EPI_ISL_1037009, EPI_ISL_1037086, EPI_ISL_1037167, EPI_ISL_1037195, EPI_ISL_1037232, EPI_ISL_1037250, EPI_ISL_1037255, EPI_ISL_1037306, EPI_ISL_1037475, EPI_ISL_1037497, EPI_ISL_1037510, EPI_ISL_1037640, EPI_ISL_1037650, EPI_ISL_1037655, EPI_ISL_1037790, EPI_ISL_1037859, EPI_ISL_1037868, EPI_ISL_1037980, EPI_ISL_1037988, EPI_ISL_1038042, EPI_ISL_1038074, EPI_ISL_1038121, EPI_ISL_1038179, EPI_ISL_1038215, EPI_ISL_1038223, EPI_ISL_1038235, EPI_ISL_1038302, EPI_ISL_1038344, EPI_ISL_1038361, EPI_ISL_1038367, EPI_ISL_1038368, EPI_ISL_1038406, EPI_ISL_1038485, EPI_ISL_1038490, EPI_ISL_1038491, EPI_ISL_1038492, EPI_ISL_1038493, EPI_ISL_1038784, EPI_ISL_1038785 | | | |
| see above | Laboratory Corporation of America | Respiratory Viruses Branch, Division of Viral Diseases, Centers for Disease Control and Prevention | Peter W. Cook, Dakota Howard, Dhwani Batra, Ben L. Rambo-Martin, Clinton R. Paden, Suxiang Tong, Duncan MacCannell |
| EPI_ISL_1038971, EPI_ISL_1038972, EPI_ISL_1038973, EPI_ISL_1038974, EPI_ISL_1038975, EPI_ISL_1038976, EPI_ISL_1038977, EPI_ISL_1038978, EPI_ISL_1038979, EPI_ISL_1038980, EPI_ISL_1038981 | | | |

| | | | |
|---|---|---|---|
| see above | Tempus | Grubaugh Lab - Yale School of Public Health | Joseph Fauver, Tara Alpert, Anderson Brito, Mallery Breban, Anne Wyllie, Chantal Vogels, Mary Petrone, Annie Watkins, Chaney Kalinich, Isabel Ott, Nathan Grubaugh |
| EPI_ISL_1038996 | Murphy Medical Associates | Grubaugh Lab - Yale School of Public Health | Joseph Fauver, Tara Alpert, Anderson Brito, Mallery Breban, Anne Wyllie, Chantal Vogels, Mary Petrone, Annie Watkins, Chaney Kalinich, Isabel Ott, Nathan Grubaugh |
| EPI_ISL_1040480 | Pandemic Response Lab - NYC | Pandemic Response Lab, R&D | Henry Lee, Michael Hammerling, Melissa Hopkins, Cybill del Castillo, William Ward, Pradeep Bugga, Haiping Hao, Jon Laurent |
| EPI_ISL_1040853, EPI_ISL_1040854, EPI_ISL_1040855, EPI_ISL_1040856, EPI_ISL_1040857, EPI_ISL_1040858, EPI_ISL_1040859, EPI_ISL_1040861, EPI_ISL_1040862, EPI_ISL_1040863, EPI_ISL_1040864, EPI_ISL_1040865, EPI_ISL_1040866, EPI_ISL_1040867, EPI_ISL_1040869, EPI_ISL_1040870, EPI_ISL_1040871, EPI_ISL_1040872, EPI_ISL_1040873, EPI_ISL_1040874, EPI_ISL_1040875, EPI_ISL_1040876, EPI_ISL_1040877, EPI_ISL_1040878, EPI_ISL_1040879, EPI_ISL_1040880, EPI_ISL_1040881, EPI_ISL_1040882, EPI_ISL_1040883, EPI_ISL_1040884, EPI_ISL_1040885, EPI_ISL_1040886, EPI_ISL_1040887, EPI_ISL_1040888, EPI_ISL_1040889, EPI_ISL_1040890, EPI_ISL_1040891, EPI_ISL_1040892, EPI_ISL_1040893, EPI_ISL_1040894, EPI_ISL_1040895, EPI_ISL_1040896, EPI_ISL_1040897, EPI_ISL_1040898, EPI_ISL_1040899, EPI_ISL_1040900, EPI_ISL_1040901, EPI_ISL_1040902, EPI_ISL_1040903, EPI_ISL_1040904, EPI_ISL_1040905, EPI_ISL_1040906, EPI_ISL_1040908, EPI_ISL_1040909, EPI_ISL_1040910, EPI_ISL_1040911, EPI_ISL_1040913, EPI_ISL_1040914 | | | |
| see above | NYU Langone Health | Departments of Pathology and Medicine, New York University School of Medicine | Adriana Heguy, Dacia Dimartino, Emily Guzman, Christian Marier, Peter Meyn, Sitharam Ramaswami, Gael Westby, Paul Zappile, Yutong Zhang, Paolo Cotzia, Guiqing Wang |
| EPI_ISL_1041213, EPI_ISL_1041214, EPI_ISL_1041215, EPI_ISL_1041216, EPI_ISL_1041217, EPI_ISL_1041218, EPI_ISL_1041219, EPI_ISL_1041220, EPI_ISL_1041221, EPI_ISL_1041222, EPI_ISL_1041223, EPI_ISL_1041224, EPI_ISL_1041225, EPI_ISL_1041226, EPI_ISL_1041227, EPI_ISL_1041228, EPI_ISL_1041229, EPI_ISL_1041230, EPI_ISL_1041231, EPI_ISL_1041232, EPI_ISL_1041233, EPI_ISL_1041234, EPI_ISL_1041235, EPI_ISL_1041236, EPI_ISL_1041237, EPI_ISL_1041238, EPI_ISL_1041239, EPI_ISL_1041240, EPI_ISL_1041241, EPI_ISL_1041242, EPI_ISL_1041243, EPI_ISL_1041244, EPI_ISL_1041245, EPI_ISL_1041246, EPI_ISL_1041247, EPI_ISL_1041248, EPI_ISL_1041249, EPI_ISL_1041250, EPI_ISL_1041251, EPI_ISL_1041252, EPI_ISL_1041253, EPI_ISL_1041254, EPI_ISL_1041255, EPI_ISL_1041256, EPI_ISL_1041257, EPI_ISL_1041258, EPI_ISL_1041259, EPI_ISL_1041260, EPI_ISL_1041261, EPI_ISL_1041262, EPI_ISL_1041263, EPI_ISL_1041264, EPI_ISL_1041265, EPI_ISL_1041266, EPI_ISL_1041267, EPI_ISL_1041268, EPI_ISL_1041269, EPI_ISL_1041270, EPI_ISL_1041271, EPI_ISL_1041272, EPI_ISL_1041273, EPI_ISL_1041274, EPI_ISL_1041275, EPI_ISL_1041276, EPI_ISL_1041277, EPI_ISL_1041278, EPI_ISL_1041279, EPI_ISL_1041280, EPI_ISL_1041281, EPI_ISL_1041282, EPI_ISL_1041283, EPI_ISL_1041284, EPI_ISL_1041285, EPI_ISL_1041286, EPI_ISL_1041287, EPI_ISL_1041288, EPI_ISL_1041289, EPI_ISL_1041290, EPI_ISL_1041291, EPI_ISL_1041292, EPI_ISL_1041293, EPI_ISL_1041294, EPI_ISL_1041295, EPI_ISL_1041296, EPI_ISL_1041297, EPI_ISL_1041298, EPI_ISL_1041299, EPI_ISL_1041300, EPI_ISL_1041301, EPI_ISL_1041302, EPI_ISL_1041303, EPI_ISL_1041304, EPI_ISL_1041305, EPI_ISL_1041306, EPI_ISL_1041307, EPI_ISL_1041308, EPI_ISL_1041309, EPI_ISL_1041310, EPI_ISL_1041311, EPI_ISL_1041312, EPI_ISL_1041313, EPI_ISL_1041314, EPI_ISL_1041315, EPI_ISL_1041316, EPI_ISL_1041317, EPI_ISL_1041318, EPI_ISL_1041319, EPI_ISL_1041320, EPI_ISL_1041321, EPI_ISL_1041322, EPI_ISL_1041323, EPI_ISL_1041324, EPI_ISL_1041325, EPI_ISL_1041326, EPI_ISL_1041327, EPI_ISL_1041328, EPI_ISL_1041329, EPI_ISL_1041330, EPI_ISL_1041331, EPI_ISL_1041332, EPI_ISL_1041333, EPI_ISL_1041334, EPI_ISL_1041335, EPI_ISL_1041336, EPI_ISL_1041337, EPI_ISL_1041338, EPI_ISL_1041339, EPI_ISL_1041340, EPI_ISL_1041341, EPI_ISL_1041342, EPI_ISL_1041343, EPI_ISL_1041344, EPI_ISL_1041345, EPI_ISL_1041346, EPI_ISL_1041347, EPI_ISL_1041348, EPI_ISL_1041349, EPI_ISL_1041350, EPI_ISL_1041351, EPI_ISL_1041352, EPI_ISL_1041353, EPI_ISL_1041354, EPI_ISL_1041355, EPI_ISL_1041356, EPI_ISL_1041357, EPI_ISL_1041358, EPI_ISL_1041359, EPI_ISL_1041360, EPI_ISL_1041361, EPI_ISL_1041362, EPI_ISL_1041363, EPI_ISL_1041364, EPI_ISL_1041365, EPI_ISL_1041366, EPI_ISL_1041367, EPI_ISL_1041368, EPI_ISL_1041369, EPI_ISL_1041370, EPI_ISL_1041371, EPI_ISL_1041372, EPI_ISL_1041373, EPI_ISL_1041374, EPI_ISL_1041375, EPI_ISL_1041376, EPI_ISL_1041377, EPI_ISL_1041378, EPI_ISL_1041379, EPI_ISL_1041380, EPI_ISL_1041381, EPI_ISL_1041382, EPI_ISL_1041383, EPI_ISL_1041384, EPI_ISL_1041385, EPI_ISL_1041386, EPI_ISL_1041387, EPI_ISL_1041388, EPI_ISL_1041389, EPI_ISL_1041390, EPI_ISL_1041391, EPI_ISL_1041392, EPI_ISL_1041393, EPI_ISL_1041394, EPI_ISL_1041395, EPI_ISL_1041396, EPI_ISL_1041397, EPI_ISL_1041398, EPI_ISL_1041399, EPI_ISL_1041400, EPI_ISL_1041401, EPI_ISL_1041402, EPI_ISL_1041403, EPI_ISL_1041404, EPI_ISL_1041405, EPI_ISL_1041406, EPI_ISL_1041407, EPI_ISL_1041408, EPI_ISL_1041409, EPI_ISL_1041410, EPI_ISL_1041411, EPI_ISL_1041412, EPI_ISL_1041413, EPI_ISL_1041414, EPI_ISL_1041415, EPI_ISL_1041416, EPI_ISL_1041417, EPI_ISL_1041418, EPI_ISL_1041419, EPI_ISL_1041420, EPI_ISL_1041421, EPI_ISL_1041422, EPI_ISL_1041423, EPI_ISL_1041424, EPI_ISL_1041425, EPI_ISL_1041426, EPI_ISL_1041427, EPI_ISL_1041428, EPI_ISL_1041429, EPI_ISL_1041430, EPI_ISL_1041431, EPI_ISL_1041432, EPI_ISL_1041433, EPI_ISL_1041434, EPI_ISL_1041435, EPI_ISL_1041436, EPI_ISL_1041437, EPI_ISL_1041438, EPI_ISL_1041439, EPI_ISL_1041440, EPI_ISL_1041441, EPI_ISL_1041442, EPI_ISL_1041443, EPI_ISL_1041444, EPI_ISL_1041445, EPI_ISL_1041446, EPI_ISL_1041447, EPI_ISL_1041448, EPI_ISL_1041449, EPI_ISL_1041450, EPI_ISL_1041451, EPI_ISL_1041452, EPI_ISL_1041453, EPI_ISL_1041454, EPI_ISL_1041455, EPI_ISL_1041456, EPI_ISL_1041457, EPI_ISL_1041458, EPI_ISL_1041459, EPI_ISL_1041460, EPI_ISL_1041461, EPI_ISL_1041462, EPI_ISL_1041463, EPI_ISL_1041464, EPI_ISL_1041465, EPI_ISL_1041466, EPI_ISL_1041467, EPI_ISL_1041468, EPI_ISL_1041469, EPI_ISL_1041470, EPI_ISL_1041471, EPI_ISL_1041472, EPI_ISL_1041473, EPI_ISL_1041474, EPI_ISL_1041475, EPI_ISL_1041476, EPI_ISL_1041477, EPI_ISL_1041478, EPI_ISL_1041479, EPI_ISL_1041480, EPI_ISL_1041481, EPI_ISL_1041482, EPI_ISL_1041483, EPI_ISL_1041484, EPI_ISL_1041485, EPI_ISL_1041486, EPI_ISL_1041487, EPI_ISL_1041488, EPI_ISL_1041489, EPI_ISL_1041490, EPI_ISL_1041491, EPI_ISL_1041492, EPI_ISL_1041493, EPI_ISL_1041494, EPI_ISL_1041495, EPI_ISL_1041496, EPI_ISL_1041497, EPI_ISL_1041498, EPI_ISL_1041499, EPI_ISL_1041500, EPI_ISL_1041501, EPI_ISL_1041502, EPI_ISL_1041503, EPI_ISL_1041504, EPI_ISL_1041505, EPI_ISL_1041506, EPI_ISL_1041507, EPI_ISL_1041508, EPI_ISL_1041510, EPI_ISL_1041511, EPI_ISL_1041512, EPI_ISL_1041513, EPI_ISL_1041514, EPI_ISL_1041515, EPI_ISL_1041516, EPI_ISL_1041517, EPI_ISL_1041518, EPI_ISL_1041519, EPI_ISL_1041520, EPI_ISL_1041521, EPI_ISL_1041522, EPI_ISL_1041523, EPI_ISL_1041524, EPI_ISL_1041525, EPI_ISL_1041526, EPI_ISL_1041527, EPI_ISL_1041528, EPI_ISL_1041529, EPI_ISL_1041530, EPI_ISL_1041531, EPI_ISL_1041532, EPI_ISL_1041533, EPI_ISL_1041534, EPI_ISL_1041535, EPI_ISL_1041536, EPI_ISL_1041537, EPI_ISL_1041538, EPI_ISL_1041539, EPI_ISL_1041540, EPI_ISL_1041541, EPI_ISL_1041542, EPI_ISL_1041543, EPI_ISL_1041544, EPI_ISL_1041545, EPI_ISL_1041546, EPI_ISL_1041547, EPI_ISL_1041548, EPI_ISL_1041549, EPI_ISL_1041550, EPI_ISL_1041551, EPI_ISL_1041552, EPI_ISL_1041553, EPI_ISL_1041554, EPI_ISL_1041555, EPI_ISL_1041556, EPI_ISL_1041557, EPI_ISL_1041558, EPI_ISL_1041559, EPI_ISL_1041560, EPI_ISL_1041561, EPI_ISL_1041562, EPI_ISL_1041563, EPI_ISL_1041564, EPI_ISL_1041565, EPI_ISL_1041566, EPI_ISL_1041567, EPI_ISL_1041568, EPI_ISL_1041569, EPI_ISL_1041570, EPI_ISL_1041571, EPI_ISL_1041572, EPI_ISL_1041573, EPI_ISL_1041574, EPI_ISL_1041575, EPI_ISL_1041576, EPI_ISL_1041577, EPI_ISL_1041578, EPI_ISL_1041579, EPI_ISL_1041580, EPI_ISL_1041581, EPI_ISL_1041582, EPI_ISL_1041583, EPI_ISL_1041584, EPI_ISL_1041585, EPI_ISL_1041586, EPI_ISL_1041587, EPI_ISL_1041588, EPI_ISL_1041589, EPI_ISL_1041590, EPI_ISL_1041591, EPI_ISL_1041592, EPI_ISL_1041593, EPI_ISL_1041594, EPI_ISL_1041595, EPI_ISL_1041596, EPI_ISL_1041597, EPI_ISL_1041598, EPI_ISL_1041599, EPI_ISL_1041600, EPI_ISL_1041601, EPI_ISL_1041602, EPI_ISL_1041603, EPI_ISL_1041604, EPI_ISL_1041605, EPI_ISL_1041606, EPI_ISL_1041607, EPI_ISL_1041608, EPI_ISL_1041609, EPI_ISL_1041610, EPI_ISL_1041611, EPI_ISL_1041612, EPI_ISL_1041613, EPI_ISL_1041614, EPI_ISL_1041615, EPI_ISL_1041616, EPI_ISL_1041617, EPI_ISL_1041618, EPI_ISL_1041619, EPI_ISL_1041620, EPI_ISL_1041621, EPI_ISL_1041622, EPI_ISL_1041623, EPI_ISL_1041624, EPI_ISL_1041625, EPI_ISL_1041626, EPI_ISL_1041627, EPI_ISL_1041628, EPI_ISL_1041629, EPI_ISL_1041630, EPI_ISL_1041631, EPI_ISL_1041632, EPI_ISL_1041633, EPI_ISL_1041634, EPI_ISL_1041635, EPI_ISL_1041636, EPI_ISL_1041637, EPI_ISL_1041638, EPI_ISL_1041639, EPI_ISL_1041640, EPI_ISL_1041641, EPI_ISL_1041642, EPI_ISL_1041643, EPI_ISL_1041644, EPI_ISL_1041645, EPI_ISL_1041646, EPI_ISL_1041647, EPI_ISL_1041648, EPI_ISL_1041649, EPI_ISL_1041650, EPI_ISL_1041651, EPI_ISL_1041652, EPI_ISL_1041653, EPI_ISL_1041654, EPI_ISL_1041655, EPI_ISL_1041656, EPI_ISL_1041657, EPI_ISL_1041658, EPI_ISL_1041659, EPI_ISL_1041660, EPI_ISL_1041661, EPI_ISL_1041662, EPI_ISL_1041663, EPI_ISL_1041664, EPI_ISL_1041665, EPI_ISL_1041666, EPI_ISL_1041667, EPI_ISL_1041668, EPI_ISL_1041669, EPI_ISL_1041670, EPI_ISL_1041671, EPI_ISL_1041672, EPI_ISL_1041673, EPI_ISL_1041674, EPI_ISL_1041675, EPI_ISL_1041676, EPI_ISL_1041677, EPI_ISL_1041678, EPI_ISL_1041679, EPI_ISL_1041680, EPI_ISL_1041681, EPI_ISL_1041682, EPI_ISL_1041683, EPI_ISL_1041684, EPI_ISL_1041685, EPI_ISL_1041686, EPI_ISL_1041687, EPI_ISL_1041688, EPI_ISL_1041689, EPI_ISL_1041690, EPI_ISL_1041691, EPI_ISL_1041692, EPI_ISL_1041693, EPI_ISL_1041694, EPI_ISL_1041695, EPI_ISL_1041696, EPI_ISL_1041697, EPI_ISL_1041698, EPI_ISL_1041699, EPI_ISL_1041700, EPI_ISL_1041701, EPI_ISL_1041702, EPI_ISL_1041703, EPI_ISL_1041704, EPI_ISL_1041705, EPI_ISL_1041706, EPI_ISL_1041707, EPI_ISL_1041708, EPI_ISL_1041709, EPI_ISL_1041710, EPI_ISL_1041711, EPI_ISL_1041712, EPI_ISL_1041713, EPI_ISL_1041714, EPI_ISL_1041715, EPI_ISL_1041716, EPI_ISL_1041717, EPI_ISL_1041718, EPI_ISL_1041719, EPI_ISL_1041720, EPI_ISL_1041721, EPI_ISL_1041722, EPI_ISL_1041723, EPI_ISL_1041724, EPI_ISL_1041725, EPI_ISL_1041726, EPI_ISL_1041727, EPI_ISL_1041728, EPI_ISL_1041729, EPI_ISL_1041730, EPI_ISL_1041731, EPI_ISL_1041732, EPI_ISL_1041733, EPI_ISL_1041734, EPI_ISL_1041735, EPI_ISL_1041736, EPI_ISL_1041737, EPI_ISL_1041738, EPI_ISL_1041739, EPI_ISL_1041740, EPI_ISL_1041741, EPI_ISL_1041742, EPI_ISL_1041743, EPI_ISL_1041744, EPI_ISL_1041745, EPI_ISL_1041746, EPI_ISL_1041747, EPI_ISL_1041748, EPI_ISL_1041749, EPI_ISL_1041750, EPI_ISL_1041751, EPI_ISL_1041752, EPI_ISL_1041753, EPI_ISL_1041754, EPI_ISL_1041755, EPI_ISL_1041756, EPI_ISL_1041757, EPI_ISL_1041758, EPI_ISL_1041759, EPI_ISL_1041760, EPI_ISL_1041761, EPI_ISL_1041762, EPI_ISL_1041763, EPI_ISL_1041764, EPI_ISL_1041765, EPI_ISL_1041766, EPI_ISL_1041767, EPI_ISL_1041768, EPI_ISL_1041769, EPI_ISL_1041770, EPI_ISL_1041771, EPI_ISL_1041772, EPI_ISL_1041773, EPI_ISL_1041774, EPI_ISL_1041775, EPI_ISL_1041776, EPI_ISL_1041777, EPI_ISL_1041778, EPI_ISL_1041779, EPI_ISL_1041780, EPI_ISL_1041781, EPI_ISL_1041782, EPI_ISL_1041783, EPI_ISL_1041784, EPI_ISL_1041785, EPI_ISL_1041786, EPI_ISL_1041787, EPI_ISL_1041788, EPI_ISL_1041789, EPI_ISL_1041790, EPI_ISL_1041791, EPI_ISL_1041792, EPI_ISL_1041793, EPI_ISL_1041794, EPI_ISL_1041795, EPI_ISL_1041796, EPI_ISL_1041797, EPI_ISL_1041798, EPI_ISL_1041799, EPI_ISL_1041800, EPI_ISL_1041801, EPI_ISL_1041802, EPI_ISL_1041803, EPI_ISL_1041804, EPI_ISL_1041805, EPI_ISL_1041806, EPI_ISL_1041807, EPI_ISL_1041808, EPI_ISL_1041809, EPI_ISL_1041810, EPI_ISL_1041811, EPI_ISL_1041812, EPI_ISL_1041813, EPI_ISL_1041814, EPI_ISL_1041815, EPI_ISL_1041816, EPI_ISL_1041817, EPI_ISL_1041818, EPI_ISL_1041819, EPI_ISL_1041820, EPI_ISL_1041821, EPI_ISL_1041822, EPI_ISL_1041823, EPI_ISL_1041824, EPI_ISL_1041825, EPI_ISL_1041826, EPI_ISL_1041827, EPI_ISL_1041828, EPI_ISL_1041829, EPI_ISL_1041830, EPI_ISL_1041831, EPI_ISL_1041832, EPI_ISL_1041833, EPI_ISL_1041834, EPI_ISL_1041835, EPI_ISL_1041836, EPI_ISL_1041837, EPI_ISL_1041838, EPI_ISL_1041839, EPI_ISL_1041840, EPI_ISL_1041841, EPI_ISL_1041842, EPI_ISL_1041843, EPI_ISL_1041844, EPI_ISL_1041845, EPI_ISL_1041846, EPI_ISL_1041847, EPI_ISL_1041848, EPI_ISL_1041849, EPI_ISL_1041850, EPI_ISL_1041851, EPI_ISL_1041852, EPI_ISL_1041853, EPI_ISL_1041854, EPI_ISL_1041855, EPI_ISL_1041856, EPI_ISL_1041857, EPI_ISL_1041858, EPI_ISL_1041859, EPI_ISL_1041860, EPI_ISL_1041861, EPI_ISL_1041862, EPI_ISL_1041863, EPI_ISL_1041864, EPI_ISL_1041865, EPI_ISL_1041866, EPI_ISL_1041867, EPI_ISL_1041868, EPI_ISL_1041869, EPI_ISL_1041870, EPI_ISL_1041871, EPI_ISL_1041872, EPI_ISL_1041873, EPI_ISL_1041874, EPI_ISL_1041875, EPI_ISL_1041876, EPI_ISL_1041877, EPI_ISL_1041878, EPI_ISL_1041879, EPI_ISL_1041880, EPI_ISL_1041881, EPI_ISL_1041882, EPI_ISL_1041883, EPI_ISL_1041884, EPI_ISL_1041885, EPI_ISL_1041886, EPI_ISL_1041887, EPI_ISL_1041888, EPI_ISL_1041889, EPI_ISL_1041890, EPI_ISL_1041891, EPI_ISL_1041892, EPI_ISL_1041893, EPI_ISL_1041894, EPI_ISL_1041895, EPI_ISL_1041896, EPI_ISL_1041897, EPI_ISL_1041898, EPI_ISL_1041899, EPI_ISL_1041900, EPI_ISL_1041901, EPI_ISL_1041902, EPI_ISL_1041903, EPI_ISL_1041904, EPI_ISL_1041905, EPI_ISL_1041906, EPI_ISL_1041907, EPI_ISL_1041908, EPI_ISL_1041909, EPI_ISL_1041910, EPI_ISL_1041911, EPI_ISL_1041912, EPI_ISL_1041913, EPI_ISL_1041914, EPI_ISL_1041915, EPI_ISL_1041916, EPI_ISL_1041917, EPI_ISL_1041918, EPI_ISL_1041919, EPI_ISL_1041920, EPI_ISL_1041921, EPI_ISL_1041922, EPI_ISL_1041923, EPI_ISL_1041924, EPI_ISL_1041925, EPI_ISL_1041926, EPI_ISL_1041927, EPI_ISL_1041928, EPI_ISL_1041929, EPI_ISL_1041930 | | | |
| see above | Pandemic Response Lab - NYC | Pandemic Response Lab, R&D | Henry Lee, Michael Hammerling, Melissa Hopkins, Cybill del Castillo, William Ward, Pradeep Bugga, Haiping Hao, Jon Laurent |
| EPI_ISL_1049338, EPI_ISL_1049339 | Wadsworth Center, New York State Department.of Health | Wadsworth Center, New York State Department.of Health | Kirsten St. George, Daryl M. Lamson, Alexis Russel, Matthew Shudt, Melissa A Leisner, Jonathan Plitnick, Navjot Singh, John Kelly, Erasmus Schneider, Erica Lasek-Nesselquist |
| EPI_ISL_1049482 | MSHS Clinical Microbiology Laboratories | MSHS Pathogen Surveillance Program | Ana S. Gonzalez-Reiche, Hala Alshammary, Mitchell J. Sullivan, Brianne Ciferri, Ajay Obla, Angela Amoako, Mahmoud Awawda, Daniel Floda, Julia Matthews, Ashley Salimbangon, Levy Sominsky, Katherine Beach, Kayla Russo, Charles Gleason, Shelcie Fabre, Giulio Kleiner, Zenab Khan, Bremy Alburquerque, Adriana van de Guchte, Komal Srivastava, Matthew M. Hernandez, Jayeeta Dutta, Denise Jurczyszak, Nancy Francoeur, Betsaida Salom Melo, Irina Oussenko, Gintaras Deikus, Juan Soto, Shwetha Hara Sridhar, Ying-Chih Wang, Kathryn Twyman, Deena R. Altman, Robert Sebra, Adolfo García-Sastre, Marta Luksza, Gopi Patel, Sarah Schaefer, Melissa Gitman, Michael D. Nowak, Alberto Paniz-Mondolfi, Emilia Mia Sordillo, Viviana Simon, Harm van Bakel |
| EPI_ISL_1050008, EPI_ISL_1050009, EPI_ISL_1050010, EPI_ISL_1050011, EPI_ISL_1050012, EPI_ISL_1050013, EPI_ISL_1050014, EPI_ISL_1050015, EPI_ISL_1050016, EPI_ISL_1050017, EPI_ISL_1050018, EPI_ISL_1050019, EPI_ISL_1050020, EPI_ISL_1050021, EPI_ISL_1050022, EPI_ISL_1050023, EPI_ISL_1050024, EPI_ISL_1050025, EPI_ISL_1050026, EPI_ISL_1050027, EPI_ISL_1050028, EPI_ISL_1050029, EPI_ISL_1050030, EPI_ISL_1050031, EPI_ISL_1050032, EPI_ISL_1050033, EPI_ISL_1050034, EPI_ISL_1050035, EPI_ISL_1050036, EPI_ISL_1050037, EPI_ISL_1050038, EPI_ISL_1050039, EPI_ISL_1050040, EPI_ISL_1050041, EPI_ISL_1050043, EPI_ISL_1050044, EPI_ISL_1050045, EPI_ISL_1050046, EPI_ISL_1050048, EPI_ISL_1050049, EPI_ISL_1050050, EPI_ISL_1050051, EPI_ISL_1050052, EPI_ISL_1050053, EPI_ISL_1050054, EPI_ISL_1050055, EPI_ISL_1050056, EPI_ISL_1050057, EPI_ISL_1050058, EPI_ISL_1050059, EPI_ISL_1050060, EPI_ISL_1050061, EPI_ISL_1050062, EPI_ISL_1050063, EPI_ISL_1050064, EPI_ISL_1060616, EPI_ISL_1060617, EPI_ISL_1060618, EPI_ISL_1060619, EPI_ISL_1060620, EPI_ISL_1060621, EPI_ISL_1060622, EPI_ISL_1060623, EPI_ISL_1060624, EPI_ISL_1060625, EPI_ISL_1060627, EPI_ISL_1060628, EPI_ISL_1060629, EPI_ISL_1060630, EPI_ISL_1060631, EPI_ISL_1060632, EPI_ISL_1060633, EPI_ISL_1060634, EPI_ISL_1060635, EPI_ISL_1060636, EPI_ISL_1060637, EPI_ISL_1060638, EPI_ISL_1060639, EPI_ISL_1060640, EPI_ISL_1060641, EPI_ISL_1060642, EPI_ISL_1060643, EPI_ISL_1060644, EPI_ISL_1060645, EPI_ISL_1060646, EPI_ISL_1060648, EPI_ISL_1060649, EPI_ISL_1060650, EPI_ISL_1060651, EPI_ISL_1060652, EPI_ISL_1060653, EPI_ISL_1060654, EPI_ISL_1060656, EPI_ISL_1060657, EPI_ISL_1060658, EPI_ISL_1060660, EPI_ISL_1060661, EPI_ISL_1060662, EPI_ISL_1060663, EPI_ISL_1060664, EPI_ISL_1060665, EPI_ISL_1060666, EPI_ISL_1060667, EPI_ISL_1060668, EPI_ISL_1060669, EPI_ISL_1060670, EPI_ISL_1060672, EPI_ISL_1060674, EPI_ISL_1060675 | | | |
| see above | NYU Langone Health | Departments of Pathology and Medicine, New York University School of Medicine | Adriana Heguy, Dacia Dimartino, Emily Guzman, Christian Marier, Peter Meyn, Sitharam Ramaswami, Gael Westby, Paul Zappile, Yutong Zhang, Paolo Cotzia, Guiqing Wang |
| EPI_ISL_1064272 | University of Wisconsin-Madison AIDS Vaccine Research Laboratories | University of Wisconsin-Madison AIDS Vaccine Research Laboratories | Gage Moreno, Katarina Braun, et al. AIDS Vaccine Research Laboratories |
| EPI_ISL_1064751, EPI_ISL_1064752, EPI_ISL_1064753, EPI_ISL_1064754 | OCME Office Of Chief Medical Examiner | New York City Public Health Laboratory | Jade Wang, et al. |
| EPI_ISL_1064755 | DOHMH Jamaica | New York City Public Health Laboratory | Jade Wang, et al. |
| EPI_ISL_1064756 | DOHMH PHL | New York City Public Health Laboratory | Jade Wang, et al. |
| EPI_ISL_1064757 | DOHMH Corona | New York City Public Health Laboratory | Jade Wang, et al. |

| | | | |
|---|---|---|---|
| EPI_ISL_1064758 | Department of Homeless Services | New York City Public Health Laboratory | Jade Wang, et al. |
| EPI_ISL_1064759 | DOHMH Jamaica | New York City Public Health Laboratory | Jade Wang, et al. |
| EPI_ISL_1064760, EPI_ISL_1064761 | DOHMH Corona | New York City Public Health Laboratory | Jade Wang, et al. |
| EPI_ISL_1064762 | DOHMH Fort Greene | New York City Public Health Laboratory | Jade Wang, et al. |
| EPI_ISL_1064763, EPI_ISL_1064764, EPI_ISL_1064765 | OCME Office Of Chief Medical Examiner | New York City Public Health Laboratory | Jade Wang, et al. |
| EPI_ISL_1064766 | DOHMH Chelsea | New York City Public Health Laboratory | Jade Wang, et al. |
| EPI_ISL_1064767 | DOHMH Corona | New York City Public Health Laboratory | Jade Wang, et al. |
| EPI_ISL_1064768 | Department of Homeless Services | New York City Public Health Laboratory | Jade Wang, et al. |
| EPI_ISL_1064769 | DOHMH PHL | New York City Public Health Laboratory | Jade Wang, et al. |
| EPI_ISL_1064770 | OCME Office Of Chief Medical Examiner | New York City Public Health Laboratory | Jade Wang, et al. |
| EPI_ISL_1064771 | DOHMH Fort Greene | New York City Public Health Laboratory | Jade Wang, et al. |
| EPI_ISL_1064772, EPI_ISL_1064773, EPI_ISL_1064774 | OCME Office Of Chief Medical Examiner | New York City Public Health Laboratory | Jade Wang, et al. |
| EPI_ISL_1064775 | DOHMH Central Harlem | New York City Public Health Laboratory | Jade Wang, et al. |
| EPI_ISL_1064776 | OCME Office Of Chief Medical Examiner | New York City Public Health Laboratory | Jade Wang, et al. |
| EPI_ISL_1064777 | DOHMH Crown Heights | New York City Public Health Laboratory | Jade Wang, et al. |
| EPI_ISL_1064778 | OCME Office Of Chief Medical Examiner | New York City Public Health Laboratory | Jade Wang, et al. |
| EPI_ISL_1064779 | DOHMH Chelsea | New York City Public Health Laboratory | Jade Wang, et al. |
| EPI_ISL_1064780 | DOHMH Fort Greene | New York City Public Health Laboratory | Jade Wang, et al. |
| EPI_ISL_1064781 | DOHMH Jamaica | New York City Public Health Laboratory | Jade Wang, et al. |
| EPI_ISL_1064782 | DOHMH Chelsea | New York City Public Health Laboratory | Jade Wang, et al. |
| EPI_ISL_1064783 | OCME Office Of Chief Medical Examiner | New York City Public Health Laboratory | Jade Wang, et al. |
| EPI_ISL_1064784 | DOHMH Corona | New York City Public Health Laboratory | Jade Wang, et al. |
| EPI_ISL_1064785 | DOHMH Fort Greene | New York City Public Health Laboratory | Jade Wang, et al. |
| EPI_ISL_1064786 | DOHMH Corona | New York City Public Health Laboratory | Jade Wang, et al. |
| EPI_ISL_1064787 | DOHMH Morrisania | New York City Public Health Laboratory | Jade Wang, et al. |
| EPI_ISL_1064788 | DOHMH Jamaica | New York City Public Health Laboratory | Jade Wang, et al. |
| EPI_ISL_1064789 | DOHMH Corona | New York City Public Health Laboratory | Jade Wang, et al. |
| EPI_ISL_1064790 | DOHMH Fort Greene | New York City Public Health Laboratory | Jade Wang, et al. |
| EPI_ISL_1064791 | DOHMH Crown Heights | New York City Public Health Laboratory | Jade Wang, et al. |
| EPI_ISL_1064792, EPI_ISL_1064793 | OCME Office Of Chief Medical Examiner | New York City Public Health Laboratory | Jade Wang, et al. |
| EPI_ISL_1064794 | DOHMH Fort Greene | New York City Public Health Laboratory | Jade Wang, et al. |
| EPI_ISL_1064795, EPI_ISL_1064796 | OCME Office Of Chief Medical Examiner | New York City Public Health Laboratory | Jade Wang, et al. |
| EPI_ISL_1064797 | DOHMH Chelsea | New York City Public Health Laboratory | Jade Wang, et al. |
| EPI_ISL_1064798 | DOHMH Crown Heights | New York City Public Health Laboratory | Jade Wang, et al. |
| EPI_ISL_1064799 | DOHMH PHL | New York City Public Health Laboratory | Jade Wang, et al. |
| EPI_ISL_1064800 | NYC HH Elmhurst Hospital Medical Center | New York City Public Health Laboratory | Jade Wang, et al. |
| EPI_ISL_1064801, EPI_ISL_1064802 | DOHMH Morrisania | New York City Public Health Laboratory | Jade Wang, et al. |
| EPI_ISL_1064803 | DOHMH Central Harlem | New York City Public Health Laboratory | Jade Wang, et al. |
| EPI_ISL_1064804 | OCME Office Of Chief Medical Examiner | New York City Public Health Laboratory | Jade Wang, et al. |
| EPI_ISL_1064805, EPI_ISL_1064806 | DOHMH PHL | New York City Public Health Laboratory | Jade Wang, et al. |
| EPI_ISL_1064807 | DOHMH Jamaica | New York City Public Health Laboratory | Jade Wang, et al. |
| EPI_ISL_1064808 | DOHMH Corona | New York City Public Health Laboratory | Jade Wang, et al. |
| EPI_ISL_1064809 | DOHMH Central Harlem | New York City Public Health Laboratory | Jade Wang, et al. |
| EPI_ISL_1064810 | Department of Homeless Services | New York City Public Health Laboratory | Jade Wang, et al. |
| EPI_ISL_1064811 | DOHMH Corona | New York City Public Health Laboratory | Jade Wang, et al. |
| EPI_ISL_1064812 | DOHMH Morrisania | New York City Public Health Laboratory | Jade Wang, et al. |
| EPI_ISL_1064813, EPI_ISL_1064814, EPI_ISL_1064815, EPI_ISL_1064816, EPI_ISL_1064817, EPI_ISL_1064818 | OCME Office Of Chief Medical Examiner | New York City Public Health Laboratory | Jade Wang, et al. |
| EPI_ISL_1064819 | DOHMH Jamaica | New York City Public Health Laboratory | Jade Wang, et al. |
| EPI_ISL_1064820 | DOHMH Central Harlem | New York City Public Health Laboratory | Jade Wang, et al. |
| EPI_ISL_1064821, EPI_ISL_1064822 | DOHMH PHL | New York City Public Health Laboratory | Jade Wang, et al. |
| EPI_ISL_1064823 | DOHMH Riverside | New York City Public Health Laboratory | Jade Wang, et al. |
| EPI_ISL_1064824 | DOHMH Crown Heights | New York City Public Health Laboratory | Jade Wang, et al. |
| EPI_ISL_1064825 | DOHMH Central Harlem | New York City Public Health Laboratory | Jade Wang, et al. |
| EPI_ISL_1064826 | DOHMH PHL | New York City Public Health Laboratory | Jade Wang, et al. |
| EPI_ISL_1064827, EPI_ISL_1064828 | Department of Homeless Services | New York City Public Health Laboratory | Jade Wang, et al. |
| EPI_ISL_1064829 | OCME Office Of Chief Medical Examiner | New York City Public Health Laboratory | Jade Wang, et al. |

| | | | |
|---|---|---|---|
| EPI_ISL_1064830 | DOHMH Jamaica | New York City Public Health Laboratory | Jade Wang, et al. |
| EPI_ISL_1064831 | DOHMH Central Harlem | New York City Public Health Laboratory | Jade Wang, et al. |
| EPI_ISL_1064832, EPI_ISL_1064833 | OCME Office Of Chief Medical Examiner | New York City Public Health Laboratory | Jade Wang, et al. |
| EPI_ISL_1064834 | DOHMH Morrisania | New York City Public Health Laboratory | Jade Wang, et al. |
| EPI_ISL_1064835 | DOHMH Crown Heights | New York City Public Health Laboratory | Jade Wang, et al. |
| EPI_ISL_1064836 | DOHMH Fort Greene | New York City Public Health Laboratory | Jade Wang, et al. |
| EPI_ISL_1064837, EPI_ISL_1064838, EPI_ISL_1064839, EPI_ISL_1064840 | DOHMH Jamaica | New York City Public Health Laboratory | Jade Wang, et al. |
| EPI_ISL_1064841, EPI_ISL_1064842 | DOHMH Morrisania | New York City Public Health Laboratory | Jade Wang, et al. |
| EPI_ISL_1064843, EPI_ISL_1064844, EPI_ISL_1064845, EPI_ISL_1064846 | OCME Office Of Chief Medical Examiner | New York City Public Health Laboratory | Jade Wang, et al. |
| EPI_ISL_1064847 | DOHMH Corona | New York City Public Health Laboratory | Jade Wang, et al. |
| EPI_ISL_1064848 | DOHMH Jamaica | New York City Public Health Laboratory | Jade Wang, et al. |
| EPI_ISL_1064849, EPI_ISL_1064850 | DOHMH Morrisania | New York City Public Health Laboratory | Jade Wang, et al. |
| EPI_ISL_1064851, EPI_ISL_1064852 | Department of Homeless Services | New York City Public Health Laboratory | Jade Wang, et al. |
| EPI_ISL_1064853, EPI_ISL_1064854, EPI_ISL_1064855, EPI_ISL_1064856, EPI_ISL_1064857 | OCME Office Of Chief Medical Examiner | New York City Public Health Laboratory | Jade Wang, et al. |
| EPI_ISL_1064858 | DOHMH Jamaica | New York City Public Health Laboratory | Jade Wang, et al. |
| EPI_ISL_1064859, EPI_ISL_1064860 | DOHMH PHL | New York City Public Health Laboratory | Jade Wang, et al. |
| EPI_ISL_1064861, EPI_ISL_1064862 | DOHMH Morrisania | New York City Public Health Laboratory | Jade Wang, et al. |
| EPI_ISL_1064863, EPI_ISL_1064864 | OCME Office Of Chief Medical Examiner | New York City Public Health Laboratory | Jade Wang, et al. |
| EPI_ISL_1064865, EPI_ISL_1064866, EPI_ISL_1064867, EPI_ISL_1064868, EPI_ISL_1064869 | DOHMH Jamaica | New York City Public Health Laboratory | Jade Wang, et al. |
| EPI_ISL_1064870, EPI_ISL_1064871 | DOHMH Corona | New York City Public Health Laboratory | Jade Wang, et al. |
| EPI_ISL_1064872 | DOHMH Chelsea | New York City Public Health Laboratory | Jade Wang, et al. |
| EPI_ISL_1064873, EPI_ISL_1064874 | DOHMH Corona | New York City Public Health Laboratory | Jade Wang, et al. |
| EPI_ISL_1064875 | DOHMH Morrisania | New York City Public Health Laboratory | Jade Wang, et al. |
| EPI_ISL_1064876 | DOHMH Central Harlem | New York City Public Health Laboratory | Jade Wang, et al. |
| EPI_ISL_1064877, EPI_ISL_1064878, EPI_ISL_1064879, EPI_ISL_1064880, EPI_ISL_1064881, EPI_ISL_1064882 | OCME Office Of Chief Medical Examiner | New York City Public Health Laboratory | Jade Wang, et al. |
| EPI_ISL_1064883 | DOHMH Crown Heights | New York City Public Health Laboratory | Jade Wang, et al. |
| EPI_ISL_1064884 | DOHMH Jamaica | New York City Public Health Laboratory | Jade Wang, et al. |
| EPI_ISL_1064885 | DOHMH Fort Greene | New York City Public Health Laboratory | Jade Wang, et al. |
| EPI_ISL_1064886 | DOHMH Jamaica | New York City Public Health Laboratory | Jade Wang, et al. |
| EPI_ISL_1064887 | DOHMH PHL | New York City Public Health Laboratory | Jade Wang, et al. |
| EPI_ISL_1064888 | DOHMH Corona | New York City Public Health Laboratory | Jade Wang, et al. |
| EPI_ISL_1064889 | Department of Homeless Services | New York City Public Health Laboratory | Jade Wang, et al. |
| EPI_ISL_1064890 | DOHMH Crown Heights | New York City Public Health Laboratory | Jade Wang, et al. |
| EPI_ISL_1064891 | DOHMH Fort Greene | New York City Public Health Laboratory | Jade Wang, et al. |
| EPI_ISL_1064892 | DOHMH Jamaica | New York City Public Health Laboratory | Jade Wang, et al. |
| EPI_ISL_1064893 | DOHMH Corona | New York City Public Health Laboratory | Jade Wang, et al. |
| EPI_ISL_1064894 | DOHMH PHL | New York City Public Health Laboratory | Jade Wang, et al. |
| EPI_ISL_1064895, EPI_ISL_1064896 | DOHMH Chelsea | New York City Public Health Laboratory | Jade Wang, et al. |
| EPI_ISL_1064897 | DOHMH Corona | New York City Public Health Laboratory | Jade Wang, et al. |
| EPI_ISL_1064898 | Department of Homeless Services | New York City Public Health Laboratory | Jade Wang, et al. |
| EPI_ISL_1064899, EPI_ISL_1064900, EPI_ISL_1064901, EPI_ISL_1064902, EPI_ISL_1064903 | OCME Office Of Chief Medical Examiner | New York City Public Health Laboratory | Jade Wang, et al. |
| EPI_ISL_1064904 | DOHMH Crown Heights | New York City Public Health Laboratory | Jade Wang, et al. |
| EPI_ISL_1064905 | DOHMH Corona | New York City Public Health Laboratory | Jade Wang, et al. |
| EPI_ISL_1064906, EPI_ISL_1064907 | DOHMH Jamaica | New York City Public Health Laboratory | Jade Wang, et al. |
| EPI_ISL_1064908 | DOHMH Riverside | New York City Public Health Laboratory | Jade Wang, et al. |
| EPI_ISL_1064909 | DOHMH Central Harlem | New York City Public Health Laboratory | Jade Wang, et al. |
| EPI_ISL_1064910, EPI_ISL_1064911, EPI_ISL_1064912, EPI_ISL_1064913, EPI_ISL_1064914 | OCME Office Of Chief Medical Examiner | New York City Public Health Laboratory | Jade Wang, et al. |
| EPI_ISL_1067638, EPI_ISL_1067640, EPI_ISL_1067645, EPI_ISL_1067653, EPI_ISL_1067674, EPI_ISL_1067677, EPI_ISL_1067688, EPI_ISL_1080456 | Murphy Medical Associates | Grubaugh Lab - Yale School of Public Health | Mary Petrone, Joseph Fauver, Caleb Neal, Steven Murphy, Chantal Vogels, Mallery Breban, Annie Watkins, Tara Alpert, Nathan Grubaugh |

EPI_ISL_1080751, EPI_ISL_1080752, EPI_ISL_1080753, EPI_ISL_1080754, EPI_ISL_1080755, EPI_ISL_1080756, EPI_ISL_1080757, EPI_ISL_1080758, EPI_ISL_1080759, EPI_ISL_1080760, EPI_ISL_1080761, EPI_ISL_1080762, EPI_ISL_1080763, EPI_ISL_1080764, EPI_ISL_1080765, EPI_ISL_1080766, EPI_ISL_1080767,

| | | | |
|---|---|---|---|
| EPI_ISL_1080768, EPI_ISL_1080769, EPI_ISL_1080770, EPI_ISL_1080771, EPI_ISL_1080772, EPI_ISL_1080773, EPI_ISL_1080774, EPI_ISL_1080775, EPI_ISL_1080776, EPI_ISL_1080777, EPI_ISL_1080778, EPI_ISL_1080779, EPI_ISL_1080780, EPI_ISL_1080781, EPI_ISL_1080782, EPI_ISL_1080783, EPI_ISL_1080784, EPI_ISL_1080785, EPI_ISL_1080786, EPI_ISL_1080787, EPI_ISL_1080788, EPI_ISL_1080789, EPI_ISL_1080790, EPI_ISL_1080791, EPI_ISL_1080792, EPI_ISL_1080793, EPI_ISL_1080794, EPI_ISL_1080795, EPI_ISL_1080796, EPI_ISL_1080797, EPI_ISL_1080798, EPI_ISL_1080799, EPI_ISL_1080800, EPI_ISL_1080801, EPI_ISL_1080802, EPI_ISL_1080803, EPI_ISL_1080804, EPI_ISL_1080805, EPI_ISL_1080806, EPI_ISL_1080807, EPI_ISL_1080808, EPI_ISL_1080809, EPI_ISL_1080810 | | | |
| see above | Clinical Microbiology Laboratory, NewYork Presbyterian Hospital/Columbia University Irving Medical Center | Uhlemann Laboratory, Columbia University Irving Medical Center | Medini K. Annavajhala, Anne-Catrin Uhlemann, Hiroshi Mohri, David Ho |
| EPI_ISL_1081047 | Helix/Illumina | Respiratory Viruses Branch, Division of Viral Diseases, Centers for Disease Control and Prevention | Peter W. Cook, Dakota Howard, Dhwani Batra, Ben L. Rambo-Martin, Eileen de Feo, Jan Antico, Christine Tran, Matthew Tolentino, Shannon Wickline, Kim Gietzen, Brad Sickler, Jingtao Liu, Eric Allen, Phil Febbo, Summer Galloway, Nicole L. Washington, Simon White, Geraint Levan, Kelly Schiabor Barrett, Elizabeth Cirulli, Alexandre Bolze, Ary Ascencio, Charlotte Rivera-Garcia, Ryan Cho, Jason Nguyen, Sherry Wang, Jimmy Ramirez, Tyler Cassens, Efren Sandoval, Magnus Isaksson, William Lee, David Becker, Marc Laurent, James Lu, Clinton R. Paden, Suxiang Tong, Duncan MacCannell |
| EPI_ISL_1081200, EPI_ISL_1081208, EPI_ISL_1081215, EPI_ISL_1081270, EPI_ISL_1081271 | Laboratory Corporation of America | Respiratory Viruses Branch, Division of Viral Diseases, Centers for Disease Control and Prevention | Peter W. Cook, Dakota Howard, Dhwani Batra, Ben L. Rambo-Martin, Eileen de Feo, Jan Antico, Christine Tran, Matthew Tolentino, Shannon Wickline, Kim Gietzen, Brad Sickler, Jingtao Liu, Eric Allen, Phil Febbo, Summer Galloway, Nicole L. Washington, Simon White, Geraint Levan, Kelly Schiabor Barrett, Elizabeth Cirulli, Alexandre Bolze, Ary Ascencio, Charlotte Rivera-Garcia, Ryan Cho, Jason Nguyen, Sherry Wang, Jimmy Ramirez, Tyler Cassens, Efren Sandoval, Magnus Isaksson, William Lee, David Becker, Marc Laurent, James Lu, Clinton R. Paden, Suxiang Tong, Duncan MacCannell |
| EPI_ISL_1085093, EPI_ISL_1085161, EPI_ISL_1085166, EPI_ISL_1085168, EPI_ISL_1085171, EPI_ISL_1085175 | NYU Langone Health | Departments of Pathology and Medicine, New York University School of Medicine | Adriana Heguy, Dacia Dimartino, Emily Guzman, Christian Marier, Peter Meyn, Sitharam Ramaswami, Gael Westby, Paul Zappile, Yutong Zhang, Paolo Cotzia, Guiqing Wang |
| EPI_ISL_1086395, EPI_ISL_1086555, EPI_ISL_1086562, EPI_ISL_1086567, EPI_ISL_1086581, EPI_ISL_1086611, EPI_ISL_1086614, EPI_ISL_1086627, EPI_ISL_1086643, EPI_ISL_1086647, EPI_ISL_1086652, EPI_ISL_1086656, EPI_ISL_1086718, EPI_ISL_1086862, EPI_ISL_1086880, EPI_ISL_1087014, EPI_ISL_1087022, EPI_ISL_1087023, EPI_ISL_1087040, EPI_ISL_1087046, EPI_ISL_1087067, EPI_ISL_1087118, EPI_ISL_1087130, EPI_ISL_1087205, EPI_ISL_1087206, EPI_ISL_1087244, EPI_ISL_1087253 | | | |
| see above | Quest Diagnostics Incorporated | Respiratory Viruses Branch, Division of Viral Diseases, Centers for Disease Control and Prevention | Peter W. Cook, Dakota Howard, Dhwani Batra, Ben L. Rambo-Martin, S. H. Rosenthal, A. Gerasimova, R. M. Kagan, B. Anderson, M. Hua, Y. Liu, L.E. Bernstein, K.E. Livingston, A. Perez, I. A. Shlyakhter, R. V. Rolando, R. Owen, P. Tanpaiboon, F. Lacbawan, Clinton R. Paden, Suxiang Tong, Duncan MacCannell |
| EPI_ISL_1087272, EPI_ISL_1087273, EPI_ISL_1087274, EPI_ISL_1087275, EPI_ISL_1087276, EPI_ISL_1087277, EPI_ISL_1087278, EPI_ISL_1087279, EPI_ISL_1087280, EPI_ISL_1087281, EPI_ISL_1087282, EPI_ISL_1087283, EPI_ISL_1087284, EPI_ISL_1087285, EPI_ISL_1087287, EPI_ISL_1087288, EPI_ISL_1087289, EPI_ISL_1087290, EPI_ISL_1087291, EPI_ISL_1087292, EPI_ISL_1087293, EPI_ISL_1087294, EPI_ISL_1087295, EPI_ISL_1087296, EPI_ISL_1087297, EPI_ISL_1087298, EPI_ISL_1087299, EPI_ISL_1087300, EPI_ISL_1087301, EPI_ISL_1087302, EPI_ISL_1087303, EPI_ISL_1087304, EPI_ISL_1087305, EPI_ISL_1087306, EPI_ISL_1087307, EPI_ISL_1087308, EPI_ISL_1087309, EPI_ISL_1087310, EPI_ISL_1087311, EPI_ISL_1087312, EPI_ISL_1087313, EPI_ISL_1087314, EPI_ISL_1087315, EPI_ISL_1087316, EPI_ISL_1087317, EPI_ISL_1087318, EPI_ISL_1087319, EPI_ISL_1087320, EPI_ISL_1087321, EPI_ISL_1087322, EPI_ISL_1087323, EPI_ISL_1087324, EPI_ISL_1087325, EPI_ISL_1087326, EPI_ISL_1087327, EPI_ISL_1087328, EPI_ISL_1087329, EPI_ISL_1087330, EPI_ISL_1087331, EPI_ISL_1087332, EPI_ISL_1087333, EPI_ISL_1087334, EPI_ISL_1087335, EPI_ISL_1087336, EPI_ISL_1087337, EPI_ISL_1087338, EPI_ISL_1087339, EPI_ISL_1087340, EPI_ISL_1087341, EPI_ISL_1087342, EPI_ISL_1087343, EPI_ISL_1087344, EPI_ISL_1087345, EPI_ISL_1087346, EPI_ISL_1087347, EPI_ISL_1087348, EPI_ISL_1087349, EPI_ISL_1087350, EPI_ISL_1087351, EPI_ISL_1087352 | | | |
| see above | NYU Langone Health | Departments of Pathology and Medicine, New York University School of Medicine | Adriana Heguy, Dacia Dimartino, Emily Guzman, Christian Marier, Peter Meyn, Sitharam Ramaswami, Gael Westby, Paul Zappile, Yutong Zhang, Paolo Cotzia, Guiqing Wang |
| EPI_ISL_1087538, EPI_ISL_1087540, EPI_ISL_1087725, EPI_ISL_1087797, EPI_ISL_1087803, EPI_ISL_1087805, EPI_ISL_1087873, EPI_ISL_1087874, EPI_ISL_1087877, EPI_ISL_1087878, EPI_ISL_1087879, EPI_ISL_1087888, EPI_ISL_1087891, EPI_ISL_1087960, EPI_ISL_1087988, EPI_ISL_1087989, EPI_ISL_1087992, EPI_ISL_1088098, EPI_ISL_1088116, EPI_ISL_1088118, EPI_ISL_1088133, EPI_ISL_1088134, EPI_ISL_1088147, EPI_ISL_1088169, EPI_ISL_1088177, EPI_ISL_1088220, EPI_ISL_1088233 | | | |
| see above | Quest Diagnostics Incorporated | Respiratory Viruses Branch, Division of Viral Diseases, Centers for Disease Control and Prevention | Peter W. Cook, Dakota Howard, Dhwani Batra, Ben L. Rambo-Martin, S. H. Rosenthal, A. Gerasimova, R. M. Kagan, B. Anderson, M. Hua, Y. Liu, L.E. Bernstein, K.E. Livingston, A. Perez, I. A. Shlyakhter, R. V. Rolando, R. Owen, P. Tanpaiboon, F. Lacbawan, Clinton R. Paden, Suxiang Tong, Duncan MacCannell |
| EPI_ISL_1088840, EPI_ISL_1089387 | Helix / Illumina | Respiratory Viruses Branch, Division of Viral Diseases, Centers for Disease Control and Prevention | Peter W. Cook, Dakota Howard, Dhwani Batra, Ben L. Rambo-Martin, Eileen de Feo, Jan Antico, Christine Tran, Matthew Tolentino, Shannon Wickline, Kim Gietzen, Brad Sickler, Jingtao Liu, Eric Allen, Phil Febbo, Summer Galloway, Nicole L. Washington, Simon White, Geraint Levan, Kelly Schiabor Barrett, Elizabeth Cirulli, Alexandre Bolze, Ary Ascencio, Charlotte Rivera-Garcia, Ryan Cho, Jason Nguyen, Sherry Wang, Jimmy Ramirez, Tyler Cassens, Efren Sandoval, Magnus Isaksson, William Lee, David Becker, Marc Laurent, James Lu, Clinton R. Paden, Suxiang Tong, Duncan MacCannell |
| EPI_ISL_1090341, EPI_ISL_1090405, EPI_ISL_1090414, EPI_ISL_1090429, EPI_ISL_1090456, EPI_ISL_1090461, EPI_ISL_1090465, EPI_ISL_1090466, EPI_ISL_1090469, EPI_ISL_1090518, EPI_ISL_1090578, EPI_ISL_1090599, EPI_ISL_1090617, EPI_ISL_1090720, EPI_ISL_1090722, EPI_ISL_1090755, EPI_ISL_1090757, EPI_ISL_1090870, EPI_ISL_1090901, EPI_ISL_1090958 | | | |
| see above | Quest Diagnostics Incorporated | Respiratory Viruses Branch, Division of Viral Diseases, Centers for Disease Control and Prevention | Peter W. Cook, Dakota Howard, Dhwani Batra, Ben L. Rambo-Martin, S. H. Rosenthal, A. Gerasimova, R. M. Kagan, B. Anderson, M. Hua, Y. Liu, L.E. Bernstein, K.E. Livingston, A. Perez, I. A. Shlyakhter, R. V. Rolando, R. Owen, P. Tanpaiboon, F. Lacbawan, Clinton R. Paden, Suxiang Tong, Duncan MacCannell |
| EPI_ISL_1091898, EPI_ISL_1091899, EPI_ISL_1091900 | New York Presbetyrian / Weill Cornell Medicine | Grubaugh Lab - Yale School of Public Health | Mary Petrone, Joseph Fauver, Tara Alpert, Anderson Brito, Mallery Breban, Anne Wyllie, Chantal Vogels, Annie Watkins, Chaney Kalinich, Isabel Ott, Nathan Grubaugh |
| EPI_ISL_1091901, EPI_ISL_1091902 | Murphy Medical Associates | Grubaugh Lab - Yale School of Public Health | Mary Petrone, Joseph Fauver, Caleb Neal, Steven Murphy, Chantal Vogels, Mallery Breban, Annie Watkins, Tara Alpert, Nathan Grubaugh |
| EPI_ISL_1091903, EPI_ISL_1091904, EPI_ISL_1091905, EPI_ISL_1091906, EPI_ISL_1091907, EPI_ISL_1091908, EPI_ISL_1091909, EPI_ISL_1091910, EPI_ISL_1091911, EPI_ISL_1091912, EPI_ISL_1091913, EPI_ISL_1091914, EPI_ISL_1091915 | | | |
| see above | New York Presbetyrian / Weill Cornell Medicine | Grubaugh Lab - Yale School of Public Health | Mary Petrone, Joseph Fauver, Tara Alpert, Anderson Brito, Mallery Breban, Anne Wyllie, Chantal Vogels, Annie Watkins, Chaney Kalinich, Isabel Ott, Nathan Grubaugh |
| EPI_ISL_1091916, EPI_ISL_1091917, EPI_ISL_1091918 | Murphy Medical Associates | Grubaugh Lab - Yale School of Public Health | Mary Petrone, Joseph Fauver, Caleb Neal, Steven Murphy, Chantal Vogels, Mallery Breban, Annie Watkins, Tara Alpert, Nathan Grubaugh |
| EPI_ISL_1094217, EPI_ISL_1094219, EPI_ISL_1094227, EPI_ISL_1094285, EPI_ISL_1094286, EPI_ISL_1094307, EPI_ISL_1094322, EPI_ISL_1094359, EPI_ISL_1094368, EPI_ISL_1094370, EPI_ISL_1094378, EPI_ISL_1094433, EPI_ISL_1094434, EPI_ISL_1094435, EPI_ISL_1094436, EPI_ISL_1094437, EPI_ISL_1094438, EPI_ISL_1094439, EPI_ISL_1094440, EPI_ISL_1094441, EPI_ISL_1094443, EPI_ISL_1094444, EPI_ISL_1095131, EPI_ISL_1095132, EPI_ISL_1095134, EPI_ISL_1095135, EPI_ISL_1095136, EPI_ISL_1095138, EPI_ISL_1095139, EPI_ISL_1095140, EPI_ISL_1095141, EPI_ISL_1095143, EPI_ISL_1095144, EPI_ISL_1095145, EPI_ISL_1095146, EPI_ISL_1095147, EPI_ISL_1095148, EPI_ISL_1095150, EPI_ISL_1095151, EPI_ISL_1095152, EPI_ISL_1095153, EPI_ISL_1095154, EPI_ISL_1095155, EPI_ISL_1095156, EPI_ISL_1095157, EPI_ISL_1095158, EPI_ISL_1095159, EPI_ISL_1095160, EPI_ISL_1095161, EPI_ISL_1095162, EPI_ISL_1095163, EPI_ISL_1095164, EPI_ISL_1095165, EPI_ISL_1095166, EPI_ISL_1095167, EPI_ISL_1095168 | | | |
| see above | NYSDOH Wadsworth Center, Virology Lab | Respiratory Viruses Branch, Division of Viral Diseases, Centers for Disease Control and Prevention | Krista Queen, Yan Li, Ying Tao, Jing Zhang, Anna Uehara, Anna Montmayeur, Clinton R. Paden, Peter W. Cook, Rachel Marine, Mili Sheth, Jasmine Padilla, Sarah Nobles, Mark Burroughs, Lori Rowe, Haibin Wang, Ben L. Rambo-Martin, Dhwani Batra, Justin Lee, Suxiang Tong |
| EPI_ISL_1097781, EPI_ISL_1097782, EPI_ISL_1097783, EPI_ISL_1097784, EPI_ISL_1097785, EPI_ISL_1097786, EPI_ISL_1097787, EPI_ISL_1097788, EPI_ISL_1097789, EPI_ISL_1097790, EPI_ISL_1097791, EPI_ISL_1097792, EPI_ISL_1097793, EPI_ISL_1097794, EPI_ISL_1097795, EPI_ISL_1097796, EPI_ISL_1097797, EPI_ISL_1097798, EPI_ISL_1097799, EPI_ISL_1097800, EPI_ISL_1097801, EPI_ISL_1097802, EPI_ISL_1097803, EPI_ISL_1097804, EPI_ISL_1097805, EPI_ISL_1097806, EPI_ISL_1097807, EPI_ISL_1097808, EPI_ISL_1097809, EPI_ISL_1097810, EPI_ISL_1097811, EPI_ISL_1097812, EPI_ISL_1097813, EPI_ISL_1097814, EPI_ISL_1097815, EPI_ISL_1097816, EPI_ISL_1097817, EPI_ISL_1097818, EPI_ISL_1097819, EPI_ISL_1097820, EPI_ISL_1097821, EPI_ISL_1097822, EPI_ISL_1097823, EPI_ISL_1097824, EPI_ISL_1097825, EPI_ISL_1097826, EPI_ISL_1097827, EPI_ISL_1097828, EPI_ISL_1097829, EPI_ISL_1097830, EPI_ISL_1097831, EPI_ISL_1097832, EPI_ISL_1097833, EPI_ISL_1097834, EPI_ISL_1097835, EPI_ISL_1097836, EPI_ISL_1097837, EPI_ISL_1097838, EPI_ISL_1097839, EPI_ISL_1097840, EPI_ISL_1097841, EPI_ISL_1097842, EPI_ISL_1097843, EPI_ISL_1097844, EPI_ISL_1097845, EPI_ISL_1097846, EPI_ISL_1097847, EPI_ISL_1097848, EPI_ISL_1097849, EPI_ISL_1097850, EPI_ISL_1097851, EPI_ISL_1097852, EPI_ISL_1097853, EPI_ISL_1097854, EPI_ISL_1097855, EPI_ISL_1097856, EPI_ISL_1097857, EPI_ISL_1097858, EPI_ISL_1097859, EPI_ISL_1097860, EPI_ISL_1097861, EPI_ISL_1097862, EPI_ISL_1097863, EPI_ISL_1097864, EPI_ISL_1097865, EPI_ISL_1097866, EPI_ISL_1097867, EPI_ISL_1097868, EPI_ISL_1097869, EPI_ISL_1097870, EPI_ISL_1097871, EPI_ISL_1097872, EPI_ISL_1097873, EPI_ISL_1097874, EPI_ISL_1097875, EPI_ISL_1097876, EPI_ISL_1097877, EPI_ISL_1097878, EPI_ISL_1097879, EPI_ISL_1097880, EPI_ISL_1097881, EPI_ISL_1097882, EPI_ISL_1097883, EPI_ISL_1097884, EPI_ISL_1097885, EPI_ISL_1097886, EPI_ISL_1097887, EPI_ISL_1097888, EPI_ISL_1097889, EPI_ISL_1097890, EPI_ISL_1097891, EPI_ISL_1097892, EPI_ISL_1097893, EPI_ISL_1097894, EPI_ISL_1097895, EPI_ISL_1097896, EPI_ISL_1097897, EPI_ISL_1097898, EPI_ISL_1097899, EPI_ISL_1097900, EPI_ISL_1097901, EPI_ISL_1097902, EPI_ISL_1097903, EPI_ISL_1097904, EPI_ISL_1097905, EPI_ISL_1097906, EPI_ISL_1097907, EPI_ISL_1097908, EPI_ISL_1097909, EPI_ISL_1097910, EPI_ISL_1097911, EPI_ISL_1097912, EPI_ISL_1097913, EPI_ISL_1097914, EPI_ISL_1097915, EPI_ISL_1097916, EPI_ISL_1097917, EPI_ISL_1097918, EPI_ISL_1097919, EPI_ISL_1097920, EPI_ISL_1097921, EPI_ISL_1097922, EPI_ISL_1097923, EPI_ISL_1097924, EPI_ISL_1097925, EPI_ISL_1097926, EPI_ISL_1097927, EPI_ISL_1097928, EPI_ISL_1097929, EPI_ISL_1097930, EPI_ISL_1097931, EPI_ISL_1097932, EPI_ISL_1097933, EPI_ISL_1097934, EPI_ISL_1097935, EPI_ISL_1097936, EPI_ISL_1097937, EPI_ISL_1097938, EPI_ISL_1097939, EPI_ISL_1097940, EPI_ISL_1097941, EPI_ISL_1097942, EPI_ISL_1097943, EPI_ISL_1097944, EPI_ISL_1097945, EPI_ISL_1097946, EPI_ISL_1097947, EPI_ISL_1097948, EPI_ISL_1097949, EPI_ISL_1097950, EPI_ISL_1097951, EPI_ISL_1097952, EPI_ISL_1097953, EPI_ISL_1097954, EPI_ISL_1097955, EPI_ISL_1097956, EPI_ISL_1097957, EPI_ISL_1097958, EPI_ISL_1097959, EPI_ISL_1097960, EPI_ISL_1097961, EPI_ISL_1097962, EPI_ISL_1097963, EPI_ISL_1097964, EPI_ISL_1097965, EPI_ISL_1097966, EPI_ISL_1097967, EPI_ISL_1097968, EPI_ISL_1097969, EPI_ISL_1097970, EPI_ISL_1097971, EPI_ISL_1097972, EPI_ISL_1097973, EPI_ISL_1097974, EPI_ISL_1097975, EPI_ISL_1097976, EPI_ISL_1097977, EPI_ISL_1097978, EPI_ISL_1097979, EPI_ISL_1097980, EPI_ISL_1097981, EPI_ISL_1097982, EPI_ISL_1097983, EPI_ISL_1097984, EPI_ISL_1097985, EPI_ISL_1097986, EPI_ISL_1097987, EPI_ISL_1097988, EPI_ISL_1097989, EPI_ISL_1097990, EPI_ISL_1097991, EPI_ISL_1097992, EPI_ISL_1097993, EPI_ISL_1097994, EPI_ISL_1097995, EPI_ISL_1097996, EPI_ISL_1097997, EPI_ISL_1097998, EPI_ISL_1097999, EPI_ISL_1098000, EPI_ISL_1098001, EPI_ISL_1098002, EPI_ISL_1098003, EPI_ISL_1098004, EPI_ISL_1098005, EPI_ISL_1098006, EPI_ISL_1098007, EPI_ISL_1098008, EPI_ISL_1098009, EPI_ISL_1098010, EPI_ISL_1098011, EPI_ISL_1098012, EPI_ISL_1098013, EPI_ISL_1098014, EPI_ISL_1098015, EPI_ISL_1098016, EPI_ISL_1098017, EPI_ISL_1098018, EPI_ISL_1098019, EPI_ISL_1098020, EPI_ISL_1098021, EPI_ISL_1098022, EPI_ISL_1098023, EPI_ISL_1098024, EPI_ISL_1098025, EPI_ISL_1098026, EPI_ISL_1098027, EPI_ISL_1098028, EPI_ISL_1098029, EPI_ISL_1098030, EPI_ISL_1098031, EPI_ISL_1098032, EPI_ISL_1098033, EPI_ISL_1098034, EPI_ISL_1098035, EPI_ISL_1098036, EPI_ISL_1098037, EPI_ISL_1098038, EPI_ISL_1098039, EPI_ISL_1098040, EPI_ISL_1098041, EPI_ISL_1098042, EPI_ISL_1098043, EPI_ISL_1098044, EPI_ISL_1098045, EPI_ISL_1098046, EPI_ISL_1098047, EPI_ISL_1098048, EPI_ISL_1098049, EPI_ISL_1098050, EPI_ISL_1098051, EPI_ISL_1098052, EPI_ISL_1098053, EPI_ISL_1098054, EPI_ISL_1098055, EPI_ISL_1098056, EPI_ISL_1098057, EPI_ISL_1098058, EPI_ISL_1098059, EPI_ISL_1098060, EPI_ISL_1098061, EPI_ISL_1098062, EPI_ISL_1098063, EPI_ISL_1098064, EPI_ISL_1098065, EPI_ISL_1098066, EPI_ISL_1098067, EPI_ISL_1098068, EPI_ISL_1098069, EPI_ISL_1098070, EPI_ISL_1098071, EPI_ISL_1098072, EPI_ISL_1098073, EPI_ISL_1098074, EPI_ISL_1098075, EPI_ISL_1098076, EPI_ISL_1098077, EPI_ISL_1098078, EPI_ISL_1098079, EPI_ISL_1098080, EPI_ISL_1098081, EPI_ISL_1098082, EPI_ISL_1098083, EPI_ISL_1098084, EPI_ISL_1098085, EPI_ISL_1098086, EPI_ISL_1098087, EPI_ISL_1098088, EPI_ISL_1098089, EPI_ISL_1098090, EPI_ISL_1098091, EPI_ISL_1098092, EPI_ISL_1098093, EPI_ISL_1098094, EPI_ISL_1098095, EPI_ISL_1098096, EPI_ISL_1098097, EPI_ISL_1098098, EPI_ISL_1098099, EPI_ISL_1098100, EPI_ISL_1098101, EPI_ISL_1098102, EPI_ISL_1098103, EPI_ISL_1098104, EPI_ISL_1098105, EPI_ISL_1098106, EPI_ISL_1098107, EPI_ISL_1098108, EPI_ISL_1098109, EPI_ISL_1098110, EPI_ISL_1098111, EPI_ISL_1098112, EPI_ISL_1098113, EPI_ISL_1098114, EPI_ISL_1098115, EPI_ISL_1098116, EPI_ISL_1098117, EPI_ISL_1098118, EPI_ISL_1098119, EPI_ISL_1098120, EPI_ISL_1098121, EPI_ISL_1098122, EPI_ISL_1098123, EPI_ISL_1098124, EPI_ISL_1098125, EPI_ISL_1098126, EPI_ISL_1098127, EPI_ISL_1098128, EPI_ISL_1098129, EPI_ISL_1098130, EPI_ISL_1098131, EPI_ISL_1098132, EPI_ISL_1098133, EPI_ISL_1098134, EPI_ISL_1098135, EPI_ISL_1098136, EPI_ISL_1098137, | | | |

EPI_ISL_1098138, EPI_ISL_1098139, EPI_ISL_1098140, EPI_ISL_1098141, EPI_ISL_1098142, EPI_ISL_1098143, EPI_ISL_1098144, EPI_ISL_1098145, EPI_ISL_1098146, EPI_ISL_1098147, EPI_ISL_1098148, EPI_ISL_1098149, EPI_ISL_1098150, EPI_ISL_1098151, EPI_ISL_1098152, EPI_ISL_1098153, EPI_ISL_1098154, EPI_ISL_1098155, EPI_ISL_1098156, EPI_ISL_1098157, EPI_ISL_1098158, EPI_ISL_1098159, EPI_ISL_1098160, EPI_ISL_1098161, EPI_ISL_1098162, EPI_ISL_1098163, EPI_ISL_1098164, EPI_ISL_1098165, EPI_ISL_1098166, EPI_ISL_1098167, EPI_ISL_1098168, EPI_ISL_1098169, EPI_ISL_1098170, EPI_ISL_1098171, EPI_ISL_1098172, EPI_ISL_1098173, EPI_ISL_1098174, EPI_ISL_1098175, EPI_ISL_1098176, EPI_ISL_1098177, EPI_ISL_1098178, EPI_ISL_1098179, EPI_ISL_1098180, EPI_ISL_1098181, EPI_ISL_1098182, EPI_ISL_1098183, EPI_ISL_1098184, EPI_ISL_1098185, EPI_ISL_1098186, EPI_ISL_1098187, EPI_ISL_1098188, EPI_ISL_1098189, EPI_ISL_1098190, EPI_ISL_1098191, EPI_ISL_1098192, EPI_ISL_1098193, EPI_ISL_1098194, EPI_ISL_1098195, EPI_ISL_1098196, EPI_ISL_1098197, EPI_ISL_1098198, EPI_ISL_1098199, EPI_ISL_1098200, EPI_ISL_1098201, EPI_ISL_1098202, EPI_ISL_1098203, EPI_ISL_1098204, EPI_ISL_1098205, EPI_ISL_1098206, EPI_ISL_1098207, EPI_ISL_1098208, EPI_ISL_1098209, EPI_ISL_1098210, EPI_ISL_1098211, EPI_ISL_1098212, EPI_ISL_1098213, EPI_ISL_1098214, EPI_ISL_1098215, EPI_ISL_1098216, EPI_ISL_1098217, EPI_ISL_1098218, EPI_ISL_1098219, EPI_ISL_1098220, EPI_ISL_1098221, EPI_ISL_1098222, EPI_ISL_1098223, EPI_ISL_1098224, EPI_ISL_1098225, EPI_ISL_1098226, EPI_ISL_1098227, EPI_ISL_1098228, EPI_ISL_1098229, EPI_ISL_1098230, EPI_ISL_1098231, EPI_ISL_1098232, EPI_ISL_1098233, EPI_ISL_1098234, EPI_ISL_1098235, EPI_ISL_1098236, EPI_ISL_1098237, EPI_ISL_1098238, EPI_ISL_1098239, EPI_ISL_1098240, EPI_ISL_1098241, EPI_ISL_1098242, EPI_ISL_1098243, EPI_ISL_1098244, EPI_ISL_1098245, EPI_ISL_1098246, EPI_ISL_1098247, EPI_ISL_1098248, EPI_ISL_1098249, EPI_ISL_1098250, EPI_ISL_1098251, EPI_ISL_1098252, EPI_ISL_1098253, EPI_ISL_1098254, EPI_ISL_1098255, EPI_ISL_1098256, EPI_ISL_1098257, EPI_ISL_1098258, EPI_ISL_1098259, EPI_ISL_1098260, EPI_ISL_1098261, EPI_ISL_1098262, EPI_ISL_1098263, EPI_ISL_1098264, EPI_ISL_1098265, EPI_ISL_1098266, EPI_ISL_1098267, EPI_ISL_1098268, EPI_ISL_1098269, EPI_ISL_1098270, EPI_ISL_1098271, EPI_ISL_1098272, EPI_ISL_1098273, EPI_ISL_1098274, EPI_ISL_1098275, EPI_ISL_1098276, EPI_ISL_1098277, EPI_ISL_1098278, EPI_ISL_1098279, EPI_ISL_1098280, EPI_ISL_1098281, EPI_ISL_1098282, EPI_ISL_1098283, EPI_ISL_1098284, EPI_ISL_1098285, EPI_ISL_1098286, EPI_ISL_1098287, EPI_ISL_1098288, EPI_ISL_1098289, EPI_ISL_1098290, EPI_ISL_1098291, EPI_ISL_1098292, EPI_ISL_1098293, EPI_ISL_1098294, EPI_ISL_1098295, EPI_ISL_1098296, EPI_ISL_1098297, EPI_ISL_1098298, EPI_ISL_1098299, EPI_ISL_1098300, EPI_ISL_1098301, EPI_ISL_1098302, EPI_ISL_1098303, EPI_ISL_1098304, EPI_ISL_1098305, EPI_ISL_1098306, EPI_ISL_1098307, EPI_ISL_1098308, EPI_ISL_1098309, EPI_ISL_1098310, EPI_ISL_1098311, EPI_ISL_1098312, EPI_ISL_1098313, EPI_ISL_1098314, EPI_ISL_1098315, EPI_ISL_1098316, EPI_ISL_1098317, EPI_ISL_1098318, EPI_ISL_1098319, EPI_ISL_1098320, EPI_ISL_1098321, EPI_ISL_1098322, EPI_ISL_1098323, EPI_ISL_1098324, EPI_ISL_1098325, EPI_ISL_1098326, EPI_ISL_1098327, EPI_ISL_1098328, EPI_ISL_1098329, EPI_ISL_1098330, EPI_ISL_1098331, EPI_ISL_1098332, EPI_ISL_1098333, EPI_ISL_1098334, EPI_ISL_1098335, EPI_ISL_1098336, EPI_ISL_1098337, EPI_ISL_1098338, EPI_ISL_1098339, EPI_ISL_1098340, EPI_ISL_1098341, EPI_ISL_1098342, EPI_ISL_1098343, EPI_ISL_1098344, EPI_ISL_1098345, EPI_ISL_1098346, EPI_ISL_1098347, EPI_ISL_1098348, EPI_ISL_1098349, EPI_ISL_1098350, EPI_ISL_1098351, EPI_ISL_1098352, EPI_ISL_1098353, EPI_ISL_1098354, EPI_ISL_1098355, EPI_ISL_1098356, EPI_ISL_1098357, EPI_ISL_1098358, EPI_ISL_1098359, EPI_ISL_1098360, EPI_ISL_1098361, EPI_ISL_1098362, EPI_ISL_1098363, EPI_ISL_1098364, EPI_ISL_1098365, EPI_ISL_1098366, EPI_ISL_1098367, EPI_ISL_1098368, EPI_ISL_1098369, EPI_ISL_1098370, EPI_ISL_1098371, EPI_ISL_1098372, EPI_ISL_1098373, EPI_ISL_1098374, EPI_ISL_1098375, EPI_ISL_1098376, EPI_ISL_1098377, EPI_ISL_1098378, EPI_ISL_1098379, EPI_ISL_1098380, EPI_ISL_1098381, EPI_ISL_1098382, EPI_ISL_1098383, EPI_ISL_1098384, EPI_ISL_1098385, EPI_ISL_1098386, EPI_ISL_1098387, EPI_ISL_1098388, EPI_ISL_1098389, EPI_ISL_1098390, EPI_ISL_1098391, EPI_ISL_1098392, EPI_ISL_1098393, EPI_ISL_1098394, EPI_ISL_1098395, EPI_ISL_1098396, EPI_ISL_1098397, EPI_ISL_1098398, EPI_ISL_1098399, EPI_ISL_1098400, EPI_ISL_1098401, EPI_ISL_1098402, EPI_ISL_1098403, EPI_ISL_1098404, EPI_ISL_1098405, EPI_ISL_1098406, EPI_ISL_1098407, EPI_ISL_1098408, EPI_ISL_1098409, EPI_ISL_1098410, EPI_ISL_1098411, EPI_ISL_1098412, EPI_ISL_1098413, EPI_ISL_1098414, EPI_ISL_1098415, EPI_ISL_1098416, EPI_ISL_1098417, EPI_ISL_1098418, EPI_ISL_1098419, EPI_ISL_1098420, EPI_ISL_1098421, EPI_ISL_1098422, EPI_ISL_1098423, EPI_ISL_1098424, EPI_ISL_1098425, EPI_ISL_1098426, EPI_ISL_1098427, EPI_ISL_1098428, EPI_ISL_1098429, EPI_ISL_1098430, EPI_ISL_1098431, EPI_ISL_1098432, EPI_ISL_1098433, EPI_ISL_1098434, EPI_ISL_1098435, EPI_ISL_1098436, EPI_ISL_1098437, EPI_ISL_1098438, EPI_ISL_1098439, EPI_ISL_1098440, EPI_ISL_1098441, EPI_ISL_1098442, EPI_ISL_1098443, EPI_ISL_1098444, EPI_ISL_1098445, EPI_ISL_1098446, EPI_ISL_1098447, EPI_ISL_1098448, EPI_ISL_1098449, EPI_ISL_1098450, EPI_ISL_1098451, EPI_ISL_1098452, EPI_ISL_1098453, EPI_ISL_1098454, EPI_ISL_1098455, EPI_ISL_1098456, EPI_ISL_1098457, EPI_ISL_1098458, EPI_ISL_1098459, EPI_ISL_1098460, EPI_ISL_1098461, EPI_ISL_1098462, EPI_ISL_1098463, EPI_ISL_1098464, EPI_ISL_1098465, EPI_ISL_1098466, EPI_ISL_1098467, EPI_ISL_1098468, EPI_ISL_1098469, EPI_ISL_1098470, EPI_ISL_1098471, EPI_ISL_1098472, EPI_ISL_1098473, EPI_ISL_1098474, EPI_ISL_1098475, EPI_ISL_1098476, EPI_ISL_1098477, EPI_ISL_1098478, EPI_ISL_1098479, EPI_ISL_1098480, EPI_ISL_1098481, EPI_ISL_1098482, EPI_ISL_1098483, EPI_ISL_1098484, EPI_ISL_1098485, EPI_ISL_1098486, EPI_ISL_1098487, EPI_ISL_1098488, EPI_ISL_1098489, EPI_ISL_1098490, EPI_ISL_1098491, EPI_ISL_1098492, EPI_ISL_1098493, EPI_ISL_1098494, EPI_ISL_1098495, EPI_ISL_1098496, EPI_ISL_1098497, EPI_ISL_1098498, EPI_ISL_1098499, EPI_ISL_1098500, EPI_ISL_1098501, EPI_ISL_1098502, EPI_ISL_1098503, EPI_ISL_1098504, EPI_ISL_1098505, EPI_ISL_1098506, EPI_ISL_1098507, EPI_ISL_1098508, EPI_ISL_1098509, EPI_ISL_1098510, EPI_ISL_1098511, EPI_ISL_1098512, EPI_ISL_1098513, EPI_ISL_1098514, EPI_ISL_1098515, EPI_ISL_1098516, EPI_ISL_1098517, EPI_ISL_1098518, EPI_ISL_1098519, EPI_ISL_1098520, EPI_ISL_1098521, EPI_ISL_1098522, EPI_ISL_1098523, EPI_ISL_1098524, EPI_ISL_1098525, EPI_ISL_1098526, EPI_ISL_1098527, EPI_ISL_1098528, EPI_ISL_1098529, EPI_ISL_1098530, EPI_ISL_1098531, EPI_ISL_1098532, EPI_ISL_1098533, EPI_ISL_1098534, EPI_ISL_1098535, EPI_ISL_1098536, EPI_ISL_1098537, EPI_ISL_1098538, EPI_ISL_1098539, EPI_ISL_1098540, EPI_ISL_1098541, EPI_ISL_1098542, EPI_ISL_1098543, EPI_ISL_1098544, EPI_ISL_1098545, EPI_ISL_1098546, EPI_ISL_1098547, EPI_ISL_1098548, EPI_ISL_1098549, EPI_ISL_1098550, EPI_ISL_1098551, EPI_ISL_1098552, EPI_ISL_1098553, EPI_ISL_1098554, EPI_ISL_1098555, EPI_ISL_1098556, EPI_ISL_1098557, EPI_ISL_1098558, EPI_ISL_1098559, EPI_ISL_1098560, EPI_ISL_1098561, EPI_ISL_1098562, EPI_ISL_1098563, EPI_ISL_1098564, EPI_ISL_1098565, EPI_ISL_1098566, EPI_ISL_1098567, EPI_ISL_1098568, EPI_ISL_1098569, EPI_ISL_1098570, EPI_ISL_1098571, EPI_ISL_1098572, EPI_ISL_1098573, EPI_ISL_1098574, EPI_ISL_1098575, EPI_ISL_1098576, EPI_ISL_1098577, EPI_ISL_1098578, EPI_ISL_1098579, EPI_ISL_1098580, EPI_ISL_1098581, EPI_ISL_1098582, EPI_ISL_1098583, EPI_ISL_1098584, EPI_ISL_1098585, EPI_ISL_1098586, EPI_ISL_1098587, EPI_ISL_1098588, EPI_ISL_1098589, EPI_ISL_1098590, EPI_ISL_1098591, EPI_ISL_1098592, EPI_ISL_1098593, EPI_ISL_1098594, EPI_ISL_1098595, EPI_ISL_1098596, EPI_ISL_1098597, EPI_ISL_1098598, EPI_ISL_1098599, EPI_ISL_1098600, EPI_ISL_1098601

| | | | |
|---|---|---|---|
| see above | Pandemic Response Lab - NYC | Pandemic Response Lab, R&D | Henry Lee, Michael Hammerling, Melissa Hopkins, Cybill del Castillo, Shinyoung Clair Kang, William Ward, Pradeep Bugga, Haiping Hao, Jon Laurent |
| EPI_ISL_1110651 | Helix/Illumina | Respiratory Viruses Branch, Division of Viral Diseases, Centers for Disease Control and Prevention | Peter W. Cook, Dakota Howard, Dhwani Batra, Ben L. Rambo-Martin, Eileen de Feo, Jan Antico, Christine Tran, Matthew Tolentino, Shannon Wickline, Kim Gietzen, Brad Sickler, Jingtao Liu, Eric Allen, Phil Febbo, Summer Galloway, Nicole L. Washington, Simon White, Geraint Levan, Kelly Schiabor Barrett, Elizabeth Cirulli, Alexandre Bolze, Ary Ascencio, Charlotte Rivera-Garcia, Ryan Cho, Jason Nguyen, Sherry Wang, Jimmy Ramirez, Tyler Cassens, Efren Sandoval, Magnus Isaksson, William Lee, David Becker, Marc Laurent, James Lu, Clinton R. Paden, Suxiang Tong, Duncan MacCannell |
| EPI_ISL_1112936 | Murphy Medical Associates | Grubaugh Lab - Yale School of Public Health | Mary Petrone, Joseph Fauver, Caleb Neal, Steven Murphy, Chantal Vogels, Mallery Breban, Annie Watkins, Tara Alpert, Nathan Grubaugh |
| EPI_ISL_1113774, EPI_ISL_1113827, EPI_ISL_1113838, EPI_ISL_1113857, EPI_ISL_1114014, EPI_ISL_1114067, EPI_ISL_1114130, EPI_ISL_1114133 | Quest Diagnostics Incorporated | Respiratory Viruses Branch, Division of Viral Diseases, Centers for Disease Control and Prevention | Peter W. Cook, Dakota Howard, Dhwani Batra, Ben L. Rambo-Martin, S. H. Rosenthal, A. Gerasimova, R. M. Kagan, B. Anderson, M. Hua, Y. Liu, L.E. Bernstein, K.E. Livingston, A. Perez, I. A. Shlyakhter, R. V. Rolando, R. Owen, P. Tanpaiboon, F. Lacbawan, Clinton R. Paden, Suxiang Tong, Duncan MacCannell |
| EPI_ISL_1114469 | Department of Health and Mental Hygiene Public Health Laboratory | New York City Public Health Laboratory | Jade Wang, et al. |
| EPI_ISL_1114470, EPI_ISL_1114471 | Department of Health and Mental Hygiene Central Harlem | New York City Public Health Laboratory | Jade Wang, et al. |
| EPI_ISL_1114472, EPI_ISL_1114473, EPI_ISL_1114474, EPI_ISL_1114475 | Department of Health and Mental Hygiene Corona | New York City Public Health Laboratory | Jade Wang, et al. |
| EPI_ISL_1114476 | Department of Health and Mental Hygiene Public Health Laboratory | New York City Public Health Laboratory | Jade Wang, et al. |
| EPI_ISL_1114477, EPI_ISL_1114478, EPI_ISL_1114479, EPI_ISL_1114480, EPI_ISL_1114481 | Department of Health and Mental Hygiene Jamaica | New York City Public Health Laboratory | Jade Wang, et al. |
| EPI_ISL_1114482, EPI_ISL_1114483, EPI_ISL_1114484 | Department of Health and Mental Hygiene Public Health Laboratory | New York City Public Health Laboratory | Jade Wang, et al. |
| EPI_ISL_1114485 | Department of Health and Mental Hygiene Crown Heights | New York City Public Health Laboratory | Jade Wang, et al. |
| EPI_ISL_1114486 | Department of Health and Mental Hygiene Public Health Laboratory | New York City Public Health Laboratory | Jade Wang, et al. |
| EPI_ISL_1114487, EPI_ISL_1114488, EPI_ISL_1114489, EPI_ISL_1114490, EPI_ISL_1114491 | Department of Health and Mental Hygiene Morrisania | New York City Public Health Laboratory | Jade Wang, et al. |
| EPI_ISL_1114492, EPI_ISL_1114493 | Department of Health and Mental Hygiene Central Harlem | New York City Public Health Laboratory | Jade Wang, et al. |
| EPI_ISL_1114494, EPI_ISL_1114495, EPI_ISL_1114496 | Department of Health and Mental Hygiene Fort Greene | New York City Public Health Laboratory | Jade Wang, et al. |
| EPI_ISL_1114497, EPI_ISL_1114498 | Department of Health and Mental Hygiene Riverside | New York City Public Health Laboratory | Jade Wang, et al. |
| EPI_ISL_1114499 | Department of Health and Mental Hygiene Crown Heights | New York City Public Health Laboratory | Jade Wang, et al. |
| EPI_ISL_1114500, EPI_ISL_1114501 | Department of Health and Mental Hygiene Chelsea | New York City Public Health Laboratory | Jade Wang, et al. |
| EPI_ISL_1114502 | Department of Health and Mental Hygiene Crown Heights | New York City Public Health Laboratory | Jade Wang, et al. |
| EPI_ISL_1114503 | Department of Health and Mental Hygiene Riverside | New York City Public Health Laboratory | Jade Wang, et al. |
| EPI_ISL_1114504, EPI_ISL_1114505, EPI_ISL_1114506, EPI_ISL_1114507 | Department of Health and Mental Hygiene Corona | New York City Public Health Laboratory | Jade Wang, et al. |
| EPI_ISL_1114508 | Department of Health and Mental Hygiene Jamaica | New York City Public Health Laboratory | Jade Wang, et al. |
| EPI_ISL_1114509 | Department of Health and Mental Hygiene Chelsea | New York City Public Health Laboratory | Jade Wang, et al. |
| EPI_ISL_1114510 | Department of Health and Mental Hygiene Public Health Laboratory | New York City Public Health Laboratory | Jade Wang, et al. |
| EPI_ISL_1114511, EPI_ISL_1114512 | Department of Homeless Services | New York City Public Health Laboratory | Jade Wang, et al. |
| EPI_ISL_1114513, EPI_ISL_1114514, EPI_ISL_1114515 | Department of Health and Mental Hygiene Corona | New York City Public Health Laboratory | Jade Wang, et al. |
| EPI_ISL_1114516, EPI_ISL_1114517 | Department of Health and Mental Hygiene Crown Heights | New York City Public Health Laboratory | Jade Wang, et al. |

| | | | |
|---|---|---|---|
| EPI_ISL_1114518 | Department of Health and Mental Hygiene Jamaica | New York City Public Health Laboratory | Jade Wang, et al. |
| EPI_ISL_1114519, EPI_ISL_1114520, EPI_ISL_1114521, EPI_ISL_1114522, EPI_ISL_1114523 | Department of Health and Mental Hygiene Public Health Laboratory | New York City Public Health Laboratory | Jade Wang, et al. |
| EPI_ISL_1114524 | Department of Health and Mental Hygiene Chelsea | New York City Public Health Laboratory | Jade Wang, et al. |
| EPI_ISL_1114525, EPI_ISL_1114526, EPI_ISL_1114527, EPI_ISL_1114528, EPI_ISL_1114529, EPI_ISL_1114530 | OCME Office Of Chief Medical Examiner | New York City Public Health Laboratory | Jade Wang, et al. |
| EPI_ISL_1114531, EPI_ISL_1114535, EPI_ISL_1114536, EPI_ISL_1114548, EPI_ISL_1114561, EPI_ISL_1114564, EPI_ISL_1114571, EPI_ISL_1114575, EPI_ISL_1114576, EPI_ISL_1114578, EPI_ISL_1114582, EPI_ISL_1114583, EPI_ISL_1114587, EPI_ISL_1114593, EPI_ISL_1114596, EPI_ISL_1114601, EPI_ISL_1114604, EPI_ISL_1114606, EPI_ISL_1114607, EPI_ISL_1114610, EPI_ISL_1114617, EPI_ISL_1114620, EPI_ISL_1114623, EPI_ISL_1114640, EPI_ISL_1114655, EPI_ISL_1114661, EPI_ISL_1114663, EPI_ISL_1114670 | | | |
| see above | Quest Diagnostics Incorporated | Respiratory Viruses Branch, Division of Viral Diseases, Centers for Disease Control and Prevention | Peter W. Cook, Dakota Howard, Dhwani Batra, Ben L. Rambo-Martin, S. H. Rosenthal, A. Gerasimova, R. M. Kagan, B. Anderson, M. Hua, Y. Liu, L.E. Bernstein, K.E. Livingston, A. Perez, I. A. Shlyakhter, R. V. Rolando, R. Owen, P. Tanpaiboon, F. Lacbawan, Clinton R. Paden, Suxiang Tong, Duncan MacCannell |
| EPI_ISL_1116476, EPI_ISL_1116477, EPI_ISL_1116478, EPI_ISL_1116479, EPI_ISL_1116480, EPI_ISL_1116481, EPI_ISL_1116482, EPI_ISL_1116483, EPI_ISL_1116484, EPI_ISL_1116485, EPI_ISL_1116486, EPI_ISL_1116487, EPI_ISL_1116488, EPI_ISL_1116489, EPI_ISL_1116490, EPI_ISL_1116491, EPI_ISL_1116492, EPI_ISL_1116493, EPI_ISL_1116494, EPI_ISL_1116495, EPI_ISL_1116496, EPI_ISL_1116497, EPI_ISL_1116498, EPI_ISL_1116499, EPI_ISL_1116500, EPI_ISL_1116501, EPI_ISL_1116502, EPI_ISL_1116504, EPI_ISL_1116505, EPI_ISL_1116506, EPI_ISL_1116507, EPI_ISL_1116508, EPI_ISL_1116509, EPI_ISL_1116510, EPI_ISL_1116511, EPI_ISL_1116512, EPI_ISL_1116513, EPI_ISL_1116514, EPI_ISL_1116515, EPI_ISL_1116517, EPI_ISL_1116518, EPI_ISL_1116519, EPI_ISL_1116520, EPI_ISL_1116521, EPI_ISL_1116522, EPI_ISL_1116523, EPI_ISL_1116524, EPI_ISL_1116525, EPI_ISL_1116526, EPI_ISL_1116527, EPI_ISL_1116528, EPI_ISL_1116529, EPI_ISL_1116530, EPI_ISL_1116531, EPI_ISL_1116532, EPI_ISL_1116533, EPI_ISL_1116534, EPI_ISL_1116535, EPI_ISL_1116536, EPI_ISL_1116537, EPI_ISL_1116538, EPI_ISL_1116539, EPI_ISL_1116540, EPI_ISL_1116541, EPI_ISL_1116542, EPI_ISL_1116543, EPI_ISL_1116544, EPI_ISL_1116545, EPI_ISL_1116546, EPI_ISL_1116547, EPI_ISL_1116548, EPI_ISL_1116549, EPI_ISL_1116550, EPI_ISL_1116551, EPI_ISL_1116552, EPI_ISL_1116553, EPI_ISL_1116554 | | | |
| see above | NYU Langone Health | Departments of Pathology and Medicine, New York University School of Medicine | Adriana Heguy, Dacia Dimartino, Emily Guzman, Christian Marier, Peter Meyn, Sitharam Ramaswami, Gael Westby, Paul Zappile, Yutong Zhang, Paolo Cotzia, Guiqing Wang |
| EPI_ISL_1121887 | Department of Health & Mental Hygiene Corona | New York City Public Health Laboratory | Jade Wang, et al. |
| EPI_ISL_1121888 | Department of Health & Mental Hygiene Morrisania | New York City Public Health Laboratory | Jade Wang, et al. |
| EPI_ISL_1121889 | Department of Health & Mental Hygiene Fort Greene | New York City Public Health Laboratory | Jade Wang, et al. |
| EPI_ISL_1121890, EPI_ISL_1121891, EPI_ISL_1121892 | Department of Health & Mental Hygiene Crown Heights | New York City Public Health Laboratory | Jade Wang, et al. |
| EPI_ISL_1121893 | Department of Health & Mental Hygiene Central Harlem | New York City Public Health Laboratory | Jade Wang, et al. |
| EPI_ISL_1121894 | Department of Health & Mental Hygiene PHL | New York City Public Health Laboratory | Jade Wang, et al. |
| EPI_ISL_1121895, EPI_ISL_1121896, EPI_ISL_1121897, EPI_ISL_1121898 | OCME Office Of Chief Medical Examiner | New York City Public Health Laboratory | Jade Wang, et al. |
| EPI_ISL_1121899 | Department of Health & Mental Hygiene Corona | New York City Public Health Laboratory | Jade Wang, et al. |
| EPI_ISL_1121900 | Department of Health & Mental Hygiene Central Harlem | New York City Public Health Laboratory | Jade Wang, et al. |
| EPI_ISL_1121901, EPI_ISL_1121902, EPI_ISL_1121903 | Department of Health & Mental Hygiene Fort Greene | New York City Public Health Laboratory | Jade Wang, et al. |
| EPI_ISL_1121904 | Department of Health & Mental Hygiene Crown Heights | New York City Public Health Laboratory | Jade Wang, et al. |
| EPI_ISL_1121905, EPI_ISL_1121906, EPI_ISL_1121907, EPI_ISL_1121908 | Department of Health & Mental Hygiene Corona | New York City Public Health Laboratory | Jade Wang, et al. |
| EPI_ISL_1121909, EPI_ISL_1121910, EPI_ISL_1121911, EPI_ISL_1121912 | Department of Health & Mental Hygiene PHL | New York City Public Health Laboratory | Jade Wang, et al. |
| EPI_ISL_1121913 | Department of Health & Mental Hygiene Morrisania | New York City Public Health Laboratory | Jade Wang, et al. |
| EPI_ISL_1121914 | Department of Health & Mental Hygiene Riverside | New York City Public Health Laboratory | Jade Wang, et al. |
| EPI_ISL_1121915, EPI_ISL_1121916, EPI_ISL_1121917, EPI_ISL_1121918 | Department of Health & Mental Hygiene Jamaica | New York City Public Health Laboratory | Jade Wang, et al. |
| EPI_ISL_1121919, EPI_ISL_1121920, EPI_ISL_1121921, EPI_ISL_1121922, EPI_ISL_1121923 | Department of Health & Mental Hygiene Morrisania | New York City Public Health Laboratory | Jade Wang, et al. |
| EPI_ISL_1121924 | Department of Health & Mental Hygiene Jamaica | New York City Public Health Laboratory | Jade Wang, et al. |
| EPI_ISL_1121925 | Department of Health & Mental Hygiene Chelsea | New York City Public Health Laboratory | Jade Wang, et al. |
| EPI_ISL_1121926 | Department of Health & Mental Hygiene Morrisania | New York City Public Health Laboratory | Jade Wang, et al. |
| EPI_ISL_1121927, EPI_ISL_1121928 | Department of Health & Mental Hygiene Chelsea | New York City Public Health Laboratory | Jade Wang, et al. |
| EPI_ISL_1121929, EPI_ISL_1121930, EPI_ISL_1121931 | Department of Health & Mental Hygiene PHL | New York City Public Health Laboratory | Jade Wang, et al. |
| EPI_ISL_1121932, EPI_ISL_1121933, EPI_ISL_1121934, EPI_ISL_1121935, EPI_ISL_1121936 | Department of Health & Mental Hygiene Corona | New York City Public Health Laboratory | Jade Wang, et al. |
| EPI_ISL_1121937 | Department of Health & Mental Hygiene Fort Greene | New York City Public Health Laboratory | Jade Wang, et al. |
| EPI_ISL_1121938, EPI_ISL_1121939 | Department of Health & Mental Hygiene Crown Heights | New York City Public Health Laboratory | Jade Wang, et al. |
| EPI_ISL_1121940, EPI_ISL_1121941, EPI_ISL_1121942 | Department of Health & Mental Hygiene Morrisania | New York City Public Health Laboratory | Jade Wang, et al. |
| EPI_ISL_1121943 | Department of Health & Mental Hygiene Riverside | New York City Public Health Laboratory | Jade Wang, et al. |
| EPI_ISL_1121944 | Department of Health & Mental Hygiene Morrisania | New York City Public Health Laboratory | Jade Wang, et al. |
| EPI_ISL_1121945, EPI_ISL_1121946 | Department of Health & Mental Hygiene Central Harlem | New York City Public Health Laboratory | Jade Wang, et al. |
| EPI_ISL_1121947, EPI_ISL_1121948, EPI_ISL_1121949 | Department of Health & Mental Hygiene Morrisania | New York City Public Health Laboratory | Jade Wang, et al. |
| EPI_ISL_1121950 | Department of Health & Mental Hygiene Central Harlem | New York City Public Health Laboratory | Jade Wang, et al. |
| EPI_ISL_1121951, EPI_ISL_1121952, EPI_ISL_1121953, EPI_ISL_1121954, | Department of Health & Mental Hygiene Jamaica | New York City Public Health Laboratory | Jade Wang, et al. |

| | | | |
|---|---|---|---|
| EPI_ISL_1121955, EPI_ISL_1121956, EPI_ISL_1121957, EPI_ISL_1121958, EPI_ISL_1121959 | | | |
| EPI_ISL_1121960 | Department of Health & Mental Hygiene Chelsea | New York City Public Health Laboratory | Jade Wang, et al. |
| EPI_ISL_1133941, EPI_ISL_1134090, EPI_ISL_1134455, EPI_ISL_1134696 | Helix/Illumina | Respiratory Viruses Branch, Division of Viral Diseases, Centers for Disease Control and Prevention | Peter W. Cook, Dakota Howard, Dhwani Batra, Ben L. Rambo-Martin, Eileen de Feo, Jan Antico, Christine Tran, Matthew Tolentino, Shannon Wickline, Kim Gietzen, Brad Sickler, Jingtao Liu, Eric Allen, Phil Febbo, Summer Galloway, Nicole L. Washington, Simon White, Geraint Levan, Kelly Schiabor Barrett, Elizabeth Cirulli, Alexandre Bolze, Ary Ascencio, Charlotte Rivera-Garcia, Ryan Cho, Jason Nguyen, Sherry Wang, Jimmy Ramirez, Tyler Cassens, Efren Sandoval, Magnus Isaksson, William Lee, David Becker, Marc Laurent, James Lu, Clinton R. Paden, Suxiang Tong, Duncan MacCannell |
| EPI_ISL_1135292, EPI_ISL_1135297, EPI_ISL_1135299, EPI_ISL_1139206, EPI_ISL_1139211, EPI_ISL_1139216, EPI_ISL_1139242, EPI_ISL_1139248, EPI_ISL_1139270, EPI_ISL_1139338, EPI_ISL_1139340, EPI_ISL_1139373, EPI_ISL_1139666, EPI_ISL_1139759, EPI_ISL_1139818, EPI_ISL_1158932, EPI_ISL_1158939, EPI_ISL_1158947, EPI_ISL_1158948, EPI_ISL_1158955, EPI_ISL_1158956, EPI_ISL_1158961, EPI_ISL_1158963, EPI_ISL_1158970, EPI_ISL_1158977, EPI_ISL_1158992, EPI_ISL_1158995, EPI_ISL_1158996, EPI_ISL_1158998, EPI_ISL_1159010, EPI_ISL_1159013, EPI_ISL_1159014, EPI_ISL_1159016, EPI_ISL_1159018, EPI_ISL_1159024, EPI_ISL_1159035, EPI_ISL_1159036, EPI_ISL_1159042, EPI_ISL_1159045, EPI_ISL_1159056, EPI_ISL_1159059, EPI_ISL_1159062, EPI_ISL_1159066 | | | |
| see above | Quest Diagnostics Incorporated | Respiratory Viruses Branch, Division of Viral Diseases, Centers for Disease Control and Prevention | Peter W. Cook, Dakota Howard, Dhwani Batra, Ben L. Rambo-Martin, S. H. Rosenthal, A. Gerasimova, R. M. Kagan, B. Anderson, M. Hua, Y. Liu, L.E. Bernstein, K.E. Livingston, A. Perez, I. A. Shlyakhter, R. V. Rolando, R. Owen, P. Tanpaiboon, F. Lacbawan, Clinton R. Paden, Suxiang Tong, Duncan MacCannell |
| EPI_ISL_1159904, EPI_ISL_1159919, EPI_ISL_1159978, EPI_ISL_1160029, EPI_ISL_1160030, EPI_ISL_1160031, EPI_ISL_1160038, EPI_ISL_1160045, EPI_ISL_1160097, EPI_ISL_1160115, EPI_ISL_1160181, EPI_ISL_1160251, EPI_ISL_1160298, EPI_ISL_1160318, EPI_ISL_1160325, EPI_ISL_1160377, EPI_ISL_1160400, EPI_ISL_1160444, EPI_ISL_1160465, EPI_ISL_1160491, EPI_ISL_1160494, EPI_ISL_1160513, EPI_ISL_1160545, EPI_ISL_1160546, EPI_ISL_1160556, EPI_ISL_1160565, EPI_ISL_1160590, EPI_ISL_1160593, EPI_ISL_1160600, EPI_ISL_1160628, EPI_ISL_1160656, EPI_ISL_1160697, EPI_ISL_1160700, EPI_ISL_1160705, EPI_ISL_1160708, EPI_ISL_1160711, EPI_ISL_1160732, EPI_ISL_1160733, EPI_ISL_1160735, EPI_ISL_1160751, EPI_ISL_1160802, EPI_ISL_1160815, EPI_ISL_1160844, EPI_ISL_1160846, EPI_ISL_1160847, EPI_ISL_1160905, EPI_ISL_1160914, EPI_ISL_1160919, EPI_ISL_1160925, EPI_ISL_1160943, EPI_ISL_1160944, EPI_ISL_1160953, EPI_ISL_1160966, EPI_ISL_1161138, EPI_ISL_1161139, EPI_ISL_1161140, EPI_ISL_1161141, EPI_ISL_1161144, EPI_ISL_1161146, EPI_ISL_1161157, EPI_ISL_1161168, EPI_ISL_1161190, EPI_ISL_1161363, EPI_ISL_1161367, EPI_ISL_1161380, EPI_ISL_1161381, EPI_ISL_1161382, EPI_ISL_1161440, EPI_ISL_1161448, EPI_ISL_1161451, EPI_ISL_1161468, EPI_ISL_1161473, EPI_ISL_1161530, EPI_ISL_1161558, EPI_ISL_1161621, EPI_ISL_1161627, EPI_ISL_1161631, EPI_ISL_1161673, EPI_ISL_1161680, EPI_ISL_1161683, EPI_ISL_1161690, EPI_ISL_1161764, EPI_ISL_1161777, EPI_ISL_1161778, EPI_ISL_1161780, EPI_ISL_1161782, EPI_ISL_1161800, EPI_ISL_1161808, EPI_ISL_1161818, EPI_ISL_1161824, EPI_ISL_1161828, EPI_ISL_1161890, EPI_ISL_1161909, EPI_ISL_1161921, EPI_ISL_1161941, EPI_ISL_1161956, EPI_ISL_1162030, EPI_ISL_1162073, EPI_ISL_1162080, EPI_ISL_1162091, EPI_ISL_1162111, EPI_ISL_1162132, EPI_ISL_1162147, EPI_ISL_1162153, EPI_ISL_1162165, EPI_ISL_1162230, EPI_ISL_1162268, EPI_ISL_1162277, EPI_ISL_1162294, EPI_ISL_1162297, EPI_ISL_1162306, EPI_ISL_1162348, EPI_ISL_1162387, EPI_ISL_1162403, EPI_ISL_1162478, EPI_ISL_1162484, EPI_ISL_1162519, EPI_ISL_1162531, EPI_ISL_1162532, EPI_ISL_1162603, EPI_ISL_1162626, EPI_ISL_1162663, EPI_ISL_1162664, EPI_ISL_1162682, EPI_ISL_1162685, EPI_ISL_1162699, EPI_ISL_1162773, EPI_ISL_1162784, EPI_ISL_1162806, EPI_ISL_1162816, EPI_ISL_1162827, EPI_ISL_1162836, EPI_ISL_1162845, EPI_ISL_1162860, EPI_ISL_1162881, EPI_ISL_1162883, EPI_ISL_1162912, EPI_ISL_1162922, EPI_ISL_1162928, EPI_ISL_1162942, EPI_ISL_1162953, EPI_ISL_1162996, EPI_ISL_1163009, EPI_ISL_1163010, EPI_ISL_1163012, EPI_ISL_1163024, EPI_ISL_1163043, EPI_ISL_1163082, EPI_ISL_1163132, EPI_ISL_1163158, EPI_ISL_1163161, EPI_ISL_1163198, EPI_ISL_1163227, EPI_ISL_1163228, EPI_ISL_1163229, EPI_ISL_1163234, EPI_ISL_1163235, EPI_ISL_1163268, EPI_ISL_1163272, EPI_ISL_1163292, EPI_ISL_1163293, EPI_ISL_1163294, EPI_ISL_1163295, EPI_ISL_1163349, EPI_ISL_1163373, EPI_ISL_1163387, EPI_ISL_1163393, EPI_ISL_1163440, EPI_ISL_1163441, EPI_ISL_1163450, EPI_ISL_1163463, EPI_ISL_1163483 | | | |
| see above | Laboratory Corporation of America | Respiratory Viruses Branch, Division of Viral Diseases, Centers for Disease Control and Prevention | Peter W. Cook, Dakota Howard, Dhwani Batra, Ben L. Rambo-Martin, Minoo Agarwal, Eyad Almasri Debbie Boles, Ayla Burns, Nuthawin Charoensri, Oren Cohen, Susan Countryman, Mary Ann Cristobal, Bobbi Croy, Suzanne Dale, Hrushikesh Deshmukh, Amanda Douglas, Vincent Drouillon, Marcia Eisenberg, Howard Engler, Rama Ghatti, Prashant Gupta, Susan Hicks, Jake Humphrey, Lax Iyer, Manoj Jain, Mohan Kolli, Brian Krueger, Tim Kuphal, Stanley Letovsky, Michael Levandoski, Craig Lukasik, Jonathan Meltzer, Brian Norvell, Mindy Nye, Scott Parker, Christos Petropoulos, John Pruitt, Steven Ragan, Scott Ryan, Mike Sapeta, Jana Schroth, Suresh Babu Selvaraju, Goran Stevovic, Amanda Suchanek, Andrea Throop, Lyndon Tilson, Thomas Urban, Joe Voshell, Kimberly Wagner, Jonathan Williams, Mary Williamson, Qian Zeng, Tricia Zwiefelhofer, Clinton R. Paden, Suxiang Tong, Duncan MacCannell |
| EPI_ISL_1163665, EPI_ISL_1163666 | Tempus | Grubaugh Lab - Yale School of Public Health | Joseph Fauver, Tara Alpert, Anderson Brito, Mallery Breban, Anne Wyllie, Chantal Vogels, Mary Petrone, Annie Watkins, Chaney Kalinich, Isabel Ott, Nathan Grubaugh |
| EPI_ISL_1168728, EPI_ISL_1168729, EPI_ISL_1168730, EPI_ISL_1168734, EPI_ISL_1168735 | Biotia | Biotia | Dorottya Nagy-Szakal, Mara Couto-Rodriguez, Heather Wells, Joseph Barrows, Marilyne Debieu, Kristin Butcher, Siyuan Chen, Agnes Berki, Courteny Hager, Robert Boorstein, Mariah Taylor, Colleen Jonsson, Christopher Mason, Niamh O'Hara |
| EPI_ISL_1171856 | MSHS Clinical Microbiology Laboratories | MSHS Pathogen Surveillance Program | Fatima Amanat, Mahima Thapa, Tinting Lei, Shaza M. Sayed Ahmed, Daniel C. Adelsberg, Juan Manuel Carreno, Shirin Strohmeier, Aaron J. Schmitz, Sarah Zafar, Julian Q Zhou, Willemijn Rijnink, Hala Alshammary, Nicholas Borcherding, Ana Gonzalez Reiche, Komal Srivastava, Emilia Mia Sordillo, Harm van Bakel, The Personalized Virology Initiative, Jackson S. Turner, Goran Bajic, Viviana Simon, Ali H. Ellebedy, Florian Krammer |
| EPI_ISL_1171857 | Department of Microbiology, Icahn School of Medicine at Mount Sinai | MSHS Pathogen Surveillance Program | Fatima Amanat, Mahima Thapa, Tinting Lei, Shaza M. Sayed Ahmed, Daniel C. Adelsberg, Juan Manuel Carreno, Shirin Strohmeier, Aaron J. Schmitz, Sarah Zafar, Julian Q Zhou, Willemijn Rijnink, Hala Alshammary, Nicholas Borcherding, Ana Gonzalez Reiche, Komal Srivastava, Emilia Mia Sordillo, Harm van Bakel, The Personalized Virology Initiative, Jackson S. Turner, Goran Bajic, Viviana Simon, Ali H. Ellebedy, Florian Krammer |
| EPI_ISL_1172110, EPI_ISL_1172111, EPI_ISL_1172112, EPI_ISL_1172113, EPI_ISL_1172114, EPI_ISL_1172115, EPI_ISL_1172116, EPI_ISL_1172117, EPI_ISL_1172118, EPI_ISL_1172119, EPI_ISL_1172120, EPI_ISL_1172121, EPI_ISL_1172122, EPI_ISL_1172123, EPI_ISL_1172124, EPI_ISL_1172125, EPI_ISL_1172126, EPI_ISL_1172127, EPI_ISL_1172128, EPI_ISL_1172129, EPI_ISL_1172130, EPI_ISL_1172131, EPI_ISL_1172132, EPI_ISL_1172133, EPI_ISL_1172134, EPI_ISL_1172135, EPI_ISL_1172136, EPI_ISL_1172137, EPI_ISL_1172138, EPI_ISL_1172139, EPI_ISL_1172140, EPI_ISL_1172141, EPI_ISL_1172142, EPI_ISL_1172143, EPI_ISL_1172144, EPI_ISL_1172145, EPI_ISL_1172146, EPI_ISL_1172147, EPI_ISL_1172148, EPI_ISL_1172149, EPI_ISL_1172150, EPI_ISL_1172151, EPI_ISL_1172152, EPI_ISL_1172153, EPI_ISL_1172154, EPI_ISL_1172155, EPI_ISL_1172156, EPI_ISL_1172157, EPI_ISL_1172158, EPI_ISL_1172159, EPI_ISL_1172160, EPI_ISL_1172161, EPI_ISL_1172162, EPI_ISL_1172163, EPI_ISL_1172164, EPI_ISL_1172165, EPI_ISL_1172166, EPI_ISL_1172167, EPI_ISL_1172168, EPI_ISL_1172169, EPI_ISL_1172170, EPI_ISL_1172171, EPI_ISL_1172172, EPI_ISL_1172173, EPI_ISL_1172174, EPI_ISL_1172175, EPI_ISL_1172176, EPI_ISL_1172177, EPI_ISL_1172178, EPI_ISL_1172179, EPI_ISL_1172180, EPI_ISL_1172181, EPI_ISL_1172182, EPI_ISL_1172183, EPI_ISL_1172184, EPI_ISL_1172185, EPI_ISL_1172186, EPI_ISL_1172187, EPI_ISL_1172188, EPI_ISL_1172189, EPI_ISL_1172190, EPI_ISL_1172191, EPI_ISL_1172192, EPI_ISL_1172193, EPI_ISL_1172194, EPI_ISL_1172195, EPI_ISL_1172196, EPI_ISL_1172197, EPI_ISL_1172198, EPI_ISL_1172199, EPI_ISL_1172200, EPI_ISL_1172201, EPI_ISL_1172202, EPI_ISL_1172203, EPI_ISL_1172204, EPI_ISL_1172205, EPI_ISL_1172206, EPI_ISL_1172207, EPI_ISL_1172208, EPI_ISL_1172209, EPI_ISL_1172210, EPI_ISL_1172211, EPI_ISL_1172212, EPI_ISL_1172213, EPI_ISL_1172214, EPI_ISL_1172215, EPI_ISL_1172216, EPI_ISL_1172217, EPI_ISL_1172218, EPI_ISL_1172219, EPI_ISL_1172220, EPI_ISL_1172221, EPI_ISL_1172222, EPI_ISL_1172223, EPI_ISL_1172224, EPI_ISL_1172225, EPI_ISL_1172226, EPI_ISL_1172227, EPI_ISL_1172228, EPI_ISL_1172229, EPI_ISL_1172230, EPI_ISL_1172231, EPI_ISL_1172232, EPI_ISL_1172233, EPI_ISL_1172234, EPI_ISL_1172235, EPI_ISL_1172236, EPI_ISL_1172237, EPI_ISL_1172238, EPI_ISL_1172239, EPI_ISL_1172240, EPI_ISL_1172241, EPI_ISL_1172242, EPI_ISL_1172243, EPI_ISL_1172244, EPI_ISL_1172245, EPI_ISL_1172246, EPI_ISL_1172247, EPI_ISL_1172248, EPI_ISL_1172249, EPI_ISL_1172250, EPI_ISL_1172251, EPI_ISL_1172252, EPI_ISL_1172253, EPI_ISL_1172254, EPI_ISL_1172255, EPI_ISL_1172256, EPI_ISL_1172257, EPI_ISL_1172258, EPI_ISL_1172259, EPI_ISL_1172260, EPI_ISL_1172261, EPI_ISL_1172262, EPI_ISL_1172263, EPI_ISL_1172264, EPI_ISL_1172265, EPI_ISL_1172266, EPI_ISL_1172267, EPI_ISL_1172268, EPI_ISL_1172269, EPI_ISL_1172270, EPI_ISL_1172271, EPI_ISL_1172272, EPI_ISL_1172273, EPI_ISL_1172274, EPI_ISL_1172275, EPI_ISL_1172276, EPI_ISL_1172277, EPI_ISL_1172278, EPI_ISL_1172279, EPI_ISL_1172280, EPI_ISL_1172281, EPI_ISL_1172282, EPI_ISL_1172283, EPI_ISL_1172284, EPI_ISL_1172285, EPI_ISL_1172286, EPI_ISL_1172287, EPI_ISL_1172288, EPI_ISL_1172289, EPI_ISL_1172290, EPI_ISL_1172291, EPI_ISL_1172292, EPI_ISL_1172293, EPI_ISL_1172294, EPI_ISL_1172295, EPI_ISL_1172296, EPI_ISL_1172297, EPI_ISL_1172298, EPI_ISL_1172299, EPI_ISL_1172300, EPI_ISL_1172301, EPI_ISL_1172302, EPI_ISL_1172303, EPI_ISL_1172304, EPI_ISL_1172305, EPI_ISL_1172306, EPI_ISL_1172307, EPI_ISL_1172308, EPI_ISL_1172309, EPI_ISL_1172310, EPI_ISL_1172311, EPI_ISL_1172312, EPI_ISL_1172313, EPI_ISL_1172314, EPI_ISL_1172315, EPI_ISL_1172316, EPI_ISL_1172317, EPI_ISL_1172318, EPI_ISL_1172319, EPI_ISL_1172320, EPI_ISL_1172321, EPI_ISL_1172322, EPI_ISL_1172323, EPI_ISL_1172324, EPI_ISL_1172325, EPI_ISL_1172326, EPI_ISL_1172327, EPI_ISL_1172328, EPI_ISL_1172329, EPI_ISL_1172330, EPI_ISL_1172331, EPI_ISL_1172332, EPI_ISL_1172333, EPI_ISL_1172334, EPI_ISL_1172335, EPI_ISL_1172336, EPI_ISL_1172337, EPI_ISL_1172338, EPI_ISL_1172339, EPI_ISL_1172340, EPI_ISL_1172341, EPI_ISL_1172342, EPI_ISL_1172343, EPI_ISL_1172344, EPI_ISL_1172345, EPI_ISL_1172346, EPI_ISL_1172347, EPI_ISL_1172348, EPI_ISL_1172349, EPI_ISL_1172350, EPI_ISL_1172351, EPI_ISL_1172352, EPI_ISL_1172353, EPI_ISL_1172354, EPI_ISL_1172355, EPI_ISL_1172356, EPI_ISL_1172357, EPI_ISL_1172358, EPI_ISL_1172359, EPI_ISL_1172360, EPI_ISL_1172361, EPI_ISL_1172362, EPI_ISL_1172363, EPI_ISL_1172364, EPI_ISL_1172365, EPI_ISL_1172366, EPI_ISL_1172367, EPI_ISL_1172368, EPI_ISL_1172369, EPI_ISL_1172370, EPI_ISL_1172371, EPI_ISL_1172372, EPI_ISL_1172373, EPI_ISL_1172374, EPI_ISL_1172375, EPI_ISL_1172376, EPI_ISL_1172377, EPI_ISL_1172378, EPI_ISL_1172379, EPI_ISL_1172380, EPI_ISL_1172381, EPI_ISL_1172382, EPI_ISL_1172383, EPI_ISL_1172384, EPI_ISL_1172385, EPI_ISL_1172386, EPI_ISL_1172387, EPI_ISL_1172388, EPI_ISL_1172389, EPI_ISL_1172390, EPI_ISL_1172391, EPI_ISL_1172392, EPI_ISL_1172393, EPI_ISL_1172394, EPI_ISL_1172395, EPI_ISL_1172396, EPI_ISL_1172397, EPI_ISL_1172398, EPI_ISL_1172399, EPI_ISL_1172400, EPI_ISL_1172401, EPI_ISL_1172402, EPI_ISL_1172403, EPI_ISL_1172404, EPI_ISL_1172405, EPI_ISL_1172406, EPI_ISL_1172407, EPI_ISL_1172408, EPI_ISL_1172409, EPI_ISL_1172410, EPI_ISL_1172411, EPI_ISL_1172412, EPI_ISL_1172413, EPI_ISL_1172414, EPI_ISL_1172415, EPI_ISL_1172416, EPI_ISL_1172417, EPI_ISL_1172418, EPI_ISL_1172419, EPI_ISL_1172420, EPI_ISL_1172421, EPI_ISL_1172422, EPI_ISL_1172423, EPI_ISL_1172424, EPI_ISL_1172425, EPI_ISL_1172426, EPI_ISL_1172427, EPI_ISL_1172428, EPI_ISL_1172429, EPI_ISL_1172430, EPI_ISL_1172431, EPI_ISL_1172432, EPI_ISL_1172433, EPI_ISL_1172434, EPI_ISL_1172435, EPI_ISL_1172436, EPI_ISL_1172437, EPI_ISL_1172438, EPI_ISL_1172439, EPI_ISL_1172440, EPI_ISL_1172441, EPI_ISL_1172442, EPI_ISL_1172443, EPI_ISL_1172444, EPI_ISL_1172445, EPI_ISL_1172446, EPI_ISL_1172447, EPI_ISL_1172448, EPI_ISL_1172449, EPI_ISL_1172450, EPI_ISL_1172451, EPI_ISL_1172452, EPI_ISL_1172453, EPI_ISL_1172454, EPI_ISL_1172455, EPI_ISL_1172456, EPI_ISL_1172457, EPI_ISL_1172458, EPI_ISL_1172459, EPI_ISL_1172460, EPI_ISL_1172461, EPI_ISL_1172462, EPI_ISL_1172463, EPI_ISL_1172464, EPI_ISL_1172465, EPI_ISL_1172466, EPI_ISL_1172467, EPI_ISL_1172468, EPI_ISL_1172469, EPI_ISL_1172470, EPI_ISL_1172471, EPI_ISL_1172472, EPI_ISL_1172473, EPI_ISL_1172474, EPI_ISL_1172475, EPI_ISL_1172476, EPI_ISL_1172477, EPI_ISL_1172478, EPI_ISL_1172479, EPI_ISL_1172480, EPI_ISL_1172481, EPI_ISL_1172482, EPI_ISL_1172483, EPI_ISL_1172484, EPI_ISL_1172485, EPI_ISL_1172486, EPI_ISL_1172487, EPI_ISL_1172488, EPI_ISL_1172489, EPI_ISL_1172490, EPI_ISL_1172491, EPI_ISL_1172492, EPI_ISL_1172493, EPI_ISL_1172494, EPI_ISL_1172495, EPI_ISL_1172496, EPI_ISL_1172497, EPI_ISL_1172498, EPI_ISL_1172499, EPI_ISL_1172500, EPI_ISL_1172501, EPI_ISL_1172502, EPI_ISL_1172503, EPI_ISL_1172504, EPI_ISL_1172505, EPI_ISL_1172506, EPI_ISL_1172507, EPI_ISL_1172508, EPI_ISL_1172509, EPI_ISL_1172510, EPI_ISL_1172511, EPI_ISL_1172512, EPI_ISL_1172513, EPI_ISL_1172514, EPI_ISL_1172515, EPI_ISL_1172516, EPI_ISL_1172517, EPI_ISL_1172518, EPI_ISL_1172519, EPI_ISL_1172520, EPI_ISL_1172521, EPI_ISL_1172522, EPI_ISL_1172523, EPI_ISL_1172524, EPI_ISL_1172525, EPI_ISL_1172526, EPI_ISL_1172527, EPI_ISL_1172528, EPI_ISL_1172529, EPI_ISL_1172530, EPI_ISL_1172531, EPI_ISL_1172532, EPI_ISL_1172533, EPI_ISL_1172534, EPI_ISL_1172535, EPI_ISL_1172536, EPI_ISL_1172537, EPI_ISL_1172538, EPI_ISL_1172539, EPI_ISL_1172540, EPI_ISL_1172541, EPI_ISL_1172542, EPI_ISL_1172543, EPI_ISL_1172544, EPI_ISL_1172545, EPI_ISL_1172546, EPI_ISL_1172547, EPI_ISL_1172548, EPI_ISL_1172549, EPI_ISL_1172550, EPI_ISL_1172551, EPI_ISL_1172552, EPI_ISL_1172553, EPI_ISL_1172554, EPI_ISL_1172555, EPI_ISL_1172556, EPI_ISL_1172557, EPI_ISL_1172558, EPI_ISL_1172559, EPI_ISL_1172560, EPI_ISL_1172561, EPI_ISL_1172562, EPI_ISL_1172563, EPI_ISL_1172564, EPI_ISL_1172565, EPI_ISL_1172566, EPI_ISL_1172567, EPI_ISL_1172568, EPI_ISL_1172569, EPI_ISL_1172570, EPI_ISL_1172571, EPI_ISL_1172572, EPI_ISL_1172573, EPI_ISL_1172574, EPI_ISL_1172575, EPI_ISL_1172576, EPI_ISL_1172577, EPI_ISL_1172578, EPI_ISL_1172579, EPI_ISL_1172580, EPI_ISL_1172581, EPI_ISL_1172582, EPI_ISL_1172583, EPI_ISL_1172584, EPI_ISL_1172585, EPI_ISL_1172586, EPI_ISL_1172587, EPI_ISL_1172588, EPI_ISL_1172589, EPI_ISL_1172590, EPI_ISL_1172591, EPI_ISL_1172592, EPI_ISL_1172593, EPI_ISL_1172594, EPI_ISL_1172595, EPI_ISL_1172596, EPI_ISL_1172597, EPI_ISL_1172598, EPI_ISL_1172599, EPI_ISL_1172600, EPI_ISL_1172601, EPI_ISL_1172602, EPI_ISL_1172603, EPI_ISL_1172604, EPI_ISL_1172605, EPI_ISL_1172606, EPI_ISL_1172607, EPI_ISL_1172608, EPI_ISL_1172609, EPI_ISL_1172610, EPI_ISL_1172611, EPI_ISL_1172612, EPI_ISL_1172613, EPI_ISL_1172614, EPI_ISL_1172615, EPI_ISL_1172616, EPI_ISL_1172617, EPI_ISL_1172618, EPI_ISL_1172619, EPI_ISL_1172620, EPI_ISL_1172621, EPI_ISL_1172622, EPI_ISL_1172623, EPI_ISL_1172624, EPI_ISL_1172625, EPI_ISL_1172626, EPI_ISL_1172627, EPI_ISL_1172628, EPI_ISL_1172629, EPI_ISL_1172630, EPI_ISL_1172631, EPI_ISL_1172632, EPI_ISL_1172633, EPI_ISL_1172634, EPI_ISL_1172635, EPI_ISL_1172636, EPI_ISL_1172637, EPI_ISL_1172638, EPI_ISL_1172639, EPI_ISL_1172640, EPI_ISL_1172641, EPI_ISL_1172642, EPI_ISL_1172643, EPI_ISL_1172644, EPI_ISL_1172645, EPI_ISL_1172646, EPI_ISL_1172647, EPI_ISL_1172648, EPI_ISL_1172649, EPI_ISL_1172650, EPI_ISL_1172651, EPI_ISL_1172652, EPI_ISL_1172653, EPI_ISL_1172654, EPI_ISL_1172655, EPI_ISL_1172656, EPI_ISL_1172657, EPI_ISL_1172658, EPI_ISL_1172659, EPI_ISL_1172660, EPI_ISL_1172661, EPI_ISL_1172662, EPI_ISL_1172663, EPI_ISL_1172664, EPI_ISL_1172665, EPI_ISL_1172666, EPI_ISL_1172667, EPI_ISL_1172668, EPI_ISL_1172669, EPI_ISL_1172670, EPI_ISL_1172671, EPI_ISL_1172672, EPI_ISL_1172673, EPI_ISL_1172674, EPI_ISL_1172675, EPI_ISL_1172676, EPI_ISL_1172677, EPI_ISL_1172678, EPI_ISL_1172679, EPI_ISL_1172680, EPI_ISL_1172681, EPI_ISL_1172682, EPI_ISL_1172683, EPI_ISL_1172684, EPI_ISL_1172685, EPI_ISL_1172686, EPI_ISL_1172687, EPI_ISL_1172688, EPI_ISL_1172689, EPI_ISL_1172690, EPI_ISL_1172691, EPI_ISL_1172692, EPI_ISL_1172693, EPI_ISL_1172694, EPI_ISL_1172695, EPI_ISL_1172696, EPI_ISL_1172697, EPI_ISL_1172698, EPI_ISL_1172699, EPI_ISL_1172700, EPI_ISL_1172701, EPI_ISL_1172702, EPI_ISL_1172703, EPI_ISL_1172704, EPI_ISL_1172705, EPI_ISL_1172706, EPI_ISL_1172707, EPI_ISL_1172708, EPI_ISL_1172709, EPI_ISL_1172710, EPI_ISL_1172711, EPI_ISL_1172712, EPI_ISL_1172713, EPI_ISL_1172714, EPI_ISL_1172715, EPI_ISL_1172716, EPI_ISL_1172717, EPI_ISL_1172718, EPI_ISL_1172719, EPI_ISL_1172720, EPI_ISL_1172721, EPI_ISL_1172722, EPI_ISL_1172723, EPI_ISL_1172724, EPI_ISL_1172725, EPI_ISL_1172726, EPI_ISL_1172727, EPI_ISL_1172728, EPI_ISL_1172729, EPI_ISL_1172730, EPI_ISL_1172731, EPI_ISL_1172732, EPI_ISL_1172733, EPI_ISL_1172734, EPI_ISL_1172735, EPI_ISL_1172736, EPI_ISL_1172737, EPI_ISL_1172738, EPI_ISL_1172739, EPI_ISL_1172740, EPI_ISL_1172741, EPI_ISL_1172742, EPI_ISL_1172743, EPI_ISL_1172744, EPI_ISL_1172745, EPI_ISL_1172746, EPI_ISL_1172747, EPI_ISL_1172748, EPI_ISL_1172749, EPI_ISL_1172750, EPI_ISL_1172751, EPI_ISL_1172752, EPI_ISL_1172753, EPI_ISL_1172754, EPI_ISL_1172755, EPI_ISL_1172756, EPI_ISL_1172757, EPI_ISL_1172758, EPI_ISL_1172759, EPI_ISL_1172760, EPI_ISL_1172761, EPI_ISL_1172762, EPI_ISL_1172763, EPI_ISL_1172764, EPI_ISL_1172765, EPI_ISL_1172766, EPI_ISL_1172767, EPI_ISL_1172768, EPI_ISL_1172769, EPI_ISL_1172770, EPI_ISL_1172771, EPI_ISL_1172772, | | | |

| | | | |
|---|---|---|---|
| EPI_ISL_1172773, EPI_ISL_1172774, EPI_ISL_1172775, EPI_ISL_1172776, EPI_ISL_1172777, EPI_ISL_1172778, EPI_ISL_1172779, EPI_ISL_1172780, EPI_ISL_1172781, EPI_ISL_1172782, EPI_ISL_1172783, EPI_ISL_1172784, EPI_ISL_1172785, EPI_ISL_1172786, EPI_ISL_1172787, EPI_ISL_1172788, EPI_ISL_1172789, EPI_ISL_1172790, EPI_ISL_1172791, EPI_ISL_1172792, EPI_ISL_1172793, EPI_ISL_1172794, EPI_ISL_1172795, EPI_ISL_1172796, EPI_ISL_1172797, EPI_ISL_1172798, EPI_ISL_1172799, EPI_ISL_1172800, EPI_ISL_1172801, EPI_ISL_1172802, EPI_ISL_1172803, EPI_ISL_1172804, EPI_ISL_1172805, EPI_ISL_1172806, EPI_ISL_1172807, EPI_ISL_1172808, EPI_ISL_1172809, EPI_ISL_1172810, EPI_ISL_1172811, EPI_ISL_1172812, EPI_ISL_1172813, EPI_ISL_1172814, EPI_ISL_1172815, EPI_ISL_1172816, EPI_ISL_1172817, EPI_ISL_1172818, EPI_ISL_1172819, EPI_ISL_1172820, EPI_ISL_1172821, EPI_ISL_1172822, EPI_ISL_1172823, EPI_ISL_1172824, EPI_ISL_1172825, EPI_ISL_1172826, EPI_ISL_1172827, EPI_ISL_1172828, EPI_ISL_1172829, EPI_ISL_1172830, EPI_ISL_1172831, EPI_ISL_1172832, EPI_ISL_1172833, EPI_ISL_1172834, EPI_ISL_1172835, EPI_ISL_1172836, EPI_ISL_1172837, EPI_ISL_1172838, EPI_ISL_1172839, EPI_ISL_1172840, EPI_ISL_1172841, EPI_ISL_1172842, EPI_ISL_1172843, EPI_ISL_1172844, EPI_ISL_1172845, EPI_ISL_1172846, EPI_ISL_1172847, EPI_ISL_1172848, EPI_ISL_1172849, EPI_ISL_1172850, EPI_ISL_1172851, EPI_ISL_1172852, EPI_ISL_1172853, EPI_ISL_1172854, EPI_ISL_1172855, EPI_ISL_1172856, EPI_ISL_1172857, EPI_ISL_1172858, EPI_ISL_1172859, EPI_ISL_1172860, EPI_ISL_1172861, EPI_ISL_1172862, EPI_ISL_1172863, EPI_ISL_1172864, EPI_ISL_1172865, EPI_ISL_1172866, EPI_ISL_1172867, EPI_ISL_1172868, EPI_ISL_1172869, EPI_ISL_1172870, EPI_ISL_1172871, EPI_ISL_1172872, EPI_ISL_1172873, EPI_ISL_1172874, EPI_ISL_1172875, EPI_ISL_1172876, EPI_ISL_1172877, EPI_ISL_1172878, EPI_ISL_1172879, EPI_ISL_1172880, EPI_ISL_1172881, EPI_ISL_1172882, EPI_ISL_1172883, EPI_ISL_1172884, EPI_ISL_1172885, EPI_ISL_1172886, EPI_ISL_1172887, EPI_ISL_1172888, EPI_ISL_1172889, EPI_ISL_1172890, EPI_ISL_1172891, EPI_ISL_1172892, EPI_ISL_1172893, EPI_ISL_1172894, EPI_ISL_1172895, EPI_ISL_1172896, EPI_ISL_1172897, EPI_ISL_1172898, EPI_ISL_1172899, EPI_ISL_1172900, EPI_ISL_1172901, EPI_ISL_1172902, EPI_ISL_1172903, EPI_ISL_1172904, EPI_ISL_1172905, EPI_ISL_1172906, EPI_ISL_1172907, EPI_ISL_1172908, EPI_ISL_1172909, EPI_ISL_1172910, EPI_ISL_1172911, EPI_ISL_1172912, EPI_ISL_1172913, EPI_ISL_1172914, EPI_ISL_1172915, EPI_ISL_1172916, EPI_ISL_1172917, EPI_ISL_1172918, EPI_ISL_1172919, EPI_ISL_1172920, EPI_ISL_1172921, EPI_ISL_1172922, EPI_ISL_1172923, EPI_ISL_1172924, EPI_ISL_1172925, EPI_ISL_1172926, EPI_ISL_1172927, EPI_ISL_1172928, EPI_ISL_1172929, EPI_ISL_1172930, EPI_ISL_1172931, EPI_ISL_1172932, EPI_ISL_1172933, EPI_ISL_1172934, EPI_ISL_1172935, EPI_ISL_1172936, EPI_ISL_1172937, EPI_ISL_1172938, EPI_ISL_1172939, EPI_ISL_1172940, EPI_ISL_1172941, EPI_ISL_1172942, EPI_ISL_1172943, EPI_ISL_1172944, EPI_ISL_1172945, EPI_ISL_1172946, EPI_ISL_1172947, EPI_ISL_1172948, EPI_ISL_1172949, EPI_ISL_1172950, EPI_ISL_1172951, EPI_ISL_1172952, EPI_ISL_1172953, EPI_ISL_1172954, EPI_ISL_1172955, EPI_ISL_1172956, EPI_ISL_1172957, EPI_ISL_1172958, EPI_ISL_1172959, EPI_ISL_1172960, EPI_ISL_1172961, EPI_ISL_1172962, EPI_ISL_1172963, EPI_ISL_1172964, EPI_ISL_1172965, EPI_ISL_1172966, EPI_ISL_1172967, EPI_ISL_1172968, EPI_ISL_1172969, EPI_ISL_1172970, EPI_ISL_1172971, EPI_ISL_1172972, EPI_ISL_1172973, EPI_ISL_1172974, EPI_ISL_1172975, EPI_ISL_1172976, EPI_ISL_1172977, EPI_ISL_1172978, EPI_ISL_1172979, EPI_ISL_1172980, EPI_ISL_1172981, EPI_ISL_1172982, EPI_ISL_1172983, EPI_ISL_1172984, EPI_ISL_1172985, EPI_ISL_1172986, EPI_ISL_1172987, EPI_ISL_1172988, EPI_ISL_1172989, EPI_ISL_1172990, EPI_ISL_1172991, EPI_ISL_1172992, EPI_ISL_1172993, EPI_ISL_1172994, EPI_ISL_1172995, EPI_ISL_1172996, EPI_ISL_1172997, EPI_ISL_1172998, EPI_ISL_1172999, EPI_ISL_1173000, EPI_ISL_1173001, EPI_ISL_1173002, EPI_ISL_1173003, EPI_ISL_1173004, EPI_ISL_1173005, EPI_ISL_1173006, EPI_ISL_1173007, EPI_ISL_1173008, EPI_ISL_1173009, EPI_ISL_1173010, EPI_ISL_1173011, EPI_ISL_1173012, EPI_ISL_1173013, EPI_ISL_1173014, EPI_ISL_1173015, EPI_ISL_1173016, EPI_ISL_1173017, EPI_ISL_1173018, EPI_ISL_1173019, EPI_ISL_1173020, EPI_ISL_1173021, EPI_ISL_1173022, EPI_ISL_1173023, EPI_ISL_1173024, EPI_ISL_1173025, EPI_ISL_1173026, EPI_ISL_1173027, EPI_ISL_1173028, EPI_ISL_1173029, EPI_ISL_1173030, EPI_ISL_1173031, EPI_ISL_1173032, EPI_ISL_1173033, EPI_ISL_1173034, EPI_ISL_1173035, EPI_ISL_1173036, EPI_ISL_1173037, EPI_ISL_1173038, EPI_ISL_1173039, EPI_ISL_1173040, EPI_ISL_1173041, EPI_ISL_1173042, EPI_ISL_1173043, EPI_ISL_1173044, EPI_ISL_1173045, EPI_ISL_1173046, EPI_ISL_1173047, EPI_ISL_1173048, EPI_ISL_1173049, EPI_ISL_1173050, EPI_ISL_1173051, EPI_ISL_1173052, EPI_ISL_1173053, EPI_ISL_1173054, EPI_ISL_1173055, EPI_ISL_1173056, EPI_ISL_1173057, EPI_ISL_1173058, EPI_ISL_1173059, EPI_ISL_1173060, EPI_ISL_1173061, EPI_ISL_1173062, EPI_ISL_1173063, EPI_ISL_1173064, EPI_ISL_1173065, EPI_ISL_1173066, EPI_ISL_1173067, EPI_ISL_1173068, EPI_ISL_1173069, EPI_ISL_1173070, EPI_ISL_1173071, EPI_ISL_1173072, EPI_ISL_1173073, EPI_ISL_1173074, EPI_ISL_1173075, EPI_ISL_1173076, EPI_ISL_1173077, EPI_ISL_1173078, EPI_ISL_1173079, EPI_ISL_1173080, EPI_ISL_1173081, EPI_ISL_1173082, EPI_ISL_1173083, EPI_ISL_1173084, EPI_ISL_1173085, EPI_ISL_1173086, EPI_ISL_1173087, EPI_ISL_1173088, EPI_ISL_1173089, EPI_ISL_1173090, EPI_ISL_1173091, EPI_ISL_1173092, EPI_ISL_1173093, EPI_ISL_1173094, EPI_ISL_1173095, EPI_ISL_1173096, EPI_ISL_1173097, EPI_ISL_1173098, EPI_ISL_1173099, EPI_ISL_1173100, EPI_ISL_1173101, EPI_ISL_1173102, EPI_ISL_1173103, EPI_ISL_1173104, EPI_ISL_1173105, EPI_ISL_1173106, EPI_ISL_1173107, EPI_ISL_1173108, EPI_ISL_1173109, EPI_ISL_1173110, EPI_ISL_1173111, EPI_ISL_1173112, EPI_ISL_1173113, EPI_ISL_1173114, EPI_ISL_1173115, EPI_ISL_1173116, EPI_ISL_1173117, EPI_ISL_1173118, EPI_ISL_1173119, EPI_ISL_1173120, EPI_ISL_1173121, EPI_ISL_1173122, EPI_ISL_1173123, EPI_ISL_1173124, EPI_ISL_1173125, EPI_ISL_1173126, EPI_ISL_1173127, EPI_ISL_1173128, EPI_ISL_1173129, EPI_ISL_1173130, EPI_ISL_1173131, EPI_ISL_1173132, EPI_ISL_1173133, EPI_ISL_1173134, EPI_ISL_1173135, EPI_ISL_1173136, EPI_ISL_1173137, EPI_ISL_1173138, EPI_ISL_1173139, EPI_ISL_1173140, EPI_ISL_1173141, EPI_ISL_1173142, EPI_ISL_1173143, EPI_ISL_1173144, EPI_ISL_1173145, EPI_ISL_1173146, EPI_ISL_1173147, EPI_ISL_1173148, EPI_ISL_1173149, EPI_ISL_1173150, EPI_ISL_1173151, EPI_ISL_1173152, EPI_ISL_1173153, EPI_ISL_1173154, EPI_ISL_1173155, EPI_ISL_1173156, EPI_ISL_1173157, EPI_ISL_1173158 | | | |
| see above | Pandemic Response Lab - NYC | Pandemic Response Lab, R&D | Henry Lee, Michael Hammerling, Melissa Hopkins, Cybill del Castillo, Shinyoung Clair Kang, William Ward, Pradeep Bugga, Haiping Hao, Jon Laurent |
| EPI_ISL_1182091, EPI_ISL_1182092 | Murphy Medical Association | Grubaugh Lab - Yale School of Public Health | Joseph Fauver, Mallery Breban, Isabell Ott, Tara Alpert, Mary Petrone, Anderson Brito, Chantal Vogels, Annie Watkins, Chaney Kalinich, Caleb Neal, Eva Laszlo, Steven Murphy, Nathan Grubaugh |
| EPI_ISL_1184113, EPI_ISL_1184114, EPI_ISL_1184135, EPI_ISL_1184136, EPI_ISL_1184137 | Rochester Regional Hospital | Multidrug-Resistant Organism Repository and Surveillance Network (MRSN) | Emil Lesho, Brendan Corey, Francois Lebreton, Ana C. Ong, Brett E. Swierczewski, Jason W. Bennett, Edward E. Walsh, and Patrick Mc Gann |
| EPI_ISL_1193261, EPI_ISL_1193327, EPI_ISL_1193344, EPI_ISL_1193368 | Laboratory Corporation of America | Centers for Disease Control and Prevention Division of Viral Diseases, Pathogen Discovery | Peter W. Cook, Dakota Howard, Dhwani Batra, Ben L. Rambo-Martin, Minoo Agarwal, Eyad Almasri Debbie Boles, Ayla Burns, Nuthawin Charoensri, Oren Cohen, Susan Countryman, Mary Ann Cristobal, Bobbi Croy, Suzanne Dale, Hrushikesh Deshmukh, Amanda Douglas, Vincent Drouillon, Marcia Eisenberg, Howard Engler, Rama Ghatti, Prashant Gupta, Susan Hicks, Jake Humphrey, Lax Iyer, Manoj Jain, Mohan Kolli, Brian Krueger, Tim Kuphal, Stanley Letovsky, Michael Levandoski, Craig Lukasik, Jonathan Meltzer, Brian Norvell, Mindy Nye, Scott Parker, Christos Petropoulos, John Pruitt, Steven Ragan, Scott Ryan, Mike Sapeta, Jana Schroth, Suresh Babu Selvaraju, Goran Stevovic, Amanda Suchanek, Andrea Throop, Lyndon Tilson, Thomas Urban, Joe Voshell, Kimberly Wagner, Jonathan Williams, Mary Williamson, Qian Zeng, Tricia Zwiefelhofer, Clinton R. Paden, Suxiang Tong, Duncan MacCannell |
| EPI_ISL_1193643, EPI_ISL_1193692, EPI_ISL_1193695, EPI_ISL_1193713, EPI_ISL_1193729, EPI_ISL_1193749, EPI_ISL_1193765, EPI_ISL_1193812, EPI_ISL_1193827, EPI_ISL_1193829, EPI_ISL_1193830, EPI_ISL_1193836, EPI_ISL_1193880, EPI_ISL_1193889, EPI_ISL_1193891, EPI_ISL_1193896, EPI_ISL_1193909, EPI_ISL_1194044, EPI_ISL_1194045, EPI_ISL_1194128, EPI_ISL_1194140, EPI_ISL_1194157, EPI_ISL_1194178, EPI_ISL_1194179, EPI_ISL_1194185, EPI_ISL_1194191, EPI_ISL_1194194, EPI_ISL_1194211, EPI_ISL_1194212, EPI_ISL_1194213, EPI_ISL_1194218, EPI_ISL_1194222, EPI_ISL_1194225, EPI_ISL_1194342, EPI_ISL_1194374, EPI_ISL_1194390, EPI_ISL_1194405, EPI_ISL_1194417, EPI_ISL_1194421, EPI_ISL_1194435, EPI_ISL_1194461 | | | |
| see above | Quest Diagnostics Incorporated | Centers for Disease Control and Prevention Division of Viral Diseases, Pathogen Discovery | Peter W. Cook, Dakota Howard, Dhwani Batra, Ben L. Rambo-Martin, S. H. Rosenthal, A. Gerasimova, R. M. Kagan, B. Anderson, M. Hua, Y. Liu, L.E. Bernstein, K.E. Livingston, A. Perez, I. A. Shlyakhter, R. V. Rolando, R. Owen, P. Tanpaiboon, F. Lacbawan, Clinton R. Paden, Suxiang Tong, Duncan MacCannell |
| EPI_ISL_1194777 | Helix/Illumina | Centers for Disease Control and Prevention Division of Viral Diseases, Pathogen Discovery | Peter W. Cook, Dakota Howard, Dhwani Batra, Ben L. Rambo-Martin, Eileen de Feo, Jan Antico, Christine Tran, Matthew Tolentino, Shannon Wickline, Kim Gietzen, Brad Sickler, Jingtao Liu, Eric Allen, Phil Febbo, Summer Galloway, Nicole L. Washington, Simon White, Geraint Levan, Kelly Schiabor Barrett, Elizabeth Cirulli, Alexandre Bolze, Ary Ascencio, Charlotte Rivera-Garcia, Ryan Cho, Jason Nguyen, Sherry Wang, Jimmy Ramirez, Tyler Cassens, Efren Sandoval, Magnus Isaksson, William Lee, David Becker, Marc Laurent, James Lu, Clinton R. Paden, Suxiang Tong, Duncan MacCannell |
| EPI_ISL_1195744, EPI_ISL_1195745, EPI_ISL_1195746, EPI_ISL_1195747, EPI_ISL_1195748, EPI_ISL_1195749, EPI_ISL_1195750, EPI_ISL_1195751, EPI_ISL_1195752, EPI_ISL_1195753, EPI_ISL_1195754, EPI_ISL_1195755, EPI_ISL_1195756, EPI_ISL_1195757, EPI_ISL_1195758, EPI_ISL_1195759, EPI_ISL_1195760, EPI_ISL_1195761, EPI_ISL_1195762, EPI_ISL_1195763, EPI_ISL_1195764, EPI_ISL_1195765, EPI_ISL_1195766, EPI_ISL_1195767, EPI_ISL_1195768, EPI_ISL_1195770, EPI_ISL_1195771, EPI_ISL_1195772, EPI_ISL_1195773, EPI_ISL_1195774, EPI_ISL_1195775, EPI_ISL_1195776, EPI_ISL_1195777, EPI_ISL_1195778, EPI_ISL_1195779, EPI_ISL_1195780, EPI_ISL_1195781, EPI_ISL_1195782, EPI_ISL_1195783, EPI_ISL_1195784, EPI_ISL_1195785, EPI_ISL_1195786, EPI_ISL_1195787, EPI_ISL_1195788, EPI_ISL_1195789, EPI_ISL_1195790, EPI_ISL_1195791, EPI_ISL_1195792, EPI_ISL_1195793, EPI_ISL_1195794, EPI_ISL_1195795, EPI_ISL_1195796, EPI_ISL_1195798, EPI_ISL_1195799, EPI_ISL_1195801, EPI_ISL_1195802, EPI_ISL_1195803, EPI_ISL_1195804, EPI_ISL_1195805, EPI_ISL_1195806, EPI_ISL_1195807, EPI_ISL_1195808, EPI_ISL_1195809, EPI_ISL_1195810, EPI_ISL_1195811, EPI_ISL_1195812, EPI_ISL_1195813, EPI_ISL_1195814, EPI_ISL_1195815, EPI_ISL_1195816, EPI_ISL_1195817, EPI_ISL_1195818, EPI_ISL_1195819, EPI_ISL_1195820, EPI_ISL_1195821, EPI_ISL_1195822, EPI_ISL_1195823, EPI_ISL_1195824, EPI_ISL_1195825, EPI_ISL_1195826, EPI_ISL_1195827, EPI_ISL_1195828, EPI_ISL_1195829, EPI_ISL_1195830, EPI_ISL_1195831, EPI_ISL_1195832, EPI_ISL_1195833, EPI_ISL_1195834, EPI_ISL_1195835, EPI_ISL_1195836, EPI_ISL_1195837, EPI_ISL_1195838, EPI_ISL_1195839, EPI_ISL_1195840, EPI_ISL_1195841, EPI_ISL_1195842, EPI_ISL_1195843, EPI_ISL_1195845, EPI_ISL_1195846, EPI_ISL_1195848, EPI_ISL_1195849, EPI_ISL_1195850, EPI_ISL_1195851, EPI_ISL_1195852, EPI_ISL_1195853, EPI_ISL_1195854, EPI_ISL_1195855, EPI_ISL_1195856, EPI_ISL_1195857, EPI_ISL_1195858, EPI_ISL_1195859, EPI_ISL_1195860, EPI_ISL_1195861, EPI_ISL_1195862, EPI_ISL_1195864, EPI_ISL_1195865, EPI_ISL_1195866, EPI_ISL_1195867, EPI_ISL_1195868, EPI_ISL_1195869, EPI_ISL_1195870, EPI_ISL_1195871, EPI_ISL_1195872, EPI_ISL_1195873, EPI_ISL_1195874, EPI_ISL_1195875, EPI_ISL_1195876, EPI_ISL_1195877, EPI_ISL_1195878, EPI_ISL_1195879, EPI_ISL_1195880, EPI_ISL_1195881, EPI_ISL_1195882, EPI_ISL_1195883, EPI_ISL_1195884, EPI_ISL_1195885, EPI_ISL_1195886, EPI_ISL_1195887, EPI_ISL_1195888, EPI_ISL_1195889, EPI_ISL_1195890, EPI_ISL_1195892, EPI_ISL_1195893, EPI_ISL_1195894, EPI_ISL_1195895, EPI_ISL_1195896, EPI_ISL_1195897, EPI_ISL_1195898, EPI_ISL_1195899, EPI_ISL_1195900, EPI_ISL_1195901, EPI_ISL_1195902 | | | |
| see above | NYU Langone Health | Departments of Pathology and Medicine, New York University School of Medicine | Adriana Heguy, Dacia Dimartino, Emily Guzman, Christian Marier, Peter Meyn, Sitharam Ramaswami, Gael Westby, Paul Zappile, Yutong Zhang, Paolo Cotzia, Guiqing Wang |
| EPI_ISL_1200518 | DOHMH Chelsea | New York City Public Health Laboratory | Jade Wang, et al. |
| EPI_ISL_1200519 | HealthQuest Esoterics | New York City Public Health Laboratory | Jade Wang, et al. |
| EPI_ISL_1200520, EPI_ISL_1200521 | DOHMH Jamaica | New York City Public Health Laboratory | Jade Wang, et al. |
| EPI_ISL_1200522 | DOHMH Central Harlem | New York City Public Health Laboratory | Jade Wang, et al. |
| EPI_ISL_1200523 | DOHMH Fort Greene | New York City Public Health Laboratory | Jade Wang, et al. |
| EPI_ISL_1200524 | DOHMH Crown Heights | New York City Public Health Laboratory | Jade Wang, et al. |
| EPI_ISL_1200525 | DOHMH Riverside | New York City Public Health Laboratory | Jade Wang, et al. |
| EPI_ISL_1200526, EPI_ISL_1200527 | OCME Office Of Chief Medical Examiner | New York City Public Health Laboratory | Jade Wang, et al. |
| EPI_ISL_1200528, EPI_ISL_1200529, EPI_ISL_1200530 | DOHMH Chelsea | New York City Public Health Laboratory | Jade Wang, et al. |
| EPI_ISL_1200531, EPI_ISL_1200532 | OCME Office Of Chief Medical Examiner | New York City Public Health Laboratory | Jade Wang, et al. |
| EPI_ISL_1200533 | DOHMH Corona | New York City Public Health Laboratory | Jade Wang, et al. |
| EPI_ISL_1200534 | DOHMH Chelsea | New York City Public Health Laboratory | Jade Wang, et al. |
| EPI_ISL_1200535, EPI_ISL_1200536 | DOHMH Jamaica | New York City Public Health Laboratory | Jade Wang, et al. |
| EPI_ISL_1200537 | DOHMH Riverside | New York City Public Health Laboratory | Jade Wang, et al. |
| EPI_ISL_1200538 | DOHMH PHL | New York City Public Health Laboratory | Jade Wang, et al. |
| EPI_ISL_1200539 | OCME Office Of Chief Medical Examiner | New York City Public Health Laboratory | Jade Wang, et al. |

| | | | |
|---|---|---|---|
| EPI_ISL_1200540 | DOHMH Corona | New York City Public Health Laboratory | Jade Wang, et al. |
| EPI_ISL_1200541 | DOHMH Central Harlem | New York City Public Health Laboratory | Jade Wang, et al. |
| EPI_ISL_1200542 | DOHMH Jamaica | New York City Public Health Laboratory | Jade Wang, et al. |
| EPI_ISL_1200543 | DOHMH Crown Heights | New York City Public Health Laboratory | Jade Wang, et al. |
| EPI_ISL_1200544 | DOHMH Jamaica | New York City Public Health Laboratory | Jade Wang, et al. |
| EPI_ISL_1200545, EPI_ISL_1200546 | DOHMH Corona | New York City Public Health Laboratory | Jade Wang, et al. |
| EPI_ISL_1200547 | DOHMH Jamaica | New York City Public Health Laboratory | Jade Wang, et al. |
| EPI_ISL_1200548 | DOHMH PHL | New York City Public Health Laboratory | Jade Wang, et al. |
| EPI_ISL_1200549 | OCME Office Of Chief Medical Examiner | New York City Public Health Laboratory | Jade Wang, et al. |
| EPI_ISL_1200550 | DOHMH Morrisania | New York City Public Health Laboratory | Jade Wang, et al. |
| EPI_ISL_1200551 | DOHMH Jamaica | New York City Public Health Laboratory | Jade Wang, et al. |
| EPI_ISL_1200552 | DOHMH Central Harlem | New York City Public Health Laboratory | Jade Wang, et al. |
| EPI_ISL_1200553 | DOHMH Jamaica | New York City Public Health Laboratory | Jade Wang, et al. |
| EPI_ISL_1200554 | DOHMH Crown Heights | New York City Public Health Laboratory | Jade Wang, et al. |
| EPI_ISL_1200555 | OCME Office Of Chief Medical Examiner | New York City Public Health Laboratory | Jade Wang, et al. |
| EPI_ISL_1200556 | DOHMH Central Harlem | New York City Public Health Laboratory | Jade Wang, et al. |
| EPI_ISL_1200557, EPI_ISL_1200558, EPI_ISL_1200559 | DOHMH Morrisania | New York City Public Health Laboratory | Jade Wang, et al. |
| EPI_ISL_1200560 | DOHMH Chelsea | New York City Public Health Laboratory | Jade Wang, et al. |
| EPI_ISL_1200561 | Rockefeller University Hospital | New York City Public Health Laboratory | Jade Wang, et al. |
| EPI_ISL_1200562 | Department of Homeless Services | New York City Public Health Laboratory | Jade Wang, et al. |
| EPI_ISL_1200563 | DOHMH Jamaica | New York City Public Health Laboratory | Jade Wang, et al. |
| EPI_ISL_1200564 | DOHMH Chelsea | New York City Public Health Laboratory | Jade Wang, et al. |
| EPI_ISL_1200565 | DOHMH Corona | New York City Public Health Laboratory | Jade Wang, et al. |
| EPI_ISL_1200566 | DOHMH PHL | New York City Public Health Laboratory | Jade Wang, et al. |
| EPI_ISL_1200567 | DOHMH Morrisania | New York City Public Health Laboratory | Jade Wang, et al. |
| EPI_ISL_1200568, EPI_ISL_1200569, EPI_ISL_1200570 | DOHMH Corona | New York City Public Health Laboratory | Jade Wang, et al. |
| EPI_ISL_1200571 | DOHMH Crown Heights | New York City Public Health Laboratory | Jade Wang, et al. |
| EPI_ISL_1200572, EPI_ISL_1200573 | DOHMH Jamaica | New York City Public Health Laboratory | Jade Wang, et al. |
| EPI_ISL_1200574 | DOHMH Chelsea | New York City Public Health Laboratory | Jade Wang, et al. |
| EPI_ISL_1200575 | DOHMH Morrisania | New York City Public Health Laboratory | Jade Wang, et al. |
| EPI_ISL_1200576 | OCME Office Of Chief Medical Examiner | New York City Public Health Laboratory | Jade Wang, et al. |
| EPI_ISL_1200577 | DOHMH Central Harlem | New York City Public Health Laboratory | Jade Wang, et al. |
| EPI_ISL_1200578, EPI_ISL_1200579, EPI_ISL_1200580 | DOHMH Morrisania | New York City Public Health Laboratory | Jade Wang, et al. |
| EPI_ISL_1200581 | DOHMH Jamaica | New York City Public Health Laboratory | Jade Wang, et al. |
| EPI_ISL_1200582 | DOHMH Morrisania | New York City Public Health Laboratory | Jade Wang, et al. |
| EPI_ISL_1200583 | DOHMH Jamaica | New York City Public Health Laboratory | Jade Wang, et al. |
| EPI_ISL_1200584 | DOHMH Chelsea | New York City Public Health Laboratory | Jade Wang, et al. |
| EPI_ISL_1200585 | DOHMH Jamaica | New York City Public Health Laboratory | Jade Wang, et al. |
| EPI_ISL_1200586, EPI_ISL_1200587 | DOHMH PHL | New York City Public Health Laboratory | Jade Wang, et al. |
| EPI_ISL_1200588, EPI_ISL_1200589 | DOHMH Morrisania | New York City Public Health Laboratory | Jade Wang, et al. |
| EPI_ISL_1200590 | DOHMH Jamaica | New York City Public Health Laboratory | Jade Wang, et al. |
| EPI_ISL_1200591, EPI_ISL_1200592, EPI_ISL_1200593 | DOHMH Corona | New York City Public Health Laboratory | Jade Wang, et al. |
| EPI_ISL_1200594 | DOHMH Central Harlem | New York City Public Health Laboratory | Jade Wang, et al. |
| EPI_ISL_1200595, EPI_ISL_1200596 | DOHMH Jamaica | New York City Public Health Laboratory | Jade Wang, et al. |
| EPI_ISL_1200597, EPI_ISL_1200598 | DOHMH Corona | New York City Public Health Laboratory | Jade Wang, et al. |
| EPI_ISL_1200599 | Department of Homeless Services | New York City Public Health Laboratory | Jade Wang, et al. |
| EPI_ISL_1200600 | DOHMH Morrisania | New York City Public Health Laboratory | Jade Wang, et al. |
| EPI_ISL_1200601, EPI_ISL_1200602 | DOHMH PHL | New York City Public Health Laboratory | Jade Wang, et al. |
| EPI_ISL_1200603 | DOHMH Corona | New York City Public Health Laboratory | Jade Wang, et al. |
| EPI_ISL_1200604 | DOHMH Morrisania | New York City Public Health Laboratory | Jade Wang, et al. |
| EPI_ISL_1200605 | DOHMH Crown Heights | New York City Public Health Laboratory | Jade Wang, et al. |
| EPI_ISL_1200606 | DOHMH Riverside | New York City Public Health Laboratory | Jade Wang, et al. |
| EPI_ISL_1200607, EPI_ISL_1200608, EPI_ISL_1200609, EPI_ISL_1200610 | OCME Office Of Chief Medical Examiner | New York City Public Health Laboratory | Jade Wang, et al. |
| EPI_ISL_1200611 | DOHMH Riverside | New York City Public Health Laboratory | Jade Wang, et al. |
| EPI_ISL_1200612 | DOHMH Morrisania | New York City Public Health Laboratory | Jade Wang, et al. |

| | | | |
|---|---|---|---|
| EPI_ISL_1200613, EPI_ISL_1200614 | DOHMH Corona | New York City Public Health Laboratory | Jade Wang, et al. |
| EPI_ISL_1200615, EPI_ISL_1200616, EPI_ISL_1200617, EPI_ISL_1200618 | DOHMH Jamaica | New York City Public Health Laboratory | Jade Wang, et al. |
| EPI_ISL_1200619 | DOHMH PHL | New York City Public Health Laboratory | Jade Wang, et al. |
| EPI_ISL_1200620 | DOHMH Fort Greene | New York City Public Health Laboratory | Jade Wang, et al. |
| EPI_ISL_1200621, EPI_ISL_1200622 | DOHMH Central Harlem | New York City Public Health Laboratory | Jade Wang, et al. |
| EPI_ISL_1200623 | DOHMH PHL | New York City Public Health Laboratory | Jade Wang, et al. |
| EPI_ISL_1200624, EPI_ISL_1200625 | DOHMH Central Harlem | New York City Public Health Laboratory | Jade Wang, et al. |
| EPI_ISL_1200626 | DOHMH Corona | New York City Public Health Laboratory | Jade Wang, et al. |
| EPI_ISL_1200627 | OCME Office Of Chief Medical Examiner | New York City Public Health Laboratory | Jade Wang, et al. |
| EPI_ISL_1220388, EPI_ISL_1220390, EPI_ISL_1220411, EPI_ISL_1220420, EPI_ISL_1220517, EPI_ISL_1220548, EPI_ISL_1220578, EPI_ISL_1220619, EPI_ISL_1220646, EPI_ISL_1220655, EPI_ISL_1220669, EPI_ISL_1220676, EPI_ISL_1220685, EPI_ISL_1220697, EPI_ISL_1220710, EPI_ISL_1220711 | | | |
| see above | Quest Diagnostics Incorporated | Centers for Disease Control and Prevention Division of Viral Diseases, Pathogen Discovery | Peter W. Cook, Dakota Howard, Dhwani Batra, Ben L. Rambo-Martin, S. H. Rosenthal, A. Gerasimova, R. M. Kagan, B. Anderson, M. Hua, Y. Liu, L.E. Bernstein, K.E. Livingston, A. Perez, I. A. Shlyakhter, R. V. Rolando, R. Owen, P. Tanpaiboon, F. Lacbawan, Clinton R. Paden, Suxiang Tong, Duncan MacCannell |
| EPI_ISL_1220913, EPI_ISL_1220914, EPI_ISL_1220915, EPI_ISL_1220916, EPI_ISL_1220917, EPI_ISL_1220951, EPI_ISL_1220953, EPI_ISL_1221009, EPI_ISL_1221010, EPI_ISL_1221011, EPI_ISL_1221014, EPI_ISL_1221015, EPI_ISL_1221138, EPI_ISL_1221161, EPI_ISL_1221270, EPI_ISL_1221271, EPI_ISL_1221549, EPI_ISL_1221551, EPI_ISL_1221754, EPI_ISL_1221755, EPI_ISL_1221756, EPI_ISL_1221757, EPI_ISL_1221758, EPI_ISL_1221759, EPI_ISL_1221760, EPI_ISL_1221767, EPI_ISL_1221768, EPI_ISL_1221769, EPI_ISL_1221770, EPI_ISL_1221782, EPI_ISL_1222050, EPI_ISL_1222051, EPI_ISL_1222052, EPI_ISL_1222053, EPI_ISL_1222054, EPI_ISL_1222055, EPI_ISL_1222056, EPI_ISL_1222057, EPI_ISL_1222058, EPI_ISL_1222059, EPI_ISL_1222060, EPI_ISL_1222064, EPI_ISL_1222065, EPI_ISL_1222067, EPI_ISL_1222068, EPI_ISL_1222069, EPI_ISL_1222139, EPI_ISL_1222140, EPI_ISL_1222141, EPI_ISL_1222142, EPI_ISL_1222143 | | | |
| see above | Laboratory Corporation of America | Centers for Disease Control and Prevention Division of Viral Diseases, Pathogen Discovery | Peter W. Cook, Dakota Howard, Dhwani Batra, Ben L. Rambo-Martin, Minoo Agarwal, Eyad Almasri, Debbie Boles, Ayla Burns, Nuthawin Charoensri, Oren Cohen, Susan Countryman, Mary Ann Cristobal, Bobbi Croy, Suzanne Dale, Hrushikesh Deshmukh, Amanda Douglas, Vincent Drouillon, Marcia Eisenberg, Howard Engler, Rama Ghatti, Prashant Gupta, Susan Hicks, Jake Humphrey, Lax Iyer, Manoj Jain, Mohan Kolli, Brian Krueger, Tim Kuphal, Stanley Letovsky, Michael Levandoski, Craig Lukasik, Jonathan Meltzer, Brian Norvell, Mindy Nye, Scott Parker, Christos Petropoulos, John Pruitt, Steven Ragan, Scott Ryan, Mike Sapeta, Jana Schroth, Suresh Babu Selvaraju, Goran Stevovic, Amanda Suchanek, Andrea Throop, Lyndon Tilson, Thomas Urban, Joe Voshell, Kimberly Wagner, Jonathan Williams, Mary Williamson, Qian Zeng, Tricia Zwiefelhofer, Clinton R. Paden, Suxiang Tong, Duncan MacCannell |
| EPI_ISL_1225664 | Murphy Medical Association | Grubaugh Lab - Yale School of Public Health | Joseph Fauver, Mallery Breban, Isabell Ott, Tara Alpert, Mary Petrone, Anderson Brito, Chantal Vogels, Annie Watkins, Chaney Kalinich, Caleb Neal, Eva Laszlo, Steven Murphy, Nathan Grubaugh |
| EPI_ISL_1225669 | Yale Clinical Virology Lab | Grubaugh Lab - Yale School of Public Health | Joseph Fauver, Mallery Breban, Isabell Ott, Tara Alpert, Mary Petrone, Anderson Brito, Chantal Vogels, Annie Watkins, Chaney Kalinich, Marie L. Landry, Nathan Grubaugh |
| EPI_ISL_1225678, EPI_ISL_1225699 | Murphy Medical Association | Grubaugh Lab - Yale School of Public Health | Joseph Fauver, Mallery Breban, Isabell Ott, Tara Alpert, Mary Petrone, Anderson Brito, Chantal Vogels, Annie Watkins, Chaney Kalinich, Caleb Neal, Eva Laszlo, Steven Murphy, Nathan Grubaugh |
| EPI_ISL_1225706 | Yale Clinical Virology Lab | Grubaugh Lab - Yale School of Public Health | Joseph Fauver, Mallery Breban, Isabell Ott, Tara Alpert, Mary Petrone, Anderson Brito, Chantal Vogels, Annie Watkins, Chaney Kalinich, Marie L. Landry, Nathan Grubaugh |
| EPI_ISL_1226003, EPI_ISL_1226004, EPI_ISL_1226005, EPI_ISL_1226006, EPI_ISL_1226048, EPI_ISL_1226049, EPI_ISL_1226050, EPI_ISL_1226051, EPI_ISL_1226052, EPI_ISL_1226053, EPI_ISL_1226054, EPI_ISL_1226055, EPI_ISL_1226056, EPI_ISL_1226057, EPI_ISL_1226058, EPI_ISL_1226059, EPI_ISL_1226060, EPI_ISL_1226061, EPI_ISL_1226062, EPI_ISL_1226063, EPI_ISL_1226064, EPI_ISL_1226065, EPI_ISL_1226066, EPI_ISL_1226067, EPI_ISL_1226068, EPI_ISL_1226069, EPI_ISL_1226070, EPI_ISL_1226071, EPI_ISL_1226072, EPI_ISL_1226073, EPI_ISL_1226074, EPI_ISL_1226075, EPI_ISL_1226076, EPI_ISL_1226077, EPI_ISL_1226078, EPI_ISL_1226079, EPI_ISL_1226080, EPI_ISL_1226081 | | | |
| see above | "NYSDOH Wadsworth Center, Virology Lab" | Genomics and Discovery, Respiratory Viruses Branch, Division of Viral Diseases, Centers for Disease Control and Prevention | Krista Queen, Yan Li, Ying Tao, Jing Zhang, Anna Uehara, Anna Montmayeur, Clinton R. Paden, Kristen Knipe, Matthew Schmerer, Shoshona Le, Katie Dillon, Peter W. Cook, Rachel Marine, Mili Sheth, Jasmine Padilla, Sarah Nobles, Mark Burroughs, Lori Rowe, Haibin Wang, Ben L. Rambo-Martin, Kristine Lacek, Sam Shepard, Dhwani Batra, Suxiang Tong, Justin Lee |
| EPI_ISL_1226417 | WESTCHESTER MEDICAL CENTER | Wadsworth Center, New York State Department of Health | Kirsten St. George, Daryl M. Lamson, Alexis Russel, Matthew Shudt, Melissa A Leisner, Jonathan Plitnick, Navjot Singh, John Kelly, Erasmus Schneider, Erica Lasek-Nesselquist |
| EPI_ISL_1226418 | GLENS FALLS HOSPITAL LABORATORY | Wadsworth Center, New York State Department of Health | Kirsten St. George, Daryl M. Lamson, Alexis Russel, Matthew Shudt, Melissa A Leisner, Jonathan Plitnick, Navjot Singh, John Kelly, Erasmus Schneider, Erica Lasek-Nesselquist |
| EPI_ISL_1226419 | NYC Pandemic Response Lab | Wadsworth Center, New York State Department of Health | Kirsten St. George, Daryl M. Lamson, Alexis Russel, Matthew Shudt, Melissa A Leisner, Jonathan Plitnick, Navjot Singh, John Kelly, Erasmus Schneider, Erica Lasek-Nesselquist |
| EPI_ISL_1226420 | NORTH SHORE UNIVERSITY HOSPITAL | Wadsworth Center, New York State Department of Health | Kirsten St. George, Daryl M. Lamson, Alexis Russel, Matthew Shudt, Melissa A Leisner, Jonathan Plitnick, Navjot Singh, John Kelly, Erasmus Schneider, Erica Lasek-Nesselquist |
| EPI_ISL_1226421 | URMC LABS | Wadsworth Center, New York State Department of Health | Kirsten St. George, Daryl M. Lamson, Alexis Russel, Matthew Shudt, Melissa A Leisner, Jonathan Plitnick, Navjot Singh, John Kelly, Erasmus Schneider, Erica Lasek-Nesselquist |
| EPI_ISL_1226422 | Masonic Care Community of NY | Wadsworth Center, New York State Department of Health | Kirsten St. George, Daryl M. Lamson, Alexis Russel, Matthew Shudt, Melissa A Leisner, Jonathan Plitnick, Navjot Singh, John Kelly, Erasmus Schneider, Erica Lasek-Nesselquist |
| EPI_ISL_1226423 | THE MARY IMOGENE BASSETT HOSPITAL | Wadsworth Center, New York State Department of Health | Kirsten St. George, Daryl M. Lamson, Alexis Russel, Matthew Shudt, Melissa A Leisner, Jonathan Plitnick, Navjot Singh, John Kelly, Erasmus Schneider, Erica Lasek-Nesselquist |
| EPI_ISL_1226424 | MONTEFIORE MEDICAL CENTER LABORATORIES | Wadsworth Center, New York State Department of Health | Kirsten St. George, Daryl M. Lamson, Alexis Russel, Matthew Shudt, Melissa A Leisner, Jonathan Plitnick, Navjot Singh, John Kelly, Erasmus Schneider, Erica Lasek-Nesselquist |
| EPI_ISL_1226425 | NORTHWELL HEALTH LABORATORIES | Wadsworth Center, New York State Department of Health | Kirsten St. George, Daryl M. Lamson, Alexis Russel, Matthew Shudt, Melissa A Leisner, Jonathan Plitnick, Navjot Singh, John Kelly, Erasmus Schneider, Erica Lasek-Nesselquist |
| EPI_ISL_1226426 | MONTEFIORE MEDICAL CENTER LABORATORIES | Wadsworth Center, New York State Department of Health | Kirsten St. George, Daryl M. Lamson, Alexis Russel, Matthew Shudt, Melissa A Leisner, Jonathan Plitnick, Navjot Singh, John Kelly, Erasmus Schneider, Erica Lasek-Nesselquist |
| EPI_ISL_1226427 | MEMORIAL SLOAN KETTERING CANCER CENTER | Wadsworth Center, New York State Department of Health | Kirsten St. George, Daryl M. Lamson, Alexis Russel, Matthew Shudt, Melissa A Leisner, Jonathan Plitnick, Navjot Singh, John Kelly, Erasmus Schneider, Erica Lasek-Nesselquist |
| EPI_ISL_1226428 | URMC LABS | Wadsworth Center, New York State Department of Health | Kirsten St. George, Daryl M. Lamson, Alexis Russel, Matthew Shudt, Melissa A Leisner, Jonathan Plitnick, Navjot Singh, John Kelly, Erasmus Schneider, Erica Lasek-Nesselquist |
| EPI_ISL_1226429 | Masonic Care Community of NY | Wadsworth Center, New York State Department of Health | Kirsten St. George, Daryl M. Lamson, Alexis Russel, Matthew Shudt, Melissa A Leisner, Jonathan Plitnick, Navjot Singh, John Kelly, Erasmus Schneider, Erica Lasek-Nesselquist |
| EPI_ISL_1226430 | NYC Pandemic Response Lab | Wadsworth Center, New York State Department of Health | Kirsten St. George, Daryl M. Lamson, Alexis Russel, Matthew Shudt, Melissa A Leisner, Jonathan Plitnick, Navjot Singh, John Kelly, Erasmus Schneider, Erica Lasek-Nesselquist |
| EPI_ISL_1226431 | MONTEFIORE MEDICAL CENTER LABORATORIES | Wadsworth Center, New York State Department of Health | Kirsten St. George, Daryl M. Lamson, Alexis Russel, Matthew Shudt, Melissa A Leisner, Jonathan Plitnick, Navjot Singh, John Kelly, Erasmus Schneider, Erica Lasek-Nesselquist |
| EPI_ISL_1226432 | URMC LABS | Wadsworth Center, New York State Department of Health | Kirsten St. George, Daryl M. Lamson, Alexis Russel, Matthew Shudt, Melissa A Leisner, Jonathan Plitnick, Navjot Singh, John Kelly, Erasmus Schneider, Erica Lasek-Nesselquist |

| | | | |
|---|---|---|---|
| EPI_ISL_1226433 | MONTEFIORE MEDICAL CENTER LABORATORIES | Wadsworth Center, New York State Department of Health | Kirsten St. George, Daryl M. Lamson, Alexis Russel, Matthew Shudt, Melissa A Leisner, Jonathan Plitnick, Navjot Singh, John Kelly, Erasmus Schneider, Erica Lasek-Nesselquist |
| EPI_ISL_1226434 | New York Presbyterian Hospital | Wadsworth Center, New York State Department of Health | Kirsten St. George, Daryl M. Lamson, Alexis Russel, Matthew Shudt, Melissa A Leisner, Jonathan Plitnick, Navjot Singh, John Kelly, Erasmus Schneider, Erica Lasek-Nesselquist |
| EPI_ISL_1226435 | URMC LABS | Wadsworth Center, New York State Department of Health | Kirsten St. George, Daryl M. Lamson, Alexis Russel, Matthew Shudt, Melissa A Leisner, Jonathan Plitnick, Navjot Singh, John Kelly, Erasmus Schneider, Erica Lasek-Nesselquist |
| EPI_ISL_1226436 | KALEIDA CENTER FOR LABORATORY MEDICINE | Wadsworth Center, New York State Department of Health | Kirsten St. George, Daryl M. Lamson, Alexis Russel, Matthew Shudt, Melissa A Leisner, Jonathan Plitnick, Navjot Singh, John Kelly, Erasmus Schneider, Erica Lasek-Nesselquist |
| EPI_ISL_1226437 | MEMORIAL SLOAN KETTERING CANCER CENTER | Wadsworth Center, New York State Department of Health | Kirsten St. George, Daryl M. Lamson, Alexis Russel, Matthew Shudt, Melissa A Leisner, Jonathan Plitnick, Navjot Singh, John Kelly, Erasmus Schneider, Erica Lasek-Nesselquist |
| EPI_ISL_1226438 | WESTCHESTER MEDICAL CENTER | Wadsworth Center, New York State Department of Health | Kirsten St. George, Daryl M. Lamson, Alexis Russel, Matthew Shudt, Melissa A Leisner, Jonathan Plitnick, Navjot Singh, John Kelly, Erasmus Schneider, Erica Lasek-Nesselquist |
| EPI_ISL_1226439, EPI_ISL_1226440, EPI_ISL_1226441 | NYC Pandemic Response Lab | Wadsworth Center, New York State Department of Health | Kirsten St. George, Daryl M. Lamson, Alexis Russel, Matthew Shudt, Melissa A Leisner, Jonathan Plitnick, Navjot Singh, John Kelly, Erasmus Schneider, Erica Lasek-Nesselquist |
| EPI_ISL_1226442 | MEMORIAL SLOAN KETTERING CANCER CENTER | Wadsworth Center, New York State Department of Health | Kirsten St. George, Daryl M. Lamson, Alexis Russel, Matthew Shudt, Melissa A Leisner, Jonathan Plitnick, Navjot Singh, John Kelly, Erasmus Schneider, Erica Lasek-Nesselquist |
| EPI_ISL_1226443 | NYC Pandemic Response Lab | Wadsworth Center, New York State Department of Health | Kirsten St. George, Daryl M. Lamson, Alexis Russel, Matthew Shudt, Melissa A Leisner, Jonathan Plitnick, Navjot Singh, John Kelly, Erasmus Schneider, Erica Lasek-Nesselquist |
| EPI_ISL_1226444 | NORTHWELL HEALTH LABORATORIES | Wadsworth Center, New York State Department of Health | Kirsten St. George, Daryl M. Lamson, Alexis Russel, Matthew Shudt, Melissa A Leisner, Jonathan Plitnick, Navjot Singh, John Kelly, Erasmus Schneider, Erica Lasek-Nesselquist |
| EPI_ISL_1226445 | URMC LABS | Wadsworth Center, New York State Department of Health | Kirsten St. George, Daryl M. Lamson, Alexis Russel, Matthew Shudt, Melissa A Leisner, Jonathan Plitnick, Navjot Singh, John Kelly, Erasmus Schneider, Erica Lasek-Nesselquist |
| EPI_ISL_1226446 | MONTEFIORE MEDICAL CENTER LABORATORIES | Wadsworth Center, New York State Department of Health | Kirsten St. George, Daryl M. Lamson, Alexis Russel, Matthew Shudt, Melissa A Leisner, Jonathan Plitnick, Navjot Singh, John Kelly, Erasmus Schneider, Erica Lasek-Nesselquist |
| EPI_ISL_1226447 | NORTH SHORE UNIVERSITY HOSPITAL | Wadsworth Center, New York State Department of Health | Kirsten St. George, Daryl M. Lamson, Alexis Russel, Matthew Shudt, Melissa A Leisner, Jonathan Plitnick, Navjot Singh, John Kelly, Erasmus Schneider, Erica Lasek-Nesselquist |
| EPI_ISL_1226448 | NYC Pandemic Response Lab | Wadsworth Center, New York State Department of Health | Kirsten St. George, Daryl M. Lamson, Alexis Russel, Matthew Shudt, Melissa A Leisner, Jonathan Plitnick, Navjot Singh, John Kelly, Erasmus Schneider, Erica Lasek-Nesselquist |
| EPI_ISL_1226449 | TEMPUS LABS INC | Wadsworth Center, New York State Department of Health | Kirsten St. George, Daryl M. Lamson, Alexis Russel, Matthew Shudt, Melissa A Leisner, Jonathan Plitnick, Navjot Singh, John Kelly, Erasmus Schneider, Erica Lasek-Nesselquist |
| EPI_ISL_1226450 | MONTEFIORE MEDICAL CENTER LABORATORIES | Wadsworth Center, New York State Department of Health | Kirsten St. George, Daryl M. Lamson, Alexis Russel, Matthew Shudt, Melissa A Leisner, Jonathan Plitnick, Navjot Singh, John Kelly, Erasmus Schneider, Erica Lasek-Nesselquist |
| EPI_ISL_1226451 | URMC LABS | Wadsworth Center, New York State Department of Health | Kirsten St. George, Daryl M. Lamson, Alexis Russel, Matthew Shudt, Melissa A Leisner, Jonathan Plitnick, Navjot Singh, John Kelly, Erasmus Schneider, Erica Lasek-Nesselquist |
| EPI_ISL_1226452 | GLENS FALLS HOSPITAL LABORATORY | Wadsworth Center, New York State Department of Health | Kirsten St. George, Daryl M. Lamson, Alexis Russel, Matthew Shudt, Melissa A Leisner, Jonathan Plitnick, Navjot Singh, John Kelly, Erasmus Schneider, Erica Lasek-Nesselquist |
| EPI_ISL_1226453 | WESTCHESTER MEDICAL CENTER | Wadsworth Center, New York State Department of Health | Kirsten St. George, Daryl M. Lamson, Alexis Russel, Matthew Shudt, Melissa A Leisner, Jonathan Plitnick, Navjot Singh, John Kelly, Erasmus Schneider, Erica Lasek-Nesselquist |
| EPI_ISL_1226454 | MONTEFIORE MEDICAL CENTER LABORATORIES | Wadsworth Center, New York State Department of Health | Kirsten St. George, Daryl M. Lamson, Alexis Russel, Matthew Shudt, Melissa A Leisner, Jonathan Plitnick, Navjot Singh, John Kelly, Erasmus Schneider, Erica Lasek-Nesselquist |
| EPI_ISL_1226455 | MEMORIAL SLOAN KETTERING CANCER CENTER | Wadsworth Center, New York State Department of Health | Kirsten St. George, Daryl M. Lamson, Alexis Russel, Matthew Shudt, Melissa A Leisner, Jonathan Plitnick, Navjot Singh, John Kelly, Erasmus Schneider, Erica Lasek-Nesselquist |
| EPI_ISL_1226456 | Columbia University Irving Medical Center | Wadsworth Center, New York State Department of Health | Kirsten St. George, Daryl M. Lamson, Alexis Russel, Matthew Shudt, Melissa A Leisner, Jonathan Plitnick, Navjot Singh, John Kelly, Erasmus Schneider, Erica Lasek-Nesselquist |
| EPI_ISL_1226457 | MONTEFIORE MEDICAL CENTER LABORATORIES | Wadsworth Center, New York State Department of Health | Kirsten St. George, Daryl M. Lamson, Alexis Russel, Matthew Shudt, Melissa A Leisner, Jonathan Plitnick, Navjot Singh, John Kelly, Erasmus Schneider, Erica Lasek-Nesselquist |
| EPI_ISL_1226458 | KALEIDA CENTER FOR LABORATORY MEDICINE | Wadsworth Center, New York State Department of Health | Kirsten St. George, Daryl M. Lamson, Alexis Russel, Matthew Shudt, Melissa A Leisner, Jonathan Plitnick, Navjot Singh, John Kelly, Erasmus Schneider, Erica Lasek-Nesselquist |
| EPI_ISL_1226459 | NYC Pandemic Response Lab | Wadsworth Center, New York State Department of Health | Kirsten St. George, Daryl M. Lamson, Alexis Russel, Matthew Shudt, Melissa A Leisner, Jonathan Plitnick, Navjot Singh, John Kelly, Erasmus Schneider, Erica Lasek-Nesselquist |
| EPI_ISL_1226460 | URMC LABS | Wadsworth Center, New York State Department of Health | Kirsten St. George, Daryl M. Lamson, Alexis Russel, Matthew Shudt, Melissa A Leisner, Jonathan Plitnick, Navjot Singh, John Kelly, Erasmus Schneider, Erica Lasek-Nesselquist |
| EPI_ISL_1226461 | Columbia University Irving Medical Center | Wadsworth Center, New York State Department of Health | Kirsten St. George, Daryl M. Lamson, Alexis Russel, Matthew Shudt, Melissa A Leisner, Jonathan Plitnick, Navjot Singh, John Kelly, Erasmus Schneider, Erica Lasek-Nesselquist |
| EPI_ISL_1226462 | WESTCHESTER MEDICAL CENTER | Wadsworth Center, New York State Department of Health | Kirsten St. George, Daryl M. Lamson, Alexis Russel, Matthew Shudt, Melissa A Leisner, Jonathan Plitnick, Navjot Singh, John Kelly, Erasmus Schneider, Erica Lasek-Nesselquist |
| EPI_ISL_1226463 | Columbia University Irving Medical Center | Wadsworth Center, New York State Department of Health | Kirsten St. George, Daryl M. Lamson, Alexis Russel, Matthew Shudt, Melissa A Leisner, Jonathan Plitnick, Navjot Singh, John Kelly, Erasmus Schneider, Erica Lasek-Nesselquist |
| EPI_ISL_1226464, EPI_ISL_1226465 | MONTEFIORE MEDICAL CENTER LABORATORIES | Wadsworth Center, New York State Department of Health | Kirsten St. George, Daryl M. Lamson, Alexis Russel, Matthew Shudt, Melissa A Leisner, Jonathan Plitnick, Navjot Singh, John Kelly, Erasmus Schneider, Erica Lasek-Nesselquist |
| EPI_ISL_1226466 | Columbia University Irving Medical Center | Wadsworth Center, New York State Department of Health | Kirsten St. George, Daryl M. Lamson, Alexis Russel, Matthew Shudt, Melissa A Leisner, Jonathan Plitnick, Navjot Singh, John Kelly, Erasmus Schneider, Erica Lasek-Nesselquist |
| EPI_ISL_1226467 | MEMORIAL SLOAN KETTERING CANCER CENTER | Wadsworth Center, New York State Department of Health | Kirsten St. George, Daryl M. Lamson, Alexis Russel, Matthew Shudt, Melissa A Leisner, Jonathan Plitnick, Navjot Singh, John Kelly, Erasmus Schneider, Erica Lasek-Nesselquist |
| EPI_ISL_1226468, EPI_ISL_1226469 | Wadsworth Center, New York State Department of Health | Wadsworth Center, New York State Department of Health | Kirsten St. George, Daryl M. Lamson, Alexis Russel, Matthew Shudt, Melissa A Leisner, Jonathan Plitnick, Navjot Singh, John Kelly, Erasmus Schneider, Erica Lasek-Nesselquist |
| EPI_ISL_1226470 | Masonic Care Community of NY | Wadsworth Center, New York State Department of Health | Kirsten St. George, Daryl M. Lamson, Alexis Russel, Matthew Shudt, Melissa A Leisner, Jonathan Plitnick, Navjot Singh, John Kelly, Erasmus Schneider, Erica Lasek-Nesselquist |
| EPI_ISL_1226471 | MEMORIAL SLOAN KETTERING CANCER CENTER | Wadsworth Center, New York State Department of Health | Kirsten St. George, Daryl M. Lamson, Alexis Russel, Matthew Shudt, Melissa A Leisner, Jonathan Plitnick, Navjot Singh, John Kelly, Erasmus Schneider, Erica Lasek-Nesselquist |

| | | | |
|---|---|---|---|
| EPI_ISL_1226472 | NORTHWELL HEALTH LABORATORIES | Wadsworth Center, New York State Department of Health | Kirsten St. George, Daryl M. Lamson, Alexis Russel, Matthew Shudt, Melissa A Leisner, Jonathan Plitnick, Navjot Singh, John Kelly, Erasmus Schneider, Erica Lasek-Nesselquist |
| EPI_ISL_1226473 | Masonic Care Community of NY | Wadsworth Center, New York State Department of Health | Kirsten St. George, Daryl M. Lamson, Alexis Russel, Matthew Shudt, Melissa A Leisner, Jonathan Plitnick, Navjot Singh, John Kelly, Erasmus Schneider, Erica Lasek-Nesselquist |
| EPI_ISL_1226474, EPI_ISL_1226475 | MEMORIAL SLOAN KETTERING CANCER CENTER | Wadsworth Center, New York State Department of Health | Kirsten St. George, Daryl M. Lamson, Alexis Russel, Matthew Shudt, Melissa A Leisner, Jonathan Plitnick, Navjot Singh, John Kelly, Erasmus Schneider, Erica Lasek-Nesselquist |
| EPI_ISL_1226476 | NYC Pandemic Response Lab | Wadsworth Center, New York State Department of Health | Kirsten St. George, Daryl M. Lamson, Alexis Russel, Matthew Shudt, Melissa A Leisner, Jonathan Plitnick, Navjot Singh, John Kelly, Erasmus Schneider, Erica Lasek-Nesselquist |
| EPI_ISL_1226477, EPI_ISL_1226478, EPI_ISL_1226479, EPI_ISL_1226480 | URMC LABS | Wadsworth Center, New York State Department of Health | Kirsten St. George, Daryl M. Lamson, Alexis Russel, Matthew Shudt, Melissa A Leisner, Jonathan Plitnick, Navjot Singh, John Kelly, Erasmus Schneider, Erica Lasek-Nesselquist |
| EPI_ISL_1226481 | MEMORIAL SLOAN KETTERING CANCER CENTER | Wadsworth Center, New York State Department of Health | Kirsten St. George, Daryl M. Lamson, Alexis Russel, Matthew Shudt, Melissa A Leisner, Jonathan Plitnick, Navjot Singh, John Kelly, Erasmus Schneider, Erica Lasek-Nesselquist |
| EPI_ISL_1226482, EPI_ISL_1226483, EPI_ISL_1226484 | Wadsworth Center, New York State Department of Health | Wadsworth Center, New York State Department of Health | Kirsten St. George, Daryl M. Lamson, Alexis Russel, Matthew Shudt, Melissa A Leisner, Jonathan Plitnick, Navjot Singh, John Kelly, Erasmus Schneider, Erica Lasek-Nesselquist |
| EPI_ISL_1226485 | ALBANY MEDICAL CENTER | Wadsworth Center, New York State Department of Health | Kirsten St. George, Daryl M. Lamson, Alexis Russel, Matthew Shudt, Melissa A Leisner, Jonathan Plitnick, Navjot Singh, John Kelly, Erasmus Schneider, Erica Lasek-Nesselquist |
| EPI_ISL_1226486 | URMC LABS | Wadsworth Center, New York State Department of Health | Kirsten St. George, Daryl M. Lamson, Alexis Russel, Matthew Shudt, Melissa A Leisner, Jonathan Plitnick, Navjot Singh, John Kelly, Erasmus Schneider, Erica Lasek-Nesselquist |
| EPI_ISL_1226487 | TEMPUS LABS INC | Wadsworth Center, New York State Department of Health | Kirsten St. George, Daryl M. Lamson, Alexis Russel, Matthew Shudt, Melissa A Leisner, Jonathan Plitnick, Navjot Singh, John Kelly, Erasmus Schneider, Erica Lasek-Nesselquist |
| EPI_ISL_1226488 | KALEIDA CENTER FOR LABORATORY MEDICINE | Wadsworth Center, New York State Department of Health | Kirsten St. George, Daryl M. Lamson, Alexis Russel, Matthew Shudt, Melissa A Leisner, Jonathan Plitnick, Navjot Singh, John Kelly, Erasmus Schneider, Erica Lasek-Nesselquist |
| EPI_ISL_1226489 | MONTEFIORE MEDICAL CENTER LABORATORIES | Wadsworth Center, New York State Department of Health | Kirsten St. George, Daryl M. Lamson, Alexis Russel, Matthew Shudt, Melissa A Leisner, Jonathan Plitnick, Navjot Singh, John Kelly, Erasmus Schneider, Erica Lasek-Nesselquist |
| EPI_ISL_1226490 | THE MARY IMOGENE BASSETT HOSPITAL | Wadsworth Center, New York State Department of Health | Kirsten St. George, Daryl M. Lamson, Alexis Russel, Matthew Shudt, Melissa A Leisner, Jonathan Plitnick, Navjot Singh, John Kelly, Erasmus Schneider, Erica Lasek-Nesselquist |
| EPI_ISL_1226491 | WESTCHESTER MEDICAL CENTER | Wadsworth Center, New York State Department of Health | Kirsten St. George, Daryl M. Lamson, Alexis Russel, Matthew Shudt, Melissa A Leisner, Jonathan Plitnick, Navjot Singh, John Kelly, Erasmus Schneider, Erica Lasek-Nesselquist |
| EPI_ISL_1226492, EPI_ISL_1226493 | MONTEFIORE MEDICAL CENTER LABORATORIES | Wadsworth Center, New York State Department of Health | Kirsten St. George, Daryl M. Lamson, Alexis Russel, Matthew Shudt, Melissa A Leisner, Jonathan Plitnick, Navjot Singh, John Kelly, Erasmus Schneider, Erica Lasek-Nesselquist |
| EPI_ISL_1226494 | MEMORIAL SLOAN KETTERING CANCER CENTER | Wadsworth Center, New York State Department of Health | Kirsten St. George, Daryl M. Lamson, Alexis Russel, Matthew Shudt, Melissa A Leisner, Jonathan Plitnick, Navjot Singh, John Kelly, Erasmus Schneider, Erica Lasek-Nesselquist |
| EPI_ISL_1226495 | TEMPUS LABS INC | Wadsworth Center, New York State Department of Health | Kirsten St. George, Daryl M. Lamson, Alexis Russel, Matthew Shudt, Melissa A Leisner, Jonathan Plitnick, Navjot Singh, John Kelly, Erasmus Schneider, Erica Lasek-Nesselquist |
| EPI_ISL_1226496, EPI_ISL_1226497 | WESTCHESTER MEDICAL CENTER | Wadsworth Center, New York State Department of Health | Kirsten St. George, Daryl M. Lamson, Alexis Russel, Matthew Shudt, Melissa A Leisner, Jonathan Plitnick, Navjot Singh, John Kelly, Erasmus Schneider, Erica Lasek-Nesselquist |
| EPI_ISL_1226498 | MONTEFIORE MEDICAL CENTER LABORATORIES | Wadsworth Center, New York State Department of Health | Kirsten St. George, Daryl M. Lamson, Alexis Russel, Matthew Shudt, Melissa A Leisner, Jonathan Plitnick, Navjot Singh, John Kelly, Erasmus Schneider, Erica Lasek-Nesselquist |
| EPI_ISL_1226499 | KALEIDA CENTER FOR LABORATORY MEDICINE | Wadsworth Center, New York State Department of Health | Kirsten St. George, Daryl M. Lamson, Alexis Russel, Matthew Shudt, Melissa A Leisner, Jonathan Plitnick, Navjot Singh, John Kelly, Erasmus Schneider, Erica Lasek-Nesselquist |
| EPI_ISL_1226500 | NYC Pandemic Response Lab | Wadsworth Center, New York State Department of Health | Kirsten St. George, Daryl M. Lamson, Alexis Russel, Matthew Shudt, Melissa A Leisner, Jonathan Plitnick, Navjot Singh, John Kelly, Erasmus Schneider, Erica Lasek-Nesselquist |
| EPI_ISL_1226501 | TEMPUS LABS INC | Wadsworth Center, New York State Department of Health | Kirsten St. George, Daryl M. Lamson, Alexis Russel, Matthew Shudt, Melissa A Leisner, Jonathan Plitnick, Navjot Singh, John Kelly, Erasmus Schneider, Erica Lasek-Nesselquist |
| EPI_ISL_1226502, EPI_ISL_1226503 | Columbia University Irving Medical Center | Wadsworth Center, New York State Department of Health | Kirsten St. George, Daryl M. Lamson, Alexis Russel, Matthew Shudt, Melissa A Leisner, Jonathan Plitnick, Navjot Singh, John Kelly, Erasmus Schneider, Erica Lasek-Nesselquist |
| EPI_ISL_1226504 | MEMORIAL SLOAN KETTERING CANCER CENTER | Wadsworth Center, New York State Department of Health | Kirsten St. George, Daryl M. Lamson, Alexis Russel, Matthew Shudt, Melissa A Leisner, Jonathan Plitnick, Navjot Singh, John Kelly, Erasmus Schneider, Erica Lasek-Nesselquist |
| EPI_ISL_1226505 | URMC LABS | Wadsworth Center, New York State Department of Health | Kirsten St. George, Daryl M. Lamson, Alexis Russel, Matthew Shudt, Melissa A Leisner, Jonathan Plitnick, Navjot Singh, John Kelly, Erasmus Schneider, Erica Lasek-Nesselquist |
| EPI_ISL_1226506 | NORTHWELL HEALTH LABORATORIES | Wadsworth Center, New York State Department of Health | Kirsten St. George, Daryl M. Lamson, Alexis Russel, Matthew Shudt, Melissa A Leisner, Jonathan Plitnick, Navjot Singh, John Kelly, Erasmus Schneider, Erica Lasek-Nesselquist |
| EPI_ISL_1226507, EPI_ISL_1226508 | NYC Pandemic Response Lab | Wadsworth Center, New York State Department of Health | Kirsten St. George, Daryl M. Lamson, Alexis Russel, Matthew Shudt, Melissa A Leisner, Jonathan Plitnick, Navjot Singh, John Kelly, Erasmus Schneider, Erica Lasek-Nesselquist |
| EPI_ISL_1226509 | MONTEFIORE MEDICAL CENTER LABORATORIES | Wadsworth Center, New York State Department of Health | Kirsten St. George, Daryl M. Lamson, Alexis Russel, Matthew Shudt, Melissa A Leisner, Jonathan Plitnick, Navjot Singh, John Kelly, Erasmus Schneider, Erica Lasek-Nesselquist |
| EPI_ISL_1226510 | NYC Pandemic Response Lab | Wadsworth Center, New York State Department of Health | Kirsten St. George, Daryl M. Lamson, Alexis Russel, Matthew Shudt, Melissa A Leisner, Jonathan Plitnick, Navjot Singh, John Kelly, Erasmus Schneider, Erica Lasek-Nesselquist |
| EPI_ISL_1226511 | ALBANY MEDICAL CENTER | Wadsworth Center, New York State Department of Health | Kirsten St. George, Daryl M. Lamson, Alexis Russel, Matthew Shudt, Melissa A Leisner, Jonathan Plitnick, Navjot Singh, John Kelly, Erasmus Schneider, Erica Lasek-Nesselquist |
| EPI_ISL_1226512 | Columbia University Irving Medical Center | Wadsworth Center, New York State Department of Health | Kirsten St. George, Daryl M. Lamson, Alexis Russel, Matthew Shudt, Melissa A Leisner, Jonathan Plitnick, Navjot Singh, John Kelly, Erasmus Schneider, Erica Lasek-Nesselquist |
| EPI_ISL_1226513 | NYC Pandemic Response Lab | Wadsworth Center, New York State Department of Health | Kirsten St. George, Daryl M. Lamson, Alexis Russel, Matthew Shudt, Melissa A Leisner, Jonathan Plitnick, Navjot Singh, John Kelly, Erasmus Schneider, Erica Lasek-Nesselquist |
| EPI_ISL_1226514 | URMC LABS | Wadsworth Center, New York State Department of Health | Kirsten St. George, Daryl M. Lamson, Alexis Russel, Matthew Shudt, Melissa A Leisner, Jonathan Plitnick, Navjot Singh, John Kelly, Erasmus Schneider, Erica Lasek-Nesselquist |
| EPI_ISL_1226515 | MONTEFIORE MEDICAL CENTER LABORATORIES | Wadsworth Center, New York State Department of Health | Kirsten St. George, Daryl M. Lamson, Alexis Russel, Matthew Shudt, Melissa A Leisner, Jonathan Plitnick, Navjot Singh, John Kelly, Erasmus Schneider, Erica Lasek-Nesselquist |
| EPI_ISL_1226516, EPI_ISL_1226517 | URMC LABS | Wadsworth Center, New York State Department of Health | Kirsten St. George, Daryl M. Lamson, Alexis Russel, Matthew Shudt, Melissa A Leisner, Jonathan Plitnick, Navjot Singh, John Kelly, Erasmus Schneider, Erica Lasek-Nesselquist |

| | | | |
|---|---|---|---|
| EPI_ISL_1226518 | Columbia University Irving Medical Center | Wadsworth Center, New York State Department of Health | Kirsten St. George, Daryl M. Lamson, Alexis Russel, Matthew Shudt, Melissa A Leisner, Jonathan Plitnick, Navjot Singh, John Kelly, Erasmus Schneider, Erica Lasek-Nesselquist |
| EPI_ISL_1226519 | Masonic Care Community of NY | Wadsworth Center, New York State Department of Health | Kirsten St. George, Daryl M. Lamson, Alexis Russel, Matthew Shudt, Melissa A Leisner, Jonathan Plitnick, Navjot Singh, John Kelly, Erasmus Schneider, Erica Lasek-Nesselquist |
| EPI_ISL_1226520 | URMC LABS | Wadsworth Center, New York State Department of Health | Kirsten St. George, Daryl M. Lamson, Alexis Russel, Matthew Shudt, Melissa A Leisner, Jonathan Plitnick, Navjot Singh, John Kelly, Erasmus Schneider, Erica Lasek-Nesselquist |
| EPI_ISL_1226521 | NYC Pandemic Response Lab | Wadsworth Center, New York State Department of Health | Kirsten St. George, Daryl M. Lamson, Alexis Russel, Matthew Shudt, Melissa A Leisner, Jonathan Plitnick, Navjot Singh, John Kelly, Erasmus Schneider, Erica Lasek-Nesselquist |
| EPI_ISL_1226522 | MEMORIAL SLOAN KETTERING CANCER CENTER | Wadsworth Center, New York State Department of Health | Kirsten St. George, Daryl M. Lamson, Alexis Russel, Matthew Shudt, Melissa A Leisner, Jonathan Plitnick, Navjot Singh, John Kelly, Erasmus Schneider, Erica Lasek-Nesselquist |
| EPI_ISL_1226523 | MONTEFIORE MEDICAL CENTER LABORATORIES | Wadsworth Center, New York State Department of Health | Kirsten St. George, Daryl M. Lamson, Alexis Russel, Matthew Shudt, Melissa A Leisner, Jonathan Plitnick, Navjot Singh, John Kelly, Erasmus Schneider, Erica Lasek-Nesselquist |
| EPI_ISL_1226524 | THE MARY IMOGENE BASSETT HOSPITAL | Wadsworth Center, New York State Department of Health | Kirsten St. George, Daryl M. Lamson, Alexis Russel, Matthew Shudt, Melissa A Leisner, Jonathan Plitnick, Navjot Singh, John Kelly, Erasmus Schneider, Erica Lasek-Nesselquist |
| EPI_ISL_1226525, EPI_ISL_1226526 | MEMORIAL SLOAN KETTERING CANCER CENTER | Wadsworth Center, New York State Department of Health | Kirsten St. George, Daryl M. Lamson, Alexis Russel, Matthew Shudt, Melissa A Leisner, Jonathan Plitnick, Navjot Singh, John Kelly, Erasmus Schneider, Erica Lasek-Nesselquist |
| EPI_ISL_1226527, EPI_ISL_1226528 | Wadsworth Center, New York State Department of Health | Wadsworth Center, New York State Department of Health | Kirsten St. George, Daryl M. Lamson, Alexis Russel, Matthew Shudt, Melissa A Leisner, Jonathan Plitnick, Navjot Singh, John Kelly, Erasmus Schneider, Erica Lasek-Nesselquist |
| EPI_ISL_1226529, EPI_ISL_1226530, EPI_ISL_1226531, EPI_ISL_1226532 | MONTEFIORE MEDICAL CENTER LABORATORIES | Wadsworth Center, New York State Department of Health | Kirsten St. George, Daryl M. Lamson, Alexis Russel, Matthew Shudt, Melissa A Leisner, Jonathan Plitnick, Navjot Singh, John Kelly, Erasmus Schneider, Erica Lasek-Nesselquist |
| EPI_ISL_1226533, EPI_ISL_1226534, EPI_ISL_1226535, EPI_ISL_1226536, EPI_ISL_1226537 | TEMPUS LABS INC | Wadsworth Center, New York State Department of Health | Kirsten St. George, Daryl M. Lamson, Alexis Russel, Matthew Shudt, Melissa A Leisner, Jonathan Plitnick, Navjot Singh, John Kelly, Erasmus Schneider, Erica Lasek-Nesselquist |
| EPI_ISL_1226538, EPI_ISL_1226539 | WESTCHESTER MEDICAL CENTER | Wadsworth Center, New York State Department of Health | Kirsten St. George, Daryl M. Lamson, Alexis Russel, Matthew Shudt, Melissa A Leisner, Jonathan Plitnick, Navjot Singh, John Kelly, Erasmus Schneider, Erica Lasek-Nesselquist |
| EPI_ISL_1226540 | BOSTON HEART DIAGNOSTICS CORP | Wadsworth Center, New York State Department of Health | Kirsten St. George, Daryl M. Lamson, Alexis Russel, Matthew Shudt, Melissa A Leisner, Jonathan Plitnick, Navjot Singh, John Kelly, Erasmus Schneider, Erica Lasek-Nesselquist |
| EPI_ISL_1226541, EPI_ISL_1226542 | KALEIDA CENTER FOR LABORATORY MEDICINE | Wadsworth Center, New York State Department of Health | Kirsten St. George, Daryl M. Lamson, Alexis Russel, Matthew Shudt, Melissa A Leisner, Jonathan Plitnick, Navjot Singh, John Kelly, Erasmus Schneider, Erica Lasek-Nesselquist |
| EPI_ISL_1226543 | MEMORIAL SLOAN KETTERING CANCER CENTER | Wadsworth Center, New York State Department of Health | Kirsten St. George, Daryl M. Lamson, Alexis Russel, Matthew Shudt, Melissa A Leisner, Jonathan Plitnick, Navjot Singh, John Kelly, Erasmus Schneider, Erica Lasek-Nesselquist |
| EPI_ISL_1226544 | New York Presbyterian Hospital | Wadsworth Center, New York State Department of Health | Kirsten St. George, Daryl M. Lamson, Alexis Russel, Matthew Shudt, Melissa A Leisner, Jonathan Plitnick, Navjot Singh, John Kelly, Erasmus Schneider, Erica Lasek-Nesselquist |
| EPI_ISL_1226545, EPI_ISL_1226546, EPI_ISL_1226547, EPI_ISL_1226548, EPI_ISL_1226549 | NYC Pandemic Response Lab | Wadsworth Center, New York State Department of Health | Kirsten St. George, Daryl M. Lamson, Alexis Russel, Matthew Shudt, Melissa A Leisner, Jonathan Plitnick, Navjot Singh, John Kelly, Erasmus Schneider, Erica Lasek-Nesselquist |
| EPI_ISL_1226550 | TEMPUS LABS INC | Wadsworth Center, New York State Department of Health | Kirsten St. George, Daryl M. Lamson, Alexis Russel, Matthew Shudt, Melissa A Leisner, Jonathan Plitnick, Navjot Singh, John Kelly, Erasmus Schneider, Erica Lasek-Nesselquist |
| EPI_ISL_1226551 | BIO-REFERENCE LABORATORIES | Wadsworth Center, New York State Department of Health | Kirsten St. George, Daryl M. Lamson, Alexis Russel, Matthew Shudt, Melissa A Leisner, Jonathan Plitnick, Navjot Singh, John Kelly, Erasmus Schneider, Erica Lasek-Nesselquist |
| EPI_ISL_1226552 | URMC LABS | Wadsworth Center, New York State Department of Health | Kirsten St. George, Daryl M. Lamson, Alexis Russel, Matthew Shudt, Melissa A Leisner, Jonathan Plitnick, Navjot Singh, John Kelly, Erasmus Schneider, Erica Lasek-Nesselquist |
| EPI_ISL_1226553 | NORTHWELL HEALTH LABORATORIES | Wadsworth Center, New York State Department of Health | Kirsten St. George, Daryl M. Lamson, Alexis Russel, Matthew Shudt, Melissa A Leisner, Jonathan Plitnick, Navjot Singh, John Kelly, Erasmus Schneider, Erica Lasek-Nesselquist |
| EPI_ISL_1226554 | NYC Pandemic Response Lab | Wadsworth Center, New York State Department of Health | Kirsten St. George, Daryl M. Lamson, Alexis Russel, Matthew Shudt, Melissa A Leisner, Jonathan Plitnick, Navjot Singh, John Kelly, Erasmus Schneider, Erica Lasek-Nesselquist |
| EPI_ISL_1226555 | TEMPUS LABS INC | Wadsworth Center, New York State Department of Health | Kirsten St. George, Daryl M. Lamson, Alexis Russel, Matthew Shudt, Melissa A Leisner, Jonathan Plitnick, Navjot Singh, John Kelly, Erasmus Schneider, Erica Lasek-Nesselquist |
| EPI_ISL_1226556 | BIO-REFERENCE LABORATORIES | Wadsworth Center, New York State Department of Health | Kirsten St. George, Daryl M. Lamson, Alexis Russel, Matthew Shudt, Melissa A Leisner, Jonathan Plitnick, Navjot Singh, John Kelly, Erasmus Schneider, Erica Lasek-Nesselquist |
| EPI_ISL_1226557 | TEMPUS LABS INC | Wadsworth Center, New York State Department of Health | Kirsten St. George, Daryl M. Lamson, Alexis Russel, Matthew Shudt, Melissa A Leisner, Jonathan Plitnick, Navjot Singh, John Kelly, Erasmus Schneider, Erica Lasek-Nesselquist |
| EPI_ISL_1226558 | WESTCHESTER MEDICAL CENTER | Wadsworth Center, New York State Department of Health | Kirsten St. George, Daryl M. Lamson, Alexis Russel, Matthew Shudt, Melissa A Leisner, Jonathan Plitnick, Navjot Singh, John Kelly, Erasmus Schneider, Erica Lasek-Nesselquist |
| EPI_ISL_1226559 | New York Presbyterian Hospital | Wadsworth Center, New York State Department of Health | Kirsten St. George, Daryl M. Lamson, Alexis Russel, Matthew Shudt, Melissa A Leisner, Jonathan Plitnick, Navjot Singh, John Kelly, Erasmus Schneider, Erica Lasek-Nesselquist |
| EPI_ISL_1226560, EPI_ISL_1226561, EPI_ISL_1226562 | NYC Pandemic Response Lab | Wadsworth Center, New York State Department of Health | Kirsten St. George, Daryl M. Lamson, Alexis Russel, Matthew Shudt, Melissa A Leisner, Jonathan Plitnick, Navjot Singh, John Kelly, Erasmus Schneider, Erica Lasek-Nesselquist |
| EPI_ISL_1226563 | ALBANY MEDICAL CENTER | Wadsworth Center, New York State Department of Health | Kirsten St. George, Daryl M. Lamson, Alexis Russel, Matthew Shudt, Melissa A Leisner, Jonathan Plitnick, Navjot Singh, John Kelly, Erasmus Schneider, Erica Lasek-Nesselquist |
| EPI_ISL_1226564, EPI_ISL_1226565, EPI_ISL_1226566 | NYC Pandemic Response Lab | Wadsworth Center, New York State Department of Health | Kirsten St. George, Daryl M. Lamson, Alexis Russel, Matthew Shudt, Melissa A Leisner, Jonathan Plitnick, Navjot Singh, John Kelly, Erasmus Schneider, Erica Lasek-Nesselquist |
| EPI_ISL_1226567 | TEMPUS LABS INC | Wadsworth Center, New York State Department of Health | Kirsten St. George, Daryl M. Lamson, Alexis Russel, Matthew Shudt, Melissa A Leisner, Jonathan Plitnick, Navjot Singh, John Kelly, Erasmus Schneider, Erica Lasek-Nesselquist |
| EPI_ISL_1226568 | WESTCHESTER MEDICAL CENTER | Wadsworth Center, New York State Department of Health | Kirsten St. George, Daryl M. Lamson, Alexis Russel, Matthew Shudt, Melissa A Leisner, Jonathan Plitnick, Navjot Singh, John Kelly, Erasmus Schneider, Erica Lasek-Nesselquist |
| EPI_ISL_1226569, EPI_ISL_1226570 | TEMPUS LABS INC | Wadsworth Center, New York State Department of Health | Kirsten St. George, Daryl M. Lamson, Alexis Russel, Matthew Shudt, Melissa A Leisner, Jonathan Plitnick, Navjot Singh, John Kelly, Erasmus Schneider, Erica Lasek-Nesselquist |
| EPI_ISL_1226571, EPI_ISL_1226572 | BIO-REFERENCE LABORATORIES | Wadsworth Center, New York State Department of Health | Kirsten St. George, Daryl M. Lamson, Alexis Russel, Matthew Shudt, Melissa A Leisner, Jonathan Plitnick, Navjot Singh, John Kelly, Erasmus Schneider, Erica Lasek-Nesselquist |
| EPI_ISL_1226573 | MONTEFIORE MEDICAL CENTER LABORATORIES | Wadsworth Center, New York State Department of Health | Kirsten St. George, Daryl M. Lamson, Alexis Russel, Matthew Shudt, Melissa A Leisner, Jonathan Plitnick, Navjot Singh, John Kelly, Erasmus Schneider, |

| | | | |
|---|---|---|---|
| | | | Erica Lasek-Nesselquist |
| EPI_ISL_1226574 | BIO-REFERENCE LABORATORIES | Wadsworth Center, New York State Department of Health | Kirsten St. George, Daryl M. Lamson, Alexis Russel, Matthew Shudt, Melissa A Leisner, Jonathan Plitnick, Navjot Singh, John Kelly, Erasmus Schneider, Erica Lasek-Nesselquist |
| EPI_ISL_1226575 | TEMPUS LABS INC | Wadsworth Center, New York State Department of Health | Kirsten St. George, Daryl M. Lamson, Alexis Russel, Matthew Shudt, Melissa A Leisner, Jonathan Plitnick, Navjot Singh, John Kelly, Erasmus Schneider, Erica Lasek-Nesselquist |
| EPI_ISL_1226576 | MONTEFIORE MEDICAL CENTER LABORATORIES | Wadsworth Center, New York State Department of Health | Kirsten St. George, Daryl M. Lamson, Alexis Russel, Matthew Shudt, Melissa A Leisner, Jonathan Plitnick, Navjot Singh, John Kelly, Erasmus Schneider, Erica Lasek-Nesselquist |
| EPI_ISL_1226577 | BIO-REFERENCE LABORATORIES | Wadsworth Center, New York State Department of Health | Kirsten St. George, Daryl M. Lamson, Alexis Russel, Matthew Shudt, Melissa A Leisner, Jonathan Plitnick, Navjot Singh, John Kelly, Erasmus Schneider, Erica Lasek-Nesselquist |
| EPI_ISL_1226578, EPI_ISL_1226579 | TEMPUS LABS INC | Wadsworth Center, New York State Department of Health | Kirsten St. George, Daryl M. Lamson, Alexis Russel, Matthew Shudt, Melissa A Leisner, Jonathan Plitnick, Navjot Singh, John Kelly, Erasmus Schneider, Erica Lasek-Nesselquist |
| EPI_ISL_1226580, EPI_ISL_1226581, EPI_ISL_1226582 | NYC Pandemic Response Lab | Wadsworth Center, New York State Department of Health | Kirsten St. George, Daryl M. Lamson, Alexis Russel, Matthew Shudt, Melissa A Leisner, Jonathan Plitnick, Navjot Singh, John Kelly, Erasmus Schneider, Erica Lasek-Nesselquist |
| EPI_ISL_1226583 | BIO-REFERENCE LABORATORIES | Wadsworth Center, New York State Department of Health | Kirsten St. George, Daryl M. Lamson, Alexis Russel, Matthew Shudt, Melissa A Leisner, Jonathan Plitnick, Navjot Singh, John Kelly, Erasmus Schneider, Erica Lasek-Nesselquist |
| EPI_ISL_1226584, EPI_ISL_1226585, EPI_ISL_1226586 | TEMPUS LABS INC | Wadsworth Center, New York State Department of Health | Kirsten St. George, Daryl M. Lamson, Alexis Russel, Matthew Shudt, Melissa A Leisner, Jonathan Plitnick, Navjot Singh, John Kelly, Erasmus Schneider, Erica Lasek-Nesselquist |
| EPI_ISL_1226587, EPI_ISL_1226588 | WESTCHESTER MEDICAL CENTER | Wadsworth Center, New York State Department of Health | Kirsten St. George, Daryl M. Lamson, Alexis Russel, Matthew Shudt, Melissa A Leisner, Jonathan Plitnick, Navjot Singh, John Kelly, Erasmus Schneider, Erica Lasek-Nesselquist |
| EPI_ISL_1226589 | TEMPUS LABS INC | Wadsworth Center, New York State Department of Health | Kirsten St. George, Daryl M. Lamson, Alexis Russel, Matthew Shudt, Melissa A Leisner, Jonathan Plitnick, Navjot Singh, John Kelly, Erasmus Schneider, Erica Lasek-Nesselquist |
| EPI_ISL_1226590, EPI_ISL_1226591 | WESTCHESTER MEDICAL CENTER | Wadsworth Center, New York State Department of Health | Kirsten St. George, Daryl M. Lamson, Alexis Russel, Matthew Shudt, Melissa A Leisner, Jonathan Plitnick, Navjot Singh, John Kelly, Erasmus Schneider, Erica Lasek-Nesselquist |
| EPI_ISL_1226592 | TEMPUS LABS INC | Wadsworth Center, New York State Department of Health | Kirsten St. George, Daryl M. Lamson, Alexis Russel, Matthew Shudt, Melissa A Leisner, Jonathan Plitnick, Navjot Singh, John Kelly, Erasmus Schneider, Erica Lasek-Nesselquist |
| EPI_ISL_1226593 | SUNY UPSTATE MEDICAL UNIVERSITY | Wadsworth Center, New York State Department of Health | Kirsten St. George, Daryl M. Lamson, Alexis Russel, Matthew Shudt, Melissa A Leisner, Jonathan Plitnick, Navjot Singh, John Kelly, Erasmus Schneider, Erica Lasek-Nesselquist |
| EPI_ISL_1226594 | MONTEFIORE MEDICAL CENTER LABORATORIES | Wadsworth Center, New York State Department of Health | Kirsten St. George, Daryl M. Lamson, Alexis Russel, Matthew Shudt, Melissa A Leisner, Jonathan Plitnick, Navjot Singh, John Kelly, Erasmus Schneider, Erica Lasek-Nesselquist |
| EPI_ISL_1226595 | NYC Pandemic Response Lab | Wadsworth Center, New York State Department of Health | Kirsten St. George, Daryl M. Lamson, Alexis Russel, Matthew Shudt, Melissa A Leisner, Jonathan Plitnick, Navjot Singh, John Kelly, Erasmus Schneider, Erica Lasek-Nesselquist |
| EPI_ISL_1226596 | Columbia University Irving Medical Center | Wadsworth Center, New York State Department of Health | Kirsten St. George, Daryl M. Lamson, Alexis Russel, Matthew Shudt, Melissa A Leisner, Jonathan Plitnick, Navjot Singh, John Kelly, Erasmus Schneider, Erica Lasek-Nesselquist |
| EPI_ISL_1226597 | MONTEFIORE MEDICAL CENTER LABORATORIES | Wadsworth Center, New York State Department of Health | Kirsten St. George, Daryl M. Lamson, Alexis Russel, Matthew Shudt, Melissa A Leisner, Jonathan Plitnick, Navjot Singh, John Kelly, Erasmus Schneider, Erica Lasek-Nesselquist |
| EPI_ISL_1226598 | URMC LABS | Wadsworth Center, New York State Department of Health | Kirsten St. George, Daryl M. Lamson, Alexis Russel, Matthew Shudt, Melissa A Leisner, Jonathan Plitnick, Navjot Singh, John Kelly, Erasmus Schneider, Erica Lasek-Nesselquist |
| EPI_ISL_1226599 | Columbia University Irving Medical Center | Wadsworth Center, New York State Department of Health | Kirsten St. George, Daryl M. Lamson, Alexis Russel, Matthew Shudt, Melissa A Leisner, Jonathan Plitnick, Navjot Singh, John Kelly, Erasmus Schneider, Erica Lasek-Nesselquist |
| EPI_ISL_1226600 | NYC Pandemic Response Lab | Wadsworth Center, New York State Department of Health | Kirsten St. George, Daryl M. Lamson, Alexis Russel, Matthew Shudt, Melissa A Leisner, Jonathan Plitnick, Navjot Singh, John Kelly, Erasmus Schneider, Erica Lasek-Nesselquist |
| EPI_ISL_1226601 | MONTEFIORE MEDICAL CENTER LABORATORIES | Wadsworth Center, New York State Department of Health | Kirsten St. George, Daryl M. Lamson, Alexis Russel, Matthew Shudt, Melissa A Leisner, Jonathan Plitnick, Navjot Singh, John Kelly, Erasmus Schneider, Erica Lasek-Nesselquist |
| EPI_ISL_1226602 | URMC LABS | Wadsworth Center, New York State Department of Health | Kirsten St. George, Daryl M. Lamson, Alexis Russel, Matthew Shudt, Melissa A Leisner, Jonathan Plitnick, Navjot Singh, John Kelly, Erasmus Schneider, Erica Lasek-Nesselquist |
| EPI_ISL_1226603 | TEMPUS LABS INC | Wadsworth Center, New York State Department of Health | Kirsten St. George, Daryl M. Lamson, Alexis Russel, Matthew Shudt, Melissa A Leisner, Jonathan Plitnick, Navjot Singh, John Kelly, Erasmus Schneider, Erica Lasek-Nesselquist |
| EPI_ISL_1226604, EPI_ISL_1226605, EPI_ISL_1226606, EPI_ISL_1226607, EPI_ISL_1226608, EPI_ISL_1226609, EPI_ISL_1226610, EPI_ISL_1226611, EPI_ISL_1226612, EPI_ISL_1226613, EPI_ISL_1226614 | | | |
| see above | MONTEFIORE MEDICAL CENTER LABORATORIES | Wadsworth Center, New York State Department of Health | Kirsten St. George, Daryl M. Lamson, Alexis Russel, Matthew Shudt, Melissa A Leisner, Jonathan Plitnick, Navjot Singh, John Kelly, Erasmus Schneider, Erica Lasek-Nesselquist |
| EPI_ISL_1226615 | URMC LABS | Wadsworth Center, New York State Department of Health | Kirsten St. George, Daryl M. Lamson, Alexis Russel, Matthew Shudt, Melissa A Leisner, Jonathan Plitnick, Navjot Singh, John Kelly, Erasmus Schneider, Erica Lasek-Nesselquist |
| EPI_ISL_1226616 | WESTCHESTER MEDICAL CENTER | Wadsworth Center, New York State Department of Health | Kirsten St. George, Daryl M. Lamson, Alexis Russel, Matthew Shudt, Melissa A Leisner, Jonathan Plitnick, Navjot Singh, John Kelly, Erasmus Schneider, Erica Lasek-Nesselquist |
| EPI_ISL_1226617 | KALEIDA CENTER FOR LABORATORY MEDICINE | Wadsworth Center, New York State Department of Health | Kirsten St. George, Daryl M. Lamson, Alexis Russel, Matthew Shudt, Melissa A Leisner, Jonathan Plitnick, Navjot Singh, John Kelly, Erasmus Schneider, Erica Lasek-Nesselquist |
| EPI_ISL_1226618, EPI_ISL_1226619 | TEMPUS LABS INC | Wadsworth Center, New York State Department of Health | Kirsten St. George, Daryl M. Lamson, Alexis Russel, Matthew Shudt, Melissa A Leisner, Jonathan Plitnick, Navjot Singh, John Kelly, Erasmus Schneider, Erica Lasek-Nesselquist |
| EPI_ISL_1226620, EPI_ISL_1226621 | NYC Pandemic Response Lab | Wadsworth Center, New York State Department of Health | Kirsten St. George, Daryl M. Lamson, Alexis Russel, Matthew Shudt, Melissa A Leisner, Jonathan Plitnick, Navjot Singh, John Kelly, Erasmus Schneider, Erica Lasek-Nesselquist |
| EPI_ISL_1226622 | BIO-REFERENCE LABORATORIES | Wadsworth Center, New York State Department of Health | Kirsten St. George, Daryl M. Lamson, Alexis Russel, Matthew Shudt, Melissa A Leisner, Jonathan Plitnick, Navjot Singh, John Kelly, Erasmus Schneider, Erica Lasek-Nesselquist |
| EPI_ISL_1226623 | URMC LABS | Wadsworth Center, New York State Department of Health | Kirsten St. George, Daryl M. Lamson, Alexis Russel, Matthew Shudt, Melissa A Leisner, Jonathan Plitnick, Navjot Singh, John Kelly, Erasmus Schneider, Erica Lasek-Nesselquist |
| EPI_ISL_1226624 | New York Presbyterian Hospital | Wadsworth Center, New York State Department of Health | Kirsten St. George, Daryl M. Lamson, Alexis Russel, Matthew Shudt, Melissa A Leisner, Jonathan Plitnick, Navjot Singh, John Kelly, Erasmus Schneider, Erica Lasek-Nesselquist |
| EPI_ISL_1226625 | NYC Pandemic Response Lab | Wadsworth Center, New York State Department of Health | Kirsten St. George, Daryl M. Lamson, Alexis Russel, Matthew Shudt, Melissa A Leisner, Jonathan Plitnick, Navjot Singh, John Kelly, Erasmus Schneider, Erica Lasek-Nesselquist |
| EPI_ISL_1226626, EPI_ISL_1226627, EPI_ISL_1226628, EPI_ISL_1226629, | TEMPUS LABS INC | Wadsworth Center, New York State Department of Health | Kirsten St. George, Daryl M. Lamson, Alexis Russel, Matthew Shudt, Melissa A Leisner, Jonathan Plitnick, Navjot Singh, John Kelly, Erasmus Schneider, Erica Lasek-Nesselquist |

| | | | |
|---|---|---|---|
| EPI_ISL_1226630, EPI_ISL_1226631, EPI_ISL_1226632, EPI_ISL_1226633, EPI_ISL_1226634, EPI_ISL_1226635 | | | |
| EPI_ISL_1226636 | NYC Pandemic Response Lab | Wadsworth Center, New York State Department of Health | Kirsten St. George, Daryl M. Lamson, Alexis Russel, Matthew Shudt, Melissa A Leisner, Jonathan Plitnick, Navjot Singh, John Kelly, Erasmus Schneider, Erica Lasek-Nesselquist |
| EPI_ISL_1226637, EPI_ISL_1226638 | Columbia University Irving Medical Center | Wadsworth Center, New York State Department of Health | Kirsten St. George, Daryl M. Lamson, Alexis Russel, Matthew Shudt, Melissa A Leisner, Jonathan Plitnick, Navjot Singh, John Kelly, Erasmus Schneider, Erica Lasek-Nesselquist |
| EPI_ISL_1226639, EPI_ISL_1226640, EPI_ISL_1226641, EPI_ISL_1226642, EPI_ISL_1226643, EPI_ISL_1226644 | NYC Pandemic Response Lab | Wadsworth Center, New York State Department of Health | Kirsten St. George, Daryl M. Lamson, Alexis Russel, Matthew Shudt, Melissa A Leisner, Jonathan Plitnick, Navjot Singh, John Kelly, Erasmus Schneider, Erica Lasek-Nesselquist |
| EPI_ISL_1226645, EPI_ISL_1226646 | BIO-REFERENCE LABORATORIES | Wadsworth Center, New York State Department of Health | Kirsten St. George, Daryl M. Lamson, Alexis Russel, Matthew Shudt, Melissa A Leisner, Jonathan Plitnick, Navjot Singh, John Kelly, Erasmus Schneider, Erica Lasek-Nesselquist |
| EPI_ISL_1226647, EPI_ISL_1226648 | NYC Pandemic Response Lab | Wadsworth Center, New York State Department of Health | Kirsten St. George, Daryl M. Lamson, Alexis Russel, Matthew Shudt, Melissa A Leisner, Jonathan Plitnick, Navjot Singh, John Kelly, Erasmus Schneider, Erica Lasek-Nesselquist |
| EPI_ISL_1226649, EPI_ISL_1226650 | TEMPUS LABS INC | Wadsworth Center, New York State Department of Health | Kirsten St. George, Daryl M. Lamson, Alexis Russel, Matthew Shudt, Melissa A Leisner, Jonathan Plitnick, Navjot Singh, John Kelly, Erasmus Schneider, Erica Lasek-Nesselquist |
| EPI_ISL_1226651, EPI_ISL_1226652 | NYC Pandemic Response Lab | Wadsworth Center, New York State Department of Health | Kirsten St. George, Daryl M. Lamson, Alexis Russel, Matthew Shudt, Melissa A Leisner, Jonathan Plitnick, Navjot Singh, John Kelly, Erasmus Schneider, Erica Lasek-Nesselquist |
| EPI_ISL_1226653 | TEMPUS LABS INC | Wadsworth Center, New York State Department of Health | Kirsten St. George, Daryl M. Lamson, Alexis Russel, Matthew Shudt, Melissa A Leisner, Jonathan Plitnick, Navjot Singh, John Kelly, Erasmus Schneider, Erica Lasek-Nesselquist |
| EPI_ISL_1226654 | MONTEFIORE MEDICAL CENTER LABORATORIES | Wadsworth Center, New York State Department of Health | Kirsten St. George, Daryl M. Lamson, Alexis Russel, Matthew Shudt, Melissa A Leisner, Jonathan Plitnick, Navjot Singh, John Kelly, Erasmus Schneider, Erica Lasek-Nesselquist |
| EPI_ISL_1226655, EPI_ISL_1226656, EPI_ISL_1226657 | WESTCHESTER MEDICAL CENTER | Wadsworth Center, New York State Department of Health | Kirsten St. George, Daryl M. Lamson, Alexis Russel, Matthew Shudt, Melissa A Leisner, Jonathan Plitnick, Navjot Singh, John Kelly, Erasmus Schneider, Erica Lasek-Nesselquist |
| EPI_ISL_1226658 | BIO-REFERENCE LABORATORIES | Wadsworth Center, New York State Department of Health | Kirsten St. George, Daryl M. Lamson, Alexis Russel, Matthew Shudt, Melissa A Leisner, Jonathan Plitnick, Navjot Singh, John Kelly, Erasmus Schneider, Erica Lasek-Nesselquist |
| EPI_ISL_1226659 | TEMPUS LABS INC | Wadsworth Center, New York State Department of Health | Kirsten St. George, Daryl M. Lamson, Alexis Russel, Matthew Shudt, Melissa A Leisner, Jonathan Plitnick, Navjot Singh, John Kelly, Erasmus Schneider, Erica Lasek-Nesselquist |
| EPI_ISL_1226660, EPI_ISL_1226661 | NYC Pandemic Response Lab | Wadsworth Center, New York State Department of Health | Kirsten St. George, Daryl M. Lamson, Alexis Russel, Matthew Shudt, Melissa A Leisner, Jonathan Plitnick, Navjot Singh, John Kelly, Erasmus Schneider, Erica Lasek-Nesselquist |
| EPI_ISL_1226662, EPI_ISL_1226663 | URMC LABS | Wadsworth Center, New York State Department of Health | Kirsten St. George, Daryl M. Lamson, Alexis Russel, Matthew Shudt, Melissa A Leisner, Jonathan Plitnick, Navjot Singh, John Kelly, Erasmus Schneider, Erica Lasek-Nesselquist |
| EPI_ISL_1226664 | MEMORIAL SLOAN KETTERING CANCER CENTER | Wadsworth Center, New York State Department of Health | Kirsten St. George, Daryl M. Lamson, Alexis Russel, Matthew Shudt, Melissa A Leisner, Jonathan Plitnick, Navjot Singh, John Kelly, Erasmus Schneider, Erica Lasek-Nesselquist |
| EPI_ISL_1226665 | NYC Pandemic Response Lab | Wadsworth Center, New York State Department of Health | Kirsten St. George, Daryl M. Lamson, Alexis Russel, Matthew Shudt, Melissa A Leisner, Jonathan Plitnick, Navjot Singh, John Kelly, Erasmus Schneider, Erica Lasek-Nesselquist |
| EPI_ISL_1226666, EPI_ISL_1226667 | TEMPUS LABS INC | Wadsworth Center, New York State Department of Health | Kirsten St. George, Daryl M. Lamson, Alexis Russel, Matthew Shudt, Melissa A Leisner, Jonathan Plitnick, Navjot Singh, John Kelly, Erasmus Schneider, Erica Lasek-Nesselquist |
| EPI_ISL_1226668 | WESTCHESTER MEDICAL CENTER | Wadsworth Center, New York State Department of Health | Kirsten St. George, Daryl M. Lamson, Alexis Russel, Matthew Shudt, Melissa A Leisner, Jonathan Plitnick, Navjot Singh, John Kelly, Erasmus Schneider, Erica Lasek-Nesselquist |
| EPI_ISL_1226669 | BIO-REFERENCE LABORATORIES | Wadsworth Center, New York State Department of Health | Kirsten St. George, Daryl M. Lamson, Alexis Russel, Matthew Shudt, Melissa A Leisner, Jonathan Plitnick, Navjot Singh, John Kelly, Erasmus Schneider, Erica Lasek-Nesselquist |
| EPI_ISL_1226670 | TEMPUS LABS INC | Wadsworth Center, New York State Department of Health | Kirsten St. George, Daryl M. Lamson, Alexis Russel, Matthew Shudt, Melissa A Leisner, Jonathan Plitnick, Navjot Singh, John Kelly, Erasmus Schneider, Erica Lasek-Nesselquist |
| EPI_ISL_1226671 | Columbia University Irving Medical Center | Wadsworth Center, New York State Department of Health | Kirsten St. George, Daryl M. Lamson, Alexis Russel, Matthew Shudt, Melissa A Leisner, Jonathan Plitnick, Navjot Singh, John Kelly, Erasmus Schneider, Erica Lasek-Nesselquist |
| EPI_ISL_1226672 | NORTHWELL HEALTH LABORATORIES | Wadsworth Center, New York State Department of Health | Kirsten St. George, Daryl M. Lamson, Alexis Russel, Matthew Shudt, Melissa A Leisner, Jonathan Plitnick, Navjot Singh, John Kelly, Erasmus Schneider, Erica Lasek-Nesselquist |
| EPI_ISL_1226673 | NYC Pandemic Response Lab | Wadsworth Center, New York State Department of Health | Kirsten St. George, Daryl M. Lamson, Alexis Russel, Matthew Shudt, Melissa A Leisner, Jonathan Plitnick, Navjot Singh, John Kelly, Erasmus Schneider, Erica Lasek-Nesselquist |
| EPI_ISL_1226674 | Columbia University Irving Medical Center | Wadsworth Center, New York State Department of Health | Kirsten St. George, Daryl M. Lamson, Alexis Russel, Matthew Shudt, Melissa A Leisner, Jonathan Plitnick, Navjot Singh, John Kelly, Erasmus Schneider, Erica Lasek-Nesselquist |
| EPI_ISL_1226675, EPI_ISL_1226676, EPI_ISL_1226677 | URMC LABS | Wadsworth Center, New York State Department of Health | Kirsten St. George, Daryl M. Lamson, Alexis Russel, Matthew Shudt, Melissa A Leisner, Jonathan Plitnick, Navjot Singh, John Kelly, Erasmus Schneider, Erica Lasek-Nesselquist |
| EPI_ISL_1226678, EPI_ISL_1226679 | WESTCHESTER MEDICAL CENTER | Wadsworth Center, New York State Department of Health | Kirsten St. George, Daryl M. Lamson, Alexis Russel, Matthew Shudt, Melissa A Leisner, Jonathan Plitnick, Navjot Singh, John Kelly, Erasmus Schneider, Erica Lasek-Nesselquist |
| EPI_ISL_1226680, EPI_ISL_1226681 | MONTEFIORE MEDICAL CENTER LABORATORIES | Wadsworth Center, New York State Department of Health | Kirsten St. George, Daryl M. Lamson, Alexis Russel, Matthew Shudt, Melissa A Leisner, Jonathan Plitnick, Navjot Singh, John Kelly, Erasmus Schneider, Erica Lasek-Nesselquist |
| EPI_ISL_1226682 | WESTCHESTER MEDICAL CENTER | Wadsworth Center, New York State Department of Health | Kirsten St. George, Daryl M. Lamson, Alexis Russel, Matthew Shudt, Melissa A Leisner, Jonathan Plitnick, Navjot Singh, John Kelly, Erasmus Schneider, Erica Lasek-Nesselquist |
| EPI_ISL_1226683 | SUNY UPSTATE MEDICAL UNIVERSITY | Wadsworth Center, New York State Department of Health | Kirsten St. George, Daryl M. Lamson, Alexis Russel, Matthew Shudt, Melissa A Leisner, Jonathan Plitnick, Navjot Singh, John Kelly, Erasmus Schneider, Erica Lasek-Nesselquist |
| EPI_ISL_1226684 | KALEIDA CENTER FOR LABORATORY MEDICINE | Wadsworth Center, New York State Department of Health | Kirsten St. George, Daryl M. Lamson, Alexis Russel, Matthew Shudt, Melissa A Leisner, Jonathan Plitnick, Navjot Singh, John Kelly, Erasmus Schneider, Erica Lasek-Nesselquist |
| EPI_ISL_1226685 | NYC Pandemic Response Lab | Wadsworth Center, New York State Department of Health | Kirsten St. George, Daryl M. Lamson, Alexis Russel, Matthew Shudt, Melissa A Leisner, Jonathan Plitnick, Navjot Singh, John Kelly, Erasmus Schneider, Erica Lasek-Nesselquist |
| EPI_ISL_1226686, EPI_ISL_1226687, EPI_ISL_1226688 | WESTCHESTER MEDICAL CENTER | Wadsworth Center, New York State Department of Health | Kirsten St. George, Daryl M. Lamson, Alexis Russel, Matthew Shudt, Melissa A Leisner, Jonathan Plitnick, Navjot Singh, John Kelly, Erasmus Schneider, Erica Lasek-Nesselquist |
| EPI_ISL_1226689, EPI_ISL_1226690, EPI_ISL_1226691 | MONTEFIORE MEDICAL CENTER LABORATORIES | Wadsworth Center, New York State Department of Health | Kirsten St. George, Daryl M. Lamson, Alexis Russel, Matthew Shudt, Melissa A Leisner, Jonathan Plitnick, Navjot Singh, John Kelly, Erasmus Schneider, Erica Lasek-Nesselquist |
| EPI_ISL_1226692 | NYC Pandemic Response Lab | Wadsworth Center, New York State Department of Health | Kirsten St. George, Daryl M. Lamson, Alexis Russel, Matthew Shudt, Melissa A Leisner, Jonathan Plitnick, Navjot Singh, John Kelly, Erasmus Schneider, |

| | | | |
|---|---|---|---|
| | | | Erica Lasek-Nesselquist |
| EPI_ISL_1226693, EPI_ISL_1226694, EPI_ISL_1226695 | WESTCHESTER MEDICAL CENTER | Wadsworth Center, New York State Department of Health | Kirsten St. George, Daryl M. Lamson, Alexis Russel, Matthew Shudt, Melissa A Leisner, Jonathan Plitnick, Navjot Singh, John Kelly, Erasmus Schneider, Erica Lasek-Nesselquist |
| EPI_ISL_1226696 | KALEIDA CENTER FOR LABORATORY MEDICINE | Wadsworth Center, New York State Department of Health | Kirsten St. George, Daryl M. Lamson, Alexis Russel, Matthew Shudt, Melissa A Leisner, Jonathan Plitnick, Navjot Singh, John Kelly, Erasmus Schneider, Erica Lasek-Nesselquist |
| EPI_ISL_1226697 | WESTCHESTER MEDICAL CENTER | Wadsworth Center, New York State Department of Health | Kirsten St. George, Daryl M. Lamson, Alexis Russel, Matthew Shudt, Melissa A Leisner, Jonathan Plitnick, Navjot Singh, John Kelly, Erasmus Schneider, Erica Lasek-Nesselquist |
| EPI_ISL_1226698 | Wadsworth Center, New York State Department of Health | Wadsworth Center, New York State Department of Health | Kirsten St. George, Daryl M. Lamson, Alexis Russel, Matthew Shudt, Melissa A Leisner, Jonathan Plitnick, Navjot Singh, John Kelly, Erasmus Schneider, Erica Lasek-Nesselquist |
| EPI_ISL_1226699 | NYC Pandemic Response Lab | Wadsworth Center, New York State Department of Health | Kirsten St. George, Daryl M. Lamson, Alexis Russel, Matthew Shudt, Melissa A Leisner, Jonathan Plitnick, Navjot Singh, John Kelly, Erasmus Schneider, Erica Lasek-Nesselquist |
| EPI_ISL_1226700, EPI_ISL_1226701 | MEMORIAL SLOAN KETTERING CANCER CENTER | Wadsworth Center, New York State Department of Health | Kirsten St. George, Daryl M. Lamson, Alexis Russel, Matthew Shudt, Melissa A Leisner, Jonathan Plitnick, Navjot Singh, John Kelly, Erasmus Schneider, Erica Lasek-Nesselquist |
| EPI_ISL_1226702 | WESTCHESTER MEDICAL CENTER | Wadsworth Center, New York State Department of Health | Kirsten St. George, Daryl M. Lamson, Alexis Russel, Matthew Shudt, Melissa A Leisner, Jonathan Plitnick, Navjot Singh, John Kelly, Erasmus Schneider, Erica Lasek-Nesselquist |
| EPI_ISL_1226703 | NYC Pandemic Response Lab | Wadsworth Center, New York State Department of Health | Kirsten St. George, Daryl M. Lamson, Alexis Russel, Matthew Shudt, Melissa A Leisner, Jonathan Plitnick, Navjot Singh, John Kelly, Erasmus Schneider, Erica Lasek-Nesselquist |
| EPI_ISL_1226704, EPI_ISL_1226705, EPI_ISL_1226706 | ALBANY MEDICAL CENTER | Wadsworth Center, New York State Department of Health | Kirsten St. George, Daryl M. Lamson, Alexis Russel, Matthew Shudt, Melissa A Leisner, Jonathan Plitnick, Navjot Singh, John Kelly, Erasmus Schneider, Erica Lasek-Nesselquist |
| EPI_ISL_1226707 | NORTHWELL HEALTH LABORATORIES | Wadsworth Center, New York State Department of Health | Kirsten St. George, Daryl M. Lamson, Alexis Russel, Matthew Shudt, Melissa A Leisner, Jonathan Plitnick, Navjot Singh, John Kelly, Erasmus Schneider, Erica Lasek-Nesselquist |
| EPI_ISL_1226708 | WESTCHESTER MEDICAL CENTER | Wadsworth Center, New York State Department of Health | Kirsten St. George, Daryl M. Lamson, Alexis Russel, Matthew Shudt, Melissa A Leisner, Jonathan Plitnick, Navjot Singh, John Kelly, Erasmus Schneider, Erica Lasek-Nesselquist |
| EPI_ISL_1226709 | TEMPUS LABS INC | Wadsworth Center, New York State Department of Health | Kirsten St. George, Daryl M. Lamson, Alexis Russel, Matthew Shudt, Melissa A Leisner, Jonathan Plitnick, Navjot Singh, John Kelly, Erasmus Schneider, Erica Lasek-Nesselquist |
| EPI_ISL_1226710 | MEMORIAL SLOAN KETTERING CANCER CENTER | Wadsworth Center, New York State Department of Health | Kirsten St. George, Daryl M. Lamson, Alexis Russel, Matthew Shudt, Melissa A Leisner, Jonathan Plitnick, Navjot Singh, John Kelly, Erasmus Schneider, Erica Lasek-Nesselquist |
| EPI_ISL_1226711 | NYC Pandemic Response Lab | Wadsworth Center, New York State Department of Health | Kirsten St. George, Daryl M. Lamson, Alexis Russel, Matthew Shudt, Melissa A Leisner, Jonathan Plitnick, Navjot Singh, John Kelly, Erasmus Schneider, Erica Lasek-Nesselquist |
| EPI_ISL_1226712 | TEMPUS LABS INC | Wadsworth Center, New York State Department of Health | Kirsten St. George, Daryl M. Lamson, Alexis Russel, Matthew Shudt, Melissa A Leisner, Jonathan Plitnick, Navjot Singh, John Kelly, Erasmus Schneider, Erica Lasek-Nesselquist |
| EPI_ISL_1226713 | URMC LABS | Wadsworth Center, New York State Department of Health | Kirsten St. George, Daryl M. Lamson, Alexis Russel, Matthew Shudt, Melissa A Leisner, Jonathan Plitnick, Navjot Singh, John Kelly, Erasmus Schneider, Erica Lasek-Nesselquist |
| EPI_ISL_1226714, EPI_ISL_1226715 | MONTEFIORE MEDICAL CENTER LABORATORIES | Wadsworth Center, New York State Department of Health | Kirsten St. George, Daryl M. Lamson, Alexis Russel, Matthew Shudt, Melissa A Leisner, Jonathan Plitnick, Navjot Singh, John Kelly, Erasmus Schneider, Erica Lasek-Nesselquist |
| EPI_ISL_1226716 | Wadsworth Center, New York State Department of Health | Wadsworth Center, New York State Department of Health | Kirsten St. George, Daryl M. Lamson, Alexis Russel, Matthew Shudt, Melissa A Leisner, Jonathan Plitnick, Navjot Singh, John Kelly, Erasmus Schneider, Erica Lasek-Nesselquist |
| EPI_ISL_1226717 | NORTHWELL HEALTH LABORATORIES | Wadsworth Center, New York State Department of Health | Kirsten St. George, Daryl M. Lamson, Alexis Russel, Matthew Shudt, Melissa A Leisner, Jonathan Plitnick, Navjot Singh, John Kelly, Erasmus Schneider, Erica Lasek-Nesselquist |
| EPI_ISL_1226718, EPI_ISL_1226719 | NYC Pandemic Response Lab | Wadsworth Center, New York State Department of Health | Kirsten St. George, Daryl M. Lamson, Alexis Russel, Matthew Shudt, Melissa A Leisner, Jonathan Plitnick, Navjot Singh, John Kelly, Erasmus Schneider, Erica Lasek-Nesselquist |
| EPI_ISL_1226720 | NORTHWELL HEALTH LABORATORIES | Wadsworth Center, New York State Department of Health | Kirsten St. George, Daryl M. Lamson, Alexis Russel, Matthew Shudt, Melissa A Leisner, Jonathan Plitnick, Navjot Singh, John Kelly, Erasmus Schneider, Erica Lasek-Nesselquist |
| EPI_ISL_1226721 | WESTCHESTER MEDICAL CENTER | Wadsworth Center, New York State Department of Health | Kirsten St. George, Daryl M. Lamson, Alexis Russel, Matthew Shudt, Melissa A Leisner, Jonathan Plitnick, Navjot Singh, John Kelly, Erasmus Schneider, Erica Lasek-Nesselquist |
| EPI_ISL_1226722 | MONTEFIORE MEDICAL CENTER LABORATORIES | Wadsworth Center, New York State Department of Health | Kirsten St. George, Daryl M. Lamson, Alexis Russel, Matthew Shudt, Melissa A Leisner, Jonathan Plitnick, Navjot Singh, John Kelly, Erasmus Schneider, Erica Lasek-Nesselquist |
| EPI_ISL_1226723 | WESTCHESTER MEDICAL CENTER | Wadsworth Center, New York State Department of Health | Kirsten St. George, Daryl M. Lamson, Alexis Russel, Matthew Shudt, Melissa A Leisner, Jonathan Plitnick, Navjot Singh, John Kelly, Erasmus Schneider, Erica Lasek-Nesselquist |
| EPI_ISL_1226724 | MONTEFIORE MEDICAL CENTER LABORATORIES | Wadsworth Center, New York State Department of Health | Kirsten St. George, Daryl M. Lamson, Alexis Russel, Matthew Shudt, Melissa A Leisner, Jonathan Plitnick, Navjot Singh, John Kelly, Erasmus Schneider, Erica Lasek-Nesselquist |
| EPI_ISL_1226725, EPI_ISL_1226726 | MEMORIAL SLOAN KETTERING CANCER CENTER | Wadsworth Center, New York State Department of Health | Kirsten St. George, Daryl M. Lamson, Alexis Russel, Matthew Shudt, Melissa A Leisner, Jonathan Plitnick, Navjot Singh, John Kelly, Erasmus Schneider, Erica Lasek-Nesselquist |
| EPI_ISL_1226727 | WESTCHESTER MEDICAL CENTER | Wadsworth Center, New York State Department of Health | Kirsten St. George, Daryl M. Lamson, Alexis Russel, Matthew Shudt, Melissa A Leisner, Jonathan Plitnick, Navjot Singh, John Kelly, Erasmus Schneider, Erica Lasek-Nesselquist |
| EPI_ISL_1226728 | Columbia University Irving Medical Center | Wadsworth Center, New York State Department of Health | Kirsten St. George, Daryl M. Lamson, Alexis Russel, Matthew Shudt, Melissa A Leisner, Jonathan Plitnick, Navjot Singh, John Kelly, Erasmus Schneider, Erica Lasek-Nesselquist |
| EPI_ISL_1226729 | NYC Pandemic Response Lab | Wadsworth Center, New York State Department of Health | Kirsten St. George, Daryl M. Lamson, Alexis Russel, Matthew Shudt, Melissa A Leisner, Jonathan Plitnick, Navjot Singh, John Kelly, Erasmus Schneider, Erica Lasek-Nesselquist |
| EPI_ISL_1226730 | WESTCHESTER MEDICAL CENTER | Wadsworth Center, New York State Department of Health | Kirsten St. George, Daryl M. Lamson, Alexis Russel, Matthew Shudt, Melissa A Leisner, Jonathan Plitnick, Navjot Singh, John Kelly, Erasmus Schneider, Erica Lasek-Nesselquist |
| EPI_ISL_1226731 | MEMORIAL SLOAN KETTERING CANCER CENTER | Wadsworth Center, New York State Department of Health | Kirsten St. George, Daryl M. Lamson, Alexis Russel, Matthew Shudt, Melissa A Leisner, Jonathan Plitnick, Navjot Singh, John Kelly, Erasmus Schneider, Erica Lasek-Nesselquist |
| EPI_ISL_1226732 | SUNY UPSTATE MEDICAL UNIVERSITY | Wadsworth Center, New York State Department of Health | Kirsten St. George, Daryl M. Lamson, Alexis Russel, Matthew Shudt, Melissa A Leisner, Jonathan Plitnick, Navjot Singh, John Kelly, Erasmus Schneider, Erica Lasek-Nesselquist |
| EPI_ISL_1226733 | MEMORIAL SLOAN KETTERING CANCER CENTER | Wadsworth Center, New York State Department of Health | Kirsten St. George, Daryl M. Lamson, Alexis Russel, Matthew Shudt, Melissa A Leisner, Jonathan Plitnick, Navjot Singh, John Kelly, Erasmus Schneider, Erica Lasek-Nesselquist |
| EPI_ISL_1226734, EPI_ISL_1226735 | MONTEFIORE MEDICAL CENTER LABORATORIES | Wadsworth Center, New York State Department of Health | Kirsten St. George, Daryl M. Lamson, Alexis Russel, Matthew Shudt, Melissa A Leisner, Jonathan Plitnick, Navjot Singh, John Kelly, Erasmus Schneider, Erica Lasek-Nesselquist |
| EPI_ISL_1226736 | MEMORIAL SLOAN KETTERING CANCER CENTER | Wadsworth Center, New York State Department of Health | Kirsten St. George, Daryl M. Lamson, Alexis Russel, Matthew Shudt, Melissa A Leisner, Jonathan Plitnick, Navjot Singh, John Kelly, Erasmus Schneider, |

| | | | |
|---|---|---|---|
| | | | Erica Lasek-Nesselquist |
| EPI_ISL_1226737, EPI_ISL_1226738 | MONTEFIORE MEDICAL CENTER LABORATORIES | Wadsworth Center, New York State Department of Health | Kirsten St. George, Daryl M. Lamson, Alexis Russel, Matthew Shudt, Melissa A Leisner, Jonathan Plitnick, Navjot Singh, John Kelly, Erasmus Schneider, Erica Lasek-Nesselquist |
| EPI_ISL_1226739 | NYC Pandemic Response Lab | Wadsworth Center, New York State Department of Health | Kirsten St. George, Daryl M. Lamson, Alexis Russel, Matthew Shudt, Melissa A Leisner, Jonathan Plitnick, Navjot Singh, John Kelly, Erasmus Schneider, Erica Lasek-Nesselquist |
| EPI_ISL_1226740, EPI_ISL_1226741, EPI_ISL_1226742 | MONTEFIORE MEDICAL CENTER LABORATORIES | Wadsworth Center, New York State Department of Health | Kirsten St. George, Daryl M. Lamson, Alexis Russel, Matthew Shudt, Melissa A Leisner, Jonathan Plitnick, Navjot Singh, John Kelly, Erasmus Schneider, Erica Lasek-Nesselquist |
| EPI_ISL_1226743 | Columbia University Irving Medical Center | Wadsworth Center, New York State Department of Health | Kirsten St. George, Daryl M. Lamson, Alexis Russel, Matthew Shudt, Melissa A Leisner, Jonathan Plitnick, Navjot Singh, John Kelly, Erasmus Schneider, Erica Lasek-Nesselquist |
| EPI_ISL_1226744, EPI_ISL_1226745, EPI_ISL_1226746, EPI_ISL_1226747, EPI_ISL_1226748, EPI_ISL_1226749, EPI_ISL_1226750, EPI_ISL_1226751, EPI_ISL_1226752, EPI_ISL_1226753, EPI_ISL_1226754, EPI_ISL_1226755 | | | |
| see above | MONTEFIORE MEDICAL CENTER LABORATORIES | Wadsworth Center, New York State Department of Health | Kirsten St. George, Daryl M. Lamson, Alexis Russel, Matthew Shudt, Melissa A Leisner, Jonathan Plitnick, Navjot Singh, John Kelly, Erasmus Schneider, Erica Lasek-Nesselquist |
| EPI_ISL_1226756 | WESTCHESTER MEDICAL CENTER | Wadsworth Center, New York State Department of Health | Kirsten St. George, Daryl M. Lamson, Alexis Russel, Matthew Shudt, Melissa A Leisner, Jonathan Plitnick, Navjot Singh, John Kelly, Erasmus Schneider, Erica Lasek-Nesselquist |
| EPI_ISL_1226757 | New York Presbyterian Hospital | Wadsworth Center, New York State Department of Health | Kirsten St. George, Daryl M. Lamson, Alexis Russel, Matthew Shudt, Melissa A Leisner, Jonathan Plitnick, Navjot Singh, John Kelly, Erasmus Schneider, Erica Lasek-Nesselquist |
| EPI_ISL_1226758 | MONTEFIORE MEDICAL CENTER LABORATORIES | Wadsworth Center, New York State Department of Health | Kirsten St. George, Daryl M. Lamson, Alexis Russel, Matthew Shudt, Melissa A Leisner, Jonathan Plitnick, Navjot Singh, John Kelly, Erasmus Schneider, Erica Lasek-Nesselquist |
| EPI_ISL_1226759 | Columbia University Irving Medical Center | Wadsworth Center, New York State Department of Health | Kirsten St. George, Daryl M. Lamson, Alexis Russel, Matthew Shudt, Melissa A Leisner, Jonathan Plitnick, Navjot Singh, John Kelly, Erasmus Schneider, Erica Lasek-Nesselquist |
| EPI_ISL_1226760, EPI_ISL_1226761, EPI_ISL_1226762, EPI_ISL_1226763, EPI_ISL_1226764 | MONTEFIORE MEDICAL CENTER LABORATORIES | Wadsworth Center, New York State Department of Health | Kirsten St. George, Daryl M. Lamson, Alexis Russel, Matthew Shudt, Melissa A Leisner, Jonathan Plitnick, Navjot Singh, John Kelly, Erasmus Schneider, Erica Lasek-Nesselquist |
| EPI_ISL_1226765 | Columbia University Irving Medical Center | Wadsworth Center, New York State Department of Health | Kirsten St. George, Daryl M. Lamson, Alexis Russel, Matthew Shudt, Melissa A Leisner, Jonathan Plitnick, Navjot Singh, John Kelly, Erasmus Schneider, Erica Lasek-Nesselquist |
| EPI_ISL_1226766, EPI_ISL_1226767 | MONTEFIORE MEDICAL CENTER LABORATORIES | Wadsworth Center, New York State Department of Health | Kirsten St. George, Daryl M. Lamson, Alexis Russel, Matthew Shudt, Melissa A Leisner, Jonathan Plitnick, Navjot Singh, John Kelly, Erasmus Schneider, Erica Lasek-Nesselquist |
| EPI_ISL_1226768, EPI_ISL_1226769 | NYC Pandemic Response Lab | Wadsworth Center, New York State Department of Health | Kirsten St. George, Daryl M. Lamson, Alexis Russel, Matthew Shudt, Melissa A Leisner, Jonathan Plitnick, Navjot Singh, John Kelly, Erasmus Schneider, Erica Lasek-Nesselquist |
| EPI_ISL_1226770, EPI_ISL_1226771 | MONTEFIORE MEDICAL CENTER LABORATORIES | Wadsworth Center, New York State Department of Health | Kirsten St. George, Daryl M. Lamson, Alexis Russel, Matthew Shudt, Melissa A Leisner, Jonathan Plitnick, Navjot Singh, John Kelly, Erasmus Schneider, Erica Lasek-Nesselquist |
| EPI_ISL_1226772 | WESTCHESTER MEDICAL CENTER | Wadsworth Center, New York State Department of Health | Kirsten St. George, Daryl M. Lamson, Alexis Russel, Matthew Shudt, Melissa A Leisner, Jonathan Plitnick, Navjot Singh, John Kelly, Erasmus Schneider, Erica Lasek-Nesselquist |
| EPI_ISL_1226773, EPI_ISL_1226774, EPI_ISL_1226775, EPI_ISL_1226776, EPI_ISL_1226777, EPI_ISL_1226778, EPI_ISL_1226779, EPI_ISL_1226780 | MONTEFIORE MEDICAL CENTER LABORATORIES | Wadsworth Center, New York State Department of Health | Kirsten St. George, Daryl M. Lamson, Alexis Russel, Matthew Shudt, Melissa A Leisner, Jonathan Plitnick, Navjot Singh, John Kelly, Erasmus Schneider, Erica Lasek-Nesselquist |
| EPI_ISL_1226781, EPI_ISL_1226782, EPI_ISL_1226783, EPI_ISL_1226784, EPI_ISL_1226785, EPI_ISL_1226786, EPI_ISL_1226787, EPI_ISL_1226788 | WESTCHESTER MEDICAL CENTER | Wadsworth Center, New York State Department of Health | Kirsten St. George, Daryl M. Lamson, Alexis Russel, Matthew Shudt, Melissa A Leisner, Jonathan Plitnick, Navjot Singh, John Kelly, Erasmus Schneider, Erica Lasek-Nesselquist |
| EPI_ISL_1226789, EPI_ISL_1226790, EPI_ISL_1226791, EPI_ISL_1226792, EPI_ISL_1226793, EPI_ISL_1226794, EPI_ISL_1226795, EPI_ISL_1226796, EPI_ISL_1226797 | NYC Pandemic Response Lab | Wadsworth Center, New York State Department of Health | Kirsten St. George, Daryl M. Lamson, Alexis Russel, Matthew Shudt, Melissa A Leisner, Jonathan Plitnick, Navjot Singh, John Kelly, Erasmus Schneider, Erica Lasek-Nesselquist |
| EPI_ISL_1226798 | NORTH SHORE UNIVERSITY HOSPITAL | Wadsworth Center, New York State Department of Health | Kirsten St. George, Daryl M. Lamson, Alexis Russel, Matthew Shudt, Melissa A Leisner, Jonathan Plitnick, Navjot Singh, John Kelly, Erasmus Schneider, Erica Lasek-Nesselquist |
| EPI_ISL_1226799 | MEMORIAL SLOAN KETTERING CANCER CENTER | Wadsworth Center, New York State Department of Health | Kirsten St. George, Daryl M. Lamson, Alexis Russel, Matthew Shudt, Melissa A Leisner, Jonathan Plitnick, Navjot Singh, John Kelly, Erasmus Schneider, Erica Lasek-Nesselquist |
| EPI_ISL_1226800, EPI_ISL_1226801, EPI_ISL_1226802, EPI_ISL_1226803, EPI_ISL_1226804, EPI_ISL_1226805, EPI_ISL_1226806, EPI_ISL_1226807 | Columbia University Irving Medical Center | Wadsworth Center, New York State Department of Health | Kirsten St. George, Daryl M. Lamson, Alexis Russel, Matthew Shudt, Melissa A Leisner, Jonathan Plitnick, Navjot Singh, John Kelly, Erasmus Schneider, Erica Lasek-Nesselquist |
| EPI_ISL_1226808, EPI_ISL_1226809, EPI_ISL_1226810 | NYC Pandemic Response Lab | Wadsworth Center, New York State Department of Health | Kirsten St. George, Daryl M. Lamson, Alexis Russel, Matthew Shudt, Melissa A Leisner, Jonathan Plitnick, Navjot Singh, John Kelly, Erasmus Schneider, Erica Lasek-Nesselquist |
| EPI_ISL_1226811, EPI_ISL_1226812 | New York Presbyterian Hospital | Wadsworth Center, New York State Department of Health | Kirsten St. George, Daryl M. Lamson, Alexis Russel, Matthew Shudt, Melissa A Leisner, Jonathan Plitnick, Navjot Singh, John Kelly, Erasmus Schneider, Erica Lasek-Nesselquist |
| EPI_ISL_1226813, EPI_ISL_1226814 | NYC Pandemic Response Lab | Wadsworth Center, New York State Department of Health | Kirsten St. George, Daryl M. Lamson, Alexis Russel, Matthew Shudt, Melissa A Leisner, Jonathan Plitnick, Navjot Singh, John Kelly, Erasmus Schneider, Erica Lasek-Nesselquist |
| EPI_ISL_1226815 | NORTH SHORE UNIVERSITY HOSPITAL | Wadsworth Center, New York State Department of Health | Kirsten St. George, Daryl M. Lamson, Alexis Russel, Matthew Shudt, Melissa A Leisner, Jonathan Plitnick, Navjot Singh, John Kelly, Erasmus Schneider, Erica Lasek-Nesselquist |
| EPI_ISL_1226816, EPI_ISL_1226817 | NYC Pandemic Response Lab | Wadsworth Center, New York State Department of Health | Kirsten St. George, Daryl M. Lamson, Alexis Russel, Matthew Shudt, Melissa A Leisner, Jonathan Plitnick, Navjot Singh, John Kelly, Erasmus Schneider, Erica Lasek-Nesselquist |
| EPI_ISL_1226818 | WESTCHESTER MEDICAL CENTER | Wadsworth Center, New York State Department of Health | Kirsten St. George, Daryl M. Lamson, Alexis Russel, Matthew Shudt, Melissa A Leisner, Jonathan Plitnick, Navjot Singh, John Kelly, Erasmus Schneider, Erica Lasek-Nesselquist |
| EPI_ISL_1226819 | ALBANY MEDICAL CENTER | Wadsworth Center, New York State Department of Health | Kirsten St. George, Daryl M. Lamson, Alexis Russel, Matthew Shudt, Melissa A Leisner, Jonathan Plitnick, Navjot Singh, John Kelly, Erasmus Schneider, Erica Lasek-Nesselquist |
| EPI_ISL_1226820 | MEMORIAL SLOAN KETTERING CANCER CENTER | Wadsworth Center, New York State Department of Health | Kirsten St. George, Daryl M. Lamson, Alexis Russel, Matthew Shudt, Melissa A Leisner, Jonathan Plitnick, Navjot Singh, John Kelly, Erasmus Schneider, Erica Lasek-Nesselquist |
| EPI_ISL_1226821, EPI_ISL_1226822 | WESTCHESTER MEDICAL CENTER | Wadsworth Center, New York State Department of Health | Kirsten St. George, Daryl M. Lamson, Alexis Russel, Matthew Shudt, Melissa A Leisner, Jonathan Plitnick, Navjot Singh, John Kelly, Erasmus Schneider, Erica Lasek-Nesselquist |

| | | | |
|---|---|---|---|
| EPI_ISL_1226823, EPI_ISL_1226824 | Columbia University Irving Medical Center | Wadsworth Center, New York State Department of Health | Kirsten St. George, Daryl M. Lamson, Alexis Russel, Matthew Shudt, Melissa A Leisner, Jonathan Plitnick, Navjot Singh, John Kelly, Erasmus Schneider, Erica Lasek-Nesselquist |
| EPI_ISL_1226825, EPI_ISL_1226826, EPI_ISL_1226827, EPI_ISL_1226828, EPI_ISL_1226829, EPI_ISL_1226830 | WESTCHESTER MEDICAL CENTER | Wadsworth Center, New York State Department of Health | Kirsten St. George, Daryl M. Lamson, Alexis Russel, Matthew Shudt, Melissa A Leisner, Jonathan Plitnick, Navjot Singh, John Kelly, Erasmus Schneider, Erica Lasek-Nesselquist |
| EPI_ISL_1226831 | MONTEFIORE MEDICAL CENTER LABORATORIES | Wadsworth Center, New York State Department of Health | Kirsten St. George, Daryl M. Lamson, Alexis Russel, Matthew Shudt, Melissa A Leisner, Jonathan Plitnick, Navjot Singh, John Kelly, Erasmus Schneider, Erica Lasek-Nesselquist |
| EPI_ISL_1226832, EPI_ISL_1226833 | NYC Pandemic Response Lab | Wadsworth Center, New York State Department of Health | Kirsten St. George, Daryl M. Lamson, Alexis Russel, Matthew Shudt, Melissa A Leisner, Jonathan Plitnick, Navjot Singh, John Kelly, Erasmus Schneider, Erica Lasek-Nesselquist |
| EPI_ISL_1226834 | WESTCHESTER MEDICAL CENTER | Wadsworth Center, New York State Department of Health | Kirsten St. George, Daryl M. Lamson, Alexis Russel, Matthew Shudt, Melissa A Leisner, Jonathan Plitnick, Navjot Singh, John Kelly, Erasmus Schneider, Erica Lasek-Nesselquist |
| EPI_ISL_1226835 | MONTEFIORE MEDICAL CENTER LABORATORIES | Wadsworth Center, New York State Department of Health | Kirsten St. George, Daryl M. Lamson, Alexis Russel, Matthew Shudt, Melissa A Leisner, Jonathan Plitnick, Navjot Singh, John Kelly, Erasmus Schneider, Erica Lasek-Nesselquist |
| EPI_ISL_1226836, EPI_ISL_1226837, EPI_ISL_1226838 | WESTCHESTER MEDICAL CENTER | Wadsworth Center, New York State Department of Health | Kirsten St. George, Daryl M. Lamson, Alexis Russel, Matthew Shudt, Melissa A Leisner, Jonathan Plitnick, Navjot Singh, John Kelly, Erasmus Schneider, Erica Lasek-Nesselquist |
| EPI_ISL_1226839 | MONTEFIORE MEDICAL CENTER LABORATORIES | Wadsworth Center, New York State Department of Health | Kirsten St. George, Daryl M. Lamson, Alexis Russel, Matthew Shudt, Melissa A Leisner, Jonathan Plitnick, Navjot Singh, John Kelly, Erasmus Schneider, Erica Lasek-Nesselquist |
| EPI_ISL_1226840, EPI_ISL_1226841 | WESTCHESTER MEDICAL CENTER | Wadsworth Center, New York State Department of Health | Kirsten St. George, Daryl M. Lamson, Alexis Russel, Matthew Shudt, Melissa A Leisner, Jonathan Plitnick, Navjot Singh, John Kelly, Erasmus Schneider, Erica Lasek-Nesselquist |
| EPI_ISL_1226842 | MONTEFIORE MEDICAL CENTER LABORATORIES | Wadsworth Center, New York State Department of Health | Kirsten St. George, Daryl M. Lamson, Alexis Russel, Matthew Shudt, Melissa A Leisner, Jonathan Plitnick, Navjot Singh, John Kelly, Erasmus Schneider, Erica Lasek-Nesselquist |
| EPI_ISL_1226843 | New York Presbyterian Hospital | Wadsworth Center, New York State Department of Health | Kirsten St. George, Daryl M. Lamson, Alexis Russel, Matthew Shudt, Melissa A Leisner, Jonathan Plitnick, Navjot Singh, John Kelly, Erasmus Schneider, Erica Lasek-Nesselquist |
| EPI_ISL_1226844 | MONTEFIORE MEDICAL CENTER LABORATORIES | Wadsworth Center, New York State Department of Health | Kirsten St. George, Daryl M. Lamson, Alexis Russel, Matthew Shudt, Melissa A Leisner, Jonathan Plitnick, Navjot Singh, John Kelly, Erasmus Schneider, Erica Lasek-Nesselquist |
| EPI_ISL_1226845 | WESTCHESTER MEDICAL CENTER | Wadsworth Center, New York State Department of Health | Kirsten St. George, Daryl M. Lamson, Alexis Russel, Matthew Shudt, Melissa A Leisner, Jonathan Plitnick, Navjot Singh, John Kelly, Erasmus Schneider, Erica Lasek-Nesselquist |
| EPI_ISL_1226846, EPI_ISL_1226847, EPI_ISL_1226848 | KALEIDA CENTER FOR LABORATORY MEDICINE | Wadsworth Center, New York State Department of Health | Kirsten St. George, Daryl M. Lamson, Alexis Russel, Matthew Shudt, Melissa A Leisner, Jonathan Plitnick, Navjot Singh, John Kelly, Erasmus Schneider, Erica Lasek-Nesselquist |
| EPI_ISL_1226849 | NORTHWELL HEALTH LABORATORIES | Wadsworth Center, New York State Department of Health | Kirsten St. George, Daryl M. Lamson, Alexis Russel, Matthew Shudt, Melissa A Leisner, Jonathan Plitnick, Navjot Singh, John Kelly, Erasmus Schneider, Erica Lasek-Nesselquist |
| EPI_ISL_1226850 | New York Presbyterian Hospital | Wadsworth Center, New York State Department of Health | Kirsten St. George, Daryl M. Lamson, Alexis Russel, Matthew Shudt, Melissa A Leisner, Jonathan Plitnick, Navjot Singh, John Kelly, Erasmus Schneider, Erica Lasek-Nesselquist |
| EPI_ISL_1226851, EPI_ISL_1226852 | URMC LABS | Wadsworth Center, New York State Department of Health | Kirsten St. George, Daryl M. Lamson, Alexis Russel, Matthew Shudt, Melissa A Leisner, Jonathan Plitnick, Navjot Singh, John Kelly, Erasmus Schneider, Erica Lasek-Nesselquist |
| EPI_ISL_1226853 | NORTHWELL HEALTH LABORATORIES | Wadsworth Center, New York State Department of Health | Kirsten St. George, Daryl M. Lamson, Alexis Russel, Matthew Shudt, Melissa A Leisner, Jonathan Plitnick, Navjot Singh, John Kelly, Erasmus Schneider, Erica Lasek-Nesselquist |
| EPI_ISL_1226854, EPI_ISL_1226855 | NYC Pandemic Response Lab | Wadsworth Center, New York State Department of Health | Kirsten St. George, Daryl M. Lamson, Alexis Russel, Matthew Shudt, Melissa A Leisner, Jonathan Plitnick, Navjot Singh, John Kelly, Erasmus Schneider, Erica Lasek-Nesselquist |
| EPI_ISL_1226856 | NORTHWELL HEALTH LABORATORIES | Wadsworth Center, New York State Department of Health | Kirsten St. George, Daryl M. Lamson, Alexis Russel, Matthew Shudt, Melissa A Leisner, Jonathan Plitnick, Navjot Singh, John Kelly, Erasmus Schneider, Erica Lasek-Nesselquist |
| EPI_ISL_1226857, EPI_ISL_1226858 | MEMORIAL SLOAN KETTERING CANCER CENTER | Wadsworth Center, New York State Department of Health | Kirsten St. George, Daryl M. Lamson, Alexis Russel, Matthew Shudt, Melissa A Leisner, Jonathan Plitnick, Navjot Singh, John Kelly, Erasmus Schneider, Erica Lasek-Nesselquist |
| EPI_ISL_1226859 | NORTHWELL HEALTH LABORATORIES | Wadsworth Center, New York State Department of Health | Kirsten St. George, Daryl M. Lamson, Alexis Russel, Matthew Shudt, Melissa A Leisner, Jonathan Plitnick, Navjot Singh, John Kelly, Erasmus Schneider, Erica Lasek-Nesselquist |
| EPI_ISL_1226860 | NYC Pandemic Response Lab | Wadsworth Center, New York State Department of Health | Kirsten St. George, Daryl M. Lamson, Alexis Russel, Matthew Shudt, Melissa A Leisner, Jonathan Plitnick, Navjot Singh, John Kelly, Erasmus Schneider, Erica Lasek-Nesselquist |
| EPI_ISL_1226861 | MEMORIAL SLOAN KETTERING CANCER CENTER | Wadsworth Center, New York State Department of Health | Kirsten St. George, Daryl M. Lamson, Alexis Russel, Matthew Shudt, Melissa A Leisner, Jonathan Plitnick, Navjot Singh, John Kelly, Erasmus Schneider, Erica Lasek-Nesselquist |
| EPI_ISL_1226862 | NYC Pandemic Response Lab | Wadsworth Center, New York State Department of Health | Kirsten St. George, Daryl M. Lamson, Alexis Russel, Matthew Shudt, Melissa A Leisner, Jonathan Plitnick, Navjot Singh, John Kelly, Erasmus Schneider, Erica Lasek-Nesselquist |
| EPI_ISL_1226863 | WESTCHESTER MEDICAL CENTER | Wadsworth Center, New York State Department of Health | Kirsten St. George, Daryl M. Lamson, Alexis Russel, Matthew Shudt, Melissa A Leisner, Jonathan Plitnick, Navjot Singh, John Kelly, Erasmus Schneider, Erica Lasek-Nesselquist |
| EPI_ISL_1226864 | MONTEFIORE MEDICAL CENTER LABORATORIES | Wadsworth Center, New York State Department of Health | Kirsten St. George, Daryl M. Lamson, Alexis Russel, Matthew Shudt, Melissa A Leisner, Jonathan Plitnick, Navjot Singh, John Kelly, Erasmus Schneider, Erica Lasek-Nesselquist |
| EPI_ISL_1226865 | NORTHWELL HEALTH LABORATORIES | Wadsworth Center, New York State Department of Health | Kirsten St. George, Daryl M. Lamson, Alexis Russel, Matthew Shudt, Melissa A Leisner, Jonathan Plitnick, Navjot Singh, John Kelly, Erasmus Schneider, Erica Lasek-Nesselquist |
| EPI_ISL_1226866 | URMC LABS | Wadsworth Center, New York State Department of Health | Kirsten St. George, Daryl M. Lamson, Alexis Russel, Matthew Shudt, Melissa A Leisner, Jonathan Plitnick, Navjot Singh, John Kelly, Erasmus Schneider, Erica Lasek-Nesselquist |
| EPI_ISL_1226867 | NYC Pandemic Response Lab | Wadsworth Center, New York State Department of Health | Kirsten St. George, Daryl M. Lamson, Alexis Russel, Matthew Shudt, Melissa A Leisner, Jonathan Plitnick, Navjot Singh, John Kelly, Erasmus Schneider, Erica Lasek-Nesselquist |
| EPI_ISL_1226868 | THE MARY IMOGENE BASSETT HOSPITAL | Wadsworth Center, New York State Department of Health | Kirsten St. George, Daryl M. Lamson, Alexis Russel, Matthew Shudt, Melissa A Leisner, Jonathan Plitnick, Navjot Singh, John Kelly, Erasmus Schneider, Erica Lasek-Nesselquist |
| EPI_ISL_1226869 | NYC Pandemic Response Lab | Wadsworth Center, New York State Department of Health | Kirsten St. George, Daryl M. Lamson, Alexis Russel, Matthew Shudt, Melissa A Leisner, Jonathan Plitnick, Navjot Singh, John Kelly, Erasmus Schneider, Erica Lasek-Nesselquist |
| EPI_ISL_1226870 | WESTCHESTER MEDICAL CENTER | Wadsworth Center, New York State Department of Health | Kirsten St. George, Daryl M. Lamson, Alexis Russel, Matthew Shudt, Melissa A Leisner, Jonathan Plitnick, Navjot Singh, John Kelly, Erasmus Schneider, Erica Lasek-Nesselquist |
| EPI_ISL_1226871 | MONTEFIORE MEDICAL CENTER LABORATORIES | Wadsworth Center, New York State Department of Health | Kirsten St. George, Daryl M. Lamson, Alexis Russel, Matthew Shudt, Melissa A Leisner, Jonathan Plitnick, Navjot Singh, John Kelly, Erasmus Schneider, Erica Lasek-Nesselquist |
| EPI_ISL_1226872, EPI_ISL_1226873 | SUNY UPSTATE MEDICAL UNIVERSITY | Wadsworth Center, New York State Department of Health | Kirsten St. George, Daryl M. Lamson, Alexis Russel, Matthew Shudt, Melissa A Leisner, Jonathan Plitnick, Navjot Singh, John Kelly, Erasmus Schneider, Erica Lasek-Nesselquist |

| | | | |
|---|---|---|---|
| EPI_ISL_1226874, EPI_ISL_1226875 | Columbia University Irving Medical Center | Wadsworth Center, New York State Department of Health | Kirsten St. George, Daryl M. Lamson, Alexis Russel, Matthew Shudt, Melissa A Leisner, Jonathan Plitnick, Navjot Singh, John Kelly, Erasmus Schneider, Erica Lasek-Nesselquist |
| EPI_ISL_1226876, EPI_ISL_1226877, EPI_ISL_1226878, EPI_ISL_1226879, EPI_ISL_1226880, EPI_ISL_1226881, EPI_ISL_1226882, EPI_ISL_1226883, EPI_ISL_1226884, EPI_ISL_1226885, EPI_ISL_1226886, EPI_ISL_1226887, EPI_ISL_1226888, EPI_ISL_1226889, EPI_ISL_1226890 | | | |
| see above | MONTEFIORE MEDICAL CENTER LABORATORIES | Wadsworth Center, New York State Department of Health | Kirsten St. George, Daryl M. Lamson, Alexis Russel, Matthew Shudt, Melissa A Leisner, Jonathan Plitnick, Navjot Singh, John Kelly, Erasmus Schneider, Erica Lasek-Nesselquist |
| EPI_ISL_1226891, EPI_ISL_1226892 | WESTCHESTER MEDICAL CENTER | Wadsworth Center, New York State Department of Health | Kirsten St. George, Daryl M. Lamson, Alexis Russel, Matthew Shudt, Melissa A Leisner, Jonathan Plitnick, Navjot Singh, John Kelly, Erasmus Schneider, Erica Lasek-Nesselquist |
| EPI_ISL_1226893, EPI_ISL_1226894, EPI_ISL_1226895, EPI_ISL_1226896 | NYC Pandemic Response Lab | Wadsworth Center, New York State Department of Health | Kirsten St. George, Daryl M. Lamson, Alexis Russel, Matthew Shudt, Melissa A Leisner, Jonathan Plitnick, Navjot Singh, John Kelly, Erasmus Schneider, Erica Lasek-Nesselquist |
| EPI_ISL_1226897 | URMC LABS | Wadsworth Center, New York State Department of Health | Kirsten St. George, Daryl M. Lamson, Alexis Russel, Matthew Shudt, Melissa A Leisner, Jonathan Plitnick, Navjot Singh, John Kelly, Erasmus Schneider, Erica Lasek-Nesselquist |
| EPI_ISL_1226898, EPI_ISL_1226899 | NORTHWELL HEALTH LABORATORIES | Wadsworth Center, New York State Department of Health | Kirsten St. George, Daryl M. Lamson, Alexis Russel, Matthew Shudt, Melissa A Leisner, Jonathan Plitnick, Navjot Singh, John Kelly, Erasmus Schneider, Erica Lasek-Nesselquist |
| EPI_ISL_1226900 | NYC Pandemic Response Lab | Wadsworth Center, New York State Department of Health | Kirsten St. George, Daryl M. Lamson, Alexis Russel, Matthew Shudt, Melissa A Leisner, Jonathan Plitnick, Navjot Singh, John Kelly, Erasmus Schneider, Erica Lasek-Nesselquist |
| EPI_ISL_1226901, EPI_ISL_1226902, EPI_ISL_1226903, EPI_ISL_1226904, EPI_ISL_1226905, EPI_ISL_1226906 | Columbia University Irving Medical Center | Wadsworth Center, New York State Department of Health | Kirsten St. George, Daryl M. Lamson, Alexis Russel, Matthew Shudt, Melissa A Leisner, Jonathan Plitnick, Navjot Singh, John Kelly, Erasmus Schneider, Erica Lasek-Nesselquist |
| EPI_ISL_1226907 | MEMORIAL SLOAN KETTERING CANCER CENTER | Wadsworth Center, New York State Department of Health | Kirsten St. George, Daryl M. Lamson, Alexis Russel, Matthew Shudt, Melissa A Leisner, Jonathan Plitnick, Navjot Singh, John Kelly, Erasmus Schneider, Erica Lasek-Nesselquist |
| EPI_ISL_1226908 | WESTCHESTER MEDICAL CENTER | Wadsworth Center, New York State Department of Health | Kirsten St. George, Daryl M. Lamson, Alexis Russel, Matthew Shudt, Melissa A Leisner, Jonathan Plitnick, Navjot Singh, John Kelly, Erasmus Schneider, Erica Lasek-Nesselquist |
| EPI_ISL_1226909 | MONTEFIORE MEDICAL CENTER LABORATORIES | Wadsworth Center, New York State Department of Health | Kirsten St. George, Daryl M. Lamson, Alexis Russel, Matthew Shudt, Melissa A Leisner, Jonathan Plitnick, Navjot Singh, John Kelly, Erasmus Schneider, Erica Lasek-Nesselquist |
| EPI_ISL_1226910 | MEMORIAL SLOAN KETTERING CANCER CENTER | Wadsworth Center, New York State Department of Health | Kirsten St. George, Daryl M. Lamson, Alexis Russel, Matthew Shudt, Melissa A Leisner, Jonathan Plitnick, Navjot Singh, John Kelly, Erasmus Schneider, Erica Lasek-Nesselquist |
| EPI_ISL_1226911, EPI_ISL_1226912 | NORTHWELL HEALTH LABORATORIES | Wadsworth Center, New York State Department of Health | Kirsten St. George, Daryl M. Lamson, Alexis Russel, Matthew Shudt, Melissa A Leisner, Jonathan Plitnick, Navjot Singh, John Kelly, Erasmus Schneider, Erica Lasek-Nesselquist |
| EPI_ISL_1226913, EPI_ISL_1226914 | NYC Pandemic Response Lab | Wadsworth Center, New York State Department of Health | Kirsten St. George, Daryl M. Lamson, Alexis Russel, Matthew Shudt, Melissa A Leisner, Jonathan Plitnick, Navjot Singh, John Kelly, Erasmus Schneider, Erica Lasek-Nesselquist |
| EPI_ISL_1226915 | New York Presbyterian Hospital | Wadsworth Center, New York State Department of Health | Kirsten St. George, Daryl M. Lamson, Alexis Russel, Matthew Shudt, Melissa A Leisner, Jonathan Plitnick, Navjot Singh, John Kelly, Erasmus Schneider, Erica Lasek-Nesselquist |
| EPI_ISL_1226916, EPI_ISL_1226917, EPI_ISL_1226918, EPI_ISL_1226919, EPI_ISL_1226920, EPI_ISL_1226921, EPI_ISL_1226922, EPI_ISL_1226923 | NYC Pandemic Response Lab | Wadsworth Center, New York State Department of Health | Kirsten St. George, Daryl M. Lamson, Alexis Russel, Matthew Shudt, Melissa A Leisner, Jonathan Plitnick, Navjot Singh, John Kelly, Erasmus Schneider, Erica Lasek-Nesselquist |
| EPI_ISL_1226924, EPI_ISL_1226925 | WESTCHESTER MEDICAL CENTER | Wadsworth Center, New York State Department of Health | Kirsten St. George, Daryl M. Lamson, Alexis Russel, Matthew Shudt, Melissa A Leisner, Jonathan Plitnick, Navjot Singh, John Kelly, Erasmus Schneider, Erica Lasek-Nesselquist |
| EPI_ISL_1226926 | Columbia University Irving Medical Center | Wadsworth Center, New York State Department of Health | Kirsten St. George, Daryl M. Lamson, Alexis Russel, Matthew Shudt, Melissa A Leisner, Jonathan Plitnick, Navjot Singh, John Kelly, Erasmus Schneider, Erica Lasek-Nesselquist |
| EPI_ISL_1226927, EPI_ISL_1226928, EPI_ISL_1226929 | WESTCHESTER MEDICAL CENTER | Wadsworth Center, New York State Department of Health | Kirsten St. George, Daryl M. Lamson, Alexis Russel, Matthew Shudt, Melissa A Leisner, Jonathan Plitnick, Navjot Singh, John Kelly, Erasmus Schneider, Erica Lasek-Nesselquist |
| EPI_ISL_1226930 | Columbia University Irving Medical Center | Wadsworth Center, New York State Department of Health | Kirsten St. George, Daryl M. Lamson, Alexis Russel, Matthew Shudt, Melissa A Leisner, Jonathan Plitnick, Navjot Singh, John Kelly, Erasmus Schneider, Erica Lasek-Nesselquist |
| EPI_ISL_1226931 | MONTEFIORE MEDICAL CENTER LABORATORIES | Wadsworth Center, New York State Department of Health | Kirsten St. George, Daryl M. Lamson, Alexis Russel, Matthew Shudt, Melissa A Leisner, Jonathan Plitnick, Navjot Singh, John Kelly, Erasmus Schneider, Erica Lasek-Nesselquist |
| EPI_ISL_1226932 | BIO-REFERENCE LABORATORIES | Wadsworth Center, New York State Department of Health | Kirsten St. George, Daryl M. Lamson, Alexis Russel, Matthew Shudt, Melissa A Leisner, Jonathan Plitnick, Navjot Singh, John Kelly, Erasmus Schneider, Erica Lasek-Nesselquist |
| EPI_ISL_1226933 | Wadsworth Center, New York State Department of Health | Wadsworth Center, New York State Department of Health | Kirsten St. George, Daryl M. Lamson, Alexis Russel, Matthew Shudt, Melissa A Leisner, Jonathan Plitnick, Navjot Singh, John Kelly, Erasmus Schneider, Erica Lasek-Nesselquist |
| EPI_ISL_1226934 | MONTEFIORE MEDICAL CENTER LABORATORIES | Wadsworth Center, New York State Department of Health | Kirsten St. George, Daryl M. Lamson, Alexis Russel, Matthew Shudt, Melissa A Leisner, Jonathan Plitnick, Navjot Singh, John Kelly, Erasmus Schneider, Erica Lasek-Nesselquist |
| EPI_ISL_1226935 | NYC Pandemic Response Lab | Wadsworth Center, New York State Department of Health | Kirsten St. George, Daryl M. Lamson, Alexis Russel, Matthew Shudt, Melissa A Leisner, Jonathan Plitnick, Navjot Singh, John Kelly, Erasmus Schneider, Erica Lasek-Nesselquist |
| EPI_ISL_1226936, EPI_ISL_1226937 | MONTEFIORE MEDICAL CENTER LABORATORIES | Wadsworth Center, New York State Department of Health | Kirsten St. George, Daryl M. Lamson, Alexis Russel, Matthew Shudt, Melissa A Leisner, Jonathan Plitnick, Navjot Singh, John Kelly, Erasmus Schneider, Erica Lasek-Nesselquist |
| EPI_ISL_1226938 | Wadsworth Center, New York State Department of Health | Wadsworth Center, New York State Department of Health | Kirsten St. George, Daryl M. Lamson, Alexis Russel, Matthew Shudt, Melissa A Leisner, Jonathan Plitnick, Navjot Singh, John Kelly, Erasmus Schneider, Erica Lasek-Nesselquist |
| EPI_ISL_1226939, EPI_ISL_1226940, EPI_ISL_1226941, EPI_ISL_1226942, EPI_ISL_1226943, EPI_ISL_1226944, EPI_ISL_1226945, EPI_ISL_1226946, EPI_ISL_1226947, EPI_ISL_1226948, EPI_ISL_1226949, EPI_ISL_1226950, EPI_ISL_1226951, EPI_ISL_1226952, EPI_ISL_1226953, EPI_ISL_1226954, EPI_ISL_1226955, EPI_ISL_1226956, EPI_ISL_1226957, EPI_ISL_1226958, EPI_ISL_1226959, EPI_ISL_1226960, EPI_ISL_1226961, EPI_ISL_1226962 | | | |
| see above | MONTEFIORE MEDICAL CENTER LABORATORIES | Wadsworth Center, New York State Department of Health | Kirsten St. George, Daryl M. Lamson, Alexis Russel, Matthew Shudt, Melissa A Leisner, Jonathan Plitnick, Navjot Singh, John Kelly, Erasmus Schneider, Erica Lasek-Nesselquist |
| EPI_ISL_1226963 | NYC Pandemic Response Lab | Wadsworth Center, New York State Department of Health | Kirsten St. George, Daryl M. Lamson, Alexis Russel, Matthew Shudt, Melissa A Leisner, Jonathan Plitnick, Navjot Singh, John Kelly, Erasmus Schneider, Erica Lasek-Nesselquist |
| EPI_ISL_1226964, EPI_ISL_1226965, EPI_ISL_1226966, EPI_ISL_1226967, EPI_ISL_1226968, EPI_ISL_1226969 | MONTEFIORE MEDICAL CENTER LABORATORIES | Wadsworth Center, New York State Department of Health | Kirsten St. George, Daryl M. Lamson, Alexis Russel, Matthew Shudt, Melissa A Leisner, Jonathan Plitnick, Navjot Singh, John Kelly, Erasmus Schneider, Erica Lasek-Nesselquist |
| EPI_ISL_1226970 | WESTCHESTER MEDICAL CENTER | Wadsworth Center, New York State Department of Health | Kirsten St. George, Daryl M. Lamson, Alexis Russel, Matthew Shudt, Melissa A Leisner, Jonathan Plitnick, Navjot Singh, John Kelly, Erasmus Schneider, Erica Lasek-Nesselquist |
| EPI_ISL_1226971, EPI_ISL_1226972 | MONTEFIORE MEDICAL CENTER LABORATORIES | Wadsworth Center, New York State Department of Health | Kirsten St. George, Daryl M. Lamson, Alexis Russel, Matthew Shudt, Melissa A Leisner, Jonathan Plitnick, Navjot Singh, John Kelly, Erasmus Schneider, Erica Lasek-Nesselquist |

| | | | |
|---|---|---|---|
| EPI_ISL_1226973 | WESTCHESTER MEDICAL CENTER | Wadsworth Center, New York State Department of Health | Kirsten St. George, Daryl M. Lamson, Alexis Russel, Matthew Shudt, Melissa A Leisner, Jonathan Plitnick, Navjot Singh, John Kelly, Erasmus Schneider, Erica Lasek-Nesselquist |
| EPI_ISL_1226974, EPI_ISL_1226975, EPI_ISL_1226976, EPI_ISL_1226977, EPI_ISL_1226978, EPI_ISL_1226979 | MONTEFIORE MEDICAL CENTER LABORATORIES | Wadsworth Center, New York State Department of Health | Kirsten St. George, Daryl M. Lamson, Alexis Russel, Matthew Shudt, Melissa A Leisner, Jonathan Plitnick, Navjot Singh, John Kelly, Erasmus Schneider, Erica Lasek-Nesselquist |
| EPI_ISL_1226980 | MEMORIAL SLOAN KETTERING CANCER CENTER | Wadsworth Center, New York State Department of Health | Kirsten St. George, Daryl M. Lamson, Alexis Russel, Matthew Shudt, Melissa A Leisner, Jonathan Plitnick, Navjot Singh, John Kelly, Erasmus Schneider, Erica Lasek-Nesselquist |
| EPI_ISL_1226981 | SUNY UPSTATE MEDICAL UNIVERSITY | Wadsworth Center, New York State Department of Health | Kirsten St. George, Daryl M. Lamson, Alexis Russel, Matthew Shudt, Melissa A Leisner, Jonathan Plitnick, Navjot Singh, John Kelly, Erasmus Schneider, Erica Lasek-Nesselquist |
| EPI_ISL_1226982, EPI_ISL_1226983, EPI_ISL_1226984 | NYC Pandemic Response Lab | Wadsworth Center, New York State Department of Health | Kirsten St. George, Daryl M. Lamson, Alexis Russel, Matthew Shudt, Melissa A Leisner, Jonathan Plitnick, Navjot Singh, John Kelly, Erasmus Schneider, Erica Lasek-Nesselquist |
| EPI_ISL_1226985 | BIO-REFERENCE LABORATORIES | Wadsworth Center, New York State Department of Health | Kirsten St. George, Daryl M. Lamson, Alexis Russel, Matthew Shudt, Melissa A Leisner, Jonathan Plitnick, Navjot Singh, John Kelly, Erasmus Schneider, Erica Lasek-Nesselquist |
| EPI_ISL_1226986 | MEMORIAL SLOAN KETTERING CANCER CENTER | Wadsworth Center, New York State Department of Health | Kirsten St. George, Daryl M. Lamson, Alexis Russel, Matthew Shudt, Melissa A Leisner, Jonathan Plitnick, Navjot Singh, John Kelly, Erasmus Schneider, Erica Lasek-Nesselquist |
| EPI_ISL_1226987 | NYC Pandemic Response Lab | Wadsworth Center, New York State Department of Health | Kirsten St. George, Daryl M. Lamson, Alexis Russel, Matthew Shudt, Melissa A Leisner, Jonathan Plitnick, Navjot Singh, John Kelly, Erasmus Schneider, Erica Lasek-Nesselquist |
| EPI_ISL_1226988, EPI_ISL_1226989, EPI_ISL_1226990, EPI_ISL_1226991, EPI_ISL_1226992, EPI_ISL_1226993, EPI_ISL_1226994, EPI_ISL_1226995, EPI_ISL_1226996, EPI_ISL_1226997 | Columbia University Irving Medical Center | Wadsworth Center, New York State Department of Health | Kirsten St. George, Daryl M. Lamson, Alexis Russel, Matthew Shudt, Melissa A Leisner, Jonathan Plitnick, Navjot Singh, John Kelly, Erasmus Schneider, Erica Lasek-Nesselquist |
| EPI_ISL_1226998, EPI_ISL_1226999 | NYC Pandemic Response Lab | Wadsworth Center, New York State Department of Health | Kirsten St. George, Daryl M. Lamson, Alexis Russel, Matthew Shudt, Melissa A Leisner, Jonathan Plitnick, Navjot Singh, John Kelly, Erasmus Schneider, Erica Lasek-Nesselquist |
| EPI_ISL_1227000 | New York Presbyterian Hospital | Wadsworth Center, New York State Department of Health | Kirsten St. George, Daryl M. Lamson, Alexis Russel, Matthew Shudt, Melissa A Leisner, Jonathan Plitnick, Navjot Singh, John Kelly, Erasmus Schneider, Erica Lasek-Nesselquist |
| EPI_ISL_1227001 | NYC Pandemic Response Lab | Wadsworth Center, New York State Department of Health | Kirsten St. George, Daryl M. Lamson, Alexis Russel, Matthew Shudt, Melissa A Leisner, Jonathan Plitnick, Navjot Singh, John Kelly, Erasmus Schneider, Erica Lasek-Nesselquist |
| EPI_ISL_1227002, EPI_ISL_1227003, EPI_ISL_1227004 | Columbia University Irving Medical Center | Wadsworth Center, New York State Department of Health | Kirsten St. George, Daryl M. Lamson, Alexis Russel, Matthew Shudt, Melissa A Leisner, Jonathan Plitnick, Navjot Singh, John Kelly, Erasmus Schneider, Erica Lasek-Nesselquist |
| EPI_ISL_1227005, EPI_ISL_1227006 | NYC Pandemic Response Lab | Wadsworth Center, New York State Department of Health | Kirsten St. George, Daryl M. Lamson, Alexis Russel, Matthew Shudt, Melissa A Leisner, Jonathan Plitnick, Navjot Singh, John Kelly, Erasmus Schneider, Erica Lasek-Nesselquist |
| EPI_ISL_1227007 | WESTCHESTER MEDICAL CENTER | Wadsworth Center, New York State Department of Health | Kirsten St. George, Daryl M. Lamson, Alexis Russel, Matthew Shudt, Melissa A Leisner, Jonathan Plitnick, Navjot Singh, John Kelly, Erasmus Schneider, Erica Lasek-Nesselquist |
| EPI_ISL_1227008 | NORTH SHORE UNIVERSITY HOSPITAL | Wadsworth Center, New York State Department of Health | Kirsten St. George, Daryl M. Lamson, Alexis Russel, Matthew Shudt, Melissa A Leisner, Jonathan Plitnick, Navjot Singh, John Kelly, Erasmus Schneider, Erica Lasek-Nesselquist |
| EPI_ISL_1227009, EPI_ISL_1227010, EPI_ISL_1227011, EPI_ISL_1227012, EPI_ISL_1227013 | NYC Pandemic Response Lab | Wadsworth Center, New York State Department of Health | Kirsten St. George, Daryl M. Lamson, Alexis Russel, Matthew Shudt, Melissa A Leisner, Jonathan Plitnick, Navjot Singh, John Kelly, Erasmus Schneider, Erica Lasek-Nesselquist |
| EPI_ISL_1227014 | NORTH SHORE UNIVERSITY HOSPITAL | Wadsworth Center, New York State Department of Health | Kirsten St. George, Daryl M. Lamson, Alexis Russel, Matthew Shudt, Melissa A Leisner, Jonathan Plitnick, Navjot Singh, John Kelly, Erasmus Schneider, Erica Lasek-Nesselquist |
| EPI_ISL_1227015 | NYC Pandemic Response Lab | Wadsworth Center, New York State Department of Health | Kirsten St. George, Daryl M. Lamson, Alexis Russel, Matthew Shudt, Melissa A Leisner, Jonathan Plitnick, Navjot Singh, John Kelly, Erasmus Schneider, Erica Lasek-Nesselquist |
| EPI_ISL_1227016, EPI_ISL_1227017, EPI_ISL_1227018 | MONTEFIORE MEDICAL CENTER LABORATORIES | Wadsworth Center, New York State Department of Health | Kirsten St. George, Daryl M. Lamson, Alexis Russel, Matthew Shudt, Melissa A Leisner, Jonathan Plitnick, Navjot Singh, John Kelly, Erasmus Schneider, Erica Lasek-Nesselquist |
| EPI_ISL_1227019, EPI_ISL_1227020 | WESTCHESTER MEDICAL CENTER | Wadsworth Center, New York State Department of Health | Kirsten St. George, Daryl M. Lamson, Alexis Russel, Matthew Shudt, Melissa A Leisner, Jonathan Plitnick, Navjot Singh, John Kelly, Erasmus Schneider, Erica Lasek-Nesselquist |
| EPI_ISL_1227021, EPI_ISL_1227022 | MONTEFIORE MEDICAL CENTER LABORATORIES | Wadsworth Center, New York State Department of Health | Kirsten St. George, Daryl M. Lamson, Alexis Russel, Matthew Shudt, Melissa A Leisner, Jonathan Plitnick, Navjot Singh, John Kelly, Erasmus Schneider, Erica Lasek-Nesselquist |
| EPI_ISL_1227023 | WESTCHESTER MEDICAL CENTER | Wadsworth Center, New York State Department of Health | Kirsten St. George, Daryl M. Lamson, Alexis Russel, Matthew Shudt, Melissa A Leisner, Jonathan Plitnick, Navjot Singh, John Kelly, Erasmus Schneider, Erica Lasek-Nesselquist |
| EPI_ISL_1227024, EPI_ISL_1227025 | MONTEFIORE MEDICAL CENTER LABORATORIES | Wadsworth Center, New York State Department of Health | Kirsten St. George, Daryl M. Lamson, Alexis Russel, Matthew Shudt, Melissa A Leisner, Jonathan Plitnick, Navjot Singh, John Kelly, Erasmus Schneider, Erica Lasek-Nesselquist |
| EPI_ISL_1227026 | NORTH SHORE UNIVERSITY HOSPITAL | Wadsworth Center, New York State Department of Health | Kirsten St. George, Daryl M. Lamson, Alexis Russel, Matthew Shudt, Melissa A Leisner, Jonathan Plitnick, Navjot Singh, John Kelly, Erasmus Schneider, Erica Lasek-Nesselquist |
| EPI_ISL_1227027, EPI_ISL_1227028 | NYC Pandemic Response Lab | Wadsworth Center, New York State Department of Health | Kirsten St. George, Daryl M. Lamson, Alexis Russel, Matthew Shudt, Melissa A Leisner, Jonathan Plitnick, Navjot Singh, John Kelly, Erasmus Schneider, Erica Lasek-Nesselquist |
| EPI_ISL_1227029 | ALBANY MEDICAL CENTER | Wadsworth Center, New York State Department of Health | Kirsten St. George, Daryl M. Lamson, Alexis Russel, Matthew Shudt, Melissa A Leisner, Jonathan Plitnick, Navjot Singh, John Kelly, Erasmus Schneider, Erica Lasek-Nesselquist |
| EPI_ISL_1227030 | Wadsworth Center, New York State Department of Health | Wadsworth Center, New York State Department of Health | Kirsten St. George, Daryl M. Lamson, Alexis Russel, Matthew Shudt, Melissa A Leisner, Jonathan Plitnick, Navjot Singh, John Kelly, Erasmus Schneider, Erica Lasek-Nesselquist |
| EPI_ISL_1227031, EPI_ISL_1227032 | ALBANY MEDICAL CENTER | Wadsworth Center, New York State Department of Health | Kirsten St. George, Daryl M. Lamson, Alexis Russel, Matthew Shudt, Melissa A Leisner, Jonathan Plitnick, Navjot Singh, John Kelly, Erasmus Schneider, Erica Lasek-Nesselquist |
| EPI_ISL_1227033, EPI_ISL_1227034 | MEMORIAL SLOAN KETTERING CANCER CENTER | Wadsworth Center, New York State Department of Health | Kirsten St. George, Daryl M. Lamson, Alexis Russel, Matthew Shudt, Melissa A Leisner, Jonathan Plitnick, Navjot Singh, John Kelly, Erasmus Schneider, Erica Lasek-Nesselquist |
| EPI_ISL_1227035, EPI_ISL_1227036, EPI_ISL_1227037, EPI_ISL_1227038, EPI_ISL_1227039 | MONTEFIORE MEDICAL CENTER LABORATORIES | Wadsworth Center, New York State Department of Health | Kirsten St. George, Daryl M. Lamson, Alexis Russel, Matthew Shudt, Melissa A Leisner, Jonathan Plitnick, Navjot Singh, John Kelly, Erasmus Schneider, Erica Lasek-Nesselquist |
| EPI_ISL_1227040 | WESTCHESTER MEDICAL CENTER | Wadsworth Center, New York State Department of Health | Kirsten St. George, Daryl M. Lamson, Alexis Russel, Matthew Shudt, Melissa A Leisner, Jonathan Plitnick, Navjot Singh, John Kelly, Erasmus Schneider, Erica Lasek-Nesselquist |

EPI_ISL_1227041, EPI_ISL_1227042, EPI_ISL_1227043, EPI_ISL_1227044, EPI_ISL_1227045, EPI_ISL_1227046, EPI_ISL_1227047, EPI_ISL_1227048, EPI_ISL_1227049, EPI_ISL_1227050, EPI_ISL_1227051, EPI_ISL_1227052, EPI_ISL_1227053, EPI_ISL_1227054, EPI_ISL_1227055, EPI_ISL_1227056, EPI_ISL_1227057,

| | | | |
|---|---|---|---|
| EPI_ISL_1227058, EPI_ISL_1227059, EPI_ISL_1227060, EPI_ISL_1227061, EPI_ISL_1227062, EPI_ISL_1227063, EPI_ISL_1227064, EPI_ISL_1227065, EPI_ISL_1227066, EPI_ISL_1227067 | | | |
| see above | MONTEFIORE MEDICAL CENTER LABORATORIES | Wadsworth Center, New York State Department of Health | Kirsten St. George, Daryl M. Lamson, Alexis Russel, Matthew Shudt, Melissa A Leisner, Jonathan Plitnick, Navjot Singh, John Kelly, Erasmus Schneider, Erica Lasek-Nesselquist |
| EPI_ISL_1227068, EPI_ISL_1227069 | Columbia University Irving Medical Center | Wadsworth Center, New York State Department of Health | Kirsten St. George, Daryl M. Lamson, Alexis Russel, Matthew Shudt, Melissa A Leisner, Jonathan Plitnick, Navjot Singh, John Kelly, Erasmus Schneider, Erica Lasek-Nesselquist |
| EPI_ISL_1227070, EPI_ISL_1227071, EPI_ISL_1227072, EPI_ISL_1227073, EPI_ISL_1227074, EPI_ISL_1227075, EPI_ISL_1227076, EPI_ISL_1227077, EPI_ISL_1227078, EPI_ISL_1227079, EPI_ISL_1227080 | | | |
| see above | MONTEFIORE MEDICAL CENTER LABORATORIES | Wadsworth Center, New York State Department of Health | Kirsten St. George, Daryl M. Lamson, Alexis Russel, Matthew Shudt, Melissa A Leisner, Jonathan Plitnick, Navjot Singh, John Kelly, Erasmus Schneider, Erica Lasek-Nesselquist |
| EPI_ISL_1227081 | NYC Pandemic Response Lab | Wadsworth Center, New York State Department of Health | Kirsten St. George, Daryl M. Lamson, Alexis Russel, Matthew Shudt, Melissa A Leisner, Jonathan Plitnick, Navjot Singh, John Kelly, Erasmus Schneider, Erica Lasek-Nesselquist |
| EPI_ISL_1227082, EPI_ISL_1227083, EPI_ISL_1227084, EPI_ISL_1227085, EPI_ISL_1227086 | MONTEFIORE MEDICAL CENTER LABORATORIES | Wadsworth Center, New York State Department of Health | Kirsten St. George, Daryl M. Lamson, Alexis Russel, Matthew Shudt, Melissa A Leisner, Jonathan Plitnick, Navjot Singh, John Kelly, Erasmus Schneider, Erica Lasek-Nesselquist |
| EPI_ISL_1227087 | NYC Pandemic Response Lab | Wadsworth Center, New York State Department of Health | Kirsten St. George, Daryl M. Lamson, Alexis Russel, Matthew Shudt, Melissa A Leisner, Jonathan Plitnick, Navjot Singh, John Kelly, Erasmus Schneider, Erica Lasek-Nesselquist |
| EPI_ISL_1227088, EPI_ISL_1227089 | Columbia University Irving Medical Center | Wadsworth Center, New York State Department of Health | Kirsten St. George, Daryl M. Lamson, Alexis Russel, Matthew Shudt, Melissa A Leisner, Jonathan Plitnick, Navjot Singh, John Kelly, Erasmus Schneider, Erica Lasek-Nesselquist |
| EPI_ISL_1227090, EPI_ISL_1227091, EPI_ISL_1227092, EPI_ISL_1227093, EPI_ISL_1227094 | MONTEFIORE MEDICAL CENTER LABORATORIES | Wadsworth Center, New York State Department of Health | Kirsten St. George, Daryl M. Lamson, Alexis Russel, Matthew Shudt, Melissa A Leisner, Jonathan Plitnick, Navjot Singh, John Kelly, Erasmus Schneider, Erica Lasek-Nesselquist |
| EPI_ISL_1227095 | NYC Pandemic Response Lab | Wadsworth Center, New York State Department of Health | Kirsten St. George, Daryl M. Lamson, Alexis Russel, Matthew Shudt, Melissa A Leisner, Jonathan Plitnick, Navjot Singh, John Kelly, Erasmus Schneider, Erica Lasek-Nesselquist |
| EPI_ISL_1227096 | Columbia University Irving Medical Center | Wadsworth Center, New York State Department of Health | Kirsten St. George, Daryl M. Lamson, Alexis Russel, Matthew Shudt, Melissa A Leisner, Jonathan Plitnick, Navjot Singh, John Kelly, Erasmus Schneider, Erica Lasek-Nesselquist |
| EPI_ISL_1227097, EPI_ISL_1227098, EPI_ISL_1227099, EPI_ISL_1227100, EPI_ISL_1227101, EPI_ISL_1227102, EPI_ISL_1227103, EPI_ISL_1227104, EPI_ISL_1227105, EPI_ISL_1227106, EPI_ISL_1227107, EPI_ISL_1227108 | | | |
| see above | MONTEFIORE MEDICAL CENTER LABORATORIES | Wadsworth Center, New York State Department of Health | Kirsten St. George, Daryl M. Lamson, Alexis Russel, Matthew Shudt, Melissa A Leisner, Jonathan Plitnick, Navjot Singh, John Kelly, Erasmus Schneider, Erica Lasek-Nesselquist |
| EPI_ISL_1227109 | NYC Pandemic Response Lab | Wadsworth Center, New York State Department of Health | Kirsten St. George, Daryl M. Lamson, Alexis Russel, Matthew Shudt, Melissa A Leisner, Jonathan Plitnick, Navjot Singh, John Kelly, Erasmus Schneider, Erica Lasek-Nesselquist |
| EPI_ISL_1227110, EPI_ISL_1227111, EPI_ISL_1227112, EPI_ISL_1227113, EPI_ISL_1227114, EPI_ISL_1227115, EPI_ISL_1227116, EPI_ISL_1227117, EPI_ISL_1227118, EPI_ISL_1227119, EPI_ISL_1227120, EPI_ISL_1227121, EPI_ISL_1227122, EPI_ISL_1227123, EPI_ISL_1227124, EPI_ISL_1227125, EPI_ISL_1227126, EPI_ISL_1227127, EPI_ISL_1227128 | | | |
| see above | MONTEFIORE MEDICAL CENTER LABORATORIES | Wadsworth Center, New York State Department of Health | Kirsten St. George, Daryl M. Lamson, Alexis Russel, Matthew Shudt, Melissa A Leisner, Jonathan Plitnick, Navjot Singh, John Kelly, Erasmus Schneider, Erica Lasek-Nesselquist |
| EPI_ISL_1227129 | SUNY UPSTATE MEDICAL UNIVERSITY | Wadsworth Center, New York State Department of Health | Kirsten St. George, Daryl M. Lamson, Alexis Russel, Matthew Shudt, Melissa A Leisner, Jonathan Plitnick, Navjot Singh, John Kelly, Erasmus Schneider, Erica Lasek-Nesselquist |
| EPI_ISL_1227130, EPI_ISL_1227131, EPI_ISL_1227132, EPI_ISL_1227133 | WESTCHESTER MEDICAL CENTER | Wadsworth Center, New York State Department of Health | Kirsten St. George, Daryl M. Lamson, Alexis Russel, Matthew Shudt, Melissa A Leisner, Jonathan Plitnick, Navjot Singh, John Kelly, Erasmus Schneider, Erica Lasek-Nesselquist |
| EPI_ISL_1227134, EPI_ISL_1227135, EPI_ISL_1227136, EPI_ISL_1227137, EPI_ISL_1227138, EPI_ISL_1227139, EPI_ISL_1227140, EPI_ISL_1227141, EPI_ISL_1227142, EPI_ISL_1227143, EPI_ISL_1227144, EPI_ISL_1227145, EPI_ISL_1227146, EPI_ISL_1227147, EPI_ISL_1227148, EPI_ISL_1227149 | | | |
| see above | NYC Pandemic Response Lab | Wadsworth Center, New York State Department of Health | Kirsten St. George, Daryl M. Lamson, Alexis Russel, Matthew Shudt, Melissa A Leisner, Jonathan Plitnick, Navjot Singh, John Kelly, Erasmus Schneider, Erica Lasek-Nesselquist |
| EPI_ISL_1227150 | MONTEFIORE MEDICAL CENTER LABORATORIES | Wadsworth Center, New York State Department of Health | Kirsten St. George, Daryl M. Lamson, Alexis Russel, Matthew Shudt, Melissa A Leisner, Jonathan Plitnick, Navjot Singh, John Kelly, Erasmus Schneider, Erica Lasek-Nesselquist |
| EPI_ISL_1227151 | NORTH SHORE UNIVERSITY HOSPITAL | Wadsworth Center, New York State Department of Health | Kirsten St. George, Daryl M. Lamson, Alexis Russel, Matthew Shudt, Melissa A Leisner, Jonathan Plitnick, Navjot Singh, John Kelly, Erasmus Schneider, Erica Lasek-Nesselquist |
| EPI_ISL_1227152 | NYC Pandemic Response Lab | Wadsworth Center, New York State Department of Health | Kirsten St. George, Daryl M. Lamson, Alexis Russel, Matthew Shudt, Melissa A Leisner, Jonathan Plitnick, Navjot Singh, John Kelly, Erasmus Schneider, Erica Lasek-Nesselquist |
| EPI_ISL_1227153, EPI_ISL_1227154 | NORTHWELL HEALTH LABORATORIES | Wadsworth Center, New York State Department of Health | Kirsten St. George, Daryl M. Lamson, Alexis Russel, Matthew Shudt, Melissa A Leisner, Jonathan Plitnick, Navjot Singh, John Kelly, Erasmus Schneider, Erica Lasek-Nesselquist |
| EPI_ISL_1227155, EPI_ISL_1227156, EPI_ISL_1227157, EPI_ISL_1227158, EPI_ISL_1227159, EPI_ISL_1227160, EPI_ISL_1227161 | NYC Pandemic Response Lab | Wadsworth Center, New York State Department of Health | Kirsten St. George, Daryl M. Lamson, Alexis Russel, Matthew Shudt, Melissa A Leisner, Jonathan Plitnick, Navjot Singh, John Kelly, Erasmus Schneider, Erica Lasek-Nesselquist |
| EPI_ISL_1227162 | TEMPUS LABS INC | Wadsworth Center, New York State Department of Health | Kirsten St. George, Daryl M. Lamson, Alexis Russel, Matthew Shudt, Melissa A Leisner, Jonathan Plitnick, Navjot Singh, John Kelly, Erasmus Schneider, Erica Lasek-Nesselquist |
| EPI_ISL_1227163 | Columbia University Irving Medical Center | Wadsworth Center, New York State Department of Health | Kirsten St. George, Daryl M. Lamson, Alexis Russel, Matthew Shudt, Melissa A Leisner, Jonathan Plitnick, Navjot Singh, John Kelly, Erasmus Schneider, Erica Lasek-Nesselquist |
| EPI_ISL_1227164, EPI_ISL_1227165 | New York Presbyterian Hospital | Wadsworth Center, New York State Department of Health | Kirsten St. George, Daryl M. Lamson, Alexis Russel, Matthew Shudt, Melissa A Leisner, Jonathan Plitnick, Navjot Singh, John Kelly, Erasmus Schneider, Erica Lasek-Nesselquist |
| EPI_ISL_1227166 | Columbia University Irving Medical Center | Wadsworth Center, New York State Department of Health | Kirsten St. George, Daryl M. Lamson, Alexis Russel, Matthew Shudt, Melissa A Leisner, Jonathan Plitnick, Navjot Singh, John Kelly, Erasmus Schneider, Erica Lasek-Nesselquist |
| EPI_ISL_1227167, EPI_ISL_1227168, EPI_ISL_1227169, EPI_ISL_1227170, EPI_ISL_1227171, EPI_ISL_1227172 | NYC Pandemic Response Lab | Wadsworth Center, New York State Department of Health | Kirsten St. George, Daryl M. Lamson, Alexis Russel, Matthew Shudt, Melissa A Leisner, Jonathan Plitnick, Navjot Singh, John Kelly, Erasmus Schneider, Erica Lasek-Nesselquist |
| EPI_ISL_1227173 | SUNY UPSTATE MEDICAL UNIVERSITY | Wadsworth Center, New York State Department of Health | Kirsten St. George, Daryl M. Lamson, Alexis Russel, Matthew Shudt, Melissa A Leisner, Jonathan Plitnick, Navjot Singh, John Kelly, Erasmus Schneider, Erica Lasek-Nesselquist |
| EPI_ISL_1227174 | WESTCHESTER MEDICAL CENTER | Wadsworth Center, New York State Department of Health | Kirsten St. George, Daryl M. Lamson, Alexis Russel, Matthew Shudt, Melissa A Leisner, Jonathan Plitnick, Navjot Singh, John Kelly, Erasmus Schneider, Erica Lasek-Nesselquist |
| EPI_ISL_1227175 | MEMORIAL SLOAN KETTERING CANCER CENTER | Wadsworth Center, New York State Department of Health | Kirsten St. George, Daryl M. Lamson, Alexis Russel, Matthew Shudt, Melissa A Leisner, Jonathan Plitnick, Navjot Singh, John Kelly, Erasmus Schneider, Erica Lasek-Nesselquist |
| EPI_ISL_1227176, EPI_ISL_1227177 | WESTCHESTER MEDICAL CENTER | Wadsworth Center, New York State Department of Health | Kirsten St. George, Daryl M. Lamson, Alexis Russel, Matthew Shudt, Melissa A Leisner, Jonathan Plitnick, Navjot Singh, John Kelly, Erasmus Schneider, Erica Lasek-Nesselquist |

| | | | |
|---|---|---|---|
| EPI_ISL_1227178, EPI_ISL_1227179, EPI_ISL_1227180 | MEMORIAL SLOAN KETTERING CANCER CENTER | Wadsworth Center, New York State Department of Health | Kirsten St. George, Daryl M. Lamson, Alexis Russel, Matthew Shudt, Melissa A Leisner, Jonathan Plitnick, Navjot Singh, John Kelly, Erasmus Schneider, Erica Lasek-Nesselquist |
| EPI_ISL_1227181 | NORTHWELL HEALTH LABORATORIES | Wadsworth Center, New York State Department of Health | Kirsten St. George, Daryl M. Lamson, Alexis Russel, Matthew Shudt, Melissa A Leisner, Jonathan Plitnick, Navjot Singh, John Kelly, Erasmus Schneider, Erica Lasek-Nesselquist |
| EPI_ISL_1227182, EPI_ISL_1227183 | New York Presbyterian Hospital | Wadsworth Center, New York State Department of Health | Kirsten St. George, Daryl M. Lamson, Alexis Russel, Matthew Shudt, Melissa A Leisner, Jonathan Plitnick, Navjot Singh, John Kelly, Erasmus Schneider, Erica Lasek-Nesselquist |
| EPI_ISL_1227184 | NYC Pandemic Response Lab | Wadsworth Center, New York State Department of Health | Kirsten St. George, Daryl M. Lamson, Alexis Russel, Matthew Shudt, Melissa A Leisner, Jonathan Plitnick, Navjot Singh, John Kelly, Erasmus Schneider, Erica Lasek-Nesselquist |
| EPI_ISL_1227185 | New York Presbyterian Hospital | Wadsworth Center, New York State Department of Health | Kirsten St. George, Daryl M. Lamson, Alexis Russel, Matthew Shudt, Melissa A Leisner, Jonathan Plitnick, Navjot Singh, John Kelly, Erasmus Schneider, Erica Lasek-Nesselquist |
| EPI_ISL_1227186, EPI_ISL_1227187, EPI_ISL_1227188 | NYC Pandemic Response Lab | Wadsworth Center, New York State Department of Health | Kirsten St. George, Daryl M. Lamson, Alexis Russel, Matthew Shudt, Melissa A Leisner, Jonathan Plitnick, Navjot Singh, John Kelly, Erasmus Schneider, Erica Lasek-Nesselquist |
| EPI_ISL_1227189 | WESTCHESTER MEDICAL CENTER | Wadsworth Center, New York State Department of Health | Kirsten St. George, Daryl M. Lamson, Alexis Russel, Matthew Shudt, Melissa A Leisner, Jonathan Plitnick, Navjot Singh, John Kelly, Erasmus Schneider, Erica Lasek-Nesselquist |
| EPI_ISL_1227190, EPI_ISL_1227191, EPI_ISL_1227192, EPI_ISL_1227193, EPI_ISL_1227194, EPI_ISL_1227195, EPI_ISL_1227196, EPI_ISL_1227197, EPI_ISL_1227198, EPI_ISL_1227199, EPI_ISL_1227200, EPI_ISL_1227201, EPI_ISL_1227202, EPI_ISL_1227203, EPI_ISL_1227204, EPI_ISL_1227205 | | | |
| see above | NYC Pandemic Response Lab | Wadsworth Center, New York State Department of Health | Kirsten St. George, Daryl M. Lamson, Alexis Russel, Matthew Shudt, Melissa A Leisner, Jonathan Plitnick, Navjot Singh, John Kelly, Erasmus Schneider, Erica Lasek-Nesselquist |
| EPI_ISL_1227206, EPI_ISL_1227207 | WESTCHESTER MEDICAL CENTER | Wadsworth Center, New York State Department of Health | Kirsten St. George, Daryl M. Lamson, Alexis Russel, Matthew Shudt, Melissa A Leisner, Jonathan Plitnick, Navjot Singh, John Kelly, Erasmus Schneider, Erica Lasek-Nesselquist |
| EPI_ISL_1227208, EPI_ISL_1227209 | ALBANY MEDICAL CENTER | Wadsworth Center, New York State Department of Health | Kirsten St. George, Daryl M. Lamson, Alexis Russel, Matthew Shudt, Melissa A Leisner, Jonathan Plitnick, Navjot Singh, John Kelly, Erasmus Schneider, Erica Lasek-Nesselquist |
| EPI_ISL_1227210 | THE MARY IMOGENE BASSETT HOSPITAL | Wadsworth Center, New York State Department of Health | Kirsten St. George, Daryl M. Lamson, Alexis Russel, Matthew Shudt, Melissa A Leisner, Jonathan Plitnick, Navjot Singh, John Kelly, Erasmus Schneider, Erica Lasek-Nesselquist |
| EPI_ISL_1227211 | MEMORIAL SLOAN KETTERING CANCER CENTER | Wadsworth Center, New York State Department of Health | Kirsten St. George, Daryl M. Lamson, Alexis Russel, Matthew Shudt, Melissa A Leisner, Jonathan Plitnick, Navjot Singh, John Kelly, Erasmus Schneider, Erica Lasek-Nesselquist |
| EPI_ISL_1227212, EPI_ISL_1227213 | NYC Pandemic Response Lab | Wadsworth Center, New York State Department of Health | Kirsten St. George, Daryl M. Lamson, Alexis Russel, Matthew Shudt, Melissa A Leisner, Jonathan Plitnick, Navjot Singh, John Kelly, Erasmus Schneider, Erica Lasek-Nesselquist |
| EPI_ISL_1227214 | MEMORIAL SLOAN KETTERING CANCER CENTER | Wadsworth Center, New York State Department of Health | Kirsten St. George, Daryl M. Lamson, Alexis Russel, Matthew Shudt, Melissa A Leisner, Jonathan Plitnick, Navjot Singh, John Kelly, Erasmus Schneider, Erica Lasek-Nesselquist |
| EPI_ISL_1227215, EPI_ISL_1227216, EPI_ISL_1227217, EPI_ISL_1227218, EPI_ISL_1227219, EPI_ISL_1227220 | WESTCHESTER MEDICAL CENTER | Wadsworth Center, New York State Department of Health | Kirsten St. George, Daryl M. Lamson, Alexis Russel, Matthew Shudt, Melissa A Leisner, Jonathan Plitnick, Navjot Singh, John Kelly, Erasmus Schneider, Erica Lasek-Nesselquist |
| EPI_ISL_1227221 | NYC Pandemic Response Lab | Wadsworth Center, New York State Department of Health | Kirsten St. George, Daryl M. Lamson, Alexis Russel, Matthew Shudt, Melissa A Leisner, Jonathan Plitnick, Navjot Singh, John Kelly, Erasmus Schneider, Erica Lasek-Nesselquist |
| EPI_ISL_1227222 | Columbia University Irving Medical Center | Wadsworth Center, New York State Department of Health | Kirsten St. George, Daryl M. Lamson, Alexis Russel, Matthew Shudt, Melissa A Leisner, Jonathan Plitnick, Navjot Singh, John Kelly, Erasmus Schneider, Erica Lasek-Nesselquist |
| EPI_ISL_1227223, EPI_ISL_1227224, EPI_ISL_1227225 | WESTCHESTER MEDICAL CENTER | Wadsworth Center, New York State Department of Health | Kirsten St. George, Daryl M. Lamson, Alexis Russel, Matthew Shudt, Melissa A Leisner, Jonathan Plitnick, Navjot Singh, John Kelly, Erasmus Schneider, Erica Lasek-Nesselquist |
| EPI_ISL_1227226, EPI_ISL_1227227 | MONTEFIORE MEDICAL CENTER LABORATORIES | Wadsworth Center, New York State Department of Health | Kirsten St. George, Daryl M. Lamson, Alexis Russel, Matthew Shudt, Melissa A Leisner, Jonathan Plitnick, Navjot Singh, John Kelly, Erasmus Schneider, Erica Lasek-Nesselquist |
| EPI_ISL_1227228, EPI_ISL_1227229 | WESTCHESTER MEDICAL CENTER | Wadsworth Center, New York State Department of Health | Kirsten St. George, Daryl M. Lamson, Alexis Russel, Matthew Shudt, Melissa A Leisner, Jonathan Plitnick, Navjot Singh, John Kelly, Erasmus Schneider, Erica Lasek-Nesselquist |
| EPI_ISL_1227230 | MEMORIAL SLOAN KETTERING CANCER CENTER | Wadsworth Center, New York State Department of Health | Kirsten St. George, Daryl M. Lamson, Alexis Russel, Matthew Shudt, Melissa A Leisner, Jonathan Plitnick, Navjot Singh, John Kelly, Erasmus Schneider, Erica Lasek-Nesselquist |
| EPI_ISL_1227231 | URMC LABS | Wadsworth Center, New York State Department of Health | Kirsten St. George, Daryl M. Lamson, Alexis Russel, Matthew Shudt, Melissa A Leisner, Jonathan Plitnick, Navjot Singh, John Kelly, Erasmus Schneider, Erica Lasek-Nesselquist |
| EPI_ISL_1227232, EPI_ISL_1227233 | ALBANY MEDICAL CENTER | Wadsworth Center, New York State Department of Health | Kirsten St. George, Daryl M. Lamson, Alexis Russel, Matthew Shudt, Melissa A Leisner, Jonathan Plitnick, Navjot Singh, John Kelly, Erasmus Schneider, Erica Lasek-Nesselquist |
| EPI_ISL_1227234 | Wadsworth Center, New York State Department of Health | Wadsworth Center, New York State Department of Health | Kirsten St. George, Daryl M. Lamson, Alexis Russel, Matthew Shudt, Melissa A Leisner, Jonathan Plitnick, Navjot Singh, John Kelly, Erasmus Schneider, Erica Lasek-Nesselquist |
| EPI_ISL_1227235 | ALBANY MEDICAL CENTER | Wadsworth Center, New York State Department of Health | Kirsten St. George, Daryl M. Lamson, Alexis Russel, Matthew Shudt, Melissa A Leisner, Jonathan Plitnick, Navjot Singh, John Kelly, Erasmus Schneider, Erica Lasek-Nesselquist |
| EPI_ISL_1227236 | MEMORIAL SLOAN KETTERING CANCER CENTER | Wadsworth Center, New York State Department of Health | Kirsten St. George, Daryl M. Lamson, Alexis Russel, Matthew Shudt, Melissa A Leisner, Jonathan Plitnick, Navjot Singh, John Kelly, Erasmus Schneider, Erica Lasek-Nesselquist |
| EPI_ISL_1227237 | Columbia University Irving Medical Center | Wadsworth Center, New York State Department of Health | Kirsten St. George, Daryl M. Lamson, Alexis Russel, Matthew Shudt, Melissa A Leisner, Jonathan Plitnick, Navjot Singh, John Kelly, Erasmus Schneider, Erica Lasek-Nesselquist |
| EPI_ISL_1227238 | NYC Pandemic Response Lab | Wadsworth Center, New York State Department of Health | Kirsten St. George, Daryl M. Lamson, Alexis Russel, Matthew Shudt, Melissa A Leisner, Jonathan Plitnick, Navjot Singh, John Kelly, Erasmus Schneider, Erica Lasek-Nesselquist |
| EPI_ISL_1227239, EPI_ISL_1227240, EPI_ISL_1227241 | SUNY UPSTATE MEDICAL UNIVERSITY | Wadsworth Center, New York State Department of Health | Kirsten St. George, Daryl M. Lamson, Alexis Russel, Matthew Shudt, Melissa A Leisner, Jonathan Plitnick, Navjot Singh, John Kelly, Erasmus Schneider, Erica Lasek-Nesselquist |
| EPI_ISL_1227242, EPI_ISL_1227243 | MEMORIAL SLOAN KETTERING CANCER CENTER | Wadsworth Center, New York State Department of Health | Kirsten St. George, Daryl M. Lamson, Alexis Russel, Matthew Shudt, Melissa A Leisner, Jonathan Plitnick, Navjot Singh, John Kelly, Erasmus Schneider, Erica Lasek-Nesselquist |
| EPI_ISL_1227244 | NYC Pandemic Response Lab | Wadsworth Center, New York State Department of Health | Kirsten St. George, Daryl M. Lamson, Alexis Russel, Matthew Shudt, Melissa A Leisner, Jonathan Plitnick, Navjot Singh, John Kelly, Erasmus Schneider, Erica Lasek-Nesselquist |
| EPI_ISL_1227245 | SARATOGA HOSPITAL LABORATORY | Wadsworth Center, New York State Department of Health | Kirsten St. George, Daryl M. Lamson, Alexis Russel, Matthew Shudt, Melissa A Leisner, Jonathan Plitnick, Navjot Singh, John Kelly, Erasmus Schneider, Erica Lasek-Nesselquist |
| EPI_ISL_1227246 | Wadsworth Center, New York State Department of Health | Wadsworth Center, New York State Department of Health | Kirsten St. George, Daryl M. Lamson, Alexis Russel, Matthew Shudt, Melissa A Leisner, Jonathan Plitnick, Navjot Singh, John Kelly, Erasmus Schneider, Erica Lasek-Nesselquist |
| EPI_ISL_1227247, EPI_ISL_1227248 | WESTCHESTER MEDICAL CENTER | Wadsworth Center, New York State Department of Health | Kirsten St. George, Daryl M. Lamson, Alexis Russel, Matthew Shudt, Melissa A Leisner, Jonathan Plitnick, Navjot Singh, John Kelly, Erasmus Schneider, Erica Lasek-Nesselquist |

| | | | |
|---|---|---|---|
| EPI_ISL_1227249, EPI_ISL_1227250, EPI_ISL_1227251, EPI_ISL_1227252, EPI_ISL_1227253 | ALBANY MEDICAL CENTER | Wadsworth Center, New York State Department of Health | Kirsten St. George, Daryl M. Lamson, Alexis Russel, Matthew Shudt, Melissa A Leisner, Jonathan Plitnick, Navjot Singh, John Kelly, Erasmus Schneider, Erica Lasek-Nesselquist |
| EPI_ISL_1227254, EPI_ISL_1227255 | Wadsworth Center, New York State Department of Health | Wadsworth Center, New York State Department of Health | Kirsten St. George, Daryl M. Lamson, Alexis Russel, Matthew Shudt, Melissa A Leisner, Jonathan Plitnick, Navjot Singh, John Kelly, Erasmus Schneider, Erica Lasek-Nesselquist |
| EPI_ISL_1227256 | ALBANY MEDICAL CENTER | Wadsworth Center, New York State Department of Health | Kirsten St. George, Daryl M. Lamson, Alexis Russel, Matthew Shudt, Melissa A Leisner, Jonathan Plitnick, Navjot Singh, John Kelly, Erasmus Schneider, Erica Lasek-Nesselquist |
| EPI_ISL_1227257, EPI_ISL_1227258, EPI_ISL_1227259, EPI_ISL_1227260, EPI_ISL_1227261, EPI_ISL_1227262 | Wadsworth Center, New York State Department of Health | Wadsworth Center, New York State Department of Health | Kirsten St. George, Daryl M. Lamson, Alexis Russel, Matthew Shudt, Melissa A Leisner, Jonathan Plitnick, Navjot Singh, John Kelly, Erasmus Schneider, Erica Lasek-Nesselquist |
| EPI_ISL_1227263, EPI_ISL_1227264, EPI_ISL_1227265, EPI_ISL_1227266, EPI_ISL_1227267, EPI_ISL_1227268, EPI_ISL_1227269, EPI_ISL_1227270, EPI_ISL_1227271 | ALBANY MEDICAL CENTER | Wadsworth Center, New York State Department of Health | Kirsten St. George, Daryl M. Lamson, Alexis Russel, Matthew Shudt, Melissa A Leisner, Jonathan Plitnick, Navjot Singh, John Kelly, Erasmus Schneider, Erica Lasek-Nesselquist |
| EPI_ISL_1227272, EPI_ISL_1227273, EPI_ISL_1227274, EPI_ISL_1227275, EPI_ISL_1227276, EPI_ISL_1227277 | URMC LABS | Wadsworth Center, New York State Department of Health | Kirsten St. George, Daryl M. Lamson, Alexis Russel, Matthew Shudt, Melissa A Leisner, Jonathan Plitnick, Navjot Singh, John Kelly, Erasmus Schneider, Erica Lasek-Nesselquist |
| EPI_ISL_1227278, EPI_ISL_1227279, EPI_ISL_1227280, EPI_ISL_1227281, EPI_ISL_1227282 | MEMORIAL SLOAN KETTERING CANCER CENTER | Wadsworth Center, New York State Department of Health | Kirsten St. George, Daryl M. Lamson, Alexis Russel, Matthew Shudt, Melissa A Leisner, Jonathan Plitnick, Navjot Singh, John Kelly, Erasmus Schneider, Erica Lasek-Nesselquist |
| EPI_ISL_1227283 | Columbia University Irving Medical Center | Wadsworth Center, New York State Department of Health | Kirsten St. George, Daryl M. Lamson, Alexis Russel, Matthew Shudt, Melissa A Leisner, Jonathan Plitnick, Navjot Singh, John Kelly, Erasmus Schneider, Erica Lasek-Nesselquist |
| EPI_ISL_1227284, EPI_ISL_1227285, EPI_ISL_1227286, EPI_ISL_1227287, EPI_ISL_1227288 | MEMORIAL SLOAN KETTERING CANCER CENTER | Wadsworth Center, New York State Department of Health | Kirsten St. George, Daryl M. Lamson, Alexis Russel, Matthew Shudt, Melissa A Leisner, Jonathan Plitnick, Navjot Singh, John Kelly, Erasmus Schneider, Erica Lasek-Nesselquist |
| EPI_ISL_1227289, EPI_ISL_1227290 | Columbia University Irving Medical Center | Wadsworth Center, New York State Department of Health | Kirsten St. George, Daryl M. Lamson, Alexis Russel, Matthew Shudt, Melissa A Leisner, Jonathan Plitnick, Navjot Singh, John Kelly, Erasmus Schneider, Erica Lasek-Nesselquist |
| EPI_ISL_1227291, EPI_ISL_1227292 | MEMORIAL SLOAN KETTERING CANCER CENTER | Wadsworth Center, New York State Department of Health | Kirsten St. George, Daryl M. Lamson, Alexis Russel, Matthew Shudt, Melissa A Leisner, Jonathan Plitnick, Navjot Singh, John Kelly, Erasmus Schneider, Erica Lasek-Nesselquist |
| EPI_ISL_1227293 | Columbia University Irving Medical Center | Wadsworth Center, New York State Department of Health | Kirsten St. George, Daryl M. Lamson, Alexis Russel, Matthew Shudt, Melissa A Leisner, Jonathan Plitnick, Navjot Singh, John Kelly, Erasmus Schneider, Erica Lasek-Nesselquist |
| EPI_ISL_1227294 | WESTCHESTER MEDICAL CENTER | Wadsworth Center, New York State Department of Health | Kirsten St. George, Daryl M. Lamson, Alexis Russel, Matthew Shudt, Melissa A Leisner, Jonathan Plitnick, Navjot Singh, John Kelly, Erasmus Schneider, Erica Lasek-Nesselquist |
| EPI_ISL_1227295, EPI_ISL_1227296 | Columbia University Irving Medical Center | Wadsworth Center, New York State Department of Health | Kirsten St. George, Daryl M. Lamson, Alexis Russel, Matthew Shudt, Melissa A Leisner, Jonathan Plitnick, Navjot Singh, John Kelly, Erasmus Schneider, Erica Lasek-Nesselquist |
| EPI_ISL_1227297, EPI_ISL_1227298, EPI_ISL_1227299, EPI_ISL_1227300, EPI_ISL_1227301 | MONTEFIORE MEDICAL CENTER LABORATORIES | Wadsworth Center, New York State Department of Health | Kirsten St. George, Daryl M. Lamson, Alexis Russel, Matthew Shudt, Melissa A Leisner, Jonathan Plitnick, Navjot Singh, John Kelly, Erasmus Schneider, Erica Lasek-Nesselquist |
| EPI_ISL_1227302, EPI_ISL_1227303 | WESTCHESTER MEDICAL CENTER | Wadsworth Center, New York State Department of Health | Kirsten St. George, Daryl M. Lamson, Alexis Russel, Matthew Shudt, Melissa A Leisner, Jonathan Plitnick, Navjot Singh, John Kelly, Erasmus Schneider, Erica Lasek-Nesselquist |
| EPI_ISL_1227304, EPI_ISL_1227305 | Columbia University Irving Medical Center | Wadsworth Center, New York State Department of Health | Kirsten St. George, Daryl M. Lamson, Alexis Russel, Matthew Shudt, Melissa A Leisner, Jonathan Plitnick, Navjot Singh, John Kelly, Erasmus Schneider, Erica Lasek-Nesselquist |
| EPI_ISL_1227306, EPI_ISL_1227307, EPI_ISL_1227308, EPI_ISL_1227309, EPI_ISL_1227310, EPI_ISL_1227311, EPI_ISL_1227312, EPI_ISL_1227313 | MONTEFIORE MEDICAL CENTER LABORATORIES | Wadsworth Center, New York State Department of Health | Kirsten St. George, Daryl M. Lamson, Alexis Russel, Matthew Shudt, Melissa A Leisner, Jonathan Plitnick, Navjot Singh, John Kelly, Erasmus Schneider, Erica Lasek-Nesselquist |
| EPI_ISL_1227314 | Columbia University Irving Medical Center | Wadsworth Center, New York State Department of Health | Kirsten St. George, Daryl M. Lamson, Alexis Russel, Matthew Shudt, Melissa A Leisner, Jonathan Plitnick, Navjot Singh, John Kelly, Erasmus Schneider, Erica Lasek-Nesselquist |
| EPI_ISL_1227315, EPI_ISL_1227316, EPI_ISL_1227317, EPI_ISL_1227318, EPI_ISL_1227319 | MONTEFIORE MEDICAL CENTER LABORATORIES | Wadsworth Center, New York State Department of Health | Kirsten St. George, Daryl M. Lamson, Alexis Russel, Matthew Shudt, Melissa A Leisner, Jonathan Plitnick, Navjot Singh, John Kelly, Erasmus Schneider, Erica Lasek-Nesselquist |
| EPI_ISL_1227320 | Columbia University Irving Medical Center | Wadsworth Center, New York State Department of Health | Kirsten St. George, Daryl M. Lamson, Alexis Russel, Matthew Shudt, Melissa A Leisner, Jonathan Plitnick, Navjot Singh, John Kelly, Erasmus Schneider, Erica Lasek-Nesselquist |
| EPI_ISL_1227321, EPI_ISL_1227322, EPI_ISL_1227323, EPI_ISL_1227324, EPI_ISL_1227325 | MONTEFIORE MEDICAL CENTER LABORATORIES | Wadsworth Center, New York State Department of Health | Kirsten St. George, Daryl M. Lamson, Alexis Russel, Matthew Shudt, Melissa A Leisner, Jonathan Plitnick, Navjot Singh, John Kelly, Erasmus Schneider, Erica Lasek-Nesselquist |
| EPI_ISL_1227326 | Columbia University Irving Medical Center | Wadsworth Center, New York State Department of Health | Kirsten St. George, Daryl M. Lamson, Alexis Russel, Matthew Shudt, Melissa A Leisner, Jonathan Plitnick, Navjot Singh, John Kelly, Erasmus Schneider, Erica Lasek-Nesselquist |
| EPI_ISL_1227327, EPI_ISL_1227328, EPI_ISL_1227329, EPI_ISL_1227330, EPI_ISL_1227331, EPI_ISL_1227332, EPI_ISL_1227333, EPI_ISL_1227334, EPI_ISL_1227335, EPI_ISL_1227336 | MONTEFIORE MEDICAL CENTER LABORATORIES | Wadsworth Center, New York State Department of Health | Kirsten St. George, Daryl M. Lamson, Alexis Russel, Matthew Shudt, Melissa A Leisner, Jonathan Plitnick, Navjot Singh, John Kelly, Erasmus Schneider, Erica Lasek-Nesselquist |
| EPI_ISL_1227337 | WESTCHESTER MEDICAL CENTER | Wadsworth Center, New York State Department of Health | Kirsten St. George, Daryl M. Lamson, Alexis Russel, Matthew Shudt, Melissa A Leisner, Jonathan Plitnick, Navjot Singh, John Kelly, Erasmus Schneider, Erica Lasek-Nesselquist |
| EPI_ISL_1227338, EPI_ISL_1227339, EPI_ISL_1227340, EPI_ISL_1227341, EPI_ISL_1227342, EPI_ISL_1227343, EPI_ISL_1227344, EPI_ISL_1227345, EPI_ISL_1227346, EPI_ISL_1227347, EPI_ISL_1227348, EPI_ISL_1227349, EPI_ISL_1227350, EPI_ISL_1227351, EPI_ISL_1227352, EPI_ISL_1227353, EPI_ISL_1227354, EPI_ISL_1227355, EPI_ISL_1227356 | | | |
| see above | MONTEFIORE MEDICAL CENTER LABORATORIES | Wadsworth Center, New York State Department of Health | Kirsten St. George, Daryl M. Lamson, Alexis Russel, Matthew Shudt, Melissa A Leisner, Jonathan Plitnick, Navjot Singh, John Kelly, Erasmus Schneider, Erica Lasek-Nesselquist |
| EPI_ISL_1227357 | Columbia University Irving Medical Center | Wadsworth Center, New York State Department of Health | Kirsten St. George, Daryl M. Lamson, Alexis Russel, Matthew Shudt, Melissa A Leisner, Jonathan Plitnick, Navjot Singh, John Kelly, Erasmus Schneider, Erica Lasek-Nesselquist |
| EPI_ISL_1227358, EPI_ISL_1227359 | Wadsworth Center, New York State Department of Health | Wadsworth Center, New York State Department of Health | Kirsten St. George, Daryl M. Lamson, Alexis Russel, Matthew Shudt, Melissa A Leisner, Jonathan Plitnick, Navjot Singh, John Kelly, Erasmus Schneider, |

| | | | |
|---|---|---|---|
| | | | Erica Lasek-Nesselquist |
| EPI_ISL_1227360, EPI_ISL_1227361, EPI_ISL_1227362, EPI_ISL_1227363 | Columbia University Irving Medical Center | Wadsworth Center, New York State Department of Health | Kirsten St. George, Daryl M. Lamson, Alexis Russel, Matthew Shudt, Melissa A Leisner, Jonathan Plitnick, Navjot Singh, John Kelly, Erasmus Schneider, Erica Lasek-Nesselquist |
| EPI_ISL_1227364, EPI_ISL_1227365, EPI_ISL_1227366, EPI_ISL_1227367, EPI_ISL_1227368, EPI_ISL_1227369, EPI_ISL_1227370, EPI_ISL_1227371, EPI_ISL_1227372, EPI_ISL_1227373, EPI_ISL_1227374, EPI_ISL_1227375, EPI_ISL_1227376, EPI_ISL_1227377, EPI_ISL_1227378 | | | |
| see above | MONTEFIORE MEDICAL CENTER LABORATORIES | Wadsworth Center, New York State Department of Health | Kirsten St. George, Daryl M. Lamson, Alexis Russel, Matthew Shudt, Melissa A Leisner, Jonathan Plitnick, Navjot Singh, John Kelly, Erasmus Schneider, Erica Lasek-Nesselquist |
| EPI_ISL_1227379 | Wadsworth Center, New York State Department of Health | Wadsworth Center, New York State Department of Health | Kirsten St. George, Daryl M. Lamson, Alexis Russel, Matthew Shudt, Melissa A Leisner, Jonathan Plitnick, Navjot Singh, John Kelly, Erasmus Schneider, Erica Lasek-Nesselquist |
| EPI_ISL_1227380, EPI_ISL_1227381, EPI_ISL_1227382, EPI_ISL_1227383, EPI_ISL_1227384, EPI_ISL_1227385, EPI_ISL_1227386, EPI_ISL_1227387, EPI_ISL_1227388, EPI_ISL_1227389, EPI_ISL_1227390, EPI_ISL_1227391, EPI_ISL_1227392, EPI_ISL_1227393, EPI_ISL_1227394, EPI_ISL_1227395, EPI_ISL_1227396, EPI_ISL_1227397, EPI_ISL_1227398, EPI_ISL_1227399, EPI_ISL_1227400 | | | |
| see above | MONTEFIORE MEDICAL CENTER LABORATORIES | Wadsworth Center, New York State Department of Health | Kirsten St. George, Daryl M. Lamson, Alexis Russel, Matthew Shudt, Melissa A Leisner, Jonathan Plitnick, Navjot Singh, John Kelly, Erasmus Schneider, Erica Lasek-Nesselquist |
| EPI_ISL_1227401, EPI_ISL_1227402 | Columbia University Irving Medical Center | Wadsworth Center, New York State Department of Health | Kirsten St. George, Daryl M. Lamson, Alexis Russel, Matthew Shudt, Melissa A Leisner, Jonathan Plitnick, Navjot Singh, John Kelly, Erasmus Schneider, Erica Lasek-Nesselquist |
| EPI_ISL_1227403, EPI_ISL_1227404, EPI_ISL_1227405 | MONTEFIORE MEDICAL CENTER LABORATORIES | Wadsworth Center, New York State Department of Health | Kirsten St. George, Daryl M. Lamson, Alexis Russel, Matthew Shudt, Melissa A Leisner, Jonathan Plitnick, Navjot Singh, John Kelly, Erasmus Schneider, Erica Lasek-Nesselquist |
| EPI_ISL_1227406, EPI_ISL_1227407, EPI_ISL_1227408, EPI_ISL_1227409 | Columbia University Irving Medical Center | Wadsworth Center, New York State Department of Health | Kirsten St. George, Daryl M. Lamson, Alexis Russel, Matthew Shudt, Melissa A Leisner, Jonathan Plitnick, Navjot Singh, John Kelly, Erasmus Schneider, Erica Lasek-Nesselquist |
| EPI_ISL_1227410 | New York Presbyterian Hospital | Wadsworth Center, New York State Department of Health | Kirsten St. George, Daryl M. Lamson, Alexis Russel, Matthew Shudt, Melissa A Leisner, Jonathan Plitnick, Navjot Singh, John Kelly, Erasmus Schneider, Erica Lasek-Nesselquist |
| EPI_ISL_1227411, EPI_ISL_1227412, EPI_ISL_1227413, EPI_ISL_1227414, EPI_ISL_1227415 | MONTEFIORE MEDICAL CENTER LABORATORIES | Wadsworth Center, New York State Department of Health | Kirsten St. George, Daryl M. Lamson, Alexis Russel, Matthew Shudt, Melissa A Leisner, Jonathan Plitnick, Navjot Singh, John Kelly, Erasmus Schneider, Erica Lasek-Nesselquist |
| EPI_ISL_1227416 | Columbia University Irving Medical Center | Wadsworth Center, New York State Department of Health | Kirsten St. George, Daryl M. Lamson, Alexis Russel, Matthew Shudt, Melissa A Leisner, Jonathan Plitnick, Navjot Singh, John Kelly, Erasmus Schneider, Erica Lasek-Nesselquist |
| EPI_ISL_1227417 | New York Presbyterian Hospital | Wadsworth Center, New York State Department of Health | Kirsten St. George, Daryl M. Lamson, Alexis Russel, Matthew Shudt, Melissa A Leisner, Jonathan Plitnick, Navjot Singh, John Kelly, Erasmus Schneider, Erica Lasek-Nesselquist |
| EPI_ISL_1227418, EPI_ISL_1227419, EPI_ISL_1227420, EPI_ISL_1227421 | MONTEFIORE MEDICAL CENTER LABORATORIES | Wadsworth Center, New York State Department of Health | Kirsten St. George, Daryl M. Lamson, Alexis Russel, Matthew Shudt, Melissa A Leisner, Jonathan Plitnick, Navjot Singh, John Kelly, Erasmus Schneider, Erica Lasek-Nesselquist |
| EPI_ISL_1227422, EPI_ISL_1227423 | NYC Pandemic Response Lab | Wadsworth Center, New York State Department of Health | Kirsten St. George, Daryl M. Lamson, Alexis Russel, Matthew Shudt, Melissa A Leisner, Jonathan Plitnick, Navjot Singh, John Kelly, Erasmus Schneider, Erica Lasek-Nesselquist |
| EPI_ISL_1227424, EPI_ISL_1227425, EPI_ISL_1227426, EPI_ISL_1227427 | MONTEFIORE MEDICAL CENTER LABORATORIES | Wadsworth Center, New York State Department of Health | Kirsten St. George, Daryl M. Lamson, Alexis Russel, Matthew Shudt, Melissa A Leisner, Jonathan Plitnick, Navjot Singh, John Kelly, Erasmus Schneider, Erica Lasek-Nesselquist |
| EPI_ISL_1227428, EPI_ISL_1227429 | NYC Pandemic Response Lab | Wadsworth Center, New York State Department of Health | Kirsten St. George, Daryl M. Lamson, Alexis Russel, Matthew Shudt, Melissa A Leisner, Jonathan Plitnick, Navjot Singh, John Kelly, Erasmus Schneider, Erica Lasek-Nesselquist |
| EPI_ISL_1227430, EPI_ISL_1227431, EPI_ISL_1227432, EPI_ISL_1227433, EPI_ISL_1227434 | MONTEFIORE MEDICAL CENTER LABORATORIES | Wadsworth Center, New York State Department of Health | Kirsten St. George, Daryl M. Lamson, Alexis Russel, Matthew Shudt, Melissa A Leisner, Jonathan Plitnick, Navjot Singh, John Kelly, Erasmus Schneider, Erica Lasek-Nesselquist |
| EPI_ISL_1227435 | Columbia University Irving Medical Center | Wadsworth Center, New York State Department of Health | Kirsten St. George, Daryl M. Lamson, Alexis Russel, Matthew Shudt, Melissa A Leisner, Jonathan Plitnick, Navjot Singh, John Kelly, Erasmus Schneider, Erica Lasek-Nesselquist |
| EPI_ISL_1227436, EPI_ISL_1227437, EPI_ISL_1227438, EPI_ISL_1227439 | MONTEFIORE MEDICAL CENTER LABORATORIES | Wadsworth Center, New York State Department of Health | Kirsten St. George, Daryl M. Lamson, Alexis Russel, Matthew Shudt, Melissa A Leisner, Jonathan Plitnick, Navjot Singh, John Kelly, Erasmus Schneider, Erica Lasek-Nesselquist |
| EPI_ISL_1227440 | NYC Pandemic Response Lab | Wadsworth Center, New York State Department of Health | Kirsten St. George, Daryl M. Lamson, Alexis Russel, Matthew Shudt, Melissa A Leisner, Jonathan Plitnick, Navjot Singh, John Kelly, Erasmus Schneider, Erica Lasek-Nesselquist |
| EPI_ISL_1227441 | WESTCHESTER MEDICAL CENTER | Wadsworth Center, New York State Department of Health | Kirsten St. George, Daryl M. Lamson, Alexis Russel, Matthew Shudt, Melissa A Leisner, Jonathan Plitnick, Navjot Singh, John Kelly, Erasmus Schneider, Erica Lasek-Nesselquist |
| EPI_ISL_1227442, EPI_ISL_1227443, EPI_ISL_1227444, EPI_ISL_1227445, EPI_ISL_1227446 | MONTEFIORE MEDICAL CENTER LABORATORIES | Wadsworth Center, New York State Department of Health | Kirsten St. George, Daryl M. Lamson, Alexis Russel, Matthew Shudt, Melissa A Leisner, Jonathan Plitnick, Navjot Singh, John Kelly, Erasmus Schneider, Erica Lasek-Nesselquist |
| EPI_ISL_1227447 | SUNY UPSTATE MEDICAL UNIVERSITY | Wadsworth Center, New York State Department of Health | Kirsten St. George, Daryl M. Lamson, Alexis Russel, Matthew Shudt, Melissa A Leisner, Jonathan Plitnick, Navjot Singh, John Kelly, Erasmus Schneider, Erica Lasek-Nesselquist |
| EPI_ISL_1227448, EPI_ISL_1227449, EPI_ISL_1227450, EPI_ISL_1227451, EPI_ISL_1227452, EPI_ISL_1227453, EPI_ISL_1227454, EPI_ISL_1227455 | URMC LABS | Wadsworth Center, New York State Department of Health | Kirsten St. George, Daryl M. Lamson, Alexis Russel, Matthew Shudt, Melissa A Leisner, Jonathan Plitnick, Navjot Singh, John Kelly, Erasmus Schneider, Erica Lasek-Nesselquist |
| EPI_ISL_1227456, EPI_ISL_1227457 | THE MARY IMOGENE BASSETT HOSPITAL | Wadsworth Center, New York State Department of Health | Kirsten St. George, Daryl M. Lamson, Alexis Russel, Matthew Shudt, Melissa A Leisner, Jonathan Plitnick, Navjot Singh, John Kelly, Erasmus Schneider, Erica Lasek-Nesselquist |
| EPI_ISL_1227458, EPI_ISL_1227459, EPI_ISL_1227460, EPI_ISL_1227461, EPI_ISL_1227462 | ALBANY MEDICAL CENTER | Wadsworth Center, New York State Department of Health | Kirsten St. George, Daryl M. Lamson, Alexis Russel, Matthew Shudt, Melissa A Leisner, Jonathan Plitnick, Navjot Singh, John Kelly, Erasmus Schneider, Erica Lasek-Nesselquist |
| EPI_ISL_1227463, EPI_ISL_1227464, EPI_ISL_1227465 | SUNY UPSTATE MEDICAL UNIVERSITY | Wadsworth Center, New York State Department of Health | Kirsten St. George, Daryl M. Lamson, Alexis Russel, Matthew Shudt, Melissa A Leisner, Jonathan Plitnick, Navjot Singh, John Kelly, Erasmus Schneider, Erica Lasek-Nesselquist |
| EPI_ISL_1227466 | THE MARY IMOGENE BASSETT HOSPITAL | Wadsworth Center, New York State Department of Health | Kirsten St. George, Daryl M. Lamson, Alexis Russel, Matthew Shudt, Melissa A Leisner, Jonathan Plitnick, Navjot Singh, John Kelly, Erasmus Schneider, Erica Lasek-Nesselquist |
| EPI_ISL_1227467 | WESTCHESTER MEDICAL CENTER | Wadsworth Center, New York State Department of Health | Kirsten St. George, Daryl M. Lamson, Alexis Russel, Matthew Shudt, Melissa A Leisner, Jonathan Plitnick, Navjot Singh, John Kelly, Erasmus Schneider, Erica Lasek-Nesselquist |
| EPI_ISL_1227468, EPI_ISL_1227469 | MEMORIAL SLOAN KETTERING CANCER CENTER | Wadsworth Center, New York State Department of Health | Kirsten St. George, Daryl M. Lamson, Alexis Russel, Matthew Shudt, Melissa A Leisner, Jonathan Plitnick, Navjot Singh, John Kelly, Erasmus Schneider, Erica Lasek-Nesselquist |
| EPI_ISL_1227470 | WESTCHESTER MEDICAL CENTER | Wadsworth Center, New York State Department of Health | Kirsten St. George, Daryl M. Lamson, Alexis Russel, Matthew Shudt, Melissa A Leisner, Jonathan Plitnick, Navjot Singh, John Kelly, Erasmus Schneider, Erica Lasek-Nesselquist |
| EPI_ISL_1227471 | MEMORIAL SLOAN KETTERING CANCER CENTER | Wadsworth Center, New York State Department of Health | Kirsten St. George, Daryl M. Lamson, Alexis Russel, Matthew Shudt, Melissa A Leisner, Jonathan Plitnick, Navjot Singh, John Kelly, Erasmus Schneider, |

| | | | |
|---|---|---|---|
| | | | Erica Lasek-Nesselquist |
| EPI_ISL_1227472, EPI_ISL_1227473, EPI_ISL_1227474, EPI_ISL_1227475, EPI_ISL_1227476, EPI_ISL_1227477, EPI_ISL_1227478, EPI_ISL_1227479, EPI_ISL_1227480, EPI_ISL_1227481, EPI_ISL_1227482, EPI_ISL_1227483, EPI_ISL_1227484, EPI_ISL_1227485, EPI_ISL_1227486, EPI_ISL_1227487, EPI_ISL_1227488, EPI_ISL_1227489, EPI_ISL_1227490, EPI_ISL_1227491, EPI_ISL_1227492, EPI_ISL_1227493, EPI_ISL_1227494, EPI_ISL_1227495, EPI_ISL_1227496, EPI_ISL_1227497, EPI_ISL_1227498, EPI_ISL_1227499, EPI_ISL_1227500, EPI_ISL_1227501, EPI_ISL_1227502 | | | |
| see above | WESTCHESTER MEDICAL CENTER | Wadsworth Center, New York State Department of Health | Kirsten St. George, Daryl M. Lamson, Alexis Russel, Matthew Shudt, Melissa A Leisner, Jonathan Plitnick, Navjot Singh, John Kelly, Erasmus Schneider, Erica Lasek-Nesselquist |
| EPI_ISL_1227503 | MONTEFIORE MEDICAL CENTER LABORATORIES | Wadsworth Center, New York State Department of Health | Kirsten St. George, Daryl M. Lamson, Alexis Russel, Matthew Shudt, Melissa A Leisner, Jonathan Plitnick, Navjot Singh, John Kelly, Erasmus Schneider, Erica Lasek-Nesselquist |
| EPI_ISL_1227504, EPI_ISL_1227505, EPI_ISL_1227506, EPI_ISL_1227507, EPI_ISL_1227508, EPI_ISL_1227509, EPI_ISL_1227510, EPI_ISL_1227511, EPI_ISL_1227512, EPI_ISL_1227513, EPI_ISL_1227514, EPI_ISL_1227515, EPI_ISL_1227516, EPI_ISL_1227517, EPI_ISL_1227518, EPI_ISL_1227519, EPI_ISL_1227520, EPI_ISL_1227521, EPI_ISL_1227522, EPI_ISL_1227523, EPI_ISL_1227524, EPI_ISL_1227525, EPI_ISL_1227526, EPI_ISL_1227527, EPI_ISL_1227528, EPI_ISL_1227529, EPI_ISL_1227530, EPI_ISL_1227531, EPI_ISL_1227532, EPI_ISL_1227533, EPI_ISL_1227534, EPI_ISL_1227535 | | | |
| see above | WESTCHESTER MEDICAL CENTER | Wadsworth Center, New York State Department of Health | Kirsten St. George, Daryl M. Lamson, Alexis Russel, Matthew Shudt, Melissa A Leisner, Jonathan Plitnick, Navjot Singh, John Kelly, Erasmus Schneider, Erica Lasek-Nesselquist |
| EPI_ISL_1227536 | Columbia University Irving Medical Center | Wadsworth Center, New York State Department of Health | Kirsten St. George, Daryl M. Lamson, Alexis Russel, Matthew Shudt, Melissa A Leisner, Jonathan Plitnick, Navjot Singh, John Kelly, Erasmus Schneider, Erica Lasek-Nesselquist |
| EPI_ISL_1227537, EPI_ISL_1227538, EPI_ISL_1227539, EPI_ISL_1227540 | WESTCHESTER MEDICAL CENTER | Wadsworth Center, New York State Department of Health | Kirsten St. George, Daryl M. Lamson, Alexis Russel, Matthew Shudt, Melissa A Leisner, Jonathan Plitnick, Navjot Singh, John Kelly, Erasmus Schneider, Erica Lasek-Nesselquist |
| EPI_ISL_1227541, EPI_ISL_1227542, EPI_ISL_1227543, EPI_ISL_1227544, EPI_ISL_1227545 | THE MARY IMOGENE BASSETT HOSPITAL | Wadsworth Center, New York State Department of Health | Kirsten St. George, Daryl M. Lamson, Alexis Russel, Matthew Shudt, Melissa A Leisner, Jonathan Plitnick, Navjot Singh, John Kelly, Erasmus Schneider, Erica Lasek-Nesselquist |
| EPI_ISL_1227546, EPI_ISL_1227547, EPI_ISL_1227548, EPI_ISL_1227549, EPI_ISL_1227550, EPI_ISL_1227551, EPI_ISL_1227552, EPI_ISL_1227553, EPI_ISL_1227554 | KALEIDA CENTER FOR LABORATORY MEDICINE | Wadsworth Center, New York State Department of Health | Kirsten St. George, Daryl M. Lamson, Alexis Russel, Matthew Shudt, Melissa A Leisner, Jonathan Plitnick, Navjot Singh, John Kelly, Erasmus Schneider, Erica Lasek-Nesselquist |
| EPI_ISL_1227555 | ADIRONDACK MEDICAL CENTER | Wadsworth Center, New York State Department of Health | Kirsten St. George, Daryl M. Lamson, Alexis Russel, Matthew Shudt, Melissa A Leisner, Jonathan Plitnick, Navjot Singh, John Kelly, Erasmus Schneider, Erica Lasek-Nesselquist |
| EPI_ISL_1227556 | ALBANY MEDICAL CENTER | Wadsworth Center, New York State Department of Health | Kirsten St. George, Daryl M. Lamson, Alexis Russel, Matthew Shudt, Melissa A Leisner, Jonathan Plitnick, Navjot Singh, John Kelly, Erasmus Schneider, Erica Lasek-Nesselquist |
| EPI_ISL_1227557 | Wadsworth Center, New York State Department of Health | Wadsworth Center, New York State Department of Health | Kirsten St. George, Daryl M. Lamson, Alexis Russel, Matthew Shudt, Melissa A Leisner, Jonathan Plitnick, Navjot Singh, John Kelly, Erasmus Schneider, Erica Lasek-Nesselquist |
| EPI_ISL_1227558 | ALBANY MEDICAL CENTER | Wadsworth Center, New York State Department of Health | Kirsten St. George, Daryl M. Lamson, Alexis Russel, Matthew Shudt, Melissa A Leisner, Jonathan Plitnick, Navjot Singh, John Kelly, Erasmus Schneider, Erica Lasek-Nesselquist |
| EPI_ISL_1227559 | Wadsworth Center, New York State Department of Health | Wadsworth Center, New York State Department of Health | Kirsten St. George, Daryl M. Lamson, Alexis Russel, Matthew Shudt, Melissa A Leisner, Jonathan Plitnick, Navjot Singh, John Kelly, Erasmus Schneider, Erica Lasek-Nesselquist |
| EPI_ISL_1227560, EPI_ISL_1227561, EPI_ISL_1227562, EPI_ISL_1227563 | URMC LABS | Wadsworth Center, New York State Department of Health | Kirsten St. George, Daryl M. Lamson, Alexis Russel, Matthew Shudt, Melissa A Leisner, Jonathan Plitnick, Navjot Singh, John Kelly, Erasmus Schneider, Erica Lasek-Nesselquist |
| EPI_ISL_1227564, EPI_ISL_1227565 | ALBANY MEDICAL CENTER | Wadsworth Center, New York State Department of Health | Kirsten St. George, Daryl M. Lamson, Alexis Russel, Matthew Shudt, Melissa A Leisner, Jonathan Plitnick, Navjot Singh, John Kelly, Erasmus Schneider, Erica Lasek-Nesselquist |
| EPI_ISL_1227566, EPI_ISL_1227567, EPI_ISL_1227568 | Wadsworth Center, New York State Department of Health | Wadsworth Center, New York State Department of Health | Kirsten St. George, Daryl M. Lamson, Alexis Russel, Matthew Shudt, Melissa A Leisner, Jonathan Plitnick, Navjot Singh, John Kelly, Erasmus Schneider, Erica Lasek-Nesselquist |
| EPI_ISL_1227569 | WESTCHESTER MEDICAL CENTER | Wadsworth Center, New York State Department of Health | Kirsten St. George, Daryl M. Lamson, Alexis Russel, Matthew Shudt, Melissa A Leisner, Jonathan Plitnick, Navjot Singh, John Kelly, Erasmus Schneider, Erica Lasek-Nesselquist |
| EPI_ISL_1227570, EPI_ISL_1227571 | Wadsworth Center, New York State Department of Health | Wadsworth Center, New York State Department of Health | Kirsten St. George, Daryl M. Lamson, Alexis Russel, Matthew Shudt, Melissa A Leisner, Jonathan Plitnick, Navjot Singh, John Kelly, Erasmus Schneider, Erica Lasek-Nesselquist |
| EPI_ISL_1227572, EPI_ISL_1227573 | ALBANY MEDICAL CENTER | Wadsworth Center, New York State Department of Health | Kirsten St. George, Daryl M. Lamson, Alexis Russel, Matthew Shudt, Melissa A Leisner, Jonathan Plitnick, Navjot Singh, John Kelly, Erasmus Schneider, Erica Lasek-Nesselquist |
| EPI_ISL_1227574, EPI_ISL_1227575, EPI_ISL_1227576, EPI_ISL_1227577, EPI_ISL_1227578, EPI_ISL_1227579, EPI_ISL_1227580, EPI_ISL_1227581, EPI_ISL_1227582, EPI_ISL_1227583, EPI_ISL_1227584, EPI_ISL_1227585, EPI_ISL_1227586, EPI_ISL_1227587, EPI_ISL_1227588, EPI_ISL_1227589, EPI_ISL_1227590, EPI_ISL_1227591, EPI_ISL_1227592, EPI_ISL_1227593, EPI_ISL_1227594, EPI_ISL_1227595, EPI_ISL_1227596, EPI_ISL_1227597, EPI_ISL_1227598, EPI_ISL_1227599, EPI_ISL_1227600, EPI_ISL_1227601, EPI_ISL_1227602, EPI_ISL_1227603, EPI_ISL_1227604, EPI_ISL_1227605, EPI_ISL_1227606, EPI_ISL_1227607, EPI_ISL_1227608, EPI_ISL_1227609, EPI_ISL_1227610, EPI_ISL_1227611, EPI_ISL_1227612, EPI_ISL_1227613, EPI_ISL_1227614, EPI_ISL_1227615, EPI_ISL_1227616 | | | |
| see above | Wadsworth Center, New York State Department of Health | Wadsworth Center, New York State Department of Health | Kirsten St. George, Daryl M. Lamson, Alexis Russel, Matthew Shudt, Melissa A Leisner, Jonathan Plitnick, Navjot Singh, John Kelly, Erasmus Schneider, Erica Lasek-Nesselquist |
| EPI_ISL_1227617, EPI_ISL_1227618, EPI_ISL_1227619, EPI_ISL_1227620, EPI_ISL_1227621, EPI_ISL_1227622, EPI_ISL_1227623, EPI_ISL_1227624, EPI_ISL_1227625, EPI_ISL_1227626, EPI_ISL_1227627, EPI_ISL_1227628, EPI_ISL_1227629, EPI_ISL_1227630, EPI_ISL_1227631, EPI_ISL_1227632, EPI_ISL_1227633, EPI_ISL_1227634, EPI_ISL_1227635, EPI_ISL_1227636, EPI_ISL_1227637, EPI_ISL_1227638, EPI_ISL_1227639, EPI_ISL_1227640 | | | |
| see above | THE MARY IMOGENE BASSETT HOSPITAL | Wadsworth Center, New York State Department of Health | Kirsten St. George, Daryl M. Lamson, Alexis Russel, Matthew Shudt, Melissa A Leisner, Jonathan Plitnick, Navjot Singh, John Kelly, Erasmus Schneider, Erica Lasek-Nesselquist |
| EPI_ISL_1227641, EPI_ISL_1227642 | SUNY UPSTATE MEDICAL UNIVERSITY | Wadsworth Center, New York State Department of Health | Kirsten St. George, Daryl M. Lamson, Alexis Russel, Matthew Shudt, Melissa A Leisner, Jonathan Plitnick, Navjot Singh, John Kelly, Erasmus Schneider, Erica Lasek-Nesselquist |
| EPI_ISL_1227643, EPI_ISL_1227644, EPI_ISL_1227645, EPI_ISL_1227646, EPI_ISL_1227647, EPI_ISL_1227648, EPI_ISL_1227649, EPI_ISL_1227650, EPI_ISL_1227651, EPI_ISL_1227652, EPI_ISL_1227653, EPI_ISL_1227654, EPI_ISL_1227655, EPI_ISL_1227656, EPI_ISL_1227657, EPI_ISL_1227658, EPI_ISL_1227659, EPI_ISL_1227660, EPI_ISL_1227661, EPI_ISL_1227662 | | | |
| see above | MEMORIAL SLOAN KETTERING CANCER CENTER | Wadsworth Center, New York State Department of Health | Kirsten St. George, Daryl M. Lamson, Alexis Russel, Matthew Shudt, Melissa A Leisner, Jonathan Plitnick, Navjot Singh, John Kelly, Erasmus Schneider, Erica Lasek-Nesselquist |
| EPI_ISL_1227663 | NORTH SHORE UNIVERSITY HOSPITAL | Wadsworth Center, New York State Department of Health | Kirsten St. George, Daryl M. Lamson, Alexis Russel, Matthew Shudt, Melissa A Leisner, Jonathan Plitnick, Navjot Singh, John Kelly, Erasmus Schneider, Erica Lasek-Nesselquist |
| EPI_ISL_1227664 | NORTHWELL HEALTH LABORATORIES | Wadsworth Center, New York State Department of Health | Kirsten St. George, Daryl M. Lamson, Alexis Russel, Matthew Shudt, Melissa A Leisner, Jonathan Plitnick, Navjot Singh, John Kelly, Erasmus Schneider, Erica Lasek-Nesselquist |
| EPI_ISL_1227665 | MONTEFIORE MEDICAL CENTER LABORATORIES | Wadsworth Center, New York State Department of Health | Kirsten St. George, Daryl M. Lamson, Alexis Russel, Matthew Shudt, Melissa A Leisner, Jonathan Plitnick, Navjot Singh, John Kelly, Erasmus Schneider, Erica Lasek-Nesselquist |
| EPI_ISL_1227666, EPI_ISL_1227667, EPI_ISL_1227668, EPI_ISL_1227669, EPI_ISL_1227670, EPI_ISL_1227671, EPI_ISL_1227672, EPI_ISL_1227673, EPI_ISL_1227674 | NYC Pandemic Response Lab | Wadsworth Center, New York State Department of Health | Kirsten St. George, Daryl M. Lamson, Alexis Russel, Matthew Shudt, Melissa A Leisner, Jonathan Plitnick, Navjot Singh, John Kelly, Erasmus Schneider, Erica Lasek-Nesselquist |
| EPI_ISL_1227675 | Columbia University Irving Medical Center | Wadsworth Center, New York State Department of Health | Kirsten St. George, Daryl M. Lamson, Alexis Russel, Matthew Shudt, Melissa A Leisner, Jonathan Plitnick, Navjot Singh, John Kelly, Erasmus Schneider, |

| | | | |
|---|---|---|---|
| | | | Erica Lasek-Nesselquist |
| EPI_ISL_1227676 | WESTCHESTER MEDICAL CENTER | Wadsworth Center, New York State Department of Health | Kirsten St. George, Daryl M. Lamson, Alexis Russel, Matthew Shudt, Melissa A Leisner, Jonathan Plitnick, Navjot Singh, John Kelly, Erasmus Schneider, Erica Lasek-Nesselquist |
| EPI_ISL_1227677, EPI_ISL_1227678 | NORTHWELL HEALTH LABORATORIES | Wadsworth Center, New York State Department of Health | Kirsten St. George, Daryl M. Lamson, Alexis Russel, Matthew Shudt, Melissa A Leisner, Jonathan Plitnick, Navjot Singh, John Kelly, Erasmus Schneider, Erica Lasek-Nesselquist |
| EPI_ISL_1227679 | New York Presbyterian Hospital | Wadsworth Center, New York State Department of Health | Kirsten St. George, Daryl M. Lamson, Alexis Russel, Matthew Shudt, Melissa A Leisner, Jonathan Plitnick, Navjot Singh, John Kelly, Erasmus Schneider, Erica Lasek-Nesselquist |
| EPI_ISL_1227680, EPI_ISL_1227681, EPI_ISL_1227682, EPI_ISL_1227683 | NYC Pandemic Response Lab | Wadsworth Center, New York State Department of Health | Kirsten St. George, Daryl M. Lamson, Alexis Russel, Matthew Shudt, Melissa A Leisner, Jonathan Plitnick, Navjot Singh, John Kelly, Erasmus Schneider, Erica Lasek-Nesselquist |
| EPI_ISL_1227684, EPI_ISL_1227685 | New York Presbyterian Hospital | Wadsworth Center, New York State Department of Health | Kirsten St. George, Daryl M. Lamson, Alexis Russel, Matthew Shudt, Melissa A Leisner, Jonathan Plitnick, Navjot Singh, John Kelly, Erasmus Schneider, Erica Lasek-Nesselquist |
| EPI_ISL_1227686, EPI_ISL_1227687, EPI_ISL_1227688, EPI_ISL_1227689, EPI_ISL_1227690, EPI_ISL_1227691, EPI_ISL_1227692, EPI_ISL_1227693, EPI_ISL_1227694, EPI_ISL_1227695, EPI_ISL_1227696, EPI_ISL_1227697, EPI_ISL_1227698, EPI_ISL_1227699, EPI_ISL_1227700, EPI_ISL_1227701, EPI_ISL_1227702, EPI_ISL_1227703, EPI_ISL_1227704 | | | |
| see above | NYC Pandemic Response Lab | Wadsworth Center, New York State Department of Health | Kirsten St. George, Daryl M. Lamson, Alexis Russel, Matthew Shudt, Melissa A Leisner, Jonathan Plitnick, Navjot Singh, John Kelly, Erasmus Schneider, Erica Lasek-Nesselquist |
| EPI_ISL_1227705 | MONTEFIORE MEDICAL CENTER LABORATORIES | Wadsworth Center, New York State Department of Health | Kirsten St. George, Daryl M. Lamson, Alexis Russel, Matthew Shudt, Melissa A Leisner, Jonathan Plitnick, Navjot Singh, John Kelly, Erasmus Schneider, Erica Lasek-Nesselquist |
| EPI_ISL_1227706 | NYC Pandemic Response Lab | Wadsworth Center, New York State Department of Health | Kirsten St. George, Daryl M. Lamson, Alexis Russel, Matthew Shudt, Melissa A Leisner, Jonathan Plitnick, Navjot Singh, John Kelly, Erasmus Schneider, Erica Lasek-Nesselquist |
| EPI_ISL_1227707 | Masonic Care Community of NY | Wadsworth Center, New York State Department of Health | Kirsten St. George, Daryl M. Lamson, Alexis Russel, Matthew Shudt, Melissa A Leisner, Jonathan Plitnick, Navjot Singh, John Kelly, Erasmus Schneider, Erica Lasek-Nesselquist |
| EPI_ISL_1227708 | SUNY UPSTATE MEDICAL UNIVERSITY | Wadsworth Center, New York State Department of Health | Kirsten St. George, Daryl M. Lamson, Alexis Russel, Matthew Shudt, Melissa A Leisner, Jonathan Plitnick, Navjot Singh, John Kelly, Erasmus Schneider, Erica Lasek-Nesselquist |
| EPI_ISL_1227709, EPI_ISL_1227710, EPI_ISL_1227711, EPI_ISL_1227712, EPI_ISL_1227713, EPI_ISL_1227714, EPI_ISL_1227715, EPI_ISL_1227716, EPI_ISL_1227717, EPI_ISL_1227718, EPI_ISL_1227719 | | | |
| see above | URMC LABS | Wadsworth Center, New York State Department of Health | Kirsten St. George, Daryl M. Lamson, Alexis Russel, Matthew Shudt, Melissa A Leisner, Jonathan Plitnick, Navjot Singh, John Kelly, Erasmus Schneider, Erica Lasek-Nesselquist |
| EPI_ISL_1227720, EPI_ISL_1227721, EPI_ISL_1227722, EPI_ISL_1227723, EPI_ISL_1227724 | Masonic Care Community of NY | Wadsworth Center, New York State Department of Health | Kirsten St. George, Daryl M. Lamson, Alexis Russel, Matthew Shudt, Melissa A Leisner, Jonathan Plitnick, Navjot Singh, John Kelly, Erasmus Schneider, Erica Lasek-Nesselquist |
| EPI_ISL_1227725, EPI_ISL_1227726, EPI_ISL_1227727 | SUNY UPSTATE MEDICAL UNIVERSITY | Wadsworth Center, New York State Department of Health | Kirsten St. George, Daryl M. Lamson, Alexis Russel, Matthew Shudt, Melissa A Leisner, Jonathan Plitnick, Navjot Singh, John Kelly, Erasmus Schneider, Erica Lasek-Nesselquist |
| EPI_ISL_1227728, EPI_ISL_1227729, EPI_ISL_1227730, EPI_ISL_1227731, EPI_ISL_1227732, EPI_ISL_1227733, EPI_ISL_1227734, EPI_ISL_1227735, EPI_ISL_1227736, EPI_ISL_1227737, EPI_ISL_1227738, EPI_ISL_1227739, EPI_ISL_1227740, EPI_ISL_1227741, EPI_ISL_1227742, EPI_ISL_1227743, EPI_ISL_1227744, EPI_ISL_1227745, EPI_ISL_1227746, EPI_ISL_1227747, EPI_ISL_1227748, EPI_ISL_1227749, EPI_ISL_1227750, EPI_ISL_1227751, EPI_ISL_1227752, EPI_ISL_1227753, EPI_ISL_1227754, EPI_ISL_1227755, EPI_ISL_1227756, EPI_ISL_1227757, EPI_ISL_1227758, EPI_ISL_1227759, EPI_ISL_1227760, EPI_ISL_1227761, EPI_ISL_1227762, EPI_ISL_1227763, EPI_ISL_1227764, EPI_ISL_1227765, EPI_ISL_1227766, EPI_ISL_1227767, EPI_ISL_1227768, EPI_ISL_1227769, EPI_ISL_1227770, EPI_ISL_1227771, EPI_ISL_1227772, EPI_ISL_1227773, EPI_ISL_1227774, EPI_ISL_1227775, EPI_ISL_1227776, EPI_ISL_1227777, EPI_ISL_1227778, EPI_ISL_1227779, EPI_ISL_1227780, EPI_ISL_1227781, EPI_ISL_1227782, EPI_ISL_1227783, EPI_ISL_1227784, EPI_ISL_1227785, EPI_ISL_1227786, EPI_ISL_1227787, EPI_ISL_1227788, EPI_ISL_1227789 | | | |
| see above | URMC LABS | Wadsworth Center, New York State Department of Health | Kirsten St. George, Daryl M. Lamson, Alexis Russel, Matthew Shudt, Melissa A Leisner, Jonathan Plitnick, Navjot Singh, John Kelly, Erasmus Schneider, Erica Lasek-Nesselquist |
| EPI_ISL_1227790 | ALBANY MEDICAL CENTER | Wadsworth Center, New York State Department of Health | Kirsten St. George, Daryl M. Lamson, Alexis Russel, Matthew Shudt, Melissa A Leisner, Jonathan Plitnick, Navjot Singh, John Kelly, Erasmus Schneider, Erica Lasek-Nesselquist |
| EPI_ISL_1227791, EPI_ISL_1227792, EPI_ISL_1227793, EPI_ISL_1227794, EPI_ISL_1227795, EPI_ISL_1227796, EPI_ISL_1227797, EPI_ISL_1227798, EPI_ISL_1227799, EPI_ISL_1227800, EPI_ISL_1227801, EPI_ISL_1227802, EPI_ISL_1227803 | | | |
| see above | MEMORIAL SLOAN KETTERING CANCER CENTER | Wadsworth Center, New York State Department of Health | Kirsten St. George, Daryl M. Lamson, Alexis Russel, Matthew Shudt, Melissa A Leisner, Jonathan Plitnick, Navjot Singh, John Kelly, Erasmus Schneider, Erica Lasek-Nesselquist |
| EPI_ISL_1227804 | NORTH SHORE UNIVERSITY HOSPITAL | Wadsworth Center, New York State Department of Health | Kirsten St. George, Daryl M. Lamson, Alexis Russel, Matthew Shudt, Melissa A Leisner, Jonathan Plitnick, Navjot Singh, John Kelly, Erasmus Schneider, Erica Lasek-Nesselquist |
| EPI_ISL_1227805, EPI_ISL_1227806, EPI_ISL_1227807, EPI_ISL_1227808, EPI_ISL_1227809, EPI_ISL_1227810, EPI_ISL_1227811, EPI_ISL_1227812 | MEMORIAL SLOAN KETTERING CANCER CENTER | Wadsworth Center, New York State Department of Health | Kirsten St. George, Daryl M. Lamson, Alexis Russel, Matthew Shudt, Melissa A Leisner, Jonathan Plitnick, Navjot Singh, John Kelly, Erasmus Schneider, Erica Lasek-Nesselquist |
| EPI_ISL_1227813 | NORTH SHORE UNIVERSITY HOSPITAL | Wadsworth Center, New York State Department of Health | Kirsten St. George, Daryl M. Lamson, Alexis Russel, Matthew Shudt, Melissa A Leisner, Jonathan Plitnick, Navjot Singh, John Kelly, Erasmus Schneider, Erica Lasek-Nesselquist |
| EPI_ISL_1227814, EPI_ISL_1227815 | MEMORIAL SLOAN KETTERING CANCER CENTER | Wadsworth Center, New York State Department of Health | Kirsten St. George, Daryl M. Lamson, Alexis Russel, Matthew Shudt, Melissa A Leisner, Jonathan Plitnick, Navjot Singh, John Kelly, Erasmus Schneider, Erica Lasek-Nesselquist |
| EPI_ISL_1227816 | NORTH SHORE UNIVERSITY HOSPITAL | Wadsworth Center, New York State Department of Health | Kirsten St. George, Daryl M. Lamson, Alexis Russel, Matthew Shudt, Melissa A Leisner, Jonathan Plitnick, Navjot Singh, John Kelly, Erasmus Schneider, Erica Lasek-Nesselquist |
| EPI_ISL_1227817, EPI_ISL_1227818 | MEMORIAL SLOAN KETTERING CANCER CENTER | Wadsworth Center, New York State Department of Health | Kirsten St. George, Daryl M. Lamson, Alexis Russel, Matthew Shudt, Melissa A Leisner, Jonathan Plitnick, Navjot Singh, John Kelly, Erasmus Schneider, Erica Lasek-Nesselquist |
| EPI_ISL_1227819 | NORTHWELL HEALTH LABORATORIES | Wadsworth Center, New York State Department of Health | Kirsten St. George, Daryl M. Lamson, Alexis Russel, Matthew Shudt, Melissa A Leisner, Jonathan Plitnick, Navjot Singh, John Kelly, Erasmus Schneider, Erica Lasek-Nesselquist |
| EPI_ISL_1227820 | MEMORIAL SLOAN KETTERING CANCER CENTER | Wadsworth Center, New York State Department of Health | Kirsten St. George, Daryl M. Lamson, Alexis Russel, Matthew Shudt, Melissa A Leisner, Jonathan Plitnick, Navjot Singh, John Kelly, Erasmus Schneider, Erica Lasek-Nesselquist |
| EPI_ISL_1227821, EPI_ISL_1227822, EPI_ISL_1227823, EPI_ISL_1227824, EPI_ISL_1227825, EPI_ISL_1227826 | NORTHWELL HEALTH LABORATORIES | Wadsworth Center, New York State Department of Health | Kirsten St. George, Daryl M. Lamson, Alexis Russel, Matthew Shudt, Melissa A Leisner, Jonathan Plitnick, Navjot Singh, John Kelly, Erasmus Schneider, Erica Lasek-Nesselquist |
| EPI_ISL_1227827, EPI_ISL_1227828, EPI_ISL_1227829 | MEMORIAL SLOAN KETTERING CANCER CENTER | Wadsworth Center, New York State Department of Health | Kirsten St. George, Daryl M. Lamson, Alexis Russel, Matthew Shudt, Melissa A Leisner, Jonathan Plitnick, Navjot Singh, John Kelly, Erasmus Schneider, Erica Lasek-Nesselquist |
| EPI_ISL_1227830 | NORTH SHORE UNIVERSITY HOSPITAL | Wadsworth Center, New York State Department of Health | Kirsten St. George, Daryl M. Lamson, Alexis Russel, Matthew Shudt, Melissa A Leisner, Jonathan Plitnick, Navjot Singh, John Kelly, Erasmus Schneider, Erica Lasek-Nesselquist |
| EPI_ISL_1227831, EPI_ISL_1227832, EPI_ISL_1227833, EPI_ISL_1227834, EPI_ISL_1227835, EPI_ISL_1227836 | NORTHWELL HEALTH LABORATORIES | Wadsworth Center, New York State Department of Health | Kirsten St. George, Daryl M. Lamson, Alexis Russel, Matthew Shudt, Melissa A Leisner, Jonathan Plitnick, Navjot Singh, John Kelly, Erasmus Schneider, Erica Lasek-Nesselquist |
| EPI_ISL_1227837, EPI_ISL_1227838 | MEMORIAL SLOAN KETTERING CANCER CENTER | Wadsworth Center, New York State Department of Health | Kirsten St. George, Daryl M. Lamson, Alexis Russel, Matthew Shudt, Melissa A Leisner, Jonathan Plitnick, Navjot Singh, John Kelly, Erasmus Schneider, |

| | | | |
|---|---|---|---|
| | | | Erica Lasek-Nesselquist |
| EPI_ISL_1227839, EPI_ISL_1227840, EPI_ISL_1227841, EPI_ISL_1227842 | NORTHWELL HEALTH LABORATORIES | Wadsworth Center, New York State Department of Health | Kirsten St. George, Daryl M. Lamson, Alexis Russel, Matthew Shudt, Melissa A Leisner, Jonathan Plitnick, Navjot Singh, John Kelly, Erasmus Schneider, Erica Lasek-Nesselquist |
| EPI_ISL_1227843, EPI_ISL_1227844, EPI_ISL_1227845, EPI_ISL_1227846, EPI_ISL_1227847 | MEMORIAL SLOAN KETTERING CANCER CENTER | Wadsworth Center, New York State Department of Health | Kirsten St. George, Daryl M. Lamson, Alexis Russel, Matthew Shudt, Melissa A Leisner, Jonathan Plitnick, Navjot Singh, John Kelly, Erasmus Schneider, Erica Lasek-Nesselquist |
| EPI_ISL_1227848, EPI_ISL_1227849, EPI_ISL_1227850 | NORTHWELL HEALTH LABORATORIES | Wadsworth Center, New York State Department of Health | Kirsten St. George, Daryl M. Lamson, Alexis Russel, Matthew Shudt, Melissa A Leisner, Jonathan Plitnick, Navjot Singh, John Kelly, Erasmus Schneider, Erica Lasek-Nesselquist |
| EPI_ISL_1227851 | MEMORIAL SLOAN KETTERING CANCER CENTER | Wadsworth Center, New York State Department of Health | Kirsten St. George, Daryl M. Lamson, Alexis Russel, Matthew Shudt, Melissa A Leisner, Jonathan Plitnick, Navjot Singh, John Kelly, Erasmus Schneider, Erica Lasek-Nesselquist |
| EPI_ISL_1227852 | NORTH SHORE UNIVERSITY HOSPITAL | Wadsworth Center, New York State Department of Health | Kirsten St. George, Daryl M. Lamson, Alexis Russel, Matthew Shudt, Melissa A Leisner, Jonathan Plitnick, Navjot Singh, John Kelly, Erasmus Schneider, Erica Lasek-Nesselquist |
| EPI_ISL_1227853, EPI_ISL_1227854, EPI_ISL_1227855, EPI_ISL_1227856, EPI_ISL_1227857, EPI_ISL_1227858, EPI_ISL_1227859, EPI_ISL_1227860 | NORTHWELL HEALTH LABORATORIES | Wadsworth Center, New York State Department of Health | Kirsten St. George, Daryl M. Lamson, Alexis Russel, Matthew Shudt, Melissa A Leisner, Jonathan Plitnick, Navjot Singh, John Kelly, Erasmus Schneider, Erica Lasek-Nesselquist |
| EPI_ISL_1227861, EPI_ISL_1227862, EPI_ISL_1227863, EPI_ISL_1227864, EPI_ISL_1227865, EPI_ISL_1227866 | NYC Pandemic Response Lab | Wadsworth Center, New York State Department of Health | Kirsten St. George, Daryl M. Lamson, Alexis Russel, Matthew Shudt, Melissa A Leisner, Jonathan Plitnick, Navjot Singh, John Kelly, Erasmus Schneider, Erica Lasek-Nesselquist |
| EPI_ISL_1227867 | New York Presbyterian Hospital | Wadsworth Center, New York State Department of Health | Kirsten St. George, Daryl M. Lamson, Alexis Russel, Matthew Shudt, Melissa A Leisner, Jonathan Plitnick, Navjot Singh, John Kelly, Erasmus Schneider, Erica Lasek-Nesselquist |
| EPI_ISL_1227868, EPI_ISL_1227869 | NYC Pandemic Response Lab | Wadsworth Center, New York State Department of Health | Kirsten St. George, Daryl M. Lamson, Alexis Russel, Matthew Shudt, Melissa A Leisner, Jonathan Plitnick, Navjot Singh, John Kelly, Erasmus Schneider, Erica Lasek-Nesselquist |
| EPI_ISL_1227870 | NORTH SHORE UNIVERSITY HOSPITAL | Wadsworth Center, New York State Department of Health | Kirsten St. George, Daryl M. Lamson, Alexis Russel, Matthew Shudt, Melissa A Leisner, Jonathan Plitnick, Navjot Singh, John Kelly, Erasmus Schneider, Erica Lasek-Nesselquist |
| EPI_ISL_1227871, EPI_ISL_1227872, EPI_ISL_1227873, EPI_ISL_1227874, EPI_ISL_1227875, EPI_ISL_1227876, EPI_ISL_1227877, EPI_ISL_1227878, EPI_ISL_1227879 | NYC Pandemic Response Lab | Wadsworth Center, New York State Department of Health | Kirsten St. George, Daryl M. Lamson, Alexis Russel, Matthew Shudt, Melissa A Leisner, Jonathan Plitnick, Navjot Singh, John Kelly, Erasmus Schneider, Erica Lasek-Nesselquist |
| EPI_ISL_1227880, EPI_ISL_1227881, EPI_ISL_1227882, EPI_ISL_1227883, EPI_ISL_1227884, EPI_ISL_1227885, EPI_ISL_1227886, EPI_ISL_1227887, EPI_ISL_1227888, EPI_ISL_1227889, EPI_ISL_1227890, EPI_ISL_1227891, EPI_ISL_1227892, EPI_ISL_1227893, EPI_ISL_1227894 | | | |
| see above | MEMORIAL SLOAN KETTERING CANCER CENTER | Wadsworth Center, New York State Department of Health | Kirsten St. George, Daryl M. Lamson, Alexis Russel, Matthew Shudt, Melissa A Leisner, Jonathan Plitnick, Navjot Singh, John Kelly, Erasmus Schneider, Erica Lasek-Nesselquist |
| EPI_ISL_1227895, EPI_ISL_1227896, EPI_ISL_1227897, EPI_ISL_1227898, EPI_ISL_1227899, EPI_ISL_1227900, EPI_ISL_1227901, EPI_ISL_1227902 | Columbia University Irving Medical Center | Wadsworth Center, New York State Department of Health | Kirsten St. George, Daryl M. Lamson, Alexis Russel, Matthew Shudt, Melissa A Leisner, Jonathan Plitnick, Navjot Singh, John Kelly, Erasmus Schneider, Erica Lasek-Nesselquist |
| EPI_ISL_1227903 | NORTHWELL HEALTH LABORATORIES | Wadsworth Center, New York State Department of Health | Kirsten St. George, Daryl M. Lamson, Alexis Russel, Matthew Shudt, Melissa A Leisner, Jonathan Plitnick, Navjot Singh, John Kelly, Erasmus Schneider, Erica Lasek-Nesselquist |
| EPI_ISL_1227904, EPI_ISL_1227905, EPI_ISL_1227906, EPI_ISL_1227907, EPI_ISL_1227908 | Columbia University Irving Medical Center | Wadsworth Center, New York State Department of Health | Kirsten St. George, Daryl M. Lamson, Alexis Russel, Matthew Shudt, Melissa A Leisner, Jonathan Plitnick, Navjot Singh, John Kelly, Erasmus Schneider, Erica Lasek-Nesselquist |
| EPI_ISL_1227909 | MONTEFIORE MEDICAL CENTER LABORATORIES | Wadsworth Center, New York State Department of Health | Kirsten St. George, Daryl M. Lamson, Alexis Russel, Matthew Shudt, Melissa A Leisner, Jonathan Plitnick, Navjot Singh, John Kelly, Erasmus Schneider, Erica Lasek-Nesselquist |
| EPI_ISL_1227910 | Columbia University Irving Medical Center | Wadsworth Center, New York State Department of Health | Kirsten St. George, Daryl M. Lamson, Alexis Russel, Matthew Shudt, Melissa A Leisner, Jonathan Plitnick, Navjot Singh, John Kelly, Erasmus Schneider, Erica Lasek-Nesselquist |
| EPI_ISL_1227911, EPI_ISL_1227912 | MONTEFIORE MEDICAL CENTER LABORATORIES | Wadsworth Center, New York State Department of Health | Kirsten St. George, Daryl M. Lamson, Alexis Russel, Matthew Shudt, Melissa A Leisner, Jonathan Plitnick, Navjot Singh, John Kelly, Erasmus Schneider, Erica Lasek-Nesselquist |
| EPI_ISL_1227913, EPI_ISL_1227914, EPI_ISL_1227915, EPI_ISL_1227916, EPI_ISL_1227917, EPI_ISL_1227918, EPI_ISL_1227919 | Columbia University Irving Medical Center | Wadsworth Center, New York State Department of Health | Kirsten St. George, Daryl M. Lamson, Alexis Russel, Matthew Shudt, Melissa A Leisner, Jonathan Plitnick, Navjot Singh, John Kelly, Erasmus Schneider, Erica Lasek-Nesselquist |
| EPI_ISL_1227920 | NYC Pandemic Response Lab | Wadsworth Center, New York State Department of Health | Kirsten St. George, Daryl M. Lamson, Alexis Russel, Matthew Shudt, Melissa A Leisner, Jonathan Plitnick, Navjot Singh, John Kelly, Erasmus Schneider, Erica Lasek-Nesselquist |
| EPI_ISL_1227921, EPI_ISL_1227922, EPI_ISL_1227923, EPI_ISL_1227924, EPI_ISL_1227925, EPI_ISL_1227926, EPI_ISL_1227927, EPI_ISL_1227928, EPI_ISL_1227929, EPI_ISL_1227930, EPI_ISL_1227931 | | | |
| see above | Columbia University Irving Medical Center | Wadsworth Center, New York State Department of Health | Kirsten St. George, Daryl M. Lamson, Alexis Russel, Matthew Shudt, Melissa A Leisner, Jonathan Plitnick, Navjot Singh, John Kelly, Erasmus Schneider, Erica Lasek-Nesselquist |
| EPI_ISL_1227932, EPI_ISL_1227933 | MONTEFIORE MEDICAL CENTER LABORATORIES | Wadsworth Center, New York State Department of Health | Kirsten St. George, Daryl M. Lamson, Alexis Russel, Matthew Shudt, Melissa A Leisner, Jonathan Plitnick, Navjot Singh, John Kelly, Erasmus Schneider, Erica Lasek-Nesselquist |
| EPI_ISL_1227934 | Columbia University Irving Medical Center | Wadsworth Center, New York State Department of Health | Kirsten St. George, Daryl M. Lamson, Alexis Russel, Matthew Shudt, Melissa A Leisner, Jonathan Plitnick, Navjot Singh, John Kelly, Erasmus Schneider, Erica Lasek-Nesselquist |
| EPI_ISL_1227935, EPI_ISL_1227936, EPI_ISL_1227937 | New York Presbyterian Hospital | Wadsworth Center, New York State Department of Health | Kirsten St. George, Daryl M. Lamson, Alexis Russel, Matthew Shudt, Melissa A Leisner, Jonathan Plitnick, Navjot Singh, John Kelly, Erasmus Schneider, Erica Lasek-Nesselquist |
| EPI_ISL_1227938, EPI_ISL_1227939, EPI_ISL_1227940, EPI_ISL_1227941, EPI_ISL_1227942 | NYC Pandemic Response Lab | Wadsworth Center, New York State Department of Health | Kirsten St. George, Daryl M. Lamson, Alexis Russel, Matthew Shudt, Melissa A Leisner, Jonathan Plitnick, Navjot Singh, John Kelly, Erasmus Schneider, Erica Lasek-Nesselquist |
| EPI_ISL_1227943, EPI_ISL_1227944, EPI_ISL_1227945 | New York Presbyterian Hospital | Wadsworth Center, New York State Department of Health | Kirsten St. George, Daryl M. Lamson, Alexis Russel, Matthew Shudt, Melissa A Leisner, Jonathan Plitnick, Navjot Singh, John Kelly, Erasmus Schneider, Erica Lasek-Nesselquist |
| EPI_ISL_1227946, EPI_ISL_1227947, EPI_ISL_1227948, EPI_ISL_1227949, EPI_ISL_1227950, EPI_ISL_1227951, EPI_ISL_1227952, EPI_ISL_1227953, EPI_ISL_1227954, EPI_ISL_1227955, EPI_ISL_1227956, EPI_ISL_1227957, EPI_ISL_1227958 | | | |
| see above | NYC Pandemic Response Lab | Wadsworth Center, New York State Department of Health | Kirsten St. George, Daryl M. Lamson, Alexis Russel, Matthew Shudt, Melissa A Leisner, Jonathan Plitnick, Navjot Singh, John Kelly, Erasmus Schneider, Erica Lasek-Nesselquist |
| EPI_ISL_1227959 | MONTEFIORE MEDICAL CENTER LABORATORIES | Wadsworth Center, New York State Department of Health | Kirsten St. George, Daryl M. Lamson, Alexis Russel, Matthew Shudt, Melissa A Leisner, Jonathan Plitnick, Navjot Singh, John Kelly, Erasmus Schneider, |

| | | | |
|---|---|---|---|
| | | | Erica Lasek-Nesselquist |
| EPI_ISL_1227960 | Wadsworth Center, New York State Department of Health | Wadsworth Center, New York State Department of Health | Kirsten St. George, Daryl M. Lamson, Alexis Russel, Matthew Shudt, Melissa A Leisner, Jonathan Plitnick, Navjot Singh, John Kelly, Erasmus Schneider, Erica Lasek-Nesselquist |
| EPI_ISL_1227961 | Masonic Care Community of NY | Wadsworth Center, New York State Department of Health | Kirsten St. George, Daryl M. Lamson, Alexis Russel, Matthew Shudt, Melissa A Leisner, Jonathan Plitnick, Navjot Singh, John Kelly, Erasmus Schneider, Erica Lasek-Nesselquist |
| EPI_ISL_1227962, EPI_ISL_1227963, EPI_ISL_1227964 | THE MARY IMOGENE BASSETT HOSPITAL | Wadsworth Center, New York State Department of Health | Kirsten St. George, Daryl M. Lamson, Alexis Russel, Matthew Shudt, Melissa A Leisner, Jonathan Plitnick, Navjot Singh, John Kelly, Erasmus Schneider, Erica Lasek-Nesselquist |
| EPI_ISL_1227965, EPI_ISL_1227966, EPI_ISL_1227967, EPI_ISL_1227968, EPI_ISL_1227969, EPI_ISL_1227970, EPI_ISL_1227971, EPI_ISL_1227972 | KALEIDA CENTER FOR LABORATORY MEDICINE | Wadsworth Center, New York State Department of Health | Kirsten St. George, Daryl M. Lamson, Alexis Russel, Matthew Shudt, Melissa A Leisner, Jonathan Plitnick, Navjot Singh, John Kelly, Erasmus Schneider, Erica Lasek-Nesselquist |
| EPI_ISL_1227973 | URMC LABS | Wadsworth Center, New York State Department of Health | Kirsten St. George, Daryl M. Lamson, Alexis Russel, Matthew Shudt, Melissa A Leisner, Jonathan Plitnick, Navjot Singh, John Kelly, Erasmus Schneider, Erica Lasek-Nesselquist |
| EPI_ISL_1227974, EPI_ISL_1227975 | THE MARY IMOGENE BASSETT HOSPITAL | Wadsworth Center, New York State Department of Health | Kirsten St. George, Daryl M. Lamson, Alexis Russel, Matthew Shudt, Melissa A Leisner, Jonathan Plitnick, Navjot Singh, John Kelly, Erasmus Schneider, Erica Lasek-Nesselquist |
| EPI_ISL_1227976, EPI_ISL_1227977, EPI_ISL_1227978, EPI_ISL_1227979, EPI_ISL_1227980, EPI_ISL_1227981, EPI_ISL_1227982, EPI_ISL_1227983, EPI_ISL_1227984, EPI_ISL_1227985, EPI_ISL_1227986, EPI_ISL_1227987, EPI_ISL_1227988, EPI_ISL_1227989, EPI_ISL_1227990 | | | |
| see above | Masonic Care Community of NY | Wadsworth Center, New York State Department of Health | Kirsten St. George, Daryl M. Lamson, Alexis Russel, Matthew Shudt, Melissa A Leisner, Jonathan Plitnick, Navjot Singh, John Kelly, Erasmus Schneider, Erica Lasek-Nesselquist |
| EPI_ISL_1227991, EPI_ISL_1227992, EPI_ISL_1227993, EPI_ISL_1227994, EPI_ISL_1227995, EPI_ISL_1227996, EPI_ISL_1227997, EPI_ISL_1227998, EPI_ISL_1227999, EPI_ISL_1228000, EPI_ISL_1228001, EPI_ISL_1228002, EPI_ISL_1228003, EPI_ISL_1228004, EPI_ISL_1228005, EPI_ISL_1228006, EPI_ISL_1228007, EPI_ISL_1228008, EPI_ISL_1228009, EPI_ISL_1228010, EPI_ISL_1228011, EPI_ISL_1228012, EPI_ISL_1228013, EPI_ISL_1228014, EPI_ISL_1228015, EPI_ISL_1228016, EPI_ISL_1228017, EPI_ISL_1228018, EPI_ISL_1228019, EPI_ISL_1228020, EPI_ISL_1228021, EPI_ISL_1228022 | | | |
| see above | SUNY UPSTATE MEDICAL UNIVERSITY | Wadsworth Center, New York State Department of Health | Kirsten St. George, Daryl M. Lamson, Alexis Russel, Matthew Shudt, Melissa A Leisner, Jonathan Plitnick, Navjot Singh, John Kelly, Erasmus Schneider, Erica Lasek-Nesselquist |
| EPI_ISL_1228023, EPI_ISL_1228024, EPI_ISL_1228025, EPI_ISL_1228026, EPI_ISL_1228027, EPI_ISL_1228028, EPI_ISL_1228029, EPI_ISL_1228030, EPI_ISL_1228031 | URMC LABS | Wadsworth Center, New York State Department of Health | Kirsten St. George, Daryl M. Lamson, Alexis Russel, Matthew Shudt, Melissa A Leisner, Jonathan Plitnick, Navjot Singh, John Kelly, Erasmus Schneider, Erica Lasek-Nesselquist |
| EPI_ISL_1228032, EPI_ISL_1228033, EPI_ISL_1228034, EPI_ISL_1228035 | MEMORIAL SLOAN KETTERING CANCER CENTER | Wadsworth Center, New York State Department of Health | Kirsten St. George, Daryl M. Lamson, Alexis Russel, Matthew Shudt, Melissa A Leisner, Jonathan Plitnick, Navjot Singh, John Kelly, Erasmus Schneider, Erica Lasek-Nesselquist |
| EPI_ISL_1228036, EPI_ISL_1228037, EPI_ISL_1228038, EPI_ISL_1228039 | WESTCHESTER MEDICAL CENTER | Wadsworth Center, New York State Department of Health | Kirsten St. George, Daryl M. Lamson, Alexis Russel, Matthew Shudt, Melissa A Leisner, Jonathan Plitnick, Navjot Singh, John Kelly, Erasmus Schneider, Erica Lasek-Nesselquist |
| EPI_ISL_1228040, EPI_ISL_1228041 | URMC LABS | Wadsworth Center, New York State Department of Health | Kirsten St. George, Daryl M. Lamson, Alexis Russel, Matthew Shudt, Melissa A Leisner, Jonathan Plitnick, Navjot Singh, John Kelly, Erasmus Schneider, Erica Lasek-Nesselquist |
| EPI_ISL_1228042 | KALEIDA CENTER FOR LABORATORY MEDICINE | Wadsworth Center, New York State Department of Health | Kirsten St. George, Daryl M. Lamson, Alexis Russel, Matthew Shudt, Melissa A Leisner, Jonathan Plitnick, Navjot Singh, John Kelly, Erasmus Schneider, Erica Lasek-Nesselquist |
| EPI_ISL_1228043, EPI_ISL_1228044, EPI_ISL_1228045 | URMC LABS | Wadsworth Center, New York State Department of Health | Kirsten St. George, Daryl M. Lamson, Alexis Russel, Matthew Shudt, Melissa A Leisner, Jonathan Plitnick, Navjot Singh, John Kelly, Erasmus Schneider, Erica Lasek-Nesselquist |
| EPI_ISL_1228046 | Masonic Care Community of NY | Wadsworth Center, New York State Department of Health | Kirsten St. George, Daryl M. Lamson, Alexis Russel, Matthew Shudt, Melissa A Leisner, Jonathan Plitnick, Navjot Singh, John Kelly, Erasmus Schneider, Erica Lasek-Nesselquist |
| EPI_ISL_1228047, EPI_ISL_1228048 | SUNY UPSTATE MEDICAL UNIVERSITY | Wadsworth Center, New York State Department of Health | Kirsten St. George, Daryl M. Lamson, Alexis Russel, Matthew Shudt, Melissa A Leisner, Jonathan Plitnick, Navjot Singh, John Kelly, Erasmus Schneider, Erica Lasek-Nesselquist |
| EPI_ISL_1228049, EPI_ISL_1228050, EPI_ISL_1228051 | THE MARY IMOGENE BASSETT HOSPITAL | Wadsworth Center, New York State Department of Health | Kirsten St. George, Daryl M. Lamson, Alexis Russel, Matthew Shudt, Melissa A Leisner, Jonathan Plitnick, Navjot Singh, John Kelly, Erasmus Schneider, Erica Lasek-Nesselquist |
| EPI_ISL_1228052, EPI_ISL_1228053, EPI_ISL_1228054, EPI_ISL_1228055, EPI_ISL_1228056, EPI_ISL_1228057, EPI_ISL_1228058, EPI_ISL_1228059, EPI_ISL_1228060, EPI_ISL_1228061, EPI_ISL_1228062, EPI_ISL_1228063, EPI_ISL_1228064, EPI_ISL_1228065, EPI_ISL_1228066, EPI_ISL_1228067, EPI_ISL_1228068, EPI_ISL_1228069, EPI_ISL_1228070, EPI_ISL_1228071, EPI_ISL_1228072, EPI_ISL_1228073, EPI_ISL_1228074, EPI_ISL_1228075, EPI_ISL_1228076, EPI_ISL_1228077, EPI_ISL_1228078, EPI_ISL_1228079, EPI_ISL_1228080, EPI_ISL_1228081, EPI_ISL_1228082, EPI_ISL_1228083, EPI_ISL_1228084, EPI_ISL_1228085, EPI_ISL_1228086, EPI_ISL_1228087, EPI_ISL_1228088, EPI_ISL_1228089, EPI_ISL_1228090, EPI_ISL_1228091, EPI_ISL_1228092, EPI_ISL_1228093, EPI_ISL_1228094, EPI_ISL_1228095, EPI_ISL_1228096, EPI_ISL_1228097, EPI_ISL_1228098, EPI_ISL_1228099, EPI_ISL_1228100, EPI_ISL_1228101, EPI_ISL_1228102, EPI_ISL_1228103, EPI_ISL_1228104, EPI_ISL_1228105, EPI_ISL_1228106, EPI_ISL_1228107, EPI_ISL_1228108, EPI_ISL_1228109, EPI_ISL_1228110, EPI_ISL_1228111, EPI_ISL_1228112, EPI_ISL_1228113 | | | |
| see above | MEMORIAL SLOAN KETTERING CANCER CENTER | Wadsworth Center, New York State Department of Health | Kirsten St. George, Daryl M. Lamson, Alexis Russel, Matthew Shudt, Melissa A Leisner, Jonathan Plitnick, Navjot Singh, John Kelly, Erasmus Schneider, Erica Lasek-Nesselquist |
| EPI_ISL_1228114 | MONTEFIORE MEDICAL CENTER LABORATORIES | Wadsworth Center, New York State Department of Health | Kirsten St. George, Daryl M. Lamson, Alexis Russel, Matthew Shudt, Melissa A Leisner, Jonathan Plitnick, Navjot Singh, John Kelly, Erasmus Schneider, Erica Lasek-Nesselquist |
| EPI_ISL_1228115, EPI_ISL_1228116 | URMC LABS | Wadsworth Center, New York State Department of Health | Kirsten St. George, Daryl M. Lamson, Alexis Russel, Matthew Shudt, Melissa A Leisner, Jonathan Plitnick, Navjot Singh, John Kelly, Erasmus Schneider, Erica Lasek-Nesselquist |
| EPI_ISL_1228117, EPI_ISL_1228118 | MEMORIAL SLOAN KETTERING CANCER CENTER | Wadsworth Center, New York State Department of Health | Kirsten St. George, Daryl M. Lamson, Alexis Russel, Matthew Shudt, Melissa A Leisner, Jonathan Plitnick, Navjot Singh, John Kelly, Erasmus Schneider, Erica Lasek-Nesselquist |
| EPI_ISL_1228119, EPI_ISL_1228120, EPI_ISL_1228121 | WESTCHESTER MEDICAL CENTER | Wadsworth Center, New York State Department of Health | Kirsten St. George, Daryl M. Lamson, Alexis Russel, Matthew Shudt, Melissa A Leisner, Jonathan Plitnick, Navjot Singh, John Kelly, Erasmus Schneider, Erica Lasek-Nesselquist |
| EPI_ISL_1228122, EPI_ISL_1228123, EPI_ISL_1228124, EPI_ISL_1228125, EPI_ISL_1228126, EPI_ISL_1228127, EPI_ISL_1228128, EPI_ISL_1228129 | MEMORIAL SLOAN KETTERING CANCER CENTER | Wadsworth Center, New York State Department of Health | Kirsten St. George, Daryl M. Lamson, Alexis Russel, Matthew Shudt, Melissa A Leisner, Jonathan Plitnick, Navjot Singh, John Kelly, Erasmus Schneider, Erica Lasek-Nesselquist |
| EPI_ISL_1228130 | NORTH SHORE UNIVERSITY HOSPITAL | Wadsworth Center, New York State Department of Health | Kirsten St. George, Daryl M. Lamson, Alexis Russel, Matthew Shudt, Melissa A Leisner, Jonathan Plitnick, Navjot Singh, John Kelly, Erasmus Schneider, Erica Lasek-Nesselquist |
| EPI_ISL_1228131, EPI_ISL_1228132, EPI_ISL_1228133, EPI_ISL_1228134 | MEMORIAL SLOAN KETTERING CANCER CENTER | Wadsworth Center, New York State Department of Health | Kirsten St. George, Daryl M. Lamson, Alexis Russel, Matthew Shudt, Melissa A Leisner, Jonathan Plitnick, Navjot Singh, John Kelly, Erasmus Schneider, Erica Lasek-Nesselquist |
| EPI_ISL_1228135 | NORTH SHORE UNIVERSITY HOSPITAL | Wadsworth Center, New York State Department of Health | Kirsten St. George, Daryl M. Lamson, Alexis Russel, Matthew Shudt, Melissa A Leisner, Jonathan Plitnick, Navjot Singh, John Kelly, Erasmus Schneider, Erica Lasek-Nesselquist |
| EPI_ISL_1228136, EPI_ISL_1228137, EPI_ISL_1228138 | MEMORIAL SLOAN KETTERING CANCER CENTER | Wadsworth Center, New York State Department of Health | Kirsten St. George, Daryl M. Lamson, Alexis Russel, Matthew Shudt, Melissa A Leisner, Jonathan Plitnick, Navjot Singh, John Kelly, Erasmus Schneider, Erica Lasek-Nesselquist |
| EPI_ISL_1228139, EPI_ISL_1228140 | NORTH SHORE UNIVERSITY HOSPITAL | Wadsworth Center, New York State Department of Health | Kirsten St. George, Daryl M. Lamson, Alexis Russel, Matthew Shudt, Melissa A Leisner, Jonathan Plitnick, Navjot Singh, John Kelly, Erasmus Schneider, Erica Lasek-Nesselquist |
| EPI_ISL_1228141, EPI_ISL_1228142 | MEMORIAL SLOAN KETTERING CANCER CENTER | Wadsworth Center, New York State Department of Health | Kirsten St. George, Daryl M. Lamson, Alexis Russel, Matthew Shudt, Melissa A Leisner, Jonathan Plitnick, Navjot Singh, John Kelly, Erasmus Schneider, |

| | | | |
|---|---|---|---|
| | | | Erica Lasek-Nesselquist |
| EPI_ISL_1228143 | NYC Pandemic Response Lab | Wadsworth Center, New York State Department of Health | Kirsten St. George, Daryl M. Lamson, Alexis Russel, Matthew Shudt, Melissa A Leisner, Jonathan Plitnick, Navjot Singh, John Kelly, Erasmus Schneider, Erica Lasek-Nesselquist |
| EPI_ISL_1228144, EPI_ISL_1228145, EPI_ISL_1228146 | MEMORIAL SLOAN KETTERING CANCER CENTER | Wadsworth Center, New York State Department of Health | Kirsten St. George, Daryl M. Lamson, Alexis Russel, Matthew Shudt, Melissa A Leisner, Jonathan Plitnick, Navjot Singh, John Kelly, Erasmus Schneider, Erica Lasek-Nesselquist |
| EPI_ISL_1228147, EPI_ISL_1228148, EPI_ISL_1228149 | NORTH SHORE UNIVERSITY HOSPITAL | Wadsworth Center, New York State Department of Health | Kirsten St. George, Daryl M. Lamson, Alexis Russel, Matthew Shudt, Melissa A Leisner, Jonathan Plitnick, Navjot Singh, John Kelly, Erasmus Schneider, Erica Lasek-Nesselquist |
| EPI_ISL_1228150, EPI_ISL_1228151, EPI_ISL_1228152, EPI_ISL_1228153 | NORTHWELL HEALTH LABORATORIES | Wadsworth Center, New York State Department of Health | Kirsten St. George, Daryl M. Lamson, Alexis Russel, Matthew Shudt, Melissa A Leisner, Jonathan Plitnick, Navjot Singh, John Kelly, Erasmus Schneider, Erica Lasek-Nesselquist |
| EPI_ISL_1228154 | WESTCHESTER MEDICAL CENTER | Wadsworth Center, New York State Department of Health | Kirsten St. George, Daryl M. Lamson, Alexis Russel, Matthew Shudt, Melissa A Leisner, Jonathan Plitnick, Navjot Singh, John Kelly, Erasmus Schneider, Erica Lasek-Nesselquist |
| EPI_ISL_1228155, EPI_ISL_1228156, EPI_ISL_1228157 | NORTHWELL HEALTH LABORATORIES | Wadsworth Center, New York State Department of Health | Kirsten St. George, Daryl M. Lamson, Alexis Russel, Matthew Shudt, Melissa A Leisner, Jonathan Plitnick, Navjot Singh, John Kelly, Erasmus Schneider, Erica Lasek-Nesselquist |
| EPI_ISL_1228158 | Columbia University Irving Medical Center | Wadsworth Center, New York State Department of Health | Kirsten St. George, Daryl M. Lamson, Alexis Russel, Matthew Shudt, Melissa A Leisner, Jonathan Plitnick, Navjot Singh, John Kelly, Erasmus Schneider, Erica Lasek-Nesselquist |
| EPI_ISL_1228159, EPI_ISL_1228160, EPI_ISL_1228161 | NORTH SHORE UNIVERSITY HOSPITAL | Wadsworth Center, New York State Department of Health | Kirsten St. George, Daryl M. Lamson, Alexis Russel, Matthew Shudt, Melissa A Leisner, Jonathan Plitnick, Navjot Singh, John Kelly, Erasmus Schneider, Erica Lasek-Nesselquist |
| EPI_ISL_1228162, EPI_ISL_1228163, EPI_ISL_1228164, EPI_ISL_1228165, EPI_ISL_1228166, EPI_ISL_1228167, EPI_ISL_1228168, EPI_ISL_1228169 | NYC Pandemic Response Lab | Wadsworth Center, New York State Department of Health | Kirsten St. George, Daryl M. Lamson, Alexis Russel, Matthew Shudt, Melissa A Leisner, Jonathan Plitnick, Navjot Singh, John Kelly, Erasmus Schneider, Erica Lasek-Nesselquist |
| EPI_ISL_1228170 | NORTHWELL HEALTH LABORATORIES | Wadsworth Center, New York State Department of Health | Kirsten St. George, Daryl M. Lamson, Alexis Russel, Matthew Shudt, Melissa A Leisner, Jonathan Plitnick, Navjot Singh, John Kelly, Erasmus Schneider, Erica Lasek-Nesselquist |
| EPI_ISL_1228171, EPI_ISL_1228172 | NORTH SHORE UNIVERSITY HOSPITAL | Wadsworth Center, New York State Department of Health | Kirsten St. George, Daryl M. Lamson, Alexis Russel, Matthew Shudt, Melissa A Leisner, Jonathan Plitnick, Navjot Singh, John Kelly, Erasmus Schneider, Erica Lasek-Nesselquist |
| EPI_ISL_1228173 | MONTEFIORE MEDICAL CENTER LABORATORIES | Wadsworth Center, New York State Department of Health | Kirsten St. George, Daryl M. Lamson, Alexis Russel, Matthew Shudt, Melissa A Leisner, Jonathan Plitnick, Navjot Singh, John Kelly, Erasmus Schneider, Erica Lasek-Nesselquist |
| EPI_ISL_1228174, EPI_ISL_1228175 | NORTH SHORE UNIVERSITY HOSPITAL | Wadsworth Center, New York State Department of Health | Kirsten St. George, Daryl M. Lamson, Alexis Russel, Matthew Shudt, Melissa A Leisner, Jonathan Plitnick, Navjot Singh, John Kelly, Erasmus Schneider, Erica Lasek-Nesselquist |
| EPI_ISL_1228176 | MONTEFIORE MEDICAL CENTER LABORATORIES | Wadsworth Center, New York State Department of Health | Kirsten St. George, Daryl M. Lamson, Alexis Russel, Matthew Shudt, Melissa A Leisner, Jonathan Plitnick, Navjot Singh, John Kelly, Erasmus Schneider, Erica Lasek-Nesselquist |
| EPI_ISL_1228177, EPI_ISL_1228178, EPI_ISL_1228179, EPI_ISL_1228180 | NYC Pandemic Response Lab | Wadsworth Center, New York State Department of Health | Kirsten St. George, Daryl M. Lamson, Alexis Russel, Matthew Shudt, Melissa A Leisner, Jonathan Plitnick, Navjot Singh, John Kelly, Erasmus Schneider, Erica Lasek-Nesselquist |
| EPI_ISL_1228181 | New York Presbyterian Hospital | Wadsworth Center, New York State Department of Health | Kirsten St. George, Daryl M. Lamson, Alexis Russel, Matthew Shudt, Melissa A Leisner, Jonathan Plitnick, Navjot Singh, John Kelly, Erasmus Schneider, Erica Lasek-Nesselquist |
| EPI_ISL_1228182 | NYC Pandemic Response Lab | Wadsworth Center, New York State Department of Health | Kirsten St. George, Daryl M. Lamson, Alexis Russel, Matthew Shudt, Melissa A Leisner, Jonathan Plitnick, Navjot Singh, John Kelly, Erasmus Schneider, Erica Lasek-Nesselquist |
| EPI_ISL_1228183, EPI_ISL_1228184 | New York Presbyterian Hospital | Wadsworth Center, New York State Department of Health | Kirsten St. George, Daryl M. Lamson, Alexis Russel, Matthew Shudt, Melissa A Leisner, Jonathan Plitnick, Navjot Singh, John Kelly, Erasmus Schneider, Erica Lasek-Nesselquist |
| EPI_ISL_1228185, EPI_ISL_1228186, EPI_ISL_1228187 | NYC Pandemic Response Lab | Wadsworth Center, New York State Department of Health | Kirsten St. George, Daryl M. Lamson, Alexis Russel, Matthew Shudt, Melissa A Leisner, Jonathan Plitnick, Navjot Singh, John Kelly, Erasmus Schneider, Erica Lasek-Nesselquist |
| EPI_ISL_1228188 | MONTEFIORE MEDICAL CENTER LABORATORIES | Wadsworth Center, New York State Department of Health | Kirsten St. George, Daryl M. Lamson, Alexis Russel, Matthew Shudt, Melissa A Leisner, Jonathan Plitnick, Navjot Singh, John Kelly, Erasmus Schneider, Erica Lasek-Nesselquist |
| EPI_ISL_1228189, EPI_ISL_1228190, EPI_ISL_1228191, EPI_ISL_1228192, EPI_ISL_1228193, EPI_ISL_1228194, EPI_ISL_1228195, EPI_ISL_1228196, EPI_ISL_1228197, EPI_ISL_1228198, EPI_ISL_1228199, EPI_ISL_1228200, EPI_ISL_1228201, EPI_ISL_1228202, EPI_ISL_1228203, EPI_ISL_1228204, EPI_ISL_1228205, EPI_ISL_1228206, EPI_ISL_1228207, EPI_ISL_1228208, EPI_ISL_1228209, EPI_ISL_1228210, EPI_ISL_1228211, EPI_ISL_1228212, EPI_ISL_1228213, EPI_ISL_1228214, EPI_ISL_1228215, EPI_ISL_1228216, EPI_ISL_1228217 | | | |
| see above | NYC Pandemic Response Lab | Wadsworth Center, New York State Department of Health | Kirsten St. George, Daryl M. Lamson, Alexis Russel, Matthew Shudt, Melissa A Leisner, Jonathan Plitnick, Navjot Singh, John Kelly, Erasmus Schneider, Erica Lasek-Nesselquist |
| EPI_ISL_1228218, EPI_ISL_1228219, EPI_ISL_1228220, EPI_ISL_1228221 | ALBANY MEDICAL CENTER | Wadsworth Center, New York State Department of Health | Kirsten St. George, Daryl M. Lamson, Alexis Russel, Matthew Shudt, Melissa A Leisner, Jonathan Plitnick, Navjot Singh, John Kelly, Erasmus Schneider, Erica Lasek-Nesselquist |
| EPI_ISL_1228222, EPI_ISL_1228223 | Wadsworth Center, New York State Department of Health | Wadsworth Center, New York State Department of Health | Kirsten St. George, Daryl M. Lamson, Alexis Russel, Matthew Shudt, Melissa A Leisner, Jonathan Plitnick, Navjot Singh, John Kelly, Erasmus Schneider, Erica Lasek-Nesselquist |
| EPI_ISL_1228224, EPI_ISL_1228225, EPI_ISL_1228226, EPI_ISL_1228227 | MEMORIAL SLOAN KETTERING CANCER CENTER | Wadsworth Center, New York State Department of Health | Kirsten St. George, Daryl M. Lamson, Alexis Russel, Matthew Shudt, Melissa A Leisner, Jonathan Plitnick, Navjot Singh, John Kelly, Erasmus Schneider, Erica Lasek-Nesselquist |
| EPI_ISL_1228228 | NYC Pandemic Response Lab | Wadsworth Center, New York State Department of Health | Kirsten St. George, Daryl M. Lamson, Alexis Russel, Matthew Shudt, Melissa A Leisner, Jonathan Plitnick, Navjot Singh, John Kelly, Erasmus Schneider, Erica Lasek-Nesselquist |
| EPI_ISL_1228229 | MEMORIAL SLOAN KETTERING CANCER CENTER | Wadsworth Center, New York State Department of Health | Kirsten St. George, Daryl M. Lamson, Alexis Russel, Matthew Shudt, Melissa A Leisner, Jonathan Plitnick, Navjot Singh, John Kelly, Erasmus Schneider, Erica Lasek-Nesselquist |
| EPI_ISL_1228230, EPI_ISL_1228231, EPI_ISL_1228232, EPI_ISL_1228233, EPI_ISL_1228234 | NYC Pandemic Response Lab | Wadsworth Center, New York State Department of Health | Kirsten St. George, Daryl M. Lamson, Alexis Russel, Matthew Shudt, Melissa A Leisner, Jonathan Plitnick, Navjot Singh, John Kelly, Erasmus Schneider, Erica Lasek-Nesselquist |
| EPI_ISL_1228235 | NORTHWELL HEALTH LABORATORIES | Wadsworth Center, New York State Department of Health | Kirsten St. George, Daryl M. Lamson, Alexis Russel, Matthew Shudt, Melissa A Leisner, Jonathan Plitnick, Navjot Singh, John Kelly, Erasmus Schneider, Erica Lasek-Nesselquist |
| EPI_ISL_1228236 | MEMORIAL SLOAN KETTERING CANCER CENTER | Wadsworth Center, New York State Department of Health | Kirsten St. George, Daryl M. Lamson, Alexis Russel, Matthew Shudt, Melissa A Leisner, Jonathan Plitnick, Navjot Singh, John Kelly, Erasmus Schneider, Erica Lasek-Nesselquist |
| EPI_ISL_1228237, EPI_ISL_1228238, EPI_ISL_1228239 | NYC Pandemic Response Lab | Wadsworth Center, New York State Department of Health | Kirsten St. George, Daryl M. Lamson, Alexis Russel, Matthew Shudt, Melissa A Leisner, Jonathan Plitnick, Navjot Singh, John Kelly, Erasmus Schneider, Erica Lasek-Nesselquist |
| EPI_ISL_1228240, EPI_ISL_1228241, EPI_ISL_1228242, EPI_ISL_1228243, EPI_ISL_1228244, EPI_ISL_1228245, EPI_ISL_1228246, EPI_ISL_1228247, EPI_ISL_1228248, EPI_ISL_1228249, EPI_ISL_1228250, EPI_ISL_1228251 | | | |
| see above | MEMORIAL SLOAN KETTERING CANCER CENTER | Wadsworth Center, New York State Department of Health | Kirsten St. George, Daryl M. Lamson, Alexis Russel, Matthew Shudt, Melissa A Leisner, Jonathan Plitnick, Navjot Singh, John Kelly, Erasmus Schneider, Erica Lasek-Nesselquist |
| EPI_ISL_1228252 | WESTCHESTER MEDICAL CENTER | Wadsworth Center, New York State Department of Health | Kirsten St. George, Daryl M. Lamson, Alexis Russel, Matthew Shudt, Melissa A Leisner, Jonathan Plitnick, Navjot Singh, John Kelly, Erasmus Schneider, Erica Lasek-Nesselquist |

| | | | |
|---|---|---|---|
| EPI_ISL_1228253 | Columbia University Irving Medical Center | Wadsworth Center, New York State Department of Health | Kirsten St. George, Daryl M. Lamson, Alexis Russel, Matthew Shudt, Melissa A Leisner, Jonathan Plitnick, Navjot Singh, John Kelly, Erasmus Schneider, Erica Lasek-Nesselquist |
| EPI_ISL_1228254 | MEMORIAL SLOAN KETTERING CANCER CENTER | Wadsworth Center, New York State Department of Health | Kirsten St. George, Daryl M. Lamson, Alexis Russel, Matthew Shudt, Melissa A Leisner, Jonathan Plitnick, Navjot Singh, John Kelly, Erasmus Schneider, Erica Lasek-Nesselquist |
| EPI_ISL_1228255 | WESTCHESTER MEDICAL CENTER | Wadsworth Center, New York State Department of Health | Kirsten St. George, Daryl M. Lamson, Alexis Russel, Matthew Shudt, Melissa A Leisner, Jonathan Plitnick, Navjot Singh, John Kelly, Erasmus Schneider, Erica Lasek-Nesselquist |
| EPI_ISL_1228256 | MONTEFIORE MEDICAL CENTER LABORATORIES | Wadsworth Center, New York State Department of Health | Kirsten St. George, Daryl M. Lamson, Alexis Russel, Matthew Shudt, Melissa A Leisner, Jonathan Plitnick, Navjot Singh, John Kelly, Erasmus Schneider, Erica Lasek-Nesselquist |
| EPI_ISL_1228257, EPI_ISL_1228258, EPI_ISL_1228259 | WESTCHESTER MEDICAL CENTER | Wadsworth Center, New York State Department of Health | Kirsten St. George, Daryl M. Lamson, Alexis Russel, Matthew Shudt, Melissa A Leisner, Jonathan Plitnick, Navjot Singh, John Kelly, Erasmus Schneider, Erica Lasek-Nesselquist |
| EPI_ISL_1228260 | Wadsworth Center, New York State Department of Health | Wadsworth Center, New York State Department of Health | Kirsten St. George, Daryl M. Lamson, Alexis Russel, Matthew Shudt, Melissa A Leisner, Jonathan Plitnick, Navjot Singh, John Kelly, Erasmus Schneider, Erica Lasek-Nesselquist |
| EPI_ISL_1228261 | WESTCHESTER MEDICAL CENTER | Wadsworth Center, New York State Department of Health | Kirsten St. George, Daryl M. Lamson, Alexis Russel, Matthew Shudt, Melissa A Leisner, Jonathan Plitnick, Navjot Singh, John Kelly, Erasmus Schneider, Erica Lasek-Nesselquist |
| EPI_ISL_1228262 | MONTEFIORE MEDICAL CENTER LABORATORIES | Wadsworth Center, New York State Department of Health | Kirsten St. George, Daryl M. Lamson, Alexis Russel, Matthew Shudt, Melissa A Leisner, Jonathan Plitnick, Navjot Singh, John Kelly, Erasmus Schneider, Erica Lasek-Nesselquist |
| EPI_ISL_1228263, EPI_ISL_1228264 | WESTCHESTER MEDICAL CENTER | Wadsworth Center, New York State Department of Health | Kirsten St. George, Daryl M. Lamson, Alexis Russel, Matthew Shudt, Melissa A Leisner, Jonathan Plitnick, Navjot Singh, John Kelly, Erasmus Schneider, Erica Lasek-Nesselquist |
| EPI_ISL_1228265 | NYC Pandemic Response Lab | Wadsworth Center, New York State Department of Health | Kirsten St. George, Daryl M. Lamson, Alexis Russel, Matthew Shudt, Melissa A Leisner, Jonathan Plitnick, Navjot Singh, John Kelly, Erasmus Schneider, Erica Lasek-Nesselquist |
| EPI_ISL_1228266 | MONTEFIORE MEDICAL CENTER LABORATORIES | Wadsworth Center, New York State Department of Health | Kirsten St. George, Daryl M. Lamson, Alexis Russel, Matthew Shudt, Melissa A Leisner, Jonathan Plitnick, Navjot Singh, John Kelly, Erasmus Schneider, Erica Lasek-Nesselquist |
| EPI_ISL_1228267 | WESTCHESTER MEDICAL CENTER | Wadsworth Center, New York State Department of Health | Kirsten St. George, Daryl M. Lamson, Alexis Russel, Matthew Shudt, Melissa A Leisner, Jonathan Plitnick, Navjot Singh, John Kelly, Erasmus Schneider, Erica Lasek-Nesselquist |
| EPI_ISL_1228268, EPI_ISL_1228269 | GLENS FALLS HOSPITAL LABORATORY | Wadsworth Center, New York State Department of Health | Kirsten St. George, Daryl M. Lamson, Alexis Russel, Matthew Shudt, Melissa A Leisner, Jonathan Plitnick, Navjot Singh, John Kelly, Erasmus Schneider, Erica Lasek-Nesselquist |
| EPI_ISL_1228270 | ALBANY MEDICAL CENTER | Wadsworth Center, New York State Department of Health | Kirsten St. George, Daryl M. Lamson, Alexis Russel, Matthew Shudt, Melissa A Leisner, Jonathan Plitnick, Navjot Singh, John Kelly, Erasmus Schneider, Erica Lasek-Nesselquist |
| EPI_ISL_1228271 | Wadsworth Center, New York State Department of Health | Wadsworth Center, New York State Department of Health | Kirsten St. George, Daryl M. Lamson, Alexis Russel, Matthew Shudt, Melissa A Leisner, Jonathan Plitnick, Navjot Singh, John Kelly, Erasmus Schneider, Erica Lasek-Nesselquist |
| EPI_ISL_1228272, EPI_ISL_1228273, EPI_ISL_1228274 | GLENS FALLS HOSPITAL LABORATORY | Wadsworth Center, New York State Department of Health | Kirsten St. George, Daryl M. Lamson, Alexis Russel, Matthew Shudt, Melissa A Leisner, Jonathan Plitnick, Navjot Singh, John Kelly, Erasmus Schneider, Erica Lasek-Nesselquist |
| EPI_ISL_1228275, EPI_ISL_1228276, EPI_ISL_1228277 | ALBANY MEDICAL CENTER | Wadsworth Center, New York State Department of Health | Kirsten St. George, Daryl M. Lamson, Alexis Russel, Matthew Shudt, Melissa A Leisner, Jonathan Plitnick, Navjot Singh, John Kelly, Erasmus Schneider, Erica Lasek-Nesselquist |
| EPI_ISL_1228278 | SARATOGA HOSPITAL LABORATORY | Wadsworth Center, New York State Department of Health | Kirsten St. George, Daryl M. Lamson, Alexis Russel, Matthew Shudt, Melissa A Leisner, Jonathan Plitnick, Navjot Singh, John Kelly, Erasmus Schneider, Erica Lasek-Nesselquist |
| EPI_ISL_1228279 | ALBANY MEDICAL CENTER | Wadsworth Center, New York State Department of Health | Kirsten St. George, Daryl M. Lamson, Alexis Russel, Matthew Shudt, Melissa A Leisner, Jonathan Plitnick, Navjot Singh, John Kelly, Erasmus Schneider, Erica Lasek-Nesselquist |
| EPI_ISL_1228280 | Masonic Care Community of NY | Wadsworth Center, New York State Department of Health | Kirsten St. George, Daryl M. Lamson, Alexis Russel, Matthew Shudt, Melissa A Leisner, Jonathan Plitnick, Navjot Singh, John Kelly, Erasmus Schneider, Erica Lasek-Nesselquist |
| EPI_ISL_1228281 | Wadsworth Center, New York State Department of Health | Wadsworth Center, New York State Department of Health | Kirsten St. George, Daryl M. Lamson, Alexis Russel, Matthew Shudt, Melissa A Leisner, Jonathan Plitnick, Navjot Singh, John Kelly, Erasmus Schneider, Erica Lasek-Nesselquist |
| EPI_ISL_1228282, EPI_ISL_1228283 | ALBANY MEDICAL CENTER | Wadsworth Center, New York State Department of Health | Kirsten St. George, Daryl M. Lamson, Alexis Russel, Matthew Shudt, Melissa A Leisner, Jonathan Plitnick, Navjot Singh, John Kelly, Erasmus Schneider, Erica Lasek-Nesselquist |
| EPI_ISL_1228284, EPI_ISL_1228285, EPI_ISL_1228286, EPI_ISL_1228287, EPI_ISL_1228288, EPI_ISL_1228289, EPI_ISL_1228290, EPI_ISL_1228291 | THE MARY IMOGENE BASSETT HOSPITAL | Wadsworth Center, New York State Department of Health | Kirsten St. George, Daryl M. Lamson, Alexis Russel, Matthew Shudt, Melissa A Leisner, Jonathan Plitnick, Navjot Singh, John Kelly, Erasmus Schneider, Erica Lasek-Nesselquist |
| EPI_ISL_1228292 | ADIRONDACK MEDICAL CENTER | Wadsworth Center, New York State Department of Health | Kirsten St. George, Daryl M. Lamson, Alexis Russel, Matthew Shudt, Melissa A Leisner, Jonathan Plitnick, Navjot Singh, John Kelly, Erasmus Schneider, Erica Lasek-Nesselquist |
| EPI_ISL_1228293 | SUNY UPSTATE MEDICAL UNIVERSITY | Wadsworth Center, New York State Department of Health | Kirsten St. George, Daryl M. Lamson, Alexis Russel, Matthew Shudt, Melissa A Leisner, Jonathan Plitnick, Navjot Singh, John Kelly, Erasmus Schneider, Erica Lasek-Nesselquist |
| EPI_ISL_1228294, EPI_ISL_1228295 | Wadsworth Center, New York State Department of Health | Wadsworth Center, New York State Department of Health | Kirsten St. George, Daryl M. Lamson, Alexis Russel, Matthew Shudt, Melissa A Leisner, Jonathan Plitnick, Navjot Singh, John Kelly, Erasmus Schneider, Erica Lasek-Nesselquist |
| EPI_ISL_1228296, EPI_ISL_1228297, EPI_ISL_1228298, EPI_ISL_1228299, EPI_ISL_1228300, EPI_ISL_1228301, EPI_ISL_1228302, EPI_ISL_1228303, EPI_ISL_1228304, EPI_ISL_1228305, EPI_ISL_1228306 | | | |
| see above | URMC LABS | Wadsworth Center, New York State Department of Health | Kirsten St. George, Daryl M. Lamson, Alexis Russel, Matthew Shudt, Melissa A Leisner, Jonathan Plitnick, Navjot Singh, John Kelly, Erasmus Schneider, Erica Lasek-Nesselquist |
| EPI_ISL_1228307, EPI_ISL_1228308, EPI_ISL_1228309, EPI_ISL_1228310, EPI_ISL_1228311, EPI_ISL_1228312, EPI_ISL_1228313, EPI_ISL_1228314, EPI_ISL_1228315, EPI_ISL_1228316, EPI_ISL_1228317, EPI_ISL_1228318, EPI_ISL_1228319, EPI_ISL_1228320, EPI_ISL_1228321, EPI_ISL_1228322, EPI_ISL_1228323, EPI_ISL_1228324, EPI_ISL_1228325, EPI_ISL_1228326, EPI_ISL_1228327, EPI_ISL_1228328, EPI_ISL_1228329, EPI_ISL_1228330, EPI_ISL_1228331 | | | |
| see above | MEMORIAL SLOAN KETTERING CANCER CENTER | Wadsworth Center, New York State Department of Health | Kirsten St. George, Daryl M. Lamson, Alexis Russel, Matthew Shudt, Melissa A Leisner, Jonathan Plitnick, Navjot Singh, John Kelly, Erasmus Schneider, Erica Lasek-Nesselquist |
| EPI_ISL_1228332 | NORTH SHORE UNIVERSITY HOSPITAL | Wadsworth Center, New York State Department of Health | Kirsten St. George, Daryl M. Lamson, Alexis Russel, Matthew Shudt, Melissa A Leisner, Jonathan Plitnick, Navjot Singh, John Kelly, Erasmus Schneider, Erica Lasek-Nesselquist |
| EPI_ISL_1228333, EPI_ISL_1228334, EPI_ISL_1228335, EPI_ISL_1228336, EPI_ISL_1228337, EPI_ISL_1228338, EPI_ISL_1228339, EPI_ISL_1228340, EPI_ISL_1228341 | MEMORIAL SLOAN KETTERING CANCER CENTER | Wadsworth Center, New York State Department of Health | Kirsten St. George, Daryl M. Lamson, Alexis Russel, Matthew Shudt, Melissa A Leisner, Jonathan Plitnick, Navjot Singh, John Kelly, Erasmus Schneider, Erica Lasek-Nesselquist |
| EPI_ISL_1228342, EPI_ISL_1228343, EPI_ISL_1228344, EPI_ISL_1228345 | NORTHWELL HEALTH LABORATORIES | Wadsworth Center, New York State Department of Health | Kirsten St. George, Daryl M. Lamson, Alexis Russel, Matthew Shudt, Melissa A Leisner, Jonathan Plitnick, Navjot Singh, John Kelly, Erasmus Schneider, Erica Lasek-Nesselquist |
| EPI_ISL_1228346, EPI_ISL_1228347 | MEMORIAL SLOAN KETTERING CANCER CENTER | Wadsworth Center, New York State Department of Health | Kirsten St. George, Daryl M. Lamson, Alexis Russel, Matthew Shudt, Melissa A Leisner, Jonathan Plitnick, Navjot Singh, John Kelly, Erasmus Schneider, |

| | | | |
|---|---|---|---|
| | | | Erica Lasek-Nesselquist |
| EPI_ISL_1228348 | NORTHWELL HEALTH LABORATORIES | Wadsworth Center, New York State Department of Health | Kirsten St. George, Daryl M. Lamson, Alexis Russel, Matthew Shudt, Melissa A Leisner, Jonathan Plitnick, Navjot Singh, John Kelly, Erasmus Schneider, Erica Lasek-Nesselquist |
| EPI_ISL_1228349, EPI_ISL_1228350, EPI_ISL_1228351, EPI_ISL_1228352, EPI_ISL_1228353, EPI_ISL_1228354, EPI_ISL_1228355 | MEMORIAL SLOAN KETTERING CANCER CENTER | Wadsworth Center, New York State Department of Health | Kirsten St. George, Daryl M. Lamson, Alexis Russel, Matthew Shudt, Melissa A Leisner, Jonathan Plitnick, Navjot Singh, John Kelly, Erasmus Schneider, Erica Lasek-Nesselquist |
| EPI_ISL_1228356 | NORTH SHORE UNIVERSITY HOSPITAL | Wadsworth Center, New York State Department of Health | Kirsten St. George, Daryl M. Lamson, Alexis Russel, Matthew Shudt, Melissa A Leisner, Jonathan Plitnick, Navjot Singh, John Kelly, Erasmus Schneider, Erica Lasek-Nesselquist |
| EPI_ISL_1228357, EPI_ISL_1228358, EPI_ISL_1228359 | NORTHWELL HEALTH LABORATORIES | Wadsworth Center, New York State Department of Health | Kirsten St. George, Daryl M. Lamson, Alexis Russel, Matthew Shudt, Melissa A Leisner, Jonathan Plitnick, Navjot Singh, John Kelly, Erasmus Schneider, Erica Lasek-Nesselquist |
| EPI_ISL_1228360 | MEMORIAL SLOAN KETTERING CANCER CENTER | Wadsworth Center, New York State Department of Health | Kirsten St. George, Daryl M. Lamson, Alexis Russel, Matthew Shudt, Melissa A Leisner, Jonathan Plitnick, Navjot Singh, John Kelly, Erasmus Schneider, Erica Lasek-Nesselquist |
| EPI_ISL_1228361 | NORTHWELL HEALTH LABORATORIES | Wadsworth Center, New York State Department of Health | Kirsten St. George, Daryl M. Lamson, Alexis Russel, Matthew Shudt, Melissa A Leisner, Jonathan Plitnick, Navjot Singh, John Kelly, Erasmus Schneider, Erica Lasek-Nesselquist |
| EPI_ISL_1228362, EPI_ISL_1228363, EPI_ISL_1228364, EPI_ISL_1228365, EPI_ISL_1228366, EPI_ISL_1228367, EPI_ISL_1228368 | MEMORIAL SLOAN KETTERING CANCER CENTER | Wadsworth Center, New York State Department of Health | Kirsten St. George, Daryl M. Lamson, Alexis Russel, Matthew Shudt, Melissa A Leisner, Jonathan Plitnick, Navjot Singh, John Kelly, Erasmus Schneider, Erica Lasek-Nesselquist |
| EPI_ISL_1228369, EPI_ISL_1228370, EPI_ISL_1228371, EPI_ISL_1228372, EPI_ISL_1228373 | NYC Pandemic Response Lab | Wadsworth Center, New York State Department of Health | Kirsten St. George, Daryl M. Lamson, Alexis Russel, Matthew Shudt, Melissa A Leisner, Jonathan Plitnick, Navjot Singh, John Kelly, Erasmus Schneider, Erica Lasek-Nesselquist |
| EPI_ISL_1228374 | WESTCHESTER MEDICAL CENTER | Wadsworth Center, New York State Department of Health | Kirsten St. George, Daryl M. Lamson, Alexis Russel, Matthew Shudt, Melissa A Leisner, Jonathan Plitnick, Navjot Singh, John Kelly, Erasmus Schneider, Erica Lasek-Nesselquist |
| EPI_ISL_1228375, EPI_ISL_1228376, EPI_ISL_1228377 | DEPARTMENT OF VETERANS AFFAIR MEDICAL CENTER | Wadsworth Center, New York State Department of Health | Kirsten St. George, Daryl M. Lamson, Alexis Russel, Matthew Shudt, Melissa A Leisner, Jonathan Plitnick, Navjot Singh, John Kelly, Erasmus Schneider, Erica Lasek-Nesselquist |
| EPI_ISL_1228378, EPI_ISL_1228379 | WESTCHESTER MEDICAL CENTER | Wadsworth Center, New York State Department of Health | Kirsten St. George, Daryl M. Lamson, Alexis Russel, Matthew Shudt, Melissa A Leisner, Jonathan Plitnick, Navjot Singh, John Kelly, Erasmus Schneider, Erica Lasek-Nesselquist |
| EPI_ISL_1228380 | SUNY UPSTATE MEDICAL UNIVERSITY | Wadsworth Center, New York State Department of Health | Kirsten St. George, Daryl M. Lamson, Alexis Russel, Matthew Shudt, Melissa A Leisner, Jonathan Plitnick, Navjot Singh, John Kelly, Erasmus Schneider, Erica Lasek-Nesselquist |
| EPI_ISL_1228381, EPI_ISL_1228382, EPI_ISL_1228383, EPI_ISL_1228384, EPI_ISL_1228385, EPI_ISL_1228386, EPI_ISL_1228387, EPI_ISL_1228388, EPI_ISL_1228389, EPI_ISL_1228390, EPI_ISL_1228391, EPI_ISL_1228392, EPI_ISL_1228393, EPI_ISL_1228394, EPI_ISL_1228395, EPI_ISL_1228396, EPI_ISL_1228397, EPI_ISL_1228398, EPI_ISL_1228399, EPI_ISL_1228400, EPI_ISL_1228401, EPI_ISL_1228402, EPI_ISL_1228403, EPI_ISL_1228404, EPI_ISL_1228405, EPI_ISL_1228406, EPI_ISL_1228407, EPI_ISL_1228408, EPI_ISL_1228409, EPI_ISL_1228410, EPI_ISL_1228411, EPI_ISL_1228412, EPI_ISL_1228413, EPI_ISL_1228414, EPI_ISL_1228415, EPI_ISL_1228416, EPI_ISL_1228417, EPI_ISL_1228418, EPI_ISL_1228419, EPI_ISL_1228420, EPI_ISL_1228421, EPI_ISL_1228422, EPI_ISL_1228423, EPI_ISL_1228424, EPI_ISL_1228425, EPI_ISL_1228426, EPI_ISL_1228427, EPI_ISL_1228428, EPI_ISL_1228429 | | | |
| see above | WESTCHESTER MEDICAL CENTER | Wadsworth Center, New York State Department of Health | Kirsten St. George, Daryl M. Lamson, Alexis Russel, Matthew Shudt, Melissa A Leisner, Jonathan Plitnick, Navjot Singh, John Kelly, Erasmus Schneider, Erica Lasek-Nesselquist |
| EPI_ISL_1228430 | ALBANY MEDICAL CENTER | Wadsworth Center, New York State Department of Health | Kirsten St. George, Daryl M. Lamson, Alexis Russel, Matthew Shudt, Melissa A Leisner, Jonathan Plitnick, Navjot Singh, John Kelly, Erasmus Schneider, Erica Lasek-Nesselquist |
| EPI_ISL_1228431, EPI_ISL_1228432, EPI_ISL_1228433 | WESTCHESTER MEDICAL CENTER | Wadsworth Center, New York State Department of Health | Kirsten St. George, Daryl M. Lamson, Alexis Russel, Matthew Shudt, Melissa A Leisner, Jonathan Plitnick, Navjot Singh, John Kelly, Erasmus Schneider, Erica Lasek-Nesselquist |
| EPI_ISL_1228434, EPI_ISL_1228435, EPI_ISL_1228436, EPI_ISL_1228437, EPI_ISL_1228438, EPI_ISL_1228439, EPI_ISL_1228440, EPI_ISL_1228441, EPI_ISL_1228442, EPI_ISL_1228443 | GLENS FALLS HOSPITAL LABORATORY | Wadsworth Center, New York State Department of Health | Kirsten St. George, Daryl M. Lamson, Alexis Russel, Matthew Shudt, Melissa A Leisner, Jonathan Plitnick, Navjot Singh, John Kelly, Erasmus Schneider, Erica Lasek-Nesselquist |
| EPI_ISL_1228444, EPI_ISL_1228445, EPI_ISL_1228446 | Wadsworth Center, New York State Department of Health | Wadsworth Center, New York State Department of Health | Kirsten St. George, Daryl M. Lamson, Alexis Russel, Matthew Shudt, Melissa A Leisner, Jonathan Plitnick, Navjot Singh, John Kelly, Erasmus Schneider, Erica Lasek-Nesselquist |
| EPI_ISL_1228447, EPI_ISL_1228448, EPI_ISL_1228449 | GLENS FALLS HOSPITAL LABORATORY | Wadsworth Center, New York State Department of Health | Kirsten St. George, Daryl M. Lamson, Alexis Russel, Matthew Shudt, Melissa A Leisner, Jonathan Plitnick, Navjot Singh, John Kelly, Erasmus Schneider, Erica Lasek-Nesselquist |
| EPI_ISL_1228450 | Wadsworth Center, New York State Department of Health | Wadsworth Center, New York State Department of Health | Kirsten St. George, Daryl M. Lamson, Alexis Russel, Matthew Shudt, Melissa A Leisner, Jonathan Plitnick, Navjot Singh, John Kelly, Erasmus Schneider, Erica Lasek-Nesselquist |
| EPI_ISL_1228451 | GLENS FALLS HOSPITAL LABORATORY | Wadsworth Center, New York State Department of Health | Kirsten St. George, Daryl M. Lamson, Alexis Russel, Matthew Shudt, Melissa A Leisner, Jonathan Plitnick, Navjot Singh, John Kelly, Erasmus Schneider, Erica Lasek-Nesselquist |
| EPI_ISL_1228452 | Wadsworth Center, New York State Department of Health | Wadsworth Center, New York State Department of Health | Kirsten St. George, Daryl M. Lamson, Alexis Russel, Matthew Shudt, Melissa A Leisner, Jonathan Plitnick, Navjot Singh, John Kelly, Erasmus Schneider, Erica Lasek-Nesselquist |
| EPI_ISL_1228453 | THE MARY IMOGENE BASSETT HOSPITAL | Wadsworth Center, New York State Department of Health | Kirsten St. George, Daryl M. Lamson, Alexis Russel, Matthew Shudt, Melissa A Leisner, Jonathan Plitnick, Navjot Singh, John Kelly, Erasmus Schneider, Erica Lasek-Nesselquist |
| EPI_ISL_1228454, EPI_ISL_1228455, EPI_ISL_1228456, EPI_ISL_1228457, EPI_ISL_1228458, EPI_ISL_1228459 | GLENS FALLS HOSPITAL LABORATORY | Wadsworth Center, New York State Department of Health | Kirsten St. George, Daryl M. Lamson, Alexis Russel, Matthew Shudt, Melissa A Leisner, Jonathan Plitnick, Navjot Singh, John Kelly, Erasmus Schneider, Erica Lasek-Nesselquist |
| EPI_ISL_1228460, EPI_ISL_1228461, EPI_ISL_1228462 | Wadsworth Center, New York State Department of Health | Wadsworth Center, New York State Department of Health | Kirsten St. George, Daryl M. Lamson, Alexis Russel, Matthew Shudt, Melissa A Leisner, Jonathan Plitnick, Navjot Singh, John Kelly, Erasmus Schneider, Erica Lasek-Nesselquist |
| EPI_ISL_1228463, EPI_ISL_1228464 | GLENS FALLS HOSPITAL LABORATORY | Wadsworth Center, New York State Department of Health | Kirsten St. George, Daryl M. Lamson, Alexis Russel, Matthew Shudt, Melissa A Leisner, Jonathan Plitnick, Navjot Singh, John Kelly, Erasmus Schneider, Erica Lasek-Nesselquist |
| EPI_ISL_1228465, EPI_ISL_1228466 | Wadsworth Center, New York State Department of Health | Wadsworth Center, New York State Department of Health | Kirsten St. George, Daryl M. Lamson, Alexis Russel, Matthew Shudt, Melissa A Leisner, Jonathan Plitnick, Navjot Singh, John Kelly, Erasmus Schneider, Erica Lasek-Nesselquist |
| EPI_ISL_1228467, EPI_ISL_1228468 | GLENS FALLS HOSPITAL LABORATORY | Wadsworth Center, New York State Department of Health | Kirsten St. George, Daryl M. Lamson, Alexis Russel, Matthew Shudt, Melissa A Leisner, Jonathan Plitnick, Navjot Singh, John Kelly, Erasmus Schneider, Erica Lasek-Nesselquist |
| EPI_ISL_1228469 | Wadsworth Center, New York State Department of Health | Wadsworth Center, New York State Department of Health | Kirsten St. George, Daryl M. Lamson, Alexis Russel, Matthew Shudt, Melissa A Leisner, Jonathan Plitnick, Navjot Singh, John Kelly, Erasmus Schneider, Erica Lasek-Nesselquist |
| EPI_ISL_1228470, EPI_ISL_1228471, EPI_ISL_1228472, EPI_ISL_1228473 | GLENS FALLS HOSPITAL LABORATORY | Wadsworth Center, New York State Department of Health | Kirsten St. George, Daryl M. Lamson, Alexis Russel, Matthew Shudt, Melissa A Leisner, Jonathan Plitnick, Navjot Singh, John Kelly, Erasmus Schneider, Erica Lasek-Nesselquist |
| EPI_ISL_1228474, EPI_ISL_1228475 | Wadsworth Center, New York State Department of Health | Wadsworth Center, New York State Department of Health | Kirsten St. George, Daryl M. Lamson, Alexis Russel, Matthew Shudt, Melissa A Leisner, Jonathan Plitnick, Navjot Singh, John Kelly, Erasmus Schneider, |

| | | | |
|---|---|---|---|
| | | | Erica Lasek-Nesselquist |
| EPI_ISL_1228476 | GLENS FALLS HOSPITAL LABORATORY | Wadsworth Center, New York State Department of Health | Kirsten St. George, Daryl M. Lamson, Alexis Russel, Matthew Shudt, Melissa A Leisner, Jonathan Plitnick, Navjot Singh, John Kelly, Erasmus Schneider, Erica Lasek-Nesselquist |
| EPI_ISL_1228477, EPI_ISL_1228478, EPI_ISL_1228479 | ALBANY MEDICAL CENTER | Wadsworth Center, New York State Department of Health | Kirsten St. George, Daryl M. Lamson, Alexis Russel, Matthew Shudt, Melissa A Leisner, Jonathan Plitnick, Navjot Singh, John Kelly, Erasmus Schneider, Erica Lasek-Nesselquist |
| EPI_ISL_1228480, EPI_ISL_1228481, EPI_ISL_1228482, EPI_ISL_1228483 | URMC LABS | Wadsworth Center, New York State Department of Health | Kirsten St. George, Daryl M. Lamson, Alexis Russel, Matthew Shudt, Melissa A Leisner, Jonathan Plitnick, Navjot Singh, John Kelly, Erasmus Schneider, Erica Lasek-Nesselquist |
| EPI_ISL_1228484, EPI_ISL_1228485, EPI_ISL_1228486, EPI_ISL_1228487, EPI_ISL_1228488, EPI_ISL_1228489, EPI_ISL_1228490, EPI_ISL_1228491, EPI_ISL_1228492, EPI_ISL_1228493, EPI_ISL_1228494, EPI_ISL_1228495, EPI_ISL_1228496, EPI_ISL_1228497, EPI_ISL_1228498, EPI_ISL_1228499, EPI_ISL_1228500 | | | |
| see above | MEMORIAL SLOAN KETTERING CANCER CENTER | Wadsworth Center, New York State Department of Health | Kirsten St. George, Daryl M. Lamson, Alexis Russel, Matthew Shudt, Melissa A Leisner, Jonathan Plitnick, Navjot Singh, John Kelly, Erasmus Schneider, Erica Lasek-Nesselquist |
| EPI_ISL_1228501 | WESTCHESTER MEDICAL CENTER | Wadsworth Center, New York State Department of Health | Kirsten St. George, Daryl M. Lamson, Alexis Russel, Matthew Shudt, Melissa A Leisner, Jonathan Plitnick, Navjot Singh, John Kelly, Erasmus Schneider, Erica Lasek-Nesselquist |
| EPI_ISL_1228502, EPI_ISL_1228503 | MEMORIAL SLOAN KETTERING CANCER CENTER | Wadsworth Center, New York State Department of Health | Kirsten St. George, Daryl M. Lamson, Alexis Russel, Matthew Shudt, Melissa A Leisner, Jonathan Plitnick, Navjot Singh, John Kelly, Erasmus Schneider, Erica Lasek-Nesselquist |
| EPI_ISL_1228504, EPI_ISL_1228505, EPI_ISL_1228506, EPI_ISL_1228507, EPI_ISL_1228508 | WESTCHESTER MEDICAL CENTER | Wadsworth Center, New York State Department of Health | Kirsten St. George, Daryl M. Lamson, Alexis Russel, Matthew Shudt, Melissa A Leisner, Jonathan Plitnick, Navjot Singh, John Kelly, Erasmus Schneider, Erica Lasek-Nesselquist |
| EPI_ISL_1228509 | Columbia University Irving Medical Center | Wadsworth Center, New York State Department of Health | Kirsten St. George, Daryl M. Lamson, Alexis Russel, Matthew Shudt, Melissa A Leisner, Jonathan Plitnick, Navjot Singh, John Kelly, Erasmus Schneider, Erica Lasek-Nesselquist |
| EPI_ISL_1228510, EPI_ISL_1228511, EPI_ISL_1228512, EPI_ISL_1228513 | WESTCHESTER MEDICAL CENTER | Wadsworth Center, New York State Department of Health | Kirsten St. George, Daryl M. Lamson, Alexis Russel, Matthew Shudt, Melissa A Leisner, Jonathan Plitnick, Navjot Singh, John Kelly, Erasmus Schneider, Erica Lasek-Nesselquist |
| EPI_ISL_1228514 | Columbia University Irving Medical Center | Wadsworth Center, New York State Department of Health | Kirsten St. George, Daryl M. Lamson, Alexis Russel, Matthew Shudt, Melissa A Leisner, Jonathan Plitnick, Navjot Singh, John Kelly, Erasmus Schneider, Erica Lasek-Nesselquist |
| EPI_ISL_1228515, EPI_ISL_1228516, EPI_ISL_1228517, EPI_ISL_1228518, EPI_ISL_1228519 | WESTCHESTER MEDICAL CENTER | Wadsworth Center, New York State Department of Health | Kirsten St. George, Daryl M. Lamson, Alexis Russel, Matthew Shudt, Melissa A Leisner, Jonathan Plitnick, Navjot Singh, John Kelly, Erasmus Schneider, Erica Lasek-Nesselquist |
| EPI_ISL_1228520 | MONTEFIORE MEDICAL CENTER LABORATORIES | Wadsworth Center, New York State Department of Health | Kirsten St. George, Daryl M. Lamson, Alexis Russel, Matthew Shudt, Melissa A Leisner, Jonathan Plitnick, Navjot Singh, John Kelly, Erasmus Schneider, Erica Lasek-Nesselquist |
| EPI_ISL_1228521, EPI_ISL_1228522, EPI_ISL_1228523, EPI_ISL_1228524, EPI_ISL_1228525, EPI_ISL_1228526, EPI_ISL_1228527, EPI_ISL_1228528 | WESTCHESTER MEDICAL CENTER | Wadsworth Center, New York State Department of Health | Kirsten St. George, Daryl M. Lamson, Alexis Russel, Matthew Shudt, Melissa A Leisner, Jonathan Plitnick, Navjot Singh, John Kelly, Erasmus Schneider, Erica Lasek-Nesselquist |
| EPI_ISL_1228529 | Columbia University Irving Medical Center | Wadsworth Center, New York State Department of Health | Kirsten St. George, Daryl M. Lamson, Alexis Russel, Matthew Shudt, Melissa A Leisner, Jonathan Plitnick, Navjot Singh, John Kelly, Erasmus Schneider, Erica Lasek-Nesselquist |
| EPI_ISL_1228530, EPI_ISL_1228531 | MONTEFIORE MEDICAL CENTER LABORATORIES | Wadsworth Center, New York State Department of Health | Kirsten St. George, Daryl M. Lamson, Alexis Russel, Matthew Shudt, Melissa A Leisner, Jonathan Plitnick, Navjot Singh, John Kelly, Erasmus Schneider, Erica Lasek-Nesselquist |
| EPI_ISL_1228532, EPI_ISL_1228533, EPI_ISL_1228534, EPI_ISL_1228535 | WESTCHESTER MEDICAL CENTER | Wadsworth Center, New York State Department of Health | Kirsten St. George, Daryl M. Lamson, Alexis Russel, Matthew Shudt, Melissa A Leisner, Jonathan Plitnick, Navjot Singh, John Kelly, Erasmus Schneider, Erica Lasek-Nesselquist |
| EPI_ISL_1228536 | MONTEFIORE MEDICAL CENTER LABORATORIES | Wadsworth Center, New York State Department of Health | Kirsten St. George, Daryl M. Lamson, Alexis Russel, Matthew Shudt, Melissa A Leisner, Jonathan Plitnick, Navjot Singh, John Kelly, Erasmus Schneider, Erica Lasek-Nesselquist |
| EPI_ISL_1228537, EPI_ISL_1228538, EPI_ISL_1228539, EPI_ISL_1228540 | WESTCHESTER MEDICAL CENTER | Wadsworth Center, New York State Department of Health | Kirsten St. George, Daryl M. Lamson, Alexis Russel, Matthew Shudt, Melissa A Leisner, Jonathan Plitnick, Navjot Singh, John Kelly, Erasmus Schneider, Erica Lasek-Nesselquist |
| EPI_ISL_1228541, EPI_ISL_1228542 | MONTEFIORE MEDICAL CENTER LABORATORIES | Wadsworth Center, New York State Department of Health | Kirsten St. George, Daryl M. Lamson, Alexis Russel, Matthew Shudt, Melissa A Leisner, Jonathan Plitnick, Navjot Singh, John Kelly, Erasmus Schneider, Erica Lasek-Nesselquist |
| EPI_ISL_1228543, EPI_ISL_1228544 | Columbia University Irving Medical Center | Wadsworth Center, New York State Department of Health | Kirsten St. George, Daryl M. Lamson, Alexis Russel, Matthew Shudt, Melissa A Leisner, Jonathan Plitnick, Navjot Singh, John Kelly, Erasmus Schneider, Erica Lasek-Nesselquist |
| EPI_ISL_1228545, EPI_ISL_1228546, EPI_ISL_1228547, EPI_ISL_1228548, EPI_ISL_1228549 | WESTCHESTER MEDICAL CENTER | Wadsworth Center, New York State Department of Health | Kirsten St. George, Daryl M. Lamson, Alexis Russel, Matthew Shudt, Melissa A Leisner, Jonathan Plitnick, Navjot Singh, John Kelly, Erasmus Schneider, Erica Lasek-Nesselquist |
| EPI_ISL_1228550 | MONTEFIORE MEDICAL CENTER LABORATORIES | Wadsworth Center, New York State Department of Health | Kirsten St. George, Daryl M. Lamson, Alexis Russel, Matthew Shudt, Melissa A Leisner, Jonathan Plitnick, Navjot Singh, John Kelly, Erasmus Schneider, Erica Lasek-Nesselquist |
| EPI_ISL_1228551, EPI_ISL_1228552 | WESTCHESTER MEDICAL CENTER | Wadsworth Center, New York State Department of Health | Kirsten St. George, Daryl M. Lamson, Alexis Russel, Matthew Shudt, Melissa A Leisner, Jonathan Plitnick, Navjot Singh, John Kelly, Erasmus Schneider, Erica Lasek-Nesselquist |
| EPI_ISL_1228553 | MONTEFIORE MEDICAL CENTER LABORATORIES | Wadsworth Center, New York State Department of Health | Kirsten St. George, Daryl M. Lamson, Alexis Russel, Matthew Shudt, Melissa A Leisner, Jonathan Plitnick, Navjot Singh, John Kelly, Erasmus Schneider, Erica Lasek-Nesselquist |
| EPI_ISL_1228554, EPI_ISL_1228555, EPI_ISL_1228556, EPI_ISL_1228557 | WESTCHESTER MEDICAL CENTER | Wadsworth Center, New York State Department of Health | Kirsten St. George, Daryl M. Lamson, Alexis Russel, Matthew Shudt, Melissa A Leisner, Jonathan Plitnick, Navjot Singh, John Kelly, Erasmus Schneider, Erica Lasek-Nesselquist |
| EPI_ISL_1228558, EPI_ISL_1228559, EPI_ISL_1228560 | MONTEFIORE MEDICAL CENTER LABORATORIES | Wadsworth Center, New York State Department of Health | Kirsten St. George, Daryl M. Lamson, Alexis Russel, Matthew Shudt, Melissa A Leisner, Jonathan Plitnick, Navjot Singh, John Kelly, Erasmus Schneider, Erica Lasek-Nesselquist |
| EPI_ISL_1228561, EPI_ISL_1228562 | WESTCHESTER MEDICAL CENTER | Wadsworth Center, New York State Department of Health | Kirsten St. George, Daryl M. Lamson, Alexis Russel, Matthew Shudt, Melissa A Leisner, Jonathan Plitnick, Navjot Singh, John Kelly, Erasmus Schneider, Erica Lasek-Nesselquist |
| EPI_ISL_1228563 | NYC Pandemic Response Lab | Wadsworth Center, New York State Department of Health | Kirsten St. George, Daryl M. Lamson, Alexis Russel, Matthew Shudt, Melissa A Leisner, Jonathan Plitnick, Navjot Singh, John Kelly, Erasmus Schneider, Erica Lasek-Nesselquist |
| EPI_ISL_1228564 | New York Presbyterian Hospital | Wadsworth Center, New York State Department of Health | Kirsten St. George, Daryl M. Lamson, Alexis Russel, Matthew Shudt, Melissa A Leisner, Jonathan Plitnick, Navjot Singh, John Kelly, Erasmus Schneider, Erica Lasek-Nesselquist |
| EPI_ISL_1228565, EPI_ISL_1228566 | WESTCHESTER MEDICAL CENTER | Wadsworth Center, New York State Department of Health | Kirsten St. George, Daryl M. Lamson, Alexis Russel, Matthew Shudt, Melissa A Leisner, Jonathan Plitnick, Navjot Singh, John Kelly, Erasmus Schneider, Erica Lasek-Nesselquist |
| EPI_ISL_1228567 | MONTEFIORE MEDICAL CENTER LABORATORIES | Wadsworth Center, New York State Department of Health | Kirsten St. George, Daryl M. Lamson, Alexis Russel, Matthew Shudt, Melissa A Leisner, Jonathan Plitnick, Navjot Singh, John Kelly, Erasmus Schneider, Erica Lasek-Nesselquist |
| EPI_ISL_1228568 | New York Presbyterian Hospital | Wadsworth Center, New York State Department of Health | Kirsten St. George, Daryl M. Lamson, Alexis Russel, Matthew Shudt, Melissa A Leisner, Jonathan Plitnick, Navjot Singh, John Kelly, Erasmus Schneider, Erica Lasek-Nesselquist |

| | | | |
|---|---|---|---|
| EPI_ISL_1228569 | MONTEFIORE MEDICAL CENTER LABORATORIES | Wadsworth Center, New York State Department of Health | Kirsten St. George, Daryl M. Lamson, Alexis Russel, Matthew Shudt, Melissa A Leisner, Jonathan Plitnick, Navjot Singh, John Kelly, Erasmus Schneider, Erica Lasek-Nesselquist |
| EPI_ISL_1228570 | NYC Pandemic Response Lab | Wadsworth Center, New York State Department of Health | Kirsten St. George, Daryl M. Lamson, Alexis Russel, Matthew Shudt, Melissa A Leisner, Jonathan Plitnick, Navjot Singh, John Kelly, Erasmus Schneider, Erica Lasek-Nesselquist |
| EPI_ISL_1228571 | WESTCHESTER MEDICAL CENTER | Wadsworth Center, New York State Department of Health | Kirsten St. George, Daryl M. Lamson, Alexis Russel, Matthew Shudt, Melissa A Leisner, Jonathan Plitnick, Navjot Singh, John Kelly, Erasmus Schneider, Erica Lasek-Nesselquist |
| EPI_ISL_1228572 | MONTEFIORE MEDICAL CENTER LABORATORIES | Wadsworth Center, New York State Department of Health | Kirsten St. George, Daryl M. Lamson, Alexis Russel, Matthew Shudt, Melissa A Leisner, Jonathan Plitnick, Navjot Singh, John Kelly, Erasmus Schneider, Erica Lasek-Nesselquist |
| EPI_ISL_1228573 | WESTCHESTER MEDICAL CENTER | Wadsworth Center, New York State Department of Health | Kirsten St. George, Daryl M. Lamson, Alexis Russel, Matthew Shudt, Melissa A Leisner, Jonathan Plitnick, Navjot Singh, John Kelly, Erasmus Schneider, Erica Lasek-Nesselquist |
| EPI_ISL_1228574, EPI_ISL_1228575 | NYC Pandemic Response Lab | Wadsworth Center, New York State Department of Health | Kirsten St. George, Daryl M. Lamson, Alexis Russel, Matthew Shudt, Melissa A Leisner, Jonathan Plitnick, Navjot Singh, John Kelly, Erasmus Schneider, Erica Lasek-Nesselquist |
| EPI_ISL_1228576 | Columbia University Irving Medical Center | Wadsworth Center, New York State Department of Health | Kirsten St. George, Daryl M. Lamson, Alexis Russel, Matthew Shudt, Melissa A Leisner, Jonathan Plitnick, Navjot Singh, John Kelly, Erasmus Schneider, Erica Lasek-Nesselquist |
| EPI_ISL_1228577 | WESTCHESTER MEDICAL CENTER | Wadsworth Center, New York State Department of Health | Kirsten St. George, Daryl M. Lamson, Alexis Russel, Matthew Shudt, Melissa A Leisner, Jonathan Plitnick, Navjot Singh, John Kelly, Erasmus Schneider, Erica Lasek-Nesselquist |
| EPI_ISL_1228578 | MONTEFIORE MEDICAL CENTER LABORATORIES | Wadsworth Center, New York State Department of Health | Kirsten St. George, Daryl M. Lamson, Alexis Russel, Matthew Shudt, Melissa A Leisner, Jonathan Plitnick, Navjot Singh, John Kelly, Erasmus Schneider, Erica Lasek-Nesselquist |
| EPI_ISL_1228579, EPI_ISL_1228580, EPI_ISL_1228581, EPI_ISL_1228582, EPI_ISL_1228583 | WESTCHESTER MEDICAL CENTER | Wadsworth Center, New York State Department of Health | Kirsten St. George, Daryl M. Lamson, Alexis Russel, Matthew Shudt, Melissa A Leisner, Jonathan Plitnick, Navjot Singh, John Kelly, Erasmus Schneider, Erica Lasek-Nesselquist |
| EPI_ISL_1228584 | MONTEFIORE MEDICAL CENTER LABORATORIES | Wadsworth Center, New York State Department of Health | Kirsten St. George, Daryl M. Lamson, Alexis Russel, Matthew Shudt, Melissa A Leisner, Jonathan Plitnick, Navjot Singh, John Kelly, Erasmus Schneider, Erica Lasek-Nesselquist |
| EPI_ISL_1228585, EPI_ISL_1228586 | WESTCHESTER MEDICAL CENTER | Wadsworth Center, New York State Department of Health | Kirsten St. George, Daryl M. Lamson, Alexis Russel, Matthew Shudt, Melissa A Leisner, Jonathan Plitnick, Navjot Singh, John Kelly, Erasmus Schneider, Erica Lasek-Nesselquist |
| EPI_ISL_1233139, EPI_ISL_1233182, EPI_ISL_1233183, EPI_ISL_1233184, EPI_ISL_1233185, EPI_ISL_1233186 | UW Virology Lab | UW Virology Lab | Pavitra Roychoudhury, Hong Xie, Lasata Shrestha, Shah Mohamed Bakhash, Michelle Lin, Margaret Mills, Noah Baker, Sean Ellis, Saraswathi Sathees, Meei-Li Huang, Keith R Jerome, Alexander Greninger |
| EPI_ISL_1252933, EPI_ISL_1252953, EPI_ISL_1252965, EPI_ISL_1252976, EPI_ISL_1253437, EPI_ISL_1254016, EPI_ISL_1254017, EPI_ISL_1254028, EPI_ISL_1254039, EPI_ISL_1254040, EPI_ISL_1254067, EPI_ISL_1254083, EPI_ISL_1254087, EPI_ISL_1254105, EPI_ISL_1254111, EPI_ISL_1254120, EPI_ISL_1254124, EPI_ISL_1254128, EPI_ISL_1254132, EPI_ISL_1254153, EPI_ISL_1254155, EPI_ISL_1254165, EPI_ISL_1254170, EPI_ISL_1254171, EPI_ISL_1254184, EPI_ISL_1254190, EPI_ISL_1254195, EPI_ISL_1254201, EPI_ISL_1254203, EPI_ISL_1254208, EPI_ISL_1254211, EPI_ISL_1254238, EPI_ISL_1254246, EPI_ISL_1254249, EPI_ISL_1254251, EPI_ISL_1254284, EPI_ISL_1254290, EPI_ISL_1254295, EPI_ISL_1254303, EPI_ISL_1254438, EPI_ISL_1254445, EPI_ISL_1254447 | | | |
| see above | Quest Diagnostics Incorporated | Centers for Disease Control and Prevention Division of Viral Diseases, Pathogen Discovery | Peter W. Cook, Dakota Howard, Dhwani Batra, Ben L. Rambo-Martin, S. H. Rosenthal, A. Gerasimova, R. M. Kagan, B. Anderson, M. Hua, Y. Liu, L.E. Bernstein, K.E. Livingston, A. Perez, I. A. Shlyakhter, R. V. Rolando, R. Owen, P. Tanpaiboon, F. Lacbawan, Clinton R. Paden, Suxiang Tong, Duncan MacCannell |
| EPI_ISL_1255301 | Laboratory Corporation of America | Respiratory Viruses Branch, Division of Viral Diseases, Centers for Disease Control and Prevention | Peter W. Cook, Dakota Howard, Dhwani Batra, Ben L. Rambo-Martin, Minoo Agarwal, Eyad Almasri Debbie Boles, Ayla Burns, Nuthawin Charoensri, Oren Cohen, Susan Countryman, Mary Ann Cristobal, Bobbi Croy, Suzanne Dale, Hrushikesh Deshmukh, Amanda Douglas, Vincent Drouillon, Marcia Eisenberg, Howard Engler, Rama Ghatti, Prashant Gupta, Susan Hicks, Jake Humphrey, Lax Iyer, Manoj Jain, Mohan Kolli, Brian Krueger, Tim Kuphal, Stanley Letovsky, Michael Levandoski, Craig Lukasik, Jonathan Meltzer, Brian Norvell, Mindy Nye, Scott Parker, Christos Petropoulos, John Pruitt, Steven Ragan, Scott Ryan, Mike Sapeta, Jana Schroth, Suresh Babu Selvaraju, Goran Stevovic, Amanda Suchanek, Andrea Throop, Lyndon Tilson, Thomas Urban, Joe Voshell, Kimberly Wagner, Jonathan Williams, Mary Williamson, Qian Zeng, Tricia Zwiefelhofer, Clinton R. Paden, Suxiang Tong, Duncan MacCannell |
| EPI_ISL_1255415 | Helix/Illumina | Respiratory Viruses Branch, Division of Viral Diseases, Centers for Disease Control and Prevention | Peter W. Cook, Dakota Howard, Dhwani Batra, Ben L. Rambo-Martin, Eileen de Feo, Jan Antico, Christine Tran, Matthew Tolentino, Shannon Wickline, Kim Gietzen, Brad Sickler, Jingtao Liu, Eric Allen, Phil Febbo, Summer Galloway, Nicole L. Washington, Simon White, Geraint Levan, Kelly Schiabor Barrett, Elizabeth Cirulli, Alexandre Bolze, Ary Ascencio, Charlotte Rivera-Garcia, Ryan Cho, Jason Nguyen, Sherry Wang, Jimmy Ramirez, Tyler Cassens, Efren Sandoval, Magnus Isaksson, William Lee, David Becker, Marc Laurent, James Lu, Clinton R. Paden, Suxiang Tong, Duncan MacCannell |
| EPI_ISL_1258466, EPI_ISL_1258467, EPI_ISL_1258468, EPI_ISL_1258469, EPI_ISL_1258470, EPI_ISL_1258471, EPI_ISL_1258472, EPI_ISL_1258473, EPI_ISL_1258474, EPI_ISL_1258475, EPI_ISL_1258476, EPI_ISL_1258477, EPI_ISL_1258478, EPI_ISL_1258479, EPI_ISL_1258480, EPI_ISL_1258481, EPI_ISL_1258482, EPI_ISL_1258483, EPI_ISL_1258484, EPI_ISL_1258485, EPI_ISL_1258486, EPI_ISL_1258487, EPI_ISL_1258488, EPI_ISL_1258489, EPI_ISL_1258490, EPI_ISL_1258491, EPI_ISL_1258492, EPI_ISL_1258493, EPI_ISL_1258494, EPI_ISL_1258495, EPI_ISL_1258496, EPI_ISL_1258497, EPI_ISL_1258498, EPI_ISL_1258499, EPI_ISL_1258500, EPI_ISL_1258501, EPI_ISL_1258502, EPI_ISL_1258503, EPI_ISL_1258504, EPI_ISL_1258505, EPI_ISL_1258506, EPI_ISL_1258507, EPI_ISL_1258508, EPI_ISL_1258509, EPI_ISL_1258510, EPI_ISL_1258511, EPI_ISL_1258512, EPI_ISL_1258513, EPI_ISL_1258514, EPI_ISL_1258515, EPI_ISL_1258516, EPI_ISL_1258517, EPI_ISL_1258518, EPI_ISL_1258519, EPI_ISL_1258520, EPI_ISL_1258521, EPI_ISL_1258522, EPI_ISL_1258523, EPI_ISL_1258524, EPI_ISL_1258525, EPI_ISL_1258526, EPI_ISL_1258527, EPI_ISL_1258528, EPI_ISL_1258529, EPI_ISL_1258530, EPI_ISL_1258531, EPI_ISL_1258532, EPI_ISL_1258533, EPI_ISL_1258534, EPI_ISL_1258535, EPI_ISL_1258536, EPI_ISL_1258537, EPI_ISL_1258538, EPI_ISL_1258539, EPI_ISL_1258540, EPI_ISL_1258541, EPI_ISL_1258542, EPI_ISL_1258543, EPI_ISL_1258544, EPI_ISL_1258545, EPI_ISL_1258546, EPI_ISL_1258547, EPI_ISL_1258548, EPI_ISL_1258549, EPI_ISL_1258550, EPI_ISL_1258551, EPI_ISL_1258552, EPI_ISL_1258553, EPI_ISL_1258554, EPI_ISL_1258555, EPI_ISL_1258556, EPI_ISL_1258557, EPI_ISL_1258558, EPI_ISL_1258559, EPI_ISL_1258560, EPI_ISL_1258561, EPI_ISL_1258562, EPI_ISL_1258563, EPI_ISL_1258564, EPI_ISL_1258565, EPI_ISL_1258566, EPI_ISL_1258567, EPI_ISL_1258568, EPI_ISL_1258569, EPI_ISL_1258570, EPI_ISL_1258571, EPI_ISL_1258572, EPI_ISL_1258573, EPI_ISL_1258574, EPI_ISL_1258575, EPI_ISL_1258576, EPI_ISL_1258577, EPI_ISL_1258578, EPI_ISL_1258579, EPI_ISL_1258580, EPI_ISL_1258581, EPI_ISL_1258582, EPI_ISL_1258583, EPI_ISL_1258584, EPI_ISL_1258585, EPI_ISL_1258586, EPI_ISL_1258587, EPI_ISL_1258588, EPI_ISL_1258589, EPI_ISL_1258590, EPI_ISL_1258591, EPI_ISL_1258592, EPI_ISL_1258593, EPI_ISL_1258594, EPI_ISL_1258595, EPI_ISL_1258596, EPI_ISL_1258597, EPI_ISL_1258598, EPI_ISL_1258599, EPI_ISL_1258600, EPI_ISL_1258601, EPI_ISL_1258602, EPI_ISL_1258603, EPI_ISL_1258604, EPI_ISL_1258605, EPI_ISL_1258606, EPI_ISL_1258607, EPI_ISL_1258608, EPI_ISL_1258609, EPI_ISL_1258610, EPI_ISL_1258611, EPI_ISL_1258612, EPI_ISL_1258613, EPI_ISL_1258614, EPI_ISL_1258615, EPI_ISL_1258616, EPI_ISL_1258617, EPI_ISL_1258618, EPI_ISL_1258619, EPI_ISL_1258620, EPI_ISL_1258621, EPI_ISL_1258622, EPI_ISL_1258623, EPI_ISL_1258624, EPI_ISL_1258625, EPI_ISL_1258626, EPI_ISL_1258627, EPI_ISL_1258628, EPI_ISL_1258629, EPI_ISL_1258630, EPI_ISL_1258631, EPI_ISL_1258632, EPI_ISL_1258633, EPI_ISL_1258634, EPI_ISL_1258635, EPI_ISL_1258636, EPI_ISL_1258637, EPI_ISL_1258638, EPI_ISL_1258639, EPI_ISL_1258640, EPI_ISL_1258641, EPI_ISL_1258642, EPI_ISL_1258643, EPI_ISL_1258644, EPI_ISL_1258645, EPI_ISL_1258646, EPI_ISL_1258647, EPI_ISL_1258648, EPI_ISL_1258649, EPI_ISL_1258650, EPI_ISL_1258651, EPI_ISL_1258652, EPI_ISL_1258653, EPI_ISL_1258654, EPI_ISL_1258655, EPI_ISL_1258656, EPI_ISL_1258657, EPI_ISL_1258658, EPI_ISL_1258659, EPI_ISL_1258660, EPI_ISL_1258661, EPI_ISL_1258662, EPI_ISL_1258663, EPI_ISL_1258664, EPI_ISL_1258665, EPI_ISL_1258666, EPI_ISL_1258667, EPI_ISL_1258668, EPI_ISL_1258669, EPI_ISL_1258670, EPI_ISL_1258671, EPI_ISL_1258672, EPI_ISL_1258673, EPI_ISL_1258674, EPI_ISL_1258675, EPI_ISL_1258676, EPI_ISL_1258677, EPI_ISL_1258678, EPI_ISL_1258679, EPI_ISL_1258680, EPI_ISL_1258681, EPI_ISL_1258682, EPI_ISL_1258683, EPI_ISL_1258684, EPI_ISL_1258685, EPI_ISL_1258686, EPI_ISL_1258687, EPI_ISL_1258688, EPI_ISL_1258689, EPI_ISL_1258690, EPI_ISL_1258691, EPI_ISL_1258692, EPI_ISL_1258693, EPI_ISL_1258694, EPI_ISL_1258695, EPI_ISL_1258696, EPI_ISL_1258697, EPI_ISL_1258698, EPI_ISL_1258699, EPI_ISL_1258700, EPI_ISL_1258701, EPI_ISL_1258702, EPI_ISL_1258703, EPI_ISL_1258704, EPI_ISL_1258705, EPI_ISL_1258706, EPI_ISL_1258707, EPI_ISL_1258708, EPI_ISL_1258709, EPI_ISL_1258710, EPI_ISL_1258711, EPI_ISL_1258712, EPI_ISL_1258713, EPI_ISL_1258714, EPI_ISL_1258715, EPI_ISL_1258716, EPI_ISL_1258717, EPI_ISL_1258718, EPI_ISL_1258719, EPI_ISL_1258720, EPI_ISL_1258721, EPI_ISL_1258722, EPI_ISL_1258723, EPI_ISL_1258724, EPI_ISL_1258725, EPI_ISL_1258726, EPI_ISL_1258727, EPI_ISL_1258728, EPI_ISL_1258729, EPI_ISL_1258730, EPI_ISL_1258731, EPI_ISL_1258732, EPI_ISL_1258733, EPI_ISL_1258734, EPI_ISL_1258735, EPI_ISL_1258736, EPI_ISL_1258737, EPI_ISL_1258738, EPI_ISL_1258739, EPI_ISL_1258740, EPI_ISL_1258741, EPI_ISL_1258742, EPI_ISL_1258743, EPI_ISL_1258744, EPI_ISL_1258745, EPI_ISL_1258746, EPI_ISL_1258747, EPI_ISL_1258748, EPI_ISL_1258749, EPI_ISL_1258750, EPI_ISL_1258751, EPI_ISL_1258752, EPI_ISL_1258753, EPI_ISL_1258754, EPI_ISL_1258755, EPI_ISL_1258756, EPI_ISL_1258757, EPI_ISL_1258758, EPI_ISL_1258759, EPI_ISL_1258760, EPI_ISL_1258761, EPI_ISL_1258762, EPI_ISL_1258763, EPI_ISL_1258764, EPI_ISL_1258765, EPI_ISL_1258766, EPI_ISL_1258767, EPI_ISL_1258768, EPI_ISL_1258769, EPI_ISL_1258770, EPI_ISL_1258771, EPI_ISL_1258772, EPI_ISL_1258773, EPI_ISL_1258774, EPI_ISL_1258775, EPI_ISL_1258776, EPI_ISL_1258777, EPI_ISL_1258778, EPI_ISL_1258779, EPI_ISL_1258780, EPI_ISL_1258781, EPI_ISL_1258782, EPI_ISL_1258783, EPI_ISL_1258784, EPI_ISL_1258785, EPI_ISL_1258786, EPI_ISL_1258787, EPI_ISL_1258788, EPI_ISL_1258789, EPI_ISL_1258790, EPI_ISL_1258791, EPI_ISL_1258792, EPI_ISL_1258793, EPI_ISL_1258794, EPI_ISL_1258795, EPI_ISL_1258796, EPI_ISL_1258797, EPI_ISL_1258798, EPI_ISL_1258799, EPI_ISL_1258800, EPI_ISL_1258801, EPI_ISL_1258802, EPI_ISL_1258803, EPI_ISL_1258804, EPI_ISL_1258805, EPI_ISL_1258806, EPI_ISL_1258807, EPI_ISL_1258808, EPI_ISL_1258809, EPI_ISL_1258810, EPI_ISL_1258811, EPI_ISL_1258812, EPI_ISL_1258813, EPI_ISL_1258814, EPI_ISL_1258815, EPI_ISL_1258816, EPI_ISL_1258817, EPI_ISL_1258818, EPI_ISL_1258819, EPI_ISL_1258820, EPI_ISL_1258821, EPI_ISL_1258822, EPI_ISL_1258823, EPI_ISL_1258824, EPI_ISL_1258825, EPI_ISL_1258826, EPI_ISL_1258827, EPI_ISL_1258828, EPI_ISL_1258829, EPI_ISL_1258830, EPI_ISL_1258831, EPI_ISL_1258832, EPI_ISL_1258833, EPI_ISL_1258834, EPI_ISL_1258835, EPI_ISL_1258836, EPI_ISL_1258837, EPI_ISL_1258838, EPI_ISL_1258839, EPI_ISL_1258840, EPI_ISL_1258841, EPI_ISL_1258842, EPI_ISL_1258843, EPI_ISL_1258844, EPI_ISL_1258845, EPI_ISL_1258846, EPI_ISL_1258847, EPI_ISL_1258848, EPI_ISL_1258849, EPI_ISL_1258850, EPI_ISL_1258851, EPI_ISL_1258852, EPI_ISL_1258853, EPI_ISL_1258854, EPI_ISL_1258855, EPI_ISL_1258856, EPI_ISL_1258857, EPI_ISL_1258858, EPI_ISL_1258859, EPI_ISL_1258860, EPI_ISL_1258861, EPI_ISL_1258862, EPI_ISL_1258863, EPI_ISL_1258864, EPI_ISL_1258865, EPI_ISL_1258866, EPI_ISL_1258867, EPI_ISL_1258868, EPI_ISL_1258869, EPI_ISL_1258870, EPI_ISL_1258871, EPI_ISL_1258872, EPI_ISL_1258873, EPI_ISL_1258874, EPI_ISL_1258875, EPI_ISL_1258876, EPI_ISL_1258877, EPI_ISL_1258878, EPI_ISL_1258879, EPI_ISL_1258880, EPI_ISL_1258881, EPI_ISL_1258882, EPI_ISL_1258883, EPI_ISL_1258884, EPI_ISL_1258885, EPI_ISL_1258886, EPI_ISL_1258887, EPI_ISL_1258888, EPI_ISL_1258889, EPI_ISL_1258890, EPI_ISL_1258891, EPI_ISL_1258892, EPI_ISL_1258893, EPI_ISL_1258894, EPI_ISL_1258895, EPI_ISL_1258896, EPI_ISL_1258897, EPI_ISL_1258898, EPI_ISL_1258899, EPI_ISL_1258900, EPI_ISL_1258901, EPI_ISL_1258902, EPI_ISL_1258903, EPI_ISL_1258904, EPI_ISL_1258905, EPI_ISL_1258906, EPI_ISL_1258907, EPI_ISL_1258908, EPI_ISL_1258909, EPI_ISL_1258910, EPI_ISL_1258911, EPI_ISL_1258912, EPI_ISL_1258913, EPI_ISL_1258914, EPI_ISL_1258915, EPI_ISL_1258916, EPI_ISL_1258917, EPI_ISL_1258918, EPI_ISL_1258919, EPI_ISL_1258920, EPI_ISL_1258921, EPI_ISL_1258922, EPI_ISL_1258923, EPI_ISL_1258924, EPI_ISL_1258925, EPI_ISL_1258926, EPI_ISL_1258927, EPI_ISL_1258928, EPI_ISL_1258929, EPI_ISL_1258930, EPI_ISL_1258931, EPI_ISL_1258932, EPI_ISL_1258933, EPI_ISL_1258934, EPI_ISL_1258935, EPI_ISL_1258936, EPI_ISL_1258937, EPI_ISL_1258938, EPI_ISL_1258939, EPI_ISL_1258940, EPI_ISL_1258941, EPI_ISL_1258942, EPI_ISL_1258943, EPI_ISL_1258944, EPI_ISL_1258945, EPI_ISL_1258946, EPI_ISL_1258947, EPI_ISL_1258948, EPI_ISL_1258949, EPI_ISL_1258950, EPI_ISL_1258951, EPI_ISL_1258952, EPI_ISL_1258953, EPI_ISL_1258954, EPI_ISL_1258955, EPI_ISL_1258956, EPI_ISL_1258957, EPI_ISL_1258958, EPI_ISL_1258959, EPI_ISL_1258960, EPI_ISL_1258961, EPI_ISL_1258962, EPI_ISL_1258963, EPI_ISL_1258964, EPI_ISL_1258965, EPI_ISL_1258966, EPI_ISL_1258967, EPI_ISL_1258968, EPI_ISL_1258969, EPI_ISL_1258970, EPI_ISL_1258971, EPI_ISL_1258972, EPI_ISL_1258973, EPI_ISL_1258974, EPI_ISL_1258975, EPI_ISL_1258976, EPI_ISL_1258977, EPI_ISL_1258978, EPI_ISL_1258979, EPI_ISL_1258980, EPI_ISL_1258981, EPI_ISL_1258982, EPI_ISL_1258983, EPI_ISL_1258984, EPI_ISL_1258985, EPI_ISL_1258986, EPI_ISL_1258987 | | | |
| see above | Pandemic Response Lab - NYC | Pandemic Response Lab, R&D | Henry Lee, Michael Hammerling, Melissa Hopkins, Cybill del Castillo, Shinyoung Clair Kang, William Ward, Pradeep Bugga, Haiping Hao, Jon Laurent |
| EPI_ISL_1261790, EPI_ISL_1261791, | DOHMH Jamaica | New York City Public Health Laboratory | Jade Wang, et al. |

| | | | |
|---|---|---|---|
| EPI_ISL_1261792, EPI_ISL_1261793, EPI_ISL_1261794 | | | |
| EPI_ISL_1261795 | DOHMH Riverside | New York City Public Health Laboratory | Jade Wang, et al. |
| EPI_ISL_1261796, EPI_ISL_1261797 | OCME Office Of Chief Medical Examiner | New York City Public Health Laboratory | Jade Wang, et al. |
| EPI_ISL_1261798 | DOHMH PHL | New York City Public Health Laboratory | Jade Wang, et al. |
| EPI_ISL_1261799 | DOHMH Riverside | New York City Public Health Laboratory | Jade Wang, et al. |
| EPI_ISL_1261800 | Department of Homeless Services | New York City Public Health Laboratory | Jade Wang, et al. |
| EPI_ISL_1261801 | DOHMH Central Harlem | New York City Public Health Laboratory | Jade Wang, et al. |
| EPI_ISL_1261802 | DOHMH Jamaica | New York City Public Health Laboratory | Jade Wang, et al. |
| EPI_ISL_1261803 | DOHMH Crown Heights | New York City Public Health Laboratory | Jade Wang, et al. |
| EPI_ISL_1261804 | DOHMH Central Harlem | New York City Public Health Laboratory | Jade Wang, et al. |
| EPI_ISL_1261805 | DOHMH PHL | New York City Public Health Laboratory | Jade Wang, et al. |
| EPI_ISL_1261806 | DOHMH Corona | New York City Public Health Laboratory | Jade Wang, et al. |
| EPI_ISL_1261807, EPI_ISL_1261808 | DOHMH Central Harlem | New York City Public Health Laboratory | Jade Wang, et al. |
| EPI_ISL_1261809 | DOHMH Fort Greene | New York City Public Health Laboratory | Jade Wang, et al. |
| EPI_ISL_1261810 | DOHMH Corona | New York City Public Health Laboratory | Jade Wang, et al. |
| EPI_ISL_1261811 | OCME Office Of Chief Medical Examiner | New York City Public Health Laboratory | Jade Wang, et al. |
| EPI_ISL_1261812, EPI_ISL_1261813 | DOHMH Corona | New York City Public Health Laboratory | Jade Wang, et al. |
| EPI_ISL_1261814 | DOHMH Morrisania | New York City Public Health Laboratory | Jade Wang, et al. |
| EPI_ISL_1261815 | DOHMH Jamaica | New York City Public Health Laboratory | Jade Wang, et al. |
| EPI_ISL_1261816 | DOHMH Riverside | New York City Public Health Laboratory | Jade Wang, et al. |
| EPI_ISL_1261817 | DOHMH Chelsea | New York City Public Health Laboratory | Jade Wang, et al. |
| EPI_ISL_1261818 | DOHMH Fort Greene | New York City Public Health Laboratory | Jade Wang, et al. |
| EPI_ISL_1261819 | OCME Office Of Chief Medical Examiner | New York City Public Health Laboratory | Jade Wang, et al. |
| EPI_ISL_1261820 | DOHMH Jamaica | New York City Public Health Laboratory | Jade Wang, et al. |
| EPI_ISL_1261821 | DOHMH Crown Heights | New York City Public Health Laboratory | Jade Wang, et al. |
| EPI_ISL_1261822 | DOHMH Jamaica | New York City Public Health Laboratory | Jade Wang, et al. |
| EPI_ISL_1261823 | DOHMH PHL | New York City Public Health Laboratory | Jade Wang, et al. |
| EPI_ISL_1261824 | DOHMH Central Harlem | New York City Public Health Laboratory | Jade Wang, et al. |
| EPI_ISL_1261825 | DOHMH Crown Heights | New York City Public Health Laboratory | Jade Wang, et al. |
| EPI_ISL_1261826 | DOHMH Morrisania | New York City Public Health Laboratory | Jade Wang, et al. |
| EPI_ISL_1261827, EPI_ISL_1261828, EPI_ISL_1261829 | OCME Office Of Chief Medical Examiner | New York City Public Health Laboratory | Jade Wang, et al. |
| EPI_ISL_1261830 | Department of Homeless Services | New York City Public Health Laboratory | Jade Wang, et al. |
| EPI_ISL_1261831 | DOHMH Jamaica | New York City Public Health Laboratory | Jade Wang, et al. |
| EPI_ISL_1261832, EPI_ISL_1261833 | DOHMH Corona | New York City Public Health Laboratory | Jade Wang, et al. |
| EPI_ISL_1261834 | DOHMH Crown Heights | New York City Public Health Laboratory | Jade Wang, et al. |
| EPI_ISL_1261835, EPI_ISL_1261836, EPI_ISL_1261837 | DOHMH Corona | New York City Public Health Laboratory | Jade Wang, et al. |
| EPI_ISL_1261838, EPI_ISL_1261839 | DOHMH Morrisania | New York City Public Health Laboratory | Jade Wang, et al. |
| EPI_ISL_1261840, EPI_ISL_1261841, EPI_ISL_1261842 | DOHMH Riverside | New York City Public Health Laboratory | Jade Wang, et al. |
| EPI_ISL_1261843 | DOHMH Central Harlem | New York City Public Health Laboratory | Jade Wang, et al. |
| EPI_ISL_1261844 | DOHMH Corona | New York City Public Health Laboratory | Jade Wang, et al. |
| EPI_ISL_1261845 | DOHMH Riverside | New York City Public Health Laboratory | Jade Wang, et al. |
| EPI_ISL_1261846 | OCME Office Of Chief Medical Examiner | New York City Public Health Laboratory | Jade Wang, et al. |
| EPI_ISL_1261847 | DOHMH Fort Greene | New York City Public Health Laboratory | Jade Wang, et al. |
| EPI_ISL_1261848 | DOHMH Morrisania | New York City Public Health Laboratory | Jade Wang, et al. |
| EPI_ISL_1261849 | DOHMH Riverside | New York City Public Health Laboratory | Jade Wang, et al. |
| EPI_ISL_1261850 | DOHMH Fort Greene | New York City Public Health Laboratory | Jade Wang, et al. |
| EPI_ISL_1261851 | DOHMH Morrisania | New York City Public Health Laboratory | Jade Wang, et al. |
| EPI_ISL_1261852 | DOHMH Chelsea | New York City Public Health Laboratory | Jade Wang, et al. |
| EPI_ISL_1261853 | Department of Homeless Services | New York City Public Health Laboratory | Jade Wang, et al. |
| EPI_ISL_1261854 | OCME Office Of Chief Medical Examiner | New York City Public Health Laboratory | Jade Wang, et al. |
| EPI_ISL_1261855 | DOHMH Chelsea | New York City Public Health Laboratory | Jade Wang, et al. |
| EPI_ISL_1261856 | DOHMH Central Harlem | New York City Public Health Laboratory | Jade Wang, et al. |
| EPI_ISL_1261857 | DOHMH Jamaica | New York City Public Health Laboratory | Jade Wang, et al. |
| EPI_ISL_1261858 | DOHMH Central Harlem | New York City Public Health Laboratory | Jade Wang, et al. |
| EPI_ISL_1261859 | DOHMH Corona | New York City Public Health Laboratory | Jade Wang, et al. |
| EPI_ISL_1261860 | DOHMH Jamaica | New York City Public Health Laboratory | Jade Wang, et al. |

| | | | |
|---|---|---|---|
| EPI_ISL_1261861 | DOHMH Crown Heights | New York City Public Health Laboratory | Jade Wang, et al. |
| EPI_ISL_1261862, EPI_ISL_1261863 | DOHMH Chelsea | New York City Public Health Laboratory | Jade Wang, et al. |
| EPI_ISL_1261864 | DOHMH Central Harlem | New York City Public Health Laboratory | Jade Wang, et al. |
| EPI_ISL_1261865 | OCME Office Of Chief Medical Examiner | New York City Public Health Laboratory | Jade Wang, et al. |
| EPI_ISL_1261866 | DOHMH Jamaica | New York City Public Health Laboratory | Jade Wang, et al. |
| EPI_ISL_1261867 | Department of Homeless Services | New York City Public Health Laboratory | Jade Wang, et al. |
| EPI_ISL_1261929, EPI_ISL_1261938 | Helix/Illumina | Centers for Disease Control and Prevention Division of Viral Diseases, Pathogen Discovery | Peter W. Cook, Dakota Howard, Dhwani Batra, Ben L. Rambo-Martin, Eileen de Feo, Jan Antico, Christine Tran, Matthew Tolentino, Shannon Wickline, Kim Gietzen, Brad Sickler, Jingtao Liu, Eric Allen, Phil Febbo, Summer Galloway, Nicole L. Washington, Simon White, Geraint Levan, Kelly Schiabor Barrett, Elizabeth Cirulli, Alexandre Bolze, Ary Ascencio, Charlotte Rivera-Garcia, Ryan Cho, Jason Nguyen, Sherry Wang, Jimmy Ramirez, Tyler Cassens, Efren Sandoval, Magnus Isaksson, William Lee, David Becker, Marc Laurent, James Lu, Clinton R. Paden, Suxiang Tong, Duncan MacCannell |
| EPI_ISL_1262501, EPI_ISL_1262502, EPI_ISL_1262503, EPI_ISL_1262504, EPI_ISL_1262505, EPI_ISL_1262506, EPI_ISL_1262507, EPI_ISL_1262508, EPI_ISL_1262509, EPI_ISL_1262510, EPI_ISL_1262511, EPI_ISL_1262512, EPI_ISL_1262513, EPI_ISL_1262514, EPI_ISL_1262515, EPI_ISL_1262516, EPI_ISL_1262517, EPI_ISL_1262518, EPI_ISL_1262519, EPI_ISL_1262520, EPI_ISL_1262521, EPI_ISL_1262522, EPI_ISL_1262523, EPI_ISL_1262524, EPI_ISL_1262525, EPI_ISL_1262526, EPI_ISL_1262527, EPI_ISL_1262528, EPI_ISL_1262529, EPI_ISL_1262530, EPI_ISL_1262531, EPI_ISL_1262532, EPI_ISL_1262533, EPI_ISL_1262534, EPI_ISL_1262535, EPI_ISL_1262536, EPI_ISL_1262537, EPI_ISL_1262538, EPI_ISL_1262539, EPI_ISL_1262540, EPI_ISL_1262541, EPI_ISL_1262542, EPI_ISL_1262543, EPI_ISL_1262544, EPI_ISL_1262545, EPI_ISL_1262546, EPI_ISL_1262547, EPI_ISL_1262548, EPI_ISL_1262549, EPI_ISL_1262550, EPI_ISL_1262551, EPI_ISL_1262552, EPI_ISL_1262553, EPI_ISL_1262554, EPI_ISL_1262555, EPI_ISL_1262556, EPI_ISL_1262557, EPI_ISL_1262558, EPI_ISL_1262559, EPI_ISL_1262560, EPI_ISL_1262561, EPI_ISL_1262562, EPI_ISL_1262563, EPI_ISL_1262564, EPI_ISL_1262565, EPI_ISL_1262566, EPI_ISL_1262567, EPI_ISL_1262568, EPI_ISL_1262569, EPI_ISL_1262570, EPI_ISL_1262571, EPI_ISL_1262572, EPI_ISL_1262573, EPI_ISL_1262574, EPI_ISL_1262575, EPI_ISL_1262576, EPI_ISL_1262577, EPI_ISL_1262578, EPI_ISL_1262579, EPI_ISL_1262580, EPI_ISL_1262581, EPI_ISL_1262582, EPI_ISL_1262583, EPI_ISL_1262584, EPI_ISL_1262585 | | | |
| see above | NYU Langone Health | Departments of Pathology and Medicine, New York University School of Medicine | Adriana Heguy, Dacia Dimartino, Emily Guzman, Christian Marier, Peter Meyn, Sitharam Ramaswami, Gael Westby, Paul Zappile, Yutong Zhang, Paolo Cotzia, Guiqing Wang |
| EPI_ISL_1265950, EPI_ISL_1265965, EPI_ISL_1265968, EPI_ISL_1265969, EPI_ISL_1265988, EPI_ISL_1265999, EPI_ISL_1266188, EPI_ISL_1266559, EPI_ISL_1266584, EPI_ISL_1266796, EPI_ISL_1266804, EPI_ISL_1267348, EPI_ISL_1267379, EPI_ISL_1267456 | | | |
| see above | Quest Diagnostics Incorporated | Centers for Disease Control and Prevention Division of Viral Diseases, Pathogen Discovery | Peter W. Cook, Dakota Howard, Dhwani Batra, Ben L. Rambo-Martin, S. H. Rosenthal, A. Gerasimova, R. M. Kagan, B. Anderson, M. Hua, Y. Liu, L.E. Bernstein, K.E. Livingston, A. Perez, I. A. Shlyakhter, R. V. Rolando, R. Owen, P. Tanpaiboon, F. Lacbawan, Clinton R. Paden, Suxiang Tong, Duncan MacCannell |
| EPI_ISL_1268329, EPI_ISL_1268980, EPI_ISL_1268992, EPI_ISL_1269067, EPI_ISL_1270016, EPI_ISL_1270051, EPI_ISL_1270165, EPI_ISL_1270213 | Helix/Illumina | Centers for Disease Control and Prevention Division of Viral Diseases, Pathogen Discovery | Peter W. Cook, Dakota Howard, Dhwani Batra, Ben L. Rambo-Martin, Eileen de Feo, Jan Antico, Christine Tran, Matthew Tolentino, Shannon Wickline, Kim Gietzen, Brad Sickler, Jingtao Liu, Eric Allen, Phil Febbo, Summer Galloway, Nicole L. Washington, Simon White, Geraint Levan, Kelly Schiabor Barrett, Elizabeth Cirulli, Alexandre Bolze, Ary Ascencio, Charlotte Rivera-Garcia, Ryan Cho, Jason Nguyen, Sherry Wang, Jimmy Ramirez, Tyler Cassens, Efren Sandoval, Magnus Isaksson, William Lee, David Becker, Marc Laurent, James Lu, Clinton R. Paden, Suxiang Tong, Duncan MacCannell |
| EPI_ISL_1272922, EPI_ISL_1272923, EPI_ISL_1272924, EPI_ISL_1272925, EPI_ISL_1272926, EPI_ISL_1272927 | NYSDOH Wadsworth Center, Virology Lab | Centers for Disease Control and Prevention Division of Viral Diseases, Pathogen Discovery | Krista Queen, Yan Li, Ying Tao, Jing Zhang, Anna Uehara, Anna Montmayeur, Clinton R. Paden, Kristen Knipe, Matthew Schmerer, Shoshona Le, Katie Dillon, Peter W. Cook, Rachel Marine, Mili Sheth, Jasmine Padilla, Sarah Nobles, Mark Burroughs, Lori Rowe, Haibin Wang, Ben L. Rambo-Martin, Kristine Lacek, Sam Shepard, Dhwani Batra, Suxiang Tong, Justin Lee |
| EPI_ISL_1273043, EPI_ISL_1273044, EPI_ISL_1273045, EPI_ISL_1273046, EPI_ISL_1273047 | NYC Department of Health and Mental Hygiene | Centers for Disease Control and Prevention Division of Viral Diseases, Pathogen Discovery | Krista Queen, Yan Li, Ying Tao, Jing Zhang, Anna Uehara, Anna Montmayeur, Clinton R. Paden, Kristen Knipe, Matthew Schmerer, Shoshona Le, Katie Dillon, Peter W. Cook, Rachel Marine, Mili Sheth, Jasmine Padilla, Sarah Nobles, Mark Burroughs, Lori Rowe, Haibin Wang, Ben L. Rambo-Martin, Kristine Lacek, Sam Shepard, Dhwani Batra, Suxiang Tong, Justin Lee |
| EPI_ISL_1290859 | Quest Diagnostics Incorporated | Respiratory Viruses Branch, Division of Viral Diseases, Centers for Disease Control and Prevention | Peter W. Cook, Dakota Howard, Dhwani Batra, Ben L. Rambo-Martin, S. H. Rosenthal, A. Gerasimova, R. M. Kagan, B. Anderson, M. Hua, Y. Liu, L.E. Bernstein, K.E. Livingston, A. Perez, I. A. Shlyakhter, R. V. Rolando, R. Owen, P. Tanpaiboon, F. Lacbawan, Clinton R. Paden, Suxiang Tong, Duncan MacCannell |
| EPI_ISL_1291205, EPI_ISL_1291217, EPI_ISL_1291248, EPI_ISL_1291252, EPI_ISL_1291253 | Laboratory Corporation of America | Centers for Disease Control and Prevention Division of Viral Diseases, Pathogen Discovery | Peter W. Cook, Dakota Howard, Dhwani Batra, Ben L. Rambo-Martin, Minoo Agarwal, Eyad Almasri Debbie Boles, Ayla Burns, Nuthawin Charoensri, Oren Cohen, Susan Countryman, Mary Ann Cristobal, Bobbi Croy, Suzanne Dale, Hrushikesh Deshmukh, Amanda Douglas, Vincent Drouillon, Marcia Eisenberg, Howard Engler, Rama Ghatti, Prashant Gupta, Susan Hicks, Jake Humphrey, Lax Iyer, Manoj Jain, Mohan Kolli, Brian Krueger, Tim Kuphal, Stanley Letovsky, Michael Levandoski, Craig Lukasik, Jonathan Meltzer, Brian Norvell, Mindy Nye, Scott Parker, Christos Petropoulos, John Pruitt, Steven Ragan, Scott Ryan, Mike Sapeta, Jana Schroth, Suresh Babu Selvaraju, Goran Stevovic, Amanda Suchanek, Andrea Throop, Lyndon Tilson, Thomas Urban, Joe Voshell, Kimberly Wagner, Jonathan Williams, Mary Williamson, Qian Zeng, Tricia Zwiefelhofer, Clinton R. Paden, Suxiang Tong, Duncan MacCannell |
| EPI_ISL_1291532, EPI_ISL_1291558, EPI_ISL_1291560, EPI_ISL_1291564 | Quest Diagnostics Incorporated | Centers for Disease Control and Prevention Division of Viral Diseases, Pathogen Discovery | Peter W. Cook, Dakota Howard, Dhwani Batra, Ben L. Rambo-Martin, S. H. Rosenthal, A. Gerasimova, R. M. Kagan, B. Anderson, M. Hua, Y. Liu, L.E. Bernstein, K.E. Livingston, A. Perez, I. A. Shlyakhter, R. V. Rolando, R. Owen, P. Tanpaiboon, F. Lacbawan, Clinton R. Paden, Suxiang Tong, Duncan MacCannell |
| EPI_ISL_1292992 | Yale Clinical Virology Lab | Grubaugh Lab - Yale School of Public Health | Joseph Fauver, Mallery Breban, Isabell Ott, Tara Alpert, Mary Petrone, Anderson Brito, Chantal Vogels, Annie Watkins, Chaney Kalinich, Jessica Rothman, Marie L. Landry, Nathan Grubaugh |
| EPI_ISL_1293138 | Murphy Medical Association | Grubaugh Lab - Yale School of Public Health | Joseph Fauver, Mallery Breban, Isabell Ott, Tara Alpert, Mary Petrone, Anderson Brito, Chantal Vogels, Annie Watkins, Chaney Kalinich, Jessica Rothman, Caleb Neal, Eva Laszlo, Steven Murphy, Nathan Grubaugh |
| EPI_ISL_1297766, EPI_ISL_1297769, EPI_ISL_1298226, EPI_ISL_1298231, EPI_ISL_1298234, EPI_ISL_1298242 | Quest Diagnostics Incorporated | Centers for Disease Control and Prevention Division of Viral Diseases, Pathogen Discovery | Peter W. Cook, Dakota Howard, Dhwani Batra, Ben L. Rambo-Martin, S. H. Rosenthal, A. Gerasimova, R. M. Kagan, B. Anderson, M. Hua, Y. Liu, L.E. Bernstein, K.E. Livingston, A. Perez, I. A. Shlyakhter, R. V. Rolando, R. Owen, P. Tanpaiboon, F. Lacbawan, Clinton R. Paden, Suxiang Tong, Duncan MacCannell |
| EPI_ISL_1298252, EPI_ISL_1298253, EPI_ISL_1298306, EPI_ISL_1298327, EPI_ISL_1298354, EPI_ISL_1298361 | Laboratory Corporation of America | Centers for Disease Control and Prevention Division of Viral Diseases, Pathogen Discovery | Peter W. Cook, Dakota Howard, Dhwani Batra, Ben L. Rambo-Martin, Minoo Agarwal, Eyad Almasri, Debbie Boles, Ayla Burns, Nuthawin Charoensri, Oren Cohen, Susan Countryman, Mary Ann Cristobal, Bobbi Croy, Suzanne Dale, Hrushikesh Deshmukh, Amanda Douglas, Vincent Drouillon, Marcia Eisenberg, Howard Engler, Rama Ghatti, Prashant Gupta, Susan Hicks, Jake Humphrey, Lax Iyer, Manoj Jain, Mohan Kolli, Brian Krueger, Tim Kuphal, Stanley Letovsky, Michael Levandoski, Craig Lukasik, Jonathan Meltzer, Brian Norvell, Mindy Nye, Scott Parker, Christos Petropoulos, John Pruitt, Steven Ragan, Scott Ryan, Mike Sapeta, Jana Schroth, Suresh Babu Selvaraju, Goran Stevovic, Amanda Suchanek, Andrea Throop, Lyndon Tilson, Thomas Urban, Joe Voshell, Kimberly Wagner, Jonathan Williams, Mary Williamson, Qian Zeng, Tricia Zwiefelhofer, Clinton R. Paden, Suxiang Tong, Duncan MacCannell |
| EPI_ISL_1300175, EPI_ISL_1300176, EPI_ISL_1300177, EPI_ISL_1300178, EPI_ISL_1300179, EPI_ISL_1300180, EPI_ISL_1300181, EPI_ISL_1300183, EPI_ISL_1300184, EPI_ISL_1300185, EPI_ISL_1300186, EPI_ISL_1300187, EPI_ISL_1300188, EPI_ISL_1300189, EPI_ISL_1300190, EPI_ISL_1300191, EPI_ISL_1300192, EPI_ISL_1300193, EPI_ISL_1300194, EPI_ISL_1300195, EPI_ISL_1300196, EPI_ISL_1300197, EPI_ISL_1300198, EPI_ISL_1300199, EPI_ISL_1300200, EPI_ISL_1300201, EPI_ISL_1300202, EPI_ISL_1300203, EPI_ISL_1300204, EPI_ISL_1300205, EPI_ISL_1300206, EPI_ISL_1300207, EPI_ISL_1300208, EPI_ISL_1300209, EPI_ISL_1300210, EPI_ISL_1300211, EPI_ISL_1300212, EPI_ISL_1300213, EPI_ISL_1300214, EPI_ISL_1300215, EPI_ISL_1300216, EPI_ISL_1300217, EPI_ISL_1300218, EPI_ISL_1300219, EPI_ISL_1300220, EPI_ISL_1300221, EPI_ISL_1300222, EPI_ISL_1300223, EPI_ISL_1300224, EPI_ISL_1300225, EPI_ISL_1300226, EPI_ISL_1300227, EPI_ISL_1300228, EPI_ISL_1300229, EPI_ISL_1300230, EPI_ISL_1300231, EPI_ISL_1300232, EPI_ISL_1300233, EPI_ISL_1300234, EPI_ISL_1300235, EPI_ISL_1300236, EPI_ISL_1300237, EPI_ISL_1300238, EPI_ISL_1300239, EPI_ISL_1300240, EPI_ISL_1300241, EPI_ISL_1300242, EPI_ISL_1300243, EPI_ISL_1300244, EPI_ISL_1300245, EPI_ISL_1300246, EPI_ISL_1300247, EPI_ISL_1300248, EPI_ISL_1300249, EPI_ISL_1300250, EPI_ISL_1300251, EPI_ISL_1300252, EPI_ISL_1300253, EPI_ISL_1300254, EPI_ISL_1300255, EPI_ISL_1300256, EPI_ISL_1300257, EPI_ISL_1300258, EPI_ISL_1300259, EPI_ISL_1300261, EPI_ISL_1300262, EPI_ISL_1300263, EPI_ISL_1300264, EPI_ISL_1300265, EPI_ISL_1300266, EPI_ISL_1300267, EPI_ISL_1300268, EPI_ISL_1300269, EPI_ISL_1300270, EPI_ISL_1300271, EPI_ISL_1300272, EPI_ISL_1300273, EPI_ISL_1300274, EPI_ISL_1300275, EPI_ISL_1300276 | | | |
| see above | NYU Langone Health | Departments of Pathology and Medicine, New York University School of Medicine | Adriana Heguy, Dacia Dimartino, Emily Guzman, Christian Marier, Peter Meyn, Sitharam Ramaswami, Gael Westby, Paul Zappile, Yutong Zhang, Paolo Cotzia, Guiqing Wang |
| EPI_ISL_1300756, EPI_ISL_1300757, EPI_ISL_1300758, EPI_ISL_1300759, EPI_ISL_1300760, EPI_ISL_1300761, EPI_ISL_1300762, EPI_ISL_1300763, EPI_ISL_1300764, EPI_ISL_1300765, EPI_ISL_1300766, EPI_ISL_1300767, EPI_ISL_1300768, EPI_ISL_1300769, EPI_ISL_1300770, EPI_ISL_1300771, EPI_ISL_1300772, EPI_ISL_1300773, EPI_ISL_1300774, EPI_ISL_1300776, EPI_ISL_1300777, EPI_ISL_1300778, EPI_ISL_1300779, EPI_ISL_1300780, EPI_ISL_1300781, EPI_ISL_1300782, EPI_ISL_1300783, EPI_ISL_1300784, EPI_ISL_1300785, EPI_ISL_1300786, EPI_ISL_1300787, EPI_ISL_1300788, EPI_ISL_1300789, EPI_ISL_1300790, EPI_ISL_1300791, EPI_ISL_1300792, EPI_ISL_1300793, EPI_ISL_1300794, EPI_ISL_1300795, EPI_ISL_1300796, EPI_ISL_1300797, EPI_ISL_1300798, EPI_ISL_1300799, EPI_ISL_1300800, EPI_ISL_1300801, EPI_ISL_1300802, EPI_ISL_1300803, EPI_ISL_1300804, EPI_ISL_1300805, EPI_ISL_1300806, EPI_ISL_1300807, EPI_ISL_1300808, EPI_ISL_1300809, EPI_ISL_1300810, EPI_ISL_1300811, EPI_ISL_1300812, EPI_ISL_1300813, EPI_ISL_1300814, EPI_ISL_1300815, EPI_ISL_1300816, EPI_ISL_1300817, EPI_ISL_1300818, EPI_ISL_1300819, EPI_ISL_1300820, EPI_ISL_1300821, EPI_ISL_1300822, EPI_ISL_1300823, EPI_ISL_1300824, EPI_ISL_1300825, EPI_ISL_1300826, EPI_ISL_1300827, EPI_ISL_1300828, EPI_ISL_1300829, EPI_ISL_1300830, EPI_ISL_1300831, EPI_ISL_1300832, EPI_ISL_1300833, EPI_ISL_1300834, EPI_ISL_1300835, EPI_ISL_1300836, EPI_ISL_1300837, EPI_ISL_1300838, EPI_ISL_1300839, EPI_ISL_1300840, EPI_ISL_1300841, EPI_ISL_1300842, EPI_ISL_1300843, EPI_ISL_1300844, EPI_ISL_1300845, EPI_ISL_1300846, EPI_ISL_1300847, EPI_ISL_1300848, EPI_ISL_1300849, EPI_ISL_1300850, EPI_ISL_1300851, EPI_ISL_1300852, EPI_ISL_1300853, EPI_ISL_1300854, EPI_ISL_1300856, EPI_ISL_1300857, EPI_ISL_1300858, EPI_ISL_1300859, EPI_ISL_1300860, EPI_ISL_1300861, EPI_ISL_1300862, EPI_ISL_1300863, EPI_ISL_1300864, EPI_ISL_1300865, EPI_ISL_1300866, EPI_ISL_1300867, EPI_ISL_1300868, EPI_ISL_1300869, EPI_ISL_1300870, EPI_ISL_1300871, EPI_ISL_1300872, EPI_ISL_1300873, EPI_ISL_1300874, EPI_ISL_1300875, EPI_ISL_1300877, EPI_ISL_1300878, EPI_ISL_1300879, EPI_ISL_1300880, EPI_ISL_1300881, EPI_ISL_1300882, EPI_ISL_1300883, EPI_ISL_1300884, EPI_ISL_1300885, EPI_ISL_1300886, EPI_ISL_1300887, EPI_ISL_1300888, EPI_ISL_1300889, EPI_ISL_1300890, EPI_ISL_1300891, EPI_ISL_1300892, EPI_ISL_1300893, EPI_ISL_1300894, EPI_ISL_1300895, EPI_ISL_1300896, EPI_ISL_1300897, EPI_ISL_1300898, EPI_ISL_1300899, EPI_ISL_1300900, EPI_ISL_1300901, EPI_ISL_1300902, EPI_ISL_1300903, EPI_ISL_1300904, EPI_ISL_1300905, EPI_ISL_1300906, EPI_ISL_1300907, EPI_ISL_1300909, EPI_ISL_1300910, EPI_ISL_1300911, EPI_ISL_1300912, EPI_ISL_1300913, EPI_ISL_1300914, EPI_ISL_1300915, EPI_ISL_1300916, EPI_ISL_1300917, EPI_ISL_1300918, EPI_ISL_1300919, EPI_ISL_1300920, EPI_ISL_1300921, EPI_ISL_1300922, EPI_ISL_1300923, EPI_ISL_1300924, EPI_ISL_1300925, EPI_ISL_1300926, EPI_ISL_1300927, EPI_ISL_1300928, EPI_ISL_1300929, | | | |

EPI_ISL_1300930, EPI_ISL_1300931, EPI_ISL_1300932, EPI_ISL_1300933, EPI_ISL_1300934, EPI_ISL_1300935, EPI_ISL_1300936, EPI_ISL_1300937, EPI_ISL_1300938, EPI_ISL_1300939, EPI_ISL_1300940, EPI_ISL_1300941, EPI_ISL_1300942, EPI_ISL_1300943, EPI_ISL_1300944, EPI_ISL_1300945, EPI_ISL_1300946, EPI_ISL_1300947, EPI_ISL_1300948, EPI_ISL_1300949, EPI_ISL_1300950, EPI_ISL_1300951, EPI_ISL_1300952, EPI_ISL_1300953, EPI_ISL_1300954, EPI_ISL_1300955, EPI_ISL_1300956, EPI_ISL_1300957, EPI_ISL_1300958, EPI_ISL_1300959, EPI_ISL_1300960, EPI_ISL_1300961, EPI_ISL_1300963, EPI_ISL_1300964, EPI_ISL_1300965, EPI_ISL_1300966, EPI_ISL_1300967, EPI_ISL_1300968, EPI_ISL_1300969, EPI_ISL_1300970, EPI_ISL_1300971, EPI_ISL_1300972, EPI_ISL_1300973, EPI_ISL_1300974, EPI_ISL_1300975, EPI_ISL_1300976, EPI_ISL_1300977, EPI_ISL_1300978, EPI_ISL_1300979, EPI_ISL_1300980, EPI_ISL_1300981, EPI_ISL_1300982, EPI_ISL_1300983, EPI_ISL_1300984, EPI_ISL_1300985, EPI_ISL_1300986, EPI_ISL_1300987, EPI_ISL_1300988, EPI_ISL_1300989, EPI_ISL_1300990, EPI_ISL_1300991, EPI_ISL_1300992, EPI_ISL_1300993, EPI_ISL_1300994, EPI_ISL_1300995, EPI_ISL_1300996, EPI_ISL_1300997, EPI_ISL_1300998, EPI_ISL_1300999, EPI_ISL_1301000, EPI_ISL_1301001, EPI_ISL_1301002, EPI_ISL_1301003, EPI_ISL_1301004, EPI_ISL_1301005, EPI_ISL_1301006, EPI_ISL_1301007, EPI_ISL_1301008, EPI_ISL_1301009, EPI_ISL_1301010, EPI_ISL_1301011, EPI_ISL_1301012, EPI_ISL_1301013, EPI_ISL_1301014, EPI_ISL_1301015, EPI_ISL_1301016, EPI_ISL_1301018, EPI_ISL_1301019, EPI_ISL_1301020, EPI_ISL_1301021, EPI_ISL_1301022, EPI_ISL_1301023, EPI_ISL_1301024, EPI_ISL_1301025, EPI_ISL_1301026, EPI_ISL_1301027, EPI_ISL_1301028, EPI_ISL_1301029, EPI_ISL_1301030, EPI_ISL_1301031, EPI_ISL_1301032, EPI_ISL_1301034, EPI_ISL_1301035, EPI_ISL_1301036, EPI_ISL_1301037, EPI_ISL_1301038, EPI_ISL_1301039, EPI_ISL_1301040, EPI_ISL_1301041, EPI_ISL_1301042, EPI_ISL_1301043, EPI_ISL_1301044, EPI_ISL_1301045, EPI_ISL_1301046, EPI_ISL_1301047, EPI_ISL_1301048, EPI_ISL_1301049, EPI_ISL_1301050, EPI_ISL_1301051, EPI_ISL_1301053, EPI_ISL_1301054, EPI_ISL_1301055, EPI_ISL_1301056, EPI_ISL_1301057, EPI_ISL_1301058, EPI_ISL_1301059, EPI_ISL_1301060, EPI_ISL_1301061, EPI_ISL_1301062, EPI_ISL_1301063, EPI_ISL_1301064, EPI_ISL_1301065, EPI_ISL_1301066, EPI_ISL_1301067, EPI_ISL_1301068, EPI_ISL_1301069, EPI_ISL_1301070, EPI_ISL_1301071, EPI_ISL_1301072, EPI_ISL_1301073, EPI_ISL_1301074, EPI_ISL_1301075, EPI_ISL_1301077, EPI_ISL_1301078, EPI_ISL_1301079, EPI_ISL_1301080, EPI_ISL_1301081, EPI_ISL_1301082, EPI_ISL_1301083, EPI_ISL_1301084, EPI_ISL_1301085, EPI_ISL_1301086, EPI_ISL_1301087, EPI_ISL_1301088, EPI_ISL_1301089, EPI_ISL_1301090, EPI_ISL_1301091, EPI_ISL_1301093, EPI_ISL_1301095, EPI_ISL_1301097, EPI_ISL_1301098, EPI_ISL_1301099, EPI_ISL_1301100, EPI_ISL_1301101, EPI_ISL_1301102, EPI_ISL_1301103, EPI_ISL_1301104, EPI_ISL_1301105, EPI_ISL_1301106, EPI_ISL_1301107, EPI_ISL_1301108, EPI_ISL_1301109, EPI_ISL_1301110, EPI_ISL_1301111, EPI_ISL_1301112, EPI_ISL_1301113, EPI_ISL_1301114, EPI_ISL_1301115, EPI_ISL_1301116, EPI_ISL_1301117, EPI_ISL_1301118, EPI_ISL_1301119, EPI_ISL_1301120, EPI_ISL_1301121, EPI_ISL_1301122, EPI_ISL_1301123, EPI_ISL_1301124, EPI_ISL_1301125, EPI_ISL_1301126, EPI_ISL_1301127, EPI_ISL_1301128, EPI_ISL_1301129, EPI_ISL_1301130, EPI_ISL_1301131, EPI_ISL_1301132, EPI_ISL_1301133, EPI_ISL_1301134, EPI_ISL_1301135, EPI_ISL_1301136, EPI_ISL_1301137, EPI_ISL_1301138, EPI_ISL_1301139, EPI_ISL_1301140, EPI_ISL_1301141, EPI_ISL_1301142, EPI_ISL_1301143, EPI_ISL_1301144, EPI_ISL_1301145, EPI_ISL_1301146, EPI_ISL_1301147, EPI_ISL_1301148, EPI_ISL_1301149, EPI_ISL_1301150, EPI_ISL_1301151, EPI_ISL_1301152, EPI_ISL_1301153, EPI_ISL_1301154, EPI_ISL_1301155, EPI_ISL_1301156, EPI_ISL_1301157, EPI_ISL_1301158, EPI_ISL_1301159, EPI_ISL_1301160, EPI_ISL_1301161, EPI_ISL_1301162, EPI_ISL_1301163, EPI_ISL_1301164, EPI_ISL_1301165, EPI_ISL_1301166, EPI_ISL_1301167, EPI_ISL_1301168, EPI_ISL_1301169, EPI_ISL_1301170, EPI_ISL_1301171, EPI_ISL_1301172, EPI_ISL_1301173, EPI_ISL_1301174, EPI_ISL_1301175, EPI_ISL_1301176, EPI_ISL_1301177, EPI_ISL_1301178, EPI_ISL_1301179, EPI_ISL_1301180, EPI_ISL_1301181, EPI_ISL_1301182, EPI_ISL_1301183, EPI_ISL_1301184, EPI_ISL_1301185, EPI_ISL_1301186, EPI_ISL_1301187, EPI_ISL_1301188, EPI_ISL_1301189, EPI_ISL_1301190, EPI_ISL_1301191, EPI_ISL_1301193, EPI_ISL_1301194, EPI_ISL_1301195, EPI_ISL_1301196, EPI_ISL_1301197, EPI_ISL_1301198, EPI_ISL_1301199, EPI_ISL_1301200, EPI_ISL_1301201, EPI_ISL_1301202, EPI_ISL_1301203, EPI_ISL_1301204, EPI_ISL_1301205, EPI_ISL_1301206, EPI_ISL_1301207, EPI_ISL_1301208, EPI_ISL_1301209, EPI_ISL_1301210, EPI_ISL_1301211, EPI_ISL_1301212, EPI_ISL_1301213, EPI_ISL_1301214, EPI_ISL_1301215, EPI_ISL_1301216, EPI_ISL_1301217, EPI_ISL_1301218, EPI_ISL_1301219, EPI_ISL_1301220, EPI_ISL_1301221, EPI_ISL_1301222, EPI_ISL_1301223, EPI_ISL_1301224, EPI_ISL_1301225, EPI_ISL_1301226, EPI_ISL_1301227, EPI_ISL_1301228, EPI_ISL_1301229, EPI_ISL_1301230, EPI_ISL_1301231, EPI_ISL_1301232, EPI_ISL_1301233, EPI_ISL_1301234, EPI_ISL_1301235, EPI_ISL_1301236, EPI_ISL_1301237, EPI_ISL_1301238, EPI_ISL_1301239, EPI_ISL_1301240, EPI_ISL_1301241, EPI_ISL_1301242, EPI_ISL_1301243, EPI_ISL_1301244, EPI_ISL_1301245, EPI_ISL_1301246, EPI_ISL_1301247, EPI_ISL_1301248, EPI_ISL_1301249, EPI_ISL_1301250, EPI_ISL_1301251, EPI_ISL_1301252, EPI_ISL_1301253, EPI_ISL_1301254, EPI_ISL_1301255, EPI_ISL_1301257, EPI_ISL_1301258, EPI_ISL_1301259, EPI_ISL_1301260, EPI_ISL_1301261, EPI_ISL_1301262, EPI_ISL_1301263, EPI_ISL_1301264, EPI_ISL_1301265, EPI_ISL_1301266, EPI_ISL_1301267, EPI_ISL_1301268, EPI_ISL_1301269, EPI_ISL_1301270, EPI_ISL_1301271, EPI_ISL_1301272, EPI_ISL_1301273, EPI_ISL_1301274, EPI_ISL_1301275, EPI_ISL_1301276, EPI_ISL_1301277, EPI_ISL_1301278, EPI_ISL_1301279, EPI_ISL_1301280, EPI_ISL_1301281, EPI_ISL_1301282, EPI_ISL_1301283, EPI_ISL_1301284, EPI_ISL_1301285, EPI_ISL_1301286, EPI_ISL_1301287, EPI_ISL_1301288, EPI_ISL_1301289, EPI_ISL_1301290, EPI_ISL_1301291, EPI_ISL_1301292, EPI_ISL_1301293, EPI_ISL_1301294, EPI_ISL_1301295, EPI_ISL_1301296, EPI_ISL_1301297, EPI_ISL_1301298, EPI_ISL_1301299, EPI_ISL_1301300, EPI_ISL_1301301, EPI_ISL_1301302, EPI_ISL_1301303, EPI_ISL_1301304, EPI_ISL_1301305, EPI_ISL_1301306, EPI_ISL_1301307, EPI_ISL_1301308, EPI_ISL_1301309, EPI_ISL_1301310, EPI_ISL_1301311, EPI_ISL_1301312, EPI_ISL_1301313, EPI_ISL_1301314, EPI_ISL_1301315, EPI_ISL_1301316, EPI_ISL_1301317, EPI_ISL_1301318, EPI_ISL_1301319, EPI_ISL_1301320, EPI_ISL_1301321, EPI_ISL_1301322, EPI_ISL_1301323, EPI_ISL_1301324, EPI_ISL_1301325, EPI_ISL_1301326, EPI_ISL_1301327, EPI_ISL_1301328, EPI_ISL_1301329, EPI_ISL_1301330, EPI_ISL_1301331, EPI_ISL_1301332, EPI_ISL_1301333, EPI_ISL_1301334, EPI_ISL_1301335, EPI_ISL_1301336, EPI_ISL_1301337, EPI_ISL_1301338, EPI_ISL_1301339, EPI_ISL_1301340, EPI_ISL_1301341, EPI_ISL_1301342, EPI_ISL_1301343, EPI_ISL_1301344, EPI_ISL_1301345, EPI_ISL_1301346, EPI_ISL_1301347, EPI_ISL_1301348, EPI_ISL_1301349, EPI_ISL_1301350, EPI_ISL_1301351, EPI_ISL_1301352, EPI_ISL_1301353, EPI_ISL_1301354, EPI_ISL_1301355, EPI_ISL_1301356, EPI_ISL_1301357, EPI_ISL_1301358, EPI_ISL_1301359, EPI_ISL_1301360, EPI_ISL_1301361, EPI_ISL_1301362, EPI_ISL_1301363, EPI_ISL_1301364, EPI_ISL_1301365, EPI_ISL_1301366, EPI_ISL_1301367, EPI_ISL_1301368, EPI_ISL_1301369, EPI_ISL_1301370, EPI_ISL_1301371, EPI_ISL_1301372, EPI_ISL_1301373, EPI_ISL_1301374, EPI_ISL_1301375, EPI_ISL_1301376, EPI_ISL_1301377, EPI_ISL_1301378, EPI_ISL_1301379, EPI_ISL_1301380, EPI_ISL_1301381, EPI_ISL_1301382, EPI_ISL_1301383, EPI_ISL_1301384, EPI_ISL_1301385, EPI_ISL_1301386, EPI_ISL_1301387, EPI_ISL_1301388, EPI_ISL_1301389, EPI_ISL_1301390, EPI_ISL_1301392, EPI_ISL_1301393, EPI_ISL_1301394, EPI_ISL_1301395, EPI_ISL_1301396, EPI_ISL_1301397, EPI_ISL_1301398, EPI_ISL_1301400, EPI_ISL_1301401, EPI_ISL_1301402, EPI_ISL_1301403, EPI_ISL_1301404, EPI_ISL_1301405, EPI_ISL_1301406, EPI_ISL_1301407, EPI_ISL_1301408, EPI_ISL_1301409, EPI_ISL_1301410, EPI_ISL_1301411, EPI_ISL_1301412, EPI_ISL_1301413, EPI_ISL_1301414, EPI_ISL_1301415, EPI_ISL_1301416, EPI_ISL_1301417, EPI_ISL_1301418, EPI_ISL_1301419, EPI_ISL_1301420, EPI_ISL_1301421, EPI_ISL_1301422, EPI_ISL_1301423, EPI_ISL_1301424, EPI_ISL_1301425, EPI_ISL_1301426, EPI_ISL_1301427, EPI_ISL_1301428, EPI_ISL_1301429, EPI_ISL_1301430, EPI_ISL_1301431, EPI_ISL_1301432, EPI_ISL_1301433, EPI_ISL_1301434, EPI_ISL_1301435, EPI_ISL_1301436, EPI_ISL_1301437, EPI_ISL_1301438, EPI_ISL_1301439

| | | | |
|---|---|---|---|
| see above | MSHS Clinical Microbiology Laboratories | MSHS Pathogen Surveillance Program | Ana S. Gonzalez-Reiche, Hala Alshammary, Mitchell J. Sullivan, Brianne Ciferri, Ajay Obla, Angela Amoako, Mahmoud Awawda, Daniel Floda, Julia Matthews, Ashley Salimbangon, Levy Sominsky, Katherine Beach, Kayla Russo, Charles Gleason, Shelcie Fabre, Giulio Kleiner, Zenab Khan, Bremy Alburquerque, Adriana van de Guchte, Komal Srivastava, Matthew M. Hernandez, Jayeeta Dutta, Denise Jurczyszak, Nancy Francoeur, Betsaida Salom Melo, Irina Oussenko, Gintaras Deikus, Juan Soto, Shwetha Hara Sridhar, Ying-Chih Wang, Kathryn Twyman, Deena R. Altman, Robert Sebra, Adolfo García-Sastre, Marta Luksza, Gopi Patel, Sarah Schaefer, Melissa Gitman, Michael D. Nowak, Alberto Paniz-Mondolfi, Emilia Mia Sordillo, Viviana Simon, Harm van Bakel |
| EPI_ISL_1301440 | MSHS Clinical Microbiology Laboratories | NBACC | Michael Schuit, Jennifer Biryukov, Katie Beck, Jason Yolitz, Jordan Bohannon, Wade Weaver, David Miller, Brian Holland, Melissa Krause, Denise Freeburger, Gregory Williams, Stewart Wood, Amanda Graham, M.J. Rosovitz, Adam Bazinet, Aaron Phillips, Sean Lovett, Karla Garcia, Elyse Abbott, Victoria Wahl, Shanna Ratnesar-Shumate, Paul Dabisch |
