## Supplemental acknowledgement table #3 for "Genomic Surveillance of SARS-CoV-2 in Erie County, New York"

We gratefully acknowledge the following Authors from the Originating laboratories responsible for obtaining the specimens, as well as the Submitting laboratories where the genome data were generated and shared via GISAID, on which this research is based.

All Submitters of data may be contacted directly via www.gisaid.org

Authors are sorted alphabetically.

| Accession ID | Originating Laboratory | Submitting Laboratory | Authors |
| --- | --- | --- | --- |

EPI_ISL_1306253, EPI_ISL_1306254, EPI_ISL_1306255, EPI_ISL_1306256, EPI_ISL_1306257, EPI_ISL_1306258, EPI_ISL_1306259, EPI_ISL_1306260, EPI_ISL_1306261, EPI_ISL_1306262, EPI_ISL_1306263, EPI_ISL_1306264, EPI_ISL_1306265, EPI_ISL_1306266, EPI_ISL_1306267, EPI_ISL_1306268, EPI_ISL_1306269,
EPI_ISL_1306270, EPI_ISL_1306271, EPI_ISL_1306272, EPI_ISL_1306273, EPI_ISL_1306274, EPI_ISL_1306275, EPI_ISL_1306276, EPI_ISL_1306277, EPI_ISL_1306278, EPI_ISL_1306279, EPI_ISL_1306280, EPI_ISL_1306281, EPI_ISL_1306282, EPI_ISL_1306283, EPI_ISL_1306284, EPI_ISL_1306285, EPI_ISL_1306286,
EPI_ISL_1306287, EPI_ISL_1306288, EPI_ISL_1306289, EPI_ISL_1306290, EPI_ISL_1306291, EPI_ISL_1306292, EPI_ISL_1306293, EPI_ISL_1306294, EPI_ISL_1306295, EPI_ISL_1306296, EPI_ISL_1306297, EPI_ISL_1306298, EPI_ISL_1306299, EPI_ISL_1306300, EPI_ISL_1306301, EPI_ISL_1306302, EPI_ISL_1306303,
EPI_ISL_1306304, EPI_ISL_1306305, EPI_ISL_1306306, EPI_ISL_1306307, EPI_ISL_1306308, EPI_ISL_1306309, EPI_ISL_1306310, EPI_ISL_1306311, EPI_ISL_1306312, EPI_ISL_1306313, EPI_ISL_1306314, EPI_ISL_1306315, EPI_ISL_1306316, EPI_ISL_1306317, EPI_ISL_1306318, EPI_ISL_1306319, EPI_ISL_1306320,
EPI_ISL_1306321, EPI_ISL_1306322, EPI_ISL_1306323, EPI_ISL_1306324, EPI_ISL_1306325, EPI_ISL_1306326, EPI_ISL_1306327, EPI_ISL_1306328, EPI_ISL_1306329, EPI_ISL_1306330, EPI_ISL_1306331, EPI_ISL_1306332, EPI_ISL_1306333, EPI_ISL_1306334, EPI_ISL_1306335, EPI_ISL_1306336, EPI_ISL_1306337,
EPI_ISL_1306338, EPI_ISL_1306339, EPI_ISL_1306340, EPI_ISL_1306341, EPI_ISL_1306342, EPI_ISL_1306343, EPI_ISL_1306344, EPI_ISL_1306345, EPI_ISL_1306346, EPI_ISL_1306347, EPI_ISL_1306348, EPI_ISL_1306349, EPI_ISL_1306350, EPI_ISL_1306351, EPI_ISL_1306352, EPI_ISL_1306353, EPI_ISL_1306354,
EPI_ISL_1306355, EPI_ISL_1306356, EPI_ISL_1306357, EPI_ISL_1306358, EPI_ISL_1306359, EPI_ISL_1306360, EPI_ISL_1306361, EPI_ISL_1306362, EPI_ISL_1306363, EPI_ISL_1306364, EPI_ISL_1306365, EPI_ISL_1306366, EPI_ISL_1306367, EPI_ISL_1306368, EPI_ISL_1306369, EPI_ISL_1306370, EPI_ISL_1306371,
EPI_ISL_1306372, EPI_ISL_1306373, EPI_ISL_1306374, EPI_ISL_1306375, EPI_ISL_1306376, EPI_ISL_1306377, EPI_ISL_1306378, EPI_ISL_1306379, EPI_ISL_1306380, EPI_ISL_1306381, EPI_ISL_1306382, EPI_ISL_1306383, EPI_ISL_1306384, EPI_ISL_1306385, EPI_ISL_1306386, EPI_ISL_1306387, EPI_ISL_1306388,
EPI_ISL_1306389, EPI_ISL_1306390, EPI_ISL_1306391, EPI_ISL_1306392, EPI_ISL_1306393, EPI_ISL_1306394, EPI_ISL_1306395, EPI_ISL_1306396, EPI_ISL_1306397, EPI_ISL_1306398, EPI_ISL_1306399, EPI_ISL_1306400, EPI_ISL_1306401, EPI_ISL_1306402, EPI_ISL_1306403, EPI_ISL_1306404, EPI_ISL_1306405,
EPI_ISL_1306406, EPI_ISL_1306407, EPI_ISL_1306408, EPI_ISL_1306409, EPI_ISL_1306410, EPI_ISL_1306411, EPI_ISL_1306412, EPI_ISL_1306413, EPI_ISL_1306414, EPI_ISL_1306415, EPI_ISL_1306416, EPI_ISL_1306417, EPI_ISL_1306418, EPI_ISL_1306419, EPI_ISL_1306420, EPI_ISL_1306421, EPI_ISL_1306422,
EPI_ISL_1306423, EPI_ISL_1306424, EPI_ISL_1306425, EPI_ISL_1306426, EPI_ISL_1306427, EPI_ISL_1306428, EPI_ISL_1306429, EPI_ISL_1306430, EPI_ISL_1306431, EPI_ISL_1306432, EPI_ISL_1306433, EPI_ISL_1306434, EPI_ISL_1306435, EPI_ISL_1306436, EPI_ISL_1306437, EPI_ISL_1306438, EPI_ISL_1306439,
EPI_ISL_1306440, EPI_ISL_1306441, EPI_ISL_1306442, EPI_ISL_1306443, EPI_ISL_1306444, EPI_ISL_1306445, EPI_ISL_1306446, EPI_ISL_1306447, EPI_ISL_1306448, EPI_ISL_1306449, EPI_ISL_1306450, EPI_ISL_1306451, EPI_ISL_1306452, EPI_ISL_1306453, EPI_ISL_1306454, EPI_ISL_1306455, EPI_ISL_1306456,
EPI_ISL_1306457, EPI_ISL_1306458, EPI_ISL_1306459, EPI_ISL_1306460, EPI_ISL_1306461, EPI_ISL_1306462, EPI_ISL_1306463, EPI_ISL_1306464, EPI_ISL_1306465, EPI_ISL_1306466, EPI_ISL_1306467, EPI_ISL_1306468, EPI_ISL_1306469, EPI_ISL_1306470, EPI_ISL_1306471, EPI_ISL_1306472, EPI_ISL_1306473,
EPI_ISL_1306474, EPI_ISL_1306475, EPI_ISL_1306476, EPI_ISL_1306477, EPI_ISL_1306478, EPI_ISL_1306479, EPI_ISL_1306480, EPI_ISL_1306481, EPI_ISL_1306482, EPI_ISL_1306483, EPI_ISL_1306484, EPI_ISL_1306485, EPI_ISL_1306486, EPI_ISL_1306487, EPI_ISL_1306488, EPI_ISL_1306489, EPI_ISL_1306490,
EPI_ISL_1306491, EPI_ISL_1306492, EPI_ISL_1306493, EPI_ISL_1306494, EPI_ISL_1306495, EPI_ISL_1306496, EPI_ISL_1306497, EPI_ISL_1306498, EPI_ISL_1306499, EPI_ISL_1306500, EPI_ISL_1306501, EPI_ISL_1306502, EPI_ISL_1306503, EPI_ISL_1306504, EPI_ISL_1306505, EPI_ISL_1306506, EPI_ISL_1306507,
EPI_ISL_1306508, EPI_ISL_1306509, EPI_ISL_1306510, EPI_ISL_1306511, EPI_ISL_1306512, EPI_ISL_1306513, EPI_ISL_1306514, EPI_ISL_1306515, EPI_ISL_1306516, EPI_ISL_1306517, EPI_ISL_1306518, EPI_ISL_1306519, EPI_ISL_1306520, EPI_ISL_1306521, EPI_ISL_1306522, EPI_ISL_1306523, EPI_ISL_1306524,
EPI_ISL_1306525, EPI_ISL_1306526, EPI_ISL_1306527, EPI_ISL_1306528, EPI_ISL_1306529, EPI_ISL_1306530, EPI_ISL_1306531, EPI_ISL_1306532, EPI_ISL_1306533, EPI_ISL_1306534, EPI_ISL_1306535, EPI_ISL_1306536, EPI_ISL_1306537, EPI_ISL_1306538, EPI_ISL_1306539, EPI_ISL_1306540, EPI_ISL_1306541,
EPI_ISL_1306542, EPI_ISL_1306543, EPI_ISL_1306544, EPI_ISL_1306545, EPI_ISL_1306546, EPI_ISL_1306547, EPI_ISL_1306548, EPI_ISL_1306549, EPI_ISL_1306550, EPI_ISL_1306551, EPI_ISL_1306552, EPI_ISL_1306553, EPI_ISL_1306554, EPI_ISL_1306555, EPI_ISL_1306556, EPI_ISL_1306557, EPI_ISL_1306558,
EPI_ISL_1306559, EPI_ISL_1306560, EPI_ISL_1306561, EPI_ISL_1306562, EPI_ISL_1306563, EPI_ISL_1306564, EPI_ISL_1306565, EPI_ISL_1306566, EPI_ISL_1306567, EPI_ISL_1306568, EPI_ISL_1306569, EPI_ISL_1306570, EPI_ISL_1306571, EPI_ISL_1306572, EPI_ISL_1306573, EPI_ISL_1306574, EPI_ISL_1306575,
EPI_ISL_1306576, EPI_ISL_1306577, EPI_ISL_1306578, EPI_ISL_1306579, EPI_ISL_1306580, EPI_ISL_1306581, EPI_ISL_1306582, EPI_ISL_1306583, EPI_ISL_1306584, EPI_ISL_1306585, EPI_ISL_1306586, EPI_ISL_1306587, EPI_ISL_1306588, EPI_ISL_1306589, EPI_ISL_1306590, EPI_ISL_1306591, EPI_ISL_1306592,
EPI_ISL_1306593, EPI_ISL_1306594, EPI_ISL_1306595, EPI_ISL_1306596, EPI_ISL_1306597, EPI_ISL_1306598, EPI_ISL_1306599, EPI_ISL_1306600, EPI_ISL_1306601, EPI_ISL_1306602, EPI_ISL_1306603, EPI_ISL_1306604, EPI_ISL_1306605, EPI_ISL_1306606, EPI_ISL_1306607, EPI_ISL_1306608, EPI_ISL_1306609,
EPI_ISL_1306610, EPI_ISL_1306611, EPI_ISL_1306612, EPI_ISL_1306613, EPI_ISL_1306614, EPI_ISL_1306615, EPI_ISL_1306616, EPI_ISL_1306617, EPI_ISL_1306618, EPI_ISL_1306619, EPI_ISL_1306620, EPI_ISL_1306621, EPI_ISL_1306622, EPI_ISL_1306623, EPI_ISL_1306624, EPI_ISL_1306625, EPI_ISL_1306626,
EPI_ISL_1306627, EPI_ISL_1306628, EPI_ISL_1306629, EPI_ISL_1306630, EPI_ISL_1306631, EPI_ISL_1306632, EPI_ISL_1306633, EPI_ISL_1306634, EPI_ISL_1306635, EPI_ISL_1306636, EPI_ISL_1306637, EPI_ISL_1306638, EPI_ISL_1306639, EPI_ISL_1306640, EPI_ISL_1306641, EPI_ISL_1306642, EPI_ISL_1306643,
EPI_ISL_1306644, EPI_ISL_1306645, EPI_ISL_1306646, EPI_ISL_1306647, EPI_ISL_1306648, EPI_ISL_1306649, EPI_ISL_1306650, EPI_ISL_1306651, EPI_ISL_1306652, EPI_ISL_1306653, EPI_ISL_1306654, EPI_ISL_1306655, EPI_ISL_1306656, EPI_ISL_1306657, EPI_ISL_1306658, EPI_ISL_1306659, EPI_ISL_1306660,
EPI_ISL_1306661, EPI_ISL_1306662, EPI_ISL_1306663, EPI_ISL_1306664, EPI_ISL_1306665, EPI_ISL_1306666, EPI_ISL_1306667, EPI_ISL_1306668, EPI_ISL_1306669, EPI_ISL_1306670, EPI_ISL_1306671, EPI_ISL_1306672, EPI_ISL_1306673, EPI_ISL_1306674, EPI_ISL_1306675, EPI_ISL_1306676, EPI_ISL_1306677,
EPI_ISL_1306678, EPI_ISL_1306679, EPI_ISL_1306680, EPI_ISL_1306681, EPI_ISL_1306682, EPI_ISL_1306683, EPI_ISL_1306684, EPI_ISL_1306685, EPI_ISL_1306686, EPI_ISL_1306687, EPI_ISL_1306688, EPI_ISL_1306689, EPI_ISL_1306690, EPI_ISL_1306691, EPI_ISL_1306692, EPI_ISL_1306693, EPI_ISL_1306694,
EPI_ISL_1306695, EPI_ISL_1306696, EPI_ISL_1306697, EPI_ISL_1306698, EPI_ISL_1306699, EPI_ISL_1306700, EPI_ISL_1306701, EPI_ISL_1306702, EPI_ISL_1306703, EPI_ISL_1306704, EPI_ISL_1306705, EPI_ISL_1306706, EPI_ISL_1306707, EPI_ISL_1306708, EPI_ISL_1306709, EPI_ISL_1306710, EPI_ISL_1306711,
EPI_ISL_1306712, EPI_ISL_1306713, EPI_ISL_1306714, EPI_ISL_1306715, EPI_ISL_1306716, EPI_ISL_1306717, EPI_ISL_1306718, EPI_ISL_1306719, EPI_ISL_1306720, EPI_ISL_1306721, EPI_ISL_1306722, EPI_ISL_1306723, EPI_ISL_1306724, EPI_ISL_1306725, EPI_ISL_1306726, EPI_ISL_1306727, EPI_ISL_1306728,
EPI_ISL_1306729, EPI_ISL_1306730, EPI_ISL_1306731, EPI_ISL_1306732, EPI_ISL_1306733, EPI_ISL_1306734, EPI_ISL_1306735, EPI_ISL_1306736, EPI_ISL_1306737, EPI_ISL_1306738, EPI_ISL_1306739, EPI_ISL_1306740, EPI_ISL_1306741, EPI_ISL_1306742, EPI_ISL_1306743, EPI_ISL_1306744, EPI_ISL_1306745,
EPI_ISL_1306746, EPI_ISL_1306747, EPI_ISL_1306748, EPI_ISL_1306749, EPI_ISL_1306750, EPI_ISL_1306751, EPI_ISL_1306752, EPI_ISL_1306753, EPI_ISL_1306754, EPI_ISL_1306755, EPI_ISL_1306756, EPI_ISL_1306757, EPI_ISL_1306758, EPI_ISL_1306759, EPI_ISL_1306760, EPI_ISL_1306761, EPI_ISL_1306762,
EPI_ISL_1306763, EPI_ISL_1306764, EPI_ISL_1306765, EPI_ISL_1306766, EPI_ISL_1306767, EPI_ISL_1306768, EPI_ISL_1306769, EPI_ISL_1306770, EPI_ISL_1306771, EPI_ISL_1306772, EPI_ISL_1306773, EPI_ISL_1306774, EPI_ISL_1306775, EPI_ISL_1306776, EPI_ISL_1306777, EPI_ISL_1306778, EPI_ISL_1306779,
EPI_ISL_1306780, EPI_ISL_1306781, EPI_ISL_1306782, EPI_ISL_1306783, EPI_ISL_1306784, EPI_ISL_1306785, EPI_ISL_1306786, EPI_ISL_1306787, EPI_ISL_1306788, EPI_ISL_1306789, EPI_ISL_1306790, EPI_ISL_1306791, EPI_ISL_1306792, EPI_ISL_1306793, EPI_ISL_1306794, EPI_ISL_1306795, EPI_ISL_1306796,
EPI_ISL_1306797, EPI_ISL_1306798, EPI_ISL_1306799, EPI_ISL_1306800, EPI_ISL_1306801, EPI_ISL_1306802, EPI_ISL_1306803, EPI_ISL_1306804, EPI_ISL_1306805, EPI_ISL_1306806, EPI_ISL_1306807, EPI_ISL_1306808, EPI_ISL_1306809, EPI_ISL_1306810, EPI_ISL_1306811, EPI_ISL_1306812, EPI_ISL_1306813,
EPI_ISL_1306814, EPI_ISL_1306815, EPI_ISL_1306816, EPI_ISL_1306817, EPI_ISL_1306818, EPI_ISL_1306819, EPI_ISL_1306820, EPI_ISL_1306821, EPI_ISL_1306822, EPI_ISL_1306823, EPI_ISL_1306824, EPI_ISL_1306825, EPI_ISL_1306826, EPI_ISL_1306827, EPI_ISL_1306828, EPI_ISL_1306829, EPI_ISL_1306830,
EPI_ISL_1306831, EPI_ISL_1306832, EPI_ISL_1306833, EPI_ISL_1306834, EPI_ISL_1306835, EPI_ISL_1306836, EPI_ISL_1306837, EPI_ISL_1306838, EPI_ISL_1306839, EPI_ISL_1306840, EPI_ISL_1306841, EPI_ISL_1306842, EPI_ISL_1306843, EPI_ISL_1306844, EPI_ISL_1306845, EPI_ISL_1306846, EPI_ISL_1306847,
EPI_ISL_1306848, EPI_ISL_1306849, EPI_ISL_1306850, EPI_ISL_1306851, EPI_ISL_1306852, EPI_ISL_1306853, EPI_ISL_1306854, EPI_ISL_1306855, EPI_ISL_1306856, EPI_ISL_1306857, EPI_ISL_1306858, EPI_ISL_1306859, EPI_ISL_1306860, EPI_ISL_1306861, EPI_ISL_1306862, EPI_ISL_1306863, EPI_ISL_1306864,
EPI_ISL_1306865, EPI_ISL_1306866, EPI_ISL_1306867, EPI_ISL_1306868, EPI_ISL_1306869, EPI_ISL_1306870, EPI_ISL_1306871, EPI_ISL_1306872, EPI_ISL_1306873, EPI_ISL_1306874, EPI_ISL_1306875, EPI_ISL_1306876, EPI_ISL_1306877, EPI_ISL_1306878, EPI_ISL_1306879, EPI_ISL_1306880, EPI_ISL_1306881,
EPI_ISL_1306882, EPI_ISL_1306883, EPI_ISL_1306884, EPI_ISL_1306885, EPI_ISL_1306886, EPI_ISL_1306887, EPI_ISL_1306888, EPI_ISL_1306889, EPI_ISL_1306890, EPI_ISL_1306891, EPI_ISL_1306892, EPI_ISL_1306893, EPI_ISL_1306894, EPI_ISL_1306895, EPI_ISL_1306896, EPI_ISL_1306897, EPI_ISL_1306898,
EPI_ISL_1306899, EPI_ISL_1306900, EPI_ISL_1306901, EPI_ISL_1306902, EPI_ISL_1306903, EPI_ISL_1306904, EPI_ISL_1306905, EPI_ISL_1306906, EPI_ISL_1306907, EPI_ISL_1306908, EPI_ISL_1306909, EPI_ISL_1306910, EPI_ISL_1306911, EPI_ISL_1306912, EPI_ISL_1306913, EPI_ISL_1306914, EPI_ISL_1306915,
EPI_ISL_1306916, EPI_ISL_1306917, EPI_ISL_1306918, EPI_ISL_1306919, EPI_ISL_1306920, EPI_ISL_1306921, EPI_ISL_1306922, EPI_ISL_1306923, EPI_ISL_1306924, EPI_ISL_1306925, EPI_ISL_1306926, EPI_ISL_1306927, EPI_ISL_1306928, EPI_ISL_1306929, EPI_ISL_1306930, EPI_ISL_1306931, EPI_ISL_1306932,
EPI_ISL_1306933, EPI_ISL_1306934, EPI_ISL_1306935, EPI_ISL_1306936, EPI_ISL_1306937, EPI_ISL_1306938, EPI_ISL_1306939, EPI_ISL_1306940, EPI_ISL_1306941, EPI_ISL_1306942, EPI_ISL_1306943, EPI_ISL_1306944, EPI_ISL_1306945, EPI_ISL_1306946, EPI_ISL_1306947, EPI_ISL_1306948, EPI_ISL_1306949,
EPI_ISL_1306950, EPI_ISL_1306951, EPI_ISL_1306952, EPI_ISL_1306953, EPI_ISL_1306954, EPI_ISL_1306955, EPI_ISL_1306956, EPI_ISL_1306957, EPI_ISL_1306958, EPI_ISL_1306959, EPI_ISL_1306960, EPI_ISL_1306961, EPI_ISL_1306962, EPI_ISL_1306963, EPI_ISL_1306964, EPI_ISL_1306965, EPI_ISL_1306966,
EPI_ISL_1306967, EPI_ISL_1306968, EPI_ISL_1306969, EPI_ISL_1306970, EPI_ISL_1306971, EPI_ISL_1306972, EPI_ISL_1306973, EPI_ISL_1306974, EPI_ISL_1306975, EPI_ISL_1306976, EPI_ISL_1306977, EPI_ISL_1306978, EPI_ISL_1306979, EPI_ISL_1306980, EPI_ISL_1306981, EPI_ISL_1306982, EPI_ISL_1306983,
EPI_ISL_1306984, EPI_ISL_1306985, EPI_ISL_1306986, EPI_ISL_1306987, EPI_ISL_1306988, EPI_ISL_1306989, EPI_ISL_1306990, EPI_ISL_1306991, EPI_ISL_1306992, EPI_ISL_1306993, EPI_ISL_1306994, EPI_ISL_1306995, EPI_ISL_1306996, EPI_ISL_1306997, EPI_ISL_1306998, EPI_ISL_1306999, EPI_ISL_1307000,
EPI_ISL_1307001, EPI_ISL_1307002, EPI_ISL_1307003, EPI_ISL_1307004, EPI_ISL_1307005, EPI_ISL_1307006, EPI_ISL_1307007, EPI_ISL_1307008, EPI_ISL_1307009, EPI_ISL_1307010, EPI_ISL_1307011, EPI_ISL_1307012, EPI_ISL_1307013, EPI_ISL_1307014, EPI_ISL_1307015, EPI_ISL_1307016, EPI_ISL_1307017,
EPI_ISL_1307018, EPI_ISL_1307019, EPI_ISL_1307020, EPI_ISL_1307021, EPI_ISL_1307022, EPI_ISL_1307023, EPI_ISL_1307024, EPI_ISL_1307025, EPI_ISL_1307026, EPI_ISL_1307027, EPI_ISL_1307028, EPI_ISL_1307029, EPI_ISL_1307030, EPI_ISL_1307031, EPI_ISL_1307032, EPI_ISL_1307033, EPI_ISL_1307034,
EPI_ISL_1307035, EPI_ISL_1307036, EPI_ISL_1307037, EPI_ISL_1307038, EPI_ISL_1307039, EPI_ISL_1307040, EPI_ISL_1307041, EPI_ISL_1307042, EPI_ISL_1307043, EPI_ISL_1307044, EPI_ISL_1307045, EPI_ISL_1307046, EPI_ISL_1307047, EPI_ISL_1307048, EPI_ISL_1307049, EPI_ISL_1307050, EPI_ISL_1307051,
EPI_ISL_1307052, EPI_ISL_1307053, EPI_ISL_1307054, EPI_ISL_1307055, EPI_ISL_1307056, EPI_ISL_1307057, EPI_ISL_1307058, EPI_ISL_1307059, EPI_ISL_1307060, EPI_ISL_1307061, EPI_ISL_1307062, EPI_ISL_1307063, EPI_ISL_1307064, EPI_ISL_1307065, EPI_ISL_1307066, EPI_ISL_1307067, EPI_ISL_1307068,
EPI_ISL_1307069, EPI_ISL_1307070, EPI_ISL_1307071, EPI_ISL_1307072, EPI_ISL_1307073, EPI_ISL_1307074, EPI_ISL_1307075, EPI_ISL_1307076, EPI_ISL_1307077, EPI_ISL_1307078, EPI_ISL_1307079, EPI_ISL_1307080, EPI_ISL_1307081, EPI_ISL_1307082, EPI_ISL_1307083, EPI_ISL_1307084, EPI_ISL_1307085,
EPI_ISL_1307086, EPI_ISL_1307087, EPI_ISL_1307088, EPI_ISL_1307089, EPI_ISL_1307090, EPI_ISL_1307091, EPI_ISL_1307092, EPI_ISL_1307093, EPI_ISL_1307094, EPI_ISL_1307095, EPI_ISL_1307096, EPI_ISL_1307097, EPI_ISL_1307098, EPI_ISL_1307099, EPI_ISL_1307100, EPI_ISL_1307101, EPI_ISL_1307102,
EPI_ISL_1307103, EPI_ISL_1307104, EPI_ISL_1307105, EPI_ISL_1307106, EPI_ISL_1307107, EPI_ISL_1307108, EPI_ISL_1307109, EPI_ISL_1307110, EPI_ISL_1307111, EPI_ISL_1307112, EPI_ISL_1307113, EPI_ISL_1307114, EPI_ISL_1307115, EPI_ISL_1307116, EPI_ISL_1307117, EPI_ISL_1307118, EPI_ISL_1307119,
EPI_ISL_1307120, EPI_ISL_1307121, EPI_ISL_1307122, EPI_ISL_1307123, EPI_ISL_1307124, EPI_ISL_1307125, EPI_ISL_1307126, EPI_ISL_1307127, EPI_ISL_1307128, EPI_ISL_1307129, EPI_ISL_1307130, EPI_ISL_1307131, EPI_ISL_1307132, EPI_ISL_1307133, EPI_ISL_1307134, EPI_ISL_1307135, EPI_ISL_1307136,
EPI_ISL_1307137, EPI_ISL_1307138, EPI_ISL_1307139, EPI_ISL_1307140, EPI_ISL_1307141, EPI_ISL_1307142, EPI_ISL_1307143, EPI_ISL_1307144, EPI_ISL_1307145, EPI_ISL_1307146, EPI_ISL_1307147, EPI_ISL_1307148, EPI_ISL_1307149, EPI_ISL_1307150, EPI_ISL_1307151, EPI_ISL_1307152, EPI_ISL_1307153,
EPI_ISL_1307154, EPI_ISL_1307155, EPI_ISL_1307156, EPI_ISL_1307157, EPI_ISL_1307158, EPI_ISL_1307159, EPI_ISL_1307160, EPI_ISL_1307161, EPI_ISL_1307162, EPI_ISL_1307163, EPI_ISL_1307164, EPI_ISL_1307165, EPI_ISL_1307166, EPI_ISL_1307167, EPI_ISL_1307168, EPI_ISL_1307169, EPI_ISL_1307170,
EPI_ISL_1307171, EPI_ISL_1307172, EPI_ISL_1307173, EPI_ISL_1307174, EPI_ISL_1307175, EPI_ISL_1307176, EPI_ISL_1307178, EPI_ISL_1307179, EPI_ISL_1307180, EPI_ISL_1307181, EPI_ISL_1307182, EPI_ISL_1307183, EPI_ISL_1307184, EPI_ISL_1307185, EPI_ISL_1307186, EPI_ISL_1307187, EPI_ISL_1307188,
EPI_ISL_1307189, EPI_ISL_1307190, EPI_ISL_1307191, EPI_ISL_1307192, EPI_ISL_1307193, EPI_ISL_1307194, EPI_ISL_1307195, EPI_ISL_1307196, EPI_ISL_1307197, EPI_ISL_1307198, EPI_ISL_1307199, EPI_ISL_1307200, EPI_ISL_1307201, EPI_ISL_1307202, EPI_ISL_1307203, EPI_ISL_1307204, EPI_ISL_1307205,
EPI_ISL_1307206, EPI_ISL_1307207, EPI_ISL_1307208, EPI_ISL_1307209, EPI_ISL_1307210, EPI_ISL_1307211, EPI_ISL_1307212, EPI_ISL_1307213, EPI_ISL_1307214, EPI_ISL_1307215, EPI_ISL_1307216, EPI_ISL_1307217, EPI_ISL_1307218, EPI_ISL_1307219, EPI_ISL_1307220, EPI_ISL_1307221, EPI_ISL_1307222,
EPI_ISL_1307223, EPI_ISL_1307224, EPI_ISL_1307225, EPI_ISL_1307227, EPI_ISL_1307228, EPI_ISL_1307229, EPI_ISL_1307230, EPI_ISL_1307231, EPI_ISL_1307232, EPI_ISL_1307233, EPI_ISL_1307234, EPI_ISL_1307235, EPI_ISL_1307236, EPI_ISL_1307237, EPI_ISL_1307238, EPI_ISL_1307239,
EPI_ISL_1307240, EPI_ISL_1307241, EPI_ISL_1307242, EPI_ISL_1307243, EPI_ISL_1307244, EPI_ISL_1307245, EPI_ISL_1307246, EPI_ISL_1307247, EPI_ISL_1307248, EPI_ISL_1307249, EPI_ISL_1307250, EPI_ISL_1307251, EPI_ISL_1307252, EPI_ISL_1307253, EPI_ISL_1307254, EPI_ISL_1307255, EPI_ISL_1307256,
EPI_ISL_1307257, EPI_ISL_1307258, EPI_ISL_1307259, EPI_ISL_1307260, EPI_ISL_1307261, EPI_ISL_1307262, EPI_ISL_1307263, EPI_ISL_1307264, EPI_ISL_1307265, EPI_ISL_1307266, EPI_ISL_1307268, EPI_ISL_1307269, EPI_ISL_1307270, EPI_ISL_1307271, EPI_ISL_1307272, EPI_ISL_1307273,
EPI_ISL_1307274, EPI_ISL_1307275, EPI_ISL_1307276, EPI_ISL_1307277, EPI_ISL_1307278, EPI_ISL_1307279, EPI_ISL_1307280, EPI_ISL_1307281, EPI_ISL_1307282, EPI_ISL_1307283, EPI_ISL_1307284, EPI_ISL_1307285, EPI_ISL_1307286, EPI_ISL_1307287, EPI_ISL_1307288, EPI_ISL_1307289, EPI_ISL_1307290,
EPI_ISL_1307291, EPI_ISL_1307292, EPI_ISL_1307293, EPI_ISL_1307294, EPI_ISL_1307295, EPI_ISL_1307296, EPI_ISL_1307297, EPI_ISL_1307298, EPI_ISL_1307299, EPI_ISL_1307300, EPI_ISL_1307301, EPI_ISL_1307302, EPI_ISL_1307303, EPI_ISL_1307304, EPI_ISL_1307305, EPI_ISL_1307306, EPI_ISL_1307307,
EPI_ISL_1307308, EPI_ISL_1307309, EPI_ISL_1307310, EPI_ISL_1307311, EPI_ISL_1307312, EPI_ISL_1307313, EPI_ISL_1307314, EPI_ISL_1307315, EPI_ISL_1307316, EPI_ISL_1307317, EPI_ISL_1307318, EPI_ISL_1307319, EPI_ISL_1307320, EPI_ISL_1307321, EPI_ISL_1307322, EPI_ISL_1307323, EPI_ISL_1307324,
EPI_ISL_1307325, EPI_ISL_1307326, EPI_ISL_1307327, EPI_ISL_1307328, EPI_ISL_1307329, EPI_ISL_1307330, EPI_ISL_1307331, EPI_ISL_1307332, EPI_ISL_1307333, EPI_ISL_1307334, EPI_ISL_1307335, EPI_ISL_1307336, EPI_ISL_1307337, EPI_ISL_1307338, EPI_ISL_1307339, EPI_ISL_1307340, EPI_ISL_1307341,
EPI_ISL_1307342, EPI_ISL_1307343, EPI_ISL_1307344, EPI_ISL_1307345, EPI_ISL_1307346, EPI_ISL_1307347, EPI_ISL_1307348, EPI_ISL_1307349, EPI_ISL_1307350, EPI_ISL_1307351, EPI_ISL_1307352, EPI_ISL_1307353, EPI_ISL_1307354, EPI_ISL_1307355, EPI_ISL_1307356, EPI_ISL_1307357, EPI_ISL_1307358,
EPI_ISL_1307359, EPI_ISL_1307360, EPI_ISL_1307361, EPI_ISL_1307362, EPI_ISL_1307363, EPI_ISL_1307364, EPI_ISL_1307365, EPI_ISL_1307366, EPI_ISL_1307367, EPI_ISL_1307368, EPI_ISL_1307369, EPI_ISL_1307370, EPI_ISL_1307371, EPI_ISL_1307372, EPI_ISL_1307373, EPI_ISL_1307374, EPI_ISL_1307375,
EPI_ISL_1307376, EPI_ISL_1307377, EPI_ISL_1307378, EPI_ISL_1307379, EPI_ISL_1307380, EPI_ISL_1307381, EPI_ISL_1307382, EPI_ISL_1307383, EPI_ISL_1307384, EPI_ISL_1307385, EPI_ISL_1307386, EPI_ISL_1307387, EPI_ISL_1307388, EPI_ISL_1307389, EPI_ISL_1307390, EPI_ISL_1307391, EPI_ISL_1307392,
EPI_ISL_1307393, EPI_ISL_1307394, EPI_ISL_1307395, EPI_ISL_1307396, EPI_ISL_1307397, EPI_ISL_1307398, EPI_ISL_1307399, EPI_ISL_1307400, EPI_ISL_1307401, EPI_ISL_1307402, EPI_ISL_1307403, EPI_ISL_1307404, EPI_ISL_1307405, EPI_ISL_1307406, EPI_ISL_1307407, EPI_ISL_1307408, EPI_ISL_1307409,
EPI_ISL_1307410, EPI_ISL_1307411, EPI_ISL_1307412, EPI_ISL_1307413, EPI_ISL_1307414, EPI_ISL_1307415, EPI_ISL_1307416, EPI_ISL_1307417, EPI_ISL_1307418, EPI_ISL_1307419, EPI_ISL_1307420, EPI_ISL_1307421, EPI_ISL_1307422, EPI_ISL_1307423, EPI_ISL_1307424, EPI_ISL_1307425, EPI_ISL_1307426,
EPI_ISL_1307427, EPI_ISL_1307428, EPI_ISL_1307429, EPI_ISL_1307430, EPI_ISL_1307431, EPI_ISL_1307432, EPI_ISL_1307433, EPI_ISL_1307434, EPI_ISL_1307435, EPI_ISL_1307436, EPI_ISL_1307437, EPI_ISL_1307438, EPI_ISL_1307439, EPI_ISL_1307440, EPI_ISL_1307441, EPI_ISL_1307442, EPI_ISL_1307443,
EPI_ISL_1307444, EPI_ISL_1307445, EPI_ISL_1307446, EPI_ISL_1307447, EPI_ISL_1307448, EPI_ISL_1307449, EPI_ISL_1307450, EPI_ISL_1307451, EPI_ISL_1307452, EPI_ISL_1307453, EPI_ISL_1307454, EPI_ISL_1307455, EPI_ISL_1307456, EPI_ISL_1307457, EPI_ISL_1307458, EPI_ISL_1307459, EPI_ISL_1307460,
EPI_ISL_1307461, EPI_ISL_1307462, EPI_ISL_1307463, EPI_ISL_1307464, EPI_ISL_1307465, EPI_ISL_1307466, EPI_ISL_1307467, EPI_ISL_1307468, EPI_ISL_1307469, EPI_ISL_1307470, EPI_ISL_1307471, EPI_ISL_1307472, EPI_ISL_1307473, EPI_ISL_1307474, EPI_ISL_1307475, EPI_ISL_1307476, EPI_ISL_1307477,
EPI_ISL_1307478, EPI_ISL_1307479, EPI_ISL_1307480, EPI_ISL_1307481, EPI_ISL_1307482, EPI_ISL_1307483, EPI_ISL_1307484, EPI_ISL_1307485, EPI_ISL_1307486, EPI_ISL_1307487, EPI_ISL_1307488, EPI_ISL_1307489, EPI_ISL_1307490, EPI_ISL_1307491, EPI_ISL_1307492, EPI_ISL_1307493, EPI_ISL_1307494,
EPI_ISL_1307495, EPI_ISL_1307496, EPI_ISL_1307497, EPI_ISL_1307498, EPI_ISL_1307499, EPI_ISL_1307500, EPI_ISL_1307501, EPI_ISL_1307502, EPI_ISL_1307503, EPI_ISL_1307504, EPI_ISL_1307505, EPI_ISL_1307506, EPI_ISL_1307507, EPI_ISL_1307508, EPI_ISL_1307509, EPI_ISL_1307510, EPI_ISL_1307511,
EPI_ISL_1307512, EPI_ISL_1307513, EPI_ISL_1307514, EPI_ISL_1307515, EPI_ISL_1307516, EPI_ISL_1307517, EPI_ISL_1307518, EPI_ISL_1307519, EPI_ISL_1307520, EPI_ISL_1307521, EPI_ISL_1307522, EPI_ISL_1307523, EPI_ISL_1307524, EPI_ISL_1307525, EPI_ISL_1307526, EPI_ISL_1307527, EPI_ISL_1307528,

| | | | |
|---|---|---|---|
| EPI_ISL_1307529, EPI_ISL_1307530, EPI_ISL_1307531, EPI_ISL_1307532, EPI_ISL_1307533, EPI_ISL_1307534, EPI_ISL_1307535, EPI_ISL_1307536, EPI_ISL_1307537, EPI_ISL_1307538, EPI_ISL_1307539, EPI_ISL_1307540, EPI_ISL_1307541, EPI_ISL_1307542, EPI_ISL_1307543, EPI_ISL_1307544, EPI_ISL_1307545, EPI_ISL_1307546, EPI_ISL_1307547, EPI_ISL_1307548, EPI_ISL_1307549, EPI_ISL_1307550, EPI_ISL_1307551, EPI_ISL_1307552, EPI_ISL_1307553, EPI_ISL_1307554, EPI_ISL_1307555, EPI_ISL_1307556, EPI_ISL_1307557, EPI_ISL_1307558, EPI_ISL_1307559, EPI_ISL_1307560, EPI_ISL_1307561, EPI_ISL_1307562, EPI_ISL_1307563, EPI_ISL_1307564, EPI_ISL_1307565, EPI_ISL_1307566, EPI_ISL_1307567, EPI_ISL_1307568, EPI_ISL_1307569, EPI_ISL_1307570, EPI_ISL_1307571, EPI_ISL_1307572, EPI_ISL_1307573, EPI_ISL_1307574, EPI_ISL_1307575, EPI_ISL_1307576, EPI_ISL_1307577, EPI_ISL_1307578, EPI_ISL_1307579, EPI_ISL_1307580, EPI_ISL_1307581, EPI_ISL_1307582, EPI_ISL_1307583, EPI_ISL_1307584, EPI_ISL_1307585, EPI_ISL_1307586, EPI_ISL_1307587, EPI_ISL_1307588, EPI_ISL_1307589, EPI_ISL_1307590, EPI_ISL_1307591, EPI_ISL_1307592, EPI_ISL_1307593, EPI_ISL_1307594, EPI_ISL_1307595, EPI_ISL_1307596, EPI_ISL_1307597, EPI_ISL_1307598, EPI_ISL_1307599, EPI_ISL_1307600, EPI_ISL_1307601, EPI_ISL_1307602, EPI_ISL_1307603, EPI_ISL_1307604, EPI_ISL_1307605, EPI_ISL_1307606, EPI_ISL_1307607, EPI_ISL_1307608, EPI_ISL_1307609, EPI_ISL_1307610, EPI_ISL_1307611, EPI_ISL_1307612, EPI_ISL_1307613, EPI_ISL_1307614, EPI_ISL_1307615, EPI_ISL_1307616, EPI_ISL_1307617, EPI_ISL_1307618, EPI_ISL_1307619, EPI_ISL_1307620, EPI_ISL_1307621, EPI_ISL_1307622, EPI_ISL_1307623, EPI_ISL_1307624, EPI_ISL_1307625, EPI_ISL_1307626, EPI_ISL_1307627, EPI_ISL_1307628, EPI_ISL_1307629, EPI_ISL_1307630, EPI_ISL_1307631, EPI_ISL_1307632, EPI_ISL_1307633, EPI_ISL_1307634, EPI_ISL_1307635 | | | |
| see above | Pandemic Response Lab - NYC | Pandemic Response Lab, R&D | Henry Lee, Michael Hammerling, Melissa Hopkins, Cybill del Castillo, Shinyoung Clair Kang, William Ward, Pradeep Bugga, Sol Rey, Dylan Law, Haiping Hao, Jon Laurent |
| EPI_ISL_1314552, EPI_ISL_1314553, EPI_ISL_1314556, EPI_ISL_1314568, EPI_ISL_1314573, EPI_ISL_1314589, EPI_ISL_1314590, EPI_ISL_1314608, EPI_ISL_1314613, EPI_ISL_1314624, EPI_ISL_1314630, EPI_ISL_1314643, EPI_ISL_1314646, EPI_ISL_1314659, EPI_ISL_1314664, EPI_ISL_1314677, EPI_ISL_1314700, EPI_ISL_1314708, EPI_ISL_1314709, EPI_ISL_1314739, EPI_ISL_1314760, EPI_ISL_1314782, EPI_ISL_1314809, EPI_ISL_1314812, EPI_ISL_1314813, EPI_ISL_1314829, EPI_ISL_1314830, EPI_ISL_1314914, EPI_ISL_1314922 | | | |
| see above | Quest Diagnostics Incorporated | Centers for Disease Control and Prevention Division of Viral Diseases, Pathogen Discovery | Peter W. Cook, Dakota Howard, Dhwani Batra, Ben L. Rambo-Martin, S. H. Rosenthal, A. Gerasimova, R. M. Kagan, B. Anderson, M. Hua, Y. Liu, L.E. Bernstein, K.E. Livingston, A. Perez, I. A. Shlyakhter, R. V. Rolando, R. Owen, P. Tanpaiboon, F. Lacbawan, Clinton R. Paden, Suxiang Tong, Duncan MacCannell |
| EPI_ISL_1318347, EPI_ISL_1318348, EPI_ISL_1318349 | NYC Department of Health and Mental Hygiene | Centers for Disease Control and Prevention Division of Viral Diseases, Pathogen Discovery | Krista Queen, Yan Li, Ying Tao, Jing Zhang, Anna Uehara, Anna Montmayeur, Clinton R. Paden, Kristen Knipe, Matthew Schmerer, Shoshona Le, Katie Dillon, Peter W. Cook, Rachel Marine, Mili Sheth, Jasmine Padilla, Sarah Nobles, Mark Burroughs, Lori Rowe, Haibin Wang, Ben L. Rambo-Martin, Kristine Lacek, Sam Shepard, Dhwani Batra, Suxiang Tong, Justin Lee |
| EPI_ISL_1319111, EPI_ISL_1319176, EPI_ISL_1319180, EPI_ISL_1319189, EPI_ISL_1319190, EPI_ISL_1319473, EPI_ISL_1319474, EPI_ISL_1319489, EPI_ISL_1319493, EPI_ISL_1319494, EPI_ISL_1319495, EPI_ISL_1319496, EPI_ISL_1319509, EPI_ISL_1319537, EPI_ISL_1319538, EPI_ISL_1319539, EPI_ISL_1319560, EPI_ISL_1319608, EPI_ISL_1319609, EPI_ISL_1319610, EPI_ISL_1319613, EPI_ISL_1319616, EPI_ISL_1319622, EPI_ISL_1319638, EPI_ISL_1319646, EPI_ISL_1319647, EPI_ISL_1319648, EPI_ISL_1319653, EPI_ISL_1319654, EPI_ISL_1319679, EPI_ISL_1319683, EPI_ISL_1319686, EPI_ISL_1319687, EPI_ISL_1319704, EPI_ISL_1319720, EPI_ISL_1319740, EPI_ISL_1319779, EPI_ISL_1319797, EPI_ISL_1319799, EPI_ISL_1319807, EPI_ISL_1319824, EPI_ISL_1319826, EPI_ISL_1319828, EPI_ISL_1319829, EPI_ISL_1319839, EPI_ISL_1319840, EPI_ISL_1319847, EPI_ISL_1319857, EPI_ISL_1319884, EPI_ISL_1319892, EPI_ISL_1319925, EPI_ISL_1319929, EPI_ISL_1319930, EPI_ISL_1319932, EPI_ISL_1319936, EPI_ISL_1319937, EPI_ISL_1319951, EPI_ISL_1319972, EPI_ISL_1320000, EPI_ISL_1320007, EPI_ISL_1320332, EPI_ISL_1320341, EPI_ISL_1320347, EPI_ISL_1320348, EPI_ISL_1320349, EPI_ISL_1320350, EPI_ISL_1320356, EPI_ISL_1320384, EPI_ISL_1320392, EPI_ISL_1320421, EPI_ISL_1320425, EPI_ISL_1320435, EPI_ISL_1320531, EPI_ISL_1320544, EPI_ISL_1320550, EPI_ISL_1320551, EPI_ISL_1320553, EPI_ISL_1320554, EPI_ISL_1320564, EPI_ISL_1320586, EPI_ISL_1320610, EPI_ISL_1320611, EPI_ISL_1320618, EPI_ISL_1320622, EPI_ISL_1320632, EPI_ISL_1320633, EPI_ISL_1320644, EPI_ISL_1320723, EPI_ISL_1320724, EPI_ISL_1320725, EPI_ISL_1320735, EPI_ISL_1320747, EPI_ISL_1320751, EPI_ISL_1320756, EPI_ISL_1320767, EPI_ISL_1320768, EPI_ISL_1320769, EPI_ISL_1320770, EPI_ISL_1320776, EPI_ISL_1320777, EPI_ISL_1320786, EPI_ISL_1320792, EPI_ISL_1320798, EPI_ISL_1320821, EPI_ISL_1320822, EPI_ISL_1320823, EPI_ISL_1320890, EPI_ISL_1320891, EPI_ISL_1320892, EPI_ISL_1320893, EPI_ISL_1320894, EPI_ISL_1320895, EPI_ISL_1320896, EPI_ISL_1320897, EPI_ISL_1320898, EPI_ISL_1320899, EPI_ISL_1320900, EPI_ISL_1320901, EPI_ISL_1320902, EPI_ISL_1320903, EPI_ISL_1320906, EPI_ISL_1320907, EPI_ISL_1320919, EPI_ISL_1320952, EPI_ISL_1320953, EPI_ISL_1320954, EPI_ISL_1320955, EPI_ISL_1320956, EPI_ISL_1320957, EPI_ISL_1320958, EPI_ISL_1320959, EPI_ISL_1320960, EPI_ISL_1320961, EPI_ISL_1320962, EPI_ISL_1320966, EPI_ISL_1320984, EPI_ISL_1321226 | | | |
| see above | Laboratory Corporation of America | Centers for Disease Control and Prevention Division of Viral Diseases, Pathogen Discovery | Peter W. Cook, Dakota Howard, Dhwani Batra, Ben L. Rambo-Martin, Minoo Agarwal, Eyad Almasri, Debbie Boles, Ayla Burns, Nuthawin Charoensri, Oren Cohen, Susan Countryman, Mary Ann Cristobal, Bobbi Croy, Suzanne Dale, Hrushikesh Deshmukh, Amanda Douglas, Vincent Drouillon, Marcia Eisenberg, Howard Engler, Rama Ghatti, Prashant Gupta, Susan Hicks, Jake Humphrey, Lax Iyer, Manoj Jain, Mohan Kolli, Brian Krueger, Tim Kuphal, Stanley Letovsky, Michael Levandoski, Craig Lukasik, Jonathan Meltzer, Brian Norvell, Mindy Nye, Scott Parker, Christos Petropoulos, John Pruitt, Steven Ragan, Scott Ryan, Mike Sapeta, Jana Schroth, Suresh Babu Selvaraju, Goran Stevovic, Amanda Suchanek, Andrea Throop, Lyndon Tilson, Thomas Urban, Joe Voshell, Kimberly Wagner, Jonathan Williams, Mary Williamson, Qian Zeng, Tricia Zwiefelhofer, Clinton R. Paden, Suxiang Tong, Duncan MacCannell |
| EPI_ISL_1322745, EPI_ISL_1322746, EPI_ISL_1322747, EPI_ISL_1322748, EPI_ISL_1322749, EPI_ISL_1322750, EPI_ISL_1322751, EPI_ISL_1322752, EPI_ISL_1322753, EPI_ISL_1322754, EPI_ISL_1322755, EPI_ISL_1322756, EPI_ISL_1322757, EPI_ISL_1322758, EPI_ISL_1322759, EPI_ISL_1322760, EPI_ISL_1322761, EPI_ISL_1322762, EPI_ISL_1322763, EPI_ISL_1322764, EPI_ISL_1322765, EPI_ISL_1322766, EPI_ISL_1322767, EPI_ISL_1322768, EPI_ISL_1322769, EPI_ISL_1322770, EPI_ISL_1322771, EPI_ISL_1322772, EPI_ISL_1322773, EPI_ISL_1322774, EPI_ISL_1322775, EPI_ISL_1322776, EPI_ISL_1322777, EPI_ISL_1322778, EPI_ISL_1322779, EPI_ISL_1322780, EPI_ISL_1322781, EPI_ISL_1322782, EPI_ISL_1322783, EPI_ISL_1322784, EPI_ISL_1322785, EPI_ISL_1322786, EPI_ISL_1322787, EPI_ISL_1322788, EPI_ISL_1322789, EPI_ISL_1322790, EPI_ISL_1322791, EPI_ISL_1322792, EPI_ISL_1322793, EPI_ISL_1322794, EPI_ISL_1322795, EPI_ISL_1322796, EPI_ISL_1322797, EPI_ISL_1322798, EPI_ISL_1322799, EPI_ISL_1322800, EPI_ISL_1322801, EPI_ISL_1322802, EPI_ISL_1322803, EPI_ISL_1322804, EPI_ISL_1322805, EPI_ISL_1322806, EPI_ISL_1322807, EPI_ISL_1322808, EPI_ISL_1322809, EPI_ISL_1322810, EPI_ISL_1322811, EPI_ISL_1322812, EPI_ISL_1322813, EPI_ISL_1322814, EPI_ISL_1322815, EPI_ISL_1322816, EPI_ISL_1322817, EPI_ISL_1322818, EPI_ISL_1322819, EPI_ISL_1322821, EPI_ISL_1322822, EPI_ISL_1322823, EPI_ISL_1322824, EPI_ISL_1322825, EPI_ISL_1322826, EPI_ISL_1322827, EPI_ISL_1322828, EPI_ISL_1322829, EPI_ISL_1322830, EPI_ISL_1322831, EPI_ISL_1322832, EPI_ISL_1322833, EPI_ISL_1322834 | | | |
| see above | NYU Langone Health | Departments of Pathology and Medicine, New York University School of Medicine | Adriana Heguy, Dacia Dimartino, Emily Guzman, Christian Marier, Peter Meyn, Sitharam Ramaswami, Gael Westby, Paul Zappile, Yutong Zhang, Paolo Cotzia, Guiqing Wang |
| EPI_ISL_1335914, EPI_ISL_1335930, EPI_ISL_1336026, EPI_ISL_1336102 | Quest Diagnostics Incorporated | Centers for Disease Control and Prevention Division of Viral Diseases, Pathogen Discovery | Peter W. Cook, Dakota Howard, Dhwani Batra, Ben L. Rambo-Martin, S. H. Rosenthal, A. Gerasimova, R. M. Kagan, B. Anderson, M. Hua, Y. Liu, L.E. Bernstein, K.E. Livingston, A. Perez, I. A. Shlyakhter, R. V. Rolando, R. Owen, P. Tanpaiboon, F. Lacbawan, Clinton R. Paden, Suxiang Tong, Duncan MacCannell |
| EPI_ISL_1337041 | Helix/Illumina | Centers for Disease Control and Prevention Division of Viral Diseases, Pathogen Discovery | Peter W. Cook, Dakota Howard, Dhwani Batra, Ben L. Rambo-Martin, Eileen de Feo, Jan Antico, Christine Tran, Matthew Tolentino, Shannon Wickline, Kim Gietzen, Brad Sickler, Jingtao Liu, Eric Allen, Phil Febbo, Summer Galloway, Nicole L. Washington, Simon White, Geraint Levan, Kelly Schiabor Barrett, Elizabeth Cirulli, Alexandre Bolze, Ary Ascencio, Charlotte Rivera-Garcia, Ryan Cho, Jason Nguyen, Sherry Wang, Jimmy Ramirez, Tyler Cassens, Efren Sandoval, Magnus Isaksson, William Lee, David Becker, Marc Laurent, James Lu, Clinton R. Paden, Suxiang Tong, Duncan MacCannell |
| EPI_ISL_1337463 | DOHMH Morrisania | New York City Public Health Laboratory | Jade Wang, et al. |
| EPI_ISL_1337464 | DOHMH Chelsea | New York City Public Health Laboratory | Jade Wang, et al. |
| EPI_ISL_1337465 | DOHMH Central Harlem | New York City Public Health Laboratory | Jade Wang, et al. |
| EPI_ISL_1337466 | Department of Homeless Services | New York City Public Health Laboratory | Jade Wang, et al. |
| EPI_ISL_1337467 | DOHMH Corona | New York City Public Health Laboratory | Jade Wang, et al. |
| EPI_ISL_1337468 | DOHMH Fort Greene | New York City Public Health Laboratory | Jade Wang, et al. |
| EPI_ISL_1337469 | DOHMH Jamaica | New York City Public Health Laboratory | Jade Wang, et al. |
| EPI_ISL_1337470 | DOHMH Morrisania | New York City Public Health Laboratory | Jade Wang, et al. |
| EPI_ISL_1337471 | DOHMH Fort Greene | New York City Public Health Laboratory | Jade Wang, et al. |
| EPI_ISL_1337472 | Department of Homeless Services | New York City Public Health Laboratory | Jade Wang, et al. |
| EPI_ISL_1337473 | DOHMH Morrisania | New York City Public Health Laboratory | Jade Wang, et al. |
| EPI_ISL_1337474 | DOHMH Crown Heights | New York City Public Health Laboratory | Jade Wang, et al. |
| EPI_ISL_1337475 | DOHMH Corona | New York City Public Health Laboratory | Jade Wang, et al. |
| EPI_ISL_1337476 | DOHMH Central Harlem | New York City Public Health Laboratory | Jade Wang, et al. |
| EPI_ISL_1337477 | DOHMH Corona | New York City Public Health Laboratory | Jade Wang, et al. |
| EPI_ISL_1337478 | DOHMH Morrisania | New York City Public Health Laboratory | Jade Wang, et al. |
| EPI_ISL_1337479 | DOHMH Chelsea | New York City Public Health Laboratory | Jade Wang, et al. |
| EPI_ISL_1337480 | DOHMH Riverside | New York City Public Health Laboratory | Jade Wang, et al. |
| EPI_ISL_1337481 | DOHMH Morrisania | New York City Public Health Laboratory | Jade Wang, et al. |
| EPI_ISL_1337482 | OCME Office Of Chief Medical Examiner | New York City Public Health Laboratory | Jade Wang, et al. |
| EPI_ISL_1337483 | DOHMH Riverside | New York City Public Health Laboratory | Jade Wang, et al. |
| EPI_ISL_1337484 | DOHMH Corona | New York City Public Health Laboratory | Jade Wang, et al. |
| EPI_ISL_1337485 | DOHMH Morrisania | New York City Public Health Laboratory | Jade Wang, et al. |

| | | | |
|---|---|---|---|
| EPI_ISL_1337486, EPI_ISL_1337487 | DOHMH Jamaica | New York City Public Health Laboratory | Jade Wang, et al. |
| EPI_ISL_1337488 | DOHMH Crown Heights | New York City Public Health Laboratory | Jade Wang, et al. |
| EPI_ISL_1337489 | DOHMH Central Harlem | New York City Public Health Laboratory | Jade Wang, et al. |
| EPI_ISL_1337490 | DOHMH Crown Heights | New York City Public Health Laboratory | Jade Wang, et al. |
| EPI_ISL_1337491 | DOHMH Corona | New York City Public Health Laboratory | Jade Wang, et al. |
| EPI_ISL_1337492, EPI_ISL_1337493, EPI_ISL_1337494 | DOHMH Crown Heights | New York City Public Health Laboratory | Jade Wang, et al. |
| EPI_ISL_1337495, EPI_ISL_1337496, EPI_ISL_1337497, EPI_ISL_1337498 | DOHMH Jamaica | New York City Public Health Laboratory | Jade Wang, et al. |
| EPI_ISL_1337499 | DOHMH Crown Heights | New York City Public Health Laboratory | Jade Wang, et al. |
| EPI_ISL_1337500 | DOHMH Chelsea | New York City Public Health Laboratory | Jade Wang, et al. |
| EPI_ISL_1337501 | DOHMH Jamaica | New York City Public Health Laboratory | Jade Wang, et al. |
| EPI_ISL_1337502 | DOHMH Morrisania | New York City Public Health Laboratory | Jade Wang, et al. |
| EPI_ISL_1337503 | DOHMH Riverside | New York City Public Health Laboratory | Jade Wang, et al. |
| EPI_ISL_1337504, EPI_ISL_1337505 | DOHMH Jamaica | New York City Public Health Laboratory | Jade Wang, et al. |
| EPI_ISL_1337506 | DOHMH Morrisania | New York City Public Health Laboratory | Jade Wang, et al. |
| EPI_ISL_1337507 | DOHMH Corona | New York City Public Health Laboratory | Jade Wang, et al. |
| EPI_ISL_1337508, EPI_ISL_1337509 | DOHMH PHL | New York City Public Health Laboratory | Jade Wang, et al. |
| EPI_ISL_1337510 | DOHMH Fort Greene | New York City Public Health Laboratory | Jade Wang, et al. |
| EPI_ISL_1337511, EPI_ISL_1337512 | DOHMH Corona | New York City Public Health Laboratory | Jade Wang, et al. |
| EPI_ISL_1337513 | DOHMH Jamaica | New York City Public Health Laboratory | Jade Wang, et al. |
| EPI_ISL_1337514, EPI_ISL_1337515 | DOHMH Corona | New York City Public Health Laboratory | Jade Wang, et al. |
| EPI_ISL_1337516 | DOHMH PHL | New York City Public Health Laboratory | Jade Wang, et al. |
| EPI_ISL_1337517 | DOHMH Central Harlem | New York City Public Health Laboratory | Jade Wang, et al. |
| EPI_ISL_1337518, EPI_ISL_1337519 | OCME Office Of Chief Medical Examiner | New York City Public Health Laboratory | Jade Wang, et al. |
| EPI_ISL_1337520 | DOHMH Central Harlem | New York City Public Health Laboratory | Jade Wang, et al. |
| EPI_ISL_1337521, EPI_ISL_1337522 | DOHMH Jamaica | New York City Public Health Laboratory | Jade Wang, et al. |
| EPI_ISL_1337523 | DOHMH Morrisania | New York City Public Health Laboratory | Jade Wang, et al. |
| EPI_ISL_1337524 | OCME Office Of Chief Medical Examiner | New York City Public Health Laboratory | Jade Wang, et al. |
| EPI_ISL_1337525 | DOHMH Jamaica | New York City Public Health Laboratory | Jade Wang, et al. |
| EPI_ISL_1337526 | DOHMH Riverside | New York City Public Health Laboratory | Jade Wang, et al. |
| EPI_ISL_1337527 | OCME Office Of Chief Medical Examiner | New York City Public Health Laboratory | Jade Wang, et al. |
| EPI_ISL_1337528 | DOHMH Crown Heights | New York City Public Health Laboratory | Jade Wang, et al. |
| EPI_ISL_1337529 | OCME Office Of Chief Medical Examiner | New York City Public Health Laboratory | Jade Wang, et al. |
| EPI_ISL_1337530 | DOHMH PHL | New York City Public Health Laboratory | Jade Wang, et al. |
| EPI_ISL_1337531 | DOHMH Corona | New York City Public Health Laboratory | Jade Wang, et al. |
| EPI_ISL_1337532, EPI_ISL_1337533 | DOHMH Jamaica | New York City Public Health Laboratory | Jade Wang, et al. |
| EPI_ISL_1337534, EPI_ISL_1337535 | DOHMH Crown Heights | New York City Public Health Laboratory | Jade Wang, et al. |
| EPI_ISL_1337536 | DOHMH Central Harlem | New York City Public Health Laboratory | Jade Wang, et al. |
| EPI_ISL_1337537, EPI_ISL_1337538 | DOHMH Jamaica | New York City Public Health Laboratory | Jade Wang, et al. |
| EPI_ISL_1337539 | OCME Office Of Chief Medical Examiner | New York City Public Health Laboratory | Jade Wang, et al. |
| EPI_ISL_1337540 | DOHMH Morrisania | New York City Public Health Laboratory | Jade Wang, et al. |
| EPI_ISL_1337541 | OCME Office Of Chief Medical Examiner | New York City Public Health Laboratory | Jade Wang, et al. |
| EPI_ISL_1337542 | DOHMH Chelsea | New York City Public Health Laboratory | Jade Wang, et al. |
| EPI_ISL_1337543 | DOHMH Riverside | New York City Public Health Laboratory | Jade Wang, et al. |
| EPI_ISL_1337544 | DOHMH Crown Heights | New York City Public Health Laboratory | Jade Wang, et al. |
| EPI_ISL_1337545 | DOHMH Morrisania | New York City Public Health Laboratory | Jade Wang, et al. |
| EPI_ISL_1337546 | OCME Office Of Chief Medical Examiner | New York City Public Health Laboratory | Jade Wang, et al. |
| EPI_ISL_1337547 | DOHMH Riverside | New York City Public Health Laboratory | Jade Wang, et al. |
| EPI_ISL_1337548, EPI_ISL_1337549 | DOHMH Central Harlem | New York City Public Health Laboratory | Jade Wang, et al. |
| EPI_ISL_1337550 | DOHMH Jamaica | New York City Public Health Laboratory | Jade Wang, et al. |
| EPI_ISL_1337551 | DOHMH Corona | New York City Public Health Laboratory | Jade Wang, et al. |
| EPI_ISL_1338149, EPI_ISL_1338229, EPI_ISL_1338263 | Helix/Illumina | Centers for Disease Control and Prevention Division of Viral Diseases, Pathogen Discovery | Peter W. Cook, Dakota Howard, Dhwani Batra, Ben L. Rambo-Martin, Eileen de Feo, Jan Antico, Christine Tran, Matthew Tolentino, Shannon Wickline, Kim Gietzen, Brad Sickler, Jingtao Liu, Eric Allen, Phil Febbo, Summer Galloway, Nicole L. Washington, Simon White, Geraint Levan, Kelly Schiabor Barrett, Elizabeth Cirulli, Alexandre Bolze, Ary Ascencio, Charlotte Rivera-Garcia, Ryan Cho, Jason Nguyen, Sherry Wang, Jimmy Ramirez, Tyler Cassens, Efren Sandoval, Magnus Isaksson, William Lee, David Becker, Marc Laurent, James Lu, Clinton R. Paden, Suxiang Tong, Duncan MacCannell |
| EPI_ISL_1338400, EPI_ISL_1338401, EPI_ISL_1338521, EPI_ISL_1338525, EPI_ISL_1338526, EPI_ISL_1338527, EPI_ISL_1338530, EPI_ISL_1338531, EPI_ISL_1338532, EPI_ISL_1338533, EPI_ISL_1338534, EPI_ISL_1338535, EPI_ISL_1338536, EPI_ISL_1338541, EPI_ISL_1338625, EPI_ISL_1338627, EPI_ISL_1338629, EPI_ISL_1338644, EPI_ISL_1338649, EPI_ISL_1338653, EPI_ISL_1338658, EPI_ISL_1338660, EPI_ISL_1338680, EPI_ISL_1338681, EPI_ISL_1338682, EPI_ISL_1338683, EPI_ISL_1338687, EPI_ISL_1338705, EPI_ISL_1338720, EPI_ISL_1338721, EPI_ISL_1338730, EPI_ISL_1338737, EPI_ISL_1338846, EPI_ISL_1338847, EPI_ISL_1339180, EPI_ISL_1339237, EPI_ISL_1339238, EPI_ISL_1339241, EPI_ISL_1339242, EPI_ISL_1339247, EPI_ISL_1339498, EPI_ISL_1339505, EPI_ISL_1339522, EPI_ISL_1339526, EPI_ISL_1339529, EPI_ISL_1339535, EPI_ISL_1339536, EPI_ISL_1339543, EPI_ISL_1339544, EPI_ISL_1339573, EPI_ISL_1339577, EPI_ISL_1339584, EPI_ISL_1339585, EPI_ISL_1339591, EPI_ISL_1339628, EPI_ISL_1339638, EPI_ISL_1339639, EPI_ISL_1339642 | | | |
| see above | Laboratory Corporation of America | Centers for Disease Control and Prevention Division of Viral Diseases, Pathogen Discovery | Peter W. Cook, Dakota Howard, Dhwani Batra, Ben L. Rambo-Martin, Minoo Agarwal, Eyad Almasri, Debbie Boles, Ayla Burns, Nuthawin Charoensri, Oren Cohen, Susan Countryman, Mary Ann Cristobal, Bobbi Croy, Suzanne Dale, Hrushikesh Deshmukh, Amanda Douglas, Vincent Drouillon, Marcia |

| | | | |
|---|---|---|---|
| | | | Eisenberg, Howard Engler, Rama Ghatti, Prashant Gupta, Susan Hicks, Jake Humphrey, Lax Iyer, Manoj Jain, Mohan Kolli, Brian Krueger, Tim Kuphal, Stanley Letovsky, Michael Levandoski, Craig Lukasik, Jonathan Meltzer, Brian Norvell, Mindy Nye, Scott Parker, Christos Petropoulos, John Pruitt, Steven Ragan, Scott Ryan, Mike Sapeta, Jana Schroth, Suresh Babu Selvaraju, Goran Stevovic, Amanda Suchanek, Andrea Throop, Lyndon Tilson, Thomas Urban, Joe Voshell, Kimberly Wagner, Jonathan Williams, Mary Williamson, Qian Zeng, Tricia Zwiefelhofer, Clinton R. Paden, Suxiang Tong, Duncan MacCannell |
| EPI_ISL_1339842 | Quest Diagnostics Incorporated | Centers for Disease Control and Prevention Division of Viral Diseases, Pathogen Discovery | Peter W. Cook, Dakota Howard, Dhwani Batra, Ben L. Rambo-Martin, S. H. Rosenthal, A. Gerasimova, R. M. Kagan, B. Anderson, M. Hua, Y. Liu, L.E. Bernstein, K.E. Livingston, A. Perez, I. A. Shlyakhter, R. V. Rolando, R. Owen, P. Tanpaiboon, F. Lacbawan, Clinton R. Paden, Suxiang Tong, Duncan MacCannell |
| EPI_ISL_1340378, EPI_ISL_1340381, EPI_ISL_1340392, EPI_ISL_1340441 | Helix/Illumina | Centers for Disease Control and Prevention Division of Viral Diseases, Pathogen Discovery | Peter W. Cook, Dakota Howard, Dhwani Batra, Ben L. Rambo-Martin, Eileen de Feo, Jan Antico, Christine Tran, Matthew Tolentino, Shannon Wickline, Kim Gietzen, Brad Sickler, Jingtao Liu, Eric Allen, Phil Febbo, Summer Galloway, Nicole L. Washington, Simon White, Geraint Levan, Kelly Schiabor Barrett, Elizabeth Cirulli, Alexandre Bolze, Ary Ascencio, Charlotte Rivera-Garcia, Ryan Cho, Jason Nguyen, Sherry Wang, Jimmy Ramirez, Tyler Cassens, Efren Sandoval, Magnus Isaksson, William Lee, David Becker, Marc Laurent, James Lu, Clinton R. Paden, Suxiang Tong, Duncan MacCannell |
| EPI_ISL_1340452, EPI_ISL_1340472, EPI_ISL_1340475, EPI_ISL_1340478, EPI_ISL_1340511, EPI_ISL_1340521, EPI_ISL_1340735, EPI_ISL_1367423, EPI_ISL_1367441, EPI_ISL_1367453, EPI_ISL_1367467, EPI_ISL_1367479 | | | |
| see above | Quest Diagnostics Incorporated | Centers for Disease Control and Prevention Division of Viral Diseases, Pathogen Discovery | Peter W. Cook, Dakota Howard, Dhwani Batra, Ben L. Rambo-Martin, S. H. Rosenthal, A. Gerasimova, R. M. Kagan, B. Anderson, M. Hua, Y. Liu, L.E. Bernstein, K.E. Livingston, A. Perez, I. A. Shlyakhter, R. V. Rolando, R. Owen, P. Tanpaiboon, F. Lacbawan, Clinton R. Paden, Suxiang Tong, Duncan MacCannell |
| EPI_ISL_1378687 | Yale Clinical Virology Lab | Grubaugh Lab - Yale School of Public Health | Joseph Fauver, Mallery Breban, Isabell Ott, Tara Alpert, Mary Petrone, Anderson Brito, Chantal Vogels, Annie Watkins, Chaney Kalinich, Jessica Rothman, Marie L. Landry, Nathan Grubaugh |
| EPI_ISL_1378790, EPI_ISL_1378791, EPI_ISL_1378792, EPI_ISL_1378793, EPI_ISL_1378794, EPI_ISL_1378795, EPI_ISL_1378796 | Tempus | Grubaugh Lab - Yale School of Public Health | Joseph Fauver, Mallery Breban, Isabell Ott, Tara Alpert, Mary Petrone, Anderson Brito, Chantal Vogels, Annie Watkins, Chaney Kalinich, Jessica Rothman, Matthew J. MacKay, Gaurav Khullar, Jessica Metti, Joel T. Dudley, Megan Nash, Nike Beaubier, Christopher E. Mason, Nathan Grubaugh |
| EPI_ISL_1378824, EPI_ISL_1378825, EPI_ISL_1378826 | Murphy Medical Association | Grubaugh Lab - Yale School of Public Health | Joseph Fauver, Mallery Breban, Isabell Ott, Tara Alpert, Mary Petrone, Anderson Brito, Chantal Vogels, Annie Watkins, Chaney Kalinich, Jessica Rothman, Caleb Neal, Eva Laszlo, Steven Murphy, Nathan Grubaugh |
| EPI_ISL_1379198, EPI_ISL_1379199, EPI_ISL_1379200, EPI_ISL_1379201, EPI_ISL_1379202, EPI_ISL_1379203, EPI_ISL_1379204, EPI_ISL_1379205, EPI_ISL_1379206, EPI_ISL_1379207, EPI_ISL_1379208, EPI_ISL_1379209, EPI_ISL_1379210, EPI_ISL_1379211, EPI_ISL_1379212, EPI_ISL_1379213, EPI_ISL_1379214, EPI_ISL_1379215, EPI_ISL_1379216, EPI_ISL_1379217, EPI_ISL_1379218, EPI_ISL_1379219, EPI_ISL_1379220, EPI_ISL_1379221, EPI_ISL_1379222, EPI_ISL_1379223, EPI_ISL_1379224, EPI_ISL_1379225, EPI_ISL_1379226, EPI_ISL_1379227, EPI_ISL_1379228, EPI_ISL_1379229, EPI_ISL_1379230, EPI_ISL_1379231, EPI_ISL_1379232, EPI_ISL_1379233, EPI_ISL_1379234, EPI_ISL_1379235, EPI_ISL_1379236, EPI_ISL_1379237, EPI_ISL_1379238, EPI_ISL_1379239, EPI_ISL_1379240, EPI_ISL_1379241, EPI_ISL_1379242, EPI_ISL_1379243, EPI_ISL_1379244, EPI_ISL_1379245, EPI_ISL_1379246, EPI_ISL_1379247, EPI_ISL_1379248, EPI_ISL_1379249, EPI_ISL_1379250, EPI_ISL_1379251, EPI_ISL_1379252, EPI_ISL_1379253, EPI_ISL_1379254, EPI_ISL_1379255, EPI_ISL_1379256, EPI_ISL_1379257, EPI_ISL_1379258, EPI_ISL_1379259, EPI_ISL_1379260, EPI_ISL_1379261, EPI_ISL_1379262, EPI_ISL_1379263, EPI_ISL_1379264, EPI_ISL_1379265, EPI_ISL_1379266, EPI_ISL_1379339, EPI_ISL_1379340, EPI_ISL_1379341, EPI_ISL_1379342, EPI_ISL_1379343, EPI_ISL_1379344, EPI_ISL_1379345, EPI_ISL_1379346, EPI_ISL_1379347, EPI_ISL_1379348, EPI_ISL_1379349, EPI_ISL_1379350, EPI_ISL_1379351, EPI_ISL_1379352, EPI_ISL_1379353, EPI_ISL_1379354, EPI_ISL_1379355, EPI_ISL_1379356, EPI_ISL_1379357, EPI_ISL_1379358, EPI_ISL_1379359, EPI_ISL_1379360, EPI_ISL_1379361, EPI_ISL_1379362, EPI_ISL_1379363, EPI_ISL_1379364, EPI_ISL_1379365, EPI_ISL_1379366, EPI_ISL_1379367, EPI_ISL_1379368, EPI_ISL_1379369, EPI_ISL_1379370, EPI_ISL_1379371, EPI_ISL_1379372, EPI_ISL_1379373, EPI_ISL_1379374, EPI_ISL_1379375, EPI_ISL_1379376, EPI_ISL_1379377, EPI_ISL_1379378, EPI_ISL_1379379, EPI_ISL_1379380, EPI_ISL_1379381, EPI_ISL_1379382, EPI_ISL_1379383, EPI_ISL_1379384, EPI_ISL_1379385, EPI_ISL_1379386, EPI_ISL_1379387, EPI_ISL_1379388, EPI_ISL_1379389, EPI_ISL_1379390 | | | |
| see above | NYP-WCM | New York Genome Center | Michael Zody, Andre Corvelo, Dayna M. Oschwald, Samantha Fennessey, Tom Maniatis, Melissa Cushing, Olivier Elemento, Margaret Elizabeth Ross, Chris Mason, Priya Velu, Hanna Rennert, Arryn Craney, Lars F Westblade |
| EPI_ISL_1384885, EPI_ISL_1384886, EPI_ISL_1384887, EPI_ISL_1384888, EPI_ISL_1384889, EPI_ISL_1384890, EPI_ISL_1384891, EPI_ISL_1384892, EPI_ISL_1384893, EPI_ISL_1384894, EPI_ISL_1384895, EPI_ISL_1384896, EPI_ISL_1384897, EPI_ISL_1384898, EPI_ISL_1384899, EPI_ISL_1384900, EPI_ISL_1384901, EPI_ISL_1384902, EPI_ISL_1384903, EPI_ISL_1384904, EPI_ISL_1384905, EPI_ISL_1384906, EPI_ISL_1384907, EPI_ISL_1384908, EPI_ISL_1384909, EPI_ISL_1384910, EPI_ISL_1384911, EPI_ISL_1384912, EPI_ISL_1384913, EPI_ISL_1384914, EPI_ISL_1384915, EPI_ISL_1384916, EPI_ISL_1384917, EPI_ISL_1384918, EPI_ISL_1384919, EPI_ISL_1384920, EPI_ISL_1384921, EPI_ISL_1384922, EPI_ISL_1384923, EPI_ISL_1384924, EPI_ISL_1384925, EPI_ISL_1384926, EPI_ISL_1384927, EPI_ISL_1384928, EPI_ISL_1384929, EPI_ISL_1384930, EPI_ISL_1384931, EPI_ISL_1384932, EPI_ISL_1384933, EPI_ISL_1384934, EPI_ISL_1384935, EPI_ISL_1384936, EPI_ISL_1384937, EPI_ISL_1384938, EPI_ISL_1384939, EPI_ISL_1384940, EPI_ISL_1384941, EPI_ISL_1384942, EPI_ISL_1384943, EPI_ISL_1384944, EPI_ISL_1384945, EPI_ISL_1384946, EPI_ISL_1384947, EPI_ISL_1384948, EPI_ISL_1384949, EPI_ISL_1384950, EPI_ISL_1384951, EPI_ISL_1384952, EPI_ISL_1384953, EPI_ISL_1384954, EPI_ISL_1384955, EPI_ISL_1384956, EPI_ISL_1384957, EPI_ISL_1384958, EPI_ISL_1384959, EPI_ISL_1384960, EPI_ISL_1384961, EPI_ISL_1384962, EPI_ISL_1384963, EPI_ISL_1384964, EPI_ISL_1384965, EPI_ISL_1384966, EPI_ISL_1384967, EPI_ISL_1384968, EPI_ISL_1384969, EPI_ISL_1384970, EPI_ISL_1384971, EPI_ISL_1384972, EPI_ISL_1384973, EPI_ISL_1384974, EPI_ISL_1384975, EPI_ISL_1384976, EPI_ISL_1384977, EPI_ISL_1384978, EPI_ISL_1384979, EPI_ISL_1384980, EPI_ISL_1384981, EPI_ISL_1384982, EPI_ISL_1384983, EPI_ISL_1384984, EPI_ISL_1384985, EPI_ISL_1384986, EPI_ISL_1384987, EPI_ISL_1384988, EPI_ISL_1384989, EPI_ISL_1384990, EPI_ISL_1384991, EPI_ISL_1384992, EPI_ISL_1384993, EPI_ISL_1384994, EPI_ISL_1384995, EPI_ISL_1384996, EPI_ISL_1384997, EPI_ISL_1384998, EPI_ISL_1384999, EPI_ISL_1385000, EPI_ISL_1385001, EPI_ISL_1385002, EPI_ISL_1385003, EPI_ISL_1385004, EPI_ISL_1385005, EPI_ISL_1385006, EPI_ISL_1385007, EPI_ISL_1385008, EPI_ISL_1385009, EPI_ISL_1385010, EPI_ISL_1385011, EPI_ISL_1385012, EPI_ISL_1385013, EPI_ISL_1385014, EPI_ISL_1385015, EPI_ISL_1385016, EPI_ISL_1385017, EPI_ISL_1385018, EPI_ISL_1385019, EPI_ISL_1385020, EPI_ISL_1385021, EPI_ISL_1385022, EPI_ISL_1385023, EPI_ISL_1385024, EPI_ISL_1385025, EPI_ISL_1385026, EPI_ISL_1385027, EPI_ISL_1385028, EPI_ISL_1385029, EPI_ISL_1385030, EPI_ISL_1385031, EPI_ISL_1385032, EPI_ISL_1385033, EPI_ISL_1385034, EPI_ISL_1385035, EPI_ISL_1385036, EPI_ISL_1385037, EPI_ISL_1385038, EPI_ISL_1385039, EPI_ISL_1385040, EPI_ISL_1385041, EPI_ISL_1385042, EPI_ISL_1385043, EPI_ISL_1385044, EPI_ISL_1385045, EPI_ISL_1385046, EPI_ISL_1385047, EPI_ISL_1385048, EPI_ISL_1385049, EPI_ISL_1385050, EPI_ISL_1385051, EPI_ISL_1385052, EPI_ISL_1385053, EPI_ISL_1385054, EPI_ISL_1385055, EPI_ISL_1385056, EPI_ISL_1385057, EPI_ISL_1385058, EPI_ISL_1385059, EPI_ISL_1385060, EPI_ISL_1385061, EPI_ISL_1385062, EPI_ISL_1385063, EPI_ISL_1385064, EPI_ISL_1385065, EPI_ISL_1385066, EPI_ISL_1385067, EPI_ISL_1385068, EPI_ISL_1385069, EPI_ISL_1385070, EPI_ISL_1385071, EPI_ISL_1385072, EPI_ISL_1385073, EPI_ISL_1385074, EPI_ISL_1385075, EPI_ISL_1385076, EPI_ISL_1385077, EPI_ISL_1385078, EPI_ISL_1385079, EPI_ISL_1385080, EPI_ISL_1385081, EPI_ISL_1385082, EPI_ISL_1385083, EPI_ISL_1385084, EPI_ISL_1385085, EPI_ISL_1385086, EPI_ISL_1385087, EPI_ISL_1385088, EPI_ISL_1385089, EPI_ISL_1385090, EPI_ISL_1385091, EPI_ISL_1385092, EPI_ISL_1385093, EPI_ISL_1385094, EPI_ISL_1385095, EPI_ISL_1385096, EPI_ISL_1385097, EPI_ISL_1385098, EPI_ISL_1385099, EPI_ISL_1385100, EPI_ISL_1385101, EPI_ISL_1385102, EPI_ISL_1385103, EPI_ISL_1385104, EPI_ISL_1385105, EPI_ISL_1385106, EPI_ISL_1385107, EPI_ISL_1385108, EPI_ISL_1385109, EPI_ISL_1385110, EPI_ISL_1385111, EPI_ISL_1385112, EPI_ISL_1385113, EPI_ISL_1385114, EPI_ISL_1385115, EPI_ISL_1385116, EPI_ISL_1385117, EPI_ISL_1385118, EPI_ISL_1385119, EPI_ISL_1385120, EPI_ISL_1385121, EPI_ISL_1385122, EPI_ISL_1385123, EPI_ISL_1385124, EPI_ISL_1385125, EPI_ISL_1385126, EPI_ISL_1385127, EPI_ISL_1385128, EPI_ISL_1385129, EPI_ISL_1385130, EPI_ISL_1385131, EPI_ISL_1385132, EPI_ISL_1385133, EPI_ISL_1385134, EPI_ISL_1385135, EPI_ISL_1385136, EPI_ISL_1385137, EPI_ISL_1385138, EPI_ISL_1385139, EPI_ISL_1385140, EPI_ISL_1385141, EPI_ISL_1385142, EPI_ISL_1385143, EPI_ISL_1385144, EPI_ISL_1385145, EPI_ISL_1385146, EPI_ISL_1385147, EPI_ISL_1385148, EPI_ISL_1385149, EPI_ISL_1385150, EPI_ISL_1385151, EPI_ISL_1385152, EPI_ISL_1385153, EPI_ISL_1385154, EPI_ISL_1385155, EPI_ISL_1385156, EPI_ISL_1385157, EPI_ISL_1385158, EPI_ISL_1385159, EPI_ISL_1385160, EPI_ISL_1385161, EPI_ISL_1385162, EPI_ISL_1385163, EPI_ISL_1385164, EPI_ISL_1385165, EPI_ISL_1385166, EPI_ISL_1385167, EPI_ISL_1385168, EPI_ISL_1385169, EPI_ISL_1385170, EPI_ISL_1385171, EPI_ISL_1385172, EPI_ISL_1385173, EPI_ISL_1385174, EPI_ISL_1385175, EPI_ISL_1385176, EPI_ISL_1385177, EPI_ISL_1385178, EPI_ISL_1385179, EPI_ISL_1385180, EPI_ISL_1385181, EPI_ISL_1385182, EPI_ISL_1385183, EPI_ISL_1385184, EPI_ISL_1385185, EPI_ISL_1385186, EPI_ISL_1385187, EPI_ISL_1385188, EPI_ISL_1385189, EPI_ISL_1385190, EPI_ISL_1385191, EPI_ISL_1385192, EPI_ISL_1385193, EPI_ISL_1385194, EPI_ISL_1385195, EPI_ISL_1385196, EPI_ISL_1385197, EPI_ISL_1385198, EPI_ISL_1385199, EPI_ISL_1385200, EPI_ISL_1385201, EPI_ISL_1385202, EPI_ISL_1385203, EPI_ISL_1385204, EPI_ISL_1385205, EPI_ISL_1385206, EPI_ISL_1385207, EPI_ISL_1385208, EPI_ISL_1385209, EPI_ISL_1385210, EPI_ISL_1385211, EPI_ISL_1385212, EPI_ISL_1385213, EPI_ISL_1385214, EPI_ISL_1385215, EPI_ISL_1385216, EPI_ISL_1385217, EPI_ISL_1385218, EPI_ISL_1385219, EPI_ISL_1385220, EPI_ISL_1385221, EPI_ISL_1385222, EPI_ISL_1385223, EPI_ISL_1385224, EPI_ISL_1385225, EPI_ISL_1385226, EPI_ISL_1385227, EPI_ISL_1385228, EPI_ISL_1385229, EPI_ISL_1385230, EPI_ISL_1385231, EPI_ISL_1385232, EPI_ISL_1385233, EPI_ISL_1385234, EPI_ISL_1385235, EPI_ISL_1385236, EPI_ISL_1385237, EPI_ISL_1385238, EPI_ISL_1385239, EPI_ISL_1385240, EPI_ISL_1385241, EPI_ISL_1385242, EPI_ISL_1385243, EPI_ISL_1385244, EPI_ISL_1385245, EPI_ISL_1385246, EPI_ISL_1385247, EPI_ISL_1385248, EPI_ISL_1385249, EPI_ISL_1385250, EPI_ISL_1385251, EPI_ISL_1385252, EPI_ISL_1385253, EPI_ISL_1385254, EPI_ISL_1385255, EPI_ISL_1385256, EPI_ISL_1385257, EPI_ISL_1385258, EPI_ISL_1385259, EPI_ISL_1385260, EPI_ISL_1385261, EPI_ISL_1385262, EPI_ISL_1385263, EPI_ISL_1385264, EPI_ISL_1385265, EPI_ISL_1385266, EPI_ISL_1385267, EPI_ISL_1385268, EPI_ISL_1385269, EPI_ISL_1385270, EPI_ISL_1385271, EPI_ISL_1385272, EPI_ISL_1385273, EPI_ISL_1385274, EPI_ISL_1385275, EPI_ISL_1385276, EPI_ISL_1385277, EPI_ISL_1385278, EPI_ISL_1385279, EPI_ISL_1385280, EPI_ISL_1385281, EPI_ISL_1385282, EPI_ISL_1385283, EPI_ISL_1385284, EPI_ISL_1385285, EPI_ISL_1385286, EPI_ISL_1385287, EPI_ISL_1385288, EPI_ISL_1385289, EPI_ISL_1385290, EPI_ISL_1385291, EPI_ISL_1385292, EPI_ISL_1385293, EPI_ISL_1385294, EPI_ISL_1385295, EPI_ISL_1385296, EPI_ISL_1385297, EPI_ISL_1385298, EPI_ISL_1385299, EPI_ISL_1385300, EPI_ISL_1385301, EPI_ISL_1385302, EPI_ISL_1385303, EPI_ISL_1385304, EPI_ISL_1385305, EPI_ISL_1385306, EPI_ISL_1385307, EPI_ISL_1385308, EPI_ISL_1385309, EPI_ISL_1385310, EPI_ISL_1385311, EPI_ISL_1385312, EPI_ISL_1385313, EPI_ISL_1385314, EPI_ISL_1385315, EPI_ISL_1385316, EPI_ISL_1385317, EPI_ISL_1385318, EPI_ISL_1385319, EPI_ISL_1385320, EPI_ISL_1385321, EPI_ISL_1385322, EPI_ISL_1385323, EPI_ISL_1385324, EPI_ISL_1385325, EPI_ISL_1385326, EPI_ISL_1385327, EPI_ISL_1385328, EPI_ISL_1385329, EPI_ISL_1385330, EPI_ISL_1385331, EPI_ISL_1385332, EPI_ISL_1385333, EPI_ISL_1385334, EPI_ISL_1385335, EPI_ISL_1385336, EPI_ISL_1385337, EPI_ISL_1385338, EPI_ISL_1385339, EPI_ISL_1385340, EPI_ISL_1385341, EPI_ISL_1385342, EPI_ISL_1385343, EPI_ISL_1385344, EPI_ISL_1385345, EPI_ISL_1385346, EPI_ISL_1385347, EPI_ISL_1385348, EPI_ISL_1385349, EPI_ISL_1385350, EPI_ISL_1385351, EPI_ISL_1385352, EPI_ISL_1385353, EPI_ISL_1385354, EPI_ISL_1385355, EPI_ISL_1385356, EPI_ISL_1385357, EPI_ISL_1385358, EPI_ISL_1385359, EPI_ISL_1385360, EPI_ISL_1385361, EPI_ISL_1385362, EPI_ISL_1385363, EPI_ISL_1385364, EPI_ISL_1385365, EPI_ISL_1385366, EPI_ISL_1385367, EPI_ISL_1385368, EPI_ISL_1385369, EPI_ISL_1385370, EPI_ISL_1385371, EPI_ISL_1385372, EPI_ISL_1385373, EPI_ISL_1385374, EPI_ISL_1385375, EPI_ISL_1385376, EPI_ISL_1385377, EPI_ISL_1385378, EPI_ISL_1385379, EPI_ISL_1385380, EPI_ISL_1385381, EPI_ISL_1385382, EPI_ISL_1385383, EPI_ISL_1385384, EPI_ISL_1385385, EPI_ISL_1385386, EPI_ISL_1385387, EPI_ISL_1385388, EPI_ISL_1385389, EPI_ISL_1385390, EPI_ISL_1385391, EPI_ISL_1385392, EPI_ISL_1385393, EPI_ISL_1385394, EPI_ISL_1385395, EPI_ISL_1385396, EPI_ISL_1385397, EPI_ISL_1385398, EPI_ISL_1385399, EPI_ISL_1385400, EPI_ISL_1385401, EPI_ISL_1385402, EPI_ISL_1385403, EPI_ISL_1385404, EPI_ISL_1385405, EPI_ISL_1385406, EPI_ISL_1385407, EPI_ISL_1385408, EPI_ISL_1385409, EPI_ISL_1385410, EPI_ISL_1385411, EPI_ISL_1385412, EPI_ISL_1385413, EPI_ISL_1385414, EPI_ISL_1385415, EPI_ISL_1385416, EPI_ISL_1385417, EPI_ISL_1385418, EPI_ISL_1385419, EPI_ISL_1385420, EPI_ISL_1385421, EPI_ISL_1385422, EPI_ISL_1385423, EPI_ISL_1385424, EPI_ISL_1385425, EPI_ISL_1385426, EPI_ISL_1385427, EPI_ISL_1385428, EPI_ISL_1385429, EPI_ISL_1385430, EPI_ISL_1385431, EPI_ISL_1385432, EPI_ISL_1385433, EPI_ISL_1385434, EPI_ISL_1385435, EPI_ISL_1385436, EPI_ISL_1385437, EPI_ISL_1385438, EPI_ISL_1385439, EPI_ISL_1385440, EPI_ISL_1385441, EPI_ISL_1385442, EPI_ISL_1385443, EPI_ISL_1385444, EPI_ISL_1385445, EPI_ISL_1385446, EPI_ISL_1385447, EPI_ISL_1385448, EPI_ISL_1385449, EPI_ISL_1385450, EPI_ISL_1385451, EPI_ISL_1385452, EPI_ISL_1385453, EPI_ISL_1385454, EPI_ISL_1385455, EPI_ISL_1385456, EPI_ISL_1385457, EPI_ISL_1385458, EPI_ISL_1385459, EPI_ISL_1385460, EPI_ISL_1385461, EPI_ISL_1385462, EPI_ISL_1385463, EPI_ISL_1385464, EPI_ISL_1385465, EPI_ISL_1385466, EPI_ISL_1385467, EPI_ISL_1385468, EPI_ISL_1385469, EPI_ISL_1385470, EPI_ISL_1385471, EPI_ISL_1385472, EPI_ISL_1385473, EPI_ISL_1385474, EPI_ISL_1385475, EPI_ISL_1385476, EPI_ISL_1385477, EPI_ISL_1385478, EPI_ISL_1385479, EPI_ISL_1385480, EPI_ISL_1385481, EPI_ISL_1385482, EPI_ISL_1385483, EPI_ISL_1385484, EPI_ISL_1385485, EPI_ISL_1385486, EPI_ISL_1385487, EPI_ISL_1385488, EPI_ISL_1385489, EPI_ISL_1385490, EPI_ISL_1385491, EPI_ISL_1385492, EPI_ISL_1385493, EPI_ISL_1385494, EPI_ISL_1385495, EPI_ISL_1385496, EPI_ISL_1385497, EPI_ISL_1385498, EPI_ISL_1385499, EPI_ISL_1385500, EPI_ISL_1385501, EPI_ISL_1385502, EPI_ISL_1385503, EPI_ISL_1385504, EPI_ISL_1385505, EPI_ISL_1385506, EPI_ISL_1385507, EPI_ISL_1385508, EPI_ISL_1385509, EPI_ISL_1385510, EPI_ISL_1385511, EPI_ISL_1385512, EPI_ISL_1385513, EPI_ISL_1385514, EPI_ISL_1385515, EPI_ISL_1385516, EPI_ISL_1385517, EPI_ISL_1385518, EPI_ISL_1385519, EPI_ISL_1385520, EPI_ISL_1385521, EPI_ISL_1385522, EPI_ISL_1385523, EPI_ISL_1385524, EPI_ISL_1385525, EPI_ISL_1385526, EPI_ISL_1385527, EPI_ISL_1385528, EPI_ISL_1385529, EPI_ISL_1385530, EPI_ISL_1385531, EPI_ISL_1385532, EPI_ISL_1385533, EPI_ISL_1385534, EPI_ISL_1385535, EPI_ISL_1385536, EPI_ISL_1385537, EPI_ISL_1385538, EPI_ISL_1385539, EPI_ISL_1385540, EPI_ISL_1385541, EPI_ISL_1385542, EPI_ISL_1385543, EPI_ISL_1385544, EPI_ISL_1385545, EPI_ISL_1385546, EPI_ISL_1385547, EPI_ISL_1385548, EPI_ISL_1385549, EPI_ISL_1385550, EPI_ISL_1385551, EPI_ISL_1385552, EPI_ISL_1385553, EPI_ISL_1385554, EPI_ISL_1385555, EPI_ISL_1385556, EPI_ISL_1385557, EPI_ISL_1385558, EPI_ISL_1385559, EPI_ISL_1385560, EPI_ISL_1385561, EPI_ISL_1385562, EPI_ISL_1385563, EPI_ISL_1385564, EPI_ISL_1385565, EPI_ISL_1385566, EPI_ISL_1385567, EPI_ISL_1385568, EPI_ISL_1385569, EPI_ISL_1385570, EPI_ISL_1385571, EPI_ISL_1385572, EPI_ISL_1385573, EPI_ISL_1385574, EPI_ISL_1385575, EPI_ISL_1385576, EPI_ISL_1385577, EPI_ISL_1385578, EPI_ISL_1385579, EPI_ISL_1385580, EPI_ISL_1385581, EPI_ISL_1385582, EPI_ISL_1385583, EPI_ISL_1385584, EPI_ISL_1385585, EPI_ISL_1385586, EPI_ISL_1385587, EPI_ISL_1385588, EPI_ISL_1385589, EPI_ISL_1385590, EPI_ISL_1385591, EPI_ISL_1385592, EPI_ISL_1385593, EPI_ISL_1385594, EPI_ISL_1385595, EPI_ISL_1385596, EPI_ISL_1385597, EPI_ISL_1385598, EPI_ISL_1385599, EPI_ISL_1385600, EPI_ISL_1385601, EPI_ISL_1385602, EPI_ISL_1385603, EPI_ISL_1385604, EPI_ISL_1385605, EPI_ISL_1385606, EPI_ISL_1385607, EPI_ISL_1385608, EPI_ISL_1385609, EPI_ISL_1385610, EPI_ISL_1385611, EPI_ISL_1385612, EPI_ISL_1385613, EPI_ISL_1385614, EPI_ISL_1385615, EPI_ISL_1385616, EPI_ISL_1385617, EPI_ISL_1385618, EPI_ISL_1385619, EPI_ISL_1385620, EPI_ISL_1385621, EPI_ISL_1385622, EPI_ISL_1385623, EPI_ISL_1385624, EPI_ISL_1385625, EPI_ISL_1385626, EPI_ISL_1385627, EPI_ISL_1385628, EPI_ISL_1385629, EPI_ISL_1385630, EPI_ISL_1385631, EPI_ISL_1385632, EPI_ISL_1385633, EPI_ISL_1385634, EPI_ISL_1385635, EPI_ISL_1385636, EPI_ISL_1385637, EPI_ISL_1385638, EPI_ISL_1385639, EPI_ISL_1385640, EPI_ISL_1385641, EPI_ISL_1385642, EPI_ISL_1385643, EPI_ISL_1385644, EPI_ISL_1385645, EPI_ISL_1385646, EPI_ISL_1385647, EPI_ISL_1385648, EPI_ISL_1385649, EPI_ISL_1385650, EPI_ISL_1385651, EPI_ISL_1385652, EPI_ISL_1385653, EPI_ISL_1385654, EPI_ISL_1385655, EPI_ISL_1385656, EPI_ISL_1385657, EPI_ISL_1385658, EPI_ISL_1385659, EPI_ISL_1385660, EPI_ISL_1385661, EPI_ISL_1385662, EPI_ISL_1385663, EPI_ISL_1385664, EPI_ISL_1385665, EPI_ISL_1385666, EPI_ISL_1385667, EPI_ISL_1385668, EPI_ISL_1385669, EPI_ISL_1385670, EPI_ISL_1385671, EPI_ISL_1385672, EPI_ISL_1385673, EPI_ISL_1385674, EPI_ISL_1385675, EPI_ISL_1385676, EPI_ISL_1385677, EPI_ISL_1385678, EPI_ISL_1385679, EPI_ISL_1385680, EPI_ISL_1385681, EPI_ISL_1385682, EPI_ISL_1385683, EPI_ISL_1385684, EPI_ISL_1385685, EPI_ISL_1385686, EPI_ISL_1385687, EPI_ISL_1385688, EPI_ISL_1385689, EPI_ISL_1385690, EPI_ISL_1385691, EPI_ISL_1385692, EPI_ISL_1385693, EPI_ISL_1385694, EPI_ISL_1385695, EPI_ISL_1385696, EPI_ISL_1385697, EPI_ISL_1385698, EPI_ISL_1385699, EPI_ISL_1385700, | | | |

| | | | |
|---|---|---|---|
| EPI_ISL_1385701, EPI_ISL_1385702, EPI_ISL_1385703, EPI_ISL_1385704, EPI_ISL_1385705, EPI_ISL_1385706, EPI_ISL_1385707, EPI_ISL_1385708, EPI_ISL_1385709, EPI_ISL_1385710, EPI_ISL_1385711, EPI_ISL_1385712, EPI_ISL_1385713, EPI_ISL_1385714, EPI_ISL_1385715, EPI_ISL_1385716, EPI_ISL_1385717, EPI_ISL_1385718, EPI_ISL_1385719, EPI_ISL_1385720, EPI_ISL_1385721, EPI_ISL_1385722, EPI_ISL_1385723, EPI_ISL_1385724, EPI_ISL_1385725, EPI_ISL_1385726, EPI_ISL_1385727, EPI_ISL_1385728, EPI_ISL_1385729, EPI_ISL_1385730, EPI_ISL_1385731, EPI_ISL_1385732, EPI_ISL_1385733, EPI_ISL_1385734, EPI_ISL_1385735, EPI_ISL_1385736, EPI_ISL_1385737, EPI_ISL_1385738, EPI_ISL_1385739, EPI_ISL_1385740, EPI_ISL_1385741, EPI_ISL_1385742, EPI_ISL_1385743, EPI_ISL_1385744, EPI_ISL_1385745, EPI_ISL_1385746, EPI_ISL_1385747, EPI_ISL_1385748, EPI_ISL_1385749, EPI_ISL_1385750, EPI_ISL_1385751, EPI_ISL_1385752, EPI_ISL_1385753, EPI_ISL_1385754, EPI_ISL_1385755, EPI_ISL_1385756, EPI_ISL_1385757, EPI_ISL_1385758, EPI_ISL_1385759, EPI_ISL_1385760, EPI_ISL_1385761, EPI_ISL_1385762, EPI_ISL_1385763, EPI_ISL_1385764, EPI_ISL_1385765, EPI_ISL_1385766, EPI_ISL_1385767, EPI_ISL_1385768, EPI_ISL_1385769, EPI_ISL_1385770, EPI_ISL_1385771, EPI_ISL_1385772, EPI_ISL_1385773, EPI_ISL_1385774, EPI_ISL_1385775, EPI_ISL_1385776, EPI_ISL_1385777, EPI_ISL_1385778, EPI_ISL_1385779, EPI_ISL_1385780, EPI_ISL_1385781, EPI_ISL_1385782, EPI_ISL_1385783 | | | |
| see above | Pandemic Response Lab - NYC | Pandemic Response Lab, R&D | Henry Lee, Michael Hammerling, Melissa Hopkins, Cybill del Castillo, Shinyoung Clair Kang, William Ward, Pradeep Bugga, Sol Rey, Dylan Law, Haiping Hao, Jon Laurent |
| EPI_ISL_1391780 | Helix/Illumina | Centers for Disease Control and Prevention Division of Viral Diseases, Pathogen Discovery | Peter W. Cook, Dakota Howard, Dhwani Batra, Ben L. Rambo-Martin, Eileen de Feo, Jan Antico, Christine Tran, Matthew Tolentino, Shannon Wickline, Kim Gietzen, Brad Sickler, Jingtao Liu, Eric Allen, Phil Febbo, Summer Galloway, Nicole L. Washington, Simon White, Geraint Levan, Kelly Schiabor Barrett, Elizabeth Cirulli, Alexandre Bolze, Ary Ascencio, Charlotte Rivera-Garcia, Ryan Cho, Jason Nguyen, Sherry Wang, Jimmy Ramirez, Tyler Cassens, Efren Sandoval, Magnus Isaksson, William Lee, David Becker, Marc Laurent, James Lu, Clinton R. Paden, Suxiang Tong, Duncan MacCannell |
| EPI_ISL_1394593, EPI_ISL_1394594, EPI_ISL_1394595, EPI_ISL_1394596, EPI_ISL_1394597, EPI_ISL_1394598, EPI_ISL_1394599, EPI_ISL_1394600, EPI_ISL_1394601, EPI_ISL_1394602, EPI_ISL_1394603, EPI_ISL_1394604, EPI_ISL_1394605, EPI_ISL_1394606, EPI_ISL_1394607, EPI_ISL_1394608, EPI_ISL_1394609, EPI_ISL_1394610, EPI_ISL_1394611, EPI_ISL_1394612, EPI_ISL_1394613, EPI_ISL_1394614, EPI_ISL_1394615, EPI_ISL_1394616, EPI_ISL_1394617, EPI_ISL_1394618, EPI_ISL_1394619, EPI_ISL_1394620, EPI_ISL_1394621, EPI_ISL_1394622, EPI_ISL_1394623, EPI_ISL_1394624, EPI_ISL_1394625, EPI_ISL_1394626, EPI_ISL_1394627, EPI_ISL_1394628 | | | |
| see above | WESTCHESTER MEDICAL CENTER | Wadsworth Center, New York State Department of Health | Kirsten St. George, Daryl M. Lamson, Alexis Russel, Matthew Shudt, Melissa A Leisner, Jonathan Plitnick, Navjot Singh, John Kelly, Erasmus Schneider, Erica Lasek-Nesselquist |
| EPI_ISL_1394629, EPI_ISL_1394630, EPI_ISL_1394631, EPI_ISL_1394632, EPI_ISL_1394633, EPI_ISL_1394634, EPI_ISL_1394635, EPI_ISL_1394636, EPI_ISL_1394637, EPI_ISL_1394638, EPI_ISL_1394639, EPI_ISL_1394640, EPI_ISL_1394641 | | | |
| see above | URMC LABS | Wadsworth Center, New York State Department of Health | Kirsten St. George, Daryl M. Lamson, Alexis Russel, Matthew Shudt, Melissa A Leisner, Jonathan Plitnick, Navjot Singh, John Kelly, Erasmus Schneider, Erica Lasek-Nesselquist |
| EPI_ISL_1394642 | New York Presbyterian Hospital | Wadsworth Center, New York State Department of Health | Kirsten St. George, Daryl M. Lamson, Alexis Russel, Matthew Shudt, Melissa A Leisner, Jonathan Plitnick, Navjot Singh, John Kelly, Erasmus Schneider, Erica Lasek-Nesselquist |
| EPI_ISL_1394643 | URMC LABS | Wadsworth Center, New York State Department of Health | Kirsten St. George, Daryl M. Lamson, Alexis Russel, Matthew Shudt, Melissa A Leisner, Jonathan Plitnick, Navjot Singh, John Kelly, Erasmus Schneider, Erica Lasek-Nesselquist |
| EPI_ISL_1394644, EPI_ISL_1394645, EPI_ISL_1394646, EPI_ISL_1394647, EPI_ISL_1394648, EPI_ISL_1394649, EPI_ISL_1394650, EPI_ISL_1394651, EPI_ISL_1394652, EPI_ISL_1394653, EPI_ISL_1394654, EPI_ISL_1394655, EPI_ISL_1394656, EPI_ISL_1394657, EPI_ISL_1394658, EPI_ISL_1394659, EPI_ISL_1394660, EPI_ISL_1394661, EPI_ISL_1394662, EPI_ISL_1394663, EPI_ISL_1394664 | | | |
| see above | New York Presbyterian Hospital | Wadsworth Center, New York State Department of Health | Kirsten St. George, Daryl M. Lamson, Alexis Russel, Matthew Shudt, Melissa A Leisner, Jonathan Plitnick, Navjot Singh, John Kelly, Erasmus Schneider, Erica Lasek-Nesselquist |
| EPI_ISL_1395706 | NYC Pandemic Response Lab | Wadsworth Center, New York State Department of Health | Kirsten St. George, Daryl M. Lamson, Alexis Russel, Matthew Shudt, Melissa A Leisner, Jonathan Plitnick, Navjot Singh, John Kelly, Erasmus Schneider, Erica Lasek-Nesselquist |
| EPI_ISL_1395707 | NORTHWELL HEALTH LABORATORIES | Wadsworth Center, New York State Department of Health | Kirsten St. George, Daryl M. Lamson, Alexis Russel, Matthew Shudt, Melissa A Leisner, Jonathan Plitnick, Navjot Singh, John Kelly, Erasmus Schneider, Erica Lasek-Nesselquist |
| EPI_ISL_1395708, EPI_ISL_1395709 | NYC Pandemic Response Lab | Wadsworth Center, New York State Department of Health | Kirsten St. George, Daryl M. Lamson, Alexis Russel, Matthew Shudt, Melissa A Leisner, Jonathan Plitnick, Navjot Singh, John Kelly, Erasmus Schneider, Erica Lasek-Nesselquist |
| EPI_ISL_1395710, EPI_ISL_1395711, EPI_ISL_1395712, EPI_ISL_1395713, EPI_ISL_1395714, EPI_ISL_1395715, EPI_ISL_1395716, EPI_ISL_1395717, EPI_ISL_1395718, EPI_ISL_1395719, EPI_ISL_1395720, EPI_ISL_1395721, EPI_ISL_1395722, EPI_ISL_1395723, EPI_ISL_1395724, EPI_ISL_1395725 | | | |
| see above | MEMORIAL SLOAN KETTERING CANCER CENTER | Wadsworth Center, New York State Department of Health | Kirsten St. George, Daryl M. Lamson, Alexis Russel, Matthew Shudt, Melissa A Leisner, Jonathan Plitnick, Navjot Singh, John Kelly, Erasmus Schneider, Erica Lasek-Nesselquist |
| EPI_ISL_1395726, EPI_ISL_1395727, EPI_ISL_1395728, EPI_ISL_1395729, EPI_ISL_1395730, EPI_ISL_1395731, EPI_ISL_1395732, EPI_ISL_1395733, EPI_ISL_1395734, EPI_ISL_1395735, EPI_ISL_1395736, EPI_ISL_1395737, EPI_ISL_1395738, EPI_ISL_1395739, EPI_ISL_1395740, EPI_ISL_1395741, EPI_ISL_1395742, EPI_ISL_1395743, EPI_ISL_1395744, EPI_ISL_1395745, EPI_ISL_1395746, EPI_ISL_1395747, EPI_ISL_1395748, EPI_ISL_1395749 | | | |
| see above | Columbia University Irving Medical Center | Wadsworth Center, New York State Department of Health | Kirsten St. George, Daryl M. Lamson, Alexis Russel, Matthew Shudt, Melissa A Leisner, Jonathan Plitnick, Navjot Singh, John Kelly, Erasmus Schneider, Erica Lasek-Nesselquist |
| EPI_ISL_1395750 | NORTH SHORE UNIVERSITY HOSPITAL | Wadsworth Center, New York State Department of Health | Kirsten St. George, Daryl M. Lamson, Alexis Russel, Matthew Shudt, Melissa A Leisner, Jonathan Plitnick, Navjot Singh, John Kelly, Erasmus Schneider, Erica Lasek-Nesselquist |
| EPI_ISL_1395751, EPI_ISL_1395752 | SUNY UPSTATE MEDICAL UNIVERSITY | Wadsworth Center, New York State Department of Health | Kirsten St. George, Daryl M. Lamson, Alexis Russel, Matthew Shudt, Melissa A Leisner, Jonathan Plitnick, Navjot Singh, John Kelly, Erasmus Schneider, Erica Lasek-Nesselquist |
| EPI_ISL_1395753, EPI_ISL_1395754 | NORTH SHORE UNIVERSITY HOSPITAL | Wadsworth Center, New York State Department of Health | Kirsten St. George, Daryl M. Lamson, Alexis Russel, Matthew Shudt, Melissa A Leisner, Jonathan Plitnick, Navjot Singh, John Kelly, Erasmus Schneider, Erica Lasek-Nesselquist |
| EPI_ISL_1395755 | SUNY UPSTATE MEDICAL UNIVERSITY | Wadsworth Center, New York State Department of Health | Kirsten St. George, Daryl M. Lamson, Alexis Russel, Matthew Shudt, Melissa A Leisner, Jonathan Plitnick, Navjot Singh, John Kelly, Erasmus Schneider, Erica Lasek-Nesselquist |
| EPI_ISL_1395756, EPI_ISL_1395757, EPI_ISL_1395758 | NORTH SHORE UNIVERSITY HOSPITAL | Wadsworth Center, New York State Department of Health | Kirsten St. George, Daryl M. Lamson, Alexis Russel, Matthew Shudt, Melissa A Leisner, Jonathan Plitnick, Navjot Singh, John Kelly, Erasmus Schneider, Erica Lasek-Nesselquist |
| EPI_ISL_1395759 | SUNY UPSTATE MEDICAL UNIVERSITY | Wadsworth Center, New York State Department of Health | Kirsten St. George, Daryl M. Lamson, Alexis Russel, Matthew Shudt, Melissa A Leisner, Jonathan Plitnick, Navjot Singh, John Kelly, Erasmus Schneider, Erica Lasek-Nesselquist |
| EPI_ISL_1395760 | NORTH SHORE UNIVERSITY HOSPITAL | Wadsworth Center, New York State Department of Health | Kirsten St. George, Daryl M. Lamson, Alexis Russel, Matthew Shudt, Melissa A Leisner, Jonathan Plitnick, Navjot Singh, John Kelly, Erasmus Schneider, Erica Lasek-Nesselquist |
| EPI_ISL_1395761, EPI_ISL_1395762 | SUNY UPSTATE MEDICAL UNIVERSITY | Wadsworth Center, New York State Department of Health | Kirsten St. George, Daryl M. Lamson, Alexis Russel, Matthew Shudt, Melissa A Leisner, Jonathan Plitnick, Navjot Singh, John Kelly, Erasmus Schneider, Erica Lasek-Nesselquist |
| EPI_ISL_1395763 | NORTH SHORE UNIVERSITY HOSPITAL | Wadsworth Center, New York State Department of Health | Kirsten St. George, Daryl M. Lamson, Alexis Russel, Matthew Shudt, Melissa A Leisner, Jonathan Plitnick, Navjot Singh, John Kelly, Erasmus Schneider, Erica Lasek-Nesselquist |
| EPI_ISL_1395764, EPI_ISL_1395765 | SUNY UPSTATE MEDICAL UNIVERSITY | Wadsworth Center, New York State Department of Health | Kirsten St. George, Daryl M. Lamson, Alexis Russel, Matthew Shudt, Melissa A Leisner, Jonathan Plitnick, Navjot Singh, John Kelly, Erasmus Schneider, Erica Lasek-Nesselquist |
| EPI_ISL_1395766 | THE MARY IMOGENE BASSETT HOSPITAL | Wadsworth Center, New York State Department of Health | Kirsten St. George, Daryl M. Lamson, Alexis Russel, Matthew Shudt, Melissa A Leisner, Jonathan Plitnick, Navjot Singh, John Kelly, Erasmus Schneider, Erica Lasek-Nesselquist |
| EPI_ISL_1395767 | SUNY UPSTATE MEDICAL UNIVERSITY | Wadsworth Center, New York State Department of Health | Kirsten St. George, Daryl M. Lamson, Alexis Russel, Matthew Shudt, Melissa A Leisner, Jonathan Plitnick, Navjot Singh, John Kelly, Erasmus Schneider, Erica Lasek-Nesselquist |
| EPI_ISL_1395768, EPI_ISL_1395769 | THE MARY IMOGENE BASSETT HOSPITAL | Wadsworth Center, New York State Department of Health | Kirsten St. George, Daryl M. Lamson, Alexis Russel, Matthew Shudt, Melissa A Leisner, Jonathan Plitnick, Navjot Singh, John Kelly, Erasmus Schneider, Erica Lasek-Nesselquist |
| EPI_ISL_1395770 | SUNY UPSTATE MEDICAL UNIVERSITY | Wadsworth Center, New York State Department of Health | Kirsten St. George, Daryl M. Lamson, Alexis Russel, Matthew Shudt, Melissa A Leisner, Jonathan Plitnick, Navjot Singh, John Kelly, Erasmus Schneider, Erica Lasek-Nesselquist |
| EPI_ISL_1395771 | THE MARY IMOGENE BASSETT HOSPITAL | Wadsworth Center, New York State Department of Health | Kirsten St. George, Daryl M. Lamson, Alexis Russel, Matthew Shudt, Melissa A Leisner, Jonathan Plitnick, Navjot Singh, John Kelly, Erasmus Schneider, Erica Lasek-Nesselquist |
| EPI_ISL_1395772 | SUNY UPSTATE MEDICAL UNIVERSITY | Wadsworth Center, New York State Department of Health | Kirsten St. George, Daryl M. Lamson, Alexis Russel, Matthew Shudt, Melissa A Leisner, Jonathan Plitnick, Navjot Singh, John Kelly, Erasmus Schneider, |

| | | | |
|---|---|---|---|
| | | | Erica Lasek-Nesselquist |
| EPI_ISL_1395773 | THE MARY IMOGENE BASSETT HOSPITAL | Wadsworth Center, New York State Department of Health | Kirsten St. George, Daryl M. Lamson, Alexis Russel, Matthew Shudt, Melissa A Leisner, Jonathan Plitnick, Navjot Singh, John Kelly, Erasmus Schneider, Erica Lasek-Nesselquist |
| EPI_ISL_1395774, EPI_ISL_1395775, EPI_ISL_1395776 | SUNY UPSTATE MEDICAL UNIVERSITY | Wadsworth Center, New York State Department of Health | Kirsten St. George, Daryl M. Lamson, Alexis Russel, Matthew Shudt, Melissa A Leisner, Jonathan Plitnick, Navjot Singh, John Kelly, Erasmus Schneider, Erica Lasek-Nesselquist |
| EPI_ISL_1397383, EPI_ISL_1397384, EPI_ISL_1397385, EPI_ISL_1397386, EPI_ISL_1397387, EPI_ISL_1397388 | MEMORIAL SLOAN KETTERING CANCER CENTER | Wadsworth Center, New York State Department of Health | Kirsten St. George, Daryl M. Lamson, Alexis Russel, Matthew Shudt, Melissa A Leisner, Jonathan Plitnick, Navjot Singh, John Kelly, Erasmus Schneider, Erica Lasek-Nesselquist |
| EPI_ISL_1397389, EPI_ISL_1397390, EPI_ISL_1397391, EPI_ISL_1397392 | SUNY UPSTATE MEDICAL UNIVERSITY | Wadsworth Center, New York State Department of Health | Kirsten St. George, Daryl M. Lamson, Alexis Russel, Matthew Shudt, Melissa A Leisner, Jonathan Plitnick, Navjot Singh, John Kelly, Erasmus Schneider, Erica Lasek-Nesselquist |
| EPI_ISL_1397393 | NORTH SHORE UNIVERSITY HOSPITAL | Wadsworth Center, New York State Department of Health | Kirsten St. George, Daryl M. Lamson, Alexis Russel, Matthew Shudt, Melissa A Leisner, Jonathan Plitnick, Navjot Singh, John Kelly, Erasmus Schneider, Erica Lasek-Nesselquist |
| EPI_ISL_1397394 | SUNY UPSTATE MEDICAL UNIVERSITY | Wadsworth Center, New York State Department of Health | Kirsten St. George, Daryl M. Lamson, Alexis Russel, Matthew Shudt, Melissa A Leisner, Jonathan Plitnick, Navjot Singh, John Kelly, Erasmus Schneider, Erica Lasek-Nesselquist |
| EPI_ISL_1397395 | NORTH SHORE UNIVERSITY HOSPITAL | Wadsworth Center, New York State Department of Health | Kirsten St. George, Daryl M. Lamson, Alexis Russel, Matthew Shudt, Melissa A Leisner, Jonathan Plitnick, Navjot Singh, John Kelly, Erasmus Schneider, Erica Lasek-Nesselquist |
| EPI_ISL_1397396, EPI_ISL_1397397, EPI_ISL_1397398, EPI_ISL_1397399, EPI_ISL_1397400, EPI_ISL_1397401, EPI_ISL_1397402 | SUNY UPSTATE MEDICAL UNIVERSITY | Wadsworth Center, New York State Department of Health | Kirsten St. George, Daryl M. Lamson, Alexis Russel, Matthew Shudt, Melissa A Leisner, Jonathan Plitnick, Navjot Singh, John Kelly, Erasmus Schneider, Erica Lasek-Nesselquist |
| EPI_ISL_1397403 | NORTH SHORE UNIVERSITY HOSPITAL | Wadsworth Center, New York State Department of Health | Kirsten St. George, Daryl M. Lamson, Alexis Russel, Matthew Shudt, Melissa A Leisner, Jonathan Plitnick, Navjot Singh, John Kelly, Erasmus Schneider, Erica Lasek-Nesselquist |
| EPI_ISL_1397404 | SUNY UPSTATE MEDICAL UNIVERSITY | Wadsworth Center, New York State Department of Health | Kirsten St. George, Daryl M. Lamson, Alexis Russel, Matthew Shudt, Melissa A Leisner, Jonathan Plitnick, Navjot Singh, John Kelly, Erasmus Schneider, Erica Lasek-Nesselquist |
| EPI_ISL_1397405 | NORTH SHORE UNIVERSITY HOSPITAL | Wadsworth Center, New York State Department of Health | Kirsten St. George, Daryl M. Lamson, Alexis Russel, Matthew Shudt, Melissa A Leisner, Jonathan Plitnick, Navjot Singh, John Kelly, Erasmus Schneider, Erica Lasek-Nesselquist |
| EPI_ISL_1397406 | SUNY UPSTATE MEDICAL UNIVERSITY | Wadsworth Center, New York State Department of Health | Kirsten St. George, Daryl M. Lamson, Alexis Russel, Matthew Shudt, Melissa A Leisner, Jonathan Plitnick, Navjot Singh, John Kelly, Erasmus Schneider, Erica Lasek-Nesselquist |
| EPI_ISL_1397407 | NORTH SHORE UNIVERSITY HOSPITAL | Wadsworth Center, New York State Department of Health | Kirsten St. George, Daryl M. Lamson, Alexis Russel, Matthew Shudt, Melissa A Leisner, Jonathan Plitnick, Navjot Singh, John Kelly, Erasmus Schneider, Erica Lasek-Nesselquist |
| EPI_ISL_1397408 | SUNY UPSTATE MEDICAL UNIVERSITY | Wadsworth Center, New York State Department of Health | Kirsten St. George, Daryl M. Lamson, Alexis Russel, Matthew Shudt, Melissa A Leisner, Jonathan Plitnick, Navjot Singh, John Kelly, Erasmus Schneider, Erica Lasek-Nesselquist |
| EPI_ISL_1397409 | NORTH SHORE UNIVERSITY HOSPITAL | Wadsworth Center, New York State Department of Health | Kirsten St. George, Daryl M. Lamson, Alexis Russel, Matthew Shudt, Melissa A Leisner, Jonathan Plitnick, Navjot Singh, John Kelly, Erasmus Schneider, Erica Lasek-Nesselquist |
| EPI_ISL_1397410, EPI_ISL_1397411, EPI_ISL_1397412, EPI_ISL_1397413, EPI_ISL_1397414, EPI_ISL_1397415, EPI_ISL_1397416, EPI_ISL_1397417, EPI_ISL_1397418, EPI_ISL_1397419, EPI_ISL_1397420, EPI_ISL_1397421, EPI_ISL_1397422, EPI_ISL_1397423 | | | |
| see above | SUNY UPSTATE MEDICAL UNIVERSITY | Wadsworth Center, New York State Department of Health | Kirsten St. George, Daryl M. Lamson, Alexis Russel, Matthew Shudt, Melissa A Leisner, Jonathan Plitnick, Navjot Singh, John Kelly, Erasmus Schneider, Erica Lasek-Nesselquist |
| EPI_ISL_1397424, EPI_ISL_1397425, EPI_ISL_1397426, EPI_ISL_1397427, EPI_ISL_1397428, EPI_ISL_1397429, EPI_ISL_1397430, EPI_ISL_1397431, EPI_ISL_1397432, EPI_ISL_1397433, EPI_ISL_1397434, EPI_ISL_1397435, EPI_ISL_1397436, EPI_ISL_1397437, EPI_ISL_1397438, EPI_ISL_1397439, EPI_ISL_1397440, EPI_ISL_1397441, EPI_ISL_1397442, EPI_ISL_1397443 | | | |
| see above | ADIRONDACK MEDICAL CENTER | Wadsworth Center, New York State Department of Health | Kirsten St. George, Daryl M. Lamson, Alexis Russel, Matthew Shudt, Melissa A Leisner, Jonathan Plitnick, Navjot Singh, John Kelly, Erasmus Schneider, Erica Lasek-Nesselquist |
| EPI_ISL_1397444, EPI_ISL_1397445, EPI_ISL_1397446, EPI_ISL_1397447, EPI_ISL_1397448, EPI_ISL_1397449, EPI_ISL_1397450, EPI_ISL_1397451, EPI_ISL_1397452, EPI_ISL_1397453, EPI_ISL_1397454, EPI_ISL_1397455, EPI_ISL_1397456, EPI_ISL_1397457, EPI_ISL_1397458 | | | |
| see above | WESTCHESTER MEDICAL CENTER | Wadsworth Center, New York State Department of Health | Kirsten St. George, Daryl M. Lamson, Alexis Russel, Matthew Shudt, Melissa A Leisner, Jonathan Plitnick, Navjot Singh, John Kelly, Erasmus Schneider, Erica Lasek-Nesselquist |
| EPI_ISL_1397459 | SUNY UPSTATE MEDICAL UNIVERSITY | Wadsworth Center, New York State Department of Health | Kirsten St. George, Daryl M. Lamson, Alexis Russel, Matthew Shudt, Melissa A Leisner, Jonathan Plitnick, Navjot Singh, John Kelly, Erasmus Schneider, Erica Lasek-Nesselquist |
| EPI_ISL_1397460, EPI_ISL_1397461, EPI_ISL_1397462, EPI_ISL_1397463, EPI_ISL_1397464, EPI_ISL_1397465, EPI_ISL_1397466, EPI_ISL_1397467, EPI_ISL_1397468, EPI_ISL_1397469, EPI_ISL_1397470, EPI_ISL_1397471, EPI_ISL_1397472, EPI_ISL_1397473, EPI_ISL_1397474, EPI_ISL_1397475, EPI_ISL_1397476, EPI_ISL_1397477, EPI_ISL_1397478, EPI_ISL_1397479, EPI_ISL_1397480, EPI_ISL_1397481, EPI_ISL_1397482, EPI_ISL_1397483, EPI_ISL_1397484, EPI_ISL_1397485, EPI_ISL_1397486, EPI_ISL_1397487, EPI_ISL_1397488, EPI_ISL_1397489, EPI_ISL_1397490, EPI_ISL_1397491, EPI_ISL_1397492, EPI_ISL_1397493, EPI_ISL_1397494, EPI_ISL_1397495, EPI_ISL_1397496, EPI_ISL_1397497, EPI_ISL_1397498, EPI_ISL_1397499, EPI_ISL_1397500, EPI_ISL_1397501, EPI_ISL_1397502, EPI_ISL_1397503, EPI_ISL_1397504, EPI_ISL_1397505, EPI_ISL_1397506, EPI_ISL_1397507, EPI_ISL_1397508, EPI_ISL_1397509, EPI_ISL_1397510, EPI_ISL_1397511, EPI_ISL_1397512, EPI_ISL_1397513, EPI_ISL_1397514, EPI_ISL_1397515, EPI_ISL_1397516, EPI_ISL_1397517, EPI_ISL_1397518, EPI_ISL_1397519, EPI_ISL_1397520, EPI_ISL_1397521, EPI_ISL_1397522, EPI_ISL_1397523, EPI_ISL_1397524, EPI_ISL_1397525, EPI_ISL_1397526, EPI_ISL_1397527, EPI_ISL_1397528, EPI_ISL_1397529, EPI_ISL_1397530, EPI_ISL_1397531, EPI_ISL_1397532, EPI_ISL_1397533, EPI_ISL_1397534, EPI_ISL_1397535, EPI_ISL_1397536, EPI_ISL_1397537, EPI_ISL_1397538, EPI_ISL_1397539, EPI_ISL_1397540, EPI_ISL_1397541 | | | |
| see above | New York Presbyterian Hospital | Wadsworth Center, New York State Department of Health | Kirsten St. George, Daryl M. Lamson, Alexis Russel, Matthew Shudt, Melissa A Leisner, Jonathan Plitnick, Navjot Singh, John Kelly, Erasmus Schneider, Erica Lasek-Nesselquist |
| EPI_ISL_1397542, EPI_ISL_1397543, EPI_ISL_1397544, EPI_ISL_1397545 | ALBANY MEDICAL CENTER | Wadsworth Center, New York State Department of Health | Kirsten St. George, Daryl M. Lamson, Alexis Russel, Matthew Shudt, Melissa A Leisner, Jonathan Plitnick, Navjot Singh, John Kelly, Erasmus Schneider, Erica Lasek-Nesselquist |
| EPI_ISL_1397546, EPI_ISL_1397547, EPI_ISL_1397548, EPI_ISL_1397549 | GLENS FALLS HOSPITAL LABORATORY | Wadsworth Center, New York State Department of Health | Kirsten St. George, Daryl M. Lamson, Alexis Russel, Matthew Shudt, Melissa A Leisner, Jonathan Plitnick, Navjot Singh, John Kelly, Erasmus Schneider, Erica Lasek-Nesselquist |
| EPI_ISL_1397550 | WESTCHESTER MEDICAL CENTER | Wadsworth Center, New York State Department of Health | Kirsten St. George, Daryl M. Lamson, Alexis Russel, Matthew Shudt, Melissa A Leisner, Jonathan Plitnick, Navjot Singh, John Kelly, Erasmus Schneider, Erica Lasek-Nesselquist |
| EPI_ISL_1397551 | ALBANY MEDICAL CENTER | Wadsworth Center, New York State Department of Health | Kirsten St. George, Daryl M. Lamson, Alexis Russel, Matthew Shudt, Melissa A Leisner, Jonathan Plitnick, Navjot Singh, John Kelly, Erasmus Schneider, Erica Lasek-Nesselquist |
| EPI_ISL_1397552 | URMC LABS | Wadsworth Center, New York State Department of Health | Kirsten St. George, Daryl M. Lamson, Alexis Russel, Matthew Shudt, Melissa A Leisner, Jonathan Plitnick, Navjot Singh, John Kelly, Erasmus Schneider, Erica Lasek-Nesselquist |
| EPI_ISL_1397553, EPI_ISL_1397554 | TEMPUS LABS INC | Wadsworth Center, New York State Department of Health | Kirsten St. George, Daryl M. Lamson, Alexis Russel, Matthew Shudt, Melissa A Leisner, Jonathan Plitnick, Navjot Singh, John Kelly, Erasmus Schneider, Erica Lasek-Nesselquist |
| EPI_ISL_1397555, EPI_ISL_1397556, EPI_ISL_1397557, EPI_ISL_1397558, EPI_ISL_1397559, EPI_ISL_1397560, EPI_ISL_1397561, EPI_ISL_1397562, EPI_ISL_1397563, EPI_ISL_1397564, EPI_ISL_1397565, EPI_ISL_1397566, EPI_ISL_1397567, EPI_ISL_1397568, EPI_ISL_1397569, EPI_ISL_1397570, EPI_ISL_1397571, EPI_ISL_1397572, EPI_ISL_1397573, EPI_ISL_1397574, EPI_ISL_1397575, EPI_ISL_1397576, EPI_ISL_1397577, EPI_ISL_1397578, EPI_ISL_1397579, EPI_ISL_1397580 | | | |
| see above | URMC LABS | Wadsworth Center, New York State Department of Health | Kirsten St. George, Daryl M. Lamson, Alexis Russel, Matthew Shudt, Melissa A Leisner, Jonathan Plitnick, Navjot Singh, John Kelly, Erasmus Schneider, Erica Lasek-Nesselquist |
| EPI_ISL_1397581, EPI_ISL_1397582, EPI_ISL_1397583, EPI_ISL_1397584, EPI_ISL_1397585, EPI_ISL_1397586, EPI_ISL_1397587, EPI_ISL_1397588, EPI_ISL_1397589, EPI_ISL_1397590, EPI_ISL_1397591 | | | |
| see above | TEMPUS LABS INC | Wadsworth Center, New York State Department of Health | Kirsten St. George, Daryl M. Lamson, Alexis Russel, Matthew Shudt, Melissa A Leisner, Jonathan Plitnick, Navjot Singh, John Kelly, Erasmus Schneider, |

| | | | |
|---|---|---|---|
| | | | Erica Lasek-Nesselquist |
| EPI_ISL_1397592, EPI_ISL_1397593, EPI_ISL_1397594, EPI_ISL_1397595, EPI_ISL_1397596, EPI_ISL_1397597, EPI_ISL_1397598, EPI_ISL_1397599, EPI_ISL_1397600, EPI_ISL_1397601, EPI_ISL_1397602, EPI_ISL_1397603, EPI_ISL_1397604, EPI_ISL_1397605, EPI_ISL_1397606, EPI_ISL_1397607, EPI_ISL_1397608, EPI_ISL_1397609, EPI_ISL_1397610, EPI_ISL_1397611 | | | |
| see above | SUNY UPSTATE MEDICAL UNIVERSITY | Wadsworth Center, New York State Department of Health | Kirsten St. George, Daryl M. Lamson, Alexis Russel, Matthew Shudt, Melissa A Leisner, Jonathan Plitnick, Navjot Singh, John Kelly, Erasmus Schneider, Erica Lasek-Nesselquist |
| EPI_ISL_1397612, EPI_ISL_1397613, EPI_ISL_1397614, EPI_ISL_1397615, EPI_ISL_1397616, EPI_ISL_1397617 | THE MARY IMOGENE BASSETT HOSPITAL | Wadsworth Center, New York State Department of Health | Kirsten St. George, Daryl M. Lamson, Alexis Russel, Matthew Shudt, Melissa A Leisner, Jonathan Plitnick, Navjot Singh, John Kelly, Erasmus Schneider, Erica Lasek-Nesselquist |
| EPI_ISL_1397618, EPI_ISL_1397619, EPI_ISL_1397620, EPI_ISL_1397621, EPI_ISL_1397622 | MONTEFIORE MEDICAL CENTER LABORATORIES | Wadsworth Center, New York State Department of Health | Kirsten St. George, Daryl M. Lamson, Alexis Russel, Matthew Shudt, Melissa A Leisner, Jonathan Plitnick, Navjot Singh, John Kelly, Erasmus Schneider, Erica Lasek-Nesselquist |
| EPI_ISL_1397623, EPI_ISL_1397624, EPI_ISL_1397625, EPI_ISL_1397626, EPI_ISL_1397627, EPI_ISL_1397628 | NYC Pandemic Response Lab | Wadsworth Center, New York State Department of Health | Kirsten St. George, Daryl M. Lamson, Alexis Russel, Matthew Shudt, Melissa A Leisner, Jonathan Plitnick, Navjot Singh, John Kelly, Erasmus Schneider, Erica Lasek-Nesselquist |
| EPI_ISL_1397629 | SUNY UPSTATE MEDICAL UNIVERSITY | Wadsworth Center, New York State Department of Health | Kirsten St. George, Daryl M. Lamson, Alexis Russel, Matthew Shudt, Melissa A Leisner, Jonathan Plitnick, Navjot Singh, John Kelly, Erasmus Schneider, Erica Lasek-Nesselquist |
| EPI_ISL_1397630, EPI_ISL_1397631, EPI_ISL_1397632, EPI_ISL_1397633, EPI_ISL_1397634, EPI_ISL_1397635 | URMC LABS | Wadsworth Center, New York State Department of Health | Kirsten St. George, Daryl M. Lamson, Alexis Russel, Matthew Shudt, Melissa A Leisner, Jonathan Plitnick, Navjot Singh, John Kelly, Erasmus Schneider, Erica Lasek-Nesselquist |
| EPI_ISL_1397636, EPI_ISL_1397637, EPI_ISL_1397638, EPI_ISL_1397639, EPI_ISL_1397640, EPI_ISL_1397641, EPI_ISL_1397642, EPI_ISL_1397643, EPI_ISL_1397644, EPI_ISL_1397645, EPI_ISL_1397646, EPI_ISL_1397647, EPI_ISL_1397648, EPI_ISL_1397649 | | | |
| see above | Columbia University Irving Medical Center | Wadsworth Center, New York State Department of Health | Kirsten St. George, Daryl M. Lamson, Alexis Russel, Matthew Shudt, Melissa A Leisner, Jonathan Plitnick, Navjot Singh, John Kelly, Erasmus Schneider, Erica Lasek-Nesselquist |
| EPI_ISL_1397650, EPI_ISL_1397651, EPI_ISL_1397652, EPI_ISL_1397653, EPI_ISL_1397654, EPI_ISL_1397655, EPI_ISL_1397656, EPI_ISL_1397657, EPI_ISL_1397658, EPI_ISL_1397659, EPI_ISL_1397660, EPI_ISL_1397661, EPI_ISL_1397662, EPI_ISL_1397663, EPI_ISL_1397664, EPI_ISL_1397665, EPI_ISL_1397666, EPI_ISL_1397667, EPI_ISL_1397668, EPI_ISL_1397669, EPI_ISL_1397670, EPI_ISL_1397671, EPI_ISL_1397672, EPI_ISL_1397673, EPI_ISL_1397674, EPI_ISL_1397675, EPI_ISL_1397676, EPI_ISL_1397677, EPI_ISL_1397678, EPI_ISL_1397679, EPI_ISL_1397680, EPI_ISL_1397681, EPI_ISL_1397682, EPI_ISL_1397683, EPI_ISL_1397684, EPI_ISL_1397685, EPI_ISL_1397686, EPI_ISL_1397687, EPI_ISL_1397688, EPI_ISL_1397689, EPI_ISL_1397690, EPI_ISL_1397691, EPI_ISL_1397692, EPI_ISL_1397693 | | | |
| see above | NORTH SHORE UNIVERSITY HOSPITAL | Wadsworth Center, New York State Department of Health | Kirsten St. George, Daryl M. Lamson, Alexis Russel, Matthew Shudt, Melissa A Leisner, Jonathan Plitnick, Navjot Singh, John Kelly, Erasmus Schneider, Erica Lasek-Nesselquist |
| EPI_ISL_1397694, EPI_ISL_1397695, EPI_ISL_1397696, EPI_ISL_1397697, EPI_ISL_1397698, EPI_ISL_1397699 | BIO-REFERENCE LABORATORIES | Wadsworth Center, New York State Department of Health | Kirsten St. George, Daryl M. Lamson, Alexis Russel, Matthew Shudt, Melissa A Leisner, Jonathan Plitnick, Navjot Singh, John Kelly, Erasmus Schneider, Erica Lasek-Nesselquist |
| EPI_ISL_1397700, EPI_ISL_1397701, EPI_ISL_1397702, EPI_ISL_1397703, EPI_ISL_1397704, EPI_ISL_1397705 | TEMPUS LABS INC | Wadsworth Center, New York State Department of Health | Kirsten St. George, Daryl M. Lamson, Alexis Russel, Matthew Shudt, Melissa A Leisner, Jonathan Plitnick, Navjot Singh, John Kelly, Erasmus Schneider, Erica Lasek-Nesselquist |
| EPI_ISL_1397706 | SUNY UPSTATE MEDICAL UNIVERSITY | Wadsworth Center, New York State Department of Health | Kirsten St. George, Daryl M. Lamson, Alexis Russel, Matthew Shudt, Melissa A Leisner, Jonathan Plitnick, Navjot Singh, John Kelly, Erasmus Schneider, Erica Lasek-Nesselquist |
| EPI_ISL_1397707, EPI_ISL_1397708, EPI_ISL_1397709, EPI_ISL_1397710 | ALBANY MEDICAL CENTER | Wadsworth Center, New York State Department of Health | Kirsten St. George, Daryl M. Lamson, Alexis Russel, Matthew Shudt, Melissa A Leisner, Jonathan Plitnick, Navjot Singh, John Kelly, Erasmus Schneider, Erica Lasek-Nesselquist |
| EPI_ISL_1397711 | ADIRONDACK MEDICAL CENTER | Wadsworth Center, New York State Department of Health | Kirsten St. George, Daryl M. Lamson, Alexis Russel, Matthew Shudt, Melissa A Leisner, Jonathan Plitnick, Navjot Singh, John Kelly, Erasmus Schneider, Erica Lasek-Nesselquist |
| EPI_ISL_1397712, EPI_ISL_1397713, EPI_ISL_1397714, EPI_ISL_1397715, EPI_ISL_1397716, EPI_ISL_1397717, EPI_ISL_1397718, EPI_ISL_1397719, EPI_ISL_1397720 | ALBANY MEDICAL CENTER | Wadsworth Center, New York State Department of Health | Kirsten St. George, Daryl M. Lamson, Alexis Russel, Matthew Shudt, Melissa A Leisner, Jonathan Plitnick, Navjot Singh, John Kelly, Erasmus Schneider, Erica Lasek-Nesselquist |
| EPI_ISL_1397721 | ADIRONDACK MEDICAL CENTER | Wadsworth Center, New York State Department of Health | Kirsten St. George, Daryl M. Lamson, Alexis Russel, Matthew Shudt, Melissa A Leisner, Jonathan Plitnick, Navjot Singh, John Kelly, Erasmus Schneider, Erica Lasek-Nesselquist |
| EPI_ISL_1397722 | ALBANY MEDICAL CENTER | Wadsworth Center, New York State Department of Health | Kirsten St. George, Daryl M. Lamson, Alexis Russel, Matthew Shudt, Melissa A Leisner, Jonathan Plitnick, Navjot Singh, John Kelly, Erasmus Schneider, Erica Lasek-Nesselquist |
| EPI_ISL_1397723 | ADIRONDACK MEDICAL CENTER | Wadsworth Center, New York State Department of Health | Kirsten St. George, Daryl M. Lamson, Alexis Russel, Matthew Shudt, Melissa A Leisner, Jonathan Plitnick, Navjot Singh, John Kelly, Erasmus Schneider, Erica Lasek-Nesselquist |
| EPI_ISL_1397724, EPI_ISL_1397725, EPI_ISL_1397726, EPI_ISL_1397727, EPI_ISL_1397728, EPI_ISL_1397729, EPI_ISL_1397730 | ALBANY MEDICAL CENTER | Wadsworth Center, New York State Department of Health | Kirsten St. George, Daryl M. Lamson, Alexis Russel, Matthew Shudt, Melissa A Leisner, Jonathan Plitnick, Navjot Singh, John Kelly, Erasmus Schneider, Erica Lasek-Nesselquist |
| EPI_ISL_1397731 | ADIRONDACK MEDICAL CENTER | Wadsworth Center, New York State Department of Health | Kirsten St. George, Daryl M. Lamson, Alexis Russel, Matthew Shudt, Melissa A Leisner, Jonathan Plitnick, Navjot Singh, John Kelly, Erasmus Schneider, Erica Lasek-Nesselquist |
| EPI_ISL_1397732 | ALBANY MEDICAL CENTER | Wadsworth Center, New York State Department of Health | Kirsten St. George, Daryl M. Lamson, Alexis Russel, Matthew Shudt, Melissa A Leisner, Jonathan Plitnick, Navjot Singh, John Kelly, Erasmus Schneider, Erica Lasek-Nesselquist |
| EPI_ISL_1397733, EPI_ISL_1397734, EPI_ISL_1397735 | ADIRONDACK MEDICAL CENTER | Wadsworth Center, New York State Department of Health | Kirsten St. George, Daryl M. Lamson, Alexis Russel, Matthew Shudt, Melissa A Leisner, Jonathan Plitnick, Navjot Singh, John Kelly, Erasmus Schneider, Erica Lasek-Nesselquist |
| EPI_ISL_1397736, EPI_ISL_1397737 | ALBANY MEDICAL CENTER | Wadsworth Center, New York State Department of Health | Kirsten St. George, Daryl M. Lamson, Alexis Russel, Matthew Shudt, Melissa A Leisner, Jonathan Plitnick, Navjot Singh, John Kelly, Erasmus Schneider, Erica Lasek-Nesselquist |
| EPI_ISL_1397738, EPI_ISL_1397739 | ADIRONDACK MEDICAL CENTER | Wadsworth Center, New York State Department of Health | Kirsten St. George, Daryl M. Lamson, Alexis Russel, Matthew Shudt, Melissa A Leisner, Jonathan Plitnick, Navjot Singh, John Kelly, Erasmus Schneider, Erica Lasek-Nesselquist |
| EPI_ISL_1397740 | ALBANY MEDICAL CENTER | Wadsworth Center, New York State Department of Health | Kirsten St. George, Daryl M. Lamson, Alexis Russel, Matthew Shudt, Melissa A Leisner, Jonathan Plitnick, Navjot Singh, John Kelly, Erasmus Schneider, Erica Lasek-Nesselquist |
| EPI_ISL_1397741 | ADIRONDACK MEDICAL CENTER | Wadsworth Center, New York State Department of Health | Kirsten St. George, Daryl M. Lamson, Alexis Russel, Matthew Shudt, Melissa A Leisner, Jonathan Plitnick, Navjot Singh, John Kelly, Erasmus Schneider, Erica Lasek-Nesselquist |
| EPI_ISL_1397742 | ALBANY MEDICAL CENTER | Wadsworth Center, New York State Department of Health | Kirsten St. George, Daryl M. Lamson, Alexis Russel, Matthew Shudt, Melissa A Leisner, Jonathan Plitnick, Navjot Singh, John Kelly, Erasmus Schneider, Erica Lasek-Nesselquist |
| EPI_ISL_1397743 | ADIRONDACK MEDICAL CENTER | Wadsworth Center, New York State Department of Health | Kirsten St. George, Daryl M. Lamson, Alexis Russel, Matthew Shudt, Melissa A Leisner, Jonathan Plitnick, Navjot Singh, John Kelly, Erasmus Schneider, Erica Lasek-Nesselquist |

| | | | |
|---|---|---|---|
| EPI_ISL_1397744 | ALBANY MEDICAL CENTER | Wadsworth Center, New York State Department of Health | Kirsten St. George, Daryl M. Lamson, Alexis Russel, Matthew Shudt, Melissa A Leisner, Jonathan Plitnick, Navjot Singh, John Kelly, Erasmus Schneider, Erica Lasek-Nesselquist |
| EPI_ISL_1397745, EPI_ISL_1397746, EPI_ISL_1397747, EPI_ISL_1397748, EPI_ISL_1397749, EPI_ISL_1397750, EPI_ISL_1397751 | ADIRONDACK MEDICAL CENTER | Wadsworth Center, New York State Department of Health | Kirsten St. George, Daryl M. Lamson, Alexis Russel, Matthew Shudt, Melissa A Leisner, Jonathan Plitnick, Navjot Singh, John Kelly, Erasmus Schneider, Erica Lasek-Nesselquist |
| EPI_ISL_1397752, EPI_ISL_1397753, EPI_ISL_1397754, EPI_ISL_1397755, EPI_ISL_1397756, EPI_ISL_1397757, EPI_ISL_1397758, EPI_ISL_1397759, EPI_ISL_1397760, EPI_ISL_1397761, EPI_ISL_1397762, EPI_ISL_1397763, EPI_ISL_1397764, EPI_ISL_1397765 | | | |
| see above | ALBANY MEDICAL CENTER | Wadsworth Center, New York State Department of Health | Kirsten St. George, Daryl M. Lamson, Alexis Russel, Matthew Shudt, Melissa A Leisner, Jonathan Plitnick, Navjot Singh, John Kelly, Erasmus Schneider, Erica Lasek-Nesselquist |
| EPI_ISL_1397766 | ADIRONDACK MEDICAL CENTER | Wadsworth Center, New York State Department of Health | Kirsten St. George, Daryl M. Lamson, Alexis Russel, Matthew Shudt, Melissa A Leisner, Jonathan Plitnick, Navjot Singh, John Kelly, Erasmus Schneider, Erica Lasek-Nesselquist |
| EPI_ISL_1397767 | ALBANY MEDICAL CENTER | Wadsworth Center, New York State Department of Health | Kirsten St. George, Daryl M. Lamson, Alexis Russel, Matthew Shudt, Melissa A Leisner, Jonathan Plitnick, Navjot Singh, John Kelly, Erasmus Schneider, Erica Lasek-Nesselquist |
| EPI_ISL_1397768, EPI_ISL_1397769, EPI_ISL_1397770, EPI_ISL_1397771, EPI_ISL_1397772, EPI_ISL_1397773, EPI_ISL_1397774, EPI_ISL_1397775, EPI_ISL_1397776, EPI_ISL_1397777, EPI_ISL_1397778, EPI_ISL_1397779, EPI_ISL_1397780, EPI_ISL_1397781, EPI_ISL_1397782 | | | |
| see above | WESTCHESTER MEDICAL CENTER | Wadsworth Center, New York State Department of Health | Kirsten St. George, Daryl M. Lamson, Alexis Russel, Matthew Shudt, Melissa A Leisner, Jonathan Plitnick, Navjot Singh, John Kelly, Erasmus Schneider, Erica Lasek-Nesselquist |
| EPI_ISL_1397783 | THE MARY IMOGENE BASSETT HOSPITAL | Wadsworth Center, New York State Department of Health | Kirsten St. George, Daryl M. Lamson, Alexis Russel, Matthew Shudt, Melissa A Leisner, Jonathan Plitnick, Navjot Singh, John Kelly, Erasmus Schneider, Erica Lasek-Nesselquist |
| EPI_ISL_1397784, EPI_ISL_1397785, EPI_ISL_1397786, EPI_ISL_1397787 | SUNY UPSTATE MEDICAL UNIVERSITY | Wadsworth Center, New York State Department of Health | Kirsten St. George, Daryl M. Lamson, Alexis Russel, Matthew Shudt, Melissa A Leisner, Jonathan Plitnick, Navjot Singh, John Kelly, Erasmus Schneider, Erica Lasek-Nesselquist |
| EPI_ISL_1397788, EPI_ISL_1397789, EPI_ISL_1397790, EPI_ISL_1397791, EPI_ISL_1397792, EPI_ISL_1397793, EPI_ISL_1397794, EPI_ISL_1397795, EPI_ISL_1397796, EPI_ISL_1397797, EPI_ISL_1397798, EPI_ISL_1397799, EPI_ISL_1397800, EPI_ISL_1397801, EPI_ISL_1397802, EPI_ISL_1397803, EPI_ISL_1397804, EPI_ISL_1397805, EPI_ISL_1397806, EPI_ISL_1397807, EPI_ISL_1397808, EPI_ISL_1397809 | | | |
| see above | MONTEFIORE MEDICAL CENTER LABORATORIES | Wadsworth Center, New York State Department of Health | Kirsten St. George, Daryl M. Lamson, Alexis Russel, Matthew Shudt, Melissa A Leisner, Jonathan Plitnick, Navjot Singh, John Kelly, Erasmus Schneider, Erica Lasek-Nesselquist |
| EPI_ISL_1397810, EPI_ISL_1397811, EPI_ISL_1397812, EPI_ISL_1397813, EPI_ISL_1397814, EPI_ISL_1397815, EPI_ISL_1397816, EPI_ISL_1397817, EPI_ISL_1397818, EPI_ISL_1397819, EPI_ISL_1397820, EPI_ISL_1397821, EPI_ISL_1397822, EPI_ISL_1397823 | | | |
| see above | URMC LABS | Wadsworth Center, New York State Department of Health | Kirsten St. George, Daryl M. Lamson, Alexis Russel, Matthew Shudt, Melissa A Leisner, Jonathan Plitnick, Navjot Singh, John Kelly, Erasmus Schneider, Erica Lasek-Nesselquist |
| EPI_ISL_1397824, EPI_ISL_1397825, EPI_ISL_1397826, EPI_ISL_1397827, EPI_ISL_1397828 | Columbia University Irving Medical Center | Wadsworth Center, New York State Department of Health | Kirsten St. George, Daryl M. Lamson, Alexis Russel, Matthew Shudt, Melissa A Leisner, Jonathan Plitnick, Navjot Singh, John Kelly, Erasmus Schneider, Erica Lasek-Nesselquist |
| EPI_ISL_1397829, EPI_ISL_1397830 | NORTHWELL HEALTH LABORATORIES | Wadsworth Center, New York State Department of Health | Kirsten St. George, Daryl M. Lamson, Alexis Russel, Matthew Shudt, Melissa A Leisner, Jonathan Plitnick, Navjot Singh, John Kelly, Erasmus Schneider, Erica Lasek-Nesselquist |
| EPI_ISL_1397831, EPI_ISL_1397832 | STONY BROOK UNIVERSITY HOSPITAL | Wadsworth Center, New York State Department of Health | Kirsten St. George, Daryl M. Lamson, Alexis Russel, Matthew Shudt, Melissa A Leisner, Jonathan Plitnick, Navjot Singh, John Kelly, Erasmus Schneider, Erica Lasek-Nesselquist |
| EPI_ISL_1397833 | Wadsworth Center, New York State Department of Health | Wadsworth Center, New York State Department of Health | Kirsten St. George, Daryl M. Lamson, Alexis Russel, Matthew Shudt, Melissa A Leisner, Jonathan Plitnick, Navjot Singh, John Kelly, Erasmus Schneider, Erica Lasek-Nesselquist |
| EPI_ISL_1397834, EPI_ISL_1397835 | ALBANY MEDICAL CENTER | Wadsworth Center, New York State Department of Health | Kirsten St. George, Daryl M. Lamson, Alexis Russel, Matthew Shudt, Melissa A Leisner, Jonathan Plitnick, Navjot Singh, John Kelly, Erasmus Schneider, Erica Lasek-Nesselquist |
| EPI_ISL_1397836 | TEMPUS LABS INC | Wadsworth Center, New York State Department of Health | Kirsten St. George, Daryl M. Lamson, Alexis Russel, Matthew Shudt, Melissa A Leisner, Jonathan Plitnick, Navjot Singh, John Kelly, Erasmus Schneider, Erica Lasek-Nesselquist |
| EPI_ISL_1397837 | ALBANY MEDICAL CENTER | Wadsworth Center, New York State Department of Health | Kirsten St. George, Daryl M. Lamson, Alexis Russel, Matthew Shudt, Melissa A Leisner, Jonathan Plitnick, Navjot Singh, John Kelly, Erasmus Schneider, Erica Lasek-Nesselquist |
| EPI_ISL_1397838, EPI_ISL_1397839, EPI_ISL_1397840, EPI_ISL_1397841, EPI_ISL_1397842 | TEMPUS LABS INC | Wadsworth Center, New York State Department of Health | Kirsten St. George, Daryl M. Lamson, Alexis Russel, Matthew Shudt, Melissa A Leisner, Jonathan Plitnick, Navjot Singh, John Kelly, Erasmus Schneider, Erica Lasek-Nesselquist |
| EPI_ISL_1397843 | ALBANY MEDICAL CENTER | Wadsworth Center, New York State Department of Health | Kirsten St. George, Daryl M. Lamson, Alexis Russel, Matthew Shudt, Melissa A Leisner, Jonathan Plitnick, Navjot Singh, John Kelly, Erasmus Schneider, Erica Lasek-Nesselquist |
| EPI_ISL_1397844, EPI_ISL_1397845, EPI_ISL_1397846, EPI_ISL_1397847, EPI_ISL_1397848, EPI_ISL_1397849, EPI_ISL_1397850, EPI_ISL_1397851, EPI_ISL_1397852 | TEMPUS LABS INC | Wadsworth Center, New York State Department of Health | Kirsten St. George, Daryl M. Lamson, Alexis Russel, Matthew Shudt, Melissa A Leisner, Jonathan Plitnick, Navjot Singh, John Kelly, Erasmus Schneider, Erica Lasek-Nesselquist |
| EPI_ISL_1397853, EPI_ISL_1397854, EPI_ISL_1397855, EPI_ISL_1397856 | ALBANY MEDICAL CENTER | Wadsworth Center, New York State Department of Health | Kirsten St. George, Daryl M. Lamson, Alexis Russel, Matthew Shudt, Melissa A Leisner, Jonathan Plitnick, Navjot Singh, John Kelly, Erasmus Schneider, Erica Lasek-Nesselquist |
| EPI_ISL_1397857 | TEMPUS LABS INC | Wadsworth Center, New York State Department of Health | Kirsten St. George, Daryl M. Lamson, Alexis Russel, Matthew Shudt, Melissa A Leisner, Jonathan Plitnick, Navjot Singh, John Kelly, Erasmus Schneider, Erica Lasek-Nesselquist |
| EPI_ISL_1397858 | ALBANY MEDICAL CENTER | Wadsworth Center, New York State Department of Health | Kirsten St. George, Daryl M. Lamson, Alexis Russel, Matthew Shudt, Melissa A Leisner, Jonathan Plitnick, Navjot Singh, John Kelly, Erasmus Schneider, Erica Lasek-Nesselquist |
| EPI_ISL_1397859, EPI_ISL_1397860 | TEMPUS LABS INC | Wadsworth Center, New York State Department of Health | Kirsten St. George, Daryl M. Lamson, Alexis Russel, Matthew Shudt, Melissa A Leisner, Jonathan Plitnick, Navjot Singh, John Kelly, Erasmus Schneider, Erica Lasek-Nesselquist |
| EPI_ISL_1397861 | ALBANY MEDICAL CENTER | Wadsworth Center, New York State Department of Health | Kirsten St. George, Daryl M. Lamson, Alexis Russel, Matthew Shudt, Melissa A Leisner, Jonathan Plitnick, Navjot Singh, John Kelly, Erasmus Schneider, Erica Lasek-Nesselquist |
| EPI_ISL_1397862, EPI_ISL_1397863, EPI_ISL_1397864, EPI_ISL_1397865 | TEMPUS LABS INC | Wadsworth Center, New York State Department of Health | Kirsten St. George, Daryl M. Lamson, Alexis Russel, Matthew Shudt, Melissa A Leisner, Jonathan Plitnick, Navjot Singh, John Kelly, Erasmus Schneider, Erica Lasek-Nesselquist |
| EPI_ISL_1397866, EPI_ISL_1397867, EPI_ISL_1397868, EPI_ISL_1397869 | ALBANY MEDICAL CENTER | Wadsworth Center, New York State Department of Health | Kirsten St. George, Daryl M. Lamson, Alexis Russel, Matthew Shudt, Melissa A Leisner, Jonathan Plitnick, Navjot Singh, John Kelly, Erasmus Schneider, Erica Lasek-Nesselquist |
| EPI_ISL_1397870, EPI_ISL_1397871, EPI_ISL_1397872, EPI_ISL_1397873, EPI_ISL_1397874, EPI_ISL_1397875, EPI_ISL_1397876, EPI_ISL_1397877, EPI_ISL_1397878, EPI_ISL_1397879, EPI_ISL_1397880, EPI_ISL_1397881, EPI_ISL_1397882, EPI_ISL_1397883, EPI_ISL_1397884, EPI_ISL_1397885 | | | |
| see above | MONTEFIORE MEDICAL CENTER LABORATORIES | Wadsworth Center, New York State Department of Health | Kirsten St. George, Daryl M. Lamson, Alexis Russel, Matthew Shudt, Melissa A Leisner, Jonathan Plitnick, Navjot Singh, John Kelly, Erasmus Schneider, Erica Lasek-Nesselquist |
| EPI_ISL_1397886, EPI_ISL_1397887, EPI_ISL_1397888, EPI_ISL_1397889, EPI_ISL_1397890, EPI_ISL_1397891, EPI_ISL_1397892, EPI_ISL_1397893, EPI_ISL_1397894, EPI_ISL_1397895, EPI_ISL_1397896, EPI_ISL_1397897, EPI_ISL_1397898, EPI_ISL_1397899, EPI_ISL_1397900, EPI_ISL_1397901, EPI_ISL_1397902, EPI_ISL_1397903, EPI_ISL_1397904, EPI_ISL_1397905, EPI_ISL_1397906 | | | |

| | | | |
|---|---|---|---|
| see above | Columbia University Irving Medical Center | Wadsworth Center, New York State Department of Health | Kirsten St. George, Daryl M. Lamson, Alexis Russel, Matthew Shudt, Melissa A Leisner, Jonathan Plitnick, Navjot Singh, John Kelly, Erasmus Schneider, Erica Lasek-Nesselquist |
| EPI_ISL_1397907, EPI_ISL_1397908, EPI_ISL_1397909, EPI_ISL_1397910, EPI_ISL_1397911 | NORTHWELL HEALTH LABORATORIES | Wadsworth Center, New York State Department of Health | Kirsten St. George, Daryl M. Lamson, Alexis Russel, Matthew Shudt, Melissa A Leisner, Jonathan Plitnick, Navjot Singh, John Kelly, Erasmus Schneider, Erica Lasek-Nesselquist |
| EPI_ISL_1397912, EPI_ISL_1397913 | BIO-REFERENCE LABORATORIES | Wadsworth Center, New York State Department of Health | Kirsten St. George, Daryl M. Lamson, Alexis Russel, Matthew Shudt, Melissa A Leisner, Jonathan Plitnick, Navjot Singh, John Kelly, Erasmus Schneider, Erica Lasek-Nesselquist |
| EPI_ISL_1397914, EPI_ISL_1397915, EPI_ISL_1397916, EPI_ISL_1397917, EPI_ISL_1397918, EPI_ISL_1397919, EPI_ISL_1397920, EPI_ISL_1397921, EPI_ISL_1397922, EPI_ISL_1397923, EPI_ISL_1397924, EPI_ISL_1397925, EPI_ISL_1397926, EPI_ISL_1397927, EPI_ISL_1397928, EPI_ISL_1397929, EPI_ISL_1397930, EPI_ISL_1397931, EPI_ISL_1397932, EPI_ISL_1397933, EPI_ISL_1397934, EPI_ISL_1397935, EPI_ISL_1397936, EPI_ISL_1397937, EPI_ISL_1397938, EPI_ISL_1397939, EPI_ISL_1397940, EPI_ISL_1397941, EPI_ISL_1397942, EPI_ISL_1397943, EPI_ISL_1397944, EPI_ISL_1397945, EPI_ISL_1397946, EPI_ISL_1397947, EPI_ISL_1397948, EPI_ISL_1397949, EPI_ISL_1397950, EPI_ISL_1397951, EPI_ISL_1397952 | | | |
| see above | WESTCHESTER MEDICAL CENTER | Wadsworth Center, New York State Department of Health | Kirsten St. George, Daryl M. Lamson, Alexis Russel, Matthew Shudt, Melissa A Leisner, Jonathan Plitnick, Navjot Singh, John Kelly, Erasmus Schneider, Erica Lasek-Nesselquist |
| EPI_ISL_1397961 | ALBANY MEDICAL CENTER | Wadsworth Center, New York State Department of Health | Kirsten St. George, Daryl M. Lamson, Alexis Russel, Matthew Shudt, Melissa A Leisner, Jonathan Plitnick, Navjot Singh, John Kelly, Erasmus Schneider, Erica Lasek-Nesselquist |
| EPI_ISL_1397962, EPI_ISL_1397963, EPI_ISL_1397964, EPI_ISL_1397965 | GLENS FALLS HOSPITAL LABORATORY | Wadsworth Center, New York State Department of Health | Kirsten St. George, Daryl M. Lamson, Alexis Russel, Matthew Shudt, Melissa A Leisner, Jonathan Plitnick, Navjot Singh, John Kelly, Erasmus Schneider, Erica Lasek-Nesselquist |
| EPI_ISL_1397966, EPI_ISL_1397967 | THE MARY IMOGENE BASSETT HOSPITAL | Wadsworth Center, New York State Department of Health | Kirsten St. George, Daryl M. Lamson, Alexis Russel, Matthew Shudt, Melissa A Leisner, Jonathan Plitnick, Navjot Singh, John Kelly, Erasmus Schneider, Erica Lasek-Nesselquist |
| EPI_ISL_1397968, EPI_ISL_1397969, EPI_ISL_1397970 | SUNY UPSTATE MEDICAL UNIVERSITY | Wadsworth Center, New York State Department of Health | Kirsten St. George, Daryl M. Lamson, Alexis Russel, Matthew Shudt, Melissa A Leisner, Jonathan Plitnick, Navjot Singh, John Kelly, Erasmus Schneider, Erica Lasek-Nesselquist |
| EPI_ISL_1397971, EPI_ISL_1397972, EPI_ISL_1397973, EPI_ISL_1397974, EPI_ISL_1397975, EPI_ISL_1397976, EPI_ISL_1397977, EPI_ISL_1397978, EPI_ISL_1397979, EPI_ISL_1397980, EPI_ISL_1397981, EPI_ISL_1397982 | | | |
| see above | URMC LABS | Wadsworth Center, New York State Department of Health | Kirsten St. George, Daryl M. Lamson, Alexis Russel, Matthew Shudt, Melissa A Leisner, Jonathan Plitnick, Navjot Singh, John Kelly, Erasmus Schneider, Erica Lasek-Nesselquist |
| EPI_ISL_1397983 | SUNY UPSTATE MEDICAL UNIVERSITY | Wadsworth Center, New York State Department of Health | Kirsten St. George, Daryl M. Lamson, Alexis Russel, Matthew Shudt, Melissa A Leisner, Jonathan Plitnick, Navjot Singh, John Kelly, Erasmus Schneider, Erica Lasek-Nesselquist |
| EPI_ISL_1397984, EPI_ISL_1397985, EPI_ISL_1397986, EPI_ISL_1397987 | URMC LABS | Wadsworth Center, New York State Department of Health | Kirsten St. George, Daryl M. Lamson, Alexis Russel, Matthew Shudt, Melissa A Leisner, Jonathan Plitnick, Navjot Singh, John Kelly, Erasmus Schneider, Erica Lasek-Nesselquist |
| EPI_ISL_1397988 | THE MARY IMOGENE BASSETT HOSPITAL | Wadsworth Center, New York State Department of Health | Kirsten St. George, Daryl M. Lamson, Alexis Russel, Matthew Shudt, Melissa A Leisner, Jonathan Plitnick, Navjot Singh, John Kelly, Erasmus Schneider, Erica Lasek-Nesselquist |
| EPI_ISL_1397989, EPI_ISL_1397990, EPI_ISL_1397991, EPI_ISL_1397992, EPI_ISL_1397993, EPI_ISL_1397994, EPI_ISL_1397995, EPI_ISL_1397996, EPI_ISL_1397997, EPI_ISL_1397998, EPI_ISL_1397999, EPI_ISL_1398000, EPI_ISL_1398001 | | | |
| see above | URMC LABS | Wadsworth Center, New York State Department of Health | Kirsten St. George, Daryl M. Lamson, Alexis Russel, Matthew Shudt, Melissa A Leisner, Jonathan Plitnick, Navjot Singh, John Kelly, Erasmus Schneider, Erica Lasek-Nesselquist |
| EPI_ISL_1398002, EPI_ISL_1398003, EPI_ISL_1398004 | SUNY UPSTATE MEDICAL UNIVERSITY | Wadsworth Center, New York State Department of Health | Kirsten St. George, Daryl M. Lamson, Alexis Russel, Matthew Shudt, Melissa A Leisner, Jonathan Plitnick, Navjot Singh, John Kelly, Erasmus Schneider, Erica Lasek-Nesselquist |
| EPI_ISL_1398005, EPI_ISL_1398006, EPI_ISL_1398007, EPI_ISL_1398008, EPI_ISL_1398009, EPI_ISL_1398010, EPI_ISL_1398011, EPI_ISL_1398012, EPI_ISL_1398013, EPI_ISL_1398014, EPI_ISL_1398015, EPI_ISL_1398016, EPI_ISL_1398017, EPI_ISL_1398018, EPI_ISL_1398019, EPI_ISL_1398020 | | | |
| see above | URMC LABS | Wadsworth Center, New York State Department of Health | Kirsten St. George, Daryl M. Lamson, Alexis Russel, Matthew Shudt, Melissa A Leisner, Jonathan Plitnick, Navjot Singh, John Kelly, Erasmus Schneider, Erica Lasek-Nesselquist |
| EPI_ISL_1398021, EPI_ISL_1398022, EPI_ISL_1398023, EPI_ISL_1398024, EPI_ISL_1398025, EPI_ISL_1398026, EPI_ISL_1398027 | ALBANY MEDICAL CENTER | Wadsworth Center, New York State Department of Health | Kirsten St. George, Daryl M. Lamson, Alexis Russel, Matthew Shudt, Melissa A Leisner, Jonathan Plitnick, Navjot Singh, John Kelly, Erasmus Schneider, Erica Lasek-Nesselquist |
| EPI_ISL_1398028, EPI_ISL_1398029, EPI_ISL_1398030, EPI_ISL_1398031, EPI_ISL_1398032, EPI_ISL_1398033 | WESTCHESTER MEDICAL CENTER | Wadsworth Center, New York State Department of Health | Kirsten St. George, Daryl M. Lamson, Alexis Russel, Matthew Shudt, Melissa A Leisner, Jonathan Plitnick, Navjot Singh, John Kelly, Erasmus Schneider, Erica Lasek-Nesselquist |
| EPI_ISL_1398034, EPI_ISL_1398035, EPI_ISL_1398036, EPI_ISL_1398037, EPI_ISL_1398038 | MONTEFIORE MEDICAL CENTER LABORATORIES | Wadsworth Center, New York State Department of Health | Kirsten St. George, Daryl M. Lamson, Alexis Russel, Matthew Shudt, Melissa A Leisner, Jonathan Plitnick, Navjot Singh, John Kelly, Erasmus Schneider, Erica Lasek-Nesselquist |
| EPI_ISL_1398040, EPI_ISL_1398041 | MEMORIAL SLOAN KETTERING CANCER CENTER | Wadsworth Center, New York State Department of Health | Kirsten St. George, Daryl M. Lamson, Alexis Russel, Matthew Shudt, Melissa A Leisner, Jonathan Plitnick, Navjot Singh, John Kelly, Erasmus Schneider, Erica Lasek-Nesselquist |
| EPI_ISL_1398042 | NORTHWELL HEALTH LABORATORIES | Wadsworth Center, New York State Department of Health | Kirsten St. George, Daryl M. Lamson, Alexis Russel, Matthew Shudt, Melissa A Leisner, Jonathan Plitnick, Navjot Singh, John Kelly, Erasmus Schneider, Erica Lasek-Nesselquist |
| EPI_ISL_1398043 | MEMORIAL SLOAN KETTERING CANCER CENTER | Wadsworth Center, New York State Department of Health | Kirsten St. George, Daryl M. Lamson, Alexis Russel, Matthew Shudt, Melissa A Leisner, Jonathan Plitnick, Navjot Singh, John Kelly, Erasmus Schneider, Erica Lasek-Nesselquist |
| EPI_ISL_1398044, EPI_ISL_1398045, EPI_ISL_1398046, EPI_ISL_1398047 | Columbia University Irving Medical Center | Wadsworth Center, New York State Department of Health | Kirsten St. George, Daryl M. Lamson, Alexis Russel, Matthew Shudt, Melissa A Leisner, Jonathan Plitnick, Navjot Singh, John Kelly, Erasmus Schneider, Erica Lasek-Nesselquist |
| EPI_ISL_1398048, EPI_ISL_1398049, EPI_ISL_1398050, EPI_ISL_1398051, EPI_ISL_1398052, EPI_ISL_1398053, EPI_ISL_1398054, EPI_ISL_1398055, EPI_ISL_1398056, EPI_ISL_1398057, EPI_ISL_1398058, EPI_ISL_1398059, EPI_ISL_1398060, EPI_ISL_1398061, EPI_ISL_1398062, EPI_ISL_1398063, EPI_ISL_1398064, EPI_ISL_1398065, EPI_ISL_1398066, EPI_ISL_1398067 | | | |
| see above | MONTEFIORE MEDICAL CENTER LABORATORIES | Wadsworth Center, New York State Department of Health | Kirsten St. George, Daryl M. Lamson, Alexis Russel, Matthew Shudt, Melissa A Leisner, Jonathan Plitnick, Navjot Singh, John Kelly, Erasmus Schneider, Erica Lasek-Nesselquist |
| EPI_ISL_1398068, EPI_ISL_1398069, EPI_ISL_1398070, EPI_ISL_1398071, EPI_ISL_1398072, EPI_ISL_1398073, EPI_ISL_1398074, EPI_ISL_1398075, EPI_ISL_1398076, EPI_ISL_1398077, EPI_ISL_1398078, EPI_ISL_1398079, EPI_ISL_1398080, EPI_ISL_1398081, EPI_ISL_1398082, EPI_ISL_1398083, EPI_ISL_1398084, EPI_ISL_1398085, EPI_ISL_1398086, EPI_ISL_1398087, EPI_ISL_1398088, EPI_ISL_1398089, EPI_ISL_1398090, EPI_ISL_1398091, EPI_ISL_1398092, EPI_ISL_1398093, EPI_ISL_1398094, EPI_ISL_1398095, EPI_ISL_1398096, EPI_ISL_1398097, EPI_ISL_1398098, EPI_ISL_1398099, EPI_ISL_1398100, EPI_ISL_1398101, EPI_ISL_1398102, EPI_ISL_1398103, EPI_ISL_1398104, EPI_ISL_1398105, EPI_ISL_1398106, EPI_ISL_1398107, EPI_ISL_1398108, EPI_ISL_1398109, EPI_ISL_1398110, EPI_ISL_1398111, EPI_ISL_1398112, EPI_ISL_1398113, EPI_ISL_1398114, EPI_ISL_1398115, EPI_ISL_1398116, EPI_ISL_1398117, EPI_ISL_1398118, EPI_ISL_1398119 | | | |
| see above | NORTHWELL HEALTH LABORATORIES | Wadsworth Center, New York State Department of Health | Kirsten St. George, Daryl M. Lamson, Alexis Russel, Matthew Shudt, Melissa A Leisner, Jonathan Plitnick, Navjot Singh, John Kelly, Erasmus Schneider, Erica Lasek-Nesselquist |
| EPI_ISL_1398120, EPI_ISL_1398121, EPI_ISL_1398122 | MONTEFIORE MEDICAL CENTER LABORATORIES | Wadsworth Center, New York State Department of Health | Kirsten St. George, Daryl M. Lamson, Alexis Russel, Matthew Shudt, Melissa A Leisner, Jonathan Plitnick, Navjot Singh, John Kelly, Erasmus Schneider, Erica Lasek-Nesselquist |
| EPI_ISL_1398123, EPI_ISL_1398124, EPI_ISL_1398125, EPI_ISL_1398126, EPI_ISL_1398127, EPI_ISL_1398128, EPI_ISL_1398129, EPI_ISL_1398130, EPI_ISL_1398131 | URMC LABS | Wadsworth Center, New York State Department of Health | Kirsten St. George, Daryl M. Lamson, Alexis Russel, Matthew Shudt, Melissa A Leisner, Jonathan Plitnick, Navjot Singh, John Kelly, Erasmus Schneider, Erica Lasek-Nesselquist |

| | | | |
|---|---|---|---|
| EPI_ISL_1398132, EPI_ISL_1398133, EPI_ISL_1398134, EPI_ISL_1398135, EPI_ISL_1398136 | MONTEFIORE MEDICAL CENTER LABORATORIES | Wadsworth Center, New York State Department of Health | Kirsten St. George, Daryl M. Lamson, Alexis Russel, Matthew Shudt, Melissa A Leisner, Jonathan Plitnick, Navjot Singh, John Kelly, Erasmus Schneider, Erica Lasek-Nesselquist |
| EPI_ISL_1398137, EPI_ISL_1398138, EPI_ISL_1398139, EPI_ISL_1398140, EPI_ISL_1398141, EPI_ISL_1398142, EPI_ISL_1398143 | URMC LABS | Wadsworth Center, New York State Department of Health | Kirsten St. George, Daryl M. Lamson, Alexis Russel, Matthew Shudt, Melissa A Leisner, Jonathan Plitnick, Navjot Singh, John Kelly, Erasmus Schneider, Erica Lasek-Nesselquist |
| EPI_ISL_1398144, EPI_ISL_1398145, EPI_ISL_1398146 | Columbia University Irving Medical Center | Wadsworth Center, New York State Department of Health | Kirsten St. George, Daryl M. Lamson, Alexis Russel, Matthew Shudt, Melissa A Leisner, Jonathan Plitnick, Navjot Singh, John Kelly, Erasmus Schneider, Erica Lasek-Nesselquist |
| EPI_ISL_1398147, EPI_ISL_1398148, EPI_ISL_1398149, EPI_ISL_1398150, EPI_ISL_1398151, EPI_ISL_1398152, EPI_ISL_1398153 | ALBANY MEDICAL CENTER | Wadsworth Center, New York State Department of Health | Kirsten St. George, Daryl M. Lamson, Alexis Russel, Matthew Shudt, Melissa A Leisner, Jonathan Plitnick, Navjot Singh, John Kelly, Erasmus Schneider, Erica Lasek-Nesselquist |
| EPI_ISL_1398154, EPI_ISL_1398155 | GLENS FALLS HOSPITAL LABORATORY | Wadsworth Center, New York State Department of Health | Kirsten St. George, Daryl M. Lamson, Alexis Russel, Matthew Shudt, Melissa A Leisner, Jonathan Plitnick, Navjot Singh, John Kelly, Erasmus Schneider, Erica Lasek-Nesselquist |
| EPI_ISL_1398156, EPI_ISL_1398157, EPI_ISL_1398158, EPI_ISL_1398159, EPI_ISL_1398160, EPI_ISL_1398161, EPI_ISL_1398162 | SUNY UPSTATE MEDICAL UNIVERSITY | Wadsworth Center, New York State Department of Health | Kirsten St. George, Daryl M. Lamson, Alexis Russel, Matthew Shudt, Melissa A Leisner, Jonathan Plitnick, Navjot Singh, John Kelly, Erasmus Schneider, Erica Lasek-Nesselquist |
| EPI_ISL_1398163, EPI_ISL_1398164, EPI_ISL_1398165, EPI_ISL_1398166, EPI_ISL_1398167, EPI_ISL_1398168, EPI_ISL_1398169, EPI_ISL_1398170, EPI_ISL_1398171, EPI_ISL_1398172, EPI_ISL_1398173, EPI_ISL_1398174, EPI_ISL_1398175, EPI_ISL_1398176, EPI_ISL_1398177, EPI_ISL_1398178, EPI_ISL_1398179, EPI_ISL_1398180, EPI_ISL_1398181, EPI_ISL_1398182, EPI_ISL_1398183, EPI_ISL_1398184, EPI_ISL_1398185, EPI_ISL_1398186, EPI_ISL_1398187, EPI_ISL_1398188, EPI_ISL_1398189, EPI_ISL_1398190, EPI_ISL_1398191, EPI_ISL_1398192, EPI_ISL_1398193, EPI_ISL_1398194, EPI_ISL_1398195, EPI_ISL_1398196, EPI_ISL_1398197, EPI_ISL_1398198, EPI_ISL_1398199, EPI_ISL_1398200, EPI_ISL_1398201, EPI_ISL_1398202, EPI_ISL_1398203 | | | |
| see above | Columbia University Irving Medical Center | Wadsworth Center, New York State Department of Health | Kirsten St. George, Daryl M. Lamson, Alexis Russel, Matthew Shudt, Melissa A Leisner, Jonathan Plitnick, Navjot Singh, John Kelly, Erasmus Schneider, Erica Lasek-Nesselquist |
| EPI_ISL_1398204, EPI_ISL_1398205, EPI_ISL_1398206, EPI_ISL_1398207, EPI_ISL_1398208 | LENOX HILL HOSPITAL | Wadsworth Center, New York State Department of Health | Kirsten St. George, Daryl M. Lamson, Alexis Russel, Matthew Shudt, Melissa A Leisner, Jonathan Plitnick, Navjot Singh, John Kelly, Erasmus Schneider, Erica Lasek-Nesselquist |
| EPI_ISL_1398209, EPI_ISL_1398210, EPI_ISL_1398211, EPI_ISL_1398212, EPI_ISL_1398213, EPI_ISL_1398214, EPI_ISL_1398215, EPI_ISL_1398216, EPI_ISL_1398217, EPI_ISL_1398218, EPI_ISL_1398219, EPI_ISL_1398220, EPI_ISL_1398221, EPI_ISL_1398222, EPI_ISL_1398223, EPI_ISL_1398224, EPI_ISL_1398225, EPI_ISL_1398226, EPI_ISL_1398227, EPI_ISL_1398228, EPI_ISL_1398229, EPI_ISL_1398230, EPI_ISL_1398231, EPI_ISL_1398232, EPI_ISL_1398233, EPI_ISL_1398234, EPI_ISL_1398235, EPI_ISL_1398236, EPI_ISL_1398237, EPI_ISL_1398238, EPI_ISL_1398239, EPI_ISL_1398240, EPI_ISL_1398241, EPI_ISL_1398242, EPI_ISL_1398243, EPI_ISL_1398244, EPI_ISL_1398245, EPI_ISL_1398246, EPI_ISL_1398247, EPI_ISL_1398248, EPI_ISL_1398249, EPI_ISL_1398250, EPI_ISL_1398251, EPI_ISL_1398252, EPI_ISL_1398253, EPI_ISL_1398254, EPI_ISL_1398255, EPI_ISL_1398256, EPI_ISL_1398257, EPI_ISL_1398258 | | | |
| see above | KALEIDA CENTER FOR LABORATORY MEDICINE | Wadsworth Center, New York State Department of Health | Kirsten St. George, Daryl M. Lamson, Alexis Russel, Matthew Shudt, Melissa A Leisner, Jonathan Plitnick, Navjot Singh, John Kelly, Erasmus Schneider, Erica Lasek-Nesselquist |
| EPI_ISL_1398259, EPI_ISL_1398260, EPI_ISL_1398261, EPI_ISL_1398262, EPI_ISL_1398263 | THE MARY IMOGENE BASSETT HOSPITAL | Wadsworth Center, New York State Department of Health | Kirsten St. George, Daryl M. Lamson, Alexis Russel, Matthew Shudt, Melissa A Leisner, Jonathan Plitnick, Navjot Singh, John Kelly, Erasmus Schneider, Erica Lasek-Nesselquist |
| EPI_ISL_1398268 | WHITE PLAINS HOSPITAL CENTER LABORATORY | Wadsworth Center, New York State Department of Health | Kirsten St. George, Daryl M. Lamson, Alexis Russel, Matthew Shudt, Melissa A Leisner, Jonathan Plitnick, Navjot Singh, John Kelly, Erasmus Schneider, Erica Lasek-Nesselquist |
| EPI_ISL_1398269 | SARATOGA HOSPITAL LABORATORY | Wadsworth Center, New York State Department of Health | Kirsten St. George, Daryl M. Lamson, Alexis Russel, Matthew Shudt, Melissa A Leisner, Jonathan Plitnick, Navjot Singh, John Kelly, Erasmus Schneider, Erica Lasek-Nesselquist |
| EPI_ISL_1398270 | GLENS FALLS HOSPITAL LABORATORY | Wadsworth Center, New York State Department of Health | Kirsten St. George, Daryl M. Lamson, Alexis Russel, Matthew Shudt, Melissa A Leisner, Jonathan Plitnick, Navjot Singh, John Kelly, Erasmus Schneider, Erica Lasek-Nesselquist |
| EPI_ISL_1398271, EPI_ISL_1398272, EPI_ISL_1398273 | URMC LABS | Wadsworth Center, New York State Department of Health | Kirsten St. George, Daryl M. Lamson, Alexis Russel, Matthew Shudt, Melissa A Leisner, Jonathan Plitnick, Navjot Singh, John Kelly, Erasmus Schneider, Erica Lasek-Nesselquist |
| EPI_ISL_1398274, EPI_ISL_1398275, EPI_ISL_1398276, EPI_ISL_1398277, EPI_ISL_1398278, EPI_ISL_1398279, EPI_ISL_1398280, EPI_ISL_1398281, EPI_ISL_1398282, EPI_ISL_1398283 | ALBANY MEDICAL CENTER | Wadsworth Center, New York State Department of Health | Kirsten St. George, Daryl M. Lamson, Alexis Russel, Matthew Shudt, Melissa A Leisner, Jonathan Plitnick, Navjot Singh, John Kelly, Erasmus Schneider, Erica Lasek-Nesselquist |
| EPI_ISL_1398284, EPI_ISL_1398285, EPI_ISL_1398286, EPI_ISL_1398287, EPI_ISL_1398288, EPI_ISL_1398289, EPI_ISL_1398290, EPI_ISL_1398291, EPI_ISL_1398292, EPI_ISL_1398293, EPI_ISL_1398294, EPI_ISL_1398295, EPI_ISL_1398296, EPI_ISL_1398297, EPI_ISL_1398298, EPI_ISL_1398299, EPI_ISL_1398300, EPI_ISL_1398301, EPI_ISL_1398302, EPI_ISL_1398303, EPI_ISL_1398304, EPI_ISL_1398305, EPI_ISL_1398306, EPI_ISL_1398307, EPI_ISL_1398308, EPI_ISL_1398309, EPI_ISL_1398310, EPI_ISL_1398311 | | | |
| see above | NYC Pandemic Response Lab | Wadsworth Center, New York State Department of Health | Kirsten St. George, Daryl M. Lamson, Alexis Russel, Matthew Shudt, Melissa A Leisner, Jonathan Plitnick, Navjot Singh, John Kelly, Erasmus Schneider, Erica Lasek-Nesselquist |
| EPI_ISL_1398312, EPI_ISL_1398313, EPI_ISL_1398314, EPI_ISL_1398315, EPI_ISL_1398316, EPI_ISL_1398317 | URMC LABS | Wadsworth Center, New York State Department of Health | Kirsten St. George, Daryl M. Lamson, Alexis Russel, Matthew Shudt, Melissa A Leisner, Jonathan Plitnick, Navjot Singh, John Kelly, Erasmus Schneider, Erica Lasek-Nesselquist |
| EPI_ISL_1398318, EPI_ISL_1398319, EPI_ISL_1398320, EPI_ISL_1398321, EPI_ISL_1398322, EPI_ISL_1398323, EPI_ISL_1398324, EPI_ISL_1398325, EPI_ISL_1398326, EPI_ISL_1398327, EPI_ISL_1398328, EPI_ISL_1398329, EPI_ISL_1398330 | | | |
| see above | NYC Pandemic Response Lab | Wadsworth Center, New York State Department of Health | Kirsten St. George, Daryl M. Lamson, Alexis Russel, Matthew Shudt, Melissa A Leisner, Jonathan Plitnick, Navjot Singh, John Kelly, Erasmus Schneider, Erica Lasek-Nesselquist |
| EPI_ISL_1398331, EPI_ISL_1398333, EPI_ISL_1398334, EPI_ISL_1398335, EPI_ISL_1398336, EPI_ISL_1398337, EPI_ISL_1398338, EPI_ISL_1398339 | MONTEFIORE MEDICAL CENTER LABORATORIES | Wadsworth Center, New York State Department of Health | Kirsten St. George, Daryl M. Lamson, Alexis Russel, Matthew Shudt, Melissa A Leisner, Jonathan Plitnick, Navjot Singh, John Kelly, Erasmus Schneider, Erica Lasek-Nesselquist |
| EPI_ISL_1398340, EPI_ISL_1398341, EPI_ISL_1398342, EPI_ISL_1398343, EPI_ISL_1398344, EPI_ISL_1398345, EPI_ISL_1398346, EPI_ISL_1398347, EPI_ISL_1398348, EPI_ISL_1398349, EPI_ISL_1398350, EPI_ISL_1398351, EPI_ISL_1398352 | | | |
| see above | NYC Pandemic Response Lab | Wadsworth Center, New York State Department of Health | Kirsten St. George, Daryl M. Lamson, Alexis Russel, Matthew Shudt, Melissa A Leisner, Jonathan Plitnick, Navjot Singh, John Kelly, Erasmus Schneider, Erica Lasek-Nesselquist |
| EPI_ISL_1400985, EPI_ISL_1400986, EPI_ISL_1400987, EPI_ISL_1400988, EPI_ISL_1400989, EPI_ISL_1400990, EPI_ISL_1400991, EPI_ISL_1400992, EPI_ISL_1400993, EPI_ISL_1400994, EPI_ISL_1400995, EPI_ISL_1400996, EPI_ISL_1400997, EPI_ISL_1400998, EPI_ISL_1400999, EPI_ISL_1401000, EPI_ISL_1401001, EPI_ISL_1401002, EPI_ISL_1401003, EPI_ISL_1401004, EPI_ISL_1401005, EPI_ISL_1401006, EPI_ISL_1401007, EPI_ISL_1401008, EPI_ISL_1401009, EPI_ISL_1401010, EPI_ISL_1401011, EPI_ISL_1401012, EPI_ISL_1401013, EPI_ISL_1401014, EPI_ISL_1401015, EPI_ISL_1401016, EPI_ISL_1401017, EPI_ISL_1401018, EPI_ISL_1401019, EPI_ISL_1401020, EPI_ISL_1401021, EPI_ISL_1401022, EPI_ISL_1401023, EPI_ISL_1401024, EPI_ISL_1401025, EPI_ISL_1401026, EPI_ISL_1401027, EPI_ISL_1401028, EPI_ISL_1401029, EPI_ISL_1401030, EPI_ISL_1401031, EPI_ISL_1401032, EPI_ISL_1401033, EPI_ISL_1401034, EPI_ISL_1401035, EPI_ISL_1401036, EPI_ISL_1401037, EPI_ISL_1401038, EPI_ISL_1401039, EPI_ISL_1401040, EPI_ISL_1401041, EPI_ISL_1401042, EPI_ISL_1401043, EPI_ISL_1401044, EPI_ISL_1401045, EPI_ISL_1401046, EPI_ISL_1401047, EPI_ISL_1401048, EPI_ISL_1401049, EPI_ISL_1401050, EPI_ISL_1401051, EPI_ISL_1401052, EPI_ISL_1401053, EPI_ISL_1401054, EPI_ISL_1401055, EPI_ISL_1401056, EPI_ISL_1401057, EPI_ISL_1401058, EPI_ISL_1401059, EPI_ISL_1401060, EPI_ISL_1401061, EPI_ISL_1401062, EPI_ISL_1401063, EPI_ISL_1401064, EPI_ISL_1401065, EPI_ISL_1401066, EPI_ISL_1401067, EPI_ISL_1401068, EPI_ISL_1401069 | | | |
| see above | Erie County Public Health (ECPHL) | University at Buffalo Genomics and Bioinformatics Core | Jonathan Bard, Natalie Lamb, Alyssa Pohlman, Brandon Marzullo, Amanda Boccolucci, Norma Nowak, Donald Yergeau, Jennifer Surtees |
| EPI_ISL_1413241, EPI_ISL_1413243, EPI_ISL_1413246, EPI_ISL_1413263, EPI_ISL_1413350 | Broad Institute Clinical Research Sequencing Platform | Infectious Disease Program, Broad Institute of Harvard and MIT | Siddle,K.J., Adams,G., Pearlman,L., Gladden-Young,A., Vicente,G., Blumenstiel,B., DeFelice,M., Lee,M., McGovern,S., Lagerborg,K., Rudy,M., DeRuff,K., Carter,A., Normandin,E., Bauer,M., Reilly,S., Tomkins-Tinch,C., Loreth,C., Chaluvadi,S., Meldrim,J., Granger,B., Lemieux,J.E., Birren,B.W., Sabeti,P.C., Larkin,K., Dodge,S., Lennon,N., Madoff,L., Brown,C., Gallagher,G., Smole,S., Park,D.J., Gabriel,S., and MacInnis,B.L. |

| | | | |
|---|---|---|---|
| EPI_ISL_1420931 | Helix/Illumina | Centers for Disease Control and Prevention Division of Viral Diseases, Pathogen Discovery | Peter W. Cook, Dakota Howard, Dhwani Batra, Ben L. Rambo-Martin, Eileen de Feo, Jan Antico, Christine Tran, Matthew Tolentino, Shannon Wickline, Kim Gietzen, Brad Sickler, Jingtao Liu, Eric Allen, Phil Febbo, Summer Galloway, Nicole L. Washington, Simon White, Geraint Levan, Kelly Schiabor Barrett, Elizabeth Cirulli, Alexandre Bolze, Ary Ascencio, Charlotte Rivera-Garcia, Ryan Cho, Jason Nguyen, Sherry Wang, Jimmy Ramirez, Tyler Cassens, Efren Sandoval, Magnus Isaksson, William Lee, David Becker, Marc Laurent, James Lu, Clinton R. Paden, Suxiang Tong, Duncan MacCannell |
| EPI_ISL_1421765, EPI_ISL_1421766, EPI_ISL_1421853, EPI_ISL_1421854, EPI_ISL_1421861, EPI_ISL_1421871, EPI_ISL_1421879, EPI_ISL_1421885, EPI_ISL_1421886, EPI_ISL_1421894, EPI_ISL_1421895, EPI_ISL_1421897, EPI_ISL_1421925, EPI_ISL_1421926, EPI_ISL_1421927, EPI_ISL_1421931, EPI_ISL_1421934, EPI_ISL_1421939, EPI_ISL_1421941, EPI_ISL_1421942, EPI_ISL_1421955, EPI_ISL_1421956, EPI_ISL_1421958, EPI_ISL_1421961, EPI_ISL_1421965, EPI_ISL_1421967, EPI_ISL_1421976, EPI_ISL_1421977, EPI_ISL_1421995, EPI_ISL_1421997, EPI_ISL_1422001, EPI_ISL_1422004, EPI_ISL_1422025, EPI_ISL_1422044, EPI_ISL_1422046, EPI_ISL_1422077, EPI_ISL_1422083, EPI_ISL_1422086, EPI_ISL_1422094, EPI_ISL_1422095, EPI_ISL_1422106, EPI_ISL_1422107, EPI_ISL_1422108, EPI_ISL_1422109, EPI_ISL_1422110, EPI_ISL_1422111, EPI_ISL_1422112, EPI_ISL_1422113, EPI_ISL_1422114, EPI_ISL_1422115, EPI_ISL_1422116, EPI_ISL_1422146, EPI_ISL_1422147, EPI_ISL_1422148, EPI_ISL_1422182, EPI_ISL_1422186, EPI_ISL_1422187 | | | |
| see above | Laboratory Corporation of America | Centers for Disease Control and Prevention Division of Viral Diseases, Pathogen Discovery | Peter W. Cook, Dakota Howard, Dhwani Batra, Ben L. Rambo-Martin, Minoo Agarwal, Eyad Almasri, Debbie Boles, Ayla Burns, Nuthawin Charoensri, Oren Cohen, Susan Countryman, Mary Ann Cristobal, Bobbi Croy, Suzanne Dale, Hrushikesh Deshmukh, Amanda Douglas, Vincent Drouillon, Marcia Eisenberg, Howard Engler, Rama Ghatti, Prashant Gupta, Susan Hicks, Jake Humphrey, Lax Iyer, Manoj Jain, Mohan Kolli, Brian Krueger, Tim Kuphal, Stanley Letovsky, Michael Levandoski, Craig Lukasik, Jonathan Meltzer, Brian Norvell, Mindy Nye, Scott Parker, Christos Petropoulos, John Pruitt, Steven Ragan, Scott Ryan, Mike Sapeta, Jana Schroth, Suresh Babu Selvaraju, Goran Stevovic, Amanda Suchanek, Andrea Throop, Lyndon Tilson, Thomas Urban, Joe Voshell, Kimberly Wagner, Jonathan Williams, Mary Williamson, Qian Zeng, Tricia Zwiefelhofer, Clinton R. Paden, Suxiang Tong, Duncan MacCannell |
| EPI_ISL_1422334, EPI_ISL_1422337, EPI_ISL_1422350, EPI_ISL_1422481, EPI_ISL_1422804, EPI_ISL_1422813, EPI_ISL_1422814, EPI_ISL_1422823, EPI_ISL_1422859, EPI_ISL_1422864, EPI_ISL_1422886, EPI_ISL_1422888, EPI_ISL_1422902, EPI_ISL_1422905, EPI_ISL_1422953, EPI_ISL_1422954, EPI_ISL_1422959, EPI_ISL_1422974 | | | |
| see above | Quest Diagnostics Incorporated | Centers for Disease Control and Prevention Division of Viral Diseases, Pathogen Discovery | Peter W. Cook, Dakota Howard, Dhwani Batra, Ben L. Rambo-Martin, S. H. Rosenthal, A. Gerasimova, R. M. Kagan, B. Anderson, M. Hua, Y. Liu, L.E. Bernstein, K.E. Livingston, A. Perez, I. A. Shlyakhter, R. V. Rolando, R. Owen, P. Tanpaiboon, F. Lacbawan, Clinton R. Paden, Suxiang Tong, Duncan MacCannell |
| EPI_ISL_1423376, EPI_ISL_1423377, EPI_ISL_1423378, EPI_ISL_1423380, EPI_ISL_1423381, EPI_ISL_1423382, EPI_ISL_1423383, EPI_ISL_1423384, EPI_ISL_1423385, EPI_ISL_1423386, EPI_ISL_1423387, EPI_ISL_1423388, EPI_ISL_1423389, EPI_ISL_1423390, EPI_ISL_1423391, EPI_ISL_1423392, EPI_ISL_1423393, EPI_ISL_1423394, EPI_ISL_1423395, EPI_ISL_1423396, EPI_ISL_1423397, EPI_ISL_1423398, EPI_ISL_1423399, EPI_ISL_1423400, EPI_ISL_1423401, EPI_ISL_1423402, EPI_ISL_1423403, EPI_ISL_1423404, EPI_ISL_1423405, EPI_ISL_1423406, EPI_ISL_1423407, EPI_ISL_1423408, EPI_ISL_1423409, EPI_ISL_1423410, EPI_ISL_1423411, EPI_ISL_1423412, EPI_ISL_1423413, EPI_ISL_1423414, EPI_ISL_1423415, EPI_ISL_1423416, EPI_ISL_1423417, EPI_ISL_1423418, EPI_ISL_1423419, EPI_ISL_1423420, EPI_ISL_1423421, EPI_ISL_1423422, EPI_ISL_1423423, EPI_ISL_1423424, EPI_ISL_1423425, EPI_ISL_1423426, EPI_ISL_1423427, EPI_ISL_1423428, EPI_ISL_1423429, EPI_ISL_1423430, EPI_ISL_1423431, EPI_ISL_1423432, EPI_ISL_1423433, EPI_ISL_1423434, EPI_ISL_1423435, EPI_ISL_1423436, EPI_ISL_1423437, EPI_ISL_1423438, EPI_ISL_1423439, EPI_ISL_1423440, EPI_ISL_1423441, EPI_ISL_1423442, EPI_ISL_1423443, EPI_ISL_1423444, EPI_ISL_1423447, EPI_ISL_1423448, EPI_ISL_1423449, EPI_ISL_1423450, EPI_ISL_1423451, EPI_ISL_1423452, EPI_ISL_1423453, EPI_ISL_1423454, EPI_ISL_1423455, EPI_ISL_1423456, EPI_ISL_1423457, EPI_ISL_1423458, EPI_ISL_1423459, EPI_ISL_1423460, EPI_ISL_1423461, EPI_ISL_1423462, EPI_ISL_1423463, EPI_ISL_1423464, EPI_ISL_1423465, EPI_ISL_1423466, EPI_ISL_1423467, EPI_ISL_1423468, EPI_ISL_1423469, EPI_ISL_1423470, EPI_ISL_1423471, EPI_ISL_1423472, EPI_ISL_1423473, EPI_ISL_1423474, EPI_ISL_1423475, EPI_ISL_1423476, EPI_ISL_1423477, EPI_ISL_1423478, EPI_ISL_1423479, EPI_ISL_1423480, EPI_ISL_1423481, EPI_ISL_1423483, EPI_ISL_1423484, EPI_ISL_1423485, EPI_ISL_1423486, EPI_ISL_1423487, EPI_ISL_1423488, EPI_ISL_1423489, EPI_ISL_1423490, EPI_ISL_1423492, EPI_ISL_1423493, EPI_ISL_1423494, EPI_ISL_1423495, EPI_ISL_1423496, EPI_ISL_1423497, EPI_ISL_1423498, EPI_ISL_1423499, EPI_ISL_1423500, EPI_ISL_1423501, EPI_ISL_1423502, EPI_ISL_1423503 | | | |
| see above | NYU Langone Health | Departments of Pathology and Medicine, New York University School of Medicine | Adriana Heguy, Dacia Dimartino, Emily Guzman, Christian Marier, Peter Meyn, Sitharam Ramaswami, Gael Westby, Paul Zappile, Yutong Zhang, Paolo Cotzia, Guiqing Wang |
| EPI_ISL_1443911 | Helix/Illumina | Centers for Disease Control and Prevention Division of Viral Diseases, Pathogen Discovery | Peter W. Cook, Dakota Howard, Dhwani Batra, Ben L. Rambo-Martin, Eileen de Feo, Jan Antico, Christine Tran, Matthew Tolentino, Shannon Wickline, Kim Gietzen, Brad Sickler, Jingtao Liu, Eric Allen, Phil Febbo, Summer Galloway, Nicole L. Washington, Simon White, Geraint Levan, Kelly Schiabor Barrett, Elizabeth Cirulli, Alexandre Bolze, Ary Ascencio, Charlotte Rivera-Garcia, Ryan Cho, Jason Nguyen, Sherry Wang, Jimmy Ramirez, Tyler Cassens, Efren Sandoval, Magnus Isaksson, William Lee, David Becker, Marc Laurent, James Lu, Clinton R. Paden, Suxiang Tong, Duncan MacCannell |
| EPI_ISL_1444909, EPI_ISL_1444910, EPI_ISL_1444911, EPI_ISL_1444912, EPI_ISL_1444913, EPI_ISL_1444914, EPI_ISL_1444915, EPI_ISL_1444916, EPI_ISL_1444917, EPI_ISL_1444918, EPI_ISL_1444919, EPI_ISL_1444920, EPI_ISL_1444921, EPI_ISL_1444922, EPI_ISL_1444923, EPI_ISL_1444924, EPI_ISL_1444925, EPI_ISL_1444926, EPI_ISL_1444927, EPI_ISL_1444928, EPI_ISL_1444929, EPI_ISL_1444930, EPI_ISL_1444931, EPI_ISL_1444932, EPI_ISL_1444933, EPI_ISL_1444934, EPI_ISL_1444935, EPI_ISL_1444936, EPI_ISL_1444937, EPI_ISL_1444938, EPI_ISL_1444939, EPI_ISL_1444940, EPI_ISL_1444941, EPI_ISL_1444942, EPI_ISL_1444943, EPI_ISL_1444944, EPI_ISL_1444945, EPI_ISL_1444946, EPI_ISL_1444947, EPI_ISL_1444948, EPI_ISL_1444949, EPI_ISL_1444950, EPI_ISL_1444951, EPI_ISL_1444952, EPI_ISL_1444953, EPI_ISL_1444954, EPI_ISL_1444955, EPI_ISL_1444956, EPI_ISL_1444957, EPI_ISL_1444958, EPI_ISL_1444959, EPI_ISL_1444960, EPI_ISL_1444961, EPI_ISL_1444962, EPI_ISL_1444963, EPI_ISL_1444964, EPI_ISL_1444965, EPI_ISL_1444966, EPI_ISL_1444967, EPI_ISL_1444968, EPI_ISL_1444969, EPI_ISL_1444970, EPI_ISL_1444971, EPI_ISL_1444972, EPI_ISL_1444973, EPI_ISL_1444974, EPI_ISL_1444975, EPI_ISL_1444976, EPI_ISL_1444977, EPI_ISL_1444978, EPI_ISL_1444979, EPI_ISL_1444980, EPI_ISL_1444981, EPI_ISL_1444982, EPI_ISL_1444983, EPI_ISL_1444984, EPI_ISL_1444985, EPI_ISL_1444986, EPI_ISL_1444987, EPI_ISL_1444988, EPI_ISL_1444989, EPI_ISL_1444990, EPI_ISL_1444991, EPI_ISL_1444992, EPI_ISL_1444993, EPI_ISL_1444994, EPI_ISL_1444995, EPI_ISL_1444996, EPI_ISL_1444997, EPI_ISL_1444998, EPI_ISL_1444999, EPI_ISL_1445000, EPI_ISL_1445001, EPI_ISL_1445002, EPI_ISL_1445003, EPI_ISL_1445004, EPI_ISL_1445005, EPI_ISL_1445006, EPI_ISL_1445007, EPI_ISL_1445008, EPI_ISL_1445009, EPI_ISL_1445010, EPI_ISL_1445011, EPI_ISL_1445012, EPI_ISL_1445013, EPI_ISL_1445014, EPI_ISL_1445015, EPI_ISL_1445016, EPI_ISL_1445017, EPI_ISL_1445018, EPI_ISL_1445019 | | | |
| see above | Erie County Public Health (ECPHL) | University at Buffalo Genomics and Bioinformatics Core | Jonathan Bard, Natalie Lamb, Alyssa Pohlman, Brandon Marzullo, Amanda Boccolucci, Norma Nowak, Donald Yergeau, Jennifer Surtees |
| EPI_ISL_1445307, EPI_ISL_1445308, EPI_ISL_1445380, EPI_ISL_1445424, EPI_ISL_1445443, EPI_ISL_1445702, EPI_ISL_1445705, EPI_ISL_1445878, EPI_ISL_1445881, EPI_ISL_1445889, EPI_ISL_1445906, EPI_ISL_1445928, EPI_ISL_1445943, EPI_ISL_1446075, EPI_ISL_1446114, EPI_ISL_1446139, EPI_ISL_1446140, EPI_ISL_1446144, EPI_ISL_1446148, EPI_ISL_1446152, EPI_ISL_1446190 | | | |
| see above | Aegis Sciences Corporation | Centers for Disease Control and Prevention Division of Viral Diseases, Pathogen Discovery | Dakota Howard, Dhwani Batra, Peter W. Cook, Kara Moser, Adrian Paskey, Jason Caravas, Benjamin Rambo-Martin, Shatavia Morrison, Christopher Gulvick, Scott Sammons, Yvette Unoarumhi, Darlene Wagner, Matthew Schmerer, Cyndi Clark, Patrick Campbell, Rob Case, Vikramsinha Ghorpade, Holly Houdeshell, Ola Kvalvaag, Dillon Nall, Ethan Sanders, Alec Vest, Shaun Westlund, Matthew Hardison, Clinton R. Paden, Duncan MacCannell |
| EPI_ISL_1446676, EPI_ISL_1446677 | "NYSDOH Wadsworth Center, Virology Lab" | Centers for Disease Control and Prevention Division of Viral Diseases, Pathogen Discovery | Mili Sheth, Sarah Nobles, Jasmine Padilla, Mark Burroughs, Shoshona Le, Katie Dillon, Peter Cook, Clinton R. Paden, Dhwani Batra, Krista Queen, Kristen Knipe, Dakota Howard, Yvette Unoarumhi, Darlene Wagner, Matthew Schmerer, Ben L. Rambo-Martin, Kristine Lacek, Sam Shepard, Alison Laufer Halpin, Dave Wentworth, Vivien Dugan, Suxiang Tong, Justin Lee |
| EPI_ISL_1446974, EPI_ISL_1446975 | NYC Department of Health and Mental Hygiene | Centers for Disease Control and Prevention Division of Viral Diseases, Pathogen Discovery | Mili Sheth, Sarah Nobles, Jasmine Padilla, Mark Burroughs, Shoshona Le, Katie Dillon, Peter Cook, Clinton R. Paden, Dhwani Batra, Krista Queen, Kristen Knipe, Dakota Howard, Yvette Unoarumhi, Darlene Wagner, Matthew Schmerer, Ben L. Rambo-Martin, Kristine Lacek, Sam Shepard, Alison Laufer Halpin, Dave Wentworth, Vivien Dugan, Suxiang Tong, Justin Lee |
| EPI_ISL_1447053 | "NYSDOH Wadsworth Center, Virology Lab" | Centers for Disease Control and Prevention Division of Viral Diseases, Pathogen Discovery | Mili Sheth, Sarah Nobles, Jasmine Padilla, Mark Burroughs, Shoshona Le, Katie Dillon, Peter Cook, Clinton R. Paden, Dhwani Batra, Krista Queen, Kristen Knipe, Dakota Howard, Yvette Unoarumhi, Darlene Wagner, Matthew Schmerer, Ben L. Rambo-Martin, Kristine Lacek, Sam Shepard, Alison Laufer Halpin, Dave Wentworth, Vivien Dugan, Suxiang Tong, Justin Lee |
| EPI_ISL_1447060, EPI_ISL_1447061, EPI_ISL_1447062 | NYC Department of Health and Mental Hygiene | Centers for Disease Control and Prevention Division of Viral Diseases, Pathogen Discovery | Mili Sheth, Sarah Nobles, Jasmine Padilla, Mark Burroughs, Shoshona Le, Katie Dillon, Peter Cook, Clinton R. Paden, Dhwani Batra, Krista Queen, Kristen Knipe, Dakota Howard, Yvette Unoarumhi, Darlene Wagner, Matthew Schmerer, Ben L. Rambo-Martin, Kristine Lacek, Sam Shepard, Alison Laufer Halpin, Dave Wentworth, Vivien Dugan, Suxiang Tong, Justin Lee |
| EPI_ISL_1447087 | DOHMH Morrisania | New York City Public Health Laboratory | Jade Wang, et al. |
| EPI_ISL_1447088 | DOHMH Corona | New York City Public Health Laboratory | Jade Wang, et al. |
| EPI_ISL_1447089 | DOHMH Crown Heights | New York City Public Health Laboratory | Jade Wang, et al. |
| EPI_ISL_1447090 | DOHMH Morrisania | New York City Public Health Laboratory | Jade Wang, et al. |
| EPI_ISL_1447091 | OCME Office Of Chief Medical Examiner | New York City Public Health Laboratory | Jade Wang, et al. |
| EPI_ISL_1447092 | DOHMH Jamaica | New York City Public Health Laboratory | Jade Wang, et al. |
| EPI_ISL_1447093 | DOHMH Chelsea | New York City Public Health Laboratory | Jade Wang, et al. |
| EPI_ISL_1447094 | DOHMH Jamaica | New York City Public Health Laboratory | Jade Wang, et al. |
| EPI_ISL_1447095 | DOHMH Corona | New York City Public Health Laboratory | Jade Wang, et al. |
| EPI_ISL_1447096 | DOHMH Chelsea | New York City Public Health Laboratory | Jade Wang, et al. |
| EPI_ISL_1447097 | DOHMH Morrisania | New York City Public Health Laboratory | Jade Wang, et al. |
| EPI_ISL_1447098 | DOHMH Fort Greene | New York City Public Health Laboratory | Jade Wang, et al. |
| EPI_ISL_1447099 | DOHMH PHL | New York City Public Health Laboratory | Jade Wang, et al. |
| EPI_ISL_1447100 | OCME Office Of Chief Medical Examiner | New York City Public Health Laboratory | Jade Wang, et al. |

| | | | |
|---|---|---|---|
| EPI_ISL_1447101 | DOHMH Central Harlem | New York City Public Health Laboratory | Jade Wang, et al. |
| EPI_ISL_1447102 | Department of Homeless Services | New York City Public Health Laboratory | Jade Wang, et al. |
| EPI_ISL_1447103 | DOHMH Jamaica | New York City Public Health Laboratory | Jade Wang, et al. |
| EPI_ISL_1447104 | OCME Office Of Chief Medical Examiner | New York City Public Health Laboratory | Jade Wang, et al. |
| EPI_ISL_1447105 | DOHMH Riverside | New York City Public Health Laboratory | Jade Wang, et al. |
| EPI_ISL_1447106 | DOHMH Jamaica | New York City Public Health Laboratory | Jade Wang, et al. |
| EPI_ISL_1447107 | DOHMH PHL | New York City Public Health Laboratory | Jade Wang, et al. |
| EPI_ISL_1447108 | DOHMH Morrisania | New York City Public Health Laboratory | Jade Wang, et al. |
| EPI_ISL_1447109 | DOHMH Central Harlem | New York City Public Health Laboratory | Jade Wang, et al. |
| EPI_ISL_1447110 | DOHMH Morrisania | New York City Public Health Laboratory | Jade Wang, et al. |
| EPI_ISL_1447111 | OCME Office Of Chief Medical Examiner | New York City Public Health Laboratory | Jade Wang, et al. |
| EPI_ISL_1447112 | DOHMH Fort Greene | New York City Public Health Laboratory | Jade Wang, et al. |
| EPI_ISL_1447113 | DOHMH Crown Heights | New York City Public Health Laboratory | Jade Wang, et al. |
| EPI_ISL_1447114 | OCME Office Of Chief Medical Examiner | New York City Public Health Laboratory | Jade Wang, et al. |
| EPI_ISL_1447115 | DOHMH Fort Greene | New York City Public Health Laboratory | Jade Wang, et al. |
| EPI_ISL_1447116 | DOHMH Central Harlem | New York City Public Health Laboratory | Jade Wang, et al. |
| EPI_ISL_1447117 | DOHMH Jamaica | New York City Public Health Laboratory | Jade Wang, et al. |
| EPI_ISL_1447118 | DOHMH Chelsea | New York City Public Health Laboratory | Jade Wang, et al. |
| EPI_ISL_1447119 | OCME Office Of Chief Medical Examiner | New York City Public Health Laboratory | Jade Wang, et al. |
| EPI_ISL_1447120, EPI_ISL_1447121, EPI_ISL_1447122 | DOHMH Jamaica | New York City Public Health Laboratory | Jade Wang, et al. |
| EPI_ISL_1447123 | DOHMH Chelsea | New York City Public Health Laboratory | Jade Wang, et al. |
| EPI_ISL_1447124, EPI_ISL_1447125 | DOHMH Jamaica | New York City Public Health Laboratory | Jade Wang, et al. |
| EPI_ISL_1447126 | DOHMH Corona | New York City Public Health Laboratory | Jade Wang, et al. |
| EPI_ISL_1447127 | DOHMH Fort Greene | New York City Public Health Laboratory | Jade Wang, et al. |
| EPI_ISL_1447128 | DOHMH Crown Heights | New York City Public Health Laboratory | Jade Wang, et al. |
| EPI_ISL_1447129, EPI_ISL_1447130 | DOHMH Central Harlem | New York City Public Health Laboratory | Jade Wang, et al. |
| EPI_ISL_1447131, EPI_ISL_1447132 | DOHMH Jamaica | New York City Public Health Laboratory | Jade Wang, et al. |
| EPI_ISL_1447133 | DOHMH Chelsea | New York City Public Health Laboratory | Jade Wang, et al. |
| EPI_ISL_1447134 | DOHMH Central Harlem | New York City Public Health Laboratory | Jade Wang, et al. |
| EPI_ISL_1447135 | DOHMH Fort Greene | New York City Public Health Laboratory | Jade Wang, et al. |
| EPI_ISL_1447136, EPI_ISL_1447137 | DOHMH Corona | New York City Public Health Laboratory | Jade Wang, et al. |
| EPI_ISL_1447138 | OCME Office Of Chief Medical Examiner | New York City Public Health Laboratory | Jade Wang, et al. |
| EPI_ISL_1447139, EPI_ISL_1447140, EPI_ISL_1447141, EPI_ISL_1447142 | DOHMH Corona | New York City Public Health Laboratory | Jade Wang, et al. |
| EPI_ISL_1447143 | Rockefeller University Hospital | New York City Public Health Laboratory | Jade Wang, et al. |
| EPI_ISL_1447144 | DOHMH Corona | New York City Public Health Laboratory | Jade Wang, et al. |
| EPI_ISL_1447145 | DOHMH PHL | New York City Public Health Laboratory | Jade Wang, et al. |
| EPI_ISL_1447146 | DOHMH Jamaica | New York City Public Health Laboratory | Jade Wang, et al. |
| EPI_ISL_1447147 | DOHMH Corona | New York City Public Health Laboratory | Jade Wang, et al. |
| EPI_ISL_1447148 | DOHMH Crown Heights | New York City Public Health Laboratory | Jade Wang, et al. |
| EPI_ISL_1447149, EPI_ISL_1447150 | DOHMH PHL | New York City Public Health Laboratory | Jade Wang, et al. |
| EPI_ISL_1447151, EPI_ISL_1447152, EPI_ISL_1447153 | DOHMH Jamaica | New York City Public Health Laboratory | Jade Wang, et al. |
| EPI_ISL_1447154 | DOHMH Riverside | New York City Public Health Laboratory | Jade Wang, et al. |
| EPI_ISL_1447155 | DOHMH Corona | New York City Public Health Laboratory | Jade Wang, et al. |
| EPI_ISL_1447156, EPI_ISL_1447157 | DOHMH Chelsea | New York City Public Health Laboratory | Jade Wang, et al. |
| EPI_ISL_1447158, EPI_ISL_1447159 | DOHMH Jamaica | New York City Public Health Laboratory | Jade Wang, et al. |
| EPI_ISL_1447160 | DOHMH Corona | New York City Public Health Laboratory | Jade Wang, et al. |
| EPI_ISL_1447161, EPI_ISL_1447162 | DOHMH Crown Heights | New York City Public Health Laboratory | Jade Wang, et al. |
| EPI_ISL_1447163, EPI_ISL_1447164 | DOHMH Jamaica | New York City Public Health Laboratory | Jade Wang, et al. |
| EPI_ISL_1447165 | DOHMH Corona | New York City Public Health Laboratory | Jade Wang, et al. |
| EPI_ISL_1447166 | DOHMH Crown Heights | New York City Public Health Laboratory | Jade Wang, et al. |
| EPI_ISL_1447167, EPI_ISL_1447168 | DOHMH Central Harlem | New York City Public Health Laboratory | Jade Wang, et al. |
| EPI_ISL_1447169 | DOHMH Chelsea | New York City Public Health Laboratory | Jade Wang, et al. |
| EPI_ISL_1447170 | DOHMH Crown Heights | New York City Public Health Laboratory | Jade Wang, et al. |
| EPI_ISL_1447171, EPI_ISL_1447172 | DOHMH Corona | New York City Public Health Laboratory | Jade Wang, et al. |
| EPI_ISL_1447173 | DOHMH Riverside | New York City Public Health Laboratory | Jade Wang, et al. |
| EPI_ISL_1447174 | Department of Homeless Services | New York City Public Health Laboratory | Jade Wang, et al. |
| EPI_ISL_1447175 | DOHMH Jamaica | New York City Public Health Laboratory | Jade Wang, et al. |

| | | | |
|---|---|---|---|
| EPI_ISL_1447176 | DOHMH Fort Greene | New York City Public Health Laboratory | Jade Wang, et al. |
| EPI_ISL_1447177, EPI_ISL_1447178 | OCME Office Of Chief Medical Examiner | New York City Public Health Laboratory | Jade Wang, et al. |
| EPI_ISL_1447179, EPI_ISL_1447180 | DOHMH Jamaica | New York City Public Health Laboratory | Jade Wang, et al. |
| EPI_ISL_1447181 | DOHMH Morrisania | New York City Public Health Laboratory | Jade Wang, et al. |
| EPI_ISL_1447182 | Department of Homeless Services | New York City Public Health Laboratory | Jade Wang, et al. |
| EPI_ISL_1447183 | OCME Office Of Chief Medical Examiner | New York City Public Health Laboratory | Jade Wang, et al. |
| EPI_ISL_1447184 | DOHMH PHL | New York City Public Health Laboratory | Jade Wang, et al. |
| EPI_ISL_1447185 | DOHMH Corona | New York City Public Health Laboratory | Jade Wang, et al. |
| EPI_ISL_1447186 | DOHMH Central Harlem | New York City Public Health Laboratory | Jade Wang, et al. |
| EPI_ISL_1447187 | DOHMH Fort Greene | New York City Public Health Laboratory | Jade Wang, et al. |
| EPI_ISL_1447188, EPI_ISL_1447189, EPI_ISL_1447190 | DOHMH Jamaica | New York City Public Health Laboratory | Jade Wang, et al. |
| EPI_ISL_1447191 | DOHMH Crown Heights | New York City Public Health Laboratory | Jade Wang, et al. |
| EPI_ISL_1447192 | DOHMH Jamaica | New York City Public Health Laboratory | Jade Wang, et al. |
| EPI_ISL_1447193 | DOHMH Chelsea | New York City Public Health Laboratory | Jade Wang, et al. |
| EPI_ISL_1447572, EPI_ISL_1447577 | Yale Clinical Virology Lab | Grubaugh Lab - Yale School of Public Health | Joseph Fauver, Mallery Breban, Isabel Ott, Tara Alpert, Mary Petrone, Anderson Brito, Chantal Vogels, Annie Watkins, Chaney Kalinich, Jessica Rothman, Marie L. Landry, Nathan Grubaugh |
| EPI_ISL_1461290, EPI_ISL_1461291, EPI_ISL_1461292 | URMC LABS | Wadsworth Center, New York State Department of Health | Kirsten St. George, Daryl M. Lamson, Alexis Russell, Matthew Shudt, Melissa A Leisner, Jonathan Plitnick, Catharine Prussing, Navjot Singh, John Kelly, Erasmus Schneider, Erica Lasek-Nesselquist |
| EPI_ISL_1461293 | GLENS FALLS HOSPITAL LABORATORY | Wadsworth Center, New York State Department of Health | Kirsten St. George, Daryl M. Lamson, Alexis Russell, Matthew Shudt, Melissa A Leisner, Jonathan Plitnick, Catharine Prussing, Navjot Singh, John Kelly, Erasmus Schneider, Erica Lasek-Nesselquist |
| EPI_ISL_1461294 | SUNY UPSTATE MEDICAL UNIVERSITY | Wadsworth Center, New York State Department of Health | Kirsten St. George, Daryl M. Lamson, Alexis Russell, Matthew Shudt, Melissa A Leisner, Jonathan Plitnick, Catharine Prussing, Navjot Singh, John Kelly, Erasmus Schneider, Erica Lasek-Nesselquist |
| EPI_ISL_1461295, EPI_ISL_1461296, EPI_ISL_1461297, EPI_ISL_1461298, EPI_ISL_1461299 | ALBANY MEDICAL CENTER | Wadsworth Center, New York State Department of Health | Kirsten St. George, Daryl M. Lamson, Alexis Russell, Matthew Shudt, Melissa A Leisner, Jonathan Plitnick, Catharine Prussing, Navjot Singh, John Kelly, Erasmus Schneider, Erica Lasek-Nesselquist |
| EPI_ISL_1461300, EPI_ISL_1461301, EPI_ISL_1461302, EPI_ISL_1461303, EPI_ISL_1461304, EPI_ISL_1461305, EPI_ISL_1461306, EPI_ISL_1461307, EPI_ISL_1461308, EPI_ISL_1461309 | URMC LABS | Wadsworth Center, New York State Department of Health | Kirsten St. George, Daryl M. Lamson, Alexis Russell, Matthew Shudt, Melissa A Leisner, Jonathan Plitnick, Catharine Prussing, Navjot Singh, John Kelly, Erasmus Schneider, Erica Lasek-Nesselquist |
| EPI_ISL_1461310, EPI_ISL_1461311, EPI_ISL_1461312, EPI_ISL_1461313, EPI_ISL_1461314, EPI_ISL_1461315, EPI_ISL_1461316, EPI_ISL_1461317 | SUNY UPSTATE MEDICAL UNIVERSITY | Wadsworth Center, New York State Department of Health | Kirsten St. George, Daryl M. Lamson, Alexis Russell, Matthew Shudt, Melissa A Leisner, Jonathan Plitnick, Catharine Prussing, Navjot Singh, John Kelly, Erasmus Schneider, Erica Lasek-Nesselquist |
| EPI_ISL_1461318 | URMC LABS | Wadsworth Center, New York State Department of Health | Kirsten St. George, Daryl M. Lamson, Alexis Russell, Matthew Shudt, Melissa A Leisner, Jonathan Plitnick, Catharine Prussing, Navjot Singh, John Kelly, Erasmus Schneider, Erica Lasek-Nesselquist |
| EPI_ISL_1461319, EPI_ISL_1461320, EPI_ISL_1461321, EPI_ISL_1461323, EPI_ISL_1461324, EPI_ISL_1461325, EPI_ISL_1461326, EPI_ISL_1461327, EPI_ISL_1461328, EPI_ISL_1461329, EPI_ISL_1461330, EPI_ISL_1461331, EPI_ISL_1461332, EPI_ISL_1461333, EPI_ISL_1461334, EPI_ISL_1461335, EPI_ISL_1461336, EPI_ISL_1461337, EPI_ISL_1461338, EPI_ISL_1461339, EPI_ISL_1461340, EPI_ISL_1461341, EPI_ISL_1461342, EPI_ISL_1461343, EPI_ISL_1461344, EPI_ISL_1461345, EPI_ISL_1461346, EPI_ISL_1461347, EPI_ISL_1461348, EPI_ISL_1461349, EPI_ISL_1461350, EPI_ISL_1461351, EPI_ISL_1461352, EPI_ISL_1461353, EPI_ISL_1461354, EPI_ISL_1461355 | | | |
| see above | NYC Pandemic Response Lab | Wadsworth Center, New York State Department of Health | Kirsten St. George, Daryl M. Lamson, Alexis Russell, Matthew Shudt, Melissa A Leisner, Jonathan Plitnick, Catharine Prussing, Navjot Singh, John Kelly, Erasmus Schneider, Erica Lasek-Nesselquist |
| EPI_ISL_1461356, EPI_ISL_1461357, EPI_ISL_1461358, EPI_ISL_1461359, EPI_ISL_1461360, EPI_ISL_1461361, EPI_ISL_1461362, EPI_ISL_1461363, EPI_ISL_1461364, EPI_ISL_1461365, EPI_ISL_1461366, EPI_ISL_1461367, EPI_ISL_1461368, EPI_ISL_1461369, EPI_ISL_1461370, EPI_ISL_1461371, EPI_ISL_1461372, EPI_ISL_1461373, EPI_ISL_1461374, EPI_ISL_1461375, EPI_ISL_1461376, EPI_ISL_1461377 | | | |
| see above | TEMPUS LABS INC | Wadsworth Center, New York State Department of Health | Kirsten St. George, Daryl M. Lamson, Alexis Russell, Matthew Shudt, Melissa A Leisner, Jonathan Plitnick, Catharine Prussing, Navjot Singh, John Kelly, Erasmus Schneider, Erica Lasek-Nesselquist |
| EPI_ISL_1461826, EPI_ISL_1461842, EPI_ISL_1461983, EPI_ISL_1461996, EPI_ISL_1461997, EPI_ISL_1461998, EPI_ISL_1462005, EPI_ISL_1462238, EPI_ISL_1462239, EPI_ISL_1462242, EPI_ISL_1462357, EPI_ISL_1462358, EPI_ISL_1462359, EPI_ISL_1462360, EPI_ISL_1462361, EPI_ISL_1462362, EPI_ISL_1462363, EPI_ISL_1462371, EPI_ISL_1462372, EPI_ISL_1462375, EPI_ISL_1462376, EPI_ISL_1462377, EPI_ISL_1462425, EPI_ISL_1462426, EPI_ISL_1462427, EPI_ISL_1462428, EPI_ISL_1462429, EPI_ISL_1462430, EPI_ISL_1462431, EPI_ISL_1462432, EPI_ISL_1462433, EPI_ISL_1462434, EPI_ISL_1462440, EPI_ISL_1462441, EPI_ISL_1462473, EPI_ISL_1462474, EPI_ISL_1462475, EPI_ISL_1462478, EPI_ISL_1462479, EPI_ISL_1462480, EPI_ISL_1462483, EPI_ISL_1462484, EPI_ISL_1462485, EPI_ISL_1462497, EPI_ISL_1462498, EPI_ISL_1462499 | | | |
| see above | Laboratory Corporation of America | Centers for Disease Control and Prevention Division of Viral Diseases, Pathogen Discovery | Dakota Howard, Dhwani Batra, Peter W. Cook, Kara Moser, Adrian Paskey, Jason Caravas, Benjamin Rambo-Martin, Shatavia Morrison, Christopher Gulvick, Scott Sammons, Yvette Unoarumhi, Darlene Wagner, Matthew Schmerer, Minoo Agarwal, Eyad Almasri, Debbie Boles, Ayla Burns, Nuthawin Charoensri, Oren Cohen, Susan Countryman, Mary Ann Cristobal, Bobbi Croy, Suzanne Dale, Hrushikesh Deshmukh, Amanda Douglas, Vincent Drouillon, Marcia Eisenberg, Howard Engler, Rama Ghatti, Prashant Gupta, Susan Hicks, Jake Humphrey, Lax Iyer, Manoj Jain, Mohan Kolli, Brian Krueger, Tim Kuphal, Stanley Letovsky, Michael Levandoski, Craig Lukasik, Jonathan Meltzer, Brian Norvell, Mindy Nye, Scott Parker, Christos Petropoulos, John Pruitt, Steven Ragan, Scott Ryan, Mike Sapeta, Jana Schroth, Suresh Babu Selvaraju, Goran Stevovic, Amanda Suchanek, Andrea Throop, Lyndon Tilson, Thomas Urban, Joe Voshell, Kimberly Wagner, Jonathan Williams, Mary Williamson, Qian Zeng, Tricia Zwiefelhofer, Clinton R. Paden, Duncan MacCannell |
| EPI_ISL_1462500 | SUNY UPSTATE MEDICAL UNIVERSITY | Wadsworth Center, New York State Department of Health | Kirsten St. George, Daryl M. Lamson, Alexis Russell, Matthew Shudt, Melissa A Leisner, Jonathan Plitnick, Catharine Prussing, Navjot Singh, John Kelly, Erasmus Schneider, Erica Lasek-Nesselquist |
| EPI_ISL_1462501 | Laboratory Corporation of America | Centers for Disease Control and Prevention Division of Viral Diseases, Pathogen Discovery | Dakota Howard, Dhwani Batra, Peter W. Cook, Kara Moser, Adrian Paskey, Jason Caravas, Benjamin Rambo-Martin, Shatavia Morrison, Christopher Gulvick, Scott Sammons, Yvette Unoarumhi, Darlene Wagner, Matthew Schmerer, Minoo Agarwal, Eyad Almasri, Debbie Boles, Ayla Burns, Nuthawin Charoensri, Oren Cohen, Susan Countryman, Mary Ann Cristobal, Bobbi Croy, Suzanne Dale, Hrushikesh Deshmukh, Amanda Douglas, Vincent Drouillon, Marcia Eisenberg, Howard Engler, Rama Ghatti, Prashant Gupta, Susan Hicks, Jake Humphrey, Lax Iyer, Manoj Jain, Mohan Kolli, Brian Krueger, Tim Kuphal, Stanley Letovsky, Michael Levandoski, Craig Lukasik, Jonathan Meltzer, Brian Norvell, Mindy Nye, Scott Parker, Christos Petropoulos, John Pruitt, Steven Ragan, Scott Ryan, Mike Sapeta, Jana Schroth, Suresh Babu Selvaraju, Goran Stevovic, Amanda Suchanek, Andrea Throop, Lyndon Tilson, Thomas Urban, Joe Voshell, Kimberly Wagner, Jonathan Williams, Mary Williamson, Qian Zeng, Tricia Zwiefelhofer, Clinton R. Paden, Duncan MacCannell |
| EPI_ISL_1462502, EPI_ISL_1462503 | SUNY UPSTATE MEDICAL UNIVERSITY | Wadsworth Center, New York State Department of Health | Kirsten St. George, Daryl M. Lamson, Alexis Russell, Matthew Shudt, Melissa A Leisner, Jonathan Plitnick, Catharine Prussing, Navjot Singh, John Kelly, Erasmus Schneider, Erica Lasek-Nesselquist |
| EPI_ISL_1462504 | Laboratory Corporation of America | Centers for Disease Control and Prevention Division of Viral Diseases, Pathogen Discovery | Dakota Howard, Dhwani Batra, Peter W. Cook, Kara Moser, Adrian Paskey, Jason Caravas, Benjamin Rambo-Martin, Shatavia Morrison, Christopher Gulvick, Scott Sammons, Yvette Unoarumhi, Darlene Wagner, Matthew Schmerer, Minoo Agarwal, Eyad Almasri, Debbie Boles, Ayla Burns, Nuthawin Charoensri, Oren Cohen, Susan Countryman, Mary Ann Cristobal, Bobbi Croy, Suzanne Dale, Hrushikesh Deshmukh, Amanda Douglas, Vincent Drouillon, Marcia Eisenberg, Howard Engler, Rama Ghatti, Prashant Gupta, Susan Hicks, Jake Humphrey, Lax Iyer, Manoj Jain, Mohan Kolli, Brian Krueger, Tim Kuphal, Stanley Letovsky, Michael Levandoski, Craig Lukasik, Jonathan Meltzer, Brian Norvell, Mindy Nye, Scott Parker, Christos Petropoulos, John Pruitt, |

| | | | |
|---|---|---|---|
| | | | Steven Ragan, Scott Ryan, Mike Sapeta, Jana Schroth, Suresh Babu Selvaraju, Goran Stevovic, Amanda Suchanek, Andrea Throop, Lyndon Tilson, Thomas Urban, Joe Voshell, Kimberly Wagner, Jonathan Williams, Mary Williamson, Qian Zeng, Tricia Zwiefelhofer, Clinton R. Paden, Duncan MacCannell |
| EPI_ISL_1462505 | SUNY UPSTATE MEDICAL UNIVERSITY | Wadsworth Center, New York State Department of Health | Kirsten St. George, Daryl M. Lamson, Alexis Russell, Matthew Shudt, Melissa A Leisner, Jonathan Plitnick, Catharine Prussing, Navjot Singh, John Kelly, Erasmus Schneider, Erica Lasek-Nesselquist |
| EPI_ISL_1462506 | ALBANY MEDICAL CENTER | Wadsworth Center, New York State Department of Health | Kirsten St. George, Daryl M. Lamson, Alexis Russell, Matthew Shudt, Melissa A Leisner, Jonathan Plitnick, Catharine Prussing, Navjot Singh, John Kelly, Erasmus Schneider, Erica Lasek-Nesselquist |
| EPI_ISL_1462507 | Laboratory Corporation of America | Centers for Disease Control and Prevention Division of Viral Diseases, Pathogen Discovery | Dakota Howard, Dhwani Batra, Peter W. Cook, Kara Moser, Adrian Paskey, Jason Caravas, Benjamin Rambo-Martin, Shatavia Morrison, Christopher Gulvick, Scott Sammons, Yvette Unoarumhi, Darlene Wagner, Matthew Schmerer, Minoo Agarwal, Eyad Almasri, Debbie Boles, Ayla Burns, Nuthawin Charoensri, Oren Cohen, Susan Countryman, Mary Ann Cristobal, Bobbi Croy, Suzanne Dale, Hrushikesh Deshmukh, Amanda Douglas, Vincent Drouillon, Marcia Eisenberg, Howard Engler, Rama Ghatti, Prashant Gupta, Susan Hicks, Jake Humphrey, Lax Iyer, Manoj Jain, Mohan Kolli, Brian Krueger, Tim Kuphal, Stanley Letovsky, Michael Levandoski, Craig Lukasik, Jonathan Meltzer, Brian Norvell, Mindy Nye, Scott Parker, Christos Petropoulos, John Pruitt, Steven Ragan, Scott Ryan, Mike Sapeta, Jana Schroth, Suresh Babu Selvaraju, Goran Stevovic, Amanda Suchanek, Andrea Throop, Lyndon Tilson, Thomas Urban, Joe Voshell, Kimberly Wagner, Jonathan Williams, Mary Williamson, Qian Zeng, Tricia Zwiefelhofer, Clinton R. Paden, Duncan MacCannell |
| EPI_ISL_1462508 | ALBANY MEDICAL CENTER | Wadsworth Center, New York State Department of Health | Kirsten St. George, Daryl M. Lamson, Alexis Russell, Matthew Shudt, Melissa A Leisner, Jonathan Plitnick, Catharine Prussing, Navjot Singh, John Kelly, Erasmus Schneider, Erica Lasek-Nesselquist |
| EPI_ISL_1462509, EPI_ISL_1462510, EPI_ISL_1462512, EPI_ISL_1462513, EPI_ISL_1462515, EPI_ISL_1462516 | URMC LABS | Wadsworth Center, New York State Department of Health | Kirsten St. George, Daryl M. Lamson, Alexis Russell, Matthew Shudt, Melissa A Leisner, Jonathan Plitnick, Catharine Prussing, Navjot Singh, John Kelly, Erasmus Schneider, Erica Lasek-Nesselquist |
| EPI_ISL_1462517 | Laboratory Corporation of America | Centers for Disease Control and Prevention Division of Viral Diseases, Pathogen Discovery | Dakota Howard, Dhwani Batra, Peter W. Cook, Kara Moser, Adrian Paskey, Jason Caravas, Benjamin Rambo-Martin, Shatavia Morrison, Christopher Gulvick, Scott Sammons, Yvette Unoarumhi, Darlene Wagner, Matthew Schmerer, Minoo Agarwal, Eyad Almasri, Debbie Boles, Ayla Burns, Nuthawin Charoensri, Oren Cohen, Susan Countryman, Mary Ann Cristobal, Bobbi Croy, Suzanne Dale, Hrushikesh Deshmukh, Amanda Douglas, Vincent Drouillon, Marcia Eisenberg, Howard Engler, Rama Ghatti, Prashant Gupta, Susan Hicks, Jake Humphrey, Lax Iyer, Manoj Jain, Mohan Kolli, Brian Krueger, Tim Kuphal, Stanley Letovsky, Michael Levandoski, Craig Lukasik, Jonathan Meltzer, Brian Norvell, Mindy Nye, Scott Parker, Christos Petropoulos, John Pruitt, Steven Ragan, Scott Ryan, Mike Sapeta, Jana Schroth, Suresh Babu Selvaraju, Goran Stevovic, Amanda Suchanek, Andrea Throop, Lyndon Tilson, Thomas Urban, Joe Voshell, Kimberly Wagner, Jonathan Williams, Mary Williamson, Qian Zeng, Tricia Zwiefelhofer, Clinton R. Paden, Duncan MacCannell |
| EPI_ISL_1462518, EPI_ISL_1462519 | URMC LABS | Wadsworth Center, New York State Department of Health | Kirsten St. George, Daryl M. Lamson, Alexis Russell, Matthew Shudt, Melissa A Leisner, Jonathan Plitnick, Catharine Prussing, Navjot Singh, John Kelly, Erasmus Schneider, Erica Lasek-Nesselquist |
| EPI_ISL_1462520 | Laboratory Corporation of America | Centers for Disease Control and Prevention Division of Viral Diseases, Pathogen Discovery | Dakota Howard, Dhwani Batra, Peter W. Cook, Kara Moser, Adrian Paskey, Jason Caravas, Benjamin Rambo-Martin, Shatavia Morrison, Christopher Gulvick, Scott Sammons, Yvette Unoarumhi, Darlene Wagner, Matthew Schmerer, Minoo Agarwal, Eyad Almasri, Debbie Boles, Ayla Burns, Nuthawin Charoensri, Oren Cohen, Susan Countryman, Mary Ann Cristobal, Bobbi Croy, Suzanne Dale, Hrushikesh Deshmukh, Amanda Douglas, Vincent Drouillon, Marcia Eisenberg, Howard Engler, Rama Ghatti, Prashant Gupta, Susan Hicks, Jake Humphrey, Lax Iyer, Manoj Jain, Mohan Kolli, Brian Krueger, Tim Kuphal, Stanley Letovsky, Michael Levandoski, Craig Lukasik, Jonathan Meltzer, Brian Norvell, Mindy Nye, Scott Parker, Christos Petropoulos, John Pruitt, Steven Ragan, Scott Ryan, Mike Sapeta, Jana Schroth, Suresh Babu Selvaraju, Goran Stevovic, Amanda Suchanek, Andrea Throop, Lyndon Tilson, Thomas Urban, Joe Voshell, Kimberly Wagner, Jonathan Williams, Mary Williamson, Qian Zeng, Tricia Zwiefelhofer, Clinton R. Paden, Duncan MacCannell |
| EPI_ISL_1462521, EPI_ISL_1462523, EPI_ISL_1462524, EPI_ISL_1462525, EPI_ISL_1462527, EPI_ISL_1462528, EPI_ISL_1462530, EPI_ISL_1462531, EPI_ISL_1462533, EPI_ISL_1462534 | URMC LABS | Wadsworth Center, New York State Department of Health | Kirsten St. George, Daryl M. Lamson, Alexis Russell, Matthew Shudt, Melissa A Leisner, Jonathan Plitnick, Catharine Prussing, Navjot Singh, John Kelly, Erasmus Schneider, Erica Lasek-Nesselquist |
| EPI_ISL_1462536, EPI_ISL_1462537, EPI_ISL_1462539, EPI_ISL_1462540, EPI_ISL_1462542, EPI_ISL_1462543, EPI_ISL_1462545, EPI_ISL_1462546, EPI_ISL_1462548, EPI_ISL_1462549 | SUNY UPSTATE MEDICAL UNIVERSITY | Wadsworth Center, New York State Department of Health | Kirsten St. George, Daryl M. Lamson, Alexis Russell, Matthew Shudt, Melissa A Leisner, Jonathan Plitnick, Catharine Prussing, Navjot Singh, John Kelly, Erasmus Schneider, Erica Lasek-Nesselquist |
| EPI_ISL_1462551 | MONTEFIORE MEDICAL CENTER LABORATORIES | Wadsworth Center, New York State Department of Health | Kirsten St. George, Daryl M. Lamson, Alexis Russell, Matthew Shudt, Melissa A Leisner, Jonathan Plitnick, Catharine Prussing, Navjot Singh, John Kelly, Erasmus Schneider, Erica Lasek-Nesselquist |
| EPI_ISL_1462552 | TEMPUS LABS INC | Wadsworth Center, New York State Department of Health | Kirsten St. George, Daryl M. Lamson, Alexis Russell, Matthew Shudt, Melissa A Leisner, Jonathan Plitnick, Catharine Prussing, Navjot Singh, John Kelly, Erasmus Schneider, Erica Lasek-Nesselquist |
| EPI_ISL_1462553, EPI_ISL_1462555 | WESTCHESTER MEDICAL CENTER | Wadsworth Center, New York State Department of Health | Kirsten St. George, Daryl M. Lamson, Alexis Russell, Matthew Shudt, Melissa A Leisner, Jonathan Plitnick, Catharine Prussing, Navjot Singh, John Kelly, Erasmus Schneider, Erica Lasek-Nesselquist |
| EPI_ISL_1462556, EPI_ISL_1462558, EPI_ISL_1462559, EPI_ISL_1462561, EPI_ISL_1462563, EPI_ISL_1462564, EPI_ISL_1462566, EPI_ISL_1462567, EPI_ISL_1462569, EPI_ISL_1462570 | NORTHWELL HEALTH LABORATORIES | Wadsworth Center, New York State Department of Health | Kirsten St. George, Daryl M. Lamson, Alexis Russell, Matthew Shudt, Melissa A Leisner, Jonathan Plitnick, Catharine Prussing, Navjot Singh, John Kelly, Erasmus Schneider, Erica Lasek-Nesselquist |
| EPI_ISL_1462571 | Laboratory Corporation of America | Centers for Disease Control and Prevention Division of Viral Diseases, Pathogen Discovery | Dakota Howard, Dhwani Batra, Peter W. Cook, Kara Moser, Adrian Paskey, Jason Caravas, Benjamin Rambo-Martin, Shatavia Morrison, Christopher Gulvick, Scott Sammons, Yvette Unoarumhi, Darlene Wagner, Matthew Schmerer, Minoo Agarwal, Eyad Almasri, Debbie Boles, Ayla Burns, Nuthawin Charoensri, Oren Cohen, Susan Countryman, Mary Ann Cristobal, Bobbi Croy, Suzanne Dale, Hrushikesh Deshmukh, Amanda Douglas, Vincent Drouillon, Marcia Eisenberg, Howard Engler, Rama Ghatti, Prashant Gupta, Susan Hicks, Jake Humphrey, Lax Iyer, Manoj Jain, Mohan Kolli, Brian Krueger, Tim Kuphal, Stanley Letovsky, Michael Levandoski, Craig Lukasik, Jonathan Meltzer, Brian Norvell, Mindy Nye, Scott Parker, Christos Petropoulos, John Pruitt, Steven Ragan, Scott Ryan, Mike Sapeta, Jana Schroth, Suresh Babu Selvaraju, Goran Stevovic, Amanda Suchanek, Andrea Throop, Lyndon Tilson, Thomas Urban, Joe Voshell, Kimberly Wagner, Jonathan Williams, Mary Williamson, Qian Zeng, Tricia Zwiefelhofer, Clinton R. Paden, Duncan MacCannell |
| EPI_ISL_1462572, EPI_ISL_1462573, EPI_ISL_1462575 | THE MARY IMOGENE BASSETT HOSPITAL | Wadsworth Center, New York State Department of Health | Kirsten St. George, Daryl M. Lamson, Alexis Russell, Matthew Shudt, Melissa A Leisner, Jonathan Plitnick, Catharine Prussing, Navjot Singh, John Kelly, Erasmus Schneider, Erica Lasek-Nesselquist |
| EPI_ISL_1462576, EPI_ISL_1462578, EPI_ISL_1462579, EPI_ISL_1462581, EPI_ISL_1462582, EPI_ISL_1462584, EPI_ISL_1462585, EPI_ISL_1462587, EPI_ISL_1462588, EPI_ISL_1462589, EPI_ISL_1462591, EPI_ISL_1462593, EPI_ISL_1462594, EPI_ISL_1462596, EPI_ISL_1462597, EPI_ISL_1462599, EPI_ISL_1462600, EPI_ISL_1462601, EPI_ISL_1462603, EPI_ISL_1462604, EPI_ISL_1462606, EPI_ISL_1462607, EPI_ISL_1462609, EPI_ISL_1462610, EPI_ISL_1462612, EPI_ISL_1462613, EPI_ISL_1462615, EPI_ISL_1462617, EPI_ISL_1462618, EPI_ISL_1462620, EPI_ISL_1462621, EPI_ISL_1462623, EPI_ISL_1462624 | | | |
| see above | NORTHWELL HEALTH LABORATORIES | Wadsworth Center, New York State Department of Health | Kirsten St. George, Daryl M. Lamson, Alexis Russell, Matthew Shudt, Melissa A Leisner, Jonathan Plitnick, Catharine Prussing, Navjot Singh, John Kelly, Erasmus Schneider, Erica Lasek-Nesselquist |
| EPI_ISL_1462838, EPI_ISL_1463082, EPI_ISL_1463106, EPI_ISL_1463109, EPI_ISL_1463110, EPI_ISL_1463112, EPI_ISL_1463121, EPI_ISL_1463122, EPI_ISL_1463125, EPI_ISL_1463126, EPI_ISL_1463139, EPI_ISL_1463140, EPI_ISL_1463141, EPI_ISL_1463142, EPI_ISL_1463173, EPI_ISL_1463187, EPI_ISL_1463188, EPI_ISL_1463189, EPI_ISL_1463191, EPI_ISL_1463194, EPI_ISL_1463207, EPI_ISL_1463208, EPI_ISL_1463209, EPI_ISL_1463210, EPI_ISL_1463211, EPI_ISL_1463212, EPI_ISL_1463213, EPI_ISL_1463214, EPI_ISL_1463215, EPI_ISL_1463216, EPI_ISL_1463217, EPI_ISL_1463218, EPI_ISL_1463222, EPI_ISL_1463229, EPI_ISL_1463260, EPI_ISL_1463261, EPI_ISL_1463262, EPI_ISL_1463263, EPI_ISL_1463264, EPI_ISL_1463265, EPI_ISL_1463266, EPI_ISL_1463267, EPI_ISL_1463268, EPI_ISL_1463269, EPI_ISL_1463270, EPI_ISL_1463271, EPI_ISL_1463272, EPI_ISL_1463278, EPI_ISL_1463281, EPI_ISL_1463282, EPI_ISL_1463307, EPI_ISL_1463308, EPI_ISL_1463309, EPI_ISL_1463310, EPI_ISL_1463311, EPI_ISL_1463312, EPI_ISL_1463313, EPI_ISL_1463314, EPI_ISL_1463316, EPI_ISL_1463317, EPI_ISL_1463318, EPI_ISL_1463319, EPI_ISL_1463320, EPI_ISL_1463321, EPI_ISL_1463335, EPI_ISL_1463355, EPI_ISL_1463356, EPI_ISL_1463357, EPI_ISL_1463358, EPI_ISL_1463359, EPI_ISL_1463360, EPI_ISL_1463362, EPI_ISL_1463392, EPI_ISL_1463393, EPI_ISL_1463394, EPI_ISL_1463395, EPI_ISL_1463399, EPI_ISL_1463400 | | | |
| see above | Laboratory Corporation of America | Centers for Disease Control and Prevention Division of Viral Diseases, Pathogen Discovery | Dakota Howard, Dhwani Batra, Peter W. Cook, Kara Moser, Adrian Paskey, Jason Caravas, Benjamin Rambo-Martin, Shatavia Morrison, Christopher Gulvick, Scott Sammons, Yvette Unoarumhi, Darlene Wagner, Matthew Schmerer, Minoo Agarwal, Eyad Almasri, Debbie Boles, Ayla Burns, Nuthawin Charoensri, Oren Cohen, Susan Countryman, Mary Ann Cristobal, Bobbi Croy, Suzanne Dale, Hrushikesh Deshmukh, Amanda Douglas, Vincent Drouillon, Marcia Eisenberg, Howard Engler, Rama Ghatti, Prashant Gupta, Susan Hicks, Jake Humphrey, Lax Iyer, Manoj Jain, Mohan Kolli, Brian Krueger, Tim |

| | | | |
|---|---|---|---|
| | | | Kuphal, Stanley Letovsky, Michael Levandoski, Craig Lukasik, Jonathan Meltzer, Brian Norvell, Mindy Nye, Scott Parker, Christos Petropoulos, John Pruitt, Steven Ragan, Scott Ryan, Mike Sapeta, Jana Schroth, Suresh Babu Selvaraju, Goran Stevovic, Amanda Suchanek, Andrea Throop, Lyndon Tilson, Thomas Urban, Joe Voshell, Kimberly Wagner, Jonathan Williams, Mary Williamson, Qian Zeng, Tricia Zwiefelhofer, Clinton R. Paden, Duncan MacCannell |
| EPI_ISL_1465615, EPI_ISL_1465616 | ALBANY MEDICAL CENTER | Wadsworth Center, New York State Department of Health | Kirsten St. George, Daryl M. Lamson, Alexis Russell, Matthew Shudt, Melissa A Leisner, Jonathan Plitnick, Catharine Prussing, Navjot Singh, John Kelly, Erasmus Schneider, Erica Lasek-Nesselquist |
| EPI_ISL_1465617 | GLENS FALLS HOSPITAL LABORATORY | Wadsworth Center, New York State Department of Health | Kirsten St. George, Daryl M. Lamson, Alexis Russell, Matthew Shudt, Melissa A Leisner, Jonathan Plitnick, Catharine Prussing, Navjot Singh, John Kelly, Erasmus Schneider, Erica Lasek-Nesselquist |
| EPI_ISL_1465618, EPI_ISL_1465619, EPI_ISL_1465620, EPI_ISL_1465621 | URMC LABS | Wadsworth Center, New York State Department of Health | Kirsten St. George, Daryl M. Lamson, Alexis Russell, Matthew Shudt, Melissa A Leisner, Jonathan Plitnick, Catharine Prussing, Navjot Singh, John Kelly, Erasmus Schneider, Erica Lasek-Nesselquist |
| EPI_ISL_1465622 | WESTCHESTER MEDICAL CENTER | Wadsworth Center, New York State Department of Health | Kirsten St. George, Daryl M. Lamson, Alexis Russell, Matthew Shudt, Melissa A Leisner, Jonathan Plitnick, Catharine Prussing, Navjot Singh, John Kelly, Erasmus Schneider, Erica Lasek-Nesselquist |
| EPI_ISL_1465623, EPI_ISL_1465624, EPI_ISL_1465625, EPI_ISL_1465626, EPI_ISL_1465627, EPI_ISL_1465628, EPI_ISL_1465629, EPI_ISL_1465630, EPI_ISL_1465631, EPI_ISL_1465632 | URMC LABS | Wadsworth Center, New York State Department of Health | Kirsten St. George, Daryl M. Lamson, Alexis Russell, Matthew Shudt, Melissa A Leisner, Jonathan Plitnick, Catharine Prussing, Navjot Singh, John Kelly, Erasmus Schneider, Erica Lasek-Nesselquist |
| EPI_ISL_1465633 | Columbia University Irving Medical Center | Wadsworth Center, New York State Department of Health | Kirsten St. George, Daryl M. Lamson, Alexis Russell, Matthew Shudt, Melissa A Leisner, Jonathan Plitnick, Catharine Prussing, Navjot Singh, John Kelly, Erasmus Schneider, Erica Lasek-Nesselquist |
| EPI_ISL_1465634, EPI_ISL_1465635, EPI_ISL_1465636 | ALBANY MEDICAL CENTER | Wadsworth Center, New York State Department of Health | Kirsten St. George, Daryl M. Lamson, Alexis Russell, Matthew Shudt, Melissa A Leisner, Jonathan Plitnick, Catharine Prussing, Navjot Singh, John Kelly, Erasmus Schneider, Erica Lasek-Nesselquist |
| EPI_ISL_1465637 | WESTCHESTER MEDICAL CENTER | Wadsworth Center, New York State Department of Health | Kirsten St. George, Daryl M. Lamson, Alexis Russell, Matthew Shudt, Melissa A Leisner, Jonathan Plitnick, Catharine Prussing, Navjot Singh, John Kelly, Erasmus Schneider, Erica Lasek-Nesselquist |
| EPI_ISL_1465638 | SUNY UPSTATE MEDICAL UNIVERSITY | Wadsworth Center, New York State Department of Health | Kirsten St. George, Daryl M. Lamson, Alexis Russell, Matthew Shudt, Melissa A Leisner, Jonathan Plitnick, Catharine Prussing, Navjot Singh, John Kelly, Erasmus Schneider, Erica Lasek-Nesselquist |
| EPI_ISL_1465639 | ALBANY MEDICAL CENTER | Wadsworth Center, New York State Department of Health | Kirsten St. George, Daryl M. Lamson, Alexis Russell, Matthew Shudt, Melissa A Leisner, Jonathan Plitnick, Catharine Prussing, Navjot Singh, John Kelly, Erasmus Schneider, Erica Lasek-Nesselquist |
| EPI_ISL_1465640, EPI_ISL_1465641, EPI_ISL_1465642, EPI_ISL_1465643, EPI_ISL_1465644, EPI_ISL_1465645, EPI_ISL_1465646, EPI_ISL_1465647, EPI_ISL_1465648, EPI_ISL_1465649, EPI_ISL_1465650, EPI_ISL_1465651, EPI_ISL_1465652, EPI_ISL_1465653, EPI_ISL_1465654, EPI_ISL_1465655, EPI_ISL_1465656, EPI_ISL_1465657, EPI_ISL_1465658, EPI_ISL_1465659, EPI_ISL_1465660, EPI_ISL_1465661, EPI_ISL_1465662, EPI_ISL_1465663, EPI_ISL_1465664 | | | |
| see above | URMC LABS | Wadsworth Center, New York State Department of Health | Kirsten St. George, Daryl M. Lamson, Alexis Russell, Matthew Shudt, Melissa A Leisner, Jonathan Plitnick, Catharine Prussing, Navjot Singh, John Kelly, Erasmus Schneider, Erica Lasek-Nesselquist |
| EPI_ISL_1465665, EPI_ISL_1465666 | SUNY UPSTATE MEDICAL UNIVERSITY | Wadsworth Center, New York State Department of Health | Kirsten St. George, Daryl M. Lamson, Alexis Russell, Matthew Shudt, Melissa A Leisner, Jonathan Plitnick, Catharine Prussing, Navjot Singh, John Kelly, Erasmus Schneider, Erica Lasek-Nesselquist |
| EPI_ISL_1465667 | URMC LABS | Wadsworth Center, New York State Department of Health | Kirsten St. George, Daryl M. Lamson, Alexis Russell, Matthew Shudt, Melissa A Leisner, Jonathan Plitnick, Catharine Prussing, Navjot Singh, John Kelly, Erasmus Schneider, Erica Lasek-Nesselquist |
| EPI_ISL_1465668, EPI_ISL_1465669, EPI_ISL_1465670, EPI_ISL_1465671, EPI_ISL_1465672 | MONTEFIORE MEDICAL CENTER LABORATORIES | Wadsworth Center, New York State Department of Health | Kirsten St. George, Daryl M. Lamson, Alexis Russell, Matthew Shudt, Melissa A Leisner, Jonathan Plitnick, Catharine Prussing, Navjot Singh, John Kelly, Erasmus Schneider, Erica Lasek-Nesselquist |
| EPI_ISL_1465673, EPI_ISL_1465674, EPI_ISL_1465675, EPI_ISL_1465676, EPI_ISL_1465677 | THE MARY IMOGENE BASSETT HOSPITAL | Wadsworth Center, New York State Department of Health | Kirsten St. George, Daryl M. Lamson, Alexis Russell, Matthew Shudt, Melissa A Leisner, Jonathan Plitnick, Catharine Prussing, Navjot Singh, John Kelly, Erasmus Schneider, Erica Lasek-Nesselquist |
| EPI_ISL_1465678, EPI_ISL_1465679, EPI_ISL_1465680, EPI_ISL_1465681, EPI_ISL_1465682 | NORTHWELL HEALTH LABORATORIES | Wadsworth Center, New York State Department of Health | Kirsten St. George, Daryl M. Lamson, Alexis Russell, Matthew Shudt, Melissa A Leisner, Jonathan Plitnick, Catharine Prussing, Navjot Singh, John Kelly, Erasmus Schneider, Erica Lasek-Nesselquist |
| EPI_ISL_1465689, EPI_ISL_1465690, EPI_ISL_1465691, EPI_ISL_1465692 | SUNY UPSTATE MEDICAL UNIVERSITY | Wadsworth Center, New York State Department of Health | Kirsten St. George, Daryl M. Lamson, Alexis Russell, Matthew Shudt, Melissa A Leisner, Jonathan Plitnick, Catharine Prussing, Navjot Singh, John Kelly, Erasmus Schneider, Erica Lasek-Nesselquist |
| EPI_ISL_1465693, EPI_ISL_1465694, EPI_ISL_1465695 | Columbia University Irving Medical Center | Wadsworth Center, New York State Department of Health | Kirsten St. George, Daryl M. Lamson, Alexis Russell, Matthew Shudt, Melissa A Leisner, Jonathan Plitnick, Catharine Prussing, Navjot Singh, John Kelly, Erasmus Schneider, Erica Lasek-Nesselquist |
| EPI_ISL_1465696 | URMC LABS | Wadsworth Center, New York State Department of Health | Kirsten St. George, Daryl M. Lamson, Alexis Russell, Matthew Shudt, Melissa A Leisner, Jonathan Plitnick, Catharine Prussing, Navjot Singh, John Kelly, Erasmus Schneider, Erica Lasek-Nesselquist |
| EPI_ISL_1465697, EPI_ISL_1465698, EPI_ISL_1465699, EPI_ISL_1465700 | SUNY UPSTATE MEDICAL UNIVERSITY | Wadsworth Center, New York State Department of Health | Kirsten St. George, Daryl M. Lamson, Alexis Russell, Matthew Shudt, Melissa A Leisner, Jonathan Plitnick, Catharine Prussing, Navjot Singh, John Kelly, Erasmus Schneider, Erica Lasek-Nesselquist |
| EPI_ISL_1465701, EPI_ISL_1465702, EPI_ISL_1465703, EPI_ISL_1465704, EPI_ISL_1465705, EPI_ISL_1465706, EPI_ISL_1465707, EPI_ISL_1465708, EPI_ISL_1465709, EPI_ISL_1465710, EPI_ISL_1465711, EPI_ISL_1465712, EPI_ISL_1465713 | | | |
| see above | MONTEFIORE MEDICAL CENTER LABORATORIES | Wadsworth Center, New York State Department of Health | Kirsten St. George, Daryl M. Lamson, Alexis Russell, Matthew Shudt, Melissa A Leisner, Jonathan Plitnick, Catharine Prussing, Navjot Singh, John Kelly, Erasmus Schneider, Erica Lasek-Nesselquist |
| EPI_ISL_1465714, EPI_ISL_1465715, EPI_ISL_1465716, EPI_ISL_1465717, EPI_ISL_1465718, EPI_ISL_1465719, EPI_ISL_1465720, EPI_ISL_1465721, EPI_ISL_1465722 | NYC Pandemic Response Lab | Wadsworth Center, New York State Department of Health | Kirsten St. George, Daryl M. Lamson, Alexis Russell, Matthew Shudt, Melissa A Leisner, Jonathan Plitnick, Catharine Prussing, Navjot Singh, John Kelly, Erasmus Schneider, Erica Lasek-Nesselquist |
| EPI_ISL_1465723, EPI_ISL_1465724, EPI_ISL_1465725, EPI_ISL_1465726, EPI_ISL_1465727, EPI_ISL_1465728, EPI_ISL_1465729, EPI_ISL_1465730, EPI_ISL_1465731, EPI_ISL_1465732, EPI_ISL_1465733, EPI_ISL_1465734, EPI_ISL_1465735, EPI_ISL_1465736, EPI_ISL_1465737, EPI_ISL_1465738, EPI_ISL_1465739, EPI_ISL_1465740, EPI_ISL_1465741, EPI_ISL_1465742, EPI_ISL_1465743, EPI_ISL_1465744, EPI_ISL_1465745, EPI_ISL_1465746, EPI_ISL_1465747, EPI_ISL_1465748, EPI_ISL_1465749, EPI_ISL_1465750, EPI_ISL_1465751, EPI_ISL_1465752, EPI_ISL_1465753, EPI_ISL_1465754 | | | |
| see above | URMC LABS | Wadsworth Center, New York State Department of Health | Kirsten St. George, Daryl M. Lamson, Alexis Russell, Matthew Shudt, Melissa A Leisner, Jonathan Plitnick, Catharine Prussing, Navjot Singh, John Kelly, Erasmus Schneider, Erica Lasek-Nesselquist |
| EPI_ISL_1465755, EPI_ISL_1465756, EPI_ISL_1465757, EPI_ISL_1465758, EPI_ISL_1465759 | NORTHWELL HEALTH LABORATORIES | Wadsworth Center, New York State Department of Health | Kirsten St. George, Daryl M. Lamson, Alexis Russell, Matthew Shudt, Melissa A Leisner, Jonathan Plitnick, Catharine Prussing, Navjot Singh, John Kelly, Erasmus Schneider, Erica Lasek-Nesselquist |
| EPI_ISL_1465760 | THE MARY IMOGENE BASSETT HOSPITAL | Wadsworth Center, New York State Department of Health | Kirsten St. George, Daryl M. Lamson, Alexis Russell, Matthew Shudt, Melissa A Leisner, Jonathan Plitnick, Catharine Prussing, Navjot Singh, John Kelly, Erasmus Schneider, Erica Lasek-Nesselquist |
| EPI_ISL_1465761, EPI_ISL_1465762, EPI_ISL_1465763, EPI_ISL_1465764, EPI_ISL_1465765, EPI_ISL_1465766 | NORTHWELL HEALTH LABORATORIES | Wadsworth Center, New York State Department of Health | Kirsten St. George, Daryl M. Lamson, Alexis Russell, Matthew Shudt, Melissa A Leisner, Jonathan Plitnick, Catharine Prussing, Navjot Singh, John Kelly, Erasmus Schneider, Erica Lasek-Nesselquist |
| EPI_ISL_1465794, EPI_ISL_1465795, EPI_ISL_1465796, EPI_ISL_1465797, | MONTEFIORE MEDICAL CENTER LABORATORIES | Wadsworth Center, New York State Department of Health | Kirsten St. George, Daryl M. Lamson, Alexis Russell, Matthew Shudt, Melissa A Leisner, Jonathan Plitnick, Catharine Prussing, Navjot Singh, John Kelly, Erasmus Schneider, Erica Lasek-Nesselquist |

| | | | |
|---|---|---|---|
| EPI_ISL_1465798, EPI_ISL_1465799, EPI_ISL_1465800, EPI_ISL_1465801 | | | |
| EPI_ISL_1465802 | MONTEFIORE NEW ROCHELLE HOSPITAL LABORATORY | Wadsworth Center, New York State Department of Health | Kirsten St. George, Daryl M. Lamson, Alexis Russell, Matthew Shudt, Melissa A Leisner, Jonathan Plitnick, Catharine Prussing, Navjot Singh, John Kelly, Erasmus Schneider, Erica Lasek-Nesselquist |
| EPI_ISL_1465803, EPI_ISL_1465804 | Columbia University Irving Medical Center | Wadsworth Center, New York State Department of Health | Kirsten St. George, Daryl M. Lamson, Alexis Russell, Matthew Shudt, Melissa A Leisner, Jonathan Plitnick, Catharine Prussing, Navjot Singh, John Kelly, Erasmus Schneider, Erica Lasek-Nesselquist |
| EPI_ISL_1465805, EPI_ISL_1465806, EPI_ISL_1465807, EPI_ISL_1465808, EPI_ISL_1465809, EPI_ISL_1465810, EPI_ISL_1465811, EPI_ISL_1465812, EPI_ISL_1465813, EPI_ISL_1465814, EPI_ISL_1465815, EPI_ISL_1465816, EPI_ISL_1465817, EPI_ISL_1465818, EPI_ISL_1465819, EPI_ISL_1465820, EPI_ISL_1465821, EPI_ISL_1465822, EPI_ISL_1465823, EPI_ISL_1465824, EPI_ISL_1465825, EPI_ISL_1465826, EPI_ISL_1465827, EPI_ISL_1465828, EPI_ISL_1465829, EPI_ISL_1465830, EPI_ISL_1465831, EPI_ISL_1465832, EPI_ISL_1465833, EPI_ISL_1465834 | | | |
| see above | NYC Pandemic Response Lab | Wadsworth Center, New York State Department of Health | Kirsten St. George, Daryl M. Lamson, Alexis Russell, Matthew Shudt, Melissa A Leisner, Jonathan Plitnick, Catharine Prussing, Navjot Singh, John Kelly, Erasmus Schneider, Erica Lasek-Nesselquist |
| EPI_ISL_1465835, EPI_ISL_1465836, EPI_ISL_1465837 | NORTHWELL HEALTH LABORATORIES | Wadsworth Center, New York State Department of Health | Kirsten St. George, Daryl M. Lamson, Alexis Russell, Matthew Shudt, Melissa A Leisner, Jonathan Plitnick, Catharine Prussing, Navjot Singh, John Kelly, Erasmus Schneider, Erica Lasek-Nesselquist |
| EPI_ISL_1465838, EPI_ISL_1465839, EPI_ISL_1465840, EPI_ISL_1465841, EPI_ISL_1465842, EPI_ISL_1465843, EPI_ISL_1465844, EPI_ISL_1465845, EPI_ISL_1465846, EPI_ISL_1465847 | STONY BROOK UNIVERSITY HOSPITAL | Wadsworth Center, New York State Department of Health | Kirsten St. George, Daryl M. Lamson, Alexis Russell, Matthew Shudt, Melissa A Leisner, Jonathan Plitnick, Catharine Prussing, Navjot Singh, John Kelly, Erasmus Schneider, Erica Lasek-Nesselquist |
| EPI_ISL_1465848 | GOOD SAMARITAN HOSPITAL LABORATORY | Wadsworth Center, New York State Department of Health | Kirsten St. George, Daryl M. Lamson, Alexis Russell, Matthew Shudt, Melissa A Leisner, Jonathan Plitnick, Catharine Prussing, Navjot Singh, John Kelly, Erasmus Schneider, Erica Lasek-Nesselquist |
| EPI_ISL_1465849, EPI_ISL_1465850, EPI_ISL_1465851, EPI_ISL_1465852 | MONTEFIORE NEW ROCHELLE HOSPITAL LABORATORY | Wadsworth Center, New York State Department of Health | Kirsten St. George, Daryl M. Lamson, Alexis Russell, Matthew Shudt, Melissa A Leisner, Jonathan Plitnick, Catharine Prussing, Navjot Singh, John Kelly, Erasmus Schneider, Erica Lasek-Nesselquist |
| EPI_ISL_1465853, EPI_ISL_1465854, EPI_ISL_1465855, EPI_ISL_1465856 | GLENS FALLS HOSPITAL LABORATORY | Wadsworth Center, New York State Department of Health | Kirsten St. George, Daryl M. Lamson, Alexis Russell, Matthew Shudt, Melissa A Leisner, Jonathan Plitnick, Catharine Prussing, Navjot Singh, John Kelly, Erasmus Schneider, Erica Lasek-Nesselquist |
| EPI_ISL_1465857 | ITHACA COLLEGE LABORATORY HAMMOND HEALTH CENTER | Wadsworth Center, New York State Department of Health | Kirsten St. George, Daryl M. Lamson, Alexis Russell, Matthew Shudt, Melissa A Leisner, Jonathan Plitnick, Catharine Prussing, Navjot Singh, John Kelly, Erasmus Schneider, Erica Lasek-Nesselquist |
| EPI_ISL_1465858 | GOOD SAMARITAN HOSPITAL LABORATORY | Wadsworth Center, New York State Department of Health | Kirsten St. George, Daryl M. Lamson, Alexis Russell, Matthew Shudt, Melissa A Leisner, Jonathan Plitnick, Catharine Prussing, Navjot Singh, John Kelly, Erasmus Schneider, Erica Lasek-Nesselquist |
| EPI_ISL_1465859, EPI_ISL_1465860, EPI_ISL_1465861, EPI_ISL_1465862, EPI_ISL_1465863, EPI_ISL_1465864, EPI_ISL_1465865, EPI_ISL_1465866, EPI_ISL_1465867, EPI_ISL_1465868, EPI_ISL_1465869, EPI_ISL_1465870, EPI_ISL_1465871, EPI_ISL_1465872, EPI_ISL_1465873, EPI_ISL_1465874, EPI_ISL_1465875, EPI_ISL_1465876, EPI_ISL_1465877, EPI_ISL_1465878 | | | |
| see above | TEMPUS LABS INC | Wadsworth Center, New York State Department of Health | Kirsten St. George, Daryl M. Lamson, Alexis Russell, Matthew Shudt, Melissa A Leisner, Jonathan Plitnick, Catharine Prussing, Navjot Singh, John Kelly, Erasmus Schneider, Erica Lasek-Nesselquist |
| EPI_ISL_1470626 | The Jackson Laboratory | The Jackson Laboratory | Bergeron D, Renzette N, Adams M, Omerza G, Kelly K, Li L |
| EPI_ISL_1470741, EPI_ISL_1470742, EPI_ISL_1470743, EPI_ISL_1470744, EPI_ISL_1470745, EPI_ISL_1470746, EPI_ISL_1470747, EPI_ISL_1470748, EPI_ISL_1470749, EPI_ISL_1470750, EPI_ISL_1470751, EPI_ISL_1470752, EPI_ISL_1470753, EPI_ISL_1470754, EPI_ISL_1470755, EPI_ISL_1470756, EPI_ISL_1470757, EPI_ISL_1470758, EPI_ISL_1470759, EPI_ISL_1470760, EPI_ISL_1470761, EPI_ISL_1470762, EPI_ISL_1470763, EPI_ISL_1470764, EPI_ISL_1470765, EPI_ISL_1470766, EPI_ISL_1470767, EPI_ISL_1470768, EPI_ISL_1470769, EPI_ISL_1470770, EPI_ISL_1470771, EPI_ISL_1470772, EPI_ISL_1470773, EPI_ISL_1470774, EPI_ISL_1470775, EPI_ISL_1470776, EPI_ISL_1470777, EPI_ISL_1470778, EPI_ISL_1470779, EPI_ISL_1470780, EPI_ISL_1470781, EPI_ISL_1470782, EPI_ISL_1470783, EPI_ISL_1470784, EPI_ISL_1470785, EPI_ISL_1470786, EPI_ISL_1470787, EPI_ISL_1470788, EPI_ISL_1470789, EPI_ISL_1470790, EPI_ISL_1470791, EPI_ISL_1470792, EPI_ISL_1470793, EPI_ISL_1470794, EPI_ISL_1470795, EPI_ISL_1470796, EPI_ISL_1470797, EPI_ISL_1470798, EPI_ISL_1470799, EPI_ISL_1470800, EPI_ISL_1470801, EPI_ISL_1470802, EPI_ISL_1470803, EPI_ISL_1470804, EPI_ISL_1470805, EPI_ISL_1470806, EPI_ISL_1470807, EPI_ISL_1470808, EPI_ISL_1470809, EPI_ISL_1470810, EPI_ISL_1470811, EPI_ISL_1470812, EPI_ISL_1470813, EPI_ISL_1470814, EPI_ISL_1470815, EPI_ISL_1470816, EPI_ISL_1470817, EPI_ISL_1470818, EPI_ISL_1470819, EPI_ISL_1470820, EPI_ISL_1470821, EPI_ISL_1470822, EPI_ISL_1470823, EPI_ISL_1470824, EPI_ISL_1470825, EPI_ISL_1470826, EPI_ISL_1470827, EPI_ISL_1470828, EPI_ISL_1470829, EPI_ISL_1470830, EPI_ISL_1470831, EPI_ISL_1470832, EPI_ISL_1470833, EPI_ISL_1470834, EPI_ISL_1470835, EPI_ISL_1470836, EPI_ISL_1470837, EPI_ISL_1470838, EPI_ISL_1470839, EPI_ISL_1470840, EPI_ISL_1470841, EPI_ISL_1470842, EPI_ISL_1470843, EPI_ISL_1470844, EPI_ISL_1470845, EPI_ISL_1470846, EPI_ISL_1470847, EPI_ISL_1470848, EPI_ISL_1470849, EPI_ISL_1470850, EPI_ISL_1470851, EPI_ISL_1470852, EPI_ISL_1470853, EPI_ISL_1470854, EPI_ISL_1470855, EPI_ISL_1470856, EPI_ISL_1470857, EPI_ISL_1470858, EPI_ISL_1470859, EPI_ISL_1470860, EPI_ISL_1470861, EPI_ISL_1470862, EPI_ISL_1470863, EPI_ISL_1470864, EPI_ISL_1470865, EPI_ISL_1470866, EPI_ISL_1470867, EPI_ISL_1470868, EPI_ISL_1470869, EPI_ISL_1470870, EPI_ISL_1470871, EPI_ISL_1470872, EPI_ISL_1470873, EPI_ISL_1470874, EPI_ISL_1470875, EPI_ISL_1470876, EPI_ISL_1470877, EPI_ISL_1470878, EPI_ISL_1470879, EPI_ISL_1470880, EPI_ISL_1470881, EPI_ISL_1470882, EPI_ISL_1470883, EPI_ISL_1470884, EPI_ISL_1470885, EPI_ISL_1470886, EPI_ISL_1470887, EPI_ISL_1470888, EPI_ISL_1470889, EPI_ISL_1470890, EPI_ISL_1470891, EPI_ISL_1470892, EPI_ISL_1470893, EPI_ISL_1470894, EPI_ISL_1470895, EPI_ISL_1470896, EPI_ISL_1470897, EPI_ISL_1470898, EPI_ISL_1470899, EPI_ISL_1470900, EPI_ISL_1470901, EPI_ISL_1470902, EPI_ISL_1470903, EPI_ISL_1470904, EPI_ISL_1470905, EPI_ISL_1470906, EPI_ISL_1470907, EPI_ISL_1470908, EPI_ISL_1470909, EPI_ISL_1470910, EPI_ISL_1470911, EPI_ISL_1470912, EPI_ISL_1470913, EPI_ISL_1470914, EPI_ISL_1470915, EPI_ISL_1470916, EPI_ISL_1470917, EPI_ISL_1470918, EPI_ISL_1470919, EPI_ISL_1470920, EPI_ISL_1470921, EPI_ISL_1470922, EPI_ISL_1470923, EPI_ISL_1470924, EPI_ISL_1470925, EPI_ISL_1470926, EPI_ISL_1470927, EPI_ISL_1470928, EPI_ISL_1470929, EPI_ISL_1470930, EPI_ISL_1470931, EPI_ISL_1470932, EPI_ISL_1470933, EPI_ISL_1470934, EPI_ISL_1470935, EPI_ISL_1470936, EPI_ISL_1470937, EPI_ISL_1470938, EPI_ISL_1470939, EPI_ISL_1470940, EPI_ISL_1470941, EPI_ISL_1470942, EPI_ISL_1470943, EPI_ISL_1470944, EPI_ISL_1470945, EPI_ISL_1470946, EPI_ISL_1470947, EPI_ISL_1470948, EPI_ISL_1470949, EPI_ISL_1470950, EPI_ISL_1470951, EPI_ISL_1470952, EPI_ISL_1470953, EPI_ISL_1470954, EPI_ISL_1470955, EPI_ISL_1470956, EPI_ISL_1470957, EPI_ISL_1470958, EPI_ISL_1470959, EPI_ISL_1470960, EPI_ISL_1470961, EPI_ISL_1470962, EPI_ISL_1470963, EPI_ISL_1470964, EPI_ISL_1470965, EPI_ISL_1470966, EPI_ISL_1470967, EPI_ISL_1470968, EPI_ISL_1470969, EPI_ISL_1470970, EPI_ISL_1470971, EPI_ISL_1470972, EPI_ISL_1470973, EPI_ISL_1470974, EPI_ISL_1470975, EPI_ISL_1470976, EPI_ISL_1470977, EPI_ISL_1470978, EPI_ISL_1470979, EPI_ISL_1470980, EPI_ISL_1470981, EPI_ISL_1470982, EPI_ISL_1470983, EPI_ISL_1470984, EPI_ISL_1470985, EPI_ISL_1470986, EPI_ISL_1470987, EPI_ISL_1470988, EPI_ISL_1470989, EPI_ISL_1470990, EPI_ISL_1470991, EPI_ISL_1470992, EPI_ISL_1470993, EPI_ISL_1470994, EPI_ISL_1470995, EPI_ISL_1470996, EPI_ISL_1470997, EPI_ISL_1470998, EPI_ISL_1470999, EPI_ISL_1471000, EPI_ISL_1471001, EPI_ISL_1471002, EPI_ISL_1471003, EPI_ISL_1471004, EPI_ISL_1471005, EPI_ISL_1471006, EPI_ISL_1471007, EPI_ISL_1471008, EPI_ISL_1471009, EPI_ISL_1471010, EPI_ISL_1471011, EPI_ISL_1471012, EPI_ISL_1471013, EPI_ISL_1471014, EPI_ISL_1471015, EPI_ISL_1471016, EPI_ISL_1471017, EPI_ISL_1471018, EPI_ISL_1471019, EPI_ISL_1471020, EPI_ISL_1471021, EPI_ISL_1471022, EPI_ISL_1471023, EPI_ISL_1471024, EPI_ISL_1471025, EPI_ISL_1471026, EPI_ISL_1471027, EPI_ISL_1471028, EPI_ISL_1471029, EPI_ISL_1471030, EPI_ISL_1471031, EPI_ISL_1471032, EPI_ISL_1471033, EPI_ISL_1471034, EPI_ISL_1471035, EPI_ISL_1471036, EPI_ISL_1471037, EPI_ISL_1471038, EPI_ISL_1471039, EPI_ISL_1471040, EPI_ISL_1471041, EPI_ISL_1471042, EPI_ISL_1471043, EPI_ISL_1471044, EPI_ISL_1471045, EPI_ISL_1471046, EPI_ISL_1471047, EPI_ISL_1471048, EPI_ISL_1471049, EPI_ISL_1471050, EPI_ISL_1471051, EPI_ISL_1471052, EPI_ISL_1471053, EPI_ISL_1471054, EPI_ISL_1471055, EPI_ISL_1471056, EPI_ISL_1471057, EPI_ISL_1471058, EPI_ISL_1471059, EPI_ISL_1471060, EPI_ISL_1471061, EPI_ISL_1471062, EPI_ISL_1471063, EPI_ISL_1471064, EPI_ISL_1471065, EPI_ISL_1471066, EPI_ISL_1471067, EPI_ISL_1471068, EPI_ISL_1471069, EPI_ISL_1471070, EPI_ISL_1471071, EPI_ISL_1471072, EPI_ISL_1471073, EPI_ISL_1471074, EPI_ISL_1471075, EPI_ISL_1471076, EPI_ISL_1471077, EPI_ISL_1471078, EPI_ISL_1471079, EPI_ISL_1471080, EPI_ISL_1471081, EPI_ISL_1471082, EPI_ISL_1471083, EPI_ISL_1471084, EPI_ISL_1471085, EPI_ISL_1471086, EPI_ISL_1471087, EPI_ISL_1471088, EPI_ISL_1471089, EPI_ISL_1471090, EPI_ISL_1471091, EPI_ISL_1471092, EPI_ISL_1471093, EPI_ISL_1471094, EPI_ISL_1471095, EPI_ISL_1471096, EPI_ISL_1471097, EPI_ISL_1471098, EPI_ISL_1471099, EPI_ISL_1471100, EPI_ISL_1471101, EPI_ISL_1471102, EPI_ISL_1471103, EPI_ISL_1471104, EPI_ISL_1471105, EPI_ISL_1471106, EPI_ISL_1471107, EPI_ISL_1471108, EPI_ISL_1471109, EPI_ISL_1471110, EPI_ISL_1471111, EPI_ISL_1471112, EPI_ISL_1471113, EPI_ISL_1471114, EPI_ISL_1471115, EPI_ISL_1471116, EPI_ISL_1471117, EPI_ISL_1471118, EPI_ISL_1471119, EPI_ISL_1471120, EPI_ISL_1471121, EPI_ISL_1471122, EPI_ISL_1471123, EPI_ISL_1471124, EPI_ISL_1471125, EPI_ISL_1471126, EPI_ISL_1471127, EPI_ISL_1471128, EPI_ISL_1471129, EPI_ISL_1471130, EPI_ISL_1471131, EPI_ISL_1471132, EPI_ISL_1471133, EPI_ISL_1471134, EPI_ISL_1471135, EPI_ISL_1471136, EPI_ISL_1471137, EPI_ISL_1471138, EPI_ISL_1471139, EPI_ISL_1471140, EPI_ISL_1471141, EPI_ISL_1471142, EPI_ISL_1471143, EPI_ISL_1471144, EPI_ISL_1471145, EPI_ISL_1471146, EPI_ISL_1471147, EPI_ISL_1471148, EPI_ISL_1471149, EPI_ISL_1471150, EPI_ISL_1471151, EPI_ISL_1471152, EPI_ISL_1471153, EPI_ISL_1471154, EPI_ISL_1471155, EPI_ISL_1471156, EPI_ISL_1471157, EPI_ISL_1471158, EPI_ISL_1471159, EPI_ISL_1471160, EPI_ISL_1471161, EPI_ISL_1471162, EPI_ISL_1471163, EPI_ISL_1471164, EPI_ISL_1471165, EPI_ISL_1471166, EPI_ISL_1471167, EPI_ISL_1471168, EPI_ISL_1471169, EPI_ISL_1471170, EPI_ISL_1471171, EPI_ISL_1471172, EPI_ISL_1471173, EPI_ISL_1471174, EPI_ISL_1471175, EPI_ISL_1471176, EPI_ISL_1471177, EPI_ISL_1471178, EPI_ISL_1471179, EPI_ISL_1471180, EPI_ISL_1471181, EPI_ISL_1471182, EPI_ISL_1471183, EPI_ISL_1471184, EPI_ISL_1471185, EPI_ISL_1471186, EPI_ISL_1471187, EPI_ISL_1471188, EPI_ISL_1471189, EPI_ISL_1471190, EPI_ISL_1471191, EPI_ISL_1471192, EPI_ISL_1471193, EPI_ISL_1471194, EPI_ISL_1471195, EPI_ISL_1471196, EPI_ISL_1471197, EPI_ISL_1471198, EPI_ISL_1471199, EPI_ISL_1471200, EPI_ISL_1471201, EPI_ISL_1471202, EPI_ISL_1471203, EPI_ISL_1471204, EPI_ISL_1471205, EPI_ISL_1471206, EPI_ISL_1471207, EPI_ISL_1471208, EPI_ISL_1471209, EPI_ISL_1471210, EPI_ISL_1471211, EPI_ISL_1471212, EPI_ISL_1471213, EPI_ISL_1471214, EPI_ISL_1471215, EPI_ISL_1471216, EPI_ISL_1471217, EPI_ISL_1471218, EPI_ISL_1471219, EPI_ISL_1471220, EPI_ISL_1471221, EPI_ISL_1471222, EPI_ISL_1471223, EPI_ISL_1471224, EPI_ISL_1471225, EPI_ISL_1471226, EPI_ISL_1471227, EPI_ISL_1471228, EPI_ISL_1471229, EPI_ISL_1471230, EPI_ISL_1471231, EPI_ISL_1471232, EPI_ISL_1471233, EPI_ISL_1471234, EPI_ISL_1471235, EPI_ISL_1471236, EPI_ISL_1471237, EPI_ISL_1471238, EPI_ISL_1471239, EPI_ISL_1471240, EPI_ISL_1471241, EPI_ISL_1471242, EPI_ISL_1471243, EPI_ISL_1471244, EPI_ISL_1471245, EPI_ISL_1471246, EPI_ISL_1471247, EPI_ISL_1471248, EPI_ISL_1471249, EPI_ISL_1471250, EPI_ISL_1471251, EPI_ISL_1471252, EPI_ISL_1471253, EPI_ISL_1471254, EPI_ISL_1471255, EPI_ISL_1471256, EPI_ISL_1471257, EPI_ISL_1471258, EPI_ISL_1471259, EPI_ISL_1471260, EPI_ISL_1471261, EPI_ISL_1471262, EPI_ISL_1471263, EPI_ISL_1471264, EPI_ISL_1471265, EPI_ISL_1471266, EPI_ISL_1471267, EPI_ISL_1471268, EPI_ISL_1471269, EPI_ISL_1471270, EPI_ISL_1471271, EPI_ISL_1471272, EPI_ISL_1471273, EPI_ISL_1471274, EPI_ISL_1471275, EPI_ISL_1471276, EPI_ISL_1471277, EPI_ISL_1471278, EPI_ISL_1471279, EPI_ISL_1471280, EPI_ISL_1471281, EPI_ISL_1471282, EPI_ISL_1471283, EPI_ISL_1471284, EPI_ISL_1471285, EPI_ISL_1471286, EPI_ISL_1471287, EPI_ISL_1471288, EPI_ISL_1471289, EPI_ISL_1471290, EPI_ISL_1471291, EPI_ISL_1471292, EPI_ISL_1471293, EPI_ISL_1471294, EPI_ISL_1471295, EPI_ISL_1471296, EPI_ISL_1471297, EPI_ISL_1471298, EPI_ISL_1471299, EPI_ISL_1471300, EPI_ISL_1471301, EPI_ISL_1471302, EPI_ISL_1471303, EPI_ISL_1471304, EPI_ISL_1471305, EPI_ISL_1471306, EPI_ISL_1471307, EPI_ISL_1471308, EPI_ISL_1471309, EPI_ISL_1471310, EPI_ISL_1471311, EPI_ISL_1471312, EPI_ISL_1471313, EPI_ISL_1471314, EPI_ISL_1471315, EPI_ISL_1471316, EPI_ISL_1471317, EPI_ISL_1471318, EPI_ISL_1471319, EPI_ISL_1471320, EPI_ISL_1471321, EPI_ISL_1471322, EPI_ISL_1471323, EPI_ISL_1471324, EPI_ISL_1471325, EPI_ISL_1471326, EPI_ISL_1471327, EPI_ISL_1471328, EPI_ISL_1471329, EPI_ISL_1471330, EPI_ISL_1471331, EPI_ISL_1471332, EPI_ISL_1471333, EPI_ISL_1471334, EPI_ISL_1471335, EPI_ISL_1471336, EPI_ISL_1471337, EPI_ISL_1471338, EPI_ISL_1471339, EPI_ISL_1471340, EPI_ISL_1471341, EPI_ISL_1471342, EPI_ISL_1471343, EPI_ISL_1471344, EPI_ISL_1471345, EPI_ISL_1471346, EPI_ISL_1471347, EPI_ISL_1471348, EPI_ISL_1471349, EPI_ISL_1471350, EPI_ISL_1471351, EPI_ISL_1471352, EPI_ISL_1471353, EPI_ISL_1471354, EPI_ISL_1471355, EPI_ISL_1471356, EPI_ISL_1471357, EPI_ISL_1471358, EPI_ISL_1471359, EPI_ISL_1471360, EPI_ISL_1471361, EPI_ISL_1471362, EPI_ISL_1471363, EPI_ISL_1471364, EPI_ISL_1471365, EPI_ISL_1471366, EPI_ISL_1471367, EPI_ISL_1471368, EPI_ISL_1471369, EPI_ISL_1471370, EPI_ISL_1471371, EPI_ISL_1471372, EPI_ISL_1471373, EPI_ISL_1471374, EPI_ISL_1471375, EPI_ISL_1471376, EPI_ISL_1471377, EPI_ISL_1471378, EPI_ISL_1471379, EPI_ISL_1471380, EPI_ISL_1471381, EPI_ISL_1471382, EPI_ISL_1471383, EPI_ISL_1471384, EPI_ISL_1471385, EPI_ISL_1471386, EPI_ISL_1471387, EPI_ISL_1471388, EPI_ISL_1471389, EPI_ISL_1471390, EPI_ISL_1471391, EPI_ISL_1471392, EPI_ISL_1471393, EPI_ISL_1471394, EPI_ISL_1471395, EPI_ISL_1471396, EPI_ISL_1471397, EPI_ISL_1471398, EPI_ISL_1471399, EPI_ISL_1471400, EPI_ISL_1471401, EPI_ISL_1471402, EPI_ISL_1471403, EPI_ISL_1471404, EPI_ISL_1471405, EPI_ISL_1471406, EPI_ISL_1471407, EPI_ISL_1471408, EPI_ISL_1471409, EPI_ISL_1471410, EPI_ISL_1471411, EPI_ISL_1471412, EPI_ISL_1471413, EPI_ISL_1471414, EPI_ISL_1471415, EPI_ISL_1471416, EPI_ISL_1471417, EPI_ISL_1471418, EPI_ISL_1471419, EPI_ISL_1471420, EPI_ISL_1471421, EPI_ISL_1471422, EPI_ISL_1471423, EPI_ISL_1471424, EPI_ISL_1471425, EPI_ISL_1471426, EPI_ISL_1471427, EPI_ISL_1471428, EPI_ISL_1471429, EPI_ISL_1471430, EPI_ISL_1471431, EPI_ISL_1471432, EPI_ISL_1471433, EPI_ISL_1471434, EPI_ISL_1471435, EPI_ISL_1471436, EPI_ISL_1471437, EPI_ISL_1471438, EPI_ISL_1471439, EPI_ISL_1471440, EPI_ISL_1471441, EPI_ISL_1471442, EPI_ISL_1471443, EPI_ISL_1471444, EPI_ISL_1471445, EPI_ISL_1471446, EPI_ISL_1471447, EPI_ISL_1471448, EPI_ISL_1471449, EPI_ISL_1471450, EPI_ISL_1471451, EPI_ISL_1471452, EPI_ISL_1471453, EPI_ISL_1471454, EPI_ISL_1471455, EPI_ISL_1471456, EPI_ISL_1471457, EPI_ISL_1471458, EPI_ISL_1471459, EPI_ISL_1471460, EPI_ISL_1471461, EPI_ISL_1471462, EPI_ISL_1471463, EPI_ISL_1471464, EPI_ISL_1471465, EPI_ISL_1471466, EPI_ISL_1471467, EPI_ISL_1471468, EPI_ISL_1471469, EPI_ISL_1471470, EPI_ISL_1471471, EPI_ISL_1471472, EPI_ISL_1471473, EPI_ISL_1471474, EPI_ISL_1471475, EPI_ISL_1471476, EPI_ISL_1471477, EPI_ISL_1471478, EPI_ISL_1471479, EPI_ISL_1471480, EPI_ISL_1471481, EPI_ISL_1471482, EPI_ISL_1471483, EPI_ISL_1471484, EPI_ISL_1471485, EPI_ISL_1471486, EPI_ISL_1471487, EPI_ISL_1471488, EPI_ISL_1471489, EPI_ISL_1471490, EPI_ISL_1471491, EPI_ISL_1471492, EPI_ISL_1471493, EPI_ISL_1471494, EPI_ISL_1471495, EPI_ISL_1471496, EPI_ISL_1471497, EPI_ISL_1471498, EPI_ISL_1471499, EPI_ISL_1471500, EPI_ISL_1471501, EPI_ISL_1471502, EPI_ISL_1471503, EPI_ISL_1471504, EPI_ISL_1471505, EPI_ISL_1471506, EPI_ISL_1471507, EPI_ISL_1471508, EPI_ISL_1471509, EPI_ISL_1471510, EPI_ISL_1471511, EPI_ISL_1471512, EPI_ISL_1471513, EPI_ISL_1471514, EPI_ISL_1471515, EPI_ISL_1471516, EPI_ISL_1471517, EPI_ISL_1471518, EPI_ISL_1471519, EPI_ISL_1471520, EPI_ISL_1471521, EPI_ISL_1471522, EPI_ISL_1471523, EPI_ISL_1471524, EPI_ISL_1471525, EPI_ISL_1471526, EPI_ISL_1471527, EPI_ISL_1471528, EPI_ISL_1471529, EPI_ISL_1471530, EPI_ISL_1471531, EPI_ISL_1471532, EPI_ISL_1471533, EPI_ISL_1471534, EPI_ISL_1471535, EPI_ISL_1471536, EPI_ISL_1471537, EPI_ISL_1471538, EPI_ISL_1471539, EPI_ISL_1471540, EPI_ISL_1471541, EPI_ISL_1471542, EPI_ISL_1471543, EPI_ISL_1471544, EPI_ISL_1471545, EPI_ISL_1471546, EPI_ISL_1471547, EPI_ISL_1471548, EPI_ISL_1471549, EPI_ISL_1471550, EPI_ISL_1471551, EPI_ISL_1471552, EPI_ISL_1471553, EPI_ISL_1471554, EPI_ISL_1471555, EPI_ISL_1471556, | | | |

| | | | |
|---|---|---|---|
| EPI_ISL_1471557, EPI_ISL_1471558, EPI_ISL_1471559, EPI_ISL_1471560, EPI_ISL_1471561, EPI_ISL_1471562, EPI_ISL_1471563, EPI_ISL_1471564, EPI_ISL_1471565, EPI_ISL_1471566, EPI_ISL_1471567, EPI_ISL_1471568, EPI_ISL_1471569, EPI_ISL_1471570, EPI_ISL_1471571, EPI_ISL_1471572, EPI_ISL_1471573, EPI_ISL_1471574, EPI_ISL_1471575, EPI_ISL_1471576, EPI_ISL_1471577, EPI_ISL_1471578, EPI_ISL_1471579, EPI_ISL_1471580, EPI_ISL_1471581, EPI_ISL_1471582, EPI_ISL_1471583, EPI_ISL_1471584, EPI_ISL_1471585, EPI_ISL_1471586, EPI_ISL_1471587, EPI_ISL_1471588, EPI_ISL_1471589, EPI_ISL_1471590, EPI_ISL_1471591, EPI_ISL_1471592, EPI_ISL_1471593, EPI_ISL_1471594, EPI_ISL_1471595, EPI_ISL_1471596, EPI_ISL_1471597, EPI_ISL_1471598, EPI_ISL_1471599, EPI_ISL_1471600, EPI_ISL_1471601, EPI_ISL_1471602, EPI_ISL_1471603, EPI_ISL_1471604, EPI_ISL_1471605, EPI_ISL_1471606, EPI_ISL_1471607, EPI_ISL_1471608, EPI_ISL_1471609, EPI_ISL_1471610, EPI_ISL_1471611, EPI_ISL_1471612, EPI_ISL_1471613, EPI_ISL_1471614, EPI_ISL_1471615, EPI_ISL_1471616, EPI_ISL_1471617, EPI_ISL_1471618, EPI_ISL_1471619, EPI_ISL_1471620, EPI_ISL_1471621, EPI_ISL_1471622, EPI_ISL_1471623, EPI_ISL_1471624, EPI_ISL_1471625, EPI_ISL_1471626, EPI_ISL_1471627, EPI_ISL_1471628, EPI_ISL_1471629, EPI_ISL_1471630, EPI_ISL_1471631, EPI_ISL_1471632, EPI_ISL_1471633, EPI_ISL_1471634, EPI_ISL_1471635, EPI_ISL_1471636, EPI_ISL_1471637, EPI_ISL_1471638, EPI_ISL_1471639, EPI_ISL_1471640, EPI_ISL_1471641, EPI_ISL_1471642, EPI_ISL_1471643, EPI_ISL_1471644, EPI_ISL_1471645, EPI_ISL_1471646, EPI_ISL_1471647, EPI_ISL_1471648, EPI_ISL_1471649, EPI_ISL_1471650, EPI_ISL_1471651, EPI_ISL_1471652, EPI_ISL_1471653, EPI_ISL_1471654, EPI_ISL_1471655, EPI_ISL_1471656, EPI_ISL_1471657, EPI_ISL_1471658, EPI_ISL_1471659, EPI_ISL_1471660, EPI_ISL_1471661, EPI_ISL_1471662, EPI_ISL_1471663, EPI_ISL_1471664, EPI_ISL_1471665, EPI_ISL_1471666, EPI_ISL_1471667, EPI_ISL_1471668, EPI_ISL_1471669, EPI_ISL_1471670, EPI_ISL_1471671, EPI_ISL_1471672, EPI_ISL_1471673, EPI_ISL_1471674, EPI_ISL_1471675, EPI_ISL_1471676, EPI_ISL_1471677, EPI_ISL_1471678, EPI_ISL_1471679, EPI_ISL_1471680, EPI_ISL_1471681, EPI_ISL_1471682, EPI_ISL_1471683, EPI_ISL_1471684, EPI_ISL_1471685, EPI_ISL_1471686, EPI_ISL_1471687, EPI_ISL_1471688, EPI_ISL_1471689, EPI_ISL_1471690, EPI_ISL_1471691, EPI_ISL_1471692, EPI_ISL_1471693, EPI_ISL_1471694, EPI_ISL_1471695, EPI_ISL_1471696, EPI_ISL_1471697, EPI_ISL_1471698, EPI_ISL_1471699, EPI_ISL_1471700, EPI_ISL_1471701, EPI_ISL_1471702, EPI_ISL_1471703, EPI_ISL_1471704, EPI_ISL_1471705, EPI_ISL_1471706, EPI_ISL_1471707, EPI_ISL_1471708, EPI_ISL_1471709, EPI_ISL_1471710, EPI_ISL_1471711, EPI_ISL_1471712, EPI_ISL_1471713, EPI_ISL_1471714, EPI_ISL_1471715, EPI_ISL_1471716, EPI_ISL_1471717, EPI_ISL_1471718, EPI_ISL_1471719, EPI_ISL_1471720, EPI_ISL_1471721, EPI_ISL_1471722, EPI_ISL_1471723, EPI_ISL_1471724, EPI_ISL_1471725, EPI_ISL_1471726, EPI_ISL_1471727, EPI_ISL_1471728, EPI_ISL_1471729, EPI_ISL_1471730, EPI_ISL_1471731, EPI_ISL_1471732, EPI_ISL_1471733, EPI_ISL_1471734, EPI_ISL_1471735, EPI_ISL_1471736, EPI_ISL_1471737, EPI_ISL_1471738, EPI_ISL_1471739, EPI_ISL_1471740, EPI_ISL_1471741, EPI_ISL_1471742, EPI_ISL_1471743, EPI_ISL_1471744, EPI_ISL_1471745, EPI_ISL_1471746, EPI_ISL_1471747, EPI_ISL_1471748, EPI_ISL_1471749, EPI_ISL_1471750, EPI_ISL_1471751, EPI_ISL_1471752, EPI_ISL_1471753, EPI_ISL_1471754, EPI_ISL_1471755, EPI_ISL_1471756, EPI_ISL_1471757, EPI_ISL_1471758, EPI_ISL_1471759, EPI_ISL_1471760, EPI_ISL_1471761, EPI_ISL_1471762, EPI_ISL_1471763, EPI_ISL_1471764, EPI_ISL_1471765, EPI_ISL_1471766, EPI_ISL_1471767, EPI_ISL_1471768, EPI_ISL_1471769, EPI_ISL_1471770, EPI_ISL_1471771, EPI_ISL_1471772, EPI_ISL_1471773, EPI_ISL_1471774, EPI_ISL_1471775, EPI_ISL_1471776, EPI_ISL_1471777, EPI_ISL_1471778, EPI_ISL_1471779, EPI_ISL_1471780, EPI_ISL_1471781, EPI_ISL_1471782, EPI_ISL_1471783, EPI_ISL_1471784, EPI_ISL_1471785, EPI_ISL_1471786, EPI_ISL_1471787, EPI_ISL_1471788, EPI_ISL_1471789, EPI_ISL_1471790, EPI_ISL_1471791, EPI_ISL_1471792, EPI_ISL_1471793, EPI_ISL_1471794, EPI_ISL_1471795, EPI_ISL_1471796, EPI_ISL_1471797, EPI_ISL_1471798, EPI_ISL_1471799, EPI_ISL_1471800, EPI_ISL_1471801, EPI_ISL_1471802, EPI_ISL_1471803, EPI_ISL_1471804, EPI_ISL_1471805, EPI_ISL_1471806, EPI_ISL_1471807, EPI_ISL_1471808, EPI_ISL_1471809, EPI_ISL_1471810, EPI_ISL_1471811, EPI_ISL_1471812, EPI_ISL_1471813, EPI_ISL_1471814, EPI_ISL_1471815, EPI_ISL_1471816, EPI_ISL_1471817, EPI_ISL_1471818, EPI_ISL_1471819, EPI_ISL_1471820, EPI_ISL_1471821, EPI_ISL_1471822, EPI_ISL_1471823, EPI_ISL_1471824, EPI_ISL_1471825, EPI_ISL_1471826, EPI_ISL_1471827, EPI_ISL_1471828, EPI_ISL_1471829, EPI_ISL_1471830, EPI_ISL_1471831, EPI_ISL_1471832, EPI_ISL_1471833, EPI_ISL_1471834, EPI_ISL_1471835, EPI_ISL_1471836, EPI_ISL_1471837, EPI_ISL_1471838, EPI_ISL_1471839, EPI_ISL_1471840, EPI_ISL_1471841, EPI_ISL_1471842, EPI_ISL_1471843, EPI_ISL_1471844, EPI_ISL_1471845, EPI_ISL_1471846, EPI_ISL_1471847, EPI_ISL_1471848, EPI_ISL_1471849, EPI_ISL_1471850, EPI_ISL_1471851, EPI_ISL_1471852, EPI_ISL_1471853, EPI_ISL_1471854, EPI_ISL_1471855, EPI_ISL_1471856, EPI_ISL_1471857, EPI_ISL_1471858, EPI_ISL_1471859, EPI_ISL_1471860, EPI_ISL_1471861, EPI_ISL_1471862, EPI_ISL_1471863, EPI_ISL_1471864, EPI_ISL_1471865, EPI_ISL_1471866, EPI_ISL_1471867, EPI_ISL_1471868, EPI_ISL_1471869, EPI_ISL_1471870, EPI_ISL_1471871, EPI_ISL_1471872, EPI_ISL_1471873, EPI_ISL_1471874, EPI_ISL_1471875, EPI_ISL_1471876, EPI_ISL_1471877, EPI_ISL_1471878, EPI_ISL_1471879, EPI_ISL_1471880, EPI_ISL_1471881, EPI_ISL_1471882, EPI_ISL_1471883, EPI_ISL_1471884, EPI_ISL_1471885, EPI_ISL_1471886, EPI_ISL_1471887, EPI_ISL_1471888, EPI_ISL_1471889, EPI_ISL_1471890, EPI_ISL_1471891, EPI_ISL_1471892, EPI_ISL_1471893, EPI_ISL_1471894, EPI_ISL_1471895, EPI_ISL_1471896, EPI_ISL_1471897, EPI_ISL_1471898, EPI_ISL_1471899, EPI_ISL_1471900, EPI_ISL_1471901, EPI_ISL_1471902, EPI_ISL_1471903, EPI_ISL_1471904, EPI_ISL_1471905, EPI_ISL_1471906, EPI_ISL_1471907, EPI_ISL_1471908, EPI_ISL_1471909, EPI_ISL_1471910, EPI_ISL_1471911, EPI_ISL_1471912, EPI_ISL_1471913, EPI_ISL_1471914, EPI_ISL_1471915, EPI_ISL_1471916, EPI_ISL_1471917, EPI_ISL_1471918, EPI_ISL_1471919, EPI_ISL_1471920, EPI_ISL_1471921, EPI_ISL_1471922, EPI_ISL_1471923, EPI_ISL_1471924, EPI_ISL_1471925, EPI_ISL_1471926, EPI_ISL_1471927, EPI_ISL_1471928, EPI_ISL_1471929, EPI_ISL_1471930, EPI_ISL_1471931, EPI_ISL_1471932, EPI_ISL_1471933, EPI_ISL_1471934, EPI_ISL_1471935, EPI_ISL_1471936, EPI_ISL_1471937, EPI_ISL_1471938, EPI_ISL_1471939, EPI_ISL_1471940, EPI_ISL_1471941, EPI_ISL_1471942, EPI_ISL_1471943, EPI_ISL_1471944, EPI_ISL_1471945, EPI_ISL_1471946, EPI_ISL_1471947, EPI_ISL_1471948, EPI_ISL_1471949, EPI_ISL_1471950, EPI_ISL_1471951, EPI_ISL_1471952, EPI_ISL_1471953, EPI_ISL_1471954, EPI_ISL_1471955, EPI_ISL_1471956, EPI_ISL_1471957, EPI_ISL_1471958, EPI_ISL_1471959, EPI_ISL_1471960, EPI_ISL_1471961, EPI_ISL_1471962, EPI_ISL_1471963, EPI_ISL_1471964, EPI_ISL_1471965, EPI_ISL_1471966, EPI_ISL_1471967, EPI_ISL_1471968, EPI_ISL_1471969, EPI_ISL_1471970, EPI_ISL_1471971, EPI_ISL_1471972, EPI_ISL_1471973, EPI_ISL_1471974, EPI_ISL_1471975, EPI_ISL_1471976, EPI_ISL_1471977, EPI_ISL_1471978, EPI_ISL_1471979, EPI_ISL_1471980, EPI_ISL_1471981, EPI_ISL_1471982, EPI_ISL_1471983, EPI_ISL_1471984, EPI_ISL_1471985, EPI_ISL_1471986, EPI_ISL_1471987, EPI_ISL_1471988, EPI_ISL_1471989, EPI_ISL_1471990, EPI_ISL_1471991, EPI_ISL_1471992, EPI_ISL_1471993, EPI_ISL_1471994, EPI_ISL_1471995, EPI_ISL_1471996, EPI_ISL_1471997, EPI_ISL_1471998, EPI_ISL_1471999, EPI_ISL_1472000, EPI_ISL_1472001, EPI_ISL_1472002, EPI_ISL_1472003, EPI_ISL_1472004, EPI_ISL_1472005, EPI_ISL_1472006, EPI_ISL_1472007, EPI_ISL_1472008, EPI_ISL_1472009, EPI_ISL_1472010, EPI_ISL_1472011, EPI_ISL_1472012, EPI_ISL_1472013, EPI_ISL_1472014, EPI_ISL_1472015, EPI_ISL_1472016, EPI_ISL_1472017, EPI_ISL_1472018, EPI_ISL_1472019, EPI_ISL_1472020, EPI_ISL_1472021, EPI_ISL_1472022, EPI_ISL_1472023, EPI_ISL_1472024, EPI_ISL_1472025, EPI_ISL_1472026, EPI_ISL_1472027, EPI_ISL_1472028, EPI_ISL_1472029, EPI_ISL_1472030, EPI_ISL_1472031, EPI_ISL_1472032, EPI_ISL_1472033, EPI_ISL_1472034, EPI_ISL_1472035, EPI_ISL_1472036, EPI_ISL_1472037, EPI_ISL_1472038, EPI_ISL_1472039, EPI_ISL_1472040, EPI_ISL_1472041, EPI_ISL_1472042, EPI_ISL_1472043, EPI_ISL_1472044, EPI_ISL_1472045, EPI_ISL_1472046, EPI_ISL_1472047, EPI_ISL_1472048, EPI_ISL_1472049, EPI_ISL_1472050, EPI_ISL_1472051, EPI_ISL_1472052, EPI_ISL_1472053, EPI_ISL_1472054, EPI_ISL_1472055, EPI_ISL_1472056, EPI_ISL_1472057, EPI_ISL_1472058, EPI_ISL_1472059, EPI_ISL_1472060, EPI_ISL_1472061, EPI_ISL_1472062, EPI_ISL_1472063, EPI_ISL_1472064, EPI_ISL_1472065, EPI_ISL_1472066, EPI_ISL_1472067, EPI_ISL_1472068, EPI_ISL_1472069, EPI_ISL_1472070, EPI_ISL_1472071, EPI_ISL_1472072, EPI_ISL_1472073, EPI_ISL_1472074, EPI_ISL_1472075, EPI_ISL_1472076 | | | |
| see above | Pandemic Response Lab - NYC | Pandemic Response Lab, R&D | Henry Lee, Michael Hammerling, Melissa Hopkins, Cybill del Castillo, Shinyoung Clair Kang, William Ward, Pradeep Bugga, Sol Rey, Dylan Law, Haiping Hao, Jon Laurent |
| EPI_ISL_1479386 | Helix/Illumina | Centers for Disease Control and Prevention Division of Viral Diseases, Pathogen Discovery | Dakota Howard, Dhwani Batra, Peter W. Cook, Kara Moser, Adrian Paskey, Jason Caravas, Benjamin Rambo-Martin, Shatavia Morrison, Christopher Gulvick, Scott Sammons, Yvette Unoarumhi, Darlene Wagner, Matthew Schmerer, Eileen de Feo, Jan Antico, Christine Tran, Matthew Tolentino, Shannon Wickline, Kim Gietzen, Brad Sickler, Jingtao Liu, Eric Allen, Phil Febbo, Nicole L. Washington, Simon White, Geraint Levan, Kelly Schiabor Barrett, Elizabeth Cirulli, Alexandre Bolze, Ary Ascencio, Charlotte Rivera-Garcia, Ryan Cho, Jason Nguyen, Sherry Wang, Jimmy Ramirez, Tyler Cassens, Efren Sandoval, Magnus Isaksson, William Lee, David Becker, Marc Laurent, James Lu, Clinton R. Paden, Duncan MacCannell |
| EPI_ISL_1479449, EPI_ISL_1479456, EPI_ISL_1479460, EPI_ISL_1479461, EPI_ISL_1479477, EPI_ISL_1479549, EPI_ISL_1479604, EPI_ISL_1479823 | Aegis Sciences Corporation | Centers for Disease Control and Prevention Division of Viral Diseases, Pathogen Discovery | Dakota Howard, Dhwani Batra, Peter W. Cook, Kara Moser, Adrian Paskey, Jason Caravas, Benjamin Rambo-Martin, Shatavia Morrison, Christopher Gulvick, Scott Sammons, Yvette Unoarumhi, Darlene Wagner, Matthew Schmerer, Cyndi Clark, Patrick Campbell, Rob Case, Vikramsinha Ghorpade, Holly Houdeshell, Ola Kvalvaag, Dillon Nall, Ethan Sanders, Alec Vest, Shaun Westlund, Matthew Hardison, Clinton R. Paden, Duncan MacCannell |
| EPI_ISL_1479876, EPI_ISL_1479881, EPI_ISL_1479891, EPI_ISL_1479911 | Quest Diagnostics Incorporated | Centers for Disease Control and Prevention Division of Viral Diseases, Pathogen Discovery | Dakota Howard, Dhwani Batra, Peter W. Cook, Kara Moser, Adrian Paskey, Jason Caravas, Benjamin Rambo-Martin, Shatavia Morrison, Christopher Gulvick, Scott Sammons, Yvette Unoarumhi, Darlene Wagner, Matthew Schmerer, S. H. Rosenthal, A. Gerasimova, R. M. Kagan, B. Anderson, M. Hua, Y. Liu, L.E. Bernstein, K.E. Livingston, A. Perez, I. A. Shlyakhter, R. V. Rolando, R. Owen, P. Tanpaiboon, F. Lacbawan, Clinton R. Paden, Duncan MacCannell |
| EPI_ISL_1480025, EPI_ISL_1480043, EPI_ISL_1480059, EPI_ISL_1480155, EPI_ISL_1480290, EPI_ISL_1480429 | Helix/Illumina | Centers for Disease Control and Prevention Division of Viral Diseases, Pathogen Discovery | Dakota Howard, Dhwani Batra, Peter W. Cook, Kara Moser, Adrian Paskey, Jason Caravas, Benjamin Rambo-Martin, Shatavia Morrison, Christopher Gulvick, Scott Sammons, Yvette Unoarumhi, Darlene Wagner, Matthew Schmerer, Eileen de Feo, Jan Antico, Christine Tran, Matthew Tolentino, Shannon Wickline, Kim Gietzen, Brad Sickler, Jingtao Liu, Eric Allen, Phil Febbo, Nicole L. Washington, Simon White, Geraint Levan, Kelly Schiabor Barrett, Elizabeth Cirulli, Alexandre Bolze, Ary Ascencio, Charlotte Rivera-Garcia, Ryan Cho, Jason Nguyen, Sherry Wang, Jimmy Ramirez, Tyler Cassens, Efren Sandoval, Magnus Isaksson, William Lee, David Becker, Marc Laurent, James Lu, Clinton R. Paden, Duncan MacCannell |
| EPI_ISL_1480604, EPI_ISL_1480608, EPI_ISL_1480617, EPI_ISL_1480632, EPI_ISL_1480654, EPI_ISL_1480859, EPI_ISL_1480943, EPI_ISL_1480971, EPI_ISL_1480976 | Quest Diagnostics Incorporated | Centers for Disease Control and Prevention Division of Viral Diseases, Pathogen Discovery | Dakota Howard, Dhwani Batra, Peter W. Cook, Kara Moser, Adrian Paskey, Jason Caravas, Benjamin Rambo-Martin, Shatavia Morrison, Christopher Gulvick, Scott Sammons, Yvette Unoarumhi, Darlene Wagner, Matthew Schmerer, S. H. Rosenthal, A. Gerasimova, R. M. Kagan, B. Anderson, M. Hua, Y. Liu, L.E. Bernstein, K.E. Livingston, A. Perez, I. A. Shlyakhter, R. V. Rolando, R. Owen, P. Tanpaiboon, F. Lacbawan, Clinton R. Paden, Duncan MacCannell |
| EPI_ISL_1480977, EPI_ISL_1481057, EPI_ISL_1481058, EPI_ISL_1481061, EPI_ISL_1481067, EPI_ISL_1481105, EPI_ISL_1481110, EPI_ISL_1481111, EPI_ISL_1481132, EPI_ISL_1481136, EPI_ISL_1481137, EPI_ISL_1481138, EPI_ISL_1481154, EPI_ISL_1481182, EPI_ISL_1481187, EPI_ISL_1481373, EPI_ISL_1481388, EPI_ISL_1481392, EPI_ISL_1481396, EPI_ISL_1481398, EPI_ISL_1481400, EPI_ISL_1481404, EPI_ISL_1481405, EPI_ISL_1481409, EPI_ISL_1481429, EPI_ISL_1481450, EPI_ISL_1481451, EPI_ISL_1481475, EPI_ISL_1481480, EPI_ISL_1481481 | | | |
| see above | Laboratory Corporation of America | Centers for Disease Control and Prevention Division of Viral Diseases, Pathogen Discovery | Dakota Howard, Dhwani Batra, Peter W. Cook, Kara Moser, Adrian Paskey, Jason Caravas, Benjamin Rambo-Martin, Shatavia Morrison, Christopher Gulvick, Scott Sammons, Yvette Unoarumhi, Darlene Wagner, Matthew Schmerer, Minoo Agarwal, Eyad Almasri, Debbie Boles, Ayla Burns, Nuthawin Charoensri, Oren Cohen, Susan Countryman, Mary Ann Cristobal, Bobbi Croy, Suzanne Dale, Hrushikesh Deshmukh, Amanda Douglas, Vincent Drouillon, Marcia Eisenberg, Howard Engler, Rama Ghatti, Prashant Gupta, Susan Hicks, Jake Humphrey, Lax Iyer, Manoj Jain, Mohan Kolli, Brian Krueger, Tim Kuphal, Stanley Letovsky, Michael Levandoski, Craig Lukasik, Jonathan Meltzer, Brian Norvell, Mindy Nye, Scott Parker, Christos Petropoulos, John Pruitt, Steven Ragan, Scott Ryan, Mike Sapeta, Jana Schroth, Suresh Babu Selvaraju, Goran Stevovic, Amanda Suchanek, Andrea Throop, Lyndon Tilson, Thomas Urban, Joe Voshell, Kimberly Wagner, Jonathan Williams, Mary Williamson, Qian Zeng, Tricia Zwiefelhofer, Clinton R. Paden, Duncan MacCannell |
| EPI_ISL_1481835 | Fulgent Genetics | Centers for Disease Control and Prevention Division of Viral Diseases, Pathogen Discovery | Dakota Howard, Dhwani Batra, Peter W. Cook, Kara Moser, Adrian Paskey, Jason Caravas, Benjamin Rambo-Martin, Shatavia Morrison, Christopher Gulvick, Scott Sammons, Yvette Unoarumhi, Darlene Wagner, Matthew Schmerer, Harry Gao, Mickey Li, John Gao, Joseph Fierro, Benafsh Sapra, Becky Tsai, Yan Meng, Doreen Ng, James Xie, Clinton R. Paden, Duncan MacCannell |
| EPI_ISL_1482274, EPI_ISL_1482275, EPI_ISL_1482276 | SUNY UPSTATE MEDICAL UNIVERSITY | Wadsworth Center, New York State Department of Health | Kirsten St. George, Daryl M. Lamson, Alexis Russell, Matthew Shudt, Melissa A Leisner, Jonathan Plitnick, Catharine Prussing, Navjot Singh, John Kelly, Erasmus Schneider, Erica Lasek-Nesselquist |
| EPI_ISL_1482277 | Columbia University Irving Medical Center | Wadsworth Center, New York State Department of Health | Kirsten St. George, Daryl M. Lamson, Alexis Russell, Matthew Shudt, Melissa A Leisner, Jonathan Plitnick, Catharine Prussing, Navjot Singh, John Kelly, Erasmus Schneider, Erica Lasek-Nesselquist |
| EPI_ISL_1482278, EPI_ISL_1482279, EPI_ISL_1482280, EPI_ISL_1482281, EPI_ISL_1482282, EPI_ISL_1482283, EPI_ISL_1482284, EPI_ISL_1482285, EPI_ISL_1482286 | URMC LABS | Wadsworth Center, New York State Department of Health | Kirsten St. George, Daryl M. Lamson, Alexis Russell, Matthew Shudt, Melissa A Leisner, Jonathan Plitnick, Catharine Prussing, Navjot Singh, John Kelly, Erasmus Schneider, Erica Lasek-Nesselquist |
| EPI_ISL_1482287, EPI_ISL_1482288, EPI_ISL_1482289, EPI_ISL_1482290, | NYC Pandemic Response Lab | Wadsworth Center, New York State Department of Health | Kirsten St. George, Daryl M. Lamson, Alexis Russell, Matthew Shudt, Melissa A Leisner, Jonathan Plitnick, Catharine Prussing, Navjot Singh, John Kelly, Erasmus Schneider, Erica Lasek-Nesselquist |

| | | | |
|---|---|---|---|
| EPI_ISL_1482291, EPI_ISL_1482292, EPI_ISL_1482293, EPI_ISL_1482294, EPI_ISL_1482295 | | | |
| EPI_ISL_1482296, EPI_ISL_1482297, EPI_ISL_1482298, EPI_ISL_1482299, EPI_ISL_1482300, EPI_ISL_1482301, EPI_ISL_1482302, EPI_ISL_1482303, EPI_ISL_1482304, EPI_ISL_1482305, EPI_ISL_1482306, EPI_ISL_1482307, EPI_ISL_1482308, EPI_ISL_1482309, EPI_ISL_1482310, EPI_ISL_1482311, EPI_ISL_1482312, EPI_ISL_1482313, EPI_ISL_1482314, EPI_ISL_1482315, EPI_ISL_1482316, EPI_ISL_1482317, EPI_ISL_1482318, EPI_ISL_1482319, EPI_ISL_1482320, EPI_ISL_1482321, EPI_ISL_1482322, EPI_ISL_1482323, EPI_ISL_1482324, EPI_ISL_1482325, EPI_ISL_1482326, EPI_ISL_1482327, EPI_ISL_1482328, EPI_ISL_1482329, EPI_ISL_1482330, EPI_ISL_1482331, EPI_ISL_1482332 | | | |
| see above | URMC LABS | Wadsworth Center, New York State Department of Health | Kirsten St. George, Daryl M. Lamson, Alexis Russell, Matthew Shudt, Melissa A Leisner, Jonathan Plitnick, Catharine Prussing, Navjot Singh, John Kelly, Erasmus Schneider, Erica Lasek-Nesselquist |
| EPI_ISL_1482333, EPI_ISL_1482334, EPI_ISL_1482335 | GLENS FALLS HOSPITAL LABORATORY | Wadsworth Center, New York State Department of Health | Kirsten St. George, Daryl M. Lamson, Alexis Russell, Matthew Shudt, Melissa A Leisner, Jonathan Plitnick, Catharine Prussing, Navjot Singh, John Kelly, Erasmus Schneider, Erica Lasek-Nesselquist |
| EPI_ISL_1482336, EPI_ISL_1482337, EPI_ISL_1482338, EPI_ISL_1482339, EPI_ISL_1482340, EPI_ISL_1482341, EPI_ISL_1482342, EPI_ISL_1482343, EPI_ISL_1482344, EPI_ISL_1482345 | MONTEFIORE NEW ROCHELLE HOSPITAL LABORATORY | Wadsworth Center, New York State Department of Health | Kirsten St. George, Daryl M. Lamson, Alexis Russell, Matthew Shudt, Melissa A Leisner, Jonathan Plitnick, Catharine Prussing, Navjot Singh, John Kelly, Erasmus Schneider, Erica Lasek-Nesselquist |
| EPI_ISL_1482346, EPI_ISL_1482347, EPI_ISL_1482348, EPI_ISL_1482349, EPI_ISL_1482350, EPI_ISL_1482351, EPI_ISL_1482352, EPI_ISL_1482353 | NORTH SHORE UNIVERSITY HOSPITAL | Wadsworth Center, New York State Department of Health | Kirsten St. George, Daryl M. Lamson, Alexis Russell, Matthew Shudt, Melissa A Leisner, Jonathan Plitnick, Catharine Prussing, Navjot Singh, John Kelly, Erasmus Schneider, Erica Lasek-Nesselquist |
| EPI_ISL_1482394, EPI_ISL_1482395, EPI_ISL_1482396, EPI_ISL_1482397, EPI_ISL_1482398, EPI_ISL_1482399, EPI_ISL_1482400, EPI_ISL_1482401, EPI_ISL_1482402, EPI_ISL_1482403, EPI_ISL_1482404, EPI_ISL_1482405 | | | |
| see above | WESTCHESTER MEDICAL CENTER | Wadsworth Center, New York State Department of Health | Kirsten St. George, Daryl M. Lamson, Alexis Russell, Matthew Shudt, Melissa A Leisner, Jonathan Plitnick, Catharine Prussing, Navjot Singh, John Kelly, Erasmus Schneider, Erica Lasek-Nesselquist |
| EPI_ISL_1482406, EPI_ISL_1482407 | SUNY UPSTATE MEDICAL UNIVERSITY | Wadsworth Center, New York State Department of Health | Kirsten St. George, Daryl M. Lamson, Alexis Russell, Matthew Shudt, Melissa A Leisner, Jonathan Plitnick, Catharine Prussing, Navjot Singh, John Kelly, Erasmus Schneider, Erica Lasek-Nesselquist |
| EPI_ISL_1482408, EPI_ISL_1482409 | BOSTON HEART DIAGNOSTICS CORP | Wadsworth Center, New York State Department of Health | Kirsten St. George, Daryl M. Lamson, Alexis Russell, Matthew Shudt, Melissa A Leisner, Jonathan Plitnick, Catharine Prussing, Navjot Singh, John Kelly, Erasmus Schneider, Erica Lasek-Nesselquist |
| EPI_ISL_1482410 | Columbia University Irving Medical Center | Wadsworth Center, New York State Department of Health | Kirsten St. George, Daryl M. Lamson, Alexis Russell, Matthew Shudt, Melissa A Leisner, Jonathan Plitnick, Catharine Prussing, Navjot Singh, John Kelly, Erasmus Schneider, Erica Lasek-Nesselquist |
| EPI_ISL_1482411, EPI_ISL_1482412, EPI_ISL_1482413, EPI_ISL_1482414, EPI_ISL_1482415, EPI_ISL_1482416, EPI_ISL_1482417, EPI_ISL_1482418, EPI_ISL_1482419, EPI_ISL_1482420, EPI_ISL_1482421, EPI_ISL_1482422, EPI_ISL_1482423, EPI_ISL_1482424, EPI_ISL_1482425, EPI_ISL_1482426, EPI_ISL_1482427, EPI_ISL_1482428, EPI_ISL_1482429, EPI_ISL_1482430, EPI_ISL_1482431, EPI_ISL_1482432, EPI_ISL_1482433, EPI_ISL_1482434, EPI_ISL_1482435, EPI_ISL_1482436, EPI_ISL_1482437, EPI_ISL_1482438, EPI_ISL_1482439, EPI_ISL_1482440, EPI_ISL_1482441, EPI_ISL_1482442, EPI_ISL_1482443, EPI_ISL_1482444, EPI_ISL_1482445 | | | |
| see above | WESTCHESTER MEDICAL CENTER | Wadsworth Center, New York State Department of Health | Kirsten St. George, Daryl M. Lamson, Alexis Russell, Matthew Shudt, Melissa A Leisner, Jonathan Plitnick, Catharine Prussing, Navjot Singh, John Kelly, Erasmus Schneider, Erica Lasek-Nesselquist |
| EPI_ISL_1482446, EPI_ISL_1482447, EPI_ISL_1482448, EPI_ISL_1482449, EPI_ISL_1482450, EPI_ISL_1482451, EPI_ISL_1482452, EPI_ISL_1482453 | BOSTON HEART DIAGNOSTICS CORP | Wadsworth Center, New York State Department of Health | Kirsten St. George, Daryl M. Lamson, Alexis Russell, Matthew Shudt, Melissa A Leisner, Jonathan Plitnick, Catharine Prussing, Navjot Singh, John Kelly, Erasmus Schneider, Erica Lasek-Nesselquist |
| EPI_ISL_1482454 | ST. FRANCIS HOSPITAL LABORATORY | Wadsworth Center, New York State Department of Health | Kirsten St. George, Daryl M. Lamson, Alexis Russell, Matthew Shudt, Melissa A Leisner, Jonathan Plitnick, Catharine Prussing, Navjot Singh, John Kelly, Erasmus Schneider, Erica Lasek-Nesselquist |
| EPI_ISL_1482455, EPI_ISL_1482456, EPI_ISL_1482457, EPI_ISL_1482458, EPI_ISL_1482459, EPI_ISL_1482460, EPI_ISL_1482461, EPI_ISL_1482462, EPI_ISL_1482463, EPI_ISL_1482464, EPI_ISL_1482465, EPI_ISL_1482466, EPI_ISL_1482467, EPI_ISL_1482468, EPI_ISL_1482469 | | | |
| see above | ALBANY MEDICAL CENTER | Wadsworth Center, New York State Department of Health | Kirsten St. George, Daryl M. Lamson, Alexis Russell, Matthew Shudt, Melissa A Leisner, Jonathan Plitnick, Catharine Prussing, Navjot Singh, John Kelly, Erasmus Schneider, Erica Lasek-Nesselquist |
| EPI_ISL_1482470 | Good Samaritan Hospital Laboratory | Wadsworth Center, New York State Department of Health | Kirsten St. George, Daryl M. Lamson, Alexis Russell, Matthew Shudt, Melissa A Leisner, Jonathan Plitnick, Catharine Prussing, Navjot Singh, John Kelly, Erasmus Schneider, Erica Lasek-Nesselquist |
| EPI_ISL_1482539, EPI_ISL_1482540, EPI_ISL_1482541, EPI_ISL_1482542, EPI_ISL_1482543, EPI_ISL_1482544, EPI_ISL_1482545, EPI_ISL_1482546, EPI_ISL_1482547, EPI_ISL_1482548, EPI_ISL_1482549, EPI_ISL_1482550, EPI_ISL_1482551, EPI_ISL_1482552, EPI_ISL_1482553, EPI_ISL_1482554, EPI_ISL_1482555 | | | |
| see above | WESTCHESTER MEDICAL CENTER | Wadsworth Center, New York State Department of Health | Kirsten St. George, Daryl M. Lamson, Alexis Russell, Matthew Shudt, Melissa A Leisner, Jonathan Plitnick, Catharine Prussing, Navjot Singh, John Kelly, Erasmus Schneider, Erica Lasek-Nesselquist |
| EPI_ISL_1482556, EPI_ISL_1482557, EPI_ISL_1482558, EPI_ISL_1482559, EPI_ISL_1482560, EPI_ISL_1482561, EPI_ISL_1482562, EPI_ISL_1482563, EPI_ISL_1482564, EPI_ISL_1482565, EPI_ISL_1482566, EPI_ISL_1482567, EPI_ISL_1482568, EPI_ISL_1482569, EPI_ISL_1482570, EPI_ISL_1482571, EPI_ISL_1482572 | | | |
| see above | NORTHWELL HEALTH LABORATORIES | Wadsworth Center, New York State Department of Health | Kirsten St. George, Daryl M. Lamson, Alexis Russell, Matthew Shudt, Melissa A Leisner, Jonathan Plitnick, Catharine Prussing, Navjot Singh, John Kelly, Erasmus Schneider, Erica Lasek-Nesselquist |
| EPI_ISL_1482573, EPI_ISL_1482574, EPI_ISL_1482575, EPI_ISL_1482576, EPI_ISL_1482577, EPI_ISL_1482578, EPI_ISL_1482579 | WHITE PLAINS HOSPITAL CENTER LABORATORY | Wadsworth Center, New York State Department of Health | Kirsten St. George, Daryl M. Lamson, Alexis Russell, Matthew Shudt, Melissa A Leisner, Jonathan Plitnick, Catharine Prussing, Navjot Singh, John Kelly, Erasmus Schneider, Erica Lasek-Nesselquist |
| EPI_ISL_1482580, EPI_ISL_1482581, EPI_ISL_1482582, EPI_ISL_1482583, EPI_ISL_1482584, EPI_ISL_1482585, EPI_ISL_1482586, EPI_ISL_1482587, EPI_ISL_1482588, EPI_ISL_1482589, EPI_ISL_1482590, EPI_ISL_1482591, EPI_ISL_1482592, EPI_ISL_1482593, EPI_ISL_1482594, EPI_ISL_1482595, EPI_ISL_1482596, EPI_ISL_1482597, EPI_ISL_1482598, EPI_ISL_1482599, EPI_ISL_1482600, EPI_ISL_1482601, EPI_ISL_1482602, EPI_ISL_1482603, EPI_ISL_1482604, EPI_ISL_1482605, EPI_ISL_1482606, EPI_ISL_1482607, EPI_ISL_1482608, EPI_ISL_1482609, EPI_ISL_1482610, EPI_ISL_1482611, EPI_ISL_1482612, EPI_ISL_1482613, EPI_ISL_1482614, EPI_ISL_1482615, EPI_ISL_1482616 | | | |
| see above | NORTHWELL HEALTH LABORATORIES | Wadsworth Center, New York State Department of Health | Kirsten St. George, Daryl M. Lamson, Alexis Russell, Matthew Shudt, Melissa A Leisner, Jonathan Plitnick, Catharine Prussing, Navjot Singh, John Kelly, Erasmus Schneider, Erica Lasek-Nesselquist |
| EPI_ISL_1482617, EPI_ISL_1482618, EPI_ISL_1482619, EPI_ISL_1482620, EPI_ISL_1482621, EPI_ISL_1482622, EPI_ISL_1482623, EPI_ISL_1482624 | WESTCHESTER MEDICAL CENTER | Wadsworth Center, New York State Department of Health | Kirsten St. George, Daryl M. Lamson, Alexis Russell, Matthew Shudt, Melissa A Leisner, Jonathan Plitnick, Catharine Prussing, Navjot Singh, John Kelly, Erasmus Schneider, Erica Lasek-Nesselquist |
| EPI_ISL_1482625, EPI_ISL_1482626 | URMC LABS | Wadsworth Center, New York State Department of Health | Kirsten St. George, Daryl M. Lamson, Alexis Russell, Matthew Shudt, Melissa A Leisner, Jonathan Plitnick, Catharine Prussing, Navjot Singh, John Kelly, Erasmus Schneider, Erica Lasek-Nesselquist |
| EPI_ISL_1491743, EPI_ISL_1491783, EPI_ISL_1491784, EPI_ISL_1491815, EPI_ISL_1491849, EPI_ISL_1491887, EPI_ISL_1491937 | Aegis Sciences Corporation | Centers for Disease Control and Prevention Division of Viral Diseases, Pathogen Discovery | Dakota Howard, Dhwani Batra, Peter W. Cook, Kara Moser, Adrian Paskey, Jason Caravas, Benjamin Rambo-Martin, Shatavia Morrison, Christopher Gulvick, Scott Sammons, Yvette Unoarumhi, Darlene Wagner, Matthew Schmerer, Cyndi Clark, Patrick Campbell, Rob Case, Vikramsinha Ghorpade, Holly Houdeshell, Ola Kvalvaag, Dillon Nall, Ethan Sanders, Alec Vest, Shaun Westlund, Matthew Hardison, Clinton R. Paden, Duncan MacCannell |
| EPI_ISL_1491965, EPI_ISL_1491986, EPI_ISL_1491994, EPI_ISL_1492112 | Quest Diagnostics Incorporated | Centers for Disease Control and Prevention Division of Viral Diseases, Pathogen Discovery | Dakota Howard, Dhwani Batra, Peter W. Cook, Kara Moser, Adrian Paskey, Jason Caravas, Benjamin Rambo-Martin, Shatavia Morrison, Christopher Gulvick, Scott Sammons, Yvette Unoarumhi, Darlene Wagner, Matthew Schmerer, S. H. Rosenthal, A. Gerasimova, R. M. Kagan, B. Anderson, M. Hua, Y. Liu, L.E. Bernstein, K.E. Livingston, A. Perez, I. A. Shlyakhter, R. V. Rolando, R. Owen, P. Tanpaiboon, F. Lacbawan, Clinton R. Paden, Duncan MacCannell |

| | | | |
|---|---|---|---|
| EPI_ISL_1493507, EPI_ISL_1493519, EPI_ISL_1493520, EPI_ISL_1493534, EPI_ISL_1493540, EPI_ISL_1493559 | Aegis Sciences Corporation | Centers for Disease Control and Prevention Division of Viral Diseases, Pathogen Discovery | Dakota Howard, Dhwani Batra, Peter W. Cook, Kara Moser, Adrian Paskey, Jason Caravas, Benjamin Rambo-Martin, Shatavia Morrison, Christopher Gulvick, Scott Sammons, Yvette Unoarumhi, Darlene Wagner, Matthew Schmerer, Cyndi Clark, Patrick Campbell, Rob Case, Vikramsinha Ghorpade, Holly Houdeshell, Ola Kvalvaag, Dillon Nall, Ethan Sanders, Alec Vest, Shaun Westlund, Matthew Hardison, Clinton R. Paden, Duncan MacCannell |
| EPI_ISL_1493627 | Quest Diagnostics Incorporated | Centers for Disease Control and Prevention Division of Viral Diseases, Pathogen Discovery | Dakota Howard, Dhwani Batra, Peter W. Cook, Kara Moser, Adrian Paskey, Jason Caravas, Benjamin Rambo-Martin, Shatavia Morrison, Christopher Gulvick, Scott Sammons, Yvette Unoarumhi, Darlene Wagner, Matthew Schmerer, S. H. Rosenthal, A. Gerasimova, R. M. Kagan, B. Anderson, M. Hua, Y. Liu, L.E. Bernstein, K.E. Livingston, A. Perez, I. A. Shlyakhter, R. V. Rolando, R. Owen, P. Tanpaiboon, F. Lacbawan, Clinton R. Paden, Duncan MacCannell |
| EPI_ISL_1493799 | Helix/Illumina | Centers for Disease Control and Prevention Division of Viral Diseases, Pathogen Discovery | Dakota Howard, Dhwani Batra, Peter W. Cook, Kara Moser, Adrian Paskey, Jason Caravas, Benjamin Rambo-Martin, Shatavia Morrison, Christopher Gulvick, Scott Sammons, Yvette Unoarumhi, Darlene Wagner, Matthew Schmerer, Eileen de Feo, Jan Antico, Christine Tran, Matthew Tolentino, Shannon Wickline, Kim Gietzen, Brad Sickler, Jingtao Liu, Eric Allen, Phil Febbo, Nicole L. Washington, Simon White, Geraint Levan, Kelly Schiabor Barrett, Elizabeth Cirulli, Alexandre Bolze, Ary Ascencio, Charlotte Rivera-Garcia, Ryan Cho, Jason Nguyen, Sherry Wang, Jimmy Ramirez, Tyler Cassens, Efren Sandoval, Magnus Isaksson, William Lee, David Becker, Marc Laurent, James Lu, Clinton R. Paden, Duncan MacCannell |
| EPI_ISL_1499395, EPI_ISL_1499396, EPI_ISL_1499397, EPI_ISL_1499398, EPI_ISL_1499399, EPI_ISL_1499400, EPI_ISL_1499401, EPI_ISL_1499402, EPI_ISL_1499403, EPI_ISL_1499404, EPI_ISL_1499405, EPI_ISL_1499406, EPI_ISL_1499407, EPI_ISL_1499408, EPI_ISL_1499409, EPI_ISL_1499410, EPI_ISL_1499411, EPI_ISL_1499412, EPI_ISL_1499413, EPI_ISL_1499414, EPI_ISL_1499415, EPI_ISL_1499416, EPI_ISL_1499417, EPI_ISL_1499418, EPI_ISL_1499419, EPI_ISL_1499420, EPI_ISL_1499421, EPI_ISL_1499422, EPI_ISL_1499423, EPI_ISL_1499424, EPI_ISL_1499425, EPI_ISL_1499426, EPI_ISL_1499427, EPI_ISL_1499428, EPI_ISL_1499429, EPI_ISL_1499430, EPI_ISL_1499431, EPI_ISL_1499432, EPI_ISL_1499433, EPI_ISL_1499434, EPI_ISL_1499435, EPI_ISL_1499436 | | | |
| see above | URMC LABS | Wadsworth Center, New York State Department of Health | Kirsten St. George, Daryl M. Lamson, Alexis Russel, Matthew Shudt, Melissa A Leisner, Jonathan Plitnick, Navjot Singh, John Kelly, Erasmus Schneider, Erica Lasek-Nesselquist |
| EPI_ISL_1499437, EPI_ISL_1499438, EPI_ISL_1499439, EPI_ISL_1499440, EPI_ISL_1499441, EPI_ISL_1499442, EPI_ISL_1499443, EPI_ISL_1499444, EPI_ISL_1499445, EPI_ISL_1499446, EPI_ISL_1499447, EPI_ISL_1499448, EPI_ISL_1499449, EPI_ISL_1499450, EPI_ISL_1499451, EPI_ISL_1499452, EPI_ISL_1499453, EPI_ISL_1499454, EPI_ISL_1499455, EPI_ISL_1499456 | | | |
| see above | SUNY UPSTATE MEDICAL UNIVERSITY | Wadsworth Center, New York State Department of Health | Kirsten St. George, Daryl M. Lamson, Alexis Russel, Matthew Shudt, Melissa A Leisner, Jonathan Plitnick, Navjot Singh, John Kelly, Erasmus Schneider, Erica Lasek-Nesselquist |
| EPI_ISL_1499457, EPI_ISL_1499458, EPI_ISL_1499459, EPI_ISL_1499460, EPI_ISL_1499461, EPI_ISL_1499462, EPI_ISL_1499463, EPI_ISL_1499464, EPI_ISL_1499465, EPI_ISL_1499466, EPI_ISL_1499467, EPI_ISL_1499468, EPI_ISL_1499469, EPI_ISL_1499470, EPI_ISL_1499471, EPI_ISL_1499472, EPI_ISL_1499473 | | | |
| see above | ALBANY MEDICAL CENTER HOSPITAL CLINICAL LABORATORIES | Wadsworth Center, New York State Department of Health | Kirsten St. George, Daryl M. Lamson, Alexis Russel, Matthew Shudt, Melissa A Leisner, Jonathan Plitnick, Navjot Singh, John Kelly, Erasmus Schneider, Erica Lasek-Nesselquist |
| EPI_ISL_1500472, EPI_ISL_1500473, EPI_ISL_1500474, EPI_ISL_1500475, EPI_ISL_1500476, EPI_ISL_1500477, EPI_ISL_1500478, EPI_ISL_1500479, EPI_ISL_1500480, EPI_ISL_1500481, EPI_ISL_1500482, EPI_ISL_1500483, EPI_ISL_1500484, EPI_ISL_1500485, EPI_ISL_1500486, EPI_ISL_1500487, EPI_ISL_1500488, EPI_ISL_1500489, EPI_ISL_1500490, EPI_ISL_1500491, EPI_ISL_1500492, EPI_ISL_1500493, EPI_ISL_1500494, EPI_ISL_1500495, EPI_ISL_1500496, EPI_ISL_1500497, EPI_ISL_1500498, EPI_ISL_1500499, EPI_ISL_1500500, EPI_ISL_1500501, EPI_ISL_1500502, EPI_ISL_1500503, EPI_ISL_1500504, EPI_ISL_1500505, EPI_ISL_1500506, EPI_ISL_1500507, EPI_ISL_1500508, EPI_ISL_1500509, EPI_ISL_1500510, EPI_ISL_1500511, EPI_ISL_1500512, EPI_ISL_1500513, EPI_ISL_1500514, EPI_ISL_1500516, EPI_ISL_1500517, EPI_ISL_1500518, EPI_ISL_1500519, EPI_ISL_1500520, EPI_ISL_1500521, EPI_ISL_1500522, EPI_ISL_1500523, EPI_ISL_1500524, EPI_ISL_1500525, EPI_ISL_1500526, EPI_ISL_1500527, EPI_ISL_1500528, EPI_ISL_1500529, EPI_ISL_1500530, EPI_ISL_1500531, EPI_ISL_1500532, EPI_ISL_1500533, EPI_ISL_1500534, EPI_ISL_1500535, EPI_ISL_1500536, EPI_ISL_1500537, EPI_ISL_1500538, EPI_ISL_1500539, EPI_ISL_1500540, EPI_ISL_1500541, EPI_ISL_1500542, EPI_ISL_1500543, EPI_ISL_1500544, EPI_ISL_1500545, EPI_ISL_1500546, EPI_ISL_1500547, EPI_ISL_1500548, EPI_ISL_1500549, EPI_ISL_1500550, EPI_ISL_1500551, EPI_ISL_1500552, EPI_ISL_1500553, EPI_ISL_1500554, EPI_ISL_1500555, EPI_ISL_1500556, EPI_ISL_1500557, EPI_ISL_1500558, EPI_ISL_1500559, EPI_ISL_1500560, EPI_ISL_1500561, EPI_ISL_1500562, EPI_ISL_1500563, EPI_ISL_1500564, EPI_ISL_1500565, EPI_ISL_1500566, EPI_ISL_1500567, EPI_ISL_1500568, EPI_ISL_1500569, EPI_ISL_1500570, EPI_ISL_1500571, EPI_ISL_1500572, EPI_ISL_1500573, EPI_ISL_1500574, EPI_ISL_1500575, EPI_ISL_1500576, EPI_ISL_1500577, EPI_ISL_1500578, EPI_ISL_1500579, EPI_ISL_1500580, EPI_ISL_1500581, EPI_ISL_1500582, EPI_ISL_1500583, EPI_ISL_1500584, EPI_ISL_1500585, EPI_ISL_1500586, EPI_ISL_1500587, EPI_ISL_1500588, EPI_ISL_1500589, EPI_ISL_1500590, EPI_ISL_1500591, EPI_ISL_1500592, EPI_ISL_1500593, EPI_ISL_1500594, EPI_ISL_1500595, EPI_ISL_1500596, EPI_ISL_1500597, EPI_ISL_1500598, EPI_ISL_1500599, EPI_ISL_1500600, EPI_ISL_1500601, EPI_ISL_1500602, EPI_ISL_1500603, EPI_ISL_1500604, EPI_ISL_1500605, EPI_ISL_1500606, EPI_ISL_1500607, EPI_ISL_1500608, EPI_ISL_1500609, EPI_ISL_1500610, EPI_ISL_1500611, EPI_ISL_1500612, EPI_ISL_1500613, EPI_ISL_1500614, EPI_ISL_1500615, EPI_ISL_1500616, EPI_ISL_1500617, EPI_ISL_1500618, EPI_ISL_1500619, EPI_ISL_1500620, EPI_ISL_1500621, EPI_ISL_1500622, EPI_ISL_1500623, EPI_ISL_1500624, EPI_ISL_1500625, EPI_ISL_1500626, EPI_ISL_1500627, EPI_ISL_1500628, EPI_ISL_1500629, EPI_ISL_1500630, EPI_ISL_1500631, EPI_ISL_1500632, EPI_ISL_1500633, EPI_ISL_1500634, EPI_ISL_1500635, EPI_ISL_1500636, EPI_ISL_1500637 | | | |
| see above | Clinical Microbiology Laboratory, NewYork Presbyterian Hospital/Columbia University Irving Medical Center | Uhlemann Laboratory, Columbia University Irving Medical Center | Medini K. Annavajhala, Anne-Catrin Uhlemann, Hiroshi Mohri, David Ho |
| EPI_ISL_1509180, EPI_ISL_1509238 | Yale Clinical Virology Lab | Grubaugh Lab - Yale School of Public Health | Joseph Fauver, Mallery Breban, Isabell Ott, Tara Alpert, Mary Petrone, Anderson Brito, Chantal Vogels, Annie Watkins, Chaney Kalinich, Jessica Rothman, Marie L. Landry, Nathan Grubaugh |
| EPI_ISL_1511714, EPI_ISL_1511768, EPI_ISL_1511769, EPI_ISL_1511770, EPI_ISL_1511774, EPI_ISL_1511776, EPI_ISL_1511779, EPI_ISL_1511781, EPI_ISL_1511782, EPI_ISL_1511783, EPI_ISL_1511789, EPI_ISL_1511791, EPI_ISL_1511793, EPI_ISL_1511794, EPI_ISL_1511797, EPI_ISL_1511802, EPI_ISL_1511805, EPI_ISL_1511809, EPI_ISL_1511813, EPI_ISL_1511814, EPI_ISL_1511815, EPI_ISL_1511816, EPI_ISL_1511817, EPI_ISL_1511820, EPI_ISL_1511821, EPI_ISL_1511822, EPI_ISL_1511824, EPI_ISL_1511829, EPI_ISL_1511830, EPI_ISL_1511831, EPI_ISL_1511832, EPI_ISL_1511834, EPI_ISL_1511836, EPI_ISL_1511838, EPI_ISL_1511842, EPI_ISL_1511846, EPI_ISL_1511848, EPI_ISL_1511850, EPI_ISL_1511852, EPI_ISL_1511856, EPI_ISL_1511858 | | | |
| see above | Quest Diagnostics Incorporated | Centers for Disease Control and Prevention Division of Viral Diseases, Pathogen Discovery | Dakota Howard, Dhwani Batra, Peter W. Cook, Kara Moser, Adrian Paskey, Jason Caravas, Benjamin Rambo-Martin, Shatavia Morrison, Christopher Gulvick, Scott Sammons, Yvette Unoarumhi, Darlene Wagner, Matthew Schmerer, S. H. Rosenthal, A. Gerasimova, R. M. Kagan, B. Anderson, M. Hua, Y. Liu, L.E. Bernstein, K.E. Livingston, A. Perez, I. A. Shlyakhter, R. V. Rolando, R. Owen, P. Tanpaiboon, F. Lacbawan, Clinton R. Paden, Duncan MacCannell |
| EPI_ISL_1511926, EPI_ISL_1512010, EPI_ISL_1512135, EPI_ISL_1512243, EPI_ISL_1512415, EPI_ISL_1512449, EPI_ISL_1512483 | Helix/Illumina | Centers for Disease Control and Prevention Division of Viral Diseases, Pathogen Discovery | Dakota Howard, Dhwani Batra, Peter W. Cook, Kara Moser, Adrian Paskey, Jason Caravas, Benjamin Rambo-Martin, Shatavia Morrison, Christopher Gulvick, Scott Sammons, Yvette Unoarumhi, Darlene Wagner, Matthew Schmerer, Eileen de Feo, Jan Antico, Christine Tran, Matthew Tolentino, Shannon Wickline, Kim Gietzen, Brad Sickler, Jingtao Liu, Eric Allen, Phil Febbo, Nicole L. Washington, Simon White, Geraint Levan, Kelly Schiabor Barrett, Elizabeth Cirulli, Alexandre Bolze, Ary Ascencio, Charlotte Rivera-Garcia, Ryan Cho, Jason Nguyen, Sherry Wang, Jimmy Ramirez, Tyler Cassens, Efren Sandoval, Magnus Isaksson, William Lee, David Becker, Marc Laurent, James Lu, Clinton R. Paden, Duncan MacCannell |
| EPI_ISL_1512832, EPI_ISL_1512837, EPI_ISL_1512853, EPI_ISL_1512854, EPI_ISL_1512855, EPI_ISL_1512857, EPI_ISL_1512862, EPI_ISL_1512896, EPI_ISL_1512917, EPI_ISL_1512935, EPI_ISL_1512948, EPI_ISL_1512949, EPI_ISL_1512950, EPI_ISL_1512952, EPI_ISL_1513036, EPI_ISL_1513039, EPI_ISL_1513044, EPI_ISL_1513074, EPI_ISL_1513092, EPI_ISL_1513094, EPI_ISL_1513096, EPI_ISL_1513101, EPI_ISL_1513102, EPI_ISL_1513131 | | | |
| see above | Infinity Biologix | Centers for Disease Control and Prevention Division of Viral Diseases, Pathogen Discovery | Dakota Howard, Dhwani Batra, Peter W. Cook, Kara Moser, Adrian Paskey, Jason Caravas, Benjamin Rambo-Martin, Shatavia Morrison, Christopher Gulvick, Scott Sammons, Yvette Unoarumhi, Darlene Wagner, Matthew Schmerer, Christian Bixby, Yihe Wang, Jonathan Schultz, Chirayu Goswami, Russ Hager, Robin Grimwood, Clinton R. Paden, Duncan MacCannell |
| EPI_ISL_1513202, EPI_ISL_1513204, EPI_ISL_1513205, EPI_ISL_1513229, EPI_ISL_1513231, EPI_ISL_1513233, EPI_ISL_1513257, EPI_ISL_1513258, EPI_ISL_1513259, EPI_ISL_1513261, EPI_ISL_1513337, EPI_ISL_1513339, EPI_ISL_1513368, EPI_ISL_1513473, EPI_ISL_1513482, EPI_ISL_1513562, EPI_ISL_1513564, EPI_ISL_1513567, EPI_ISL_1513590, EPI_ISL_1513592 | | | |
| see above | Aegis Sciences Corporation | Centers for Disease Control and Prevention Division of Viral Diseases, Pathogen Discovery | Dakota Howard, Dhwani Batra, Peter W. Cook, Kara Moser, Adrian Paskey, Jason Caravas, Benjamin Rambo-Martin, Shatavia Morrison, Christopher Gulvick, Scott Sammons, Yvette Unoarumhi, Darlene Wagner, Matthew Schmerer, Cyndi Clark, Patrick Campbell, Rob Case, Vikramsinha Ghorpade, Holly Houdeshell, Ola Kvalvaag, Dillon Nall, Ethan Sanders, Alec Vest, Shaun Westlund, Matthew Hardison, Clinton R. Paden, Duncan MacCannell |
| EPI_ISL_1513637, EPI_ISL_1513640, EPI_ISL_1513641, EPI_ISL_1513642, EPI_ISL_1513647, EPI_ISL_1513652, EPI_ISL_1513653, EPI_ISL_1513989, EPI_ISL_1513990, EPI_ISL_1513992, EPI_ISL_1513993, EPI_ISL_1513994, EPI_ISL_1513996, EPI_ISL_1513997, EPI_ISL_1514220, EPI_ISL_1514224, EPI_ISL_1514284, EPI_ISL_1514329, EPI_ISL_1514364, EPI_ISL_1514575, EPI_ISL_1514582, EPI_ISL_1514583, EPI_ISL_1514585, EPI_ISL_1514587, EPI_ISL_1514588, EPI_ISL_1514590, EPI_ISL_1514591, EPI_ISL_1514592, EPI_ISL_1514604, EPI_ISL_1514605, EPI_ISL_1514606, EPI_ISL_1514621, EPI_ISL_1514624, EPI_ISL_1514625, EPI_ISL_1514627, EPI_ISL_1514628, EPI_ISL_1514632, EPI_ISL_1514633, EPI_ISL_1514634, EPI_ISL_1514635, EPI_ISL_1514636, EPI_ISL_1514637, EPI_ISL_1514638, EPI_ISL_1514647, EPI_ISL_1514648, EPI_ISL_1514649, EPI_ISL_1514653, EPI_ISL_1514657, EPI_ISL_1514659, EPI_ISL_1514660, EPI_ISL_1514661, EPI_ISL_1514663, EPI_ISL_1514692, EPI_ISL_1514693, EPI_ISL_1514702, EPI_ISL_1514706, EPI_ISL_1514707, EPI_ISL_1514708, EPI_ISL_1514709, EPI_ISL_1514710, EPI_ISL_1514711, EPI_ISL_1514716, EPI_ISL_1514718, EPI_ISL_1514719, EPI_ISL_1514721, EPI_ISL_1515063, EPI_ISL_1515076, EPI_ISL_1515077, EPI_ISL_1515079, EPI_ISL_1515083, EPI_ISL_1515084, EPI_ISL_1515085, EPI_ISL_1515086, EPI_ISL_1515087, EPI_ISL_1515088, EPI_ISL_1515089, EPI_ISL_1515125, EPI_ISL_1515128, EPI_ISL_1515129, EPI_ISL_1515132, EPI_ISL_1515137, EPI_ISL_1515138, EPI_ISL_1515154, EPI_ISL_1515323, EPI_ISL_1515326, EPI_ISL_1515327, EPI_ISL_1515329, EPI_ISL_1515520 | | | |
| see above | Laboratory Corporation of America | Centers for Disease Control and Prevention Division of Viral Diseases, Pathogen Discovery | Dakota Howard, Dhwani Batra, Peter W. Cook, Kara Moser, Adrian Paskey, Jason Caravas, Benjamin Rambo-Martin, Shatavia Morrison, Christopher Gulvick, Scott Sammons, Yvette Unoarumhi, Darlene Wagner, Matthew Schmerer, Minoo Agarwal, Eyad Almasri, Debbie Boles, Ayla Burns, Nuthawin Charoensri, Oren Cohen, Susan Countryman, Mary Ann Cristobal, Bobbi Croy, Suzanne Dale, Hrushikesh Deshmukh, Amanda Douglas, Vincent Drouillon, Marcia Eisenberg, Howard Engler, Rama Ghatti, Prashant Gupta, Susan Hicks, Jake Humphrey, Lax Iyer, Manoj Jain, Mohan Kolli, Brian Krueger, Tim Kuphal, Stanley Letovsky, Michael Levandoski, Craig Lukasik, Jonathan Meltzer, Brian Norvell, Mindy Nye, Scott Parker, Christos Petropoulos, John Pruitt, Steven Ragan, Scott Ryan, Mike Sapeta, Jana Schroth, Suresh Babu Selvaraju, Goran Stevovic, Amanda Suchanek, Andrea Throop, Lyndon Tilson, Thomas Urban, Joe Voshell, Kimberly Wagner, Jonathan Williams, Mary Williamson, Qian Zeng, Tricia Zwiefelhofer, Clinton R. Paden, Duncan MacCannell |
| EPI_ISL_1516205, EPI_ISL_1516206, EPI_ISL_1516207, EPI_ISL_1516208, EPI_ISL_1516209, EPI_ISL_1516210, EPI_ISL_1516211, EPI_ISL_1516212 | NYC Department of Health and Mental Hygiene | Centers for Disease Control and Prevention Division of Viral Diseases, Pathogen Discovery | Mili Sheth, Sarah Nobles, Jasmine Padilla, Mark Burroughs, Shoshona Le, Katie Dillon, Peter Cook, Clinton R. Paden, Dhwani Batra, Krista Queen, Kristen Knipe, Dakota Howard, Yvette Unoarumhi, Darlene Wagner, Matthew Schmerer, Ben L. Rambo-Martin, Kristine Lacek, Sam Shepard, Alison Laufer Halpin, Dave Wentworth, Vivien Dugan, Suxiang Tong, Justin Lee |

| | | | |
|---|---|---|---|
| EPI_ISL_1516525, EPI_ISL_1516526, EPI_ISL_1516527, EPI_ISL_1516528, EPI_ISL_1516530, EPI_ISL_1516531, EPI_ISL_1516532, EPI_ISL_1516533, EPI_ISL_1516536, EPI_ISL_1516537, EPI_ISL_1516538, EPI_ISL_1516539, EPI_ISL_1516541, EPI_ISL_1516542, EPI_ISL_1516543, EPI_ISL_1516544, EPI_ISL_1516563, EPI_ISL_1516571 | | | |
| see above | Broad Institute Clinical Research Sequencing Platform | Infectious Disease Program, Broad Institute of Harvard and MIT | Siddle,K.J., Adams,G., Pearlman,L., Gladden-Young,A., Vicente,G., Blumenstiel,B., DeFelice,M., Lee,M., McGovern,S., Lagerborg,K., Rudy,M., DeRuff,K., Carter,A., Normandin,E., Bauer,M., Reilly,S., Tomkins-Tinch,C., Loreth,C., Chaluvadi,S., Meldrim,J., Granger,B., Lemieux,J.E., Birren,B.W., Sabeti,P.C., Larkin,K., Dodge,S., Lennon,N., Madoff,L., Brown,C., Gallagher,G., Smole,S., Park,D.J., Gabriel,S., and MacInnis,B.L. |
| EPI_ISL_1525019, EPI_ISL_1525020, EPI_ISL_1525037, EPI_ISL_1525038, EPI_ISL_1525043, EPI_ISL_1525050, EPI_ISL_1525057, EPI_ISL_1525058, EPI_ISL_1525108, EPI_ISL_1525137, EPI_ISL_1525181, EPI_ISL_1525183, EPI_ISL_1525218, EPI_ISL_1525239, EPI_ISL_1525243 | | | |
| see above | Aegis Sciences Corporation | Centers for Disease Control and Prevention Division of Viral Diseases, Pathogen Discovery | Dakota Howard, Dhwani Batra, Peter W. Cook, Kara Moser, Adrian Paskey, Jason Caravas, Benjamin Rambo-Martin, Shatavia Morrison, Christopher Gulvick, Scott Sammons, Yvette Unoarumhi, Darlene Wagner, Matthew Schmerer, Cyndi Clark, Patrick Campbell, Rob Case, Vikramsinha Ghorpade, Holly Houdeshell, Ola Kvalvaag, Dillon Nall, Ethan Sanders, Alec Vest, Shaun Westlund, Matthew Hardison, Clinton R. Paden, Duncan MacCannell |
| EPI_ISL_1526833, EPI_ISL_1526834, EPI_ISL_1526835 | UW Virology Lab | UW Virology Lab | Pavitra Roychoudhury, Hong Xie, Lasata Shrestha, Shah Mohamed Bakhash, Michelle Lin, Margaret Mills, Noah Baker, Sean Ellis, Saraswathi Sathees, Meei-Li Huang, Keith R Jerome, Alexander Greninger |
| EPI_ISL_1531475 | US Air Force School of Aerospace Medicine | US Air Force School of Aerospace Medicine | Anthony Fries, Jennifer Meyer, William Gruner, William Buggele, Amanda Javorina, Sarah Purves, Clarise Starr, Elizabeth Macias |
| EPI_ISL_1532731, EPI_ISL_1532732, EPI_ISL_1532733, EPI_ISL_1532734, EPI_ISL_1532735, EPI_ISL_1532736, EPI_ISL_1532737, EPI_ISL_1532738, EPI_ISL_1532739, EPI_ISL_1532740, EPI_ISL_1532741, EPI_ISL_1532742, EPI_ISL_1532743, EPI_ISL_1532744, EPI_ISL_1532746, EPI_ISL_1532747, EPI_ISL_1532748, EPI_ISL_1532749, EPI_ISL_1532750, EPI_ISL_1532751, EPI_ISL_1532752, EPI_ISL_1532753, EPI_ISL_1532754, EPI_ISL_1532755, EPI_ISL_1532756, EPI_ISL_1532757, EPI_ISL_1532758, EPI_ISL_1532759, EPI_ISL_1532760, EPI_ISL_1532761, EPI_ISL_1532762, EPI_ISL_1532763, EPI_ISL_1532765, EPI_ISL_1532766, EPI_ISL_1532767, EPI_ISL_1532769, EPI_ISL_1532770, EPI_ISL_1532771, EPI_ISL_1532772, EPI_ISL_1532773, EPI_ISL_1532774, EPI_ISL_1532775, EPI_ISL_1532776, EPI_ISL_1532777, EPI_ISL_1532778, EPI_ISL_1532779, EPI_ISL_1532780, EPI_ISL_1532781, EPI_ISL_1532783, EPI_ISL_1532784, EPI_ISL_1532785 | | | |
| see above | NYU Langone Health | Departments of Pathology and Medicine, New York University School of Medicine | Adriana Heguy, Dacia Dimartino, Emily Guzman, Christian Marier, Peter Meyn, Sitharam Ramaswami, Gael Westby, Paul Zappile, Yutong Zhang, Paolo Cotzia, Guiqing Wang |
| EPI_ISL_1540718 | The Jackson Laboratory | The Jackson Laboratory | Bergeron D, Renzette N, Adams M, Omerza G, Kelly K, Long J, Li L |
| EPI_ISL_1542074, EPI_ISL_1542075, EPI_ISL_1542076, EPI_ISL_1542077, EPI_ISL_1542078, EPI_ISL_1542079, EPI_ISL_1542080, EPI_ISL_1542081, EPI_ISL_1542082, EPI_ISL_1542083, EPI_ISL_1542084, EPI_ISL_1542085, EPI_ISL_1542086, EPI_ISL_1542087, EPI_ISL_1542088, EPI_ISL_1542089, EPI_ISL_1542090, EPI_ISL_1542091, EPI_ISL_1542092, EPI_ISL_1542093, EPI_ISL_1542094, EPI_ISL_1542095, EPI_ISL_1542096, EPI_ISL_1542097, EPI_ISL_1542098, EPI_ISL_1542099, EPI_ISL_1542100, EPI_ISL_1542101, EPI_ISL_1542102, EPI_ISL_1542103, EPI_ISL_1542104, EPI_ISL_1542105, EPI_ISL_1542106, EPI_ISL_1542107, EPI_ISL_1542108, EPI_ISL_1542109, EPI_ISL_1542110, EPI_ISL_1542111, EPI_ISL_1542112, EPI_ISL_1542113, EPI_ISL_1542114, EPI_ISL_1542115, EPI_ISL_1542116, EPI_ISL_1542117, EPI_ISL_1542118, EPI_ISL_1542119, EPI_ISL_1542120, EPI_ISL_1542121, EPI_ISL_1542122, EPI_ISL_1542123, EPI_ISL_1542124, EPI_ISL_1542125, EPI_ISL_1542126, EPI_ISL_1542127, EPI_ISL_1542128, EPI_ISL_1542129, EPI_ISL_1542130, EPI_ISL_1542131, EPI_ISL_1542132, EPI_ISL_1542133, EPI_ISL_1542134, EPI_ISL_1542135, EPI_ISL_1542136, EPI_ISL_1542137, EPI_ISL_1542138, EPI_ISL_1542139, EPI_ISL_1542140, EPI_ISL_1542141, EPI_ISL_1542142, EPI_ISL_1542143, EPI_ISL_1542144, EPI_ISL_1542145, EPI_ISL_1542146, EPI_ISL_1542147, EPI_ISL_1542148, EPI_ISL_1542149, EPI_ISL_1542150, EPI_ISL_1542151, EPI_ISL_1542152, EPI_ISL_1542153, EPI_ISL_1542154, EPI_ISL_1542155, EPI_ISL_1542156, EPI_ISL_1542157, EPI_ISL_1542158, EPI_ISL_1542159, EPI_ISL_1542160, EPI_ISL_1542161, EPI_ISL_1542162, EPI_ISL_1542163, EPI_ISL_1542164, EPI_ISL_1542165, EPI_ISL_1542166, EPI_ISL_1542167, EPI_ISL_1542168, EPI_ISL_1542169, EPI_ISL_1542170, EPI_ISL_1542171, EPI_ISL_1542172, EPI_ISL_1542173, EPI_ISL_1542174, EPI_ISL_1542175, EPI_ISL_1542176, EPI_ISL_1542177, EPI_ISL_1542178, EPI_ISL_1542179, EPI_ISL_1542180, EPI_ISL_1542181, EPI_ISL_1542182, EPI_ISL_1542183, EPI_ISL_1542184, EPI_ISL_1542185, EPI_ISL_1542186, EPI_ISL_1542187, EPI_ISL_1542188, EPI_ISL_1542189, EPI_ISL_1542190, EPI_ISL_1542191, EPI_ISL_1542192, EPI_ISL_1542193, EPI_ISL_1542194, EPI_ISL_1542195, EPI_ISL_1542196, EPI_ISL_1542197, EPI_ISL_1542198, EPI_ISL_1542199, EPI_ISL_1542200, EPI_ISL_1542201, EPI_ISL_1542202, EPI_ISL_1542203, EPI_ISL_1542204, EPI_ISL_1542205, EPI_ISL_1542206, EPI_ISL_1542207, EPI_ISL_1542208, EPI_ISL_1542209, EPI_ISL_1542210, EPI_ISL_1542211, EPI_ISL_1542212, EPI_ISL_1542213, EPI_ISL_1542214, EPI_ISL_1542215, EPI_ISL_1542216, EPI_ISL_1542217, EPI_ISL_1542218, EPI_ISL_1542219, EPI_ISL_1542220, EPI_ISL_1542221, EPI_ISL_1542222, EPI_ISL_1542223, EPI_ISL_1542224, EPI_ISL_1542225, EPI_ISL_1542226, EPI_ISL_1542227, EPI_ISL_1542228, EPI_ISL_1542229, EPI_ISL_1542230, EPI_ISL_1542231, EPI_ISL_1542232, EPI_ISL_1542233, EPI_ISL_1542234, EPI_ISL_1542235, EPI_ISL_1542236, EPI_ISL_1542237, EPI_ISL_1542238, EPI_ISL_1542239, EPI_ISL_1542240, EPI_ISL_1542241, EPI_ISL_1542242, EPI_ISL_1542243, EPI_ISL_1542244, EPI_ISL_1542245, EPI_ISL_1542246, EPI_ISL_1542247, EPI_ISL_1542248, EPI_ISL_1542249, EPI_ISL_1542250, EPI_ISL_1542251, EPI_ISL_1542252, EPI_ISL_1542253, EPI_ISL_1542254, EPI_ISL_1542255, EPI_ISL_1542256, EPI_ISL_1542257, EPI_ISL_1542258, EPI_ISL_1542259, EPI_ISL_1542260, EPI_ISL_1542261, EPI_ISL_1542262, EPI_ISL_1542263, EPI_ISL_1542264, EPI_ISL_1542265, EPI_ISL_1542266, EPI_ISL_1542267, EPI_ISL_1542268, EPI_ISL_1542269, EPI_ISL_1542270, EPI_ISL_1542271, EPI_ISL_1542272, EPI_ISL_1542273, EPI_ISL_1542274, EPI_ISL_1542275, EPI_ISL_1542276, EPI_ISL_1542277, EPI_ISL_1542278, EPI_ISL_1542279, EPI_ISL_1542280, EPI_ISL_1542281, EPI_ISL_1542282, EPI_ISL_1542283, EPI_ISL_1542284, EPI_ISL_1542285, EPI_ISL_1542286, EPI_ISL_1542287, EPI_ISL_1542288, EPI_ISL_1542289, EPI_ISL_1542290, EPI_ISL_1542291, EPI_ISL_1542292, EPI_ISL_1542293, EPI_ISL_1542294, EPI_ISL_1542295, EPI_ISL_1542296, EPI_ISL_1542297, EPI_ISL_1542298, EPI_ISL_1542299, EPI_ISL_1542300, EPI_ISL_1542301, EPI_ISL_1542302, EPI_ISL_1542303, EPI_ISL_1542304, EPI_ISL_1542305, EPI_ISL_1542306, EPI_ISL_1542307, EPI_ISL_1542308, EPI_ISL_1542309, EPI_ISL_1542310, EPI_ISL_1542311, EPI_ISL_1542312, EPI_ISL_1542313, EPI_ISL_1542314, EPI_ISL_1542315, EPI_ISL_1542316, EPI_ISL_1542317, EPI_ISL_1542318, EPI_ISL_1542319, EPI_ISL_1542320, EPI_ISL_1542321, EPI_ISL_1542322, EPI_ISL_1542323, EPI_ISL_1542324, EPI_ISL_1542325, EPI_ISL_1542326, EPI_ISL_1542327, EPI_ISL_1542328, EPI_ISL_1542329, EPI_ISL_1542330, EPI_ISL_1542331, EPI_ISL_1542332, EPI_ISL_1542333, EPI_ISL_1542334, EPI_ISL_1542335, EPI_ISL_1542336, EPI_ISL_1542337, EPI_ISL_1542338, EPI_ISL_1542339, EPI_ISL_1542340, EPI_ISL_1542341, EPI_ISL_1542342, EPI_ISL_1542343, EPI_ISL_1542344, EPI_ISL_1542345, EPI_ISL_1542346, EPI_ISL_1542347, EPI_ISL_1542348, EPI_ISL_1542349, EPI_ISL_1542350, EPI_ISL_1542351, EPI_ISL_1542352, EPI_ISL_1542353, EPI_ISL_1542354, EPI_ISL_1542355, EPI_ISL_1542356, EPI_ISL_1542357, EPI_ISL_1542358, EPI_ISL_1542359, EPI_ISL_1542360, EPI_ISL_1542361, EPI_ISL_1542362, EPI_ISL_1542363, EPI_ISL_1542364, EPI_ISL_1542365, EPI_ISL_1542366, EPI_ISL_1542367, EPI_ISL_1542368, EPI_ISL_1542369, EPI_ISL_1542370, EPI_ISL_1542371, EPI_ISL_1542372, EPI_ISL_1542373, EPI_ISL_1542374, EPI_ISL_1542375, EPI_ISL_1542376, EPI_ISL_1542377, EPI_ISL_1542378, EPI_ISL_1542379, EPI_ISL_1542380, EPI_ISL_1542381, EPI_ISL_1542382, EPI_ISL_1542383, EPI_ISL_1542384, EPI_ISL_1542385, EPI_ISL_1542386, EPI_ISL_1542387, EPI_ISL_1542388, EPI_ISL_1542389, EPI_ISL_1542390, EPI_ISL_1542391, EPI_ISL_1542392, EPI_ISL_1542393, EPI_ISL_1542394, EPI_ISL_1542395, EPI_ISL_1542396, EPI_ISL_1542397, EPI_ISL_1542398, EPI_ISL_1542399, EPI_ISL_1542400, EPI_ISL_1542401, EPI_ISL_1542402, EPI_ISL_1542403, EPI_ISL_1542404, EPI_ISL_1542405, EPI_ISL_1542406, EPI_ISL_1542407, EPI_ISL_1542408, EPI_ISL_1542409, EPI_ISL_1542410, EPI_ISL_1542411, EPI_ISL_1542412, EPI_ISL_1542413, EPI_ISL_1542414, EPI_ISL_1542415, EPI_ISL_1542416, EPI_ISL_1542417, EPI_ISL_1542418, EPI_ISL_1542419, EPI_ISL_1542420, EPI_ISL_1542421, EPI_ISL_1542422, EPI_ISL_1542423, EPI_ISL_1542424, EPI_ISL_1542425, EPI_ISL_1542426, EPI_ISL_1542427, EPI_ISL_1542428, EPI_ISL_1542429, EPI_ISL_1542430, EPI_ISL_1542431, EPI_ISL_1542432, EPI_ISL_1542433, EPI_ISL_1542434, EPI_ISL_1542435, EPI_ISL_1542436, EPI_ISL_1542437, EPI_ISL_1542438, EPI_ISL_1542439, EPI_ISL_1542440, EPI_ISL_1542441, EPI_ISL_1542442, EPI_ISL_1542443, EPI_ISL_1542444, EPI_ISL_1542445, EPI_ISL_1542446, EPI_ISL_1542447, EPI_ISL_1542448, EPI_ISL_1542449, EPI_ISL_1542450, EPI_ISL_1542451, EPI_ISL_1542452, EPI_ISL_1542453, EPI_ISL_1542454, EPI_ISL_1542455, EPI_ISL_1542456, EPI_ISL_1542457, EPI_ISL_1542458, EPI_ISL_1542459, EPI_ISL_1542460, EPI_ISL_1542461, EPI_ISL_1542462, EPI_ISL_1542463, EPI_ISL_1542464, EPI_ISL_1542465, EPI_ISL_1542466, EPI_ISL_1542467, EPI_ISL_1542468, EPI_ISL_1542469, EPI_ISL_1542470, EPI_ISL_1542471, EPI_ISL_1542472, EPI_ISL_1542473, EPI_ISL_1542474, EPI_ISL_1542475, EPI_ISL_1542476, EPI_ISL_1542477, EPI_ISL_1542478, EPI_ISL_1542479, EPI_ISL_1542480, EPI_ISL_1542481, EPI_ISL_1542482, EPI_ISL_1542483, EPI_ISL_1542484, EPI_ISL_1542485, EPI_ISL_1542486, EPI_ISL_1542487, EPI_ISL_1542488, EPI_ISL_1542489, EPI_ISL_1542490, EPI_ISL_1542491, EPI_ISL_1542492, EPI_ISL_1542493, EPI_ISL_1542494, EPI_ISL_1542495, EPI_ISL_1542496, EPI_ISL_1542497, EPI_ISL_1542498, EPI_ISL_1542499, EPI_ISL_1542500, EPI_ISL_1542501, EPI_ISL_1542502, EPI_ISL_1542503, EPI_ISL_1542504, EPI_ISL_1542505, EPI_ISL_1542506, EPI_ISL_1542507, EPI_ISL_1542508, EPI_ISL_1542509, EPI_ISL_1542510, EPI_ISL_1542511, EPI_ISL_1542512, EPI_ISL_1542513, EPI_ISL_1542514, EPI_ISL_1542515, EPI_ISL_1542516, EPI_ISL_1542517, EPI_ISL_1542518, EPI_ISL_1542519, EPI_ISL_1542520, EPI_ISL_1542521, EPI_ISL_1542522, EPI_ISL_1542523, EPI_ISL_1542524, EPI_ISL_1542525, EPI_ISL_1542526, EPI_ISL_1542527, EPI_ISL_1542528, EPI_ISL_1542529, EPI_ISL_1542530, EPI_ISL_1542531, EPI_ISL_1542532, EPI_ISL_1542533, EPI_ISL_1542534, EPI_ISL_1542535, EPI_ISL_1542536, EPI_ISL_1542537, EPI_ISL_1542538, EPI_ISL_1542539, EPI_ISL_1542540, EPI_ISL_1542541, EPI_ISL_1542542, EPI_ISL_1542543, EPI_ISL_1542544, EPI_ISL_1542545, EPI_ISL_1542546, EPI_ISL_1542547, EPI_ISL_1542548, EPI_ISL_1542549, EPI_ISL_1542550, EPI_ISL_1542551, EPI_ISL_1542552, EPI_ISL_1542553, EPI_ISL_1542554, EPI_ISL_1542555, EPI_ISL_1542556, EPI_ISL_1542557, EPI_ISL_1542558, EPI_ISL_1542559, EPI_ISL_1542560, EPI_ISL_1542561, EPI_ISL_1542562, EPI_ISL_1542563, EPI_ISL_1542564, EPI_ISL_1542565, EPI_ISL_1542566, EPI_ISL_1542567, EPI_ISL_1542568, EPI_ISL_1542569, EPI_ISL_1542570, EPI_ISL_1542571, EPI_ISL_1542572, EPI_ISL_1542573, EPI_ISL_1542574, EPI_ISL_1542575, EPI_ISL_1542576, EPI_ISL_1542577, EPI_ISL_1542578, EPI_ISL_1542579, EPI_ISL_1542580, EPI_ISL_1542581, EPI_ISL_1542582, EPI_ISL_1542583, EPI_ISL_1542584, EPI_ISL_1542585, EPI_ISL_1542586, EPI_ISL_1542587, EPI_ISL_1542588, EPI_ISL_1542589, EPI_ISL_1542590, EPI_ISL_1542591, EPI_ISL_1542592, EPI_ISL_1542593, EPI_ISL_1542594, EPI_ISL_1542595, EPI_ISL_1542596, EPI_ISL_1542597, EPI_ISL_1542598, EPI_ISL_1542599, EPI_ISL_1542600, EPI_ISL_1542601, EPI_ISL_1542602, EPI_ISL_1542603, EPI_ISL_1542604, EPI_ISL_1542605, EPI_ISL_1542606, EPI_ISL_1542607, EPI_ISL_1542608, EPI_ISL_1542609, EPI_ISL_1542610, EPI_ISL_1542611, EPI_ISL_1542612, EPI_ISL_1542613, EPI_ISL_1542614, EPI_ISL_1542615, EPI_ISL_1542616, EPI_ISL_1542617, EPI_ISL_1542618, EPI_ISL_1542619, EPI_ISL_1542620, EPI_ISL_1542621, EPI_ISL_1542622, EPI_ISL_1542623, EPI_ISL_1542624, EPI_ISL_1542625, EPI_ISL_1542626, EPI_ISL_1542627, EPI_ISL_1542628, EPI_ISL_1542629, EPI_ISL_1542630, EPI_ISL_1542631, EPI_ISL_1542632, EPI_ISL_1542633, EPI_ISL_1542634, EPI_ISL_1542635, EPI_ISL_1542636, EPI_ISL_1542637, EPI_ISL_1542638, EPI_ISL_1542639, EPI_ISL_1542640, EPI_ISL_1542641, EPI_ISL_1542642, EPI_ISL_1542643, EPI_ISL_1542644, EPI_ISL_1542645, EPI_ISL_1542646, EPI_ISL_1542647, EPI_ISL_1542648, EPI_ISL_1542649, EPI_ISL_1542650, EPI_ISL_1542651, EPI_ISL_1542652, EPI_ISL_1542653, EPI_ISL_1542654, EPI_ISL_1542655, EPI_ISL_1542656, EPI_ISL_1542657, EPI_ISL_1542658, EPI_ISL_1542659, EPI_ISL_1542660, EPI_ISL_1542661, EPI_ISL_1542662, EPI_ISL_1542663, EPI_ISL_1542664, EPI_ISL_1542665, EPI_ISL_1542666, EPI_ISL_1542667, EPI_ISL_1542668, EPI_ISL_1542669, EPI_ISL_1542670, EPI_ISL_1542671, EPI_ISL_1542672, EPI_ISL_1542673, EPI_ISL_1542674, EPI_ISL_1542675, EPI_ISL_1542676, EPI_ISL_1542677, EPI_ISL_1542678, EPI_ISL_1542679, EPI_ISL_1542680, EPI_ISL_1542681, EPI_ISL_1542682, EPI_ISL_1542683, EPI_ISL_1542684, EPI_ISL_1542685, EPI_ISL_1542686, EPI_ISL_1542687, EPI_ISL_1542688, EPI_ISL_1542689, EPI_ISL_1542690, EPI_ISL_1542691, EPI_ISL_1542692, EPI_ISL_1542693, EPI_ISL_1542694, EPI_ISL_1542695, EPI_ISL_1542696, EPI_ISL_1542697, EPI_ISL_1542698, EPI_ISL_1542699, EPI_ISL_1542700, EPI_ISL_1542701, EPI_ISL_1542702, EPI_ISL_1542703, EPI_ISL_1542704, EPI_ISL_1542705, EPI_ISL_1542706, EPI_ISL_1542707, EPI_ISL_1542708, EPI_ISL_1542709, EPI_ISL_1542710, EPI_ISL_1542711, EPI_ISL_1542712, EPI_ISL_1542713, EPI_ISL_1542714, EPI_ISL_1542715, EPI_ISL_1542716, EPI_ISL_1542717, EPI_ISL_1542718, EPI_ISL_1542719, EPI_ISL_1542720, EPI_ISL_1542721, EPI_ISL_1542722, EPI_ISL_1542723, EPI_ISL_1542724, EPI_ISL_1542725, EPI_ISL_1542726, EPI_ISL_1542727, EPI_ISL_1542728, EPI_ISL_1542729, EPI_ISL_1542730, EPI_ISL_1542731, EPI_ISL_1542732, EPI_ISL_1542733, EPI_ISL_1542734, EPI_ISL_1542735, EPI_ISL_1542736, EPI_ISL_1542737, EPI_ISL_1542738, EPI_ISL_1542739, EPI_ISL_1542740, EPI_ISL_1542741, EPI_ISL_1542742, EPI_ISL_1542743, EPI_ISL_1542744, EPI_ISL_1542745, EPI_ISL_1542746, EPI_ISL_1542747, EPI_ISL_1542748, EPI_ISL_1542749, EPI_ISL_1542750, EPI_ISL_1542751, EPI_ISL_1542752, EPI_ISL_1542753, EPI_ISL_1542754, EPI_ISL_1542755, EPI_ISL_1542756, EPI_ISL_1542757, EPI_ISL_1542758, EPI_ISL_1542759, EPI_ISL_1542760, EPI_ISL_1542761, EPI_ISL_1542762, EPI_ISL_1542763, EPI_ISL_1542764, EPI_ISL_1542765, EPI_ISL_1542766, EPI_ISL_1542767, EPI_ISL_1542768, EPI_ISL_1542769, EPI_ISL_1542770, EPI_ISL_1542771, EPI_ISL_1542772, EPI_ISL_1542773, EPI_ISL_1542774, EPI_ISL_1542775, EPI_ISL_1542776, EPI_ISL_1542777, EPI_ISL_1542778, EPI_ISL_1542779, EPI_ISL_1542780, EPI_ISL_1542781, EPI_ISL_1542782, EPI_ISL_1542783, EPI_ISL_1542784, EPI_ISL_1542785, EPI_ISL_1542786, EPI_ISL_1542787, EPI_ISL_1542788, EPI_ISL_1542789, EPI_ISL_1542790, EPI_ISL_1542791, EPI_ISL_1542792, EPI_ISL_1542793, EPI_ISL_1542794, EPI_ISL_1542795, EPI_ISL_1542796, EPI_ISL_1542797, EPI_ISL_1542798, EPI_ISL_1542799, EPI_ISL_1542800, EPI_ISL_1542801, EPI_ISL_1542802, EPI_ISL_1542803, EPI_ISL_1542804, EPI_ISL_1542805, EPI_ISL_1542806, EPI_ISL_1542807, EPI_ISL_1542808, EPI_ISL_1542809, EPI_ISL_1542810, EPI_ISL_1542811, EPI_ISL_1542812, EPI_ISL_1542813, EPI_ISL_1542814, EPI_ISL_1542815, EPI_ISL_1542816, EPI_ISL_1542817, EPI_ISL_1542818, EPI_ISL_1542819, EPI_ISL_1542820, EPI_ISL_1542821, EPI_ISL_1542822, EPI_ISL_1542823, EPI_ISL_1542824, EPI_ISL_1542825, EPI_ISL_1542826, EPI_ISL_1542827, EPI_ISL_1542828, EPI_ISL_1542829, EPI_ISL_1542830, EPI_ISL_1542831, EPI_ISL_1542832, EPI_ISL_1542833, EPI_ISL_1542834, EPI_ISL_1542835, EPI_ISL_1542836, EPI_ISL_1542837, EPI_ISL_1542838, EPI_ISL_1542839, EPI_ISL_1542840, EPI_ISL_1542841, EPI_ISL_1542842, EPI_ISL_1542843, EPI_ISL_1542844, EPI_ISL_1542845, EPI_ISL_1542846, EPI_ISL_1542847, EPI_ISL_1542848, EPI_ISL_1542849, EPI_ISL_1542850, EPI_ISL_1542851, EPI_ISL_1542852, EPI_ISL_1542853, EPI_ISL_1542854, EPI_ISL_1542855, EPI_ISL_1542856, EPI_ISL_1542857, EPI_ISL_1542858, EPI_ISL_1542859, EPI_ISL_1542860, EPI_ISL_1542861, EPI_ISL_1542862, EPI_ISL_1542863, EPI_ISL_1542864, EPI_ISL_1542865, EPI_ISL_1542866, EPI_ISL_1542867, EPI_ISL_1542868, EPI_ISL_1542869, EPI_ISL_1542870, EPI_ISL_1542871, EPI_ISL_1542872, EPI_ISL_1542873, EPI_ISL_1542874, EPI_ISL_1542875, EPI_ISL_1542876, EPI_ISL_1542877, EPI_ISL_1542878, EPI_ISL_1542879, EPI_ISL_1542880, EPI_ISL_1542881, EPI_ISL_1542882, EPI_ISL_1542883, EPI_ISL_1542884, EPI_ISL_1542885, EPI_ISL_1542886, EPI_ISL_1542887, EPI_ISL_1542888, EPI_ISL_1542889, EPI_ISL_1542890, EPI_ISL_1542891, EPI_ISL_1542892, EPI_ISL_1542893, EPI_ISL_1542894, EPI_ISL_1542895, EPI_ISL_1542896, EPI_ISL_1542897, EPI_ISL_1542898, EPI_ISL_1542899, EPI_ISL_1542900, EPI_ISL_1542901, EPI_ISL_1542902, EPI_ISL_1542903, EPI_ISL_1542904, EPI_ISL_1542905, EPI_ISL_1542906, EPI_ISL_1542907, EPI_ISL_1542908, EPI_ISL_1542909, EPI_ISL_1542910, EPI_ISL_1542911, EPI_ISL_1542912, EPI_ISL_1542913, EPI_ISL_1542914, EPI_ISL_1542915, EPI_ISL_1542916, EPI_ISL_1542917, EPI_ISL_1542919, EPI_ISL_1542920, EPI_ISL_1542921, EPI_ISL_1542922, EPI_ISL_1542923, EPI_ISL_1542924, EPI_ISL_1542925, EPI_ISL_1542926, EPI_ISL_1542927, EPI_ISL_1542928, EPI_ISL_1542929, EPI_ISL_1542930, EPI_ISL_1542931, EPI_ISL_1542932, EPI_ISL_1542933, EPI_ISL_1542934, EPI_ISL_1542935, EPI_ISL_1542936, EPI_ISL_1542937, EPI_ISL_1542938, EPI_ISL_1542939, EPI_ISL_1542940, EPI_ISL_1542941, EPI_ISL_1542942, EPI_ISL_1542943, EPI_ISL_1542944, EPI_ISL_1542945, EPI_ISL_1542946, EPI_ISL_1542947, EPI_ISL_1542948, EPI_ISL_1542949, EPI_ISL_1542950, EPI_ISL_1542951, EPI_ISL_1542952, EPI_ISL_1542953, EPI_ISL_1542954, EPI_ISL_1542955, EPI_ISL_1542956, EPI_ISL_1542957, EPI_ISL_1542958, EPI_ISL_1542959, EPI_ISL_1542960, EPI_ISL_1542961, EPI_ISL_1542962, EPI_ISL_1542963, EPI_ISL_1542964, EPI_ISL_1542965, EPI_ISL_1542966, EPI_ISL_1542967, EPI_ISL_1542968, EPI_ISL_1542969, EPI_ISL_1542970, EPI_ISL_1542971, EPI_ISL_1542972, EPI_ISL_1542973, EPI_ISL_1542974, EPI_ISL_1542975, EPI_ISL_1542976, EPI_ISL_1542977, EPI_ISL_1542978, EPI_ISL_1542979, EPI_ISL_1542980, EPI_ISL_1542981, EPI_ISL_1542982, EPI_ISL_1542983, EPI_ISL_1542984, EPI_ISL_1542985, EPI_ISL_1542986, EPI_ISL_1542987, EPI_ISL_1542988, EPI_ISL_1542989, EPI_ISL_1542990, EPI_ISL_1542991, EPI_ISL_1542992, EPI_ISL_1542993, EPI_ISL_1542994, EPI_ISL_1542995, EPI_ISL_1542996, EPI_ISL_1542997, EPI_ISL_1542998, EPI_ISL_1542999, EPI_ISL_1543000, EPI_ISL_1543001, EPI_ISL_1543002, EPI_ISL_1543003, EPI_ISL_1543004, EPI_ISL_1543005, EPI_ISL_1543006, EPI_ISL_1543007, EPI_ISL_1543008, EPI_ISL_1543009, EPI_ISL_1543010, EPI_ISL_1543011, EPI_ISL_1543012, EPI_ISL_1543013, EPI_ISL_1543014, EPI_ISL_1543015, EPI_ISL_1543016, EPI_ISL_1543017, EPI_ISL_1543018, EPI_ISL_1543019, EPI_ISL_1543020, EPI_ISL_1543021, EPI_ISL_1543022, EPI_ISL_1543023, EPI_ISL_1543024, EPI_ISL_1543025, EPI_ISL_1543026, EPI_ISL_1543027, EPI_ISL_1543028, EPI_ISL_1543029, EPI_ISL_1543030, EPI_ISL_1543031, EPI_ISL_1543032, EPI_ISL_1543033, EPI_ISL_1543034, EPI_ISL_1543035, EPI_ISL_1543036, EPI_ISL_1543037, EPI_ISL_1543038, EPI_ISL_1543039, EPI_ISL_1543040, EPI_ISL_1543041, EPI_ISL_1543042, EPI_ISL_1543043, EPI_ISL_1543044, EPI_ISL_1543045, EPI_ISL_1543046, EPI_ISL_1543047, EPI_ISL_1543048, EPI_ISL_1543049, EPI_ISL_1543050, EPI_ISL_1543051, EPI_ISL_1543052, EPI_ISL_1543053, EPI_ISL_1543054, EPI_ISL_1543055, EPI_ISL_1543056, EPI_ISL_1543057, EPI_ISL_1543058, EPI_ISL_1543059, EPI_ISL_1543060, EPI_ISL_1543061, EPI_ISL_1543062, EPI_ISL_1543063, EPI_ISL_1543064, EPI_ISL_1543065, EPI_ISL_1543066, EPI_ISL_1543067, EPI_ISL_1543068, EPI_ISL_1543069, EPI_ISL_1543070, EPI_ISL_1543071, EPI_ISL_1543072, EPI_ISL_1543073, EPI_ISL_1543074, EPI_ISL_1543075, EPI_ISL_1543076, EPI_ISL_1543077, EPI_ISL_1543078, EPI_ISL_1543079, EPI_ISL_1543080, EPI_ISL_1543081, EPI_ISL_1543082, EPI_ISL_1543083, EPI_ISL_1543084, EPI_ISL_1543085, EPI_ISL_1543086, EPI_ISL_1543087, EPI_ISL_1543088, EPI_ISL_1543089, EPI_ISL_1543090, EPI_ISL_1543091, EPI_ISL_1543092, EPI_ISL_1543093, EPI_ISL_1543094, EPI_ISL_1543095, EPI_ISL_1543096, EPI_ISL_1543097, EPI_ISL_1543098, EPI_ISL_1543099, EPI_ISL_1543100, EPI_ISL_1543101, EPI_ISL_1543102, EPI_ISL_1543103, EPI_ISL_1543104, EPI_ISL_1543105, EPI_ISL_1543106, EPI_ISL_1543107, EPI_ISL_1543108, EPI_ISL_1543109, EPI_ISL_1543110, EPI_ISL_1543111, EPI_ISL_1543112, EPI_ISL_1543113, EPI_ISL_1543114, EPI_ISL_1543115, EPI_ISL_1543116, EPI_ISL_1543117, EPI_ISL_1543118, EPI_ISL_1543119, EPI_ISL_1543120, EPI_ISL_1543121, EPI_ISL_1543122, EPI_ISL_1543123, EPI_ISL_1543124, EPI_ISL_1543125, EPI_ISL_1543126, EPI_ISL_1543127, EPI_ISL_1543128, EPI_ISL_1543129, EPI_ISL_1543130, EPI_ISL_1543131, EPI_ISL_1543132, EPI_ISL_1543133, EPI_ISL_1543134, EPI_ISL_1543135, EPI_ISL_1543136, EPI_ISL_1543137, EPI_ISL_1543138, EPI_ISL_1543139, EPI_ISL_1543140, EPI_ISL_1543141, EPI_ISL_1543142, EPI_ISL_1543143, EPI_ISL_1543144, EPI_ISL_1543145, EPI_ISL_1543146, EPI_ISL_1543147, EPI_ISL_1543148, EPI_ISL_1543149, EPI_ISL_1543150, EPI_ISL_1543151, EPI_ISL_1543152, EPI_ISL_1543153, EPI_ISL_1543154, EPI_ISL_1543155, EPI_ISL_1543156, EPI_ISL_1543157, EPI_ISL_1543158, EPI_ISL_1543159, EPI_ISL_1543160, EPI_ISL_1543161, EPI_ISL_1543162, EPI_ISL_1543163, EPI_ISL_1543164, EPI_ISL_1543165, EPI_ISL_1543166, EPI_ISL_1543167, EPI_ISL_1543168, EPI_ISL_1543169, EPI_ISL_1543170, EPI_ISL_1543171, EPI_ISL_1543172, EPI_ISL_1543173, EPI_ISL_1543174, EPI_ISL_1543175, EPI_ISL_1543176, EPI_ISL_1543177, EPI_ISL_1543178, EPI_ISL_1543179, | | | |

| | | | |
|---|---|---|---|
| EPI_ISL_1543180, EPI_ISL_1543181, EPI_ISL_1543182, EPI_ISL_1543183, EPI_ISL_1543184, EPI_ISL_1543185, EPI_ISL_1543186, EPI_ISL_1543187, EPI_ISL_1543188, EPI_ISL_1543189, EPI_ISL_1543190, EPI_ISL_1543191, EPI_ISL_1543192, EPI_ISL_1543193, EPI_ISL_1543194, EPI_ISL_1543195, EPI_ISL_1543196, EPI_ISL_1543197, EPI_ISL_1543198, EPI_ISL_1543199, EPI_ISL_1543200, EPI_ISL_1543201, EPI_ISL_1543202, EPI_ISL_1543203, EPI_ISL_1543204, EPI_ISL_1543205, EPI_ISL_1543206, EPI_ISL_1543207, EPI_ISL_1543208, EPI_ISL_1543209, EPI_ISL_1543210, EPI_ISL_1543211, EPI_ISL_1543212, EPI_ISL_1543213, EPI_ISL_1543214, EPI_ISL_1543215, EPI_ISL_1543216, EPI_ISL_1543217, EPI_ISL_1543218, EPI_ISL_1543219, EPI_ISL_1543220, EPI_ISL_1543221, EPI_ISL_1543222, EPI_ISL_1543223, EPI_ISL_1543224, EPI_ISL_1543225, EPI_ISL_1543226, EPI_ISL_1543227, EPI_ISL_1543228, EPI_ISL_1543229, EPI_ISL_1543230, EPI_ISL_1543231, EPI_ISL_1543232, EPI_ISL_1543233, EPI_ISL_1543234, EPI_ISL_1543235, EPI_ISL_1543236, EPI_ISL_1543237, EPI_ISL_1543238, EPI_ISL_1543239, EPI_ISL_1543240, EPI_ISL_1543241, EPI_ISL_1543242, EPI_ISL_1543243, EPI_ISL_1543244, EPI_ISL_1543245, EPI_ISL_1543246, EPI_ISL_1543247, EPI_ISL_1543248, EPI_ISL_1543249, EPI_ISL_1543250, EPI_ISL_1543251, EPI_ISL_1543252, EPI_ISL_1543253, EPI_ISL_1543254, EPI_ISL_1543255, EPI_ISL_1543256, EPI_ISL_1543257, EPI_ISL_1543258, EPI_ISL_1543259, EPI_ISL_1543260, EPI_ISL_1543261, EPI_ISL_1543262, EPI_ISL_1543263, EPI_ISL_1543264, EPI_ISL_1543265, EPI_ISL_1543266, EPI_ISL_1543267, EPI_ISL_1543268, EPI_ISL_1543269, EPI_ISL_1543270, EPI_ISL_1543271, EPI_ISL_1543272, EPI_ISL_1543273, EPI_ISL_1543274, EPI_ISL_1543275, EPI_ISL_1543276, EPI_ISL_1543277, EPI_ISL_1543278, EPI_ISL_1543279, EPI_ISL_1543280, EPI_ISL_1543281, EPI_ISL_1543282, EPI_ISL_1543283, EPI_ISL_1543284, EPI_ISL_1543285, EPI_ISL_1543286, EPI_ISL_1543287, EPI_ISL_1543288, EPI_ISL_1543289, EPI_ISL_1543290, EPI_ISL_1543291, EPI_ISL_1543292, EPI_ISL_1543293, EPI_ISL_1543294, EPI_ISL_1543295, EPI_ISL_1543296, EPI_ISL_1543297, EPI_ISL_1543298, EPI_ISL_1543299, EPI_ISL_1543300, EPI_ISL_1543301, EPI_ISL_1543302, EPI_ISL_1543303, EPI_ISL_1543304, EPI_ISL_1543305, EPI_ISL_1543306, EPI_ISL_1543307, EPI_ISL_1543308, EPI_ISL_1543309, EPI_ISL_1543310, EPI_ISL_1543311, EPI_ISL_1543312, EPI_ISL_1543313, EPI_ISL_1543314, EPI_ISL_1543315, EPI_ISL_1543316, EPI_ISL_1543317, EPI_ISL_1543318, EPI_ISL_1543319, EPI_ISL_1543320, EPI_ISL_1543321, EPI_ISL_1543322, EPI_ISL_1543323, EPI_ISL_1543324, EPI_ISL_1543325, EPI_ISL_1543326, EPI_ISL_1543327, EPI_ISL_1543328, EPI_ISL_1543329, EPI_ISL_1543330, EPI_ISL_1543331, EPI_ISL_1543332, EPI_ISL_1543333, EPI_ISL_1543334, EPI_ISL_1543335, EPI_ISL_1543336, EPI_ISL_1543337, EPI_ISL_1543338, EPI_ISL_1543339, EPI_ISL_1543340, EPI_ISL_1543341, EPI_ISL_1543342, EPI_ISL_1543343, EPI_ISL_1543344, EPI_ISL_1543345, EPI_ISL_1543346, EPI_ISL_1543347, EPI_ISL_1543348, EPI_ISL_1543349, EPI_ISL_1543350, EPI_ISL_1543351, EPI_ISL_1543352, EPI_ISL_1543353, EPI_ISL_1543354, EPI_ISL_1543355, EPI_ISL_1543356, EPI_ISL_1543357, EPI_ISL_1543358, EPI_ISL_1543359, EPI_ISL_1543360, EPI_ISL_1543361, EPI_ISL_1543362, EPI_ISL_1543363, EPI_ISL_1543364, EPI_ISL_1543365, EPI_ISL_1543366, EPI_ISL_1543367, EPI_ISL_1543368, EPI_ISL_1543369, EPI_ISL_1543370, EPI_ISL_1543371, EPI_ISL_1543372, EPI_ISL_1543373, EPI_ISL_1543374, EPI_ISL_1543375, EPI_ISL_1543376, EPI_ISL_1543377, EPI_ISL_1543378, EPI_ISL_1543379, EPI_ISL_1543380, EPI_ISL_1543381, EPI_ISL_1543382, EPI_ISL_1543383, EPI_ISL_1543384, EPI_ISL_1543385, EPI_ISL_1543386, EPI_ISL_1543387, EPI_ISL_1543388, EPI_ISL_1543389, EPI_ISL_1543390, EPI_ISL_1543391, EPI_ISL_1543392, EPI_ISL_1543393, EPI_ISL_1543394, EPI_ISL_1543395, EPI_ISL_1543396, EPI_ISL_1543397, EPI_ISL_1543398, EPI_ISL_1543399, EPI_ISL_1543400, EPI_ISL_1543401, EPI_ISL_1543402, EPI_ISL_1543403, EPI_ISL_1543404, EPI_ISL_1543405, EPI_ISL_1543406, EPI_ISL_1543407, EPI_ISL_1543408, EPI_ISL_1543409, EPI_ISL_1543410, EPI_ISL_1543411, EPI_ISL_1543412, EPI_ISL_1543413, EPI_ISL_1543414, EPI_ISL_1543415, EPI_ISL_1543416, EPI_ISL_1543417, EPI_ISL_1543418, EPI_ISL_1543419, EPI_ISL_1543420, EPI_ISL_1543421, EPI_ISL_1543422, EPI_ISL_1543423, EPI_ISL_1543424, EPI_ISL_1543425, EPI_ISL_1543426, EPI_ISL_1543427, EPI_ISL_1543428, EPI_ISL_1543429, EPI_ISL_1543430, EPI_ISL_1543431, EPI_ISL_1543432, EPI_ISL_1543433, EPI_ISL_1543434, EPI_ISL_1543435, EPI_ISL_1543436, EPI_ISL_1543437, EPI_ISL_1543438, EPI_ISL_1543439, EPI_ISL_1543440, EPI_ISL_1543441, EPI_ISL_1543442, EPI_ISL_1543443, EPI_ISL_1543444, EPI_ISL_1543445, EPI_ISL_1543446, EPI_ISL_1543447, EPI_ISL_1543448, EPI_ISL_1543449, EPI_ISL_1543450, EPI_ISL_1543451, EPI_ISL_1543452, EPI_ISL_1543453, EPI_ISL_1543454, EPI_ISL_1543455, EPI_ISL_1543456, EPI_ISL_1543457, EPI_ISL_1543458, EPI_ISL_1543459, EPI_ISL_1543460, EPI_ISL_1543461, EPI_ISL_1543462, EPI_ISL_1543463, EPI_ISL_1543464, EPI_ISL_1543465, EPI_ISL_1543466, EPI_ISL_1543467, EPI_ISL_1543468, EPI_ISL_1543469, EPI_ISL_1543470, EPI_ISL_1543471, EPI_ISL_1543472, EPI_ISL_1543473, EPI_ISL_1543474, EPI_ISL_1543475, EPI_ISL_1543476, EPI_ISL_1543477, EPI_ISL_1543478, EPI_ISL_1543479, EPI_ISL_1543480, EPI_ISL_1543481, EPI_ISL_1543482, EPI_ISL_1543483, EPI_ISL_1543484, EPI_ISL_1543485, EPI_ISL_1543486, EPI_ISL_1543487, EPI_ISL_1543488, EPI_ISL_1543489, EPI_ISL_1543490, EPI_ISL_1543491, EPI_ISL_1543492, EPI_ISL_1543493, EPI_ISL_1543494, EPI_ISL_1543495, EPI_ISL_1543496, EPI_ISL_1543497, EPI_ISL_1543498, EPI_ISL_1543499, EPI_ISL_1543500, EPI_ISL_1543501, EPI_ISL_1543502, EPI_ISL_1543503, EPI_ISL_1543504, EPI_ISL_1543505, EPI_ISL_1543506, EPI_ISL_1543507, EPI_ISL_1543508, EPI_ISL_1543509, EPI_ISL_1543510, EPI_ISL_1543511, EPI_ISL_1543512, EPI_ISL_1543513, EPI_ISL_1543514, EPI_ISL_1543515, EPI_ISL_1543516, EPI_ISL_1543517, EPI_ISL_1543518, EPI_ISL_1543519, EPI_ISL_1543520, EPI_ISL_1543521, EPI_ISL_1543522, EPI_ISL_1543523, EPI_ISL_1543524, EPI_ISL_1543525, EPI_ISL_1543526, EPI_ISL_1543527, EPI_ISL_1543528, EPI_ISL_1543529, EPI_ISL_1543530, EPI_ISL_1543531, EPI_ISL_1543532, EPI_ISL_1543533, EPI_ISL_1543534, EPI_ISL_1543535, EPI_ISL_1543536, EPI_ISL_1543537, EPI_ISL_1543538, EPI_ISL_1543539, EPI_ISL_1543540, EPI_ISL_1543541, EPI_ISL_1543542, EPI_ISL_1543543, EPI_ISL_1543544, EPI_ISL_1543545, EPI_ISL_1543546, EPI_ISL_1543547, EPI_ISL_1543548, EPI_ISL_1543549, EPI_ISL_1543550, EPI_ISL_1543551, EPI_ISL_1543552, EPI_ISL_1543553, EPI_ISL_1543554, EPI_ISL_1543555, EPI_ISL_1543556, EPI_ISL_1543557, EPI_ISL_1543558, EPI_ISL_1543559, EPI_ISL_1543560, EPI_ISL_1543561, EPI_ISL_1543562, EPI_ISL_1543563, EPI_ISL_1543564, EPI_ISL_1543565, EPI_ISL_1543566, EPI_ISL_1543567, EPI_ISL_1543568, EPI_ISL_1543569, EPI_ISL_1543570, EPI_ISL_1543571, EPI_ISL_1543572, EPI_ISL_1543573, EPI_ISL_1543574, EPI_ISL_1543575, EPI_ISL_1543576, EPI_ISL_1543577, EPI_ISL_1543578, EPI_ISL_1543579, EPI_ISL_1543580, EPI_ISL_1543581, EPI_ISL_1543582, EPI_ISL_1543583, EPI_ISL_1543584, EPI_ISL_1543585, EPI_ISL_1543586, EPI_ISL_1543587, EPI_ISL_1543588, EPI_ISL_1543589, EPI_ISL_1543590, EPI_ISL_1543591, EPI_ISL_1543592, EPI_ISL_1543593, EPI_ISL_1543594, EPI_ISL_1543595, EPI_ISL_1543596, EPI_ISL_1543597, EPI_ISL_1543598, EPI_ISL_1543599, EPI_ISL_1543600, EPI_ISL_1543601, EPI_ISL_1543602, EPI_ISL_1543603, EPI_ISL_1543604, EPI_ISL_1543605, EPI_ISL_1543606, EPI_ISL_1543607, EPI_ISL_1543608, EPI_ISL_1543609, EPI_ISL_1543610, EPI_ISL_1543611, EPI_ISL_1543612, EPI_ISL_1543613, EPI_ISL_1543614, EPI_ISL_1543615, EPI_ISL_1543616, EPI_ISL_1543617, EPI_ISL_1543618, EPI_ISL_1543619, EPI_ISL_1543620, EPI_ISL_1543621, EPI_ISL_1543622, EPI_ISL_1543623, EPI_ISL_1543624, EPI_ISL_1543625, EPI_ISL_1543626, EPI_ISL_1543627, EPI_ISL_1543628, EPI_ISL_1543629, EPI_ISL_1543630, EPI_ISL_1543631, EPI_ISL_1543632, EPI_ISL_1543633, EPI_ISL_1543634, EPI_ISL_1543635, EPI_ISL_1543636, EPI_ISL_1543637, EPI_ISL_1543638, EPI_ISL_1543639, EPI_ISL_1543640, EPI_ISL_1543641, EPI_ISL_1543642, EPI_ISL_1543643, EPI_ISL_1543644, EPI_ISL_1543645, EPI_ISL_1543646, EPI_ISL_1543647, EPI_ISL_1543648, EPI_ISL_1543649, EPI_ISL_1543650, EPI_ISL_1543651, EPI_ISL_1543652, EPI_ISL_1543653, EPI_ISL_1543654, EPI_ISL_1543655, EPI_ISL_1543656, EPI_ISL_1543657, EPI_ISL_1543658, EPI_ISL_1543659, EPI_ISL_1543660, EPI_ISL_1543661, EPI_ISL_1543662, EPI_ISL_1543663, EPI_ISL_1543664, EPI_ISL_1543665, EPI_ISL_1543666, EPI_ISL_1543667, EPI_ISL_1543668, EPI_ISL_1543669, EPI_ISL_1543670, EPI_ISL_1543671, EPI_ISL_1543672, EPI_ISL_1543673, EPI_ISL_1543674, EPI_ISL_1543675, EPI_ISL_1543676, EPI_ISL_1543677, EPI_ISL_1543678, EPI_ISL_1543679, EPI_ISL_1543680, EPI_ISL_1543681, EPI_ISL_1543682, EPI_ISL_1543683, EPI_ISL_1543684, EPI_ISL_1543685, EPI_ISL_1543686, EPI_ISL_1543687, EPI_ISL_1543688, EPI_ISL_1543689, EPI_ISL_1543690, EPI_ISL_1543691, EPI_ISL_1543692, EPI_ISL_1543693, EPI_ISL_1543694, EPI_ISL_1543695, EPI_ISL_1543696, EPI_ISL_1543697, EPI_ISL_1543698, EPI_ISL_1543699, EPI_ISL_1543700, EPI_ISL_1543701, EPI_ISL_1543702, EPI_ISL_1543703, EPI_ISL_1543704, EPI_ISL_1543705, EPI_ISL_1543706, EPI_ISL_1543707, EPI_ISL_1543708, EPI_ISL_1543709, EPI_ISL_1543710, EPI_ISL_1543711, EPI_ISL_1543712, EPI_ISL_1543713, EPI_ISL_1543714, EPI_ISL_1543715, EPI_ISL_1543716, EPI_ISL_1543717, EPI_ISL_1543718, EPI_ISL_1543719, EPI_ISL_1543720, EPI_ISL_1543721, EPI_ISL_1543722, EPI_ISL_1543723, EPI_ISL_1543724, EPI_ISL_1543725, EPI_ISL_1543726, EPI_ISL_1543727, EPI_ISL_1543728, EPI_ISL_1543729, EPI_ISL_1543730, EPI_ISL_1543731, EPI_ISL_1543732, EPI_ISL_1543733, EPI_ISL_1543734, EPI_ISL_1543735, EPI_ISL_1543736, EPI_ISL_1543737, EPI_ISL_1543738, EPI_ISL_1543739, EPI_ISL_1543740, EPI_ISL_1543741, EPI_ISL_1543742, EPI_ISL_1543743, EPI_ISL_1543744, EPI_ISL_1543745, EPI_ISL_1543746, EPI_ISL_1543747, EPI_ISL_1543748, EPI_ISL_1543749, EPI_ISL_1543750, EPI_ISL_1543751, EPI_ISL_1543752, EPI_ISL_1543753, EPI_ISL_1543754, EPI_ISL_1543755, EPI_ISL_1543756, EPI_ISL_1543757, EPI_ISL_1543758, EPI_ISL_1543759, EPI_ISL_1543760, EPI_ISL_1543761, EPI_ISL_1543762, EPI_ISL_1543763, EPI_ISL_1543764, EPI_ISL_1543765, EPI_ISL_1543766, EPI_ISL_1543767, EPI_ISL_1543768, EPI_ISL_1543769, EPI_ISL_1543770, EPI_ISL_1543771, EPI_ISL_1543772, EPI_ISL_1543773, EPI_ISL_1543774, EPI_ISL_1543775, EPI_ISL_1543776, EPI_ISL_1543777, EPI_ISL_1543778, EPI_ISL_1543779, EPI_ISL_1543780, EPI_ISL_1543781, EPI_ISL_1543782, EPI_ISL_1543783, EPI_ISL_1543784, EPI_ISL_1543785, EPI_ISL_1543786, EPI_ISL_1543787, EPI_ISL_1543788, EPI_ISL_1543789, EPI_ISL_1543790, EPI_ISL_1543791, EPI_ISL_1543792, EPI_ISL_1543793, EPI_ISL_1543794, EPI_ISL_1543795, EPI_ISL_1543796, EPI_ISL_1543797, EPI_ISL_1543798, EPI_ISL_1543799, EPI_ISL_1543800, EPI_ISL_1543801, EPI_ISL_1543802, EPI_ISL_1543803, EPI_ISL_1543804, EPI_ISL_1543805, EPI_ISL_1543806, EPI_ISL_1543807, EPI_ISL_1543808, EPI_ISL_1543809, EPI_ISL_1543810, EPI_ISL_1543811, EPI_ISL_1543812, EPI_ISL_1543813, EPI_ISL_1543814, EPI_ISL_1543815, EPI_ISL_1543816, EPI_ISL_1543817, EPI_ISL_1543818, EPI_ISL_1543819, EPI_ISL_1543820, EPI_ISL_1543821, EPI_ISL_1543822, EPI_ISL_1543823, EPI_ISL_1543824, EPI_ISL_1543825, EPI_ISL_1543826, EPI_ISL_1543827, EPI_ISL_1543828, EPI_ISL_1543829, EPI_ISL_1543830, EPI_ISL_1543831, EPI_ISL_1543832, EPI_ISL_1543833, EPI_ISL_1543834, EPI_ISL_1543835, EPI_ISL_1543836, EPI_ISL_1543837, EPI_ISL_1543838, EPI_ISL_1543839, EPI_ISL_1543840, EPI_ISL_1543841, EPI_ISL_1543842, EPI_ISL_1543843, EPI_ISL_1543844, EPI_ISL_1543845, EPI_ISL_1543846, EPI_ISL_1543847, EPI_ISL_1543848, EPI_ISL_1543849, EPI_ISL_1543850, EPI_ISL_1543851, EPI_ISL_1543852, EPI_ISL_1543853, EPI_ISL_1543854, EPI_ISL_1543855, EPI_ISL_1543856, EPI_ISL_1543857, EPI_ISL_1543858, EPI_ISL_1543859, EPI_ISL_1543860, EPI_ISL_1543861, EPI_ISL_1543862, EPI_ISL_1543863, EPI_ISL_1543864, EPI_ISL_1543865, EPI_ISL_1543866, EPI_ISL_1543867, EPI_ISL_1543868, EPI_ISL_1543869, EPI_ISL_1543870, EPI_ISL_1543871, EPI_ISL_1543872, EPI_ISL_1543873, EPI_ISL_1543874, EPI_ISL_1543875, EPI_ISL_1543876, EPI_ISL_1543877, EPI_ISL_1543878, EPI_ISL_1543879, EPI_ISL_1543880, EPI_ISL_1543881, EPI_ISL_1543882, EPI_ISL_1543883, EPI_ISL_1543884, EPI_ISL_1543885, EPI_ISL_1543886, EPI_ISL_1543887, EPI_ISL_1543888, EPI_ISL_1543889, EPI_ISL_1543890, EPI_ISL_1543891, EPI_ISL_1543892, EPI_ISL_1543893, EPI_ISL_1543894, EPI_ISL_1543895, EPI_ISL_1543896, EPI_ISL_1543897, EPI_ISL_1543898, EPI_ISL_1543899, EPI_ISL_1543900, EPI_ISL_1543901, EPI_ISL_1543902, EPI_ISL_1543903, EPI_ISL_1543904, EPI_ISL_1543905, EPI_ISL_1543906, EPI_ISL_1543907, EPI_ISL_1543908, EPI_ISL_1543909, EPI_ISL_1543910, EPI_ISL_1543911, EPI_ISL_1543912, EPI_ISL_1543913, EPI_ISL_1543914, EPI_ISL_1543915, EPI_ISL_1543916, EPI_ISL_1543917, EPI_ISL_1543918, EPI_ISL_1543919, EPI_ISL_1543920 | | | |
| see above | Pandemic Response Lab - NYC | Pandemic Response Lab, R&D | Henry Lee, Michael Hammerling, Melissa Hopkins, Cybill del Castillo, Shinyoung Clair Kang, William Ward, Pradeep Bugga, Sol Rey, Dylan Law, Katharine Nelson, Haiping Hao, Jon Laurent |
| EPI_ISL_1548152, EPI_ISL_1548166, EPI_ISL_1548207, EPI_ISL_1548208, EPI_ISL_1548210, EPI_ISL_1548211, EPI_ISL_1548213, EPI_ISL_1548235, EPI_ISL_1548237, EPI_ISL_1548241, EPI_ISL_1548242, EPI_ISL_1548248, EPI_ISL_1548249, EPI_ISL_1548250, EPI_ISL_1548260, EPI_ISL_1548262, EPI_ISL_1548375, EPI_ISL_1548382, EPI_ISL_1548384, EPI_ISL_1548438, EPI_ISL_1548439, EPI_ISL_1548440, EPI_ISL_1548441, EPI_ISL_1548442, EPI_ISL_1548445, EPI_ISL_1548446, EPI_ISL_1548447, EPI_ISL_1548474, EPI_ISL_1548475, EPI_ISL_1548476, EPI_ISL_1548477, EPI_ISL_1548478, EPI_ISL_1548479, EPI_ISL_1548506, EPI_ISL_1548507, EPI_ISL_1548508, EPI_ISL_1548509, EPI_ISL_1548952, EPI_ISL_1548979, EPI_ISL_1548986, EPI_ISL_1548993, EPI_ISL_1549000, EPI_ISL_1549017, EPI_ISL_1549019, EPI_ISL_1549024, EPI_ISL_1549032, EPI_ISL_1549033, EPI_ISL_1549034, EPI_ISL_1549038, EPI_ISL_1549044, EPI_ISL_1549045, EPI_ISL_1549054, EPI_ISL_1549063, EPI_ISL_1549064, EPI_ISL_1549065, EPI_ISL_1549066, EPI_ISL_1549067, EPI_ISL_1549068, EPI_ISL_1549080, EPI_ISL_1549081, EPI_ISL_1549082, EPI_ISL_1549102, EPI_ISL_1549106, EPI_ISL_1549173, EPI_ISL_1549376, EPI_ISL_1549377, EPI_ISL_1549378, EPI_ISL_1549379, EPI_ISL_1549380, EPI_ISL_1549558, EPI_ISL_1549564, EPI_ISL_1549632, EPI_ISL_1549635, EPI_ISL_1549645, EPI_ISL_1549650, EPI_ISL_1549674, EPI_ISL_1549758, EPI_ISL_1549760, EPI_ISL_1549768, EPI_ISL_1549769, EPI_ISL_1549771, EPI_ISL_1549776, EPI_ISL_1549777, EPI_ISL_1549779, EPI_ISL_1549790, EPI_ISL_1549791, EPI_ISL_1549919, EPI_ISL_1549940, EPI_ISL_1549942, EPI_ISL_1549943, EPI_ISL_1550039, EPI_ISL_1550040 | | | |
| see above | Laboratory Corporation of America | Centers for Disease Control and Prevention Division of Viral Diseases, Pathogen Discovery | Dakota Howard, Dhwani Batra, Peter W. Cook, Kara Moser, Adrian Paskey, Jason Caravas, Benjamin Rambo-Martin, Shatavia Morrison, Christopher Gulvick, Scott Sammons, Yvette Unoarumhi, Darlene Wagner, Matthew Schmerer, Minoo Agarwal, Eyad Almasri, Debbie Boles, Ayla Burns, Nuthawin Charoensri, Oren Cohen, Susan Countryman, Mary Ann Cristobal, Bobbi Croy, Suzanne Dale, Hrushikesh Deshmukh, Amanda Douglas, Vincent Drouillon, Marcia Eisenberg, Howard Engler, Rama Ghatti, Prashant Gupta, Susan Hicks, Jake Humphrey, Lax Iyer, Manoj Jain, Mohan Kolli, Brian Krueger, Tim Kuphal, Stanley Letovsky, Michael Levandoski, Craig Lukasik, Jonathan Meltzer, Brian Norvell, Mindy Nye, Scott Parker, Christos Petropoulos, John Pruitt, Steven Ragan, Scott Ryan, Mike Sapeta, Jana Schroth, Suresh Babu Selvaraju, Goran Stevovic, Amanda Suchanek, Andrea Throop, Lyndon Tilson, Thomas Urban, Joe Voshell, Kimberly Wagner, Jonathan Williams, Mary Williamson, Qian Zeng, Tricia Zwiefelhofer, Clinton R. Paden, Duncan MacCannell |
| EPI_ISL_1550397, EPI_ISL_1550503, EPI_ISL_1550549, EPI_ISL_1550668, EPI_ISL_1550684, EPI_ISL_1550694, EPI_ISL_1550820, EPI_ISL_1550871, EPI_ISL_1550989, EPI_ISL_1551037, EPI_ISL_1551052, EPI_ISL_1551082, EPI_ISL_1551091, EPI_ISL_1551103, EPI_ISL_1551116, EPI_ISL_1551213, EPI_ISL_1551330, EPI_ISL_1551343, EPI_ISL_1551344, EPI_ISL_1551390, EPI_ISL_1551457, EPI_ISL_1551499, EPI_ISL_1551502, EPI_ISL_1551504, EPI_ISL_1551511, EPI_ISL_1551512, EPI_ISL_1551612, EPI_ISL_1551626 | | | |
| see above | Aegis Sciences Corporation | Centers for Disease Control and Prevention Division of Viral Diseases, Pathogen Discovery | Dakota Howard, Dhwani Batra, Peter W. Cook, Kara Moser, Adrian Paskey, Jason Caravas, Benjamin Rambo-Martin, Shatavia Morrison, Christopher Gulvick, Scott Sammons, Yvette Unoarumhi, Darlene Wagner, Matthew Schmerer, Cyndi Clark, Patrick Campbell, Rob Case, Vikramsinha Ghorpade, Holly Houdeshell, Ola Kvalvaag, Dillon Nall, Ethan Sanders, Alec Vest, Shaun Westlund, Matthew Hardison, Clinton R. Paden, Duncan MacCannell |
| EPI_ISL_1551720, EPI_ISL_1551723, EPI_ISL_1551725, EPI_ISL_1551741, EPI_ISL_1551746, EPI_ISL_1551766, EPI_ISL_1551772 | Quest Diagnostics Incorporated | Centers for Disease Control and Prevention Division of Viral Diseases, Pathogen Discovery | Peter W. Cook, Dakota Howard, Dhwani Batra, Ben L. Rambo-Martin, S. H. Rosenthal, A. Gerasimova, R. M. Kagan, B. Anderson, M. Hua, Y. Liu, L.E. Bernstein, K.E. Livingston, A. Perez, I. A. Shlyakhter, R. V. Rolando, R. Owen, P. Tanpaiboon, F. Lacbawan, Clinton R. Paden, Suxiang Tong, Duncan MacCannell |
| EPI_ISL_1552235, EPI_ISL_1552259, EPI_ISL_1552275, EPI_ISL_1552278, EPI_ISL_1552301, EPI_ISL_1552713, EPI_ISL_1552918, EPI_ISL_1553024, EPI_ISL_1553038, EPI_ISL_1553044, EPI_ISL_1553049, EPI_ISL_1553053, EPI_ISL_1553056, EPI_ISL_1553061, EPI_ISL_1553064, EPI_ISL_1553069, EPI_ISL_1553070, EPI_ISL_1553071, EPI_ISL_1553075, EPI_ISL_1553076, EPI_ISL_1553080, EPI_ISL_1553082, EPI_ISL_1553085, EPI_ISL_1553086, EPI_ISL_1553087, EPI_ISL_1553092, EPI_ISL_1553093 | | | |
| see above | Quest Diagnostics Incorporated | Centers for Disease Control and Prevention Division of Viral Diseases, Pathogen Discovery | Dakota Howard, Dhwani Batra, Peter W. Cook, Kara Moser, Adrian Paskey, Jason Caravas, Benjamin Rambo-Martin, Shatavia Morrison, Christopher Gulvick, Scott Sammons, Yvette Unoarumhi, Darlene Wagner, Matthew Schmerer, S. H. Rosenthal, A. Gerasimova, R. M. Kagan, B. Anderson, M. Hua, Y. Liu, L.E. Bernstein, K.E. Livingston, A. Perez, I. A. Shlyakhter, R. V. Rolando, R. Owen, P. Tanpaiboon, F. Lacbawan, Clinton R. Paden, Duncan MacCannell |
| EPI_ISL_1553421, EPI_ISL_1553555, EPI_ISL_1554114, EPI_ISL_1554306 | Helix/Illumina | Centers for Disease Control and Prevention Division of Viral Diseases, Pathogen Discovery | Dakota Howard, Dhwani Batra, Peter W. Cook, Kara Moser, Adrian Paskey, Jason Caravas, Benjamin Rambo-Martin, Shatavia Morrison, Christopher Gulvick, Scott Sammons, Yvette Unoarumhi, Darlene Wagner, Matthew Schmerer, Eileen de Feo, Jan Antico, Christine Tran, Matthew Tolentino, Shannon Wickline, Kim Gietzen, Brad Sickler, Jingtao Liu, Eric Allen, Phil Febbo, Nicole L. Washington, Simon White, Geraint Levan, Kelly Schiabor Barrett, Elizabeth Cirulli, Alexandre Bolze, Ary Ascencio, Charlotte Rivera-Garcia, Ryan Cho, Jason Nguyen, Sherry Wang, Jimmy Ramirez, Tyler Cassens, Efren Sandoval, Magnus Isaksson, William Lee, David Becker, Marc Laurent, James Lu, Clinton R. Paden, Duncan MacCannell |
| EPI_ISL_1559420, EPI_ISL_1559430, EPI_ISL_1559442, EPI_ISL_1559557 | Quest Diagnostics Incorporated | Centers for Disease Control and Prevention Division of Viral Diseases, Pathogen Discovery | Dakota Howard, Dhwani Batra, Peter W. Cook, Kara Moser, Adrian Paskey, Jason Caravas, Benjamin Rambo-Martin, Shatavia Morrison, Christopher Gulvick, Scott Sammons, Yvette Unoarumhi, Darlene Wagner, Matthew Schmerer, S. H. Rosenthal, A. Gerasimova, R. M. Kagan, B. Anderson, M. Hua, Y. Liu, L.E. Bernstein, K.E. Livingston, A. Perez, I. A. Shlyakhter, R. V. Rolando, R. Owen, P. Tanpaiboon, F. Lacbawan, Clinton R. Paden, Duncan |

| | | | |
|---|---|---|---|
| | | | MacCannell |
| EPI_ISL_1559638, EPI_ISL_1559639, EPI_ISL_1559646, EPI_ISL_1559688, EPI_ISL_1559691, EPI_ISL_1559698, EPI_ISL_1559704, EPI_ISL_1559705, EPI_ISL_1559715, EPI_ISL_1559717, EPI_ISL_1559751, EPI_ISL_1559791, EPI_ISL_1559800, EPI_ISL_1559817, EPI_ISL_1559826, EPI_ISL_1559884, EPI_ISL_1559888, EPI_ISL_1559890, EPI_ISL_1559892, EPI_ISL_1559893, EPI_ISL_1559920, EPI_ISL_1559929, EPI_ISL_1559946, EPI_ISL_1559947, EPI_ISL_1559948, EPI_ISL_1559949, EPI_ISL_1560017, EPI_ISL_1560022, EPI_ISL_1560024, EPI_ISL_1560048, EPI_ISL_1560049, EPI_ISL_1560071, EPI_ISL_1560186, EPI_ISL_1560187, EPI_ISL_1560199, EPI_ISL_1560244, EPI_ISL_1560265, EPI_ISL_1560269, EPI_ISL_1560287, EPI_ISL_1560291, EPI_ISL_1560358, EPI_ISL_1560402, EPI_ISL_1560403, EPI_ISL_1560462, EPI_ISL_1560493, EPI_ISL_1560494, EPI_ISL_1560667, EPI_ISL_1560668, EPI_ISL_1560698, EPI_ISL_1560855, EPI_ISL_1560859, EPI_ISL_1560871, EPI_ISL_1560875, EPI_ISL_1560901, EPI_ISL_1560902, EPI_ISL_1560907, EPI_ISL_1560968, EPI_ISL_1560971, EPI_ISL_1561025, EPI_ISL_1561031, EPI_ISL_1561124, EPI_ISL_1561127, EPI_ISL_1561228, EPI_ISL_1561230, EPI_ISL_1561257, EPI_ISL_1561259, EPI_ISL_1561270, EPI_ISL_1561273, EPI_ISL_1561274, EPI_ISL_1561275, EPI_ISL_1561276, EPI_ISL_1561292, EPI_ISL_1561293, EPI_ISL_1561388, EPI_ISL_1561408, EPI_ISL_1561415, EPI_ISL_1561424, EPI_ISL_1561503, EPI_ISL_1561512, EPI_ISL_1561513, EPI_ISL_1561532, EPI_ISL_1561653, EPI_ISL_1561654, EPI_ISL_1561655, EPI_ISL_1561670, EPI_ISL_1561671, EPI_ISL_1561672, EPI_ISL_1561691, EPI_ISL_1561720, EPI_ISL_1561732, EPI_ISL_1561743, EPI_ISL_1561766, EPI_ISL_1561771, EPI_ISL_1561775, EPI_ISL_1561776, EPI_ISL_1561777, EPI_ISL_1561778, EPI_ISL_1561835, EPI_ISL_1561837, EPI_ISL_1561839, EPI_ISL_1561840, EPI_ISL_1561843, EPI_ISL_1561854, EPI_ISL_1561865, EPI_ISL_1561890, EPI_ISL_1561905, EPI_ISL_1561909, EPI_ISL_1561913, EPI_ISL_1562066, EPI_ISL_1562068, EPI_ISL_1562150, EPI_ISL_1562154, EPI_ISL_1562220, EPI_ISL_1562234, EPI_ISL_1562245, EPI_ISL_1562246, EPI_ISL_1562251, EPI_ISL_1562275, EPI_ISL_1562276, EPI_ISL_1562293, EPI_ISL_1562303, EPI_ISL_1562316, EPI_ISL_1562369, EPI_ISL_1562370, EPI_ISL_1562372, EPI_ISL_1562417, EPI_ISL_1562452, EPI_ISL_1562496, EPI_ISL_1562507, EPI_ISL_1562510, EPI_ISL_1562545, EPI_ISL_1562555, EPI_ISL_1562569, EPI_ISL_1562609, EPI_ISL_1562614, EPI_ISL_1562615, EPI_ISL_1562622, EPI_ISL_1562623, EPI_ISL_1562625, EPI_ISL_1562752, EPI_ISL_1562784, EPI_ISL_1562785, EPI_ISL_1562786, EPI_ISL_1562787, EPI_ISL_1562788, EPI_ISL_1562789, EPI_ISL_1562814, EPI_ISL_1562937, EPI_ISL_1562949, EPI_ISL_1562952, EPI_ISL_1562956, EPI_ISL_1562959, EPI_ISL_1562968, EPI_ISL_1562969, EPI_ISL_1562994, EPI_ISL_1563070, EPI_ISL_1563156, EPI_ISL_1563184, EPI_ISL_1563241, EPI_ISL_1563249, EPI_ISL_1563295, EPI_ISL_1563299, EPI_ISL_1563330, EPI_ISL_1563332, EPI_ISL_1563355, EPI_ISL_1563356, EPI_ISL_1563377, EPI_ISL_1563381, EPI_ISL_1563578, EPI_ISL_1563587, EPI_ISL_1563593, EPI_ISL_1563606, EPI_ISL_1563617, EPI_ISL_1563618 | | | |
| see above | Aegis Sciences Corporation | Centers for Disease Control and Prevention Division of Viral Diseases, Pathogen Discovery | Dakota Howard, Dhwani Batra, Peter W. Cook, Kara Moser, Adrian Paskey, Jason Caravas, Benjamin Rambo-Martin, Shatavia Morrison, Christopher Gulvick, Scott Sammons, Yvette Unoarumhi, Darlene Wagner, Matthew Schmerer, Cyndi Clark, Patrick Campbell, Rob Case, Vikramsinha Ghorpade, Holly Houdeshell, Ola Kvalvaag, Dillon Nall, Ethan Sanders, Alec Vest, Shaun Westlund, Matthew Hardison, Clinton R. Paden, Duncan MacCannell |
| EPI_ISL_1575445 | SUNY UPSTATE MEDICAL UNIVERSITY | Wadsworth Center, New York State Department of Health | Kirsten St. George, Daryl M. Lamson, Alexis Russell, Matthew Shudt, Melissa A Leisner, Jonathan Plitnick, Catharine Prussing, Navjot Singh, John Kelly, Erasmus Schneider, Erica Lasek-Nesselquist |
| EPI_ISL_1575446, EPI_ISL_1575447, EPI_ISL_1575448 | URMC LABS | Wadsworth Center, New York State Department of Health | Kirsten St. George, Daryl M. Lamson, Alexis Russell, Matthew Shudt, Melissa A Leisner, Jonathan Plitnick, Catharine Prussing, Navjot Singh, John Kelly, Erasmus Schneider, Erica Lasek-Nesselquist |
| EPI_ISL_1575449 | NORTHWELL HEALTH LABORATORIES | Wadsworth Center, New York State Department of Health | Kirsten St. George, Daryl M. Lamson, Alexis Russell, Matthew Shudt, Melissa A Leisner, Jonathan Plitnick, Catharine Prussing, Navjot Singh, John Kelly, Erasmus Schneider, Erica Lasek-Nesselquist |
| EPI_ISL_1575450, EPI_ISL_1575451, EPI_ISL_1575452 | URMC LABS | Wadsworth Center, New York State Department of Health | Kirsten St. George, Daryl M. Lamson, Alexis Russell, Matthew Shudt, Melissa A Leisner, Jonathan Plitnick, Catharine Prussing, Navjot Singh, John Kelly, Erasmus Schneider, Erica Lasek-Nesselquist |
| EPI_ISL_1575453, EPI_ISL_1575454 | WESTCHESTER MEDICAL CENTER | Wadsworth Center, New York State Department of Health | Kirsten St. George, Daryl M. Lamson, Alexis Russell, Matthew Shudt, Melissa A Leisner, Jonathan Plitnick, Catharine Prussing, Navjot Singh, John Kelly, Erasmus Schneider, Erica Lasek-Nesselquist |
| EPI_ISL_1575455, EPI_ISL_1575456, EPI_ISL_1575457, EPI_ISL_1575458 | URMC LABS | Wadsworth Center, New York State Department of Health | Kirsten St. George, Daryl M. Lamson, Alexis Russell, Matthew Shudt, Melissa A Leisner, Jonathan Plitnick, Catharine Prussing, Navjot Singh, John Kelly, Erasmus Schneider, Erica Lasek-Nesselquist |
| EPI_ISL_1575459 | NYC Pandemic Response Lab | Wadsworth Center, New York State Department of Health | Kirsten St. George, Daryl M. Lamson, Alexis Russell, Matthew Shudt, Melissa A Leisner, Jonathan Plitnick, Catharine Prussing, Navjot Singh, John Kelly, Erasmus Schneider, Erica Lasek-Nesselquist |
| EPI_ISL_1575460, EPI_ISL_1575461, EPI_ISL_1575462, EPI_ISL_1575463, EPI_ISL_1575464, EPI_ISL_1575465, EPI_ISL_1575466, EPI_ISL_1575467, EPI_ISL_1575468 | URMC LABS | Wadsworth Center, New York State Department of Health | Kirsten St. George, Daryl M. Lamson, Alexis Russell, Matthew Shudt, Melissa A Leisner, Jonathan Plitnick, Catharine Prussing, Navjot Singh, John Kelly, Erasmus Schneider, Erica Lasek-Nesselquist |
| EPI_ISL_1575469 | NORTHWELL HEALTH LABORATORIES | Wadsworth Center, New York State Department of Health | Kirsten St. George, Daryl M. Lamson, Alexis Russell, Matthew Shudt, Melissa A Leisner, Jonathan Plitnick, Catharine Prussing, Navjot Singh, John Kelly, Erasmus Schneider, Erica Lasek-Nesselquist |
| EPI_ISL_1575470 | MONTEFIORE MEDICAL CENTER LABORATORIES | Wadsworth Center, New York State Department of Health | Kirsten St. George, Daryl M. Lamson, Alexis Russell, Matthew Shudt, Melissa A Leisner, Jonathan Plitnick, Catharine Prussing, Navjot Singh, John Kelly, Erasmus Schneider, Erica Lasek-Nesselquist |
| EPI_ISL_1575471 | TEMPUS LABS INC | Wadsworth Center, New York State Department of Health | Kirsten St. George, Daryl M. Lamson, Alexis Russell, Matthew Shudt, Melissa A Leisner, Jonathan Plitnick, Catharine Prussing, Navjot Singh, John Kelly, Erasmus Schneider, Erica Lasek-Nesselquist |
| EPI_ISL_1575472 | Columbia University Irving Medical Center | Wadsworth Center, New York State Department of Health | Kirsten St. George, Daryl M. Lamson, Alexis Russell, Matthew Shudt, Melissa A Leisner, Jonathan Plitnick, Catharine Prussing, Navjot Singh, John Kelly, Erasmus Schneider, Erica Lasek-Nesselquist |
| EPI_ISL_1575473, EPI_ISL_1575474, EPI_ISL_1575475, EPI_ISL_1575476 | URMC LABS | Wadsworth Center, New York State Department of Health | Kirsten St. George, Daryl M. Lamson, Alexis Russell, Matthew Shudt, Melissa A Leisner, Jonathan Plitnick, Catharine Prussing, Navjot Singh, John Kelly, Erasmus Schneider, Erica Lasek-Nesselquist |
| EPI_ISL_1575477 | BOSTON HEART DIAGNOSTICS CORP | Wadsworth Center, New York State Department of Health | Kirsten St. George, Daryl M. Lamson, Alexis Russell, Matthew Shudt, Melissa A Leisner, Jonathan Plitnick, Catharine Prussing, Navjot Singh, John Kelly, Erasmus Schneider, Erica Lasek-Nesselquist |
| EPI_ISL_1575478 | NORTHWELL HEALTH LABORATORIES | Wadsworth Center, New York State Department of Health | Kirsten St. George, Daryl M. Lamson, Alexis Russell, Matthew Shudt, Melissa A Leisner, Jonathan Plitnick, Catharine Prussing, Navjot Singh, John Kelly, Erasmus Schneider, Erica Lasek-Nesselquist |
| EPI_ISL_1575510, EPI_ISL_1575846, EPI_ISL_1575913, EPI_ISL_1576513, EPI_ISL_1576617, EPI_ISL_1576668 | Helix/Illumina | Centers for Disease Control and Prevention Division of Viral Diseases, Pathogen Discovery | Dakota Howard, Dhwani Batra, Peter W. Cook, Kara Moser, Adrian Paskey, Jason Caravas, Benjamin Rambo-Martin, Shatavia Morrison, Christopher Gulvick, Scott Sammons, Yvette Unoarumhi, Darlene Wagner, Matthew Schmerer, Eileen de Feo, Jan Antico, Christine Tran, Matthew Tolentino, Shannon Wickline, Kim Gietzen, Brad Sickler, Jingtao Liu, Eric Allen, Phil Febbo, Nicole L. Washington, Simon White, Geraint Levan, Kelly Schiabor Barrett, Elizabeth Cirulli, Alexandre Bolze, Ary Ascencio, Charlotte Rivera-Garcia, Ryan Cho, Jason Nguyen, Sherry Wang, Jimmy Ramirez, Tyler Cassens, Efren Sandoval, Magnus Isaksson, William Lee, David Becker, Marc Laurent, James Lu, Clinton R. Paden, Duncan MacCannell |
| EPI_ISL_1576812, EPI_ISL_1576813 | SUNY UPSTATE MEDICAL UNIVERSITY | Wadsworth Center, New York State Department of Health | Kirsten St. George, Daryl M. Lamson, Alexis Russell, Matthew Shudt, Melissa A Leisner, Jonathan Plitnick, Catharine Prussing, Navjot Singh, John Kelly, Erasmus Schneider, Erica Lasek-Nesselquist |
| EPI_ISL_1576814 | TEMPUS LABS INC | Wadsworth Center, New York State Department of Health | Kirsten St. George, Daryl M. Lamson, Alexis Russell, Matthew Shudt, Melissa A Leisner, Jonathan Plitnick, Catharine Prussing, Navjot Singh, John Kelly, Erasmus Schneider, Erica Lasek-Nesselquist |
| EPI_ISL_1576815 | NORTHWELL HEALTH LABORATORIES | Wadsworth Center, New York State Department of Health | Kirsten St. George, Daryl M. Lamson, Alexis Russell, Matthew Shudt, Melissa A Leisner, Jonathan Plitnick, Catharine Prussing, Navjot Singh, John Kelly, Erasmus Schneider, Erica Lasek-Nesselquist |
| EPI_ISL_1576816 | MONTEFIORE MEDICAL CENTER LABORATORIES | Wadsworth Center, New York State Department of Health | Kirsten St. George, Daryl M. Lamson, Alexis Russell, Matthew Shudt, Melissa A Leisner, Jonathan Plitnick, Catharine Prussing, Navjot Singh, John Kelly, Erasmus Schneider, Erica Lasek-Nesselquist |
| EPI_ISL_1576817, EPI_ISL_1576818 | URMC LABS | Wadsworth Center, New York State Department of Health | Kirsten St. George, Daryl M. Lamson, Alexis Russell, Matthew Shudt, Melissa A Leisner, Jonathan Plitnick, Catharine Prussing, Navjot Singh, John Kelly, Erasmus Schneider, Erica Lasek-Nesselquist |
| EPI_ISL_1576819 | MONTEFIORE MEDICAL CENTER LABORATORIES | Wadsworth Center, New York State Department of Health | Kirsten St. George, Daryl M. Lamson, Alexis Russell, Matthew Shudt, Melissa A Leisner, Jonathan Plitnick, Catharine Prussing, Navjot Singh, John Kelly, Erasmus Schneider, Erica Lasek-Nesselquist |
| EPI_ISL_1576820 | Columbia University Irving Medical Center | Wadsworth Center, New York State Department of Health | Kirsten St. George, Daryl M. Lamson, Alexis Russell, Matthew Shudt, Melissa A Leisner, Jonathan Plitnick, Catharine Prussing, Navjot Singh, John Kelly, Erasmus Schneider, Erica Lasek-Nesselquist |
| EPI_ISL_1576821, EPI_ISL_1576822 | TEMPUS LABS INC | Wadsworth Center, New York State Department of Health | Kirsten St. George, Daryl M. Lamson, Alexis Russell, Matthew Shudt, Melissa A Leisner, Jonathan Plitnick, Catharine Prussing, Navjot Singh, John Kelly, Erasmus Schneider, Erica Lasek-Nesselquist |
| EPI_ISL_1576823 | URMC LABS | Wadsworth Center, New York State Department of Health | Kirsten St. George, Daryl M. Lamson, Alexis Russell, Matthew Shudt, Melissa A Leisner, Jonathan Plitnick, Catharine Prussing, Navjot Singh, John Kelly, Erasmus Schneider, Erica Lasek-Nesselquist |
| EPI_ISL_1576824 | ALBANY MEDICAL CENTER | Wadsworth Center, New York State Department of Health | Kirsten St. George, Daryl M. Lamson, Alexis Russell, Matthew Shudt, Melissa A Leisner, Jonathan Plitnick, Catharine Prussing, Navjot Singh, John Kelly, Erasmus Schneider, Erica Lasek-Nesselquist |

| | | | |
|---|---|---|---|
| EPI_ISL_1576825, EPI_ISL_1576826 | NORTHWELL HEALTH LABORATORIES | Wadsworth Center, New York State Department of Health | Kirsten St. George, Daryl M. Lamson, Alexis Russell, Matthew Shudt, Melissa A Leisner, Jonathan Plitnick, Catharine Prussing, Navjot Singh, John Kelly, Erasmus Schneider, Erica Lasek-Nesselquist |
| EPI_ISL_1576862, EPI_ISL_1576863, EPI_ISL_1576864, EPI_ISL_1576865, EPI_ISL_1576866, EPI_ISL_1576867, EPI_ISL_1576868, EPI_ISL_1576869, EPI_ISL_1576870, EPI_ISL_1576871, EPI_ISL_1576872, EPI_ISL_1576873, EPI_ISL_1576874, EPI_ISL_1576875, EPI_ISL_1576876, EPI_ISL_1576877, EPI_ISL_1576878, EPI_ISL_1576879, EPI_ISL_1576880, EPI_ISL_1576881, EPI_ISL_1576882, EPI_ISL_1576883, EPI_ISL_1576884, EPI_ISL_1576885, EPI_ISL_1576886, EPI_ISL_1576887, EPI_ISL_1576888, EPI_ISL_1576889, EPI_ISL_1576890, EPI_ISL_1576891, EPI_ISL_1576892, EPI_ISL_1576893, EPI_ISL_1576894, EPI_ISL_1576895, EPI_ISL_1576896, EPI_ISL_1576897, EPI_ISL_1576898, EPI_ISL_1576899, EPI_ISL_1576900, EPI_ISL_1576901, EPI_ISL_1576902, EPI_ISL_1576904, EPI_ISL_1576906, EPI_ISL_1576907, EPI_ISL_1576908, EPI_ISL_1576909, EPI_ISL_1576910, EPI_ISL_1576911, EPI_ISL_1576912, EPI_ISL_1576913, EPI_ISL_1576914, EPI_ISL_1576915, EPI_ISL_1576916, EPI_ISL_1576917, EPI_ISL_1576918, EPI_ISL_1576919, EPI_ISL_1576920, EPI_ISL_1576921, EPI_ISL_1576922, EPI_ISL_1576923, EPI_ISL_1576924, EPI_ISL_1576925, EPI_ISL_1576926, EPI_ISL_1576927, EPI_ISL_1576928, EPI_ISL_1576929, EPI_ISL_1576930, EPI_ISL_1576931, EPI_ISL_1576932, EPI_ISL_1576934, EPI_ISL_1576935, EPI_ISL_1576936, EPI_ISL_1576937, EPI_ISL_1576938, EPI_ISL_1576939, EPI_ISL_1576940, EPI_ISL_1576941, EPI_ISL_1576942, EPI_ISL_1576944, EPI_ISL_1576945, EPI_ISL_1576946, EPI_ISL_1576948 | | | |
| see above | NYU Langone Health | Departments of Pathology and Medicine, New York University School of Medicine | Adriana Heguy, Dacia Dimartino, Emily Guzman, Christian Marier, Peter Meyn, Sitharam Ramaswami, Gael Westby, Paul Zappile, Yutong Zhang, Paolo Cotzia, Guiqing Wang |
| EPI_ISL_1578133, EPI_ISL_1578152, EPI_ISL_1578170, EPI_ISL_1578181, EPI_ISL_1578201, EPI_ISL_1578202, EPI_ISL_1578280, EPI_ISL_1578316, EPI_ISL_1578318, EPI_ISL_1578319, EPI_ISL_1578320, EPI_ISL_1578321, EPI_ISL_1578325, EPI_ISL_1578334, EPI_ISL_1578374, EPI_ISL_1578386, EPI_ISL_1578396, EPI_ISL_1578398, EPI_ISL_1578401, EPI_ISL_1578405, EPI_ISL_1578406, EPI_ISL_1578432 | | | |
| see above | Broad Institute Clinical Research Sequencing Platform | Infectious Disease Program, Broad Institute of Harvard and MIT | Siddle,K.J., Adams,G., Pearlman,L., Gladden-Young,A., Vicente,G., Blumenstiel,B., DeFelice,M., Lee,M., McGovern,S., Lagerborg,K., Rudy,M., DeRuff,K., Carter,A., Normandin,E., Bauer,M., Reilly,S., Tomkins-Tinch,C., Loreth,C., Chaluvadi,S., Meldrim,J., Granger,B., Lemieux,J.E., Birren,B.W., Sabeti,P.C., Larkin,K., Dodge,S., Lennon,N., Madoff,L., Brown,C., Gallagher,G., Smole,S., Park,D.J., Gabriel,S., and MacInnis,B.L. |
| EPI_ISL_1580667, EPI_ISL_1580668, EPI_ISL_1580669, EPI_ISL_1580670 | BOSTON HEART DIAGNOSTICS CORP | Wadsworth Center, New York State Department of Health | Kirsten St. George, Daryl M. Lamson, Alexis Russell, Matthew Shudt, Melissa A Leisner, Jonathan Plitnick, Catharine Prussing, Navjot Singh, John Kelly, Erasmus Schneider, Erica Lasek-Nesselquist |
| EPI_ISL_1580671, EPI_ISL_1580672 | URMC LABS | Wadsworth Center, New York State Department of Health | Kirsten St. George, Daryl M. Lamson, Alexis Russell, Matthew Shudt, Melissa A Leisner, Jonathan Plitnick, Catharine Prussing, Navjot Singh, John Kelly, Erasmus Schneider, Erica Lasek-Nesselquist |
| EPI_ISL_1580673, EPI_ISL_1580674, EPI_ISL_1580675, EPI_ISL_1580676, EPI_ISL_1580677 | WESTCHESTER MEDICAL CENTER | Wadsworth Center, New York State Department of Health | Kirsten St. George, Daryl M. Lamson, Alexis Russell, Matthew Shudt, Melissa A Leisner, Jonathan Plitnick, Catharine Prussing, Navjot Singh, John Kelly, Erasmus Schneider, Erica Lasek-Nesselquist |
| EPI_ISL_1580678, EPI_ISL_1580679, EPI_ISL_1580680, EPI_ISL_1580681, EPI_ISL_1580682, EPI_ISL_1580683, EPI_ISL_1580684, EPI_ISL_1580685 | SUNY UPSTATE MEDICAL UNIVERSITY | Wadsworth Center, New York State Department of Health | Kirsten St. George, Daryl M. Lamson, Alexis Russell, Matthew Shudt, Melissa A Leisner, Jonathan Plitnick, Catharine Prussing, Navjot Singh, John Kelly, Erasmus Schneider, Erica Lasek-Nesselquist |
| EPI_ISL_1580686 | MONTEFIORE MEDICAL CENTER LABORATORIES | Wadsworth Center, New York State Department of Health | Kirsten St. George, Daryl M. Lamson, Alexis Russell, Matthew Shudt, Melissa A Leisner, Jonathan Plitnick, Catharine Prussing, Navjot Singh, John Kelly, Erasmus Schneider, Erica Lasek-Nesselquist |
| EPI_ISL_1580687, EPI_ISL_1580688, EPI_ISL_1580689, EPI_ISL_1580690, EPI_ISL_1580691, EPI_ISL_1580692, EPI_ISL_1580693, EPI_ISL_1580694, EPI_ISL_1580695, EPI_ISL_1580696, EPI_ISL_1580697, EPI_ISL_1580698 | | | |
| see above | Columbia University Irving Medical Center | Wadsworth Center, New York State Department of Health | Kirsten St. George, Daryl M. Lamson, Alexis Russell, Matthew Shudt, Melissa A Leisner, Jonathan Plitnick, Catharine Prussing, Navjot Singh, John Kelly, Erasmus Schneider, Erica Lasek-Nesselquist |
| EPI_ISL_1580699 | WESTCHESTER MEDICAL CENTER | Wadsworth Center, New York State Department of Health | Kirsten St. George, Daryl M. Lamson, Alexis Russell, Matthew Shudt, Melissa A Leisner, Jonathan Plitnick, Catharine Prussing, Navjot Singh, John Kelly, Erasmus Schneider, Erica Lasek-Nesselquist |
| EPI_ISL_1580700 | ALBANY MEDICAL CENTER | Wadsworth Center, New York State Department of Health | Kirsten St. George, Daryl M. Lamson, Alexis Russell, Matthew Shudt, Melissa A Leisner, Jonathan Plitnick, Catharine Prussing, Navjot Singh, John Kelly, Erasmus Schneider, Erica Lasek-Nesselquist |
| EPI_ISL_1580701, EPI_ISL_1580702, EPI_ISL_1580703, EPI_ISL_1580704, EPI_ISL_1580705, EPI_ISL_1580706, EPI_ISL_1580707, EPI_ISL_1580708, EPI_ISL_1580709, EPI_ISL_1580710, EPI_ISL_1580711, EPI_ISL_1580712, EPI_ISL_1580713, EPI_ISL_1580714, EPI_ISL_1580715, EPI_ISL_1580716, EPI_ISL_1580717, EPI_ISL_1580718, EPI_ISL_1580719, EPI_ISL_1580720, EPI_ISL_1580721, EPI_ISL_1580722 | | | |
| see above | URMC LABS | Wadsworth Center, New York State Department of Health | Kirsten St. George, Daryl M. Lamson, Alexis Russell, Matthew Shudt, Melissa A Leisner, Jonathan Plitnick, Catharine Prussing, Navjot Singh, John Kelly, Erasmus Schneider, Erica Lasek-Nesselquist |
| EPI_ISL_1580723, EPI_ISL_1580724, EPI_ISL_1580725, EPI_ISL_1580726, EPI_ISL_1580727, EPI_ISL_1580728, EPI_ISL_1580729, EPI_ISL_1580730, EPI_ISL_1580731, EPI_ISL_1580732, EPI_ISL_1580733, EPI_ISL_1580734, EPI_ISL_1580735, EPI_ISL_1580736, EPI_ISL_1580737, EPI_ISL_1580738 | | | |
| see above | NORTHWELL HEALTH LABORATORIES | Wadsworth Center, New York State Department of Health | Kirsten St. George, Daryl M. Lamson, Alexis Russell, Matthew Shudt, Melissa A Leisner, Jonathan Plitnick, Catharine Prussing, Navjot Singh, John Kelly, Erasmus Schneider, Erica Lasek-Nesselquist |
| EPI_ISL_1580739, EPI_ISL_1580740 | SARATOGA HOSPITAL LABORATORY | Wadsworth Center, New York State Department of Health | Kirsten St. George, Daryl M. Lamson, Alexis Russell, Matthew Shudt, Melissa A Leisner, Jonathan Plitnick, Catharine Prussing, Navjot Singh, John Kelly, Erasmus Schneider, Erica Lasek-Nesselquist |
| EPI_ISL_1580741 | ALBANY MEDICAL CENTER | Wadsworth Center, New York State Department of Health | Kirsten St. George, Daryl M. Lamson, Alexis Russell, Matthew Shudt, Melissa A Leisner, Jonathan Plitnick, Catharine Prussing, Navjot Singh, John Kelly, Erasmus Schneider, Erica Lasek-Nesselquist |
| EPI_ISL_1580918, EPI_ISL_1581703 | Helix/Illumina | Centers for Disease Control and Prevention Division of Viral Diseases, Pathogen Discovery | Dakota Howard, Dhwani Batra, Peter W. Cook, Kara Moser, Adrian Paskey, Jason Caravas, Benjamin Rambo-Martin, Shatavia Morrison, Christopher Gulvick, Scott Sammons, Yvette Unoarumhi, Darlene Wagner, Matthew Schmerer, Eileen de Feo, Jan Antico, Christine Tran, Matthew Tolentino, Shannon Wickline, Kim Gietzen, Brad Sickler, Jingtao Liu, Eric Allen, Phil Febbo, Nicole L. Washington, Simon White, Geraint Levan, Kelly Schiabor Barrett, Elizabeth Cirulli, Alexandre Bolze, Ary Ascencio, Charlotte Rivera-Garcia, Ryan Cho, Jason Nguyen, Sherry Wang, Jimmy Ramirez, Tyler Cassens, Efren Sandoval, Magnus Isaksson, William Lee, David Becker, Marc Laurent, James Lu, Clinton R. Paden, Duncan MacCannell |
| EPI_ISL_1581871, EPI_ISL_1581898, EPI_ISL_1581931, EPI_ISL_1581934, EPI_ISL_1581962, EPI_ISL_1582034, EPI_ISL_1582039, EPI_ISL_1582051, EPI_ISL_1582058, EPI_ISL_1582066, EPI_ISL_1582100, EPI_ISL_1582109, EPI_ISL_1582116, EPI_ISL_1582119, EPI_ISL_1582133, EPI_ISL_1582142, EPI_ISL_1582148 | | | |
| see above | Quest Diagnostics Incorporated | Centers for Disease Control and Prevention Division of Viral Diseases, Pathogen Discovery | Dakota Howard, Dhwani Batra, Peter W. Cook, Kara Moser, Adrian Paskey, Jason Caravas, Benjamin Rambo-Martin, Shatavia Morrison, Christopher Gulvick, Scott Sammons, Yvette Unoarumhi, Darlene Wagner, Matthew Schmerer, S. H. Rosenthal, A. Gerasimova, R. M. Kagan, B. Anderson, M. Hua, Y. Liu, L.E. Bernstein, K.E. Livingston, A. Perez, I. A. Shlyakhter, R. V. Rolando, R. Owen, P. Tanpaiboon, F. Lacbawan, Clinton R. Paden, Duncan MacCannell |
| EPI_ISL_1582149 | SUNY UPSTATE MEDICAL UNIVERSITY | Wadsworth Center, New York State Department of Health | Kirsten St. George, Daryl M. Lamson, Alexis Russell, Matthew Shudt, Melissa A Leisner, Jonathan Plitnick, Catharine Prussing, Navjot Singh, John Kelly, Erasmus Schneider, Erica Lasek-Nesselquist |
| EPI_ISL_1582151 | URMC LABS | Wadsworth Center, New York State Department of Health | Kirsten St. George, Daryl M. Lamson, Alexis Russell, Matthew Shudt, Melissa A Leisner, Jonathan Plitnick, Catharine Prussing, Navjot Singh, John Kelly, Erasmus Schneider, Erica Lasek-Nesselquist |
| EPI_ISL_1582154 | ALBANY MEDICAL CENTER | Wadsworth Center, New York State Department of Health | Kirsten St. George, Daryl M. Lamson, Alexis Russell, Matthew Shudt, Melissa A Leisner, Jonathan Plitnick, Catharine Prussing, Navjot Singh, John Kelly, Erasmus Schneider, Erica Lasek-Nesselquist |
| EPI_ISL_1582156 | NORTHWELL HEALTH LABORATORIES | Wadsworth Center, New York State Department of Health | Kirsten St. George, Daryl M. Lamson, Alexis Russell, Matthew Shudt, Melissa A Leisner, Jonathan Plitnick, Catharine Prussing, Navjot Singh, John Kelly, Erasmus Schneider, Erica Lasek-Nesselquist |
| EPI_ISL_1582159 | MONTEFIORE MEDICAL CENTER LABORATORIES | Wadsworth Center, New York State Department of Health | Kirsten St. George, Daryl M. Lamson, Alexis Russell, Matthew Shudt, Melissa A Leisner, Jonathan Plitnick, Catharine Prussing, Navjot Singh, John Kelly, Erasmus Schneider, Erica Lasek-Nesselquist |
| EPI_ISL_1582161 | Quest Diagnostics Incorporated | Centers for Disease Control and Prevention Division of Viral Diseases, Pathogen Discovery | Dakota Howard, Dhwani Batra, Peter W. Cook, Kara Moser, Adrian Paskey, Jason Caravas, Benjamin Rambo-Martin, Shatavia Morrison, Christopher Gulvick, Scott Sammons, Yvette Unoarumhi, Darlene Wagner, Matthew Schmerer, S. H. Rosenthal, A. Gerasimova, R. M. Kagan, B. Anderson, M. Hua, Y. Liu, L.E. Bernstein, K.E. Livingston, A. Perez, I. A. Shlyakhter, R. V. Rolando, R. Owen, P. Tanpaiboon, F. Lacbawan, Clinton R. Paden, Duncan MacCannell |
| EPI_ISL_1582162, EPI_ISL_1582165, EPI_ISL_1582168, EPI_ISL_1582170, | MONTEFIORE MEDICAL CENTER LABORATORIES | Wadsworth Center, New York State Department of Health | Kirsten St. George, Daryl M. Lamson, Alexis Russell, Matthew Shudt, Melissa A Leisner, Jonathan Plitnick, Catharine Prussing, Navjot Singh, John Kelly, Erasmus Schneider, Erica Lasek-Nesselquist |

| | | | |
|---|---|---|---|
| EPI_ISL_1582173, EPI_ISL_1582174, EPI_ISL_1582175, EPI_ISL_1582176, EPI_ISL_1582177, EPI_ISL_1582178 | | | |
| EPI_ISL_1582179, EPI_ISL_1582180, EPI_ISL_1582181, EPI_ISL_1582182, EPI_ISL_1582183, EPI_ISL_1582184, EPI_ISL_1582185, EPI_ISL_1582186, EPI_ISL_1582187, EPI_ISL_1582188, EPI_ISL_1582189, EPI_ISL_1582190, EPI_ISL_1582191, EPI_ISL_1582192, EPI_ISL_1582193, EPI_ISL_1582194, EPI_ISL_1582195, EPI_ISL_1582196, EPI_ISL_1582197, EPI_ISL_1582198, EPI_ISL_1582199, EPI_ISL_1582200, EPI_ISL_1582201, EPI_ISL_1582202, EPI_ISL_1582203, EPI_ISL_1582204, EPI_ISL_1582205, EPI_ISL_1582206, EPI_ISL_1582207, EPI_ISL_1582208, EPI_ISL_1582209, EPI_ISL_1582210, EPI_ISL_1582211, EPI_ISL_1582212, EPI_ISL_1582213 | | | |
| see above | NYC Pandemic Response Lab | Wadsworth Center, New York State Department of Health | Kirsten St. George, Daryl M. Lamson, Alexis Russell, Matthew Shudt, Melissa A Leisner, Jonathan Plitnick, Catharine Prussing, Navjot Singh, John Kelly, Erasmus Schneider, Erica Lasek-Nesselquist |
| EPI_ISL_1582214, EPI_ISL_1582215, EPI_ISL_1582216, EPI_ISL_1582217 | Columbia University Irving Medical Center | Wadsworth Center, New York State Department of Health | Kirsten St. George, Daryl M. Lamson, Alexis Russell, Matthew Shudt, Melissa A Leisner, Jonathan Plitnick, Catharine Prussing, Navjot Singh, John Kelly, Erasmus Schneider, Erica Lasek-Nesselquist |
| EPI_ISL_1582218 | ALBANY MEDICAL CENTER | Wadsworth Center, New York State Department of Health | Kirsten St. George, Daryl M. Lamson, Alexis Russell, Matthew Shudt, Melissa A Leisner, Jonathan Plitnick, Catharine Prussing, Navjot Singh, John Kelly, Erasmus Schneider, Erica Lasek-Nesselquist |
| EPI_ISL_1582219, EPI_ISL_1582220, EPI_ISL_1582221, EPI_ISL_1582222, EPI_ISL_1582223, EPI_ISL_1582224 | URMC LABS | Wadsworth Center, New York State Department of Health | Kirsten St. George, Daryl M. Lamson, Alexis Russell, Matthew Shudt, Melissa A Leisner, Jonathan Plitnick, Catharine Prussing, Navjot Singh, John Kelly, Erasmus Schneider, Erica Lasek-Nesselquist |
| EPI_ISL_1582225, EPI_ISL_1582226 | NORTHWELL HEALTH LABORATORIES | Wadsworth Center, New York State Department of Health | Kirsten St. George, Daryl M. Lamson, Alexis Russell, Matthew Shudt, Melissa A Leisner, Jonathan Plitnick, Catharine Prussing, Navjot Singh, John Kelly, Erasmus Schneider, Erica Lasek-Nesselquist |
| EPI_ISL_1582227, EPI_ISL_1582228, EPI_ISL_1582229 | SARATOGA HOSPITAL LABORATORY | Wadsworth Center, New York State Department of Health | Kirsten St. George, Daryl M. Lamson, Alexis Russell, Matthew Shudt, Melissa A Leisner, Jonathan Plitnick, Catharine Prussing, Navjot Singh, John Kelly, Erasmus Schneider, Erica Lasek-Nesselquist |
| EPI_ISL_1582230 | NYC Pandemic Response Lab | Wadsworth Center, New York State Department of Health | Kirsten St. George, Daryl M. Lamson, Alexis Russell, Matthew Shudt, Melissa A Leisner, Jonathan Plitnick, Catharine Prussing, Navjot Singh, John Kelly, Erasmus Schneider, Erica Lasek-Nesselquist |
| EPI_ISL_1582231 | WESTCHESTER MEDICAL CENTER | Wadsworth Center, New York State Department of Health | Kirsten St. George, Daryl M. Lamson, Alexis Russell, Matthew Shudt, Melissa A Leisner, Jonathan Plitnick, Catharine Prussing, Navjot Singh, John Kelly, Erasmus Schneider, Erica Lasek-Nesselquist |
| EPI_ISL_1582232, EPI_ISL_1582233, EPI_ISL_1582234, EPI_ISL_1582235, EPI_ISL_1582236, EPI_ISL_1582237, EPI_ISL_1582238, EPI_ISL_1582239, EPI_ISL_1582240, EPI_ISL_1582241, EPI_ISL_1582242, EPI_ISL_1582243, EPI_ISL_1582244 | | | |
| see above | TEMPUS LABS INC | Wadsworth Center, New York State Department of Health | Kirsten St. George, Daryl M. Lamson, Alexis Russell, Matthew Shudt, Melissa A Leisner, Jonathan Plitnick, Catharine Prussing, Navjot Singh, John Kelly, Erasmus Schneider, Erica Lasek-Nesselquist |
| EPI_ISL_1582245, EPI_ISL_1582246, EPI_ISL_1582247, EPI_ISL_1582248, EPI_ISL_1582249, EPI_ISL_1582250, EPI_ISL_1582251, EPI_ISL_1582252, EPI_ISL_1582253, EPI_ISL_1582254 | STONY BROOK UNIVERSITY HOSPITAL | Wadsworth Center, New York State Department of Health | Kirsten St. George, Daryl M. Lamson, Alexis Russell, Matthew Shudt, Melissa A Leisner, Jonathan Plitnick, Catharine Prussing, Navjot Singh, John Kelly, Erasmus Schneider, Erica Lasek-Nesselquist |
| EPI_ISL_1582255 | GOOD SAMARITAN HOSPITAL LABORATORY | Wadsworth Center, New York State Department of Health | Kirsten St. George, Daryl M. Lamson, Alexis Russell, Matthew Shudt, Melissa A Leisner, Jonathan Plitnick, Catharine Prussing, Navjot Singh, John Kelly, Erasmus Schneider, Erica Lasek-Nesselquist |
| EPI_ISL_1582256, EPI_ISL_1582257, EPI_ISL_1582258, EPI_ISL_1582259 | NORTH SHORE UNIVERSITY HOSPITAL | Wadsworth Center, New York State Department of Health | Kirsten St. George, Daryl M. Lamson, Alexis Russell, Matthew Shudt, Melissa A Leisner, Jonathan Plitnick, Catharine Prussing, Navjot Singh, John Kelly, Erasmus Schneider, Erica Lasek-Nesselquist |
| EPI_ISL_1582260 | ALBANY MEDICAL CENTER | Wadsworth Center, New York State Department of Health | Kirsten St. George, Daryl M. Lamson, Alexis Russell, Matthew Shudt, Melissa A Leisner, Jonathan Plitnick, Catharine Prussing, Navjot Singh, John Kelly, Erasmus Schneider, Erica Lasek-Nesselquist |
| EPI_ISL_1582261, EPI_ISL_1582262, EPI_ISL_1582263, EPI_ISL_1582264, EPI_ISL_1582265, EPI_ISL_1582266, EPI_ISL_1582267, EPI_ISL_1582268, EPI_ISL_1582269, EPI_ISL_1582270, EPI_ISL_1582271 | | | |
| see above | Columbia University Irving Medical Center | Wadsworth Center, New York State Department of Health | Kirsten St. George, Daryl M. Lamson, Alexis Russell, Matthew Shudt, Melissa A Leisner, Jonathan Plitnick, Catharine Prussing, Navjot Singh, John Kelly, Erasmus Schneider, Erica Lasek-Nesselquist |
| EPI_ISL_1582272 | ST. FRANCIS HOSPITAL LABORATORY | Wadsworth Center, New York State Department of Health | Kirsten St. George, Daryl M. Lamson, Alexis Russell, Matthew Shudt, Melissa A Leisner, Jonathan Plitnick, Catharine Prussing, Navjot Singh, John Kelly, Erasmus Schneider, Erica Lasek-Nesselquist |
| EPI_ISL_1582273, EPI_ISL_1582274, EPI_ISL_1582275, EPI_ISL_1582276, EPI_ISL_1582277, EPI_ISL_1582278, EPI_ISL_1582279, EPI_ISL_1582280, EPI_ISL_1582281, EPI_ISL_1582282 | WHITE PLAINS HOSPITAL CENTER LABORATORY | Wadsworth Center, New York State Department of Health | Kirsten St. George, Daryl M. Lamson, Alexis Russell, Matthew Shudt, Melissa A Leisner, Jonathan Plitnick, Catharine Prussing, Navjot Singh, John Kelly, Erasmus Schneider, Erica Lasek-Nesselquist |
| EPI_ISL_1582283, EPI_ISL_1582284, EPI_ISL_1582285, EPI_ISL_1582286, EPI_ISL_1582287, EPI_ISL_1582288, EPI_ISL_1582289, EPI_ISL_1582290, EPI_ISL_1582291, EPI_ISL_1582292, EPI_ISL_1582293, EPI_ISL_1582294, EPI_ISL_1582295, EPI_ISL_1582296, EPI_ISL_1582297, EPI_ISL_1582298, EPI_ISL_1582299, EPI_ISL_1582300, EPI_ISL_1582301, EPI_ISL_1582302, EPI_ISL_1582303, EPI_ISL_1582304, EPI_ISL_1582305, EPI_ISL_1582306, EPI_ISL_1582307 | | | |
| see above | NORTHWELL HEALTH LABORATORIES | Wadsworth Center, New York State Department of Health | Kirsten St. George, Daryl M. Lamson, Alexis Russell, Matthew Shudt, Melissa A Leisner, Jonathan Plitnick, Catharine Prussing, Navjot Singh, John Kelly, Erasmus Schneider, Erica Lasek-Nesselquist |
| EPI_ISL_1582308, EPI_ISL_1582309, EPI_ISL_1582310, EPI_ISL_1582311, EPI_ISL_1582312, EPI_ISL_1582313, EPI_ISL_1582314, EPI_ISL_1582315, EPI_ISL_1582316 | WESTCHESTER MEDICAL CENTER | Wadsworth Center, New York State Department of Health | Kirsten St. George, Daryl M. Lamson, Alexis Russell, Matthew Shudt, Melissa A Leisner, Jonathan Plitnick, Catharine Prussing, Navjot Singh, John Kelly, Erasmus Schneider, Erica Lasek-Nesselquist |
| EPI_ISL_1582317 | NORTHWELL HEALTH LABORATORIES | Wadsworth Center, New York State Department of Health | Kirsten St. George, Daryl M. Lamson, Alexis Russell, Matthew Shudt, Melissa A Leisner, Jonathan Plitnick, Catharine Prussing, Navjot Singh, John Kelly, Erasmus Schneider, Erica Lasek-Nesselquist |
| EPI_ISL_1582318 | SUNY UPSTATE MEDICAL UNIVERSITY | Wadsworth Center, New York State Department of Health | Kirsten St. George, Daryl M. Lamson, Alexis Russell, Matthew Shudt, Melissa A Leisner, Jonathan Plitnick, Catharine Prussing, Navjot Singh, John Kelly, Erasmus Schneider, Erica Lasek-Nesselquist |
| EPI_ISL_1582319, EPI_ISL_1582320, EPI_ISL_1582321 | MONTEFIORE MEDICAL CENTER LABORATORIES | Wadsworth Center, New York State Department of Health | Kirsten St. George, Daryl M. Lamson, Alexis Russell, Matthew Shudt, Melissa A Leisner, Jonathan Plitnick, Catharine Prussing, Navjot Singh, John Kelly, Erasmus Schneider, Erica Lasek-Nesselquist |
| EPI_ISL_1582322, EPI_ISL_1582323, EPI_ISL_1582324, EPI_ISL_1582325, EPI_ISL_1582326, EPI_ISL_1582327, EPI_ISL_1582328, EPI_ISL_1582329, EPI_ISL_1582330, EPI_ISL_1582331, EPI_ISL_1582332, EPI_ISL_1582333, EPI_ISL_1582334, EPI_ISL_1582335, EPI_ISL_1582336 | | | |
| see above | NYC Pandemic Response Lab | Wadsworth Center, New York State Department of Health | Kirsten St. George, Daryl M. Lamson, Alexis Russell, Matthew Shudt, Melissa A Leisner, Jonathan Plitnick, Catharine Prussing, Navjot Singh, John Kelly, Erasmus Schneider, Erica Lasek-Nesselquist |
| EPI_ISL_1582337 | Columbia University Irving Medical Center | Wadsworth Center, New York State Department of Health | Kirsten St. George, Daryl M. Lamson, Alexis Russell, Matthew Shudt, Melissa A Leisner, Jonathan Plitnick, Catharine Prussing, Navjot Singh, John Kelly, Erasmus Schneider, Erica Lasek-Nesselquist |
| EPI_ISL_1582338 | WESTCHESTER MEDICAL CENTER | Wadsworth Center, New York State Department of Health | Kirsten St. George, Daryl M. Lamson, Alexis Russell, Matthew Shudt, Melissa A Leisner, Jonathan Plitnick, Catharine Prussing, Navjot Singh, John Kelly, Erasmus Schneider, Erica Lasek-Nesselquist |
| EPI_ISL_1582339, EPI_ISL_1582340, EPI_ISL_1582341, EPI_ISL_1582342, EPI_ISL_1582343, EPI_ISL_1582344, EPI_ISL_1582345, EPI_ISL_1582346, EPI_ISL_1582347, EPI_ISL_1582348, EPI_ISL_1582349, EPI_ISL_1582350, EPI_ISL_1582351, EPI_ISL_1582352, EPI_ISL_1582353 | | | |
| see above | URMC LABS | Wadsworth Center, New York State Department of Health | Kirsten St. George, Daryl M. Lamson, Alexis Russell, Matthew Shudt, Melissa A Leisner, Jonathan Plitnick, Catharine Prussing, Navjot Singh, John Kelly, Erasmus Schneider, Erica Lasek-Nesselquist |

| | | | |
|---|---|---|---|
| EPI_ISL_1582354, EPI_ISL_1582355, EPI_ISL_1582356 | NORTHWELL HEALTH LABORATORIES | Wadsworth Center, New York State Department of Health | Kirsten St. George, Daryl M. Lamson, Alexis Russell, Matthew Shudt, Melissa A Leisner, Jonathan Plitnick, Catharine Prussing, Navjot Singh, John Kelly, Erasmus Schneider, Erica Lasek-Nesselquist |
| EPI_ISL_1582357, EPI_ISL_1582358, EPI_ISL_1582359 | TEMPUS LABS INC | Wadsworth Center, New York State Department of Health | Kirsten St. George, Daryl M. Lamson, Alexis Russell, Matthew Shudt, Melissa A Leisner, Jonathan Plitnick, Catharine Prussing, Navjot Singh, John Kelly, Erasmus Schneider, Erica Lasek-Nesselquist |
| EPI_ISL_1582360, EPI_ISL_1582361, EPI_ISL_1582362, EPI_ISL_1582363, EPI_ISL_1582364, EPI_ISL_1582365, EPI_ISL_1582366, EPI_ISL_1582367 | SUNY UPSTATE MEDICAL UNIVERSITY | Wadsworth Center, New York State Department of Health | Kirsten St. George, Daryl M. Lamson, Alexis Russell, Matthew Shudt, Melissa A Leisner, Jonathan Plitnick, Catharine Prussing, Navjot Singh, John Kelly, Erasmus Schneider, Erica Lasek-Nesselquist |
| EPI_ISL_1582368 | Columbia University Irving Medical Center | Wadsworth Center, New York State Department of Health | Kirsten St. George, Daryl M. Lamson, Alexis Russell, Matthew Shudt, Melissa A Leisner, Jonathan Plitnick, Catharine Prussing, Navjot Singh, John Kelly, Erasmus Schneider, Erica Lasek-Nesselquist |
| EPI_ISL_1582369, EPI_ISL_1582370, EPI_ISL_1582371, EPI_ISL_1582372, EPI_ISL_1582373, EPI_ISL_1582374, EPI_ISL_1582375, EPI_ISL_1582376, EPI_ISL_1582377, EPI_ISL_1582378, EPI_ISL_1582379, EPI_ISL_1582380, EPI_ISL_1582381, EPI_ISL_1582382, EPI_ISL_1582383, EPI_ISL_1582384, EPI_ISL_1582385, EPI_ISL_1582386, EPI_ISL_1582387, EPI_ISL_1582388, EPI_ISL_1582389 | | | |
| see above | NORTHWELL HEALTH LABORATORIES | Wadsworth Center, New York State Department of Health | Kirsten St. George, Daryl M. Lamson, Alexis Russell, Matthew Shudt, Melissa A Leisner, Jonathan Plitnick, Catharine Prussing, Navjot Singh, John Kelly, Erasmus Schneider, Erica Lasek-Nesselquist |
| EPI_ISL_1582398 | URMC LABS | Wadsworth Center, New York State Department of Health | Kirsten St. George, Daryl M. Lamson, Alexis Russell, Matthew Shudt, Melissa A Leisner, Jonathan Plitnick, Catharine Prussing, Navjot Singh, John Kelly, Erasmus Schneider, Erica Lasek-Nesselquist |
| EPI_ISL_1582399 | SUNY UPSTATE MEDICAL UNIVERSITY | Wadsworth Center, New York State Department of Health | Kirsten St. George, Daryl M. Lamson, Alexis Russell, Matthew Shudt, Melissa A Leisner, Jonathan Plitnick, Catharine Prussing, Navjot Singh, John Kelly, Erasmus Schneider, Erica Lasek-Nesselquist |
| EPI_ISL_1582400, EPI_ISL_1582401, EPI_ISL_1582402 | NYC Pandemic Response Lab | Wadsworth Center, New York State Department of Health | Kirsten St. George, Daryl M. Lamson, Alexis Russell, Matthew Shudt, Melissa A Leisner, Jonathan Plitnick, Catharine Prussing, Navjot Singh, John Kelly, Erasmus Schneider, Erica Lasek-Nesselquist |
| EPI_ISL_1582403 | TEMPUS LABS INC | Wadsworth Center, New York State Department of Health | Kirsten St. George, Daryl M. Lamson, Alexis Russell, Matthew Shudt, Melissa A Leisner, Jonathan Plitnick, Catharine Prussing, Navjot Singh, John Kelly, Erasmus Schneider, Erica Lasek-Nesselquist |
| EPI_ISL_1582404 | URMC LABS | Wadsworth Center, New York State Department of Health | Kirsten St. George, Daryl M. Lamson, Alexis Russell, Matthew Shudt, Melissa A Leisner, Jonathan Plitnick, Catharine Prussing, Navjot Singh, John Kelly, Erasmus Schneider, Erica Lasek-Nesselquist |
| EPI_ISL_1582405 | NORTHWELL HEALTH LABORATORIES | Wadsworth Center, New York State Department of Health | Kirsten St. George, Daryl M. Lamson, Alexis Russell, Matthew Shudt, Melissa A Leisner, Jonathan Plitnick, Catharine Prussing, Navjot Singh, John Kelly, Erasmus Schneider, Erica Lasek-Nesselquist |
| EPI_ISL_1582406 | GLENS FALLS HOSPITAL LABORATORY | Wadsworth Center, New York State Department of Health | Kirsten St. George, Daryl M. Lamson, Alexis Russell, Matthew Shudt, Melissa A Leisner, Jonathan Plitnick, Catharine Prussing, Navjot Singh, John Kelly, Erasmus Schneider, Erica Lasek-Nesselquist |
| EPI_ISL_1582407 | ALBANY MEDICAL CENTER | Wadsworth Center, New York State Department of Health | Kirsten St. George, Daryl M. Lamson, Alexis Russell, Matthew Shudt, Melissa A Leisner, Jonathan Plitnick, Catharine Prussing, Navjot Singh, John Kelly, Erasmus Schneider, Erica Lasek-Nesselquist |
