## Supplemental acknowledgement table #4 for "Genomic Surveillance of SARS-CoV-2 in Erie County, New York"

We gratefully acknowledge the following Authors from the Originating laboratories responsible for obtaining the specimens, as well as the Submitting laboratories where the genome data were generated and shared via GISAID, on which this research is based.

All Submitters of data may be contacted directly via www.gisaid.org

Authors are sorted alphabetically.

| Accession ID | Originating Laboratory | Submitting Laboratory | Authors |
|---|---|---|---|
| EPI_ISL_1587343, EPI_ISL_1587390 | Yale Clinical Virology Lab | Grubaugh Lab - Yale School of Public Health | Joseph Fauver, Mallery Breban, Isabel Ott, Tara Alpert, Mary Petrone, Anderson Brito, Chantal Vogels, Annie Watkins, Chaney Kalinich, Jessica Rothman, Marie L. Landry, Nathan Grubaugh |
| EPI_ISL_1587589, EPI_ISL_1587712, EPI_ISL_1587757, EPI_ISL_1587786, EPI_ISL_1587941, EPI_ISL_1587942, EPI_ISL_1587944, EPI_ISL_1587951, EPI_ISL_1587952, EPI_ISL_1587954, EPI_ISL_1587968, EPI_ISL_1587975, EPI_ISL_1588003, EPI_ISL_1588112, EPI_ISL_1588190, EPI_ISL_1588288, EPI_ISL_1588322, EPI_ISL_1588324, EPI_ISL_1588405, EPI_ISL_1588406, EPI_ISL_1588412, EPI_ISL_1588415, EPI_ISL_1588418, EPI_ISL_1588419, EPI_ISL_1588427, EPI_ISL_1588433 | | | |
| see above | Infinity Biologix | Centers for Disease Control and Prevention Division of Viral Diseases, Pathogen Discovery | Dakota Howard, Dhwani Batra, Peter W. Cook, Kara Moser, Adrian Paskey, Jason Caravas, Benjamin Rambo-Martin, Shatavia Morrison, Christopher Gulvick, Scott Sammons, Yvette Unoarumhi, Darlene Wagner, Matthew Schmerer, Christian Bixby, Yihe Wang, Jonathan Schultz, Chirayu Goswami, Russ Hager, Robin Grimwood, Clinton R. Paden, Duncan MacCannell |
| EPI_ISL_1592417, EPI_ISL_1592426 | Helix/Illumina | Centers for Disease Control and Prevention Division of Viral Diseases, Pathogen Discovery | Dakota Howard, Dhwani Batra, Peter W. Cook, Kara Moser, Adrian Paskey, Jason Caravas, Benjamin Rambo-Martin, Shatavia Morrison, Christopher Gulvick, Scott Sammons, Yvette Unoarumhi, Darlene Wagner, Matthew Schmerer, Eileen de Feo, Jan Antico, Christine Tran, Matthew Tolentino, Shannon Wickline, Kim Gietzen, Brad Sickler, Jingtao Liu, Eric Allen, Phil Febbo, Nicole L. Washington, Simon White, Geraint Levan, Kelly Schiabor Barrett, Elizabeth Cirulli, Alexandre Bolze, Ary Ascencio, Charlotte Rivera-Garcia, Ryan Cho, Jason Nguyen, Sherry Wang, Jimmy Ramirez, Tyler Cassens, Efren Sandoval, Magnus Isaksson, William Lee, David Becker, Marc Laurent, James Lu, Clinton R. Paden, Duncan MacCannell |
| EPI_ISL_1593720 | DOHMH Central Harlem | New York City Public Health Laboratory | Jade Wang, et al. |
| EPI_ISL_1593721 | DOHMH Corona | New York City Public Health Laboratory | Jade Wang, et al. |
| EPI_ISL_1593722 | DOHMH Central Harlem | New York City Public Health Laboratory | Jade Wang, et al. |
| EPI_ISL_1593723 | Bellevue Hospital Center | New York City Public Health Laboratory | Jade Wang, et al. |
| EPI_ISL_1593724 | Brookdale University Hospital Medical Center | New York City Public Health Laboratory | Jade Wang, et al. |
| EPI_ISL_1593729 | DOHMH Crown Heights | New York City Public Health Laboratory | Jade Wang, et al. |
| EPI_ISL_1593730 | DOHMH Jamaica | New York City Public Health Laboratory | Jade Wang, et al. |
| EPI_ISL_1593731 | DOHMH PHL | New York City Public Health Laboratory | Jade Wang, et al. |
| EPI_ISL_1593732 | DOHMH Jamaica | New York City Public Health Laboratory | Jade Wang, et al. |
| EPI_ISL_1593733 | DOHMH Chelsea | New York City Public Health Laboratory | Jade Wang, et al. |
| EPI_ISL_1593734 | DOHMH PHL | New York City Public Health Laboratory | Jade Wang, et al. |
| EPI_ISL_1593735 | DOHMH Jamaica | New York City Public Health Laboratory | Jade Wang, et al. |
| EPI_ISL_1593736 | DOHMH Riverside | New York City Public Health Laboratory | Jade Wang, et al. |
| EPI_ISL_1593737 | DOHMH Corona | New York City Public Health Laboratory | Jade Wang, et al. |
| EPI_ISL_1593738, EPI_ISL_1593739 | DOHMH Jamaica | New York City Public Health Laboratory | Jade Wang, et al. |
| EPI_ISL_1593740 | DOHMH Fort Greene | New York City Public Health Laboratory | Jade Wang, et al. |
| EPI_ISL_1593741 | DOHMH Crown Heights | New York City Public Health Laboratory | Jade Wang, et al. |
| EPI_ISL_1593742, EPI_ISL_1593743, EPI_ISL_1593744, EPI_ISL_1593745 | DOHMH Central Harlem | New York City Public Health Laboratory | Jade Wang, et al. |
| EPI_ISL_1593746 | DOHMH Corona | New York City Public Health Laboratory | Jade Wang, et al. |
| EPI_ISL_1593747, EPI_ISL_1593748 | DOHMH Jamaica | New York City Public Health Laboratory | Jade Wang, et al. |
| EPI_ISL_1593749 | DOHMH Crown Heights | New York City Public Health Laboratory | Jade Wang, et al. |
| EPI_ISL_1593750 | DOHMH Fort Greene | New York City Public Health Laboratory | Jade Wang, et al. |
| EPI_ISL_1593751, EPI_ISL_1593752 | DOHMH Central Harlem | New York City Public Health Laboratory | Jade Wang, et al. |
| EPI_ISL_1593753 | DOHMH Morrisania | New York City Public Health Laboratory | Jade Wang, et al. |
| EPI_ISL_1593754, EPI_ISL_1593755 | DOHMH Chelsea | New York City Public Health Laboratory | Jade Wang, et al. |
| EPI_ISL_1593756, EPI_ISL_1593757 | OCME Office Of Chief Medical Examiner | New York City Public Health Laboratory | Jade Wang, et al. |
| EPI_ISL_1593758 | DOHMH Fort Greene | New York City Public Health Laboratory | Jade Wang, et al. |
| EPI_ISL_1593759, EPI_ISL_1593760, EPI_ISL_1593761 | DOHMH Corona | New York City Public Health Laboratory | Jade Wang, et al. |
| EPI_ISL_1593762 | DOHMH Jamaica | New York City Public Health Laboratory | Jade Wang, et al. |
| EPI_ISL_1593763, EPI_ISL_1593764, EPI_ISL_1593765, EPI_ISL_1593766, EPI_ISL_1593767 | DOHMH Central Harlem | New York City Public Health Laboratory | Jade Wang, et al. |
| EPI_ISL_1593768 | DOHMH Corona | New York City Public Health Laboratory | Jade Wang, et al. |
| EPI_ISL_1593769 | DOHMH Crown Heights | New York City Public Health Laboratory | Jade Wang, et al. |
| EPI_ISL_1593770 | DOHMH Fort Greene | New York City Public Health Laboratory | Jade Wang, et al. |
| EPI_ISL_1593771 | Department of Homeless Services | New York City Public Health Laboratory | Jade Wang, et al. |
| EPI_ISL_1593772 | OCME Office Of Chief Medical Examiner | New York City Public Health Laboratory | Jade Wang, et al. |
| EPI_ISL_1593864, EPI_ISL_1593865 | WESTCHESTER MEDICAL CENTER | Wadsworth Center, New York State Department of Health | Kirsten St. George, Daryl M. Lamson, Alexis Russell, Matthew Shudt, Melissa A Leisner, Jonathan Plitnick, Catharine Prussing, Navjot Singh, John Kelly, Erasmus Schneider, Erica Lasek-Nesselquist |
| EPI_ISL_1593866, EPI_ISL_1593867 | URMC LABS | Wadsworth Center, New York State Department of Health | Kirsten St. George, Daryl M. Lamson, Alexis Russell, Matthew Shudt, Melissa A Leisner, Jonathan Plitnick, Catharine Prussing, Navjot Singh, John Kelly, Erasmus Schneider, Erica Lasek-Nesselquist |

| | | | |
|---|---|---|---|
| EPI_ISL_1593868, EPI_ISL_1593869 | MONTEFIORE MEDICAL CENTER LABORATORIES | Wadsworth Center, New York State Department of Health | Kirsten St. George, Daryl M. Lamson, Alexis Russell, Matthew Shudt, Melissa A Leisner, Jonathan Plitnick, Catharine Prussing, Navjot Singh, John Kelly, Erasmus Schneider, Erica Lasek-Nesselquist |
| EPI_ISL_1593870, EPI_ISL_1593871 | NYC Pandemic Response Lab | Wadsworth Center, New York State Department of Health | Kirsten St. George, Daryl M. Lamson, Alexis Russell, Matthew Shudt, Melissa A Leisner, Jonathan Plitnick, Catharine Prussing, Navjot Singh, John Kelly, Erasmus Schneider, Erica Lasek-Nesselquist |
| EPI_ISL_1593872 | WESTCHESTER MEDICAL CENTER | Wadsworth Center, New York State Department of Health | Kirsten St. George, Daryl M. Lamson, Alexis Russell, Matthew Shudt, Melissa A Leisner, Jonathan Plitnick, Catharine Prussing, Navjot Singh, John Kelly, Erasmus Schneider, Erica Lasek-Nesselquist |
| EPI_ISL_1593873, EPI_ISL_1593874, EPI_ISL_1593875 | TEMPUS LABS INC | Wadsworth Center, New York State Department of Health | Kirsten St. George, Daryl M. Lamson, Alexis Russell, Matthew Shudt, Melissa A Leisner, Jonathan Plitnick, Catharine Prussing, Navjot Singh, John Kelly, Erasmus Schneider, Erica Lasek-Nesselquist |
| EPI_ISL_1593876, EPI_ISL_1593877 | NORTHWELL HEALTH LABORATORIES | Wadsworth Center, New York State Department of Health | Kirsten St. George, Daryl M. Lamson, Alexis Russell, Matthew Shudt, Melissa A Leisner, Jonathan Plitnick, Catharine Prussing, Navjot Singh, John Kelly, Erasmus Schneider, Erica Lasek-Nesselquist |
| EPI_ISL_1593878 | NYC Pandemic Response Lab | Wadsworth Center, New York State Department of Health | Kirsten St. George, Daryl M. Lamson, Alexis Russell, Matthew Shudt, Melissa A Leisner, Jonathan Plitnick, Catharine Prussing, Navjot Singh, John Kelly, Erasmus Schneider, Erica Lasek-Nesselquist |
| EPI_ISL_1593881 | MONTEFIORE MEDICAL CENTER LABORATORIES | Wadsworth Center, New York State Department of Health | Kirsten St. George, Daryl M. Lamson, Alexis Russell, Matthew Shudt, Melissa A Leisner, Jonathan Plitnick, Catharine Prussing, Navjot Singh, John Kelly, Erasmus Schneider, Erica Lasek-Nesselquist |
| EPI_ISL_1593882 | URMC LABS | Wadsworth Center, New York State Department of Health | Kirsten St. George, Daryl M. Lamson, Alexis Russell, Matthew Shudt, Melissa A Leisner, Jonathan Plitnick, Catharine Prussing, Navjot Singh, John Kelly, Erasmus Schneider, Erica Lasek-Nesselquist |
| EPI_ISL_1593883 | NYC Pandemic Response Lab | Wadsworth Center, New York State Department of Health | Kirsten St. George, Daryl M. Lamson, Alexis Russell, Matthew Shudt, Melissa A Leisner, Jonathan Plitnick, Catharine Prussing, Navjot Singh, John Kelly, Erasmus Schneider, Erica Lasek-Nesselquist |
| EPI_ISL_1593884, EPI_ISL_1593885 | TEMPUS LABS INC | Wadsworth Center, New York State Department of Health | Kirsten St. George, Daryl M. Lamson, Alexis Russell, Matthew Shudt, Melissa A Leisner, Jonathan Plitnick, Catharine Prussing, Navjot Singh, John Kelly, Erasmus Schneider, Erica Lasek-Nesselquist |
| EPI_ISL_1593886 | WHITE PLAINS HOSPITAL CENTER LABORATORY | Wadsworth Center, New York State Department of Health | Kirsten St. George, Daryl M. Lamson, Alexis Russell, Matthew Shudt, Melissa A Leisner, Jonathan Plitnick, Catharine Prussing, Navjot Singh, John Kelly, Erasmus Schneider, Erica Lasek-Nesselquist |
| EPI_ISL_1593887 | NORTHWELL HEALTH LABORATORIES | Wadsworth Center, New York State Department of Health | Kirsten St. George, Daryl M. Lamson, Alexis Russell, Matthew Shudt, Melissa A Leisner, Jonathan Plitnick, Catharine Prussing, Navjot Singh, John Kelly, Erasmus Schneider, Erica Lasek-Nesselquist |
| EPI_ISL_1597696, EPI_ISL_1597697, EPI_ISL_1597698, EPI_ISL_1597699, EPI_ISL_1597700, EPI_ISL_1597701, EPI_ISL_1597702, EPI_ISL_1597703, EPI_ISL_1597704, EPI_ISL_1597705, EPI_ISL_1597706, EPI_ISL_1597707, EPI_ISL_1597708, EPI_ISL_1597709, EPI_ISL_1597710, EPI_ISL_1597711, EPI_ISL_1597712, EPI_ISL_1597713, EPI_ISL_1597714, EPI_ISL_1597715, EPI_ISL_1597716, EPI_ISL_1597717, EPI_ISL_1597725, EPI_ISL_1597726, EPI_ISL_1597727, EPI_ISL_1597728, EPI_ISL_1597729, EPI_ISL_1597730, EPI_ISL_1597731, EPI_ISL_1597732, EPI_ISL_1597733, EPI_ISL_1597745, EPI_ISL_1597746, EPI_ISL_1597747, EPI_ISL_1597748, EPI_ISL_1597749, EPI_ISL_1597750, EPI_ISL_1597751, EPI_ISL_1597752, EPI_ISL_1597753, EPI_ISL_1597754, EPI_ISL_1597755, EPI_ISL_1597756, EPI_ISL_1597757, EPI_ISL_1597758, EPI_ISL_1597759, EPI_ISL_1597760, EPI_ISL_1597761, EPI_ISL_1597762, EPI_ISL_1597763, EPI_ISL_1597764, EPI_ISL_1597765, EPI_ISL_1597766, EPI_ISL_1597767, EPI_ISL_1597768, EPI_ISL_1597769, EPI_ISL_1597770, EPI_ISL_1597771, EPI_ISL_1597772, EPI_ISL_1597773, EPI_ISL_1597774, EPI_ISL_1597775, EPI_ISL_1597776, EPI_ISL_1597777, EPI_ISL_1597778, EPI_ISL_1597779, EPI_ISL_1597780, EPI_ISL_1597781 | | | |
| see above | Montefiore Medical Center | Abbott | Yitz Goldstein, Amy Fox, Ana Olivo, Ana Vallari, Barbara Harris, Mary Rodgers, Todd Meyer, Gavin Cloherty |
| EPI_ISL_1605699 | TEMPUS LABS INC | Wadsworth Center, New York State Department of Health | Kirsten St. George, Daryl M. Lamson, Alexis Russell, Matthew Shudt, Melissa A Leisner, Jonathan Plitnick, Catharine Prussing, Navjot Singh, John Kelly, Erasmus Schneider, Erica Lasek-Nesselquist |
| EPI_ISL_1605704 | NYC Pandemic Response Lab | Wadsworth Center, New York State Department of Health | Kirsten St. George, Daryl M. Lamson, Alexis Russell, Matthew Shudt, Melissa A Leisner, Jonathan Plitnick, Catharine Prussing, Navjot Singh, John Kelly, Erasmus Schneider, Erica Lasek-Nesselquist |
| EPI_ISL_1609076, EPI_ISL_1609079, EPI_ISL_1609080, EPI_ISL_1609139, EPI_ISL_1609147, EPI_ISL_1609151, EPI_ISL_1609174, EPI_ISL_1609178, EPI_ISL_1609179, EPI_ISL_1609180, EPI_ISL_1609216, EPI_ISL_1609238, EPI_ISL_1609239, EPI_ISL_1609240, EPI_ISL_1609241, EPI_ISL_1609249, EPI_ISL_1609250, EPI_ISL_1609251, EPI_ISL_1609314, EPI_ISL_1609315, EPI_ISL_1609316, EPI_ISL_1609319, EPI_ISL_1609328, EPI_ISL_1609332, EPI_ISL_1609333, EPI_ISL_1609345, EPI_ISL_1609346, EPI_ISL_1609348, EPI_ISL_1609351, EPI_ISL_1609352, EPI_ISL_1609353, EPI_ISL_1609354, EPI_ISL_1609355, EPI_ISL_1609356, EPI_ISL_1609357, EPI_ISL_1609368, EPI_ISL_1609369, EPI_ISL_1609406, EPI_ISL_1609430, EPI_ISL_1609432, EPI_ISL_1609433, EPI_ISL_1609434, EPI_ISL_1609437, EPI_ISL_1609438, EPI_ISL_1609439, EPI_ISL_1609444, EPI_ISL_1609446, EPI_ISL_1609454, EPI_ISL_1609457, EPI_ISL_1609464, EPI_ISL_1609465, EPI_ISL_1609471, EPI_ISL_1609472, EPI_ISL_1609475, EPI_ISL_1609476, EPI_ISL_1609477, EPI_ISL_1609495, EPI_ISL_1609499, EPI_ISL_1609504, EPI_ISL_1609513, EPI_ISL_1609527, EPI_ISL_1609535, EPI_ISL_1609537, EPI_ISL_1609538, EPI_ISL_1609539, EPI_ISL_1609554, EPI_ISL_1609556, EPI_ISL_1609557, EPI_ISL_1609558, EPI_ISL_1609559, EPI_ISL_1609583, EPI_ISL_1609584, EPI_ISL_1609585, EPI_ISL_1609586, EPI_ISL_1609604, EPI_ISL_1609608, EPI_ISL_1609609, EPI_ISL_1609610, EPI_ISL_1609611, EPI_ISL_1609612, EPI_ISL_1609613, EPI_ISL_1609614, EPI_ISL_1609619, EPI_ISL_1609620, EPI_ISL_1609623, EPI_ISL_1609624, EPI_ISL_1609633, EPI_ISL_1609635, EPI_ISL_1609636, EPI_ISL_1609640, EPI_ISL_1609641, EPI_ISL_1609666, EPI_ISL_1609667, EPI_ISL_1609668, EPI_ISL_1609669, EPI_ISL_1609670, EPI_ISL_1609683, EPI_ISL_1609687, EPI_ISL_1609689, EPI_ISL_1609690, EPI_ISL_1609698, EPI_ISL_1609704, EPI_ISL_1609706, EPI_ISL_1609714, EPI_ISL_1609716, EPI_ISL_1609719, EPI_ISL_1609720, EPI_ISL_1609731, EPI_ISL_1609733, EPI_ISL_1609741, EPI_ISL_1609751, EPI_ISL_1609754, EPI_ISL_1609757, EPI_ISL_1609770, EPI_ISL_1609778, EPI_ISL_1609779, EPI_ISL_1609785, EPI_ISL_1609786, EPI_ISL_1609789, EPI_ISL_1609792, EPI_ISL_1609799, EPI_ISL_1609800, EPI_ISL_1609801, EPI_ISL_1609802, EPI_ISL_1609803, EPI_ISL_1609804, EPI_ISL_1609805, EPI_ISL_1609806, EPI_ISL_1609807, EPI_ISL_1609808, EPI_ISL_1609809, EPI_ISL_1609810, EPI_ISL_1609811, EPI_ISL_1609812, EPI_ISL_1609813, EPI_ISL_1609814, EPI_ISL_1609815, EPI_ISL_1609874, EPI_ISL_1609880, EPI_ISL_1609883, EPI_ISL_1609887, EPI_ISL_1609889, EPI_ISL_1609890, EPI_ISL_1609891, EPI_ISL_1609893, EPI_ISL_1609894, EPI_ISL_1609900, EPI_ISL_1609904, EPI_ISL_1609909, EPI_ISL_1609910, EPI_ISL_1609912, EPI_ISL_1609917, EPI_ISL_1609921, EPI_ISL_1609922, EPI_ISL_1609923, EPI_ISL_1609924, EPI_ISL_1609926, EPI_ISL_1609927, EPI_ISL_1609938, EPI_ISL_1609946, EPI_ISL_1609949, EPI_ISL_1609952, EPI_ISL_1609954, EPI_ISL_1609955, EPI_ISL_1609957, EPI_ISL_1609964, EPI_ISL_1609967, EPI_ISL_1610041, EPI_ISL_1610047, EPI_ISL_1610236, EPI_ISL_1610237, EPI_ISL_1610238, EPI_ISL_1610239, EPI_ISL_1610240, EPI_ISL_1610241, EPI_ISL_1610242, EPI_ISL_1610243, EPI_ISL_1610245, EPI_ISL_1610246, EPI_ISL_1610247, EPI_ISL_1610248, EPI_ISL_1610249, EPI_ISL_1610293, EPI_ISL_1610294, EPI_ISL_1610295, EPI_ISL_1610296, EPI_ISL_1610297, EPI_ISL_1610298, EPI_ISL_1610299, EPI_ISL_1610300, EPI_ISL_1610302, EPI_ISL_1610303, EPI_ISL_1610321, EPI_ISL_1610322, EPI_ISL_1610323, EPI_ISL_1610401, EPI_ISL_1610404, EPI_ISL_1610406, EPI_ISL_1610408, EPI_ISL_1610410, EPI_ISL_1610411, EPI_ISL_1610432, EPI_ISL_1610441, EPI_ISL_1610467, EPI_ISL_1610494, EPI_ISL_1610568, EPI_ISL_1610569, EPI_ISL_1610571, EPI_ISL_1610573, EPI_ISL_1610595, EPI_ISL_1610597, EPI_ISL_1610599, EPI_ISL_1610601, EPI_ISL_1610602, EPI_ISL_1610604, EPI_ISL_1610623, EPI_ISL_1610625, EPI_ISL_1610900, EPI_ISL_1610901, EPI_ISL_1610902, EPI_ISL_1610903, EPI_ISL_1610904, EPI_ISL_1610905, EPI_ISL_1610906, EPI_ISL_1610907, EPI_ISL_1610908, EPI_ISL_1610920, EPI_ISL_1610949, EPI_ISL_1610951, EPI_ISL_1610953, EPI_ISL_1611111, EPI_ISL_1611173, EPI_ISL_1611206, EPI_ISL_1611213, EPI_ISL_1611219, EPI_ISL_1611220, EPI_ISL_1611221, EPI_ISL_1611242, EPI_ISL_1611243, EPI_ISL_1611244, EPI_ISL_1611268, EPI_ISL_1611282, EPI_ISL_1611297, EPI_ISL_1611300, EPI_ISL_1611308, EPI_ISL_1611311, EPI_ISL_1611314, EPI_ISL_1611315, EPI_ISL_1611316, EPI_ISL_1611363, EPI_ISL_1611364, EPI_ISL_1611366, EPI_ISL_1611368, EPI_ISL_1611371, EPI_ISL_1611374, EPI_ISL_1611375, EPI_ISL_1611376, EPI_ISL_1611385, EPI_ISL_1611398, EPI_ISL_1611399, EPI_ISL_1611417, EPI_ISL_1611420, EPI_ISL_1611433, EPI_ISL_1611436, EPI_ISL_1611464, EPI_ISL_1611472, EPI_ISL_1611474, EPI_ISL_1611475, EPI_ISL_1611476, EPI_ISL_1611477, EPI_ISL_1611479, EPI_ISL_1611480, EPI_ISL_1611482, EPI_ISL_1611526, EPI_ISL_1611527, EPI_ISL_1611528, EPI_ISL_1611534, EPI_ISL_1611535, EPI_ISL_1611542, EPI_ISL_1611543, EPI_ISL_1611544, EPI_ISL_1611547, EPI_ISL_1611549, EPI_ISL_1611564, EPI_ISL_1611565, EPI_ISL_1611566, EPI_ISL_1611567, EPI_ISL_1611568, EPI_ISL_1611570, EPI_ISL_1611572, EPI_ISL_1611624, EPI_ISL_1611625, EPI_ISL_1611628, EPI_ISL_1611632, EPI_ISL_1611640, EPI_ISL_1611644, EPI_ISL_1611647, EPI_ISL_1611650, EPI_ISL_1611651, EPI_ISL_1611653, EPI_ISL_1611654, EPI_ISL_1611659, EPI_ISL_1611660, EPI_ISL_1611667, EPI_ISL_1611668, EPI_ISL_1611690, EPI_ISL_1611693, EPI_ISL_1611695, EPI_ISL_1611719, EPI_ISL_1611721, EPI_ISL_1611722, EPI_ISL_1611738, EPI_ISL_1611739, EPI_ISL_1611741, EPI_ISL_1611743, EPI_ISL_1611744, EPI_ISL_1611768, EPI_ISL_1611769, EPI_ISL_1611776, EPI_ISL_1611784, EPI_ISL_1611785, EPI_ISL_1611792, EPI_ISL_1611795, EPI_ISL_1611796, EPI_ISL_1611813, EPI_ISL_1611814, EPI_ISL_1611825, EPI_ISL_1611839, EPI_ISL_1611911, EPI_ISL_1611913, EPI_ISL_1611917, EPI_ISL_1611919, EPI_ISL_1611948, EPI_ISL_1611952, EPI_ISL_1611954, EPI_ISL_1611955, EPI_ISL_1611957, EPI_ISL_1611959 | | | |
| see above | Laboratory Corporation of America | Centers for Disease Control and Prevention Division of Viral Diseases, Pathogen Discovery | Dakota Howard, Dhwani Batra, Peter W. Cook, Kara Moser, Adrian Paskey, Jason Caravas, Benjamin Rambo-Martin, Shatavia Morrison, Christopher Gulvick, Scott Sammons, Yvette Unoarumhi, Darlene Wagner, Matthew Schmerer, Minoo Agarwal, Eyad Almasri, Debbie Boles, Ayla Burns, Nuthawin Charoensri, Oren Cohen, Susan Countryman, Mary Ann Cristobal, Bobbi Croy, Suzanne Dale, Hrushikesh Deshmukh, Amanda Douglas, Vincent Drouillon, Marcia Eisenberg, Howard Engler, Rama Ghatti, Prashant Gupta, Susan Hicks, Jake Humphrey, Lax Iyer, Manoj Jain, Mohan Kolli, Brian Krueger, Tim Kuphal, Stanley Letovsky, Michael Levandoski, Craig Lukasik, Jonathan Meltzer, Brian Norvell, Mindy Nye, Scott Parker, Christos Petropoulos, John Pruitt, Steven Ragan, Scott Ryan, Mike Sapeta, Jana Schroth, Suresh Babu Selvaraju, Goran Stevovic, Amanda Suchanek, Andrea Throop, Lyndon Tilson, Thomas Urban, Joe Voshell, Kimberly Wagner, Jonathan Williams, Mary Williamson, Qian Zeng, Tricia Zwiefelhofer, Clinton R. Paden, Duncan MacCannell |
| EPI_ISL_1614913, EPI_ISL_1614918, EPI_ISL_1614935, EPI_ISL_1614940, EPI_ISL_1614948, EPI_ISL_1614957, EPI_ISL_1614967, EPI_ISL_1614977, EPI_ISL_1614981, EPI_ISL_1614988, EPI_ISL_1615021, EPI_ISL_1615032, EPI_ISL_1615037, EPI_ISL_1615039, EPI_ISL_1615047, EPI_ISL_1615061 | | | |
| see above | Quest Diagnostics Incorporated | Centers for Disease Control and Prevention Division of Viral Diseases, Pathogen Discovery | Dakota Howard, Dhwani Batra, Peter W. Cook, Kara Moser, Adrian Paskey, Jason Caravas, Benjamin Rambo-Martin, Shatavia Morrison, Christopher Gulvick, Scott Sammons, Yvette Unoarumhi, Darlene Wagner, Matthew Schmerer, S. H. Rosenthal, A. Gerasimova, R. M. Kagan, B. Anderson, M. Hua, Y. Liu, L.E. Bernstein, K.E. Livingston, A. Perez, I. A. Shlyakhter, R. V. Rolando, R. Owen, P. Tanpaiboon, F. Lacbawan, Clinton R. Paden, Duncan MacCannell |
| EPI_ISL_1615131 | Helix/Illumina | Centers for Disease Control and Prevention Division of Viral Diseases, Pathogen Discovery | Dakota Howard, Dhwani Batra, Peter W. Cook, Kara Moser, Adrian Paskey, Jason Caravas, Benjamin Rambo-Martin, Shatavia Morrison, Christopher Gulvick, Scott Sammons, Yvette Unoarumhi, Darlene Wagner, Matthew Schmerer, Eileen de Feo, Jan Antico, Christine Tran, Matthew Tolentino, Shannon Wickline, Kim Gietzen, Brad Sickler, Jingtao Liu, Eric Allen, Phil Febbo, Nicole L. Washington, Simon White, Geraint Levan, Kelly Schiabor Barrett, Elizabeth Cirulli, Alexandre Bolze, Ary Ascencio, Charlotte Rivera-Garcia, Ryan Cho, Jason Nguyen, Sherry Wang, Jimmy Ramirez, Tyler Cassens, Efren Sandoval, Magnus Isaksson, William Lee, David Becker, Marc Laurent, James Lu, Clinton R. Paden, Duncan MacCannell |
| EPI_ISL_1621729, EPI_ISL_1621744, EPI_ISL_1621747, EPI_ISL_1621749, EPI_ISL_1621786, EPI_ISL_1621789, EPI_ISL_1621807, EPI_ISL_1621812, EPI_ISL_1621815, EPI_ISL_1621818, EPI_ISL_1621850, EPI_ISL_1621854, EPI_ISL_1621855, EPI_ISL_1621856, EPI_ISL_1621857, EPI_ISL_1621859, EPI_ISL_1621860, EPI_ISL_1621865, EPI_ISL_1621866, EPI_ISL_1621871, EPI_ISL_1621872, EPI_ISL_1621873, EPI_ISL_1621875, EPI_ISL_1621876, EPI_ISL_1621879, EPI_ISL_1621880, EPI_ISL_1621881, EPI_ISL_1621884, EPI_ISL_1621890, EPI_ISL_1621896, EPI_ISL_1621903, EPI_ISL_1621906, EPI_ISL_1621909, EPI_ISL_1621913, EPI_ISL_1621915, EPI_ISL_1621921, EPI_ISL_1621924, EPI_ISL_1621925, EPI_ISL_1621928, EPI_ISL_1621941, EPI_ISL_1621942, EPI_ISL_1621945, EPI_ISL_1621946, EPI_ISL_1621947, EPI_ISL_1621956, EPI_ISL_1621958, EPI_ISL_1621962, EPI_ISL_1621988, EPI_ISL_1621991, EPI_ISL_1621992, EPI_ISL_1622001, EPI_ISL_1622003, EPI_ISL_1622005, EPI_ISL_1622007, EPI_ISL_1622008, EPI_ISL_1622018, EPI_ISL_1622026, EPI_ISL_1622030, EPI_ISL_1622031, EPI_ISL_1622038, EPI_ISL_1622042, EPI_ISL_1622047, EPI_ISL_1622055, EPI_ISL_1622059, EPI_ISL_1622072, EPI_ISL_1622073, EPI_ISL_1622075, EPI_ISL_1622079, EPI_ISL_1622087, EPI_ISL_1622095, EPI_ISL_1622103, EPI_ISL_1622105, EPI_ISL_1622109 | | | |
| see above | Quest Diagnostics Incorporated | Centers for Disease Control and Prevention Division of Viral | Dakota Howard, Dhwani Batra, Peter W. Cook, Kara Moser, Adrian Paskey, Jason Caravas, Benjamin Rambo-Martin, Shatavia Morrison, Christopher |

| | | | |
|---|---|---|---|
| | | Diseases, Pathogen Discovery | Gulvick, Scott Sammons, Yvette Unoarumhi, Darlene Wagner, Matthew Schmerer, S. H. Rosenthal, A. Gerasimova, R. M. Kagan, B. Anderson, M. Hua, Y. Liu, L.E. Bernstein, K.E. Livingston, A. Perez, I. A. Shlyakhter, R. V. Rolando, R. Owen, P. Tanpaiboon, F. Lacbawan, Clinton R. Paden, Duncan MacCannell |
| EPI_ISL_1623918 | NORTHWELL HEALTH LABORATORIES | Wadsworth Center, New York State Department of Health | Kirsten St. George, Daryl M. Lamson, Alexis Russell, Matthew Shudt, Melissa A Leisner, Jonathan Plitnick, Catharine Prussing, Navjot Singh, John Kelly, Erasmus Schneider, Erica Lasek-Nesselquist |
| EPI_ISL_1623919, EPI_ISL_1623920, EPI_ISL_1623921, EPI_ISL_1623922, EPI_ISL_1623923, EPI_ISL_1623924, EPI_ISL_1623925, EPI_ISL_1623926, EPI_ISL_1623927, EPI_ISL_1623928, EPI_ISL_1623929, EPI_ISL_1623930, EPI_ISL_1623931, EPI_ISL_1623932, EPI_ISL_1623933, EPI_ISL_1623934, EPI_ISL_1623935, EPI_ISL_1623936, EPI_ISL_1623937, EPI_ISL_1623938, EPI_ISL_1623939, EPI_ISL_1623940, EPI_ISL_1623941, EPI_ISL_1623942, EPI_ISL_1623943, EPI_ISL_1623944, EPI_ISL_1623945, EPI_ISL_1623946, EPI_ISL_1623947, EPI_ISL_1623948, EPI_ISL_1623949, EPI_ISL_1623950, EPI_ISL_1623951 | | | |
| see above | MEMORIAL SLOAN KETTERING CANCER CENTER | Wadsworth Center, New York State Department of Health | Kirsten St. George, Daryl M. Lamson, Alexis Russell, Matthew Shudt, Melissa A Leisner, Jonathan Plitnick, Catharine Prussing, Navjot Singh, John Kelly, Erasmus Schneider, Erica Lasek-Nesselquist |
| EPI_ISL_1623952 | Columbia University Irving Medical Center | Wadsworth Center, New York State Department of Health | Kirsten St. George, Daryl M. Lamson, Alexis Russell, Matthew Shudt, Melissa A Leisner, Jonathan Plitnick, Catharine Prussing, Navjot Singh, John Kelly, Erasmus Schneider, Erica Lasek-Nesselquist |
| EPI_ISL_1623953, EPI_ISL_1623954, EPI_ISL_1623955, EPI_ISL_1623956, EPI_ISL_1623957, EPI_ISL_1623958, EPI_ISL_1623959, EPI_ISL_1623960, EPI_ISL_1623961, EPI_ISL_1623962, EPI_ISL_1623963, EPI_ISL_1623964, EPI_ISL_1623965, EPI_ISL_1623966, EPI_ISL_1623967, EPI_ISL_1623968, EPI_ISL_1623969, EPI_ISL_1623970, EPI_ISL_1623971, EPI_ISL_1623972 | | | |
| see above | MONTEFIORE MEDICAL CENTER LABORATORIES | Wadsworth Center, New York State Department of Health | Kirsten St. George, Daryl M. Lamson, Alexis Russell, Matthew Shudt, Melissa A Leisner, Jonathan Plitnick, Catharine Prussing, Navjot Singh, John Kelly, Erasmus Schneider, Erica Lasek-Nesselquist |
| EPI_ISL_1623973, EPI_ISL_1623974 | NYC Pandemic Response Lab | Wadsworth Center, New York State Department of Health | Kirsten St. George, Daryl M. Lamson, Alexis Russell, Matthew Shudt, Melissa A Leisner, Jonathan Plitnick, Catharine Prussing, Navjot Singh, John Kelly, Erasmus Schneider, Erica Lasek-Nesselquist |
| EPI_ISL_1623975, EPI_ISL_1623976, EPI_ISL_1623977, EPI_ISL_1623978, EPI_ISL_1623979, EPI_ISL_1623980, EPI_ISL_1623981, EPI_ISL_1623982, EPI_ISL_1623983, EPI_ISL_1623984, EPI_ISL_1623985, EPI_ISL_1623986, EPI_ISL_1623987, EPI_ISL_1623988, EPI_ISL_1623989 | | | |
| see above | MONTEFIORE MEDICAL CENTER LABORATORIES | Wadsworth Center, New York State Department of Health | Kirsten St. George, Daryl M. Lamson, Alexis Russell, Matthew Shudt, Melissa A Leisner, Jonathan Plitnick, Catharine Prussing, Navjot Singh, John Kelly, Erasmus Schneider, Erica Lasek-Nesselquist |
| EPI_ISL_1623990, EPI_ISL_1623991, EPI_ISL_1623992, EPI_ISL_1623993, EPI_ISL_1623994, EPI_ISL_1623995, EPI_ISL_1623996, EPI_ISL_1623997, EPI_ISL_1623998, EPI_ISL_1623999, EPI_ISL_1624000, EPI_ISL_1624001, EPI_ISL_1624002 | | | |
| see above | MEMORIAL SLOAN KETTERING CANCER CENTER | Wadsworth Center, New York State Department of Health | Kirsten St. George, Daryl M. Lamson, Alexis Russell, Matthew Shudt, Melissa A Leisner, Jonathan Plitnick, Catharine Prussing, Navjot Singh, John Kelly, Erasmus Schneider, Erica Lasek-Nesselquist |
| EPI_ISL_1624003 | New York Presbyterian Hospital | Wadsworth Center, New York State Department of Health | Kirsten St. George, Daryl M. Lamson, Alexis Russell, Matthew Shudt, Melissa A Leisner, Jonathan Plitnick, Catharine Prussing, Navjot Singh, John Kelly, Erasmus Schneider, Erica Lasek-Nesselquist |
| EPI_ISL_1624004 | NORTHWELL HEALTH LABORATORIES | Wadsworth Center, New York State Department of Health | Kirsten St. George, Daryl M. Lamson, Alexis Russell, Matthew Shudt, Melissa A Leisner, Jonathan Plitnick, Catharine Prussing, Navjot Singh, John Kelly, Erasmus Schneider, Erica Lasek-Nesselquist |
| EPI_ISL_1624005, EPI_ISL_1624006, EPI_ISL_1624007, EPI_ISL_1624008, EPI_ISL_1624009 | MEMORIAL SLOAN KETTERING CANCER CENTER | Wadsworth Center, New York State Department of Health | Kirsten St. George, Daryl M. Lamson, Alexis Russell, Matthew Shudt, Melissa A Leisner, Jonathan Plitnick, Catharine Prussing, Navjot Singh, John Kelly, Erasmus Schneider, Erica Lasek-Nesselquist |
| EPI_ISL_1624010, EPI_ISL_1624011 | Columbia University Irving Medical Center | Wadsworth Center, New York State Department of Health | Kirsten St. George, Daryl M. Lamson, Alexis Russell, Matthew Shudt, Melissa A Leisner, Jonathan Plitnick, Catharine Prussing, Navjot Singh, John Kelly, Erasmus Schneider, Erica Lasek-Nesselquist |
| EPI_ISL_1624012, EPI_ISL_1624013, EPI_ISL_1624014, EPI_ISL_1624015, EPI_ISL_1624016, EPI_ISL_1624017, EPI_ISL_1624018, EPI_ISL_1624019, EPI_ISL_1624020, EPI_ISL_1624021, EPI_ISL_1624022, EPI_ISL_1624023, EPI_ISL_1624024, EPI_ISL_1624025, EPI_ISL_1624026, EPI_ISL_1624027, EPI_ISL_1624028, EPI_ISL_1624029, EPI_ISL_1624030, EPI_ISL_1624031, EPI_ISL_1624032, EPI_ISL_1624033, EPI_ISL_1624034, EPI_ISL_1624035, EPI_ISL_1624036, EPI_ISL_1624037, EPI_ISL_1624038, EPI_ISL_1624039, EPI_ISL_1624040, EPI_ISL_1624041, EPI_ISL_1624042, EPI_ISL_1624043, EPI_ISL_1624044, EPI_ISL_1624045, EPI_ISL_1624046, EPI_ISL_1624047, EPI_ISL_1624048, EPI_ISL_1624049, EPI_ISL_1624050, EPI_ISL_1624051, EPI_ISL_1624052, EPI_ISL_1624053, EPI_ISL_1624054, EPI_ISL_1624055, EPI_ISL_1624056, EPI_ISL_1624057, EPI_ISL_1624058, EPI_ISL_1624059, EPI_ISL_1624060, EPI_ISL_1624061, EPI_ISL_1624062, EPI_ISL_1624063, EPI_ISL_1624064, EPI_ISL_1624065, EPI_ISL_1624066, EPI_ISL_1624067, EPI_ISL_1624068, EPI_ISL_1624069, EPI_ISL_1624070, EPI_ISL_1624071, EPI_ISL_1624072, EPI_ISL_1624073, EPI_ISL_1624074, EPI_ISL_1624075, EPI_ISL_1624076, EPI_ISL_1624077, EPI_ISL_1624078, EPI_ISL_1624079, EPI_ISL_1624080, EPI_ISL_1624081, EPI_ISL_1624082 | | | |
| see above | New York Presbyterian Hospital | Wadsworth Center, New York State Department of Health | Kirsten St. George, Daryl M. Lamson, Alexis Russell, Matthew Shudt, Melissa A Leisner, Jonathan Plitnick, Catharine Prussing, Navjot Singh, John Kelly, Erasmus Schneider, Erica Lasek-Nesselquist |
| EPI_ISL_1624083, EPI_ISL_1624084, EPI_ISL_1624085, EPI_ISL_1624086, EPI_ISL_1624087, EPI_ISL_1624088 | NYC Pandemic Response Lab | Wadsworth Center, New York State Department of Health | Kirsten St. George, Daryl M. Lamson, Alexis Russell, Matthew Shudt, Melissa A Leisner, Jonathan Plitnick, Catharine Prussing, Navjot Singh, John Kelly, Erasmus Schneider, Erica Lasek-Nesselquist |
| EPI_ISL_1624089, EPI_ISL_1624090 | GOOD SAMARITAN HOSPITAL LABORATORY | Wadsworth Center, New York State Department of Health | Kirsten St. George, Daryl M. Lamson, Alexis Russell, Matthew Shudt, Melissa A Leisner, Jonathan Plitnick, Catharine Prussing, Navjot Singh, John Kelly, Erasmus Schneider, Erica Lasek-Nesselquist |
| EPI_ISL_1624273, EPI_ISL_1624274, EPI_ISL_1624275 | USNS Comfort T-AH 20 | Naval Medical Research Center Biological Defense Research Directorate | Logan Voegtly, Lindsay Glang, Francisco Malagon Bautista, Kyle Long, Regina Cer, Kimberly Bishop-Lilly |
| EPI_ISL_1624404, EPI_ISL_1624405 | Columbia University Irving Medical Center | Wadsworth Center, New York State Department of Health | Kirsten St. George, Daryl M. Lamson, Alexis Russell, Matthew Shudt, Melissa A Leisner, Jonathan Plitnick, Catharine Prussing, Navjot Singh, John Kelly, Erasmus Schneider, Erica Lasek-Nesselquist |
| EPI_ISL_1624406, EPI_ISL_1624407, EPI_ISL_1624408 | SUNY UPSTATE MEDICAL UNIVERSITY | Wadsworth Center, New York State Department of Health | Kirsten St. George, Daryl M. Lamson, Alexis Russell, Matthew Shudt, Melissa A Leisner, Jonathan Plitnick, Catharine Prussing, Navjot Singh, John Kelly, Erasmus Schneider, Erica Lasek-Nesselquist |
| EPI_ISL_1624409 | BOSTON HEART DIAGNOSTICS CORP | Wadsworth Center, New York State Department of Health | Kirsten St. George, Daryl M. Lamson, Alexis Russell, Matthew Shudt, Melissa A Leisner, Jonathan Plitnick, Catharine Prussing, Navjot Singh, John Kelly, Erasmus Schneider, Erica Lasek-Nesselquist |
| EPI_ISL_1624410, EPI_ISL_1624411, EPI_ISL_1624412, EPI_ISL_1624413, EPI_ISL_1624414, EPI_ISL_1624415, EPI_ISL_1624416, EPI_ISL_1624417, EPI_ISL_1624418, EPI_ISL_1624419 | URMC LABS | Wadsworth Center, New York State Department of Health | Kirsten St. George, Daryl M. Lamson, Alexis Russell, Matthew Shudt, Melissa A Leisner, Jonathan Plitnick, Catharine Prussing, Navjot Singh, John Kelly, Erasmus Schneider, Erica Lasek-Nesselquist |
| EPI_ISL_1624420 | SUNY UPSTATE MEDICAL UNIVERSITY | Wadsworth Center, New York State Department of Health | Kirsten St. George, Daryl M. Lamson, Alexis Russell, Matthew Shudt, Melissa A Leisner, Jonathan Plitnick, Catharine Prussing, Navjot Singh, John Kelly, Erasmus Schneider, Erica Lasek-Nesselquist |
| EPI_ISL_1624421, EPI_ISL_1624422, EPI_ISL_1624423, EPI_ISL_1624424, EPI_ISL_1624425, EPI_ISL_1624426, EPI_ISL_1624427, EPI_ISL_1624428, EPI_ISL_1624429, EPI_ISL_1624430, EPI_ISL_1624431, EPI_ISL_1624432, EPI_ISL_1624433, EPI_ISL_1624434, EPI_ISL_1624435, EPI_ISL_1624436, EPI_ISL_1624437, EPI_ISL_1624438, EPI_ISL_1624439, EPI_ISL_1624440, EPI_ISL_1624441, EPI_ISL_1624442, EPI_ISL_1624443, EPI_ISL_1624444, EPI_ISL_1624445, EPI_ISL_1624446, EPI_ISL_1624447, EPI_ISL_1624448, EPI_ISL_1624449, EPI_ISL_1624450, EPI_ISL_1624451, EPI_ISL_1624452, EPI_ISL_1624453, EPI_ISL_1624454, EPI_ISL_1624455, EPI_ISL_1624456, EPI_ISL_1624457, EPI_ISL_1624458, EPI_ISL_1624459, EPI_ISL_1624460, EPI_ISL_1624461, EPI_ISL_1624462, EPI_ISL_1624463, EPI_ISL_1624464, EPI_ISL_1624465, EPI_ISL_1624466, EPI_ISL_1624467, EPI_ISL_1624468, EPI_ISL_1624469, EPI_ISL_1624470, EPI_ISL_1624471, EPI_ISL_1624472, EPI_ISL_1624473, EPI_ISL_1624474, EPI_ISL_1624475, EPI_ISL_1624476, EPI_ISL_1624477, EPI_ISL_1624478, EPI_ISL_1624479, EPI_ISL_1624480, EPI_ISL_1624481, EPI_ISL_1624482 | | | |
| see above | URMC LABS | Wadsworth Center, New York State Department of Health | Kirsten St. George, Daryl M. Lamson, Alexis Russell, Matthew Shudt, Melissa A Leisner, Jonathan Plitnick, Catharine Prussing, Navjot Singh, John Kelly, Erasmus Schneider, Erica Lasek-Nesselquist |
| EPI_ISL_1624483, EPI_ISL_1624484, EPI_ISL_1624485, EPI_ISL_1624486 | SARATOGA HOSPITAL LABORATORY | Wadsworth Center, New York State Department of Health | Kirsten St. George, Daryl M. Lamson, Alexis Russell, Matthew Shudt, Melissa A Leisner, Jonathan Plitnick, Catharine Prussing, Navjot Singh, John Kelly, Erasmus Schneider, Erica Lasek-Nesselquist |
| EPI_ISL_1624487 | CHAMPLAIN VALLEY PHYSICIANS HOSPITAL MEDICAL CENTER | Wadsworth Center, New York State Department of Health | Kirsten St. George, Daryl M. Lamson, Alexis Russell, Matthew Shudt, Melissa A Leisner, Jonathan Plitnick, Catharine Prussing, Navjot Singh, John Kelly, Erasmus Schneider, Erica Lasek-Nesselquist |
| EPI_ISL_1625677 | The Jackson Laboratory | The Jackson Laboratory | Bergeron D, Renzette N, Adams M, Omerza G, Kelly K, Long J, Li L |
| EPI_ISL_1625770 | Pathology and Laboratory Medicine Institute, Cleveland Clinic, Ohio, USA | Pathology and Laboratory Medicine Institute, Cleveland Clinic, Ohio, USA | Jennifer Starbuck, Joy Nakitandwe, Kristen McDonnell, David Plunkett, Zheng Jin Tu, Jay Brock, Yu-Wei Cheng, Gary Procop, Daniel Rhoads, Daniel H. Farkas, David Bosler |

| | | | |
|---|---|---|---|
| EPI_ISL_1633261 | Pathology and Laboratory Medicine Institute, Cleveland Clinic, Ohio, USA | Pathology and Laboratory Medicine Institute, Cleveland Clinic, Ohio, USA | Concetta Peck, Joy Nakitandwe, Kristen McDonnell, David Plunkett, Zheng Jin Tu, Jay Brock, Yu-Wei Cheng, Gary Procop, Daniel Rhoads, Daniel H. Farkas, David Bosler |

EPI_ISL_1633685, EPI_ISL_1633686, EPI_ISL_1633687, EPI_ISL_1633688, EPI_ISL_1633689, EPI_ISL_1633690, EPI_ISL_1633691, EPI_ISL_1633692, EPI_ISL_1633693, EPI_ISL_1633694, EPI_ISL_1633695, EPI_ISL_1633696, EPI_ISL_1633697, EPI_ISL_1633698, EPI_ISL_1633699, EPI_ISL_1633700, EPI_ISL_1633701,
EPI_ISL_1633702, EPI_ISL_1633703, EPI_ISL_1633704, EPI_ISL_1633705, EPI_ISL_1633706, EPI_ISL_1633707, EPI_ISL_1633708, EPI_ISL_1633709, EPI_ISL_1633710, EPI_ISL_1633711, EPI_ISL_1633712, EPI_ISL_1633713, EPI_ISL_1633714, EPI_ISL_1633715, EPI_ISL_1633716, EPI_ISL_1633717, EPI_ISL_1633718,
EPI_ISL_1633719, EPI_ISL_1633720, EPI_ISL_1633721, EPI_ISL_1633722, EPI_ISL_1633723, EPI_ISL_1633724, EPI_ISL_1633725, EPI_ISL_1633726, EPI_ISL_1633727, EPI_ISL_1633728, EPI_ISL_1633729, EPI_ISL_1633730, EPI_ISL_1633731, EPI_ISL_1633732, EPI_ISL_1633733, EPI_ISL_1633734, EPI_ISL_1633735,
EPI_ISL_1633736, EPI_ISL_1633737, EPI_ISL_1633738, EPI_ISL_1633739, EPI_ISL_1633740, EPI_ISL_1633741, EPI_ISL_1633742, EPI_ISL_1633743, EPI_ISL_1633744, EPI_ISL_1633745, EPI_ISL_1633746, EPI_ISL_1633747, EPI_ISL_1633748, EPI_ISL_1633749, EPI_ISL_1633750, EPI_ISL_1633751, EPI_ISL_1633752,
EPI_ISL_1633753, EPI_ISL_1633754, EPI_ISL_1633755, EPI_ISL_1633756, EPI_ISL_1633757, EPI_ISL_1633758, EPI_ISL_1633759, EPI_ISL_1633760, EPI_ISL_1633761, EPI_ISL_1633762, EPI_ISL_1633763, EPI_ISL_1633764, EPI_ISL_1633765, EPI_ISL_1633766, EPI_ISL_1633767, EPI_ISL_1633768, EPI_ISL_1633769,
EPI_ISL_1633770, EPI_ISL_1633771, EPI_ISL_1633772, EPI_ISL_1633773, EPI_ISL_1633774, EPI_ISL_1633775, EPI_ISL_1633776, EPI_ISL_1633777, EPI_ISL_1633778, EPI_ISL_1633779, EPI_ISL_1633780, EPI_ISL_1633781, EPI_ISL_1633782, EPI_ISL_1633783, EPI_ISL_1633784, EPI_ISL_1633785, EPI_ISL_1633786,
EPI_ISL_1633787, EPI_ISL_1633788, EPI_ISL_1633789, EPI_ISL_1633790, EPI_ISL_1633791, EPI_ISL_1633792, EPI_ISL_1633793, EPI_ISL_1633794, EPI_ISL_1633795, EPI_ISL_1633796, EPI_ISL_1633797, EPI_ISL_1633798, EPI_ISL_1633799, EPI_ISL_1633800, EPI_ISL_1633801, EPI_ISL_1633802, EPI_ISL_1633803,
EPI_ISL_1633804, EPI_ISL_1633805, EPI_ISL_1633806, EPI_ISL_1633807, EPI_ISL_1633808, EPI_ISL_1633809, EPI_ISL_1633810, EPI_ISL_1633811, EPI_ISL_1633812, EPI_ISL_1633813, EPI_ISL_1633814, EPI_ISL_1633815, EPI_ISL_1633816, EPI_ISL_1633817, EPI_ISL_1633818, EPI_ISL_1633819, EPI_ISL_1633820,
EPI_ISL_1633821, EPI_ISL_1633822, EPI_ISL_1633823, EPI_ISL_1633824, EPI_ISL_1633825, EPI_ISL_1633826, EPI_ISL_1633827, EPI_ISL_1633828, EPI_ISL_1633829, EPI_ISL_1633830, EPI_ISL_1633831, EPI_ISL_1633832, EPI_ISL_1633833, EPI_ISL_1633834, EPI_ISL_1633835, EPI_ISL_1633836, EPI_ISL_1633837,
EPI_ISL_1633838, EPI_ISL_1633839, EPI_ISL_1633840, EPI_ISL_1633841, EPI_ISL_1633842, EPI_ISL_1633843, EPI_ISL_1633844, EPI_ISL_1633845, EPI_ISL_1633846, EPI_ISL_1633847, EPI_ISL_1633848, EPI_ISL_1633849, EPI_ISL_1633850, EPI_ISL_1633851, EPI_ISL_1633852, EPI_ISL_1633853, EPI_ISL_1633854,
EPI_ISL_1633855, EPI_ISL_1633856, EPI_ISL_1633857, EPI_ISL_1633858, EPI_ISL_1633859, EPI_ISL_1633860, EPI_ISL_1633861, EPI_ISL_1633862, EPI_ISL_1633863, EPI_ISL_1633864, EPI_ISL_1633865, EPI_ISL_1633866, EPI_ISL_1633867, EPI_ISL_1633868, EPI_ISL_1633869, EPI_ISL_1633870, EPI_ISL_1633871,
EPI_ISL_1633872, EPI_ISL_1633873, EPI_ISL_1633874, EPI_ISL_1633875, EPI_ISL_1633876, EPI_ISL_1633877, EPI_ISL_1633878, EPI_ISL_1633879, EPI_ISL_1633880, EPI_ISL_1633881, EPI_ISL_1633882, EPI_ISL_1633883, EPI_ISL_1633884, EPI_ISL_1633885, EPI_ISL_1633886, EPI_ISL_1633887, EPI_ISL_1633888,
EPI_ISL_1633889, EPI_ISL_1633890, EPI_ISL_1633891, EPI_ISL_1633892, EPI_ISL_1633893, EPI_ISL_1633894, EPI_ISL_1633895, EPI_ISL_1633896, EPI_ISL_1633897, EPI_ISL_1633899, EPI_ISL_1633901, EPI_ISL_1633902, EPI_ISL_1633903, EPI_ISL_1633905, EPI_ISL_1633906, EPI_ISL_1633907, EPI_ISL_1633909,
EPI_ISL_1633910, EPI_ISL_1633911, EPI_ISL_1633913, EPI_ISL_1633914, EPI_ISL_1633916, EPI_ISL_1633917, EPI_ISL_1633918, EPI_ISL_1633920, EPI_ISL_1633921, EPI_ISL_1633924, EPI_ISL_1633925, EPI_ISL_1633927, EPI_ISL_1633929, EPI_ISL_1633930, EPI_ISL_1633931, EPI_ISL_1633932,
EPI_ISL_1633934, EPI_ISL_1633935, EPI_ISL_1633937, EPI_ISL_1633938, EPI_ISL_1633939, EPI_ISL_1633940, EPI_ISL_1633942, EPI_ISL_1633943, EPI_ISL_1633944, EPI_ISL_1633946, EPI_ISL_1633947, EPI_ISL_1633949, EPI_ISL_1633951, EPI_ISL_1633953, EPI_ISL_1633954, EPI_ISL_1633956, EPI_ISL_1633958,
EPI_ISL_1633959, EPI_ISL_1633960, EPI_ISL_1633962, EPI_ISL_1633963, EPI_ISL_1633964, EPI_ISL_1633966, EPI_ISL_1633967, EPI_ISL_1633968, EPI_ISL_1633970, EPI_ISL_1633971, EPI_ISL_1633972, EPI_ISL_1633973, EPI_ISL_1633975, EPI_ISL_1633976, EPI_ISL_1633977, EPI_ISL_1633979, EPI_ISL_1633980,
EPI_ISL_1633981, EPI_ISL_1633983, EPI_ISL_1633984, EPI_ISL_1633985, EPI_ISL_1633986, EPI_ISL_1633988, EPI_ISL_1633989, EPI_ISL_1633990, EPI_ISL_1633992, EPI_ISL_1633993, EPI_ISL_1633994, EPI_ISL_1633996, EPI_ISL_1633997, EPI_ISL_1633998, EPI_ISL_1634000, EPI_ISL_1634001, EPI_ISL_1634002,
EPI_ISL_1634004, EPI_ISL_1634005, EPI_ISL_1634007, EPI_ISL_1634008, EPI_ISL_1634009, EPI_ISL_1634011, EPI_ISL_1634012, EPI_ISL_1634014, EPI_ISL_1634016, EPI_ISL_1634018, EPI_ISL_1634019, EPI_ISL_1634021, EPI_ISL_1634022, EPI_ISL_1634023, EPI_ISL_1634025, EPI_ISL_1634026, EPI_ISL_1634028,
EPI_ISL_1634029, EPI_ISL_1634030, EPI_ISL_1634032, EPI_ISL_1634033, EPI_ISL_1634034, EPI_ISL_1634036, EPI_ISL_1634037, EPI_ISL_1634038, EPI_ISL_1634040, EPI_ISL_1634041, EPI_ISL_1634042, EPI_ISL_1634044, EPI_ISL_1634045, EPI_ISL_1634046, EPI_ISL_1634047, EPI_ISL_1634049, EPI_ISL_1634051,
EPI_ISL_1634052, EPI_ISL_1634053, EPI_ISL_1634054, EPI_ISL_1634056, EPI_ISL_1634058, EPI_ISL_1634060, EPI_ISL_1634061, EPI_ISL_1634062, EPI_ISL_1634064, EPI_ISL_1634065, EPI_ISL_1634066, EPI_ISL_1634068, EPI_ISL_1634069, EPI_ISL_1634071, EPI_ISL_1634072, EPI_ISL_1634074, EPI_ISL_1634076,
EPI_ISL_1634077, EPI_ISL_1634079, EPI_ISL_1634080, EPI_ISL_1634082, EPI_ISL_1634083, EPI_ISL_1634084, EPI_ISL_1634086, EPI_ISL_1634087, EPI_ISL_1634088, EPI_ISL_1634089, EPI_ISL_1634091, EPI_ISL_1634092, EPI_ISL_1634093, EPI_ISL_1634095, EPI_ISL_1634096, EPI_ISL_1634097, EPI_ISL_1634099,
EPI_ISL_1634100, EPI_ISL_1634101, EPI_ISL_1634103, EPI_ISL_1634104, EPI_ISL_1634105, EPI_ISL_1634107, EPI_ISL_1634108, EPI_ISL_1634110, EPI_ISL_1634112, EPI_ISL_1634113, EPI_ISL_1634114, EPI_ISL_1634116, EPI_ISL_1634117, EPI_ISL_1634118, EPI_ISL_1634120, EPI_ISL_1634121, EPI_ISL_1634123,
EPI_ISL_1634124, EPI_ISL_1634125, EPI_ISL_1634126, EPI_ISL_1634128, EPI_ISL_1634129, EPI_ISL_1634130, EPI_ISL_1634132, EPI_ISL_1634133, EPI_ISL_1634135, EPI_ISL_1634136, EPI_ISL_1634137, EPI_ISL_1634139, EPI_ISL_1634140, EPI_ISL_1634141, EPI_ISL_1634143, EPI_ISL_1634150, EPI_ISL_1634152,
EPI_ISL_1634153, EPI_ISL_1634155, EPI_ISL_1634157, EPI_ISL_1634158, EPI_ISL_1634159, EPI_ISL_1634161, EPI_ISL_1634162, EPI_ISL_1634163, EPI_ISL_1634165, EPI_ISL_1634166, EPI_ISL_1634167, EPI_ISL_1634169, EPI_ISL_1634170, EPI_ISL_1634171, EPI_ISL_1634173, EPI_ISL_1634174, EPI_ISL_1634175,
EPI_ISL_1634176, EPI_ISL_1634177, EPI_ISL_1634178, EPI_ISL_1634179, EPI_ISL_1634180, EPI_ISL_1634181, EPI_ISL_1634182, EPI_ISL_1634183, EPI_ISL_1634184, EPI_ISL_1634185, EPI_ISL_1634186, EPI_ISL_1634187, EPI_ISL_1634188, EPI_ISL_1634189, EPI_ISL_1634190, EPI_ISL_1634191, EPI_ISL_1634192,
EPI_ISL_1634193, EPI_ISL_1634194, EPI_ISL_1634195, EPI_ISL_1634196, EPI_ISL_1634197, EPI_ISL_1634198, EPI_ISL_1634199, EPI_ISL_1634200, EPI_ISL_1634201, EPI_ISL_1634202, EPI_ISL_1634203, EPI_ISL_1634204, EPI_ISL_1634205, EPI_ISL_1634206, EPI_ISL_1634207, EPI_ISL_1634208, EPI_ISL_1634209,
EPI_ISL_1634210, EPI_ISL_1634211, EPI_ISL_1634212, EPI_ISL_1634213, EPI_ISL_1634214, EPI_ISL_1634215, EPI_ISL_1634216, EPI_ISL_1634217, EPI_ISL_1634218, EPI_ISL_1634219, EPI_ISL_1634220, EPI_ISL_1634221, EPI_ISL_1634222, EPI_ISL_1634223, EPI_ISL_1634224, EPI_ISL_1634225, EPI_ISL_1634226,
EPI_ISL_1634227, EPI_ISL_1634228, EPI_ISL_1634229, EPI_ISL_1634230, EPI_ISL_1634231, EPI_ISL_1634232, EPI_ISL_1634233, EPI_ISL_1634234, EPI_ISL_1634235, EPI_ISL_1634236, EPI_ISL_1634237, EPI_ISL_1634238, EPI_ISL_1634239, EPI_ISL_1634240, EPI_ISL_1634241, EPI_ISL_1634242, EPI_ISL_1634243,
EPI_ISL_1634244, EPI_ISL_1634245, EPI_ISL_1634246, EPI_ISL_1634247, EPI_ISL_1634248, EPI_ISL_1634249, EPI_ISL_1634250, EPI_ISL_1634251, EPI_ISL_1634252, EPI_ISL_1634253, EPI_ISL_1634254, EPI_ISL_1634255, EPI_ISL_1634256, EPI_ISL_1634257, EPI_ISL_1634258, EPI_ISL_1634259, EPI_ISL_1634260,
EPI_ISL_1634261, EPI_ISL_1634262, EPI_ISL_1634263, EPI_ISL_1634264, EPI_ISL_1634265, EPI_ISL_1634266, EPI_ISL_1634267, EPI_ISL_1634268, EPI_ISL_1634269, EPI_ISL_1634270, EPI_ISL_1634271, EPI_ISL_1634272, EPI_ISL_1634273, EPI_ISL_1634274, EPI_ISL_1634275, EPI_ISL_1634276, EPI_ISL_1634277,
EPI_ISL_1634278, EPI_ISL_1634279, EPI_ISL_1634280, EPI_ISL_1634281, EPI_ISL_1634282, EPI_ISL_1634283, EPI_ISL_1634284, EPI_ISL_1634285, EPI_ISL_1634286, EPI_ISL_1634287, EPI_ISL_1634288, EPI_ISL_1634289, EPI_ISL_1634290, EPI_ISL_1634291, EPI_ISL_1634292, EPI_ISL_1634293, EPI_ISL_1634294,
EPI_ISL_1634295, EPI_ISL_1634296, EPI_ISL_1634297, EPI_ISL_1634298, EPI_ISL_1634299, EPI_ISL_1634300, EPI_ISL_1634301, EPI_ISL_1634302, EPI_ISL_1634303, EPI_ISL_1634304, EPI_ISL_1634305, EPI_ISL_1634306, EPI_ISL_1634307, EPI_ISL_1634308, EPI_ISL_1634309, EPI_ISL_1634310, EPI_ISL_1634311,
EPI_ISL_1634312, EPI_ISL_1634313, EPI_ISL_1634314, EPI_ISL_1634315, EPI_ISL_1634316, EPI_ISL_1634317, EPI_ISL_1634318, EPI_ISL_1634319, EPI_ISL_1634320, EPI_ISL_1634321, EPI_ISL_1634322, EPI_ISL_1634323, EPI_ISL_1634324, EPI_ISL_1634325, EPI_ISL_1634326, EPI_ISL_1634327, EPI_ISL_1634328,
EPI_ISL_1634329, EPI_ISL_1634330, EPI_ISL_1634331, EPI_ISL_1634332, EPI_ISL_1634333, EPI_ISL_1634334, EPI_ISL_1634335, EPI_ISL_1634336, EPI_ISL_1634337, EPI_ISL_1634338, EPI_ISL_1634339, EPI_ISL_1635437, EPI_ISL_1635438, EPI_ISL_1635439, EPI_ISL_1635440, EPI_ISL_1635441, EPI_ISL_1635442,
EPI_ISL_1635443, EPI_ISL_1635444, EPI_ISL_1635445, EPI_ISL_1635446, EPI_ISL_1635447, EPI_ISL_1635448, EPI_ISL_1635449, EPI_ISL_1635450, EPI_ISL_1635451, EPI_ISL_1635452, EPI_ISL_1635453, EPI_ISL_1635454, EPI_ISL_1635455, EPI_ISL_1635456, EPI_ISL_1635457, EPI_ISL_1635458, EPI_ISL_1635459,
EPI_ISL_1635460, EPI_ISL_1635461, EPI_ISL_1635462, EPI_ISL_1635463, EPI_ISL_1635464, EPI_ISL_1635465, EPI_ISL_1635466, EPI_ISL_1635467, EPI_ISL_1635468, EPI_ISL_1635469, EPI_ISL_1635470, EPI_ISL_1635471, EPI_ISL_1635472, EPI_ISL_1635473, EPI_ISL_1635474, EPI_ISL_1635475, EPI_ISL_1635476,
EPI_ISL_1635477, EPI_ISL_1635478, EPI_ISL_1635479, EPI_ISL_1635480, EPI_ISL_1635481, EPI_ISL_1635482, EPI_ISL_1635483, EPI_ISL_1635484, EPI_ISL_1635485, EPI_ISL_1635486, EPI_ISL_1635487, EPI_ISL_1635488, EPI_ISL_1635489, EPI_ISL_1635490, EPI_ISL_1635491, EPI_ISL_1635492, EPI_ISL_1635493,
EPI_ISL_1635494, EPI_ISL_1635495, EPI_ISL_1635496, EPI_ISL_1635497, EPI_ISL_1635498, EPI_ISL_1635499, EPI_ISL_1635500, EPI_ISL_1635501, EPI_ISL_1635502, EPI_ISL_1635503, EPI_ISL_1635504, EPI_ISL_1635505, EPI_ISL_1635506, EPI_ISL_1635507, EPI_ISL_1635508, EPI_ISL_1635509, EPI_ISL_1635510,
EPI_ISL_1635511, EPI_ISL_1635512, EPI_ISL_1635513, EPI_ISL_1635514, EPI_ISL_1635515, EPI_ISL_1635516, EPI_ISL_1635517, EPI_ISL_1635518, EPI_ISL_1635519, EPI_ISL_1635520, EPI_ISL_1635521, EPI_ISL_1635522, EPI_ISL_1635523, EPI_ISL_1635524, EPI_ISL_1635525, EPI_ISL_1635526, EPI_ISL_1635527,
EPI_ISL_1635528, EPI_ISL_1635529, EPI_ISL_1635530, EPI_ISL_1635531, EPI_ISL_1635532, EPI_ISL_1635533, EPI_ISL_1635534, EPI_ISL_1635535, EPI_ISL_1635536, EPI_ISL_1635537, EPI_ISL_1635538, EPI_ISL_1635539, EPI_ISL_1635540, EPI_ISL_1635541, EPI_ISL_1635542, EPI_ISL_1635543, EPI_ISL_1635544,
EPI_ISL_1635545, EPI_ISL_1635546, EPI_ISL_1635547, EPI_ISL_1635548, EPI_ISL_1635549, EPI_ISL_1635550, EPI_ISL_1635551, EPI_ISL_1635552, EPI_ISL_1635553, EPI_ISL_1635554, EPI_ISL_1635555, EPI_ISL_1635556, EPI_ISL_1635557, EPI_ISL_1635558, EPI_ISL_1635559, EPI_ISL_1635560, EPI_ISL_1635561,
EPI_ISL_1635562, EPI_ISL_1635563, EPI_ISL_1635564, EPI_ISL_1635565, EPI_ISL_1635566, EPI_ISL_1635567, EPI_ISL_1635568, EPI_ISL_1635569, EPI_ISL_1635570, EPI_ISL_1635571, EPI_ISL_1635572, EPI_ISL_1635573, EPI_ISL_1635574, EPI_ISL_1635575, EPI_ISL_1635576, EPI_ISL_1635577, EPI_ISL_1635578,
EPI_ISL_1635579, EPI_ISL_1635580, EPI_ISL_1635581, EPI_ISL_1635582, EPI_ISL_1635583, EPI_ISL_1635584, EPI_ISL_1635585, EPI_ISL_1635586, EPI_ISL_1635587, EPI_ISL_1635588, EPI_ISL_1635589, EPI_ISL_1635590, EPI_ISL_1635591, EPI_ISL_1635592, EPI_ISL_1635593, EPI_ISL_1635594, EPI_ISL_1635595,
EPI_ISL_1635596, EPI_ISL_1635597, EPI_ISL_1635598, EPI_ISL_1635599, EPI_ISL_1635600, EPI_ISL_1635601, EPI_ISL_1635602, EPI_ISL_1635603, EPI_ISL_1635604, EPI_ISL_1635605, EPI_ISL_1635606, EPI_ISL_1635607, EPI_ISL_1635608, EPI_ISL_1635609, EPI_ISL_1635610, EPI_ISL_1635611, EPI_ISL_1635612,
EPI_ISL_1635613, EPI_ISL_1635614, EPI_ISL_1635615, EPI_ISL_1635616, EPI_ISL_1635617, EPI_ISL_1635618, EPI_ISL_1635619, EPI_ISL_1635620, EPI_ISL_1635621, EPI_ISL_1635622, EPI_ISL_1635623, EPI_ISL_1635624, EPI_ISL_1635625, EPI_ISL_1635626, EPI_ISL_1635627, EPI_ISL_1635628, EPI_ISL_1635629,
EPI_ISL_1635630, EPI_ISL_1635631, EPI_ISL_1635632, EPI_ISL_1635633, EPI_ISL_1635634, EPI_ISL_1635635, EPI_ISL_1635636, EPI_ISL_1635637, EPI_ISL_1635638, EPI_ISL_1635639, EPI_ISL_1635640, EPI_ISL_1635641, EPI_ISL_1635642, EPI_ISL_1635643, EPI_ISL_1635644, EPI_ISL_1635645, EPI_ISL_1635646,
EPI_ISL_1635647, EPI_ISL_1635648, EPI_ISL_1635649, EPI_ISL_1635650, EPI_ISL_1635651, EPI_ISL_1635652, EPI_ISL_1635653, EPI_ISL_1635654, EPI_ISL_1635655, EPI_ISL_1635656, EPI_ISL_1635657, EPI_ISL_1635658, EPI_ISL_1635659, EPI_ISL_1635660, EPI_ISL_1635661, EPI_ISL_1635662, EPI_ISL_1635663,
EPI_ISL_1635664, EPI_ISL_1635665, EPI_ISL_1635667, EPI_ISL_1635668, EPI_ISL_1635669, EPI_ISL_1635670, EPI_ISL_1635671, EPI_ISL_1635672, EPI_ISL_1635673, EPI_ISL_1635674, EPI_ISL_1635675, EPI_ISL_1635676, EPI_ISL_1635677, EPI_ISL_1635678, EPI_ISL_1635679, EPI_ISL_1635680, EPI_ISL_1635681,
EPI_ISL_1635682, EPI_ISL_1635683, EPI_ISL_1635684, EPI_ISL_1635685, EPI_ISL_1635686, EPI_ISL_1635687, EPI_ISL_1635688, EPI_ISL_1635689, EPI_ISL_1635690, EPI_ISL_1635691, EPI_ISL_1635692, EPI_ISL_1635693, EPI_ISL_1635694, EPI_ISL_1635695, EPI_ISL_1635696, EPI_ISL_1635697, EPI_ISL_1635698,
EPI_ISL_1635699, EPI_ISL_1635700, EPI_ISL_1635701, EPI_ISL_1635702, EPI_ISL_1635703, EPI_ISL_1635704, EPI_ISL_1635705, EPI_ISL_1635706, EPI_ISL_1635707, EPI_ISL_1635708, EPI_ISL_1635709, EPI_ISL_1635710, EPI_ISL_1635711, EPI_ISL_1635712, EPI_ISL_1635713, EPI_ISL_1635714, EPI_ISL_1635715,
EPI_ISL_1635716, EPI_ISL_1635717, EPI_ISL_1635718, EPI_ISL_1635719, EPI_ISL_1635720, EPI_ISL_1635721, EPI_ISL_1635722, EPI_ISL_1635723, EPI_ISL_1635724, EPI_ISL_1635725, EPI_ISL_1635726, EPI_ISL_1635727, EPI_ISL_1635728, EPI_ISL_1635729, EPI_ISL_1635730, EPI_ISL_1635731, EPI_ISL_1635732,
EPI_ISL_1635733, EPI_ISL_1635734, EPI_ISL_1635735, EPI_ISL_1635736, EPI_ISL_1635737, EPI_ISL_1635738, EPI_ISL_1635739, EPI_ISL_1635740, EPI_ISL_1635741, EPI_ISL_1635742, EPI_ISL_1635743, EPI_ISL_1635744, EPI_ISL_1635745, EPI_ISL_1635746, EPI_ISL_1635747, EPI_ISL_1635748, EPI_ISL_1635749,
EPI_ISL_1635750, EPI_ISL_1635751, EPI_ISL_1635752, EPI_ISL_1635753, EPI_ISL_1635755, EPI_ISL_1635756, EPI_ISL_1635757, EPI_ISL_1635758, EPI_ISL_1635759, EPI_ISL_1635760, EPI_ISL_1635761, EPI_ISL_1635762, EPI_ISL_1635763, EPI_ISL_1635764, EPI_ISL_1635765, EPI_ISL_1635766, EPI_ISL_1635767,
EPI_ISL_1635768, EPI_ISL_1635769, EPI_ISL_1635770, EPI_ISL_1635771, EPI_ISL_1635772, EPI_ISL_1635773, EPI_ISL_1635774, EPI_ISL_1635775, EPI_ISL_1635776, EPI_ISL_1635777, EPI_ISL_1635778, EPI_ISL_1635779, EPI_ISL_1635780, EPI_ISL_1635781, EPI_ISL_1635782, EPI_ISL_1635783, EPI_ISL_1635784,
EPI_ISL_1635785, EPI_ISL_1635786, EPI_ISL_1635787, EPI_ISL_1635788, EPI_ISL_1635789, EPI_ISL_1635790, EPI_ISL_1635791, EPI_ISL_1635792, EPI_ISL_1635793, EPI_ISL_1635794, EPI_ISL_1635795, EPI_ISL_1635796, EPI_ISL_1635797, EPI_ISL_1635798, EPI_ISL_1635799, EPI_ISL_1635800, EPI_ISL_1635801,
EPI_ISL_1635802, EPI_ISL_1635803, EPI_ISL_1635804, EPI_ISL_1635805, EPI_ISL_1635806, EPI_ISL_1635807, EPI_ISL_1635808, EPI_ISL_1635809, EPI_ISL_1635810, EPI_ISL_1635811, EPI_ISL_1635812, EPI_ISL_1635813, EPI_ISL_1635814, EPI_ISL_1635815, EPI_ISL_1635816, EPI_ISL_1635817, EPI_ISL_1635818,
EPI_ISL_1635819, EPI_ISL_1635820, EPI_ISL_1635821, EPI_ISL_1635822, EPI_ISL_1635823, EPI_ISL_1635824, EPI_ISL_1635825, EPI_ISL_1635826, EPI_ISL_1635827, EPI_ISL_1635828, EPI_ISL_1635829, EPI_ISL_1635830, EPI_ISL_1635831, EPI_ISL_1635832, EPI_ISL_1635833, EPI_ISL_1635834, EPI_ISL_1635835,
EPI_ISL_1635836, EPI_ISL_1635837, EPI_ISL_1635838, EPI_ISL_1635839, EPI_ISL_1635840, EPI_ISL_1635841, EPI_ISL_1635842, EPI_ISL_1635843, EPI_ISL_1635844, EPI_ISL_1635845, EPI_ISL_1635846, EPI_ISL_1635847, EPI_ISL_1635848, EPI_ISL_1635849, EPI_ISL_1635850, EPI_ISL_1635851, EPI_ISL_1635852,
EPI_ISL_1635853, EPI_ISL_1635854, EPI_ISL_1635855, EPI_ISL_1635856, EPI_ISL_1635857, EPI_ISL_1635858, EPI_ISL_1635859, EPI_ISL_1635860, EPI_ISL_1635861, EPI_ISL_1635862, EPI_ISL_1635863, EPI_ISL_1635864, EPI_ISL_1635865, EPI_ISL_1635866, EPI_ISL_1635867, EPI_ISL_1635868, EPI_ISL_1635869,
EPI_ISL_1635870, EPI_ISL_1635871, EPI_ISL_1635872, EPI_ISL_1635873, EPI_ISL_1635874, EPI_ISL_1635875, EPI_ISL_1635876, EPI_ISL_1635877, EPI_ISL_1635878, EPI_ISL_1635879, EPI_ISL_1635880, EPI_ISL_1635881, EPI_ISL_1635882, EPI_ISL_1635883, EPI_ISL_1635884, EPI_ISL_1635885, EPI_ISL_1635886,
EPI_ISL_1635887, EPI_ISL_1635888, EPI_ISL_1635889, EPI_ISL_1635890, EPI_ISL_1635891, EPI_ISL_1635892, EPI_ISL_1635893, EPI_ISL_1635894, EPI_ISL_1635895, EPI_ISL_1635896, EPI_ISL_1635897, EPI_ISL_1635898, EPI_ISL_1635899, EPI_ISL_1635900, EPI_ISL_1635901, EPI_ISL_1635902, EPI_ISL_1635903,
EPI_ISL_1635904, EPI_ISL_1635905, EPI_ISL_1635906, EPI_ISL_1635907, EPI_ISL_1635908, EPI_ISL_1635909, EPI_ISL_1635910, EPI_ISL_1635911, EPI_ISL_1635912, EPI_ISL_1635913, EPI_ISL_1635914, EPI_ISL_1635915, EPI_ISL_1635916, EPI_ISL_1635917, EPI_ISL_1635918, EPI_ISL_1635919, EPI_ISL_1635920,
EPI_ISL_1635921, EPI_ISL_1635922, EPI_ISL_1635923, EPI_ISL_1635924, EPI_ISL_1635925, EPI_ISL_1635926, EPI_ISL_1635927, EPI_ISL_1635928, EPI_ISL_1635929, EPI_ISL_1635930, EPI_ISL_1635931, EPI_ISL_1635932, EPI_ISL_1635933, EPI_ISL_1635934, EPI_ISL_1635935, EPI_ISL_1635936, EPI_ISL_1635937,
EPI_ISL_1635938, EPI_ISL_1635939, EPI_ISL_1635940, EPI_ISL_1635941, EPI_ISL_1635942, EPI_ISL_1635943, EPI_ISL_1635944, EPI_ISL_1635945, EPI_ISL_1635946, EPI_ISL_1635947, EPI_ISL_1635948, EPI_ISL_1635949, EPI_ISL_1635950, EPI_ISL_1635951, EPI_ISL_1635952, EPI_ISL_1635953, EPI_ISL_1635954,
EPI_ISL_1635955, EPI_ISL_1635956, EPI_ISL_1635957, EPI_ISL_1635958, EPI_ISL_1635959, EPI_ISL_1635960, EPI_ISL_1635961, EPI_ISL_1635962, EPI_ISL_1635963, EPI_ISL_1635965, EPI_ISL_1635966, EPI_ISL_1635967, EPI_ISL_1635968, EPI_ISL_1635969, EPI_ISL_1635970, EPI_ISL_1635971, EPI_ISL_1635972,
EPI_ISL_1635973, EPI_ISL_1635974, EPI_ISL_1635975, EPI_ISL_1635976, EPI_ISL_1635977, EPI_ISL_1635978, EPI_ISL_1635979, EPI_ISL_1635980, EPI_ISL_1635981, EPI_ISL_1635982, EPI_ISL_1635983, EPI_ISL_1635984, EPI_ISL_1635985, EPI_ISL_1635986, EPI_ISL_1635987, EPI_ISL_1635988, EPI_ISL_1635989,
EPI_ISL_1635990, EPI_ISL_1635991, EPI_ISL_1635992, EPI_ISL_1635993, EPI_ISL_1635994, EPI_ISL_1635995, EPI_ISL_1635996, EPI_ISL_1635997, EPI_ISL_1635998, EPI_ISL_1635999, EPI_ISL_1636000, EPI_ISL_1636001, EPI_ISL_1636002, EPI_ISL_1636003, EPI_ISL_1636004, EPI_ISL_1636005, EPI_ISL_1636006,
EPI_ISL_1636007, EPI_ISL_1636008, EPI_ISL_1636009, EPI_ISL_1636010, EPI_ISL_1636011, EPI_ISL_1636012, EPI_ISL_1636013, EPI_ISL_1636014, EPI_ISL_1636015, EPI_ISL_1636017, EPI_ISL_1636018, EPI_ISL_1636019, EPI_ISL_1636020, EPI_ISL_1636021, EPI_ISL_1636022, EPI_ISL_1636023,
EPI_ISL_1636024, EPI_ISL_1636025, EPI_ISL_1636026, EPI_ISL_1636027, EPI_ISL_1636028, EPI_ISL_1636029, EPI_ISL_1636030, EPI_ISL_1636031, EPI_ISL_1636032, EPI_ISL_1636033, EPI_ISL_1636034, EPI_ISL_1636035, EPI_ISL_1636036, EPI_ISL_1636037, EPI_ISL_1636038, EPI_ISL_1636039, EPI_ISL_1636040,
EPI_ISL_1636041, EPI_ISL_1636042, EPI_ISL_1636043, EPI_ISL_1636044, EPI_ISL_1636045, EPI_ISL_1636046, EPI_ISL_1636047, EPI_ISL_1636048, EPI_ISL_1636049, EPI_ISL_1636050, EPI_ISL_1636051, EPI_ISL_1636052, EPI_ISL_1636053, EPI_ISL_1636054, EPI_ISL_1636055, EPI_ISL_1636056, EPI_ISL_1636057,
EPI_ISL_1636058, EPI_ISL_1636059, EPI_ISL_1636060, EPI_ISL_1636061, EPI_ISL_1636062, EPI_ISL_1636063, EPI_ISL_1636064, EPI_ISL_1636065, EPI_ISL_1636066, EPI_ISL_1636067, EPI_ISL_1636068, EPI_ISL_1636069, EPI_ISL_1636070, EPI_ISL_1636071, EPI_ISL_1636072, EPI_ISL_1636073, EPI_ISL_1636074,
EPI_ISL_1636075, EPI_ISL_1636076, EPI_ISL_1636077, EPI_ISL_1636078, EPI_ISL_1636079, EPI_ISL_1636080, EPI_ISL_1636081, EPI_ISL_1636082, EPI_ISL_1636083, EPI_ISL_1636084, EPI_ISL_1636085, EPI_ISL_1636086, EPI_ISL_1636087, EPI_ISL_1636088, EPI_ISL_1636089, EPI_ISL_1636090, EPI_ISL_1636091,
EPI_ISL_1636092, EPI_ISL_1636093, EPI_ISL_1636094, EPI_ISL_1636095, EPI_ISL_1636096, EPI_ISL_1636097, EPI_ISL_1636098, EPI_ISL_1636099, EPI_ISL_1636100, EPI_ISL_1636101, EPI_ISL_1636102, EPI_ISL_1636103, EPI_ISL_1636104, EPI_ISL_1636105, EPI_ISL_1636106, EPI_ISL_1636107, EPI_ISL_1636108,
EPI_ISL_1636109, EPI_ISL_1636111, EPI_ISL_1636112, EPI_ISL_1636113, EPI_ISL_1636114, EPI_ISL_1636115, EPI_ISL_1636116, EPI_ISL_1636117, EPI_ISL_1636118, EPI_ISL_1636119, EPI_ISL_1636120, EPI_ISL_1636121, EPI_ISL_1636122, EPI_ISL_1636123, EPI_ISL_1636124, EPI_ISL_1636125, EPI_ISL_1636126,
EPI_ISL_1636127, EPI_ISL_1636128, EPI_ISL_1636129, EPI_ISL_1636130, EPI_ISL_1636131, EPI_ISL_1636132, EPI_ISL_1636133, EPI_ISL_1636134, EPI_ISL_1636135, EPI_ISL_1636136, EPI_ISL_1636137, EPI_ISL_1636138, EPI_ISL_1636139, EPI_ISL_1636140, EPI_ISL_1636141, EPI_ISL_1636142, EPI_ISL_1636143,
EPI_ISL_1636144, EPI_ISL_1636145, EPI_ISL_1636146, EPI_ISL_1636147, EPI_ISL_1636148, EPI_ISL_1636149, EPI_ISL_1636150, EPI_ISL_1636151, EPI_ISL_1636152, EPI_ISL_1636153, EPI_ISL_1636154, EPI_ISL_1636155, EPI_ISL_1636156, EPI_ISL_1636157, EPI_ISL_1636158, EPI_ISL_1636159, EPI_ISL_1636160,
EPI_ISL_1636161, EPI_ISL_1636162, EPI_ISL_1636163, EPI_ISL_1636164, EPI_ISL_1636165, EPI_ISL_1636166, EPI_ISL_1636167, EPI_ISL_1636168, EPI_ISL_1636169, EPI_ISL_1636170, EPI_ISL_1636171, EPI_ISL_1636172, EPI_ISL_1636173, EPI_ISL_1636174, EPI_ISL_1636175, EPI_ISL_1636176, EPI_ISL_1636177,
EPI_ISL_1636178, EPI_ISL_1636179, EPI_ISL_1636180, EPI_ISL_1636181, EPI_ISL_1636182, EPI_ISL_1636183, EPI_ISL_1636184, EPI_ISL_1636185, EPI_ISL_1636186, EPI_ISL_1636187, EPI_ISL_1636188, EPI_ISL_1636189, EPI_ISL_1636190, EPI_ISL_1636191, EPI_ISL_1636192, EPI_ISL_1636193, EPI_ISL_1636194,
EPI_ISL_1636195, EPI_ISL_1636196, EPI_ISL_1636197, EPI_ISL_1636198, EPI_ISL_1636199, EPI_ISL_1636200, EPI_ISL_1636201, EPI_ISL_1636202, EPI_ISL_1636203, EPI_ISL_1636204, EPI_ISL_1636205, EPI_ISL_1636206, EPI_ISL_1636207, EPI_ISL_1636208, EPI_ISL_1636209, EPI_ISL_1636210, EPI_ISL_1636211,
EPI_ISL_1636212, EPI_ISL_1636213, EPI_ISL_1636214, EPI_ISL_1636215, EPI_ISL_1636217, EPI_ISL_1636218, EPI_ISL_1636219, EPI_ISL_1636220, EPI_ISL_1636221, EPI_ISL_1636222, EPI_ISL_1636223, EPI_ISL_1636224, EPI_ISL_1636225, EPI_ISL_1636226, EPI_ISL_1636227, EPI_ISL_1636228, EPI_ISL_1636229,
EPI_ISL_1636230, EPI_ISL_1636231, EPI_ISL_1636232, EPI_ISL_1636233, EPI_ISL_1636234, EPI_ISL_1636235, EPI_ISL_1636236, EPI_ISL_1636237, EPI_ISL_1636238, EPI_ISL_1636239, EPI_ISL_1636240, EPI_ISL_1636241, EPI_ISL_1636242, EPI_ISL_1636243, EPI_ISL_1636244, EPI_ISL_1636245, EPI_ISL_1636246,
EPI_ISL_1636247, EPI_ISL_1636248, EPI_ISL_1636249, EPI_ISL_1636250, EPI_ISL_1636251, EPI_ISL_1636252, EPI_ISL_1636253, EPI_ISL_1636254, EPI_ISL_1636255, EPI_ISL_1636256, EPI_ISL_1636257, EPI_ISL_1636258, EPI_ISL_1636259, EPI_ISL_1636260, EPI_ISL_1636261, EPI_ISL_1636262, EPI_ISL_1636263,
EPI_ISL_1636264, EPI_ISL_1636265, EPI_ISL_1636266, EPI_ISL_1636267, EPI_ISL_1636268, EPI_ISL_1636269, EPI_ISL_1636270, EPI_ISL_1636271, EPI_ISL_1636272, EPI_ISL_1636273, EPI_ISL_1636274, EPI_ISL_1636275, EPI_ISL_1636276, EPI_ISL_1636277, EPI_ISL_1636278, EPI_ISL_1636279, EPI_ISL_1636280,
EPI_ISL_1636281, EPI_ISL_1636282, EPI_ISL_1636283, EPI_ISL_1636284, EPI_ISL_1636285, EPI_ISL_1636286, EPI_ISL_1636287, EPI_ISL_1636288, EPI_ISL_1636289, EPI_ISL_1636290, EPI_ISL_1636291, EPI_ISL_1636292, EPI_ISL_1636293, EPI_ISL_1636294, EPI_ISL_1636295, EPI_ISL_1636296, EPI_ISL_1636297,
EPI_ISL_1636298, EPI_ISL_1636299, EPI_ISL_1636300, EPI_ISL_1636301, EPI_ISL_1636302, EPI_ISL_1636303, EPI_ISL_1636304, EPI_ISL_1636305, EPI_ISL_1636306, EPI_ISL_1636307, EPI_ISL_1636308, EPI_ISL_1636309, EPI_ISL_1636310, EPI_ISL_1636311, EPI_ISL_1636312, EPI_ISL_1636313, EPI_ISL_1636314,

| | | | |
|---|---|---|---|
| EPI_ISL_1636315, EPI_ISL_1636316, EPI_ISL_1636317, EPI_ISL_1636318, EPI_ISL_1636319, EPI_ISL_1636320, EPI_ISL_1636321, EPI_ISL_1636322, EPI_ISL_1636323, EPI_ISL_1636324, EPI_ISL_1636325, EPI_ISL_1636326, EPI_ISL_1636327, EPI_ISL_1636328, EPI_ISL_1636329, EPI_ISL_1636330, EPI_ISL_1636331, EPI_ISL_1636332, EPI_ISL_1636333, EPI_ISL_1636334, EPI_ISL_1636335, EPI_ISL_1636336, EPI_ISL_1636337, EPI_ISL_1636338, EPI_ISL_1636339, EPI_ISL_1636340, EPI_ISL_1636341, EPI_ISL_1636342, EPI_ISL_1636343, EPI_ISL_1636344, EPI_ISL_1636345, EPI_ISL_1636346, EPI_ISL_1636347, EPI_ISL_1636348, EPI_ISL_1636349, EPI_ISL_1636350, EPI_ISL_1636351, EPI_ISL_1636352, EPI_ISL_1636353, EPI_ISL_1636354, EPI_ISL_1636355, EPI_ISL_1636356, EPI_ISL_1636357, EPI_ISL_1636358, EPI_ISL_1636359, EPI_ISL_1636360, EPI_ISL_1636361, EPI_ISL_1636362, EPI_ISL_1636363, EPI_ISL_1636364, EPI_ISL_1636366, EPI_ISL_1636367, EPI_ISL_1636368, EPI_ISL_1636369, EPI_ISL_1636370, EPI_ISL_1636371, EPI_ISL_1636372, EPI_ISL_1636373, EPI_ISL_1636374, EPI_ISL_1636375, EPI_ISL_1636376, EPI_ISL_1636377, EPI_ISL_1636378, EPI_ISL_1636379, EPI_ISL_1636380, EPI_ISL_1636381, EPI_ISL_1636382, EPI_ISL_1636383, EPI_ISL_1636384, EPI_ISL_1636385, EPI_ISL_1636386, EPI_ISL_1636387, EPI_ISL_1636388, EPI_ISL_1636389, EPI_ISL_1636390, EPI_ISL_1636391, EPI_ISL_1636392, EPI_ISL_1636393, EPI_ISL_1636394, EPI_ISL_1636395, EPI_ISL_1636396, EPI_ISL_1636397 | | | |
| see above | Pandemic Response Lab - NYC | Pandemic Response Lab, R&D | Henry Lee, Michael Hammerling, Melissa Hopkins, Cybill del Castillo, Shinyoung Clair Kang, William Ward, Pradeep Bugga, Sol Rey, Dylan Law, Katharine Nelson, Haiping Hao, Jon Laurent |
| EPI_ISL_1648211, EPI_ISL_1648236, EPI_ISL_1648303, EPI_ISL_1648304, EPI_ISL_1648314, EPI_ISL_1648317, EPI_ISL_1648320, EPI_ISL_1648327, EPI_ISL_1648341, EPI_ISL_1648358, EPI_ISL_1648362, EPI_ISL_1648365, EPI_ISL_1648384, EPI_ISL_1648386, EPI_ISL_1648392, EPI_ISL_1648397, EPI_ISL_1648401, EPI_ISL_1648402, EPI_ISL_1648417, EPI_ISL_1648437, EPI_ISL_1648461, EPI_ISL_1648499, EPI_ISL_1648538, EPI_ISL_1648558, EPI_ISL_1648561, EPI_ISL_1648586, EPI_ISL_1648757, EPI_ISL_1648778, EPI_ISL_1648780, EPI_ISL_1648784, EPI_ISL_1648829, EPI_ISL_1648842, EPI_ISL_1648885, EPI_ISL_1648916 | | | |
| see above | Quest Diagnostics Incorporated | Centers for Disease Control and Prevention Division of Viral Diseases, Pathogen Discovery | Dakota Howard, Dhwani Batra, Peter W. Cook, Kara Moser, Adrian Paskey, Jason Caravas, Benjamin Rambo-Martin, Shatavia Morrison, Christopher Gulvick, Scott Sammons, Yvette Unoarumhi, Darlene Wagner, Matthew Schmerer, S. H. Rosenthal, A. Gerasimova, R. M. Kagan, B. Anderson, M. Hua, Y. Liu, L.E. Bernstein, K.E. Livingston, A. Perez, I. A. Shlyakhter, R. V. Rolando, R. Owen, P. Tanpaiboon, F. Lacbawan, Clinton R. Paden, Duncan MacCannell |
| EPI_ISL_1649006, EPI_ISL_1649008, EPI_ISL_1649024, EPI_ISL_1649040, EPI_ISL_1649113, EPI_ISL_1649116, EPI_ISL_1649154, EPI_ISL_1649155, EPI_ISL_1649185, EPI_ISL_1649209, EPI_ISL_1649223, EPI_ISL_1649228, EPI_ISL_1649230, EPI_ISL_1649234, EPI_ISL_1649235, EPI_ISL_1649236, EPI_ISL_1649266, EPI_ISL_1649277, EPI_ISL_1649278, EPI_ISL_1649293, EPI_ISL_1649295, EPI_ISL_1649385, EPI_ISL_1649392, EPI_ISL_1649393, EPI_ISL_1649394, EPI_ISL_1649459, EPI_ISL_1649462, EPI_ISL_1649463, EPI_ISL_1649482, EPI_ISL_1649483, EPI_ISL_1649523, EPI_ISL_1649589, EPI_ISL_1649646, EPI_ISL_1649685, EPI_ISL_1649687, EPI_ISL_1649729, EPI_ISL_1649753, EPI_ISL_1649759, EPI_ISL_1649763, EPI_ISL_1649820, EPI_ISL_1649840, EPI_ISL_1649845, EPI_ISL_1650018, EPI_ISL_1650049, EPI_ISL_1650053, EPI_ISL_1650054, EPI_ISL_1650055, EPI_ISL_1650056, EPI_ISL_1650061, EPI_ISL_1650132, EPI_ISL_1650143, EPI_ISL_1650144, EPI_ISL_1650158, EPI_ISL_1650159, EPI_ISL_1650233, EPI_ISL_1650237, EPI_ISL_1650240, EPI_ISL_1650241, EPI_ISL_1650242, EPI_ISL_1650243, EPI_ISL_1650259, EPI_ISL_1650287, EPI_ISL_1650293, EPI_ISL_1650313, EPI_ISL_1650326, EPI_ISL_1650352, EPI_ISL_1650383, EPI_ISL_1650384, EPI_ISL_1650411, EPI_ISL_1650436, EPI_ISL_1650555, EPI_ISL_1650657, EPI_ISL_1650664, EPI_ISL_1650678, EPI_ISL_1650686, EPI_ISL_1650687, EPI_ISL_1650711, EPI_ISL_1650839, EPI_ISL_1650997, EPI_ISL_1651147, EPI_ISL_1651148 | | | |
| see above | Aegis Sciences Corporation | Centers for Disease Control and Prevention Division of Viral Diseases, Pathogen Discovery | Dakota Howard, Dhwani Batra, Peter W. Cook, Kara Moser, Adrian Paskey, Jason Caravas, Benjamin Rambo-Martin, Shatavia Morrison, Christopher Gulvick, Scott Sammons, Yvette Unoarumhi, Darlene Wagner, Matthew Schmerer, Cyndi Clark, Patrick Campbell, Rob Case, Vikramsinha Ghorpade, Holly Houdeshell, Ola Kvalvaag, Dillon Nall, Ethan Sanders, Alec Vest, Shaun Westlund, Matthew Hardison, Clinton R. Paden, Duncan MacCannell |
| EPI_ISL_1651216, EPI_ISL_1651217 | MEMORIAL SLOAN KETTERING CANCER CENTER | Wadsworth Center, New York State Department of Health | Kirsten St. George, Daryl M. Lamson, Alexis Russell, Matthew Shudt, Melissa A Leisner, Jonathan Plitnick, Catharine Prussing, Navjot Singh, John Kelly, Erasmus Schneider, Erica Lasek-Nesselquist |
| EPI_ISL_1651218 | MONTEFIORE MEDICAL CENTER LABORATORIES | Wadsworth Center, New York State Department of Health | Kirsten St. George, Daryl M. Lamson, Alexis Russell, Matthew Shudt, Melissa A Leisner, Jonathan Plitnick, Catharine Prussing, Navjot Singh, John Kelly, Erasmus Schneider, Erica Lasek-Nesselquist |
| EPI_ISL_1651219, EPI_ISL_1651220, EPI_ISL_1651221 | New York Presbyterian Hospital | Wadsworth Center, New York State Department of Health | Kirsten St. George, Daryl M. Lamson, Alexis Russell, Matthew Shudt, Melissa A Leisner, Jonathan Plitnick, Catharine Prussing, Navjot Singh, John Kelly, Erasmus Schneider, Erica Lasek-Nesselquist |
| EPI_ISL_1651222, EPI_ISL_1651223 | URMC LABS | Wadsworth Center, New York State Department of Health | Kirsten St. George, Daryl M. Lamson, Alexis Russell, Matthew Shudt, Melissa A Leisner, Jonathan Plitnick, Catharine Prussing, Navjot Singh, John Kelly, Erasmus Schneider, Erica Lasek-Nesselquist |
| EPI_ISL_1651224 | SARATOGA HOSPITAL LABORATORY | Wadsworth Center, New York State Department of Health | Kirsten St. George, Daryl M. Lamson, Alexis Russell, Matthew Shudt, Melissa A Leisner, Jonathan Plitnick, Catharine Prussing, Navjot Singh, John Kelly, Erasmus Schneider, Erica Lasek-Nesselquist |
| EPI_ISL_1651225 | HOSPITAL FOR SPECIAL SURGERY | Wadsworth Center, New York State Department of Health | Kirsten St. George, Daryl M. Lamson, Alexis Russell, Matthew Shudt, Melissa A Leisner, Jonathan Plitnick, Catharine Prussing, Navjot Singh, John Kelly, Erasmus Schneider, Erica Lasek-Nesselquist |
| EPI_ISL_1651226 | MEMORIAL SLOAN KETTERING CANCER CENTER | Wadsworth Center, New York State Department of Health | Kirsten St. George, Daryl M. Lamson, Alexis Russell, Matthew Shudt, Melissa A Leisner, Jonathan Plitnick, Catharine Prussing, Navjot Singh, John Kelly, Erasmus Schneider, Erica Lasek-Nesselquist |
| EPI_ISL_1651227, EPI_ISL_1651228 | URMC LABS | Wadsworth Center, New York State Department of Health | Kirsten St. George, Daryl M. Lamson, Alexis Russell, Matthew Shudt, Melissa A Leisner, Jonathan Plitnick, Catharine Prussing, Navjot Singh, John Kelly, Erasmus Schneider, Erica Lasek-Nesselquist |
| EPI_ISL_1654574, EPI_ISL_1654575, EPI_ISL_1654576, EPI_ISL_1654577, EPI_ISL_1654578, EPI_ISL_1654579, EPI_ISL_1654580, EPI_ISL_1654581, EPI_ISL_1654582, EPI_ISL_1654583, EPI_ISL_1654584, EPI_ISL_1654585, EPI_ISL_1654587, EPI_ISL_1654588, EPI_ISL_1654589, EPI_ISL_1654590, EPI_ISL_1654591, EPI_ISL_1654592, EPI_ISL_1654593, EPI_ISL_1654594, EPI_ISL_1654595, EPI_ISL_1654596, EPI_ISL_1654597, EPI_ISL_1654599, EPI_ISL_1654600, EPI_ISL_1654601, EPI_ISL_1654602, EPI_ISL_1654603, EPI_ISL_1654604, EPI_ISL_1654605, EPI_ISL_1654606, EPI_ISL_1654607, EPI_ISL_1654608, EPI_ISL_1654609, EPI_ISL_1654610, EPI_ISL_1654611, EPI_ISL_1654613, EPI_ISL_1654614, EPI_ISL_1654615, EPI_ISL_1654616, EPI_ISL_1654617, EPI_ISL_1654618, EPI_ISL_1654619, EPI_ISL_1654620, EPI_ISL_1654621, EPI_ISL_1654622, EPI_ISL_1654623, EPI_ISL_1654624, EPI_ISL_1654625, EPI_ISL_1654626, EPI_ISL_1654627, EPI_ISL_1654628, EPI_ISL_1654629, EPI_ISL_1654630, EPI_ISL_1654631, EPI_ISL_1654632, EPI_ISL_1654633, EPI_ISL_1654634, EPI_ISL_1654635, EPI_ISL_1654636, EPI_ISL_1654637, EPI_ISL_1654638, EPI_ISL_1654639, EPI_ISL_1654640, EPI_ISL_1654641, EPI_ISL_1654644, EPI_ISL_1654645, EPI_ISL_1654646, EPI_ISL_1654647 | | | |
| see above | NYU Langone Health | Departments of Pathology and Medicine, New York University School of Medicine | Adriana Heguy, Dacia Dimartino, Emily Guzman, Christian Marier, Peter Meyn, Sitharam Ramaswami, Gael Westby, Paul Zappile, Yutong Zhang, Paolo Cotzia, Guiqing Wang |
| EPI_ISL_1667463, EPI_ISL_1667465, EPI_ISL_1667473, EPI_ISL_1667476, EPI_ISL_1667543, EPI_ISL_1667544, EPI_ISL_1667549, EPI_ISL_1667569, EPI_ISL_1667572, EPI_ISL_1667618, EPI_ISL_1667660, EPI_ISL_1667704, EPI_ISL_1667740, EPI_ISL_1667744, EPI_ISL_1667746, EPI_ISL_1667747, EPI_ISL_1667754, EPI_ISL_1667773, EPI_ISL_1667838, EPI_ISL_1667839, EPI_ISL_1667840, EPI_ISL_1667951, EPI_ISL_1668015, EPI_ISL_1668022, EPI_ISL_1668025, EPI_ISL_1668038, EPI_ISL_1668130, EPI_ISL_1668180, EPI_ISL_1668181, EPI_ISL_1668185, EPI_ISL_1668190, EPI_ISL_1668214, EPI_ISL_1668304, EPI_ISL_1668328, EPI_ISL_1668354, EPI_ISL_1668364, EPI_ISL_1668373, EPI_ISL_1668374, EPI_ISL_1668375 | | | |
| see above | Aegis Sciences Corporation | Centers for Disease Control and Prevention Division of Viral Diseases, Pathogen Discovery | Dakota Howard, Dhwani Batra, Peter W. Cook, Kara Moser, Adrian Paskey, Jason Caravas, Benjamin Rambo-Martin, Shatavia Morrison, Christopher Gulvick, Scott Sammons, Yvette Unoarumhi, Darlene Wagner, Matthew Schmerer, Cyndi Clark, Patrick Campbell, Rob Case, Vikramsinha Ghorpade, Holly Houdeshell, Ola Kvalvaag, Dillon Nall, Ethan Sanders, Alec Vest, Shaun Westlund, Matthew Hardison, Clinton R. Paden, Duncan MacCannell |
| EPI_ISL_1668648, EPI_ISL_1668707, EPI_ISL_1668768, EPI_ISL_1669089 | Quest Diagnostics Incorporated | Centers for Disease Control and Prevention Division of Viral Diseases, Pathogen Discovery | Dakota Howard, Dhwani Batra, Peter W. Cook, Kara Moser, Adrian Paskey, Jason Caravas, Benjamin Rambo-Martin, Shatavia Morrison, Christopher Gulvick, Scott Sammons, Yvette Unoarumhi, Darlene Wagner, Matthew Schmerer, S. H. Rosenthal, A. Gerasimova, R. M. Kagan, B. Anderson, M. Hua, Y. Liu, L.E. Bernstein, K.E. Livingston, A. Perez, I. A. Shlyakhter, R. V. Rolando, R. Owen, P. Tanpaiboon, F. Lacbawan, Clinton R. Paden, Duncan MacCannell |
| EPI_ISL_1669801, EPI_ISL_1669802, EPI_ISL_1669803, EPI_ISL_1669821 | NYU Langone Health | Departments of Pathology and Medicine, New York University School of Medicine | Adriana Heguy, Dacia Dimartino, Emily Guzman, Christian Marier, Peter Meyn, Sitharam Ramaswami, Gael Westby, Paul Zappile, Yutong Zhang, Paolo Cotzia, Guiqing Wang |
| EPI_ISL_1671137 | MEMORIAL SLOAN KETTERING CANCER CENTER | Wadsworth Center, New York State Department of Health | Kirsten St. George, Daryl M. Lamson, Alexis Russell, Matthew Shudt, Melissa A Leisner, Jonathan Plitnick, Catharine Prussing, Navjot Singh, John Kelly, Erasmus Schneider, Erica Lasek-Nesselquist |
| EPI_ISL_1671138, EPI_ISL_1671139, EPI_ISL_1671140 | New York Presbyterian Hospital | Wadsworth Center, New York State Department of Health | Kirsten St. George, Daryl M. Lamson, Alexis Russell, Matthew Shudt, Melissa A Leisner, Jonathan Plitnick, Catharine Prussing, Navjot Singh, John Kelly, Erasmus Schneider, Erica Lasek-Nesselquist |
| EPI_ISL_1671141, EPI_ISL_1671142 | NYC Pandemic Response Lab | Wadsworth Center, New York State Department of Health | Kirsten St. George, Daryl M. Lamson, Alexis Russell, Matthew Shudt, Melissa A Leisner, Jonathan Plitnick, Catharine Prussing, Navjot Singh, John Kelly, Erasmus Schneider, Erica Lasek-Nesselquist |
| EPI_ISL_1671143, EPI_ISL_1671144 | WESTCHESTER MEDICAL CENTER | Wadsworth Center, New York State Department of Health | Kirsten St. George, Daryl M. Lamson, Alexis Russell, Matthew Shudt, Melissa A Leisner, Jonathan Plitnick, Catharine Prussing, Navjot Singh, John Kelly, Erasmus Schneider, Erica Lasek-Nesselquist |
| EPI_ISL_1671145 | MONTEFIORE MEDICAL CENTER LABORATORIES | Wadsworth Center, New York State Department of Health | Kirsten St. George, Daryl M. Lamson, Alexis Russell, Matthew Shudt, Melissa A Leisner, Jonathan Plitnick, Catharine Prussing, Navjot Singh, John Kelly, Erasmus Schneider, Erica Lasek-Nesselquist |
| EPI_ISL_1671146 | New York Presbyterian Hospital | Wadsworth Center, New York State Department of Health | Kirsten St. George, Daryl M. Lamson, Alexis Russell, Matthew Shudt, Melissa A Leisner, Jonathan Plitnick, Catharine Prussing, Navjot Singh, John Kelly, Erasmus Schneider, Erica Lasek-Nesselquist |
| EPI_ISL_1671147 | MEMORIAL SLOAN KETTERING CANCER CENTER | Wadsworth Center, New York State Department of Health | Kirsten St. George, Daryl M. Lamson, Alexis Russell, Matthew Shudt, Melissa A Leisner, Jonathan Plitnick, Catharine Prussing, Navjot Singh, John Kelly, Erasmus Schneider, Erica Lasek-Nesselquist |
| EPI_ISL_1671148, EPI_ISL_1671149, EPI_ISL_1671150, EPI_ISL_1671151, EPI_ISL_1671152, EPI_ISL_1671153, EPI_ISL_1671154, EPI_ISL_1671155 | NYC Pandemic Response Lab | Wadsworth Center, New York State Department of Health | Kirsten St. George, Daryl M. Lamson, Alexis Russell, Matthew Shudt, Melissa A Leisner, Jonathan Plitnick, Catharine Prussing, Navjot Singh, John Kelly, Erasmus Schneider, Erica Lasek-Nesselquist |

| | | | |
|---|---|---|---|
| EPI_ISL_1671156 | MONTEFIORE MEDICAL CENTER LABORATORIES | Wadsworth Center, New York State Department of Health | Kirsten St. George, Daryl M. Lamson, Alexis Russell, Matthew Shudt, Melissa A Leisner, Jonathan Plitnick, Catharine Prussing, Navjot Singh, John Kelly, Erasmus Schneider, Erica Lasek-Nesselquist |
| EPI_ISL_1671157 | NYC Pandemic Response Lab | Wadsworth Center, New York State Department of Health | Kirsten St. George, Daryl M. Lamson, Alexis Russell, Matthew Shudt, Melissa A Leisner, Jonathan Plitnick, Catharine Prussing, Navjot Singh, John Kelly, Erasmus Schneider, Erica Lasek-Nesselquist |
| EPI_ISL_1671158, EPI_ISL_1671159 | MONTEFIORE MEDICAL CENTER LABORATORIES | Wadsworth Center, New York State Department of Health | Kirsten St. George, Daryl M. Lamson, Alexis Russell, Matthew Shudt, Melissa A Leisner, Jonathan Plitnick, Catharine Prussing, Navjot Singh, John Kelly, Erasmus Schneider, Erica Lasek-Nesselquist |
| EPI_ISL_1671160 | Columbia University Irving Medical Center | Wadsworth Center, New York State Department of Health | Kirsten St. George, Daryl M. Lamson, Alexis Russell, Matthew Shudt, Melissa A Leisner, Jonathan Plitnick, Catharine Prussing, Navjot Singh, John Kelly, Erasmus Schneider, Erica Lasek-Nesselquist |
| EPI_ISL_1671161 | WESTCHESTER MEDICAL CENTER | Wadsworth Center, New York State Department of Health | Kirsten St. George, Daryl M. Lamson, Alexis Russell, Matthew Shudt, Melissa A Leisner, Jonathan Plitnick, Catharine Prussing, Navjot Singh, John Kelly, Erasmus Schneider, Erica Lasek-Nesselquist |
| EPI_ISL_1671162 | Columbia University Irving Medical Center | Wadsworth Center, New York State Department of Health | Kirsten St. George, Daryl M. Lamson, Alexis Russell, Matthew Shudt, Melissa A Leisner, Jonathan Plitnick, Catharine Prussing, Navjot Singh, John Kelly, Erasmus Schneider, Erica Lasek-Nesselquist |
| EPI_ISL_1671163, EPI_ISL_1671164 | MONTEFIORE MEDICAL CENTER LABORATORIES | Wadsworth Center, New York State Department of Health | Kirsten St. George, Daryl M. Lamson, Alexis Russell, Matthew Shudt, Melissa A Leisner, Jonathan Plitnick, Catharine Prussing, Navjot Singh, John Kelly, Erasmus Schneider, Erica Lasek-Nesselquist |
| EPI_ISL_1671165 | MEMORIAL SLOAN KETTERING CANCER CENTER | Wadsworth Center, New York State Department of Health | Kirsten St. George, Daryl M. Lamson, Alexis Russell, Matthew Shudt, Melissa A Leisner, Jonathan Plitnick, Catharine Prussing, Navjot Singh, John Kelly, Erasmus Schneider, Erica Lasek-Nesselquist |
| EPI_ISL_1671166 | New York Presbyterian Hospital | Wadsworth Center, New York State Department of Health | Kirsten St. George, Daryl M. Lamson, Alexis Russell, Matthew Shudt, Melissa A Leisner, Jonathan Plitnick, Catharine Prussing, Navjot Singh, John Kelly, Erasmus Schneider, Erica Lasek-Nesselquist |
| EPI_ISL_1671167, EPI_ISL_1671168 | NYC Pandemic Response Lab | Wadsworth Center, New York State Department of Health | Kirsten St. George, Daryl M. Lamson, Alexis Russell, Matthew Shudt, Melissa A Leisner, Jonathan Plitnick, Catharine Prussing, Navjot Singh, John Kelly, Erasmus Schneider, Erica Lasek-Nesselquist |
| EPI_ISL_1671169, EPI_ISL_1671170, EPI_ISL_1671171 | MONTEFIORE MEDICAL CENTER LABORATORIES | Wadsworth Center, New York State Department of Health | Kirsten St. George, Daryl M. Lamson, Alexis Russell, Matthew Shudt, Melissa A Leisner, Jonathan Plitnick, Catharine Prussing, Navjot Singh, John Kelly, Erasmus Schneider, Erica Lasek-Nesselquist |
| EPI_ISL_1671172, EPI_ISL_1671173 | NYC Pandemic Response Lab | Wadsworth Center, New York State Department of Health | Kirsten St. George, Daryl M. Lamson, Alexis Russell, Matthew Shudt, Melissa A Leisner, Jonathan Plitnick, Catharine Prussing, Navjot Singh, John Kelly, Erasmus Schneider, Erica Lasek-Nesselquist |
| EPI_ISL_1671174, EPI_ISL_1671175 | MONTEFIORE MEDICAL CENTER LABORATORIES | Wadsworth Center, New York State Department of Health | Kirsten St. George, Daryl M. Lamson, Alexis Russell, Matthew Shudt, Melissa A Leisner, Jonathan Plitnick, Catharine Prussing, Navjot Singh, John Kelly, Erasmus Schneider, Erica Lasek-Nesselquist |
| EPI_ISL_1671176 | Columbia University Irving Medical Center | Wadsworth Center, New York State Department of Health | Kirsten St. George, Daryl M. Lamson, Alexis Russell, Matthew Shudt, Melissa A Leisner, Jonathan Plitnick, Catharine Prussing, Navjot Singh, John Kelly, Erasmus Schneider, Erica Lasek-Nesselquist |
| EPI_ISL_1671177, EPI_ISL_1671178, EPI_ISL_1671179, EPI_ISL_1671180 | NYC Pandemic Response Lab | Wadsworth Center, New York State Department of Health | Kirsten St. George, Daryl M. Lamson, Alexis Russell, Matthew Shudt, Melissa A Leisner, Jonathan Plitnick, Catharine Prussing, Navjot Singh, John Kelly, Erasmus Schneider, Erica Lasek-Nesselquist |
| EPI_ISL_1671181, EPI_ISL_1671182 | MONTEFIORE MEDICAL CENTER LABORATORIES | Wadsworth Center, New York State Department of Health | Kirsten St. George, Daryl M. Lamson, Alexis Russell, Matthew Shudt, Melissa A Leisner, Jonathan Plitnick, Catharine Prussing, Navjot Singh, John Kelly, Erasmus Schneider, Erica Lasek-Nesselquist |
| EPI_ISL_1671183, EPI_ISL_1671184 | NYC Pandemic Response Lab | Wadsworth Center, New York State Department of Health | Kirsten St. George, Daryl M. Lamson, Alexis Russell, Matthew Shudt, Melissa A Leisner, Jonathan Plitnick, Catharine Prussing, Navjot Singh, John Kelly, Erasmus Schneider, Erica Lasek-Nesselquist |
| EPI_ISL_1671185, EPI_ISL_1671186, EPI_ISL_1671187, EPI_ISL_1671188, EPI_ISL_1671189, EPI_ISL_1671190, EPI_ISL_1671191, EPI_ISL_1671192 | MONTEFIORE MEDICAL CENTER LABORATORIES | Wadsworth Center, New York State Department of Health | Kirsten St. George, Daryl M. Lamson, Alexis Russell, Matthew Shudt, Melissa A Leisner, Jonathan Plitnick, Catharine Prussing, Navjot Singh, John Kelly, Erasmus Schneider, Erica Lasek-Nesselquist |
| EPI_ISL_1671193 | MEMORIAL SLOAN KETTERING CANCER CENTER | Wadsworth Center, New York State Department of Health | Kirsten St. George, Daryl M. Lamson, Alexis Russell, Matthew Shudt, Melissa A Leisner, Jonathan Plitnick, Catharine Prussing, Navjot Singh, John Kelly, Erasmus Schneider, Erica Lasek-Nesselquist |
| EPI_ISL_1671194 | New York Presbyterian Hospital | Wadsworth Center, New York State Department of Health | Kirsten St. George, Daryl M. Lamson, Alexis Russell, Matthew Shudt, Melissa A Leisner, Jonathan Plitnick, Catharine Prussing, Navjot Singh, John Kelly, Erasmus Schneider, Erica Lasek-Nesselquist |
| EPI_ISL_1671195, EPI_ISL_1671196, EPI_ISL_1671197, EPI_ISL_1671198, EPI_ISL_1671199, EPI_ISL_1671200 | NYC Pandemic Response Lab | Wadsworth Center, New York State Department of Health | Kirsten St. George, Daryl M. Lamson, Alexis Russell, Matthew Shudt, Melissa A Leisner, Jonathan Plitnick, Catharine Prussing, Navjot Singh, John Kelly, Erasmus Schneider, Erica Lasek-Nesselquist |
| EPI_ISL_1671201, EPI_ISL_1671202 | MEMORIAL SLOAN KETTERING CANCER CENTER | Wadsworth Center, New York State Department of Health | Kirsten St. George, Daryl M. Lamson, Alexis Russell, Matthew Shudt, Melissa A Leisner, Jonathan Plitnick, Catharine Prussing, Navjot Singh, John Kelly, Erasmus Schneider, Erica Lasek-Nesselquist |
| EPI_ISL_1671203, EPI_ISL_1671204 | MONTEFIORE MEDICAL CENTER LABORATORIES | Wadsworth Center, New York State Department of Health | Kirsten St. George, Daryl M. Lamson, Alexis Russell, Matthew Shudt, Melissa A Leisner, Jonathan Plitnick, Catharine Prussing, Navjot Singh, John Kelly, Erasmus Schneider, Erica Lasek-Nesselquist |
| EPI_ISL_1671205, EPI_ISL_1671206, EPI_ISL_1671207 | NYC Pandemic Response Lab | Wadsworth Center, New York State Department of Health | Kirsten St. George, Daryl M. Lamson, Alexis Russell, Matthew Shudt, Melissa A Leisner, Jonathan Plitnick, Catharine Prussing, Navjot Singh, John Kelly, Erasmus Schneider, Erica Lasek-Nesselquist |
| EPI_ISL_1671208 | MONTEFIORE MEDICAL CENTER LABORATORIES | Wadsworth Center, New York State Department of Health | Kirsten St. George, Daryl M. Lamson, Alexis Russell, Matthew Shudt, Melissa A Leisner, Jonathan Plitnick, Catharine Prussing, Navjot Singh, John Kelly, Erasmus Schneider, Erica Lasek-Nesselquist |
| EPI_ISL_1671209 | NORTHWELL HEALTH LABORATORIES | Wadsworth Center, New York State Department of Health | Kirsten St. George, Daryl M. Lamson, Alexis Russell, Matthew Shudt, Melissa A Leisner, Jonathan Plitnick, Catharine Prussing, Navjot Singh, John Kelly, Erasmus Schneider, Erica Lasek-Nesselquist |
| EPI_ISL_1671210, EPI_ISL_1671211, EPI_ISL_1671212 | Columbia University Irving Medical Center | Wadsworth Center, New York State Department of Health | Kirsten St. George, Daryl M. Lamson, Alexis Russell, Matthew Shudt, Melissa A Leisner, Jonathan Plitnick, Catharine Prussing, Navjot Singh, John Kelly, Erasmus Schneider, Erica Lasek-Nesselquist |
| EPI_ISL_1671213, EPI_ISL_1671214, EPI_ISL_1671215, EPI_ISL_1671216 | New York Presbyterian Hospital | Wadsworth Center, New York State Department of Health | Kirsten St. George, Daryl M. Lamson, Alexis Russell, Matthew Shudt, Melissa A Leisner, Jonathan Plitnick, Catharine Prussing, Navjot Singh, John Kelly, Erasmus Schneider, Erica Lasek-Nesselquist |
| EPI_ISL_1671217 | Columbia University Irving Medical Center | Wadsworth Center, New York State Department of Health | Kirsten St. George, Daryl M. Lamson, Alexis Russell, Matthew Shudt, Melissa A Leisner, Jonathan Plitnick, Catharine Prussing, Navjot Singh, John Kelly, Erasmus Schneider, Erica Lasek-Nesselquist |
| EPI_ISL_1671218, EPI_ISL_1671219, EPI_ISL_1671220 | WESTCHESTER MEDICAL CENTER | Wadsworth Center, New York State Department of Health | Kirsten St. George, Daryl M. Lamson, Alexis Russell, Matthew Shudt, Melissa A Leisner, Jonathan Plitnick, Catharine Prussing, Navjot Singh, John Kelly, Erasmus Schneider, Erica Lasek-Nesselquist |
| EPI_ISL_1671312 | TEMPUS LABS INC | Wadsworth Center, New York State Department of Health | Kirsten St. George, Daryl M. Lamson, Alexis Russell, Matthew Shudt, Melissa A Leisner, Jonathan Plitnick, Catharine Prussing, Navjot Singh, John Kelly, Erasmus Schneider, Erica Lasek-Nesselquist |
| EPI_ISL_1671313 | ALBANY MEDICAL CENTER | Wadsworth Center, New York State Department of Health | Kirsten St. George, Daryl M. Lamson, Alexis Russell, Matthew Shudt, Melissa A Leisner, Jonathan Plitnick, Catharine Prussing, Navjot Singh, John Kelly, Erasmus Schneider, Erica Lasek-Nesselquist |
| EPI_ISL_1671314 | TEMPUS LABS INC | Wadsworth Center, New York State Department of Health | Kirsten St. George, Daryl M. Lamson, Alexis Russell, Matthew Shudt, Melissa A Leisner, Jonathan Plitnick, Catharine Prussing, Navjot Singh, John Kelly, Erasmus Schneider, Erica Lasek-Nesselquist |
| EPI_ISL_1671315 | ALBANY MEDICAL CENTER | Wadsworth Center, New York State Department of Health | Kirsten St. George, Daryl M. Lamson, Alexis Russell, Matthew Shudt, Melissa A Leisner, Jonathan Plitnick, Catharine Prussing, Navjot Singh, John Kelly, Erasmus Schneider, Erica Lasek-Nesselquist |

| | | | |
|---|---|---|---|
| EPI_ISL_1671316 | STONY BROOK UNIVERSITY HOSPITAL | Wadsworth Center, New York State Department of Health | Kirsten St. George, Daryl M. Lamson, Alexis Russell, Matthew Shudt, Melissa A Leisner, Jonathan Plitnick, Catharine Prussing, Navjot Singh, John Kelly, Erasmus Schneider, Erica Lasek-Nesselquist |
| EPI_ISL_1671317, EPI_ISL_1671318, EPI_ISL_1671319, EPI_ISL_1671320, EPI_ISL_1671321 | ALBANY MEDICAL CENTER | Wadsworth Center, New York State Department of Health | Kirsten St. George, Daryl M. Lamson, Alexis Russell, Matthew Shudt, Melissa A Leisner, Jonathan Plitnick, Catharine Prussing, Navjot Singh, John Kelly, Erasmus Schneider, Erica Lasek-Nesselquist |
| EPI_ISL_1671322 | STONY BROOK UNIVERSITY HOSPITAL | Wadsworth Center, New York State Department of Health | Kirsten St. George, Daryl M. Lamson, Alexis Russell, Matthew Shudt, Melissa A Leisner, Jonathan Plitnick, Catharine Prussing, Navjot Singh, John Kelly, Erasmus Schneider, Erica Lasek-Nesselquist |
| EPI_ISL_1671323, EPI_ISL_1671324, EPI_ISL_1671325, EPI_ISL_1671326, EPI_ISL_1671327 | ALBANY MEDICAL CENTER | Wadsworth Center, New York State Department of Health | Kirsten St. George, Daryl M. Lamson, Alexis Russell, Matthew Shudt, Melissa A Leisner, Jonathan Plitnick, Catharine Prussing, Navjot Singh, John Kelly, Erasmus Schneider, Erica Lasek-Nesselquist |
| EPI_ISL_1671328, EPI_ISL_1671329, EPI_ISL_1671330 | STONY BROOK UNIVERSITY HOSPITAL | Wadsworth Center, New York State Department of Health | Kirsten St. George, Daryl M. Lamson, Alexis Russell, Matthew Shudt, Melissa A Leisner, Jonathan Plitnick, Catharine Prussing, Navjot Singh, John Kelly, Erasmus Schneider, Erica Lasek-Nesselquist |
| EPI_ISL_1671331, EPI_ISL_1671332, EPI_ISL_1671333, EPI_ISL_1671334, EPI_ISL_1671335 | TEMPUS LABS INC | Wadsworth Center, New York State Department of Health | Kirsten St. George, Daryl M. Lamson, Alexis Russell, Matthew Shudt, Melissa A Leisner, Jonathan Plitnick, Catharine Prussing, Navjot Singh, John Kelly, Erasmus Schneider, Erica Lasek-Nesselquist |
| EPI_ISL_1671336 | STONY BROOK UNIVERSITY HOSPITAL | Wadsworth Center, New York State Department of Health | Kirsten St. George, Daryl M. Lamson, Alexis Russell, Matthew Shudt, Melissa A Leisner, Jonathan Plitnick, Catharine Prussing, Navjot Singh, John Kelly, Erasmus Schneider, Erica Lasek-Nesselquist |
| EPI_ISL_1671337 | ALBANY MEDICAL CENTER | Wadsworth Center, New York State Department of Health | Kirsten St. George, Daryl M. Lamson, Alexis Russell, Matthew Shudt, Melissa A Leisner, Jonathan Plitnick, Catharine Prussing, Navjot Singh, John Kelly, Erasmus Schneider, Erica Lasek-Nesselquist |
| EPI_ISL_1671338, EPI_ISL_1671339, EPI_ISL_1671340 | TEMPUS LABS INC | Wadsworth Center, New York State Department of Health | Kirsten St. George, Daryl M. Lamson, Alexis Russell, Matthew Shudt, Melissa A Leisner, Jonathan Plitnick, Catharine Prussing, Navjot Singh, John Kelly, Erasmus Schneider, Erica Lasek-Nesselquist |
| EPI_ISL_1671341 | ALBANY MEDICAL CENTER | Wadsworth Center, New York State Department of Health | Kirsten St. George, Daryl M. Lamson, Alexis Russell, Matthew Shudt, Melissa A Leisner, Jonathan Plitnick, Catharine Prussing, Navjot Singh, John Kelly, Erasmus Schneider, Erica Lasek-Nesselquist |
| EPI_ISL_1671342, EPI_ISL_1671343, EPI_ISL_1671344, EPI_ISL_1671345, EPI_ISL_1671346, EPI_ISL_1671347, EPI_ISL_1671348 | TEMPUS LABS INC | Wadsworth Center, New York State Department of Health | Kirsten St. George, Daryl M. Lamson, Alexis Russell, Matthew Shudt, Melissa A Leisner, Jonathan Plitnick, Catharine Prussing, Navjot Singh, John Kelly, Erasmus Schneider, Erica Lasek-Nesselquist |
| EPI_ISL_1671349 | STONY BROOK UNIVERSITY HOSPITAL | Wadsworth Center, New York State Department of Health | Kirsten St. George, Daryl M. Lamson, Alexis Russell, Matthew Shudt, Melissa A Leisner, Jonathan Plitnick, Catharine Prussing, Navjot Singh, John Kelly, Erasmus Schneider, Erica Lasek-Nesselquist |
| EPI_ISL_1671350, EPI_ISL_1671351 | ALBANY MEDICAL CENTER | Wadsworth Center, New York State Department of Health | Kirsten St. George, Daryl M. Lamson, Alexis Russell, Matthew Shudt, Melissa A Leisner, Jonathan Plitnick, Catharine Prussing, Navjot Singh, John Kelly, Erasmus Schneider, Erica Lasek-Nesselquist |
| EPI_ISL_1671352, EPI_ISL_1671353 | TEMPUS LABS INC | Wadsworth Center, New York State Department of Health | Kirsten St. George, Daryl M. Lamson, Alexis Russell, Matthew Shudt, Melissa A Leisner, Jonathan Plitnick, Catharine Prussing, Navjot Singh, John Kelly, Erasmus Schneider, Erica Lasek-Nesselquist |
| EPI_ISL_1671354 | ALBANY MEDICAL CENTER | Wadsworth Center, New York State Department of Health | Kirsten St. George, Daryl M. Lamson, Alexis Russell, Matthew Shudt, Melissa A Leisner, Jonathan Plitnick, Catharine Prussing, Navjot Singh, John Kelly, Erasmus Schneider, Erica Lasek-Nesselquist |
| EPI_ISL_1671355 | TEMPUS LABS INC | Wadsworth Center, New York State Department of Health | Kirsten St. George, Daryl M. Lamson, Alexis Russell, Matthew Shudt, Melissa A Leisner, Jonathan Plitnick, Catharine Prussing, Navjot Singh, John Kelly, Erasmus Schneider, Erica Lasek-Nesselquist |
| EPI_ISL_1671356 | STONY BROOK UNIVERSITY HOSPITAL | Wadsworth Center, New York State Department of Health | Kirsten St. George, Daryl M. Lamson, Alexis Russell, Matthew Shudt, Melissa A Leisner, Jonathan Plitnick, Catharine Prussing, Navjot Singh, John Kelly, Erasmus Schneider, Erica Lasek-Nesselquist |
| EPI_ISL_1671357, EPI_ISL_1671358, EPI_ISL_1671359 | ALBANY MEDICAL CENTER | Wadsworth Center, New York State Department of Health | Kirsten St. George, Daryl M. Lamson, Alexis Russell, Matthew Shudt, Melissa A Leisner, Jonathan Plitnick, Catharine Prussing, Navjot Singh, John Kelly, Erasmus Schneider, Erica Lasek-Nesselquist |
| EPI_ISL_1671444, EPI_ISL_1671445, EPI_ISL_1671446, EPI_ISL_1671447 | BOSTON HEART DIAGNOSTICS CORP | Wadsworth Center, New York State Department of Health | Kirsten St. George, Daryl M. Lamson, Alexis Russell, Matthew Shudt, Melissa A Leisner, Jonathan Plitnick, Catharine Prussing, Navjot Singh, John Kelly, Erasmus Schneider, Erica Lasek-Nesselquist |
| EPI_ISL_1671448 | NORTH SHORE UNIVERSITY HOSPITAL | Wadsworth Center, New York State Department of Health | Kirsten St. George, Daryl M. Lamson, Alexis Russell, Matthew Shudt, Melissa A Leisner, Jonathan Plitnick, Catharine Prussing, Navjot Singh, John Kelly, Erasmus Schneider, Erica Lasek-Nesselquist |
| EPI_ISL_1671449 | WESTCHESTER MEDICAL CENTER | Wadsworth Center, New York State Department of Health | Kirsten St. George, Daryl M. Lamson, Alexis Russell, Matthew Shudt, Melissa A Leisner, Jonathan Plitnick, Catharine Prussing, Navjot Singh, John Kelly, Erasmus Schneider, Erica Lasek-Nesselquist |
| EPI_ISL_1671450, EPI_ISL_1671451 | ALBANY MEDICAL CENTER | Wadsworth Center, New York State Department of Health | Kirsten St. George, Daryl M. Lamson, Alexis Russell, Matthew Shudt, Melissa A Leisner, Jonathan Plitnick, Catharine Prussing, Navjot Singh, John Kelly, Erasmus Schneider, Erica Lasek-Nesselquist |
| EPI_ISL_1671452, EPI_ISL_1671453 | NORTH SHORE UNIVERSITY HOSPITAL | Wadsworth Center, New York State Department of Health | Kirsten St. George, Daryl M. Lamson, Alexis Russell, Matthew Shudt, Melissa A Leisner, Jonathan Plitnick, Catharine Prussing, Navjot Singh, John Kelly, Erasmus Schneider, Erica Lasek-Nesselquist |
| EPI_ISL_1671454, EPI_ISL_1671455, EPI_ISL_1671456, EPI_ISL_1671457 | BOSTON HEART DIAGNOSTICS CORP | Wadsworth Center, New York State Department of Health | Kirsten St. George, Daryl M. Lamson, Alexis Russell, Matthew Shudt, Melissa A Leisner, Jonathan Plitnick, Catharine Prussing, Navjot Singh, John Kelly, Erasmus Schneider, Erica Lasek-Nesselquist |
| EPI_ISL_1671458, EPI_ISL_1671459, EPI_ISL_1671460, EPI_ISL_1671461 | WESTCHESTER MEDICAL CENTER | Wadsworth Center, New York State Department of Health | Kirsten St. George, Daryl M. Lamson, Alexis Russell, Matthew Shudt, Melissa A Leisner, Jonathan Plitnick, Catharine Prussing, Navjot Singh, John Kelly, Erasmus Schneider, Erica Lasek-Nesselquist |
| EPI_ISL_1671462 | ALBANY MEDICAL CENTER | Wadsworth Center, New York State Department of Health | Kirsten St. George, Daryl M. Lamson, Alexis Russell, Matthew Shudt, Melissa A Leisner, Jonathan Plitnick, Catharine Prussing, Navjot Singh, John Kelly, Erasmus Schneider, Erica Lasek-Nesselquist |
| EPI_ISL_1671463, EPI_ISL_1671464, EPI_ISL_1671465 | SUNY UPSTATE MEDICAL UNIVERSITY | Wadsworth Center, New York State Department of Health | Kirsten St. George, Daryl M. Lamson, Alexis Russell, Matthew Shudt, Melissa A Leisner, Jonathan Plitnick, Catharine Prussing, Navjot Singh, John Kelly, Erasmus Schneider, Erica Lasek-Nesselquist |
| EPI_ISL_1671466 | MONTEFIORE MEDICAL CENTER LABORATORIES | Wadsworth Center, New York State Department of Health | Kirsten St. George, Daryl M. Lamson, Alexis Russell, Matthew Shudt, Melissa A Leisner, Jonathan Plitnick, Catharine Prussing, Navjot Singh, John Kelly, Erasmus Schneider, Erica Lasek-Nesselquist |
| EPI_ISL_1671467 | WESTCHESTER MEDICAL CENTER | Wadsworth Center, New York State Department of Health | Kirsten St. George, Daryl M. Lamson, Alexis Russell, Matthew Shudt, Melissa A Leisner, Jonathan Plitnick, Catharine Prussing, Navjot Singh, John Kelly, Erasmus Schneider, Erica Lasek-Nesselquist |
| EPI_ISL_1671468 | ST. FRANCIS HOSPITAL LABORATORY | Wadsworth Center, New York State Department of Health | Kirsten St. George, Daryl M. Lamson, Alexis Russell, Matthew Shudt, Melissa A Leisner, Jonathan Plitnick, Catharine Prussing, Navjot Singh, John Kelly, Erasmus Schneider, Erica Lasek-Nesselquist |
| EPI_ISL_1671469, EPI_ISL_1671470, EPI_ISL_1671471, EPI_ISL_1671472, EPI_ISL_1671473, EPI_ISL_1671474, EPI_ISL_1671475 | WESTCHESTER MEDICAL CENTER | Wadsworth Center, New York State Department of Health | Kirsten St. George, Daryl M. Lamson, Alexis Russell, Matthew Shudt, Melissa A Leisner, Jonathan Plitnick, Catharine Prussing, Navjot Singh, John Kelly, Erasmus Schneider, Erica Lasek-Nesselquist |
| EPI_ISL_1671476 | BOSTON HEART DIAGNOSTICS CORP | Wadsworth Center, New York State Department of Health | Kirsten St. George, Daryl M. Lamson, Alexis Russell, Matthew Shudt, Melissa A Leisner, Jonathan Plitnick, Catharine Prussing, Navjot Singh, John Kelly, Erasmus Schneider, Erica Lasek-Nesselquist |
| EPI_ISL_1671477, EPI_ISL_1671478, | MONTEFIORE MEDICAL CENTER LABORATORIES | Wadsworth Center, New York State Department of Health | Kirsten St. George, Daryl M. Lamson, Alexis Russell, Matthew Shudt, Melissa A Leisner, Jonathan Plitnick, Catharine Prussing, Navjot Singh, John Kelly, |

| | | | |
|---|---|---|---|
| EPI_ISL_1671479 | | | Erasmus Schneider, Erica Lasek-Nesselquist |
| EPI_ISL_1671480, EPI_ISL_1671481, EPI_ISL_1671482, EPI_ISL_1671483 | WESTCHESTER MEDICAL CENTER | Wadsworth Center, New York State Department of Health | Kirsten St. George, Daryl M. Lamson, Alexis Russell, Matthew Shudt, Melissa A Leisner, Jonathan Plitnick, Catharine Prussing, Navjot Singh, John Kelly, Erasmus Schneider, Erica Lasek-Nesselquist |
| EPI_ISL_1671484 | ST. FRANCIS HOSPITAL LABORATORY | Wadsworth Center, New York State Department of Health | Kirsten St. George, Daryl M. Lamson, Alexis Russell, Matthew Shudt, Melissa A Leisner, Jonathan Plitnick, Catharine Prussing, Navjot Singh, John Kelly, Erasmus Schneider, Erica Lasek-Nesselquist |
| EPI_ISL_1671485, EPI_ISL_1671486, EPI_ISL_1671487, EPI_ISL_1671488, EPI_ISL_1671489, EPI_ISL_1671490, EPI_ISL_1671491, EPI_ISL_1671492 | WESTCHESTER MEDICAL CENTER | Wadsworth Center, New York State Department of Health | Kirsten St. George, Daryl M. Lamson, Alexis Russell, Matthew Shudt, Melissa A Leisner, Jonathan Plitnick, Catharine Prussing, Navjot Singh, John Kelly, Erasmus Schneider, Erica Lasek-Nesselquist |
| EPI_ISL_1671493 | NORTH SHORE UNIVERSITY HOSPITAL | Wadsworth Center, New York State Department of Health | Kirsten St. George, Daryl M. Lamson, Alexis Russell, Matthew Shudt, Melissa A Leisner, Jonathan Plitnick, Catharine Prussing, Navjot Singh, John Kelly, Erasmus Schneider, Erica Lasek-Nesselquist |
| EPI_ISL_1671494 | SUNY UPSTATE MEDICAL UNIVERSITY | Wadsworth Center, New York State Department of Health | Kirsten St. George, Daryl M. Lamson, Alexis Russell, Matthew Shudt, Melissa A Leisner, Jonathan Plitnick, Catharine Prussing, Navjot Singh, John Kelly, Erasmus Schneider, Erica Lasek-Nesselquist |
| EPI_ISL_1671495, EPI_ISL_1671496 | BOSTON HEART DIAGNOSTICS CORP | Wadsworth Center, New York State Department of Health | Kirsten St. George, Daryl M. Lamson, Alexis Russell, Matthew Shudt, Melissa A Leisner, Jonathan Plitnick, Catharine Prussing, Navjot Singh, John Kelly, Erasmus Schneider, Erica Lasek-Nesselquist |
| EPI_ISL_1671497, EPI_ISL_1671498, EPI_ISL_1671499 | MONTEFIORE MEDICAL CENTER LABORATORIES | Wadsworth Center, New York State Department of Health | Kirsten St. George, Daryl M. Lamson, Alexis Russell, Matthew Shudt, Melissa A Leisner, Jonathan Plitnick, Catharine Prussing, Navjot Singh, John Kelly, Erasmus Schneider, Erica Lasek-Nesselquist |
| EPI_ISL_1671500, EPI_ISL_1671501, EPI_ISL_1671502, EPI_ISL_1671503, EPI_ISL_1671504, EPI_ISL_1671505, EPI_ISL_1671506, EPI_ISL_1671507, EPI_ISL_1671508, EPI_ISL_1671509, EPI_ISL_1671510, EPI_ISL_1671511 | | | |
| see above | WESTCHESTER MEDICAL CENTER | Wadsworth Center, New York State Department of Health | Kirsten St. George, Daryl M. Lamson, Alexis Russell, Matthew Shudt, Melissa A Leisner, Jonathan Plitnick, Catharine Prussing, Navjot Singh, John Kelly, Erasmus Schneider, Erica Lasek-Nesselquist |
| EPI_ISL_1671512, EPI_ISL_1671513, EPI_ISL_1671514 | SUNY UPSTATE MEDICAL UNIVERSITY | Wadsworth Center, New York State Department of Health | Kirsten St. George, Daryl M. Lamson, Alexis Russell, Matthew Shudt, Melissa A Leisner, Jonathan Plitnick, Catharine Prussing, Navjot Singh, John Kelly, Erasmus Schneider, Erica Lasek-Nesselquist |
| EPI_ISL_1671596 | BOSTON HEART DIAGNOSTICS CORP | Wadsworth Center, New York State Department of Health | Kirsten St. George, Daryl M. Lamson, Alexis Russell, Matthew Shudt, Melissa A Leisner, Jonathan Plitnick, Catharine Prussing, Navjot Singh, John Kelly, Erasmus Schneider, Erica Lasek-Nesselquist |
| EPI_ISL_1671597 | WESTCHESTER MEDICAL CENTER | Wadsworth Center, New York State Department of Health | Kirsten St. George, Daryl M. Lamson, Alexis Russell, Matthew Shudt, Melissa A Leisner, Jonathan Plitnick, Catharine Prussing, Navjot Singh, John Kelly, Erasmus Schneider, Erica Lasek-Nesselquist |
| EPI_ISL_1671598 | WHITE PLAINS HOSPITAL CENTER LABORATORY | Wadsworth Center, New York State Department of Health | Kirsten St. George, Daryl M. Lamson, Alexis Russell, Matthew Shudt, Melissa A Leisner, Jonathan Plitnick, Catharine Prussing, Navjot Singh, John Kelly, Erasmus Schneider, Erica Lasek-Nesselquist |
| EPI_ISL_1671599 | ALBANY MEDICAL CENTER | Wadsworth Center, New York State Department of Health | Kirsten St. George, Daryl M. Lamson, Alexis Russell, Matthew Shudt, Melissa A Leisner, Jonathan Plitnick, Catharine Prussing, Navjot Singh, John Kelly, Erasmus Schneider, Erica Lasek-Nesselquist |
| EPI_ISL_1671600 | WHITE PLAINS HOSPITAL CENTER LABORATORY | Wadsworth Center, New York State Department of Health | Kirsten St. George, Daryl M. Lamson, Alexis Russell, Matthew Shudt, Melissa A Leisner, Jonathan Plitnick, Catharine Prussing, Navjot Singh, John Kelly, Erasmus Schneider, Erica Lasek-Nesselquist |
| EPI_ISL_1671601, EPI_ISL_1671602, EPI_ISL_1671603, EPI_ISL_1671604, EPI_ISL_1671605, EPI_ISL_1671606 | NORTHWELL HEALTH LABORATORIES | Wadsworth Center, New York State Department of Health | Kirsten St. George, Daryl M. Lamson, Alexis Russell, Matthew Shudt, Melissa A Leisner, Jonathan Plitnick, Catharine Prussing, Navjot Singh, John Kelly, Erasmus Schneider, Erica Lasek-Nesselquist |
| EPI_ISL_1671607 | WHITE PLAINS HOSPITAL CENTER LABORATORY | Wadsworth Center, New York State Department of Health | Kirsten St. George, Daryl M. Lamson, Alexis Russell, Matthew Shudt, Melissa A Leisner, Jonathan Plitnick, Catharine Prussing, Navjot Singh, John Kelly, Erasmus Schneider, Erica Lasek-Nesselquist |
| EPI_ISL_1671608, EPI_ISL_1671609 | NORTHWELL HEALTH LABORATORIES | Wadsworth Center, New York State Department of Health | Kirsten St. George, Daryl M. Lamson, Alexis Russell, Matthew Shudt, Melissa A Leisner, Jonathan Plitnick, Catharine Prussing, Navjot Singh, John Kelly, Erasmus Schneider, Erica Lasek-Nesselquist |
| EPI_ISL_1671610, EPI_ISL_1671611 | WHITE PLAINS HOSPITAL CENTER LABORATORY | Wadsworth Center, New York State Department of Health | Kirsten St. George, Daryl M. Lamson, Alexis Russell, Matthew Shudt, Melissa A Leisner, Jonathan Plitnick, Catharine Prussing, Navjot Singh, John Kelly, Erasmus Schneider, Erica Lasek-Nesselquist |
| EPI_ISL_1671612 | SUNY UPSTATE MEDICAL UNIVERSITY | Wadsworth Center, New York State Department of Health | Kirsten St. George, Daryl M. Lamson, Alexis Russell, Matthew Shudt, Melissa A Leisner, Jonathan Plitnick, Catharine Prussing, Navjot Singh, John Kelly, Erasmus Schneider, Erica Lasek-Nesselquist |
| EPI_ISL_1671613 | URMC LABS | Wadsworth Center, New York State Department of Health | Kirsten St. George, Daryl M. Lamson, Alexis Russell, Matthew Shudt, Melissa A Leisner, Jonathan Plitnick, Catharine Prussing, Navjot Singh, John Kelly, Erasmus Schneider, Erica Lasek-Nesselquist |
| EPI_ISL_1671614, EPI_ISL_1671615, EPI_ISL_1671616, EPI_ISL_1671617 | NORTHWELL HEALTH LABORATORIES | Wadsworth Center, New York State Department of Health | Kirsten St. George, Daryl M. Lamson, Alexis Russell, Matthew Shudt, Melissa A Leisner, Jonathan Plitnick, Catharine Prussing, Navjot Singh, John Kelly, Erasmus Schneider, Erica Lasek-Nesselquist |
| EPI_ISL_1671618 | BOSTON HEART DIAGNOSTICS CORP | Wadsworth Center, New York State Department of Health | Kirsten St. George, Daryl M. Lamson, Alexis Russell, Matthew Shudt, Melissa A Leisner, Jonathan Plitnick, Catharine Prussing, Navjot Singh, John Kelly, Erasmus Schneider, Erica Lasek-Nesselquist |
| EPI_ISL_1671619 | WESTCHESTER MEDICAL CENTER | Wadsworth Center, New York State Department of Health | Kirsten St. George, Daryl M. Lamson, Alexis Russell, Matthew Shudt, Melissa A Leisner, Jonathan Plitnick, Catharine Prussing, Navjot Singh, John Kelly, Erasmus Schneider, Erica Lasek-Nesselquist |
| EPI_ISL_1671620, EPI_ISL_1671621 | NORTHWELL HEALTH LABORATORIES | Wadsworth Center, New York State Department of Health | Kirsten St. George, Daryl M. Lamson, Alexis Russell, Matthew Shudt, Melissa A Leisner, Jonathan Plitnick, Catharine Prussing, Navjot Singh, John Kelly, Erasmus Schneider, Erica Lasek-Nesselquist |
| EPI_ISL_1671622 | WHITE PLAINS HOSPITAL CENTER LABORATORY | Wadsworth Center, New York State Department of Health | Kirsten St. George, Daryl M. Lamson, Alexis Russell, Matthew Shudt, Melissa A Leisner, Jonathan Plitnick, Catharine Prussing, Navjot Singh, John Kelly, Erasmus Schneider, Erica Lasek-Nesselquist |
| EPI_ISL_1671623 | ALBANY MEDICAL CENTER | Wadsworth Center, New York State Department of Health | Kirsten St. George, Daryl M. Lamson, Alexis Russell, Matthew Shudt, Melissa A Leisner, Jonathan Plitnick, Catharine Prussing, Navjot Singh, John Kelly, Erasmus Schneider, Erica Lasek-Nesselquist |
| EPI_ISL_1671624, EPI_ISL_1671625 | WESTCHESTER MEDICAL CENTER | Wadsworth Center, New York State Department of Health | Kirsten St. George, Daryl M. Lamson, Alexis Russell, Matthew Shudt, Melissa A Leisner, Jonathan Plitnick, Catharine Prussing, Navjot Singh, John Kelly, Erasmus Schneider, Erica Lasek-Nesselquist |
| EPI_ISL_1671626, EPI_ISL_1671627 | URMC LABS | Wadsworth Center, New York State Department of Health | Kirsten St. George, Daryl M. Lamson, Alexis Russell, Matthew Shudt, Melissa A Leisner, Jonathan Plitnick, Catharine Prussing, Navjot Singh, John Kelly, Erasmus Schneider, Erica Lasek-Nesselquist |
| EPI_ISL_1671628, EPI_ISL_1671629, EPI_ISL_1671630, EPI_ISL_1671631, EPI_ISL_1671632, EPI_ISL_1671633 | NORTHWELL HEALTH LABORATORIES | Wadsworth Center, New York State Department of Health | Kirsten St. George, Daryl M. Lamson, Alexis Russell, Matthew Shudt, Melissa A Leisner, Jonathan Plitnick, Catharine Prussing, Navjot Singh, John Kelly, Erasmus Schneider, Erica Lasek-Nesselquist |
| EPI_ISL_1671634 | URMC LABS | Wadsworth Center, New York State Department of Health | Kirsten St. George, Daryl M. Lamson, Alexis Russell, Matthew Shudt, Melissa A Leisner, Jonathan Plitnick, Catharine Prussing, Navjot Singh, John Kelly, Erasmus Schneider, Erica Lasek-Nesselquist |
| EPI_ISL_1671635, EPI_ISL_1671636, EPI_ISL_1671637 | WESTCHESTER MEDICAL CENTER | Wadsworth Center, New York State Department of Health | Kirsten St. George, Daryl M. Lamson, Alexis Russell, Matthew Shudt, Melissa A Leisner, Jonathan Plitnick, Catharine Prussing, Navjot Singh, John Kelly, Erasmus Schneider, Erica Lasek-Nesselquist |
| EPI_ISL_1671638 | NORTHWELL HEALTH LABORATORIES | Wadsworth Center, New York State Department of Health | Kirsten St. George, Daryl M. Lamson, Alexis Russell, Matthew Shudt, Melissa A Leisner, Jonathan Plitnick, Catharine Prussing, Navjot Singh, John Kelly, Erasmus Schneider, Erica Lasek-Nesselquist |
| EPI_ISL_1671639 | WESTCHESTER MEDICAL CENTER | Wadsworth Center, New York State Department of Health | Kirsten St. George, Daryl M. Lamson, Alexis Russell, Matthew Shudt, Melissa A Leisner, Jonathan Plitnick, Catharine Prussing, Navjot Singh, John Kelly, |

| | | | |
|---|---|---|---|
| | | | Erasmus Schneider, Erica Lasek-Nesselquist |
| EPI_ISL_1671640, EPI_ISL_1671641 | NORTHWELL HEALTH LABORATORIES | Wadsworth Center, New York State Department of Health | Kirsten St. George, Daryl M. Lamson, Alexis Russell, Matthew Shudt, Melissa A Leisner, Jonathan Plitnick, Catharine Prussing, Navjot Singh, John Kelly, Erasmus Schneider, Erica Lasek-Nesselquist |
| EPI_ISL_1671642 | MONTEFIORE MEDICAL CENTER LABORATORIES | Wadsworth Center, New York State Department of Health | Kirsten St. George, Daryl M. Lamson, Alexis Russell, Matthew Shudt, Melissa A Leisner, Jonathan Plitnick, Catharine Prussing, Navjot Singh, John Kelly, Erasmus Schneider, Erica Lasek-Nesselquist |
| EPI_ISL_1671643 | TEMPUS LABS INC | Wadsworth Center, New York State Department of Health | Kirsten St. George, Daryl M. Lamson, Alexis Russell, Matthew Shudt, Melissa A Leisner, Jonathan Plitnick, Catharine Prussing, Navjot Singh, John Kelly, Erasmus Schneider, Erica Lasek-Nesselquist |
| EPI_ISL_1671644 | WHITE PLAINS HOSPITAL CENTER LABORATORY | Wadsworth Center, New York State Department of Health | Kirsten St. George, Daryl M. Lamson, Alexis Russell, Matthew Shudt, Melissa A Leisner, Jonathan Plitnick, Catharine Prussing, Navjot Singh, John Kelly, Erasmus Schneider, Erica Lasek-Nesselquist |
| EPI_ISL_1671645 | ALBANY MEDICAL CENTER | Wadsworth Center, New York State Department of Health | Kirsten St. George, Daryl M. Lamson, Alexis Russell, Matthew Shudt, Melissa A Leisner, Jonathan Plitnick, Catharine Prussing, Navjot Singh, John Kelly, Erasmus Schneider, Erica Lasek-Nesselquist |
| EPI_ISL_1671646, EPI_ISL_1671647 | WESTCHESTER MEDICAL CENTER | Wadsworth Center, New York State Department of Health | Kirsten St. George, Daryl M. Lamson, Alexis Russell, Matthew Shudt, Melissa A Leisner, Jonathan Plitnick, Catharine Prussing, Navjot Singh, John Kelly, Erasmus Schneider, Erica Lasek-Nesselquist |
| EPI_ISL_1671648, EPI_ISL_1671649, EPI_ISL_1671650, EPI_ISL_1671651, EPI_ISL_1671652, EPI_ISL_1671653 | NORTHWELL HEALTH LABORATORIES | Wadsworth Center, New York State Department of Health | Kirsten St. George, Daryl M. Lamson, Alexis Russell, Matthew Shudt, Melissa A Leisner, Jonathan Plitnick, Catharine Prussing, Navjot Singh, John Kelly, Erasmus Schneider, Erica Lasek-Nesselquist |
| EPI_ISL_1671654 | MONTEFIORE MEDICAL CENTER LABORATORIES | Wadsworth Center, New York State Department of Health | Kirsten St. George, Daryl M. Lamson, Alexis Russell, Matthew Shudt, Melissa A Leisner, Jonathan Plitnick, Catharine Prussing, Navjot Singh, John Kelly, Erasmus Schneider, Erica Lasek-Nesselquist |
| EPI_ISL_1671655, EPI_ISL_1671656 | WHITE PLAINS HOSPITAL CENTER LABORATORY | Wadsworth Center, New York State Department of Health | Kirsten St. George, Daryl M. Lamson, Alexis Russell, Matthew Shudt, Melissa A Leisner, Jonathan Plitnick, Catharine Prussing, Navjot Singh, John Kelly, Erasmus Schneider, Erica Lasek-Nesselquist |
| EPI_ISL_1671657, EPI_ISL_1671658, EPI_ISL_1671659, EPI_ISL_1671660 | WESTCHESTER MEDICAL CENTER | Wadsworth Center, New York State Department of Health | Kirsten St. George, Daryl M. Lamson, Alexis Russell, Matthew Shudt, Melissa A Leisner, Jonathan Plitnick, Catharine Prussing, Navjot Singh, John Kelly, Erasmus Schneider, Erica Lasek-Nesselquist |
| EPI_ISL_1671661, EPI_ISL_1671662, EPI_ISL_1671663 | NORTHWELL HEALTH LABORATORIES | Wadsworth Center, New York State Department of Health | Kirsten St. George, Daryl M. Lamson, Alexis Russell, Matthew Shudt, Melissa A Leisner, Jonathan Plitnick, Catharine Prussing, Navjot Singh, John Kelly, Erasmus Schneider, Erica Lasek-Nesselquist |
| EPI_ISL_1671664 | MONTEFIORE MEDICAL CENTER LABORATORIES | Wadsworth Center, New York State Department of Health | Kirsten St. George, Daryl M. Lamson, Alexis Russell, Matthew Shudt, Melissa A Leisner, Jonathan Plitnick, Catharine Prussing, Navjot Singh, John Kelly, Erasmus Schneider, Erica Lasek-Nesselquist |
| EPI_ISL_1671665 | WHITE PLAINS HOSPITAL CENTER LABORATORY | Wadsworth Center, New York State Department of Health | Kirsten St. George, Daryl M. Lamson, Alexis Russell, Matthew Shudt, Melissa A Leisner, Jonathan Plitnick, Catharine Prussing, Navjot Singh, John Kelly, Erasmus Schneider, Erica Lasek-Nesselquist |
| EPI_ISL_1671666 | WESTCHESTER MEDICAL CENTER | Wadsworth Center, New York State Department of Health | Kirsten St. George, Daryl M. Lamson, Alexis Russell, Matthew Shudt, Melissa A Leisner, Jonathan Plitnick, Catharine Prussing, Navjot Singh, John Kelly, Erasmus Schneider, Erica Lasek-Nesselquist |
| EPI_ISL_1671667 | URMC LABS | Wadsworth Center, New York State Department of Health | Kirsten St. George, Daryl M. Lamson, Alexis Russell, Matthew Shudt, Melissa A Leisner, Jonathan Plitnick, Catharine Prussing, Navjot Singh, John Kelly, Erasmus Schneider, Erica Lasek-Nesselquist |
| EPI_ISL_1671668, EPI_ISL_1671669, EPI_ISL_1671670, EPI_ISL_1671671, EPI_ISL_1671672, EPI_ISL_1671673, EPI_ISL_1671674, EPI_ISL_1671675, EPI_ISL_1671676 | NORTHWELL HEALTH LABORATORIES | Wadsworth Center, New York State Department of Health | Kirsten St. George, Daryl M. Lamson, Alexis Russell, Matthew Shudt, Melissa A Leisner, Jonathan Plitnick, Catharine Prussing, Navjot Singh, John Kelly, Erasmus Schneider, Erica Lasek-Nesselquist |
| EPI_ISL_1671677 | SARATOGA HOSPITAL LABORATORY | Wadsworth Center, New York State Department of Health | Kirsten St. George, Daryl M. Lamson, Alexis Russell, Matthew Shudt, Melissa A Leisner, Jonathan Plitnick, Catharine Prussing, Navjot Singh, John Kelly, Erasmus Schneider, Erica Lasek-Nesselquist |
| EPI_ISL_1674703 | Murphy Medical Association | Grubaugh Lab - Yale School of Public Health | Joseph Fauver, Mallery Breban, Isabel Ott, Tara Alpert, Mary Petrone, Anderson Brito, Chantal Vogels, Annie Watkins, Chaney Kalinich, Jessica Rothman, Caleb Neal, Eva Laszlo, Steven Murphy, Nathan Grubaugh |
| EPI_ISL_1678056, EPI_ISL_1678057, EPI_ISL_1678058, EPI_ISL_1678059, EPI_ISL_1678060, EPI_ISL_1678061, EPI_ISL_1678062, EPI_ISL_1678063, EPI_ISL_1678064, EPI_ISL_1678065, EPI_ISL_1678066, EPI_ISL_1678067, EPI_ISL_1678068, EPI_ISL_1678069, EPI_ISL_1678070, EPI_ISL_1678071, EPI_ISL_1678072, EPI_ISL_1678073, EPI_ISL_1678074, EPI_ISL_1678075, EPI_ISL_1678076, EPI_ISL_1678077, EPI_ISL_1678078, EPI_ISL_1678079, EPI_ISL_1678080, EPI_ISL_1678257, EPI_ISL_1678258, EPI_ISL_1678259, EPI_ISL_1678260, EPI_ISL_1678261, EPI_ISL_1678262, EPI_ISL_1678263, EPI_ISL_1678264, EPI_ISL_1678265 | | | |
| see above | NYC Department of Health and Mental Hygiene | Centers for Disease Control and Prevention Division of Viral Diseases, Pathogen Discovery | Mili Sheth, Sarah Nobles, Jasmine Padilla, Mark Burroughs, Shoshona Le, Katie Dillon, Peter Cook, Clinton R. Paden, Dhwani Batra, Krista Queen, Kristen Knipe, Dakota Howard, Yvette Unoarumhi, Darlene Wagner, Matthew Schmerer, Ben L. Rambo-Martin, Kristine Lacek, Sam Shepard, Alison Laufer Halpin, Dave Wentworth, Vivien Dugan, Suxiang Tong, Justin Lee |
| EPI_ISL_1679008, EPI_ISL_1679009, EPI_ISL_1679029, EPI_ISL_1679426, EPI_ISL_1679621, EPI_ISL_1679640, EPI_ISL_1679734 | Helix/Illumina | Centers for Disease Control and Prevention Division of Viral Diseases, Pathogen Discovery | Dakota Howard, Dhwani Batra, Peter W. Cook, Kara Moser, Adrian Paskey, Jason Caravas, Benjamin Rambo-Martin, Shatavia Morrison, Christopher Gulvick, Scott Sammons, Yvette Unoarumhi, Darlene Wagner, Matthew Schmerer, Eileen de Feo, Jan Antico, Christine Tran, Matthew Tolentino, Shannon Wickline, Kim Gietzen, Brad Sickler, Jingtao Liu, Eric Allen, Phil Febbo, Nicole L. Washington, Simon White, Geraint Levan, Kelly Schiabor Barrett, Elizabeth Cirulli, Alexandre Bolze, Ary Ascencio, Charlotte Rivera-Garcia, Ryan Cho, Jason Nguyen, Sherry Wang, Jimmy Ramirez, Tyler Cassens, Efren Sandoval, Magnus Isaksson, William Lee, David Becker, Marc Laurent, James Lu, Clinton R. Paden, Duncan MacCannell |
| EPI_ISL_1680473, EPI_ISL_1680474, EPI_ISL_1680475, EPI_ISL_1680477, EPI_ISL_1680479, EPI_ISL_1680480, EPI_ISL_1680482, EPI_ISL_1680483, EPI_ISL_1680484, EPI_ISL_1680485, EPI_ISL_1680486, EPI_ISL_1680487, EPI_ISL_1680488, EPI_ISL_1680489, EPI_ISL_1680490, EPI_ISL_1680536, EPI_ISL_1680540, EPI_ISL_1680541, EPI_ISL_1680542, EPI_ISL_1680543, EPI_ISL_1680544, EPI_ISL_1680545, EPI_ISL_1680549, EPI_ISL_1680550, EPI_ISL_1680551, EPI_ISL_1680552, EPI_ISL_1680553, EPI_ISL_1680554, EPI_ISL_1680566, EPI_ISL_1680585, EPI_ISL_1680586, EPI_ISL_1680590, EPI_ISL_1680591, EPI_ISL_1680592, EPI_ISL_1680593, EPI_ISL_1680594, EPI_ISL_1680595, EPI_ISL_1680596, EPI_ISL_1680597, EPI_ISL_1680598, EPI_ISL_1680599, EPI_ISL_1680603, EPI_ISL_1680638, EPI_ISL_1680639, EPI_ISL_1680640, EPI_ISL_1680641, EPI_ISL_1680642, EPI_ISL_1680643, EPI_ISL_1680644, EPI_ISL_1680672, EPI_ISL_1680674, EPI_ISL_1680747, EPI_ISL_1680748, EPI_ISL_1680765, EPI_ISL_1680767, EPI_ISL_1680768, EPI_ISL_1680769, EPI_ISL_1680802, EPI_ISL_1680847, EPI_ISL_1680848, EPI_ISL_1681168, EPI_ISL_1681169, EPI_ISL_1681170, EPI_ISL_1681171, EPI_ISL_1681172, EPI_ISL_1681173, EPI_ISL_1681187, EPI_ISL_1681188, EPI_ISL_1681190, EPI_ISL_1681193, EPI_ISL_1681195, EPI_ISL_1681249, EPI_ISL_1681250, EPI_ISL_1681251, EPI_ISL_1681309, EPI_ISL_1681310, EPI_ISL_1681311, EPI_ISL_1681314, EPI_ISL_1681315, EPI_ISL_1681316, EPI_ISL_1681318, EPI_ISL_1681321, EPI_ISL_1681370, EPI_ISL_1681371, EPI_ISL_1681372, EPI_ISL_1681409, EPI_ISL_1681415, EPI_ISL_1681416, EPI_ISL_1681417, EPI_ISL_1681418, EPI_ISL_1681419, EPI_ISL_1681420, EPI_ISL_1681444, EPI_ISL_1681445, EPI_ISL_1681446, EPI_ISL_1681447, EPI_ISL_1681448, EPI_ISL_1681451, EPI_ISL_1681452, EPI_ISL_1681453, EPI_ISL_1681461, EPI_ISL_1681495, EPI_ISL_1681496, EPI_ISL_1681497, EPI_ISL_1681498, EPI_ISL_1681499, EPI_ISL_1681500, EPI_ISL_1681542, EPI_ISL_1681543, EPI_ISL_1681544, EPI_ISL_1681545, EPI_ISL_1681546, EPI_ISL_1681547, EPI_ISL_1681548, EPI_ISL_1681549, EPI_ISL_1681550, EPI_ISL_1681551, EPI_ISL_1681552, EPI_ISL_1681644, EPI_ISL_1681645, EPI_ISL_1681646, EPI_ISL_1681647, EPI_ISL_1681648, EPI_ISL_1681649, EPI_ISL_1682025, EPI_ISL_1682026, EPI_ISL_1682061, EPI_ISL_1682062, EPI_ISL_1682071, EPI_ISL_1682085, EPI_ISL_1682109, EPI_ISL_1682111, EPI_ISL_1682137, EPI_ISL_1682161, EPI_ISL_1682162, EPI_ISL_1682522, EPI_ISL_1682525, EPI_ISL_1682596, EPI_ISL_1682608, EPI_ISL_1682787, EPI_ISL_1682808, EPI_ISL_1682811, EPI_ISL_1682929, EPI_ISL_1682961, EPI_ISL_1683040, EPI_ISL_1683058, EPI_ISL_1683061, EPI_ISL_1683064, EPI_ISL_1683088, EPI_ISL_1683104, EPI_ISL_1683107, EPI_ISL_1683126, EPI_ISL_1683132, EPI_ISL_1683136, EPI_ISL_1683180, EPI_ISL_1683181, EPI_ISL_1683185, EPI_ISL_1683187, EPI_ISL_1683197, EPI_ISL_1683202, EPI_ISL_1683203, EPI_ISL_1683210, EPI_ISL_1683212, EPI_ISL_1683214, EPI_ISL_1683215, EPI_ISL_1683216, EPI_ISL_1683248, EPI_ISL_1683249, EPI_ISL_1683254, EPI_ISL_1683258, EPI_ISL_1683260, EPI_ISL_1683262, EPI_ISL_1683265, EPI_ISL_1683266, EPI_ISL_1683267, EPI_ISL_1683277, EPI_ISL_1683281, EPI_ISL_1683289, EPI_ISL_1683290, EPI_ISL_1683292, EPI_ISL_1683293, EPI_ISL_1683294, EPI_ISL_1683295, EPI_ISL_1683299, EPI_ISL_1683321, EPI_ISL_1683322, EPI_ISL_1683323, EPI_ISL_1683325, EPI_ISL_1683326, EPI_ISL_1683330, EPI_ISL_1683331, EPI_ISL_1683332, EPI_ISL_1683333, EPI_ISL_1683336, EPI_ISL_1683351, EPI_ISL_1683358, EPI_ISL_1683361, EPI_ISL_1683381, EPI_ISL_1683409, EPI_ISL_1683410, EPI_ISL_1683413, EPI_ISL_1683458, EPI_ISL_1683461, EPI_ISL_1683464, EPI_ISL_1683485, EPI_ISL_1683494, EPI_ISL_1683497, EPI_ISL_1683499, EPI_ISL_1683501, EPI_ISL_1683503, EPI_ISL_1683504, EPI_ISL_1683531, EPI_ISL_1683532, EPI_ISL_1683536, EPI_ISL_1683537, EPI_ISL_1683542, EPI_ISL_1683543, EPI_ISL_1683558, EPI_ISL_1683566, EPI_ISL_1683599, EPI_ISL_1683617, EPI_ISL_1683634, EPI_ISL_1683635, EPI_ISL_1683661, EPI_ISL_1683662, EPI_ISL_1683663, EPI_ISL_1683664, EPI_ISL_1683666, EPI_ISL_1683667, EPI_ISL_1683668, EPI_ISL_1683669, EPI_ISL_1683670, EPI_ISL_1683672, EPI_ISL_1683675, EPI_ISL_1683701, EPI_ISL_1683749, EPI_ISL_1683756, EPI_ISL_1683760, EPI_ISL_1683768, EPI_ISL_1683769, EPI_ISL_1683772, EPI_ISL_1683799, EPI_ISL_1683800, EPI_ISL_1683808, EPI_ISL_1683811, EPI_ISL_1683833, EPI_ISL_1683835, EPI_ISL_1683836, EPI_ISL_1683837, EPI_ISL_1683839, EPI_ISL_1683909, EPI_ISL_1683910, EPI_ISL_1684063, EPI_ISL_1684065, EPI_ISL_1684071, EPI_ISL_1684084, EPI_ISL_1684085, EPI_ISL_1684153, EPI_ISL_1684162, EPI_ISL_1684164, EPI_ISL_1684165, EPI_ISL_1684167, EPI_ISL_1684168, EPI_ISL_1684173, EPI_ISL_1684174, EPI_ISL_1684177, EPI_ISL_1684211, EPI_ISL_1684212, EPI_ISL_1684213, EPI_ISL_1684214, EPI_ISL_1684253, EPI_ISL_1684254, EPI_ISL_1684255, EPI_ISL_1684256, EPI_ISL_1684261, EPI_ISL_1684263, EPI_ISL_1684265, EPI_ISL_1684266, EPI_ISL_1684267, EPI_ISL_1684271, EPI_ISL_1684276, EPI_ISL_1684277, EPI_ISL_1684279, EPI_ISL_1684287, EPI_ISL_1684289, EPI_ISL_1684303, EPI_ISL_1684305, EPI_ISL_1684307, EPI_ISL_1684541, EPI_ISL_1685066 | | | |
| see above | Laboratory Corporation of America | Centers for Disease Control and Prevention Division of Viral Diseases, Pathogen Discovery | Dakota Howard, Dhwani Batra, Peter W. Cook, Kara Moser, Adrian Paskey, Jason Caravas, Benjamin Rambo-Martin, Shatavia Morrison, Christopher Gulvick, Scott Sammons, Yvette Unoarumhi, Darlene Wagner, Matthew Schmerer, Minoo Agarwal, Eyad Almasri, Debbie Boles, Ayla Burns, Nuthawin Charoensri, Oren Cohen, Susan Countryman, Mary Ann Cristobal, Bobbi Croy, Suzanne Dale, Hrushikesh Deshmukh, Amanda Douglas, Vincent Drouillon, Marcia Eisenberg, Howard Engler, Rama Ghatti, Prashant Gupta, Susan Hicks, Jake Humphrey, Lax Iyer, Manoj Jain, Mohan Kolli, Brian Krueger, Tim Kuphal, Stanley Letovsky, Michael Levandoski, Craig Lukasik, Jonathan Meltzer, Brian Norvell, Mindy Nye, Scott Parker, Christos Petropoulos, John Pruitt, Steven Ragan, Scott Ryan, Mike Sapeta, Jana Schroth, Suresh Babu Selvaraju, Goran Stevovic, Amanda Suchanek, Andrea Throop, Lyndon Tilson, Thomas Urban, Joe Voshell, Kimberly Wagner, Jonathan Williams, Mary Williamson, Qian Zeng, Tricia Zwiefelhofer, Clinton R. Paden, Duncan MacCannell |

| | | | |
|---|---|---|---|
| EPI_ISL_1685483, EPI_ISL_1685561, EPI_ISL_1685578, EPI_ISL_1685586, EPI_ISL_1685611, EPI_ISL_1685612, EPI_ISL_1685689, EPI_ISL_1685703, EPI_ISL_1685713, EPI_ISL_1685759, EPI_ISL_1685824, EPI_ISL_1685834, EPI_ISL_1685835, EPI_ISL_1685842, EPI_ISL_1685928, EPI_ISL_1685938, EPI_ISL_1685948, EPI_ISL_1685956, EPI_ISL_1685966, EPI_ISL_1685981, EPI_ISL_1685983, EPI_ISL_1685992, EPI_ISL_1686008, EPI_ISL_1686100, EPI_ISL_1686169, EPI_ISL_1686170, EPI_ISL_1686173, EPI_ISL_1686207, EPI_ISL_1686213, EPI_ISL_1686217, EPI_ISL_1686220, EPI_ISL_1686224, EPI_ISL_1686227, EPI_ISL_1686264, EPI_ISL_1686281, EPI_ISL_1686282, EPI_ISL_1686294, EPI_ISL_1686373, EPI_ISL_1686480, EPI_ISL_1686515, EPI_ISL_1686522, EPI_ISL_1686525, EPI_ISL_1686527, EPI_ISL_1686535, EPI_ISL_1686536, EPI_ISL_1686538, EPI_ISL_1686691, EPI_ISL_1686760, EPI_ISL_1686769, EPI_ISL_1686771, EPI_ISL_1686772, EPI_ISL_1686773, EPI_ISL_1686774, EPI_ISL_1686776, EPI_ISL_1686778, EPI_ISL_1686782, EPI_ISL_1686783, EPI_ISL_1686786, EPI_ISL_1686792, EPI_ISL_1686793, EPI_ISL_1686795, EPI_ISL_1686817, EPI_ISL_1686863, EPI_ISL_1686865, EPI_ISL_1686867, EPI_ISL_1686869, EPI_ISL_1686879, EPI_ISL_1686958, EPI_ISL_1686972, EPI_ISL_1686973, EPI_ISL_1686975, EPI_ISL_1687061, EPI_ISL_1687120, EPI_ISL_1687230, EPI_ISL_1687262, EPI_ISL_1687299, EPI_ISL_1687316, EPI_ISL_1687343, EPI_ISL_1687358, EPI_ISL_1687387, EPI_ISL_1687426, EPI_ISL_1687491, EPI_ISL_1687502, EPI_ISL_1687506, EPI_ISL_1687507, EPI_ISL_1687508, EPI_ISL_1687579, EPI_ISL_1687587, EPI_ISL_1687615, EPI_ISL_1687650, EPI_ISL_1687651, EPI_ISL_1687674, EPI_ISL_1687680, EPI_ISL_1687681, EPI_ISL_1687736, EPI_ISL_1687737, EPI_ISL_1687738, EPI_ISL_1687748, EPI_ISL_1687781, EPI_ISL_1687782, EPI_ISL_1687783, EPI_ISL_1687809, EPI_ISL_1687824, EPI_ISL_1687832, EPI_ISL_1687833, EPI_ISL_1687834, EPI_ISL_1687835, EPI_ISL_1687838, EPI_ISL_1687842, EPI_ISL_1687848, EPI_ISL_1687856, EPI_ISL_1687858, EPI_ISL_1687871, EPI_ISL_1688064, EPI_ISL_1688065, EPI_ISL_1688071, EPI_ISL_1688132, EPI_ISL_1688159, EPI_ISL_1688306, EPI_ISL_1688421, EPI_ISL_1688548, EPI_ISL_1688551, EPI_ISL_1688609, EPI_ISL_1688616, EPI_ISL_1688620, EPI_ISL_1688634, EPI_ISL_1688638, EPI_ISL_1688689, EPI_ISL_1688937, EPI_ISL_1688963, EPI_ISL_1689021, EPI_ISL_1689086, EPI_ISL_1689186, EPI_ISL_1689197, EPI_ISL_1689200, EPI_ISL_1689202, EPI_ISL_1689207, EPI_ISL_1689214, EPI_ISL_1689389, EPI_ISL_1689391, EPI_ISL_1689393, EPI_ISL_1689396, EPI_ISL_1689397, EPI_ISL_1689596, EPI_ISL_1689599, EPI_ISL_1689603, EPI_ISL_1689608, EPI_ISL_1689612, EPI_ISL_1689629, EPI_ISL_1689631, EPI_ISL_1689732, EPI_ISL_1689750, EPI_ISL_1689816, EPI_ISL_1689843, EPI_ISL_1689847, EPI_ISL_1689848, EPI_ISL_1689853, EPI_ISL_1689862, EPI_ISL_1689864, EPI_ISL_1689871, EPI_ISL_1689883, EPI_ISL_1689977, EPI_ISL_1690008, EPI_ISL_1690013, EPI_ISL_1690021, EPI_ISL_1690039, EPI_ISL_1690040, EPI_ISL_1690052, EPI_ISL_1690079, EPI_ISL_1690081, EPI_ISL_1690087, EPI_ISL_1690099, EPI_ISL_1690100, EPI_ISL_1690101, EPI_ISL_1690115, EPI_ISL_1690121, EPI_ISL_1690189, EPI_ISL_1690197, EPI_ISL_1690203, EPI_ISL_1690204, EPI_ISL_1690250, EPI_ISL_1690267, EPI_ISL_1690268 | | | |
| see above | Aegis Sciences Corporation | Centers for Disease Control and Prevention Division of Viral Diseases, Pathogen Discovery | Dakota Howard, Dhwani Batra, Peter W. Cook, Kara Moser, Adrian Paskey, Jason Caravas, Benjamin Rambo-Martin, Shatavia Morrison, Christopher Gulvick, Scott Sammons, Yvette Unoarumhi, Darlene Wagner, Matthew Schmerer, Cyndi Clark, Patrick Campbell, Rob Case, Vikramsinha Ghorpade, Holly Houdeshell, Ola Kvalvaag, Dillon Nall, Ethan Sanders, Alec Vest, Shaun Westlund, Matthew Hardison, Clinton R. Paden, Duncan MacCannell |
| EPI_ISL_1690309, EPI_ISL_1690352, EPI_ISL_1690546, EPI_ISL_1690890, EPI_ISL_1691214, EPI_ISL_1691266, EPI_ISL_1691275 | Helix/Illumina | Centers for Disease Control and Prevention Division of Viral Diseases, Pathogen Discovery | Dakota Howard, Dhwani Batra, Peter W. Cook, Kara Moser, Adrian Paskey, Jason Caravas, Benjamin Rambo-Martin, Shatavia Morrison, Christopher Gulvick, Scott Sammons, Yvette Unoarumhi, Darlene Wagner, Matthew Schmerer, Eileen de Feo, Jan Antico, Christine Tran, Matthew Tolentino, Shannon Wickline, Kim Gietzen, Brad Sickler, Jingtao Liu, Eric Allen, Phil Febbo, Nicole L. Washington, Simon White, Geraint Levan, Kelly Schiabor Barrett, Elizabeth Cirulli, Alexandre Bolze, Ary Ascencio, Charlotte Rivera-Garcia, Ryan Cho, Jason Nguyen, Sherry Wang, Jimmy Ramirez, Tyler Cassens, Efren Sandoval, Magnus Isaksson, William Lee, David Becker, Marc Laurent, James Lu, Clinton R. Paden, Duncan MacCannell |
| EPI_ISL_1691410, EPI_ISL_1691436, EPI_ISL_1691492 | Quest Diagnostics Incorporated | Centers for Disease Control and Prevention Division of Viral Diseases, Pathogen Discovery | Dakota Howard, Dhwani Batra, Peter W. Cook, Kara Moser, Adrian Paskey, Jason Caravas, Benjamin Rambo-Martin, Shatavia Morrison, Christopher Gulvick, Scott Sammons, Yvette Unoarumhi, Darlene Wagner, Matthew Schmerer, S. H. Rosenthal, A. Gerasimova, R. M. Kagan, B. Anderson, M. Hua, Y. Liu, L.E. Bernstein, K.E. Livingston, A. Perez, I. A. Shlyakhter, R. V. Rolando, R. Owen, P. Tanpaiboon, F. Lacbawan, Clinton R. Paden, Duncan MacCannell |
| EPI_ISL_1691557, EPI_ISL_1691562, EPI_ISL_1691674, EPI_ISL_1691686, EPI_ISL_1691716, EPI_ISL_1691720, EPI_ISL_1691721, EPI_ISL_1691756, EPI_ISL_1691959, EPI_ISL_1691975, EPI_ISL_1691986, EPI_ISL_1691995, EPI_ISL_1692005, EPI_ISL_1692010, EPI_ISL_1692028, EPI_ISL_1692030, EPI_ISL_1692084, EPI_ISL_1692129, EPI_ISL_1692160, EPI_ISL_1692198, EPI_ISL_1692224, EPI_ISL_1692266, EPI_ISL_1692284, EPI_ISL_1692285, EPI_ISL_1692292, EPI_ISL_1692310, EPI_ISL_1692327, EPI_ISL_1692341, EPI_ISL_1692371, EPI_ISL_1692408, EPI_ISL_1692409, EPI_ISL_1692448, EPI_ISL_1692460, EPI_ISL_1692516, EPI_ISL_1692524, EPI_ISL_1692575, EPI_ISL_1692578, EPI_ISL_1692581, EPI_ISL_1692587, EPI_ISL_1692592, EPI_ISL_1692600, EPI_ISL_1692606, EPI_ISL_1692634, EPI_ISL_1692655, EPI_ISL_1692658, EPI_ISL_1692669, EPI_ISL_1692677, EPI_ISL_1692687, EPI_ISL_1692705, EPI_ISL_1692708, EPI_ISL_1693102, EPI_ISL_1693220, EPI_ISL_1693274, EPI_ISL_1693317, EPI_ISL_1693319, EPI_ISL_1693320, EPI_ISL_1693321, EPI_ISL_1693868, EPI_ISL_1694015, EPI_ISL_1694028, EPI_ISL_1694242, EPI_ISL_1694388, EPI_ISL_1694519 | | | |
| see above | Infinity Biologix | Centers for Disease Control and Prevention Division of Viral Diseases, Pathogen Discovery | Dakota Howard, Dhwani Batra, Peter W. Cook, Kara Moser, Adrian Paskey, Jason Caravas, Benjamin Rambo-Martin, Shatavia Morrison, Christopher Gulvick, Scott Sammons, Yvette Unoarumhi, Darlene Wagner, Matthew Schmerer, Christian Bixby, Yihe Wang, Jonathan Schultz, Chirayu Goswami, Russ Hager, Robin Grimwood, Clinton R. Paden, Duncan MacCannell |
| EPI_ISL_1694671, EPI_ISL_1694672, EPI_ISL_1694673 | Quest Diagnostics Incorporated | Centers for Disease Control and Prevention Division of Viral Diseases, Pathogen Discovery | Dakota Howard, Dhwani Batra, Peter W. Cook, Kara Moser, Adrian Paskey, Jason Caravas, Benjamin Rambo-Martin, Shatavia Morrison, Christopher Gulvick, Scott Sammons, Yvette Unoarumhi, Darlene Wagner, Matthew Schmerer, S. H. Rosenthal, A. Gerasimova, R. M. Kagan, B. Anderson, M. Hua, Y. Liu, L.E. Bernstein, K.E. Livingston, A. Perez, I. A. Shlyakhter, R. V. Rolando, R. Owen, P. Tanpaiboon, F. Lacbawan, Clinton R. Paden, Duncan MacCannell |
| EPI_ISL_1694696 | DOHMH Riverside | New York City Public Health Laboratory | Jade Wang, et al. |
| EPI_ISL_1694697, EPI_ISL_1694698 | DOHMH Chelsea | New York City Public Health Laboratory | Jade Wang, et al. |
| EPI_ISL_1694699 | DOHMH Jamaica | New York City Public Health Laboratory | Jade Wang, et al. |
| EPI_ISL_1694700, EPI_ISL_1694701 | DOHMH Riverside | New York City Public Health Laboratory | Jade Wang, et al. |
| EPI_ISL_1694702, EPI_ISL_1694703 | DOHMH Central Harlem | New York City Public Health Laboratory | Jade Wang, et al. |
| EPI_ISL_1694704, EPI_ISL_1694705 | DOHMH Morrisania | New York City Public Health Laboratory | Jade Wang, et al. |
| EPI_ISL_1694706 | DOHMH Chelsea | New York City Public Health Laboratory | Jade Wang, et al. |
| EPI_ISL_1694707 | DOHMH Corona | New York City Public Health Laboratory | Jade Wang, et al. |
| EPI_ISL_1694708 | DOHMH Jamaica | New York City Public Health Laboratory | Jade Wang, et al. |
| EPI_ISL_1694709 | DOHMH Central Harlem | New York City Public Health Laboratory | Jade Wang, et al. |
| EPI_ISL_1694710 | DOHMH Corona | New York City Public Health Laboratory | Jade Wang, et al. |
| EPI_ISL_1694711 | DOHMH Crown Heights | New York City Public Health Laboratory | Jade Wang, et al. |
| EPI_ISL_1694712 | DOHMH Jamaica | New York City Public Health Laboratory | Jade Wang, et al. |
| EPI_ISL_1694713, EPI_ISL_1694714 | DOHMH Corona | New York City Public Health Laboratory | Jade Wang, et al. |
| EPI_ISL_1694715, EPI_ISL_1694716 | DOHMH PHL | New York City Public Health Laboratory | Jade Wang, et al. |
| EPI_ISL_1694717 | DOHMH Morrisania | New York City Public Health Laboratory | Jade Wang, et al. |
| EPI_ISL_1694718 | DOHMH Central Harlem | New York City Public Health Laboratory | Jade Wang, et al. |
| EPI_ISL_1694719 | DOHMH Fort Greene | New York City Public Health Laboratory | Jade Wang, et al. |
| EPI_ISL_1694720 | Poplar Healthcare | New York City Public Health Laboratory | Jade Wang, et al. |
| EPI_ISL_1694721, EPI_ISL_1694722, EPI_ISL_1694723, EPI_ISL_1694724, EPI_ISL_1694725, EPI_ISL_1694726, EPI_ISL_1694727 | OCME Office Of Chief Medical Examiner | New York City Public Health Laboratory | Jade Wang, et al. |
| EPI_ISL_1694728 | DOHMH Crown Heights | New York City Public Health Laboratory | Jade Wang, et al. |
| EPI_ISL_1694729 | DOHMH Morrisania | New York City Public Health Laboratory | Jade Wang, et al. |
| EPI_ISL_1694730 | DOHMH Chelsea | New York City Public Health Laboratory | Jade Wang, et al. |
| EPI_ISL_1694731, EPI_ISL_1694732 | OCME Office Of Chief Medical Examiner | New York City Public Health Laboratory | Jade Wang, et al. |
| EPI_ISL_1694733 | DOHMH Fort Greene | New York City Public Health Laboratory | Jade Wang, et al. |
| EPI_ISL_1694734, EPI_ISL_1694735 | DOHMH Corona | New York City Public Health Laboratory | Jade Wang, et al. |
| EPI_ISL_1694736, EPI_ISL_1694737, EPI_ISL_1694738, EPI_ISL_1694739, EPI_ISL_1694740 | DOHMH Jamaica | New York City Public Health Laboratory | Jade Wang, et al. |
| EPI_ISL_1694741, EPI_ISL_1694742, EPI_ISL_1694743, EPI_ISL_1694744, EPI_ISL_1694745 | DOHMH PHL | New York City Public Health Laboratory | Jade Wang, et al. |
| EPI_ISL_1694746, EPI_ISL_1694747 | DOHMH Fort Greene | New York City Public Health Laboratory | Jade Wang, et al. |

| | | | |
|---|---|---|---|
| EPI_ISL_1694748, EPI_ISL_1694749 | DOHMH PHL | New York City Public Health Laboratory | Jade Wang, et al. |
| EPI_ISL_1694750, EPI_ISL_1694751, EPI_ISL_1694752, EPI_ISL_1694753 | DOHMH Corona | New York City Public Health Laboratory | Jade Wang, et al. |
| EPI_ISL_1694754, EPI_ISL_1694755, EPI_ISL_1694756, EPI_ISL_1694757, EPI_ISL_1694758 | DOHMH Jamaica | New York City Public Health Laboratory | Jade Wang, et al. |
| EPI_ISL_1694759 | DOHMH Central Harlem | New York City Public Health Laboratory | Jade Wang, et al. |
| EPI_ISL_1694760, EPI_ISL_1694761, EPI_ISL_1694762 | Department of Homeless Services | New York City Public Health Laboratory | Jade Wang, et al. |
| EPI_ISL_1694763, EPI_ISL_1694764, EPI_ISL_1694765, EPI_ISL_1694766, EPI_ISL_1694767, EPI_ISL_1694768, EPI_ISL_1694769, EPI_ISL_1694770, EPI_ISL_1694771, EPI_ISL_1694772 | OCME Office Of Chief Medical Examiner | New York City Public Health Laboratory | Jade Wang, et al. |
| EPI_ISL_1694773, EPI_ISL_1694774, EPI_ISL_1694775 | DOHMH Central Harlem | New York City Public Health Laboratory | Jade Wang, et al. |
| EPI_ISL_1694776 | DOHMH Morrisania | New York City Public Health Laboratory | Jade Wang, et al. |
| EPI_ISL_1695014, EPI_ISL_1695015 | Laboratory Corporation of America | Centers for Disease Control and Prevention Division of Viral Diseases, Pathogen Discovery | Dakota Howard, Dhwani Batra, Peter W. Cook, Kara Moser, Adrian Paskey, Jason Caravas, Benjamin Rambo-Martin, Shatavia Morrison, Christopher Gulvick, Scott Sammons, Yvette Unoarumhi, Darlene Wagner, Matthew Schmerer, Minoo Agarwal, Eyad Almasri, Debbie Boles, Ayla Burns, Nuthawin Charoensri, Oren Cohen, Susan Countryman, Mary Ann Cristobal, Bobbi Croy, Suzanne Dale, Hrushikesh Deshmukh, Amanda Douglas, Vincent Drouillon, Marcia Eisenberg, Howard Engler, Rama Ghatti, Prashant Gupta, Susan Hicks, Jake Humphrey, Lax Iyer, Manoj Jain, Mohan Kolli, Brian Krueger, Tim Kuphal, Stanley Letovsky, Michael Levandoski, Craig Lukasik, Jonathan Meltzer, Brian Norvell, Mindy Nye, Scott Parker, Christos Petropoulos, John Pruitt, Steven Ragan, Scott Ryan, Mike Sapeta, Jana Schroth, Suresh Babu Selvaraju, Goran Stevovic, Amanda Suchanek, Andrea Throop, Lyndon Tilson, Thomas Urban, Joe Voshell, Kimberly Wagner, Jonathan Williams, Mary Williamson, Qian Zeng, Tricia Zwiefelhofer, Clinton R. Paden, Duncan MacCannell |
| EPI_ISL_1695037, EPI_ISL_1695051, EPI_ISL_1695067 | Quest Diagnostics Incorporated | Centers for Disease Control and Prevention Division of Viral Diseases, Pathogen Discovery | Dakota Howard, Dhwani Batra, Peter W. Cook, Kara Moser, Adrian Paskey, Jason Caravas, Benjamin Rambo-Martin, Shatavia Morrison, Christopher Gulvick, Scott Sammons, Yvette Unoarumhi, Darlene Wagner, Matthew Schmerer, S. H. Rosenthal, A. Gerasimova, R. M. Kagan, B. Anderson, M. Hua, Y. Liu, L.E. Bernstein, K.E. Livingston, A. Perez, I. A. Shlyakhter, R. V. Rolando, R. Owen, P. Tanpaiboon, F. Lacbawan, Clinton R. Paden, Duncan MacCannell |
| EPI_ISL_1695135, EPI_ISL_1695305, EPI_ISL_1695382, EPI_ISL_1695469, EPI_ISL_1695487, EPI_ISL_1695545, EPI_ISL_1695554, EPI_ISL_1695578 | Aegis Sciences Corporation | Centers for Disease Control and Prevention Division of Viral Diseases, Pathogen Discovery | Dakota Howard, Dhwani Batra, Peter W. Cook, Kara Moser, Adrian Paskey, Jason Caravas, Benjamin Rambo-Martin, Shatavia Morrison, Christopher Gulvick, Scott Sammons, Yvette Unoarumhi, Darlene Wagner, Matthew Schmerer, Cyndi Clark, Patrick Campbell, Rob Case, Vikramsinha Ghorpade, Holly Houdeshell, Ola Kvalvaag, Dillon Nall, Ethan Sanders, Alec Vest, Shaun Westlund, Matthew Hardison, Clinton R. Paden, Duncan MacCannell |
| EPI_ISL_1695681 | Quest Diagnostics Incorporated | Centers for Disease Control and Prevention Division of Viral Diseases, Pathogen Discovery | Dakota Howard, Dhwani Batra, Peter W. Cook, Kara Moser, Adrian Paskey, Jason Caravas, Benjamin Rambo-Martin, Shatavia Morrison, Christopher Gulvick, Scott Sammons, Yvette Unoarumhi, Darlene Wagner, Matthew Schmerer, S. H. Rosenthal, A. Gerasimova, R. M. Kagan, B. Anderson, M. Hua, Y. Liu, L.E. Bernstein, K.E. Livingston, A. Perez, I. A. Shlyakhter, R. V. Rolando, R. Owen, P. Tanpaiboon, F. Lacbawan, Clinton R. Paden, Duncan MacCannell |
| EPI_ISL_1695889, EPI_ISL_1695890, EPI_ISL_1695891, EPI_ISL_1695892 | NYSDOH Wadsworth Center, Virology Lab | Centers for Disease Control and Prevention Division of Viral Diseases, Pathogen Discovery | Mili Sheth, Sarah Nobles, Jasmine Padilla, Mark Burroughs, Shoshona Le, Katie Dillon, Peter Cook, Clinton R. Paden, Dhwani Batra, Krista Queen, Kristen Knipe, Dakota Howard, Yvette Unoarumhi, Darlene Wagner, Matthew Schmerer, Ben L. Rambo-Martin, Kristine Lacek, Sam Shepard, Alison Laufer Halpin, Dave Wentworth, Vivien Dugan, Suxiang Tong, Justin Lee |
| EPI_ISL_1695970 | NYC Department of Health and Mental Hygiene | Centers for Disease Control and Prevention Division of Viral Diseases, Pathogen Discovery | Mili Sheth, Sarah Nobles, Jasmine Padilla, Mark Burroughs, Shoshona Le, Katie Dillon, Peter Cook, Clinton R. Paden, Dhwani Batra, Krista Queen, Kristen Knipe, Dakota Howard, Yvette Unoarumhi, Darlene Wagner, Matthew Schmerer, Ben L. Rambo-Martin, Kristine Lacek, Sam Shepard, Alison Laufer Halpin, Dave Wentworth, Vivien Dugan, Suxiang Tong, Justin Lee |
| EPI_ISL_1700669, EPI_ISL_1700670, EPI_ISL_1700671, EPI_ISL_1700672, EPI_ISL_1700673 | NYC Department of Health and Mental Hygiene | Pathogen Discovery, Respiratory Viruses Branch, Division of Viral Diseases, Centers for Disease Control and Prevention | Ying Tao, Yan Li, Anna Kelleher, Jing Zhang, Anna Montmayeur, Brian Lynch, Krista Queen, Anna Uehara, Peter Cook, Han Jia Justin Ng, Rachel Marine, Clinton R. Paden, Haibin Wang, Mark Burroughs, Justin Lee, Adam Retchless, Suxiang Tong |
| EPI_ISL_1701890, EPI_ISL_1701900, EPI_ISL_1701941, EPI_ISL_1701968, EPI_ISL_1701969, EPI_ISL_1701973, EPI_ISL_1701980, EPI_ISL_1701996, EPI_ISL_1701997, EPI_ISL_1702014, EPI_ISL_1702030, EPI_ISL_1702035, EPI_ISL_1702076, EPI_ISL_1702107, EPI_ISL_1702127, EPI_ISL_1702138 | | | |
| see above | Aegis Sciences Corporation | Centers for Disease Control and Prevention Division of Viral Diseases, Pathogen Discovery | Dakota Howard, Dhwani Batra, Peter W. Cook, Kara Moser, Adrian Paskey, Jason Caravas, Benjamin Rambo-Martin, Shatavia Morrison, Christopher Gulvick, Scott Sammons, Yvette Unoarumhi, Darlene Wagner, Matthew Schmerer, Cyndi Clark, Patrick Campbell, Rob Case, Vikramsinha Ghorpade, Holly Houdeshell, Ola Kvalvaag, Dillon Nall, Ethan Sanders, Alec Vest, Shaun Westlund, Matthew Hardison, Clinton R. Paden, Duncan MacCannell |
| EPI_ISL_1702185, EPI_ISL_1702186, EPI_ISL_1702187, EPI_ISL_1702255, EPI_ISL_1702267, EPI_ISL_1702281 | Infinity Biologix | Centers for Disease Control and Prevention Division of Viral Diseases, Pathogen Discovery | Dakota Howard, Dhwani Batra, Peter W. Cook, Kara Moser, Adrian Paskey, Jason Caravas, Benjamin Rambo-Martin, Shatavia Morrison, Christopher Gulvick, Scott Sammons, Yvette Unoarumhi, Darlene Wagner, Matthew Schmerer, Christian Bixby, Yihe Wang, Jonathan Schultz, Chirayu Goswami, Russ Hager, Robin Grimwood, Clinton R. Paden, Duncan MacCannell |
| EPI_ISL_1702639 | Quest Diagnostics Incorporated | Centers for Disease Control and Prevention Division of Viral Diseases, Pathogen Discovery | Dakota Howard, Dhwani Batra, Peter W. Cook, Kara Moser, Adrian Paskey, Jason Caravas, Benjamin Rambo-Martin, Shatavia Morrison, Christopher Gulvick, Scott Sammons, Yvette Unoarumhi, Darlene Wagner, Matthew Schmerer, S. H. Rosenthal, A. Gerasimova, R. M. Kagan, B. Anderson, M. Hua, Y. Liu, L.E. Bernstein, K.E. Livingston, A. Perez, I. A. Shlyakhter, R. V. Rolando, R. Owen, P. Tanpaiboon, F. Lacbawan, Clinton R. Paden, Duncan MacCannell |
| EPI_ISL_1702728, EPI_ISL_1702778, EPI_ISL_1702788, EPI_ISL_1702789, EPI_ISL_1702816, EPI_ISL_1702829, EPI_ISL_1702838, EPI_ISL_1702865, EPI_ISL_1702867, EPI_ISL_1702884, EPI_ISL_1702954, EPI_ISL_1702970 | | | |
| see above | Aegis Sciences Corporation | Centers for Disease Control and Prevention Division of Viral Diseases, Pathogen Discovery | Dakota Howard, Dhwani Batra, Peter W. Cook, Kara Moser, Adrian Paskey, Jason Caravas, Benjamin Rambo-Martin, Shatavia Morrison, Christopher Gulvick, Scott Sammons, Yvette Unoarumhi, Darlene Wagner, Matthew Schmerer, Cyndi Clark, Patrick Campbell, Rob Case, Vikramsinha Ghorpade, Holly Houdeshell, Ola Kvalvaag, Dillon Nall, Ethan Sanders, Alec Vest, Shaun Westlund, Matthew Hardison, Clinton R. Paden, Duncan MacCannell |
| EPI_ISL_1704714 | Northwestern Memorial Hospital | Northwestern University - Ozer Lab | Ramon Lorenzo-Redondo, Lacy M. Simons, Taylor J. Dean, Chad J. Achenbach, Lawrence J. Jennings, Chao Qi, Michael G. Ison, Judd F. Hultquist, Egon A. Ozer |
| EPI_ISL_1708745, EPI_ISL_1708747, EPI_ISL_1708748, EPI_ISL_1708750, EPI_ISL_1708751, EPI_ISL_1708752, EPI_ISL_1708753, EPI_ISL_1708755, EPI_ISL_1708756, EPI_ISL_1708757, EPI_ISL_1708758, EPI_ISL_1708759, EPI_ISL_1708760, EPI_ISL_1708761, EPI_ISL_1708762, EPI_ISL_1708763, EPI_ISL_1708764, EPI_ISL_1708765, EPI_ISL_1708766, EPI_ISL_1708767, EPI_ISL_1708768, EPI_ISL_1708769, EPI_ISL_1708770, EPI_ISL_1708771, EPI_ISL_1708772, EPI_ISL_1708773, EPI_ISL_1708774, EPI_ISL_1708775, EPI_ISL_1708776, EPI_ISL_1708777, EPI_ISL_1708778, EPI_ISL_1708779, EPI_ISL_1708781, EPI_ISL_1708782, EPI_ISL_1708783, EPI_ISL_1708784, EPI_ISL_1708785, EPI_ISL_1708786, EPI_ISL_1708787, EPI_ISL_1708788, EPI_ISL_1708789, EPI_ISL_1708790, EPI_ISL_1708791, EPI_ISL_1708792, EPI_ISL_1708794, EPI_ISL_1708795, EPI_ISL_1708796, EPI_ISL_1708797, EPI_ISL_1708798, EPI_ISL_1708799, EPI_ISL_1708800, EPI_ISL_1708801, EPI_ISL_1708802, EPI_ISL_1708803, EPI_ISL_1708804, EPI_ISL_1708805, EPI_ISL_1708806, EPI_ISL_1708807, EPI_ISL_1708808, EPI_ISL_1708809, EPI_ISL_1708810, EPI_ISL_1708811, EPI_ISL_1708812, EPI_ISL_1708813, EPI_ISL_1708815, EPI_ISL_1708816, EPI_ISL_1708817, EPI_ISL_1708818, EPI_ISL_1708819, EPI_ISL_1708820, EPI_ISL_1708821, EPI_ISL_1708822, EPI_ISL_1708823, EPI_ISL_1708824, EPI_ISL_1708825, EPI_ISL_1708826, EPI_ISL_1708827, EPI_ISL_1708828, EPI_ISL_1708829, EPI_ISL_1708830, EPI_ISL_1708831, EPI_ISL_1708832, EPI_ISL_1708833, EPI_ISL_1708834, EPI_ISL_1708835, EPI_ISL_1708836, EPI_ISL_1708837, EPI_ISL_1708838, EPI_ISL_1708839, EPI_ISL_1708841, EPI_ISL_1708842, EPI_ISL_1708843, EPI_ISL_1708844, EPI_ISL_1708845, EPI_ISL_1708846, EPI_ISL_1708847, EPI_ISL_1708848, EPI_ISL_1708849, EPI_ISL_1708850, EPI_ISL_1708851, EPI_ISL_1708852, EPI_ISL_1708854, EPI_ISL_1708855, EPI_ISL_1708856, EPI_ISL_1708857, EPI_ISL_1708858, EPI_ISL_1708859, EPI_ISL_1708860, EPI_ISL_1708861, EPI_ISL_1708862, EPI_ISL_1708863, EPI_ISL_1708864, EPI_ISL_1708865, EPI_ISL_1708866, EPI_ISL_1708867, EPI_ISL_1708868, EPI_ISL_1708869, EPI_ISL_1708870, EPI_ISL_1708871, EPI_ISL_1708872, EPI_ISL_1708873, EPI_ISL_1708874, EPI_ISL_1708876, EPI_ISL_1708877, EPI_ISL_1708878, EPI_ISL_1708879, EPI_ISL_1708880, EPI_ISL_1708881, EPI_ISL_1708882, EPI_ISL_1708883, EPI_ISL_1708884, EPI_ISL_1708885, EPI_ISL_1708886, EPI_ISL_1708887, EPI_ISL_1708888, EPI_ISL_1708889, EPI_ISL_1708890, EPI_ISL_1708891, EPI_ISL_1708892, EPI_ISL_1708893, EPI_ISL_1708894, EPI_ISL_1708895, EPI_ISL_1708896, EPI_ISL_1708897, EPI_ISL_1708898, EPI_ISL_1708899, EPI_ISL_1708900, EPI_ISL_1708901, EPI_ISL_1708902, EPI_ISL_1708903, EPI_ISL_1708904, EPI_ISL_1708905, EPI_ISL_1708906, EPI_ISL_1708907, EPI_ISL_1708908, EPI_ISL_1708909, EPI_ISL_1708910, EPI_ISL_1708911, EPI_ISL_1708912, EPI_ISL_1708913, EPI_ISL_1708914, EPI_ISL_1708915, EPI_ISL_1708916, EPI_ISL_1708917, EPI_ISL_1708918, EPI_ISL_1708919, EPI_ISL_1708920, EPI_ISL_1708921, EPI_ISL_1708922, EPI_ISL_1708923, EPI_ISL_1708924, EPI_ISL_1708925, EPI_ISL_1708926, EPI_ISL_1708928, EPI_ISL_1708929, EPI_ISL_1708930, EPI_ISL_1708931, EPI_ISL_1708932, EPI_ISL_1708933, EPI_ISL_1708934, EPI_ISL_1708935, EPI_ISL_1708936, EPI_ISL_1708937, EPI_ISL_1708938, EPI_ISL_1708939, EPI_ISL_1708940, EPI_ISL_1708941, EPI_ISL_1708942, EPI_ISL_1708943, EPI_ISL_1708944, EPI_ISL_1708945, EPI_ISL_1708946, EPI_ISL_1708947, EPI_ISL_1708948, EPI_ISL_1708949, EPI_ISL_1708950, EPI_ISL_1708951, EPI_ISL_1708952, EPI_ISL_1708953, EPI_ISL_1708954, EPI_ISL_1708955, EPI_ISL_1708956, EPI_ISL_1708957, EPI_ISL_1708958, EPI_ISL_1708959, EPI_ISL_1708960, EPI_ISL_1708961, EPI_ISL_1708962, EPI_ISL_1708963, EPI_ISL_1708964, EPI_ISL_1708965, EPI_ISL_1708966, EPI_ISL_1708967, EPI_ISL_1708968, EPI_ISL_1708969, EPI_ISL_1708970, EPI_ISL_1708971, EPI_ISL_1708972, EPI_ISL_1708973, EPI_ISL_1708974, EPI_ISL_1708975, EPI_ISL_1708976, EPI_ISL_1708977, EPI_ISL_1708978, EPI_ISL_1708979, EPI_ISL_1708980, EPI_ISL_1708981, EPI_ISL_1708982, EPI_ISL_1708983, EPI_ISL_1708984, EPI_ISL_1708985, EPI_ISL_1708986, EPI_ISL_1708987, EPI_ISL_1708988, EPI_ISL_1708989, EPI_ISL_1708990, EPI_ISL_1708991, EPI_ISL_1708992, | | | |

EPI_ISL_1708993, EPI_ISL_1708994, EPI_ISL_1708995, EPI_ISL_1708996, EPI_ISL_1708997, EPI_ISL_1708998, EPI_ISL_1708999, EPI_ISL_1709000, EPI_ISL_1709001, EPI_ISL_1709002, EPI_ISL_1709003, EPI_ISL_1709004, EPI_ISL_1709005, EPI_ISL_1709006, EPI_ISL_1709007, EPI_ISL_1709008, EPI_ISL_1709009, EPI_ISL_1709010, EPI_ISL_1709011, EPI_ISL_1709012, EPI_ISL_1709013, EPI_ISL_1709014, EPI_ISL_1709015, EPI_ISL_1709016, EPI_ISL_1709017, EPI_ISL_1709018, EPI_ISL_1709019, EPI_ISL_1709020, EPI_ISL_1709021, EPI_ISL_1709022, EPI_ISL_1709023, EPI_ISL_1709024, EPI_ISL_1709025, EPI_ISL_1709026, EPI_ISL_1709027, EPI_ISL_1709028, EPI_ISL_1709029, EPI_ISL_1709030, EPI_ISL_1709031, EPI_ISL_1709032, EPI_ISL_1709033, EPI_ISL_1709034, EPI_ISL_1709035, EPI_ISL_1709036, EPI_ISL_1709037, EPI_ISL_1709038, EPI_ISL_1709039, EPI_ISL_1709040, EPI_ISL_1709041, EPI_ISL_1709042, EPI_ISL_1709043, EPI_ISL_1709044, EPI_ISL_1709045, EPI_ISL_1709046, EPI_ISL_1709048, EPI_ISL_1709049, EPI_ISL_1709050, EPI_ISL_1709051, EPI_ISL_1709052, EPI_ISL_1709053, EPI_ISL_1709054, EPI_ISL_1709055, EPI_ISL_1709056, EPI_ISL_1709057, EPI_ISL_1709058, EPI_ISL_1709059, EPI_ISL_1709060, EPI_ISL_1709061, EPI_ISL_1709062, EPI_ISL_1709063, EPI_ISL_1709064, EPI_ISL_1709065, EPI_ISL_1709066, EPI_ISL_1709067, EPI_ISL_1709068, EPI_ISL_1709069, EPI_ISL_1709070, EPI_ISL_1709071, EPI_ISL_1709072, EPI_ISL_1709073, EPI_ISL_1709074, EPI_ISL_1709075, EPI_ISL_1709076, EPI_ISL_1709077, EPI_ISL_1709078, EPI_ISL_1709079, EPI_ISL_1709081, EPI_ISL_1709082, EPI_ISL_1709083, EPI_ISL_1709084, EPI_ISL_1709085, EPI_ISL_1709086, EPI_ISL_1709087, EPI_ISL_1709088, EPI_ISL_1709089, EPI_ISL_1709090, EPI_ISL_1709091, EPI_ISL_1709092, EPI_ISL_1709093, EPI_ISL_1709094, EPI_ISL_1709095, EPI_ISL_1709096, EPI_ISL_1709097, EPI_ISL_1709098, EPI_ISL_1709099, EPI_ISL_1709100, EPI_ISL_1709101, EPI_ISL_1709102, EPI_ISL_1709103, EPI_ISL_1709104, EPI_ISL_1709105, EPI_ISL_1709106, EPI_ISL_1709107, EPI_ISL_1709108, EPI_ISL_1709109, EPI_ISL_1709110, EPI_ISL_1709111, EPI_ISL_1709112, EPI_ISL_1709113, EPI_ISL_1709114, EPI_ISL_1709115, EPI_ISL_1709116, EPI_ISL_1709117, EPI_ISL_1709118, EPI_ISL_1709119, EPI_ISL_1709120, EPI_ISL_1709121, EPI_ISL_1709122, EPI_ISL_1709123, EPI_ISL_1709124, EPI_ISL_1709125, EPI_ISL_1709126, EPI_ISL_1709127, EPI_ISL_1709128, EPI_ISL_1709129, EPI_ISL_1709130, EPI_ISL_1709131, EPI_ISL_1709132, EPI_ISL_1709133, EPI_ISL_1709134, EPI_ISL_1709135, EPI_ISL_1709136, EPI_ISL_1709137, EPI_ISL_1709138, EPI_ISL_1709139, EPI_ISL_1709140, EPI_ISL_1709141, EPI_ISL_1709142, EPI_ISL_1709143, EPI_ISL_1709144, EPI_ISL_1709145, EPI_ISL_1709146, EPI_ISL_1709147, EPI_ISL_1709148, EPI_ISL_1709149, EPI_ISL_1709150, EPI_ISL_1709151, EPI_ISL_1709152, EPI_ISL_1709153, EPI_ISL_1709154, EPI_ISL_1709155, EPI_ISL_1709156, EPI_ISL_1709157, EPI_ISL_1709158, EPI_ISL_1709159, EPI_ISL_1709160, EPI_ISL_1709161, EPI_ISL_1709162, EPI_ISL_1709163, EPI_ISL_1709164, EPI_ISL_1709165, EPI_ISL_1709166, EPI_ISL_1709167, EPI_ISL_1709168, EPI_ISL_1709169, EPI_ISL_1709170, EPI_ISL_1709171, EPI_ISL_1709172, EPI_ISL_1709173, EPI_ISL_1709174, EPI_ISL_1709175, EPI_ISL_1709176, EPI_ISL_1709177, EPI_ISL_1709178, EPI_ISL_1709179, EPI_ISL_1709180, EPI_ISL_1709181, EPI_ISL_1709182, EPI_ISL_1709183, EPI_ISL_1709184, EPI_ISL_1709185, EPI_ISL_1709186, EPI_ISL_1709187, EPI_ISL_1709188, EPI_ISL_1709189, EPI_ISL_1709190, EPI_ISL_1709191, EPI_ISL_1709192, EPI_ISL_1709193, EPI_ISL_1709194, EPI_ISL_1709195, EPI_ISL_1709196, EPI_ISL_1709197, EPI_ISL_1709198, EPI_ISL_1709199, EPI_ISL_1709200, EPI_ISL_1709201, EPI_ISL_1709202, EPI_ISL_1709203, EPI_ISL_1709204, EPI_ISL_1709205, EPI_ISL_1709206, EPI_ISL_1709207, EPI_ISL_1709208, EPI_ISL_1709209, EPI_ISL_1709210, EPI_ISL_1709211, EPI_ISL_1709212, EPI_ISL_1709213, EPI_ISL_1709214, EPI_ISL_1709215, EPI_ISL_1709216, EPI_ISL_1709217, EPI_ISL_1709218, EPI_ISL_1709219, EPI_ISL_1709220, EPI_ISL_1709221, EPI_ISL_1709222, EPI_ISL_1709223, EPI_ISL_1709224, EPI_ISL_1709225, EPI_ISL_1709226, EPI_ISL_1709227, EPI_ISL_1709228, EPI_ISL_1709229, EPI_ISL_1709230, EPI_ISL_1709231, EPI_ISL_1709232, EPI_ISL_1709233, EPI_ISL_1709234, EPI_ISL_1709235, EPI_ISL_1709236, EPI_ISL_1709237, EPI_ISL_1709238, EPI_ISL_1709239, EPI_ISL_1709240, EPI_ISL_1709241, EPI_ISL_1709242, EPI_ISL_1709243, EPI_ISL_1709244, EPI_ISL_1709245, EPI_ISL_1709246, EPI_ISL_1709247, EPI_ISL_1709248, EPI_ISL_1709249, EPI_ISL_1709250, EPI_ISL_1709251, EPI_ISL_1709252, EPI_ISL_1709253, EPI_ISL_1709254, EPI_ISL_1709255, EPI_ISL_1709256, EPI_ISL_1709257, EPI_ISL_1709258, EPI_ISL_1709259, EPI_ISL_1709260, EPI_ISL_1709261, EPI_ISL_1709262, EPI_ISL_1709263, EPI_ISL_1709264, EPI_ISL_1709265, EPI_ISL_1709266, EPI_ISL_1709267, EPI_ISL_1709268, EPI_ISL_1709269, EPI_ISL_1709270, EPI_ISL_1709271, EPI_ISL_1709272, EPI_ISL_1709273, EPI_ISL_1709274, EPI_ISL_1709275, EPI_ISL_1709276, EPI_ISL_1709277, EPI_ISL_1709278, EPI_ISL_1709280, EPI_ISL_1709281, EPI_ISL_1709282, EPI_ISL_1709283, EPI_ISL_1709284, EPI_ISL_1709285, EPI_ISL_1709286, EPI_ISL_1709287, EPI_ISL_1709288, EPI_ISL_1709289, EPI_ISL_1709290, EPI_ISL_1709291, EPI_ISL_1709292, EPI_ISL_1709293, EPI_ISL_1709294, EPI_ISL_1709295, EPI_ISL_1709296, EPI_ISL_1709297, EPI_ISL_1709298, EPI_ISL_1709299, EPI_ISL_1709301, EPI_ISL_1709302, EPI_ISL_1709303, EPI_ISL_1709304, EPI_ISL_1709305, EPI_ISL_1709306, EPI_ISL_1709307, EPI_ISL_1709308, EPI_ISL_1709309, EPI_ISL_1709310, EPI_ISL_1709311, EPI_ISL_1709312, EPI_ISL_1709313, EPI_ISL_1709314, EPI_ISL_1709315, EPI_ISL_1709316, EPI_ISL_1709317, EPI_ISL_1709318, EPI_ISL_1709319, EPI_ISL_1709320, EPI_ISL_1709321, EPI_ISL_1709322, EPI_ISL_1709323, EPI_ISL_1709324, EPI_ISL_1709325, EPI_ISL_1709326, EPI_ISL_1709327, EPI_ISL_1709328, EPI_ISL_1709329, EPI_ISL_1709330, EPI_ISL_1709331, EPI_ISL_1709332, EPI_ISL_1709333, EPI_ISL_1709334, EPI_ISL_1709335, EPI_ISL_1709336, EPI_ISL_1709337, EPI_ISL_1709338, EPI_ISL_1709339, EPI_ISL_1709340, EPI_ISL_1709341, EPI_ISL_1709342, EPI_ISL_1709343, EPI_ISL_1709344, EPI_ISL_1709345, EPI_ISL_1709346, EPI_ISL_1709347, EPI_ISL_1709348, EPI_ISL_1709349, EPI_ISL_1709350, EPI_ISL_1709351, EPI_ISL_1709352, EPI_ISL_1709353, EPI_ISL_1709354, EPI_ISL_1709355, EPI_ISL_1709356, EPI_ISL_1709357, EPI_ISL_1709358, EPI_ISL_1709359, EPI_ISL_1709360, EPI_ISL_1709361, EPI_ISL_1709362, EPI_ISL_1709363, EPI_ISL_1709364, EPI_ISL_1709365, EPI_ISL_1709366, EPI_ISL_1709367, EPI_ISL_1709368, EPI_ISL_1709369, EPI_ISL_1709370, EPI_ISL_1709371, EPI_ISL_1709372, EPI_ISL_1709373, EPI_ISL_1709374, EPI_ISL_1709375, EPI_ISL_1709376, EPI_ISL_1709377, EPI_ISL_1709378, EPI_ISL_1709379, EPI_ISL_1709380, EPI_ISL_1709381, EPI_ISL_1709382, EPI_ISL_1709383, EPI_ISL_1709384, EPI_ISL_1709385, EPI_ISL_1709386, EPI_ISL_1709387, EPI_ISL_1709388, EPI_ISL_1709389, EPI_ISL_1709390, EPI_ISL_1709391, EPI_ISL_1709392, EPI_ISL_1709393, EPI_ISL_1709394, EPI_ISL_1709395, EPI_ISL_1709396, EPI_ISL_1709397, EPI_ISL_1709398, EPI_ISL_1709399, EPI_ISL_1709400, EPI_ISL_1709401, EPI_ISL_1709402, EPI_ISL_1709403, EPI_ISL_1709404, EPI_ISL_1709405, EPI_ISL_1709406, EPI_ISL_1709407, EPI_ISL_1709408, EPI_ISL_1709409, EPI_ISL_1709410, EPI_ISL_1709411, EPI_ISL_1709412, EPI_ISL_1709413, EPI_ISL_1709414, EPI_ISL_1709415, EPI_ISL_1709416, EPI_ISL_1709417, EPI_ISL_1709418, EPI_ISL_1709419, EPI_ISL_1709420, EPI_ISL_1709422, EPI_ISL_1709423, EPI_ISL_1709424, EPI_ISL_1709425, EPI_ISL_1709426, EPI_ISL_1709427, EPI_ISL_1709428, EPI_ISL_1709429, EPI_ISL_1709430, EPI_ISL_1709431, EPI_ISL_1709432, EPI_ISL_1709434, EPI_ISL_1709435, EPI_ISL_1709436, EPI_ISL_1709437, EPI_ISL_1709438, EPI_ISL_1709439, EPI_ISL_1709440, EPI_ISL_1709441, EPI_ISL_1709442, EPI_ISL_1709443, EPI_ISL_1709444, EPI_ISL_1709445, EPI_ISL_1709446, EPI_ISL_1709447, EPI_ISL_1709448, EPI_ISL_1709449, EPI_ISL_1709450, EPI_ISL_1709451, EPI_ISL_1709452, EPI_ISL_1709453, EPI_ISL_1709454, EPI_ISL_1709455, EPI_ISL_1709456, EPI_ISL_1709457, EPI_ISL_1709458, EPI_ISL_1709459, EPI_ISL_1709460, EPI_ISL_1709461, EPI_ISL_1709462, EPI_ISL_1709463, EPI_ISL_1709464, EPI_ISL_1709465, EPI_ISL_1709466, EPI_ISL_1709467, EPI_ISL_1709468, EPI_ISL_1709469, EPI_ISL_1709470, EPI_ISL_1709472, EPI_ISL_1709473, EPI_ISL_1709474, EPI_ISL_1709475, EPI_ISL_1709476, EPI_ISL_1709477, EPI_ISL_1709478, EPI_ISL_1709479, EPI_ISL_1709480, EPI_ISL_1709481, EPI_ISL_1709482, EPI_ISL_1709483, EPI_ISL_1709484, EPI_ISL_1709485, EPI_ISL_1709486, EPI_ISL_1709487, EPI_ISL_1709488, EPI_ISL_1709489, EPI_ISL_1709490, EPI_ISL_1709492, EPI_ISL_1709493, EPI_ISL_1709494, EPI_ISL_1709495, EPI_ISL_1709496, EPI_ISL_1709497, EPI_ISL_1709498, EPI_ISL_1709499, EPI_ISL_1709500, EPI_ISL_1709501, EPI_ISL_1709502, EPI_ISL_1709503, EPI_ISL_1709504, EPI_ISL_1709505, EPI_ISL_1709506, EPI_ISL_1709507, EPI_ISL_1709508, EPI_ISL_1709509, EPI_ISL_1709510, EPI_ISL_1709511, EPI_ISL_1709512, EPI_ISL_1709513, EPI_ISL_1709514, EPI_ISL_1709515, EPI_ISL_1709516, EPI_ISL_1709517, EPI_ISL_1709518, EPI_ISL_1709519, EPI_ISL_1709520, EPI_ISL_1709521, EPI_ISL_1709522, EPI_ISL_1709523, EPI_ISL_1709524, EPI_ISL_1709525, EPI_ISL_1709526, EPI_ISL_1709527, EPI_ISL_1709528, EPI_ISL_1709529, EPI_ISL_1709530, EPI_ISL_1709531, EPI_ISL_1709532, EPI_ISL_1709533, EPI_ISL_1709534, EPI_ISL_1709535, EPI_ISL_1709536, EPI_ISL_1709537, EPI_ISL_1709538, EPI_ISL_1709539, EPI_ISL_1709540, EPI_ISL_1709541, EPI_ISL_1709542, EPI_ISL_1709543, EPI_ISL_1709544, EPI_ISL_1709545, EPI_ISL_1709546, EPI_ISL_1709547, EPI_ISL_1709548, EPI_ISL_1709549, EPI_ISL_1709550, EPI_ISL_1709551, EPI_ISL_1709552, EPI_ISL_1709553, EPI_ISL_1709554, EPI_ISL_1709555, EPI_ISL_1709556, EPI_ISL_1709558, EPI_ISL_1709559, EPI_ISL_1709560, EPI_ISL_1709561, EPI_ISL_1709562, EPI_ISL_1709563, EPI_ISL_1709564, EPI_ISL_1709565, EPI_ISL_1709566, EPI_ISL_1709567, EPI_ISL_1709568, EPI_ISL_1709569, EPI_ISL_1709570, EPI_ISL_1709571, EPI_ISL_1709572, EPI_ISL_1709573, EPI_ISL_1709574, EPI_ISL_1709575, EPI_ISL_1709576, EPI_ISL_1709577, EPI_ISL_1709578, EPI_ISL_1709579, EPI_ISL_1709580, EPI_ISL_1709581, EPI_ISL_1709582, EPI_ISL_1709583, EPI_ISL_1709584, EPI_ISL_1709585, EPI_ISL_1709586, EPI_ISL_1709587, EPI_ISL_1709588, EPI_ISL_1709589, EPI_ISL_1709590, EPI_ISL_1709591, EPI_ISL_1709592, EPI_ISL_1709593, EPI_ISL_1709594, EPI_ISL_1709595, EPI_ISL_1709596, EPI_ISL_1709597, EPI_ISL_1709598, EPI_ISL_1709599, EPI_ISL_1709600, EPI_ISL_1709601, EPI_ISL_1709602, EPI_ISL_1709603, EPI_ISL_1709604, EPI_ISL_1709605, EPI_ISL_1709606, EPI_ISL_1709607, EPI_ISL_1709608, EPI_ISL_1709609, EPI_ISL_1709610, EPI_ISL_1709611, EPI_ISL_1709612, EPI_ISL_1709613, EPI_ISL_1709614, EPI_ISL_1709615, EPI_ISL_1709616, EPI_ISL_1709617, EPI_ISL_1709618, EPI_ISL_1709619, EPI_ISL_1709620, EPI_ISL_1709621, EPI_ISL_1709622, EPI_ISL_1709623, EPI_ISL_1709624, EPI_ISL_1709625, EPI_ISL_1709626, EPI_ISL_1709627, EPI_ISL_1709628, EPI_ISL_1709629, EPI_ISL_1709630, EPI_ISL_1709631, EPI_ISL_1709632, EPI_ISL_1709633, EPI_ISL_1709634, EPI_ISL_1709635, EPI_ISL_1709636, EPI_ISL_1709637, EPI_ISL_1709638, EPI_ISL_1709639, EPI_ISL_1709640, EPI_ISL_1709641, EPI_ISL_1709642, EPI_ISL_1709643, EPI_ISL_1709644, EPI_ISL_1709645, EPI_ISL_1709646, EPI_ISL_1709647, EPI_ISL_1709648, EPI_ISL_1709649, EPI_ISL_1709650, EPI_ISL_1709651, EPI_ISL_1709652, EPI_ISL_1709653, EPI_ISL_1709654, EPI_ISL_1709655, EPI_ISL_1709656, EPI_ISL_1709657, EPI_ISL_1709658, EPI_ISL_1709659, EPI_ISL_1709660, EPI_ISL_1709661, EPI_ISL_1709662, EPI_ISL_1709663, EPI_ISL_1709664, EPI_ISL_1709665, EPI_ISL_1709666, EPI_ISL_1709667, EPI_ISL_1709668, EPI_ISL_1709669, EPI_ISL_1709670, EPI_ISL_1709671, EPI_ISL_1709672, EPI_ISL_1709673, EPI_ISL_1709674, EPI_ISL_1709675, EPI_ISL_1709676, EPI_ISL_1709677, EPI_ISL_1709678, EPI_ISL_1709679, EPI_ISL_1709680, EPI_ISL_1709681, EPI_ISL_1709682, EPI_ISL_1709683, EPI_ISL_1709684, EPI_ISL_1709685, EPI_ISL_1709686, EPI_ISL_1709687, EPI_ISL_1709688, EPI_ISL_1709689, EPI_ISL_1709690, EPI_ISL_1709691, EPI_ISL_1709692, EPI_ISL_1709693, EPI_ISL_1709694, EPI_ISL_1709695, EPI_ISL_1709696, EPI_ISL_1709697, EPI_ISL_1709698, EPI_ISL_1709699, EPI_ISL_1709700, EPI_ISL_1709701, EPI_ISL_1709702, EPI_ISL_1709703, EPI_ISL_1709704, EPI_ISL_1709705, EPI_ISL_1709706, EPI_ISL_1709707, EPI_ISL_1709708, EPI_ISL_1709709, EPI_ISL_1709710, EPI_ISL_1709711, EPI_ISL_1709712, EPI_ISL_1709713, EPI_ISL_1709714, EPI_ISL_1709715, EPI_ISL_1709716, EPI_ISL_1709717, EPI_ISL_1709718, EPI_ISL_1709719, EPI_ISL_1709720, EPI_ISL_1709721, EPI_ISL_1709722, EPI_ISL_1709723, EPI_ISL_1709724, EPI_ISL_1709725, EPI_ISL_1709726, EPI_ISL_1709727, EPI_ISL_1709728, EPI_ISL_1709729, EPI_ISL_1709731, EPI_ISL_1709732, EPI_ISL_1709733, EPI_ISL_1709734, EPI_ISL_1709735, EPI_ISL_1709736, EPI_ISL_1709737, EPI_ISL_1709738, EPI_ISL_1709739, EPI_ISL_1709740, EPI_ISL_1709741, EPI_ISL_1709742, EPI_ISL_1709743, EPI_ISL_1709744, EPI_ISL_1709745, EPI_ISL_1709746, EPI_ISL_1709747, EPI_ISL_1709748, EPI_ISL_1709749, EPI_ISL_1709750, EPI_ISL_1709751, EPI_ISL_1709752, EPI_ISL_1709753, EPI_ISL_1709754, EPI_ISL_1709755, EPI_ISL_1709756, EPI_ISL_1709757, EPI_ISL_1709758, EPI_ISL_1709759, EPI_ISL_1709760, EPI_ISL_1709761, EPI_ISL_1709762, EPI_ISL_1709763, EPI_ISL_1709764, EPI_ISL_1709765, EPI_ISL_1709766, EPI_ISL_1709767, EPI_ISL_1709768, EPI_ISL_1709769, EPI_ISL_1709770, EPI_ISL_1709771, EPI_ISL_1709772, EPI_ISL_1709773, EPI_ISL_1709774, EPI_ISL_1709775, EPI_ISL_1709776, EPI_ISL_1709777, EPI_ISL_1709778, EPI_ISL_1709779, EPI_ISL_1709780, EPI_ISL_1709781, EPI_ISL_1709782, EPI_ISL_1709783, EPI_ISL_1709784, EPI_ISL_1709785, EPI_ISL_1709786, EPI_ISL_1709787, EPI_ISL_1709788, EPI_ISL_1709789, EPI_ISL_1709790, EPI_ISL_1709791, EPI_ISL_1709792, EPI_ISL_1709793, EPI_ISL_1709794, EPI_ISL_1709795, EPI_ISL_1709796, EPI_ISL_1709797, EPI_ISL_1709798, EPI_ISL_1709799, EPI_ISL_1709800, EPI_ISL_1709801, EPI_ISL_1709802, EPI_ISL_1709803, EPI_ISL_1709804, EPI_ISL_1709805, EPI_ISL_1709806, EPI_ISL_1709807, EPI_ISL_1709808, EPI_ISL_1709809, EPI_ISL_1709810, EPI_ISL_1709811, EPI_ISL_1709812, EPI_ISL_1709813, EPI_ISL_1709814, EPI_ISL_1709815, EPI_ISL_1709816, EPI_ISL_1709817, EPI_ISL_1709818, EPI_ISL_1709819, EPI_ISL_1709820, EPI_ISL_1709821, EPI_ISL_1709822, EPI_ISL_1709823, EPI_ISL_1709824, EPI_ISL_1709825, EPI_ISL_1709826, EPI_ISL_1709827, EPI_ISL_1709828, EPI_ISL_1709829, EPI_ISL_1709830, EPI_ISL_1709831, EPI_ISL_1709832, EPI_ISL_1709833, EPI_ISL_1709834, EPI_ISL_1709835, EPI_ISL_1709836, EPI_ISL_1709837, EPI_ISL_1709838, EPI_ISL_1709839, EPI_ISL_1709840, EPI_ISL_1709841, EPI_ISL_1709842, EPI_ISL_1709843, EPI_ISL_1709844, EPI_ISL_1709845, EPI_ISL_1709846, EPI_ISL_1709847, EPI_ISL_1709848, EPI_ISL_1709849, EPI_ISL_1709850, EPI_ISL_1709851, EPI_ISL_1709852, EPI_ISL_1709853, EPI_ISL_1709854, EPI_ISL_1709855, EPI_ISL_1709856, EPI_ISL_1709857, EPI_ISL_1709858, EPI_ISL_1709859, EPI_ISL_1709860, EPI_ISL_1709861, EPI_ISL_1709862, EPI_ISL_1709863, EPI_ISL_1709864, EPI_ISL_1709865, EPI_ISL_1709866, EPI_ISL_1709867, EPI_ISL_1709868, EPI_ISL_1709869, EPI_ISL_1709870, EPI_ISL_1709871, EPI_ISL_1709872, EPI_ISL_1709873, EPI_ISL_1709874, EPI_ISL_1709875, EPI_ISL_1709876, EPI_ISL_1709877, EPI_ISL_1709878, EPI_ISL_1709879, EPI_ISL_1709880, EPI_ISL_1709881, EPI_ISL_1709882, EPI_ISL_1709883, EPI_ISL_1709884, EPI_ISL_1709885, EPI_ISL_1709886, EPI_ISL_1709887, EPI_ISL_1709888, EPI_ISL_1709890, EPI_ISL_1709891, EPI_ISL_1709892, EPI_ISL_1709893, EPI_ISL_1709894, EPI_ISL_1709895, EPI_ISL_1709896, EPI_ISL_1709897, EPI_ISL_1709898, EPI_ISL_1709899, EPI_ISL_1709900, EPI_ISL_1709901, EPI_ISL_1709902, EPI_ISL_1709903, EPI_ISL_1709904, EPI_ISL_1709905, EPI_ISL_1709906, EPI_ISL_1709907, EPI_ISL_1709908, EPI_ISL_1709909, EPI_ISL_1709910, EPI_ISL_1709911, EPI_ISL_1709912, EPI_ISL_1709913, EPI_ISL_1709914, EPI_ISL_1709915, EPI_ISL_1709916, EPI_ISL_1709917, EPI_ISL_1709918, EPI_ISL_1709919, EPI_ISL_1709920, EPI_ISL_1709921, EPI_ISL_1709922, EPI_ISL_1709923, EPI_ISL_1709924, EPI_ISL_1709925, EPI_ISL_1709926, EPI_ISL_1709927, EPI_ISL_1709928, EPI_ISL_1709929, EPI_ISL_1709930, EPI_ISL_1709931, EPI_ISL_1709932, EPI_ISL_1709933, EPI_ISL_1709934, EPI_ISL_1709935, EPI_ISL_1709936, EPI_ISL_1709937, EPI_ISL_1709938, EPI_ISL_1709939, EPI_ISL_1709940, EPI_ISL_1709941, EPI_ISL_1709942, EPI_ISL_1709943, EPI_ISL_1709944, EPI_ISL_1709945, EPI_ISL_1709947, EPI_ISL_1709948, EPI_ISL_1709950, EPI_ISL_1709951, EPI_ISL_1709952, EPI_ISL_1709953, EPI_ISL_1709954, EPI_ISL_1709955, EPI_ISL_1709956, EPI_ISL_1709957, EPI_ISL_1709958, EPI_ISL_1709959, EPI_ISL_1709960, EPI_ISL_1709961, EPI_ISL_1709962, EPI_ISL_1709963, EPI_ISL_1709964, EPI_ISL_1709965, EPI_ISL_1709966, EPI_ISL_1709967, EPI_ISL_1709968, EPI_ISL_1709969, EPI_ISL_1709970, EPI_ISL_1709971, EPI_ISL_1709972, EPI_ISL_1709974, EPI_ISL_1709975, EPI_ISL_1709976, EPI_ISL_1709977, EPI_ISL_1709978, EPI_ISL_1709979, EPI_ISL_1709980, EPI_ISL_1709981, EPI_ISL_1709982, EPI_ISL_1709983, EPI_ISL_1709984, EPI_ISL_1709985, EPI_ISL_1709986, EPI_ISL_1709987, EPI_ISL_1709988, EPI_ISL_1709989, EPI_ISL_1709991, EPI_ISL_1709992, EPI_ISL_1709993, EPI_ISL_1709994, EPI_ISL_1709995, EPI_ISL_1709996, EPI_ISL_1709997, EPI_ISL_1709998, EPI_ISL_1709999, EPI_ISL_1710000, EPI_ISL_1710001, EPI_ISL_1710002, EPI_ISL_1710003, EPI_ISL_1710004, EPI_ISL_1710005, EPI_ISL_1710006, EPI_ISL_1710007, EPI_ISL_1710008, EPI_ISL_1710009, EPI_ISL_1710010, EPI_ISL_1710011, EPI_ISL_1710012, EPI_ISL_1710013, EPI_ISL_1710014

| | | | |
|---|---|---|---|
| see above | MSHS Clinical Microbiology Laboratories | MSHS Pathogen Surveillance Program | Ana S. Gonzalez-Reiche, Hala Alshammary, Mitchell J. Sullivan, Brianne Ciferri, Ajay Obla, Angela Amoako, Mahmoud Awawda, Daniel Floda, Julia Matthews, Ashley Salimbangon, Levy Sominsky, Katherine Beach, Kayla Russo, Charles Gleason, Shelcie Fabre, Giulio Kleiner, Zenab Khan, Bremy Alburquerque, Adriana van de Guchte, Komal Srivastava, Matthew M. Hernandez, Jayeeta Dutta, Denise Jurczyszak, Nancy Francoeur, Betsaida Salom Melo, Irina Oussenko, Gintaras Deikus, Juan Soto, Shwetha Hara Sridhar, Ying-Chih Wang, Kathryn Twyman, Deena R. Altman, Robert Sebra, Adolfo García-Sastre, Marta Luksza, Gopi Patel, Sarah Schaefer, Melissa Gitman, Michael D. Nowak, Alberto Paniz-Mondolfi, Emilia Mia Sordillo, Viviana Simon, Harm van Bakel |

EPI_ISL_1710048, EPI_ISL_1710050, EPI_ISL_1710060, EPI_ISL_1710064, EPI_ISL_1710067, EPI_ISL_1710075, EPI_ISL_1710081, EPI_ISL_1710085, EPI_ISL_1710086, EPI_ISL_1710095, EPI_ISL_1710100, EPI_ISL_1710102, EPI_ISL_1710103, EPI_ISL_1710105, EPI_ISL_1710136, EPI_ISL_1710138

| | | | |
|---|---|---|---|
| see above | Broad Institute Clinical Research Sequencing Platform | Infectious Disease Program, Broad Institute of Harvard and MIT | Siddle,K.J., Adams,G., Pearlman,L., Gladden-Young,A., Vicente,G., Blumenstiel,B., DeFelice,M., Lee,M., McGovern,S., Lagerborg,K., Rudy,M., DeRuff,K., Carter,A., Normandin,E., Bauer,M., Reilly,S., Tomkins-Tinch,C., Loreth,C., Chaluvadi,S., Meldrim,J., Granger,B., Lemieux,J.E., Birren,B.W., Sabeti,P.C., Larkin,K., Dodge,S., Lennon,N., Madoff,L., Brown,C., Gallagher,G., Smole,S., Park,D.J., Gabriel,S., and MacInnis,B.L. |
| EPI_ISL_1711717, EPI_ISL_1711718 | "NYSDOH Wadsworth Center, Virology Lab" | Centers for Disease Control and Prevention Division of Viral Diseases, Pathogen Discovery | Mili Sheth, Sarah Nobles, Jasmine Padilla, Mark Burroughs, Shoshona Le, Katie Dillon, Peter Cook, Clinton R. Paden, Dhwani Batra, Krista Queen, Kristen Knipe, Dakota Howard, Yvette Unoarumhi, Darlene Wagner, Matthew Schmerer, Ben L. Rambo-Martin, Kristine Lacek, Sam Shepard, Alison Laufer Halpin, Dave Wentworth, Vivien Dugan, Suxiang Tong, Justin Lee |

EPI_ISL_1717107, EPI_ISL_1717108, EPI_ISL_1717109, EPI_ISL_1717110, EPI_ISL_1717111, EPI_ISL_1717112, EPI_ISL_1717113, EPI_ISL_1717114, EPI_ISL_1717115, EPI_ISL_1717116, EPI_ISL_1717117, EPI_ISL_1717118, EPI_ISL_1717119, EPI_ISL_1717120, EPI_ISL_1717121, EPI_ISL_1717122, EPI_ISL_1717123, EPI_ISL_1717124, EPI_ISL_1717125, EPI_ISL_1717126, EPI_ISL_1717127, EPI_ISL_1717128, EPI_ISL_1717129, EPI_ISL_1717130, EPI_ISL_1717131, EPI_ISL_1717132, EPI_ISL_1717133, EPI_ISL_1717134, EPI_ISL_1717135, EPI_ISL_1717136, EPI_ISL_1717137, EPI_ISL_1717138, EPI_ISL_1717139, EPI_ISL_1717140, EPI_ISL_1717141, EPI_ISL_1717142, EPI_ISL_1717143, EPI_ISL_1717144, EPI_ISL_1717145, EPI_ISL_1717146, EPI_ISL_1717147, EPI_ISL_1717148, EPI_ISL_1717149, EPI_ISL_1717150, EPI_ISL_1717151, EPI_ISL_1717152, EPI_ISL_1717153, EPI_ISL_1717154, EPI_ISL_1717155, EPI_ISL_1717156, EPI_ISL_1717157, EPI_ISL_1717158, EPI_ISL_1717159, EPI_ISL_1717160, EPI_ISL_1717161, EPI_ISL_1717162, EPI_ISL_1717163, EPI_ISL_1717164, EPI_ISL_1717165, EPI_ISL_1717166, EPI_ISL_1717167, EPI_ISL_1717168, EPI_ISL_1717169, EPI_ISL_1717170, EPI_ISL_1717171, EPI_ISL_1717172, EPI_ISL_1717173, EPI_ISL_1717174, EPI_ISL_1717175, EPI_ISL_1717176, EPI_ISL_1717177, EPI_ISL_1717178, EPI_ISL_1717179, EPI_ISL_1717180, EPI_ISL_1717181, EPI_ISL_1717182, EPI_ISL_1717183, EPI_ISL_1717184, EPI_ISL_1717185, EPI_ISL_1717186, EPI_ISL_1717187, EPI_ISL_1717188, EPI_ISL_1717189, EPI_ISL_1717190, EPI_ISL_1717191, EPI_ISL_1717192, EPI_ISL_1717193, EPI_ISL_1717194, EPI_ISL_1717195, EPI_ISL_1717196, EPI_ISL_1717197, EPI_ISL_1717198, EPI_ISL_1717199, EPI_ISL_1717200, EPI_ISL_1717201, EPI_ISL_1717202, EPI_ISL_1717203, EPI_ISL_1717204, EPI_ISL_1717205, EPI_ISL_1717206, EPI_ISL_1717207, EPI_ISL_1717208, EPI_ISL_1717209, EPI_ISL_1717210, EPI_ISL_1717211, EPI_ISL_1717212, EPI_ISL_1717213, EPI_ISL_1717214, EPI_ISL_1717215, EPI_ISL_1717216, EPI_ISL_1717217, EPI_ISL_1717218, EPI_ISL_1717219, EPI_ISL_1717220, EPI_ISL_1717221, EPI_ISL_1717222, EPI_ISL_1717223, EPI_ISL_1717224, EPI_ISL_1717225, EPI_ISL_1717226, EPI_ISL_1717227, EPI_ISL_1717228, EPI_ISL_1717229, EPI_ISL_1717230, EPI_ISL_1717231, EPI_ISL_1717232, EPI_ISL_1717233, EPI_ISL_1717234, EPI_ISL_1717235, EPI_ISL_1717236, EPI_ISL_1717237, EPI_ISL_1717238, EPI_ISL_1717239, EPI_ISL_1717240, EPI_ISL_1717241, EPI_ISL_1717242, EPI_ISL_1717243, EPI_ISL_1717244, EPI_ISL_1717245, EPI_ISL_1717246, EPI_ISL_1717247, EPI_ISL_1717248, EPI_ISL_1717249, EPI_ISL_1717250, EPI_ISL_1717251, EPI_ISL_1717252, EPI_ISL_1717253, EPI_ISL_1717254, EPI_ISL_1717255, EPI_ISL_1717256, EPI_ISL_1717257, EPI_ISL_1717258, EPI_ISL_1717259, EPI_ISL_1717260, EPI_ISL_1717261, EPI_ISL_1717262, EPI_ISL_1717263, EPI_ISL_1717264, EPI_ISL_1717265, EPI_ISL_1717266, EPI_ISL_1717267, EPI_ISL_1717268, EPI_ISL_1717269, EPI_ISL_1717270, EPI_ISL_1717271, EPI_ISL_1717272, EPI_ISL_1717273, EPI_ISL_1717274, EPI_ISL_1717275, EPI_ISL_1717276, EPI_ISL_1717277, EPI_ISL_1717278, EPI_ISL_1717279, EPI_ISL_1717280, EPI_ISL_1717281, EPI_ISL_1717282, EPI_ISL_1717283, EPI_ISL_1717284, EPI_ISL_1717285, EPI_ISL_1717286, EPI_ISL_1717287, EPI_ISL_1717288, EPI_ISL_1717289, EPI_ISL_1717290, EPI_ISL_1717291, EPI_ISL_1717292, EPI_ISL_1717293, EPI_ISL_1717294, EPI_ISL_1717295, EPI_ISL_1717296, EPI_ISL_1717297, EPI_ISL_1717298, EPI_ISL_1717299, EPI_ISL_1717300, EPI_ISL_1717301, EPI_ISL_1717302, EPI_ISL_1717303, EPI_ISL_1717304, EPI_ISL_1717305, EPI_ISL_1717306, EPI_ISL_1717307, EPI_ISL_1717308, EPI_ISL_1717309, EPI_ISL_1717310,

| | | | |
|---|---|---|---|
| EPI_ISL_1717311, EPI_ISL_1717312, EPI_ISL_1717313, EPI_ISL_1717314, EPI_ISL_1717315, EPI_ISL_1717316, EPI_ISL_1717317, EPI_ISL_1717318, EPI_ISL_1717319, EPI_ISL_1717320, EPI_ISL_1717321, EPI_ISL_1717322, EPI_ISL_1717323, EPI_ISL_1717324, EPI_ISL_1717325, EPI_ISL_1717326, EPI_ISL_1717327, EPI_ISL_1717328, EPI_ISL_1717329, EPI_ISL_1717330, EPI_ISL_1717331, EPI_ISL_1717332, EPI_ISL_1717333, EPI_ISL_1717334, EPI_ISL_1717335, EPI_ISL_1717336, EPI_ISL_1717337, EPI_ISL_1717338, EPI_ISL_1717339, EPI_ISL_1717340, EPI_ISL_1717341, EPI_ISL_1717342, EPI_ISL_1717343, EPI_ISL_1717344, EPI_ISL_1717345, EPI_ISL_1717346, EPI_ISL_1717347, EPI_ISL_1717348, EPI_ISL_1717349, EPI_ISL_1717350, EPI_ISL_1717351, EPI_ISL_1717352, EPI_ISL_1717353, EPI_ISL_1717354, EPI_ISL_1717355, EPI_ISL_1717356, EPI_ISL_1717357, EPI_ISL_1717358, EPI_ISL_1717359, EPI_ISL_1717360, EPI_ISL_1717361, EPI_ISL_1717362, EPI_ISL_1717363, EPI_ISL_1717364, EPI_ISL_1717365, EPI_ISL_1717366, EPI_ISL_1717367, EPI_ISL_1717368, EPI_ISL_1717369, EPI_ISL_1717370, EPI_ISL_1717371, EPI_ISL_1717372, EPI_ISL_1717373, EPI_ISL_1717374, EPI_ISL_1717375, EPI_ISL_1717376, EPI_ISL_1717377, EPI_ISL_1717378, EPI_ISL_1717379, EPI_ISL_1717380, EPI_ISL_1717381, EPI_ISL_1717382, EPI_ISL_1717383, EPI_ISL_1717384, EPI_ISL_1717385, EPI_ISL_1717386, EPI_ISL_1717387, EPI_ISL_1717388, EPI_ISL_1717389, EPI_ISL_1717390, EPI_ISL_1717391, EPI_ISL_1717392, EPI_ISL_1717393, EPI_ISL_1717394, EPI_ISL_1717395, EPI_ISL_1717396, EPI_ISL_1717397, EPI_ISL_1717398, EPI_ISL_1717399, EPI_ISL_1717400, EPI_ISL_1717401, EPI_ISL_1717402, EPI_ISL_1717403, EPI_ISL_1717404, EPI_ISL_1717405, EPI_ISL_1717406, EPI_ISL_1717407, EPI_ISL_1717408, EPI_ISL_1717409, EPI_ISL_1717410, EPI_ISL_1717411, EPI_ISL_1717412, EPI_ISL_1717413, EPI_ISL_1717414, EPI_ISL_1717415, EPI_ISL_1717416, EPI_ISL_1717417, EPI_ISL_1717418, EPI_ISL_1717419, EPI_ISL_1717420, EPI_ISL_1717421, EPI_ISL_1717422, EPI_ISL_1717423, EPI_ISL_1717424, EPI_ISL_1717425, EPI_ISL_1717426, EPI_ISL_1717427, EPI_ISL_1717428, EPI_ISL_1717429, EPI_ISL_1717430, EPI_ISL_1717431, EPI_ISL_1717432, EPI_ISL_1717433, EPI_ISL_1717434, EPI_ISL_1717435, EPI_ISL_1717436, EPI_ISL_1717437, EPI_ISL_1717438, EPI_ISL_1717439, EPI_ISL_1717440, EPI_ISL_1717441, EPI_ISL_1717442, EPI_ISL_1717443, EPI_ISL_1717444, EPI_ISL_1717445, EPI_ISL_1717446, EPI_ISL_1717447, EPI_ISL_1717448, EPI_ISL_1717449, EPI_ISL_1717450, EPI_ISL_1717451, EPI_ISL_1717452, EPI_ISL_1717453, EPI_ISL_1717454, EPI_ISL_1717455, EPI_ISL_1717456, EPI_ISL_1717457, EPI_ISL_1717458, EPI_ISL_1717459, EPI_ISL_1717460, EPI_ISL_1717461, EPI_ISL_1717462, EPI_ISL_1717463, EPI_ISL_1717464, EPI_ISL_1717465, EPI_ISL_1717466, EPI_ISL_1717467, EPI_ISL_1717468, EPI_ISL_1717469, EPI_ISL_1717470, EPI_ISL_1717471, EPI_ISL_1717472, EPI_ISL_1717473, EPI_ISL_1717474, EPI_ISL_1717475, EPI_ISL_1717476, EPI_ISL_1717477, EPI_ISL_1717478, EPI_ISL_1717479, EPI_ISL_1717480, EPI_ISL_1717481, EPI_ISL_1717482, EPI_ISL_1717483, EPI_ISL_1717484, EPI_ISL_1717485, EPI_ISL_1717486, EPI_ISL_1717487, EPI_ISL_1717488, EPI_ISL_1717489, EPI_ISL_1717490, EPI_ISL_1717491, EPI_ISL_1717492, EPI_ISL_1717493, EPI_ISL_1717494, EPI_ISL_1717495, EPI_ISL_1717496, EPI_ISL_1717497, EPI_ISL_1717498, EPI_ISL_1717499, EPI_ISL_1717500, EPI_ISL_1717501, EPI_ISL_1717502, EPI_ISL_1717503, EPI_ISL_1717504, EPI_ISL_1717505, EPI_ISL_1717506, EPI_ISL_1717507, EPI_ISL_1717508, EPI_ISL_1717509, EPI_ISL_1717510, EPI_ISL_1717511, EPI_ISL_1717512, EPI_ISL_1717513, EPI_ISL_1717514, EPI_ISL_1717515, EPI_ISL_1717516, EPI_ISL_1717517, EPI_ISL_1717518, EPI_ISL_1717519, EPI_ISL_1717520, EPI_ISL_1717521, EPI_ISL_1717522, EPI_ISL_1717523, EPI_ISL_1717524, EPI_ISL_1717525, EPI_ISL_1717526, EPI_ISL_1717527, EPI_ISL_1717528, EPI_ISL_1717529, EPI_ISL_1717530, EPI_ISL_1717531, EPI_ISL_1717532, EPI_ISL_1717533, EPI_ISL_1717534, EPI_ISL_1717535, EPI_ISL_1717536, EPI_ISL_1717537, EPI_ISL_1717538, EPI_ISL_1717539, EPI_ISL_1717540, EPI_ISL_1717541, EPI_ISL_1717542, EPI_ISL_1717543, EPI_ISL_1717544, EPI_ISL_1717545, EPI_ISL_1717546, EPI_ISL_1717547, EPI_ISL_1717548, EPI_ISL_1717549, EPI_ISL_1717550, EPI_ISL_1717551, EPI_ISL_1717552, EPI_ISL_1717553, EPI_ISL_1717554, EPI_ISL_1717555, EPI_ISL_1717556, EPI_ISL_1717557, EPI_ISL_1717558, EPI_ISL_1717559, EPI_ISL_1717560, EPI_ISL_1717561, EPI_ISL_1717562, EPI_ISL_1717563, EPI_ISL_1717564, EPI_ISL_1717565, EPI_ISL_1717566, EPI_ISL_1717567, EPI_ISL_1717568, EPI_ISL_1717569, EPI_ISL_1717570, EPI_ISL_1717571, EPI_ISL_1717572, EPI_ISL_1717573, EPI_ISL_1717574, EPI_ISL_1717575, EPI_ISL_1717576, EPI_ISL_1717577, EPI_ISL_1717578, EPI_ISL_1717579, EPI_ISL_1717580, EPI_ISL_1717581, EPI_ISL_1717582, EPI_ISL_1717583, EPI_ISL_1717584, EPI_ISL_1717585, EPI_ISL_1717586, EPI_ISL_1717587, EPI_ISL_1717588, EPI_ISL_1717589, EPI_ISL_1717590, EPI_ISL_1717591, EPI_ISL_1717592, EPI_ISL_1717593, EPI_ISL_1717594, EPI_ISL_1717595, EPI_ISL_1717596, EPI_ISL_1717597, EPI_ISL_1717598, EPI_ISL_1717599, EPI_ISL_1717600, EPI_ISL_1717601, EPI_ISL_1717602, EPI_ISL_1717603, EPI_ISL_1717604, EPI_ISL_1717605, EPI_ISL_1717606, EPI_ISL_1717607, EPI_ISL_1717608, EPI_ISL_1717609, EPI_ISL_1717610, EPI_ISL_1717611, EPI_ISL_1717612, EPI_ISL_1717613, EPI_ISL_1717614, EPI_ISL_1717615, EPI_ISL_1717616, EPI_ISL_1717617, EPI_ISL_1717618, EPI_ISL_1717619, EPI_ISL_1717620, EPI_ISL_1717621, EPI_ISL_1717622, EPI_ISL_1717623, EPI_ISL_1717624, EPI_ISL_1717625, EPI_ISL_1717626, EPI_ISL_1717627, EPI_ISL_1717628, EPI_ISL_1717629, EPI_ISL_1717630, EPI_ISL_1717631, EPI_ISL_1717632, EPI_ISL_1717633, EPI_ISL_1717634, EPI_ISL_1717635, EPI_ISL_1717636, EPI_ISL_1717637, EPI_ISL_1717638, EPI_ISL_1717639, EPI_ISL_1717640, EPI_ISL_1717641, EPI_ISL_1717642, EPI_ISL_1717643, EPI_ISL_1717644, EPI_ISL_1717645, EPI_ISL_1717646, EPI_ISL_1717647, EPI_ISL_1717648, EPI_ISL_1717649, EPI_ISL_1717650, EPI_ISL_1717651, EPI_ISL_1717652, EPI_ISL_1717653, EPI_ISL_1717654, EPI_ISL_1717655, EPI_ISL_1717656, EPI_ISL_1717657, EPI_ISL_1717658, EPI_ISL_1717659, EPI_ISL_1717660, EPI_ISL_1717661, EPI_ISL_1717662, EPI_ISL_1717663, EPI_ISL_1717664, EPI_ISL_1717665, EPI_ISL_1717666, EPI_ISL_1717667, EPI_ISL_1717668, EPI_ISL_1717669, EPI_ISL_1717670, EPI_ISL_1717671, EPI_ISL_1717672, EPI_ISL_1717673, EPI_ISL_1717674, EPI_ISL_1717675, EPI_ISL_1717676, EPI_ISL_1717677, EPI_ISL_1717678, EPI_ISL_1717679, EPI_ISL_1717680, EPI_ISL_1717681, EPI_ISL_1717682, EPI_ISL_1717683, EPI_ISL_1717684, EPI_ISL_1717685, EPI_ISL_1717686, EPI_ISL_1717687, EPI_ISL_1717688, EPI_ISL_1717689, EPI_ISL_1717690, EPI_ISL_1717691, EPI_ISL_1717692, EPI_ISL_1717693, EPI_ISL_1717694, EPI_ISL_1717695, EPI_ISL_1717696, EPI_ISL_1717697, EPI_ISL_1717698, EPI_ISL_1717699, EPI_ISL_1717700, EPI_ISL_1717701, EPI_ISL_1717702, EPI_ISL_1717703, EPI_ISL_1717704, EPI_ISL_1717705, EPI_ISL_1717706, EPI_ISL_1717707, EPI_ISL_1717708, EPI_ISL_1717709, EPI_ISL_1717710, EPI_ISL_1717711, EPI_ISL_1717712, EPI_ISL_1717713, EPI_ISL_1717714, EPI_ISL_1717715, EPI_ISL_1717716, EPI_ISL_1717717, EPI_ISL_1717718, EPI_ISL_1717719, EPI_ISL_1717720, EPI_ISL_1717721, EPI_ISL_1717722, EPI_ISL_1717723, EPI_ISL_1717724, EPI_ISL_1717725, EPI_ISL_1717726, EPI_ISL_1717727, EPI_ISL_1717728, EPI_ISL_1717729, EPI_ISL_1717730, EPI_ISL_1717731, EPI_ISL_1717732, EPI_ISL_1717733, EPI_ISL_1717734, EPI_ISL_1717735, EPI_ISL_1717736, EPI_ISL_1717737, EPI_ISL_1717738, EPI_ISL_1717739, EPI_ISL_1717740, EPI_ISL_1717741, EPI_ISL_1717742, EPI_ISL_1717743, EPI_ISL_1717744, EPI_ISL_1717745, EPI_ISL_1717746, EPI_ISL_1717747, EPI_ISL_1717748, EPI_ISL_1717749, EPI_ISL_1717750, EPI_ISL_1717751, EPI_ISL_1717752, EPI_ISL_1717753, EPI_ISL_1717754, EPI_ISL_1717755, EPI_ISL_1717756, EPI_ISL_1717757, EPI_ISL_1717758, EPI_ISL_1717759, EPI_ISL_1717760, EPI_ISL_1717761, EPI_ISL_1717762, EPI_ISL_1717763, EPI_ISL_1717764, EPI_ISL_1717765, EPI_ISL_1717766, EPI_ISL_1717767, EPI_ISL_1717768, EPI_ISL_1717769, EPI_ISL_1717770, EPI_ISL_1717771, EPI_ISL_1717772, EPI_ISL_1717773, EPI_ISL_1717774, EPI_ISL_1717775, EPI_ISL_1717776, EPI_ISL_1717777, EPI_ISL_1717778, EPI_ISL_1717779, EPI_ISL_1717780, EPI_ISL_1717781, EPI_ISL_1717782, EPI_ISL_1717783, EPI_ISL_1717784, EPI_ISL_1717785, EPI_ISL_1717786, EPI_ISL_1717787, EPI_ISL_1717788, EPI_ISL_1717789, EPI_ISL_1717790, EPI_ISL_1717791, EPI_ISL_1717792, EPI_ISL_1717793, EPI_ISL_1717794, EPI_ISL_1717795, EPI_ISL_1717796, EPI_ISL_1717797, EPI_ISL_1717798, EPI_ISL_1717799, EPI_ISL_1717800, EPI_ISL_1717801, EPI_ISL_1717802, EPI_ISL_1717803, EPI_ISL_1717804, EPI_ISL_1717805, EPI_ISL_1717806, EPI_ISL_1717807, EPI_ISL_1717808, EPI_ISL_1717809, EPI_ISL_1717810, EPI_ISL_1717811, EPI_ISL_1717812, EPI_ISL_1717813, EPI_ISL_1717814, EPI_ISL_1717815, EPI_ISL_1717816, EPI_ISL_1717817, EPI_ISL_1717818, EPI_ISL_1717819, EPI_ISL_1717820, EPI_ISL_1717821, EPI_ISL_1717822, EPI_ISL_1717823, EPI_ISL_1717824, EPI_ISL_1717825, EPI_ISL_1717826, EPI_ISL_1717827, EPI_ISL_1717828, EPI_ISL_1717829, EPI_ISL_1717830, EPI_ISL_1717831, EPI_ISL_1717832, EPI_ISL_1717833, EPI_ISL_1717834, EPI_ISL_1717835, EPI_ISL_1717836, EPI_ISL_1717837, EPI_ISL_1717838, EPI_ISL_1717839, EPI_ISL_1717840, EPI_ISL_1717841, EPI_ISL_1717842, EPI_ISL_1717843, EPI_ISL_1717844, EPI_ISL_1717845, EPI_ISL_1717846, EPI_ISL_1717847, EPI_ISL_1717848, EPI_ISL_1717849, EPI_ISL_1717850, EPI_ISL_1717851, EPI_ISL_1717852, EPI_ISL_1717853, EPI_ISL_1717854, EPI_ISL_1717855, EPI_ISL_1717856, EPI_ISL_1717857, EPI_ISL_1717858, EPI_ISL_1717859, EPI_ISL_1717860, EPI_ISL_1717861, EPI_ISL_1717862, EPI_ISL_1717863, EPI_ISL_1717864, EPI_ISL_1717865, EPI_ISL_1717866, EPI_ISL_1717867, EPI_ISL_1717868, EPI_ISL_1717869, EPI_ISL_1717870, EPI_ISL_1717871, EPI_ISL_1717872, EPI_ISL_1717873, EPI_ISL_1717874, EPI_ISL_1717875, EPI_ISL_1717876, EPI_ISL_1717877, EPI_ISL_1717878, EPI_ISL_1717879, EPI_ISL_1717880, EPI_ISL_1717881, EPI_ISL_1717882, EPI_ISL_1717883, EPI_ISL_1717884, EPI_ISL_1717885, EPI_ISL_1717886, EPI_ISL_1717887, EPI_ISL_1717888, EPI_ISL_1717889, EPI_ISL_1717890, EPI_ISL_1717891, EPI_ISL_1717892, EPI_ISL_1717893, EPI_ISL_1717894, EPI_ISL_1717895, EPI_ISL_1717896, EPI_ISL_1717897, EPI_ISL_1717898, EPI_ISL_1717899, EPI_ISL_1717900, EPI_ISL_1717901, EPI_ISL_1717902, EPI_ISL_1717903, EPI_ISL_1717904, EPI_ISL_1717905, EPI_ISL_1717906, EPI_ISL_1717907, EPI_ISL_1717908, EPI_ISL_1717909, EPI_ISL_1717910, EPI_ISL_1717911, EPI_ISL_1717912, EPI_ISL_1717913, EPI_ISL_1717914, EPI_ISL_1717915, EPI_ISL_1717916, EPI_ISL_1717917, EPI_ISL_1717918, EPI_ISL_1717919, EPI_ISL_1717920, EPI_ISL_1717921, EPI_ISL_1717922, EPI_ISL_1717923, EPI_ISL_1717924, EPI_ISL_1717925, EPI_ISL_1717926, EPI_ISL_1717927, EPI_ISL_1717928, EPI_ISL_1717929, EPI_ISL_1717930, EPI_ISL_1717931, EPI_ISL_1717932, EPI_ISL_1717933, EPI_ISL_1717934, EPI_ISL_1717935, EPI_ISL_1717936, EPI_ISL_1717937, EPI_ISL_1717938, EPI_ISL_1717939, EPI_ISL_1717940, EPI_ISL_1717941, EPI_ISL_1717942, EPI_ISL_1717943, EPI_ISL_1717944, EPI_ISL_1717945, EPI_ISL_1717946, EPI_ISL_1717947, EPI_ISL_1717948, EPI_ISL_1717949, EPI_ISL_1717950, EPI_ISL_1717951, EPI_ISL_1717952, EPI_ISL_1717953, EPI_ISL_1717954, EPI_ISL_1717955, EPI_ISL_1717956, EPI_ISL_1717957, EPI_ISL_1717958, EPI_ISL_1717959, EPI_ISL_1717960, EPI_ISL_1717961, EPI_ISL_1717962, EPI_ISL_1717963, EPI_ISL_1717964, EPI_ISL_1717965, EPI_ISL_1717966, EPI_ISL_1717967, EPI_ISL_1717968, EPI_ISL_1717969, EPI_ISL_1717970, EPI_ISL_1717971, EPI_ISL_1717972, EPI_ISL_1717973, EPI_ISL_1717974, EPI_ISL_1717975, EPI_ISL_1717976, EPI_ISL_1717977, EPI_ISL_1717978, EPI_ISL_1717979, EPI_ISL_1717980, EPI_ISL_1717981, EPI_ISL_1717982, EPI_ISL_1717983, EPI_ISL_1717984, EPI_ISL_1717985, EPI_ISL_1717986, EPI_ISL_1717987, EPI_ISL_1717988, EPI_ISL_1717989, EPI_ISL_1717990, EPI_ISL_1717991, EPI_ISL_1717992, EPI_ISL_1717993, EPI_ISL_1717994, EPI_ISL_1717995, EPI_ISL_1717996, EPI_ISL_1717997, EPI_ISL_1717998, EPI_ISL_1717999, EPI_ISL_1718000, EPI_ISL_1718001, EPI_ISL_1718002, EPI_ISL_1718003, EPI_ISL_1718004, EPI_ISL_1718005, EPI_ISL_1718006, EPI_ISL_1718007, EPI_ISL_1718008, EPI_ISL_1718009, EPI_ISL_1718010, EPI_ISL_1718011, EPI_ISL_1718012, EPI_ISL_1718013, EPI_ISL_1718014, EPI_ISL_1718015, EPI_ISL_1718016, EPI_ISL_1718017, EPI_ISL_1718018, EPI_ISL_1718019, EPI_ISL_1718020, EPI_ISL_1718021, EPI_ISL_1718022, EPI_ISL_1718023, EPI_ISL_1718024, EPI_ISL_1718025, EPI_ISL_1718026, EPI_ISL_1718027, EPI_ISL_1718028, EPI_ISL_1718029, EPI_ISL_1718030, EPI_ISL_1718031, EPI_ISL_1718032, EPI_ISL_1718033, EPI_ISL_1718034, EPI_ISL_1718035, EPI_ISL_1718036, EPI_ISL_1718037, EPI_ISL_1718038, EPI_ISL_1718039, EPI_ISL_1718040, EPI_ISL_1718041, EPI_ISL_1718042, EPI_ISL_1718043, EPI_ISL_1718044, EPI_ISL_1718045, EPI_ISL_1718046, EPI_ISL_1718047, EPI_ISL_1718048, EPI_ISL_1718060, EPI_ISL_1718061, EPI_ISL_1718062, EPI_ISL_1718063, EPI_ISL_1718064, EPI_ISL_1718065, EPI_ISL_1718066, EPI_ISL_1718067, EPI_ISL_1718068, EPI_ISL_1718069, EPI_ISL_1718070, EPI_ISL_1718071, EPI_ISL_1718072, EPI_ISL_1718073, EPI_ISL_1718074, EPI_ISL_1718075, EPI_ISL_1718076, EPI_ISL_1718077, EPI_ISL_1718078, EPI_ISL_1718079, EPI_ISL_1718080, EPI_ISL_1718081, EPI_ISL_1718082, EPI_ISL_1718083, EPI_ISL_1718084, EPI_ISL_1718085, EPI_ISL_1718086, EPI_ISL_1718087, EPI_ISL_1718088, EPI_ISL_1718089, EPI_ISL_1718090, EPI_ISL_1718091, EPI_ISL_1718092, EPI_ISL_1718093, EPI_ISL_1718094, EPI_ISL_1718095, EPI_ISL_1718096, EPI_ISL_1718097, EPI_ISL_1718098, EPI_ISL_1718099, EPI_ISL_1718100, EPI_ISL_1718101, EPI_ISL_1718102, EPI_ISL_1718103, EPI_ISL_1718104, EPI_ISL_1718105, EPI_ISL_1718106, EPI_ISL_1718107, EPI_ISL_1718108, EPI_ISL_1718109, EPI_ISL_1718110, EPI_ISL_1718111, EPI_ISL_1718112, EPI_ISL_1718113, EPI_ISL_1718114, EPI_ISL_1718115, EPI_ISL_1718116, EPI_ISL_1718117, EPI_ISL_1718118, EPI_ISL_1718119, EPI_ISL_1718120, EPI_ISL_1718121, EPI_ISL_1718122, EPI_ISL_1718123, EPI_ISL_1718124, EPI_ISL_1718125, EPI_ISL_1718126, EPI_ISL_1718127, EPI_ISL_1718128, EPI_ISL_1718129, EPI_ISL_1718130, EPI_ISL_1718131, EPI_ISL_1718132, EPI_ISL_1718133, EPI_ISL_1718134, EPI_ISL_1718135, EPI_ISL_1718136, EPI_ISL_1718137, EPI_ISL_1718138, EPI_ISL_1718139, EPI_ISL_1718140, EPI_ISL_1718141, EPI_ISL_1718142, EPI_ISL_1718143, EPI_ISL_1718144, EPI_ISL_1718145, EPI_ISL_1718146, EPI_ISL_1718147, EPI_ISL_1718148, EPI_ISL_1718149, EPI_ISL_1718150, EPI_ISL_1718151, EPI_ISL_1718152, EPI_ISL_1718153, EPI_ISL_1718154, EPI_ISL_1718155, EPI_ISL_1718156, EPI_ISL_1718157, EPI_ISL_1718158, EPI_ISL_1718159, EPI_ISL_1718160, EPI_ISL_1718161, EPI_ISL_1718162, EPI_ISL_1718163, EPI_ISL_1718164, EPI_ISL_1718165, EPI_ISL_1718166, EPI_ISL_1718167, EPI_ISL_1718168, EPI_ISL_1718169, EPI_ISL_1718170, EPI_ISL_1718171, EPI_ISL_1718172, EPI_ISL_1718173, EPI_ISL_1718174, EPI_ISL_1718175, EPI_ISL_1718176, EPI_ISL_1718177, EPI_ISL_1718178, EPI_ISL_1718179, EPI_ISL_1718180, EPI_ISL_1718181, EPI_ISL_1718182, EPI_ISL_1718183, EPI_ISL_1718184, EPI_ISL_1718185, EPI_ISL_1718186, EPI_ISL_1718187, EPI_ISL_1718188, EPI_ISL_1718189, EPI_ISL_1718190, EPI_ISL_1718191, EPI_ISL_1718192, EPI_ISL_1718193, EPI_ISL_1718194, EPI_ISL_1718195, EPI_ISL_1718196, EPI_ISL_1718197, EPI_ISL_1718198, EPI_ISL_1718199, EPI_ISL_1718200, EPI_ISL_1718201, EPI_ISL_1718202, EPI_ISL_1718203, EPI_ISL_1718204, EPI_ISL_1718205, EPI_ISL_1718206, EPI_ISL_1718207, EPI_ISL_1718208, EPI_ISL_1718209, EPI_ISL_1718210, EPI_ISL_1718211, EPI_ISL_1718212, EPI_ISL_1718213, EPI_ISL_1718214, EPI_ISL_1718215, EPI_ISL_1718216, EPI_ISL_1718217, EPI_ISL_1718218, EPI_ISL_1718219, EPI_ISL_1718220, EPI_ISL_1718221, EPI_ISL_1718222, EPI_ISL_1718223, EPI_ISL_1718224, EPI_ISL_1718225, EPI_ISL_1718226, EPI_ISL_1718227, EPI_ISL_1718228, EPI_ISL_1718229, EPI_ISL_1718230, EPI_ISL_1718231, EPI_ISL_1718232, EPI_ISL_1718233, EPI_ISL_1718234, EPI_ISL_1718235, EPI_ISL_1718236, EPI_ISL_1718237, EPI_ISL_1718238, EPI_ISL_1718239, EPI_ISL_1718240, EPI_ISL_1718241, EPI_ISL_1718242, EPI_ISL_1718243, EPI_ISL_1718244, EPI_ISL_1718245, EPI_ISL_1718246, EPI_ISL_1718247 | | | |
| see above | Pandemic Response Lab - NYC | Pandemic Response Lab, R&D | Henry Lee, Michael Hammerling, Melissa Hopkins, Cybill del Castillo, Shinyoung Clair Kang, William Ward, Pradeep Bugga, Sol Rey, Dylan Law, Katharine Nelson, Haiping Hao, Jon Laurent |
| EPI_ISL_1733367, EPI_ISL_1733640, EPI_ISL_1733721 | Infinity Biologix | Centers for Disease Control and Prevention Division of Viral Diseases, Pathogen Discovery | Dakota Howard, Dhwani Batra, Peter W. Cook, Kara Moser, Adrian Paskey, Jason Caravas, Benjamin Rambo-Martin, Shatavia Morrison, Christopher Gulvick, Scott Sammons, Yvette Unoarumhi, Darlene Wagner, Matthew Schmerer, Christian Bixby, Yihe Wang, Jonathan Schultz, Chirayu Goswami, Russ Hager, Robin Grimwood, Clinton R. Paden, Duncan MacCannell |
| EPI_ISL_1734943, EPI_ISL_1735142, EPI_ISL_1735643 | Helix/Illumina | Centers for Disease Control and Prevention Division of Viral Diseases, Pathogen Discovery | Dakota Howard, Dhwani Batra, Peter W. Cook, Kara Moser, Adrian Paskey, Jason Caravas, Benjamin Rambo-Martin, Shatavia Morrison, Christopher Gulvick, Scott Sammons, Yvette Unoarumhi, Darlene Wagner, Matthew Schmerer, Eileen de Feo, Jan Antico, Christine Tran, Matthew Tolentino, Shannon Wickline, Kim Gietzen, Brad Sickler, Jingtao Liu, Eric Allen, Phil Febbo, Nicole L. Washington, Simon White, Geraint Levan, Kelly Schiabor Barrett, Elizabeth Cirulli, Alexandre Bolze, Ary Ascencio, Charlotte Rivera-Garcia, Ryan Cho, Jason Nguyen, Sherry Wang, Jimmy Ramirez, Tyler Cassens, Efren Sandoval, Magnus Isaksson, William Lee, David Becker, Marc Laurent, James Lu, Clinton R. Paden, Duncan MacCannell |
| EPI_ISL_1735719, EPI_ISL_1735723, EPI_ISL_1735724, EPI_ISL_1735772, EPI_ISL_1735773, EPI_ISL_1735774, EPI_ISL_1735775, EPI_ISL_1735783, EPI_ISL_1735855, EPI_ISL_1735914, EPI_ISL_1735940, EPI_ISL_1736007, EPI_ISL_1736116, EPI_ISL_1736118, EPI_ISL_1736119, EPI_ISL_1736120, EPI_ISL_1736146, EPI_ISL_1736163, EPI_ISL_1736218, EPI_ISL_1736219, EPI_ISL_1736235, EPI_ISL_1736375, EPI_ISL_1736384, EPI_ISL_1736495, EPI_ISL_1736516, EPI_ISL_1736525, EPI_ISL_1736550, EPI_ISL_1736626, EPI_ISL_1736656, EPI_ISL_1736685, EPI_ISL_1736695, EPI_ISL_1736702, EPI_ISL_1736720, EPI_ISL_1736763, EPI_ISL_1736796, EPI_ISL_1736798, EPI_ISL_1736799, EPI_ISL_1736806, EPI_ISL_1736809, EPI_ISL_1736842, EPI_ISL_1736847, EPI_ISL_1736852, EPI_ISL_1736862, EPI_ISL_1736882, EPI_ISL_1736890, EPI_ISL_1736899, EPI_ISL_1736927, EPI_ISL_1736930, EPI_ISL_1737063, EPI_ISL_1737198, EPI_ISL_1737202, EPI_ISL_1737223, EPI_ISL_1737238, EPI_ISL_1737239, EPI_ISL_1737284, EPI_ISL_1737289, EPI_ISL_1737290, EPI_ISL_1737291, EPI_ISL_1737292, EPI_ISL_1737296, EPI_ISL_1737399, EPI_ISL_1737478, EPI_ISL_1737512, EPI_ISL_1737517, EPI_ISL_1737589, EPI_ISL_1737590, EPI_ISL_1737597, EPI_ISL_1737689, EPI_ISL_1737780, EPI_ISL_1737813, EPI_ISL_1737847 | | | |
| see above | Aegis Sciences Corporation | Centers for Disease Control and Prevention Division of Viral Diseases, Pathogen Discovery | Dakota Howard, Dhwani Batra, Peter W. Cook, Kara Moser, Adrian Paskey, Jason Caravas, Benjamin Rambo-Martin, Shatavia Morrison, Christopher Gulvick, Scott Sammons, Yvette Unoarumhi, Darlene Wagner, Matthew Schmerer, Cyndi Clark, Patrick Campbell, Rob Case, Vikramsinha Ghorpade, Holly Houdeshell, Ola Kvalvaag, Dillon Nall, Ethan Sanders, Alec Vest, Shaun Westlund, Matthew Hardison, Clinton R. Paden, Duncan MacCannell |
| EPI_ISL_1738119, EPI_ISL_1738187, EPI_ISL_1738235, EPI_ISL_1738591, EPI_ISL_1738671, EPI_ISL_1738750 | Quest Diagnostics Incorporated | Centers for Disease Control and Prevention Division of Viral Diseases, Pathogen Discovery | Dakota Howard, Dhwani Batra, Peter W. Cook, Kara Moser, Adrian Paskey, Jason Caravas, Benjamin Rambo-Martin, Shatavia Morrison, Christopher Gulvick, Scott Sammons, Yvette Unoarumhi, Darlene Wagner, Matthew Schmerer, S. H. Rosenthal, A. Gerasimova, R. M. Kagan, B. Anderson, M. Hua, Y. Liu, L.E. Bernstein, K.E. Livingston, A. Perez, I. A. Shlyakhter, R. V. Rolando, R. Owen, P. Tanpaiboon, F. Lacbawan, Clinton R. Paden, Duncan MacCannell |
| EPI_ISL_1738769 | Aegis Sciences Corporation | Centers for Disease Control and Prevention Division of Viral Diseases, Pathogen Discovery | Dakota Howard, Dhwani Batra, Peter W. Cook, Kara Moser, Adrian Paskey, Jason Caravas, Benjamin Rambo-Martin, Shatavia Morrison, Christopher Gulvick, Scott Sammons, Yvette Unoarumhi, Darlene Wagner, Matthew Schmerer, Cyndi Clark, Patrick Campbell, Rob Case, Vikramsinha Ghorpade, Holly Houdeshell, Ola Kvalvaag, Dillon Nall, Ethan Sanders, Alec Vest, Shaun Westlund, Matthew Hardison, Clinton R. Paden, Duncan MacCannell |
| EPI_ISL_1739440 | The Jackson Laboratory | The Jackson Laboratory | Bergeron D, Renzette N, Adams M, Omerza G, Kelly K, Long J, Li L |
| EPI_ISL_1740502 | MEMORIAL SLOAN KETTERING CANCER CENTER | Wadsworth Center, New York State Department of Health | Kirsten St. George, Daryl M. Lamson, Alexis Russell, Matthew Shudt, Melissa A Leisner, Jonathan Plitnick, Catharine Prussing, Navjot Singh, John Kelly, Erasmus Schneider, Erica Lasek-Nesselquist |

| | | | |
|---|---|---|---|
| EPI_ISL_1740503 | MONTEFIORE MEDICAL CENTER LABORATORIES | Wadsworth Center, New York State Department of Health | Kirsten St. George, Daryl M. Lamson, Alexis Russell, Matthew Shudt, Melissa A Leisner, Jonathan Plitnick, Catharine Prussing, Navjot Singh, John Kelly, Erasmus Schneider, Erica Lasek-Nesselquist |
| EPI_ISL_1740504 | NORTHWELL HEALTH LABORATORIES | Wadsworth Center, New York State Department of Health | Kirsten St. George, Daryl M. Lamson, Alexis Russell, Matthew Shudt, Melissa A Leisner, Jonathan Plitnick, Catharine Prussing, Navjot Singh, John Kelly, Erasmus Schneider, Erica Lasek-Nesselquist |
| EPI_ISL_1740505 | MONTEFIORE MEDICAL CENTER LABORATORIES | Wadsworth Center, New York State Department of Health | Kirsten St. George, Daryl M. Lamson, Alexis Russell, Matthew Shudt, Melissa A Leisner, Jonathan Plitnick, Catharine Prussing, Navjot Singh, John Kelly, Erasmus Schneider, Erica Lasek-Nesselquist |
| EPI_ISL_1740506, EPI_ISL_1740507 | TEMPUS LABS INC | Wadsworth Center, New York State Department of Health | Kirsten St. George, Daryl M. Lamson, Alexis Russell, Matthew Shudt, Melissa A Leisner, Jonathan Plitnick, Catharine Prussing, Navjot Singh, John Kelly, Erasmus Schneider, Erica Lasek-Nesselquist |
| EPI_ISL_1740508 | STONY BROOK UNIVERSITY HOSPITAL | Wadsworth Center, New York State Department of Health | Kirsten St. George, Daryl M. Lamson, Alexis Russell, Matthew Shudt, Melissa A Leisner, Jonathan Plitnick, Catharine Prussing, Navjot Singh, John Kelly, Erasmus Schneider, Erica Lasek-Nesselquist |
| EPI_ISL_1740509 | ALBANY MEDICAL CENTER | Wadsworth Center, New York State Department of Health | Kirsten St. George, Daryl M. Lamson, Alexis Russell, Matthew Shudt, Melissa A Leisner, Jonathan Plitnick, Catharine Prussing, Navjot Singh, John Kelly, Erasmus Schneider, Erica Lasek-Nesselquist |
| EPI_ISL_1740510, EPI_ISL_1740511 | BOSTON HEART DIAGNOSTICS CORP | Wadsworth Center, New York State Department of Health | Kirsten St. George, Daryl M. Lamson, Alexis Russell, Matthew Shudt, Melissa A Leisner, Jonathan Plitnick, Catharine Prussing, Navjot Singh, John Kelly, Erasmus Schneider, Erica Lasek-Nesselquist |
| EPI_ISL_1740512, EPI_ISL_1740513 | MONTEFIORE MEDICAL CENTER LABORATORIES | Wadsworth Center, New York State Department of Health | Kirsten St. George, Daryl M. Lamson, Alexis Russell, Matthew Shudt, Melissa A Leisner, Jonathan Plitnick, Catharine Prussing, Navjot Singh, John Kelly, Erasmus Schneider, Erica Lasek-Nesselquist |
| EPI_ISL_1740514, EPI_ISL_1740515 | WESTCHESTER MEDICAL CENTER | Wadsworth Center, New York State Department of Health | Kirsten St. George, Daryl M. Lamson, Alexis Russell, Matthew Shudt, Melissa A Leisner, Jonathan Plitnick, Catharine Prussing, Navjot Singh, John Kelly, Erasmus Schneider, Erica Lasek-Nesselquist |
| EPI_ISL_1740516, EPI_ISL_1740517 | URMC LABS | Wadsworth Center, New York State Department of Health | Kirsten St. George, Daryl M. Lamson, Alexis Russell, Matthew Shudt, Melissa A Leisner, Jonathan Plitnick, Catharine Prussing, Navjot Singh, John Kelly, Erasmus Schneider, Erica Lasek-Nesselquist |
| EPI_ISL_1740518, EPI_ISL_1740519, EPI_ISL_1740520, EPI_ISL_1740521, EPI_ISL_1740522 | NORTHWELL HEALTH LABORATORIES | Wadsworth Center, New York State Department of Health | Kirsten St. George, Daryl M. Lamson, Alexis Russell, Matthew Shudt, Melissa A Leisner, Jonathan Plitnick, Catharine Prussing, Navjot Singh, John Kelly, Erasmus Schneider, Erica Lasek-Nesselquist |
| EPI_ISL_1753345, EPI_ISL_1753413 | Quest Diagnostics Incorporated | Centers for Disease Control and Prevention Division of Viral Diseases, Pathogen Discovery | Dakota Howard, Dhwani Batra, Peter W. Cook, Kara Moser, Adrian Paskey, Jason Caravas, Benjamin Rambo-Martin, Shatavia Morrison, Christopher Gulvick, Scott Sammons, Yvette Unoarumhi, Darlene Wagner, Matthew Schmerer, S. H. Rosenthal, A. Gerasimova, R. M. Kagan, B. Anderson, M. Hua, Y. Liu, L.E. Bernstein, K.E. Livingston, A. Perez, I. A. Shlyakhter, R. V. Rolando, R. Owen, P. Tanpaiboon, F. Lacbawan, Clinton R. Paden, Duncan MacCannell |
| EPI_ISL_1753533, EPI_ISL_1753552, EPI_ISL_1753557, EPI_ISL_1753572, EPI_ISL_1753574, EPI_ISL_1753582 | Aegis Sciences Corporation | Centers for Disease Control and Prevention Division of Viral Diseases, Pathogen Discovery | Dakota Howard, Dhwani Batra, Peter W. Cook, Kara Moser, Adrian Paskey, Jason Caravas, Benjamin Rambo-Martin, Shatavia Morrison, Christopher Gulvick, Scott Sammons, Yvette Unoarumhi, Darlene Wagner, Matthew Schmerer, Cyndi Clark, Patrick Campbell, Rob Case, Vikramsinha Ghorpade, Holly Houdeshell, Ola Kvalvaag, Dillon Nall, Ethan Sanders, Alec Vest, Shaun Westlund, Matthew Hardison, Clinton R. Paden, Duncan MacCannell |
| EPI_ISL_1753625, EPI_ISL_1753644 | Quest Diagnostics Incorporated | Centers for Disease Control and Prevention Division of Viral Diseases, Pathogen Discovery | Dakota Howard, Dhwani Batra, Peter W. Cook, Kara Moser, Adrian Paskey, Jason Caravas, Benjamin Rambo-Martin, Shatavia Morrison, Christopher Gulvick, Scott Sammons, Yvette Unoarumhi, Darlene Wagner, Matthew Schmerer, S. H. Rosenthal, A. Gerasimova, R. M. Kagan, B. Anderson, M. Hua, Y. Liu, L.E. Bernstein, K.E. Livingston, A. Perez, I. A. Shlyakhter, R. V. Rolando, R. Owen, P. Tanpaiboon, F. Lacbawan, Clinton R. Paden, Duncan MacCannell |
| EPI_ISL_1755445, EPI_ISL_1755446, EPI_ISL_1755447, EPI_ISL_1755448, EPI_ISL_1755449, EPI_ISL_1755450, EPI_ISL_1755451, EPI_ISL_1755452, EPI_ISL_1755453, EPI_ISL_1755454, EPI_ISL_1755455, EPI_ISL_1755456, EPI_ISL_1755457, EPI_ISL_1755458, EPI_ISL_1755459, EPI_ISL_1755460, EPI_ISL_1755461, EPI_ISL_1755462, EPI_ISL_1755463, EPI_ISL_1755464, EPI_ISL_1755465, EPI_ISL_1755466, EPI_ISL_1755467, EPI_ISL_1755468, EPI_ISL_1755469, EPI_ISL_1755470, EPI_ISL_1755471, EPI_ISL_1755472, EPI_ISL_1755473, EPI_ISL_1755474, EPI_ISL_1755475, EPI_ISL_1755476, EPI_ISL_1755477, EPI_ISL_1755478, EPI_ISL_1755479, EPI_ISL_1755480, EPI_ISL_1755481, EPI_ISL_1755482, EPI_ISL_1755483, EPI_ISL_1755484, EPI_ISL_1755485, EPI_ISL_1755486 | | | |
| see above | NYC Pandemic Response Lab | Wadsworth Center, New York State Department of Health | Kirsten St. George, Daryl M. Lamson, Alexis Russell, Matthew Shudt, Melissa A Leisner, Jonathan Plitnick, Catharine Prussing, Navjot Singh, John Kelly, Erasmus Schneider, Erica Lasek-Nesselquist |
| EPI_ISL_1755487 | SUNY UPSTATE MEDICAL UNIVERSITY | Wadsworth Center, New York State Department of Health | Kirsten St. George, Daryl M. Lamson, Alexis Russell, Matthew Shudt, Melissa A Leisner, Jonathan Plitnick, Catharine Prussing, Navjot Singh, John Kelly, Erasmus Schneider, Erica Lasek-Nesselquist |
| EPI_ISL_1755488 | GLENS FALLS HOSPITAL LABORATORY | Wadsworth Center, New York State Department of Health | Kirsten St. George, Daryl M. Lamson, Alexis Russell, Matthew Shudt, Melissa A Leisner, Jonathan Plitnick, Catharine Prussing, Navjot Singh, John Kelly, Erasmus Schneider, Erica Lasek-Nesselquist |
| EPI_ISL_1755489, EPI_ISL_1755490 | SUNY UPSTATE MEDICAL UNIVERSITY | Wadsworth Center, New York State Department of Health | Kirsten St. George, Daryl M. Lamson, Alexis Russell, Matthew Shudt, Melissa A Leisner, Jonathan Plitnick, Catharine Prussing, Navjot Singh, John Kelly, Erasmus Schneider, Erica Lasek-Nesselquist |
| EPI_ISL_1755491, EPI_ISL_1755492, EPI_ISL_1755493, EPI_ISL_1755494, EPI_ISL_1755495, EPI_ISL_1755496, EPI_ISL_1755497, EPI_ISL_1755498, EPI_ISL_1755499, EPI_ISL_1755500, EPI_ISL_1755501 | | | |
| see above | NYC Pandemic Response Lab | Wadsworth Center, New York State Department of Health | Kirsten St. George, Daryl M. Lamson, Alexis Russell, Matthew Shudt, Melissa A Leisner, Jonathan Plitnick, Catharine Prussing, Navjot Singh, John Kelly, Erasmus Schneider, Erica Lasek-Nesselquist |
| EPI_ISL_1755820, EPI_ISL_1755821, EPI_ISL_1755822, EPI_ISL_1755823, EPI_ISL_1755824, EPI_ISL_1755825, EPI_ISL_1755826, EPI_ISL_1755827, EPI_ISL_1755828, EPI_ISL_1755829, EPI_ISL_1755830, EPI_ISL_1755831, EPI_ISL_1755832, EPI_ISL_1755833, EPI_ISL_1755834 | | | |
| see above | MONTEFIORE MEDICAL CENTER LABORATORIES | Wadsworth Center, New York State Department of Health | Kirsten St. George, Daryl M. Lamson, Alexis Russell, Matthew Shudt, Melissa A Leisner, Jonathan Plitnick, Catharine Prussing, Navjot Singh, John Kelly, Erasmus Schneider, Erica Lasek-Nesselquist |
| EPI_ISL_1755835 | URMC LABS | Wadsworth Center, New York State Department of Health | Kirsten St. George, Daryl M. Lamson, Alexis Russell, Matthew Shudt, Melissa A Leisner, Jonathan Plitnick, Catharine Prussing, Navjot Singh, John Kelly, Erasmus Schneider, Erica Lasek-Nesselquist |
| EPI_ISL_1755836, EPI_ISL_1755837 | MONTEFIORE MEDICAL CENTER LABORATORIES | Wadsworth Center, New York State Department of Health | Kirsten St. George, Daryl M. Lamson, Alexis Russell, Matthew Shudt, Melissa A Leisner, Jonathan Plitnick, Catharine Prussing, Navjot Singh, John Kelly, Erasmus Schneider, Erica Lasek-Nesselquist |
| EPI_ISL_1755838, EPI_ISL_1755839, EPI_ISL_1755840, EPI_ISL_1755841 | ALBANY MEDICAL CENTER | Wadsworth Center, New York State Department of Health | Kirsten St. George, Daryl M. Lamson, Alexis Russell, Matthew Shudt, Melissa A Leisner, Jonathan Plitnick, Catharine Prussing, Navjot Singh, John Kelly, Erasmus Schneider, Erica Lasek-Nesselquist |
| EPI_ISL_1755842, EPI_ISL_1755843, EPI_ISL_1755844, EPI_ISL_1755845, EPI_ISL_1755846, EPI_ISL_1755847, EPI_ISL_1755848 | SUNY UPSTATE MEDICAL UNIVERSITY | Wadsworth Center, New York State Department of Health | Kirsten St. George, Daryl M. Lamson, Alexis Russell, Matthew Shudt, Melissa A Leisner, Jonathan Plitnick, Catharine Prussing, Navjot Singh, John Kelly, Erasmus Schneider, Erica Lasek-Nesselquist |
| EPI_ISL_1755849, EPI_ISL_1755850 | BOSTON HEART DIAGNOSTICS CORP | Wadsworth Center, New York State Department of Health | Kirsten St. George, Daryl M. Lamson, Alexis Russell, Matthew Shudt, Melissa A Leisner, Jonathan Plitnick, Catharine Prussing, Navjot Singh, John Kelly, Erasmus Schneider, Erica Lasek-Nesselquist |
| EPI_ISL_1755851, EPI_ISL_1755852, EPI_ISL_1755853 | WESTCHESTER MEDICAL CENTER | Wadsworth Center, New York State Department of Health | Kirsten St. George, Daryl M. Lamson, Alexis Russell, Matthew Shudt, Melissa A Leisner, Jonathan Plitnick, Catharine Prussing, Navjot Singh, John Kelly, Erasmus Schneider, Erica Lasek-Nesselquist |
| EPI_ISL_1755854, EPI_ISL_1755855, EPI_ISL_1755856, EPI_ISL_1755857, EPI_ISL_1755858, EPI_ISL_1755859, EPI_ISL_1755860, EPI_ISL_1755861, EPI_ISL_1755862, EPI_ISL_1755863, EPI_ISL_1755864, EPI_ISL_1755865, EPI_ISL_1755866 | | | |
| see above | Columbia University Irving Medical Center | Wadsworth Center, New York State Department of Health | Kirsten St. George, Daryl M. Lamson, Alexis Russell, Matthew Shudt, Melissa A Leisner, Jonathan Plitnick, Catharine Prussing, Navjot Singh, John Kelly, Erasmus Schneider, Erica Lasek-Nesselquist |
| EPI_ISL_1755867, EPI_ISL_1755868 | WESTCHESTER MEDICAL CENTER | Wadsworth Center, New York State Department of Health | Kirsten St. George, Daryl M. Lamson, Alexis Russell, Matthew Shudt, Melissa A Leisner, Jonathan Plitnick, Catharine Prussing, Navjot Singh, John Kelly, Erasmus Schneider, Erica Lasek-Nesselquist |
| EPI_ISL_1755869, EPI_ISL_1755870, EPI_ISL_1755871, EPI_ISL_1755872, EPI_ISL_1755873, EPI_ISL_1755874, EPI_ISL_1755875, EPI_ISL_1755876, EPI_ISL_1755877, EPI_ISL_1755878, EPI_ISL_1755879, EPI_ISL_1755880, EPI_ISL_1755881, EPI_ISL_1755882, EPI_ISL_1755883, EPI_ISL_1755884, EPI_ISL_1755885, | | | |

| | | | |
|---|---|---|---|
| EPI_ISL_1755886, EPI_ISL_1755887, EPI_ISL_1755888, EPI_ISL_1755889, EPI_ISL_1755890, EPI_ISL_1755891, EPI_ISL_1755892, EPI_ISL_1755893, EPI_ISL_1755894, EPI_ISL_1755895, EPI_ISL_1755896, EPI_ISL_1755897, EPI_ISL_1755898, EPI_ISL_1755899, EPI_ISL_1755900, EPI_ISL_1755901, EPI_ISL_1755902, EPI_ISL_1755903, EPI_ISL_1755904, EPI_ISL_1755905, EPI_ISL_1755906, EPI_ISL_1755907 | | | |
| see above | URMC LABS | Wadsworth Center, New York State Department of Health | Kirsten St. George, Daryl M. Lamson, Alexis Russell, Matthew Shudt, Melissa A Leisner, Jonathan Plitnick, Catharine Prussing, Navjot Singh, John Kelly, Erasmus Schneider, Erica Lasek-Nesselquist |
| EPI_ISL_1755939 | ALBANY MEDICAL CENTER | Wadsworth Center, New York State Department of Health | Kirsten St. George, Daryl M. Lamson, Alexis Russell, Matthew Shudt, Melissa A Leisner, Jonathan Plitnick, Catharine Prussing, Navjot Singh, John Kelly, Erasmus Schneider, Erica Lasek-Nesselquist |
| EPI_ISL_1755940 | MEMORIAL SLOAN KETTERING CANCER CENTER | Wadsworth Center, New York State Department of Health | Kirsten St. George, Daryl M. Lamson, Alexis Russell, Matthew Shudt, Melissa A Leisner, Jonathan Plitnick, Catharine Prussing, Navjot Singh, John Kelly, Erasmus Schneider, Erica Lasek-Nesselquist |
| EPI_ISL_1755941, EPI_ISL_1755942 | NYC Pandemic Response Lab | Wadsworth Center, New York State Department of Health | Kirsten St. George, Daryl M. Lamson, Alexis Russell, Matthew Shudt, Melissa A Leisner, Jonathan Plitnick, Catharine Prussing, Navjot Singh, John Kelly, Erasmus Schneider, Erica Lasek-Nesselquist |
| EPI_ISL_1755943, EPI_ISL_1755944, EPI_ISL_1755945, EPI_ISL_1755946 | Columbia University Irving Medical Center | Wadsworth Center, New York State Department of Health | Kirsten St. George, Daryl M. Lamson, Alexis Russell, Matthew Shudt, Melissa A Leisner, Jonathan Plitnick, Catharine Prussing, Navjot Singh, John Kelly, Erasmus Schneider, Erica Lasek-Nesselquist |
| EPI_ISL_1755947 | NYC Pandemic Response Lab | Wadsworth Center, New York State Department of Health | Kirsten St. George, Daryl M. Lamson, Alexis Russell, Matthew Shudt, Melissa A Leisner, Jonathan Plitnick, Catharine Prussing, Navjot Singh, John Kelly, Erasmus Schneider, Erica Lasek-Nesselquist |
| EPI_ISL_1755948, EPI_ISL_1755949, EPI_ISL_1755950, EPI_ISL_1755951, EPI_ISL_1755952, EPI_ISL_1755953, EPI_ISL_1755954, EPI_ISL_1755955, EPI_ISL_1755956, EPI_ISL_1755957, EPI_ISL_1755958, EPI_ISL_1755959, EPI_ISL_1755960, EPI_ISL_1755961, EPI_ISL_1755962, EPI_ISL_1755963, EPI_ISL_1755964 | | | |
| see above | MONTEFIORE MEDICAL CENTER LABORATORIES | Wadsworth Center, New York State Department of Health | Kirsten St. George, Daryl M. Lamson, Alexis Russell, Matthew Shudt, Melissa A Leisner, Jonathan Plitnick, Catharine Prussing, Navjot Singh, John Kelly, Erasmus Schneider, Erica Lasek-Nesselquist |
| EPI_ISL_1755965, EPI_ISL_1755966, EPI_ISL_1755967 | WESTCHESTER MEDICAL CENTER | Wadsworth Center, New York State Department of Health | Kirsten St. George, Daryl M. Lamson, Alexis Russell, Matthew Shudt, Melissa A Leisner, Jonathan Plitnick, Catharine Prussing, Navjot Singh, John Kelly, Erasmus Schneider, Erica Lasek-Nesselquist |
| EPI_ISL_1755968, EPI_ISL_1755969, EPI_ISL_1755970, EPI_ISL_1755971, EPI_ISL_1755972, EPI_ISL_1755973, EPI_ISL_1755974, EPI_ISL_1755975, EPI_ISL_1755976, EPI_ISL_1755977, EPI_ISL_1755978, EPI_ISL_1755979, EPI_ISL_1755980, EPI_ISL_1755981, EPI_ISL_1755982, EPI_ISL_1755983, EPI_ISL_1755984, EPI_ISL_1755985, EPI_ISL_1755986, EPI_ISL_1755987, EPI_ISL_1755988, EPI_ISL_1755989, EPI_ISL_1755990, EPI_ISL_1755991 | | | |
| see above | TEMPUS LABS INC | Wadsworth Center, New York State Department of Health | Kirsten St. George, Daryl M. Lamson, Alexis Russell, Matthew Shudt, Melissa A Leisner, Jonathan Plitnick, Catharine Prussing, Navjot Singh, John Kelly, Erasmus Schneider, Erica Lasek-Nesselquist |
| EPI_ISL_1755992, EPI_ISL_1755993, EPI_ISL_1755994, EPI_ISL_1755995, EPI_ISL_1755996, EPI_ISL_1755997, EPI_ISL_1755998, EPI_ISL_1755999 | SARATOGA HOSPITAL LABORATORY | Wadsworth Center, New York State Department of Health | Kirsten St. George, Daryl M. Lamson, Alexis Russell, Matthew Shudt, Melissa A Leisner, Jonathan Plitnick, Catharine Prussing, Navjot Singh, John Kelly, Erasmus Schneider, Erica Lasek-Nesselquist |
| EPI_ISL_1756000 | STONY BROOK UNIVERSITY HOSPITAL | Wadsworth Center, New York State Department of Health | Kirsten St. George, Daryl M. Lamson, Alexis Russell, Matthew Shudt, Melissa A Leisner, Jonathan Plitnick, Catharine Prussing, Navjot Singh, John Kelly, Erasmus Schneider, Erica Lasek-Nesselquist |
| EPI_ISL_1756001, EPI_ISL_1756002, EPI_ISL_1756003, EPI_ISL_1756004, EPI_ISL_1756005, EPI_ISL_1756006, EPI_ISL_1756007, EPI_ISL_1756008, EPI_ISL_1756009, EPI_ISL_1756010, EPI_ISL_1756011, EPI_ISL_1756012, EPI_ISL_1756013, EPI_ISL_1756014, EPI_ISL_1756015, EPI_ISL_1756016, EPI_ISL_1756017, EPI_ISL_1756018, EPI_ISL_1756019, EPI_ISL_1756020 | | | |
| see above | ALBANY MEDICAL CENTER | Wadsworth Center, New York State Department of Health | Kirsten St. George, Daryl M. Lamson, Alexis Russell, Matthew Shudt, Melissa A Leisner, Jonathan Plitnick, Catharine Prussing, Navjot Singh, John Kelly, Erasmus Schneider, Erica Lasek-Nesselquist |
| EPI_ISL_1756326 | Columbia University Irving Medical Center | Wadsworth Center, New York State Department of Health | Kirsten St. George, Daryl M. Lamson, Alexis Russell, Matthew Shudt, Melissa A Leisner, Jonathan Plitnick, Catharine Prussing, Navjot Singh, John Kelly, Erasmus Schneider, Erica Lasek-Nesselquist |
| EPI_ISL_1756327 | NORTHWELL HEALTH LABORATORIES | Wadsworth Center, New York State Department of Health | Kirsten St. George, Daryl M. Lamson, Alexis Russell, Matthew Shudt, Melissa A Leisner, Jonathan Plitnick, Catharine Prussing, Navjot Singh, John Kelly, Erasmus Schneider, Erica Lasek-Nesselquist |
| EPI_ISL_1756328 | Columbia University Irving Medical Center | Wadsworth Center, New York State Department of Health | Kirsten St. George, Daryl M. Lamson, Alexis Russell, Matthew Shudt, Melissa A Leisner, Jonathan Plitnick, Catharine Prussing, Navjot Singh, John Kelly, Erasmus Schneider, Erica Lasek-Nesselquist |
| EPI_ISL_1756329 | ALBANY MEDICAL CENTER | Wadsworth Center, New York State Department of Health | Kirsten St. George, Daryl M. Lamson, Alexis Russell, Matthew Shudt, Melissa A Leisner, Jonathan Plitnick, Catharine Prussing, Navjot Singh, John Kelly, Erasmus Schneider, Erica Lasek-Nesselquist |
| EPI_ISL_1756330 | NORTHWELL HEALTH LABORATORIES | Wadsworth Center, New York State Department of Health | Kirsten St. George, Daryl M. Lamson, Alexis Russell, Matthew Shudt, Melissa A Leisner, Jonathan Plitnick, Catharine Prussing, Navjot Singh, John Kelly, Erasmus Schneider, Erica Lasek-Nesselquist |
| EPI_ISL_1756331, EPI_ISL_1756332, EPI_ISL_1756333, EPI_ISL_1756334, EPI_ISL_1756335, EPI_ISL_1756336, EPI_ISL_1756337 | STONY BROOK UNIVERSITY HOSPITAL | Wadsworth Center, New York State Department of Health | Kirsten St. George, Daryl M. Lamson, Alexis Russell, Matthew Shudt, Melissa A Leisner, Jonathan Plitnick, Catharine Prussing, Navjot Singh, John Kelly, Erasmus Schneider, Erica Lasek-Nesselquist |
| EPI_ISL_1756338, EPI_ISL_1756339, EPI_ISL_1756340, EPI_ISL_1756341 | SUNY UPSTATE MEDICAL UNIVERSITY | Wadsworth Center, New York State Department of Health | Kirsten St. George, Daryl M. Lamson, Alexis Russell, Matthew Shudt, Melissa A Leisner, Jonathan Plitnick, Catharine Prussing, Navjot Singh, John Kelly, Erasmus Schneider, Erica Lasek-Nesselquist |
| EPI_ISL_1756342, EPI_ISL_1756343, EPI_ISL_1756344, EPI_ISL_1756345, EPI_ISL_1756346, EPI_ISL_1756347, EPI_ISL_1756348, EPI_ISL_1756349, EPI_ISL_1756350, EPI_ISL_1756351, EPI_ISL_1756352, EPI_ISL_1756353, EPI_ISL_1756354, EPI_ISL_1756355 | | | |
| see above | MONTEFIORE MEDICAL CENTER LABORATORIES | Wadsworth Center, New York State Department of Health | Kirsten St. George, Daryl M. Lamson, Alexis Russell, Matthew Shudt, Melissa A Leisner, Jonathan Plitnick, Catharine Prussing, Navjot Singh, John Kelly, Erasmus Schneider, Erica Lasek-Nesselquist |
| EPI_ISL_1756356 | BIO-REFERENCE LABORATORIES | Wadsworth Center, New York State Department of Health | Kirsten St. George, Daryl M. Lamson, Alexis Russell, Matthew Shudt, Melissa A Leisner, Jonathan Plitnick, Catharine Prussing, Navjot Singh, John Kelly, Erasmus Schneider, Erica Lasek-Nesselquist |
| EPI_ISL_1756357, EPI_ISL_1756358, EPI_ISL_1756359, EPI_ISL_1756360, EPI_ISL_1756361, EPI_ISL_1756362, EPI_ISL_1756363, EPI_ISL_1756364, EPI_ISL_1756365, EPI_ISL_1756366, EPI_ISL_1756367 | | | |
| see above | URMC LABS | Wadsworth Center, New York State Department of Health | Kirsten St. George, Daryl M. Lamson, Alexis Russell, Matthew Shudt, Melissa A Leisner, Jonathan Plitnick, Catharine Prussing, Navjot Singh, John Kelly, Erasmus Schneider, Erica Lasek-Nesselquist |
| EPI_ISL_1756368 | ST. FRANCIS HOSPITAL LABORATORY | Wadsworth Center, New York State Department of Health | Kirsten St. George, Daryl M. Lamson, Alexis Russell, Matthew Shudt, Melissa A Leisner, Jonathan Plitnick, Catharine Prussing, Navjot Singh, John Kelly, Erasmus Schneider, Erica Lasek-Nesselquist |
| EPI_ISL_1756369, EPI_ISL_1756370, EPI_ISL_1756371, EPI_ISL_1756372, EPI_ISL_1756373, EPI_ISL_1756374, EPI_ISL_1756375, EPI_ISL_1756376, EPI_ISL_1756377, EPI_ISL_1756378, EPI_ISL_1756379, EPI_ISL_1756380, EPI_ISL_1756381, EPI_ISL_1756382, EPI_ISL_1756383, EPI_ISL_1756384, EPI_ISL_1756385, EPI_ISL_1756386, EPI_ISL_1756387, EPI_ISL_1756388, EPI_ISL_1756389, EPI_ISL_1756390, EPI_ISL_1756391, EPI_ISL_1756392, EPI_ISL_1756393, EPI_ISL_1756394, EPI_ISL_1756395, EPI_ISL_1756396, EPI_ISL_1756397, EPI_ISL_1756398, EPI_ISL_1756399, EPI_ISL_1756400, EPI_ISL_1756401, EPI_ISL_1756402, EPI_ISL_1756403, EPI_ISL_1756404, EPI_ISL_1756405, EPI_ISL_1756406, EPI_ISL_1756407, EPI_ISL_1756408 | | | |
| see above | WESTCHESTER MEDICAL CENTER | Wadsworth Center, New York State Department of Health | Kirsten St. George, Daryl M. Lamson, Alexis Russell, Matthew Shudt, Melissa A Leisner, Jonathan Plitnick, Catharine Prussing, Navjot Singh, John Kelly, Erasmus Schneider, Erica Lasek-Nesselquist |
| EPI_ISL_1757072, EPI_ISL_1757073, EPI_ISL_1757074, EPI_ISL_1757076, EPI_ISL_1757077, EPI_ISL_1757089, EPI_ISL_1757091, EPI_ISL_1757092, EPI_ISL_1757279, EPI_ISL_1757284, EPI_ISL_1757286 | | | |
| see above | Broad Institute Clinical Research Sequencing Platform | Infectious Disease Program, Broad Institute of Harvard and MIT | Siddle,K.J., Adams,G., Pearlman,L., Gladden-Young,A., Vicente,G., Blumenstiel,B., DeFelice,M., Lee,M., McGovern,S., Lagerborg,K., Rudy,M., DeRuff,K., Carter,A., Normandin,E., Bauer,M., Reilly,S., Tomkins-Tinch,C., Loreth,C., Chaluvadi,S., Meldrim,J., Granger,B., Lemieux,J.E., Birren,B.W., Sabeti,P.C., Larkin,K., Dodge,S., Lennon,N., Madoff,L., Brown,C., Gallagher,G., Smole,S., Park,D.J., Gabriel,S., and MacInnis,B.L. |
| EPI_ISL_1761906, EPI_ISL_1761913, EPI_ISL_1761936, EPI_ISL_1761939, EPI_ISL_1761949, EPI_ISL_1761951, EPI_ISL_1761953, EPI_ISL_1761963, EPI_ISL_1761971, EPI_ISL_1761977, EPI_ISL_1761985, EPI_ISL_1762034, EPI_ISL_1762038 | | | |
| see above | Quest Diagnostics Incorporated | Centers for Disease Control and Prevention Division of Viral Diseases, Pathogen Discovery | Dakota Howard, Dhwani Batra, Peter W. Cook, Kara Moser, Adrian Paskey, Jason Caravas, Benjamin Rambo-Martin, Shatavia Morrison, Christopher Gulvick, Scott Sammons, Yvette Unoarumhi, Darlene Wagner, Matthew Schmerer, S. H. Rosenthal, A. Gerasimova, R. M. Kagan, B. Anderson, M. Hua, Y. |

| | | | |
|---|---|---|---|
| | | | Liu, L.E. Bernstein, K.E. Livingston, A. Perez, I. A. Shlyakhter, R. V. Rolando, R. Owen, P. Tanpaiboon, F. Lacbawan, Clinton R. Paden, Duncan MacCannell |
| EPI_ISL_1787259, EPI_ISL_1787260, EPI_ISL_1787261 | Broad Institute Clinical Research Sequencing Platform | Infectious Disease Program, Broad Institute of Harvard and MIT | Siddle,K.J., Adams,G., Pearlman,L., Gladden-Young,A., Vicente,G., Blumenstiel,B., DeFelice,M., Lee,M., McGovern,S., Lagerborg,K., Rudy,M., DeRuff,K., Carter,A., Normandin,E., Bauer,M., Reilly,S., Tomkins-Tinch,C., Loreth,C., Chaluvadi,S., Meldrim,J., Granger,B., Lemieux,J.E., Birren,B.W., Sabeti,P.C., Larkin,K., Dodge,S., Lennon,N., Madoff,L., Brown,C., Gallagher,G., Smole,S., Park,D.J., Gabriel,S., and MacInnis,B.L. |
| EPI_ISL_1789700, EPI_ISL_1789701, EPI_ISL_1789702, EPI_ISL_1789703, EPI_ISL_1789704, EPI_ISL_1789705 | NYC Pandemic Response Lab | Wadsworth Center, New York State Department of Health | Kirsten St. George, Daryl M. Lamson, Alexis Russell, Matthew Shudt, Melissa A Leisner, Jonathan Plitnick, Catharine Prussing, Navjot Singh, John Kelly, Erasmus Schneider, Erica Lasek-Nesselquist |
| EPI_ISL_1789706, EPI_ISL_1789707 | URMC LABS | Wadsworth Center, New York State Department of Health | Kirsten St. George, Daryl M. Lamson, Alexis Russell, Matthew Shudt, Melissa A Leisner, Jonathan Plitnick, Catharine Prussing, Navjot Singh, John Kelly, Erasmus Schneider, Erica Lasek-Nesselquist |
| EPI_ISL_1789708 | NYC Pandemic Response Lab | Wadsworth Center, New York State Department of Health | Kirsten St. George, Daryl M. Lamson, Alexis Russell, Matthew Shudt, Melissa A Leisner, Jonathan Plitnick, Catharine Prussing, Navjot Singh, John Kelly, Erasmus Schneider, Erica Lasek-Nesselquist |
| EPI_ISL_1789709 | SUNY UPSTATE MEDICAL UNIVERSITY | Wadsworth Center, New York State Department of Health | Kirsten St. George, Daryl M. Lamson, Alexis Russell, Matthew Shudt, Melissa A Leisner, Jonathan Plitnick, Catharine Prussing, Navjot Singh, John Kelly, Erasmus Schneider, Erica Lasek-Nesselquist |
| EPI_ISL_1789710, EPI_ISL_1789711 | MONTEFIORE MEDICAL CENTER LABORATORIES | Wadsworth Center, New York State Department of Health | Kirsten St. George, Daryl M. Lamson, Alexis Russell, Matthew Shudt, Melissa A Leisner, Jonathan Plitnick, Catharine Prussing, Navjot Singh, John Kelly, Erasmus Schneider, Erica Lasek-Nesselquist |
| EPI_ISL_1789712 | BOSTON HEART DIAGNOSTICS CORP | Wadsworth Center, New York State Department of Health | Kirsten St. George, Daryl M. Lamson, Alexis Russell, Matthew Shudt, Melissa A Leisner, Jonathan Plitnick, Catharine Prussing, Navjot Singh, John Kelly, Erasmus Schneider, Erica Lasek-Nesselquist |
| EPI_ISL_1789713 | Columbia University Irving Medical Center | Wadsworth Center, New York State Department of Health | Kirsten St. George, Daryl M. Lamson, Alexis Russell, Matthew Shudt, Melissa A Leisner, Jonathan Plitnick, Catharine Prussing, Navjot Singh, John Kelly, Erasmus Schneider, Erica Lasek-Nesselquist |
| EPI_ISL_1789714 | New York Presbyterian Hospital | Wadsworth Center, New York State Department of Health | Kirsten St. George, Daryl M. Lamson, Alexis Russell, Matthew Shudt, Melissa A Leisner, Jonathan Plitnick, Catharine Prussing, Navjot Singh, John Kelly, Erasmus Schneider, Erica Lasek-Nesselquist |
| EPI_ISL_1789715 | MONTEFIORE MEDICAL CENTER LABORATORIES | Wadsworth Center, New York State Department of Health | Kirsten St. George, Daryl M. Lamson, Alexis Russell, Matthew Shudt, Melissa A Leisner, Jonathan Plitnick, Catharine Prussing, Navjot Singh, John Kelly, Erasmus Schneider, Erica Lasek-Nesselquist |
| EPI_ISL_1789716, EPI_ISL_1789717, EPI_ISL_1789718 | ALBANY MEDICAL CENTER | Wadsworth Center, New York State Department of Health | Kirsten St. George, Daryl M. Lamson, Alexis Russell, Matthew Shudt, Melissa A Leisner, Jonathan Plitnick, Catharine Prussing, Navjot Singh, John Kelly, Erasmus Schneider, Erica Lasek-Nesselquist |
| EPI_ISL_1789719 | STONY BROOK UNIVERSITY HOSPITAL | Wadsworth Center, New York State Department of Health | Kirsten St. George, Daryl M. Lamson, Alexis Russell, Matthew Shudt, Melissa A Leisner, Jonathan Plitnick, Catharine Prussing, Navjot Singh, John Kelly, Erasmus Schneider, Erica Lasek-Nesselquist |
| EPI_ISL_1789720, EPI_ISL_1789721 | MONTEFIORE MEDICAL CENTER LABORATORIES | Wadsworth Center, New York State Department of Health | Kirsten St. George, Daryl M. Lamson, Alexis Russell, Matthew Shudt, Melissa A Leisner, Jonathan Plitnick, Catharine Prussing, Navjot Singh, John Kelly, Erasmus Schneider, Erica Lasek-Nesselquist |
| EPI_ISL_1789722, EPI_ISL_1789723 | WESTCHESTER MEDICAL CENTER | Wadsworth Center, New York State Department of Health | Kirsten St. George, Daryl M. Lamson, Alexis Russell, Matthew Shudt, Melissa A Leisner, Jonathan Plitnick, Catharine Prussing, Navjot Singh, John Kelly, Erasmus Schneider, Erica Lasek-Nesselquist |
| EPI_ISL_1789854, EPI_ISL_1789855, EPI_ISL_1789857, EPI_ISL_1789858, EPI_ISL_1789859, EPI_ISL_1789860, EPI_ISL_1789861, EPI_ISL_1789862, EPI_ISL_1789864, EPI_ISL_1789865, EPI_ISL_1789866, EPI_ISL_1789868, EPI_ISL_1789869, EPI_ISL_1789870, EPI_ISL_1789871, EPI_ISL_1789872, EPI_ISL_1789873, EPI_ISL_1789874, EPI_ISL_1789875, EPI_ISL_1789877, EPI_ISL_1789878, EPI_ISL_1789879, EPI_ISL_1789880, EPI_ISL_1789881, EPI_ISL_1789882, EPI_ISL_1789883, EPI_ISL_1789884, EPI_ISL_1789885, EPI_ISL_1789886, EPI_ISL_1789887, EPI_ISL_1789888, EPI_ISL_1789889, EPI_ISL_1789890, EPI_ISL_1789891, EPI_ISL_1789892, EPI_ISL_1789893, EPI_ISL_1789894, EPI_ISL_1789895, EPI_ISL_1789896, EPI_ISL_1789897, EPI_ISL_1789898, EPI_ISL_1789899, EPI_ISL_1789901, EPI_ISL_1789902, EPI_ISL_1789903, EPI_ISL_1789904, EPI_ISL_1789905, EPI_ISL_1789906, EPI_ISL_1789907, EPI_ISL_1789908, EPI_ISL_1789909, EPI_ISL_1789910, EPI_ISL_1789911, EPI_ISL_1789912, EPI_ISL_1789913, EPI_ISL_1789914, EPI_ISL_1789915, EPI_ISL_1789916, EPI_ISL_1789917, EPI_ISL_1789918, EPI_ISL_1789919, EPI_ISL_1789920, EPI_ISL_1789921, EPI_ISL_1789922, EPI_ISL_1789923, EPI_ISL_1789924, EPI_ISL_1789925, EPI_ISL_1789926, EPI_ISL_1789928, EPI_ISL_1789929, EPI_ISL_1789930, EPI_ISL_1789931, EPI_ISL_1789932, EPI_ISL_1789933, EPI_ISL_1789934, EPI_ISL_1789935, EPI_ISL_1789936, EPI_ISL_1789938, EPI_ISL_1789939, EPI_ISL_1789940, EPI_ISL_1789941, EPI_ISL_1789942, EPI_ISL_1789943, EPI_ISL_1789944, EPI_ISL_1789945, EPI_ISL_1789946, EPI_ISL_1789947, EPI_ISL_1789948, EPI_ISL_1789949, EPI_ISL_1789950, EPI_ISL_1789951, EPI_ISL_1789952, EPI_ISL_1789953, EPI_ISL_1789954, EPI_ISL_1789955, EPI_ISL_1789956, EPI_ISL_1789957, EPI_ISL_1789958, EPI_ISL_1789959, EPI_ISL_1789960, EPI_ISL_1789961, EPI_ISL_1789962, EPI_ISL_1789963, EPI_ISL_1789964, EPI_ISL_1789966, EPI_ISL_1789967, EPI_ISL_1789968, EPI_ISL_1789969, EPI_ISL_1789970, EPI_ISL_1789971, EPI_ISL_1789972, EPI_ISL_1789973, EPI_ISL_1789974, EPI_ISL_1789975, EPI_ISL_1789976, EPI_ISL_1789977, EPI_ISL_1789978, EPI_ISL_1789979, EPI_ISL_1789980, EPI_ISL_1789981, EPI_ISL_1789982, EPI_ISL_1789983, EPI_ISL_1789984, EPI_ISL_1789985, EPI_ISL_1789986, EPI_ISL_1789987, EPI_ISL_1789988, EPI_ISL_1789989, EPI_ISL_1789990, EPI_ISL_1789991, EPI_ISL_1789992, EPI_ISL_1789993, EPI_ISL_1789994, EPI_ISL_1789995, EPI_ISL_1789996, EPI_ISL_1789997, EPI_ISL_1789998, EPI_ISL_1789999, EPI_ISL_1790000, EPI_ISL_1790001, EPI_ISL_1790003, EPI_ISL_1790004, EPI_ISL_1790005, EPI_ISL_1790006, EPI_ISL_1790007, EPI_ISL_1790009, EPI_ISL_1790010, EPI_ISL_1790011, EPI_ISL_1790012, EPI_ISL_1790013, EPI_ISL_1790014, EPI_ISL_1790015, EPI_ISL_1790016, EPI_ISL_1790017, EPI_ISL_1790019, EPI_ISL_1790020, EPI_ISL_1790021, EPI_ISL_1790022, EPI_ISL_1790023, EPI_ISL_1790024, EPI_ISL_1790025, EPI_ISL_1790027, EPI_ISL_1790028, EPI_ISL_1790029, EPI_ISL_1790030, EPI_ISL_1790031, EPI_ISL_1790032, EPI_ISL_1790033, EPI_ISL_1790034, EPI_ISL_1790035, EPI_ISL_1790036, EPI_ISL_1790037, EPI_ISL_1790038, EPI_ISL_1790039, EPI_ISL_1790040, EPI_ISL_1790041, EPI_ISL_1790042, EPI_ISL_1790043, EPI_ISL_1790044, EPI_ISL_1790045, EPI_ISL_1790046, EPI_ISL_1790047 | | | |
| see above | NYU Langone Health | Departments of Pathology and Medicine, New York University School of Medicine | Adriana Heguy, Dacia Dimartino, Emily Guzman, Christian Marier, Peter Meyn, Sitharam Ramaswami, Gael Westby, Paul Zappile, Yutong Zhang, Paolo Cotzia, Guiqing Wang |
| EPI_ISL_1793502 | Yale Clinical Virology Lab | Yale Center for Genomic Analysis | Shrikant Mane, Kaya Bilguvar, Curt Scharfe, Irina Tikhonova, Christopher Castaldi, Brooke Sullivan |
| EPI_ISL_1795506, EPI_ISL_1795589, EPI_ISL_1795597, EPI_ISL_1795605, EPI_ISL_1795615, EPI_ISL_1795620, EPI_ISL_1795621, EPI_ISL_1795622 | Infinity Biologix | Centers for Disease Control and Prevention Division of Viral Diseases, Pathogen Discovery | Dakota Howard, Dhwani Batra, Peter W. Cook, Kara Moser, Adrian Paskey, Jason Caravas, Benjamin Rambo-Martin, Shatavia Morrison, Christopher Gulvick, Scott Sammons, Yvette Unoarumhi, Darlene Wagner, Matthew Schmerer, Christian Bixby, Yihe Wang, Jonathan Schultz, Chirayu Goswami, Russ Hager, Robin Grimwood, Clinton R. Paden, Duncan MacCannell |
| EPI_ISL_1796130, EPI_ISL_1796436, EPI_ISL_1796534, EPI_ISL_1796840, EPI_ISL_1797051, EPI_ISL_1797367 | Helix/Illumina | Centers for Disease Control and Prevention Division of Viral Diseases, Pathogen Discovery | Dakota Howard, Dhwani Batra, Peter W. Cook, Kara Moser, Adrian Paskey, Jason Caravas, Benjamin Rambo-Martin, Shatavia Morrison, Christopher Gulvick, Scott Sammons, Yvette Unoarumhi, Darlene Wagner, Matthew Schmerer, Eileen de Feo, Jan Antico, Christine Tran, Matthew Tolentino, Shannon Wickline, Kim Gietzen, Brad Sickler, Jingtao Liu, Eric Allen, Phil Febbo, Nicole L. Washington, Simon White, Geraint Levan, Kelly Schiabor Barrett, Elizabeth Cirulli, Alexandre Bolze, Ary Ascencio, Charlotte Rivera-Garcia, Ryan Cho, Jason Nguyen, Sherry Wang, Jimmy Ramirez, Tyler Cassens, Efren Sandoval, Magnus Isaksson, William Lee, David Becker, Marc Laurent, James Lu, Clinton R. Paden, Duncan MacCannell |
| EPI_ISL_1798049, EPI_ISL_1798098, EPI_ISL_1798112, EPI_ISL_1798113, EPI_ISL_1798146, EPI_ISL_1798164 | Quest Diagnostics Incorporated | Centers for Disease Control and Prevention Division of Viral Diseases, Pathogen Discovery | Dakota Howard, Dhwani Batra, Peter W. Cook, Kara Moser, Adrian Paskey, Jason Caravas, Benjamin Rambo-Martin, Shatavia Morrison, Christopher Gulvick, Scott Sammons, Yvette Unoarumhi, Darlene Wagner, Matthew Schmerer, S. H. Rosenthal, A. Gerasimova, R. M. Kagan, B. Anderson, M. Hua, Y. Liu, L.E. Bernstein, K.E. Livingston, A. Perez, I. A. Shlyakhter, R. V. Rolando, R. Owen, P. Tanpaiboon, F. Lacbawan, Clinton R. Paden, Duncan MacCannell |
| EPI_ISL_1799429, EPI_ISL_1799495, EPI_ISL_1799693, EPI_ISL_1799740, EPI_ISL_1799741, EPI_ISL_1799744, EPI_ISL_1799746, EPI_ISL_1799747, EPI_ISL_1799769, EPI_ISL_1799781, EPI_ISL_1799784, EPI_ISL_1799790, EPI_ISL_1799840, EPI_ISL_1799841, EPI_ISL_1799842, EPI_ISL_1799877, EPI_ISL_1799938, EPI_ISL_1799950, EPI_ISL_1799970, EPI_ISL_1799974, EPI_ISL_1799975, EPI_ISL_1799980, EPI_ISL_1799983, EPI_ISL_1799984, EPI_ISL_1799985, EPI_ISL_1799986, EPI_ISL_1799992, EPI_ISL_1800344, EPI_ISL_1800349, EPI_ISL_1800355, EPI_ISL_1800357, EPI_ISL_1800359, EPI_ISL_1800402, EPI_ISL_1800404, EPI_ISL_1800405, EPI_ISL_1800406, EPI_ISL_1800407, EPI_ISL_1800409, EPI_ISL_1800413, EPI_ISL_1800455, EPI_ISL_1800457, EPI_ISL_1800460, EPI_ISL_1800461, EPI_ISL_1800470, EPI_ISL_1800471, EPI_ISL_1800472, EPI_ISL_1800473, EPI_ISL_1800475, EPI_ISL_1800479, EPI_ISL_1800488, EPI_ISL_1800489, EPI_ISL_1800490, EPI_ISL_1800491, EPI_ISL_1800492, EPI_ISL_1800513, EPI_ISL_1800515, EPI_ISL_1800516, EPI_ISL_1800517, EPI_ISL_1800527, EPI_ISL_1800533, EPI_ISL_1800541, EPI_ISL_1800542, EPI_ISL_1800666, EPI_ISL_1800695, EPI_ISL_1800701, EPI_ISL_1800709, EPI_ISL_1800747, EPI_ISL_1800753, EPI_ISL_1800765, EPI_ISL_1800766, EPI_ISL_1800773, EPI_ISL_1800774, EPI_ISL_1800778, EPI_ISL_1800780, EPI_ISL_1800781, EPI_ISL_1800799, EPI_ISL_1800816, EPI_ISL_1800817, EPI_ISL_1800822, EPI_ISL_1800824, EPI_ISL_1800825, EPI_ISL_1800828, EPI_ISL_1800829, EPI_ISL_1800859, EPI_ISL_1800860, EPI_ISL_1800866, EPI_ISL_1800889, EPI_ISL_1800891, EPI_ISL_1800892, EPI_ISL_1801293, EPI_ISL_1801320, EPI_ISL_1801321, EPI_ISL_1801323, EPI_ISL_1801324, EPI_ISL_1801325, EPI_ISL_1801327, EPI_ISL_1801403, EPI_ISL_1801437, EPI_ISL_1801487, EPI_ISL_1801488, EPI_ISL_1801507, EPI_ISL_1801508, EPI_ISL_1801509, EPI_ISL_1801510, EPI_ISL_1801513, EPI_ISL_1801514, EPI_ISL_1801516, EPI_ISL_1801517, EPI_ISL_1801518, EPI_ISL_1801519, EPI_ISL_1801520, EPI_ISL_1801521, EPI_ISL_1801522, EPI_ISL_1801523, EPI_ISL_1801524, EPI_ISL_1801525, EPI_ISL_1801526, EPI_ISL_1801527, EPI_ISL_1801528, EPI_ISL_1801574, EPI_ISL_1801575, EPI_ISL_1801576, EPI_ISL_1801577, EPI_ISL_1801579, EPI_ISL_1801580, EPI_ISL_1801609, EPI_ISL_1801610, EPI_ISL_1801611, EPI_ISL_1801612, EPI_ISL_1801634, EPI_ISL_1801663, EPI_ISL_1801664, EPI_ISL_1801666, EPI_ISL_1801667, EPI_ISL_1801668, EPI_ISL_1801669, EPI_ISL_1801670, EPI_ISL_1801671, EPI_ISL_1801672, EPI_ISL_1801673, EPI_ISL_1801762, EPI_ISL_1801846, EPI_ISL_1801852, EPI_ISL_1801860, EPI_ISL_1801866, EPI_ISL_1802295, EPI_ISL_1802304, EPI_ISL_1802307, EPI_ISL_1802310, EPI_ISL_1802312, EPI_ISL_1802324, EPI_ISL_1802512, EPI_ISL_1802513, EPI_ISL_1802574, EPI_ISL_1802576, EPI_ISL_1802663, EPI_ISL_1802664 | | | |
| see above | Laboratory Corporation of America | Centers for Disease Control and Prevention Division of Viral Diseases, Pathogen Discovery | Dakota Howard, Dhwani Batra, Peter W. Cook, Kara Moser, Adrian Paskey, Jason Caravas, Benjamin Rambo-Martin, Shatavia Morrison, Christopher Gulvick, Scott Sammons, Yvette Unoarumhi, Darlene Wagner, Matthew Schmerer, Minoo Agarwal, Eyad Almasri, Debbie Boles, Ayla Burns, Nuthawin Charoensri, Oren Cohen, Susan Countryman, Mary Ann Cristobal, Bobbi Croy, Suzanne Dale, Hrushikesh Deshmukh, Amanda Douglas, Vincent Drouillon, Marcia Eisenberg, Howard Engler, Rama Ghatti, Prashant Gupta, Susan Hicks, Jake Humphrey, Lax Iyer, Manoj Jain, Mohan Kolli, Brian Krueger, Tim Kuphal, Stanley Letovsky, Michael Levandoski, Craig Lukasik, Jonathan Meltzer, Brian Norvell, Mindy Nye, Scott Parker, Christos Petropoulos, John Pruitt, Steven Ragan, Scott Ryan, Mike Sapeta, Jana Schroth, Suresh Babu Selvaraju, Goran Stevovic, Amanda Suchanek, Andrea Throop, Lyndon Tilson, Thomas Urban, Joe Voshell, Kimberly Wagner, Jonathan Williams, Mary Williamson, Qian Zeng, Tricia Zwiefelhofer, Clinton R. Paden, Duncan MacCannell |

| | | | |
|---|---|---|---|
| EPI_ISL_1802835 | Infinity Biologix | Centers for Disease Control and Prevention Division of Viral Diseases, Pathogen Discovery | Dakota Howard, Dhwani Batra, Peter W. Cook, Kara Moser, Adrian Paskey, Jason Caravas, Benjamin Rambo-Martin, Shatavia Morrison, Christopher Gulvick, Scott Sammons, Yvette Unoarumhi, Darlene Wagner, Matthew Schmerer, Christian Bixby, Yihe Wang, Jonathan Schultz, Chirayu Goswami, Russ Hager, Robin Grimwood, Clinton R. Paden, Duncan MacCannell |
| EPI_ISL_1803619, EPI_ISL_1803731, EPI_ISL_1803819 | Helix/Illumina | Centers for Disease Control and Prevention Division of Viral Diseases, Pathogen Discovery | Dakota Howard, Dhwani Batra, Peter W. Cook, Kara Moser, Adrian Paskey, Jason Caravas, Benjamin Rambo-Martin, Shatavia Morrison, Christopher Gulvick, Scott Sammons, Yvette Unoarumhi, Darlene Wagner, Matthew Schmerer, Eileen de Feo, Jan Antico, Christine Tran, Matthew Tolentino, Shannon Wickline, Kim Gietzen, Brad Sickler, Jingtao Liu, Eric Allen, Phil Febbo, Nicole L. Washington, Simon White, Geraint Levan, Kelly Schiabor Barrett, Elizabeth Cirulli, Alexandre Bolze, Ary Ascencio, Charlotte Rivera-Garcia, Ryan Cho, Jason Nguyen, Sherry Wang, Jimmy Ramirez, Tyler Cassens, Efren Sandoval, Magnus Isaksson, William Lee, David Becker, Marc Laurent, James Lu, Clinton R. Paden, Duncan MacCannell |
| EPI_ISL_1804820 | Quest Diagnostics Incorporated | Centers for Disease Control and Prevention Division of Viral Diseases, Pathogen Discovery | Dakota Howard, Dhwani Batra, Peter W. Cook, Kara Moser, Adrian Paskey, Jason Caravas, Benjamin Rambo-Martin, Shatavia Morrison, Christopher Gulvick, Scott Sammons, Yvette Unoarumhi, Darlene Wagner, Matthew Schmerer, S. H. Rosenthal, A. Gerasimova, R. M. Kagan, B. Anderson, M. Hua, Y. Liu, L.E. Bernstein, K.E. Livingston, A. Perez, I. A. Shlyakhter, R. V. Rolando, R. Owen, P. Tanpaiboon, F. Lacbawan, Clinton R. Paden, Duncan MacCannell |
| EPI_ISL_1804941 | Department of Homeless Services | New York City Public Health Laboratory | Jade Wang, et al. |
| EPI_ISL_1804942 | DOHMH Jamaica | New York City Public Health Laboratory | Jade Wang, et al. |
| EPI_ISL_1804943 | DOHMH Corona | New York City Public Health Laboratory | Jade Wang, et al. |
| EPI_ISL_1804944 | DOHMH PHL | New York City Public Health Laboratory | Jade Wang, et al. |
| EPI_ISL_1804945 | DOHMH Central Harlem | New York City Public Health Laboratory | Jade Wang, et al. |
| EPI_ISL_1804946 | DOHMH Chelsea | New York City Public Health Laboratory | Jade Wang, et al. |
| EPI_ISL_1804947 | Department of Homeless Services | New York City Public Health Laboratory | Jade Wang, et al. |
| EPI_ISL_1804948, EPI_ISL_1804949 | OCME Office Of Chief Medical Examiner | New York City Public Health Laboratory | Jade Wang, et al. |
| EPI_ISL_1804950 | DOHMH Chelsea | New York City Public Health Laboratory | Jade Wang, et al. |
| EPI_ISL_1804951 | DOHMH PHL | New York City Public Health Laboratory | Jade Wang, et al. |
| EPI_ISL_1804952, EPI_ISL_1804953 | DOHMH Crown Heights | New York City Public Health Laboratory | Jade Wang, et al. |
| EPI_ISL_1804954, EPI_ISL_1804955 | DOHMH Central Harlem | New York City Public Health Laboratory | Jade Wang, et al. |
| EPI_ISL_1804956 | DOHMH Morrisania | New York City Public Health Laboratory | Jade Wang, et al. |
| EPI_ISL_1804957 | DOHMH Corona | New York City Public Health Laboratory | Jade Wang, et al. |
| EPI_ISL_1804958, EPI_ISL_1804959 | DOHMH Jamaica | New York City Public Health Laboratory | Jade Wang, et al. |
| EPI_ISL_1804960 | DOHMH Corona | New York City Public Health Laboratory | Jade Wang, et al. |
| EPI_ISL_1804961 | OCME Office Of Chief Medical Examiner | New York City Public Health Laboratory | Jade Wang, et al. |
| EPI_ISL_1817992 | Infinity Biologix | Centers for Disease Control and Prevention Division of Viral Diseases, Pathogen Discovery | Dakota Howard, Dhwani Batra, Peter W. Cook, Kara Moser, Adrian Paskey, Jason Caravas, Benjamin Rambo-Martin, Shatavia Morrison, Christopher Gulvick, Scott Sammons, Yvette Unoarumhi, Darlene Wagner, Matthew Schmerer, Christian Bixby, Yihe Wang, Jonathan Schultz, Chirayu Goswami, Russ Hager, Robin Grimwood, Clinton R. Paden, Duncan MacCannell |
| EPI_ISL_1818070, EPI_ISL_1818295, EPI_ISL_1818327, EPI_ISL_1818331, EPI_ISL_1818346, EPI_ISL_1818349, EPI_ISL_1818401, EPI_ISL_1818422, EPI_ISL_1818519 | Quest Diagnostics Incorporated | Centers for Disease Control and Prevention Division of Viral Diseases, Pathogen Discovery | Dakota Howard, Dhwani Batra, Peter W. Cook, Kara Moser, Adrian Paskey, Jason Caravas, Benjamin Rambo-Martin, Shatavia Morrison, Christopher Gulvick, Scott Sammons, Yvette Unoarumhi, Darlene Wagner, Matthew Schmerer, S. H. Rosenthal, A. Gerasimova, R. M. Kagan, B. Anderson, M. Hua, Y. Liu, L.E. Bernstein, K.E. Livingston, A. Perez, I. A. Shlyakhter, R. V. Rolando, R. Owen, P. Tanpaiboon, F. Lacbawan, Clinton R. Paden, Duncan MacCannell |
| EPI_ISL_1823944, EPI_ISL_1823945, EPI_ISL_1823946, EPI_ISL_1823947, EPI_ISL_1823948, EPI_ISL_1823949, EPI_ISL_1823950, EPI_ISL_1823951, EPI_ISL_1823952, EPI_ISL_1823953, EPI_ISL_1823954, EPI_ISL_1823955, EPI_ISL_1823956, EPI_ISL_1823957, EPI_ISL_1823958, EPI_ISL_1823959, EPI_ISL_1823960, EPI_ISL_1823961, EPI_ISL_1823962, EPI_ISL_1823963, EPI_ISL_1823964, EPI_ISL_1823965, EPI_ISL_1823966, EPI_ISL_1823967, EPI_ISL_1823968, EPI_ISL_1823969, EPI_ISL_1823970, EPI_ISL_1823971, EPI_ISL_1823972, EPI_ISL_1823973 | | | |
| see above | NYC Department of Health and Mental Hygiene | Centers for Disease Control and Prevention Division of Viral Diseases, Pathogen Discovery | Mili Sheth, Sarah Nobles, Jasmine Padilla, Mark Burroughs, Shoshona Le, Katie Dillon, Peter Cook, Clinton R. Paden, Dhwani Batra, Krista Queen, Kristen Knipe, Dakota Howard, Yvette Unoarumhi, Darlene Wagner, Matthew Schmerer, Ben L. Rambo-Martin, Kristine Lacek, Sam Shepard, Alison Laufer Halpin, Dave Wentworth, Vivien Dugan, Suxiang Tong, Justin Lee |
| EPI_ISL_1824033, EPI_ISL_1824034, EPI_ISL_1824035 | "NYSDOH Wadsworth Center, Virology Lab" | Centers for Disease Control and Prevention Division of Viral Diseases, Pathogen Discovery | Mili Sheth, Sarah Nobles, Jasmine Padilla, Mark Burroughs, Shoshona Le, Katie Dillon, Peter Cook, Clinton R. Paden, Dhwani Batra, Krista Queen, Kristen Knipe, Dakota Howard, Yvette Unoarumhi, Darlene Wagner, Matthew Schmerer, Ben L. Rambo-Martin, Kristine Lacek, Sam Shepard, Alison Laufer Halpin, Dave Wentworth, Vivien Dugan, Suxiang Tong, Justin Lee |
| EPI_ISL_1825233, EPI_ISL_1825481, EPI_ISL_1825483, EPI_ISL_1825484, EPI_ISL_1825487, EPI_ISL_1825488, EPI_ISL_1825492, EPI_ISL_1825494 | Broad Institute Clinical Research Sequencing Platform | Infectious Disease Program, Broad Institute of Harvard and MIT | Siddle,K.J., Adams,G., Pearlman,L., Gladden-Young,A., Vicente,G., Blumenstiel,B., DeFelice,M., Lee,M., McGovern,S., Lagerborg,K., Rudy,M., DeRuff,K., Carter,A., Normandin,E., Bauer,M., Reilly,S., Tomkins-Tinch,C., Loreth,C., Chaluvadi,S., Meldrim,J., Granger,B., Lemieux,J.E., Birren,B.W., Sabeti,P.C., Larkin,K., Dodge,S., Lennon,N., Madoff,L., Brown,C., Gallagher,G., Smole,S., Park,D.J., Gabriel,S., and MacInnis,B.L. |
| EPI_ISL_1827954, EPI_ISL_1827955, EPI_ISL_1827956, EPI_ISL_1827957, EPI_ISL_1827958, EPI_ISL_1827959, EPI_ISL_1827960, EPI_ISL_1827961, EPI_ISL_1827962, EPI_ISL_1827963, EPI_ISL_1827964, EPI_ISL_1827965, EPI_ISL_1827966, EPI_ISL_1827967, EPI_ISL_1827968, EPI_ISL_1827969, EPI_ISL_1827970, EPI_ISL_1827971, EPI_ISL_1827972, EPI_ISL_1827973, EPI_ISL_1827974, EPI_ISL_1827975, EPI_ISL_1827976, EPI_ISL_1827977, EPI_ISL_1827978, EPI_ISL_1827979, EPI_ISL_1827980, EPI_ISL_1827981, EPI_ISL_1827982, EPI_ISL_1827983, EPI_ISL_1827984, EPI_ISL_1827985, EPI_ISL_1827986, EPI_ISL_1827987, EPI_ISL_1827988, EPI_ISL_1827989, EPI_ISL_1827990, EPI_ISL_1827991, EPI_ISL_1827992, EPI_ISL_1827993, EPI_ISL_1827994, EPI_ISL_1827995, EPI_ISL_1827996, EPI_ISL_1827997, EPI_ISL_1827998, EPI_ISL_1827999, EPI_ISL_1828000, EPI_ISL_1828001, EPI_ISL_1828002, EPI_ISL_1828003, EPI_ISL_1828004, EPI_ISL_1828005, EPI_ISL_1828006, EPI_ISL_1828007, EPI_ISL_1828008, EPI_ISL_1828009, EPI_ISL_1828010, EPI_ISL_1828011, EPI_ISL_1828012, EPI_ISL_1828013, EPI_ISL_1828014, EPI_ISL_1828015, EPI_ISL_1828016, EPI_ISL_1828017, EPI_ISL_1828018, EPI_ISL_1828019, EPI_ISL_1828020, EPI_ISL_1828021, EPI_ISL_1828022, EPI_ISL_1828023, EPI_ISL_1828024, EPI_ISL_1828025, EPI_ISL_1828026, EPI_ISL_1828027, EPI_ISL_1828028, EPI_ISL_1828029, EPI_ISL_1828030, EPI_ISL_1828031, EPI_ISL_1828032, EPI_ISL_1828033, EPI_ISL_1828034, EPI_ISL_1828035, EPI_ISL_1828036, EPI_ISL_1828037, EPI_ISL_1828038, EPI_ISL_1828039, EPI_ISL_1828040, EPI_ISL_1828041, EPI_ISL_1828042, EPI_ISL_1828043, EPI_ISL_1828044, EPI_ISL_1828045, EPI_ISL_1828046, EPI_ISL_1828047, EPI_ISL_1828048, EPI_ISL_1828049, EPI_ISL_1828050, EPI_ISL_1828051, EPI_ISL_1828052, EPI_ISL_1828053, EPI_ISL_1828054, EPI_ISL_1828055, EPI_ISL_1828056, EPI_ISL_1828057, EPI_ISL_1828058, EPI_ISL_1828059, EPI_ISL_1828060, EPI_ISL_1828061, EPI_ISL_1828062, EPI_ISL_1828063, EPI_ISL_1828064, EPI_ISL_1828065, EPI_ISL_1828066, EPI_ISL_1828067, EPI_ISL_1828068, EPI_ISL_1828069, EPI_ISL_1828070, EPI_ISL_1828071, EPI_ISL_1828072, EPI_ISL_1828073, EPI_ISL_1828074, EPI_ISL_1828075, EPI_ISL_1828076, EPI_ISL_1828077, EPI_ISL_1828078, EPI_ISL_1828079, EPI_ISL_1828080, EPI_ISL_1828081, EPI_ISL_1828082, EPI_ISL_1828083, EPI_ISL_1828084, EPI_ISL_1828085, EPI_ISL_1828086, EPI_ISL_1828087, EPI_ISL_1828088, EPI_ISL_1828089, EPI_ISL_1828090, EPI_ISL_1828091, EPI_ISL_1828092, EPI_ISL_1828093, EPI_ISL_1828094, EPI_ISL_1828095, EPI_ISL_1828096, EPI_ISL_1828097, EPI_ISL_1828098, EPI_ISL_1828099, EPI_ISL_1828100, EPI_ISL_1828101, EPI_ISL_1828102, EPI_ISL_1828103, EPI_ISL_1828104, EPI_ISL_1828105, EPI_ISL_1828106, EPI_ISL_1828107, EPI_ISL_1828108, EPI_ISL_1828109, EPI_ISL_1828110, EPI_ISL_1828111, EPI_ISL_1828112, EPI_ISL_1828113, EPI_ISL_1828114, EPI_ISL_1828115, EPI_ISL_1828116, EPI_ISL_1828117, EPI_ISL_1828118, EPI_ISL_1828119, EPI_ISL_1828120, EPI_ISL_1828121, EPI_ISL_1828122, EPI_ISL_1828123, EPI_ISL_1828124, EPI_ISL_1828125, EPI_ISL_1828126, EPI_ISL_1828127, EPI_ISL_1828128, EPI_ISL_1828129, EPI_ISL_1828130, EPI_ISL_1828131, EPI_ISL_1828132, EPI_ISL_1828133, EPI_ISL_1828134, EPI_ISL_1828135, EPI_ISL_1828136, EPI_ISL_1828137, EPI_ISL_1828138, EPI_ISL_1828139, EPI_ISL_1828140, EPI_ISL_1828141, EPI_ISL_1828142, EPI_ISL_1828143, EPI_ISL_1828144, EPI_ISL_1828145, EPI_ISL_1828146, EPI_ISL_1828147, EPI_ISL_1828148, EPI_ISL_1828149, EPI_ISL_1828150, EPI_ISL_1828151, EPI_ISL_1828152, EPI_ISL_1828153, EPI_ISL_1828154, EPI_ISL_1828155, EPI_ISL_1828156, EPI_ISL_1828157, EPI_ISL_1828158, EPI_ISL_1828159, EPI_ISL_1828160, EPI_ISL_1828161, EPI_ISL_1828162, EPI_ISL_1828163, EPI_ISL_1828164, EPI_ISL_1828165, EPI_ISL_1828166, EPI_ISL_1828167, EPI_ISL_1828168, EPI_ISL_1828169, EPI_ISL_1828170, EPI_ISL_1828171, EPI_ISL_1828172, EPI_ISL_1828173, EPI_ISL_1828174, EPI_ISL_1828175, EPI_ISL_1828176, EPI_ISL_1828177, EPI_ISL_1828178, EPI_ISL_1828179, EPI_ISL_1828180, EPI_ISL_1828181, EPI_ISL_1828182, EPI_ISL_1828183, EPI_ISL_1828184, EPI_ISL_1828185, EPI_ISL_1828186, EPI_ISL_1828187, EPI_ISL_1828188, EPI_ISL_1828189, EPI_ISL_1828190, EPI_ISL_1828191, EPI_ISL_1828192, EPI_ISL_1828193, EPI_ISL_1828194, EPI_ISL_1828195, EPI_ISL_1828196, EPI_ISL_1828197, EPI_ISL_1828198, EPI_ISL_1828199, EPI_ISL_1828200, EPI_ISL_1828201, EPI_ISL_1828202, EPI_ISL_1828203, EPI_ISL_1828204, EPI_ISL_1828205, EPI_ISL_1828206, EPI_ISL_1828207, EPI_ISL_1828208, EPI_ISL_1828209, EPI_ISL_1828210, EPI_ISL_1828211, EPI_ISL_1828212, EPI_ISL_1828213, EPI_ISL_1828214, EPI_ISL_1828215, EPI_ISL_1828216, EPI_ISL_1828217, EPI_ISL_1828218, EPI_ISL_1828219, EPI_ISL_1828220, EPI_ISL_1828221, EPI_ISL_1828222, EPI_ISL_1828223, EPI_ISL_1828224, EPI_ISL_1828225, EPI_ISL_1828226, EPI_ISL_1828227, EPI_ISL_1828228, EPI_ISL_1828229, EPI_ISL_1828230, EPI_ISL_1828231, EPI_ISL_1828232, EPI_ISL_1828233, EPI_ISL_1828234, EPI_ISL_1828235, EPI_ISL_1828236, EPI_ISL_1828237, EPI_ISL_1828238, EPI_ISL_1828239, EPI_ISL_1828240, EPI_ISL_1828241, EPI_ISL_1828242, EPI_ISL_1828243, EPI_ISL_1828244, EPI_ISL_1828245, EPI_ISL_1828246, EPI_ISL_1828247, EPI_ISL_1828248, EPI_ISL_1828249, EPI_ISL_1828250, EPI_ISL_1828251, EPI_ISL_1828252, EPI_ISL_1828253, EPI_ISL_1828254, EPI_ISL_1828255, EPI_ISL_1828256, EPI_ISL_1828257, EPI_ISL_1828258, EPI_ISL_1828259, EPI_ISL_1828260, EPI_ISL_1828261, EPI_ISL_1828262, EPI_ISL_1828263, EPI_ISL_1828264, EPI_ISL_1828265, EPI_ISL_1828266, EPI_ISL_1828267, EPI_ISL_1828268, EPI_ISL_1828269, EPI_ISL_1828270, EPI_ISL_1828271, EPI_ISL_1828272, EPI_ISL_1828273, EPI_ISL_1828274, EPI_ISL_1828275, EPI_ISL_1828276, EPI_ISL_1828277, EPI_ISL_1828278, EPI_ISL_1828279, EPI_ISL_1828280, EPI_ISL_1828281, EPI_ISL_1828282, EPI_ISL_1828283, EPI_ISL_1828284, EPI_ISL_1828285, EPI_ISL_1828286, EPI_ISL_1828287, EPI_ISL_1828288, EPI_ISL_1828289, EPI_ISL_1828290, EPI_ISL_1828291, EPI_ISL_1828292, EPI_ISL_1828293, EPI_ISL_1828294, EPI_ISL_1828295, EPI_ISL_1828296, EPI_ISL_1828297, EPI_ISL_1828298, EPI_ISL_1828299, EPI_ISL_1828300, EPI_ISL_1828301, EPI_ISL_1828302, EPI_ISL_1828303, EPI_ISL_1828304, EPI_ISL_1828305, EPI_ISL_1828306, EPI_ISL_1828307, EPI_ISL_1828308, EPI_ISL_1828309, EPI_ISL_1828310, EPI_ISL_1828311, EPI_ISL_1828312, EPI_ISL_1828313, EPI_ISL_1828314, EPI_ISL_1828315, EPI_ISL_1828316, EPI_ISL_1828317, EPI_ISL_1828318, EPI_ISL_1828319, EPI_ISL_1828320, EPI_ISL_1828321, EPI_ISL_1828322, EPI_ISL_1828323, EPI_ISL_1828324, EPI_ISL_1828325, EPI_ISL_1828326, EPI_ISL_1828327, EPI_ISL_1828328, EPI_ISL_1828329, EPI_ISL_1828330, EPI_ISL_1828331, EPI_ISL_1828332, EPI_ISL_1828333, EPI_ISL_1828334, EPI_ISL_1828335, EPI_ISL_1828336, EPI_ISL_1828337, EPI_ISL_1828338, EPI_ISL_1828339, EPI_ISL_1828340, EPI_ISL_1828341, EPI_ISL_1828342, EPI_ISL_1828343, EPI_ISL_1828344, EPI_ISL_1828345, EPI_ISL_1828346, EPI_ISL_1828347, EPI_ISL_1828348, EPI_ISL_1828349, EPI_ISL_1828350, EPI_ISL_1828351, EPI_ISL_1828352, EPI_ISL_1828353, EPI_ISL_1828354, EPI_ISL_1828355, EPI_ISL_1828356, EPI_ISL_1828357, EPI_ISL_1828358, EPI_ISL_1828359, EPI_ISL_1828360, EPI_ISL_1828361, EPI_ISL_1828362, EPI_ISL_1828363, EPI_ISL_1828364, EPI_ISL_1828365, EPI_ISL_1828366, EPI_ISL_1828367, EPI_ISL_1828368, EPI_ISL_1828369, EPI_ISL_1828370, EPI_ISL_1828371, EPI_ISL_1828372, EPI_ISL_1828373, EPI_ISL_1828374, EPI_ISL_1828375, EPI_ISL_1828376, EPI_ISL_1828377, EPI_ISL_1828378, | | | |

| | | | |
|---|---|---|---|
| EPI_ISL_1828379, EPI_ISL_1828380, EPI_ISL_1828381, EPI_ISL_1828382, EPI_ISL_1828383, EPI_ISL_1828384, EPI_ISL_1828385, EPI_ISL_1828386, EPI_ISL_1828387, EPI_ISL_1828388, EPI_ISL_1828389, EPI_ISL_1828390, EPI_ISL_1828391, EPI_ISL_1828392, EPI_ISL_1828393, EPI_ISL_1828394, EPI_ISL_1828395, EPI_ISL_1828396, EPI_ISL_1828397, EPI_ISL_1828398, EPI_ISL_1828399, EPI_ISL_1828400, EPI_ISL_1828401, EPI_ISL_1828402, EPI_ISL_1828403, EPI_ISL_1828404, EPI_ISL_1828405, EPI_ISL_1828406, EPI_ISL_1828407, EPI_ISL_1828408, EPI_ISL_1828409, EPI_ISL_1828410, EPI_ISL_1828411, EPI_ISL_1828412, EPI_ISL_1828413, EPI_ISL_1828414, EPI_ISL_1828415, EPI_ISL_1828416, EPI_ISL_1828417, EPI_ISL_1828418, EPI_ISL_1828419, EPI_ISL_1828420, EPI_ISL_1828421, EPI_ISL_1828422, EPI_ISL_1828423, EPI_ISL_1828424, EPI_ISL_1828425, EPI_ISL_1828426, EPI_ISL_1828427, EPI_ISL_1828428, EPI_ISL_1828429, EPI_ISL_1828430, EPI_ISL_1828431, EPI_ISL_1828432, EPI_ISL_1828433, EPI_ISL_1828434, EPI_ISL_1828435, EPI_ISL_1828436, EPI_ISL_1828437, EPI_ISL_1828438, EPI_ISL_1828439, EPI_ISL_1828440, EPI_ISL_1828441, EPI_ISL_1828442, EPI_ISL_1828443, EPI_ISL_1828444, EPI_ISL_1828445, EPI_ISL_1828446, EPI_ISL_1828447, EPI_ISL_1828448, EPI_ISL_1828449, EPI_ISL_1828450, EPI_ISL_1828451, EPI_ISL_1828452, EPI_ISL_1828453, EPI_ISL_1828454, EPI_ISL_1828455, EPI_ISL_1828456, EPI_ISL_1828457, EPI_ISL_1828458, EPI_ISL_1828459, EPI_ISL_1828460, EPI_ISL_1828461, EPI_ISL_1828462, EPI_ISL_1828463, EPI_ISL_1828464, EPI_ISL_1828465, EPI_ISL_1828466, EPI_ISL_1828467, EPI_ISL_1828468, EPI_ISL_1828469, EPI_ISL_1828470, EPI_ISL_1828471, EPI_ISL_1828472, EPI_ISL_1828473, EPI_ISL_1828474, EPI_ISL_1828475, EPI_ISL_1828476, EPI_ISL_1828477, EPI_ISL_1828478, EPI_ISL_1828479, EPI_ISL_1828480, EPI_ISL_1828481, EPI_ISL_1828482, EPI_ISL_1828483, EPI_ISL_1828484, EPI_ISL_1828485, EPI_ISL_1828486, EPI_ISL_1828487, EPI_ISL_1828488, EPI_ISL_1828489, EPI_ISL_1828490, EPI_ISL_1828491, EPI_ISL_1828492, EPI_ISL_1828493, EPI_ISL_1828494, EPI_ISL_1828495, EPI_ISL_1828496, EPI_ISL_1828497, EPI_ISL_1828498, EPI_ISL_1828499, EPI_ISL_1828500, EPI_ISL_1828501, EPI_ISL_1828502, EPI_ISL_1828503, EPI_ISL_1828504, EPI_ISL_1828505, EPI_ISL_1828506, EPI_ISL_1828507, EPI_ISL_1828508, EPI_ISL_1828509, EPI_ISL_1828510, EPI_ISL_1828511, EPI_ISL_1828512, EPI_ISL_1828513, EPI_ISL_1828514, EPI_ISL_1828515, EPI_ISL_1828516, EPI_ISL_1828517, EPI_ISL_1828518, EPI_ISL_1828519, EPI_ISL_1828520, EPI_ISL_1828521, EPI_ISL_1828522, EPI_ISL_1828523, EPI_ISL_1828524, EPI_ISL_1828525, EPI_ISL_1828526, EPI_ISL_1828527, EPI_ISL_1828528, EPI_ISL_1828529, EPI_ISL_1828530, EPI_ISL_1828531, EPI_ISL_1828532, EPI_ISL_1828533, EPI_ISL_1828534, EPI_ISL_1828535, EPI_ISL_1828536, EPI_ISL_1828537, EPI_ISL_1828538, EPI_ISL_1828539, EPI_ISL_1828540, EPI_ISL_1828541, EPI_ISL_1828542, EPI_ISL_1828543, EPI_ISL_1828544, EPI_ISL_1828545, EPI_ISL_1828546, EPI_ISL_1828547, EPI_ISL_1828548, EPI_ISL_1828549, EPI_ISL_1828550, EPI_ISL_1828551, EPI_ISL_1828552, EPI_ISL_1828553, EPI_ISL_1828554, EPI_ISL_1828555, EPI_ISL_1828556, EPI_ISL_1828557, EPI_ISL_1828558, EPI_ISL_1828559, EPI_ISL_1828560, EPI_ISL_1828561, EPI_ISL_1828562, EPI_ISL_1828563, EPI_ISL_1828564, EPI_ISL_1828565, EPI_ISL_1828566, EPI_ISL_1828567, EPI_ISL_1828568, EPI_ISL_1828569, EPI_ISL_1828570, EPI_ISL_1828571, EPI_ISL_1828572, EPI_ISL_1828573, EPI_ISL_1828574, EPI_ISL_1828575, EPI_ISL_1828576, EPI_ISL_1828577, EPI_ISL_1828578, EPI_ISL_1828579, EPI_ISL_1828580, EPI_ISL_1828581, EPI_ISL_1828582, EPI_ISL_1828583, EPI_ISL_1828584, EPI_ISL_1828585, EPI_ISL_1828586, EPI_ISL_1828587, EPI_ISL_1828588, EPI_ISL_1828589, EPI_ISL_1828590, EPI_ISL_1828591, EPI_ISL_1828592, EPI_ISL_1828593, EPI_ISL_1828594, EPI_ISL_1828595, EPI_ISL_1828596, EPI_ISL_1828597, EPI_ISL_1828598, EPI_ISL_1828599, EPI_ISL_1828600, EPI_ISL_1828601, EPI_ISL_1828602, EPI_ISL_1828603, EPI_ISL_1828604, EPI_ISL_1828605, EPI_ISL_1828606, EPI_ISL_1828607, EPI_ISL_1828608, EPI_ISL_1828609, EPI_ISL_1828610, EPI_ISL_1828611, EPI_ISL_1828612, EPI_ISL_1828613, EPI_ISL_1828614, EPI_ISL_1828615, EPI_ISL_1828616, EPI_ISL_1828617, EPI_ISL_1828618, EPI_ISL_1828619, EPI_ISL_1828620, EPI_ISL_1828621, EPI_ISL_1828622, EPI_ISL_1828623, EPI_ISL_1828624, EPI_ISL_1828625, EPI_ISL_1828626, EPI_ISL_1828627, EPI_ISL_1828628, EPI_ISL_1828629, EPI_ISL_1828630, EPI_ISL_1828631, EPI_ISL_1828632, EPI_ISL_1828633, EPI_ISL_1828634, EPI_ISL_1828635, EPI_ISL_1828636, EPI_ISL_1828637, EPI_ISL_1828638, EPI_ISL_1828639, EPI_ISL_1828640, EPI_ISL_1828641, EPI_ISL_1828642, EPI_ISL_1828643, EPI_ISL_1828644, EPI_ISL_1828645, EPI_ISL_1828646, EPI_ISL_1828647, EPI_ISL_1828648, EPI_ISL_1828649, EPI_ISL_1828650, EPI_ISL_1828651, EPI_ISL_1828652, EPI_ISL_1828653, EPI_ISL_1828654, EPI_ISL_1828655, EPI_ISL_1828656, EPI_ISL_1828657, EPI_ISL_1828658, EPI_ISL_1828659, EPI_ISL_1828660, EPI_ISL_1828661, EPI_ISL_1828662, EPI_ISL_1828663, EPI_ISL_1828664, EPI_ISL_1828665, EPI_ISL_1828666, EPI_ISL_1828667, EPI_ISL_1828668, EPI_ISL_1828669, EPI_ISL_1828670, EPI_ISL_1828671, EPI_ISL_1828672, EPI_ISL_1828673, EPI_ISL_1828674, EPI_ISL_1828675, EPI_ISL_1828676, EPI_ISL_1828677, EPI_ISL_1828678, EPI_ISL_1828679, EPI_ISL_1828680, EPI_ISL_1828681, EPI_ISL_1828682, EPI_ISL_1828683, EPI_ISL_1828684, EPI_ISL_1828685, EPI_ISL_1828686, EPI_ISL_1828687, EPI_ISL_1828688, EPI_ISL_1828689, EPI_ISL_1828690, EPI_ISL_1828691, EPI_ISL_1828692, EPI_ISL_1828693, EPI_ISL_1828694, EPI_ISL_1828695, EPI_ISL_1828696, EPI_ISL_1828697 | | | |
| see above | Pandemic Response Lab - NYC | Pandemic Response Lab, R&D | Henry Lee, Michael Hammerling, Melissa Hopkins, Cybill del Castillo, Shinyoung Clair Kang, William Ward, Pradeep Bugga, Sol Rey, Dylan Law, Katharine Nelson, Haiping Hao, Jon Laurent |
| EPI_ISL_1834218, EPI_ISL_1834241, EPI_ISL_1834304, EPI_ISL_1834326, EPI_ISL_1834346, EPI_ISL_1834359, EPI_ISL_1834371, EPI_ISL_1834375, EPI_ISL_1834408, EPI_ISL_1834421, EPI_ISL_1834440 | | | |
| see above | Quest Diagnostics Incorporated | Centers for Disease Control and Prevention Division of Viral Diseases, Pathogen Discovery | Dakota Howard, Dhwani Batra, Peter W. Cook, Kara Moser, Adrian Paskey, Jason Caravas, Benjamin Rambo-Martin, Shatavia Morrison, Christopher Gulvick, Scott Sammons, Yvette Unoarumhi, Darlene Wagner, Matthew Schmerer, S. H. Rosenthal, A. Gerasimova, R. M. Kagan, B. Anderson, M. Hua, Y. Liu, L.E. Bernstein, K.E. Livingston, A. Perez, I. A. Shlyakhter, R. V. Rolando, R. Owen, P. Tanpaiboon, F. Lacbawan, Clinton R. Paden, Duncan MacCannell |
| EPI_ISL_1834544, EPI_ISL_1834546, EPI_ISL_1834549, EPI_ISL_1834558, EPI_ISL_1834564, EPI_ISL_1834732, EPI_ISL_1834785, EPI_ISL_1834786, EPI_ISL_1834791, EPI_ISL_1834792, EPI_ISL_1834793, EPI_ISL_1834794, EPI_ISL_1834796, EPI_ISL_1834807, EPI_ISL_1834830, EPI_ISL_1834837, EPI_ISL_1834876, EPI_ISL_1834887, EPI_ISL_1834899, EPI_ISL_1834915, EPI_ISL_1834928, EPI_ISL_1834929, EPI_ISL_1834976, EPI_ISL_1835090, EPI_ISL_1835091, EPI_ISL_1835093, EPI_ISL_1835113, EPI_ISL_1835160, EPI_ISL_1835194, EPI_ISL_1835207, EPI_ISL_1835246, EPI_ISL_1835297, EPI_ISL_1835336, EPI_ISL_1835337, EPI_ISL_1835353, EPI_ISL_1835355, EPI_ISL_1835362, EPI_ISL_1835476, EPI_ISL_1835525, EPI_ISL_1835566, EPI_ISL_1835665, EPI_ISL_1835701, EPI_ISL_1835727, EPI_ISL_1835780, EPI_ISL_1835801, EPI_ISL_1835884, EPI_ISL_1835940, EPI_ISL_1835989, EPI_ISL_1836019, EPI_ISL_1836037, EPI_ISL_1836170, EPI_ISL_1836175, EPI_ISL_1836256, EPI_ISL_1836280, EPI_ISL_1836303, EPI_ISL_1836305, EPI_ISL_1836306, EPI_ISL_1836309, EPI_ISL_1836313, EPI_ISL_1836348, EPI_ISL_1836381, EPI_ISL_1836384, EPI_ISL_1836386, EPI_ISL_1836479, EPI_ISL_1836620, EPI_ISL_1836689, EPI_ISL_1836696, EPI_ISL_1836698, EPI_ISL_1836785, EPI_ISL_1836814, EPI_ISL_1836876, EPI_ISL_1836965, EPI_ISL_1837051, EPI_ISL_1837154, EPI_ISL_1837168, EPI_ISL_1837252, EPI_ISL_1837254, EPI_ISL_1837281, EPI_ISL_1837287, EPI_ISL_1837295, EPI_ISL_1837340, EPI_ISL_1837400, EPI_ISL_1837517, EPI_ISL_1837518, EPI_ISL_1837519, EPI_ISL_1837540, EPI_ISL_1837591, EPI_ISL_1837637, EPI_ISL_1837693, EPI_ISL_1837694, EPI_ISL_1837797, EPI_ISL_1837904, EPI_ISL_1837936, EPI_ISL_1837941, EPI_ISL_1837947, EPI_ISL_1838017, EPI_ISL_1838027, EPI_ISL_1838034, EPI_ISL_1838334, EPI_ISL_1838438 | | | |
| see above | Aegis Sciences Corporation | Centers for Disease Control and Prevention Division of Viral Diseases, Pathogen Discovery | Dakota Howard, Dhwani Batra, Peter W. Cook, Kara Moser, Adrian Paskey, Jason Caravas, Benjamin Rambo-Martin, Shatavia Morrison, Christopher Gulvick, Scott Sammons, Yvette Unoarumhi, Darlene Wagner, Matthew Schmerer, Cyndi Clark, Patrick Campbell, Rob Case, Vikramsinha Ghorpade, Holly Houdeshell, Ola Kvalvaag, Dillon Nall, Ethan Sanders, Alec Vest, Shaun Westlund, Matthew Hardison, Clinton R. Paden, Duncan MacCannell |
| EPI_ISL_1838933, EPI_ISL_1838960, EPI_ISL_1838965, EPI_ISL_1838975, EPI_ISL_1838980, EPI_ISL_1838993, EPI_ISL_1838996, EPI_ISL_1839003, EPI_ISL_1839004, EPI_ISL_1839010, EPI_ISL_1839013, EPI_ISL_1839014, EPI_ISL_1839020, EPI_ISL_1839021, EPI_ISL_1839024, EPI_ISL_1839042, EPI_ISL_1839054, EPI_ISL_1839055, EPI_ISL_1840412, EPI_ISL_1840423, EPI_ISL_1840431, EPI_ISL_1840435, EPI_ISL_1840458, EPI_ISL_1840526, EPI_ISL_1840528, EPI_ISL_1840531, EPI_ISL_1840546, EPI_ISL_1840559, EPI_ISL_1840561, EPI_ISL_1840567, EPI_ISL_1840570, EPI_ISL_1840602, EPI_ISL_1840604, EPI_ISL_1840619, EPI_ISL_1840623, EPI_ISL_1840625 | | | |
| see above | Quest Diagnostics Incorporated | Centers for Disease Control and Prevention Division of Viral Diseases, Pathogen Discovery | Dakota Howard, Dhwani Batra, Peter W. Cook, Kara Moser, Adrian Paskey, Jason Caravas, Benjamin Rambo-Martin, Shatavia Morrison, Christopher Gulvick, Scott Sammons, Yvette Unoarumhi, Darlene Wagner, Matthew Schmerer, S. H. Rosenthal, A. Gerasimova, R. M. Kagan, B. Anderson, M. Hua, Y. Liu, L.E. Bernstein, K.E. Livingston, A. Perez, I. A. Shlyakhter, R. V. Rolando, R. Owen, P. Tanpaiboon, F. Lacbawan, Clinton R. Paden, Duncan MacCannell |
| EPI_ISL_1854691 | SUNY UPSTATE MEDICAL UNIVERSITY | Wadsworth Center, New York State Department of Health | Kirsten St. George, Daryl M. Lamson, Alexis Russell, Matthew Shudt, Melissa A Leisner, Jonathan Plitnick, Catharine Prussing, Navjot Singh, John Kelly, Erasmus Schneider, Erica Lasek-Nesselquist |
| EPI_ISL_1854692 | ALBANY MEDICAL CENTER | Wadsworth Center, New York State Department of Health | Kirsten St. George, Daryl M. Lamson, Alexis Russell, Matthew Shudt, Melissa A Leisner, Jonathan Plitnick, Catharine Prussing, Navjot Singh, John Kelly, Erasmus Schneider, Erica Lasek-Nesselquist |
| EPI_ISL_1854693, EPI_ISL_1854694, EPI_ISL_1854695 | URMC LABS | Wadsworth Center, New York State Department of Health | Kirsten St. George, Daryl M. Lamson, Alexis Russell, Matthew Shudt, Melissa A Leisner, Jonathan Plitnick, Catharine Prussing, Navjot Singh, John Kelly, Erasmus Schneider, Erica Lasek-Nesselquist |
| EPI_ISL_1854696, EPI_ISL_1854697, EPI_ISL_1854698, EPI_ISL_1854699, EPI_ISL_1854700, EPI_ISL_1854701, EPI_ISL_1854702, EPI_ISL_1854703, EPI_ISL_1854704, EPI_ISL_1854705, EPI_ISL_1854706, EPI_ISL_1854707, EPI_ISL_1854708, EPI_ISL_1854709, EPI_ISL_1854710, EPI_ISL_1854711, EPI_ISL_1854712, EPI_ISL_1854713, EPI_ISL_1854714, EPI_ISL_1854715, EPI_ISL_1854716, EPI_ISL_1854717, EPI_ISL_1854718, EPI_ISL_1854719, EPI_ISL_1854720, EPI_ISL_1854721, EPI_ISL_1854722, EPI_ISL_1854723, EPI_ISL_1854724, EPI_ISL_1854725, EPI_ISL_1854726, EPI_ISL_1854727, EPI_ISL_1854728, EPI_ISL_1854729, EPI_ISL_1854730, EPI_ISL_1854731, EPI_ISL_1854732, EPI_ISL_1854733, EPI_ISL_1854734, EPI_ISL_1854735, EPI_ISL_1854736, EPI_ISL_1854737, EPI_ISL_1854738, EPI_ISL_1854739 | | | |
| see above | NYC Pandemic Response Lab | Wadsworth Center, New York State Department of Health | Kirsten St. George, Daryl M. Lamson, Alexis Russell, Matthew Shudt, Melissa A Leisner, Jonathan Plitnick, Catharine Prussing, Navjot Singh, John Kelly, Erasmus Schneider, Erica Lasek-Nesselquist |
| EPI_ISL_1855103 | MONTEFIORE MEDICAL CENTER LABORATORIES | Wadsworth Center, New York State Department of Health | Kirsten St. George, Daryl M. Lamson, Alexis Russell, Matthew Shudt, Melissa A Leisner, Jonathan Plitnick, Catharine Prussing, Navjot Singh, John Kelly, Erasmus Schneider, Erica Lasek-Nesselquist |
| EPI_ISL_1855104 | TEMPUS LABS INC | Wadsworth Center, New York State Department of Health | Kirsten St. George, Daryl M. Lamson, Alexis Russell, Matthew Shudt, Melissa A Leisner, Jonathan Plitnick, Catharine Prussing, Navjot Singh, John Kelly, Erasmus Schneider, Erica Lasek-Nesselquist |
| EPI_ISL_1855105, EPI_ISL_1855106, EPI_ISL_1855107, EPI_ISL_1855108, EPI_ISL_1855109, EPI_ISL_1855110, EPI_ISL_1855111, EPI_ISL_1855112, EPI_ISL_1855113, EPI_ISL_1855114, EPI_ISL_1855115 | | | |
| see above | SUNY UPSTATE MEDICAL UNIVERSITY | Wadsworth Center, New York State Department of Health | Kirsten St. George, Daryl M. Lamson, Alexis Russell, Matthew Shudt, Melissa A Leisner, Jonathan Plitnick, Catharine Prussing, Navjot Singh, John Kelly, Erasmus Schneider, Erica Lasek-Nesselquist |
| EPI_ISL_1855116, EPI_ISL_1855117, EPI_ISL_1855118 | ALBANY MEDICAL CENTER | Wadsworth Center, New York State Department of Health | Kirsten St. George, Daryl M. Lamson, Alexis Russell, Matthew Shudt, Melissa A Leisner, Jonathan Plitnick, Catharine Prussing, Navjot Singh, John Kelly, Erasmus Schneider, Erica Lasek-Nesselquist |
| EPI_ISL_1855119, EPI_ISL_1855120, EPI_ISL_1855121, EPI_ISL_1855122, EPI_ISL_1855123, EPI_ISL_1855124, EPI_ISL_1855125, EPI_ISL_1855126, EPI_ISL_1855127, EPI_ISL_1855128, EPI_ISL_1855129, EPI_ISL_1855130, EPI_ISL_1855131, EPI_ISL_1855132, EPI_ISL_1855133, EPI_ISL_1855134, EPI_ISL_1855135, EPI_ISL_1855136, EPI_ISL_1855137, EPI_ISL_1855138, EPI_ISL_1855139, EPI_ISL_1855140, EPI_ISL_1855141, EPI_ISL_1855142, EPI_ISL_1855143 | | | |
| see above | URMC LABS | Wadsworth Center, New York State Department of Health | Kirsten St. George, Daryl M. Lamson, Alexis Russell, Matthew Shudt, Melissa A Leisner, Jonathan Plitnick, Catharine Prussing, Navjot Singh, John Kelly, Erasmus Schneider, Erica Lasek-Nesselquist |
| EPI_ISL_1855144, EPI_ISL_1855145, EPI_ISL_1855146, EPI_ISL_1855147, EPI_ISL_1855148, EPI_ISL_1855149 | GLENS FALLS HOSPITAL LABORATORY | Wadsworth Center, New York State Department of Health | Kirsten St. George, Daryl M. Lamson, Alexis Russell, Matthew Shudt, Melissa A Leisner, Jonathan Plitnick, Catharine Prussing, Navjot Singh, John Kelly, Erasmus Schneider, Erica Lasek-Nesselquist |
| EPI_ISL_1855150, EPI_ISL_1855151, EPI_ISL_1855152, EPI_ISL_1855153, EPI_ISL_1855154, EPI_ISL_1855155, EPI_ISL_1855156, EPI_ISL_1855157, EPI_ISL_1855158, EPI_ISL_1855159, EPI_ISL_1855160, EPI_ISL_1855161, EPI_ISL_1855162, EPI_ISL_1855163, EPI_ISL_1855164, EPI_ISL_1855165, EPI_ISL_1855166, EPI_ISL_1855167 | | | |
| see above | TEMPUS LABS INC | Wadsworth Center, New York State Department of Health | Kirsten St. George, Daryl M. Lamson, Alexis Russell, Matthew Shudt, Melissa A Leisner, Jonathan Plitnick, Catharine Prussing, Navjot Singh, John Kelly, Erasmus Schneider, Erica Lasek-Nesselquist |
| EPI_ISL_1855168, EPI_ISL_1855169, EPI_ISL_1855170, EPI_ISL_1855171, EPI_ISL_1855172, EPI_ISL_1855173, EPI_ISL_1855174, EPI_ISL_1855175, EPI_ISL_1855176, EPI_ISL_1855177, EPI_ISL_1855178, EPI_ISL_1855179, EPI_ISL_1855180, EPI_ISL_1855181, EPI_ISL_1855182, EPI_ISL_1855183, EPI_ISL_1855184, EPI_ISL_1855185 | | | |

| | | | |
|---|---|---|---|
| see above | URMC LABS | Wadsworth Center, New York State Department of Health | Kirsten St. George, Daryl M. Lamson, Alexis Russell, Matthew Shudt, Melissa A Leisner, Jonathan Plitnick, Catharine Prussing, Navjot Singh, John Kelly, Erasmus Schneider, Erica Lasek-Nesselquist |
| EPI_ISL_1855186, EPI_ISL_1855187 | NYC Pandemic Response Lab | Wadsworth Center, New York State Department of Health | Kirsten St. George, Daryl M. Lamson, Alexis Russell, Matthew Shudt, Melissa A Leisner, Jonathan Plitnick, Catharine Prussing, Navjot Singh, John Kelly, Erasmus Schneider, Erica Lasek-Nesselquist |
| EPI_ISL_1855188, EPI_ISL_1855189, EPI_ISL_1855190, EPI_ISL_1855191, EPI_ISL_1855192, EPI_ISL_1855193, EPI_ISL_1855194, EPI_ISL_1855195, EPI_ISL_1855196 | MONTEFIORE MEDICAL CENTER LABORATORIES | Wadsworth Center, New York State Department of Health | Kirsten St. George, Daryl M. Lamson, Alexis Russell, Matthew Shudt, Melissa A Leisner, Jonathan Plitnick, Catharine Prussing, Navjot Singh, John Kelly, Erasmus Schneider, Erica Lasek-Nesselquist |
| EPI_ISL_1855197, EPI_ISL_1855198, EPI_ISL_1855199, EPI_ISL_1855200, EPI_ISL_1855201 | NYC Pandemic Response Lab | Wadsworth Center, New York State Department of Health | Kirsten St. George, Daryl M. Lamson, Alexis Russell, Matthew Shudt, Melissa A Leisner, Jonathan Plitnick, Catharine Prussing, Navjot Singh, John Kelly, Erasmus Schneider, Erica Lasek-Nesselquist |
| EPI_ISL_1855202, EPI_ISL_1855203, EPI_ISL_1855204, EPI_ISL_1855205, EPI_ISL_1855206, EPI_ISL_1855207, EPI_ISL_1855208, EPI_ISL_1855209 | MONTEFIORE MEDICAL CENTER LABORATORIES | Wadsworth Center, New York State Department of Health | Kirsten St. George, Daryl M. Lamson, Alexis Russell, Matthew Shudt, Melissa A Leisner, Jonathan Plitnick, Catharine Prussing, Navjot Singh, John Kelly, Erasmus Schneider, Erica Lasek-Nesselquist |
| EPI_ISL_1855210 | MEMORIAL SLOAN KETTERING CANCER CENTER | Wadsworth Center, New York State Department of Health | Kirsten St. George, Daryl M. Lamson, Alexis Russell, Matthew Shudt, Melissa A Leisner, Jonathan Plitnick, Catharine Prussing, Navjot Singh, John Kelly, Erasmus Schneider, Erica Lasek-Nesselquist |
| EPI_ISL_1855211, EPI_ISL_1855212, EPI_ISL_1855213, EPI_ISL_1855214 | New York Presbyterian Hospital | Wadsworth Center, New York State Department of Health | Kirsten St. George, Daryl M. Lamson, Alexis Russell, Matthew Shudt, Melissa A Leisner, Jonathan Plitnick, Catharine Prussing, Navjot Singh, John Kelly, Erasmus Schneider, Erica Lasek-Nesselquist |
| EPI_ISL_1855215, EPI_ISL_1855216, EPI_ISL_1855217, EPI_ISL_1855218, EPI_ISL_1855219 | MONTEFIORE MEDICAL CENTER LABORATORIES | Wadsworth Center, New York State Department of Health | Kirsten St. George, Daryl M. Lamson, Alexis Russell, Matthew Shudt, Melissa A Leisner, Jonathan Plitnick, Catharine Prussing, Navjot Singh, John Kelly, Erasmus Schneider, Erica Lasek-Nesselquist |
| EPI_ISL_1855220 | SARATOGA HOSPITAL LABORATORY | Wadsworth Center, New York State Department of Health | Kirsten St. George, Daryl M. Lamson, Alexis Russell, Matthew Shudt, Melissa A Leisner, Jonathan Plitnick, Catharine Prussing, Navjot Singh, John Kelly, Erasmus Schneider, Erica Lasek-Nesselquist |
| EPI_ISL_1855221, EPI_ISL_1855222, EPI_ISL_1855223, EPI_ISL_1855224, EPI_ISL_1855225 | SUNY UPSTATE MEDICAL UNIVERSITY | Wadsworth Center, New York State Department of Health | Kirsten St. George, Daryl M. Lamson, Alexis Russell, Matthew Shudt, Melissa A Leisner, Jonathan Plitnick, Catharine Prussing, Navjot Singh, John Kelly, Erasmus Schneider, Erica Lasek-Nesselquist |
| EPI_ISL_1855226, EPI_ISL_1855227, EPI_ISL_1855228, EPI_ISL_1855229, EPI_ISL_1855230, EPI_ISL_1855231, EPI_ISL_1855232, EPI_ISL_1855233, EPI_ISL_1855234, EPI_ISL_1855235, EPI_ISL_1855236, EPI_ISL_1855237, EPI_ISL_1855238, EPI_ISL_1855239, EPI_ISL_1855240 | | | |
| see above | URMC LABS | Wadsworth Center, New York State Department of Health | Kirsten St. George, Daryl M. Lamson, Alexis Russell, Matthew Shudt, Melissa A Leisner, Jonathan Plitnick, Catharine Prussing, Navjot Singh, John Kelly, Erasmus Schneider, Erica Lasek-Nesselquist |
| EPI_ISL_1855241, EPI_ISL_1855242, EPI_ISL_1855243 | GLENS FALLS HOSPITAL LABORATORY | Wadsworth Center, New York State Department of Health | Kirsten St. George, Daryl M. Lamson, Alexis Russell, Matthew Shudt, Melissa A Leisner, Jonathan Plitnick, Catharine Prussing, Navjot Singh, John Kelly, Erasmus Schneider, Erica Lasek-Nesselquist |
| EPI_ISL_1855244, EPI_ISL_1855245, EPI_ISL_1855246, EPI_ISL_1855247, EPI_ISL_1855248 | TEMPUS LABS INC | Wadsworth Center, New York State Department of Health | Kirsten St. George, Daryl M. Lamson, Alexis Russell, Matthew Shudt, Melissa A Leisner, Jonathan Plitnick, Catharine Prussing, Navjot Singh, John Kelly, Erasmus Schneider, Erica Lasek-Nesselquist |
| EPI_ISL_1855249, EPI_ISL_1855250, EPI_ISL_1855251, EPI_ISL_1855252, EPI_ISL_1855253, EPI_ISL_1855254, EPI_ISL_1855255, EPI_ISL_1855256, EPI_ISL_1855257, EPI_ISL_1855258, EPI_ISL_1855259, EPI_ISL_1855260, EPI_ISL_1855261, EPI_ISL_1855262, EPI_ISL_1855263, EPI_ISL_1855264, EPI_ISL_1855265, EPI_ISL_1855266, EPI_ISL_1855267, EPI_ISL_1855268, EPI_ISL_1855269, EPI_ISL_1855270, EPI_ISL_1855271, EPI_ISL_1855272 | | | |
| see above | URMC LABS | Wadsworth Center, New York State Department of Health | Kirsten St. George, Daryl M. Lamson, Alexis Russell, Matthew Shudt, Melissa A Leisner, Jonathan Plitnick, Catharine Prussing, Navjot Singh, John Kelly, Erasmus Schneider, Erica Lasek-Nesselquist |
| EPI_ISL_1855273, EPI_ISL_1855274, EPI_ISL_1855275 | SARATOGA HOSPITAL LABORATORY | Wadsworth Center, New York State Department of Health | Kirsten St. George, Daryl M. Lamson, Alexis Russell, Matthew Shudt, Melissa A Leisner, Jonathan Plitnick, Catharine Prussing, Navjot Singh, John Kelly, Erasmus Schneider, Erica Lasek-Nesselquist |
| EPI_ISL_1855276, EPI_ISL_1855277, EPI_ISL_1855278, EPI_ISL_1855279, EPI_ISL_1855280, EPI_ISL_1855281, EPI_ISL_1855282, EPI_ISL_1855283, EPI_ISL_1855284, EPI_ISL_1855285 | URMC LABS | Wadsworth Center, New York State Department of Health | Kirsten St. George, Daryl M. Lamson, Alexis Russell, Matthew Shudt, Melissa A Leisner, Jonathan Plitnick, Catharine Prussing, Navjot Singh, John Kelly, Erasmus Schneider, Erica Lasek-Nesselquist |
| EPI_ISL_1855286, EPI_ISL_1855287, EPI_ISL_1855288, EPI_ISL_1855289, EPI_ISL_1855290, EPI_ISL_1855291, EPI_ISL_1855292, EPI_ISL_1855293 | Columbia University Irving Medical Center | Wadsworth Center, New York State Department of Health | Kirsten St. George, Daryl M. Lamson, Alexis Russell, Matthew Shudt, Melissa A Leisner, Jonathan Plitnick, Catharine Prussing, Navjot Singh, John Kelly, Erasmus Schneider, Erica Lasek-Nesselquist |
| EPI_ISL_1855294, EPI_ISL_1855295, EPI_ISL_1855296, EPI_ISL_1855297, EPI_ISL_1855298, EPI_ISL_1855299, EPI_ISL_1855300, EPI_ISL_1855301, EPI_ISL_1855302, EPI_ISL_1855303, EPI_ISL_1855304, EPI_ISL_1855305, EPI_ISL_1855306, EPI_ISL_1855307, EPI_ISL_1855308, EPI_ISL_1855309 | | | |
| see above | URMC LABS | Wadsworth Center, New York State Department of Health | Kirsten St. George, Daryl M. Lamson, Alexis Russell, Matthew Shudt, Melissa A Leisner, Jonathan Plitnick, Catharine Prussing, Navjot Singh, John Kelly, Erasmus Schneider, Erica Lasek-Nesselquist |
| EPI_ISL_1855310, EPI_ISL_1855311, EPI_ISL_1855312, EPI_ISL_1855313, EPI_ISL_1855314, EPI_ISL_1855315, EPI_ISL_1855316, EPI_ISL_1855317, EPI_ISL_1855318, EPI_ISL_1855319, EPI_ISL_1855320, EPI_ISL_1855321, EPI_ISL_1855322, EPI_ISL_1855323 | | | |
| see above | BOSTON HEART DIAGNOSTICS CORP | Wadsworth Center, New York State Department of Health | Kirsten St. George, Daryl M. Lamson, Alexis Russell, Matthew Shudt, Melissa A Leisner, Jonathan Plitnick, Catharine Prussing, Navjot Singh, John Kelly, Erasmus Schneider, Erica Lasek-Nesselquist |
| EPI_ISL_1855324, EPI_ISL_1855325, EPI_ISL_1855326, EPI_ISL_1855327, EPI_ISL_1855328, EPI_ISL_1855329, EPI_ISL_1855330, EPI_ISL_1855331, EPI_ISL_1855332, EPI_ISL_1855333 | MONTEFIORE MEDICAL CENTER LABORATORIES | Wadsworth Center, New York State Department of Health | Kirsten St. George, Daryl M. Lamson, Alexis Russell, Matthew Shudt, Melissa A Leisner, Jonathan Plitnick, Catharine Prussing, Navjot Singh, John Kelly, Erasmus Schneider, Erica Lasek-Nesselquist |
| EPI_ISL_1855334, EPI_ISL_1855335, EPI_ISL_1855336, EPI_ISL_1855337, EPI_ISL_1855338, EPI_ISL_1855339, EPI_ISL_1855340, EPI_ISL_1855341, EPI_ISL_1855342, EPI_ISL_1855343, EPI_ISL_1855344, EPI_ISL_1855345, EPI_ISL_1855346, EPI_ISL_1855347 | | | |
| see above | WESTCHESTER MEDICAL CENTER | Wadsworth Center, New York State Department of Health | Kirsten St. George, Daryl M. Lamson, Alexis Russell, Matthew Shudt, Melissa A Leisner, Jonathan Plitnick, Catharine Prussing, Navjot Singh, John Kelly, Erasmus Schneider, Erica Lasek-Nesselquist |
| EPI_ISL_1855348, EPI_ISL_1855349, EPI_ISL_1855350, EPI_ISL_1855351, EPI_ISL_1855352, EPI_ISL_1855353 | NORTH SHORE UNIVERSITY HOSPITAL | Wadsworth Center, New York State Department of Health | Kirsten St. George, Daryl M. Lamson, Alexis Russell, Matthew Shudt, Melissa A Leisner, Jonathan Plitnick, Catharine Prussing, Navjot Singh, John Kelly, Erasmus Schneider, Erica Lasek-Nesselquist |
| EPI_ISL_1855354, EPI_ISL_1855355, EPI_ISL_1855356, EPI_ISL_1855357, EPI_ISL_1855358, EPI_ISL_1855359, EPI_ISL_1855360 | SUNY UPSTATE MEDICAL UNIVERSITY | Wadsworth Center, New York State Department of Health | Kirsten St. George, Daryl M. Lamson, Alexis Russell, Matthew Shudt, Melissa A Leisner, Jonathan Plitnick, Catharine Prussing, Navjot Singh, John Kelly, Erasmus Schneider, Erica Lasek-Nesselquist |
| EPI_ISL_1900231, EPI_ISL_1900249, | Aegis Sciences Corporation | Centers for Disease Control and Prevention Division of Viral | Dakota Howard, Dhwani Batra, Peter W. Cook, Kara Moser, Adrian Paskey, Jason Caravas, Benjamin Rambo-Martin, Shatavia Morrison, Christopher |

| | | | |
|---|---|---|---|
| EPI_ISL_1900283, EPI_ISL_1900354, EPI_ISL_1900419 | | Diseases, Pathogen Discovery | Gulvick, Scott Sammons, Yvette Unoarumhi, Darlene Wagner, Matthew Schmerer, Cyndi Clark, Patrick Campbell, Rob Case, Vikramsinha Ghorpade, Holly Houdeshell, Ola Kvalvaag, Dillon Nall, Ethan Sanders, Alec Vest, Shaun Westlund, Matthew Hardison, Clinton R. Paden, Duncan MacCannell |
| EPI_ISL_1906183, EPI_ISL_1906197, EPI_ISL_1906206, EPI_ISL_1906210, EPI_ISL_1906215, EPI_ISL_1906217, EPI_ISL_1906223, EPI_ISL_1906244, EPI_ISL_1906253, EPI_ISL_1906258, EPI_ISL_1906268, EPI_ISL_1906270, EPI_ISL_1906272, EPI_ISL_1906274, EPI_ISL_1906276, EPI_ISL_1906286, EPI_ISL_1906290, EPI_ISL_1906308, EPI_ISL_1906318, EPI_ISL_1906329, EPI_ISL_1906345, EPI_ISL_1906347, EPI_ISL_1906349, EPI_ISL_1906353, EPI_ISL_1906361, EPI_ISL_1906372, EPI_ISL_1906400, EPI_ISL_1906402, EPI_ISL_1906407, EPI_ISL_1906420, EPI_ISL_1906422, EPI_ISL_1906437, EPI_ISL_1906440, EPI_ISL_1906452, EPI_ISL_1906455, EPI_ISL_1906461, EPI_ISL_1906474, EPI_ISL_1906480, EPI_ISL_1906482, EPI_ISL_1906486, EPI_ISL_1906491, EPI_ISL_1906504, EPI_ISL_1906511, EPI_ISL_1906516, EPI_ISL_1906523, EPI_ISL_1906526, EPI_ISL_1906530, EPI_ISL_1906532, EPI_ISL_1906538, EPI_ISL_1906542, EPI_ISL_1906546, EPI_ISL_1906549, EPI_ISL_1906555, EPI_ISL_1906557, EPI_ISL_1906562, EPI_ISL_1906564, EPI_ISL_1906567, EPI_ISL_1906573, EPI_ISL_1906593, EPI_ISL_1906595, EPI_ISL_1906600, EPI_ISL_1906619, EPI_ISL_1906623, EPI_ISL_1906634, EPI_ISL_1906639, EPI_ISL_1906657, EPI_ISL_1906659, EPI_ISL_1906665, EPI_ISL_1906714, EPI_ISL_1906721, EPI_ISL_1906747, EPI_ISL_1906761, EPI_ISL_1906774, EPI_ISL_1906781, EPI_ISL_1906788, EPI_ISL_1906797, EPI_ISL_1906806, EPI_ISL_1906814, EPI_ISL_1906823, EPI_ISL_1906828, EPI_ISL_1906832, EPI_ISL_1906844, EPI_ISL_1906850, EPI_ISL_1906851, EPI_ISL_1906852, EPI_ISL_1906855, EPI_ISL_1906857, EPI_ISL_1906858, EPI_ISL_1906861, EPI_ISL_1906866, EPI_ISL_1906868, EPI_ISL_1906872, EPI_ISL_1906879 | | | |
| see above | Quest Diagnostics Incorporated | Centers for Disease Control and Prevention Division of Viral Diseases, Pathogen Discovery | Dakota Howard, Dhwani Batra, Peter W. Cook, Kara Moser, Adrian Paskey, Jason Caravas, Benjamin Rambo-Martin, Shatavia Morrison, Christopher Gulvick, Scott Sammons, Yvette Unoarumhi, Darlene Wagner, Matthew Schmerer, S. H. Rosenthal, A. Gerasimova, R. M. Kagan, B. Anderson, M. Hua, Y. Liu, L.E. Bernstein, K.E. Livingston, A. Perez, I. A. Shlyakhter, R. V. Rolando, R. Owen, P. Tanpaiboon, F. Lacbawan, Clinton R. Paden, Duncan MacCannell |
| EPI_ISL_1906926, EPI_ISL_1907154, EPI_ISL_1907210, EPI_ISL_1907414, EPI_ISL_1907867, EPI_ISL_1907911 | Helix/Illumina | Centers for Disease Control and Prevention Division of Viral Diseases, Pathogen Discovery | Dakota Howard, Dhwani Batra, Peter W. Cook, Kara Moser, Adrian Paskey, Jason Caravas, Benjamin Rambo-Martin, Shatavia Morrison, Christopher Gulvick, Scott Sammons, Yvette Unoarumhi, Darlene Wagner, Matthew Schmerer, Eileen de Feo, Jan Antico, Christine Tran, Matthew Tolentino, Shannon Wickline, Kim Gietzen, Brad Sickler, Jingtao Liu, Eric Allen, Phil Febbo, Nicole L. Washington, Simon White, Geraint Levan, Kelly Schiabor Barrett, Elizabeth Cirulli, Alexandre Bolze, Ary Ascencio, Charlotte Rivera-Garcia, Ryan Cho, Jason Nguyen, Sherry Wang, Jimmy Ramirez, Tyler Cassens, Efren Sandoval, Magnus Isaksson, William Lee, David Becker, Marc Laurent, James Lu, Clinton R. Paden, Duncan MacCannell |
| EPI_ISL_1918967, EPI_ISL_1918968, EPI_ISL_1919089, EPI_ISL_1919090, EPI_ISL_1919091, EPI_ISL_1919092, EPI_ISL_1919093, EPI_ISL_1919094, EPI_ISL_1919095, EPI_ISL_1919096, EPI_ISL_1919097, EPI_ISL_1919098, EPI_ISL_1919099, EPI_ISL_1919100, EPI_ISL_1919101, EPI_ISL_1919102, EPI_ISL_1919103, EPI_ISL_1919104, EPI_ISL_1919105 | | | |
| see above | Broad Institute Clinical Research Sequencing Platform | Infectious Disease Program, Broad Institute of Harvard and MIT | Siddle,K.J., Adams,G., Pearlman,L., Gladden-Young,A., Vicente,G., Blumenstiel,B., DeFelice,M., Lee,M., McGovern,S., Lagerborg,K., Rudy,M., DeRuff,K., Carter,A., Normandin,E., Bauer,M., Reilly,S., Tomkins-Tinch,C., Loreth,C., Chaluvadi,S., Meldrim,J., Granger,B., Lemieux,J.E., Birren,B.W., Sabeti,P.C., Larkin,K., Dodge,S., Lennon,N., Madoff,L., Brown,C., Gallagher,G., Smole,S., Park,D.J., Gabriel,S., and MacInnis,B.L. |
| EPI_ISL_1919988, EPI_ISL_1919989 | Northwell Health Laboratories | Wadsworth Center, New York State Department of Health | Kirsten St. George, Daryl M. Lamson, Alexis Russell, Matthew Shudt, Melissa A Leisner, Jonathan Plitnick, Catharine Prussing, Navjot Singh, John Kelly, Erasmus Schneider, Erica Lasek-Nesselquist |
| EPI_ISL_1919990 | Suny Upstate Medical University | Wadsworth Center, New York State Department of Health | Kirsten St. George, Daryl M. Lamson, Alexis Russell, Matthew Shudt, Melissa A Leisner, Jonathan Plitnick, Catharine Prussing, Navjot Singh, John Kelly, Erasmus Schneider, Erica Lasek-Nesselquist |
| EPI_ISL_1919991, EPI_ISL_1919992 | Stony Brook University Hospital | Wadsworth Center, New York State Department of Health | Kirsten St. George, Daryl M. Lamson, Alexis Russell, Matthew Shudt, Melissa A Leisner, Jonathan Plitnick, Catharine Prussing, Navjot Singh, John Kelly, Erasmus Schneider, Erica Lasek-Nesselquist |
| EPI_ISL_1919993, EPI_ISL_1919994, EPI_ISL_1919995, EPI_ISL_1919996 | Northwell Health Laboratories | Wadsworth Center, New York State Department of Health | Kirsten St. George, Daryl M. Lamson, Alexis Russell, Matthew Shudt, Melissa A Leisner, Jonathan Plitnick, Catharine Prussing, Navjot Singh, John Kelly, Erasmus Schneider, Erica Lasek-Nesselquist |
| EPI_ISL_1919997, EPI_ISL_1919998, EPI_ISL_1919999, EPI_ISL_1920000, EPI_ISL_1920001, EPI_ISL_1920002, EPI_ISL_1920003, EPI_ISL_1920004, EPI_ISL_1920005, EPI_ISL_1920006 | Montefiore Medical Center Laboratories | Wadsworth Center, New York State Department of Health | Kirsten St. George, Daryl M. Lamson, Alexis Russell, Matthew Shudt, Melissa A Leisner, Jonathan Plitnick, Catharine Prussing, Navjot Singh, John Kelly, Erasmus Schneider, Erica Lasek-Nesselquist |
| EPI_ISL_1920007 | Westchester Medical Center | Wadsworth Center, New York State Department of Health | Kirsten St. George, Daryl M. Lamson, Alexis Russell, Matthew Shudt, Melissa A Leisner, Jonathan Plitnick, Catharine Prussing, Navjot Singh, John Kelly, Erasmus Schneider, Erica Lasek-Nesselquist |
| EPI_ISL_1920008 | Northwell Health Laboratories | Wadsworth Center, New York State Department of Health | Kirsten St. George, Daryl M. Lamson, Alexis Russell, Matthew Shudt, Melissa A Leisner, Jonathan Plitnick, Catharine Prussing, Navjot Singh, John Kelly, Erasmus Schneider, Erica Lasek-Nesselquist |
| EPI_ISL_1920009 | Westchester Medical Center | Wadsworth Center, New York State Department of Health | Kirsten St. George, Daryl M. Lamson, Alexis Russell, Matthew Shudt, Melissa A Leisner, Jonathan Plitnick, Catharine Prussing, Navjot Singh, John Kelly, Erasmus Schneider, Erica Lasek-Nesselquist |
| EPI_ISL_1920010, EPI_ISL_1920011, EPI_ISL_1920012, EPI_ISL_1920013, EPI_ISL_1920014, EPI_ISL_1920015, EPI_ISL_1920016, EPI_ISL_1920017, EPI_ISL_1920018, EPI_ISL_1920019, EPI_ISL_1920020, EPI_ISL_1920021, EPI_ISL_1920022, EPI_ISL_1920023, EPI_ISL_1920024, EPI_ISL_1920025, EPI_ISL_1920026, EPI_ISL_1920027, EPI_ISL_1920028 | | | |
| see above | Northwell Health Laboratories | Wadsworth Center, New York State Department of Health | Kirsten St. George, Daryl M. Lamson, Alexis Russell, Matthew Shudt, Melissa A Leisner, Jonathan Plitnick, Catharine Prussing, Navjot Singh, John Kelly, Erasmus Schneider, Erica Lasek-Nesselquist |
| EPI_ISL_1920029 | St. Francis Hospital Laboratory | Wadsworth Center, New York State Department of Health | Kirsten St. George, Daryl M. Lamson, Alexis Russell, Matthew Shudt, Melissa A Leisner, Jonathan Plitnick, Catharine Prussing, Navjot Singh, John Kelly, Erasmus Schneider, Erica Lasek-Nesselquist |
| EPI_ISL_1920030 | Stony Brook University Hospital | Wadsworth Center, New York State Department of Health | Kirsten St. George, Daryl M. Lamson, Alexis Russell, Matthew Shudt, Melissa A Leisner, Jonathan Plitnick, Catharine Prussing, Navjot Singh, John Kelly, Erasmus Schneider, Erica Lasek-Nesselquist |
| EPI_ISL_1920031, EPI_ISL_1920032, EPI_ISL_1920033, EPI_ISL_1920034 | Northwell Health Laboratories | Wadsworth Center, New York State Department of Health | Kirsten St. George, Daryl M. Lamson, Alexis Russell, Matthew Shudt, Melissa A Leisner, Jonathan Plitnick, Catharine Prussing, Navjot Singh, John Kelly, Erasmus Schneider, Erica Lasek-Nesselquist |
| EPI_ISL_1920035 | Stony Brook University Hospital | Wadsworth Center, New York State Department of Health | Kirsten St. George, Daryl M. Lamson, Alexis Russell, Matthew Shudt, Melissa A Leisner, Jonathan Plitnick, Catharine Prussing, Navjot Singh, John Kelly, Erasmus Schneider, Erica Lasek-Nesselquist |
| EPI_ISL_1920036, EPI_ISL_1920037, EPI_ISL_1920038, EPI_ISL_1920039, EPI_ISL_1920040 | Northwell Health Laboratories | Wadsworth Center, New York State Department of Health | Kirsten St. George, Daryl M. Lamson, Alexis Russell, Matthew Shudt, Melissa A Leisner, Jonathan Plitnick, Catharine Prussing, Navjot Singh, John Kelly, Erasmus Schneider, Erica Lasek-Nesselquist |
| EPI_ISL_1920041 | Stony Brook University Hospital | Wadsworth Center, New York State Department of Health | Kirsten St. George, Daryl M. Lamson, Alexis Russell, Matthew Shudt, Melissa A Leisner, Jonathan Plitnick, Catharine Prussing, Navjot Singh, John Kelly, Erasmus Schneider, Erica Lasek-Nesselquist |
| EPI_ISL_1920042, EPI_ISL_1920043 | Northwell Health Laboratories | Wadsworth Center, New York State Department of Health | Kirsten St. George, Daryl M. Lamson, Alexis Russell, Matthew Shudt, Melissa A Leisner, Jonathan Plitnick, Catharine Prussing, Navjot Singh, John Kelly, Erasmus Schneider, Erica Lasek-Nesselquist |
| EPI_ISL_1920044 | Stony Brook University Hospital | Wadsworth Center, New York State Department of Health | Kirsten St. George, Daryl M. Lamson, Alexis Russell, Matthew Shudt, Melissa A Leisner, Jonathan Plitnick, Catharine Prussing, Navjot Singh, John Kelly, Erasmus Schneider, Erica Lasek-Nesselquist |
| EPI_ISL_1920045 | Northwell Health Laboratories | Wadsworth Center, New York State Department of Health | Kirsten St. George, Daryl M. Lamson, Alexis Russell, Matthew Shudt, Melissa A Leisner, Jonathan Plitnick, Catharine Prussing, Navjot Singh, John Kelly, Erasmus Schneider, Erica Lasek-Nesselquist |
| EPI_ISL_1920046, EPI_ISL_1920047, EPI_ISL_1920048, EPI_ISL_1920049 | Stony Brook University Hospital | Wadsworth Center, New York State Department of Health | Kirsten St. George, Daryl M. Lamson, Alexis Russell, Matthew Shudt, Melissa A Leisner, Jonathan Plitnick, Catharine Prussing, Navjot Singh, John Kelly, Erasmus Schneider, Erica Lasek-Nesselquist |
| EPI_ISL_1920050, EPI_ISL_1920051, EPI_ISL_1920052, EPI_ISL_1920053, EPI_ISL_1920054, EPI_ISL_1920055, EPI_ISL_1920056, EPI_ISL_1920057, EPI_ISL_1920058, EPI_ISL_1920059, EPI_ISL_1920060 | | | |
| see above | Albany Medical Center | Wadsworth Center, New York State Department of Health | Kirsten St. George, Daryl M. Lamson, Alexis Russell, Matthew Shudt, Melissa A Leisner, Jonathan Plitnick, Catharine Prussing, Navjot Singh, John Kelly, Erasmus Schneider, Erica Lasek-Nesselquist |
| EPI_ISL_1920061 | Memorial Sloan Kettering Cancer Center | Wadsworth Center, New York State Department of Health | Kirsten St. George, Daryl M. Lamson, Alexis Russell, Matthew Shudt, Melissa A Leisner, Jonathan Plitnick, Catharine Prussing, Navjot Singh, John Kelly, Erasmus Schneider, Erica Lasek-Nesselquist |
| EPI_ISL_1920062, EPI_ISL_1920063, EPI_ISL_1920064, EPI_ISL_1920065 | Westchester Medical Center | Wadsworth Center, New York State Department of Health | Kirsten St. George, Daryl M. Lamson, Alexis Russell, Matthew Shudt, Melissa A Leisner, Jonathan Plitnick, Catharine Prussing, Navjot Singh, John Kelly, Erasmus Schneider, Erica Lasek-Nesselquist |

| | | | |
|---|---|---|---|
| EPI_ISL_1920066, EPI_ISL_1920067, EPI_ISL_1920068, EPI_ISL_1920069 | URMC LABS | Wadsworth Center, New York State Department of Health | Kirsten St. George, Daryl M. Lamson, Alexis Russell, Matthew Shudt, Melissa A Leisner, Jonathan Plitnick, Catharine Prussing, Navjot Singh, John Kelly, Erasmus Schneider, Erica Lasek-Nesselquist |
| EPI_ISL_1920070, EPI_ISL_1920071, EPI_ISL_1920072, EPI_ISL_1920073 | NYC Pandemic Response Lab | Wadsworth Center, New York State Department of Health | Kirsten St. George, Daryl M. Lamson, Alexis Russell, Matthew Shudt, Melissa A Leisner, Jonathan Plitnick, Catharine Prussing, Navjot Singh, John Kelly, Erasmus Schneider, Erica Lasek-Nesselquist |
| EPI_ISL_1920074, EPI_ISL_1920075 | Montefiore Medical Center Laboratories | Wadsworth Center, New York State Department of Health | Kirsten St. George, Daryl M. Lamson, Alexis Russell, Matthew Shudt, Melissa A Leisner, Jonathan Plitnick, Catharine Prussing, Navjot Singh, John Kelly, Erasmus Schneider, Erica Lasek-Nesselquist |
| EPI_ISL_1920076 | Memorial Sloan Kettering Cancer Center | Wadsworth Center, New York State Department of Health | Kirsten St. George, Daryl M. Lamson, Alexis Russell, Matthew Shudt, Melissa A Leisner, Jonathan Plitnick, Catharine Prussing, Navjot Singh, John Kelly, Erasmus Schneider, Erica Lasek-Nesselquist |
| EPI_ISL_1920077 | Montefiore Medical Center Laboratories | Wadsworth Center, New York State Department of Health | Kirsten St. George, Daryl M. Lamson, Alexis Russell, Matthew Shudt, Melissa A Leisner, Jonathan Plitnick, Catharine Prussing, Navjot Singh, John Kelly, Erasmus Schneider, Erica Lasek-Nesselquist |
| EPI_ISL_1920078 | Columbia University Irving Medical Center | Wadsworth Center, New York State Department of Health | Kirsten St. George, Daryl M. Lamson, Alexis Russell, Matthew Shudt, Melissa A Leisner, Jonathan Plitnick, Catharine Prussing, Navjot Singh, John Kelly, Erasmus Schneider, Erica Lasek-Nesselquist |
| EPI_ISL_1920079, EPI_ISL_1920080 | NYC Pandemic Response Lab | Wadsworth Center, New York State Department of Health | Kirsten St. George, Daryl M. Lamson, Alexis Russell, Matthew Shudt, Melissa A Leisner, Jonathan Plitnick, Catharine Prussing, Navjot Singh, John Kelly, Erasmus Schneider, Erica Lasek-Nesselquist |
| EPI_ISL_1920081, EPI_ISL_1920082 | Westchester Medical Center | Wadsworth Center, New York State Department of Health | Kirsten St. George, Daryl M. Lamson, Alexis Russell, Matthew Shudt, Melissa A Leisner, Jonathan Plitnick, Catharine Prussing, Navjot Singh, John Kelly, Erasmus Schneider, Erica Lasek-Nesselquist |
| EPI_ISL_1920083 | NYC Pandemic Response Lab | Wadsworth Center, New York State Department of Health | Kirsten St. George, Daryl M. Lamson, Alexis Russell, Matthew Shudt, Melissa A Leisner, Jonathan Plitnick, Catharine Prussing, Navjot Singh, John Kelly, Erasmus Schneider, Erica Lasek-Nesselquist |
| EPI_ISL_1920084, EPI_ISL_1920085, EPI_ISL_1920086 | Montefiore Medical Center Laboratories | Wadsworth Center, New York State Department of Health | Kirsten St. George, Daryl M. Lamson, Alexis Russell, Matthew Shudt, Melissa A Leisner, Jonathan Plitnick, Catharine Prussing, Navjot Singh, John Kelly, Erasmus Schneider, Erica Lasek-Nesselquist |
| EPI_ISL_1920087, EPI_ISL_1920088 | NYC Pandemic Response Lab | Wadsworth Center, New York State Department of Health | Kirsten St. George, Daryl M. Lamson, Alexis Russell, Matthew Shudt, Melissa A Leisner, Jonathan Plitnick, Catharine Prussing, Navjot Singh, John Kelly, Erasmus Schneider, Erica Lasek-Nesselquist |
| EPI_ISL_1920089, EPI_ISL_1920090, EPI_ISL_1920091, EPI_ISL_1920092 | Montefiore Medical Center Laboratories | Wadsworth Center, New York State Department of Health | Kirsten St. George, Daryl M. Lamson, Alexis Russell, Matthew Shudt, Melissa A Leisner, Jonathan Plitnick, Catharine Prussing, Navjot Singh, John Kelly, Erasmus Schneider, Erica Lasek-Nesselquist |
| EPI_ISL_1920093 | Westchester Medical Center | Wadsworth Center, New York State Department of Health | Kirsten St. George, Daryl M. Lamson, Alexis Russell, Matthew Shudt, Melissa A Leisner, Jonathan Plitnick, Catharine Prussing, Navjot Singh, John Kelly, Erasmus Schneider, Erica Lasek-Nesselquist |
| EPI_ISL_1920094 | Columbia University Irving Medical Center | Wadsworth Center, New York State Department of Health | Kirsten St. George, Daryl M. Lamson, Alexis Russell, Matthew Shudt, Melissa A Leisner, Jonathan Plitnick, Catharine Prussing, Navjot Singh, John Kelly, Erasmus Schneider, Erica Lasek-Nesselquist |
| EPI_ISL_1920095 | NYC Pandemic Response Lab | Wadsworth Center, New York State Department of Health | Kirsten St. George, Daryl M. Lamson, Alexis Russell, Matthew Shudt, Melissa A Leisner, Jonathan Plitnick, Catharine Prussing, Navjot Singh, John Kelly, Erasmus Schneider, Erica Lasek-Nesselquist |
| EPI_ISL_1920096 | Montefiore Medical Center Laboratories | Wadsworth Center, New York State Department of Health | Kirsten St. George, Daryl M. Lamson, Alexis Russell, Matthew Shudt, Melissa A Leisner, Jonathan Plitnick, Catharine Prussing, Navjot Singh, John Kelly, Erasmus Schneider, Erica Lasek-Nesselquist |
| EPI_ISL_1920097 | New York Presbyterian Hospital | Wadsworth Center, New York State Department of Health | Kirsten St. George, Daryl M. Lamson, Alexis Russell, Matthew Shudt, Melissa A Leisner, Jonathan Plitnick, Catharine Prussing, Navjot Singh, John Kelly, Erasmus Schneider, Erica Lasek-Nesselquist |
| EPI_ISL_1920098, EPI_ISL_1920099, EPI_ISL_1920100 | Tempus Labs Inc | Wadsworth Center, New York State Department of Health | Kirsten St. George, Daryl M. Lamson, Alexis Russell, Matthew Shudt, Melissa A Leisner, Jonathan Plitnick, Catharine Prussing, Navjot Singh, John Kelly, Erasmus Schneider, Erica Lasek-Nesselquist |
| EPI_ISL_1920101 | Northwell Health Laboratories | Wadsworth Center, New York State Department of Health | Kirsten St. George, Daryl M. Lamson, Alexis Russell, Matthew Shudt, Melissa A Leisner, Jonathan Plitnick, Catharine Prussing, Navjot Singh, John Kelly, Erasmus Schneider, Erica Lasek-Nesselquist |
| EPI_ISL_1920102, EPI_ISL_1920103 | Tempus Labs Inc | Wadsworth Center, New York State Department of Health | Kirsten St. George, Daryl M. Lamson, Alexis Russell, Matthew Shudt, Melissa A Leisner, Jonathan Plitnick, Catharine Prussing, Navjot Singh, John Kelly, Erasmus Schneider, Erica Lasek-Nesselquist |
| EPI_ISL_1920104, EPI_ISL_1920105 | Albany Medical Center | Wadsworth Center, New York State Department of Health | Kirsten St. George, Daryl M. Lamson, Alexis Russell, Matthew Shudt, Melissa A Leisner, Jonathan Plitnick, Catharine Prussing, Navjot Singh, John Kelly, Erasmus Schneider, Erica Lasek-Nesselquist |
| EPI_ISL_1920106, EPI_ISL_1920107 | Tempus Labs Inc | Wadsworth Center, New York State Department of Health | Kirsten St. George, Daryl M. Lamson, Alexis Russell, Matthew Shudt, Melissa A Leisner, Jonathan Plitnick, Catharine Prussing, Navjot Singh, John Kelly, Erasmus Schneider, Erica Lasek-Nesselquist |
| EPI_ISL_1920108, EPI_ISL_1920109, EPI_ISL_1920110, EPI_ISL_1920111, EPI_ISL_1920112 | Albany Medical Center | Wadsworth Center, New York State Department of Health | Kirsten St. George, Daryl M. Lamson, Alexis Russell, Matthew Shudt, Melissa A Leisner, Jonathan Plitnick, Catharine Prussing, Navjot Singh, John Kelly, Erasmus Schneider, Erica Lasek-Nesselquist |
| EPI_ISL_1920113, EPI_ISL_1920114 | Wadsworth Center, New York State Department of Health | Wadsworth Center, New York State Department of Health | Kirsten St. George, Daryl M. Lamson, Alexis Russell, Matthew Shudt, Melissa A Leisner, Jonathan Plitnick, Catharine Prussing, Navjot Singh, John Kelly, Erasmus Schneider, Erica Lasek-Nesselquist |
| EPI_ISL_1920115, EPI_ISL_1920116, EPI_ISL_1920117, EPI_ISL_1920118 | Tempus Labs Inc | Wadsworth Center, New York State Department of Health | Kirsten St. George, Daryl M. Lamson, Alexis Russell, Matthew Shudt, Melissa A Leisner, Jonathan Plitnick, Catharine Prussing, Navjot Singh, John Kelly, Erasmus Schneider, Erica Lasek-Nesselquist |
| EPI_ISL_1920119, EPI_ISL_1920120 | Glens Falls Hospital Laboratory | Wadsworth Center, New York State Department of Health | Kirsten St. George, Daryl M. Lamson, Alexis Russell, Matthew Shudt, Melissa A Leisner, Jonathan Plitnick, Catharine Prussing, Navjot Singh, John Kelly, Erasmus Schneider, Erica Lasek-Nesselquist |
| EPI_ISL_1920121 | Wadsworth Center, New York State Department of Health | Wadsworth Center, New York State Department of Health | Kirsten St. George, Daryl M. Lamson, Alexis Russell, Matthew Shudt, Melissa A Leisner, Jonathan Plitnick, Catharine Prussing, Navjot Singh, John Kelly, Erasmus Schneider, Erica Lasek-Nesselquist |
| EPI_ISL_1920122 | Pulaski Urgent Care | Wadsworth Center, New York State Department of Health | Kirsten St. George, Daryl M. Lamson, Alexis Russell, Matthew Shudt, Melissa A Leisner, Jonathan Plitnick, Catharine Prussing, Navjot Singh, John Kelly, Erasmus Schneider, Erica Lasek-Nesselquist |
| EPI_ISL_1920123 | Tempus Labs Inc | Wadsworth Center, New York State Department of Health | Kirsten St. George, Daryl M. Lamson, Alexis Russell, Matthew Shudt, Melissa A Leisner, Jonathan Plitnick, Catharine Prussing, Navjot Singh, John Kelly, Erasmus Schneider, Erica Lasek-Nesselquist |
| EPI_ISL_1920124, EPI_ISL_1920125, EPI_ISL_1920126, EPI_ISL_1920127, EPI_ISL_1920128, EPI_ISL_1920129, EPI_ISL_1920130, EPI_ISL_1920131 | URMC LABS | Wadsworth Center, New York State Department of Health | Kirsten St. George, Daryl M. Lamson, Alexis Russell, Matthew Shudt, Melissa A Leisner, Jonathan Plitnick, Catharine Prussing, Navjot Singh, John Kelly, Erasmus Schneider, Erica Lasek-Nesselquist |
| EPI_ISL_1920132 | Northwell Health Laboratories | Wadsworth Center, New York State Department of Health | Kirsten St. George, Daryl M. Lamson, Alexis Russell, Matthew Shudt, Melissa A Leisner, Jonathan Plitnick, Catharine Prussing, Navjot Singh, John Kelly, Erasmus Schneider, Erica Lasek-Nesselquist |
| EPI_ISL_1920133, EPI_ISL_1920134, EPI_ISL_1920135, EPI_ISL_1920136 | New York Presbyterian Hospital | Wadsworth Center, New York State Department of Health | Kirsten St. George, Daryl M. Lamson, Alexis Russell, Matthew Shudt, Melissa A Leisner, Jonathan Plitnick, Catharine Prussing, Navjot Singh, John Kelly, Erasmus Schneider, Erica Lasek-Nesselquist |
| EPI_ISL_1920137 | Columbia University Irving Medical Center | Wadsworth Center, New York State Department of Health | Kirsten St. George, Daryl M. Lamson, Alexis Russell, Matthew Shudt, Melissa A Leisner, Jonathan Plitnick, Catharine Prussing, Navjot Singh, John Kelly, Erasmus Schneider, Erica Lasek-Nesselquist |
| EPI_ISL_1920138 | New York Presbyterian Hospital | Wadsworth Center, New York State Department of Health | Kirsten St. George, Daryl M. Lamson, Alexis Russell, Matthew Shudt, Melissa A Leisner, Jonathan Plitnick, Catharine Prussing, Navjot Singh, John Kelly, Erasmus Schneider, Erica Lasek-Nesselquist |

| | | | |
|---|---|---|---|
| EPI_ISL_1920139, EPI_ISL_1920140, EPI_ISL_1920141, EPI_ISL_1920142, EPI_ISL_1920143, EPI_ISL_1920144, EPI_ISL_1920145, EPI_ISL_1920146 | Columbia University Irving Medical Center | Wadsworth Center, New York State Department of Health | Kirsten St. George, Daryl M. Lamson, Alexis Russell, Matthew Shudt, Melissa A Leisner, Jonathan Plitnick, Catharine Prussing, Navjot Singh, John Kelly, Erasmus Schneider, Erica Lasek-Nesselquist |
| EPI_ISL_1920147 | New York Presbyterian Hospital | Wadsworth Center, New York State Department of Health | Kirsten St. George, Daryl M. Lamson, Alexis Russell, Matthew Shudt, Melissa A Leisner, Jonathan Plitnick, Catharine Prussing, Navjot Singh, John Kelly, Erasmus Schneider, Erica Lasek-Nesselquist |
| EPI_ISL_1920148, EPI_ISL_1920149 | Columbia University Irving Medical Center | Wadsworth Center, New York State Department of Health | Kirsten St. George, Daryl M. Lamson, Alexis Russell, Matthew Shudt, Melissa A Leisner, Jonathan Plitnick, Catharine Prussing, Navjot Singh, John Kelly, Erasmus Schneider, Erica Lasek-Nesselquist |
| EPI_ISL_1920150 | New York Presbyterian Hospital | Wadsworth Center, New York State Department of Health | Kirsten St. George, Daryl M. Lamson, Alexis Russell, Matthew Shudt, Melissa A Leisner, Jonathan Plitnick, Catharine Prussing, Navjot Singh, John Kelly, Erasmus Schneider, Erica Lasek-Nesselquist |
| EPI_ISL_1920151 | Northwell Health Laboratories | Wadsworth Center, New York State Department of Health | Kirsten St. George, Daryl M. Lamson, Alexis Russell, Matthew Shudt, Melissa A Leisner, Jonathan Plitnick, Catharine Prussing, Navjot Singh, John Kelly, Erasmus Schneider, Erica Lasek-Nesselquist |
| EPI_ISL_1920152, EPI_ISL_1920153 | Westchester Medical Center | Wadsworth Center, New York State Department of Health | Kirsten St. George, Daryl M. Lamson, Alexis Russell, Matthew Shudt, Melissa A Leisner, Jonathan Plitnick, Catharine Prussing, Navjot Singh, John Kelly, Erasmus Schneider, Erica Lasek-Nesselquist |
| EPI_ISL_1920154 | Columbia University Irving Medical Center | Wadsworth Center, New York State Department of Health | Kirsten St. George, Daryl M. Lamson, Alexis Russell, Matthew Shudt, Melissa A Leisner, Jonathan Plitnick, Catharine Prussing, Navjot Singh, John Kelly, Erasmus Schneider, Erica Lasek-Nesselquist |
| EPI_ISL_1920155 | Westchester Medical Center | Wadsworth Center, New York State Department of Health | Kirsten St. George, Daryl M. Lamson, Alexis Russell, Matthew Shudt, Melissa A Leisner, Jonathan Plitnick, Catharine Prussing, Navjot Singh, John Kelly, Erasmus Schneider, Erica Lasek-Nesselquist |
| EPI_ISL_1920156 | Columbia University Irving Medical Center | Wadsworth Center, New York State Department of Health | Kirsten St. George, Daryl M. Lamson, Alexis Russell, Matthew Shudt, Melissa A Leisner, Jonathan Plitnick, Catharine Prussing, Navjot Singh, John Kelly, Erasmus Schneider, Erica Lasek-Nesselquist |
| EPI_ISL_1920157 | Memorial Sloan Kettering Cancer Center | Wadsworth Center, New York State Department of Health | Kirsten St. George, Daryl M. Lamson, Alexis Russell, Matthew Shudt, Melissa A Leisner, Jonathan Plitnick, Catharine Prussing, Navjot Singh, John Kelly, Erasmus Schneider, Erica Lasek-Nesselquist |
| EPI_ISL_1920158 | Westchester Medical Center | Wadsworth Center, New York State Department of Health | Kirsten St. George, Daryl M. Lamson, Alexis Russell, Matthew Shudt, Melissa A Leisner, Jonathan Plitnick, Catharine Prussing, Navjot Singh, John Kelly, Erasmus Schneider, Erica Lasek-Nesselquist |
| EPI_ISL_1920159 | Montefiore Medical Center Laboratories | Wadsworth Center, New York State Department of Health | Kirsten St. George, Daryl M. Lamson, Alexis Russell, Matthew Shudt, Melissa A Leisner, Jonathan Plitnick, Catharine Prussing, Navjot Singh, John Kelly, Erasmus Schneider, Erica Lasek-Nesselquist |
| EPI_ISL_1920160 | Columbia University Irving Medical Center | Wadsworth Center, New York State Department of Health | Kirsten St. George, Daryl M. Lamson, Alexis Russell, Matthew Shudt, Melissa A Leisner, Jonathan Plitnick, Catharine Prussing, Navjot Singh, John Kelly, Erasmus Schneider, Erica Lasek-Nesselquist |
| EPI_ISL_1920161 | Montefiore Medical Center Laboratories | Wadsworth Center, New York State Department of Health | Kirsten St. George, Daryl M. Lamson, Alexis Russell, Matthew Shudt, Melissa A Leisner, Jonathan Plitnick, Catharine Prussing, Navjot Singh, John Kelly, Erasmus Schneider, Erica Lasek-Nesselquist |
| EPI_ISL_1920162 | New York Presbyterian Hospital | Wadsworth Center, New York State Department of Health | Kirsten St. George, Daryl M. Lamson, Alexis Russell, Matthew Shudt, Melissa A Leisner, Jonathan Plitnick, Catharine Prussing, Navjot Singh, John Kelly, Erasmus Schneider, Erica Lasek-Nesselquist |
| EPI_ISL_1920163 | NYC Pandemic Response Lab | Wadsworth Center, New York State Department of Health | Kirsten St. George, Daryl M. Lamson, Alexis Russell, Matthew Shudt, Melissa A Leisner, Jonathan Plitnick, Catharine Prussing, Navjot Singh, John Kelly, Erasmus Schneider, Erica Lasek-Nesselquist |
| EPI_ISL_1920164, EPI_ISL_1920165 | New York Presbyterian Hospital | Wadsworth Center, New York State Department of Health | Kirsten St. George, Daryl M. Lamson, Alexis Russell, Matthew Shudt, Melissa A Leisner, Jonathan Plitnick, Catharine Prussing, Navjot Singh, John Kelly, Erasmus Schneider, Erica Lasek-Nesselquist |
| EPI_ISL_1920166 | Northwell Health Laboratories | Wadsworth Center, New York State Department of Health | Kirsten St. George, Daryl M. Lamson, Alexis Russell, Matthew Shudt, Melissa A Leisner, Jonathan Plitnick, Catharine Prussing, Navjot Singh, John Kelly, Erasmus Schneider, Erica Lasek-Nesselquist |
| EPI_ISL_1920167 | Westchester Medical Center | Wadsworth Center, New York State Department of Health | Kirsten St. George, Daryl M. Lamson, Alexis Russell, Matthew Shudt, Melissa A Leisner, Jonathan Plitnick, Catharine Prussing, Navjot Singh, John Kelly, Erasmus Schneider, Erica Lasek-Nesselquist |
| EPI_ISL_1920168, EPI_ISL_1920169, EPI_ISL_1920170 | Northwell Health Laboratories | Wadsworth Center, New York State Department of Health | Kirsten St. George, Daryl M. Lamson, Alexis Russell, Matthew Shudt, Melissa A Leisner, Jonathan Plitnick, Catharine Prussing, Navjot Singh, John Kelly, Erasmus Schneider, Erica Lasek-Nesselquist |
| EPI_ISL_1920171 | New York Presbyterian Hospital | Wadsworth Center, New York State Department of Health | Kirsten St. George, Daryl M. Lamson, Alexis Russell, Matthew Shudt, Melissa A Leisner, Jonathan Plitnick, Catharine Prussing, Navjot Singh, John Kelly, Erasmus Schneider, Erica Lasek-Nesselquist |
| EPI_ISL_1920172, EPI_ISL_1920173, EPI_ISL_1920174 | Westchester Medical Center | Wadsworth Center, New York State Department of Health | Kirsten St. George, Daryl M. Lamson, Alexis Russell, Matthew Shudt, Melissa A Leisner, Jonathan Plitnick, Catharine Prussing, Navjot Singh, John Kelly, Erasmus Schneider, Erica Lasek-Nesselquist |
| EPI_ISL_1920997, EPI_ISL_1920998, EPI_ISL_1920999, EPI_ISL_1921000, EPI_ISL_1921001, EPI_ISL_1921002, EPI_ISL_1921003, EPI_ISL_1921004, EPI_ISL_1921005, EPI_ISL_1921006, EPI_ISL_1921007, EPI_ISL_1921008, EPI_ISL_1921009, EPI_ISL_1921010 | | | |
| see above | URMC LABS | Wadsworth Center, New York State Department of Health | Kirsten St. George, Daryl M. Lamson, Alexis Russell, Matthew Shudt, Melissa A Leisner, Jonathan Plitnick, Catharine Prussing, Navjot Singh, John Kelly, Erasmus Schneider, Erica Lasek-Nesselquist |
| EPI_ISL_1921011, EPI_ISL_1921012, EPI_ISL_1921013, EPI_ISL_1921014, EPI_ISL_1921015, EPI_ISL_1921016, EPI_ISL_1921017 | SUNY Upstate Medical University | Wadsworth Center, New York State Department of Health | Kirsten St. George, Daryl M. Lamson, Alexis Russell, Matthew Shudt, Melissa A Leisner, Jonathan Plitnick, Catharine Prussing, Navjot Singh, John Kelly, Erasmus Schneider, Erica Lasek-Nesselquist |
| EPI_ISL_1921018 | Albany Medical Center | Wadsworth Center, New York State Department of Health | Kirsten St. George, Daryl M. Lamson, Alexis Russell, Matthew Shudt, Melissa A Leisner, Jonathan Plitnick, Catharine Prussing, Navjot Singh, John Kelly, Erasmus Schneider, Erica Lasek-Nesselquist |
| EPI_ISL_1921019, EPI_ISL_1921020, EPI_ISL_1921021 | SUNY Upstate Medical University | Wadsworth Center, New York State Department of Health | Kirsten St. George, Daryl M. Lamson, Alexis Russell, Matthew Shudt, Melissa A Leisner, Jonathan Plitnick, Catharine Prussing, Navjot Singh, John Kelly, Erasmus Schneider, Erica Lasek-Nesselquist |
| EPI_ISL_1921022 | Albany Medical Center | Wadsworth Center, New York State Department of Health | Kirsten St. George, Daryl M. Lamson, Alexis Russell, Matthew Shudt, Melissa A Leisner, Jonathan Plitnick, Catharine Prussing, Navjot Singh, John Kelly, Erasmus Schneider, Erica Lasek-Nesselquist |
| EPI_ISL_1921023, EPI_ISL_1921024, EPI_ISL_1921025, EPI_ISL_1921026, EPI_ISL_1921027 | SUNY Upstate Medical University | Wadsworth Center, New York State Department of Health | Kirsten St. George, Daryl M. Lamson, Alexis Russell, Matthew Shudt, Melissa A Leisner, Jonathan Plitnick, Catharine Prussing, Navjot Singh, John Kelly, Erasmus Schneider, Erica Lasek-Nesselquist |
| EPI_ISL_1921028, EPI_ISL_1921029, EPI_ISL_1921030, EPI_ISL_1921031, EPI_ISL_1921032, EPI_ISL_1921033, EPI_ISL_1921034, EPI_ISL_1921035, EPI_ISL_1921036 | Albany Medical Center | Wadsworth Center, New York State Department of Health | Kirsten St. George, Daryl M. Lamson, Alexis Russell, Matthew Shudt, Melissa A Leisner, Jonathan Plitnick, Catharine Prussing, Navjot Singh, John Kelly, Erasmus Schneider, Erica Lasek-Nesselquist |
| EPI_ISL_1921037 | Glens Falls Hospital Laboratory | Wadsworth Center, New York State Department of Health | Kirsten St. George, Daryl M. Lamson, Alexis Russell, Matthew Shudt, Melissa A Leisner, Jonathan Plitnick, Catharine Prussing, Navjot Singh, John Kelly, Erasmus Schneider, Erica Lasek-Nesselquist |
| EPI_ISL_1921038, EPI_ISL_1921039 | SUNY Upstate Medical University | Wadsworth Center, New York State Department of Health | Kirsten St. George, Daryl M. Lamson, Alexis Russell, Matthew Shudt, Melissa A Leisner, Jonathan Plitnick, Catharine Prussing, Navjot Singh, John Kelly, Erasmus Schneider, Erica Lasek-Nesselquist |
| EPI_ISL_1921040, EPI_ISL_1921041, EPI_ISL_1921042, EPI_ISL_1921043, EPI_ISL_1921044, EPI_ISL_1921045, EPI_ISL_1921046, EPI_ISL_1921047, EPI_ISL_1921048, EPI_ISL_1921049, EPI_ISL_1921050, EPI_ISL_1921051, EPI_ISL_1921052, EPI_ISL_1921053, EPI_ISL_1921054, EPI_ISL_1921055, EPI_ISL_1921056, | | | |

| | | | |
|---|---|---|---|
| EPI_ISL_1921057, EPI_ISL_1921058, EPI_ISL_1921059, EPI_ISL_1921060, EPI_ISL_1921061, EPI_ISL_1921062, EPI_ISL_1921063, EPI_ISL_1921064, EPI_ISL_1921065, EPI_ISL_1921066, EPI_ISL_1921067, EPI_ISL_1921068, EPI_ISL_1921069, EPI_ISL_1921070, EPI_ISL_1921071, EPI_ISL_1921072, EPI_ISL_1921073, EPI_ISL_1921074, EPI_ISL_1921075, EPI_ISL_1921076, EPI_ISL_1921077, EPI_ISL_1921078, EPI_ISL_1921079 | | | |
| see above | URMC LABS | Wadsworth Center, New York State Department of Health | Kirsten St. George, Daryl M. Lamson, Alexis Russell, Matthew Shudt, Melissa A Leisner, Jonathan Plitnick, Catharine Prussing, Navjot Singh, John Kelly, Erasmus Schneider, Erica Lasek-Nesselquist |
| EPI_ISL_1922710, EPI_ISL_1922712, EPI_ISL_1922713, EPI_ISL_1922714, EPI_ISL_1922715, EPI_ISL_1922716, EPI_ISL_1922717, EPI_ISL_1922718, EPI_ISL_1922719, EPI_ISL_1922720, EPI_ISL_1922721, EPI_ISL_1922722, EPI_ISL_1922723, EPI_ISL_1922724, EPI_ISL_1922725, EPI_ISL_1922726, EPI_ISL_1922727, EPI_ISL_1922728, EPI_ISL_1922729, EPI_ISL_1922730, EPI_ISL_1922731, EPI_ISL_1922732, EPI_ISL_1922733, EPI_ISL_1922734, EPI_ISL_1922735, EPI_ISL_1922736, EPI_ISL_1922737, EPI_ISL_1922738, EPI_ISL_1922739, EPI_ISL_1922740, EPI_ISL_1922741, EPI_ISL_1922742, EPI_ISL_1922743, EPI_ISL_1922744, EPI_ISL_1922745, EPI_ISL_1922746, EPI_ISL_1922747, EPI_ISL_1922748, EPI_ISL_1922749, EPI_ISL_1922750, EPI_ISL_1922751, EPI_ISL_1922752, EPI_ISL_1922754, EPI_ISL_1922755, EPI_ISL_1922756, EPI_ISL_1922757, EPI_ISL_1922758, EPI_ISL_1922759, EPI_ISL_1922760, EPI_ISL_1922761, EPI_ISL_1922763, EPI_ISL_1922765, EPI_ISL_1922766, EPI_ISL_1922768, EPI_ISL_1922769, EPI_ISL_1922770, EPI_ISL_1922771, EPI_ISL_1922772, EPI_ISL_1922773, EPI_ISL_1922774, EPI_ISL_1922775, EPI_ISL_1922776, EPI_ISL_1922777, EPI_ISL_1922778, EPI_ISL_1922779, EPI_ISL_1922780, EPI_ISL_1922781, EPI_ISL_1922782, EPI_ISL_1922783, EPI_ISL_1922784, EPI_ISL_1922786, EPI_ISL_1922787, EPI_ISL_1922788, EPI_ISL_1922790, EPI_ISL_1922793, EPI_ISL_1922794, EPI_ISL_1922795, EPI_ISL_1922796, EPI_ISL_1922797, EPI_ISL_1922798, EPI_ISL_1922799, EPI_ISL_1922800, EPI_ISL_1922801, EPI_ISL_1922802, EPI_ISL_1922803, EPI_ISL_1922804, EPI_ISL_1922805, EPI_ISL_1922806, EPI_ISL_1922807, EPI_ISL_1922808, EPI_ISL_1922809, EPI_ISL_1922810, EPI_ISL_1922811, EPI_ISL_1922812, EPI_ISL_1922813, EPI_ISL_1922814, EPI_ISL_1922815, EPI_ISL_1922816, EPI_ISL_1922817, EPI_ISL_1922818, EPI_ISL_1922819, EPI_ISL_1922820, EPI_ISL_1922821, EPI_ISL_1922822, EPI_ISL_1922823, EPI_ISL_1922824, EPI_ISL_1922825, EPI_ISL_1922826, EPI_ISL_1922827, EPI_ISL_1922828, EPI_ISL_1922829, EPI_ISL_1922830, EPI_ISL_1922831, EPI_ISL_1922832, EPI_ISL_1922833, EPI_ISL_1922834, EPI_ISL_1922835, EPI_ISL_1922836, EPI_ISL_1922837, EPI_ISL_1922838, EPI_ISL_1922840, EPI_ISL_1922842, EPI_ISL_1922844, EPI_ISL_1922845, EPI_ISL_1922846, EPI_ISL_1922847, EPI_ISL_1922848, EPI_ISL_1922849, EPI_ISL_1922851, EPI_ISL_1922852, EPI_ISL_1922853, EPI_ISL_1922854, EPI_ISL_1922855, EPI_ISL_1922856, EPI_ISL_1922857, EPI_ISL_1922858, EPI_ISL_1922860, EPI_ISL_1922861, EPI_ISL_1922862, EPI_ISL_1922863, EPI_ISL_1922864, EPI_ISL_1922865, EPI_ISL_1922866, EPI_ISL_1922867, EPI_ISL_1922868, EPI_ISL_1922869, EPI_ISL_1922870, EPI_ISL_1922871, EPI_ISL_1922872 | | | |
| see above | NYU Langone Health | Departments of Pathology and Medicine, New York University School of Medicine | Adriana Heguy, Dacia Dimartino, Emily Guzman, Christian Marier, Peter Meyn, Sitharam Ramaswami, Gael Westby, Paul Zappile, Yutong Zhang, Paolo Cotzia, Guiqing Wang |
| EPI_ISL_1922972, EPI_ISL_1922974, EPI_ISL_1922995, EPI_ISL_1923020, EPI_ISL_1923029, EPI_ISL_1923040, EPI_ISL_1923080, EPI_ISL_1923092, EPI_ISL_1923116, EPI_ISL_1923149 | Aegis Sciences Corporation | Centers for Disease Control and Prevention Division of Viral Diseases, Pathogen Discovery | Dakota Howard, Dhwani Batra, Peter W. Cook, Kara Moser, Adrian Paskey, Jason Caravas, Benjamin Rambo-Martin, Shatavia Morrison, Christopher Gulvick, Scott Sammons, Yvette Unoarumhi, Darlene Wagner, Matthew Schmerer, Cyndi Clark, Patrick Campbell, Rob Case, Vikramsinha Ghorpade, Holly Houdeshell, Ola Kvalvaag, Dillon Nall, Ethan Sanders, Alec Vest, Shaun Westlund, Matthew Hardison, Clinton R. Paden, Duncan MacCannell |
| EPI_ISL_1923513, EPI_ISL_1923597, EPI_ISL_1923605, EPI_ISL_1923719, EPI_ISL_1924033, EPI_ISL_1924207 | Helix/Illumina | Centers for Disease Control and Prevention Division of Viral Diseases, Pathogen Discovery | Dakota Howard, Dhwani Batra, Peter W. Cook, Kara Moser, Adrian Paskey, Jason Caravas, Benjamin Rambo-Martin, Shatavia Morrison, Christopher Gulvick, Scott Sammons, Yvette Unoarumhi, Darlene Wagner, Matthew Schmerer, Eileen de Feo, Jan Antico, Christine Tran, Matthew Tolentino, Shannon Wickline, Kim Gietzen, Brad Sickler, Jingtao Liu, Eric Allen, Phil Febbo, Nicole L. Washington, Simon White, Geraint Levan, Kelly Schiabor Barrett, Elizabeth Cirulli, Alexandre Bolze, Ary Ascencio, Charlotte Rivera-Garcia, Ryan Cho, Jason Nguyen, Sherry Wang, Jimmy Ramirez, Tyler Cassens, Efren Sandoval, Magnus Isaksson, William Lee, David Becker, Marc Laurent, James Lu, Clinton R. Paden, Duncan MacCannell |
| EPI_ISL_1924286, EPI_ISL_1924290, EPI_ISL_1924295, EPI_ISL_1924297, EPI_ISL_1924298, EPI_ISL_1924300, EPI_ISL_1924301, EPI_ISL_1924308, EPI_ISL_1924311, EPI_ISL_1924316, EPI_ISL_1924318, EPI_ISL_1924321, EPI_ISL_1924322, EPI_ISL_1924323, EPI_ISL_1924328, EPI_ISL_1924330, EPI_ISL_1924332, EPI_ISL_1924340, EPI_ISL_1924350, EPI_ISL_1924353, EPI_ISL_1924358, EPI_ISL_1924361, EPI_ISL_1924362, EPI_ISL_1924364, EPI_ISL_1924365, EPI_ISL_1924371, EPI_ISL_1924374, EPI_ISL_1924376, EPI_ISL_1924377, EPI_ISL_1924378, EPI_ISL_1924379, EPI_ISL_1924381, EPI_ISL_1924382, EPI_ISL_1924385, EPI_ISL_1924400, EPI_ISL_1924407, EPI_ISL_1924409, EPI_ISL_1924410, EPI_ISL_1924417, EPI_ISL_1924420, EPI_ISL_1924422, EPI_ISL_1924423, EPI_ISL_1924424, EPI_ISL_1924427, EPI_ISL_1924429, EPI_ISL_1924430, EPI_ISL_1924514 | | | |
| see above | Quest Diagnostics Incorporated | Centers for Disease Control and Prevention Division of Viral Diseases, Pathogen Discovery | Dakota Howard, Dhwani Batra, Peter W. Cook, Kara Moser, Adrian Paskey, Jason Caravas, Benjamin Rambo-Martin, Shatavia Morrison, Christopher Gulvick, Scott Sammons, Yvette Unoarumhi, Darlene Wagner, Matthew Schmerer, S. H. Rosenthal, A. Gerasimova, R. M. Kagan, B. Anderson, M. Hua, Y. Liu, L.E. Bernstein, K.E. Livingston, A. Perez, I. A. Shlyakhter, R. V. Rolando, R. Owen, P. Tanpaiboon, F. Lacbawan, Clinton R. Paden, Duncan MacCannell |
| EPI_ISL_1925228, EPI_ISL_1925248, EPI_ISL_1925249, EPI_ISL_1925250, EPI_ISL_1925251, EPI_ISL_1925252, EPI_ISL_1925275, EPI_ISL_1925303, EPI_ISL_1925304, EPI_ISL_1925311, EPI_ISL_1925329, EPI_ISL_1925358, EPI_ISL_1925360, EPI_ISL_1925380, EPI_ISL_1925382, EPI_ISL_1925421, EPI_ISL_1925454, EPI_ISL_1925459, EPI_ISL_1925464, EPI_ISL_1925477, EPI_ISL_1925498, EPI_ISL_1925502, EPI_ISL_1925541, EPI_ISL_1925545, EPI_ISL_1925550, EPI_ISL_1925551, EPI_ISL_1925584, EPI_ISL_1925604, EPI_ISL_1925617 | | | |
| see above | Aegis Sciences Corporation | Centers for Disease Control and Prevention Division of Viral Diseases, Pathogen Discovery | Dakota Howard, Dhwani Batra, Peter W. Cook, Kara Moser, Adrian Paskey, Jason Caravas, Benjamin Rambo-Martin, Shatavia Morrison, Christopher Gulvick, Scott Sammons, Yvette Unoarumhi, Darlene Wagner, Matthew Schmerer, Cyndi Clark, Patrick Campbell, Rob Case, Vikramsinha Ghorpade, Holly Houdeshell, Ola Kvalvaag, Dillon Nall, Ethan Sanders, Alec Vest, Shaun Westlund, Matthew Hardison, Clinton R. Paden, Duncan MacCannell |
| EPI_ISL_1925748, EPI_ISL_1925789, EPI_ISL_1925924, EPI_ISL_1925980, EPI_ISL_1925983, EPI_ISL_1925986, EPI_ISL_1926045, EPI_ISL_1926108, EPI_ISL_1926229, EPI_ISL_1926230, EPI_ISL_1926231, EPI_ISL_1926253, EPI_ISL_1926262, EPI_ISL_1926292, EPI_ISL_1926296, EPI_ISL_1926297, EPI_ISL_1926300, EPI_ISL_1926301, EPI_ISL_1926302, EPI_ISL_1926303, EPI_ISL_1926312, EPI_ISL_1926323, EPI_ISL_1926340, EPI_ISL_1926372, EPI_ISL_1926412, EPI_ISL_1926413, EPI_ISL_1926414, EPI_ISL_1926415, EPI_ISL_1926424, EPI_ISL_1926425, EPI_ISL_1926426, EPI_ISL_1926427, EPI_ISL_1926428, EPI_ISL_1926548, EPI_ISL_1926569, EPI_ISL_1926571, EPI_ISL_1926572, EPI_ISL_1926573, EPI_ISL_1926574, EPI_ISL_1926575, EPI_ISL_1926576, EPI_ISL_1926577, EPI_ISL_1926578, EPI_ISL_1927480, EPI_ISL_1927554, EPI_ISL_1927587, EPI_ISL_1927610, EPI_ISL_1927613, EPI_ISL_1927616, EPI_ISL_1927619, EPI_ISL_1927622, EPI_ISL_1927625, EPI_ISL_1927639, EPI_ISL_1927868, EPI_ISL_1927871, EPI_ISL_1927899, EPI_ISL_1927947, EPI_ISL_1927977, EPI_ISL_1927989, EPI_ISL_1927992, EPI_ISL_1927998, EPI_ISL_1928001, EPI_ISL_1928004, EPI_ISL_1928007, EPI_ISL_1928010, EPI_ISL_1928013, EPI_ISL_1928034, EPI_ISL_1928040, EPI_ISL_1928179, EPI_ISL_1928182, EPI_ISL_1928210, EPI_ISL_1928231, EPI_ISL_1928268, EPI_ISL_1928282, EPI_ISL_1928292, EPI_ISL_1928295, EPI_ISL_1928314, EPI_ISL_1928371, EPI_ISL_1928372, EPI_ISL_1928373, EPI_ISL_1928374, EPI_ISL_1928384, EPI_ISL_1928390, EPI_ISL_1928393, EPI_ISL_1928589, EPI_ISL_1928606, EPI_ISL_1928609, EPI_ISL_1928646, EPI_ISL_1928651, EPI_ISL_1928652, EPI_ISL_1928653, EPI_ISL_1928663, EPI_ISL_1928664, EPI_ISL_1928665, EPI_ISL_1928706, EPI_ISL_1928712, EPI_ISL_1928835, EPI_ISL_1928867, EPI_ISL_1928931, EPI_ISL_1928938, EPI_ISL_1928945, EPI_ISL_1929024, EPI_ISL_1929041, EPI_ISL_1929056, EPI_ISL_1929057, EPI_ISL_1929285, EPI_ISL_1929301, EPI_ISL_1929325, EPI_ISL_1929328, EPI_ISL_1929338, EPI_ISL_1929392, EPI_ISL_1929657, EPI_ISL_1929870, EPI_ISL_1929902, EPI_ISL_1930006, EPI_ISL_1930009, EPI_ISL_1930022, EPI_ISL_1930060, EPI_ISL_1930062, EPI_ISL_1930066, EPI_ISL_1930158, EPI_ISL_1930162, EPI_ISL_1930184, EPI_ISL_1930244, EPI_ISL_1930247, EPI_ISL_1930250, EPI_ISL_1930261, EPI_ISL_1930264, EPI_ISL_1930292, EPI_ISL_1930295, EPI_ISL_1930356, EPI_ISL_1930358 | | | |
| see above | Laboratory Corporation of America | Centers for Disease Control and Prevention Division of Viral Diseases, Pathogen Discovery | Dakota Howard, Dhwani Batra, Peter W. Cook, Kara Moser, Adrian Paskey, Jason Caravas, Benjamin Rambo-Martin, Shatavia Morrison, Christopher Gulvick, Scott Sammons, Yvette Unoarumhi, Darlene Wagner, Matthew Schmerer, Minoo Agarwal, Eyad Almasri, Debbie Boles, Ayla Burns, Nuthawin Charoensri, Oren Cohen, Susan Countryman, Mary Ann Cristobal, Bobbi Croy, Suzanne Dale, Hrushikesh Deshmukh, Amanda Douglas, Vincent Drouillon, Marcia Eisenberg, Howard Engler, Rama Ghatti, Prashant Gupta, Susan Hicks, Jake Humphrey, Lax Iyer, Manoj Jain, Mohan Kolli, Brian Krueger, Tim Kuphal, Stanley Letovsky, Michael Levandoski, Craig Lukasik, Jonathan Meltzer, Brian Norvell, Mindy Nye, Scott Parker, Christos Petropoulos, John Pruitt, Steven Ragan, Scott Ryan, Mike Sapeta, Jana Schroth, Suresh Babu Selvaraju, Goran Stevovic, Amanda Suchanek, Andrea Throop, Lyndon Tilson, Thomas Urban, Joe Voshell, Kimberly Wagner, Jonathan Williams, Mary Williamson, Qian Zeng, Tricia Zwiefelhofer, Clinton R. Paden, Duncan MacCannell |
| EPI_ISL_1935569, EPI_ISL_1935574 | Quest Diagnostics Incorporated | Centers for Disease Control and Prevention Division of Viral Diseases, Pathogen Discovery | Dakota Howard, Dhwani Batra, Peter W. Cook, Kara Moser, Adrian Paskey, Jason Caravas, Benjamin Rambo-Martin, Shatavia Morrison, Christopher Gulvick, Scott Sammons, Yvette Unoarumhi, Darlene Wagner, Matthew Schmerer, S. H. Rosenthal, A. Gerasimova, R. M. Kagan, B. Anderson, M. Hua, Y. Liu, L.E. Bernstein, K.E. Livingston, A. Perez, I. A. Shlyakhter, R. V. Rolando, R. Owen, P. Tanpaiboon, F. Lacbawan, Clinton R. Paden, Duncan MacCannell |
| EPI_ISL_1935607 | ALBANY MEDICAL CENTER | Wadsworth Center, New York State Department of Health | Kirsten St. George, Daryl M. Lamson, Alexis Russell, Matthew Shudt, Melissa A Leisner, Jonathan Plitnick, Catharine Prussing, Navjot Singh, John Kelly, Erasmus Schneider, Erica Lasek-Nesselquist |
| EPI_ISL_1935608, EPI_ISL_1935609 | NORTHWELL HEALTH LABORATORIES | Wadsworth Center, New York State Department of Health | Kirsten St. George, Daryl M. Lamson, Alexis Russell, Matthew Shudt, Melissa A Leisner, Jonathan Plitnick, Catharine Prussing, Navjot Singh, John Kelly, Erasmus Schneider, Erica Lasek-Nesselquist |
| EPI_ISL_1935610 | MONTEFIORE MEDICAL CENTER LABORATORIES | Wadsworth Center, New York State Department of Health | Kirsten St. George, Daryl M. Lamson, Alexis Russell, Matthew Shudt, Melissa A Leisner, Jonathan Plitnick, Catharine Prussing, Navjot Singh, John Kelly, Erasmus Schneider, Erica Lasek-Nesselquist |
| EPI_ISL_1935611 | URMC LABS | Wadsworth Center, New York State Department of Health | Kirsten St. George, Daryl M. Lamson, Alexis Russell, Matthew Shudt, Melissa A Leisner, Jonathan Plitnick, Catharine Prussing, Navjot Singh, John Kelly, Erasmus Schneider, Erica Lasek-Nesselquist |
| EPI_ISL_1935612 | WESTCHESTER MEDICAL CENTER | Wadsworth Center, New York State Department of Health | Kirsten St. George, Daryl M. Lamson, Alexis Russell, Matthew Shudt, Melissa A Leisner, Jonathan Plitnick, Catharine Prussing, Navjot Singh, John Kelly, Erasmus Schneider, Erica Lasek-Nesselquist |
| EPI_ISL_1935613 | TEMPUS LABS INC | Wadsworth Center, New York State Department of Health | Kirsten St. George, Daryl M. Lamson, Alexis Russell, Matthew Shudt, Melissa A Leisner, Jonathan Plitnick, Catharine Prussing, Navjot Singh, John Kelly, Erasmus Schneider, Erica Lasek-Nesselquist |
| EPI_ISL_1935614 | WESTCHESTER MEDICAL CENTER | Wadsworth Center, New York State Department of Health | Kirsten St. George, Daryl M. Lamson, Alexis Russell, Matthew Shudt, Melissa A Leisner, Jonathan Plitnick, Catharine Prussing, Navjot Singh, John Kelly, Erasmus Schneider, Erica Lasek-Nesselquist |
| EPI_ISL_1935615 | MONTEFIORE MEDICAL CENTER LABORATORIES | Wadsworth Center, New York State Department of Health | Kirsten St. George, Daryl M. Lamson, Alexis Russell, Matthew Shudt, Melissa A Leisner, Jonathan Plitnick, Catharine Prussing, Navjot Singh, John Kelly, Erasmus Schneider, Erica Lasek-Nesselquist |
| EPI_ISL_1935616 | URMC LABS | Wadsworth Center, New York State Department of Health | Kirsten St. George, Daryl M. Lamson, Alexis Russell, Matthew Shudt, Melissa A Leisner, Jonathan Plitnick, Catharine Prussing, Navjot Singh, John Kelly, Erasmus Schneider, Erica Lasek-Nesselquist |
| EPI_ISL_1935617 | BOSTON HEART DIAGNOSTICS CORP | Wadsworth Center, New York State Department of Health | Kirsten St. George, Daryl M. Lamson, Alexis Russell, Matthew Shudt, Melissa A Leisner, Jonathan Plitnick, Catharine Prussing, Navjot Singh, John Kelly, Erasmus Schneider, Erica Lasek-Nesselquist |

| | | | |
|---|---|---|---|
| EPI_ISL_1935618 | URMC LABS | Wadsworth Center, New York State Department of Health | Kirsten St. George, Daryl M. Lamson, Alexis Russell, Matthew Shudt, Melissa A Leisner, Jonathan Plitnick, Catharine Prussing, Navjot Singh, John Kelly, Erasmus Schneider, Erica Lasek-Nesselquist |
| EPI_ISL_1935619 | NYC Pandemic Response Lab | Wadsworth Center, New York State Department of Health | Kirsten St. George, Daryl M. Lamson, Alexis Russell, Matthew Shudt, Melissa A Leisner, Jonathan Plitnick, Catharine Prussing, Navjot Singh, John Kelly, Erasmus Schneider, Erica Lasek-Nesselquist |
| EPI_ISL_1935620 | BOSTON HEART DIAGNOSTICS CORP | Wadsworth Center, New York State Department of Health | Kirsten St. George, Daryl M. Lamson, Alexis Russell, Matthew Shudt, Melissa A Leisner, Jonathan Plitnick, Catharine Prussing, Navjot Singh, John Kelly, Erasmus Schneider, Erica Lasek-Nesselquist |
| EPI_ISL_1935621 | NORTHWELL HEALTH LABORATORIES | Wadsworth Center, New York State Department of Health | Kirsten St. George, Daryl M. Lamson, Alexis Russell, Matthew Shudt, Melissa A Leisner, Jonathan Plitnick, Catharine Prussing, Navjot Singh, John Kelly, Erasmus Schneider, Erica Lasek-Nesselquist |
| EPI_ISL_1935622, EPI_ISL_1935623 | URMC LABS | Wadsworth Center, New York State Department of Health | Kirsten St. George, Daryl M. Lamson, Alexis Russell, Matthew Shudt, Melissa A Leisner, Jonathan Plitnick, Catharine Prussing, Navjot Singh, John Kelly, Erasmus Schneider, Erica Lasek-Nesselquist |
| EPI_ISL_1935624 | NORTHWELL HEALTH LABORATORIES | Wadsworth Center, New York State Department of Health | Kirsten St. George, Daryl M. Lamson, Alexis Russell, Matthew Shudt, Melissa A Leisner, Jonathan Plitnick, Catharine Prussing, Navjot Singh, John Kelly, Erasmus Schneider, Erica Lasek-Nesselquist |
| EPI_ISL_1935625 | Columbia University Irving Medical Center | Wadsworth Center, New York State Department of Health | Kirsten St. George, Daryl M. Lamson, Alexis Russell, Matthew Shudt, Melissa A Leisner, Jonathan Plitnick, Catharine Prussing, Navjot Singh, John Kelly, Erasmus Schneider, Erica Lasek-Nesselquist |
| EPI_ISL_1935626 | NORTHWELL HEALTH LABORATORIES | Wadsworth Center, New York State Department of Health | Kirsten St. George, Daryl M. Lamson, Alexis Russell, Matthew Shudt, Melissa A Leisner, Jonathan Plitnick, Catharine Prussing, Navjot Singh, John Kelly, Erasmus Schneider, Erica Lasek-Nesselquist |
| EPI_ISL_1935627 | TEMPUS LABS INC | Wadsworth Center, New York State Department of Health | Kirsten St. George, Daryl M. Lamson, Alexis Russell, Matthew Shudt, Melissa A Leisner, Jonathan Plitnick, Catharine Prussing, Navjot Singh, John Kelly, Erasmus Schneider, Erica Lasek-Nesselquist |
| EPI_ISL_1935628 | NYC Pandemic Response Lab | Wadsworth Center, New York State Department of Health | Kirsten St. George, Daryl M. Lamson, Alexis Russell, Matthew Shudt, Melissa A Leisner, Jonathan Plitnick, Catharine Prussing, Navjot Singh, John Kelly, Erasmus Schneider, Erica Lasek-Nesselquist |
| EPI_ISL_1935629 | NORTHWELL HEALTH LABORATORIES | Wadsworth Center, New York State Department of Health | Kirsten St. George, Daryl M. Lamson, Alexis Russell, Matthew Shudt, Melissa A Leisner, Jonathan Plitnick, Catharine Prussing, Navjot Singh, John Kelly, Erasmus Schneider, Erica Lasek-Nesselquist |
| EPI_ISL_1935630 | ALBANY MEDICAL CENTER | Wadsworth Center, New York State Department of Health | Kirsten St. George, Daryl M. Lamson, Alexis Russell, Matthew Shudt, Melissa A Leisner, Jonathan Plitnick, Catharine Prussing, Navjot Singh, John Kelly, Erasmus Schneider, Erica Lasek-Nesselquist |
| EPI_ISL_1935631 | NYC Pandemic Response Lab | Wadsworth Center, New York State Department of Health | Kirsten St. George, Daryl M. Lamson, Alexis Russell, Matthew Shudt, Melissa A Leisner, Jonathan Plitnick, Catharine Prussing, Navjot Singh, John Kelly, Erasmus Schneider, Erica Lasek-Nesselquist |
| EPI_ISL_1935632, EPI_ISL_1935633 | TEMPUS LABS INC | Wadsworth Center, New York State Department of Health | Kirsten St. George, Daryl M. Lamson, Alexis Russell, Matthew Shudt, Melissa A Leisner, Jonathan Plitnick, Catharine Prussing, Navjot Singh, John Kelly, Erasmus Schneider, Erica Lasek-Nesselquist |
| EPI_ISL_1935634, EPI_ISL_1935635 | URMC LABS | Wadsworth Center, New York State Department of Health | Kirsten St. George, Daryl M. Lamson, Alexis Russell, Matthew Shudt, Melissa A Leisner, Jonathan Plitnick, Catharine Prussing, Navjot Singh, John Kelly, Erasmus Schneider, Erica Lasek-Nesselquist |
| EPI_ISL_1935636 | NYC Pandemic Response Lab | Wadsworth Center, New York State Department of Health | Kirsten St. George, Daryl M. Lamson, Alexis Russell, Matthew Shudt, Melissa A Leisner, Jonathan Plitnick, Catharine Prussing, Navjot Singh, John Kelly, Erasmus Schneider, Erica Lasek-Nesselquist |
| EPI_ISL_1935637 | NORTHWELL HEALTH LABORATORIES | Wadsworth Center, New York State Department of Health | Kirsten St. George, Daryl M. Lamson, Alexis Russell, Matthew Shudt, Melissa A Leisner, Jonathan Plitnick, Catharine Prussing, Navjot Singh, John Kelly, Erasmus Schneider, Erica Lasek-Nesselquist |
| EPI_ISL_1935638 | ALBANY MEDICAL CENTER | Wadsworth Center, New York State Department of Health | Kirsten St. George, Daryl M. Lamson, Alexis Russell, Matthew Shudt, Melissa A Leisner, Jonathan Plitnick, Catharine Prussing, Navjot Singh, John Kelly, Erasmus Schneider, Erica Lasek-Nesselquist |
| EPI_ISL_1935639, EPI_ISL_1935640, EPI_ISL_1935641, EPI_ISL_1935642 | SUNY UPSTATE MEDICAL UNIVERSITY | Wadsworth Center, New York State Department of Health | Kirsten St. George, Daryl M. Lamson, Alexis Russell, Matthew Shudt, Melissa A Leisner, Jonathan Plitnick, Catharine Prussing, Navjot Singh, John Kelly, Erasmus Schneider, Erica Lasek-Nesselquist |
| EPI_ISL_1935643 | MONTEFIORE MEDICAL CENTER LABORATORIES | Wadsworth Center, New York State Department of Health | Kirsten St. George, Daryl M. Lamson, Alexis Russell, Matthew Shudt, Melissa A Leisner, Jonathan Plitnick, Catharine Prussing, Navjot Singh, John Kelly, Erasmus Schneider, Erica Lasek-Nesselquist |
| EPI_ISL_1935644 | ALBANY MEDICAL CENTER | Wadsworth Center, New York State Department of Health | Kirsten St. George, Daryl M. Lamson, Alexis Russell, Matthew Shudt, Melissa A Leisner, Jonathan Plitnick, Catharine Prussing, Navjot Singh, John Kelly, Erasmus Schneider, Erica Lasek-Nesselquist |
| EPI_ISL_1935645 | URMC LABS | Wadsworth Center, New York State Department of Health | Kirsten St. George, Daryl M. Lamson, Alexis Russell, Matthew Shudt, Melissa A Leisner, Jonathan Plitnick, Catharine Prussing, Navjot Singh, John Kelly, Erasmus Schneider, Erica Lasek-Nesselquist |
| EPI_ISL_1935646, EPI_ISL_1935647 | SUNY UPSTATE MEDICAL UNIVERSITY | Wadsworth Center, New York State Department of Health | Kirsten St. George, Daryl M. Lamson, Alexis Russell, Matthew Shudt, Melissa A Leisner, Jonathan Plitnick, Catharine Prussing, Navjot Singh, John Kelly, Erasmus Schneider, Erica Lasek-Nesselquist |
| EPI_ISL_1935648 | URMC LABS | Wadsworth Center, New York State Department of Health | Kirsten St. George, Daryl M. Lamson, Alexis Russell, Matthew Shudt, Melissa A Leisner, Jonathan Plitnick, Catharine Prussing, Navjot Singh, John Kelly, Erasmus Schneider, Erica Lasek-Nesselquist |
| EPI_ISL_1935649, EPI_ISL_1935650, EPI_ISL_1935651 | SUNY UPSTATE MEDICAL UNIVERSITY | Wadsworth Center, New York State Department of Health | Kirsten St. George, Daryl M. Lamson, Alexis Russell, Matthew Shudt, Melissa A Leisner, Jonathan Plitnick, Catharine Prussing, Navjot Singh, John Kelly, Erasmus Schneider, Erica Lasek-Nesselquist |
| EPI_ISL_1935652 | URMC LABS | Wadsworth Center, New York State Department of Health | Kirsten St. George, Daryl M. Lamson, Alexis Russell, Matthew Shudt, Melissa A Leisner, Jonathan Plitnick, Catharine Prussing, Navjot Singh, John Kelly, Erasmus Schneider, Erica Lasek-Nesselquist |
| EPI_ISL_1937166, EPI_ISL_1937357 | Fulgent Genetics | Fulgent Genetics | Harry Gao, Mickey Li, John Gao, Joseph Fierro, Benafsh Sapra, Becky Tsai, Yan Meng, Doreen Ng, James Xie |
| EPI_ISL_1937596, EPI_ISL_1937598, EPI_ISL_1937600, EPI_ISL_1937601, EPI_ISL_1937603, EPI_ISL_1937605, EPI_ISL_1937606, EPI_ISL_1937608, EPI_ISL_1937610, EPI_ISL_1937611, EPI_ISL_1937613, EPI_ISL_1937615, EPI_ISL_1937616, EPI_ISL_1937618, EPI_ISL_1937620, EPI_ISL_1937622, EPI_ISL_1937623 | | | |
| see above | NYC Department of Health and Mental Hygiene | Centers for Disease Control and Prevention Division of Viral Diseases, Pathogen Discovery | Mili Sheth, Sarah Nobles, Jasmine Padilla, Mark Burroughs, Shoshona Le, Katie Dillon, Peter Cook, Clinton R. Paden, Dhwani Batra, Krista Queen, Kristen Knipe, Dakota Howard, Yvette Unoarumhi, Darlene Wagner, Matthew Schmerer, Ben L. Rambo-Martin, Kristine Lacek, Sam Shepard, Alison Laufer Halpin, Dave Wentworth, Vivien Dugan, Suxiang Tong, Justin Lee |
| EPI_ISL_1942142 | Aegis Sciences Corporation | Centers for Disease Control and Prevention Division of Viral Diseases, Pathogen Discovery | Dakota Howard, Dhwani Batra, Peter W. Cook, Kara Moser, Adrian Paskey, Jason Caravas, Benjamin Rambo-Martin, Shatavia Morrison, Christopher Gulvick, Scott Sammons, Yvette Unoarumhi, Darlene Wagner, Matthew Schmerer, Cyndi Clark, Patrick Campbell, Rob Case, Vikramsinha Ghorpade, Holly Houdeshell, Ola Kvalvaag, Dillon Nall, Ethan Sanders, Alec Vest, Shaun Westlund, Matthew Hardison, Clinton R. Paden, Duncan MacCannell |
| EPI_ISL_1942239 | Laboratory Corporation of America | Centers for Disease Control and Prevention Division of Viral Diseases, Pathogen Discovery | Dakota Howard, Dhwani Batra, Peter W. Cook, Kara Moser, Adrian Paskey, Jason Caravas, Benjamin Rambo-Martin, Shatavia Morrison, Christopher Gulvick, Scott Sammons, Yvette Unoarumhi, Darlene Wagner, Matthew Schmerer, Minoo Agarwal, Eyad Almasri, Debbie Boles, Ayla Burns, Nuthawin Charoensri, Oren Cohen, Susan Countryman, Mary Ann Cristobal, Bobbi Croy, Suzanne Dale, Hrushikesh Deshmukh, Amanda Douglas, Vincent Drouillon, Marcia Eisenberg, Howard Engler, Rama Ghatti, Prashant Gupta, Susan Hicks, Jake Humphrey, Lax Iyer, Manoj Jain, Mohan Kolli, Brian Krueger, Tim Kuphal, Stanley Letovsky, Michael Levandoski, Craig Lukasik, Jonathan Meltzer, Brian Norvell, Mindy Nye, Scott Parker, Christos Petropoulos, John Pruitt, Steven Ragan, Scott Ryan, Mike Sapeta, Jana Schroth, Suresh Babu Selvaraju, Goran Stevovic, Amanda Suchanek, Andrea Throop, Lyndon Tilson, Thomas Urban, Joe Voshell, Kimberly Wagner, Jonathan Williams, Mary Williamson, Qian Zeng, Tricia Zwiefelhofer, Clinton R. Paden, Duncan MacCannell |
| EPI_ISL_1963661, EPI_ISL_1963667 | Yale Clinical Virology Lab | Grubaugh Lab - Yale School of Public Health | Joseph Fauver, Mallery Breban, Isabel Ott, Tara Alpert, Mary Petrone, Anderson Brito, Chantal Vogels, Annie Watkins, Chaney Kalinich, Jessica Rothman, Marie L. Landry, Nathan Grubaugh |
| EPI_ISL_1965723, EPI_ISL_1965724, EPI_ISL_1965725, EPI_ISL_1965726, EPI_ISL_1965727, EPI_ISL_1965728, EPI_ISL_1965729, EPI_ISL_1965730, EPI_ISL_1965731, EPI_ISL_1965732, EPI_ISL_1965733, EPI_ISL_1965734, EPI_ISL_1965735, EPI_ISL_1965736, EPI_ISL_1965737, EPI_ISL_1965738, EPI_ISL_1965739, EPI_ISL_1965740, EPI_ISL_1965741, EPI_ISL_1965742, EPI_ISL_1965743, EPI_ISL_1965744, EPI_ISL_1965745, EPI_ISL_1965746, EPI_ISL_1965747, EPI_ISL_1965748, EPI_ISL_1965749, EPI_ISL_1965750, EPI_ISL_1965751, EPI_ISL_1965752, EPI_ISL_1965753, EPI_ISL_1965754, EPI_ISL_1965755, EPI_ISL_1965756, EPI_ISL_1965757, EPI_ISL_1965758, EPI_ISL_1965759, EPI_ISL_1965760, EPI_ISL_1965761, EPI_ISL_1965762, EPI_ISL_1965763, EPI_ISL_1965764, EPI_ISL_1965765, EPI_ISL_1965766, EPI_ISL_1965767, EPI_ISL_1965768, EPI_ISL_1965769, EPI_ISL_1965770, EPI_ISL_1965771, EPI_ISL_1965772, EPI_ISL_1965773, | | | |

| | | | |
|---|---|---|---|
| EPI_ISL_1965774, EPI_ISL_1965775, EPI_ISL_1965776, EPI_ISL_1965777, EPI_ISL_1965778, EPI_ISL_1965779, EPI_ISL_1965780, EPI_ISL_1965781, EPI_ISL_1965782, EPI_ISL_1965783, EPI_ISL_1965784, EPI_ISL_1965785, EPI_ISL_1965786, EPI_ISL_1965787, EPI_ISL_1965788, EPI_ISL_1965789, EPI_ISL_1965790, EPI_ISL_1965791, EPI_ISL_1965792, EPI_ISL_1965793, EPI_ISL_1965794, EPI_ISL_1965795, EPI_ISL_1965796, EPI_ISL_1965797, EPI_ISL_1965798, EPI_ISL_1965799, EPI_ISL_1965800, EPI_ISL_1965801, EPI_ISL_1965802, EPI_ISL_1965803, EPI_ISL_1965804, EPI_ISL_1965805, EPI_ISL_1965806, EPI_ISL_1965807, EPI_ISL_1965808, EPI_ISL_1965809, EPI_ISL_1965810, EPI_ISL_1965811, EPI_ISL_1965812, EPI_ISL_1965813, EPI_ISL_1965814, EPI_ISL_1965815, EPI_ISL_1965816, EPI_ISL_1965817, EPI_ISL_1965818, EPI_ISL_1965819, EPI_ISL_1965820, EPI_ISL_1965821, EPI_ISL_1965822, EPI_ISL_1965823, EPI_ISL_1965824, EPI_ISL_1965825, EPI_ISL_1965826, EPI_ISL_1965827, EPI_ISL_1965828, EPI_ISL_1965829, EPI_ISL_1965830, EPI_ISL_1965831, EPI_ISL_1965832, EPI_ISL_1965833, EPI_ISL_1965834, EPI_ISL_1965835, EPI_ISL_1965836, EPI_ISL_1965837, EPI_ISL_1965838, EPI_ISL_1965839, EPI_ISL_1965840, EPI_ISL_1965841, EPI_ISL_1965842, EPI_ISL_1965843, EPI_ISL_1965844, EPI_ISL_1965845, EPI_ISL_1965846, EPI_ISL_1965847, EPI_ISL_1965848, EPI_ISL_1965849, EPI_ISL_1965850, EPI_ISL_1965851, EPI_ISL_1965852, EPI_ISL_1965853, EPI_ISL_1965854, EPI_ISL_1965855, EPI_ISL_1965856, EPI_ISL_1965857, EPI_ISL_1965858, EPI_ISL_1965859, EPI_ISL_1965860, EPI_ISL_1965861, EPI_ISL_1965862, EPI_ISL_1965863, EPI_ISL_1965864, EPI_ISL_1965865, EPI_ISL_1965866, EPI_ISL_1965867, EPI_ISL_1965868, EPI_ISL_1965869, EPI_ISL_1965870, EPI_ISL_1965871, EPI_ISL_1965872, EPI_ISL_1965873, EPI_ISL_1965874, EPI_ISL_1965875, EPI_ISL_1965876, EPI_ISL_1965877, EPI_ISL_1965878, EPI_ISL_1965879, EPI_ISL_1965880, EPI_ISL_1965881, EPI_ISL_1965882, EPI_ISL_1965883, EPI_ISL_1965884, EPI_ISL_1965885, EPI_ISL_1965886, EPI_ISL_1965887, EPI_ISL_1965888, EPI_ISL_1965889, EPI_ISL_1965890, EPI_ISL_1965891, EPI_ISL_1965892, EPI_ISL_1965893, EPI_ISL_1965894, EPI_ISL_1965895, EPI_ISL_1965896, EPI_ISL_1965897, EPI_ISL_1965898, EPI_ISL_1965899, EPI_ISL_1965900, EPI_ISL_1965901, EPI_ISL_1965902, EPI_ISL_1965903, EPI_ISL_1965904, EPI_ISL_1965905, EPI_ISL_1965906, EPI_ISL_1965907, EPI_ISL_1965908, EPI_ISL_1965909, EPI_ISL_1965910, EPI_ISL_1965911, EPI_ISL_1965912, EPI_ISL_1965913, EPI_ISL_1965914, EPI_ISL_1965915, EPI_ISL_1965916, EPI_ISL_1965917, EPI_ISL_1965918, EPI_ISL_1965919, EPI_ISL_1965920, EPI_ISL_1965921, EPI_ISL_1965922, EPI_ISL_1965923, EPI_ISL_1965924, EPI_ISL_1965925, EPI_ISL_1965926, EPI_ISL_1965927, EPI_ISL_1965928, EPI_ISL_1965929, EPI_ISL_1965930, EPI_ISL_1965931, EPI_ISL_1965932, EPI_ISL_1965933, EPI_ISL_1965934, EPI_ISL_1965935, EPI_ISL_1965936, EPI_ISL_1965937, EPI_ISL_1965938, EPI_ISL_1965939, EPI_ISL_1965940, EPI_ISL_1965941, EPI_ISL_1965942, EPI_ISL_1965943, EPI_ISL_1965944, EPI_ISL_1965945, EPI_ISL_1965946, EPI_ISL_1965947, EPI_ISL_1965948, EPI_ISL_1965949, EPI_ISL_1965950, EPI_ISL_1965951, EPI_ISL_1965952, EPI_ISL_1965953, EPI_ISL_1965954, EPI_ISL_1965955, EPI_ISL_1965956, EPI_ISL_1965957, EPI_ISL_1965958, EPI_ISL_1965959, EPI_ISL_1965960, EPI_ISL_1965961, EPI_ISL_1965962, EPI_ISL_1965963, EPI_ISL_1965964, EPI_ISL_1965965, EPI_ISL_1965966, EPI_ISL_1965967, EPI_ISL_1965968, EPI_ISL_1965969, EPI_ISL_1965970, EPI_ISL_1965971, EPI_ISL_1965972, EPI_ISL_1965973, EPI_ISL_1965974, EPI_ISL_1965975, EPI_ISL_1965976, EPI_ISL_1965977, EPI_ISL_1965978, EPI_ISL_1965979, EPI_ISL_1965980, EPI_ISL_1965981, EPI_ISL_1965982, EPI_ISL_1965983, EPI_ISL_1965984, EPI_ISL_1965985, EPI_ISL_1965986, EPI_ISL_1965987, EPI_ISL_1965988, EPI_ISL_1965989, EPI_ISL_1965990, EPI_ISL_1965991, EPI_ISL_1965992, EPI_ISL_1965993, EPI_ISL_1965994, EPI_ISL_1965995, EPI_ISL_1965996, EPI_ISL_1965997, EPI_ISL_1965998, EPI_ISL_1965999, EPI_ISL_1966000, EPI_ISL_1966001, EPI_ISL_1966002, EPI_ISL_1966003, EPI_ISL_1966004, EPI_ISL_1966005, EPI_ISL_1966006, EPI_ISL_1966007, EPI_ISL_1966008, EPI_ISL_1966009, EPI_ISL_1966010, EPI_ISL_1966011, EPI_ISL_1966012, EPI_ISL_1966013, EPI_ISL_1966014, EPI_ISL_1966015, EPI_ISL_1966016, EPI_ISL_1966017, EPI_ISL_1966018, EPI_ISL_1966019, EPI_ISL_1966020, EPI_ISL_1966021, EPI_ISL_1966022, EPI_ISL_1966023, EPI_ISL_1966024, EPI_ISL_1966025, EPI_ISL_1966026, EPI_ISL_1966027, EPI_ISL_1966028, EPI_ISL_1966029, EPI_ISL_1966030, EPI_ISL_1966031, EPI_ISL_1966032, EPI_ISL_1966033, EPI_ISL_1966034, EPI_ISL_1966035, EPI_ISL_1966036, EPI_ISL_1966037, EPI_ISL_1966038, EPI_ISL_1966039, EPI_ISL_1966040, EPI_ISL_1966041, EPI_ISL_1966042, EPI_ISL_1966043, EPI_ISL_1966044, EPI_ISL_1966045, EPI_ISL_1966046, EPI_ISL_1966047, EPI_ISL_1966048, EPI_ISL_1966049, EPI_ISL_1966050, EPI_ISL_1966051, EPI_ISL_1966052, EPI_ISL_1966053, EPI_ISL_1966054, EPI_ISL_1966055, EPI_ISL_1966056, EPI_ISL_1966057, EPI_ISL_1966058 | | | |
| see above | Clinical Microbiology Laboratory, NewYork Presbyterian Hospital/Columbia University Irving Medical Center | Uhlemann Laboratory, Columbia University Irving Medical Center | Medini K. Annavajhala, Anne Kelley, Maya Tagliavia, Anne-Catrin Uhlemann |
| EPI_ISL_1967662, EPI_ISL_1967663, EPI_ISL_1967664, EPI_ISL_1967665, EPI_ISL_1967666, EPI_ISL_1967667, EPI_ISL_1967668, EPI_ISL_1967669, EPI_ISL_1967670, EPI_ISL_1967671, EPI_ISL_1967672, EPI_ISL_1967673, EPI_ISL_1967674, EPI_ISL_1967675, EPI_ISL_1967676, EPI_ISL_1967677, EPI_ISL_1967678, EPI_ISL_1967679, EPI_ISL_1967680, EPI_ISL_1967681, EPI_ISL_1967682, EPI_ISL_1967683, EPI_ISL_1967684 | | | |
| see above | Weill Cornell Medicine | New York Genome Center | Michael Zody, Andre Corvelo, Dayna M. Oschwald, Samantha Fennessey, Tom Maniatis, Melissa Cushing, Olivier Elemento, Margaret Elizabeth Ross, Chris Mason, Priya Velu, Hanna Rennert, Arryn Craney, Lars F Westblade |
| EPI_ISL_1967685, EPI_ISL_1967686, EPI_ISL_1967687, EPI_ISL_1967688 | Stony Brook Medicine | New York Genome Center | Michael Zody, Andre Corvelo, Dayna M. Oschwald, Samantha Fennessey, Tom Maniatis , Sarah A. Halbohn, Alaa B. Salaheldin, Sarah A. Malik, Josue M. Flores Acuna, Dimitri Gnatenko, Silvia G. Spitzer, Margaret M. McGovern, Eric D. Spitzer, Richard R. Kew, Kenneth R. Shroyer, Jiang Chen |
| EPI_ISL_1967689, EPI_ISL_1967690, EPI_ISL_1967691, EPI_ISL_1967692, EPI_ISL_1967693, EPI_ISL_1967694, EPI_ISL_1967695 | Stony Brook Medicine | New York Genome Center | Michael Zody, Andre Corvelo, Dayna M. Oschwald, Samantha Fennessey, Tom Maniatis , Sarah A. Halbohn, Alaa B. Salaheldin, Sarah A. Malik, Josue M. Flores Acuna, Dimitri Gnatenko, Silvia G. Spitzer, Margaret M. McGovern, Eric D. Spitzer, Richard R. Kew, Kenneth R. Shroyer, Jiang Chen, Susan V. Donelan |
| EPI_ISL_1968508, EPI_ISL_1968509, EPI_ISL_1968510 | Broad Institute Clinical Research Sequencing Platform | Infectious Disease Program, Broad Institute of Harvard and MIT | Siddle,K.J., Adams,G., Pearlman,L., Gladden-Young,A., Vicente,G., Blumenstiel,B., DeFelice,M., Lee,M., McGovern,S., Lagerborg,K., Rudy,M., DeRuff,K., Carter,A., Normandin,E., Bauer,M., Reilly,S., Tomkins-Tinch,C., Loreth,C., Chaluvadi,S., Meldrim,J., Granger,B., Lemieux,J.E., Birren,B.W., Sabeti,P.C., Larkin,K., Dodge,S., Lennon,N., Madoff,L., Brown,C., Gallagher,G., Smole,S., Park,D.J., Gabriel,S., and MacInnis,B.L. |
| EPI_ISL_1968561, EPI_ISL_1968562, EPI_ISL_1968563, EPI_ISL_1968564, EPI_ISL_1968565, EPI_ISL_1968566, EPI_ISL_1968567, EPI_ISL_1968568, EPI_ISL_1968569, EPI_ISL_1968570, EPI_ISL_1968571, EPI_ISL_1968572, EPI_ISL_1968573, EPI_ISL_1968574, EPI_ISL_1968575, EPI_ISL_1968576, EPI_ISL_1968577, EPI_ISL_1968578, EPI_ISL_1968579, EPI_ISL_1968580, EPI_ISL_1968581, EPI_ISL_1968582, EPI_ISL_1968583, EPI_ISL_1968584, EPI_ISL_1968585, EPI_ISL_1968586, EPI_ISL_1968587, EPI_ISL_1968588, EPI_ISL_1968589, EPI_ISL_1968590, EPI_ISL_1968591, EPI_ISL_1968592, EPI_ISL_1968593, EPI_ISL_1968594, EPI_ISL_1968595, EPI_ISL_1968596, EPI_ISL_1968597, EPI_ISL_1968598, EPI_ISL_1968599, EPI_ISL_1968600, EPI_ISL_1968601, EPI_ISL_1968602, EPI_ISL_1968603, EPI_ISL_1968604, EPI_ISL_1968605, EPI_ISL_1968606, EPI_ISL_1968607, EPI_ISL_1968608, EPI_ISL_1968609, EPI_ISL_1968610, EPI_ISL_1968611, EPI_ISL_1968612, EPI_ISL_1968613, EPI_ISL_1968614, EPI_ISL_1968615, EPI_ISL_1968616, EPI_ISL_1968617, EPI_ISL_1968618, EPI_ISL_1968619, EPI_ISL_1968620, EPI_ISL_1968621, EPI_ISL_1968622, EPI_ISL_1968623, EPI_ISL_1968624, EPI_ISL_1968625, EPI_ISL_1968626, EPI_ISL_1968627 | | | |
| see above | Weill Cornell Medicine | New York Genome Center | Michael Zody, Andre Corvelo, Dayna M. Oschwald, Samantha Fennessey, Tom Maniatis, Melissa Cushing, Olivier Elemento, Margaret Elizabeth Ross, Chris Mason, Priya Velu, Hanna Rennert, Arryn Craney, Lars F Westblade |
| EPI_ISL_1968717, EPI_ISL_1968718, EPI_ISL_1968719, EPI_ISL_1968722, EPI_ISL_1968723, EPI_ISL_1968724, EPI_ISL_1968725 | NYP-WCM | New York Genome Center | Michael Zody, Andre Corvelo, Dayna M. Oschwald, Samantha Fennessey, Tom Maniatis, Melissa Cushing, Olivier Elemento, Margaret Elizabeth Ross, Chris Mason, Priya Velu, Hanna Rennert, Arryn Craney, Lars F Westblade |
| EPI_ISL_1971088, EPI_ISL_1971089, EPI_ISL_1971090, EPI_ISL_1971091, EPI_ISL_1971092, EPI_ISL_1971093, EPI_ISL_1971094, EPI_ISL_1971095, EPI_ISL_1971096, EPI_ISL_1971097, EPI_ISL_1971098, EPI_ISL_1971099, EPI_ISL_1971100, EPI_ISL_1971101, EPI_ISL_1971102, EPI_ISL_1971103, EPI_ISL_1971104, EPI_ISL_1971105, EPI_ISL_1971106, EPI_ISL_1971107, EPI_ISL_1971108, EPI_ISL_1971109, EPI_ISL_1971110, EPI_ISL_1971111, EPI_ISL_1971112, EPI_ISL_1971113, EPI_ISL_1971114, EPI_ISL_1971115, EPI_ISL_1971116, EPI_ISL_1971117, EPI_ISL_1971118, EPI_ISL_1971119, EPI_ISL_1971120, EPI_ISL_1971121, EPI_ISL_1971122, EPI_ISL_1971123, EPI_ISL_1971124, EPI_ISL_1971125, EPI_ISL_1971126, EPI_ISL_1971127, EPI_ISL_1971128, EPI_ISL_1971129, EPI_ISL_1971130, EPI_ISL_1971131, EPI_ISL_1971132, EPI_ISL_1971133, EPI_ISL_1971134, EPI_ISL_1971135, EPI_ISL_1971136, EPI_ISL_1971137, EPI_ISL_1971138, EPI_ISL_1971139, EPI_ISL_1971140, EPI_ISL_1971141, EPI_ISL_1971142, EPI_ISL_1971143, EPI_ISL_1971144, EPI_ISL_1971145, EPI_ISL_1971146, EPI_ISL_1971147, EPI_ISL_1971148, EPI_ISL_1971149, EPI_ISL_1971150, EPI_ISL_1971151, EPI_ISL_1971152, EPI_ISL_1971153, EPI_ISL_1971154, EPI_ISL_1971155, EPI_ISL_1971156, EPI_ISL_1971157, EPI_ISL_1971158, EPI_ISL_1971159, EPI_ISL_1971160, EPI_ISL_1971161, EPI_ISL_1971162, EPI_ISL_1971163, EPI_ISL_1971164, EPI_ISL_1971165, EPI_ISL_1971166, EPI_ISL_1971167, EPI_ISL_1971168, EPI_ISL_1971169, EPI_ISL_1971170, EPI_ISL_1971171, EPI_ISL_1971172, EPI_ISL_1971173, EPI_ISL_1971174, EPI_ISL_1971175, EPI_ISL_1971176, EPI_ISL_1971177, EPI_ISL_1971178, EPI_ISL_1971179, EPI_ISL_1971180, EPI_ISL_1971181, EPI_ISL_1971182, EPI_ISL_1971183, EPI_ISL_1971184, EPI_ISL_1971185, EPI_ISL_1971186, EPI_ISL_1971187, EPI_ISL_1971188, EPI_ISL_1971189, EPI_ISL_1971190, EPI_ISL_1971191, EPI_ISL_1971192, EPI_ISL_1971193, EPI_ISL_1971194, EPI_ISL_1971195, EPI_ISL_1971196, EPI_ISL_1971197, EPI_ISL_1971198, EPI_ISL_1971199, EPI_ISL_1971200, EPI_ISL_1971201, EPI_ISL_1971202, EPI_ISL_1971203, EPI_ISL_1971204, EPI_ISL_1971205, EPI_ISL_1971206, EPI_ISL_1971207, EPI_ISL_1971208, EPI_ISL_1971209, EPI_ISL_1971210, EPI_ISL_1971211, EPI_ISL_1971212, EPI_ISL_1971213, EPI_ISL_1971214, EPI_ISL_1971215, EPI_ISL_1971216, EPI_ISL_1971217, EPI_ISL_1971218, EPI_ISL_1971219, EPI_ISL_1971220, EPI_ISL_1971221, EPI_ISL_1971222, EPI_ISL_1971223, EPI_ISL_1971224, EPI_ISL_1971225, EPI_ISL_1971226, EPI_ISL_1971227, EPI_ISL_1971228, EPI_ISL_1971229, EPI_ISL_1971230, EPI_ISL_1971231, EPI_ISL_1971369, EPI_ISL_1971370, EPI_ISL_1971371, EPI_ISL_1971372, EPI_ISL_1971373, EPI_ISL_1971374, EPI_ISL_1971375, EPI_ISL_1971376, EPI_ISL_1971377, EPI_ISL_1971378, EPI_ISL_1971379, EPI_ISL_1971380, EPI_ISL_1971381, EPI_ISL_1971382, EPI_ISL_1971383, EPI_ISL_1971384, EPI_ISL_1971385, EPI_ISL_1971386, EPI_ISL_1971387, EPI_ISL_1971388, EPI_ISL_1971389, EPI_ISL_1971390, EPI_ISL_1971391, EPI_ISL_1971392, EPI_ISL_1971393, EPI_ISL_1971394, EPI_ISL_1971395, EPI_ISL_1971396, EPI_ISL_1971397, EPI_ISL_1971398, EPI_ISL_1971399, EPI_ISL_1971400, EPI_ISL_1971401, EPI_ISL_1971402, EPI_ISL_1971403, EPI_ISL_1971404, EPI_ISL_1971405, EPI_ISL_1971406 | | | |
| see above | Weill Cornell Medicine | New York Genome Center | Michael Zody, Andre Corvelo, Dayna M. Oschwald, Samantha Fennessey, Tom Maniatis, Melissa Cushing, Olivier Elemento, Margaret Elizabeth Ross, Chris Mason, Priya Velu, Hanna Rennert, Arryn Craney, Lars F Westblade |
| EPI_ISL_1972014, EPI_ISL_1972015, EPI_ISL_1972016, EPI_ISL_1972017, EPI_ISL_1972018, EPI_ISL_1972019, EPI_ISL_1972020, EPI_ISL_1972021, EPI_ISL_1972022, EPI_ISL_1972023, EPI_ISL_1972024, EPI_ISL_1972025, EPI_ISL_1972026, EPI_ISL_1972027, EPI_ISL_1972028, EPI_ISL_1972029, EPI_ISL_1972030, EPI_ISL_1972031 | | | |
| see above | Broad Institute Clinical Research Sequencing Platform | Infectious Disease Program, Broad Institute of Harvard and MIT | Siddle,K.J., Adams,G., Pearlman,L., Gladden-Young,A., Vicente,G., Blumenstiel,B., DeFelice,M., Lee,M., McGovern,S., Lagerborg,K., Rudy,M., DeRuff,K., Carter,A., Normandin,E., Bauer,M., Reilly,S., Tomkins-Tinch,C., Loreth,C., Chaluvadi,S., Meldrim,J., Granger,B., Lemieux,J.E., Birren,B.W., Sabeti,P.C., Larkin,K., Dodge,S., Lennon,N., Madoff,L., Brown,C., Gallagher,G., Smole,S., Park,D.J., Gabriel,S., and MacInnis,B.L. |
| EPI_ISL_1989680, EPI_ISL_1989687, EPI_ISL_1989695, EPI_ISL_1989714, EPI_ISL_1989718, EPI_ISL_1989735, EPI_ISL_1989741, EPI_ISL_1989742, EPI_ISL_1989764, EPI_ISL_1989771, EPI_ISL_1989778, EPI_ISL_1989781, EPI_ISL_1989789, EPI_ISL_1989809, EPI_ISL_1989817, EPI_ISL_1989989, EPI_ISL_1990140, EPI_ISL_1990151, EPI_ISL_1990158, EPI_ISL_1990162, EPI_ISL_1990165, EPI_ISL_1990173, EPI_ISL_1990176, EPI_ISL_1990177, EPI_ISL_1990191, EPI_ISL_1990196, EPI_ISL_1990205, EPI_ISL_1990219, EPI_ISL_1990229, EPI_ISL_1990234, EPI_ISL_1990257, EPI_ISL_1990259, EPI_ISL_1990279, EPI_ISL_1990288, EPI_ISL_1990296, EPI_ISL_1990302, EPI_ISL_1990304, EPI_ISL_1990310, EPI_ISL_1990312, EPI_ISL_1990314, EPI_ISL_1990316, EPI_ISL_1990324, EPI_ISL_1990335, EPI_ISL_1990342, EPI_ISL_1990345, EPI_ISL_1990349, EPI_ISL_1990352, EPI_ISL_1990355, EPI_ISL_1990370, EPI_ISL_1990372, EPI_ISL_1990374, EPI_ISL_1990381, EPI_ISL_1990388, EPI_ISL_1990396, EPI_ISL_1990400, EPI_ISL_1990405 | | | |
| see above | Quest Diagnostics Incorporated | Centers for Disease Control and Prevention Division of Viral Diseases, Pathogen Discovery | Dakota Howard, Dhwani Batra, Peter W. Cook, Kara Moser, Adrian Paskey, Jason Caravas, Benjamin Rambo-Martin, Shatavia Morrison, Christopher Gulvick, Scott Sammons, Yvette Unoarumhi, Darlene Wagner, Matthew Schmerer, S. H. Rosenthal, A. Gerasimova, R. M. Kagan, B. Anderson, M. Hua, Y. Liu, L.E. Bernstein, K.E. Livingston, A. Perez, I. A. Shlyakhter, R. V. Rolando, R. Owen, P. Tanpaiboon, F. Lacbawan, Clinton R. Paden, Duncan MacCannell |
| EPI_ISL_1990722, EPI_ISL_1990941, EPI_ISL_1991481, EPI_ISL_1991836 | Helix/Illumina | Centers for Disease Control and Prevention Division of Viral Diseases, Pathogen Discovery | Dakota Howard, Dhwani Batra, Peter W. Cook, Kara Moser, Adrian Paskey, Jason Caravas, Benjamin Rambo-Martin, Shatavia Morrison, Christopher Gulvick, Scott Sammons, Yvette Unoarumhi, Darlene Wagner, Matthew Schmerer, Eileen de Feo, Jan Antico, Christine Tran, Matthew Tolentino, Shannon Wickline, Kim Gietzen, Brad Sickler, Jingtao Liu, Eric Allen, Phil Febbo, Nicole L. Washington, Simon White, Geraint Levan, Kelly Schiabor Barrett, Elizabeth Cirulli, Alexandre Bolze, Ary Ascencio, Charlotte Rivera-Garcia, Ryan Cho, Jason Nguyen, Sherry Wang, Jimmy Ramirez, Tyler Cassens, Efren Sandoval, Magnus Isaksson, William Lee, David Becker, Marc Laurent, James Lu, Clinton R. Paden, Duncan MacCannell |
| EPI_ISL_1994977, EPI_ISL_1994994, EPI_ISL_1995028, EPI_ISL_1995029, EPI_ISL_1995039, EPI_ISL_1995083, EPI_ISL_1995095, EPI_ISL_1995114, EPI_ISL_1995124, EPI_ISL_1995125, EPI_ISL_1995126, EPI_ISL_1995130, EPI_ISL_1995132, EPI_ISL_1995133, EPI_ISL_1995142, EPI_ISL_1995164, EPI_ISL_1995289, EPI_ISL_1995299, EPI_ISL_1995300, EPI_ISL_1995302, EPI_ISL_1995381, EPI_ISL_1995406, EPI_ISL_1995412, EPI_ISL_1995518, EPI_ISL_1995549, EPI_ISL_1995553, EPI_ISL_1995554, EPI_ISL_1995682, EPI_ISL_1995683, EPI_ISL_1995684, EPI_ISL_1995731, EPI_ISL_1995733, EPI_ISL_1995734, EPI_ISL_1995761, EPI_ISL_1995887, EPI_ISL_1995915, EPI_ISL_1995916, EPI_ISL_1995938, EPI_ISL_1995949, EPI_ISL_1995951, EPI_ISL_1995957, EPI_ISL_1996018, EPI_ISL_1996068, EPI_ISL_1996073, EPI_ISL_1996074, EPI_ISL_1996113, EPI_ISL_1996114, EPI_ISL_1996115, EPI_ISL_1996132, EPI_ISL_1996137, EPI_ISL_1996208, EPI_ISL_1996277, EPI_ISL_1996297, EPI_ISL_1996475, EPI_ISL_1996503, EPI_ISL_1996510, EPI_ISL_1996602, EPI_ISL_1996612, EPI_ISL_1996844, EPI_ISL_1996845, EPI_ISL_1996846, EPI_ISL_1996854, EPI_ISL_1996938, EPI_ISL_1996945, EPI_ISL_1996946, EPI_ISL_1996947, EPI_ISL_1996983, EPI_ISL_1996997, EPI_ISL_1997025, EPI_ISL_1997067, EPI_ISL_1997069, EPI_ISL_1997120, EPI_ISL_1997129, EPI_ISL_1997147, EPI_ISL_1997148, EPI_ISL_1997149, EPI_ISL_1997172, EPI_ISL_1997251, EPI_ISL_1997283, EPI_ISL_1997298, EPI_ISL_1997329, EPI_ISL_1997330, EPI_ISL_1997381, EPI_ISL_1997383, EPI_ISL_1997398, EPI_ISL_1997449, EPI_ISL_1997450, EPI_ISL_1997452, EPI_ISL_1997453, EPI_ISL_1997463, EPI_ISL_1997465, EPI_ISL_1997475, EPI_ISL_1997476, EPI_ISL_1997499, EPI_ISL_1997502, EPI_ISL_1997518, EPI_ISL_1997519, EPI_ISL_1997545, EPI_ISL_1997550, EPI_ISL_1997576, EPI_ISL_1997625, EPI_ISL_1997627, | | | |

EPI_ISL_1997681, EPI_ISL_1997683, EPI_ISL_1997736, EPI_ISL_1997737, EPI_ISL_1997742, EPI_ISL_1997752, EPI_ISL_1997753, EPI_ISL_1997755, EPI_ISL_1997827, EPI_ISL_1997939, EPI_ISL_1997940, EPI_ISL_1997998, EPI_ISL_1998084, EPI_ISL_1998189, EPI_ISL_1998251, EPI_ISL_1998286, EPI_ISL_1998288, EPI_ISL_1998305, EPI_ISL_1998338, EPI_ISL_1998606, EPI_ISL_1998608, EPI_ISL_1998680, EPI_ISL_1998688, EPI_ISL_1998715, EPI_ISL_1998733, EPI_ISL_1998736, EPI_ISL_1998770, EPI_ISL_1999110, EPI_ISL_1999124, EPI_ISL_1999232, EPI_ISL_1999246, EPI_ISL_1999250, EPI_ISL_1999262, EPI_ISL_1999353, EPI_ISL_1999476, EPI_ISL_1999477, EPI_ISL_1999572, EPI_ISL_1999684, EPI_ISL_1999715, EPI_ISL_1999807, EPI_ISL_1999808, EPI_ISL_1999816, EPI_ISL_1999868, EPI_ISL_1999870, EPI_ISL_1999892, EPI_ISL_1999941, EPI_ISL_1999974, EPI_ISL_1999982, EPI_ISL_2000182, EPI_ISL_2000186, EPI_ISL_2000187, EPI_ISL_2000190, EPI_ISL_2000260, EPI_ISL_2000278, EPI_ISL_2000286, EPI_ISL_2000386, EPI_ISL_2000389, EPI_ISL_2000390, EPI_ISL_2000391, EPI_ISL_2000438, EPI_ISL_2000507, EPI_ISL_2000542

| | | | |
|---|---|---|---|
| see above | Aegis Sciences Corporation | Centers for Disease Control and Prevention Division of Viral Diseases, Pathogen Discovery | Dakota Howard, Dhwani Batra, Peter W. Cook, Kara Moser, Adrian Paskey, Jason Caravas, Benjamin Rambo-Martin, Shatavia Morrison, Christopher Gulvick, Scott Sammons, Yvette Unoarumhi, Darlene Wagner, Matthew Schmerer, Cyndi Clark, Patrick Campbell, Rob Case, Vikramsinha Ghorpade, Holly Houdeshell, Ola Kvalvaag, Dillon Nall, Ethan Sanders, Alec Vest, Shaun Westlund, Matthew Hardison, Clinton R. Paden, Duncan MacCannell |
