## Supplemental acknowledgement table #5 for "Genomic Surveillance of SARS-CoV-2 in Erie County, New York"

We gratefully acknowledge the following Authors from the Originating laboratories responsible for obtaining the specimens, as well as the Submitting laboratories where the genome data were generated and shared via GISAID, on which this research is based.

All Submitters of data may be contacted directly via www.gisaid.org

Authors are sorted alphabetically.

| Accession ID | Originating Laboratory | Submitting Laboratory | Authors |
|---|---|---|---|
| EPI_ISL_2006745 | Moderna Inc. | Moderna Inc. | Yamuna Paila; Groves Dixon; Rolando Pajon |
| EPI_ISL_2009117, EPI_ISL_2009118, EPI_ISL_2009119, EPI_ISL_2009120, EPI_ISL_2009121, EPI_ISL_2009122, EPI_ISL_2009123, EPI_ISL_2009124, EPI_ISL_2009125, EPI_ISL_2009126, EPI_ISL_2009127, EPI_ISL_2009128, EPI_ISL_2009129, EPI_ISL_2009130, EPI_ISL_2009131, EPI_ISL_2009132, EPI_ISL_2009133, EPI_ISL_2009134, EPI_ISL_2009135, EPI_ISL_2009136, EPI_ISL_2009137, EPI_ISL_2009138, EPI_ISL_2009139, EPI_ISL_2009140, EPI_ISL_2009141, EPI_ISL_2009142, EPI_ISL_2009143, EPI_ISL_2009144, EPI_ISL_2009145, EPI_ISL_2009146, EPI_ISL_2009147, EPI_ISL_2009148, EPI_ISL_2009149, EPI_ISL_2009150, EPI_ISL_2009151, EPI_ISL_2009152, EPI_ISL_2009153, EPI_ISL_2009154, EPI_ISL_2009155, EPI_ISL_2009156, EPI_ISL_2009157, EPI_ISL_2009158, EPI_ISL_2009159, EPI_ISL_2009160, EPI_ISL_2009161, EPI_ISL_2009162, EPI_ISL_2009163, EPI_ISL_2009164, EPI_ISL_2009165, EPI_ISL_2009166, EPI_ISL_2009167, EPI_ISL_2009168, EPI_ISL_2009169, EPI_ISL_2009170, EPI_ISL_2009171, EPI_ISL_2009172, EPI_ISL_2009173, EPI_ISL_2009174, EPI_ISL_2009175, EPI_ISL_2009176, EPI_ISL_2009177, EPI_ISL_2009178, EPI_ISL_2009179, EPI_ISL_2009180, EPI_ISL_2009181, EPI_ISL_2009182, EPI_ISL_2009183, EPI_ISL_2009184, EPI_ISL_2009185 | | | |
| see above | MSK Microbiology Lab | MSK Microbiology Lab | Esther Babady, Tracy McMillen, Krupa Jani |
| EPI_ISL_2009437 | Moderna Inc. | Moderna Inc. | Yamuna Paila; Groves Dixon; Rolando Pajon |
| EPI_ISL_2009862, EPI_ISL_2010081, EPI_ISL_2010284, EPI_ISL_2010619, EPI_ISL_2010810, EPI_ISL_2010860 | Helix/Illumina | Centers for Disease Control and Prevention Division of Viral Diseases, Pathogen Discovery | Dakota Howard, Dhwani Batra, Peter W. Cook, Kara Moser, Adrian Paskey, Jason Caravas, Benjamin Rambo-Martin, Shatavia Morrison, Christopher Gulvick, Scott Sammons, Yvette Unoarumhi, Darlene Wagner, Matthew Schmerer, Eileen de Feo, Jan Antico, Christine Tran, Matthew Tolentino, Shannon Wickline, Kim Gietzen, Brad Sickler, Jingtao Liu, Eric Allen, Phil Febbo, Nicole L. Washington, Simon White, Geraint Levan, Kelly Schiabor Barrett, Elizabeth Cirulli, Alexandre Bolze, Ary Ascencio, Charlotte Rivera-Garcia, Ryan Cho, Jason Nguyen, Sherry Wang, Jimmy Ramirez, Tyler Cassens, Efren Sandoval, Magnus Isaksson, William Lee, David Becker, Marc Laurent, James Lu, Clinton R. Paden, Duncan MacCannell |
| EPI_ISL_2010902, EPI_ISL_2010904, EPI_ISL_2010972, EPI_ISL_2010977, EPI_ISL_2010991 | Quest Diagnostics Incorporated | Centers for Disease Control and Prevention Division of Viral Diseases, Pathogen Discovery | Dakota Howard, Dhwani Batra, Peter W. Cook, Kara Moser, Adrian Paskey, Jason Caravas, Benjamin Rambo-Martin, Shatavia Morrison, Christopher Gulvick, Scott Sammons, Yvette Unoarumhi, Darlene Wagner, Matthew Schmerer, S. H. Rosenthal, A. Gerasimova, R. M. Kagan, B. Anderson, M. Hua, Y. Liu, L.E. Bernstein, K.E. Livingston, A. Perez, I. A. Shlyakhter, R. V. Rolando, R. Owen, P. Tanpaiboon, F. Lacbawan, Clinton R. Paden, Duncan MacCannell |
| EPI_ISL_2011717 | Moderna Inc. | Moderna Inc. | Yamuna Paila; Groves Dixon; Rolando Pajon |
| EPI_ISL_2013321, EPI_ISL_2014189 | Infinity Biologix | Centers for Disease Control and Prevention Division of Viral Diseases, Pathogen Discovery | Dakota Howard, Dhwani Batra, Peter W. Cook, Kara Moser, Adrian Paskey, Jason Caravas, Benjamin Rambo-Martin, Shatavia Morrison, Christopher Gulvick, Scott Sammons, Yvette Unoarumhi, Darlene Wagner, Matthew Schmerer, Christian Bixby, Yihe Wang, Jonathan Schultz, Chirayu Goswami, Russ Hager, Robin Grimwood, Clinton R. Paden, Duncan MacCannell |
| EPI_ISL_2018243, EPI_ISL_2018244, EPI_ISL_2018245, EPI_ISL_2018246 | NYSDOH Wadsworth Center, Virology Lab | Centers for Disease Control and Prevention Division of Viral Diseases, Pathogen Discovery | Mili Sheth, Sarah Nobles, Jasmine Padilla, Mark Burroughs, Shoshona Le, Katie Dillon, Peter Cook, Clinton R. Paden, Dhwani Batra, Krista Queen, Kristen Knipe, Dakota Howard, Yvette Unoarumhi, Darlene Wagner, Matthew Schmerer, Ben L. Rambo-Martin, Kristine Lacek, Sam Shepard, Alison Laufer Halpin, Dave Wentworth, Vivien Dugan, Suxiang Tong, Justin Lee |
| EPI_ISL_2018418, EPI_ISL_2018419, EPI_ISL_2018420, EPI_ISL_2018421, EPI_ISL_2018422, EPI_ISL_2018423, EPI_ISL_2018424, EPI_ISL_2018425, EPI_ISL_2018426, EPI_ISL_2018427, EPI_ISL_2018428, EPI_ISL_2018429, EPI_ISL_2018430, EPI_ISL_2018431, EPI_ISL_2018432, EPI_ISL_2018433 | | | |
| see above | NYC Department of Health and Mental Hygiene | Centers for Disease Control and Prevention Division of Viral Diseases, Pathogen Discovery | Mili Sheth, Sarah Nobles, Jasmine Padilla, Mark Burroughs, Shoshona Le, Katie Dillon, Peter Cook, Clinton R. Paden, Dhwani Batra, Krista Queen, Kristen Knipe, Dakota Howard, Yvette Unoarumhi, Darlene Wagner, Matthew Schmerer, Ben L. Rambo-Martin, Kristine Lacek, Sam Shepard, Alison Laufer Halpin, Dave Wentworth, Vivien Dugan, Suxiang Tong, Justin Lee |
| EPI_ISL_2019648, EPI_ISL_2019655, EPI_ISL_2019672, EPI_ISL_2019686, EPI_ISL_2019701, EPI_ISL_2019790, EPI_ISL_2019854, EPI_ISL_2019933, EPI_ISL_2019956, EPI_ISL_2019961, EPI_ISL_2019966, EPI_ISL_2019976, EPI_ISL_2019994, EPI_ISL_2020004, EPI_ISL_2020005 | | | |
| see above | Moderna Inc. | Moderna Inc. | Yamuna Paila; Groves Dixon; Rolando Pajon |
| EPI_ISL_2020795, EPI_ISL_2020840 | Yale Clinical Virology Lab | Yale Center for Genomic Analysis | Shrikant Mane, Kaya Bilguvar, Curt Scharfe, Irina Tikhonova, Brooke Sullivan |
| EPI_ISL_2031642, EPI_ISL_2031643, EPI_ISL_2031647 | Columbia University | Pathogen Discovery, Respiratory Viruses Branch, Division of Viral Diseases, Centers for Disease Control and Prevention | Ying Tao, Yan Li, Jing Zhang, Anna Kelleher, Brian Lynch, Krista Queen, Anna Uehara, Peter Cook, Han Jia Justin Ng, Rachel Marine, Clinton R. Paden, Dhwani Batra, Haibin Wang, Tara Coalter, Jasmine Padilla, Morgan Davis, Mili Sheth, Sarah Nobles, Mark Burroughs, Justin Lee, Adam Retchless, Suxiang Tong |
| EPI_ISL_2031764, EPI_ISL_2031765, EPI_ISL_2031766, EPI_ISL_2031767, EPI_ISL_2031768, EPI_ISL_2031769, EPI_ISL_2031770, EPI_ISL_2031771 | Weill Cornell Medicine | New York Genome Center | Michael Zody, Andre Corvelo, Dayna M. Oschwald, Samantha Fennessey, Tom Maniatis, Melissa Cushing, Olivier Elemento, Margaret Elizabeth Ross, Chris Mason, Priya Velu, Hanna Rennert, Arryn Craney, Lars F Westblade |
| EPI_ISL_2034749 | Quest Diagnostics Incorporated | Centers for Disease Control and Prevention Division of Viral Diseases, Pathogen Discovery | Dakota Howard, Dhwani Batra, Peter W. Cook, Kara Moser, Adrian Paskey, Jason Caravas, Benjamin Rambo-Martin, Shatavia Morrison, Christopher Gulvick, Scott Sammons, Yvette Unoarumhi, Darlene Wagner, Matthew Schmerer, S. H. Rosenthal, A. Gerasimova, R. M. Kagan, B. Anderson, M. Hua, Y. Liu, L.E. Bernstein, K.E. Livingston, A. Perez, I. A. Shlyakhter, R. V. Rolando, R. Owen, P. Tanpaiboon, F. Lacbawan, Clinton R. Paden, Duncan MacCannell |
| EPI_ISL_2034813, EPI_ISL_2034818, EPI_ISL_2034886, EPI_ISL_2034934 | Yale Clinical Virology Lab | Yale Center for Genomic Analysis | Shrikant Mane, Kaya Bilguvar, Curt Scharfe, Irina Tikhonova, Brooke Sullivan |
| EPI_ISL_2035107, EPI_ISL_2035130, EPI_ISL_2035243, EPI_ISL_2035253, EPI_ISL_2035263 | Aegis Sciences Corporation | Centers for Disease Control and Prevention Division of Viral Diseases, Pathogen Discovery | Dakota Howard, Dhwani Batra, Peter W. Cook, Kara Moser, Adrian Paskey, Jason Caravas, Benjamin Rambo-Martin, Shatavia Morrison, Christopher Gulvick, Scott Sammons, Yvette Unoarumhi, Darlene Wagner, Matthew Schmerer, Cyndi Clark, Patrick Campbell, Rob Case, Vikramsinha Ghorpade, Holly Houdeshell, Ola Kvalvaag, Dillon Nall, Ethan Sanders, Alec Vest, Shaun Westlund, Matthew Hardison, Clinton R. Paden, Duncan MacCannell |
| EPI_ISL_2036759 | CT Department of Public Health | CT Department of Public Health | Claire_Pearson,Tu_N_Nguyen |
| EPI_ISL_2036964, EPI_ISL_2036965, EPI_ISL_2036966, EPI_ISL_2036967, EPI_ISL_2036968, EPI_ISL_2036969, EPI_ISL_2036970, EPI_ISL_2036971, EPI_ISL_2036972, EPI_ISL_2036973, EPI_ISL_2036974, EPI_ISL_2036975, EPI_ISL_2036976, EPI_ISL_2036977, EPI_ISL_2036978, EPI_ISL_2036979, EPI_ISL_2036980, EPI_ISL_2036981, EPI_ISL_2036982, EPI_ISL_2036983, EPI_ISL_2036984, EPI_ISL_2036985, EPI_ISL_2036986, EPI_ISL_2036987, EPI_ISL_2036988, EPI_ISL_2036989, EPI_ISL_2036990, EPI_ISL_2036991, EPI_ISL_2036992, EPI_ISL_2036993, EPI_ISL_2036994, EPI_ISL_2036995, EPI_ISL_2036996, EPI_ISL_2036997, EPI_ISL_2036998, EPI_ISL_2036999, EPI_ISL_2037000, EPI_ISL_2037001, EPI_ISL_2037002, EPI_ISL_2037003, EPI_ISL_2037004, EPI_ISL_2037005, EPI_ISL_2037006, EPI_ISL_2037007, EPI_ISL_2037008, EPI_ISL_2037009, EPI_ISL_2037010, EPI_ISL_2037011, EPI_ISL_2037012, EPI_ISL_2037013, EPI_ISL_2037014, EPI_ISL_2037015, EPI_ISL_2037016, EPI_ISL_2037017, EPI_ISL_2037018, EPI_ISL_2037019, EPI_ISL_2037020, EPI_ISL_2037021, EPI_ISL_2037022, EPI_ISL_2037023, EPI_ISL_2037024, EPI_ISL_2037025, EPI_ISL_2037026, EPI_ISL_2037027, EPI_ISL_2037028, EPI_ISL_2037029, EPI_ISL_2037030, EPI_ISL_2037031, EPI_ISL_2037032, EPI_ISL_2037033, EPI_ISL_2037034, EPI_ISL_2037035, EPI_ISL_2037036, EPI_ISL_2037037, EPI_ISL_2037038, EPI_ISL_2037039, EPI_ISL_2037040, EPI_ISL_2037041, EPI_ISL_2037042, EPI_ISL_2037043, EPI_ISL_2037044, EPI_ISL_2037045, EPI_ISL_2037046, EPI_ISL_2037047, EPI_ISL_2037048, EPI_ISL_2037049, EPI_ISL_2037050, EPI_ISL_2037051, EPI_ISL_2037052, EPI_ISL_2037053, EPI_ISL_2037054, EPI_ISL_2037055, EPI_ISL_2037056, EPI_ISL_2037057, EPI_ISL_2037058, EPI_ISL_2037059, EPI_ISL_2037060, EPI_ISL_2037061, EPI_ISL_2037062, EPI_ISL_2037063, EPI_ISL_2037064, EPI_ISL_2037065, EPI_ISL_2037066, EPI_ISL_2037067, EPI_ISL_2037068, EPI_ISL_2037069, EPI_ISL_2037070, EPI_ISL_2037071, EPI_ISL_2037072, EPI_ISL_2037073, EPI_ISL_2037074, EPI_ISL_2037075, EPI_ISL_2037076, EPI_ISL_2037077, EPI_ISL_2037078, EPI_ISL_2037079, EPI_ISL_2037080, EPI_ISL_2037081, EPI_ISL_2037082, EPI_ISL_2037083, EPI_ISL_2037084, EPI_ISL_2037085, EPI_ISL_2037086, EPI_ISL_2037087, EPI_ISL_2037088, EPI_ISL_2037089, EPI_ISL_2037090, EPI_ISL_2037091, EPI_ISL_2037092, EPI_ISL_2037093, EPI_ISL_2037094, EPI_ISL_2037095, EPI_ISL_2037096, EPI_ISL_2037097, EPI_ISL_2037098, EPI_ISL_2037099, EPI_ISL_2037100, EPI_ISL_2037101, EPI_ISL_2037102, EPI_ISL_2037103, EPI_ISL_2037104, EPI_ISL_2037105, EPI_ISL_2037106, EPI_ISL_2037107, EPI_ISL_2037108, EPI_ISL_2037109, EPI_ISL_2037110, EPI_ISL_2037111, EPI_ISL_2037112, EPI_ISL_2037113, EPI_ISL_2037114, EPI_ISL_2037115, EPI_ISL_2037116, EPI_ISL_2037117, EPI_ISL_2037118, EPI_ISL_2037119, EPI_ISL_2037120, EPI_ISL_2037121, EPI_ISL_2037122, EPI_ISL_2037123, EPI_ISL_2037124, EPI_ISL_2037125, EPI_ISL_2037126, EPI_ISL_2037127, EPI_ISL_2037128, EPI_ISL_2037129, EPI_ISL_2037130, EPI_ISL_2037131, EPI_ISL_2037132, EPI_ISL_2037133, EPI_ISL_2037134, EPI_ISL_2037135, EPI_ISL_2037136, EPI_ISL_2037137, EPI_ISL_2037138, EPI_ISL_2037139, EPI_ISL_2037140, EPI_ISL_2037141, EPI_ISL_2037142, EPI_ISL_2037143, EPI_ISL_2037144, EPI_ISL_2037145, EPI_ISL_2037146, EPI_ISL_2037147, EPI_ISL_2037148, EPI_ISL_2037149, EPI_ISL_2037150, EPI_ISL_2037151, EPI_ISL_2037152, EPI_ISL_2037153, EPI_ISL_2037154, EPI_ISL_2037155, EPI_ISL_2037156, EPI_ISL_2037157, EPI_ISL_2037158, EPI_ISL_2037159, EPI_ISL_2037160, EPI_ISL_2037161, EPI_ISL_2037162, EPI_ISL_2037163, EPI_ISL_2037164, EPI_ISL_2037165, EPI_ISL_2037166, EPI_ISL_2037167, EPI_ISL_2037168, EPI_ISL_2037169, EPI_ISL_2037170, EPI_ISL_2037171, EPI_ISL_2037172, EPI_ISL_2037173, EPI_ISL_2037174, EPI_ISL_2037175, EPI_ISL_2037176, EPI_ISL_2037177, EPI_ISL_2037178, EPI_ISL_2037179, EPI_ISL_2037180, EPI_ISL_2037181, EPI_ISL_2037182, EPI_ISL_2037183, EPI_ISL_2037184, EPI_ISL_2037185, EPI_ISL_2037186, EPI_ISL_2037187, EPI_ISL_2037188, EPI_ISL_2037189, EPI_ISL_2037190, EPI_ISL_2037191, EPI_ISL_2037192, EPI_ISL_2037193, EPI_ISL_2037194, EPI_ISL_2037195, EPI_ISL_2037196, EPI_ISL_2037197, EPI_ISL_2037198, EPI_ISL_2037199, EPI_ISL_2037200, EPI_ISL_2037201, EPI_ISL_2037202, EPI_ISL_2037203, EPI_ISL_2037204, EPI_ISL_2037205, EPI_ISL_2037206, EPI_ISL_2037207, EPI_ISL_2037208, EPI_ISL_2037209, EPI_ISL_2037210, EPI_ISL_2037211, EPI_ISL_2037212, EPI_ISL_2037213, EPI_ISL_2037214, EPI_ISL_2037215, EPI_ISL_2037216, EPI_ISL_2037217, EPI_ISL_2037218, EPI_ISL_2037219, EPI_ISL_2037220, EPI_ISL_2037221, EPI_ISL_2037222, EPI_ISL_2037223, EPI_ISL_2037224, EPI_ISL_2037225, EPI_ISL_2037226, EPI_ISL_2037227, EPI_ISL_2037228, EPI_ISL_2037229, EPI_ISL_2037230, EPI_ISL_2037231, EPI_ISL_2037232, EPI_ISL_2037233, EPI_ISL_2037234, EPI_ISL_2037235, EPI_ISL_2037236, EPI_ISL_2037237, EPI_ISL_2037238, EPI_ISL_2037239, EPI_ISL_2037240, EPI_ISL_2037241, EPI_ISL_2037242, EPI_ISL_2037243, EPI_ISL_2037244, EPI_ISL_2037245, EPI_ISL_2037246, EPI_ISL_2037247, EPI_ISL_2037248, EPI_ISL_2037249, EPI_ISL_2037250, EPI_ISL_2037251, EPI_ISL_2037252, | | | |

| | | | |
|---|---|---|---|
| EPI_ISL_2037253, EPI_ISL_2037254, EPI_ISL_2037255, EPI_ISL_2037256, EPI_ISL_2037257, EPI_ISL_2037258, EPI_ISL_2037259, EPI_ISL_2037260, EPI_ISL_2037261, EPI_ISL_2037262, EPI_ISL_2037263, EPI_ISL_2037264, EPI_ISL_2037265, EPI_ISL_2037266, EPI_ISL_2037267, EPI_ISL_2037268, EPI_ISL_2037269, EPI_ISL_2037270, EPI_ISL_2037271, EPI_ISL_2037272, EPI_ISL_2037273, EPI_ISL_2037274, EPI_ISL_2037275, EPI_ISL_2037276, EPI_ISL_2037277, EPI_ISL_2037278, EPI_ISL_2037279, EPI_ISL_2037280, EPI_ISL_2037281, EPI_ISL_2037282, EPI_ISL_2037283, EPI_ISL_2037284, EPI_ISL_2037285, EPI_ISL_2037286, EPI_ISL_2037287, EPI_ISL_2037288, EPI_ISL_2037289, EPI_ISL_2037290, EPI_ISL_2037291, EPI_ISL_2037292, EPI_ISL_2037293, EPI_ISL_2037294, EPI_ISL_2037295, EPI_ISL_2037296, EPI_ISL_2037297, EPI_ISL_2037298, EPI_ISL_2037299, EPI_ISL_2037300, EPI_ISL_2037301, EPI_ISL_2037302, EPI_ISL_2037303, EPI_ISL_2037304, EPI_ISL_2037305, EPI_ISL_2037306, EPI_ISL_2037307, EPI_ISL_2037308, EPI_ISL_2037309, EPI_ISL_2037310, EPI_ISL_2037311, EPI_ISL_2037312, EPI_ISL_2037313, EPI_ISL_2037314, EPI_ISL_2037315, EPI_ISL_2037316, EPI_ISL_2037317, EPI_ISL_2037318, EPI_ISL_2037319, EPI_ISL_2037320, EPI_ISL_2037321, EPI_ISL_2037322, EPI_ISL_2037323, EPI_ISL_2037324, EPI_ISL_2037325, EPI_ISL_2037326, EPI_ISL_2037327, EPI_ISL_2037328, EPI_ISL_2037329, EPI_ISL_2037330, EPI_ISL_2037331, EPI_ISL_2037332, EPI_ISL_2037333, EPI_ISL_2037334, EPI_ISL_2037335, EPI_ISL_2037336, EPI_ISL_2037337, EPI_ISL_2037338, EPI_ISL_2037339, EPI_ISL_2037340, EPI_ISL_2037341, EPI_ISL_2037342, EPI_ISL_2037343, EPI_ISL_2037344, EPI_ISL_2037345, EPI_ISL_2037346, EPI_ISL_2037347, EPI_ISL_2037348, EPI_ISL_2037349, EPI_ISL_2037350, EPI_ISL_2037351, EPI_ISL_2037352, EPI_ISL_2037353, EPI_ISL_2037354, EPI_ISL_2037355, EPI_ISL_2037356, EPI_ISL_2037357, EPI_ISL_2037358, EPI_ISL_2037359, EPI_ISL_2037360, EPI_ISL_2037361, EPI_ISL_2037362, EPI_ISL_2037363, EPI_ISL_2037364, EPI_ISL_2037365, EPI_ISL_2037366, EPI_ISL_2037367, EPI_ISL_2037368, EPI_ISL_2037369, EPI_ISL_2037370, EPI_ISL_2037371, EPI_ISL_2037372, EPI_ISL_2037373, EPI_ISL_2037374, EPI_ISL_2037375, EPI_ISL_2037376, EPI_ISL_2037377, EPI_ISL_2037378, EPI_ISL_2037379, EPI_ISL_2037380, EPI_ISL_2037381, EPI_ISL_2037382, EPI_ISL_2037383, EPI_ISL_2037384, EPI_ISL_2037385, EPI_ISL_2037386, EPI_ISL_2037387, EPI_ISL_2037388 | | | |
| see above | Pandemic Response Lab - NYC | Pandemic Response Lab, R&D | Henry Lee, Michael Hammerling, Melissa Hopkins, Cybill del Castillo, Shinyoung Clair Kang, William Ward, Pradeep Bugga, Sol Rey, Dylan Law, Katharine Nelson, Haiping Hao, Jon Laurent |
| EPI_ISL_2038970, EPI_ISL_2038975, EPI_ISL_2038976, EPI_ISL_2038977, EPI_ISL_2039013, EPI_ISL_2039137, EPI_ISL_2039138, EPI_ISL_2039255, EPI_ISL_2039259, EPI_ISL_2039263, EPI_ISL_2039276, EPI_ISL_2039278, EPI_ISL_2039279, EPI_ISL_2039280, EPI_ISL_2039281, EPI_ISL_2039283, EPI_ISL_2039380, EPI_ISL_2039403, EPI_ISL_2039447, EPI_ISL_2039454, EPI_ISL_2039456, EPI_ISL_2039547, EPI_ISL_2039631, EPI_ISL_2039635, EPI_ISL_2039746, EPI_ISL_2039808, EPI_ISL_2039867, EPI_ISL_2039872, EPI_ISL_2039979, EPI_ISL_2039981, EPI_ISL_2039999, EPI_ISL_2040092, EPI_ISL_2040240, EPI_ISL_2040241, EPI_ISL_2040242, EPI_ISL_2040249, EPI_ISL_2040256, EPI_ISL_2040262, EPI_ISL_2040286, EPI_ISL_2040299, EPI_ISL_2040310, EPI_ISL_2040316, EPI_ISL_2040466, EPI_ISL_2040467, EPI_ISL_2040505, EPI_ISL_2040517, EPI_ISL_2040546, EPI_ISL_2040567, EPI_ISL_2040593, EPI_ISL_2040626, EPI_ISL_2040687, EPI_ISL_2040795, EPI_ISL_2040807, EPI_ISL_2040810, EPI_ISL_2040823, EPI_ISL_2040847, EPI_ISL_2040869, EPI_ISL_2040870, EPI_ISL_2040891, EPI_ISL_2040935, EPI_ISL_2040940, EPI_ISL_2040997, EPI_ISL_2041057, EPI_ISL_2041060, EPI_ISL_2041163, EPI_ISL_2041167, EPI_ISL_2041170, EPI_ISL_2041187, EPI_ISL_2041204, EPI_ISL_2041335, EPI_ISL_2041354, EPI_ISL_2041356, EPI_ISL_2041364, EPI_ISL_2041383, EPI_ISL_2041488, EPI_ISL_2041583, EPI_ISL_2041700, EPI_ISL_2041703, EPI_ISL_2041707, EPI_ISL_2041708, EPI_ISL_2041709, EPI_ISL_2041736, EPI_ISL_2041742, EPI_ISL_2041747, EPI_ISL_2041749, EPI_ISL_2041755, EPI_ISL_2041787, EPI_ISL_2041956, EPI_ISL_2041958, EPI_ISL_2042096, EPI_ISL_2042115, EPI_ISL_2042123, EPI_ISL_2042164, EPI_ISL_2042237, EPI_ISL_2042325, EPI_ISL_2042337, EPI_ISL_2042347, EPI_ISL_2042508, EPI_ISL_2042510, EPI_ISL_2042533, EPI_ISL_2042540, EPI_ISL_2042543, EPI_ISL_2042606, EPI_ISL_2042611, EPI_ISL_2042621, EPI_ISL_2042622, EPI_ISL_2042629, EPI_ISL_2042630, EPI_ISL_2042678, EPI_ISL_2042679, EPI_ISL_2042693, EPI_ISL_2042697, EPI_ISL_2042698, EPI_ISL_2042706, EPI_ISL_2042744, EPI_ISL_2042745, EPI_ISL_2042767, EPI_ISL_2042769, EPI_ISL_2042775, EPI_ISL_2043139, EPI_ISL_2043151, EPI_ISL_2043166, EPI_ISL_2043188, EPI_ISL_2043207, EPI_ISL_2043213, EPI_ISL_2043216, EPI_ISL_2043230, EPI_ISL_2043239, EPI_ISL_2043255, EPI_ISL_2043327, EPI_ISL_2043367, EPI_ISL_2043425 | | | |
| see above | Aegis Sciences Corporation | Centers for Disease Control and Prevention Division of Viral Diseases, Pathogen Discovery | Dakota Howard, Dhwani Batra, Peter W. Cook, Kara Moser, Adrian Paskey, Jason Caravas, Benjamin Rambo-Martin, Shatavia Morrison, Christopher Gulvick, Scott Sammons, Yvette Unoarumhi, Darlene Wagner, Matthew Schmerer, Cyndi Clark, Patrick Campbell, Rob Case, Vikramsinha Ghorpade, Holly Houdeshell, Ola Kvalvaag, Dillon Nall, Ethan Sanders, Alec Vest, Shaun Westlund, Matthew Hardison, Clinton R. Paden, Duncan MacCannell |
| EPI_ISL_2044008, EPI_ISL_2044009, EPI_ISL_2044074, EPI_ISL_2044076, EPI_ISL_2044077, EPI_ISL_2044079, EPI_ISL_2044080, EPI_ISL_2044155, EPI_ISL_2044165, EPI_ISL_2044174, EPI_ISL_2044181, EPI_ISL_2044208, EPI_ISL_2044210, EPI_ISL_2044211, EPI_ISL_2044216, EPI_ISL_2044217, EPI_ISL_2044218, EPI_ISL_2044219, EPI_ISL_2044222, EPI_ISL_2044254, EPI_ISL_2044512, EPI_ISL_2044521, EPI_ISL_2044522, EPI_ISL_2044555, EPI_ISL_2044576, EPI_ISL_2044580, EPI_ISL_2044581, EPI_ISL_2044638, EPI_ISL_2044644, EPI_ISL_2044647, EPI_ISL_2044663, EPI_ISL_2044671, EPI_ISL_2044713, EPI_ISL_2044761, EPI_ISL_2044934, EPI_ISL_2044935, EPI_ISL_2044936, EPI_ISL_2044937, EPI_ISL_2044981, EPI_ISL_2044993, EPI_ISL_2044995, EPI_ISL_2044998, EPI_ISL_2045017, EPI_ISL_2045036, EPI_ISL_2045037, EPI_ISL_2045039, EPI_ISL_2045087, EPI_ISL_2045089, EPI_ISL_2045090, EPI_ISL_2045095, EPI_ISL_2045469, EPI_ISL_2045470, EPI_ISL_2045471, EPI_ISL_2045472, EPI_ISL_2045473, EPI_ISL_2045474, EPI_ISL_2045475, EPI_ISL_2045476, EPI_ISL_2045559, EPI_ISL_2045571, EPI_ISL_2045572, EPI_ISL_2045720, EPI_ISL_2045725, EPI_ISL_2045726, EPI_ISL_2045727, EPI_ISL_2045728, EPI_ISL_2045729, EPI_ISL_2045750, EPI_ISL_2045751, EPI_ISL_2045754, EPI_ISL_2045756, EPI_ISL_2045785, EPI_ISL_2045786, EPI_ISL_2045793, EPI_ISL_2045794, EPI_ISL_2045795, EPI_ISL_2045796, EPI_ISL_2045797, EPI_ISL_2045798, EPI_ISL_2045799, EPI_ISL_2045800, EPI_ISL_2045815, EPI_ISL_2045816, EPI_ISL_2045817, EPI_ISL_2045818, EPI_ISL_2045820, EPI_ISL_2045821, EPI_ISL_2045822, EPI_ISL_2045823, EPI_ISL_2045853, EPI_ISL_2045854, EPI_ISL_2045859, EPI_ISL_2045860 | | | |
| see above | Laboratory Corporation of America | Centers for Disease Control and Prevention Division of Viral Diseases, Pathogen Discovery | Dakota Howard, Dhwani Batra, Peter W. Cook, Kara Moser, Adrian Paskey, Jason Caravas, Benjamin Rambo-Martin, Shatavia Morrison, Christopher Gulvick, Scott Sammons, Yvette Unoarumhi, Darlene Wagner, Matthew Schmerer, Minoo Agarwal, Eyad Almasri, Debbie Boles, Ayla Burns, Nuthawin Charoensri, Oren Cohen, Susan Countryman, Mary Ann Cristobal, Bobbi Croy, Suzanne Dale, Hrushikesh Deshmukh, Amanda Douglas, Vincent Drouillon, Marcia Eisenberg, Howard Engler, Rama Ghatti, Prashant Gupta, Susan Hicks, Jake Humphrey, Lax Iyer, Manoj Jain, Mohan Kolli, Brian Krueger, Tim Kuphal, Stanley Letovsky, Michael Levandoski, Craig Lukasik, Jonathan Meltzer, Brian Norvell, Mindy Nye, Scott Parker, Christos Petropoulos, John Pruitt, Steven Ragan, Scott Ryan, Mike Sapeta, Jana Schroth, Suresh Babu Selvaraju, Goran Stevovic, Amanda Suchanek, Andrea Throop, Lyndon Tilson, Thomas Urban, Joe Voshell, Kimberly Wagner, Jonathan Williams, Mary Williamson, Qian Zeng, Tricia Zwiefelhofer, Clinton R. Paden, Duncan MacCannell |
| EPI_ISL_2047034, EPI_ISL_2047130, EPI_ISL_2047131, EPI_ISL_2047140, EPI_ISL_2047157, EPI_ISL_2047159, EPI_ISL_2047188, EPI_ISL_2047193, EPI_ISL_2047226, EPI_ISL_2047239, EPI_ISL_2047241, EPI_ISL_2047248, EPI_ISL_2047256, EPI_ISL_2047262, EPI_ISL_2047273, EPI_ISL_2047274, EPI_ISL_2047277 | | | |
| see above | Quest Diagnostics Incorporated | Centers for Disease Control and Prevention Division of Viral Diseases, Pathogen Discovery | Dakota Howard, Dhwani Batra, Peter W. Cook, Kara Moser, Adrian Paskey, Jason Caravas, Benjamin Rambo-Martin, Shatavia Morrison, Christopher Gulvick, Scott Sammons, Yvette Unoarumhi, Darlene Wagner, Matthew Schmerer, S. H. Rosenthal, A. Gerasimova, R. M. Kagan, B. Anderson, M. Hua, Y. Liu, L.E. Bernstein, K.E. Livingston, A. Perez, I. A. Shlyakhter, R. V. Rolando, R. Owen, P. Tanpaiboon, F. Lacbawan, Clinton R. Paden, Duncan MacCannell |
| EPI_ISL_2047506 | NYC Department of Health and Mental Hygiene | Centers for Disease Control and Prevention Division of Viral Diseases, Pathogen Discovery | Mili Sheth, Sarah Nobles, Jasmine Padilla, Mark Burroughs, Shoshona Le, Katie Dillon, Peter Cook, Clinton R. Paden, Dhwani Batra, Krista Queen, Kristen Knipe, Dakota Howard, Yvette Unoarumhi, Darlene Wagner, Matthew Schmerer, Ben L. Rambo-Martin, Kristine Lacek, Sam Shepard, Alison Laufer Halpin, Dave Wentworth, Vivien Dugan, Suxiang Tong, Justin Lee |
| EPI_ISL_2081128, EPI_ISL_2081129, EPI_ISL_2081130, EPI_ISL_2081131, EPI_ISL_2081132, EPI_ISL_2081133, EPI_ISL_2081134, EPI_ISL_2081135, EPI_ISL_2081136, EPI_ISL_2081137, EPI_ISL_2081138, EPI_ISL_2081139, EPI_ISL_2081140, EPI_ISL_2081141, EPI_ISL_2081142, EPI_ISL_2081143, EPI_ISL_2081144, EPI_ISL_2081145, EPI_ISL_2081146, EPI_ISL_2081147, EPI_ISL_2081148, EPI_ISL_2081149, EPI_ISL_2081150, EPI_ISL_2081151, EPI_ISL_2081152, EPI_ISL_2081153, EPI_ISL_2081154, EPI_ISL_2081155, EPI_ISL_2081156, EPI_ISL_2081157, EPI_ISL_2081158, EPI_ISL_2081159, EPI_ISL_2081160, EPI_ISL_2081161, EPI_ISL_2081162, EPI_ISL_2081163, EPI_ISL_2081164, EPI_ISL_2081165, EPI_ISL_2081166, EPI_ISL_2081167, EPI_ISL_2081168, EPI_ISL_2082729, EPI_ISL_2086266, EPI_ISL_2086300, EPI_ISL_2086301, EPI_ISL_2086302, EPI_ISL_2086303 | | | |
| see above | Weill Cornell Medicine | New York Genome Center | Michael Zody, Andre Corvelo, Dayna M. Oschwald, Samantha Fennessey, Tom Maniatis, Melissa Cushing, Olivier Elemento, Margaret Elizabeth Ross, Chris Mason, Priya Velu, Hanna Rennert, Arryn Craney, Lars F Westblade |
| EPI_ISL_2087865, EPI_ISL_2088170, EPI_ISL_2088271, EPI_ISL_2088913, EPI_ISL_2089185, EPI_ISL_2089216, EPI_ISL_2089237, EPI_ISL_2089245, EPI_ISL_2089329, EPI_ISL_2089442 | Infinity Biologix | Centers for Disease Control and Prevention Division of Viral Diseases, Pathogen Discovery | Dakota Howard, Dhwani Batra, Peter W. Cook, Kara Moser, Adrian Paskey, Jason Caravas, Benjamin Rambo-Martin, Shatavia Morrison, Christopher Gulvick, Scott Sammons, Yvette Unoarumhi, Darlene Wagner, Matthew Schmerer, Christian Bixby, Yihe Wang, Jonathan Schultz, Chirayu Goswami, Russ Hager, Robin Grimwood, Clinton R. Paden, Duncan MacCannell |
| EPI_ISL_2089530, EPI_ISL_2089599, EPI_ISL_2089607, EPI_ISL_2089673, EPI_ISL_2089704, EPI_ISL_2089736, EPI_ISL_2089737, EPI_ISL_2089765, EPI_ISL_2089768, EPI_ISL_2089769, EPI_ISL_2089770, EPI_ISL_2089810, EPI_ISL_2089952, EPI_ISL_2089982, EPI_ISL_2089986, EPI_ISL_2090007, EPI_ISL_2090133, EPI_ISL_2090151, EPI_ISL_2090170, EPI_ISL_2090181, EPI_ISL_2090182, EPI_ISL_2090210, EPI_ISL_2090211, EPI_ISL_2090219, EPI_ISL_2090220, EPI_ISL_2090235, EPI_ISL_2090239, EPI_ISL_2090240, EPI_ISL_2090241, EPI_ISL_2090408, EPI_ISL_2090426, EPI_ISL_2090458, EPI_ISL_2090468 | | | |
| see above | Aegis Sciences Corporation | Centers for Disease Control and Prevention Division of Viral Diseases, Pathogen Discovery | Dakota Howard, Dhwani Batra, Peter W. Cook, Kara Moser, Adrian Paskey, Jason Caravas, Benjamin Rambo-Martin, Shatavia Morrison, Christopher Gulvick, Scott Sammons, Yvette Unoarumhi, Darlene Wagner, Matthew Schmerer, Cyndi Clark, Patrick Campbell, Rob Case, Vikramsinha Ghorpade, Holly Houdeshell, Ola Kvalvaag, Dillon Nall, Ethan Sanders, Alec Vest, Shaun Westlund, Matthew Hardison, Clinton R. Paden, Duncan MacCannell |
| EPI_ISL_2096714, EPI_ISL_2096715, EPI_ISL_2096716, EPI_ISL_2096717, EPI_ISL_2096718, EPI_ISL_2096719, EPI_ISL_2096720, EPI_ISL_2096721 | Broad Institute Clinical Research Sequencing Platform | Infectious Disease Program, Broad Institute of Harvard and MIT | Siddle,K.J., Adams,G., Pearlman,L., Gladden-Young,A., Vicente,G., Blumenstiel,B., DeFelice,M., Lee,M., McGovern,S., Lagerborg,K., Rudy,M., DeRuff,K., Carter,A., Normandin,E., Bauer,M., Reilly,S., Tomkins-Tinch,C., Loreth,C., Chaluvadi,S., Meldrim,J., Granger,B., Lemieux,J.E., Birren,B.W., Sabeti,P.C., Larkin,K., Dodge,S., Lennon,N., Madoff,L., Brown,C., Gallagher,G., Smole,S., Park,D.J., Gabriel,S., and MacInnis,B.L. |
| EPI_ISL_2097232, EPI_ISL_2097233, EPI_ISL_2097234, EPI_ISL_2097235, EPI_ISL_2097236, EPI_ISL_2097237, EPI_ISL_2097238 | ALBANY MEDICAL CENTER | Wadsworth Center, New York State Department of Health | Kirsten St. George, Daryl M. Lamson, Alexis Russell, Matthew Shudt, Melissa A Leisner, Jonathan Plitnick, Catharine Prussing, Navjot Singh, John Kelly, Erasmus Schneider, Erica Lasek-Nesselquist |
| EPI_ISL_2097239, EPI_ISL_2097240, EPI_ISL_2097241, EPI_ISL_2097242, EPI_ISL_2097243, EPI_ISL_2097244, EPI_ISL_2097245, EPI_ISL_2097246, EPI_ISL_2097247, EPI_ISL_2097248, EPI_ISL_2097249, EPI_ISL_2097250, EPI_ISL_2097251 | | | |
| see above | BOSTON HEART DIAGNOSTICS CORP | Wadsworth Center, New York State Department of Health | Kirsten St. George, Daryl M. Lamson, Alexis Russell, Matthew Shudt, Melissa A Leisner, Jonathan Plitnick, Catharine Prussing, Navjot Singh, John Kelly, Erasmus Schneider, Erica Lasek-Nesselquist |
| EPI_ISL_2097252 | CENTERS LAB NJ, LLC (DBA MEDLABS DIAGNOSTICS) | Wadsworth Center, New York State Department of Health | Kirsten St. George, Daryl M. Lamson, Alexis Russell, Matthew Shudt, Melissa A Leisner, Jonathan Plitnick, Catharine Prussing, Navjot Singh, John Kelly, Erasmus Schneider, Erica Lasek-Nesselquist |
| EPI_ISL_2097253, EPI_ISL_2097254, EPI_ISL_2097255, EPI_ISL_2097256, EPI_ISL_2097257, EPI_ISL_2097258, EPI_ISL_2097259, EPI_ISL_2097260, EPI_ISL_2097261, EPI_ISL_2097262, EPI_ISL_2097263, EPI_ISL_2097264, EPI_ISL_2097265 | | | |
| see above | MONTEFIORE MEDICAL CENTER LABORATORIES | Wadsworth Center, New York State Department of Health | Kirsten St. George, Daryl M. Lamson, Alexis Russell, Matthew Shudt, Melissa A Leisner, Jonathan Plitnick, Catharine Prussing, Navjot Singh, John Kelly, Erasmus Schneider, Erica Lasek-Nesselquist |
| EPI_ISL_2097266, EPI_ISL_2097267, EPI_ISL_2097268, EPI_ISL_2097269, EPI_ISL_2097270, EPI_ISL_2097271, EPI_ISL_2097272, EPI_ISL_2097273, EPI_ISL_2097274, EPI_ISL_2097275, EPI_ISL_2097276, EPI_ISL_2097277, EPI_ISL_2097278, EPI_ISL_2097279, EPI_ISL_2097280, EPI_ISL_2097281, EPI_ISL_2097282, EPI_ISL_2097283, EPI_ISL_2097284, EPI_ISL_2097285, EPI_ISL_2097286, EPI_ISL_2097287, EPI_ISL_2097288, EPI_ISL_2097289, EPI_ISL_2097290, EPI_ISL_2097291, EPI_ISL_2097292, EPI_ISL_2097293, EPI_ISL_2097294 | | | |
| see above | NORTHWELL HEALTH LABORATORIES | Wadsworth Center, New York State Department of Health | Kirsten St. George, Daryl M. Lamson, Alexis Russell, Matthew Shudt, Melissa A Leisner, Jonathan Plitnick, Catharine Prussing, Navjot Singh, John Kelly, |

| | | | |
|---|---|---|---|
| | | | Erasmus Schneider, Erica Lasek-Nesselquist |
| EPI_ISL_2097295 | ST. CATHERINE OF SIENA HOSPITAL LABORATORY | Wadsworth Center, New York State Department of Health | Kirsten St. George, Daryl M. Lamson, Alexis Russell, Matthew Shudt, Melissa A Leisner, Jonathan Plitnick, Catharine Prussing, Navjot Singh, John Kelly, Erasmus Schneider, Erica Lasek-Nesselquist |
| EPI_ISL_2097296, EPI_ISL_2097297, EPI_ISL_2097298, EPI_ISL_2097299, EPI_ISL_2097300, EPI_ISL_2097301, EPI_ISL_2097302, EPI_ISL_2097303 | STONY BROOK UNIVERSITY HOSPITAL | Wadsworth Center, New York State Department of Health | Kirsten St. George, Daryl M. Lamson, Alexis Russell, Matthew Shudt, Melissa A Leisner, Jonathan Plitnick, Catharine Prussing, Navjot Singh, John Kelly, Erasmus Schneider, Erica Lasek-Nesselquist |
| EPI_ISL_2097304, EPI_ISL_2097305 | SUNY UPSTATE MEDICAL UNIVERSITY | Wadsworth Center, New York State Department of Health | Kirsten St. George, Daryl M. Lamson, Alexis Russell, Matthew Shudt, Melissa A Leisner, Jonathan Plitnick, Catharine Prussing, Navjot Singh, John Kelly, Erasmus Schneider, Erica Lasek-Nesselquist |
| EPI_ISL_2097306, EPI_ISL_2097307, EPI_ISL_2097308, EPI_ISL_2097309, EPI_ISL_2097310, EPI_ISL_2097311, EPI_ISL_2097312, EPI_ISL_2097313, EPI_ISL_2097314 | WESTCHESTER MEDICAL CENTER | Wadsworth Center, New York State Department of Health | Kirsten St. George, Daryl M. Lamson, Alexis Russell, Matthew Shudt, Melissa A Leisner, Jonathan Plitnick, Catharine Prussing, Navjot Singh, John Kelly, Erasmus Schneider, Erica Lasek-Nesselquist |
| EPI_ISL_2098115, EPI_ISL_2098117, EPI_ISL_2098119, EPI_ISL_2098120, EPI_ISL_2098122, EPI_ISL_2098123, EPI_ISL_2098125 | ALBANY MEDICAL CENTER | Wadsworth Center, New York State Department of Health | Kirsten St. George, Daryl M. Lamson, Alexis Russell, Matthew Shudt, Melissa A Leisner, Jonathan Plitnick, Catharine Prussing, Navjot Singh, John Kelly, Erasmus Schneider, Erica Lasek-Nesselquist |
| EPI_ISL_2098126, EPI_ISL_2098128 | BOSTON HEART DIAGNOSTICS CORP | Wadsworth Center, New York State Department of Health | Kirsten St. George, Daryl M. Lamson, Alexis Russell, Matthew Shudt, Melissa A Leisner, Jonathan Plitnick, Catharine Prussing, Navjot Singh, John Kelly, Erasmus Schneider, Erica Lasek-Nesselquist |
| EPI_ISL_2098130 | MONTEFIORE MEDICAL CENTER LABORATORIES | Wadsworth Center, New York State Department of Health | Kirsten St. George, Daryl M. Lamson, Alexis Russell, Matthew Shudt, Melissa A Leisner, Jonathan Plitnick, Catharine Prussing, Navjot Singh, John Kelly, Erasmus Schneider, Erica Lasek-Nesselquist |
| EPI_ISL_2098132 | NORTHWELL HEALTH LABORATORIES | Wadsworth Center, New York State Department of Health | Kirsten St. George, Daryl M. Lamson, Alexis Russell, Matthew Shudt, Melissa A Leisner, Jonathan Plitnick, Catharine Prussing, Navjot Singh, John Kelly, Erasmus Schneider, Erica Lasek-Nesselquist |
| EPI_ISL_2098133 | SARATOGA HOSPITAL LABORATORY | Wadsworth Center, New York State Department of Health | Kirsten St. George, Daryl M. Lamson, Alexis Russell, Matthew Shudt, Melissa A Leisner, Jonathan Plitnick, Catharine Prussing, Navjot Singh, John Kelly, Erasmus Schneider, Erica Lasek-Nesselquist |
| EPI_ISL_2098135, EPI_ISL_2098136, EPI_ISL_2098138, EPI_ISL_2098140, EPI_ISL_2098141, EPI_ISL_2098143, EPI_ISL_2098144, EPI_ISL_2098146, EPI_ISL_2098147, EPI_ISL_2098149, EPI_ISL_2098150, EPI_ISL_2098151, EPI_ISL_2098153, EPI_ISL_2098155, EPI_ISL_2098156, EPI_ISL_2098158, EPI_ISL_2098159 | | | |
| see above | SUNY UPSTATE MEDICAL UNIVERSITY | Wadsworth Center, New York State Department of Health | Kirsten St. George, Daryl M. Lamson, Alexis Russell, Matthew Shudt, Melissa A Leisner, Jonathan Plitnick, Catharine Prussing, Navjot Singh, John Kelly, Erasmus Schneider, Erica Lasek-Nesselquist |
| EPI_ISL_2098161, EPI_ISL_2098162, EPI_ISL_2098164, EPI_ISL_2098166, EPI_ISL_2098167, EPI_ISL_2098169, EPI_ISL_2098170, EPI_ISL_2098172, EPI_ISL_2098173, EPI_ISL_2098175, EPI_ISL_2098176, EPI_ISL_2098178, EPI_ISL_2098180, EPI_ISL_2098181, EPI_ISL_2098183, EPI_ISL_2098184, EPI_ISL_2098186, EPI_ISL_2098187, EPI_ISL_2098189, EPI_ISL_2098191, EPI_ISL_2098192, EPI_ISL_2098193, EPI_ISL_2098195, EPI_ISL_2098196, EPI_ISL_2098198, EPI_ISL_2098199, EPI_ISL_2098201, EPI_ISL_2098202, EPI_ISL_2098204, EPI_ISL_2098206, EPI_ISL_2098208, EPI_ISL_2098209, EPI_ISL_2098211, EPI_ISL_2098212, EPI_ISL_2098214, EPI_ISL_2098215, EPI_ISL_2098217, EPI_ISL_2098218, EPI_ISL_2098220, EPI_ISL_2098222, EPI_ISL_2098223, EPI_ISL_2098224, EPI_ISL_2098225, EPI_ISL_2098227, EPI_ISL_2098228, EPI_ISL_2098230, EPI_ISL_2098231 | | | |
| see above | URMC LABS | Wadsworth Center, New York State Department of Health | Kirsten St. George, Daryl M. Lamson, Alexis Russell, Matthew Shudt, Melissa A Leisner, Jonathan Plitnick, Catharine Prussing, Navjot Singh, John Kelly, Erasmus Schneider, Erica Lasek-Nesselquist |
| EPI_ISL_2098233 | WESTCHESTER MEDICAL CENTER | Wadsworth Center, New York State Department of Health | Kirsten St. George, Daryl M. Lamson, Alexis Russell, Matthew Shudt, Melissa A Leisner, Jonathan Plitnick, Catharine Prussing, Navjot Singh, John Kelly, Erasmus Schneider, Erica Lasek-Nesselquist |
| EPI_ISL_2098274, EPI_ISL_2098277, EPI_ISL_2098279, EPI_ISL_2098287, EPI_ISL_2098291, EPI_ISL_2098304, EPI_ISL_2098309, EPI_ISL_2098328, EPI_ISL_2098361, EPI_ISL_2098362, EPI_ISL_2098367, EPI_ISL_2098372, EPI_ISL_2098383, EPI_ISL_2098402, EPI_ISL_2098406 | | | |
| see above | Quest Diagnostics Incorporated | Centers for Disease Control and Prevention Division of Viral Diseases, Pathogen Discovery | Dakota Howard, Dhwani Batra, Peter W. Cook, Kara Moser, Adrian Paskey, Jason Caravas, Benjamin Rambo-Martin, Shatavia Morrison, Christopher Gulvick, Scott Sammons, Yvette Unoarumhi, Darlene Wagner, Matthew Schmerer, S. H. Rosenthal, A. Gerasimova, R. M. Kagan, B. Anderson, M. Hua, Y. Liu, L.E. Bernstein, K.E. Livingston, A. Perez, I. A. Shlyakhter, R. V. Rolando, R. Owen, P. Tanpaiboon, F. Lacbawan, Clinton R. Paden, Duncan MacCannell |
| EPI_ISL_2098499 | ADIRONDACK MEDICAL CENTER | Wadsworth Center, New York State Department of Health | Kirsten St. George, Daryl M. Lamson, Alexis Russell, Matthew Shudt, Melissa A Leisner, Jonathan Plitnick, Catharine Prussing, Navjot Singh, John Kelly, Erasmus Schneider, Erica Lasek-Nesselquist |
| EPI_ISL_2098500 | ALBANY MEDICAL CENTER | Wadsworth Center, New York State Department of Health | Kirsten St. George, Daryl M. Lamson, Alexis Russell, Matthew Shudt, Melissa A Leisner, Jonathan Plitnick, Catharine Prussing, Navjot Singh, John Kelly, Erasmus Schneider, Erica Lasek-Nesselquist |
| EPI_ISL_2098501, EPI_ISL_2098502, EPI_ISL_2098503, EPI_ISL_2098504, EPI_ISL_2098505, EPI_ISL_2098506, EPI_ISL_2098507, EPI_ISL_2098508, EPI_ISL_2098509, EPI_ISL_2098510, EPI_ISL_2098511, EPI_ISL_2098512, EPI_ISL_2098513, EPI_ISL_2098514, EPI_ISL_2098515, EPI_ISL_2098516, EPI_ISL_2098517, EPI_ISL_2098518, EPI_ISL_2098519, EPI_ISL_2098520, EPI_ISL_2098521, EPI_ISL_2098522, EPI_ISL_2098523, EPI_ISL_2098524, EPI_ISL_2098525, EPI_ISL_2098526, EPI_ISL_2098527, EPI_ISL_2098528, EPI_ISL_2098529, EPI_ISL_2098530, EPI_ISL_2098531, EPI_ISL_2098532, EPI_ISL_2098533, EPI_ISL_2098534, EPI_ISL_2098535, EPI_ISL_2098536, EPI_ISL_2098537, EPI_ISL_2098538, EPI_ISL_2098539, EPI_ISL_2098540, EPI_ISL_2098541, EPI_ISL_2098542, EPI_ISL_2098543, EPI_ISL_2098544, EPI_ISL_2098545, EPI_ISL_2098546, EPI_ISL_2098547, EPI_ISL_2098548, EPI_ISL_2098549, EPI_ISL_2098550, EPI_ISL_2098551, EPI_ISL_2098552, EPI_ISL_2098553, EPI_ISL_2098554, EPI_ISL_2098555, EPI_ISL_2098556, EPI_ISL_2098557, EPI_ISL_2098558, EPI_ISL_2098559, EPI_ISL_2098560, EPI_ISL_2098561, EPI_ISL_2098562, EPI_ISL_2098563, EPI_ISL_2098564, EPI_ISL_2098565, EPI_ISL_2098566, EPI_ISL_2098567, EPI_ISL_2098568, EPI_ISL_2098569, EPI_ISL_2098570, EPI_ISL_2098571, EPI_ISL_2098572, EPI_ISL_2098573, EPI_ISL_2098574, EPI_ISL_2098575, EPI_ISL_2098576, EPI_ISL_2098577, EPI_ISL_2098578, EPI_ISL_2098579, EPI_ISL_2098580, EPI_ISL_2098581, EPI_ISL_2098582, EPI_ISL_2098583, EPI_ISL_2098584, EPI_ISL_2098585, EPI_ISL_2098586, EPI_ISL_2098587 | | | |
| see above | URMC LABS | Wadsworth Center, New York State Department of Health | Kirsten St. George, Daryl M. Lamson, Alexis Russell, Matthew Shudt, Melissa A Leisner, Jonathan Plitnick, Catharine Prussing, Navjot Singh, John Kelly, Erasmus Schneider, Erica Lasek-Nesselquist |
| EPI_ISL_2098589, EPI_ISL_2098590, EPI_ISL_2098591, EPI_ISL_2098592, EPI_ISL_2098593, EPI_ISL_2098594, EPI_ISL_2098595, EPI_ISL_2098596, EPI_ISL_2098597, EPI_ISL_2098598, EPI_ISL_2098599 | | | |
| see above | BOSTON HEART DIAGNOSTICS CORP | Wadsworth Center, New York State Department of Health | Kirsten St. George, Daryl M. Lamson, Alexis Russell, Matthew Shudt, Melissa A Leisner, Jonathan Plitnick, Catharine Prussing, Navjot Singh, John Kelly, Erasmus Schneider, Erica Lasek-Nesselquist |
| EPI_ISL_2098600 | DELHI REHABILITATION AND NURSING CENTER | Wadsworth Center, New York State Department of Health | Kirsten St. George, Daryl M. Lamson, Alexis Russell, Matthew Shudt, Melissa A Leisner, Jonathan Plitnick, Catharine Prussing, Navjot Singh, John Kelly, Erasmus Schneider, Erica Lasek-Nesselquist |
| EPI_ISL_2098601, EPI_ISL_2098602, EPI_ISL_2098603, EPI_ISL_2098604, EPI_ISL_2098605, EPI_ISL_2098606, EPI_ISL_2098607, EPI_ISL_2098608, EPI_ISL_2098609, EPI_ISL_2098610, EPI_ISL_2098611, EPI_ISL_2098612, EPI_ISL_2098613, EPI_ISL_2098614, EPI_ISL_2098615, EPI_ISL_2098616, EPI_ISL_2098617, EPI_ISL_2098618, EPI_ISL_2098619, EPI_ISL_2098620, EPI_ISL_2098621 | | | |
| see above | NYC Pandemic Response Lab | Wadsworth Center, New York State Department of Health | Kirsten St. George, Daryl M. Lamson, Alexis Russell, Matthew Shudt, Melissa A Leisner, Jonathan Plitnick, Catharine Prussing, Navjot Singh, John Kelly, Erasmus Schneider, Erica Lasek-Nesselquist |
| EPI_ISL_2098622, EPI_ISL_2098623, EPI_ISL_2098624, EPI_ISL_2098625, EPI_ISL_2098626, EPI_ISL_2098627, EPI_ISL_2098628 | SARATOGA HOSPITAL LABORATORY | Wadsworth Center, New York State Department of Health | Kirsten St. George, Daryl M. Lamson, Alexis Russell, Matthew Shudt, Melissa A Leisner, Jonathan Plitnick, Catharine Prussing, Navjot Singh, John Kelly, Erasmus Schneider, Erica Lasek-Nesselquist |
| EPI_ISL_2098629, EPI_ISL_2098630 | ST. FRANCIS HOSPITAL LABORATORY | Wadsworth Center, New York State Department of Health | Kirsten St. George, Daryl M. Lamson, Alexis Russell, Matthew Shudt, Melissa A Leisner, Jonathan Plitnick, Catharine Prussing, Navjot Singh, John Kelly, Erasmus Schneider, Erica Lasek-Nesselquist |
| EPI_ISL_2098631, EPI_ISL_2098632, EPI_ISL_2098633, EPI_ISL_2098634, EPI_ISL_2098635, EPI_ISL_2098636, EPI_ISL_2098637, EPI_ISL_2098638, EPI_ISL_2098639, EPI_ISL_2098640, EPI_ISL_2098641, EPI_ISL_2098642, EPI_ISL_2098643, EPI_ISL_2098644, EPI_ISL_2098645, EPI_ISL_2098646, EPI_ISL_2098647, EPI_ISL_2098648, EPI_ISL_2098649, EPI_ISL_2098650, EPI_ISL_2098651, EPI_ISL_2098652, EPI_ISL_2098653, EPI_ISL_2098654, EPI_ISL_2098655, EPI_ISL_2098656, EPI_ISL_2098657, EPI_ISL_2098658, EPI_ISL_2098659 | | | |
| see above | SUNY UPSTATE MEDICAL UNIVERSITY | Wadsworth Center, New York State Department of Health | Kirsten St. George, Daryl M. Lamson, Alexis Russell, Matthew Shudt, Melissa A Leisner, Jonathan Plitnick, Catharine Prussing, Navjot Singh, John Kelly, Erasmus Schneider, Erica Lasek-Nesselquist |
| EPI_ISL_2098660, EPI_ISL_2098661, | URMC LABS | Wadsworth Center, New York State Department of Health | Kirsten St. George, Daryl M. Lamson, Alexis Russell, Matthew Shudt, Melissa A Leisner, Jonathan Plitnick, Catharine Prussing, Navjot Singh, John Kelly, |

| | | | |
|---|---|---|---|
| EPI_ISL_2098662, EPI_ISL_2098663, EPI_ISL_2098664, EPI_ISL_2098665 | | | Erasmus Schneider, Erica Lasek-Nesselquist |
| EPI_ISL_2101788, EPI_ISL_2101789, EPI_ISL_2101790, EPI_ISL_2101800 | Bronx Veterans Medical Research Foundation | Pathogen Discovery, Respiratory Viruses Branch, Division of Viral Diseases, Centers for Disease Control and Prevention | Ying Tao, Yan Li, Jing Zhang, Anna Kelleher, Brian Lynch, Krista Queen, Anna Uehara, Peter Cook, Han Jia Justin Ng, Rachel Marine, Clinton R. Paden, Dhwani Batra, Haibin Wang, Tara Coalter, Jasmine Padilla, Morgan Davis, Mili Sheth, Sarah Nobles, Mark Burroughs, Justin Lee, Adam Retchless, Suxiang Tong |
| EPI_ISL_2104331, EPI_ISL_2104332, EPI_ISL_2104333, EPI_ISL_2104334, EPI_ISL_2104335 | Broad Institute Clinical Research Sequencing Platform | Infectious Disease Program, Broad Institute of Harvard and MIT | Siddle,K.J., Adams,G., Pearlman,L., Gladden-Young,A., Vicente,G., Blumenstiel,B., DeFelice,M., Lee,M., McGovern,S., Lagerborg,K., Rudy,M., DeRuff,K., Carter,A., Normandin,E., Bauer,M., Reilly,S., Tomkins-Tinch,C., Loreth,C., Chaluvadi,S., Meldrim,J., Granger,B., Lemieux,J.E., Birren,B.W., Sabeti,P.C., Larkin,K., Dodge,S., Lennon,N., Madoff,L., Brown,C., Gallagher,G., Smole,S., Park,D.J., Gabriel,S., and MacInnis,B.L. |
| EPI_ISL_2106283, EPI_ISL_2106284, EPI_ISL_2106285, EPI_ISL_2106286, EPI_ISL_2106287, EPI_ISL_2106288, EPI_ISL_2106289, EPI_ISL_2106290, EPI_ISL_2106291, EPI_ISL_2106292, EPI_ISL_2106293, EPI_ISL_2106294, EPI_ISL_2106295, EPI_ISL_2106296, EPI_ISL_2106297, EPI_ISL_2106298, EPI_ISL_2106299, EPI_ISL_2106300, EPI_ISL_2106301, EPI_ISL_2106302, EPI_ISL_2106303, EPI_ISL_2106304, EPI_ISL_2106305, EPI_ISL_2106306, EPI_ISL_2106307, EPI_ISL_2106308, EPI_ISL_2106309, EPI_ISL_2106310, EPI_ISL_2106311, EPI_ISL_2106312, EPI_ISL_2106313, EPI_ISL_2106314, EPI_ISL_2106315, EPI_ISL_2106316, EPI_ISL_2106317, EPI_ISL_2106318, EPI_ISL_2106319, EPI_ISL_2106320, EPI_ISL_2106321, EPI_ISL_2106322, EPI_ISL_2106323, EPI_ISL_2106324, EPI_ISL_2106325, EPI_ISL_2106326, EPI_ISL_2106327, EPI_ISL_2106328, EPI_ISL_2106329, EPI_ISL_2106330, EPI_ISL_2106331, EPI_ISL_2106332, EPI_ISL_2106333, EPI_ISL_2106334, EPI_ISL_2106335, EPI_ISL_2106336, EPI_ISL_2106337, EPI_ISL_2106338, EPI_ISL_2106339, EPI_ISL_2106340, EPI_ISL_2106341, EPI_ISL_2106342, EPI_ISL_2106343, EPI_ISL_2106344, EPI_ISL_2106345, EPI_ISL_2106346, EPI_ISL_2106347, EPI_ISL_2106348, EPI_ISL_2106349, EPI_ISL_2106350, EPI_ISL_2106351, EPI_ISL_2106352, EPI_ISL_2106353, EPI_ISL_2106354, EPI_ISL_2106355, EPI_ISL_2106356, EPI_ISL_2106357, EPI_ISL_2106358, EPI_ISL_2106359, EPI_ISL_2106360, EPI_ISL_2106361, EPI_ISL_2106362, EPI_ISL_2106363, EPI_ISL_2106364, EPI_ISL_2106365, EPI_ISL_2106366, EPI_ISL_2106367, EPI_ISL_2106368, EPI_ISL_2106369, EPI_ISL_2106370, EPI_ISL_2106371, EPI_ISL_2106372, EPI_ISL_2106373, EPI_ISL_2106374, EPI_ISL_2106375, EPI_ISL_2106376, EPI_ISL_2106377, EPI_ISL_2106378, EPI_ISL_2106379, EPI_ISL_2106380, EPI_ISL_2106381, EPI_ISL_2106382, EPI_ISL_2106383, EPI_ISL_2106384, EPI_ISL_2106385, EPI_ISL_2106386, EPI_ISL_2106387, EPI_ISL_2106388, EPI_ISL_2106389, EPI_ISL_2106390, EPI_ISL_2106391, EPI_ISL_2106392, EPI_ISL_2106393, EPI_ISL_2106394, EPI_ISL_2106395, EPI_ISL_2106396, EPI_ISL_2106397, EPI_ISL_2106398, EPI_ISL_2106399, EPI_ISL_2106400, EPI_ISL_2106401, EPI_ISL_2106402, EPI_ISL_2106403, EPI_ISL_2106404, EPI_ISL_2106405, EPI_ISL_2106406, EPI_ISL_2106407, EPI_ISL_2106408, EPI_ISL_2106409, EPI_ISL_2106410, EPI_ISL_2106411, EPI_ISL_2106412, EPI_ISL_2106413, EPI_ISL_2106414, EPI_ISL_2106415, EPI_ISL_2106416, EPI_ISL_2106417, EPI_ISL_2106418, EPI_ISL_2106419, EPI_ISL_2106420, EPI_ISL_2106421, EPI_ISL_2106422, EPI_ISL_2106423, EPI_ISL_2106424, EPI_ISL_2106425, EPI_ISL_2106426, EPI_ISL_2106427, EPI_ISL_2106428, EPI_ISL_2106429, EPI_ISL_2106430, EPI_ISL_2106431, EPI_ISL_2106432, EPI_ISL_2106433, EPI_ISL_2106434, EPI_ISL_2106435, EPI_ISL_2106436, EPI_ISL_2106437, EPI_ISL_2106438, EPI_ISL_2106439, EPI_ISL_2106440, EPI_ISL_2106441, EPI_ISL_2106442, EPI_ISL_2106443, EPI_ISL_2106444, EPI_ISL_2106445, EPI_ISL_2106446, EPI_ISL_2106447, EPI_ISL_2106448, EPI_ISL_2106449, EPI_ISL_2106450, EPI_ISL_2106451, EPI_ISL_2106452, EPI_ISL_2106453, EPI_ISL_2106454, EPI_ISL_2106455, EPI_ISL_2106456, EPI_ISL_2106457, EPI_ISL_2106458, EPI_ISL_2106459, EPI_ISL_2106460, EPI_ISL_2106461, EPI_ISL_2106462, EPI_ISL_2106463, EPI_ISL_2106464, EPI_ISL_2106465, EPI_ISL_2106466, EPI_ISL_2106467, EPI_ISL_2106468, EPI_ISL_2106469, EPI_ISL_2106470, EPI_ISL_2106471, EPI_ISL_2106472, EPI_ISL_2106473, EPI_ISL_2106474, EPI_ISL_2106475, EPI_ISL_2106476, EPI_ISL_2106477, EPI_ISL_2106478, EPI_ISL_2106479, EPI_ISL_2106480, EPI_ISL_2106481, EPI_ISL_2106482, EPI_ISL_2106483, EPI_ISL_2106484, EPI_ISL_2106485, EPI_ISL_2106486, EPI_ISL_2106487, EPI_ISL_2106488, EPI_ISL_2106489, EPI_ISL_2106490, EPI_ISL_2106491, EPI_ISL_2106492, EPI_ISL_2106493, EPI_ISL_2106494, EPI_ISL_2106495, EPI_ISL_2106496, EPI_ISL_2106497, EPI_ISL_2106498, EPI_ISL_2106499, EPI_ISL_2106500, EPI_ISL_2106501, EPI_ISL_2106502, EPI_ISL_2106503, EPI_ISL_2106504, EPI_ISL_2106505, EPI_ISL_2106506, EPI_ISL_2106507, EPI_ISL_2106508, EPI_ISL_2106509, EPI_ISL_2106510, EPI_ISL_2106511, EPI_ISL_2106512, EPI_ISL_2106513, EPI_ISL_2106514, EPI_ISL_2106515, EPI_ISL_2106516, EPI_ISL_2106517, EPI_ISL_2106518, EPI_ISL_2106519, EPI_ISL_2106520, EPI_ISL_2106521, EPI_ISL_2106522, EPI_ISL_2106523, EPI_ISL_2106524, EPI_ISL_2106525, EPI_ISL_2106526, EPI_ISL_2106527, EPI_ISL_2106528, EPI_ISL_2106529, EPI_ISL_2106530, EPI_ISL_2106531, EPI_ISL_2106532, EPI_ISL_2106533, EPI_ISL_2106534, EPI_ISL_2106535, EPI_ISL_2106536, EPI_ISL_2106537, EPI_ISL_2106538, EPI_ISL_2106539, EPI_ISL_2106540, EPI_ISL_2106541, EPI_ISL_2106542, EPI_ISL_2106543, EPI_ISL_2106544, EPI_ISL_2106545, EPI_ISL_2106546, EPI_ISL_2106547, EPI_ISL_2106548, EPI_ISL_2106549, EPI_ISL_2106550, EPI_ISL_2106551, EPI_ISL_2106552, EPI_ISL_2106553, EPI_ISL_2106554, EPI_ISL_2106555, EPI_ISL_2106556, EPI_ISL_2106557, EPI_ISL_2106558, EPI_ISL_2106559, EPI_ISL_2106560, EPI_ISL_2106561, EPI_ISL_2106562, EPI_ISL_2106563, EPI_ISL_2106564, EPI_ISL_2106565, EPI_ISL_2106566, EPI_ISL_2106567, EPI_ISL_2106568, EPI_ISL_2106569, EPI_ISL_2106570, EPI_ISL_2106571, EPI_ISL_2106572, EPI_ISL_2106573, EPI_ISL_2106574, EPI_ISL_2106575, EPI_ISL_2106576, EPI_ISL_2106577, EPI_ISL_2106578, EPI_ISL_2106579, EPI_ISL_2106580, EPI_ISL_2106581, EPI_ISL_2106582, EPI_ISL_2106583, EPI_ISL_2106584, EPI_ISL_2106585, EPI_ISL_2106586, EPI_ISL_2106587, EPI_ISL_2106588, EPI_ISL_2106589, EPI_ISL_2106590, EPI_ISL_2106591, EPI_ISL_2106592, EPI_ISL_2106593, EPI_ISL_2106594 | | | |
| see above | Pandemic Response Lab - NYC | Pandemic Response Lab, R&D | Henry Lee, Michael Hammerling, Melissa Hopkins, Cybill del Castillo, Shinyoung Clair Kang, William Ward, Pradeep Bugga, Sol Rey, Dylan Law, Katharine Nelson, Haiping Hao, Jon Laurent |
| EPI_ISL_2132561 | Aegis Sciences Corporation | Centers for Disease Control and Prevention Division of Viral Diseases, Pathogen Discovery | Dakota Howard, Dhwani Batra, Peter W. Cook, Kara Moser, Adrian Paskey, Jason Caravas, Benjamin Rambo-Martin, Shatavia Morrison, Christopher Gulvick, Scott Sammons, Yvette Unoarumhi, Darlene Wagner, Matthew Schmerer, Cyndi Clark, Patrick Campbell, Rob Case, Vikramsinha Ghorpade, Holly Houdeshell, Ola Kvalvaag, Dillon Nall, Ethan Sanders, Alec Vest, Shaun Westlund, Matthew Hardison, Clinton R. Paden, Duncan MacCannell |
| EPI_ISL_2132626, EPI_ISL_2133297, EPI_ISL_2133301, EPI_ISL_2133580, EPI_ISL_2133608, EPI_ISL_2133615, EPI_ISL_2133638, EPI_ISL_2133693, EPI_ISL_2133749, EPI_ISL_2133770, EPI_ISL_2133820, EPI_ISL_2133842 | | | |
| see above | Quest Diagnostics Incorporated | Centers for Disease Control and Prevention Division of Viral Diseases, Pathogen Discovery | Dakota Howard, Dhwani Batra, Peter W. Cook, Kara Moser, Adrian Paskey, Jason Caravas, Benjamin Rambo-Martin, Shatavia Morrison, Christopher Gulvick, Scott Sammons, Yvette Unoarumhi, Darlene Wagner, Matthew Schmerer, S. H. Rosenthal, A. Gerasimova, R. M. Kagan, B. Anderson, M. Hua, Y. Liu, L.E. Bernstein, K.E. Livingston, A. Perez, I. A. Shlyakhter, R. V. Rolando, R. Owen, P. Tanpaiboon, F. Lacbawan, Clinton R. Paden, Duncan MacCannell |
| EPI_ISL_2134784 | BOSTON HEART DIAGNOSTICS CORP | Wadsworth Center, New York State Department of Health | Kirsten St. George, Daryl M. Lamson, Alexis Russell, Matthew Shudt, Melissa A Leisner, Jonathan Plitnick, Catharine Prussing, Navjot Singh, John Kelly, Erasmus Schneider, Erica Lasek-Nesselquist |
| EPI_ISL_2134785 | MONTEFIORE MEDICAL CENTER LABORATORIES | Wadsworth Center, New York State Department of Health | Kirsten St. George, Daryl M. Lamson, Alexis Russell, Matthew Shudt, Melissa A Leisner, Jonathan Plitnick, Catharine Prussing, Navjot Singh, John Kelly, Erasmus Schneider, Erica Lasek-Nesselquist |
| EPI_ISL_2134786, EPI_ISL_2134787 | NORTHWELL HEALTH LABORATORIES | Wadsworth Center, New York State Department of Health | Kirsten St. George, Daryl M. Lamson, Alexis Russell, Matthew Shudt, Melissa A Leisner, Jonathan Plitnick, Catharine Prussing, Navjot Singh, John Kelly, Erasmus Schneider, Erica Lasek-Nesselquist |
| EPI_ISL_2134788, EPI_ISL_2134789 | STONY BROOK UNIVERSITY HOSPITAL | Wadsworth Center, New York State Department of Health | Kirsten St. George, Daryl M. Lamson, Alexis Russell, Matthew Shudt, Melissa A Leisner, Jonathan Plitnick, Catharine Prussing, Navjot Singh, John Kelly, Erasmus Schneider, Erica Lasek-Nesselquist |
| EPI_ISL_2134790 | BOSTON HEART DIAGNOSTICS CORP | Wadsworth Center, New York State Department of Health | Kirsten St. George, Daryl M. Lamson, Alexis Russell, Matthew Shudt, Melissa A Leisner, Jonathan Plitnick, Catharine Prussing, Navjot Singh, John Kelly, Erasmus Schneider, Erica Lasek-Nesselquist |
| EPI_ISL_2134791 | MONTEFIORE MEDICAL CENTER LABORATORIES | Wadsworth Center, New York State Department of Health | Kirsten St. George, Daryl M. Lamson, Alexis Russell, Matthew Shudt, Melissa A Leisner, Jonathan Plitnick, Catharine Prussing, Navjot Singh, John Kelly, Erasmus Schneider, Erica Lasek-Nesselquist |
| EPI_ISL_2134792, EPI_ISL_2134793, EPI_ISL_2134794, EPI_ISL_2134795 | URMC LABS | Wadsworth Center, New York State Department of Health | Kirsten St. George, Daryl M. Lamson, Alexis Russell, Matthew Shudt, Melissa A Leisner, Jonathan Plitnick, Catharine Prussing, Navjot Singh, John Kelly, Erasmus Schneider, Erica Lasek-Nesselquist |
| EPI_ISL_2134796, EPI_ISL_2134797 | BOSTON HEART DIAGNOSTICS CORP | Wadsworth Center, New York State Department of Health | Kirsten St. George, Daryl M. Lamson, Alexis Russell, Matthew Shudt, Melissa A Leisner, Jonathan Plitnick, Catharine Prussing, Navjot Singh, John Kelly, Erasmus Schneider, Erica Lasek-Nesselquist |
| EPI_ISL_2134798 | SUNY UPSTATE MEDICAL UNIVERSITY | Wadsworth Center, New York State Department of Health | Kirsten St. George, Daryl M. Lamson, Alexis Russell, Matthew Shudt, Melissa A Leisner, Jonathan Plitnick, Catharine Prussing, Navjot Singh, John Kelly, Erasmus Schneider, Erica Lasek-Nesselquist |
| EPI_ISL_2134820 | PrimBio Research institute | PrimBio Research institute | Yang Liu |
| EPI_ISL_2140791, EPI_ISL_2142952, EPI_ISL_2143046, EPI_ISL_2143061, EPI_ISL_2143068, EPI_ISL_2143149, EPI_ISL_2143164, EPI_ISL_2143251, EPI_ISL_2143257 | Quest Diagnostics Incorporated | Centers for Disease Control and Prevention Division of Viral Diseases, Pathogen Discovery | Dakota Howard, Dhwani Batra, Peter W. Cook, Kara Moser, Adrian Paskey, Jason Caravas, Benjamin Rambo-Martin, Shatavia Morrison, Christopher Gulvick, Scott Sammons, Yvette Unoarumhi, Darlene Wagner, Matthew Schmerer, S. H. Rosenthal, A. Gerasimova, R. M. Kagan, B. Anderson, M. Hua, Y. Liu, L.E. Bernstein, K.E. Livingston, A. Perez, I. A. Shlyakhter, R. V. Rolando, R. Owen, P. Tanpaiboon, F. Lacbawan, Clinton R. Paden, Duncan MacCannell |
| EPI_ISL_2144378, EPI_ISL_2144388, EPI_ISL_2144820 | Helix/Illumina | Centers for Disease Control and Prevention Division of Viral Diseases, Pathogen Discovery | Dakota Howard, Dhwani Batra, Peter W. Cook, Kara Moser, Adrian Paskey, Jason Caravas, Benjamin Rambo-Martin, Shatavia Morrison, Christopher Gulvick, Scott Sammons, Yvette Unoarumhi, Darlene Wagner, Matthew Schmerer, Eileen de Feo, Jan Antico, Christine Tran, Matthew Tolentino, Shannon Wickline, Kim Gietzen, Brad Sickler, Jingtao Liu, Eric Allen, Phil Febbo, Nicole L. Washington, Simon White, Geraint Levan, Kelly Schiabor Barrett, Elizabeth Cirulli, Alexandre Bolze, Ary Ascencio, Charlotte Rivera-Garcia, Ryan Cho, Jason Nguyen, Sherry Wang, Jimmy Ramirez, Tyler Cassens, Efren Sandoval, Magnus Isaksson, William Lee, David Becker, Marc Laurent, James Lu, Clinton R. Paden, Duncan MacCannell |
| EPI_ISL_2145807, EPI_ISL_2145831, EPI_ISL_2145933, EPI_ISL_2145936, EPI_ISL_2145998, EPI_ISL_2146005, EPI_ISL_2146007, EPI_ISL_2146058, EPI_ISL_2146067, EPI_ISL_2146071, EPI_ISL_2146093, EPI_ISL_2146141, EPI_ISL_2146178, EPI_ISL_2146203, EPI_ISL_2146249, EPI_ISL_2146251, EPI_ISL_2146270, EPI_ISL_2146343, EPI_ISL_2146406, EPI_ISL_2146473, EPI_ISL_2146474, EPI_ISL_2146483, EPI_ISL_2146525, EPI_ISL_2146663, EPI_ISL_2146703, EPI_ISL_2146721, EPI_ISL_2146825, EPI_ISL_2146826, EPI_ISL_2146839, EPI_ISL_2146914, EPI_ISL_2146964, EPI_ISL_2146979, EPI_ISL_2146992, EPI_ISL_2147007, EPI_ISL_2147034, EPI_ISL_2147035, EPI_ISL_2147150, EPI_ISL_2147155, EPI_ISL_2147191, EPI_ISL_2147230, EPI_ISL_2147240, EPI_ISL_2147246, EPI_ISL_2147356, EPI_ISL_2147439, EPI_ISL_2147441, EPI_ISL_2147442, EPI_ISL_2147476, EPI_ISL_2147480, EPI_ISL_2147499, EPI_ISL_2147500, EPI_ISL_2147533, EPI_ISL_2147536, EPI_ISL_2147571, EPI_ISL_2147591, EPI_ISL_2147614, EPI_ISL_2147615, EPI_ISL_2147741, EPI_ISL_2147840, EPI_ISL_2147844, EPI_ISL_2147920, EPI_ISL_2147969, EPI_ISL_2147992, EPI_ISL_2148021, EPI_ISL_2148055, EPI_ISL_2148173, EPI_ISL_2148191, EPI_ISL_2148216, EPI_ISL_2148217, EPI_ISL_2148288, EPI_ISL_2148352, EPI_ISL_2148481, EPI_ISL_2148576, EPI_ISL_2148587, EPI_ISL_2148589, EPI_ISL_2148637, EPI_ISL_2148690, EPI_ISL_2148718, EPI_ISL_2148784, EPI_ISL_2148785, EPI_ISL_2148806, EPI_ISL_2148856, EPI_ISL_2148887, EPI_ISL_2148902, EPI_ISL_2148905, EPI_ISL_2148906, EPI_ISL_2148937, EPI_ISL_2149182, EPI_ISL_2149297, EPI_ISL_2149312, EPI_ISL_2149341, EPI_ISL_2149342, EPI_ISL_2149386, EPI_ISL_2149388, EPI_ISL_2149402, EPI_ISL_2149443, EPI_ISL_2149510, EPI_ISL_2149518, EPI_ISL_2149521, EPI_ISL_2149532, EPI_ISL_2149535, EPI_ISL_2149586, EPI_ISL_2149812, EPI_ISL_2149816, EPI_ISL_2149848, EPI_ISL_2149876, EPI_ISL_2149877, EPI_ISL_2149879, EPI_ISL_2149967, EPI_ISL_2150008, EPI_ISL_2150026, EPI_ISL_2150028, EPI_ISL_2150029, EPI_ISL_2150066, EPI_ISL_2150070, EPI_ISL_2150096, EPI_ISL_2150097, EPI_ISL_2150120, EPI_ISL_2150156, EPI_ISL_2150157, EPI_ISL_2150161, EPI_ISL_2150177, EPI_ISL_2150272, EPI_ISL_2150398, EPI_ISL_2150476, EPI_ISL_2150515, EPI_ISL_2150595, EPI_ISL_2150598, EPI_ISL_2150658, EPI_ISL_2150694, EPI_ISL_2150793, EPI_ISL_2150838, EPI_ISL_2150840, EPI_ISL_2150841 | | | |
| see above | Aegis Sciences Corporation | Centers for Disease Control and Prevention Division of Viral Diseases, Pathogen Discovery | Dakota Howard, Dhwani Batra, Peter W. Cook, Kara Moser, Adrian Paskey, Jason Caravas, Benjamin Rambo-Martin, Shatavia Morrison, Christopher Gulvick, Scott Sammons, Yvette Unoarumhi, Darlene Wagner, Matthew Schmerer, Cyndi Clark, Patrick Campbell, Rob Case, Vikramsinha Ghorpade, Holly Houdeshell, Ola Kvalvaag, Dillon Nall, Ethan Sanders, Alec Vest, Shaun Westlund, Matthew Hardison, Clinton R. Paden, Duncan MacCannell |

| | | | |
|---|---|---|---|
| EPI_ISL_2151367 | DOHMH Crown Heights | New York City Public Health Laboratory | Jade Wang, et al. |
| EPI_ISL_2151368 | DOHMH Central Harlem | New York City Public Health Laboratory | Jade Wang, et al. |
| EPI_ISL_2151369, EPI_ISL_2151370 | DOHMH Jamaica | New York City Public Health Laboratory | Jade Wang, et al. |
| EPI_ISL_2151371, EPI_ISL_2151372, EPI_ISL_2151373, EPI_ISL_2151374 | DOHMH Riverside | New York City Public Health Laboratory | Jade Wang, et al. |
| EPI_ISL_2151375, EPI_ISL_2151376, EPI_ISL_2151377 | DOHMH Crown Heights | New York City Public Health Laboratory | Jade Wang, et al. |
| EPI_ISL_2151378 | Department of Homeless Services | New York City Public Health Laboratory | Jade Wang, et al. |
| EPI_ISL_2151379, EPI_ISL_2151380 | DOHMH Crown Heights | New York City Public Health Laboratory | Jade Wang, et al. |
| EPI_ISL_2151381 | DOHMH Morrisania | New York City Public Health Laboratory | Jade Wang, et al. |
| EPI_ISL_2151382 | DOHMH Central Harlem | New York City Public Health Laboratory | Jade Wang, et al. |
| EPI_ISL_2151383 | DOHMH Chelsea | New York City Public Health Laboratory | Jade Wang, et al. |
| EPI_ISL_2151384 | DOHMH Fort Greene | New York City Public Health Laboratory | Jade Wang, et al. |
| EPI_ISL_2151385 | DOHMH Crown Heights | New York City Public Health Laboratory | Jade Wang, et al. |
| EPI_ISL_2151386 | DOHMH PHL | New York City Public Health Laboratory | Jade Wang, et al. |
| EPI_ISL_2151387 | DOHMH Central Harlem | New York City Public Health Laboratory | Jade Wang, et al. |
| EPI_ISL_2151388 | DOHMH Riverside | New York City Public Health Laboratory | Jade Wang, et al. |
| EPI_ISL_2151389 | DOHMH Jamaica | New York City Public Health Laboratory | Jade Wang, et al. |
| EPI_ISL_2151390 | DOHMH Chelsea | New York City Public Health Laboratory | Jade Wang, et al. |
| EPI_ISL_2151391 | DOHMH Central Harlem | New York City Public Health Laboratory | Jade Wang, et al. |
| EPI_ISL_2151392 | DOHMH Jamaica | New York City Public Health Laboratory | Jade Wang, et al. |
| EPI_ISL_2151393, EPI_ISL_2151394 | DOHMH PHL | New York City Public Health Laboratory | Jade Wang, et al. |
| EPI_ISL_2151395, EPI_ISL_2151396, EPI_ISL_2151397, EPI_ISL_2151398, EPI_ISL_2151399, EPI_ISL_2151400, EPI_ISL_2151401 | HealthQuest Esoterics | New York City Public Health Laboratory | Jade Wang, et al. |
| EPI_ISL_2151402, EPI_ISL_2151403 | Department of Homeless Services | New York City Public Health Laboratory | Jade Wang, et al. |
| EPI_ISL_2151404, EPI_ISL_2151405, EPI_ISL_2151406, EPI_ISL_2151407, EPI_ISL_2151408, EPI_ISL_2151409, EPI_ISL_2151410, EPI_ISL_2151411 | OCME Office Of Chief Medical Examiner | New York City Public Health Laboratory | Jade Wang, et al. |
| EPI_ISL_2151412, EPI_ISL_2151413 | DOHMH Central Harlem | New York City Public Health Laboratory | Jade Wang, et al. |
| EPI_ISL_2151414 | DOHMH Crown Heights | New York City Public Health Laboratory | Jade Wang, et al. |
| EPI_ISL_2151415 | DOHMH Jamaica | New York City Public Health Laboratory | Jade Wang, et al. |
| EPI_ISL_2151416, EPI_ISL_2151417, EPI_ISL_2151418 | DOHMH Fort Greene | New York City Public Health Laboratory | Jade Wang, et al. |
| EPI_ISL_2151419 | DOHMH Jamaica | New York City Public Health Laboratory | Jade Wang, et al. |
| EPI_ISL_2151420 | SUNY Downstate Medical Center | New York City Public Health Laboratory | Jade Wang, et al. |
| EPI_ISL_2151436, EPI_ISL_2151440 | Columbia University | Pathogen Discovery, Respiratory Viruses Branch, Division of Viral Diseases, Centers for Disease Control and Prevention | Yan Li, Jing Zhang, Anna Kelleher, Ying Tao, Brian Lynch, Krista Queen, Anna Uehara, Peter Cook, Han Jia Justin Ng, Rachel Marine, Clinton R. Paden, Dhwani Batra, Haibin Wang, Tara Coalter, Jasmine Padilla, Morgan Davis, Mili Sheth, Sarah Nobles, Mark Burroughs, Justin Lee, Adam Retchless, Suxiang Tong |
| EPI_ISL_2158971 | Aegis Sciences Corporation | Centers for Disease Control and Prevention Division of Viral Diseases, Pathogen Discovery | Dakota Howard, Dhwani Batra, Peter W. Cook, Kara Moser, Adrian Paskey, Jason Caravas, Benjamin Rambo-Martin, Shatavia Morrison, Christopher Gulvick, Scott Sammons, Yvette Unoarumhi, Darlene Wagner, Matthew Schmerer, Cyndi Clark, Patrick Campbell, Rob Case, Vikramsinha Ghorpade, Holly Houdeshell, Ola Kvalvaag, Dillon Nall, Ethan Sanders, Alec Vest, Shaun Westlund, Matthew Hardison, Clinton R. Paden, Duncan MacCannell |
| EPI_ISL_2158984 | Yale Clinical Virology Lab | Yale Center for Genomic Analysis | Shrikant Mane, Kaya Bilguvar, Curt Scharfe, Irina Tikhonova, Brooke Sullivan |
| EPI_ISL_2159089, EPI_ISL_2159170, EPI_ISL_2159187, EPI_ISL_2159198, EPI_ISL_2159201, EPI_ISL_2159213, EPI_ISL_2159242, EPI_ISL_2159253, EPI_ISL_2159429, EPI_ISL_2159457 | Aegis Sciences Corporation | Centers for Disease Control and Prevention Division of Viral Diseases, Pathogen Discovery | Dakota Howard, Dhwani Batra, Peter W. Cook, Kara Moser, Adrian Paskey, Jason Caravas, Benjamin Rambo-Martin, Shatavia Morrison, Christopher Gulvick, Scott Sammons, Yvette Unoarumhi, Darlene Wagner, Matthew Schmerer, Cyndi Clark, Patrick Campbell, Rob Case, Vikramsinha Ghorpade, Holly Houdeshell, Ola Kvalvaag, Dillon Nall, Ethan Sanders, Alec Vest, Shaun Westlund, Matthew Hardison, Clinton R. Paden, Duncan MacCannell |
| EPI_ISL_2159854, EPI_ISL_2160675 | Helix/Illumina | Centers for Disease Control and Prevention Division of Viral Diseases, Pathogen Discovery | Dakota Howard, Dhwani Batra, Peter W. Cook, Kara Moser, Adrian Paskey, Jason Caravas, Benjamin Rambo-Martin, Shatavia Morrison, Christopher Gulvick, Scott Sammons, Yvette Unoarumhi, Darlene Wagner, Matthew Schmerer, Eileen de Feo, Jan Antico, Christine Tran, Matthew Tolentino, Shannon Wickline, Kim Gietzen, Brad Sickler, Jingtao Liu, Eric Allen, Phil Febbo, Nicole L. Washington, Simon White, Geraint Levan, Kelly Schiabor Barrett, Elizabeth Cirulli, Alexandre Bolze, Ary Ascencio, Charlotte Rivera-Garcia, Ryan Cho, Jason Nguyen, Sherry Wang, Jimmy Ramirez, Tyler Cassens, Efren Sandoval, Magnus Isaksson, William Lee, David Becker, Marc Laurent, James Lu, Clinton R. Paden, Duncan MacCannell |
| EPI_ISL_2161621, EPI_ISL_2161622, EPI_ISL_2161623, EPI_ISL_2161624, EPI_ISL_2161625, EPI_ISL_2161626, EPI_ISL_2161627, EPI_ISL_2161628, EPI_ISL_2161629, EPI_ISL_2161630, EPI_ISL_2161632, EPI_ISL_2161633, EPI_ISL_2161634, EPI_ISL_2161635, EPI_ISL_2161636, EPI_ISL_2161637, EPI_ISL_2161638, EPI_ISL_2161639, EPI_ISL_2161640, EPI_ISL_2161641, EPI_ISL_2161642, EPI_ISL_2161643, EPI_ISL_2161644, EPI_ISL_2161645, EPI_ISL_2161646, EPI_ISL_2161647, EPI_ISL_2161648, EPI_ISL_2161649, EPI_ISL_2161650, EPI_ISL_2161651, EPI_ISL_2161652, EPI_ISL_2161654, EPI_ISL_2161655, EPI_ISL_2161656, EPI_ISL_2161657, EPI_ISL_2161658, EPI_ISL_2161659, EPI_ISL_2161660, EPI_ISL_2161661, EPI_ISL_2161662, EPI_ISL_2161663, EPI_ISL_2161664, EPI_ISL_2161665, EPI_ISL_2161666, EPI_ISL_2161667, EPI_ISL_2161668, EPI_ISL_2161669, EPI_ISL_2161670, EPI_ISL_2161671, EPI_ISL_2161672, EPI_ISL_2161673, EPI_ISL_2161674, EPI_ISL_2161675, EPI_ISL_2161676, EPI_ISL_2161678, EPI_ISL_2161679, EPI_ISL_2161680, EPI_ISL_2161681, EPI_ISL_2161682, EPI_ISL_2161683, EPI_ISL_2161684, EPI_ISL_2161685, EPI_ISL_2161686, EPI_ISL_2161687, EPI_ISL_2161688, EPI_ISL_2161689, EPI_ISL_2161690, EPI_ISL_2161691, EPI_ISL_2161692, EPI_ISL_2161693, EPI_ISL_2161694, EPI_ISL_2161695, EPI_ISL_2161696, EPI_ISL_2161697, EPI_ISL_2161698, EPI_ISL_2161699, EPI_ISL_2161700, EPI_ISL_2161702, EPI_ISL_2161703, EPI_ISL_2161704, EPI_ISL_2161705, EPI_ISL_2161706, EPI_ISL_2161707, EPI_ISL_2161708, EPI_ISL_2161709, EPI_ISL_2161710, EPI_ISL_2161711, EPI_ISL_2161712, EPI_ISL_2161713, EPI_ISL_2161714, EPI_ISL_2161715, EPI_ISL_2161716, EPI_ISL_2161717, EPI_ISL_2161718, EPI_ISL_2161719, EPI_ISL_2161720, EPI_ISL_2161721, EPI_ISL_2161722, EPI_ISL_2161723, EPI_ISL_2161724, EPI_ISL_2161725, EPI_ISL_2161726, EPI_ISL_2161727, EPI_ISL_2161728, EPI_ISL_2161729, EPI_ISL_2161730, EPI_ISL_2161731, EPI_ISL_2161732, EPI_ISL_2161733, EPI_ISL_2161734, EPI_ISL_2161735, EPI_ISL_2161736, EPI_ISL_2161737, EPI_ISL_2161738, EPI_ISL_2161739, EPI_ISL_2161740, EPI_ISL_2161741, EPI_ISL_2161742, EPI_ISL_2161743, EPI_ISL_2161744, EPI_ISL_2161745, EPI_ISL_2161746, EPI_ISL_2161747, EPI_ISL_2161748, EPI_ISL_2161749, EPI_ISL_2161750, EPI_ISL_2161751, EPI_ISL_2161752, EPI_ISL_2161753, EPI_ISL_2161754, EPI_ISL_2161755, EPI_ISL_2161756, EPI_ISL_2161757, EPI_ISL_2161758, EPI_ISL_2161759, EPI_ISL_2161760, EPI_ISL_2161761, EPI_ISL_2161762, EPI_ISL_2161763, EPI_ISL_2161764, EPI_ISL_2161765, EPI_ISL_2161767, EPI_ISL_2161768, EPI_ISL_2161769, EPI_ISL_2161770, EPI_ISL_2161771, EPI_ISL_2161772, EPI_ISL_2161773, EPI_ISL_2161774, EPI_ISL_2161775, EPI_ISL_2161776, EPI_ISL_2161779, EPI_ISL_2161780, EPI_ISL_2161781, EPI_ISL_2161782 | | | |
| see above | NYU Langone Health | Departments of Pathology and Medicine, New York University School of Medicine | Adriana Heguy, Dacia Dimartino, Emily Guzman, Christian Marier, Peter Meyn, Sitharam Ramaswami, Gael Westby, Paul Zappile, Yutong Zhang, Paolo Cotzia, Guiqing Wang |
| EPI_ISL_2179166 | HealthQuest Esoterics | New York City Public Health Laboratory | Jade Wang, et al. |
| EPI_ISL_2179167, EPI_ISL_2179168 | DOHMH Morrisania | New York City Public Health Laboratory | Jade Wang, et al. |

| | | | |
|---|---|---|---|
| EPI_ISL_2179169 | DOHMH Corona | New York City Public Health Laboratory | Jade Wang, et al. |
| EPI_ISL_2179170, EPI_ISL_2179171 | DOHMH PHL | New York City Public Health Laboratory | Jade Wang, et al. |
| EPI_ISL_2179172, EPI_ISL_2179173 | DOHMH Fort Greene | New York City Public Health Laboratory | Jade Wang, et al. |
| EPI_ISL_2179174 | DOHMH Crown Heights | New York City Public Health Laboratory | Jade Wang, et al. |
| EPI_ISL_2179175 | DOHMH Riverside | New York City Public Health Laboratory | Jade Wang, et al. |
| EPI_ISL_2179176 | DOHMH Morrisania | New York City Public Health Laboratory | Jade Wang, et al. |
| EPI_ISL_2179177, EPI_ISL_2179178 | DOHMH Central Harlem | New York City Public Health Laboratory | Jade Wang, et al. |
| EPI_ISL_2179179 | DOHMH PHL | New York City Public Health Laboratory | Jade Wang, et al. |
| EPI_ISL_2179180 | DOHMH Jamaica | New York City Public Health Laboratory | Jade Wang, et al. |
| EPI_ISL_2179181, EPI_ISL_2179182 | DOHMH Corona | New York City Public Health Laboratory | Jade Wang, et al. |
| EPI_ISL_2179183, EPI_ISL_2179184 | DOHMH Morrisania | New York City Public Health Laboratory | Jade Wang, et al. |
| EPI_ISL_2179185 | DOHMH Crown Heights | New York City Public Health Laboratory | Jade Wang, et al. |
| EPI_ISL_2179186, EPI_ISL_2179187 | DOHMH Jamaica | New York City Public Health Laboratory | Jade Wang, et al. |
| EPI_ISL_2179188, EPI_ISL_2179189 | Department of Homeless Services | New York City Public Health Laboratory | Jade Wang, et al. |
| EPI_ISL_2179190 | OCME Office Of Chief Medical Examiner | New York City Public Health Laboratory | Jade Wang, et al. |
| EPI_ISL_2180576, EPI_ISL_2180579, EPI_ISL_2180665, EPI_ISL_2180742, EPI_ISL_2180764, EPI_ISL_2180772, EPI_ISL_2180797, EPI_ISL_2180824, EPI_ISL_2180907, EPI_ISL_2181043, EPI_ISL_2181085, EPI_ISL_2181120, EPI_ISL_2181137, EPI_ISL_2181201, EPI_ISL_2181216, EPI_ISL_2181295, EPI_ISL_2181325, EPI_ISL_2181326, EPI_ISL_2181327, EPI_ISL_2181329, EPI_ISL_2181334, EPI_ISL_2181340, EPI_ISL_2181360, EPI_ISL_2181365, EPI_ISL_2181390, EPI_ISL_2181391, EPI_ISL_2181392, EPI_ISL_2181502, EPI_ISL_2181514, EPI_ISL_2181574, EPI_ISL_2181575, EPI_ISL_2181601, EPI_ISL_2181640, EPI_ISL_2181652, EPI_ISL_2181653 | | | |
| see above | Aegis Sciences Corporation | Centers for Disease Control and Prevention Division of Viral Diseases, Pathogen Discovery | Dakota Howard, Dhwani Batra, Peter W. Cook, Kara Moser, Adrian Paskey, Jason Caravas, Benjamin Rambo-Martin, Shatavia Morrison, Christopher Gulvick, Scott Sammons, Yvette Unoarumhi, Darlene Wagner, Matthew Schmerer, Cyndi Clark, Patrick Campbell, Rob Case, Vikramsinha Ghorpade, Holly Houdeshell, Ola Kvalvaag, Dillon Nall, Ethan Sanders, Alec Vest, Shaun Westlund, Matthew Hardison, Clinton R. Paden, Duncan MacCannell |
| EPI_ISL_2181681, EPI_ISL_2181682, EPI_ISL_2181685, EPI_ISL_2181686, EPI_ISL_2181687, EPI_ISL_2181749, EPI_ISL_2181750, EPI_ISL_2181814, EPI_ISL_2181819, EPI_ISL_2181822, EPI_ISL_2181825, EPI_ISL_2181828, EPI_ISL_2181831, EPI_ISL_2181865, EPI_ISL_2181917, EPI_ISL_2182281, EPI_ISL_2182284, EPI_ISL_2182286, EPI_ISL_2182289, EPI_ISL_2182292, EPI_ISL_2182294, EPI_ISL_2182297, EPI_ISL_2182302, EPI_ISL_2182308, EPI_ISL_2182310, EPI_ISL_2182357, EPI_ISL_2182392, EPI_ISL_2182496, EPI_ISL_2182509, EPI_ISL_2182511, EPI_ISL_2182517, EPI_ISL_2182520, EPI_ISL_2182522, EPI_ISL_2182525, EPI_ISL_2182535, EPI_ISL_2182541, EPI_ISL_2182544, EPI_ISL_2182546, EPI_ISL_2182549, EPI_ISL_2182552, EPI_ISL_2182555, EPI_ISL_2182557, EPI_ISL_2182574, EPI_ISL_2182705, EPI_ISL_2182725, EPI_ISL_2182728, EPI_ISL_2182731, EPI_ISL_2182736, EPI_ISL_2182739, EPI_ISL_2182741, EPI_ISL_2182744, EPI_ISL_2182745, EPI_ISL_2182747, EPI_ISL_2182749, EPI_ISL_2182752, EPI_ISL_2182941, EPI_ISL_2182944, EPI_ISL_2182946, EPI_ISL_2182965, EPI_ISL_2182968, EPI_ISL_2183091, EPI_ISL_2183105, EPI_ISL_2183108, EPI_ISL_2183110, EPI_ISL_2183693, EPI_ISL_2183695, EPI_ISL_2183698, EPI_ISL_2183700, EPI_ISL_2183705, EPI_ISL_2183707, EPI_ISL_2183710, EPI_ISL_2183731, EPI_ISL_2183734, EPI_ISL_2183735, EPI_ISL_2183888, EPI_ISL_2183938, EPI_ISL_2183955, EPI_ISL_2184200, EPI_ISL_2184201, EPI_ISL_2184224, EPI_ISL_2184232, EPI_ISL_2184234, EPI_ISL_2184244, EPI_ISL_2184247, EPI_ISL_2184248, EPI_ISL_2184290, EPI_ISL_2184291, EPI_ISL_2184296, EPI_ISL_2184297, EPI_ISL_2184301, EPI_ISL_2184302, EPI_ISL_2184310, EPI_ISL_2184311, EPI_ISL_2184329, EPI_ISL_2184385, EPI_ISL_2184480, EPI_ISL_2184500, EPI_ISL_2184609, EPI_ISL_2184674, EPI_ISL_2184681, EPI_ISL_2184794, EPI_ISL_2184835, EPI_ISL_2184836, EPI_ISL_2184850 | | | |
| see above | Laboratory Corporation of America | Centers for Disease Control and Prevention Division of Viral Diseases, Pathogen Discovery | Dakota Howard, Dhwani Batra, Peter W. Cook, Kara Moser, Adrian Paskey, Jason Caravas, Benjamin Rambo-Martin, Shatavia Morrison, Christopher Gulvick, Scott Sammons, Yvette Unoarumhi, Darlene Wagner, Matthew Schmerer, Minoo Agarwal, Eyad Almasri, Debbie Boles, Ayla Burns, Nuthawin Charoensri, Oren Cohen, Susan Countryman, Mary Ann Cristobal, Bobbi Croy, Suzanne Dale, Hrushikesh Deshmukh, Amanda Douglas, Vincent Drouillon, Marcia Eisenberg, Howard Engler, Rama Ghatti, Prashant Gupta, Susan Hicks, Jake Humphrey, Lax Iyer, Manoj Jain, Mohan Kolli, Brian Krueger, Tim Kuphal, Stanley Letovsky, Michael Levandoski, Craig Lukasik, Jonathan Meltzer, Brian Norvell, Mindy Nye, Scott Parker, Christos Petropoulos, John Pruitt, Steven Ragan, Scott Ryan, Mike Sapeta, Jana Schroth, Suresh Babu Selvaraju, Goran Stevovic, Amanda Suchanek, Andrea Throop, Lyndon Tilson, Thomas Urban, Joe Voshell, Kimberly Wagner, Jonathan Williams, Mary Williamson, Qian Zeng, Tricia Zwiefelhofer, Clinton R. Paden, Duncan MacCannell |
| EPI_ISL_2185809, EPI_ISL_2185920, EPI_ISL_2186066, EPI_ISL_2186121, EPI_ISL_2186197, EPI_ISL_2186211, EPI_ISL_2186248, EPI_ISL_2186249 | Aegis Sciences Corporation | Centers for Disease Control and Prevention Division of Viral Diseases, Pathogen Discovery | Dakota Howard, Dhwani Batra, Peter W. Cook, Kara Moser, Adrian Paskey, Jason Caravas, Benjamin Rambo-Martin, Shatavia Morrison, Christopher Gulvick, Scott Sammons, Yvette Unoarumhi, Darlene Wagner, Matthew Schmerer, Cyndi Clark, Patrick Campbell, Rob Case, Vikramsinha Ghorpade, Holly Houdeshell, Ola Kvalvaag, Dillon Nall, Ethan Sanders, Alec Vest, Shaun Westlund, Matthew Hardison, Clinton R. Paden, Duncan MacCannell |
| EPI_ISL_2187244 | Quest Diagnostics Incorporated | Centers for Disease Control and Prevention Division of Viral Diseases, Pathogen Discovery | Dakota Howard, Dhwani Batra, Peter W. Cook, Kara Moser, Adrian Paskey, Jason Caravas, Benjamin Rambo-Martin, Shatavia Morrison, Christopher Gulvick, Scott Sammons, Yvette Unoarumhi, Darlene Wagner, Matthew Schmerer, S. H. Rosenthal, A. Gerasimova, R. M. Kagan, B. Anderson, M. Hua, Y. Liu, L.E. Bernstein, K.E. Livingston, A. Perez, I. A. Shlyakhter, R. V. Rolando, R. Owen, P. Tanpaiboon, F. Lacbawan, Clinton R. Paden, Duncan MacCannell |
| EPI_ISL_2187370, EPI_ISL_2187445, EPI_ISL_2187452, EPI_ISL_2187491, EPI_ISL_2187508, EPI_ISL_2187591, EPI_ISL_2187596, EPI_ISL_2187607 | Aegis Sciences Corporation | Centers for Disease Control and Prevention Division of Viral Diseases, Pathogen Discovery | Dakota Howard, Dhwani Batra, Peter W. Cook, Kara Moser, Adrian Paskey, Jason Caravas, Benjamin Rambo-Martin, Shatavia Morrison, Christopher Gulvick, Scott Sammons, Yvette Unoarumhi, Darlene Wagner, Matthew Schmerer, Cyndi Clark, Patrick Campbell, Rob Case, Vikramsinha Ghorpade, Holly Houdeshell, Ola Kvalvaag, Dillon Nall, Ethan Sanders, Alec Vest, Shaun Westlund, Matthew Hardison, Clinton R. Paden, Duncan MacCannell |
| EPI_ISL_2193151, EPI_ISL_2193152, EPI_ISL_2193153 | Broad Institute Clinical Research Sequencing Platform | Infectious Disease Program, Broad Institute of Harvard and MIT | Siddle,K.J., Adams,G., Pearlman,L., Gladden-Young,A., Vicente,G., Blumenstiel,B., DeFelice,M., Lee,M., McGovern,S., Lagerborg,K., Rudy,M., DeRuff,K., Carter,A., Normandin,E., Bauer,M., Reilly,S., Tomkins-Tinch,C., Loreth,C., Chaluvadi,S., Meldrim,J., Granger,B., Lemieux,J.E., Birren,B.W., Sabeti,P.C., Larkin,K., Dodge,S., Lennon,N., Madoff,L., Brown,C., Gallagher,G., Smole,S., Park,D.J., Gabriel,S., and MacInnis,B.L. |
| EPI_ISL_2193762, EPI_ISL_2193763, EPI_ISL_2193764, EPI_ISL_2193765, EPI_ISL_2193766 | Weill Cornell Medicine | New York Genome Center | Michael Zody, Andre Corvelo, Dayna M. Oschwald, Samantha Fennessey, Tom Maniatis, Melissa Cushing, Olivier Elemento, Margaret Elizabeth Ross, Chris Mason, Priya Velu, Hanna Rennert, Arryn Craney, Lars F Westblade |
| EPI_ISL_2201199, EPI_ISL_2201298, EPI_ISL_2201341, EPI_ISL_2201517, EPI_ISL_2201532, EPI_ISL_2201538, EPI_ISL_2201582, EPI_ISL_2201612, EPI_ISL_2201763, EPI_ISL_2201772, EPI_ISL_2201785, EPI_ISL_2201843, EPI_ISL_2201849, EPI_ISL_2201862, EPI_ISL_2201911, EPI_ISL_2201949, EPI_ISL_2201955, EPI_ISL_2202009, EPI_ISL_2202165, EPI_ISL_2202169, EPI_ISL_2202277, EPI_ISL_2202350, EPI_ISL_2202459, EPI_ISL_2202467, EPI_ISL_2202579, EPI_ISL_2202581, EPI_ISL_2202583, EPI_ISL_2202585 | | | |
| see above | Aegis Sciences Corporation | Centers for Disease Control and Prevention Division of Viral Diseases, Pathogen Discovery | Dakota Howard, Dhwani Batra, Peter W. Cook, Kara Moser, Adrian Paskey, Jason Caravas, Benjamin Rambo-Martin, Shatavia Morrison, Christopher Gulvick, Scott Sammons, Yvette Unoarumhi, Darlene Wagner, Matthew Schmerer, Cyndi Clark, Patrick Campbell, Rob Case, Vikramsinha Ghorpade, Holly Houdeshell, Ola Kvalvaag, Dillon Nall, Ethan Sanders, Alec Vest, Shaun Westlund, Matthew Hardison, Clinton R. Paden, Duncan MacCannell |
| EPI_ISL_2202717, EPI_ISL_2202720, EPI_ISL_2202722, EPI_ISL_2202724, EPI_ISL_2202726, EPI_ISL_2202728, EPI_ISL_2202731, EPI_ISL_2202734, EPI_ISL_2202736, EPI_ISL_2202738, EPI_ISL_2202740, EPI_ISL_2202742, EPI_ISL_2202744, EPI_ISL_2202746, EPI_ISL_2202748, EPI_ISL_2202751, EPI_ISL_2202752, EPI_ISL_2202754 | | | |
| see above | NYU Langone Health | Departments of Pathology and Medicine, New York University School of Medicine | Adriana Heguy, Dacia Dimartino, Emily Guzman, Christian Marier, Peter Meyn, Sitharam Ramaswami, Gael Westby, Paul Zappile, Yutong Zhang, Paolo Cotzia, Guiqing Wang |
| EPI_ISL_2202756 | Aegis Sciences Corporation | Centers for Disease Control and Prevention Division of Viral Diseases, Pathogen Discovery | Dakota Howard, Dhwani Batra, Peter W. Cook, Kara Moser, Adrian Paskey, Jason Caravas, Benjamin Rambo-Martin, Shatavia Morrison, Christopher Gulvick, Scott Sammons, Yvette Unoarumhi, Darlene Wagner, Matthew Schmerer, Cyndi Clark, Patrick Campbell, Rob Case, Vikramsinha Ghorpade, Holly Houdeshell, Ola Kvalvaag, Dillon Nall, Ethan Sanders, Alec Vest, Shaun Westlund, Matthew Hardison, Clinton R. Paden, Duncan MacCannell |
| EPI_ISL_2202759, EPI_ISL_2202764, EPI_ISL_2202766, EPI_ISL_2202768, EPI_ISL_2202770, EPI_ISL_2202771, EPI_ISL_2202774, EPI_ISL_2202775, EPI_ISL_2202777, EPI_ISL_2202780, EPI_ISL_2202781, EPI_ISL_2202783, EPI_ISL_2202785, EPI_ISL_2202787, EPI_ISL_2202790, EPI_ISL_2202791, EPI_ISL_2202794, EPI_ISL_2202795, EPI_ISL_2202797, EPI_ISL_2202800, EPI_ISL_2202801, EPI_ISL_2202803, EPI_ISL_2202804, EPI_ISL_2202807, EPI_ISL_2202808, EPI_ISL_2202811 | | | |
| see above | NYU Langone Health | Departments of Pathology and Medicine, New York University School of Medicine | Adriana Heguy, Dacia Dimartino, Emily Guzman, Christian Marier, Peter Meyn, Sitharam Ramaswami, Gael Westby, Paul Zappile, Yutong Zhang, Paolo Cotzia, Guiqing Wang |
| EPI_ISL_2202812, EPI_ISL_2202814, EPI_ISL_2203113, EPI_ISL_2203116, EPI_ISL_2203120, EPI_ISL_2203121, | Aegis Sciences Corporation | Centers for Disease Control and Prevention Division of Viral Diseases, Pathogen Discovery | Dakota Howard, Dhwani Batra, Peter W. Cook, Kara Moser, Adrian Paskey, Jason Caravas, Benjamin Rambo-Martin, Shatavia Morrison, Christopher Gulvick, Scott Sammons, Yvette Unoarumhi, Darlene Wagner, Matthew Schmerer, Cyndi Clark, Patrick Campbell, Rob Case, Vikramsinha Ghorpade, Holly Houdeshell, Ola Kvalvaag, Dillon Nall, Ethan Sanders, Alec Vest, Shaun Westlund, Matthew Hardison, Clinton R. Paden, Duncan MacCannell |

| | | | |
|---|---|---|---|
| EPI_ISL_2203126 | | | |
| EPI_ISL_2203649, EPI_ISL_2203654, EPI_ISL_2203682, EPI_ISL_2203691, EPI_ISL_2203766, EPI_ISL_2203798, EPI_ISL_2203814, EPI_ISL_2203861, EPI_ISL_2203908, EPI_ISL_2203915, EPI_ISL_2203922, EPI_ISL_2203933, EPI_ISL_2203943, EPI_ISL_2203950, EPI_ISL_2203953, EPI_ISL_2203980, EPI_ISL_2203996, EPI_ISL_2204003, EPI_ISL_2204010, EPI_ISL_2204013, EPI_ISL_2204038, EPI_ISL_2204054, EPI_ISL_2204066, EPI_ISL_2204070, EPI_ISL_2204136, EPI_ISL_2204139, EPI_ISL_2204157, EPI_ISL_2204159, EPI_ISL_2204185 | | | |
| see above | Quest Diagnostics Incorporated | Centers for Disease Control and Prevention Division of Viral Diseases, Pathogen Discovery | Dakota Howard, Dhwani Batra, Peter W. Cook, Kara Moser, Adrian Paskey, Jason Caravas, Benjamin Rambo-Martin, Shatavia Morrison, Christopher Gulvick, Scott Sammons, Yvette Unoarumhi, Darlene Wagner, Matthew Schmerer, S. H. Rosenthal, A. Gerasimova, R. M. Kagan, B. Anderson, M. Hua, Y. Liu, L.E. Bernstein, K.E. Livingston, A. Perez, I. A. Shlyakhter, R. V. Rolando, R. Owen, P. Tanpaiboon, F. Lacbawan, Clinton R. Paden, Duncan MacCannell |
| EPI_ISL_2204599 | Aegis Sciences Corporation | Centers for Disease Control and Prevention Division of Viral Diseases, Pathogen Discovery | Dakota Howard, Dhwani Batra, Peter W. Cook, Kara Moser, Adrian Paskey, Jason Caravas, Benjamin Rambo-Martin, Shatavia Morrison, Christopher Gulvick, Scott Sammons, Yvette Unoarumhi, Darlene Wagner, Matthew Schmerer, Cyndi Clark, Patrick Campbell, Rob Case, Vikramsinha Ghorpade, Holly Houdeshell, Ola Kvalvaag, Dillon Nall, Ethan Sanders, Alec Vest, Shaun Westlund, Matthew Hardison, Clinton R. Paden, Duncan MacCannell |
| EPI_ISL_2229274 | "NYSDOH Wadsworth Center, Virology Lab" | Centers for Disease Control and Prevention Division of Viral Diseases, Pathogen Discovery | Mili Sheth, Sarah Nobles, Jasmine Padilla, Mark Burroughs, Shoshona Le, Katie Dillon, Peter Cook, Clinton R. Paden, Dhwani Batra, Krista Queen, Kristen Knipe, Dakota Howard, Yvette Unoarumhi, Darlene Wagner, Matthew Schmerer, Ben L. Rambo-Martin, Kristine Lacek, Sam Shepard, Alison Laufer Halpin, Dave Wentworth, Vivien Dugan, Suxiang Tong, Justin Lee |
| EPI_ISL_2229539, EPI_ISL_2229540, EPI_ISL_2229541, EPI_ISL_2229542, EPI_ISL_2229543, EPI_ISL_2229544, EPI_ISL_2229545, EPI_ISL_2229546, EPI_ISL_2229547, EPI_ISL_2229548, EPI_ISL_2229549, EPI_ISL_2229550, EPI_ISL_2229551, EPI_ISL_2229552, EPI_ISL_2229553, EPI_ISL_2229554, EPI_ISL_2229555, EPI_ISL_2229556, EPI_ISL_2229557, EPI_ISL_2229558, EPI_ISL_2229559, EPI_ISL_2229560, EPI_ISL_2229561, EPI_ISL_2229562, EPI_ISL_2229563, EPI_ISL_2229564, EPI_ISL_2229565, EPI_ISL_2229566, EPI_ISL_2229567, EPI_ISL_2229568, EPI_ISL_2229569, EPI_ISL_2229570, EPI_ISL_2229571, EPI_ISL_2229572, EPI_ISL_2229573, EPI_ISL_2229574, EPI_ISL_2229575, EPI_ISL_2229576, EPI_ISL_2229577, EPI_ISL_2229578, EPI_ISL_2229579, EPI_ISL_2229580, EPI_ISL_2229581, EPI_ISL_2229582, EPI_ISL_2229583, EPI_ISL_2229584, EPI_ISL_2229585, EPI_ISL_2229586, EPI_ISL_2229587, EPI_ISL_2229588, EPI_ISL_2229589, EPI_ISL_2229590, EPI_ISL_2229591, EPI_ISL_2229592, EPI_ISL_2229593, EPI_ISL_2229594, EPI_ISL_2229595, EPI_ISL_2229596, EPI_ISL_2229597, EPI_ISL_2229598, EPI_ISL_2229599, EPI_ISL_2229600, EPI_ISL_2229601, EPI_ISL_2229602, EPI_ISL_2229603, EPI_ISL_2229604, EPI_ISL_2229605, EPI_ISL_2229606, EPI_ISL_2229607, EPI_ISL_2229608, EPI_ISL_2229609, EPI_ISL_2229610, EPI_ISL_2229611, EPI_ISL_2229612, EPI_ISL_2229613, EPI_ISL_2229614, EPI_ISL_2229615, EPI_ISL_2229616, EPI_ISL_2229617, EPI_ISL_2229618, EPI_ISL_2229619, EPI_ISL_2229620, EPI_ISL_2229621, EPI_ISL_2229622, EPI_ISL_2229623, EPI_ISL_2229624, EPI_ISL_2229625, EPI_ISL_2229626, EPI_ISL_2229627, EPI_ISL_2229628, EPI_ISL_2229629, EPI_ISL_2229630, EPI_ISL_2229631, EPI_ISL_2229632, EPI_ISL_2229633, EPI_ISL_2229634 | | | |
| see above | Erie County Public Health (ECPHL) | University at Buffalo Genomics and Bioinformatics Core | Jonathan Bard, Natalie Lamb, Alyssa Pohlman, Brandon Marzullo, Amanda Boccolucci, Norma Nowak, Donald Yergeau, Jennifer Surtees |
| EPI_ISL_2229635, EPI_ISL_2229636, EPI_ISL_2229637, EPI_ISL_2229638, EPI_ISL_2229639, EPI_ISL_2229640, EPI_ISL_2229641, EPI_ISL_2229642, EPI_ISL_2229643, EPI_ISL_2229644, EPI_ISL_2229645, EPI_ISL_2229646, EPI_ISL_2229647, EPI_ISL_2229648, EPI_ISL_2229649, EPI_ISL_2229650, EPI_ISL_2229651, EPI_ISL_2229652, EPI_ISL_2229653, EPI_ISL_2229654, EPI_ISL_2229655, EPI_ISL_2229656, EPI_ISL_2229657, EPI_ISL_2229658, EPI_ISL_2229659, EPI_ISL_2229660, EPI_ISL_2229661, EPI_ISL_2229662, EPI_ISL_2229663, EPI_ISL_2229664, EPI_ISL_2229665, EPI_ISL_2229666, EPI_ISL_2229667, EPI_ISL_2229668, EPI_ISL_2229669, EPI_ISL_2229670, EPI_ISL_2229671, EPI_ISL_2229672, EPI_ISL_2229673, EPI_ISL_2229674, EPI_ISL_2229675, EPI_ISL_2229676, EPI_ISL_2229677, EPI_ISL_2229678, EPI_ISL_2229679, EPI_ISL_2229680, EPI_ISL_2229681, EPI_ISL_2229682, EPI_ISL_2229683, EPI_ISL_2229684, EPI_ISL_2229685, EPI_ISL_2229686, EPI_ISL_2229687, EPI_ISL_2229688, EPI_ISL_2229689, EPI_ISL_2229690, EPI_ISL_2229691, EPI_ISL_2229692, EPI_ISL_2229693, EPI_ISL_2229694, EPI_ISL_2229695, EPI_ISL_2229696, EPI_ISL_2229697, EPI_ISL_2229698, EPI_ISL_2229699, EPI_ISL_2229700, EPI_ISL_2229701, EPI_ISL_2229702, EPI_ISL_2229703, EPI_ISL_2229704, EPI_ISL_2229705, EPI_ISL_2229706, EPI_ISL_2229707, EPI_ISL_2229708, EPI_ISL_2229709, EPI_ISL_2229710, EPI_ISL_2229711, EPI_ISL_2229712, EPI_ISL_2229713, EPI_ISL_2229714, EPI_ISL_2229715, EPI_ISL_2229716, EPI_ISL_2229717, EPI_ISL_2229718, EPI_ISL_2229719, EPI_ISL_2229720, EPI_ISL_2229721, EPI_ISL_2229722, EPI_ISL_2229723, EPI_ISL_2229724, EPI_ISL_2229725, EPI_ISL_2229726, EPI_ISL_2229727, EPI_ISL_2229728, EPI_ISL_2229729, EPI_ISL_2229730, EPI_ISL_2229731, EPI_ISL_2229732, EPI_ISL_2229733, EPI_ISL_2229734, EPI_ISL_2229735, EPI_ISL_2229736, EPI_ISL_2229737, EPI_ISL_2229738, EPI_ISL_2229739, EPI_ISL_2229740, EPI_ISL_2229741, EPI_ISL_2229742, EPI_ISL_2229743, EPI_ISL_2229744, EPI_ISL_2229745, EPI_ISL_2229746, EPI_ISL_2229747, EPI_ISL_2229748, EPI_ISL_2229749, EPI_ISL_2229750, EPI_ISL_2229751, EPI_ISL_2229752, EPI_ISL_2229753, EPI_ISL_2229754, EPI_ISL_2229755, EPI_ISL_2229756, EPI_ISL_2229757, EPI_ISL_2229758, EPI_ISL_2229759, EPI_ISL_2229760, EPI_ISL_2229761, EPI_ISL_2229762, EPI_ISL_2229763, EPI_ISL_2229764 | | | |
| see above | Kaleida Health Laboratories | University at Buffalo Genomics and Bioinformatics Core | Jonathan Bard, Natalie Lamb, Alyssa Pohlman, Brandon Marzullo, Amanda Boccolucci, Norma Nowak, Donald Yergeau, Jennifer Surtees |
| EPI_ISL_2230036 | The Jackson Laboratory | The Jackson Laboratory | Long J, Bergeron D, Renzette N, Adams M, Omerza G, Kelly K, Li L |
| EPI_ISL_2234908, EPI_ISL_2234909, EPI_ISL_2234910, EPI_ISL_2234911, EPI_ISL_2234912, EPI_ISL_2234913, EPI_ISL_2234914, EPI_ISL_2234915, EPI_ISL_2234916, EPI_ISL_2234917, EPI_ISL_2234918, EPI_ISL_2234919, EPI_ISL_2234920, EPI_ISL_2234921, EPI_ISL_2234922, EPI_ISL_2234923, EPI_ISL_2234924, EPI_ISL_2234925, EPI_ISL_2234926, EPI_ISL_2234927, EPI_ISL_2234928, EPI_ISL_2234929, EPI_ISL_2234930, EPI_ISL_2234931, EPI_ISL_2234932, EPI_ISL_2234933, EPI_ISL_2234934, EPI_ISL_2234935, EPI_ISL_2234936, EPI_ISL_2234937, EPI_ISL_2234938, EPI_ISL_2234939, EPI_ISL_2234940, EPI_ISL_2234941, EPI_ISL_2234942, EPI_ISL_2234943, EPI_ISL_2234944, EPI_ISL_2234945, EPI_ISL_2234946, EPI_ISL_2234947, EPI_ISL_2234948, EPI_ISL_2234949, EPI_ISL_2234950, EPI_ISL_2234951, EPI_ISL_2234952, EPI_ISL_2234953, EPI_ISL_2234954, EPI_ISL_2234955, EPI_ISL_2234956, EPI_ISL_2234957, EPI_ISL_2234958, EPI_ISL_2234959, EPI_ISL_2234960, EPI_ISL_2234961, EPI_ISL_2234962, EPI_ISL_2234963, EPI_ISL_2234964, EPI_ISL_2234965, EPI_ISL_2234966, EPI_ISL_2234967, EPI_ISL_2234968, EPI_ISL_2234969, EPI_ISL_2234970, EPI_ISL_2234971, EPI_ISL_2234972, EPI_ISL_2234973, EPI_ISL_2234974, EPI_ISL_2234975, EPI_ISL_2234976, EPI_ISL_2234977, EPI_ISL_2234978, EPI_ISL_2234979, EPI_ISL_2234980, EPI_ISL_2234981, EPI_ISL_2234982, EPI_ISL_2234983, EPI_ISL_2234984, EPI_ISL_2234985, EPI_ISL_2234986, EPI_ISL_2234987, EPI_ISL_2234988, EPI_ISL_2234989, EPI_ISL_2234990, EPI_ISL_2234991, EPI_ISL_2234992, EPI_ISL_2234993, EPI_ISL_2234994, EPI_ISL_2234995, EPI_ISL_2234996, EPI_ISL_2234997, EPI_ISL_2234998, EPI_ISL_2234999, EPI_ISL_2235000, EPI_ISL_2235001, EPI_ISL_2235002, EPI_ISL_2235003, EPI_ISL_2235004, EPI_ISL_2235005, EPI_ISL_2235006, EPI_ISL_2235007, EPI_ISL_2235008, EPI_ISL_2235009, EPI_ISL_2235010, EPI_ISL_2235011, EPI_ISL_2235012, EPI_ISL_2235013, EPI_ISL_2235014, EPI_ISL_2235015, EPI_ISL_2235016, EPI_ISL_2235017, EPI_ISL_2235018, EPI_ISL_2235019, EPI_ISL_2235020, EPI_ISL_2235021, EPI_ISL_2235022, EPI_ISL_2235023, EPI_ISL_2235024, EPI_ISL_2235025, EPI_ISL_2235026, EPI_ISL_2235027, EPI_ISL_2235028, EPI_ISL_2235029, EPI_ISL_2235030, EPI_ISL_2235031, EPI_ISL_2235032, EPI_ISL_2235033, EPI_ISL_2235034, EPI_ISL_2235035, EPI_ISL_2235036, EPI_ISL_2235037, EPI_ISL_2235038, EPI_ISL_2235039, EPI_ISL_2235040, EPI_ISL_2235041, EPI_ISL_2235042, EPI_ISL_2235043, EPI_ISL_2235044, EPI_ISL_2235045, EPI_ISL_2235046, EPI_ISL_2235047, EPI_ISL_2235048, EPI_ISL_2235049, EPI_ISL_2235050, EPI_ISL_2235051, EPI_ISL_2235052, EPI_ISL_2235053, EPI_ISL_2235054, EPI_ISL_2235055, EPI_ISL_2235056, EPI_ISL_2235057, EPI_ISL_2235058, EPI_ISL_2235059, EPI_ISL_2235060, EPI_ISL_2235061, EPI_ISL_2235062, EPI_ISL_2235063, EPI_ISL_2235064, EPI_ISL_2235065, EPI_ISL_2235066, EPI_ISL_2235067, EPI_ISL_2235068, EPI_ISL_2235069, EPI_ISL_2235070, EPI_ISL_2235071, EPI_ISL_2235072, EPI_ISL_2235073, EPI_ISL_2235074, EPI_ISL_2235075, EPI_ISL_2235076, EPI_ISL_2235077, EPI_ISL_2235078, EPI_ISL_2235079, EPI_ISL_2235080, EPI_ISL_2235081, EPI_ISL_2235082, EPI_ISL_2235083, EPI_ISL_2235084, EPI_ISL_2235085, EPI_ISL_2235086, EPI_ISL_2235087, EPI_ISL_2235088, EPI_ISL_2235089, EPI_ISL_2235090, EPI_ISL_2235091, EPI_ISL_2235092, EPI_ISL_2235093, EPI_ISL_2235094, EPI_ISL_2235095, EPI_ISL_2235096, EPI_ISL_2235097, EPI_ISL_2235098, EPI_ISL_2235099, EPI_ISL_2235100, EPI_ISL_2235101, EPI_ISL_2235102, EPI_ISL_2235103, EPI_ISL_2235104, EPI_ISL_2235105, EPI_ISL_2235106, EPI_ISL_2235107, EPI_ISL_2235108, EPI_ISL_2235109, EPI_ISL_2235110, EPI_ISL_2235111, EPI_ISL_2235112, EPI_ISL_2235113, EPI_ISL_2235114, EPI_ISL_2235115, EPI_ISL_2235116, EPI_ISL_2235117, EPI_ISL_2235118, EPI_ISL_2235119, EPI_ISL_2235120, EPI_ISL_2235121, EPI_ISL_2235122, EPI_ISL_2235123, EPI_ISL_2235124, EPI_ISL_2235125, EPI_ISL_2235126, EPI_ISL_2235127, EPI_ISL_2235128, EPI_ISL_2235129, EPI_ISL_2235130, EPI_ISL_2235131, EPI_ISL_2235132, EPI_ISL_2235133, EPI_ISL_2235134, EPI_ISL_2235135, EPI_ISL_2235136, EPI_ISL_2235137, EPI_ISL_2235138, EPI_ISL_2235139, EPI_ISL_2235140, EPI_ISL_2235141, EPI_ISL_2235142, EPI_ISL_2235143, EPI_ISL_2235144, EPI_ISL_2235145, EPI_ISL_2235146, EPI_ISL_2235147, EPI_ISL_2235148, EPI_ISL_2235149, EPI_ISL_2235150, EPI_ISL_2235151, EPI_ISL_2235152, EPI_ISL_2235153, EPI_ISL_2235154, EPI_ISL_2235155, EPI_ISL_2235156, EPI_ISL_2235157, EPI_ISL_2235158, EPI_ISL_2235159, EPI_ISL_2235160, EPI_ISL_2235161, EPI_ISL_2235162, EPI_ISL_2235163, EPI_ISL_2235164, EPI_ISL_2235165, EPI_ISL_2235166, EPI_ISL_2235167, EPI_ISL_2235168, EPI_ISL_2235169, EPI_ISL_2235170, EPI_ISL_2235171, EPI_ISL_2235172, EPI_ISL_2235173, EPI_ISL_2235174, EPI_ISL_2235175, EPI_ISL_2235176, EPI_ISL_2235177, EPI_ISL_2235178, EPI_ISL_2235179, EPI_ISL_2235180, EPI_ISL_2235181, EPI_ISL_2235182, EPI_ISL_2235183, EPI_ISL_2235184, EPI_ISL_2235185, EPI_ISL_2235186, EPI_ISL_2235187, EPI_ISL_2235188, EPI_ISL_2235189, EPI_ISL_2235190, EPI_ISL_2235191, EPI_ISL_2235192, EPI_ISL_2235193, EPI_ISL_2235194, EPI_ISL_2235195, EPI_ISL_2235196, EPI_ISL_2235197, EPI_ISL_2235198, EPI_ISL_2235199, EPI_ISL_2235200, EPI_ISL_2235201, EPI_ISL_2235202, EPI_ISL_2235203, EPI_ISL_2235204, EPI_ISL_2235205, EPI_ISL_2235206, EPI_ISL_2235207, EPI_ISL_2235208, EPI_ISL_2235209, EPI_ISL_2235210, EPI_ISL_2235211, EPI_ISL_2235212, EPI_ISL_2235213, EPI_ISL_2235214, EPI_ISL_2235215, EPI_ISL_2235216, EPI_ISL_2235217, EPI_ISL_2235218 | | | |
| see above | Pandemic Response Lab - NYC | Pandemic Response Lab, R&D | Henry Lee, Michael Hammerling, Melissa Hopkins, Cybill del Castillo, Shinyoung Clair Kang, William Ward, Pradeep Bugga, Sol Rey, Dylan Law, Katharine Nelson, Haiping Hao, Jon Laurent |
| EPI_ISL_2241646, EPI_ISL_2241702, EPI_ISL_2241721, EPI_ISL_2241767, EPI_ISL_2241773, EPI_ISL_2241879, EPI_ISL_2241920, EPI_ISL_2241955, EPI_ISL_2241975, EPI_ISL_2242046, EPI_ISL_2242047, EPI_ISL_2242098, EPI_ISL_2242099, EPI_ISL_2242123, EPI_ISL_2242127, EPI_ISL_2242175, EPI_ISL_2242197, EPI_ISL_2242256, EPI_ISL_2242261, EPI_ISL_2242304, EPI_ISL_2242363, EPI_ISL_2242365, EPI_ISL_2242390, EPI_ISL_2242415, EPI_ISL_2242429, EPI_ISL_2242461, EPI_ISL_2242534, EPI_ISL_2242573, EPI_ISL_2242612, EPI_ISL_2242625, EPI_ISL_2242636, EPI_ISL_2242637, EPI_ISL_2242686, EPI_ISL_2242693, EPI_ISL_2242700, EPI_ISL_2242797, EPI_ISL_2242823, EPI_ISL_2242845, EPI_ISL_2242862, EPI_ISL_2242881, EPI_ISL_2242882, EPI_ISL_2242883, EPI_ISL_2242888, EPI_ISL_2242893, EPI_ISL_2242911, EPI_ISL_2242928, EPI_ISL_2242970, EPI_ISL_2242975, EPI_ISL_2242994, EPI_ISL_2242998, EPI_ISL_2243043, EPI_ISL_2243050, EPI_ISL_2243062, EPI_ISL_2243121, EPI_ISL_2243122, EPI_ISL_2243133, EPI_ISL_2243150, EPI_ISL_2243162, EPI_ISL_2243163, EPI_ISL_2243168, EPI_ISL_2243183, EPI_ISL_2243184, EPI_ISL_2243197, EPI_ISL_2243200, EPI_ISL_2243238, EPI_ISL_2243265, EPI_ISL_2243268, EPI_ISL_2243270, EPI_ISL_2243273, EPI_ISL_2243317, EPI_ISL_2243326, EPI_ISL_2243390, EPI_ISL_2243459, EPI_ISL_2243613, EPI_ISL_2243642, EPI_ISL_2243670, EPI_ISL_2243671, EPI_ISL_2243675, EPI_ISL_2243676, EPI_ISL_2243677, EPI_ISL_2243678, EPI_ISL_2243688, EPI_ISL_2243697, EPI_ISL_2243700, EPI_ISL_2243711, EPI_ISL_2243726, EPI_ISL_2243804, EPI_ISL_2243808, EPI_ISL_2243849, EPI_ISL_2243856, EPI_ISL_2243858, EPI_ISL_2243866, EPI_ISL_2243871, EPI_ISL_2243874, EPI_ISL_2243899, EPI_ISL_2243900, EPI_ISL_2243904, EPI_ISL_2243917, EPI_ISL_2243931, EPI_ISL_2243942, EPI_ISL_2243943, EPI_ISL_2243984, EPI_ISL_2243998, EPI_ISL_2244029, EPI_ISL_2244036, EPI_ISL_2244108, EPI_ISL_2244113, EPI_ISL_2244114, EPI_ISL_2244115, EPI_ISL_2244125, EPI_ISL_2244137, EPI_ISL_2244138, EPI_ISL_2244149, EPI_ISL_2244163, EPI_ISL_2244183, EPI_ISL_2244193, EPI_ISL_2244194, EPI_ISL_2244255, EPI_ISL_2244291, EPI_ISL_2244377, EPI_ISL_2244380, EPI_ISL_2244381, EPI_ISL_2244390, EPI_ISL_2244408, EPI_ISL_2244460, EPI_ISL_2244479, EPI_ISL_2244514, EPI_ISL_2244545, EPI_ISL_2244553, EPI_ISL_2244554, EPI_ISL_2244616, EPI_ISL_2244626, EPI_ISL_2244632, EPI_ISL_2244642, EPI_ISL_2244657, EPI_ISL_2244701, EPI_ISL_2244723, EPI_ISL_2244725, EPI_ISL_2244756, EPI_ISL_2244757, EPI_ISL_2244785, EPI_ISL_2244792, EPI_ISL_2244832, EPI_ISL_2244845, EPI_ISL_2244849 | | | |
| see above | Aegis Sciences Corporation | Centers for Disease Control and Prevention Division of Viral Diseases, Pathogen Discovery | Dakota Howard, Dhwani Batra, Peter W. Cook, Kara Moser, Adrian Paskey, Jason Caravas, Benjamin Rambo-Martin, Shatavia Morrison, Christopher Gulvick, Scott Sammons, Yvette Unoarumhi, Darlene Wagner, Matthew Schmerer, Cyndi Clark, Patrick Campbell, Rob Case, Vikramsinha Ghorpade, Holly Houdeshell, Ola Kvalvaag, Dillon Nall, Ethan Sanders, Alec Vest, Shaun Westlund, Matthew Hardison, Clinton R. Paden, Duncan MacCannell |
| EPI_ISL_2247107 | Quest Diagnostics Incorporated | Centers for Disease Control and Prevention Division of Viral Diseases, Pathogen Discovery | Dakota Howard, Dhwani Batra, Peter W. Cook, Kara Moser, Adrian Paskey, Jason Caravas, Benjamin Rambo-Martin, Shatavia Morrison, Christopher Gulvick, Scott Sammons, Yvette Unoarumhi, Darlene Wagner, Matthew Schmerer, S. H. Rosenthal, A. Gerasimova, R. M. Kagan, B. Anderson, M. Hua, Y. Liu, L.E. Bernstein, K.E. Livingston, A. Perez, I. A. Shlyakhter, R. V. Rolando, R. Owen, P. Tanpaiboon, F. Lacbawan, Clinton R. Paden, Duncan MacCannell |
| EPI_ISL_2248022 | Helix/Illumina | Centers for Disease Control and Prevention Division of Viral Diseases, Pathogen Discovery | Dakota Howard, Dhwani Batra, Peter W. Cook, Kara Moser, Adrian Paskey, Jason Caravas, Benjamin Rambo-Martin, Shatavia Morrison, Christopher Gulvick, Scott Sammons, Yvette Unoarumhi, Darlene Wagner, Matthew Schmerer, Eileen de Feo, Jan Antico, Christine Tran, Matthew Tolentino, Shannon Wickline, Kim Gietzen, Brad Sickler, Jingtao Liu, Eric Allen, Phil Febbo, Nicole L. Washington, Simon White, Geraint Levan, Kelly Schiabor Barrett, Elizabeth Cirulli, Alexandre Bolze, Ary Ascencio, Charlotte Rivera-Garcia, Ryan Cho, Jason Nguyen, Sherry Wang, Jimmy Ramirez, Tyler Cassens, Efren Sandoval, Magnus Isaksson, William Lee, David Becker, Marc Laurent, James Lu, Clinton R. Paden, Duncan MacCannell |
| EPI_ISL_2248467, EPI_ISL_2248570 | Quest Diagnostics Incorporated | Centers for Disease Control and Prevention Division of Viral Diseases, Pathogen Discovery | Dakota Howard, Dhwani Batra, Peter W. Cook, Kara Moser, Adrian Paskey, Jason Caravas, Benjamin Rambo-Martin, Shatavia Morrison, Christopher Gulvick, Scott Sammons, Yvette Unoarumhi, Darlene Wagner, Matthew Schmerer, S. H. Rosenthal, A. Gerasimova, R. M. Kagan, B. Anderson, M. Hua, Y. Liu, L.E. Bernstein, K.E. Livingston, A. Perez, I. A. Shlyakhter, R. V. Rolando, R. Owen, P. Tanpaiboon, F. Lacbawan, Clinton R. Paden, Duncan MacCannell |
| EPI_ISL_2254333, EPI_ISL_2254348, EPI_ISL_2254384, EPI_ISL_2254417, EPI_ISL_2254419, EPI_ISL_2254479 | Respiratory Viruses Branch, Centers for Disease Control and Prevention | Respiratory Viruses Branch, Centers for Disease Control and Prevention | Howard,D., Batra,D., Cook,P.W., Moser,K., Paskey,A., Caravas,J., Rambo-Martin,B., Morrison,S., Gulvick,C., Sammons,S., Unoarumhi,Y., Wagner,D., Schmerer,M., Clark,C., Campbell,P., Case,R., Ghorpade,V., Houdeshell,H., Kvalvaag,O., Nall,D., Sanders,E., Vest,A., Westlund,S., Hardison,M., Paden,C.R., MacCannell,D. |

| | | | |
|---|---|---|---|
| EPI_ISL_2262493, EPI_ISL_2262495 | DOHMH Riverside | New York City Public Health Laboratory | Jade Wang, et al. |
| EPI_ISL_2262497 | DOHMH Chelsea | New York City Public Health Laboratory | Jade Wang, et al. |
| EPI_ISL_2262499 | DOHMH Morrisania | New York City Public Health Laboratory | Jade Wang, et al. |
| EPI_ISL_2262500 | DOHMH Jamaica | New York City Public Health Laboratory | Jade Wang, et al. |
| EPI_ISL_2262502 | DOHMH Corona | New York City Public Health Laboratory | Jade Wang, et al. |
| EPI_ISL_2262504 | DOHMH Riverside | New York City Public Health Laboratory | Jade Wang, et al. |
| EPI_ISL_2262506, EPI_ISL_2262508 | DOHMH Crown Heights | New York City Public Health Laboratory | Jade Wang, et al. |
| EPI_ISL_2262510 | DOHMH Jamaica | New York City Public Health Laboratory | Jade Wang, et al. |
| EPI_ISL_2262511 | DOHMH Morrisania | New York City Public Health Laboratory | Jade Wang, et al. |
| EPI_ISL_2262513 | DOHMH Fort Greene | New York City Public Health Laboratory | Jade Wang, et al. |
| EPI_ISL_2262515, EPI_ISL_2262517 | DOHMH PHL | New York City Public Health Laboratory | Jade Wang, et al. |
| EPI_ISL_2262519, EPI_ISL_2262521 | DOHMH Jamaica | New York City Public Health Laboratory | Jade Wang, et al. |
| EPI_ISL_2262522 | DOHMH Corona | New York City Public Health Laboratory | Jade Wang, et al. |
| EPI_ISL_2262524 | DOHMH Central Harlem | New York City Public Health Laboratory | Jade Wang, et al. |
| EPI_ISL_2262526, EPI_ISL_2262528 | DOHMH Riverside | New York City Public Health Laboratory | Jade Wang, et al. |
| EPI_ISL_2262529 | DOHMH Central Harlem | New York City Public Health Laboratory | Jade Wang, et al. |
| EPI_ISL_2262531 | DOHMH Crown Heights | New York City Public Health Laboratory | Jade Wang, et al. |
| EPI_ISL_2262533, EPI_ISL_2262535 | DOHMH Chelsea | New York City Public Health Laboratory | Jade Wang, et al. |
| EPI_ISL_2262537, EPI_ISL_2262539 | DOHMH Jamaica | New York City Public Health Laboratory | Jade Wang, et al. |
| EPI_ISL_2262540 | DOHMH PHL | New York City Public Health Laboratory | Jade Wang, et al. |
| EPI_ISL_2262542 | DOHMH Morrisania | New York City Public Health Laboratory | Jade Wang, et al. |
| EPI_ISL_2262544 | DOHMH Central Harlem | New York City Public Health Laboratory | Jade Wang, et al. |
| EPI_ISL_2262546, EPI_ISL_2262548, EPI_ISL_2262550, EPI_ISL_2262552, EPI_ISL_2262554 | OCME Office Of Chief Medical Examiner | New York City Public Health Laboratory | Jade Wang, et al. |
| EPI_ISL_2262555 | DOHMH Jamaica | New York City Public Health Laboratory | Jade Wang, et al. |
| EPI_ISL_2262557, EPI_ISL_2262559 | DOHMH Central Harlem | New York City Public Health Laboratory | Jade Wang, et al. |
| EPI_ISL_2262561 | DOHMH Chelsea | New York City Public Health Laboratory | Jade Wang, et al. |
| EPI_ISL_2262563 | DOHMH Jamaica | New York City Public Health Laboratory | Jade Wang, et al. |
| EPI_ISL_2262564, EPI_ISL_2262566 | OCME Office Of Chief Medical Examiner | New York City Public Health Laboratory | Jade Wang, et al. |
| EPI_ISL_2262568, EPI_ISL_2262570, EPI_ISL_2262572, EPI_ISL_2262573 | Poplar Healthcare | New York City Public Health Laboratory | Jade Wang, et al. |
| EPI_ISL_2262575 | OCME Office Of Chief Medical Examiner | New York City Public Health Laboratory | Jade Wang, et al. |
| EPI_ISL_2262577, EPI_ISL_2262579, EPI_ISL_2262581 | DOHMH Chelsea | New York City Public Health Laboratory | Jade Wang, et al. |
| EPI_ISL_2262582, EPI_ISL_2262584 | DOHMH Central Harlem | New York City Public Health Laboratory | Jade Wang, et al. |
| EPI_ISL_2262586, EPI_ISL_2262588 | DOHMH Jamaica | New York City Public Health Laboratory | Jade Wang, et al. |
| EPI_ISL_2262590 | DOHMH Fort Greene | New York City Public Health Laboratory | Jade Wang, et al. |
| EPI_ISL_2262592 | DOHMH Central Harlem | New York City Public Health Laboratory | Jade Wang, et al. |
| EPI_ISL_2262594, EPI_ISL_2262595, EPI_ISL_2262597 | DOHMH Morrisania | New York City Public Health Laboratory | Jade Wang, et al. |
| EPI_ISL_2262599 | DOHMH Jamaica | New York City Public Health Laboratory | Jade Wang, et al. |
| EPI_ISL_2262601, EPI_ISL_2262602 | DOHMH Corona | New York City Public Health Laboratory | Jade Wang, et al. |
| EPI_ISL_2262604, EPI_ISL_2262606, EPI_ISL_2262607 | DOHMH PHL | New York City Public Health Laboratory | Jade Wang, et al. |
| EPI_ISL_2262609 | DOHMH Riverside | New York City Public Health Laboratory | Jade Wang, et al. |
| EPI_ISL_2262612 | DOHMH Fort Greene | New York City Public Health Laboratory | Jade Wang, et al. |
| EPI_ISL_2262613 | DOHMH Crown Heights | New York City Public Health Laboratory | Jade Wang, et al. |
| EPI_ISL_2262615 | OCME Office Of Chief Medical Examiner | New York City Public Health Laboratory | Jade Wang, et al. |
| EPI_ISL_2262617 | DOHMH Jamaica | New York City Public Health Laboratory | Jade Wang, et al. |
| EPI_ISL_2262618 | DOHMH Morrisania | New York City Public Health Laboratory | Jade Wang, et al. |
| EPI_ISL_2262620, EPI_ISL_2262622 | DOHMH PHL | New York City Public Health Laboratory | Jade Wang, et al. |
| EPI_ISL_2262624 | DOHMH Jamaica | New York City Public Health Laboratory | Jade Wang, et al. |
| EPI_ISL_2262626 | DOHMH Crown Heights | New York City Public Health Laboratory | Jade Wang, et al. |
| EPI_ISL_2262628, EPI_ISL_2262629 | DOHMH Morrisania | New York City Public Health Laboratory | Jade Wang, et al. |
| EPI_ISL_2262631, EPI_ISL_2262633 | DOHMH Central Harlem | New York City Public Health Laboratory | Jade Wang, et al. |
| EPI_ISL_2262635, EPI_ISL_2262636, EPI_ISL_2262638, EPI_ISL_2262642, EPI_ISL_2262644, EPI_ISL_2262645, EPI_ISL_2262647 | DOHMH Jamaica | New York City Public Health Laboratory | Jade Wang, et al. |
| EPI_ISL_2262649 | DOHMH Central Harlem | New York City Public Health Laboratory | Jade Wang, et al. |
| EPI_ISL_2262651, EPI_ISL_2262652 | DOHMH Fort Greene | New York City Public Health Laboratory | Jade Wang, et al. |

| | | | |
|---|---|---|---|
| EPI_ISL_2262654 | DOHMH PHL | New York City Public Health Laboratory | Jade Wang, et al. |
| EPI_ISL_2262655, EPI_ISL_2262657, EPI_ISL_2262659 | DOHMH Morrisania | New York City Public Health Laboratory | Jade Wang, et al. |
| EPI_ISL_2262661 | DOHMH Jamaica | New York City Public Health Laboratory | Jade Wang, et al. |
| EPI_ISL_2262663 | Department of Homeless Services | New York City Public Health Laboratory | Jade Wang, et al. |
| EPI_ISL_2262665 | OCME Office Of Chief Medical Examiner | New York City Public Health Laboratory | Jade Wang, et al. |
| EPI_ISL_2262666 | DOHMH PHL | New York City Public Health Laboratory | Jade Wang, et al. |
| EPI_ISL_2262668 | DOHMH Corona | New York City Public Health Laboratory | Jade Wang, et al. |
| EPI_ISL_2262670, EPI_ISL_2262672 | DOHMH Chelsea | New York City Public Health Laboratory | Jade Wang, et al. |
| EPI_ISL_2262674 | DOHMH Riverside | New York City Public Health Laboratory | Jade Wang, et al. |
| EPI_ISL_2262675, EPI_ISL_2262676, EPI_ISL_2262677 | DOHMH Morrisania | New York City Public Health Laboratory | Jade Wang, et al. |
| EPI_ISL_2262679 | DOHMH Central Harlem | New York City Public Health Laboratory | Jade Wang, et al. |
| EPI_ISL_2262680 | DOHMH Riverside | New York City Public Health Laboratory | Jade Wang, et al. |
| EPI_ISL_2262682 | DOHMH Central Harlem | New York City Public Health Laboratory | Jade Wang, et al. |
| EPI_ISL_2262684 | Department of Homeless Services | New York City Public Health Laboratory | Jade Wang, et al. |
| EPI_ISL_2262686 | OCME Office Of Chief Medical Examiner | New York City Public Health Laboratory | Jade Wang, et al. |
| EPI_ISL_2268297, EPI_ISL_2268675, EPI_ISL_2268737 | Quest Diagnostics Incorporated | Centers for Disease Control and Prevention Division of Viral Diseases, Pathogen Discovery | Dakota Howard, Dhwani Batra, Peter W. Cook, Kara Moser, Adrian Paskey, Jason Caravas, Benjamin Rambo-Martin, Shatavia Morrison, Christopher Gulvick, Scott Sammons, Yvette Unoarumhi, Darlene Wagner, Matthew Schmerer, S. H. Rosenthal, A. Gerasimova, R. M. Kagan, B. Anderson, M. Hua, Y. Liu, L.E. Bernstein, K.E. Livingston, A. Perez, I. A. Shlyakhter, R. V. Rolando, R. Owen, P. Tanpaiboon, F. Lacbawan, Clinton R. Paden, Duncan MacCannell |
| EPI_ISL_2269696, EPI_ISL_2269767, EPI_ISL_2270003 | Helix/Illumina | Centers for Disease Control and Prevention Division of Viral Diseases, Pathogen Discovery | Dakota Howard, Dhwani Batra, Peter W. Cook, Kara Moser, Adrian Paskey, Jason Caravas, Benjamin Rambo-Martin, Shatavia Morrison, Christopher Gulvick, Scott Sammons, Yvette Unoarumhi, Darlene Wagner, Matthew Schmerer, Eileen de Feo, Jan Antico, Christine Tran, Matthew Tolentino, Shannon Wickline, Kim Gietzen, Brad Sickler, Jingtao Liu, Eric Allen, Phil Febbo, Nicole L. Washington, Simon White, Geraint Levan, Kelly Schiabor Barrett, Elizabeth Cirulli, Alexandre Bolze, Ary Ascencio, Charlotte Rivera-Garcia, Ryan Cho, Jason Nguyen, Sherry Wang, Jimmy Ramirez, Tyler Cassens, Efren Sandoval, Magnus Isaksson, William Lee, David Becker, Marc Laurent, James Lu, Clinton R. Paden, Duncan MacCannell |
| EPI_ISL_2273871, EPI_ISL_2273872, EPI_ISL_2273873, EPI_ISL_2273874, EPI_ISL_2273875, EPI_ISL_2273876, EPI_ISL_2273877, EPI_ISL_2273878, EPI_ISL_2273879, EPI_ISL_2273880, EPI_ISL_2273881, EPI_ISL_2273882, EPI_ISL_2273883, EPI_ISL_2273884, EPI_ISL_2273885, EPI_ISL_2273886, EPI_ISL_2273887, EPI_ISL_2273888, EPI_ISL_2273889, EPI_ISL_2273890, EPI_ISL_2273891, EPI_ISL_2273892, EPI_ISL_2273893, EPI_ISL_2273894, EPI_ISL_2273895, EPI_ISL_2273896, EPI_ISL_2273897, EPI_ISL_2273898, EPI_ISL_2273899, EPI_ISL_2273900, EPI_ISL_2273901, EPI_ISL_2273902, EPI_ISL_2273903, EPI_ISL_2273904, EPI_ISL_2273905, EPI_ISL_2273906, EPI_ISL_2273907, EPI_ISL_2273908, EPI_ISL_2273909, EPI_ISL_2273910, EPI_ISL_2273911, EPI_ISL_2273912, EPI_ISL_2273913, EPI_ISL_2273914, EPI_ISL_2273915, EPI_ISL_2273916, EPI_ISL_2273917, EPI_ISL_2273918, EPI_ISL_2273919, EPI_ISL_2273920, EPI_ISL_2273921, EPI_ISL_2273922, EPI_ISL_2273923, EPI_ISL_2273924, EPI_ISL_2273925, EPI_ISL_2273926, EPI_ISL_2273927, EPI_ISL_2273928, EPI_ISL_2273929, EPI_ISL_2273930, EPI_ISL_2273931, EPI_ISL_2273932, EPI_ISL_2273933, EPI_ISL_2273934, EPI_ISL_2273935, EPI_ISL_2273936, EPI_ISL_2273937, EPI_ISL_2273938, EPI_ISL_2273939, EPI_ISL_2273940, EPI_ISL_2273941, EPI_ISL_2273942, EPI_ISL_2273943, EPI_ISL_2273944, EPI_ISL_2273945, EPI_ISL_2273946, EPI_ISL_2273947, EPI_ISL_2273948, EPI_ISL_2273949, EPI_ISL_2273950, EPI_ISL_2273951, EPI_ISL_2273952, EPI_ISL_2273953, EPI_ISL_2273954, EPI_ISL_2273955, EPI_ISL_2273956, EPI_ISL_2273957, EPI_ISL_2273958, EPI_ISL_2273959, EPI_ISL_2273960, EPI_ISL_2273961, EPI_ISL_2273962, EPI_ISL_2273963, EPI_ISL_2273964, EPI_ISL_2273965, EPI_ISL_2273966, EPI_ISL_2273967, EPI_ISL_2273968, EPI_ISL_2273969, EPI_ISL_2273970, EPI_ISL_2273971, EPI_ISL_2273972, EPI_ISL_2273973, EPI_ISL_2273974, EPI_ISL_2273975, EPI_ISL_2273976, EPI_ISL_2273977, EPI_ISL_2273978, EPI_ISL_2273979, EPI_ISL_2273980, EPI_ISL_2273981, EPI_ISL_2273982, EPI_ISL_2273983, EPI_ISL_2273984, EPI_ISL_2273985, EPI_ISL_2273986, EPI_ISL_2273987, EPI_ISL_2273988, EPI_ISL_2273989, EPI_ISL_2273990, EPI_ISL_2273991, EPI_ISL_2273992, EPI_ISL_2273993, EPI_ISL_2273994, EPI_ISL_2273995, EPI_ISL_2273996, EPI_ISL_2273997, EPI_ISL_2273998 | | | |
| see above | Weill Cornell Medicine | New York Genome Center | Michael Zody, Andre Corvelo, Dayna M. Oschwald, Samantha Fennessey, Tom Maniatis, Melissa Cushing, Olivier Elemento, Margaret Elizabeth Ross, Chris Mason, Priya Velu, Hanna Rennert, Arryn Craney, Lars F Westblade |
| EPI_ISL_2274574 | URMC LABS | Wadsworth Center, New York State Department of Health | Kirsten St. George, Daryl M. Lamson, Alexis Russell, Matthew Shudt, Melissa A Leisner, Jonathan Plitnick, Catharine Prussing, Navjot Singh, John Kelly, Erasmus Schneider, Erica Lasek-Nesselquist |
| EPI_ISL_2274575 | SUNY UPSTATE MEDICAL UNIVERSITY | Wadsworth Center, New York State Department of Health | Kirsten St. George, Daryl M. Lamson, Alexis Russell, Matthew Shudt, Melissa A Leisner, Jonathan Plitnick, Catharine Prussing, Navjot Singh, John Kelly, Erasmus Schneider, Erica Lasek-Nesselquist |
| EPI_ISL_2274576, EPI_ISL_2274577, EPI_ISL_2274578, EPI_ISL_2274579, EPI_ISL_2274580, EPI_ISL_2274581, EPI_ISL_2274582, EPI_ISL_2274583, EPI_ISL_2274584, EPI_ISL_2274585 | STONY BROOK UNIVERSITY HOSPITAL | Wadsworth Center, New York State Department of Health | Kirsten St. George, Daryl M. Lamson, Alexis Russell, Matthew Shudt, Melissa A Leisner, Jonathan Plitnick, Catharine Prussing, Navjot Singh, John Kelly, Erasmus Schneider, Erica Lasek-Nesselquist |
| EPI_ISL_2274586 | OSWEGO HOSPITAL LABORATORY | Wadsworth Center, New York State Department of Health | Kirsten St. George, Daryl M. Lamson, Alexis Russell, Matthew Shudt, Melissa A Leisner, Jonathan Plitnick, Catharine Prussing, Navjot Singh, John Kelly, Erasmus Schneider, Erica Lasek-Nesselquist |
| EPI_ISL_2274587, EPI_ISL_2274588, EPI_ISL_2274589, EPI_ISL_2274590, EPI_ISL_2274591, EPI_ISL_2274592, EPI_ISL_2274593, EPI_ISL_2274594, EPI_ISL_2274595, EPI_ISL_2274596, EPI_ISL_2274597, EPI_ISL_2274598, EPI_ISL_2274599, EPI_ISL_2274600, EPI_ISL_2274601, EPI_ISL_2274602, EPI_ISL_2274603, EPI_ISL_2274604, EPI_ISL_2274605, EPI_ISL_2274606 | | | |
| see above | NYC Pandemic Response Lab | Wadsworth Center, New York State Department of Health | Kirsten St. George, Daryl M. Lamson, Alexis Russell, Matthew Shudt, Melissa A Leisner, Jonathan Plitnick, Catharine Prussing, Navjot Singh, John Kelly, Erasmus Schneider, Erica Lasek-Nesselquist |
| EPI_ISL_2274607, EPI_ISL_2274608, EPI_ISL_2274609, EPI_ISL_2274610, EPI_ISL_2274611, EPI_ISL_2274612, EPI_ISL_2274613, EPI_ISL_2274614, EPI_ISL_2274615, EPI_ISL_2274616, EPI_ISL_2274617 | | | |
| see above | MONTEFIORE MEDICAL CENTER LABORATORIES | Wadsworth Center, New York State Department of Health | Kirsten St. George, Daryl M. Lamson, Alexis Russell, Matthew Shudt, Melissa A Leisner, Jonathan Plitnick, Catharine Prussing, Navjot Singh, John Kelly, Erasmus Schneider, Erica Lasek-Nesselquist |
| EPI_ISL_2274618, EPI_ISL_2274619, EPI_ISL_2274620, EPI_ISL_2274621, EPI_ISL_2274622, EPI_ISL_2274623, EPI_ISL_2274624, EPI_ISL_2274625, EPI_ISL_2274626, EPI_ISL_2274627, EPI_ISL_2274628, EPI_ISL_2274629, EPI_ISL_2274630 | | | |
| see above | ALBANY MEDICAL CENTER | Wadsworth Center, New York State Department of Health | Kirsten St. George, Daryl M. Lamson, Alexis Russell, Matthew Shudt, Melissa A Leisner, Jonathan Plitnick, Catharine Prussing, Navjot Singh, John Kelly, Erasmus Schneider, Erica Lasek-Nesselquist |
| EPI_ISL_2274631, EPI_ISL_2274632, EPI_ISL_2274633, EPI_ISL_2274634, EPI_ISL_2274635, EPI_ISL_2274636, EPI_ISL_2274637, EPI_ISL_2274638, EPI_ISL_2274639, EPI_ISL_2274640, EPI_ISL_2274641, EPI_ISL_2274642, EPI_ISL_2274643, EPI_ISL_2274644, EPI_ISL_2274645, EPI_ISL_2274646 | | | |
| see above | URMC LABS | Wadsworth Center, New York State Department of Health | Kirsten St. George, Daryl M. Lamson, Alexis Russell, Matthew Shudt, Melissa A Leisner, Jonathan Plitnick, Catharine Prussing, Navjot Singh, John Kelly, Erasmus Schneider, Erica Lasek-Nesselquist |
| EPI_ISL_2274647, EPI_ISL_2274648, EPI_ISL_2274649, EPI_ISL_2274650, EPI_ISL_2274651, EPI_ISL_2274652, EPI_ISL_2274653, EPI_ISL_2274654 | WESTCHESTER MEDICAL CENTER | Wadsworth Center, New York State Department of Health | Kirsten St. George, Daryl M. Lamson, Alexis Russell, Matthew Shudt, Melissa A Leisner, Jonathan Plitnick, Catharine Prussing, Navjot Singh, John Kelly, Erasmus Schneider, Erica Lasek-Nesselquist |
| EPI_ISL_2274657, EPI_ISL_2274658, EPI_ISL_2274659, EPI_ISL_2274660, EPI_ISL_2274661, EPI_ISL_2274662, EPI_ISL_2274663 | SUNY UPSTATE MEDICAL UNIVERSITY | Wadsworth Center, New York State Department of Health | Kirsten St. George, Daryl M. Lamson, Alexis Russell, Matthew Shudt, Melissa A Leisner, Jonathan Plitnick, Catharine Prussing, Navjot Singh, John Kelly, Erasmus Schneider, Erica Lasek-Nesselquist |
| EPI_ISL_2274664 | ALBANY MEDICAL CENTER | Wadsworth Center, New York State Department of Health | Kirsten St. George, Daryl M. Lamson, Alexis Russell, Matthew Shudt, Melissa A Leisner, Jonathan Plitnick, Catharine Prussing, Navjot Singh, John Kelly, Erasmus Schneider, Erica Lasek-Nesselquist |

| | | | |
|---|---|---|---|
| EPI_ISL_2274665, EPI_ISL_2274666, EPI_ISL_2274667, EPI_ISL_2274668, EPI_ISL_2274669, EPI_ISL_2274670, EPI_ISL_2274671, EPI_ISL_2274672, EPI_ISL_2274673, EPI_ISL_2274674, EPI_ISL_2274675 | | | |
| see above | URMC LABS | Wadsworth Center, New York State Department of Health | Kirsten St. George, Daryl M. Lamson, Alexis Russell, Matthew Shudt, Melissa A Leisner, Jonathan Plitnick, Catharine Prussing, Navjot Singh, John Kelly, Erasmus Schneider, Erica Lasek-Nesselquist |
| EPI_ISL_2274676, EPI_ISL_2274677, EPI_ISL_2274678, EPI_ISL_2274679, EPI_ISL_2274680, EPI_ISL_2274681, EPI_ISL_2274682, EPI_ISL_2274683, EPI_ISL_2274684, EPI_ISL_2274685, EPI_ISL_2274686, EPI_ISL_2274687, EPI_ISL_2274688, EPI_ISL_2274689, EPI_ISL_2274690, EPI_ISL_2274691, EPI_ISL_2274692, EPI_ISL_2274693, EPI_ISL_2274694, EPI_ISL_2274695, EPI_ISL_2274696, EPI_ISL_2274697, EPI_ISL_2274698, EPI_ISL_2274699, EPI_ISL_2274700, EPI_ISL_2274701, EPI_ISL_2274702, EPI_ISL_2274703, EPI_ISL_2274704, EPI_ISL_2274705, EPI_ISL_2274706, EPI_ISL_2274707, EPI_ISL_2274708, EPI_ISL_2274709, EPI_ISL_2274710, EPI_ISL_2274711, EPI_ISL_2274712, EPI_ISL_2274713, EPI_ISL_2274714 | | | |
| see above | NYC Pandemic Response Lab | Wadsworth Center, New York State Department of Health | Kirsten St. George, Daryl M. Lamson, Alexis Russell, Matthew Shudt, Melissa A Leisner, Jonathan Plitnick, Catharine Prussing, Navjot Singh, John Kelly, Erasmus Schneider, Erica Lasek-Nesselquist |
| EPI_ISL_2277639, EPI_ISL_2277640, EPI_ISL_2277641 | Stony Brook Medicine | New York Genome Center | Michael Zody, Andre Corvelo, Dayna M. Oschwald, Samantha Fennessey, Tom Maniatis , Sarah A. Halbohn, Alaa B. Salaheldin, Sarah A. Malik, Josue M. Flores Acuna, Dimitri Gnatenko, Silvia G. Spitzer, Margaret M. McGovern, Eric D. Spitzer, Richard R. Kew, Kenneth R. Shroyer, Jiang Chen, Susan V. Donelan |
| EPI_ISL_2278731 | MONTEFIORE MEDICAL CENTER LABORATORIES | Wadsworth Center, New York State Department of Health | Kirsten St. George, Daryl M. Lamson, Alexis Russell, Matthew Shudt, Melissa A Leisner, Jonathan Plitnick, Catharine Prussing, Navjot Singh, John Kelly, Erasmus Schneider, Erica Lasek-Nesselquist |
| EPI_ISL_2278732, EPI_ISL_2278733, EPI_ISL_2278734 | SUNY UPSTATE MEDICAL UNIVERSITY | Wadsworth Center, New York State Department of Health | Kirsten St. George, Daryl M. Lamson, Alexis Russell, Matthew Shudt, Melissa A Leisner, Jonathan Plitnick, Catharine Prussing, Navjot Singh, John Kelly, Erasmus Schneider, Erica Lasek-Nesselquist |
| EPI_ISL_2278735 | URMC LABS | Wadsworth Center, New York State Department of Health | Kirsten St. George, Daryl M. Lamson, Alexis Russell, Matthew Shudt, Melissa A Leisner, Jonathan Plitnick, Catharine Prussing, Navjot Singh, John Kelly, Erasmus Schneider, Erica Lasek-Nesselquist |
| EPI_ISL_2278736 | MONTEFIORE MEDICAL CENTER LABORATORIES | Wadsworth Center, New York State Department of Health | Kirsten St. George, Daryl M. Lamson, Alexis Russell, Matthew Shudt, Melissa A Leisner, Jonathan Plitnick, Catharine Prussing, Navjot Singh, John Kelly, Erasmus Schneider, Erica Lasek-Nesselquist |
| EPI_ISL_2278737, EPI_ISL_2278738 | ALBANY MEDICAL CENTER | Wadsworth Center, New York State Department of Health | Kirsten St. George, Daryl M. Lamson, Alexis Russell, Matthew Shudt, Melissa A Leisner, Jonathan Plitnick, Catharine Prussing, Navjot Singh, John Kelly, Erasmus Schneider, Erica Lasek-Nesselquist |
| EPI_ISL_2278739 | URMC LABS | Wadsworth Center, New York State Department of Health | Kirsten St. George, Daryl M. Lamson, Alexis Russell, Matthew Shudt, Melissa A Leisner, Jonathan Plitnick, Catharine Prussing, Navjot Singh, John Kelly, Erasmus Schneider, Erica Lasek-Nesselquist |
| EPI_ISL_2278740, EPI_ISL_2278741 | SUNY UPSTATE MEDICAL UNIVERSITY | Wadsworth Center, New York State Department of Health | Kirsten St. George, Daryl M. Lamson, Alexis Russell, Matthew Shudt, Melissa A Leisner, Jonathan Plitnick, Catharine Prussing, Navjot Singh, John Kelly, Erasmus Schneider, Erica Lasek-Nesselquist |
| EPI_ISL_2278742 | ALBANY MEDICAL CENTER | Wadsworth Center, New York State Department of Health | Kirsten St. George, Daryl M. Lamson, Alexis Russell, Matthew Shudt, Melissa A Leisner, Jonathan Plitnick, Catharine Prussing, Navjot Singh, John Kelly, Erasmus Schneider, Erica Lasek-Nesselquist |
| EPI_ISL_2278743, EPI_ISL_2278744, EPI_ISL_2278745, EPI_ISL_2278746, EPI_ISL_2278747, EPI_ISL_2278748, EPI_ISL_2278749, EPI_ISL_2278750, EPI_ISL_2278751, EPI_ISL_2278752, EPI_ISL_2278753, EPI_ISL_2278754, EPI_ISL_2278755, EPI_ISL_2278756, EPI_ISL_2278757, EPI_ISL_2278758, EPI_ISL_2278759, EPI_ISL_2278760, EPI_ISL_2278761, EPI_ISL_2278762, EPI_ISL_2278763, EPI_ISL_2278764, EPI_ISL_2278765, EPI_ISL_2278766, EPI_ISL_2278767, EPI_ISL_2278768, EPI_ISL_2278769, EPI_ISL_2278770, EPI_ISL_2278771, EPI_ISL_2278772, EPI_ISL_2278773, EPI_ISL_2278774, EPI_ISL_2278775, EPI_ISL_2278776, EPI_ISL_2278777, EPI_ISL_2278778, EPI_ISL_2278779, EPI_ISL_2278780, EPI_ISL_2278781, EPI_ISL_2278782, EPI_ISL_2278783, EPI_ISL_2278784, EPI_ISL_2278785, EPI_ISL_2278786, EPI_ISL_2278787, EPI_ISL_2278788, EPI_ISL_2278789, EPI_ISL_2278790, EPI_ISL_2278791, EPI_ISL_2278792, EPI_ISL_2278793, EPI_ISL_2278794, EPI_ISL_2278795, EPI_ISL_2278796 | | | |
| see above | URMC LABS | Wadsworth Center, New York State Department of Health | Kirsten St. George, Daryl M. Lamson, Alexis Russell, Matthew Shudt, Melissa A Leisner, Jonathan Plitnick, Catharine Prussing, Navjot Singh, John Kelly, Erasmus Schneider, Erica Lasek-Nesselquist |
| EPI_ISL_2278797 | Maimonides Medical Center | Wadsworth Center, New York State Department of Health | Kirsten St. George, Daryl M. Lamson, Alexis Russell, Matthew Shudt, Melissa A Leisner, Jonathan Plitnick, Catharine Prussing, Navjot Singh, John Kelly, Erasmus Schneider, Erica Lasek-Nesselquist |
| EPI_ISL_2278798, EPI_ISL_2278799 | URMC LABS | Wadsworth Center, New York State Department of Health | Kirsten St. George, Daryl M. Lamson, Alexis Russell, Matthew Shudt, Melissa A Leisner, Jonathan Plitnick, Catharine Prussing, Navjot Singh, John Kelly, Erasmus Schneider, Erica Lasek-Nesselquist |
| EPI_ISL_2279499, EPI_ISL_2279500, EPI_ISL_2279501, EPI_ISL_2279502 | SUNY UPSTATE MEDICAL UNIVERSITY | Wadsworth Center, New York State Department of Health | Kirsten St. George, Daryl M. Lamson, Alexis Russell, Matthew Shudt, Melissa A Leisner, Jonathan Plitnick, Catharine Prussing, Navjot Singh, John Kelly, Erasmus Schneider, Erica Lasek-Nesselquist |
| EPI_ISL_2279503, EPI_ISL_2279504, EPI_ISL_2279505 | ALBANY MEDICAL CENTER | Wadsworth Center, New York State Department of Health | Kirsten St. George, Daryl M. Lamson, Alexis Russell, Matthew Shudt, Melissa A Leisner, Jonathan Plitnick, Catharine Prussing, Navjot Singh, John Kelly, Erasmus Schneider, Erica Lasek-Nesselquist |
| EPI_ISL_2279506, EPI_ISL_2279507, EPI_ISL_2279508, EPI_ISL_2279509, EPI_ISL_2279510, EPI_ISL_2279511, EPI_ISL_2279512, EPI_ISL_2279513, EPI_ISL_2279514, EPI_ISL_2279515, EPI_ISL_2279516, EPI_ISL_2279517, EPI_ISL_2279518, EPI_ISL_2279519, EPI_ISL_2279520, EPI_ISL_2279521, EPI_ISL_2279522, EPI_ISL_2279523, EPI_ISL_2279524, EPI_ISL_2279525, EPI_ISL_2279526, EPI_ISL_2279527, EPI_ISL_2279528, EPI_ISL_2279529, EPI_ISL_2279530, EPI_ISL_2279531, EPI_ISL_2279532, EPI_ISL_2279533, EPI_ISL_2279534, EPI_ISL_2279535, EPI_ISL_2279536, EPI_ISL_2279537, EPI_ISL_2279538, EPI_ISL_2279539, EPI_ISL_2279540, EPI_ISL_2279541, EPI_ISL_2279542, EPI_ISL_2279543, EPI_ISL_2279544, EPI_ISL_2279545, EPI_ISL_2279546, EPI_ISL_2279547, EPI_ISL_2279548, EPI_ISL_2279549, EPI_ISL_2279550, EPI_ISL_2279551, EPI_ISL_2279552, EPI_ISL_2279553, EPI_ISL_2279554, EPI_ISL_2279555, EPI_ISL_2279556, EPI_ISL_2279557, EPI_ISL_2279558, EPI_ISL_2279559, EPI_ISL_2279560, EPI_ISL_2279561, EPI_ISL_2279562, EPI_ISL_2279563, EPI_ISL_2279564, EPI_ISL_2279565, EPI_ISL_2279566, EPI_ISL_2279567, EPI_ISL_2279568, EPI_ISL_2279569, EPI_ISL_2279570, EPI_ISL_2279571, EPI_ISL_2279572, EPI_ISL_2279573, EPI_ISL_2279574, EPI_ISL_2279575, EPI_ISL_2279576, EPI_ISL_2279577, EPI_ISL_2279578, EPI_ISL_2279579, EPI_ISL_2279580, EPI_ISL_2279581, EPI_ISL_2279582, EPI_ISL_2279583, EPI_ISL_2279584 | | | |
| see above | URMC LABS | Wadsworth Center, New York State Department of Health | Kirsten St. George, Daryl M. Lamson, Alexis Russell, Matthew Shudt, Melissa A Leisner, Jonathan Plitnick, Catharine Prussing, Navjot Singh, John Kelly, Erasmus Schneider, Erica Lasek-Nesselquist |
| EPI_ISL_2279585 | BIO-REFERENCE LABORATORIES | Wadsworth Center, New York State Department of Health | Kirsten St. George, Daryl M. Lamson, Alexis Russell, Matthew Shudt, Melissa A Leisner, Jonathan Plitnick, Catharine Prussing, Navjot Singh, John Kelly, Erasmus Schneider, Erica Lasek-Nesselquist |
| EPI_ISL_2280230, EPI_ISL_2280244, EPI_ISL_2280246 | Fulgent Genetics | Centers for Disease Control and Prevention Division of Viral Diseases, Pathogen Discovery | Dakota Howard, Dhwani Batra, Peter W. Cook, Kara Moser, Adrian Paskey, Jason Caravas, Benjamin Rambo-Martin, Shatavia Morrison, Christopher Gulvick, Scott Sammons, Yvette Unoarumhi, Darlene Wagner, Matthew Schmerer, Harry Gao, Mickey Li, John Gao, Joseph Fierro, Benafsh Sapra, Becky Tsai, Yan Meng, Doreen Ng, James Xie, Clinton R. Paden, Duncan MacCannell |
| EPI_ISL_2280327, EPI_ISL_2280340, EPI_ISL_2280341, EPI_ISL_2280385, EPI_ISL_2280423, EPI_ISL_2280424, EPI_ISL_2280425, EPI_ISL_2280460, EPI_ISL_2280468, EPI_ISL_2280499, EPI_ISL_2280504, EPI_ISL_2280505, EPI_ISL_2280530, EPI_ISL_2280537, EPI_ISL_2280756, EPI_ISL_2280760, EPI_ISL_2280771, EPI_ISL_2280774, EPI_ISL_2280782, EPI_ISL_2280906, EPI_ISL_2280940, EPI_ISL_2280942, EPI_ISL_2280960, EPI_ISL_2280961, EPI_ISL_2280996, EPI_ISL_2281008, EPI_ISL_2281031, EPI_ISL_2281076, EPI_ISL_2281100, EPI_ISL_2281172, EPI_ISL_2281182 | | | |
| see above | Aegis Sciences Corporation | Centers for Disease Control and Prevention Division of Viral Diseases, Pathogen Discovery | Dakota Howard, Dhwani Batra, Peter W. Cook, Kara Moser, Adrian Paskey, Jason Caravas, Benjamin Rambo-Martin, Shatavia Morrison, Christopher Gulvick, Scott Sammons, Yvette Unoarumhi, Darlene Wagner, Matthew Schmerer, Cyndi Clark, Patrick Campbell, Rob Case, Vikramsinha Ghorpade, Holly Houdeshell, Ola Kvalvaag, Dillon Nall, Ethan Sanders, Alec Vest, Shaun Westlund, Matthew Hardison, Clinton R. Paden, Duncan MacCannell |
| EPI_ISL_2282235, EPI_ISL_2282236, EPI_ISL_2282237, EPI_ISL_2282238, EPI_ISL_2282239, EPI_ISL_2282240, EPI_ISL_2282241, EPI_ISL_2282242, EPI_ISL_2282243, EPI_ISL_2282244, EPI_ISL_2282245, EPI_ISL_2282246 | | | |
| see above | Weill Cornell Medicine | New York Genome Center | Michael Zody, Andre Corvelo, Dayna M. Oschwald, Samantha Fennessey, Tom Maniatis, Melissa Cushing, Olivier Elemento, Margaret Elizabeth Ross, Chris Mason, Priya Velu, Hanna Rennert, Arryn Craney, Lars F Westblade |
| EPI_ISL_2282329, EPI_ISL_2282900, EPI_ISL_2282902 | Infinity Biologix | Centers for Disease Control and Prevention Division of Viral Diseases, Pathogen Discovery | Dakota Howard, Dhwani Batra, Peter W. Cook, Kara Moser, Adrian Paskey, Jason Caravas, Benjamin Rambo-Martin, Shatavia Morrison, Christopher Gulvick, Scott Sammons, Yvette Unoarumhi, Darlene Wagner, Matthew Schmerer, Christian Bixby, Yihe Wang, Jonathan Schultz, Chirayu Goswami, Russ Hager, Robin Grimwood, Clinton R. Paden, Duncan MacCannell |
| EPI_ISL_2283114, EPI_ISL_2283116, EPI_ISL_2283117, EPI_ISL_2283119, EPI_ISL_2283121, EPI_ISL_2283122, EPI_ISL_2283124, EPI_ISL_2283126, EPI_ISL_2283127, EPI_ISL_2283129, EPI_ISL_2283130, EPI_ISL_2283132, EPI_ISL_2283134, EPI_ISL_2283136, EPI_ISL_2283138, EPI_ISL_2283139, EPI_ISL_2283142, EPI_ISL_2283145, EPI_ISL_2283146, EPI_ISL_2283148, EPI_ISL_2283150, EPI_ISL_2283151, EPI_ISL_2283153, EPI_ISL_2283154, EPI_ISL_2283156, EPI_ISL_2283158, EPI_ISL_2283159, EPI_ISL_2283161, EPI_ISL_2283162, EPI_ISL_2283164, EPI_ISL_2283166, EPI_ISL_2283167, EPI_ISL_2283169, EPI_ISL_2283170, EPI_ISL_2283172, EPI_ISL_2283174, EPI_ISL_2283175, EPI_ISL_2283177, EPI_ISL_2283178, EPI_ISL_2283180, EPI_ISL_2283182, EPI_ISL_2283184, EPI_ISL_2283186, EPI_ISL_2283187, EPI_ISL_2283189, EPI_ISL_2283190, EPI_ISL_2283192, EPI_ISL_2283194, EPI_ISL_2283195, EPI_ISL_2283197, EPI_ISL_2283199, EPI_ISL_2283200, EPI_ISL_2283202, EPI_ISL_2283204, EPI_ISL_2283205, EPI_ISL_2283207, EPI_ISL_2283208, EPI_ISL_2283210, EPI_ISL_2283212, EPI_ISL_2283213, EPI_ISL_2283215, EPI_ISL_2283217, EPI_ISL_2283218, EPI_ISL_2283220, EPI_ISL_2283221, EPI_ISL_2283223, EPI_ISL_2283225, EPI_ISL_2283226, EPI_ISL_2283228, EPI_ISL_2283229, EPI_ISL_2283231, EPI_ISL_2283233, EPI_ISL_2283234, EPI_ISL_2283236, EPI_ISL_2283237, EPI_ISL_2283239, EPI_ISL_2283240, EPI_ISL_2283242, EPI_ISL_2283244, EPI_ISL_2283245, EPI_ISL_2283247, EPI_ISL_2283248, EPI_ISL_2283250, EPI_ISL_2283251 | | | |
| see above | Weill Cornell Medicine | New York Genome Center | Michael Zody, Andre Corvelo, Dayna M. Oschwald, Samantha Fennessey, Tom Maniatis, Melissa Cushing, Olivier Elemento, Margaret Elizabeth Ross, Chris Mason, Priya Velu, Hanna Rennert, Arryn Craney, Lars F Westblade |
| EPI_ISL_2283252 | Infinity Biologix | Centers for Disease Control and Prevention Division of Viral Diseases, Pathogen Discovery | Dakota Howard, Dhwani Batra, Peter W. Cook, Kara Moser, Adrian Paskey, Jason Caravas, Benjamin Rambo-Martin, Shatavia Morrison, Christopher Gulvick, Scott Sammons, Yvette Unoarumhi, Darlene Wagner, Matthew Schmerer, Christian Bixby, Yihe Wang, Jonathan Schultz, Chirayu Goswami, Russ |

| | | | |
|---|---|---|---|
| | | | Hager, Robin Grimwood, Clinton R. Paden, Duncan MacCannell |
| EPI_ISL_2283253, EPI_ISL_2283254, EPI_ISL_2283256, EPI_ISL_2283257, EPI_ISL_2283259, EPI_ISL_2283260, EPI_ISL_2283262, EPI_ISL_2283263, EPI_ISL_2283265, EPI_ISL_2283267, EPI_ISL_2283269, EPI_ISL_2283270, EPI_ISL_2283272, EPI_ISL_2283273, EPI_ISL_2283275, EPI_ISL_2283276, EPI_ISL_2283278, EPI_ISL_2283279, EPI_ISL_2283281, EPI_ISL_2283283 | | | |
| see above | Weill Cornell Medicine | New York Genome Center | Michael Zody, Andre Corvelo, Dayna M. Oschwald, Samantha Fennessey, Tom Maniatis, Melissa Cushing, Olivier Elemento, Margaret Elizabeth Ross, Chris Mason, Priya Velu, Hanna Rennert, Arryn Craney, Lars F Westblade |
| EPI_ISL_2283304, EPI_ISL_2283307, EPI_ISL_2283311, EPI_ISL_2283339, EPI_ISL_2283352, EPI_ISL_2283354, EPI_ISL_2283404, EPI_ISL_2283411, EPI_ISL_2283439, EPI_ISL_2283451, EPI_ISL_2283463, EPI_ISL_2283484, EPI_ISL_2283578, EPI_ISL_2283582, EPI_ISL_2283596 | | | |
| see above | Infinity Biologix | Centers for Disease Control and Prevention Division of Viral Diseases, Pathogen Discovery | Dakota Howard, Dhwani Batra, Peter W. Cook, Kara Moser, Adrian Paskey, Jason Caravas, Benjamin Rambo-Martin, Shatavia Morrison, Christopher Gulvick, Scott Sammons, Yvette Unoarumhi, Darlene Wagner, Matthew Schmerer, Christian Bixby, Yihe Wang, Jonathan Schultz, Chirayu Goswami, Russ Hager, Robin Grimwood, Clinton R. Paden, Duncan MacCannell |
| EPI_ISL_2283819, EPI_ISL_2283853, EPI_ISL_2283871 | Aegis Sciences Corporation | Centers for Disease Control and Prevention Division of Viral Diseases, Pathogen Discovery | Dakota Howard, Dhwani Batra, Peter W. Cook, Kara Moser, Adrian Paskey, Jason Caravas, Benjamin Rambo-Martin, Shatavia Morrison, Christopher Gulvick, Scott Sammons, Yvette Unoarumhi, Darlene Wagner, Matthew Schmerer, Cyndi Clark, Patrick Campbell, Rob Case, Vikramsinha Ghorpade, Holly Houdeshell, Ola Kvalvaag, Dillon Nall, Ethan Sanders, Alec Vest, Shaun Westlund, Matthew Hardison, Clinton R. Paden, Duncan MacCannell |
| EPI_ISL_2290768, EPI_ISL_2290769 | MSHS Clinical Microbiology Laboratories | MSHS Pathogen Surveillance Program | Ana S. Gonzalez-Reiche, Hala Alshammary, Mitchell J. Sullivan, Brianne Ciferri, Ajay Obla, Angela Amoako, Mahmoud Awawda, Daniel Floda, Julia Matthews, Ashley Salimbangon, Levy Sominsky, Katherine Beach, Kayla Russo, Charles Gleason, Shelcie Fabre, Giulio Kleiner, Zenab Khan, Bremy Alburquerque, Adriana van de Guchte, Komal Srivastava, Matthew M. Hernandez, Jayeeta Dutta, Denise Jurczyszak, Nancy Francoeur, Betsaida Salom Melo, Irina Oussenko, Gintaras Deikus, Juan Soto, Shwetha Hara Sridhar, Ying-Chih Wang, Kathryn Twyman, Deena R. Altman, Robert Sebra, Adolfo García-Sastre, Marta Luksza, Gopi Patel, Sarah Schaefer, Melissa Gitman, Michael D. Nowak, Alberto Paniz-Mondolfi, Emilia Mia Sordillo, Viviana Simon, Harm van Bakel |
| EPI_ISL_2304502, EPI_ISL_2304509, EPI_ISL_2304536, EPI_ISL_2304551, EPI_ISL_2304567, EPI_ISL_2304583, EPI_ISL_2304584, EPI_ISL_2304594, EPI_ISL_2304610, EPI_ISL_2304647, EPI_ISL_2304649, EPI_ISL_2304653, EPI_ISL_2304656, EPI_ISL_2304659, EPI_ISL_2304663, EPI_ISL_2304667, EPI_ISL_2304669, EPI_ISL_2304672, EPI_ISL_2304673, EPI_ISL_2304675, EPI_ISL_2304678, EPI_ISL_2304682, EPI_ISL_2304685, EPI_ISL_2304687, EPI_ISL_2304697, EPI_ISL_2304701, EPI_ISL_2304702, EPI_ISL_2304708, EPI_ISL_2304709, EPI_ISL_2304715, EPI_ISL_2304718, EPI_ISL_2304733, EPI_ISL_2304735, EPI_ISL_2304746, EPI_ISL_2304750, EPI_ISL_2304753, EPI_ISL_2304754, EPI_ISL_2304759, EPI_ISL_2304760, EPI_ISL_2304764, EPI_ISL_2304768, EPI_ISL_2304769, EPI_ISL_2304794 | | | |
| see above | Quest Diagnostics Incorporated | Centers for Disease Control and Prevention Division of Viral Diseases, Pathogen Discovery | Dakota Howard, Dhwani Batra, Peter W. Cook, Kara Moser, Adrian Paskey, Jason Caravas, Benjamin Rambo-Martin, Shatavia Morrison, Christopher Gulvick, Scott Sammons, Yvette Unoarumhi, Darlene Wagner, Matthew Schmerer, S. H. Rosenthal, A. Gerasimova, R. M. Kagan, B. Anderson, M. Hua, Y. Liu, L.E. Bernstein, K.E. Livingston, A. Perez, I. A. Shlyakhter, R. V. Rolando, R. Owen, P. Tanpaiboon, F. Lacbawan, Clinton R. Paden, Duncan MacCannell |
| EPI_ISL_2305855, EPI_ISL_2305910, EPI_ISL_2305954, EPI_ISL_2305957, EPI_ISL_2306120, EPI_ISL_2306195, EPI_ISL_2306218, EPI_ISL_2306252, EPI_ISL_2306264, EPI_ISL_2306265, EPI_ISL_2306266, EPI_ISL_2306267, EPI_ISL_2306324, EPI_ISL_2306333, EPI_ISL_2306346, EPI_ISL_2306375, EPI_ISL_2306499, EPI_ISL_2306508, EPI_ISL_2306509, EPI_ISL_2306524, EPI_ISL_2306603, EPI_ISL_2306604, EPI_ISL_2306605, EPI_ISL_2306629, EPI_ISL_2306640, EPI_ISL_2306641, EPI_ISL_2306644, EPI_ISL_2306645, EPI_ISL_2306678, EPI_ISL_2306679, EPI_ISL_2306680, EPI_ISL_2306681, EPI_ISL_2306682, EPI_ISL_2306685, EPI_ISL_2306686, EPI_ISL_2306687, EPI_ISL_2306728, EPI_ISL_2306729, EPI_ISL_2306730, EPI_ISL_2306731, EPI_ISL_2306740, EPI_ISL_2306741, EPI_ISL_2306818, EPI_ISL_2306834, EPI_ISL_2306836, EPI_ISL_2306838, EPI_ISL_2306861, EPI_ISL_2306887, EPI_ISL_2306888, EPI_ISL_2306889, EPI_ISL_2306890, EPI_ISL_2306905, EPI_ISL_2306906, EPI_ISL_2306907, EPI_ISL_2306956, EPI_ISL_2306957, EPI_ISL_2306958, EPI_ISL_2306967, EPI_ISL_2306997, EPI_ISL_2307003, EPI_ISL_2307011 | | | |
| see above | Laboratory Corporation of America | Centers for Disease Control and Prevention Division of Viral Diseases, Pathogen Discovery | Dakota Howard, Dhwani Batra, Peter W. Cook, Kara Moser, Adrian Paskey, Jason Caravas, Benjamin Rambo-Martin, Shatavia Morrison, Christopher Gulvick, Scott Sammons, Yvette Unoarumhi, Darlene Wagner, Matthew Schmerer, Minoo Agarwal, Eyad Almasri, Debbie Boles, Ayla Burns, Nuthawin Charoensri, Oren Cohen, Susan Countryman, Mary Ann Cristobal, Bobbi Croy, Suzanne Dale, Hrushikesh Deshmukh, Amanda Douglas, Vincent Drouillon, Marcia Eisenberg, Howard Engler, Rama Ghatti, Prashant Gupta, Susan Hicks, Jake Humphrey, Lax Iyer, Manoj Jain, Mohan Kolli, Brian Krueger, Tim Kuphal, Stanley Letovsky, Michael Levandoski, Craig Lukasik, Jonathan Meltzer, Brian Norvell, Mindy Nye, Scott Parker, Christos Petropoulos, John Pruitt, Steven Ragan, Scott Ryan, Mike Sapeta, Jana Schroth, Suresh Babu Selvaraju, Goran Stevovic, Amanda Suchanek, Andrea Throop, Lyndon Tilson, Thomas Urban, Joe Voshell, Kimberly Wagner, Jonathan Williams, Mary Williamson, Qian Zeng, Tricia Zwiefelhofer, Clinton R. Paden, Duncan MacCannell |
| EPI_ISL_2307143, EPI_ISL_2307160, EPI_ISL_2307161 | Quest Diagnostics Incorporated | Centers for Disease Control and Prevention Division of Viral Diseases, Pathogen Discovery | Dakota Howard, Dhwani Batra, Peter W. Cook, Kara Moser, Adrian Paskey, Jason Caravas, Benjamin Rambo-Martin, Shatavia Morrison, Christopher Gulvick, Scott Sammons, Yvette Unoarumhi, Darlene Wagner, Matthew Schmerer, S. H. Rosenthal, A. Gerasimova, R. M. Kagan, B. Anderson, M. Hua, Y. Liu, L.E. Bernstein, K.E. Livingston, A. Perez, I. A. Shlyakhter, R. V. Rolando, R. Owen, P. Tanpaiboon, F. Lacbawan, Clinton R. Paden, Duncan MacCannell |
| EPI_ISL_2308781 | New York Presbyterian Hospital | Wadsworth Center, New York State Department of Health | Kirsten St. George, Daryl M. Lamson, Alexis Russell, Matthew Shudt, Melissa A Leisner, Jonathan Plitnick, Catharine Prussing, Navjot Singh, John Kelly, Erasmus Schneider, Erica Lasek-Nesselquist |
| EPI_ISL_2308782, EPI_ISL_2308783 | WESTCHESTER MEDICAL CENTER | Wadsworth Center, New York State Department of Health | Kirsten St. George, Daryl M. Lamson, Alexis Russell, Matthew Shudt, Melissa A Leisner, Jonathan Plitnick, Catharine Prussing, Navjot Singh, John Kelly, Erasmus Schneider, Erica Lasek-Nesselquist |
| EPI_ISL_2308784, EPI_ISL_2308785 | MEMORIAL SLOAN KETTERING CANCER CENTER | Wadsworth Center, New York State Department of Health | Kirsten St. George, Daryl M. Lamson, Alexis Russell, Matthew Shudt, Melissa A Leisner, Jonathan Plitnick, Catharine Prussing, Navjot Singh, John Kelly, Erasmus Schneider, Erica Lasek-Nesselquist |
| EPI_ISL_2308786 | Columbia University Irving Medical Center | Wadsworth Center, New York State Department of Health | Kirsten St. George, Daryl M. Lamson, Alexis Russell, Matthew Shudt, Melissa A Leisner, Jonathan Plitnick, Catharine Prussing, Navjot Singh, John Kelly, Erasmus Schneider, Erica Lasek-Nesselquist |
| EPI_ISL_2308787 | NYC Pandemic Response Lab | Wadsworth Center, New York State Department of Health | Kirsten St. George, Daryl M. Lamson, Alexis Russell, Matthew Shudt, Melissa A Leisner, Jonathan Plitnick, Catharine Prussing, Navjot Singh, John Kelly, Erasmus Schneider, Erica Lasek-Nesselquist |
| EPI_ISL_2308788, EPI_ISL_2308789 | MONTEFIORE MEDICAL CENTER LABORATORIES | Wadsworth Center, New York State Department of Health | Kirsten St. George, Daryl M. Lamson, Alexis Russell, Matthew Shudt, Melissa A Leisner, Jonathan Plitnick, Catharine Prussing, Navjot Singh, John Kelly, Erasmus Schneider, Erica Lasek-Nesselquist |
| EPI_ISL_2308790, EPI_ISL_2308791 | URMC LABS | Wadsworth Center, New York State Department of Health | Kirsten St. George, Daryl M. Lamson, Alexis Russell, Matthew Shudt, Melissa A Leisner, Jonathan Plitnick, Catharine Prussing, Navjot Singh, John Kelly, Erasmus Schneider, Erica Lasek-Nesselquist |
| EPI_ISL_2308792 | WESTCHESTER MEDICAL CENTER | Wadsworth Center, New York State Department of Health | Kirsten St. George, Daryl M. Lamson, Alexis Russell, Matthew Shudt, Melissa A Leisner, Jonathan Plitnick, Catharine Prussing, Navjot Singh, John Kelly, Erasmus Schneider, Erica Lasek-Nesselquist |
| EPI_ISL_2308793, EPI_ISL_2308794 | URMC LABS | Wadsworth Center, New York State Department of Health | Kirsten St. George, Daryl M. Lamson, Alexis Russell, Matthew Shudt, Melissa A Leisner, Jonathan Plitnick, Catharine Prussing, Navjot Singh, John Kelly, Erasmus Schneider, Erica Lasek-Nesselquist |
| EPI_ISL_2308795, EPI_ISL_2308796, EPI_ISL_2308797, EPI_ISL_2308798 | NYC Pandemic Response Lab | Wadsworth Center, New York State Department of Health | Kirsten St. George, Daryl M. Lamson, Alexis Russell, Matthew Shudt, Melissa A Leisner, Jonathan Plitnick, Catharine Prussing, Navjot Singh, John Kelly, Erasmus Schneider, Erica Lasek-Nesselquist |
| EPI_ISL_2308799, EPI_ISL_2308800, EPI_ISL_2308801 | URMC LABS | Wadsworth Center, New York State Department of Health | Kirsten St. George, Daryl M. Lamson, Alexis Russell, Matthew Shudt, Melissa A Leisner, Jonathan Plitnick, Catharine Prussing, Navjot Singh, John Kelly, Erasmus Schneider, Erica Lasek-Nesselquist |
| EPI_ISL_2308802 | ALBANY MEDICAL CENTER | Wadsworth Center, New York State Department of Health | Kirsten St. George, Daryl M. Lamson, Alexis Russell, Matthew Shudt, Melissa A Leisner, Jonathan Plitnick, Catharine Prussing, Navjot Singh, John Kelly, Erasmus Schneider, Erica Lasek-Nesselquist |
| EPI_ISL_2308803, EPI_ISL_2308804, EPI_ISL_2308805, EPI_ISL_2308806 | URMC LABS | Wadsworth Center, New York State Department of Health | Kirsten St. George, Daryl M. Lamson, Alexis Russell, Matthew Shudt, Melissa A Leisner, Jonathan Plitnick, Catharine Prussing, Navjot Singh, John Kelly, Erasmus Schneider, Erica Lasek-Nesselquist |
| EPI_ISL_2312443 | Broad Institute Clinical Research Sequencing Platform | Infectious Disease Program, Broad Institute of Harvard and MIT | Siddle,K.J., Adams,G., Pearlman,L., Gladden-Young,A., Vicente,G., Blumenstiel,B., DeFelice,M., Lee,M., McGovern,S., Lagerborg,K., Rudy,M., DeRuff,K., Carter,A., Normandin,E., Bauer,M., Reilly,S., Tomkins-Tinch,C., Loreth,C., Chaluvadi,S., Meldrim,J., Granger,B., Lemieux,J.E., Birren,B.W., Sabeti,P.C., Larkin,K., Dodge,S., Lennon,N., Madoff,L., Brown,C., Gallagher,G., Smole,S., Park,D.J., Gabriel,S., and MacInnis,B.L. |
| EPI_ISL_2313517, EPI_ISL_2313606, EPI_ISL_2313617, EPI_ISL_2313633, EPI_ISL_2313635, EPI_ISL_2313637, EPI_ISL_2313667, EPI_ISL_2313705, EPI_ISL_2313707, EPI_ISL_2313797, EPI_ISL_2313844 | | | |
| see above | Respiratory Viruses Branch, Centers for Disease Control and Prevention | Respiratory Viruses Branch, Centers for Disease Control and Prevention | Howard,D., Batra,D., Cook,P.W., Moser,K., Paskey,A., Caravas,J., Rambo-Martin,B., Morrison,S., Gulvick,C., Sammons,S., Unoarumhi,Y., Wagner,D., Schmerer,M., Bixby,C., Wang,Y., Schultz,J., Goswami,C., Hager,R., Grimwood,R., Paden,C.R., MacCannell,D. |
| EPI_ISL_2320386, EPI_ISL_2320727, | Helix/Illumina | Centers for Disease Control and Prevention Division of Viral | Dakota Howard, Dhwani Batra, Peter W. Cook, Kara Moser, Adrian Paskey, Jason Caravas, Benjamin Rambo-Martin, Shatavia Morrison, Christopher |

| | | | |
|---|---|---|---|
| EPI_ISL_2321100 | | Diseases, Pathogen Discovery | Gulvick, Scott Sammons, Yvette Unoarumhi, Darlene Wagner, Matthew Schmerer, Eileen de Feo, Jan Antico, Christine Tran, Matthew Tolentino, Shannon Wickline, Kim Gietzen, Brad Sickler, Jingtao Liu, Eric Allen, Phil Febbo, Nicole L. Washington, Simon White, Geraint Levan, Kelly Schiabor Barrett, Elizabeth Cirulli, Alexandre Bolze, Ary Ascencio, Charlotte Rivera-Garcia, Ryan Cho, Jason Nguyen, Sherry Wang, Jimmy Ramirez, Tyler Cassens, Efren Sandoval, Magnus Isaksson, William Lee, David Becker, Marc Laurent, James Lu, Clinton R. Paden, Duncan MacCannell |
| EPI_ISL_2321336, EPI_ISL_2321345 | Aegis Sciences Corporation | Centers for Disease Control and Prevention Division of Viral Diseases, Pathogen Discovery | Dakota Howard, Dhwani Batra, Peter W. Cook, Kara Moser, Adrian Paskey, Jason Caravas, Benjamin Rambo-Martin, Shatavia Morrison, Christopher Gulvick, Scott Sammons, Yvette Unoarumhi, Darlene Wagner, Matthew Schmerer, Cyndi Clark, Patrick Campbell, Rob Case, Vikramsinha Ghorpade, Holly Houdeshell, Ola Kvalvaag, Dillon Nall, Ethan Sanders, Alec Vest, Shaun Westlund, Matthew Hardison, Clinton R. Paden, Duncan MacCannell |
| EPI_ISL_2322900, EPI_ISL_2322901 | DOHMH Morrisania | New York City Public Health Laboratory | Jade Wang, et al. |
| EPI_ISL_2322902 | DOHMH Fort Greene | New York City Public Health Laboratory | Jade Wang, et al. |
| EPI_ISL_2322903, EPI_ISL_2322904, EPI_ISL_2322905 | DOHMH Corona | New York City Public Health Laboratory | Jade Wang, et al. |
| EPI_ISL_2322906, EPI_ISL_2322907 | DOHMH Crown Heights | New York City Public Health Laboratory | Jade Wang, et al. |
| EPI_ISL_2322908, EPI_ISL_2322909, EPI_ISL_2322910 | DOHMH PHL | New York City Public Health Laboratory | Jade Wang, et al. |
| EPI_ISL_2322911 | DOHMH Fort Greene | New York City Public Health Laboratory | Jade Wang, et al. |
| EPI_ISL_2322912, EPI_ISL_2322913 | DOHMH Central Harlem | New York City Public Health Laboratory | Jade Wang, et al. |
| EPI_ISL_2322914, EPI_ISL_2322915, EPI_ISL_2322916, EPI_ISL_2322917, EPI_ISL_2322918 | DOHMH Jamaica | New York City Public Health Laboratory | Jade Wang, et al. |
| EPI_ISL_2322919 | DOHMH Riverside | New York City Public Health Laboratory | Jade Wang, et al. |
| EPI_ISL_2322920 | OCME Office Of Chief Medical Examiner | New York City Public Health Laboratory | Jade Wang, et al. |
| EPI_ISL_2324063, EPI_ISL_2324139, EPI_ISL_2324144, EPI_ISL_2324145, EPI_ISL_2324151, EPI_ISL_2324153, EPI_ISL_2324168 | Infinity Biologix | Centers for Disease Control and Prevention Division of Viral Diseases, Pathogen Discovery | Dakota Howard, Dhwani Batra, Peter W. Cook, Kara Moser, Adrian Paskey, Jason Caravas, Benjamin Rambo-Martin, Shatavia Morrison, Christopher Gulvick, Scott Sammons, Yvette Unoarumhi, Darlene Wagner, Matthew Schmerer, Christian Bixby, Yihe Wang, Jonathan Schultz, Chirayu Goswami, Russ Hager, Robin Grimwood, Clinton R. Paden, Duncan MacCannell |
| EPI_ISL_2324479, EPI_ISL_2324520, EPI_ISL_2324556, EPI_ISL_2324582, EPI_ISL_2324589 | Laboratory Corporation of America | Centers for Disease Control and Prevention Division of Viral Diseases, Pathogen Discovery | Dakota Howard, Dhwani Batra, Peter W. Cook, Kara Moser, Adrian Paskey, Jason Caravas, Benjamin Rambo-Martin, Shatavia Morrison, Christopher Gulvick, Scott Sammons, Yvette Unoarumhi, Darlene Wagner, Matthew Schmerer, Minoo Agarwal, Eyad Almasri, Debbie Boles, Ayla Burns, Nuthawin Charoensri, Oren Cohen, Susan Countryman, Mary Ann Cristobal, Bobbi Croy, Suzanne Dale, Hrushikesh Deshmukh, Amanda Douglas, Vincent Drouillon, Marcia Eisenberg, Howard Engler, Rama Ghatti, Prashant Gupta, Susan Hicks, Jake Humphrey, Lax Iyer, Lisa Pfefferle, Manoj Jain, Matthew Robinson, Mohan Kolli, Brian Krueger, Tim Kuphal, Stanley Letovsky, Michael Levandoski, Craig Lukasik, Jonathan Meltzer, Brian Norvell, Mindy Nye, Scott Parker, Christos Petropoulos, John Pruitt, Steven Ragan, Scott Ryan, Mike Sapeta, Jana Schroth, Suresh Babu Selvaraju, Goran Stevovic, Amanda Suchanek, Andrea Throop, Lyndon Tilson, Thomas Urban, Joe Voshell, Kimberly Wagner, Jonathan Williams, Mary Williamson, Qian Zeng, Tricia Zwiefelhofer, Clinton R. Paden, Duncan MacCannell |
| EPI_ISL_2349553, EPI_ISL_2349554, EPI_ISL_2349555, EPI_ISL_2349556, EPI_ISL_2349557, EPI_ISL_2349558, EPI_ISL_2349559, EPI_ISL_2349560, EPI_ISL_2349561, EPI_ISL_2349562, EPI_ISL_2349563, EPI_ISL_2349564, EPI_ISL_2349565, EPI_ISL_2349566, EPI_ISL_2349567, EPI_ISL_2349568, EPI_ISL_2349569, EPI_ISL_2349570, EPI_ISL_2349571, EPI_ISL_2349572, EPI_ISL_2349573, EPI_ISL_2349574, EPI_ISL_2349575, EPI_ISL_2349576, EPI_ISL_2349577, EPI_ISL_2349578, EPI_ISL_2349579, EPI_ISL_2349580, EPI_ISL_2349581, EPI_ISL_2349582, EPI_ISL_2349583, EPI_ISL_2349584, EPI_ISL_2349585, EPI_ISL_2349586, EPI_ISL_2349587, EPI_ISL_2349588, EPI_ISL_2349589, EPI_ISL_2349590, EPI_ISL_2349591, EPI_ISL_2349592, EPI_ISL_2349593, EPI_ISL_2349594, EPI_ISL_2349595, EPI_ISL_2349596, EPI_ISL_2349597, EPI_ISL_2349598, EPI_ISL_2349599, EPI_ISL_2349600, EPI_ISL_2349601, EPI_ISL_2349602, EPI_ISL_2349603, EPI_ISL_2349604, EPI_ISL_2349605, EPI_ISL_2349606, EPI_ISL_2349607, EPI_ISL_2349608, EPI_ISL_2349609, EPI_ISL_2349610, EPI_ISL_2349611, EPI_ISL_2349612, EPI_ISL_2349613, EPI_ISL_2349614, EPI_ISL_2349615, EPI_ISL_2349616, EPI_ISL_2349617, EPI_ISL_2349618, EPI_ISL_2349619, EPI_ISL_2349620, EPI_ISL_2349621, EPI_ISL_2349622, EPI_ISL_2349623, EPI_ISL_2349624, EPI_ISL_2349625, EPI_ISL_2349626, EPI_ISL_2349627, EPI_ISL_2349628, EPI_ISL_2349629, EPI_ISL_2349630, EPI_ISL_2349631, EPI_ISL_2349632, EPI_ISL_2349633, EPI_ISL_2349634, EPI_ISL_2349635, EPI_ISL_2349636, EPI_ISL_2349637, EPI_ISL_2349638, EPI_ISL_2349639, EPI_ISL_2349640, EPI_ISL_2349641, EPI_ISL_2349642, EPI_ISL_2349643, EPI_ISL_2349644, EPI_ISL_2349645, EPI_ISL_2349646, EPI_ISL_2349647, EPI_ISL_2349648, EPI_ISL_2349649, EPI_ISL_2349650, EPI_ISL_2349651, EPI_ISL_2349652, EPI_ISL_2349653, EPI_ISL_2349654, EPI_ISL_2349655, EPI_ISL_2349656, EPI_ISL_2349657, EPI_ISL_2349658, EPI_ISL_2349659, EPI_ISL_2349660, EPI_ISL_2349661, EPI_ISL_2349662, EPI_ISL_2349663, EPI_ISL_2349664, EPI_ISL_2349665, EPI_ISL_2349666, EPI_ISL_2349667, EPI_ISL_2349668, EPI_ISL_2349669, EPI_ISL_2349670, EPI_ISL_2349671, EPI_ISL_2349672, EPI_ISL_2349673, EPI_ISL_2349674, EPI_ISL_2349675, EPI_ISL_2349676, EPI_ISL_2349677, EPI_ISL_2349678, EPI_ISL_2349679, EPI_ISL_2349680, EPI_ISL_2349681, EPI_ISL_2349682, EPI_ISL_2349683, EPI_ISL_2349684, EPI_ISL_2349685, EPI_ISL_2349686, EPI_ISL_2349687, EPI_ISL_2349688, EPI_ISL_2349689, EPI_ISL_2349690, EPI_ISL_2349691 | | | |
| see above | Pandemic Response Lab - NYC | Pandemic Response Lab, R&D | Henry Lee, Michael Hammerling, Melissa Hopkins, Cybill del Castillo, Shinyoung Clair Kang, William Ward, Pradeep Bugga, Sol Rey, Dylan Law, Katharine Nelson, Haiping Hao, Jon Laurent |
| EPI_ISL_2362354 | Flow Health | Infectious Disease Program, Broad Institute of Harvard and MIT | Lemieux,J.E., Siddle,K.J., Adams,G., Gladden-Young,A., Lagerborg,K., Rudy,M., DeRuff,K., Carter,A., Normandin,E., Bauer,M., Reilly,S., Tomkins-Tinch,C., Loreth,C., Chaluvadi,S., Birren,B.W., Gallagher,G., Smole,S., Park,D.J., MacInnis,B.L., and Sabeti,P.C. |
| EPI_ISL_2367700, EPI_ISL_2367705, EPI_ISL_2367724, EPI_ISL_2367756, EPI_ISL_2367764, EPI_ISL_2367801, EPI_ISL_2367829, EPI_ISL_2367864, EPI_ISL_2367896, EPI_ISL_2367922, EPI_ISL_2367931 | | | |
| see above | Quest Diagnostics Incorporated | Centers for Disease Control and Prevention Division of Viral Diseases, Pathogen Discovery | Dakota Howard, Dhwani Batra, Peter W. Cook, Kara Moser, Adrian Paskey, Jason Caravas, Benjamin Rambo-Martin, Shatavia Morrison, Christopher Gulvick, Scott Sammons, Yvette Unoarumhi, Darlene Wagner, Matthew Schmerer, S. H. Rosenthal, A. Gerasimova, R. M. Kagan, B. Anderson, M. Hua, Y. Liu, L.E. Bernstein, K.E. Livingston, A. Perez, I. A. Shlyakhter, R. V. Rolando, R. Owen, P. Tanpaiboon, F. Lacbawan, Clinton R. Paden, Duncan MacCannell |
| EPI_ISL_2368228, EPI_ISL_2368236, EPI_ISL_2368304, EPI_ISL_2368318, EPI_ISL_2368343, EPI_ISL_2368344, EPI_ISL_2368351, EPI_ISL_2368363, EPI_ISL_2368364, EPI_ISL_2368485, EPI_ISL_2368507, EPI_ISL_2368531, EPI_ISL_2368569, EPI_ISL_2368727, EPI_ISL_2368942, EPI_ISL_2368954 | | | |
| see above | Aegis Sciences Corporation | Centers for Disease Control and Prevention Division of Viral Diseases, Pathogen Discovery | Dakota Howard, Dhwani Batra, Peter W. Cook, Kara Moser, Adrian Paskey, Jason Caravas, Benjamin Rambo-Martin, Shatavia Morrison, Christopher Gulvick, Scott Sammons, Yvette Unoarumhi, Darlene Wagner, Matthew Schmerer, Cyndi Clark, Patrick Campbell, Rob Case, Vikramsinha Ghorpade, Holly Houdeshell, Ola Kvalvaag, Dillon Nall, Ethan Sanders, Alec Vest, Shaun Westlund, Matthew Hardison, Clinton R. Paden, Duncan MacCannell |
| EPI_ISL_2369028, EPI_ISL_2369070, EPI_ISL_2369106, EPI_ISL_2369116, EPI_ISL_2369158, EPI_ISL_2369161, EPI_ISL_2369170, EPI_ISL_2369186, EPI_ISL_2369200, EPI_ISL_2369203, EPI_ISL_2369228, EPI_ISL_2369231, EPI_ISL_2369255, EPI_ISL_2369258, EPI_ISL_2369276, EPI_ISL_2369296, EPI_ISL_2369360, EPI_ISL_2369384 | | | |
| see above | Quest Diagnostics Incorporated | Centers for Disease Control and Prevention Division of Viral Diseases, Pathogen Discovery | Dakota Howard, Dhwani Batra, Peter W. Cook, Kara Moser, Adrian Paskey, Jason Caravas, Benjamin Rambo-Martin, Shatavia Morrison, Christopher Gulvick, Scott Sammons, Yvette Unoarumhi, Darlene Wagner, Matthew Schmerer, S. H. Rosenthal, A. Gerasimova, R. M. Kagan, B. Anderson, M. Hua, Y. Liu, L.E. Bernstein, K.E. Livingston, A. Perez, I. A. Shlyakhter, R. V. Rolando, R. Owen, P. Tanpaiboon, F. Lacbawan, Clinton R. Paden, Duncan MacCannell |
| EPI_ISL_2369952, EPI_ISL_2369953, EPI_ISL_2369979, EPI_ISL_2370004, EPI_ISL_2370026, EPI_ISL_2370211, EPI_ISL_2370609, EPI_ISL_2370630, EPI_ISL_2370664, EPI_ISL_2370675, EPI_ISL_2370678, EPI_ISL_2370716 | | | |
| see above | Aegis Sciences Corporation | Centers for Disease Control and Prevention Division of Viral Diseases, Pathogen Discovery | Dakota Howard, Dhwani Batra, Peter W. Cook, Kara Moser, Adrian Paskey, Jason Caravas, Benjamin Rambo-Martin, Shatavia Morrison, Christopher Gulvick, Scott Sammons, Yvette Unoarumhi, Darlene Wagner, Matthew Schmerer, Cyndi Clark, Patrick Campbell, Rob Case, Vikramsinha Ghorpade, Holly Houdeshell, Ola Kvalvaag, Dillon Nall, Ethan Sanders, Alec Vest, Shaun Westlund, Matthew Hardison, Clinton R. Paden, Duncan MacCannell |
| EPI_ISL_2371692, EPI_ISL_2371698, EPI_ISL_2371883, EPI_ISL_2371902, EPI_ISL_2371913, EPI_ISL_2371914, EPI_ISL_2371921, EPI_ISL_2371964, EPI_ISL_2371985, EPI_ISL_2371995, EPI_ISL_2371996, EPI_ISL_2371997, EPI_ISL_2371998, EPI_ISL_2372030, EPI_ISL_2372040, EPI_ISL_2372042, EPI_ISL_2372043, EPI_ISL_2372056, EPI_ISL_2372060, EPI_ISL_2372062, EPI_ISL_2372077, EPI_ISL_2372078, EPI_ISL_2372096, EPI_ISL_2372100, EPI_ISL_2372102, EPI_ISL_2372103, EPI_ISL_2372108, EPI_ISL_2372109, EPI_ISL_2372110, EPI_ISL_2372124, EPI_ISL_2372125, EPI_ISL_2372133, EPI_ISL_2372134, EPI_ISL_2372138 | | | |
| see above | Quest Diagnostics Incorporated | Centers for Disease Control and Prevention Division of Viral Diseases, Pathogen Discovery | Dakota Howard, Dhwani Batra, Peter W. Cook, Kara Moser, Adrian Paskey, Jason Caravas, Benjamin Rambo-Martin, Shatavia Morrison, Christopher Gulvick, Scott Sammons, Yvette Unoarumhi, Darlene Wagner, Matthew Schmerer, S. H. Rosenthal, A. Gerasimova, R. M. Kagan, B. Anderson, M. Hua, Y. Liu, L.E. Bernstein, K.E. Livingston, A. Perez, I. A. Shlyakhter, R. V. Rolando, R. Owen, P. Tanpaiboon, F. Lacbawan, Clinton R. Paden, Duncan MacCannell |
| EPI_ISL_2372335, EPI_ISL_2372364, EPI_ISL_2372366, EPI_ISL_2372393 | Aegis Sciences Corporation | Centers for Disease Control and Prevention Division of Viral Diseases, Pathogen Discovery | Dakota Howard, Dhwani Batra, Peter W. Cook, Kara Moser, Adrian Paskey, Jason Caravas, Benjamin Rambo-Martin, Shatavia Morrison, Christopher Gulvick, Scott Sammons, Yvette Unoarumhi, Darlene Wagner, Matthew Schmerer, Cyndi Clark, Patrick Campbell, Rob Case, Vikramsinha Ghorpade, Holly Houdeshell, Ola Kvalvaag, Dillon Nall, Ethan Sanders, Alec Vest, Shaun Westlund, Matthew Hardison, Clinton R. Paden, Duncan MacCannell |

| | | | |
|---|---|---|---|
| EPI_ISL_2372422 | Quest Diagnostics Incorporated | Centers for Disease Control and Prevention Division of Viral Diseases, Pathogen Discovery | Dakota Howard, Dhwani Batra, Peter W. Cook, Kara Moser, Adrian Paskey, Jason Caravas, Benjamin Rambo-Martin, Shatavia Morrison, Christopher Gulvick, Scott Sammons, Yvette Unoarumhi, Darlene Wagner, Matthew Schmerer, S. H. Rosenthal, A. Gerasimova, R. M. Kagan, B. Anderson, M. Hua, Y. Liu, L.E. Bernstein, K.E. Livingston, A. Perez, I. A. Shlyakhter, R. V. Rolando, R. Owen, P. Tanpaiboon, F. Lacbawan, Clinton R. Paden, Duncan MacCannell |
| EPI_ISL_2372425, EPI_ISL_2372436, EPI_ISL_2372456, EPI_ISL_2372459 | Aegis Sciences Corporation | Centers for Disease Control and Prevention Division of Viral Diseases, Pathogen Discovery | Dakota Howard, Dhwani Batra, Peter W. Cook, Kara Moser, Adrian Paskey, Jason Caravas, Benjamin Rambo-Martin, Shatavia Morrison, Christopher Gulvick, Scott Sammons, Yvette Unoarumhi, Darlene Wagner, Matthew Schmerer, Cyndi Clark, Patrick Campbell, Rob Case, Vikramsinha Ghorpade, Holly Houdeshell, Ola Kvalvaag, Dillon Nall, Ethan Sanders, Alec Vest, Shaun Westlund, Matthew Hardison, Clinton R. Paden, Duncan MacCannell |
| EPI_ISL_2372802 | Infinity Biologix | Centers for Disease Control and Prevention Division of Viral Diseases, Pathogen Discovery | Dakota Howard, Dhwani Batra, Peter W. Cook, Kara Moser, Adrian Paskey, Jason Caravas, Benjamin Rambo-Martin, Shatavia Morrison, Christopher Gulvick, Scott Sammons, Yvette Unoarumhi, Darlene Wagner, Matthew Schmerer, Christian Bixby, Yihe Wang, Jonathan Schultz, Chirayu Goswami, Russ Hager, Robin Grimwood, Clinton R. Paden, Duncan MacCannell |
| EPI_ISL_2372929, EPI_ISL_2372932 | Quest Diagnostics Incorporated | Centers for Disease Control and Prevention Division of Viral Diseases, Pathogen Discovery | Dakota Howard, Dhwani Batra, Peter W. Cook, Kara Moser, Adrian Paskey, Jason Caravas, Benjamin Rambo-Martin, Shatavia Morrison, Christopher Gulvick, Scott Sammons, Yvette Unoarumhi, Darlene Wagner, Matthew Schmerer, S. H. Rosenthal, A. Gerasimova, R. M. Kagan, B. Anderson, M. Hua, Y. Liu, L.E. Bernstein, K.E. Livingston, A. Perez, I. A. Shlyakhter, R. V. Rolando, R. Owen, P. Tanpaiboon, F. Lacbawan, Clinton R. Paden, Duncan MacCannell |
| EPI_ISL_2375782 | Infinity Biologix | Centers for Disease Control and Prevention Division of Viral Diseases, Pathogen Discovery | Dakota Howard, Dhwani Batra, Peter W. Cook, Kara Moser, Adrian Paskey, Jason Caravas, Benjamin Rambo-Martin, Shatavia Morrison, Christopher Gulvick, Scott Sammons, Yvette Unoarumhi, Darlene Wagner, Matthew Schmerer, Christian Bixby, Yihe Wang, Jonathan Schultz, Chirayu Goswami, Russ Hager, Robin Grimwood, Clinton R. Paden, Duncan MacCannell |
| EPI_ISL_2388777 | URMC LABS | URMC LABS | Dwight Hardy, Andrew Cameron, Stepan Voljehn, Nicole Pecora |
| EPI_ISL_2397416, EPI_ISL_2397433, EPI_ISL_2397439, EPI_ISL_2397459, EPI_ISL_2397464, EPI_ISL_2397475, EPI_ISL_2397493, EPI_ISL_2397509, EPI_ISL_2397546, EPI_ISL_2397563, EPI_ISL_2397572, EPI_ISL_2397577, EPI_ISL_2397605, EPI_ISL_2397606, EPI_ISL_2397630, EPI_ISL_2397635, EPI_ISL_2397640, EPI_ISL_2397648, EPI_ISL_2397668, EPI_ISL_2397696 | | | |
| see above | Quest Diagnostics Incorporated | Centers for Disease Control and Prevention Division of Viral Diseases, Pathogen Discovery | Dakota Howard, Dhwani Batra, Peter W. Cook, Kara Moser, Adrian Paskey, Jason Caravas, Benjamin Rambo-Martin, Shatavia Morrison, Christopher Gulvick, Scott Sammons, Yvette Unoarumhi, Darlene Wagner, Matthew Schmerer, S. H. Rosenthal, A. Gerasimova, R. M. Kagan, B. Anderson, M. Hua, Y. Liu, L.E. Bernstein, K.E. Livingston, A. Perez, I. A. Shlyakhter, R. V. Rolando, R. Owen, P. Tanpaiboon, F. Lacbawan, Clinton R. Paden, Duncan MacCannell |
| EPI_ISL_2398174, EPI_ISL_2398178, EPI_ISL_2398186, EPI_ISL_2398188, EPI_ISL_2398193, EPI_ISL_2398265, EPI_ISL_2398267, EPI_ISL_2398269, EPI_ISL_2398328, EPI_ISL_2398336, EPI_ISL_2398346, EPI_ISL_2398347, EPI_ISL_2398358, EPI_ISL_2398359, EPI_ISL_2398404, EPI_ISL_2398405, EPI_ISL_2398408, EPI_ISL_2398450, EPI_ISL_2398451, EPI_ISL_2398452, EPI_ISL_2398471, EPI_ISL_2398536, EPI_ISL_2398545, EPI_ISL_2398550, EPI_ISL_2398569, EPI_ISL_2398572, EPI_ISL_2398787, EPI_ISL_2398878, EPI_ISL_2398903, EPI_ISL_2398908, EPI_ISL_2398909, EPI_ISL_2398910, EPI_ISL_2399022, EPI_ISL_2399024, EPI_ISL_2399026, EPI_ISL_2399028, EPI_ISL_2399030, EPI_ISL_2399035, EPI_ISL_2399036, EPI_ISL_2399038, EPI_ISL_2399073, EPI_ISL_2399074, EPI_ISL_2399085, EPI_ISL_2399087, EPI_ISL_2399088, EPI_ISL_2399089, EPI_ISL_2399090, EPI_ISL_2399091, EPI_ISL_2399092, EPI_ISL_2399093, EPI_ISL_2399094, EPI_ISL_2399095, EPI_ISL_2399248, EPI_ISL_2399264, EPI_ISL_2399271, EPI_ISL_2399273, EPI_ISL_2399274, EPI_ISL_2399275, EPI_ISL_2399276, EPI_ISL_2399277, EPI_ISL_2399278, EPI_ISL_2399279, EPI_ISL_2399280, EPI_ISL_2399290, EPI_ISL_2399295, EPI_ISL_2399297, EPI_ISL_2399327, EPI_ISL_2399338, EPI_ISL_2399339, EPI_ISL_2399340 | | | |
| see above | Laboratory Corporation of America | Centers for Disease Control and Prevention Division of Viral Diseases, Pathogen Discovery | Dakota Howard, Dhwani Batra, Peter W. Cook, Kara Moser, Adrian Paskey, Jason Caravas, Benjamin Rambo-Martin, Shatavia Morrison, Christopher Gulvick, Scott Sammons, Yvette Unoarumhi, Darlene Wagner, Matthew Schmerer, Minoo Agarwal, Eyad Almasri, Debbie Boles, Ayla Burns, Nuthawin Charoensri, Oren Cohen, Susan Countryman, Mary Ann Cristobal, Bobbi Croy, Suzanne Dale, Hrushikesh Deshmukh, Amanda Douglas, Vincent Drouillon, Marcia Eisenberg, Howard Engler, Rama Ghatti, Prashant Gupta, Susan Hicks, Jake Humphrey, Lax Iyer, Lisa Pfefferle, Manoj Jain, Matthew Robinson, Mohan Kolli, Brian Krueger, Tim Kuphal, Stanley Letovsky, Michael Levandoski, Craig Lukasik, Jonathan Meltzer, Brian Norvell, Mindy Nye, Scott Parker, Christos Petropoulos, John Pruitt, Steven Ragan, Scott Ryan, Mike Sapeta, Jana Schroth, Suresh Babu Selvaraju, Goran Stevovic, Amanda Suchanek, Andrea Throop, Lyndon Tilson, Thomas Urban, Joe Voshell, Kimberly Wagner, Jonathan Williams, Mary Williamson, Qian Zeng, Tricia Zwiefelhofer, Clinton R. Paden, Duncan MacCannell |
| EPI_ISL_2399475 | Quest Diagnostics Incorporated | Centers for Disease Control and Prevention Division of Viral Diseases, Pathogen Discovery | Dakota Howard, Dhwani Batra, Peter W. Cook, Kara Moser, Adrian Paskey, Jason Caravas, Benjamin Rambo-Martin, Shatavia Morrison, Christopher Gulvick, Scott Sammons, Yvette Unoarumhi, Darlene Wagner, Matthew Schmerer, S. H. Rosenthal, A. Gerasimova, R. M. Kagan, B. Anderson, M. Hua, Y. Liu, L.E. Bernstein, K.E. Livingston, A. Perez, I. A. Shlyakhter, R. V. Rolando, R. Owen, P. Tanpaiboon, F. Lacbawan, Clinton R. Paden, Duncan MacCannell |
| EPI_ISL_2400287, EPI_ISL_2400336, EPI_ISL_2400382 | NYU Langone Health | Departments of Pathology and Medicine, New York University School of Medicine | Adriana Heguy, Dacia Dimartino, Emily Guzman, Christian Marier, Peter Meyn, Sitharam Ramaswami, Gael Westby, Paul Zappile, Yutong Zhang, Paolo Cotzia, Guiqing Wang |
| EPI_ISL_2400425 | DOHMH Morrisania | New York City Public Health Laboratory | Jade Wang, et al. |
| EPI_ISL_2400426 | DOHMH Chelsea | New York City Public Health Laboratory | Jade Wang, et al. |
| EPI_ISL_2400427 | Department of Homeless Services | New York City Public Health Laboratory | Jade Wang, et al. |
| EPI_ISL_2400428, EPI_ISL_2400429 | OCME Office Of Chief Medical Examiner | New York City Public Health Laboratory | Jade Wang, et al. |
| EPI_ISL_2400430 | DOHMH Crown Heights | New York City Public Health Laboratory | Jade Wang, et al. |
| EPI_ISL_2400431 | DOHMH PHL | New York City Public Health Laboratory | Jade Wang, et al. |
| EPI_ISL_2400432 | DOHMH Central Harlem | New York City Public Health Laboratory | Jade Wang, et al. |
| EPI_ISL_2400433 | DOHMH Morrisania | New York City Public Health Laboratory | Jade Wang, et al. |
| EPI_ISL_2400434, EPI_ISL_2400435 | DOHMH Corona | New York City Public Health Laboratory | Jade Wang, et al. |
| EPI_ISL_2400436 | DOHMH Chelsea | New York City Public Health Laboratory | Jade Wang, et al. |
| EPI_ISL_2400442 | OCME Office Of Chief Medical Examiner | New York City Public Health Laboratory | Jade Wang, et al. |
| EPI_ISL_2400443, EPI_ISL_2400444, EPI_ISL_2400445, EPI_ISL_2400446, EPI_ISL_2400447, EPI_ISL_2400448, EPI_ISL_2400449, EPI_ISL_2400450, EPI_ISL_2400451, EPI_ISL_2400452, EPI_ISL_2400453, EPI_ISL_2400454, EPI_ISL_2400455, EPI_ISL_2400456, EPI_ISL_2400457, EPI_ISL_2400458, EPI_ISL_2400459 | | | |
| see above | Molecular Biology Laboratory, Westchester Medical Center | Genomics Core Laboratory, New York Medical College | Weihua Huang, Utsav Pandey, Jian Zhuge, Changhong Yin |
| EPI_ISL_2402813, EPI_ISL_2402814, EPI_ISL_2402815, EPI_ISL_2402816, EPI_ISL_2402817, EPI_ISL_2402818, EPI_ISL_2402819, EPI_ISL_2402820, EPI_ISL_2402821, EPI_ISL_2402822, EPI_ISL_2402823, EPI_ISL_2402824, EPI_ISL_2402825, EPI_ISL_2402826, EPI_ISL_2402827, EPI_ISL_2402828, EPI_ISL_2402829, EPI_ISL_2402830, EPI_ISL_2402831, EPI_ISL_2402832, EPI_ISL_2402833, EPI_ISL_2402834, EPI_ISL_2402835, EPI_ISL_2402836, EPI_ISL_2402837, EPI_ISL_2402838, EPI_ISL_2402839, EPI_ISL_2402840, EPI_ISL_2402841, EPI_ISL_2402842, EPI_ISL_2402843, EPI_ISL_2402844, EPI_ISL_2402845, EPI_ISL_2402846, EPI_ISL_2402847, EPI_ISL_2402848, EPI_ISL_2402849, EPI_ISL_2402850, EPI_ISL_2402851, EPI_ISL_2402852, EPI_ISL_2402853, EPI_ISL_2402854, EPI_ISL_2402855, EPI_ISL_2402856, EPI_ISL_2402857, EPI_ISL_2402858, EPI_ISL_2402859, EPI_ISL_2402860, EPI_ISL_2402861, EPI_ISL_2402862, EPI_ISL_2402863, EPI_ISL_2402864, EPI_ISL_2402865, EPI_ISL_2402866, EPI_ISL_2402867, EPI_ISL_2402868, EPI_ISL_2402869, EPI_ISL_2402870, EPI_ISL_2402871, EPI_ISL_2402872, EPI_ISL_2402873, EPI_ISL_2402874, EPI_ISL_2402875, EPI_ISL_2402876, EPI_ISL_2402877, EPI_ISL_2402878, EPI_ISL_2402879, EPI_ISL_2402880, EPI_ISL_2402881, EPI_ISL_2402882, EPI_ISL_2402883, EPI_ISL_2402884, EPI_ISL_2402885, EPI_ISL_2402886, EPI_ISL_2402887, EPI_ISL_2402888, EPI_ISL_2402889, EPI_ISL_2402890, EPI_ISL_2402891, EPI_ISL_2402892, EPI_ISL_2402893, EPI_ISL_2402894, EPI_ISL_2402895, EPI_ISL_2402896, EPI_ISL_2402897, EPI_ISL_2402898, EPI_ISL_2402899, EPI_ISL_2402900, EPI_ISL_2402901, EPI_ISL_2402902, EPI_ISL_2402903, EPI_ISL_2402904, EPI_ISL_2402905, EPI_ISL_2402906, EPI_ISL_2402907, EPI_ISL_2402908, EPI_ISL_2402909, EPI_ISL_2402910, EPI_ISL_2402911, EPI_ISL_2402912, EPI_ISL_2402913, EPI_ISL_2402914, EPI_ISL_2402915, EPI_ISL_2402916, EPI_ISL_2402917, EPI_ISL_2402918, EPI_ISL_2402919, EPI_ISL_2402920, EPI_ISL_2402921, EPI_ISL_2402922, EPI_ISL_2402923, EPI_ISL_2402924, EPI_ISL_2402925, EPI_ISL_2402926, EPI_ISL_2402927, EPI_ISL_2402928, EPI_ISL_2402929, EPI_ISL_2402930, EPI_ISL_2402931, EPI_ISL_2402932, EPI_ISL_2402933, EPI_ISL_2402934, EPI_ISL_2402935, EPI_ISL_2404795, EPI_ISL_2404796, EPI_ISL_2404797, EPI_ISL_2404798, EPI_ISL_2404799 | | | |
| see above | Weill Cornell Medicine | New York Genome Center | Michael Zody, Andre Corvelo, Dayna M. Oschwald, Samantha Fennessey, Tom Maniatis, Melissa Cushing, Olivier Elemento, Margaret Elizabeth Ross, Chris Mason, Priya Velu, Hanna Rennert, Arryn Craney, Lars F Westblade |
| EPI_ISL_2405094, EPI_ISL_2405095, EPI_ISL_2405096, EPI_ISL_2405097, EPI_ISL_2405098, EPI_ISL_2405099, EPI_ISL_2405100, EPI_ISL_2405101, EPI_ISL_2405102, EPI_ISL_2405103, EPI_ISL_2405104, EPI_ISL_2405105, EPI_ISL_2405106, EPI_ISL_2405107, EPI_ISL_2405108 | | | |
| see above | Molecular Biology Laboratory, Westchester Medical Center | Genomics Core Laboratory, New York Medical College | Weihua Huang, Utsav Pandey, Jian Zhuge, Changhong Yin |
| EPI_ISL_2421044 | NORTHWELL HEALTH LABORATORIES | Wadsworth Center, New York State Department of Health | Kirsten St. George, Daryl M. Lamson, Alexis Russell, Matthew Shudt, Melissa A Leisner, Jonathan Plitnick, Catharine Prussing, Navjot Singh, John Kelly, Erasmus Schneider, Erica Lasek-Nesselquist |
| EPI_ISL_2421045, EPI_ISL_2421046, EPI_ISL_2421047, EPI_ISL_2421048, | STONY BROOK UNIVERSITY HOSPITAL | Wadsworth Center, New York State Department of Health | Kirsten St. George, Daryl M. Lamson, Alexis Russell, Matthew Shudt, Melissa A Leisner, Jonathan Plitnick, Catharine Prussing, Navjot Singh, John Kelly, Erasmus Schneider, Erica Lasek-Nesselquist |

| | | | |
|---|---|---|---|
| EPI_ISL_2421049, EPI_ISL_2421050, EPI_ISL_2421051, EPI_ISL_2421052, EPI_ISL_2421053 | | | |
| EPI_ISL_2421054, EPI_ISL_2421055, EPI_ISL_2421056, EPI_ISL_2421057, EPI_ISL_2421058, EPI_ISL_2421059, EPI_ISL_2421060, EPI_ISL_2421061, EPI_ISL_2421062, EPI_ISL_2421063, EPI_ISL_2421064, EPI_ISL_2421065, EPI_ISL_2421066, EPI_ISL_2421067, EPI_ISL_2421068, EPI_ISL_2421069, EPI_ISL_2421070, EPI_ISL_2421071, EPI_ISL_2421072, EPI_ISL_2421073, EPI_ISL_2421074, EPI_ISL_2421075, EPI_ISL_2421076, EPI_ISL_2421077, EPI_ISL_2421078, EPI_ISL_2421079, EPI_ISL_2421080, EPI_ISL_2421081 | | | |
| see above | SUNY UPSTATE MEDICAL UNIVERSITY | Wadsworth Center, New York State Department of Health | Kirsten St. George, Daryl M. Lamson, Alexis Russell, Matthew Shudt, Melissa A Leisner, Jonathan Plitnick, Catharine Prussing, Navjot Singh, John Kelly, Erasmus Schneider, Erica Lasek-Nesselquist |
| EPI_ISL_2421082, EPI_ISL_2421083, EPI_ISL_2421084, EPI_ISL_2421085, EPI_ISL_2421086, EPI_ISL_2421087, EPI_ISL_2421088, EPI_ISL_2421089, EPI_ISL_2421090, EPI_ISL_2421091, EPI_ISL_2421092, EPI_ISL_2421093, EPI_ISL_2421094, EPI_ISL_2421095 | | | |
| see above | BOSTON HEART DIAGNOSTICS CORP | Wadsworth Center, New York State Department of Health | Kirsten St. George, Daryl M. Lamson, Alexis Russell, Matthew Shudt, Melissa A Leisner, Jonathan Plitnick, Catharine Prussing, Navjot Singh, John Kelly, Erasmus Schneider, Erica Lasek-Nesselquist |
| EPI_ISL_2421096, EPI_ISL_2421097, EPI_ISL_2421098, EPI_ISL_2421099, EPI_ISL_2421100, EPI_ISL_2421101, EPI_ISL_2421102, EPI_ISL_2421103, EPI_ISL_2421104 | MONTEFIORE MEDICAL CENTER LABORATORIES | Wadsworth Center, New York State Department of Health | Kirsten St. George, Daryl M. Lamson, Alexis Russell, Matthew Shudt, Melissa A Leisner, Jonathan Plitnick, Catharine Prussing, Navjot Singh, John Kelly, Erasmus Schneider, Erica Lasek-Nesselquist |
| EPI_ISL_2421105, EPI_ISL_2421106, EPI_ISL_2421107 | WESTCHESTER MEDICAL CENTER | Wadsworth Center, New York State Department of Health | Kirsten St. George, Daryl M. Lamson, Alexis Russell, Matthew Shudt, Melissa A Leisner, Jonathan Plitnick, Catharine Prussing, Navjot Singh, John Kelly, Erasmus Schneider, Erica Lasek-Nesselquist |
| EPI_ISL_2421140, EPI_ISL_2421141, EPI_ISL_2421142, EPI_ISL_2421143 | ALBANY MEDICAL CENTER | Wadsworth Center, New York State Department of Health | Kirsten St. George, Daryl M. Lamson, Alexis Russell, Matthew Shudt, Melissa A Leisner, Jonathan Plitnick, Catharine Prussing, Navjot Singh, John Kelly, Erasmus Schneider, Erica Lasek-Nesselquist |
| EPI_ISL_2421144, EPI_ISL_2421145, EPI_ISL_2421146, EPI_ISL_2421147, EPI_ISL_2421148, EPI_ISL_2421149, EPI_ISL_2421150, EPI_ISL_2421151, EPI_ISL_2421152, EPI_ISL_2421153, EPI_ISL_2421154, EPI_ISL_2421155, EPI_ISL_2421156 | | | |
| see above | URMC LABS | Wadsworth Center, New York State Department of Health | Kirsten St. George, Daryl M. Lamson, Alexis Russell, Matthew Shudt, Melissa A Leisner, Jonathan Plitnick, Catharine Prussing, Navjot Singh, John Kelly, Erasmus Schneider, Erica Lasek-Nesselquist |
| EPI_ISL_2421157, EPI_ISL_2421158, EPI_ISL_2421159, EPI_ISL_2421160 | GLENS FALLS HOSPITAL LABORATORY | Wadsworth Center, New York State Department of Health | Kirsten St. George, Daryl M. Lamson, Alexis Russell, Matthew Shudt, Melissa A Leisner, Jonathan Plitnick, Catharine Prussing, Navjot Singh, John Kelly, Erasmus Schneider, Erica Lasek-Nesselquist |
| EPI_ISL_2421161, EPI_ISL_2421162, EPI_ISL_2421163, EPI_ISL_2421164, EPI_ISL_2421165, EPI_ISL_2421166, EPI_ISL_2421167, EPI_ISL_2421168, EPI_ISL_2421169, EPI_ISL_2421170, EPI_ISL_2421171, EPI_ISL_2421172, EPI_ISL_2421173, EPI_ISL_2421174, EPI_ISL_2421175, EPI_ISL_2421176, EPI_ISL_2421177, EPI_ISL_2421178, EPI_ISL_2421179, EPI_ISL_2421180, EPI_ISL_2421181, EPI_ISL_2421182, EPI_ISL_2421183, EPI_ISL_2421184, EPI_ISL_2421185, EPI_ISL_2421186, EPI_ISL_2421187, EPI_ISL_2421188 | | | |
| see above | TEMPUS LABS INC | Wadsworth Center, New York State Department of Health | Kirsten St. George, Daryl M. Lamson, Alexis Russell, Matthew Shudt, Melissa A Leisner, Jonathan Plitnick, Catharine Prussing, Navjot Singh, John Kelly, Erasmus Schneider, Erica Lasek-Nesselquist |
| EPI_ISL_2421189, EPI_ISL_2421190, EPI_ISL_2421191, EPI_ISL_2421192, EPI_ISL_2421193, EPI_ISL_2421194, EPI_ISL_2421195, EPI_ISL_2421196, EPI_ISL_2421197, EPI_ISL_2421198, EPI_ISL_2421199, EPI_ISL_2421200, EPI_ISL_2421201, EPI_ISL_2421202, EPI_ISL_2421203, EPI_ISL_2421204, EPI_ISL_2421205, EPI_ISL_2421206, EPI_ISL_2421207, EPI_ISL_2421208, EPI_ISL_2421209, EPI_ISL_2421210, EPI_ISL_2421211 | | | |
| see above | BOSTON HEART DIAGNOSTICS CORP | Wadsworth Center, New York State Department of Health | Kirsten St. George, Daryl M. Lamson, Alexis Russell, Matthew Shudt, Melissa A Leisner, Jonathan Plitnick, Catharine Prussing, Navjot Singh, John Kelly, Erasmus Schneider, Erica Lasek-Nesselquist |
| EPI_ISL_2421214 | WESTCHESTER MEDICAL CENTER | Wadsworth Center, New York State Department of Health | Kirsten St. George, Daryl M. Lamson, Alexis Russell, Matthew Shudt, Melissa A Leisner, Jonathan Plitnick, Catharine Prussing, Navjot Singh, John Kelly, Erasmus Schneider, Erica Lasek-Nesselquist |
| EPI_ISL_2421215 | NYC Pandemic Response Lab | Wadsworth Center, New York State Department of Health | Kirsten St. George, Daryl M. Lamson, Alexis Russell, Matthew Shudt, Melissa A Leisner, Jonathan Plitnick, Catharine Prussing, Navjot Singh, John Kelly, Erasmus Schneider, Erica Lasek-Nesselquist |
| EPI_ISL_2421216 | Columbia University Irving Medical Center | Wadsworth Center, New York State Department of Health | Kirsten St. George, Daryl M. Lamson, Alexis Russell, Matthew Shudt, Melissa A Leisner, Jonathan Plitnick, Catharine Prussing, Navjot Singh, John Kelly, Erasmus Schneider, Erica Lasek-Nesselquist |
| EPI_ISL_2421217 | MONTEFIORE MEDICAL CENTER LABORATORIES | Wadsworth Center, New York State Department of Health | Kirsten St. George, Daryl M. Lamson, Alexis Russell, Matthew Shudt, Melissa A Leisner, Jonathan Plitnick, Catharine Prussing, Navjot Singh, John Kelly, Erasmus Schneider, Erica Lasek-Nesselquist |
| EPI_ISL_2421218, EPI_ISL_2421219 | Columbia University Irving Medical Center | Wadsworth Center, New York State Department of Health | Kirsten St. George, Daryl M. Lamson, Alexis Russell, Matthew Shudt, Melissa A Leisner, Jonathan Plitnick, Catharine Prussing, Navjot Singh, John Kelly, Erasmus Schneider, Erica Lasek-Nesselquist |
| EPI_ISL_2421220 | SUNY UPSTATE MEDICAL UNIVERSITY | Wadsworth Center, New York State Department of Health | Kirsten St. George, Daryl M. Lamson, Alexis Russell, Matthew Shudt, Melissa A Leisner, Jonathan Plitnick, Catharine Prussing, Navjot Singh, John Kelly, Erasmus Schneider, Erica Lasek-Nesselquist |
| EPI_ISL_2421221, EPI_ISL_2421222 | NORTHWELL HEALTH LABORATORIES | Wadsworth Center, New York State Department of Health | Kirsten St. George, Daryl M. Lamson, Alexis Russell, Matthew Shudt, Melissa A Leisner, Jonathan Plitnick, Catharine Prussing, Navjot Singh, John Kelly, Erasmus Schneider, Erica Lasek-Nesselquist |
| EPI_ISL_2421223, EPI_ISL_2421224, EPI_ISL_2421225, EPI_ISL_2421226, EPI_ISL_2421227 | URMC LABS | Wadsworth Center, New York State Department of Health | Kirsten St. George, Daryl M. Lamson, Alexis Russell, Matthew Shudt, Melissa A Leisner, Jonathan Plitnick, Catharine Prussing, Navjot Singh, John Kelly, Erasmus Schneider, Erica Lasek-Nesselquist |
| EPI_ISL_2421228 | Garnet Health Medical Center Laboratory | Wadsworth Center, New York State Department of Health | Kirsten St. George, Daryl M. Lamson, Alexis Russell, Matthew Shudt, Melissa A Leisner, Jonathan Plitnick, Catharine Prussing, Navjot Singh, John Kelly, Erasmus Schneider, Erica Lasek-Nesselquist |
| EPI_ISL_2421229 | GLENS FALLS HOSPITAL LABORATORY | Wadsworth Center, New York State Department of Health | Kirsten St. George, Daryl M. Lamson, Alexis Russell, Matthew Shudt, Melissa A Leisner, Jonathan Plitnick, Catharine Prussing, Navjot Singh, John Kelly, Erasmus Schneider, Erica Lasek-Nesselquist |
| EPI_ISL_2421230, EPI_ISL_2421231, EPI_ISL_2421232, EPI_ISL_2421233, EPI_ISL_2421234, EPI_ISL_2421235, EPI_ISL_2421236, EPI_ISL_2421237, EPI_ISL_2421238, EPI_ISL_2421239, EPI_ISL_2421240, EPI_ISL_2421241, EPI_ISL_2421242, EPI_ISL_2421243, EPI_ISL_2421244, EPI_ISL_2421245, EPI_ISL_2421246, EPI_ISL_2421247, EPI_ISL_2421248 | | | |
| see above | BOSTON HEART DIAGNOSTICS CORP | Wadsworth Center, New York State Department of Health | Kirsten St. George, Daryl M. Lamson, Alexis Russell, Matthew Shudt, Melissa A Leisner, Jonathan Plitnick, Catharine Prussing, Navjot Singh, John Kelly, Erasmus Schneider, Erica Lasek-Nesselquist |
| EPI_ISL_2421249, EPI_ISL_2421250, EPI_ISL_2421251, EPI_ISL_2421252 | Saratoga Hospital Laboratory | Wadsworth Center, New York State Department of Health | Kirsten St. George, Daryl M. Lamson, Alexis Russell, Matthew Shudt, Melissa A Leisner, Jonathan Plitnick, Catharine Prussing, Navjot Singh, John Kelly, Erasmus Schneider, Erica Lasek-Nesselquist |
| EPI_ISL_2421253 | Vermont DoH | Wadsworth Center, New York State Department of Health | Kirsten St. George, Daryl M. Lamson, Alexis Russell, Matthew Shudt, Melissa A Leisner, Jonathan Plitnick, Catharine Prussing, Navjot Singh, John Kelly, Erasmus Schneider, Erica Lasek-Nesselquist |
| EPI_ISL_2421328, EPI_ISL_2421388, EPI_ISL_2421391, EPI_ISL_2421429, EPI_ISL_2421506, EPI_ISL_2421573, EPI_ISL_2421635, EPI_ISL_2421647, EPI_ISL_2421661, EPI_ISL_2421670, EPI_ISL_2421673, EPI_ISL_2421753, EPI_ISL_2421770, EPI_ISL_2421826, EPI_ISL_2421924, EPI_ISL_2421930, EPI_ISL_2421982, EPI_ISL_2421983, EPI_ISL_2421985, EPI_ISL_2422105 | | | |
| see above | Aegis Sciences Corporation | Centers for Disease Control and Prevention Division of Viral Diseases, Pathogen Discovery | Dakota Howard, Dhwani Batra, Peter W. Cook, Kara Moser, Adrian Paskey, Jason Caravas, Benjamin Rambo-Martin, Shatavia Morrison, Christopher Gulvick, Scott Sammons, Yvette Unoarumhi, Darlene Wagner, Matthew Schmerer, Cyndi Clark, Patrick Campbell, Rob Case, Vikramsinha Ghorpade, Holly Houdeshell, Ola Kvalvaag, Dillon Nall, Ethan Sanders, Alec Vest, Shaun Westlund, Matthew Hardison, Clinton R. Paden, Duncan MacCannell |
| EPI_ISL_2422283, EPI_ISL_2422284, EPI_ISL_2422285, EPI_ISL_2422286, EPI_ISL_2422287, EPI_ISL_2422288, EPI_ISL_2422289, EPI_ISL_2422290, EPI_ISL_2422291, EPI_ISL_2422292, EPI_ISL_2422293, EPI_ISL_2422294, EPI_ISL_2422295, EPI_ISL_2422296, EPI_ISL_2422297, EPI_ISL_2422298, EPI_ISL_2422299, EPI_ISL_2422300, EPI_ISL_2422301, EPI_ISL_2422302, EPI_ISL_2422303, EPI_ISL_2422304, EPI_ISL_2422305, EPI_ISL_2422306, EPI_ISL_2422307, EPI_ISL_2422308, EPI_ISL_2422309, EPI_ISL_2422310, EPI_ISL_2422311, EPI_ISL_2422312 | | | |
| see above | SUNY UPSTATE MEDICAL UNIVERSITY | Wadsworth Center, New York State Department of Health | Kirsten St. George, Daryl M. Lamson, Alexis Russell, Matthew Shudt, Melissa A Leisner, Jonathan Plitnick, Catharine Prussing, Navjot Singh, John Kelly, Erasmus Schneider, Erica Lasek-Nesselquist |
| EPI_ISL_2422313 | ALBANY MEDICAL CENTER | Wadsworth Center, New York State Department of Health | Kirsten St. George, Daryl M. Lamson, Alexis Russell, Matthew Shudt, Melissa A Leisner, Jonathan Plitnick, Catharine Prussing, Navjot Singh, John Kelly, |

| | | | |
|---|---|---|---|
| | | | Erasmus Schneider, Erica Lasek-Nesselquist |
| EPI_ISL_2422314, EPI_ISL_2422315, EPI_ISL_2422316, EPI_ISL_2422317, EPI_ISL_2422318, EPI_ISL_2422319, EPI_ISL_2422320, EPI_ISL_2422321, EPI_ISL_2422322 | STONY BROOK UNIVERSITY HOSPITAL | Wadsworth Center, New York State Department of Health | Kirsten St. George, Daryl M. Lamson, Alexis Russell, Matthew Shudt, Melissa A Leisner, Jonathan Plitnick, Catharine Prussing, Navjot Singh, John Kelly, Erasmus Schneider, Erica Lasek-Nesselquist |
| EPI_ISL_2422323, EPI_ISL_2422324, EPI_ISL_2422325, EPI_ISL_2422326 | MONTEFIORE MEDICAL CENTER LABORATORIES | Wadsworth Center, New York State Department of Health | Kirsten St. George, Daryl M. Lamson, Alexis Russell, Matthew Shudt, Melissa A Leisner, Jonathan Plitnick, Catharine Prussing, Navjot Singh, John Kelly, Erasmus Schneider, Erica Lasek-Nesselquist |
| EPI_ISL_2422327, EPI_ISL_2422328, EPI_ISL_2422329, EPI_ISL_2422330, EPI_ISL_2422331, EPI_ISL_2422332, EPI_ISL_2422333, EPI_ISL_2422334, EPI_ISL_2422335, EPI_ISL_2422336, EPI_ISL_2422337, EPI_ISL_2422338, EPI_ISL_2422339, EPI_ISL_2422340, EPI_ISL_2422341, EPI_ISL_2422342, EPI_ISL_2422343, EPI_ISL_2422344, EPI_ISL_2422345, EPI_ISL_2422346, EPI_ISL_2422347, EPI_ISL_2422348, EPI_ISL_2422349, EPI_ISL_2422350, EPI_ISL_2422351, EPI_ISL_2422352, EPI_ISL_2422353, EPI_ISL_2422354, EPI_ISL_2422355, EPI_ISL_2422356, EPI_ISL_2422357, EPI_ISL_2422358, EPI_ISL_2422359, EPI_ISL_2422360 | | | |
| see above | URMC LABS | Wadsworth Center, New York State Department of Health | Kirsten St. George, Daryl M. Lamson, Alexis Russell, Matthew Shudt, Melissa A Leisner, Jonathan Plitnick, Catharine Prussing, Navjot Singh, John Kelly, Erasmus Schneider, Erica Lasek-Nesselquist |
| EPI_ISL_2427267, EPI_ISL_2427273, EPI_ISL_2427274, EPI_ISL_2427275, EPI_ISL_2427314, EPI_ISL_2427318, EPI_ISL_2427319, EPI_ISL_2427321 | IVY3 Central Lab, Vanderbilt University Medical Center | Lauring Lab, University of Michigan, Department of Microbiology and Immunology | Gilbert |
| EPI_ISL_2427403, EPI_ISL_2427404, EPI_ISL_2427405, EPI_ISL_2427406, EPI_ISL_2427407, EPI_ISL_2427408, EPI_ISL_2427409, EPI_ISL_2427410, EPI_ISL_2427411, EPI_ISL_2427412, EPI_ISL_2427413, EPI_ISL_2427414, EPI_ISL_2427415, EPI_ISL_2427416, EPI_ISL_2427417, EPI_ISL_2427418, EPI_ISL_2427419, EPI_ISL_2427420, EPI_ISL_2427421, EPI_ISL_2427422, EPI_ISL_2427423, EPI_ISL_2427424, EPI_ISL_2427425, EPI_ISL_2427426, EPI_ISL_2427427, EPI_ISL_2427428, EPI_ISL_2427429, EPI_ISL_2427431, EPI_ISL_2427432, EPI_ISL_2427433, EPI_ISL_2427434, EPI_ISL_2427435, EPI_ISL_2427436, EPI_ISL_2427437, EPI_ISL_2427438, EPI_ISL_2427439, EPI_ISL_2427440, EPI_ISL_2427441, EPI_ISL_2427442, EPI_ISL_2427443, EPI_ISL_2427444, EPI_ISL_2427445, EPI_ISL_2427446, EPI_ISL_2427447, EPI_ISL_2427448, EPI_ISL_2427449, EPI_ISL_2427450, EPI_ISL_2427451, EPI_ISL_2427452, EPI_ISL_2427453, EPI_ISL_2427454, EPI_ISL_2427455, EPI_ISL_2427456, EPI_ISL_2427457, EPI_ISL_2427458, EPI_ISL_2427459, EPI_ISL_2427460, EPI_ISL_2427461, EPI_ISL_2427462, EPI_ISL_2427463, EPI_ISL_2427464, EPI_ISL_2427465, EPI_ISL_2427466, EPI_ISL_2427467, EPI_ISL_2427468, EPI_ISL_2427469, EPI_ISL_2427470, EPI_ISL_2427471, EPI_ISL_2427472, EPI_ISL_2427473, EPI_ISL_2427474, EPI_ISL_2427475, EPI_ISL_2427476, EPI_ISL_2427477, EPI_ISL_2427478, EPI_ISL_2427479, EPI_ISL_2427480, EPI_ISL_2427481, EPI_ISL_2427482, EPI_ISL_2427483, EPI_ISL_2427485, EPI_ISL_2427486, EPI_ISL_2427487, EPI_ISL_2427488, EPI_ISL_2427489, EPI_ISL_2427490, EPI_ISL_2427492, EPI_ISL_2427493, EPI_ISL_2427494, EPI_ISL_2427495, EPI_ISL_2427496, EPI_ISL_2427497, EPI_ISL_2427498, EPI_ISL_2427499, EPI_ISL_2427500, EPI_ISL_2427501, EPI_ISL_2427502, EPI_ISL_2427503 | | | |
| see above | NYU Langone Health | Departments of Pathology and Medicine, New York University School of Medicine | Adriana Heguy, Dacia Dimartino, Emily Guzman, Christian Marier, Peter Meyn, Sitharam Ramaswami, Gael Westby, Paul Zappile, Yutong Zhang, Paolo Cotzia, Guiqing Wang |
| EPI_ISL_2439300, EPI_ISL_2439427, EPI_ISL_2439478, EPI_ISL_2439494, EPI_ISL_2439506, EPI_ISL_2439518, EPI_ISL_2439536, EPI_ISL_2439538, EPI_ISL_2439832, EPI_ISL_2439834, EPI_ISL_2439852, EPI_ISL_2439855, EPI_ISL_2439856, EPI_ISL_2439867, EPI_ISL_2439868, EPI_ISL_2439887, EPI_ISL_2439889, EPI_ISL_2439896, EPI_ISL_2439900, EPI_ISL_2439903, EPI_ISL_2439904, EPI_ISL_2439906, EPI_ISL_2439912, EPI_ISL_2439918, EPI_ISL_2439919, EPI_ISL_2439921, EPI_ISL_2439922, EPI_ISL_2439925, EPI_ISL_2439933, EPI_ISL_2439935, EPI_ISL_2439942, EPI_ISL_2439944, EPI_ISL_2439959, EPI_ISL_2439961, EPI_ISL_2439962, EPI_ISL_2439971 | | | |
| see above | Quest Diagnostics Incorporated | Centers for Disease Control and Prevention Division of Viral Diseases, Pathogen Discovery | Dakota Howard, Dhwani Batra, Peter W. Cook, Kara Moser, Adrian Paskey, Jason Caravas, Benjamin Rambo-Martin, Shatavia Morrison, Christopher Gulvick, Scott Sammons, Yvette Unoarumhi, Darlene Wagner, Matthew Schmerer, S. H. Rosenthal, A. Gerasimova, R. M. Kagan, B. Anderson, M. Hua, Y. Liu, L.E. Bernstein, K.E. Livingston, A. Perez, I. A. Shlyakhter, R. V. Rolando, R. Owen, P. Tanpaiboon, F. Lacbawan, Clinton R. Paden, Duncan MacCannell |
| EPI_ISL_2440136, EPI_ISL_2440146 | Helix/Illumina | Centers for Disease Control and Prevention Division of Viral Diseases, Pathogen Discovery | Dakota Howard, Dhwani Batra, Peter W. Cook, Kara Moser, Adrian Paskey, Jason Caravas, Benjamin Rambo-Martin, Shatavia Morrison, Christopher Gulvick, Scott Sammons, Yvette Unoarumhi, Darlene Wagner, Matthew Schmerer, Eileen de Feo, Jan Antico, Christine Tran, Matthew Tolentino, Shannon Wickline, Kim Gietzen, Brad Sickler, Jingtao Liu, Eric Allen, Phil Febbo, Nicole L. Washington, Simon White, Geraint Levan, Kelly Schiabor Barrett, Elizabeth Cirulli, Alexandre Bolze, Ary Ascencio, Charlotte Rivera-Garcia, Ryan Cho, Jason Nguyen, Sherry Wang, Jimmy Ramirez, Tyler Cassens, Efren Sandoval, Magnus Isaksson, William Lee, David Becker, Marc Laurent, James Lu, Clinton R. Paden, Duncan MacCannell |
| EPI_ISL_2440648, EPI_ISL_2440800, EPI_ISL_2440829 | Quest Diagnostics Incorporated | Centers for Disease Control and Prevention Division of Viral Diseases, Pathogen Discovery | Dakota Howard, Dhwani Batra, Peter W. Cook, Kara Moser, Adrian Paskey, Jason Caravas, Benjamin Rambo-Martin, Shatavia Morrison, Christopher Gulvick, Scott Sammons, Yvette Unoarumhi, Darlene Wagner, Matthew Schmerer, S. H. Rosenthal, A. Gerasimova, R. M. Kagan, B. Anderson, M. Hua, Y. Liu, L.E. Bernstein, K.E. Livingston, A. Perez, I. A. Shlyakhter, R. V. Rolando, R. Owen, P. Tanpaiboon, F. Lacbawan, Clinton R. Paden, Duncan MacCannell |
| EPI_ISL_2442311, EPI_ISL_2442328 | Aegis Sciences Corporation | Centers for Disease Control and Prevention Division of Viral Diseases, Pathogen Discovery | Dakota Howard, Dhwani Batra, Peter W. Cook, Kara Moser, Adrian Paskey, Jason Caravas, Benjamin Rambo-Martin, Shatavia Morrison, Christopher Gulvick, Scott Sammons, Yvette Unoarumhi, Darlene Wagner, Matthew Schmerer, Cyndi Clark, Patrick Campbell, Rob Case, Vikramsinha Ghorpade, Holly Houdeshell, Ola Kvalvaag, Dillon Nall, Ethan Sanders, Alec Vest, Shaun Westlund, Matthew Hardison, Clinton R. Paden, Duncan MacCannell |
| EPI_ISL_2443066, EPI_ISL_2443067, EPI_ISL_2443068, EPI_ISL_2443069, EPI_ISL_2443070 | NYU Langone Health | Departments of Pathology and Medicine, New York University School of Medicine | Adriana Heguy, Dacia Dimartino, Emily Guzman, Christian Marier, Peter Meyn, Sitharam Ramaswami, Gael Westby, Paul Zappile, Yutong Zhang, Paolo Cotzia, Guiqing Wang |
| EPI_ISL_2443782 | Stony Brook Medicine | New York Genome Center | Michael Zody, Andre Corvelo, Dayna M. Oschwald, Samantha Fennessey, Tom Maniatis , Sarah A. Halbohn, Alaa B. Salaheldin, Sarah A. Malik, Josue M. Flores Acuna, Dimitri Gnatenko, Silvia G. Spitzer, Margaret M. McGovern, Eric D. Spitzer, Richard R. Kew, Kenneth R. Shroyer, Jiang Chen, Susan V. Donelan |
| EPI_ISL_2443783, EPI_ISL_2443784, EPI_ISL_2443785, EPI_ISL_2443786, EPI_ISL_2443787, EPI_ISL_2443788, EPI_ISL_2443789, EPI_ISL_2443790, EPI_ISL_2443791, EPI_ISL_2443792, EPI_ISL_2443795 | | | |
| see above | Weill Cornell Medicine | New York Genome Center | Michael Zody, Andre Corvelo, Dayna M. Oschwald, Samantha Fennessey, Tom Maniatis, Melissa Cushing, Olivier Elemento, Margaret Elizabeth Ross, Chris Mason, Priya Velu, Hanna Rennert, Arryn Craney, Lars F Westblade |
| EPI_ISL_2446035, EPI_ISL_2446036, EPI_ISL_2446037, EPI_ISL_2446038, EPI_ISL_2446039, EPI_ISL_2446040, EPI_ISL_2446041, EPI_ISL_2446042, EPI_ISL_2446043, EPI_ISL_2446044, EPI_ISL_2446045, EPI_ISL_2446046, EPI_ISL_2446047, EPI_ISL_2446048, EPI_ISL_2446049, EPI_ISL_2446050, EPI_ISL_2446051, EPI_ISL_2446052, EPI_ISL_2446053, EPI_ISL_2446054, EPI_ISL_2446055, EPI_ISL_2446056, EPI_ISL_2446057, EPI_ISL_2446058, EPI_ISL_2446059, EPI_ISL_2446060, EPI_ISL_2446061, EPI_ISL_2446062, EPI_ISL_2446063, EPI_ISL_2446064, EPI_ISL_2446065, EPI_ISL_2446066, EPI_ISL_2446067, EPI_ISL_2446068, EPI_ISL_2446069, EPI_ISL_2446070, EPI_ISL_2446071, EPI_ISL_2446072, EPI_ISL_2446073, EPI_ISL_2446074, EPI_ISL_2446075, EPI_ISL_2446076, EPI_ISL_2446077, EPI_ISL_2446078, EPI_ISL_2446079, EPI_ISL_2446080, EPI_ISL_2446081, EPI_ISL_2446082, EPI_ISL_2446083, EPI_ISL_2446084, EPI_ISL_2446085, EPI_ISL_2446086, EPI_ISL_2446087, EPI_ISL_2446088, EPI_ISL_2446089, EPI_ISL_2446090, EPI_ISL_2446091, EPI_ISL_2446092, EPI_ISL_2446093, EPI_ISL_2446094, EPI_ISL_2446096, EPI_ISL_2446099, EPI_ISL_2446102, EPI_ISL_2446105, EPI_ISL_2446107, EPI_ISL_2446111, EPI_ISL_2446114, EPI_ISL_2446117, EPI_ISL_2446120 | | | |
| see above | Pandemic Response Lab - NYC | Pandemic Response Lab, R&D | Henry Lee, Michael Hammerling, Melissa Hopkins, Cybill del Castillo, Shinyoung Clair Kang, William Ward, Pradeep Bugga, Sol Rey, Dylan Law, Katharine Nelson, Haiping Hao, Jon Laurent |
| EPI_ISL_2448298 | MONTEFIORE MEDICAL CENTER LABORATORIES | Wadsworth Center, New York State Department of Health | Kirsten St. George, Daryl M. Lamson, Alexis Russell, Matthew Shudt, Melissa A Leisner, Jonathan Plitnick, Catharine Prussing, Navjot Singh, John Kelly, Erasmus Schneider, Erica Lasek-Nesselquist |
| EPI_ISL_2448299 | SUNY UPSTATE MEDICAL UNIVERSITY | Wadsworth Center, New York State Department of Health | Kirsten St. George, Daryl M. Lamson, Alexis Russell, Matthew Shudt, Melissa A Leisner, Jonathan Plitnick, Catharine Prussing, Navjot Singh, John Kelly, Erasmus Schneider, Erica Lasek-Nesselquist |
| EPI_ISL_2448300 | TEMPUS LABS INC | Wadsworth Center, New York State Department of Health | Kirsten St. George, Daryl M. Lamson, Alexis Russell, Matthew Shudt, Melissa A Leisner, Jonathan Plitnick, Catharine Prussing, Navjot Singh, John Kelly, Erasmus Schneider, Erica Lasek-Nesselquist |
| EPI_ISL_2448301 | WESTCHESTER MEDICAL CENTER | Wadsworth Center, New York State Department of Health | Kirsten St. George, Daryl M. Lamson, Alexis Russell, Matthew Shudt, Melissa A Leisner, Jonathan Plitnick, Catharine Prussing, Navjot Singh, John Kelly, Erasmus Schneider, Erica Lasek-Nesselquist |
| EPI_ISL_2448302 | MONTEFIORE MEDICAL CENTER LABORATORIES | Wadsworth Center, New York State Department of Health | Kirsten St. George, Daryl M. Lamson, Alexis Russell, Matthew Shudt, Melissa A Leisner, Jonathan Plitnick, Catharine Prussing, Navjot Singh, John Kelly, Erasmus Schneider, Erica Lasek-Nesselquist |
| EPI_ISL_2448303 | URMC LABS | Wadsworth Center, New York State Department of Health | Kirsten St. George, Daryl M. Lamson, Alexis Russell, Matthew Shudt, Melissa A Leisner, Jonathan Plitnick, Catharine Prussing, Navjot Singh, John Kelly, Erasmus Schneider, Erica Lasek-Nesselquist |
| EPI_ISL_2448304 | BOSTON HEART DIAGNOSTICS CORP | Wadsworth Center, New York State Department of Health | Kirsten St. George, Daryl M. Lamson, Alexis Russell, Matthew Shudt, Melissa A Leisner, Jonathan Plitnick, Catharine Prussing, Navjot Singh, John Kelly, Erasmus Schneider, Erica Lasek-Nesselquist |
| EPI_ISL_2448305 | TEMPUS LABS INC | Wadsworth Center, New York State Department of Health | Kirsten St. George, Daryl M. Lamson, Alexis Russell, Matthew Shudt, Melissa A Leisner, Jonathan Plitnick, Catharine Prussing, Navjot Singh, John Kelly, |

| | | | |
|---|---|---|---|
| | | | Erasmus Schneider, Erica Lasek-Nesselquist |
| EPI_ISL_2448306, EPI_ISL_2448307 | URMC LABS | Wadsworth Center, New York State Department of Health | Kirsten St. George, Daryl M. Lamson, Alexis Russell, Matthew Shudt, Melissa A Leisner, Jonathan Plitnick, Catharine Prussing, Navjot Singh, John Kelly, Erasmus Schneider, Erica Lasek-Nesselquist |
| EPI_ISL_2448308, EPI_ISL_2448309 | BOSTON HEART DIAGNOSTICS CORP | Wadsworth Center, New York State Department of Health | Kirsten St. George, Daryl M. Lamson, Alexis Russell, Matthew Shudt, Melissa A Leisner, Jonathan Plitnick, Catharine Prussing, Navjot Singh, John Kelly, Erasmus Schneider, Erica Lasek-Nesselquist |
| EPI_ISL_2448310 | STONY BROOK UNIVERSITY HOSPITAL | Wadsworth Center, New York State Department of Health | Kirsten St. George, Daryl M. Lamson, Alexis Russell, Matthew Shudt, Melissa A Leisner, Jonathan Plitnick, Catharine Prussing, Navjot Singh, John Kelly, Erasmus Schneider, Erica Lasek-Nesselquist |
| EPI_ISL_2448311 | BOSTON HEART DIAGNOSTICS CORP | Wadsworth Center, New York State Department of Health | Kirsten St. George, Daryl M. Lamson, Alexis Russell, Matthew Shudt, Melissa A Leisner, Jonathan Plitnick, Catharine Prussing, Navjot Singh, John Kelly, Erasmus Schneider, Erica Lasek-Nesselquist |
| EPI_ISL_2448312 | ALBANY MEDICAL CENTER | Wadsworth Center, New York State Department of Health | Kirsten St. George, Daryl M. Lamson, Alexis Russell, Matthew Shudt, Melissa A Leisner, Jonathan Plitnick, Catharine Prussing, Navjot Singh, John Kelly, Erasmus Schneider, Erica Lasek-Nesselquist |
| EPI_ISL_2448313, EPI_ISL_2448314, EPI_ISL_2448315, EPI_ISL_2448316, EPI_ISL_2448317 | BOSTON HEART DIAGNOSTICS CORP | Wadsworth Center, New York State Department of Health | Kirsten St. George, Daryl M. Lamson, Alexis Russell, Matthew Shudt, Melissa A Leisner, Jonathan Plitnick, Catharine Prussing, Navjot Singh, John Kelly, Erasmus Schneider, Erica Lasek-Nesselquist |
| EPI_ISL_2448318 | URMC LABS | Wadsworth Center, New York State Department of Health | Kirsten St. George, Daryl M. Lamson, Alexis Russell, Matthew Shudt, Melissa A Leisner, Jonathan Plitnick, Catharine Prussing, Navjot Singh, John Kelly, Erasmus Schneider, Erica Lasek-Nesselquist |
| EPI_ISL_2448319, EPI_ISL_2448320 | BOSTON HEART DIAGNOSTICS CORP | Wadsworth Center, New York State Department of Health | Kirsten St. George, Daryl M. Lamson, Alexis Russell, Matthew Shudt, Melissa A Leisner, Jonathan Plitnick, Catharine Prussing, Navjot Singh, John Kelly, Erasmus Schneider, Erica Lasek-Nesselquist |
| EPI_ISL_2448321 | URMC LABS | Wadsworth Center, New York State Department of Health | Kirsten St. George, Daryl M. Lamson, Alexis Russell, Matthew Shudt, Melissa A Leisner, Jonathan Plitnick, Catharine Prussing, Navjot Singh, John Kelly, Erasmus Schneider, Erica Lasek-Nesselquist |
| EPI_ISL_2450617, EPI_ISL_2450618, EPI_ISL_2450619, EPI_ISL_2450620, EPI_ISL_2450621, EPI_ISL_2450622, EPI_ISL_2450623, EPI_ISL_2450624, EPI_ISL_2450625, EPI_ISL_2450626, EPI_ISL_2450627, EPI_ISL_2450628, EPI_ISL_2450629, EPI_ISL_2450630, EPI_ISL_2450631, EPI_ISL_2450632, EPI_ISL_2450633, EPI_ISL_2450634, EPI_ISL_2450635, EPI_ISL_2450636, EPI_ISL_2450637, EPI_ISL_2450638, EPI_ISL_2450639, EPI_ISL_2450640, EPI_ISL_2450641, EPI_ISL_2450642, EPI_ISL_2450643, EPI_ISL_2450644, EPI_ISL_2450645, EPI_ISL_2450646, EPI_ISL_2450647, EPI_ISL_2450648, EPI_ISL_2450649, EPI_ISL_2450650, EPI_ISL_2450651, EPI_ISL_2450652, EPI_ISL_2450653, EPI_ISL_2450654, EPI_ISL_2450655, EPI_ISL_2450656, EPI_ISL_2450657, EPI_ISL_2450658, EPI_ISL_2450659, EPI_ISL_2450660, EPI_ISL_2450661, EPI_ISL_2450662, EPI_ISL_2450663, EPI_ISL_2450664, EPI_ISL_2450665, EPI_ISL_2450666, EPI_ISL_2450667, EPI_ISL_2450668, EPI_ISL_2450669, EPI_ISL_2450670, EPI_ISL_2450671, EPI_ISL_2450672, EPI_ISL_2450673, EPI_ISL_2450674, EPI_ISL_2450675, EPI_ISL_2450676, EPI_ISL_2450677, EPI_ISL_2450678, EPI_ISL_2450679, EPI_ISL_2450680, EPI_ISL_2450681, EPI_ISL_2450682, EPI_ISL_2450683, EPI_ISL_2450684, EPI_ISL_2450685, EPI_ISL_2450686, EPI_ISL_2450687, EPI_ISL_2450688, EPI_ISL_2450689, EPI_ISL_2450690 | | | |
| see above | Weill Cornell Medicine | New York Genome Center | Michael Zody, Andre Corvelo, Dayna M. Oschwald, Samantha Fennessey, Tom Maniatis, Melissa Cushing, Olivier Elemento, Margaret Elizabeth Ross, Chris Mason, Priya Velu, Hanna Rennert, Arryn Craney, Lars F Westblade |
| EPI_ISL_2451376, EPI_ISL_2451388, EPI_ISL_2451404, EPI_ISL_2451424, EPI_ISL_2451433, EPI_ISL_2451478, EPI_ISL_2451488 | Quest Diagnostics Incorporated | Centers for Disease Control and Prevention Division of Viral Diseases, Pathogen Discovery | Dakota Howard, Dhwani Batra, Peter W. Cook, Kara Moser, Adrian Paskey, Jason Caravas, Benjamin Rambo-Martin, Shatavia Morrison, Christopher Gulvick, Scott Sammons, Yvette Unoarumhi, Darlene Wagner, Matthew Schmerer, S. H. Rosenthal, A. Gerasimova, R. M. Kagan, B. Anderson, M. Hua, Y. Liu, L.E. Bernstein, K.E. Livingston, A. Perez, I. A. Shlyakhter, R. V. Rolando, R. Owen, P. Tanpaiboon, F. Lacbawan, Clinton R. Paden, Duncan MacCannell |
| EPI_ISL_2452047, EPI_ISL_2452048, EPI_ISL_2452050, EPI_ISL_2452052, EPI_ISL_2452054, EPI_ISL_2452055, EPI_ISL_2452057, EPI_ISL_2452060, EPI_ISL_2452063, EPI_ISL_2452064, EPI_ISL_2452066, EPI_ISL_2452067, EPI_ISL_2452069, EPI_ISL_2452071, EPI_ISL_2452073 | | | |
| see above | NYC Department of Health and Mental Hygiene | Centers for Disease Control and Prevention Division of Viral Diseases, Pathogen Discovery | Mili Sheth, Sarah Nobles, Jasmine Padilla, Mark Burroughs, Shoshona Le, Katie Dillon, Peter Cook, Clinton R. Paden, Dhwani Batra, Krista Queen, Kristen Knipe, Dakota Howard, Yvette Unoarumhi, Darlene Wagner, Matthew Schmerer, Ben L. Rambo-Martin, Kristine Lacek, Sam Shepard, Alison Laufer Halpin, Dave Wentworth, Vivien Dugan, Suxiang Tong, Justin Lee |
| EPI_ISL_2455391, EPI_ISL_2455392, EPI_ISL_2455393, EPI_ISL_2455394, EPI_ISL_2455395 | Weill Cornell Medicine | New York Genome Center | Michael Zody, Andre Corvelo, Dayna M. Oschwald, Samantha Fennessey, Tom Maniatis, Melissa Cushing, Olivier Elemento, Margaret Elizabeth Ross, Chris Mason, Priya Velu, Hanna Rennert, Arryn Craney, Lars F Westblade |
| EPI_ISL_2458161, EPI_ISL_2458162, EPI_ISL_2458163, EPI_ISL_2458164, EPI_ISL_2458166, EPI_ISL_2458168, EPI_ISL_2458169, EPI_ISL_2458170, EPI_ISL_2458171, EPI_ISL_2458172, EPI_ISL_2458173, EPI_ISL_2458174, EPI_ISL_2458175, EPI_ISL_2458176, EPI_ISL_2458177, EPI_ISL_2458178, EPI_ISL_2458179, EPI_ISL_2458180, EPI_ISL_2458181, EPI_ISL_2458182, EPI_ISL_2458183, EPI_ISL_2458184, EPI_ISL_2458185, EPI_ISL_2458186, EPI_ISL_2458187, EPI_ISL_2458188, EPI_ISL_2458189, EPI_ISL_2458190, EPI_ISL_2458191, EPI_ISL_2458192, EPI_ISL_2458193, EPI_ISL_2458194, EPI_ISL_2458195, EPI_ISL_2458196, EPI_ISL_2458197, EPI_ISL_2458198, EPI_ISL_2458199, EPI_ISL_2458200, EPI_ISL_2458201, EPI_ISL_2458202, EPI_ISL_2458203, EPI_ISL_2458204, EPI_ISL_2458205, EPI_ISL_2458206, EPI_ISL_2458207, EPI_ISL_2458208, EPI_ISL_2458209, EPI_ISL_2458210, EPI_ISL_2458211, EPI_ISL_2458212, EPI_ISL_2458213, EPI_ISL_2458214, EPI_ISL_2458215, EPI_ISL_2458216, EPI_ISL_2458217, EPI_ISL_2458218, EPI_ISL_2458219, EPI_ISL_2458220, EPI_ISL_2458221, EPI_ISL_2458222, EPI_ISL_2458223, EPI_ISL_2458224 | | | |
| see above | NYU Langone Health | Departments of Pathology and Medicine, New York University School of Medicine | Adriana Heguy, Dacia Dimartino, Emily Guzman, Christian Marier, Peter Meyn, Sitharam Ramaswami, Gael Westby, Paul Zappile, Yutong Zhang, Paolo Cotzia, Guiqing Wang |
| EPI_ISL_2462404 | SUNY Upstate Medical University | SUNY Upstate Medical University | Frank A Middleton |
| EPI_ISL_2463182 | Quest Diagnostics Incorporated | Centers for Disease Control and Prevention Division of Viral Diseases, Pathogen Discovery | Dakota Howard, Dhwani Batra, Peter W. Cook, Kara Moser, Adrian Paskey, Jason Caravas, Benjamin Rambo-Martin, Shatavia Morrison, Christopher Gulvick, Scott Sammons, Yvette Unoarumhi, Darlene Wagner, Matthew Schmerer, S. H. Rosenthal, A. Gerasimova, R. M. Kagan, B. Anderson, M. Hua, Y. Liu, L.E. Bernstein, K.E. Livingston, A. Perez, I. A. Shlyakhter, R. V. Rolando, R. Owen, P. Tanpaiboon, F. Lacbawan, Clinton R. Paden, Duncan MacCannell |
| EPI_ISL_2463297 | NYU Langone Health | Departments of Pathology and Medicine, New York University School of Medicine | Adriana Heguy, Dacia Dimartino, Emily Guzman, Christian Marier, Peter Meyn, Sitharam Ramaswami, Gael Westby, Paul Zappile, Yutong Zhang, Paolo Cotzia, Guiqing Wang |
| EPI_ISL_2466668, EPI_ISL_2466669, EPI_ISL_2466670 | Stony Brook Medicine | New York Genome Center | Michael Zody, Andre Corvelo, Dayna M. Oschwald, Samantha Fennessey, Tom Maniatis , Sarah A. Halbohn, Alaa B. Salaheldin, Sarah A. Malik, Josue M. Flores Acuna, Dimitri Gnatenko, Silvia G. Spitzer, Margaret M. McGovern, Eric D. Spitzer, Richard R. Kew, Kenneth R. Shroyer, Jiang Chen, Susan V. Donelan |
| EPI_ISL_2466681 | DOHMH PHL | New York City Public Health Laboratory | Jade Wang, et al. |
| EPI_ISL_2466682, EPI_ISL_2466683 | DOHMH Corona | New York City Public Health Laboratory | Jade Wang, et al. |
| EPI_ISL_2466684, EPI_ISL_2466685 | DOHMH Central Harlem | New York City Public Health Laboratory | Jade Wang, et al. |
| EPI_ISL_2466686 | DOHMH Jamaica | New York City Public Health Laboratory | Jade Wang, et al. |
| EPI_ISL_2466687 | DOHMH Chelsea | New York City Public Health Laboratory | Jade Wang, et al. |
| EPI_ISL_2466688 | DOHMH Corona | New York City Public Health Laboratory | Jade Wang, et al. |
| EPI_ISL_2466689 | DOHMH Jamaica | New York City Public Health Laboratory | Jade Wang, et al. |
| EPI_ISL_2466690 | DOHMH Corona | New York City Public Health Laboratory | Jade Wang, et al. |
| EPI_ISL_2466691, EPI_ISL_2466692, EPI_ISL_2466693, EPI_ISL_2466694, EPI_ISL_2466695, EPI_ISL_2466696 | Sherman Abrams Laboratory | New York City Public Health Laboratory | Jade Wang, et al. |
| EPI_ISL_2466697 | Department of Homeless Services | New York City Public Health Laboratory | Jade Wang, et al. |
| EPI_ISL_2466700, EPI_ISL_2466701, EPI_ISL_2466702 | OCME Office Of Chief Medical Examiner | New York City Public Health Laboratory | Jade Wang, et al. |
| EPI_ISL_2480150, EPI_ISL_2480157, EPI_ISL_2480158, EPI_ISL_2480159, EPI_ISL_2480160, EPI_ISL_2480161, EPI_ISL_2480162, EPI_ISL_2480163, EPI_ISL_2480164, EPI_ISL_2480165, EPI_ISL_2480166, EPI_ISL_2480167, EPI_ISL_2480168 | | | |
| see above | Fulgent Genetics | Centers for Disease Control and Prevention Division of Viral Diseases, Pathogen Discovery | Dakota Howard, Dhwani Batra, Peter W. Cook, Kara Moser, Adrian Paskey, Jason Caravas, Benjamin Rambo-Martin, Shatavia Morrison, Christopher Gulvick, Scott Sammons, Yvette Unoarumhi, Darlene Wagner, Matthew Schmerer, Harry Gao, Mickey Li, John Gao, Joseph Fierro, Benafsh Sapra, Becky |

| | | | |
|---|---|---|---|
| | | | Tsai, Yan Meng, Doreen Ng, James Xie, Clinton R. Paden, Duncan MacCannell |
| EPI_ISL_2480230, EPI_ISL_2480231, EPI_ISL_2480232, EPI_ISL_2480485, EPI_ISL_2480486, EPI_ISL_2480487, EPI_ISL_2480488, EPI_ISL_2480489, EPI_ISL_2480491, EPI_ISL_2480499, EPI_ISL_2480500, EPI_ISL_2480507, EPI_ISL_2480508, EPI_ISL_2480516, EPI_ISL_2480974, EPI_ISL_2481063, EPI_ISL_2481084, EPI_ISL_2481113, EPI_ISL_2481114, EPI_ISL_2481141, EPI_ISL_2481143, EPI_ISL_2481144, EPI_ISL_2481145, EPI_ISL_2481154, EPI_ISL_2481178, EPI_ISL_2481220, EPI_ISL_2481301, EPI_ISL_2481456 | | | |
| see above | Laboratory Corporation of America | Centers for Disease Control and Prevention Division of Viral Diseases, Pathogen Discovery | Dakota Howard, Dhwani Batra, Peter W. Cook, Kara Moser, Adrian Paskey, Jason Caravas, Benjamin Rambo-Martin, Shatavia Morrison, Christopher Gulvick, Scott Sammons, Yvette Unoarumhi, Darlene Wagner, Matthew Schmerer, Minoo Agarwal, Eyad Almasri, Debbie Boles, Ayla Burns, Nuthawin Charoensri, Oren Cohen, Susan Countryman, Mary Ann Cristobal, Bobbi Croy, Suzanne Dale, Hrushikesh Deshmukh, Amanda Douglas, Vincent Drouillon, Marcia Eisenberg, Howard Engler, Rama Ghatti, Prashant Gupta, Susan Hicks, Jake Humphrey, Lax Iyer, Lisa Pfefferle, Manoj Jain, Matthew Robinson, Mohan Kolli, Brian Krueger, Tim Kuphal, Stanley Letovsky, Michael Levandoski, Craig Lukasik, Jonathan Meltzer, Brian Norvell, Mindy Nye, Scott Parker, Christos Petropoulos, John Pruitt, Steven Ragan, Scott Ryan, Mike Sapeta, Jana Schroth, Suresh Babu Selvaraju, Goran Stevovic, Amanda Suchanek, Andrea Throop, Lyndon Tilson, Thomas Urban, Joe Voshell, Kimberly Wagner, Jonathan Williams, Mary Williamson, Qian Zeng, Tricia Zwiefelhofer, Clinton R. Paden, Duncan MacCannell |
| EPI_ISL_2482397, EPI_ISL_2489124, EPI_ISL_2489210, EPI_ISL_2489211, EPI_ISL_2489288, EPI_ISL_2489322, EPI_ISL_2489333, EPI_ISL_2489351, EPI_ISL_2489352, EPI_ISL_2489384, EPI_ISL_2489387, EPI_ISL_2489451, EPI_ISL_2489510, EPI_ISL_2489593, EPI_ISL_2489627, EPI_ISL_2489628, EPI_ISL_2489640, EPI_ISL_2489677, EPI_ISL_2489704, EPI_ISL_2489719, EPI_ISL_2489772, EPI_ISL_2489775, EPI_ISL_2489778, EPI_ISL_2489780, EPI_ISL_2489788, EPI_ISL_2489794, EPI_ISL_2489796, EPI_ISL_2489799, EPI_ISL_2489812, EPI_ISL_2489855 | | | |
| see above | Aegis Sciences Corporation | Centers for Disease Control and Prevention Division of Viral Diseases, Pathogen Discovery | Dakota Howard, Dhwani Batra, Peter W. Cook, Kara Moser, Adrian Paskey, Jason Caravas, Benjamin Rambo-Martin, Shatavia Morrison, Christopher Gulvick, Scott Sammons, Yvette Unoarumhi, Darlene Wagner, Matthew Schmerer, Cyndi Clark, Patrick Campbell, Rob Case, Vikramsinha Ghorpade, Holly Houdeshell, Ola Kvalvaag, Dillon Nall, Ethan Sanders, Alec Vest, Shaun Westlund, Matthew Hardison, Clinton R. Paden, Duncan MacCannell |
| EPI_ISL_2497001, EPI_ISL_2497002, EPI_ISL_2497003, EPI_ISL_2497004, EPI_ISL_2497005, EPI_ISL_2497006, EPI_ISL_2497007, EPI_ISL_2497008, EPI_ISL_2497009, EPI_ISL_2497010, EPI_ISL_2497011, EPI_ISL_2497012, EPI_ISL_2497013, EPI_ISL_2497014, EPI_ISL_2497015, EPI_ISL_2497016, EPI_ISL_2497017, EPI_ISL_2497018, EPI_ISL_2497019, EPI_ISL_2497020, EPI_ISL_2497021, EPI_ISL_2497022, EPI_ISL_2497023, EPI_ISL_2497024, EPI_ISL_2497025, EPI_ISL_2497026, EPI_ISL_2497027, EPI_ISL_2497028, EPI_ISL_2497029, EPI_ISL_2497030, EPI_ISL_2497031, EPI_ISL_2497032, EPI_ISL_2497033, EPI_ISL_2497034, EPI_ISL_2497035, EPI_ISL_2497036, EPI_ISL_2497037, EPI_ISL_2497038, EPI_ISL_2497039, EPI_ISL_2497040, EPI_ISL_2497041, EPI_ISL_2497042, EPI_ISL_2497043, EPI_ISL_2497044, EPI_ISL_2497045, EPI_ISL_2497046, EPI_ISL_2497047, EPI_ISL_2497048, EPI_ISL_2497049, EPI_ISL_2497050, EPI_ISL_2497051, EPI_ISL_2497052, EPI_ISL_2497053, EPI_ISL_2497054, EPI_ISL_2497055, EPI_ISL_2497056, EPI_ISL_2497057, EPI_ISL_2497058, EPI_ISL_2497059, EPI_ISL_2497060, EPI_ISL_2497061, EPI_ISL_2497062, EPI_ISL_2497063, EPI_ISL_2497064, EPI_ISL_2497065, EPI_ISL_2497066, EPI_ISL_2497067, EPI_ISL_2497068, EPI_ISL_2497069, EPI_ISL_2497070, EPI_ISL_2497071, EPI_ISL_2497072, EPI_ISL_2497073, EPI_ISL_2497074, EPI_ISL_2497075, EPI_ISL_2497076, EPI_ISL_2497077, EPI_ISL_2497078, EPI_ISL_2497079, EPI_ISL_2497080, EPI_ISL_2497081, EPI_ISL_2497082, EPI_ISL_2497083, EPI_ISL_2497084, EPI_ISL_2497085, EPI_ISL_2497086, EPI_ISL_2497087, EPI_ISL_2497088, EPI_ISL_2497089, EPI_ISL_2497090, EPI_ISL_2497091, EPI_ISL_2497092, EPI_ISL_2497093, EPI_ISL_2497094, EPI_ISL_2497095, EPI_ISL_2497096, EPI_ISL_2497097, EPI_ISL_2497098, EPI_ISL_2497099, EPI_ISL_2497100, EPI_ISL_2497101, EPI_ISL_2497102, EPI_ISL_2497103, EPI_ISL_2497104, EPI_ISL_2497105, EPI_ISL_2497106, EPI_ISL_2497107, EPI_ISL_2497108, EPI_ISL_2497109, EPI_ISL_2497110, EPI_ISL_2497111, EPI_ISL_2497112, EPI_ISL_2497113, EPI_ISL_2497114, EPI_ISL_2497115, EPI_ISL_2497116, EPI_ISL_2497117, EPI_ISL_2497118, EPI_ISL_2497119, EPI_ISL_2497120, EPI_ISL_2497121, EPI_ISL_2497122, EPI_ISL_2497123, EPI_ISL_2497124, EPI_ISL_2497125, EPI_ISL_2497126, EPI_ISL_2497127, EPI_ISL_2497128, EPI_ISL_2497129, EPI_ISL_2497130, EPI_ISL_2497131, EPI_ISL_2497132, EPI_ISL_2497133, EPI_ISL_2497134, EPI_ISL_2497135, EPI_ISL_2497136, EPI_ISL_2497137, EPI_ISL_2497138, EPI_ISL_2497139, EPI_ISL_2497140, EPI_ISL_2497141, EPI_ISL_2497142, EPI_ISL_2497143, EPI_ISL_2497144, EPI_ISL_2497145, EPI_ISL_2497146, EPI_ISL_2497147, EPI_ISL_2497148, EPI_ISL_2497149, EPI_ISL_2497150, EPI_ISL_2497151, EPI_ISL_2497152, EPI_ISL_2497153, EPI_ISL_2497154, EPI_ISL_2497155, EPI_ISL_2497156, EPI_ISL_2497157, EPI_ISL_2497158, EPI_ISL_2497159, EPI_ISL_2497160, EPI_ISL_2497161, EPI_ISL_2497162, EPI_ISL_2497163, EPI_ISL_2497164, EPI_ISL_2497165, EPI_ISL_2497166, EPI_ISL_2497167, EPI_ISL_2497168, EPI_ISL_2497169, EPI_ISL_2497170, EPI_ISL_2497171, EPI_ISL_2497172, EPI_ISL_2497173, EPI_ISL_2497174, EPI_ISL_2497175, EPI_ISL_2497176, EPI_ISL_2497177, EPI_ISL_2497178, EPI_ISL_2497179, EPI_ISL_2497180, EPI_ISL_2497181, EPI_ISL_2497182, EPI_ISL_2497183, EPI_ISL_2497184 | | | |
| see above | Cayuga Medical Center | Cornell Covid-19 Testing Laboratory | Patrick Mitchell, Leonardo Caserta, Brittany Cronk, Renee Anderson, Melissa Laverack, Roopa Venugopalan, Elizabeth Plocharczyk, Diego Diel |
| EPI_ISL_2500399, EPI_ISL_2500400, EPI_ISL_2500401, EPI_ISL_2500402 | IVY3 Central Lab, Vanderbilt University Medical Center | Lauring Lab, University of Michigan, Department of Microbiology and Immunology | Valesano |
| EPI_ISL_2500544, EPI_ISL_2500545, EPI_ISL_2500546, EPI_ISL_2500547, EPI_ISL_2500548, EPI_ISL_2500549, EPI_ISL_2500550, EPI_ISL_2500551, EPI_ISL_2500552, EPI_ISL_2500553, EPI_ISL_2500554, EPI_ISL_2500555, EPI_ISL_2500556, EPI_ISL_2500557, EPI_ISL_2500558, EPI_ISL_2500559, EPI_ISL_2500560, EPI_ISL_2500561, EPI_ISL_2500562, EPI_ISL_2500563, EPI_ISL_2500564, EPI_ISL_2500565, EPI_ISL_2500566, EPI_ISL_2500567, EPI_ISL_2500568, EPI_ISL_2500569, EPI_ISL_2500570, EPI_ISL_2500571, EPI_ISL_2500572, EPI_ISL_2500573, EPI_ISL_2500574, EPI_ISL_2500575, EPI_ISL_2500576, EPI_ISL_2500577, EPI_ISL_2500578, EPI_ISL_2500579, EPI_ISL_2500580, EPI_ISL_2500581, EPI_ISL_2500582, EPI_ISL_2500583, EPI_ISL_2500584, EPI_ISL_2500585, EPI_ISL_2500586, EPI_ISL_2500587, EPI_ISL_2500588, EPI_ISL_2500589, EPI_ISL_2500590, EPI_ISL_2500591, EPI_ISL_2500592, EPI_ISL_2500593, EPI_ISL_2500594, EPI_ISL_2500595, EPI_ISL_2500596, EPI_ISL_2500597, EPI_ISL_2500598, EPI_ISL_2500599, EPI_ISL_2500600, EPI_ISL_2500601, EPI_ISL_2500602, EPI_ISL_2500603, EPI_ISL_2500604, EPI_ISL_2500605, EPI_ISL_2500606, EPI_ISL_2500607, EPI_ISL_2500608, EPI_ISL_2500609, EPI_ISL_2500610, EPI_ISL_2500611, EPI_ISL_2500612, EPI_ISL_2500613, EPI_ISL_2500614, EPI_ISL_2500615, EPI_ISL_2500616, EPI_ISL_2500617, EPI_ISL_2500618, EPI_ISL_2500619, EPI_ISL_2500620, EPI_ISL_2500621, EPI_ISL_2500622, EPI_ISL_2500623, EPI_ISL_2500624, EPI_ISL_2500625, EPI_ISL_2500626, EPI_ISL_2500627, EPI_ISL_2500628, EPI_ISL_2500629, EPI_ISL_2500630, EPI_ISL_2500631, EPI_ISL_2500632, EPI_ISL_2500633, EPI_ISL_2500634, EPI_ISL_2500635, EPI_ISL_2500636, EPI_ISL_2500637, EPI_ISL_2500638, EPI_ISL_2500639, EPI_ISL_2500640, EPI_ISL_2500641, EPI_ISL_2500642, EPI_ISL_2500643, EPI_ISL_2500644, EPI_ISL_2500645, EPI_ISL_2500646, EPI_ISL_2500647, EPI_ISL_2500648, EPI_ISL_2500649, EPI_ISL_2500650, EPI_ISL_2500651, EPI_ISL_2500652, EPI_ISL_2500653, EPI_ISL_2500654, EPI_ISL_2500655, EPI_ISL_2500656, EPI_ISL_2500657, EPI_ISL_2500658, EPI_ISL_2500659, EPI_ISL_2500660, EPI_ISL_2500661, EPI_ISL_2500662, EPI_ISL_2500663, EPI_ISL_2500664, EPI_ISL_2500665, EPI_ISL_2500666, EPI_ISL_2500667, EPI_ISL_2500668, EPI_ISL_2500669, EPI_ISL_2500670, EPI_ISL_2500671, EPI_ISL_2500672, EPI_ISL_2500673, EPI_ISL_2500674, EPI_ISL_2500675, EPI_ISL_2500676, EPI_ISL_2500677, EPI_ISL_2500678, EPI_ISL_2500679, EPI_ISL_2500680, EPI_ISL_2500681, EPI_ISL_2500682, EPI_ISL_2500683, EPI_ISL_2500684, EPI_ISL_2500685, EPI_ISL_2500686, EPI_ISL_2500687, EPI_ISL_2500688, EPI_ISL_2500689, EPI_ISL_2500690, EPI_ISL_2500691, EPI_ISL_2500692, EPI_ISL_2500693, EPI_ISL_2500694, EPI_ISL_2500695, EPI_ISL_2500696, EPI_ISL_2500697, EPI_ISL_2500698, EPI_ISL_2500699, EPI_ISL_2500700, EPI_ISL_2500701, EPI_ISL_2500702, EPI_ISL_2500703, EPI_ISL_2500704, EPI_ISL_2500705, EPI_ISL_2500706, EPI_ISL_2500707, EPI_ISL_2500708, EPI_ISL_2500709, EPI_ISL_2500710, EPI_ISL_2500711, EPI_ISL_2500712, EPI_ISL_2500713, EPI_ISL_2500714, EPI_ISL_2500715, EPI_ISL_2500716, EPI_ISL_2500717, EPI_ISL_2500718, EPI_ISL_2500719, EPI_ISL_2500720, EPI_ISL_2500721, EPI_ISL_2500722, EPI_ISL_2500723, EPI_ISL_2500724, EPI_ISL_2500725, EPI_ISL_2500726, EPI_ISL_2500727, EPI_ISL_2500728, EPI_ISL_2500729, EPI_ISL_2500730, EPI_ISL_2500731, EPI_ISL_2500732, EPI_ISL_2500733, EPI_ISL_2500734, EPI_ISL_2500735, EPI_ISL_2500736, EPI_ISL_2500737, EPI_ISL_2500738, EPI_ISL_2500739, EPI_ISL_2500740, EPI_ISL_2500741, EPI_ISL_2500742, EPI_ISL_2500743, EPI_ISL_2500744, EPI_ISL_2500745, EPI_ISL_2500746, EPI_ISL_2500747, EPI_ISL_2500748, EPI_ISL_2500749, EPI_ISL_2500750, EPI_ISL_2500751, EPI_ISL_2500752, EPI_ISL_2500753, EPI_ISL_2500754, EPI_ISL_2500755, EPI_ISL_2500756, EPI_ISL_2500757, EPI_ISL_2500758, EPI_ISL_2500759, EPI_ISL_2500760, EPI_ISL_2500761, EPI_ISL_2500762, EPI_ISL_2500763, EPI_ISL_2500764, EPI_ISL_2500765, EPI_ISL_2500766, EPI_ISL_2500767, EPI_ISL_2500768, EPI_ISL_2500769, EPI_ISL_2500770, EPI_ISL_2500771, EPI_ISL_2500772, EPI_ISL_2500773, EPI_ISL_2500774, EPI_ISL_2500775, EPI_ISL_2500776, EPI_ISL_2500777, EPI_ISL_2500778, EPI_ISL_2500779, EPI_ISL_2500780, EPI_ISL_2500781, EPI_ISL_2500782, EPI_ISL_2500783, EPI_ISL_2500784, EPI_ISL_2500785, EPI_ISL_2500786, EPI_ISL_2500787, EPI_ISL_2500788, EPI_ISL_2500789, EPI_ISL_2500790, EPI_ISL_2500791, EPI_ISL_2500792, EPI_ISL_2500793, EPI_ISL_2500794, EPI_ISL_2500795, EPI_ISL_2500796, EPI_ISL_2500797, EPI_ISL_2500798, EPI_ISL_2500799, EPI_ISL_2500800, EPI_ISL_2500801, EPI_ISL_2500802, EPI_ISL_2500803, EPI_ISL_2500804, EPI_ISL_2500805, EPI_ISL_2500806, EPI_ISL_2500807, EPI_ISL_2500808, EPI_ISL_2500809, EPI_ISL_2500810, EPI_ISL_2500811, EPI_ISL_2500812, EPI_ISL_2500813, EPI_ISL_2500814, EPI_ISL_2500815, EPI_ISL_2500816, EPI_ISL_2500817, EPI_ISL_2500818, EPI_ISL_2500819, EPI_ISL_2500820, EPI_ISL_2500821, EPI_ISL_2500822, EPI_ISL_2500823, EPI_ISL_2500824, EPI_ISL_2500825, EPI_ISL_2500826, EPI_ISL_2500827, EPI_ISL_2500828, EPI_ISL_2500829, EPI_ISL_2500830, EPI_ISL_2500831, EPI_ISL_2500832, EPI_ISL_2500833, EPI_ISL_2500834, EPI_ISL_2500835, EPI_ISL_2500836, EPI_ISL_2500837, EPI_ISL_2500838, EPI_ISL_2500839, EPI_ISL_2500840, EPI_ISL_2500841, EPI_ISL_2500842, EPI_ISL_2500843, EPI_ISL_2500844, EPI_ISL_2500845, EPI_ISL_2500846, EPI_ISL_2500847, EPI_ISL_2500848 | | | |
| see above | Clinical Microbiology Laboratory, NewYork Presbyterian Hospital/Columbia University Irving Medical Center | Uhlemann Laboratory, Columbia University Irving Medical Center | Medini K. Annavajhala, Anne Kelley, Maya Tagliavia, Anne-Catrin Uhlemann |
| EPI_ISL_2502795, EPI_ISL_2502796, EPI_ISL_2502797, EPI_ISL_2502798, EPI_ISL_2502799, EPI_ISL_2502800, EPI_ISL_2502801, EPI_ISL_2502802, EPI_ISL_2502803, EPI_ISL_2502804, EPI_ISL_2502805, EPI_ISL_2502806, EPI_ISL_2502807, EPI_ISL_2502808, EPI_ISL_2502809, EPI_ISL_2502810, EPI_ISL_2502811, EPI_ISL_2502812, EPI_ISL_2502828 | | | |
| see above | UW Virology Lab | UW Virology Lab | Pavitra Roychoudhury, Hong Xie, Lasata Shrestha, Shah Mohamed Bakhash, Margaret Mills, Noah Baker, Sean Ellis, Meei-Li Huang, Keith R Jerome, Alexander Greninger |
| EPI_ISL_2525982, EPI_ISL_2525987, EPI_ISL_2525991, EPI_ISL_2526002, EPI_ISL_2526015, EPI_ISL_2526017, EPI_ISL_2526024, EPI_ISL_2526028 | Fulgent Genetics | Centers for Disease Control and Prevention Division of Viral Diseases, Pathogen Discovery | Dakota Howard, Dhwani Batra, Peter W. Cook, Kara Moser, Adrian Paskey, Jason Caravas, Benjamin Rambo-Martin, Shatavia Morrison, Christopher Gulvick, Scott Sammons, Yvette Unoarumhi, Darlene Wagner, Matthew Schmerer, Harry Gao, Mickey Li, John Gao, Joseph Fierro, Benafsh Sapra, Becky Tsai, Yan Meng, Doreen Ng, James Xie, Clinton R. Paden, Duncan MacCannell |
| EPI_ISL_2526777, EPI_ISL_2526792, EPI_ISL_2526811, EPI_ISL_2526993 | Quest Diagnostics Incorporated | Centers for Disease Control and Prevention Division of Viral Diseases, Pathogen Discovery | Dakota Howard, Dhwani Batra, Peter W. Cook, Kara Moser, Adrian Paskey, Jason Caravas, Benjamin Rambo-Martin, Shatavia Morrison, Christopher Gulvick, Scott Sammons, Yvette Unoarumhi, Darlene Wagner, Matthew Schmerer, S. H. Rosenthal, A. Gerasimova, R. M. Kagan, B. Anderson, M. Hua, Y. Liu, L.E. Bernstein, K.E. Livingston, A. Perez, I. A. Shlyakhter, R. V. Rolando, R. Owen, P. Tanpaiboon, F. Lacbawan, Clinton R. Paden, Duncan MacCannell |
| EPI_ISL_2527874, EPI_ISL_2527876, EPI_ISL_2527878, EPI_ISL_2528010, EPI_ISL_2528470, EPI_ISL_2528494, EPI_ISL_2528889, EPI_ISL_2529020 | Aegis Sciences Corporation | Centers for Disease Control and Prevention Division of Viral Diseases, Pathogen Discovery | Dakota Howard, Dhwani Batra, Peter W. Cook, Kara Moser, Adrian Paskey, Jason Caravas, Benjamin Rambo-Martin, Shatavia Morrison, Christopher Gulvick, Scott Sammons, Yvette Unoarumhi, Darlene Wagner, Matthew Schmerer, Cyndi Clark, Patrick Campbell, Rob Case, Vikramsinha Ghorpade, Holly Houdeshell, Ola Kvalvaag, Dillon Nall, Ethan Sanders, Alec Vest, Shaun Westlund, Matthew Hardison, Clinton R. Paden, Duncan MacCannell |
| EPI_ISL_2529340 | Quest Diagnostics Incorporated | Centers for Disease Control and Prevention Division of Viral Diseases, Pathogen Discovery | Dakota Howard, Dhwani Batra, Peter W. Cook, Kara Moser, Adrian Paskey, Jason Caravas, Benjamin Rambo-Martin, Shatavia Morrison, Christopher Gulvick, Scott Sammons, Yvette Unoarumhi, Darlene Wagner, Matthew Schmerer, S. H. Rosenthal, A. Gerasimova, R. M. Kagan, B. Anderson, M. Hua, Y. Liu, L.E. Bernstein, K.E. Livingston, A. Perez, I. A. Shlyakhter, R. V. Rolando, R. Owen, P. Tanpaiboon, F. Lacbawan, Clinton R. Paden, Duncan MacCannell |
| EPI_ISL_2530983 | Aegis Sciences Corporation | Centers for Disease Control and Prevention Division of Viral Diseases, Pathogen Discovery | Dakota Howard, Dhwani Batra, Peter W. Cook, Kara Moser, Adrian Paskey, Jason Caravas, Benjamin Rambo-Martin, Shatavia Morrison, Christopher Gulvick, Scott Sammons, Yvette Unoarumhi, Darlene Wagner, Matthew Schmerer, Cyndi Clark, Patrick Campbell, Rob Case, Vikramsinha Ghorpade, Holly Houdeshell, Ola Kvalvaag, Dillon Nall, Ethan Sanders, Alec Vest, Shaun Westlund, Matthew Hardison, Clinton R. Paden, Duncan MacCannell |
| EPI_ISL_2534334 | IVY3 Central Lab, Vanderbilt University Medical Center | Lauring Lab, University of Michigan, Department of Microbiology and Immunology | Valesano |
| EPI_ISL_2537215 | Gravity Diagnostics, LLC | Gravity Diagnostics, LLC | Gravity Diagnostics |
| EPI_ISL_2576919, EPI_ISL_2576922, EPI_ISL_2576923, EPI_ISL_2576924 | Sonic Reference Laboratory | Sonic Reference Laboratory | "Robert Moller, Bijaya Dhakal, Krishna Maddula, John Woody, Cora Lahey, Ellen Pizarek" |

| | | | |
|---|---|---|---|
| EPI_ISL_2598282, EPI_ISL_2598283, EPI_ISL_2598284, EPI_ISL_2598285, EPI_ISL_2598286 | IVY3 Central Lab, Vanderbilt University Medical Center | Lauring Lab, University of Michigan, Department of Microbiology and Immunology | Valesano |
| EPI_ISL_2598768, EPI_ISL_2598801, EPI_ISL_2598802, EPI_ISL_2598805, EPI_ISL_2598806 | Fulgent Genetics | Centers for Disease Control and Prevention Division of Viral Diseases, Pathogen Discovery | Dakota Howard, Dhwani Batra, Peter W. Cook, Kara Moser, Adrian Paskey, Jason Caravas, Benjamin Rambo-Martin, Shatavia Morrison, Christopher Gulvick, Scott Sammons, Yvette Unoarumhi, Darlene Wagner, Matthew Schmerer, Harry Gao, Mickey Li, John Gao, Joseph Fierro, Benafsh Sapra, Becky Tsai, Yan Meng, Doreen Ng, James Xie, Clinton R. Paden, Duncan MacCannell |
| EPI_ISL_2599069 | Helix/Illumina | Centers for Disease Control and Prevention Division of Viral Diseases, Pathogen Discovery | Dakota Howard, Dhwani Batra, Peter W. Cook, Kara Moser, Adrian Paskey, Jason Caravas, Benjamin Rambo-Martin, Shatavia Morrison, Christopher Gulvick, Scott Sammons, Yvette Unoarumhi, Darlene Wagner, Matthew Schmerer, Eileen de Feo, Jan Antico, Christine Tran, Matthew Tolentino, Shannon Wickline, Kim Gietzen, Brad Sickler, Jingtao Liu, Eric Allen, Phil Febbo, Nicole L. Washington, Simon White, Geraint Levan, Kelly Schiabor Barrett, Elizabeth Cirulli, Alexandre Bolze, Ary Ascencio, Charlotte Rivera-Garcia, Ryan Cho, Jason Nguyen, Sherry Wang, Jimmy Ramirez, Tyler Cassens, Efren Sandoval, Magnus Isaksson, William Lee, David Becker, Marc Laurent, James Lu, Clinton R. Paden, Duncan MacCannell |
| EPI_ISL_2599757, EPI_ISL_2599846, EPI_ISL_2599860 | Quest Diagnostics Incorporated | Centers for Disease Control and Prevention Division of Viral Diseases, Pathogen Discovery | Dakota Howard, Dhwani Batra, Peter W. Cook, Kara Moser, Adrian Paskey, Jason Caravas, Benjamin Rambo-Martin, Shatavia Morrison, Christopher Gulvick, Scott Sammons, Yvette Unoarumhi, Darlene Wagner, Matthew Schmerer, S. H. Rosenthal, A. Gerasimova, R. M. Kagan, B. Anderson, M. Hua, Y. Liu, L.E. Bernstein, K.E. Livingston, A. Perez, I. A. Shlyakhter, R. V. Rolando, R. Owen, P. Tanpaiboon, F. Lacbawan, Clinton R. Paden, Duncan MacCannell |
| EPI_ISL_2599935, EPI_ISL_2599942, EPI_ISL_2599944, EPI_ISL_2600021, EPI_ISL_2600095, EPI_ISL_2600096, EPI_ISL_2600097, EPI_ISL_2600156, EPI_ISL_2600157 | Aegis Sciences Corporation | Centers for Disease Control and Prevention Division of Viral Diseases, Pathogen Discovery | Dakota Howard, Dhwani Batra, Peter W. Cook, Kara Moser, Adrian Paskey, Jason Caravas, Benjamin Rambo-Martin, Shatavia Morrison, Christopher Gulvick, Scott Sammons, Yvette Unoarumhi, Darlene Wagner, Matthew Schmerer, Cyndi Clark, Patrick Campbell, Rob Case, Vikramsinha Ghorpade, Holly Houdeshell, Ola Kvalvaag, Dillon Nall, Ethan Sanders, Alec Vest, Shaun Westlund, Matthew Hardison, Clinton R. Paden, Duncan MacCannell |
| EPI_ISL_2602157 | Gravity Diagnostics, LLC | Gravity Diagnostics, LLC | Gravity Diagnostics |
| EPI_ISL_2603970, EPI_ISL_2603971, EPI_ISL_2603972, EPI_ISL_2603973, EPI_ISL_2603974, EPI_ISL_2603980, EPI_ISL_2603981, EPI_ISL_2603982, EPI_ISL_2603983, EPI_ISL_2603984, EPI_ISL_2603985, EPI_ISL_2603986, EPI_ISL_2603987, EPI_ISL_2603988, EPI_ISL_2603989, EPI_ISL_2603990, EPI_ISL_2603991, EPI_ISL_2603992, EPI_ISL_2603993, EPI_ISL_2603994, EPI_ISL_2603995, EPI_ISL_2603996, EPI_ISL_2603997, EPI_ISL_2603998, EPI_ISL_2603999, EPI_ISL_2604000, EPI_ISL_2604001, EPI_ISL_2604002, EPI_ISL_2604003, EPI_ISL_2604004, EPI_ISL_2604005, EPI_ISL_2604006, EPI_ISL_2604007, EPI_ISL_2604008, EPI_ISL_2604009, EPI_ISL_2604010, EPI_ISL_2604011, EPI_ISL_2604012, EPI_ISL_2604013, EPI_ISL_2604014, EPI_ISL_2604025, EPI_ISL_2604026, EPI_ISL_2604027, EPI_ISL_2604028, EPI_ISL_2604029, EPI_ISL_2604030, EPI_ISL_2604031, EPI_ISL_2604032, EPI_ISL_2604054, EPI_ISL_2604055, EPI_ISL_2604056, EPI_ISL_2604057, EPI_ISL_2604058, EPI_ISL_2604059, EPI_ISL_2604060, EPI_ISL_2604061, EPI_ISL_2604062, EPI_ISL_2604063, EPI_ISL_2604064, EPI_ISL_2604065, EPI_ISL_2604066, EPI_ISL_2604067, EPI_ISL_2604068, EPI_ISL_2604069, EPI_ISL_2604070 | | | |
| see above | UW Virology Lab | UW Virology Lab | Pavitra Roychoudhury, Hong Xie, Lasata Shrestha, Shah Mohamed Bakhash, Margaret Mills, Noah Baker, Sean Ellis, Meei-Li Huang, Keith R Jerome, Alexander Greninger |
| EPI_ISL_2611155, EPI_ISL_2611156, EPI_ISL_2611157, EPI_ISL_2611301, EPI_ISL_2611302, EPI_ISL_2611303, EPI_ISL_2611308, EPI_ISL_2611335, EPI_ISL_2611336, EPI_ISL_2611359, EPI_ISL_2611360, EPI_ISL_2611361, EPI_ISL_2611436, EPI_ISL_2611467, EPI_ISL_2611502, EPI_ISL_2611506, EPI_ISL_2611516, EPI_ISL_2611550, EPI_ISL_2611593, EPI_ISL_2611648, EPI_ISL_2611663, EPI_ISL_2611720, EPI_ISL_2611754, EPI_ISL_2611755 | | | |
| see above | Laboratory Corporation of America | Centers for Disease Control and Prevention Division of Viral Diseases, Pathogen Discovery | Dakota Howard, Dhwani Batra, Peter W. Cook, Kara Moser, Adrian Paskey, Jason Caravas, Benjamin Rambo-Martin, Shatavia Morrison, Christopher Gulvick, Scott Sammons, Yvette Unoarumhi, Darlene Wagner, Matthew Schmerer, Minoo Agarwal, Eyad Almasri, Debbie Boles, Ayla Burns, Nuthawin Charoensri, Oren Cohen, Susan Countryman, Mary Ann Cristobal, Bobbi Croy, Suzanne Dale, Hrushikesh Deshmukh, Amanda Douglas, Vincent Drouillon, Marcia Eisenberg, Howard Engler, Rama Ghatti, Prashant Gupta, Susan Hicks, Jake Humphrey, Lax Iyer, Lisa Pfefferle, Manoj Jain, Matthew Robinson, Mohan Kolli, Brian Krueger, Tim Kuphal, Stanley Letovsky, Michael Levandoski, Craig Lukasik, Jonathan Meltzer, Brian Norvell, Mindy Nye, Scott Parker, Christos Petropoulos, John Pruitt, Steven Ragan, Scott Ryan, Mike Sapeta, Jana Schroth, Suresh Babu Selvaraju, Goran Stevovic, Amanda Suchanek, Andrea Throop, Lyndon Tilson, Thomas Urban, Joe Voshell, Kimberly Wagner, Jonathan Williams, Mary Williamson, Qian Zeng, Tricia Zwiefelhofer, Clinton R. Paden, Duncan MacCannell |
| EPI_ISL_2611828, EPI_ISL_2611830, EPI_ISL_2611834, EPI_ISL_2611841 | Quest Diagnostics Incorporated | Centers for Disease Control and Prevention Division of Viral Diseases, Pathogen Discovery | Dakota Howard, Dhwani Batra, Peter W. Cook, Kara Moser, Adrian Paskey, Jason Caravas, Benjamin Rambo-Martin, Shatavia Morrison, Christopher Gulvick, Scott Sammons, Yvette Unoarumhi, Darlene Wagner, Matthew Schmerer, S. H. Rosenthal, A. Gerasimova, R. M. Kagan, B. Anderson, M. Hua, Y. Liu, L.E. Bernstein, K.E. Livingston, A. Perez, I. A. Shlyakhter, R. V. Rolando, R. Owen, P. Tanpaiboon, F. Lacbawan, Clinton R. Paden, Duncan MacCannell |
| EPI_ISL_2612299 | IVY3 Central Lab, Vanderbilt University Medical Center | Lauring Lab, University of Michigan, Department of Microbiology and Immunology | Valesano |
| EPI_ISL_2612552 | EastGene Laboratories | New York City Public Health Laboratory | Jade Wang, et al. |
| EPI_ISL_2612553, EPI_ISL_2612554, EPI_ISL_2612555, EPI_ISL_2612556 | DOHMH Corona | New York City Public Health Laboratory | Jade Wang, et al. |
| EPI_ISL_2612557, EPI_ISL_2612558, EPI_ISL_2612559 | DOHMH Jamaica | New York City Public Health Laboratory | Jade Wang, et al. |
| EPI_ISL_2612560, EPI_ISL_2612561 | DOHMH Riverside | New York City Public Health Laboratory | Jade Wang, et al. |
| EPI_ISL_2612562 | DOHMH Jamaica | New York City Public Health Laboratory | Jade Wang, et al. |
| EPI_ISL_2612563 | OCME Office Of Chief Medical Examiner | New York City Public Health Laboratory | Jade Wang, et al. |
| EPI_ISL_2614010 | Gravity Diagnostics, LLC | Gravity Diagnostics, LLC | Gravity Diagnostics |
| EPI_ISL_2626923 | MONTEFIORE MEDICAL CENTER LABORATORIES | Wadsworth Center, New York State Department of Health | Kirsten St. George, Daryl M. Lamson, Alexis Russell, Matthew Shudt, Melissa A Leisner, Jonathan Plitnick, Catharine Prussing, Navjot Singh, John Kelly, Erasmus Schneider, Erica Lasek-Nesselquist |
| EPI_ISL_2626924 | WESTCHESTER MEDICAL CENTER | Wadsworth Center, New York State Department of Health | Kirsten St. George, Daryl M. Lamson, Alexis Russell, Matthew Shudt, Melissa A Leisner, Jonathan Plitnick, Catharine Prussing, Navjot Singh, John Kelly, Erasmus Schneider, Erica Lasek-Nesselquist |
| EPI_ISL_2626925, EPI_ISL_2626926, EPI_ISL_2626927, EPI_ISL_2626928, EPI_ISL_2626929, EPI_ISL_2626930, EPI_ISL_2626931, EPI_ISL_2626932, EPI_ISL_2626933, EPI_ISL_2626934, EPI_ISL_2626935, EPI_ISL_2626936, EPI_ISL_2626937, EPI_ISL_2626938, EPI_ISL_2626939, EPI_ISL_2626940, EPI_ISL_2626941, EPI_ISL_2626942, EPI_ISL_2626943, EPI_ISL_2626944, EPI_ISL_2626945, EPI_ISL_2626946, EPI_ISL_2626947, EPI_ISL_2626948, EPI_ISL_2626949, EPI_ISL_2626950, EPI_ISL_2626951, EPI_ISL_2626952, EPI_ISL_2626953, EPI_ISL_2626954, EPI_ISL_2626955, EPI_ISL_2626956, EPI_ISL_2626957, EPI_ISL_2626958, EPI_ISL_2626959, EPI_ISL_2626960, EPI_ISL_2626961, EPI_ISL_2626962, EPI_ISL_2626963, EPI_ISL_2626964, EPI_ISL_2626965, EPI_ISL_2626966, EPI_ISL_2626967, EPI_ISL_2626968, EPI_ISL_2626969, EPI_ISL_2626970, EPI_ISL_2626971 | | | |
| see above | TEMPUS LABS INC | Wadsworth Center, New York State Department of Health | Kirsten St. George, Daryl M. Lamson, Alexis Russell, Matthew Shudt, Melissa A Leisner, Jonathan Plitnick, Catharine Prussing, Navjot Singh, John Kelly, Erasmus Schneider, Erica Lasek-Nesselquist |
| EPI_ISL_2626972 | PECONIC BAY MEDICAL CENTER LABORATORY | Wadsworth Center, New York State Department of Health | Kirsten St. George, Daryl M. Lamson, Alexis Russell, Matthew Shudt, Melissa A Leisner, Jonathan Plitnick, Catharine Prussing, Navjot Singh, John Kelly, Erasmus Schneider, Erica Lasek-Nesselquist |
| EPI_ISL_2626973, EPI_ISL_2626974, EPI_ISL_2626975, EPI_ISL_2626976, EPI_ISL_2626977, EPI_ISL_2626978, EPI_ISL_2626979, EPI_ISL_2626980, EPI_ISL_2626981, EPI_ISL_2626982, EPI_ISL_2626983, EPI_ISL_2626984, EPI_ISL_2626985, EPI_ISL_2626986, EPI_ISL_2626987, EPI_ISL_2626988, EPI_ISL_2626989, EPI_ISL_2626990, EPI_ISL_2626991, EPI_ISL_2626992 | | | |
| see above | URMC LABS | Wadsworth Center, New York State Department of Health | Kirsten St. George, Daryl M. Lamson, Alexis Russell, Matthew Shudt, Melissa A Leisner, Jonathan Plitnick, Catharine Prussing, Navjot Singh, John Kelly, Erasmus Schneider, Erica Lasek-Nesselquist |
| EPI_ISL_2626993, EPI_ISL_2626994, EPI_ISL_2626995, EPI_ISL_2626996, EPI_ISL_2626997, EPI_ISL_2626998 | MONTEFIORE MEDICAL CENTER LABORATORIES | Wadsworth Center, New York State Department of Health | Kirsten St. George, Daryl M. Lamson, Alexis Russell, Matthew Shudt, Melissa A Leisner, Jonathan Plitnick, Catharine Prussing, Navjot Singh, John Kelly, Erasmus Schneider, Erica Lasek-Nesselquist |
| EPI_ISL_2626999, EPI_ISL_2627000, EPI_ISL_2627001, EPI_ISL_2627002 | ALBANY MEDICAL CENTER | Wadsworth Center, New York State Department of Health | Kirsten St. George, Daryl M. Lamson, Alexis Russell, Matthew Shudt, Melissa A Leisner, Jonathan Plitnick, Catharine Prussing, Navjot Singh, John Kelly, Erasmus Schneider, Erica Lasek-Nesselquist |
| EPI_ISL_2627003, EPI_ISL_2627004, | Saratoga Hospital Laboratory | Wadsworth Center, New York State Department of Health | Kirsten St. George, Daryl M. Lamson, Alexis Russell, Matthew Shudt, Melissa A Leisner, Jonathan Plitnick, Catharine Prussing, Navjot Singh, John Kelly, |

| | | | |
|---|---|---|---|
| EPI_ISL_2627005, EPI_ISL_2627006 | | | Erasmus Schneider, Erica Lasek-Nesselquist |
| EPI_ISL_2627007, EPI_ISL_2627008, EPI_ISL_2627009, EPI_ISL_2627010, EPI_ISL_2627012, EPI_ISL_2627013, EPI_ISL_2627014, EPI_ISL_2627015, EPI_ISL_2627016, EPI_ISL_2627017, EPI_ISL_2627018, EPI_ISL_2627019, EPI_ISL_2627020, EPI_ISL_2627021, EPI_ISL_2627022, EPI_ISL_2627023, EPI_ISL_2627024, EPI_ISL_2627025, EPI_ISL_2627026, EPI_ISL_2627027, EPI_ISL_2627028, EPI_ISL_2627029, EPI_ISL_2627030, EPI_ISL_2627031, EPI_ISL_2627032, EPI_ISL_2627033, EPI_ISL_2627034, EPI_ISL_2627035, EPI_ISL_2627036, EPI_ISL_2627037, EPI_ISL_2627038, EPI_ISL_2627039, EPI_ISL_2627040, EPI_ISL_2627041, EPI_ISL_2627042, EPI_ISL_2627043, EPI_ISL_2627044, EPI_ISL_2627045, EPI_ISL_2627046, EPI_ISL_2627047, EPI_ISL_2627048, EPI_ISL_2627049, EPI_ISL_2627050, EPI_ISL_2627051, EPI_ISL_2627052, EPI_ISL_2627053, EPI_ISL_2627054, EPI_ISL_2627055, EPI_ISL_2627056, EPI_ISL_2627057, EPI_ISL_2627058, EPI_ISL_2627059, EPI_ISL_2627060, EPI_ISL_2627061, EPI_ISL_2627062, EPI_ISL_2627063, EPI_ISL_2627064, EPI_ISL_2627065, EPI_ISL_2627066, EPI_ISL_2627067, EPI_ISL_2627068, EPI_ISL_2627069, EPI_ISL_2627070, EPI_ISL_2627071, EPI_ISL_2627072, EPI_ISL_2627073, EPI_ISL_2627074, EPI_ISL_2627075, EPI_ISL_2627076, EPI_ISL_2627077, EPI_ISL_2627078, EPI_ISL_2627079, EPI_ISL_2627080, EPI_ISL_2627081, EPI_ISL_2627082, EPI_ISL_2627083, EPI_ISL_2627084, EPI_ISL_2627085, EPI_ISL_2627086, EPI_ISL_2627087, EPI_ISL_2627088, EPI_ISL_2627089, EPI_ISL_2627090, EPI_ISL_2627091, EPI_ISL_2627092, EPI_ISL_2627093, EPI_ISL_2627094, EPI_ISL_2627095, EPI_ISL_2627096, EPI_ISL_2627097, EPI_ISL_2627098, EPI_ISL_2627099, EPI_ISL_2627100, EPI_ISL_2627101, EPI_ISL_2627102, EPI_ISL_2627103, EPI_ISL_2627104, EPI_ISL_2627105, EPI_ISL_2627106, EPI_ISL_2627107, EPI_ISL_2627108, EPI_ISL_2627109, EPI_ISL_2627110, EPI_ISL_2627111, EPI_ISL_2627112, EPI_ISL_2627113, EPI_ISL_2627114 | | | |
| see above | URMC LABS | Wadsworth Center, New York State Department of Health | Kirsten St. George, Daryl M. Lamson, Alexis Russell, Matthew Shudt, Melissa A Leisner, Jonathan Plitnick, Catharine Prussing, Navjot Singh, John Kelly, Erasmus Schneider, Erica Lasek-Nesselquist |
| EPI_ISL_2627115, EPI_ISL_2627116, EPI_ISL_2627117, EPI_ISL_2627118, EPI_ISL_2627119, EPI_ISL_2627120, EPI_ISL_2627121, EPI_ISL_2627122, EPI_ISL_2627123, EPI_ISL_2627124, EPI_ISL_2627125, EPI_ISL_2627126, EPI_ISL_2627127, EPI_ISL_2627128, EPI_ISL_2627129, EPI_ISL_2627130, EPI_ISL_2627131, EPI_ISL_2627132, EPI_ISL_2627133, EPI_ISL_2627134 | | | |
| see above | TEMPUS LABS INC | Wadsworth Center, New York State Department of Health | Kirsten St. George, Daryl M. Lamson, Alexis Russell, Matthew Shudt, Melissa A Leisner, Jonathan Plitnick, Catharine Prussing, Navjot Singh, John Kelly, Erasmus Schneider, Erica Lasek-Nesselquist |
| EPI_ISL_2627135, EPI_ISL_2627136, EPI_ISL_2627137 | BOSTON HEART DIAGNOSTICS CORP | Wadsworth Center, New York State Department of Health | Kirsten St. George, Daryl M. Lamson, Alexis Russell, Matthew Shudt, Melissa A Leisner, Jonathan Plitnick, Catharine Prussing, Navjot Singh, John Kelly, Erasmus Schneider, Erica Lasek-Nesselquist |
| EPI_ISL_2627138, EPI_ISL_2627139, EPI_ISL_2627140, EPI_ISL_2627141 | WESTCHESTER MEDICAL CENTER | Wadsworth Center, New York State Department of Health | Kirsten St. George, Daryl M. Lamson, Alexis Russell, Matthew Shudt, Melissa A Leisner, Jonathan Plitnick, Catharine Prussing, Navjot Singh, John Kelly, Erasmus Schneider, Erica Lasek-Nesselquist |
| EPI_ISL_2627142 | SBH Health System | Wadsworth Center, New York State Department of Health | Kirsten St. George, Daryl M. Lamson, Alexis Russell, Matthew Shudt, Melissa A Leisner, Jonathan Plitnick, Catharine Prussing, Navjot Singh, John Kelly, Erasmus Schneider, Erica Lasek-Nesselquist |
| EPI_ISL_2627143, EPI_ISL_2627144, EPI_ISL_2627145, EPI_ISL_2627146, EPI_ISL_2627147 | MONTEFIORE MEDICAL CENTER LABORATORIES | Wadsworth Center, New York State Department of Health | Kirsten St. George, Daryl M. Lamson, Alexis Russell, Matthew Shudt, Melissa A Leisner, Jonathan Plitnick, Catharine Prussing, Navjot Singh, John Kelly, Erasmus Schneider, Erica Lasek-Nesselquist |
| EPI_ISL_2627148, EPI_ISL_2627149, EPI_ISL_2627150, EPI_ISL_2627151, EPI_ISL_2627152 | STONY BROOK UNIVERSITY HOSPITAL | Wadsworth Center, New York State Department of Health | Kirsten St. George, Daryl M. Lamson, Alexis Russell, Matthew Shudt, Melissa A Leisner, Jonathan Plitnick, Catharine Prussing, Navjot Singh, John Kelly, Erasmus Schneider, Erica Lasek-Nesselquist |
| EPI_ISL_2627153, EPI_ISL_2627154, EPI_ISL_2627155, EPI_ISL_2627156 | NORTHWELL HEALTH LABORATORIES | Wadsworth Center, New York State Department of Health | Kirsten St. George, Daryl M. Lamson, Alexis Russell, Matthew Shudt, Melissa A Leisner, Jonathan Plitnick, Catharine Prussing, Navjot Singh, John Kelly, Erasmus Schneider, Erica Lasek-Nesselquist |
| EPI_ISL_2627157, EPI_ISL_2627158, EPI_ISL_2627159, EPI_ISL_2627160, EPI_ISL_2627161, EPI_ISL_2627162, EPI_ISL_2627163 | ALBANY MEDICAL CENTER | Wadsworth Center, New York State Department of Health | Kirsten St. George, Daryl M. Lamson, Alexis Russell, Matthew Shudt, Melissa A Leisner, Jonathan Plitnick, Catharine Prussing, Navjot Singh, John Kelly, Erasmus Schneider, Erica Lasek-Nesselquist |
| EPI_ISL_2627164, EPI_ISL_2627165, EPI_ISL_2627166, EPI_ISL_2627167, EPI_ISL_2627168, EPI_ISL_2627169, EPI_ISL_2627170, EPI_ISL_2627171, EPI_ISL_2627172, EPI_ISL_2627173, EPI_ISL_2627174, EPI_ISL_2627175, EPI_ISL_2627176, EPI_ISL_2627177, EPI_ISL_2627178, EPI_ISL_2627179 | | | |
| see above | NORTHWELL HEALTH LABORATORIES | Wadsworth Center, New York State Department of Health | Kirsten St. George, Daryl M. Lamson, Alexis Russell, Matthew Shudt, Melissa A Leisner, Jonathan Plitnick, Catharine Prussing, Navjot Singh, John Kelly, Erasmus Schneider, Erica Lasek-Nesselquist |
| EPI_ISL_2631338, EPI_ISL_2631339, EPI_ISL_2631340, EPI_ISL_2631341, EPI_ISL_2631342, EPI_ISL_2631343, EPI_ISL_2631344, EPI_ISL_2631345, EPI_ISL_2631346, EPI_ISL_2631347, EPI_ISL_2631348, EPI_ISL_2631349, EPI_ISL_2631350, EPI_ISL_2631351, EPI_ISL_2631352, EPI_ISL_2631353, EPI_ISL_2631354, EPI_ISL_2631355, EPI_ISL_2631356, EPI_ISL_2631357, EPI_ISL_2631358, EPI_ISL_2631359, EPI_ISL_2631360, EPI_ISL_2631361, EPI_ISL_2631362, EPI_ISL_2631363, EPI_ISL_2631364, EPI_ISL_2631365, EPI_ISL_2631366, EPI_ISL_2631367, EPI_ISL_2631368, EPI_ISL_2631369, EPI_ISL_2631370, EPI_ISL_2631371, EPI_ISL_2631372, EPI_ISL_2631373, EPI_ISL_2631374, EPI_ISL_2631375, EPI_ISL_2631376, EPI_ISL_2631377, EPI_ISL_2631378, EPI_ISL_2631379, EPI_ISL_2631380, EPI_ISL_2631381, EPI_ISL_2631382, EPI_ISL_2631383, EPI_ISL_2631384, EPI_ISL_2631385, EPI_ISL_2631386, EPI_ISL_2631387, EPI_ISL_2631388, EPI_ISL_2631389, EPI_ISL_2631390, EPI_ISL_2631391, EPI_ISL_2631392, EPI_ISL_2631393, EPI_ISL_2631394, EPI_ISL_2631395, EPI_ISL_2631396, EPI_ISL_2631397, EPI_ISL_2631398, EPI_ISL_2631399, EPI_ISL_2631400, EPI_ISL_2631401, EPI_ISL_2631402, EPI_ISL_2631403, EPI_ISL_2631404, EPI_ISL_2631405, EPI_ISL_2631406, EPI_ISL_2631407, EPI_ISL_2632876, EPI_ISL_2632880, EPI_ISL_2632884, EPI_ISL_2632887, EPI_ISL_2632890, EPI_ISL_2632893, EPI_ISL_2632897, EPI_ISL_2632900, EPI_ISL_2632904, EPI_ISL_2632907, EPI_ISL_2632911, EPI_ISL_2632915, EPI_ISL_2632918, EPI_ISL_2632922, EPI_ISL_2632925, EPI_ISL_2632929, EPI_ISL_2632933, EPI_ISL_2632936, EPI_ISL_2632940, EPI_ISL_2632944, EPI_ISL_2632949, EPI_ISL_2632953, EPI_ISL_2632957, EPI_ISL_2632961, EPI_ISL_2632965, EPI_ISL_2632969, EPI_ISL_2632973, EPI_ISL_2632976, EPI_ISL_2632979, EPI_ISL_2632983, EPI_ISL_2632986, EPI_ISL_2632990, EPI_ISL_2632994, EPI_ISL_2632998, EPI_ISL_2633001, EPI_ISL_2633005, EPI_ISL_2633009, EPI_ISL_2633013, EPI_ISL_2633017, EPI_ISL_2633021, EPI_ISL_2633025, EPI_ISL_2633028, EPI_ISL_2633031, EPI_ISL_2633035, EPI_ISL_2633039, EPI_ISL_2633043, EPI_ISL_2633048, EPI_ISL_2633051, EPI_ISL_2633056, EPI_ISL_2633060, EPI_ISL_2633064, EPI_ISL_2633067, EPI_ISL_2633072, EPI_ISL_2633076, EPI_ISL_2633079, EPI_ISL_2633081, EPI_ISL_2633085, EPI_ISL_2633089, EPI_ISL_2633093, EPI_ISL_2633097, EPI_ISL_2633101, EPI_ISL_2633105, EPI_ISL_2633107, EPI_ISL_2633109, EPI_ISL_2633112, EPI_ISL_2633116, EPI_ISL_2633119, EPI_ISL_2633122, EPI_ISL_2633125, EPI_ISL_2633129, EPI_ISL_2633133, EPI_ISL_2633135, EPI_ISL_2633136, EPI_ISL_2633137, EPI_ISL_2633138, EPI_ISL_2633140, EPI_ISL_2633143, EPI_ISL_2633147, EPI_ISL_2633151, EPI_ISL_2633154, EPI_ISL_2633158, EPI_ISL_2633162, EPI_ISL_2633165, EPI_ISL_2633169, EPI_ISL_2633173, EPI_ISL_2633176, EPI_ISL_2633179, EPI_ISL_2633183, EPI_ISL_2633186, EPI_ISL_2633189, EPI_ISL_2633193, EPI_ISL_2633196, EPI_ISL_2633200, EPI_ISL_2633204, EPI_ISL_2633207, EPI_ISL_2633210, EPI_ISL_2633214, EPI_ISL_2633217, EPI_ISL_2633221, EPI_ISL_2633224, EPI_ISL_2633227, EPI_ISL_2633230, EPI_ISL_2633234, EPI_ISL_2633237, EPI_ISL_2633240, EPI_ISL_2633244, EPI_ISL_2633248, EPI_ISL_2633251, EPI_ISL_2633254, EPI_ISL_2633256, EPI_ISL_2633258, EPI_ISL_2633260, EPI_ISL_2633262, EPI_ISL_2633263, EPI_ISL_2633265, EPI_ISL_2633267, EPI_ISL_2633269, EPI_ISL_2633271, EPI_ISL_2633274, EPI_ISL_2633278, EPI_ISL_2633280, EPI_ISL_2633282, EPI_ISL_2633284, EPI_ISL_2633285, EPI_ISL_2633287, EPI_ISL_2633289, EPI_ISL_2633291, EPI_ISL_2633293, EPI_ISL_2633295, EPI_ISL_2633297, EPI_ISL_2633300, EPI_ISL_2633305, EPI_ISL_2633309, EPI_ISL_2633315, EPI_ISL_2633320, EPI_ISL_2633326, EPI_ISL_2633332, EPI_ISL_2633338, EPI_ISL_2633344, EPI_ISL_2633347, EPI_ISL_2633351, EPI_ISL_2633354, EPI_ISL_2633358, EPI_ISL_2633361, EPI_ISL_2633365, EPI_ISL_2633368, EPI_ISL_2633370, EPI_ISL_2633372, EPI_ISL_2633374, EPI_ISL_2633378, EPI_ISL_2633382, EPI_ISL_2633385, EPI_ISL_2633389, EPI_ISL_2633393, EPI_ISL_2633396, EPI_ISL_2633400, EPI_ISL_2633402, EPI_ISL_2633404, EPI_ISL_2633405, EPI_ISL_2633407, EPI_ISL_2633411, EPI_ISL_2633414, EPI_ISL_2633418, EPI_ISL_2633422, EPI_ISL_2633426, EPI_ISL_2633430, EPI_ISL_2633434, EPI_ISL_2633438, EPI_ISL_2633442, EPI_ISL_2633447, EPI_ISL_2633448, EPI_ISL_2633449, EPI_ISL_2633450, EPI_ISL_2633451, EPI_ISL_2633454, EPI_ISL_2633458, EPI_ISL_2633459, EPI_ISL_2633461, EPI_ISL_2633463, EPI_ISL_2633467, EPI_ISL_2633471, EPI_ISL_2633472, EPI_ISL_2633474, EPI_ISL_2633476, EPI_ISL_2633480, EPI_ISL_2633484, EPI_ISL_2633486, EPI_ISL_2633488, EPI_ISL_2633490, EPI_ISL_2633494, EPI_ISL_2633497, EPI_ISL_2633501, EPI_ISL_2633505, EPI_ISL_2633508, EPI_ISL_2633513, EPI_ISL_2633517, EPI_ISL_2633522, EPI_ISL_2633527, EPI_ISL_2633530, EPI_ISL_2633534, EPI_ISL_2633538, EPI_ISL_2633541, EPI_ISL_2633544, EPI_ISL_2633546, EPI_ISL_2633548, EPI_ISL_2633550, EPI_ISL_2633552, EPI_ISL_2633553, EPI_ISL_2633555, EPI_ISL_2633556, EPI_ISL_2633560, EPI_ISL_2633564, EPI_ISL_2633567, EPI_ISL_2633571, EPI_ISL_2633575, EPI_ISL_2633579, EPI_ISL_2633582, EPI_ISL_2633587, EPI_ISL_2633589, EPI_ISL_2633593, EPI_ISL_2633597, EPI_ISL_2633600, EPI_ISL_2633604, EPI_ISL_2633609, EPI_ISL_2633611, EPI_ISL_2633615, EPI_ISL_2633619, EPI_ISL_2633622, EPI_ISL_2633626, EPI_ISL_2633628, EPI_ISL_2633629, EPI_ISL_2633631, EPI_ISL_2633633, EPI_ISL_2633634, EPI_ISL_2633636, EPI_ISL_2633638, EPI_ISL_2633640, EPI_ISL_2633642, EPI_ISL_2633644, EPI_ISL_2633645, EPI_ISL_2633647, EPI_ISL_2633649, EPI_ISL_2633651, EPI_ISL_2633653, EPI_ISL_2633654, EPI_ISL_2633656, EPI_ISL_2633657, EPI_ISL_2633659, EPI_ISL_2633663, EPI_ISL_2633666, EPI_ISL_2633669, EPI_ISL_2633671, EPI_ISL_2633673, EPI_ISL_2633675, EPI_ISL_2633677, EPI_ISL_2633678, EPI_ISL_2633680, EPI_ISL_2633682, EPI_ISL_2633684, EPI_ISL_2633685, EPI_ISL_2633689, EPI_ISL_2633691, EPI_ISL_2633694, EPI_ISL_2633697, EPI_ISL_2633700, EPI_ISL_2633704, EPI_ISL_2633707, EPI_ISL_2633710, EPI_ISL_2633714, EPI_ISL_2633717, EPI_ISL_2633721, EPI_ISL_2633724, EPI_ISL_2633727, EPI_ISL_2633729, EPI_ISL_2633730, EPI_ISL_2633732, EPI_ISL_2633736, EPI_ISL_2633740, EPI_ISL_2633743, EPI_ISL_2633745, EPI_ISL_2633746, EPI_ISL_2633748, EPI_ISL_2633750, EPI_ISL_2633752, EPI_ISL_2633753, EPI_ISL_2633755, EPI_ISL_2633757, EPI_ISL_2633759, EPI_ISL_2633761, EPI_ISL_2633763, EPI_ISL_2633764, EPI_ISL_2633766, EPI_ISL_2633768, EPI_ISL_2633770, EPI_ISL_2633774, EPI_ISL_2633778, EPI_ISL_2633781, EPI_ISL_2633783, EPI_ISL_2633785, EPI_ISL_2633788, EPI_ISL_2633792, EPI_ISL_2633796, EPI_ISL_2633799, EPI_ISL_2633803, EPI_ISL_2633807, EPI_ISL_2633810, EPI_ISL_2633814, EPI_ISL_2633820, EPI_ISL_2633822, EPI_ISL_2633826, EPI_ISL_2633830, EPI_ISL_2633833, EPI_ISL_2633837, EPI_ISL_2633840, EPI_ISL_2633842, EPI_ISL_2633844, EPI_ISL_2633848, EPI_ISL_2633851, EPI_ISL_2633855, EPI_ISL_2633858, EPI_ISL_2633861, EPI_ISL_2633865, EPI_ISL_2633869, EPI_ISL_2633872, EPI_ISL_2633874, EPI_ISL_2633875, EPI_ISL_2633879, EPI_ISL_2633883, EPI_ISL_2633887, EPI_ISL_2633890, EPI_ISL_2633894, EPI_ISL_2633897, EPI_ISL_2633902, EPI_ISL_2633905, EPI_ISL_2633907, EPI_ISL_2633909, EPI_ISL_2633911, EPI_ISL_2633912, EPI_ISL_2633917, EPI_ISL_2633919, EPI_ISL_2633921, EPI_ISL_2633922, EPI_ISL_2633924, EPI_ISL_2633928, EPI_ISL_2633931, EPI_ISL_2633935, EPI_ISL_2633937, EPI_ISL_2633939, EPI_ISL_2633940, EPI_ISL_2633942, EPI_ISL_2633944, EPI_ISL_2633946, EPI_ISL_2633948, EPI_ISL_2633949, EPI_ISL_2633950, EPI_ISL_2633952, EPI_ISL_2633954, EPI_ISL_2633957, EPI_ISL_2633960, EPI_ISL_2633963, EPI_ISL_2633967, EPI_ISL_2633970, EPI_ISL_2633974, EPI_ISL_2633976, EPI_ISL_2633978, EPI_ISL_2633979, EPI_ISL_2633981, EPI_ISL_2633983, EPI_ISL_2633985, EPI_ISL_2633987, EPI_ISL_2633989, EPI_ISL_2633990, EPI_ISL_2633992, EPI_ISL_2633994, EPI_ISL_2633996, EPI_ISL_2633997, EPI_ISL_2633999, EPI_ISL_2634003, EPI_ISL_2634006, EPI_ISL_2634010, EPI_ISL_2634014, EPI_ISL_2634017, EPI_ISL_2634021, EPI_ISL_2634024, EPI_ISL_2634028, EPI_ISL_2634032, EPI_ISL_2634036, EPI_ISL_2634040, EPI_ISL_2634043, EPI_ISL_2634047, EPI_ISL_2634050, EPI_ISL_2634054, EPI_ISL_2634057, EPI_ISL_2634061, EPI_ISL_2634065, EPI_ISL_2634068, EPI_ISL_2634071, EPI_ISL_2634074, EPI_ISL_2634076, EPI_ISL_2634080, EPI_ISL_2634084, EPI_ISL_2634088, EPI_ISL_2634090, EPI_ISL_2634091, EPI_ISL_2634093, EPI_ISL_2634095, EPI_ISL_2634099, EPI_ISL_2634103, EPI_ISL_2634106, EPI_ISL_2634109, EPI_ISL_2634112, EPI_ISL_2634116, EPI_ISL_2634118, EPI_ISL_2634120, EPI_ISL_2634122, EPI_ISL_2634123, EPI_ISL_2634125, EPI_ISL_2634127, EPI_ISL_2634129, EPI_ISL_2634130, EPI_ISL_2634132, EPI_ISL_2634134, EPI_ISL_2634136, EPI_ISL_2634137, EPI_ISL_2634139, EPI_ISL_2634141, EPI_ISL_2634143, EPI_ISL_2634145, EPI_ISL_2634147, EPI_ISL_2634148, EPI_ISL_2634150, EPI_ISL_2634152, EPI_ISL_2634154, EPI_ISL_2634156, EPI_ISL_2634158, EPI_ISL_2634160, EPI_ISL_2634163, EPI_ISL_2634165, EPI_ISL_2634166, EPI_ISL_2634168, EPI_ISL_2634170, EPI_ISL_2634171, EPI_ISL_2634173, EPI_ISL_2634175, EPI_ISL_2634176, EPI_ISL_2634178, EPI_ISL_2634180, EPI_ISL_2634181, EPI_ISL_2634183, EPI_ISL_2634185, EPI_ISL_2634187, EPI_ISL_2634189, EPI_ISL_2634191, EPI_ISL_2634193, EPI_ISL_2634194, EPI_ISL_2634196, EPI_ISL_2634198, EPI_ISL_2634201, EPI_ISL_2634203, EPI_ISL_2634205, EPI_ISL_2634206, EPI_ISL_2634208, EPI_ISL_2634210, EPI_ISL_2634211, EPI_ISL_2634213, EPI_ISL_2634214, EPI_ISL_2634216, EPI_ISL_2634218, EPI_ISL_2634219, EPI_ISL_2634221, EPI_ISL_2634223, EPI_ISL_2634224, EPI_ISL_2634226, EPI_ISL_2634228, EPI_ISL_2634229, EPI_ISL_2634231, EPI_ISL_2634233, EPI_ISL_2634235, EPI_ISL_2634236, EPI_ISL_2634238, EPI_ISL_2634240, EPI_ISL_2634242, EPI_ISL_2634243, EPI_ISL_2634245, EPI_ISL_2634247, EPI_ISL_2634248, EPI_ISL_2634250, EPI_ISL_2634251, EPI_ISL_2634252, EPI_ISL_2634254, EPI_ISL_2634256, EPI_ISL_2634258, EPI_ISL_2634260, EPI_ISL_2634261, EPI_ISL_2634263, EPI_ISL_2634265, EPI_ISL_2634267, EPI_ISL_2634268, EPI_ISL_2634270, EPI_ISL_2634272, EPI_ISL_2634273, EPI_ISL_2634275, EPI_ISL_2634276, EPI_ISL_2634277, EPI_ISL_2634279, EPI_ISL_2634280, EPI_ISL_2634282, EPI_ISL_2634284, EPI_ISL_2634286, EPI_ISL_2634288, EPI_ISL_2634289, EPI_ISL_2634291, EPI_ISL_2634293, EPI_ISL_2634295, EPI_ISL_2634296, EPI_ISL_2634298, EPI_ISL_2634300, EPI_ISL_2634302, EPI_ISL_2634304, EPI_ISL_2634305, EPI_ISL_2634307, EPI_ISL_2634309, EPI_ISL_2634311, EPI_ISL_2634313, EPI_ISL_2634314, EPI_ISL_2634316, EPI_ISL_2634318, EPI_ISL_2634320, EPI_ISL_2634322, EPI_ISL_2634324, EPI_ISL_2634325, EPI_ISL_2634327, EPI_ISL_2634329, EPI_ISL_2634331, EPI_ISL_2634333, EPI_ISL_2634334, EPI_ISL_2634336, EPI_ISL_2634338, EPI_ISL_2634340, EPI_ISL_2634342, EPI_ISL_2634344, EPI_ISL_2634346, EPI_ISL_2634347, EPI_ISL_2634349, EPI_ISL_2634351, EPI_ISL_2634353, EPI_ISL_2634355, EPI_ISL_2634356, EPI_ISL_2634358, EPI_ISL_2634360, EPI_ISL_2634362, EPI_ISL_2634364, EPI_ISL_2634365, EPI_ISL_2634367, EPI_ISL_2634369, EPI_ISL_2634370, EPI_ISL_2634372, EPI_ISL_2634374, EPI_ISL_2634375, EPI_ISL_2634377, EPI_ISL_2634379, EPI_ISL_2634381, EPI_ISL_2634382, EPI_ISL_2634384, EPI_ISL_2634386, EPI_ISL_2634388, EPI_ISL_2634389, EPI_ISL_2634391, EPI_ISL_2634393, EPI_ISL_2634394, EPI_ISL_2634396, EPI_ISL_2634398, EPI_ISL_2634400, EPI_ISL_2634402, EPI_ISL_2634403, EPI_ISL_2634405, EPI_ISL_2634407, EPI_ISL_2634412, EPI_ISL_2634416, EPI_ISL_2634420, EPI_ISL_2634422, EPI_ISL_2634423, EPI_ISL_2634425, EPI_ISL_2634427, EPI_ISL_2634429, EPI_ISL_2634431, EPI_ISL_2634433, EPI_ISL_2634434, EPI_ISL_2634436, EPI_ISL_2634437, EPI_ISL_2634438, EPI_ISL_2634440, EPI_ISL_2634442, EPI_ISL_2634444, EPI_ISL_2634445, EPI_ISL_2634447, EPI_ISL_2634449, EPI_ISL_2634451, EPI_ISL_2634453, EPI_ISL_2634454, EPI_ISL_2634456, EPI_ISL_2634458, EPI_ISL_2634460, EPI_ISL_2634462, EPI_ISL_2634464, EPI_ISL_2634465, EPI_ISL_2634467, EPI_ISL_2634469, EPI_ISL_2634471, EPI_ISL_2634473, EPI_ISL_2634475, EPI_ISL_2634476, EPI_ISL_2634478, EPI_ISL_2634480, EPI_ISL_2634482, EPI_ISL_2634484, EPI_ISL_2634485, EPI_ISL_2634487, EPI_ISL_2634489, EPI_ISL_2634491, EPI_ISL_2634493, EPI_ISL_2634494, EPI_ISL_2634496, EPI_ISL_2634498, EPI_ISL_2634502, EPI_ISL_2634505, EPI_ISL_2634507, EPI_ISL_2634509, EPI_ISL_2634511, EPI_ISL_2634512, EPI_ISL_2634514, EPI_ISL_2634516, EPI_ISL_2634517, EPI_ISL_2634519, EPI_ISL_2634521, EPI_ISL_2634523, EPI_ISL_2634525, EPI_ISL_2634527, EPI_ISL_2634528, EPI_ISL_2634530, EPI_ISL_2634532, EPI_ISL_2634533, EPI_ISL_2634535, EPI_ISL_2634537, EPI_ISL_2634539, EPI_ISL_2634541, EPI_ISL_2634542, EPI_ISL_2634544, EPI_ISL_2634546, EPI_ISL_2634547, EPI_ISL_2634548, EPI_ISL_2634549, EPI_ISL_2634550, EPI_ISL_2634552, EPI_ISL_2634554, EPI_ISL_2634555, EPI_ISL_2634557, EPI_ISL_2634559, EPI_ISL_2634560, EPI_ISL_2634562, EPI_ISL_2634564, EPI_ISL_2634566, EPI_ISL_2634568, EPI_ISL_2634570, EPI_ISL_2634571, EPI_ISL_2634573, EPI_ISL_2634575, EPI_ISL_2634576, EPI_ISL_2634578, EPI_ISL_2634580, EPI_ISL_2634582, EPI_ISL_2634584, EPI_ISL_2634586, EPI_ISL_2634587, EPI_ISL_2634589, EPI_ISL_2634591, EPI_ISL_2634593, EPI_ISL_2634595, EPI_ISL_2634597, EPI_ISL_2634598, EPI_ISL_2634600, EPI_ISL_2634602, EPI_ISL_2634604, EPI_ISL_2634606, EPI_ISL_2634607, EPI_ISL_2634609, EPI_ISL_2634611, EPI_ISL_2634613, EPI_ISL_2634615, EPI_ISL_2634616, EPI_ISL_2634618, EPI_ISL_2634620, EPI_ISL_2634622, EPI_ISL_2634624, EPI_ISL_2634626, | | | |

| | | | |
|---|---|---|---|
| EPI_ISL_2634627, EPI_ISL_2634629, EPI_ISL_2634631, EPI_ISL_2634633, EPI_ISL_2634635, EPI_ISL_2634636, EPI_ISL_2634638, EPI_ISL_2634640, EPI_ISL_2634642, EPI_ISL_2634644, EPI_ISL_2634646, EPI_ISL_2634647, EPI_ISL_2634649, EPI_ISL_2634651, EPI_ISL_2634653, EPI_ISL_2634655, EPI_ISL_2634657, EPI_ISL_2634659, EPI_ISL_2634660, EPI_ISL_2634662, EPI_ISL_2634663, EPI_ISL_2634665, EPI_ISL_2634666, EPI_ISL_2634668, EPI_ISL_2634670, EPI_ISL_2634672, EPI_ISL_2634673, EPI_ISL_2634675, EPI_ISL_2634677, EPI_ISL_2634679, EPI_ISL_2634681, EPI_ISL_2634682, EPI_ISL_2634684, EPI_ISL_2634686, EPI_ISL_2634688, EPI_ISL_2634691, EPI_ISL_2634692, EPI_ISL_2634694, EPI_ISL_2634696, EPI_ISL_2634698, EPI_ISL_2634699, EPI_ISL_2634701, EPI_ISL_2634703, EPI_ISL_2634705, EPI_ISL_2634706, EPI_ISL_2634708, EPI_ISL_2634710, EPI_ISL_2634712, EPI_ISL_2634714, EPI_ISL_2634716, EPI_ISL_2634718, EPI_ISL_2634720, EPI_ISL_2634721, EPI_ISL_2634724, EPI_ISL_2634725, EPI_ISL_2634727, EPI_ISL_2634729, EPI_ISL_2634731, EPI_ISL_2634733, EPI_ISL_2634735, EPI_ISL_2634737, EPI_ISL_2634739, EPI_ISL_2634741, EPI_ISL_2634743, EPI_ISL_2634745, EPI_ISL_2634746, EPI_ISL_2634748, EPI_ISL_2634749, EPI_ISL_2634751, EPI_ISL_2634752, EPI_ISL_2634753, EPI_ISL_2634754, EPI_ISL_2634755, EPI_ISL_2634757, EPI_ISL_2634759, EPI_ISL_2634761, EPI_ISL_2634763, EPI_ISL_2634764, EPI_ISL_2634766, EPI_ISL_2634767, EPI_ISL_2634768, EPI_ISL_2634769, EPI_ISL_2634771, EPI_ISL_2634773, EPI_ISL_2634774, EPI_ISL_2634776, EPI_ISL_2634778, EPI_ISL_2634780, EPI_ISL_2634782, EPI_ISL_2634783, EPI_ISL_2634785, EPI_ISL_2634786, EPI_ISL_2634787, EPI_ISL_2634788, EPI_ISL_2634790, EPI_ISL_2634792, EPI_ISL_2634794, EPI_ISL_2634796, EPI_ISL_2634798, EPI_ISL_2634800, EPI_ISL_2634801, EPI_ISL_2634803, EPI_ISL_2634805, EPI_ISL_2634807, EPI_ISL_2634808, EPI_ISL_2634810, EPI_ISL_2634812, EPI_ISL_2634814, EPI_ISL_2634816, EPI_ISL_2634817, EPI_ISL_2634820, EPI_ISL_2634822, EPI_ISL_2634824, EPI_ISL_2634825, EPI_ISL_2634827, EPI_ISL_2634829, EPI_ISL_2634831, EPI_ISL_2634832, EPI_ISL_2634834, EPI_ISL_2634836, EPI_ISL_2634837, EPI_ISL_2634839, EPI_ISL_2634841, EPI_ISL_2634843, EPI_ISL_2634845, EPI_ISL_2634847, EPI_ISL_2634849, EPI_ISL_2634850, EPI_ISL_2634852, EPI_ISL_2634854, EPI_ISL_2634856, EPI_ISL_2634857, EPI_ISL_2634859, EPI_ISL_2634861, EPI_ISL_2634863, EPI_ISL_2634865, EPI_ISL_2634867, EPI_ISL_2634868, EPI_ISL_2634870, EPI_ISL_2634872, EPI_ISL_2634874, EPI_ISL_2634875, EPI_ISL_2634877, EPI_ISL_2634879, EPI_ISL_2634881, EPI_ISL_2634883, EPI_ISL_2634885, EPI_ISL_2634887, EPI_ISL_2634889, EPI_ISL_2634890, EPI_ISL_2634892, EPI_ISL_2634894, EPI_ISL_2634896, EPI_ISL_2634897, EPI_ISL_2634899, EPI_ISL_2634900, EPI_ISL_2634901, EPI_ISL_2634902, EPI_ISL_2634903, EPI_ISL_2634904, EPI_ISL_2634906, EPI_ISL_2634907, EPI_ISL_2634909, EPI_ISL_2634911, EPI_ISL_2634913, EPI_ISL_2634915, EPI_ISL_2634916, EPI_ISL_2634918, EPI_ISL_2634920, EPI_ISL_2634922, EPI_ISL_2634923, EPI_ISL_2634925, EPI_ISL_2634927, EPI_ISL_2634929, EPI_ISL_2634931, EPI_ISL_2634933, EPI_ISL_2634934, EPI_ISL_2634936, EPI_ISL_2634938, EPI_ISL_2634940, EPI_ISL_2634942, EPI_ISL_2634943, EPI_ISL_2634945, EPI_ISL_2634947, EPI_ISL_2634949, EPI_ISL_2634951, EPI_ISL_2634952, EPI_ISL_2634954, EPI_ISL_2634956, EPI_ISL_2634958, EPI_ISL_2634959, EPI_ISL_2634961, EPI_ISL_2634963, EPI_ISL_2634965, EPI_ISL_2634967, EPI_ISL_2634969, EPI_ISL_2634971, EPI_ISL_2634973, EPI_ISL_2634974 | | | |
| see above | Pandemic Response Lab - NYC | Pandemic Response Lab, R&D | Henry Lee, Michael Hammerling, Melissa Hopkins, Cybill del Castillo, Shinyoung Clair Kang, William Ward, Pradeep Bugga, Sol Rey, Dylan Law, Katharine Nelson, Haiping Hao, Jon Laurent |
| EPI_ISL_2646943 | Mako Medical | Centers for Disease Control and Prevention Division of Viral Diseases, Pathogen Discovery | Dakota Howard, Dhwani Batra, Peter W. Cook, Kara Moser, Adrian Paskey, Jason Caravas, Benjamin Rambo-Martin, Shatavia Morrison, Christopher Gulvick, Scott Sammons, Yvette Unoarumhi, Darlene Wagner, Matthew Schmerer,Matthew Tugwell, Lauren Moon, Clinton R. Paden, Duncan MacCannell |
| EPI_ISL_2649967 | Quest Diagnostics Incorporated | Centers for Disease Control and Prevention Division of Viral Diseases, Pathogen Discovery | Dakota Howard, Dhwani Batra, Peter W. Cook, Kara Moser, Adrian Paskey, Jason Caravas, Benjamin Rambo-Martin, Shatavia Morrison, Christopher Gulvick, Scott Sammons, Yvette Unoarumhi, Darlene Wagner, Matthew Schmerer, S. H. Rosenthal, A. Gerasimova, R. M. Kagan, B. Anderson, M. Hua, Y. Liu, L.E. Bernstein, K.E. Livingston, A. Perez, I. A. Shlyakhter, R. V. Rolando, R. Owen, P. Tanpaiboon, F. Lacbawan, Clinton R. Paden, Duncan MacCannell |
| EPI_ISL_2652711, EPI_ISL_2652793, EPI_ISL_2652852, EPI_ISL_2652967, EPI_ISL_2652969 | Infinity Biologix | Centers for Disease Control and Prevention Division of Viral Diseases, Pathogen Discovery | Dakota Howard, Dhwani Batra, Peter W. Cook, Kara Moser, Adrian Paskey, Jason Caravas, Benjamin Rambo-Martin, Shatavia Morrison, Christopher Gulvick, Scott Sammons, Yvette Unoarumhi, Darlene Wagner, Matthew Schmerer, Christian Bixby, Yihe Wang, Jonathan Schultz, Chirayu Goswami, Russ Hager, Robin Grimwood, Clinton R. Paden, Duncan MacCannell |
| EPI_ISL_2662828, EPI_ISL_2662829, EPI_ISL_2662830, EPI_ISL_2662831 | Erie County Medical Center (ECMC) | University at Buffalo Genomics and Bioinformatics Core | Jonathan Bard, Natalie Lamb, Alyssa Pohlman, Brandon Marzullo, Amanda Boccolucci, Norma Nowak, Donald Yergeau, Jennifer Surtees |
| EPI_ISL_2662832, EPI_ISL_2662833, EPI_ISL_2662834, EPI_ISL_2662835, EPI_ISL_2662836, EPI_ISL_2662837, EPI_ISL_2662838, EPI_ISL_2662839, EPI_ISL_2662840, EPI_ISL_2662841, EPI_ISL_2662842, EPI_ISL_2662843, EPI_ISL_2662844, EPI_ISL_2662845, EPI_ISL_2662846, EPI_ISL_2662847, EPI_ISL_2662848, EPI_ISL_2662849, EPI_ISL_2662850, EPI_ISL_2662851, EPI_ISL_2662852, EPI_ISL_2662853, EPI_ISL_2662854, EPI_ISL_2662855, EPI_ISL_2662856, EPI_ISL_2662857, EPI_ISL_2662858, EPI_ISL_2662859, EPI_ISL_2662860, EPI_ISL_2662861, EPI_ISL_2662862, EPI_ISL_2662863, EPI_ISL_2662864, EPI_ISL_2662865, EPI_ISL_2662866, EPI_ISL_2662867, EPI_ISL_2662868, EPI_ISL_2662869, EPI_ISL_2662870, EPI_ISL_2662871, EPI_ISL_2662872, EPI_ISL_2662873, EPI_ISL_2662874, EPI_ISL_2662875, EPI_ISL_2662876, EPI_ISL_2662877, EPI_ISL_2662878, EPI_ISL_2662879, EPI_ISL_2662880, EPI_ISL_2662881, EPI_ISL_2662882, EPI_ISL_2662883, EPI_ISL_2662884, EPI_ISL_2662885, EPI_ISL_2662886, EPI_ISL_2662887, EPI_ISL_2662888, EPI_ISL_2662889, EPI_ISL_2662890, EPI_ISL_2662891, EPI_ISL_2662892, EPI_ISL_2662893, EPI_ISL_2662894, EPI_ISL_2662895, EPI_ISL_2662896, EPI_ISL_2662897, EPI_ISL_2662898, EPI_ISL_2662899, EPI_ISL_2662900, EPI_ISL_2662901, EPI_ISL_2662902, EPI_ISL_2662903, EPI_ISL_2662904, EPI_ISL_2662905, EPI_ISL_2662906, EPI_ISL_2662907, EPI_ISL_2662908, EPI_ISL_2662909, EPI_ISL_2662910, EPI_ISL_2662911, EPI_ISL_2662912, EPI_ISL_2662913, EPI_ISL_2662914, EPI_ISL_2662915, EPI_ISL_2662916, EPI_ISL_2662917, EPI_ISL_2662918, EPI_ISL_2662919, EPI_ISL_2662920, EPI_ISL_2662921, EPI_ISL_2662922, EPI_ISL_2662923, EPI_ISL_2662924 | | | |
| see above | Erie County Public Health (ECPHL) | University at Buffalo Genomics and Bioinformatics Core | Jonathan Bard, Natalie Lamb, Alyssa Pohlman, Brandon Marzullo, Amanda Boccolucci, Norma Nowak, Donald Yergeau, Jennifer Surtees |
| EPI_ISL_2662925, EPI_ISL_2662926, EPI_ISL_2662927, EPI_ISL_2662928, EPI_ISL_2662929, EPI_ISL_2662930, EPI_ISL_2662931, EPI_ISL_2662932, EPI_ISL_2662933, EPI_ISL_2662934, EPI_ISL_2662935, EPI_ISL_2662936, EPI_ISL_2662937, EPI_ISL_2662938, EPI_ISL_2662939, EPI_ISL_2662940, EPI_ISL_2662941, EPI_ISL_2662942, EPI_ISL_2662943, EPI_ISL_2662944, EPI_ISL_2662945, EPI_ISL_2662946, EPI_ISL_2662947, EPI_ISL_2662948, EPI_ISL_2662949, EPI_ISL_2662950, EPI_ISL_2662951, EPI_ISL_2662952, EPI_ISL_2662953, EPI_ISL_2662954, EPI_ISL_2662955, EPI_ISL_2662956, EPI_ISL_2662957, EPI_ISL_2662958, EPI_ISL_2662959, EPI_ISL_2662960, EPI_ISL_2662961, EPI_ISL_2662962, EPI_ISL_2662963, EPI_ISL_2662964, EPI_ISL_2662965, EPI_ISL_2662966, EPI_ISL_2662967, EPI_ISL_2662968, EPI_ISL_2662969, EPI_ISL_2662970, EPI_ISL_2662971, EPI_ISL_2662972, EPI_ISL_2662973, EPI_ISL_2662974, EPI_ISL_2662975, EPI_ISL_2662976, EPI_ISL_2662977, EPI_ISL_2662978, EPI_ISL_2662979, EPI_ISL_2662980, EPI_ISL_2662981, EPI_ISL_2662982, EPI_ISL_2662983, EPI_ISL_2662984, EPI_ISL_2662985, EPI_ISL_2662986, EPI_ISL_2662987, EPI_ISL_2662988, EPI_ISL_2662989, EPI_ISL_2662990, EPI_ISL_2662991, EPI_ISL_2662992, EPI_ISL_2662993, EPI_ISL_2662994 | | | |
| see above | Kaleida Health Laboratories | University at Buffalo Genomics and Bioinformatics Core | Jonathan Bard, Natalie Lamb, Alyssa Pohlman, Brandon Marzullo, Amanda Boccolucci, Norma Nowak, Donald Yergeau, Jennifer Surtees |
| EPI_ISL_2663512, EPI_ISL_2663513, EPI_ISL_2663514, EPI_ISL_2663515, EPI_ISL_2663516, EPI_ISL_2663517, EPI_ISL_2663518, EPI_ISL_2663519, EPI_ISL_2663520, EPI_ISL_2663522, EPI_ISL_2663523, EPI_ISL_2663524, EPI_ISL_2663525, EPI_ISL_2663526, EPI_ISL_2663527, EPI_ISL_2663529, EPI_ISL_2663530, EPI_ISL_2663531, EPI_ISL_2663532, EPI_ISL_2663533, EPI_ISL_2663534, EPI_ISL_2663535, EPI_ISL_2663537, EPI_ISL_2663538, EPI_ISL_2663539, EPI_ISL_2663540, EPI_ISL_2663541, EPI_ISL_2663542, EPI_ISL_2663543, EPI_ISL_2663544, EPI_ISL_2663545, EPI_ISL_2663546, EPI_ISL_2663547, EPI_ISL_2663548, EPI_ISL_2663549, EPI_ISL_2663550, EPI_ISL_2663551, EPI_ISL_2663552 | | | |
| see above | NYU Langone Health | Departments of Pathology and Medicine, New York University School of Medicine | Adriana Heguy, Dacia Dimartino, Emily Guzman, Christian Marier, Peter Meyn, Sitharam Ramaswami, Gael Westby, Paul Zappile, Yutong Zhang, Paolo Cotzia, Guiqing Wang |
| EPI_ISL_2686446, EPI_ISL_2686447, EPI_ISL_2686448, EPI_ISL_2686449, EPI_ISL_2686450, EPI_ISL_2686452, EPI_ISL_2686474, EPI_ISL_2686476, EPI_ISL_2686477, EPI_ISL_2686479, EPI_ISL_2686482, EPI_ISL_2686485, EPI_ISL_2686486, EPI_ISL_2686489, EPI_ISL_2686498, EPI_ISL_2686499, EPI_ISL_2686502, EPI_ISL_2686503, EPI_ISL_2686504, EPI_ISL_2686505, EPI_ISL_2686506, EPI_ISL_2686507, EPI_ISL_2686508, EPI_ISL_2686509, EPI_ISL_2686696, EPI_ISL_2686699, EPI_ISL_2686700, EPI_ISL_2686701, EPI_ISL_2686702, EPI_ISL_2686703, EPI_ISL_2686705, EPI_ISL_2686707, EPI_ISL_2686749, EPI_ISL_2686751, EPI_ISL_2686752, EPI_ISL_2686753, EPI_ISL_2686755, EPI_ISL_2686756, EPI_ISL_2686759, EPI_ISL_2686760, EPI_ISL_2686761 | | | |
| see above | Fulgent Genetics | Centers for Disease Control and Prevention Division of Viral Diseases, Pathogen Discovery | Dakota Howard, Dhwani Batra, Peter W. Cook, Kara Moser, Adrian Paskey, Jason Caravas, Benjamin Rambo-Martin, Shatavia Morrison, Christopher Gulvick, Scott Sammons, Yvette Unoarumhi, Darlene Wagner, Matthew Schmerer, Harry Gao, Mickey Li, John Gao, Joseph Fierro, Benafsh Sapra, Becky Tsai, Yan Meng, Doreen Ng, James Xie, Clinton R. Paden, Duncan MacCannell |
| EPI_ISL_2686896, EPI_ISL_2686897 | Infinity Biologix | Centers for Disease Control and Prevention Division of Viral Diseases, Pathogen Discovery | Dakota Howard, Dhwani Batra, Peter W. Cook, Kara Moser, Adrian Paskey, Jason Caravas, Benjamin Rambo-Martin, Shatavia Morrison, Christopher Gulvick, Scott Sammons, Yvette Unoarumhi, Darlene Wagner, Matthew Schmerer, Christian Bixby, Yihe Wang, Jonathan Schultz, Chirayu Goswami, Russ Hager, Robin Grimwood, Clinton R. Paden, Duncan MacCannell |
| EPI_ISL_2687051, EPI_ISL_2687092, EPI_ISL_2687121, EPI_ISL_2687160, EPI_ISL_2687186, EPI_ISL_2687260, EPI_ISL_2687293, EPI_ISL_2687329, EPI_ISL_2687572 | Aegis Sciences Corporation | Centers for Disease Control and Prevention Division of Viral Diseases, Pathogen Discovery | Dakota Howard, Dhwani Batra, Peter W. Cook, Kara Moser, Adrian Paskey, Jason Caravas, Benjamin Rambo-Martin, Shatavia Morrison, Christopher Gulvick, Scott Sammons, Yvette Unoarumhi, Darlene Wagner, Matthew Schmerer, Cyndi Clark, Patrick Campbell, Rob Case, Vikramsinha Ghorpade, Holly Houdeshell, Ola Kvalvaag, Dillon Nall, Ethan Sanders, Alec Vest, Shaun Westlund, Matthew Hardison, Clinton R. Paden, Duncan MacCannell |
| EPI_ISL_2689144, EPI_ISL_2689145, EPI_ISL_2689146, EPI_ISL_2689147, EPI_ISL_2689148, EPI_ISL_2689149 | MSK Microbiology Lab | MSK Microbiology Lab | Esther Babady, Tracy McMillen, Krupa Jani |
| EPI_ISL_2690057, EPI_ISL_2690058 | NYC Department of Health and Mental Hygiene | Centers for Disease Control and Prevention Division of Viral Diseases, Pathogen Discovery | Mili Sheth, Sarah Nobles, Jasmine Padilla, Mark Burroughs, Shoshona Le, Katie Dillon, Peter Cook, Clinton R. Paden, Dhwani Batra, Krista Queen, Kristen Knipe, Dakota Howard, Yvette Unoarumhi, Darlene Wagner, Matthew Schmerer, Ben L. Rambo-Martin, Kristine Lacek, Sam Shepard, Alison Laufer Halpin, Dave Wentworth, Vivien Dugan, Suxiang Tong, Justin Lee |
| EPI_ISL_2690297, EPI_ISL_2690298, EPI_ISL_2690299, EPI_ISL_2690300, EPI_ISL_2690301, EPI_ISL_2690302, EPI_ISL_2690303, EPI_ISL_2690304, EPI_ISL_2690305, EPI_ISL_2690306, EPI_ISL_2690307, EPI_ISL_2690308, EPI_ISL_2690309, EPI_ISL_2690310, EPI_ISL_2690311, EPI_ISL_2690312, EPI_ISL_2690313, EPI_ISL_2690314, EPI_ISL_2690315, EPI_ISL_2690316, EPI_ISL_2690317, EPI_ISL_2690318, EPI_ISL_2690319, EPI_ISL_2690320, EPI_ISL_2690321, EPI_ISL_2690322, EPI_ISL_2690323, EPI_ISL_2690324, EPI_ISL_2690325, EPI_ISL_2690326, EPI_ISL_2690327, EPI_ISL_2690328, EPI_ISL_2690329, EPI_ISL_2690330, EPI_ISL_2690401, EPI_ISL_2690402, EPI_ISL_2690403, EPI_ISL_2690404, EPI_ISL_2690405, EPI_ISL_2690406, EPI_ISL_2690407, EPI_ISL_2690408, EPI_ISL_2690409, EPI_ISL_2690410, EPI_ISL_2690411, EPI_ISL_2690412, EPI_ISL_2690413, EPI_ISL_2690414, EPI_ISL_2690415, EPI_ISL_2690416, EPI_ISL_2690417, EPI_ISL_2690418, EPI_ISL_2690419, EPI_ISL_2690420, EPI_ISL_2690421, EPI_ISL_2690422, EPI_ISL_2690423, EPI_ISL_2690424, EPI_ISL_2690425, EPI_ISL_2690426, EPI_ISL_2690427, EPI_ISL_2690428, EPI_ISL_2690429, EPI_ISL_2690430, EPI_ISL_2690431, EPI_ISL_2690432, EPI_ISL_2690433, EPI_ISL_2690434, EPI_ISL_2690435, EPI_ISL_2690436, EPI_ISL_2690437, EPI_ISL_2690438, EPI_ISL_2690439, EPI_ISL_2690440, EPI_ISL_2690441, EPI_ISL_2690443, EPI_ISL_2690444, EPI_ISL_2690445, EPI_ISL_2690568, EPI_ISL_2690570, EPI_ISL_2690572, EPI_ISL_2690573, EPI_ISL_2690575, EPI_ISL_2690576, EPI_ISL_2690578, EPI_ISL_2690579, EPI_ISL_2690580, EPI_ISL_2690582, EPI_ISL_2690583, EPI_ISL_2690585, EPI_ISL_2690586, EPI_ISL_2690588, EPI_ISL_2690589, EPI_ISL_2690590, EPI_ISL_2690592, EPI_ISL_2690593, EPI_ISL_2690595, EPI_ISL_2690596, EPI_ISL_2690598, EPI_ISL_2690599, EPI_ISL_2690601, EPI_ISL_2690602, EPI_ISL_2690604, EPI_ISL_2690605, EPI_ISL_2690607, EPI_ISL_2690608, EPI_ISL_2690609, EPI_ISL_2690611, EPI_ISL_2690612, EPI_ISL_2690614, EPI_ISL_2690615, EPI_ISL_2690617, EPI_ISL_2690618, EPI_ISL_2690620, EPI_ISL_2690621, EPI_ISL_2690622, EPI_ISL_2690624, EPI_ISL_2690625, EPI_ISL_2690627, EPI_ISL_2690628, EPI_ISL_2690629, EPI_ISL_2690631, EPI_ISL_2690632, EPI_ISL_2690634, EPI_ISL_2690635, EPI_ISL_2690637, EPI_ISL_2690638, EPI_ISL_2690639, EPI_ISL_2690641, EPI_ISL_2690642, EPI_ISL_2690644, EPI_ISL_2690645, EPI_ISL_2690647, EPI_ISL_2690648, EPI_ISL_2690650, EPI_ISL_2690651, EPI_ISL_2690652, EPI_ISL_2690654, EPI_ISL_2690655, EPI_ISL_2690657, EPI_ISL_2690658, EPI_ISL_2690659, EPI_ISL_2690661, EPI_ISL_2690662, EPI_ISL_2690664, EPI_ISL_2690665, EPI_ISL_2690667, EPI_ISL_2690668, EPI_ISL_2690670, EPI_ISL_2690671, EPI_ISL_2690673, EPI_ISL_2690674, EPI_ISL_2690675, EPI_ISL_2690677, EPI_ISL_2690678, EPI_ISL_2690680, EPI_ISL_2690681, EPI_ISL_2690683, EPI_ISL_2690684, EPI_ISL_2690685, EPI_ISL_2690687, EPI_ISL_2690688, EPI_ISL_2690690, EPI_ISL_2690691, EPI_ISL_2690693, EPI_ISL_2690694, EPI_ISL_2690695, EPI_ISL_2690697, EPI_ISL_2690698, EPI_ISL_2690996, EPI_ISL_2690997, EPI_ISL_2690998, EPI_ISL_2690999, EPI_ISL_2691000, EPI_ISL_2691001, EPI_ISL_2691002, EPI_ISL_2691003, EPI_ISL_2691004, EPI_ISL_2691005, EPI_ISL_2691006, EPI_ISL_2691007, EPI_ISL_2691008, EPI_ISL_2691009, EPI_ISL_2691010, EPI_ISL_2691011, EPI_ISL_2691012, EPI_ISL_2691013, EPI_ISL_2691014, EPI_ISL_2691015, EPI_ISL_2691016, EPI_ISL_2691017, EPI_ISL_2691018, EPI_ISL_2691019, EPI_ISL_2691020, EPI_ISL_2691021, EPI_ISL_2691022, EPI_ISL_2691023, EPI_ISL_2691024, EPI_ISL_2691025, EPI_ISL_2691026, EPI_ISL_2691027, EPI_ISL_2691028, EPI_ISL_2691029, EPI_ISL_2691030, EPI_ISL_2691031, EPI_ISL_2691032, EPI_ISL_2691033, EPI_ISL_2691034, EPI_ISL_2691035, EPI_ISL_2691036, EPI_ISL_2691037, EPI_ISL_2691038, EPI_ISL_2691039, EPI_ISL_2691040, EPI_ISL_2691041, EPI_ISL_2691042, EPI_ISL_2691043, EPI_ISL_2691044, EPI_ISL_2691045, EPI_ISL_2691046, | | | |

EPI_ISL_2691047, EPI_ISL_2691048, EPI_ISL_2691049, EPI_ISL_2691050, EPI_ISL_2691051, EPI_ISL_2691052, EPI_ISL_2691053, EPI_ISL_2691054, EPI_ISL_2691055, EPI_ISL_2691056, EPI_ISL_2691057, EPI_ISL_2691058, EPI_ISL_2691059, EPI_ISL_2691060, EPI_ISL_2691061, EPI_ISL_2691062, EPI_ISL_2691063, EPI_ISL_2691064, EPI_ISL_2691065, EPI_ISL_2691066, EPI_ISL_2691067, EPI_ISL_2691068, EPI_ISL_2691069, EPI_ISL_2691070, EPI_ISL_2691071, EPI_ISL_2691072, EPI_ISL_2691073, EPI_ISL_2691074, EPI_ISL_2691075, EPI_ISL_2691076, EPI_ISL_2691077, EPI_ISL_2691078, EPI_ISL_2691079, EPI_ISL_2691080, EPI_ISL_2691081, EPI_ISL_2691082, EPI_ISL_2691083, EPI_ISL_2691084, EPI_ISL_2691085, EPI_ISL_2691086, EPI_ISL_2691087, EPI_ISL_2691088

| | | | |
|---|---|---|---|
| see above | MSK Microbiology Lab | MSK Microbiology Lab | Esther Babady, Tracy McMillen, Krupa Jani |
| EPI_ISL_2696473 | Laboratory, Gravity Diagnostics | Laboratory, Gravity Diagnostics | Diagnostics,G. |

EPI_ISL_2715372, EPI_ISL_2715373, EPI_ISL_2715374, EPI_ISL_2715375, EPI_ISL_2715376, EPI_ISL_2715377, EPI_ISL_2715378, EPI_ISL_2715379, EPI_ISL_2715380, EPI_ISL_2715381, EPI_ISL_2715382, EPI_ISL_2715383, EPI_ISL_2715384, EPI_ISL_2715385, EPI_ISL_2715386, EPI_ISL_2715387, EPI_ISL_2715388, EPI_ISL_2715389, EPI_ISL_2715390, EPI_ISL_2715391, EPI_ISL_2715392, EPI_ISL_2715393, EPI_ISL_2715394, EPI_ISL_2715395, EPI_ISL_2715396, EPI_ISL_2715397, EPI_ISL_2715398, EPI_ISL_2715399, EPI_ISL_2715400, EPI_ISL_2715401, EPI_ISL_2715402, EPI_ISL_2715403, EPI_ISL_2715404, EPI_ISL_2715405, EPI_ISL_2715406, EPI_ISL_2715407, EPI_ISL_2715408, EPI_ISL_2715409, EPI_ISL_2715410, EPI_ISL_2715411, EPI_ISL_2715412, EPI_ISL_2715413, EPI_ISL_2715414, EPI_ISL_2715415, EPI_ISL_2715416, EPI_ISL_2715417, EPI_ISL_2715418, EPI_ISL_2715419, EPI_ISL_2715420, EPI_ISL_2715421, EPI_ISL_2715422, EPI_ISL_2715423, EPI_ISL_2715424, EPI_ISL_2715425, EPI_ISL_2715426, EPI_ISL_2715427, EPI_ISL_2715428, EPI_ISL_2715429, EPI_ISL_2715430, EPI_ISL_2715431, EPI_ISL_2715432, EPI_ISL_2715433, EPI_ISL_2715434, EPI_ISL_2715435, EPI_ISL_2715436, EPI_ISL_2715437, EPI_ISL_2715438, EPI_ISL_2715439, EPI_ISL_2715440, EPI_ISL_2715441, EPI_ISL_2715442, EPI_ISL_2715443, EPI_ISL_2715444, EPI_ISL_2715445, EPI_ISL_2715446, EPI_ISL_2715447, EPI_ISL_2715448, EPI_ISL_2715449, EPI_ISL_2715450, EPI_ISL_2715451, EPI_ISL_2715452, EPI_ISL_2715453, EPI_ISL_2715454, EPI_ISL_2715455, EPI_ISL_2715456, EPI_ISL_2715457, EPI_ISL_2715458, EPI_ISL_2715459, EPI_ISL_2715460, EPI_ISL_2715461, EPI_ISL_2715462, EPI_ISL_2715463, EPI_ISL_2715464, EPI_ISL_2715465, EPI_ISL_2715466, EPI_ISL_2715467, EPI_ISL_2715468, EPI_ISL_2715469, EPI_ISL_2715470, EPI_ISL_2715471, EPI_ISL_2715472, EPI_ISL_2715473, EPI_ISL_2715474, EPI_ISL_2715475, EPI_ISL_2715476, EPI_ISL_2715477, EPI_ISL_2715478, EPI_ISL_2715479, EPI_ISL_2715480, EPI_ISL_2715481, EPI_ISL_2715482, EPI_ISL_2715483, EPI_ISL_2715484, EPI_ISL_2715485, EPI_ISL_2715486, EPI_ISL_2715487, EPI_ISL_2715488, EPI_ISL_2715489, EPI_ISL_2715490, EPI_ISL_2715491, EPI_ISL_2715492, EPI_ISL_2715493, EPI_ISL_2715494, EPI_ISL_2715495, EPI_ISL_2715496, EPI_ISL_2715497, EPI_ISL_2715498, EPI_ISL_2715499, EPI_ISL_2715500, EPI_ISL_2715501, EPI_ISL_2715502, EPI_ISL_2715503, EPI_ISL_2715504, EPI_ISL_2715505, EPI_ISL_2715506, EPI_ISL_2715507, EPI_ISL_2715508, EPI_ISL_2715509, EPI_ISL_2715510, EPI_ISL_2715511, EPI_ISL_2715512, EPI_ISL_2715513, EPI_ISL_2715514, EPI_ISL_2715515, EPI_ISL_2715516, EPI_ISL_2715517, EPI_ISL_2715518, EPI_ISL_2715519, EPI_ISL_2715520, EPI_ISL_2715521, EPI_ISL_2715522, EPI_ISL_2715523, EPI_ISL_2715524, EPI_ISL_2715525, EPI_ISL_2715526, EPI_ISL_2715527, EPI_ISL_2715528, EPI_ISL_2715529, EPI_ISL_2715530, EPI_ISL_2715531, EPI_ISL_2715532, EPI_ISL_2715533, EPI_ISL_2715534, EPI_ISL_2715535, EPI_ISL_2715536, EPI_ISL_2715537, EPI_ISL_2715538, EPI_ISL_2715539, EPI_ISL_2715540, EPI_ISL_2715541, EPI_ISL_2715542, EPI_ISL_2715543, EPI_ISL_2715544, EPI_ISL_2715545, EPI_ISL_2715546, EPI_ISL_2715547, EPI_ISL_2715548, EPI_ISL_2715549, EPI_ISL_2715550, EPI_ISL_2715551, EPI_ISL_2715552, EPI_ISL_2715553, EPI_ISL_2715554, EPI_ISL_2715555, EPI_ISL_2715556, EPI_ISL_2715557, EPI_ISL_2715558, EPI_ISL_2715559, EPI_ISL_2715560, EPI_ISL_2715561, EPI_ISL_2715562, EPI_ISL_2715563, EPI_ISL_2715564, EPI_ISL_2715565, EPI_ISL_2715566, EPI_ISL_2715567, EPI_ISL_2715568, EPI_ISL_2715569, EPI_ISL_2715570, EPI_ISL_2715571, EPI_ISL_2715572, EPI_ISL_2715573, EPI_ISL_2715574, EPI_ISL_2715575, EPI_ISL_2715576, EPI_ISL_2715577, EPI_ISL_2715578, EPI_ISL_2715579, EPI_ISL_2715580, EPI_ISL_2715581, EPI_ISL_2715582, EPI_ISL_2715583, EPI_ISL_2715584, EPI_ISL_2715585, EPI_ISL_2715586, EPI_ISL_2715587, EPI_ISL_2715588, EPI_ISL_2715589, EPI_ISL_2715590, EPI_ISL_2715591, EPI_ISL_2715592, EPI_ISL_2715593, EPI_ISL_2715594, EPI_ISL_2715595, EPI_ISL_2715596, EPI_ISL_2715597, EPI_ISL_2715598, EPI_ISL_2715599, EPI_ISL_2715600, EPI_ISL_2715601, EPI_ISL_2715602, EPI_ISL_2715603, EPI_ISL_2715604, EPI_ISL_2715605, EPI_ISL_2715606, EPI_ISL_2715607, EPI_ISL_2715608, EPI_ISL_2715609, EPI_ISL_2715610, EPI_ISL_2715611, EPI_ISL_2715612, EPI_ISL_2715613, EPI_ISL_2715614, EPI_ISL_2715615, EPI_ISL_2715616, EPI_ISL_2715617, EPI_ISL_2715618, EPI_ISL_2715619, EPI_ISL_2715620, EPI_ISL_2715621, EPI_ISL_2715622, EPI_ISL_2715623, EPI_ISL_2715624, EPI_ISL_2715625, EPI_ISL_2715626, EPI_ISL_2715627, EPI_ISL_2715628, EPI_ISL_2715629, EPI_ISL_2715630, EPI_ISL_2715631, EPI_ISL_2715632, EPI_ISL_2715633, EPI_ISL_2715634, EPI_ISL_2715635, EPI_ISL_2715636, EPI_ISL_2715637, EPI_ISL_2715638, EPI_ISL_2715639, EPI_ISL_2715640, EPI_ISL_2715641, EPI_ISL_2715642, EPI_ISL_2715643, EPI_ISL_2715644, EPI_ISL_2715645, EPI_ISL_2715646, EPI_ISL_2715647, EPI_ISL_2715648, EPI_ISL_2715649, EPI_ISL_2715650, EPI_ISL_2715651, EPI_ISL_2715652, EPI_ISL_2715653, EPI_ISL_2715654, EPI_ISL_2715655, EPI_ISL_2715656, EPI_ISL_2715657, EPI_ISL_2715658, EPI_ISL_2715659, EPI_ISL_2715660, EPI_ISL_2715661, EPI_ISL_2715662, EPI_ISL_2715663, EPI_ISL_2715664, EPI_ISL_2715665, EPI_ISL_2715666, EPI_ISL_2715667, EPI_ISL_2715668, EPI_ISL_2715669, EPI_ISL_2715670, EPI_ISL_2715671, EPI_ISL_2715672, EPI_ISL_2715673, EPI_ISL_2715674, EPI_ISL_2715675, EPI_ISL_2715676, EPI_ISL_2715677, EPI_ISL_2715678, EPI_ISL_2715679, EPI_ISL_2715680, EPI_ISL_2715681, EPI_ISL_2715682, EPI_ISL_2715683, EPI_ISL_2715684, EPI_ISL_2715685, EPI_ISL_2715686, EPI_ISL_2715687, EPI_ISL_2715688, EPI_ISL_2715689, EPI_ISL_2715690, EPI_ISL_2715691, EPI_ISL_2715692, EPI_ISL_2715693, EPI_ISL_2715694, EPI_ISL_2715695, EPI_ISL_2715696, EPI_ISL_2715697, EPI_ISL_2715698, EPI_ISL_2715699, EPI_ISL_2715700, EPI_ISL_2715701, EPI_ISL_2715702, EPI_ISL_2715703, EPI_ISL_2715704, EPI_ISL_2715705, EPI_ISL_2715706, EPI_ISL_2715707, EPI_ISL_2715708, EPI_ISL_2715709, EPI_ISL_2715710, EPI_ISL_2715711, EPI_ISL_2715712, EPI_ISL_2715713, EPI_ISL_2715714, EPI_ISL_2715715, EPI_ISL_2715716, EPI_ISL_2715717, EPI_ISL_2715718, EPI_ISL_2715719, EPI_ISL_2715720, EPI_ISL_2715721, EPI_ISL_2715722, EPI_ISL_2715723, EPI_ISL_2715724, EPI_ISL_2715725, EPI_ISL_2715726, EPI_ISL_2715727, EPI_ISL_2715728, EPI_ISL_2715729, EPI_ISL_2715730, EPI_ISL_2715731, EPI_ISL_2715732, EPI_ISL_2715733, EPI_ISL_2715734, EPI_ISL_2715735, EPI_ISL_2715736, EPI_ISL_2715737, EPI_ISL_2715738, EPI_ISL_2715739, EPI_ISL_2715740, EPI_ISL_2715741, EPI_ISL_2715742, EPI_ISL_2715743, EPI_ISL_2715744, EPI_ISL_2715745, EPI_ISL_2715746, EPI_ISL_2715747, EPI_ISL_2715748, EPI_ISL_2715749, EPI_ISL_2715750, EPI_ISL_2715751, EPI_ISL_2715752, EPI_ISL_2715753, EPI_ISL_2715754, EPI_ISL_2715755, EPI_ISL_2715756, EPI_ISL_2715757, EPI_ISL_2715758, EPI_ISL_2715759, EPI_ISL_2715760, EPI_ISL_2715761, EPI_ISL_2715762, EPI_ISL_2715763, EPI_ISL_2715764, EPI_ISL_2715765, EPI_ISL_2715766, EPI_ISL_2715767, EPI_ISL_2715768, EPI_ISL_2715769, EPI_ISL_2715770, EPI_ISL_2715771, EPI_ISL_2715772, EPI_ISL_2715773, EPI_ISL_2715774, EPI_ISL_2715775, EPI_ISL_2715776, EPI_ISL_2715777, EPI_ISL_2715778, EPI_ISL_2715779, EPI_ISL_2715780, EPI_ISL_2715781, EPI_ISL_2715782, EPI_ISL_2715783, EPI_ISL_2715784, EPI_ISL_2715785, EPI_ISL_2715786, EPI_ISL_2715787, EPI_ISL_2715788, EPI_ISL_2715789, EPI_ISL_2715790, EPI_ISL_2715791, EPI_ISL_2715792, EPI_ISL_2715793, EPI_ISL_2715794, EPI_ISL_2715795, EPI_ISL_2715796, EPI_ISL_2715797, EPI_ISL_2715798, EPI_ISL_2715799, EPI_ISL_2715800, EPI_ISL_2715801, EPI_ISL_2715802, EPI_ISL_2715803, EPI_ISL_2715804, EPI_ISL_2715805, EPI_ISL_2715806, EPI_ISL_2715807, EPI_ISL_2715808, EPI_ISL_2715809, EPI_ISL_2715810, EPI_ISL_2715811, EPI_ISL_2715812, EPI_ISL_2715813, EPI_ISL_2715814, EPI_ISL_2715815, EPI_ISL_2715816, EPI_ISL_2715817, EPI_ISL_2715818, EPI_ISL_2715819, EPI_ISL_2715820, EPI_ISL_2715821, EPI_ISL_2715822, EPI_ISL_2715823, EPI_ISL_2715824, EPI_ISL_2715825, EPI_ISL_2715826, EPI_ISL_2715827, EPI_ISL_2715828, EPI_ISL_2715829, EPI_ISL_2715830, EPI_ISL_2715831, EPI_ISL_2715832, EPI_ISL_2715833, EPI_ISL_2715834, EPI_ISL_2715835, EPI_ISL_2715836, EPI_ISL_2715837, EPI_ISL_2715838, EPI_ISL_2715839, EPI_ISL_2715840, EPI_ISL_2715841, EPI_ISL_2715842, EPI_ISL_2715843, EPI_ISL_2715844, EPI_ISL_2715845, EPI_ISL_2715846, EPI_ISL_2715847, EPI_ISL_2715848, EPI_ISL_2715849, EPI_ISL_2715850, EPI_ISL_2715851, EPI_ISL_2715852, EPI_ISL_2715853, EPI_ISL_2715854, EPI_ISL_2715855, EPI_ISL_2715856, EPI_ISL_2715857, EPI_ISL_2715858, EPI_ISL_2715859, EPI_ISL_2715860, EPI_ISL_2715861, EPI_ISL_2715862, EPI_ISL_2715863, EPI_ISL_2715864, EPI_ISL_2715865, EPI_ISL_2715866, EPI_ISL_2715867, EPI_ISL_2715868, EPI_ISL_2715869, EPI_ISL_2715870, EPI_ISL_2715871, EPI_ISL_2715872, EPI_ISL_2715873, EPI_ISL_2715874, EPI_ISL_2715875, EPI_ISL_2715876, EPI_ISL_2715877, EPI_ISL_2715878, EPI_ISL_2715879, EPI_ISL_2715880, EPI_ISL_2715881, EPI_ISL_2715882, EPI_ISL_2715883, EPI_ISL_2715884, EPI_ISL_2715885, EPI_ISL_2715886, EPI_ISL_2715887, EPI_ISL_2715888, EPI_ISL_2715889, EPI_ISL_2715890, EPI_ISL_2715891, EPI_ISL_2715892, EPI_ISL_2715893, EPI_ISL_2715894, EPI_ISL_2715895, EPI_ISL_2715896, EPI_ISL_2715897, EPI_ISL_2715898, EPI_ISL_2715899, EPI_ISL_2715900, EPI_ISL_2715901, EPI_ISL_2715902, EPI_ISL_2715903, EPI_ISL_2715904, EPI_ISL_2715905, EPI_ISL_2715906, EPI_ISL_2715907, EPI_ISL_2715908, EPI_ISL_2715909, EPI_ISL_2715910, EPI_ISL_2715911, EPI_ISL_2715912, EPI_ISL_2715913, EPI_ISL_2715914, EPI_ISL_2715915, EPI_ISL_2715916, EPI_ISL_2715917, EPI_ISL_2715918, EPI_ISL_2715919, EPI_ISL_2715920, EPI_ISL_2715921, EPI_ISL_2715922, EPI_ISL_2715923, EPI_ISL_2715924, EPI_ISL_2715925, EPI_ISL_2715926, EPI_ISL_2715927, EPI_ISL_2715928, EPI_ISL_2715929, EPI_ISL_2715930, EPI_ISL_2715931, EPI_ISL_2715932, EPI_ISL_2715933, EPI_ISL_2715934, EPI_ISL_2715935, EPI_ISL_2715936, EPI_ISL_2715937, EPI_ISL_2715938, EPI_ISL_2715939, EPI_ISL_2715940, EPI_ISL_2715941, EPI_ISL_2715942, EPI_ISL_2715943, EPI_ISL_2715944, EPI_ISL_2715945, EPI_ISL_2715946, EPI_ISL_2715947, EPI_ISL_2715948, EPI_ISL_2715949, EPI_ISL_2715950, EPI_ISL_2715951, EPI_ISL_2715952, EPI_ISL_2715953, EPI_ISL_2715954, EPI_ISL_2715955, EPI_ISL_2715956, EPI_ISL_2715957, EPI_ISL_2715958, EPI_ISL_2715959, EPI_ISL_2715960, EPI_ISL_2715961, EPI_ISL_2715962, EPI_ISL_2715963, EPI_ISL_2715964, EPI_ISL_2715965, EPI_ISL_2715966, EPI_ISL_2715967, EPI_ISL_2715968, EPI_ISL_2715969, EPI_ISL_2715970, EPI_ISL_2715971, EPI_ISL_2715972, EPI_ISL_2715973, EPI_ISL_2715974, EPI_ISL_2715975, EPI_ISL_2715976, EPI_ISL_2715977, EPI_ISL_2715978, EPI_ISL_2715979, EPI_ISL_2715980, EPI_ISL_2715981, EPI_ISL_2715982, EPI_ISL_2715983, EPI_ISL_2715984, EPI_ISL_2715985, EPI_ISL_2715986, EPI_ISL_2715987, EPI_ISL_2715988, EPI_ISL_2715989, EPI_ISL_2715990, EPI_ISL_2715991, EPI_ISL_2715992, EPI_ISL_2715993, EPI_ISL_2715994, EPI_ISL_2715995, EPI_ISL_2715996, EPI_ISL_2715997, EPI_ISL_2715998, EPI_ISL_2715999, EPI_ISL_2716000, EPI_ISL_2716001, EPI_ISL_2716002, EPI_ISL_2716003, EPI_ISL_2716004, EPI_ISL_2716005, EPI_ISL_2716006, EPI_ISL_2716007, EPI_ISL_2716008, EPI_ISL_2716009, EPI_ISL_2716010, EPI_ISL_2716011, EPI_ISL_2716012, EPI_ISL_2716013, EPI_ISL_2716014, EPI_ISL_2716015, EPI_ISL_2716016, EPI_ISL_2716017, EPI_ISL_2716018, EPI_ISL_2716019, EPI_ISL_2716020, EPI_ISL_2716021, EPI_ISL_2716022, EPI_ISL_2716023, EPI_ISL_2716024, EPI_ISL_2716025, EPI_ISL_2716026, EPI_ISL_2716027, EPI_ISL_2716028, EPI_ISL_2716029, EPI_ISL_2716030, EPI_ISL_2716031, EPI_ISL_2716032, EPI_ISL_2716033, EPI_ISL_2716034, EPI_ISL_2716035, EPI_ISL_2716036, EPI_ISL_2716037, EPI_ISL_2716038, EPI_ISL_2716039, EPI_ISL_2716040, EPI_ISL_2716041, EPI_ISL_2716042, EPI_ISL_2716043, EPI_ISL_2716044, EPI_ISL_2716045, EPI_ISL_2716046, EPI_ISL_2716047, EPI_ISL_2716048, EPI_ISL_2716049, EPI_ISL_2716050, EPI_ISL_2716051, EPI_ISL_2716052, EPI_ISL_2716053, EPI_ISL_2716054, EPI_ISL_2716055, EPI_ISL_2716056, EPI_ISL_2716057, EPI_ISL_2716058, EPI_ISL_2716059, EPI_ISL_2716060, EPI_ISL_2716061, EPI_ISL_2716062, EPI_ISL_2716063, EPI_ISL_2716064, EPI_ISL_2716065, EPI_ISL_2716066, EPI_ISL_2716067, EPI_ISL_2716068, EPI_ISL_2716069, EPI_ISL_2716070, EPI_ISL_2716071, EPI_ISL_2716072, EPI_ISL_2716073, EPI_ISL_2716074, EPI_ISL_2716075, EPI_ISL_2716076, EPI_ISL_2716077, EPI_ISL_2716078, EPI_ISL_2716079, EPI_ISL_2716080, EPI_ISL_2716081, EPI_ISL_2716082, EPI_ISL_2716083, EPI_ISL_2716084, EPI_ISL_2716085, EPI_ISL_2716086, EPI_ISL_2716087, EPI_ISL_2716088, EPI_ISL_2716089, EPI_ISL_2716090, EPI_ISL_2716091, EPI_ISL_2716092, EPI_ISL_2716093, EPI_ISL_2716094, EPI_ISL_2716095, EPI_ISL_2716096, EPI_ISL_2716097, EPI_ISL_2716098, EPI_ISL_2716099, EPI_ISL_2716100, EPI_ISL_2716101, EPI_ISL_2716102, EPI_ISL_2716103, EPI_ISL_2716104, EPI_ISL_2716105, EPI_ISL_2716106, EPI_ISL_2716107, EPI_ISL_2716108, EPI_ISL_2716109, EPI_ISL_2716110, EPI_ISL_2716111, EPI_ISL_2716112, EPI_ISL_2716113, EPI_ISL_2716114, EPI_ISL_2716115, EPI_ISL_2716116, EPI_ISL_2716117, EPI_ISL_2716118, EPI_ISL_2716119, EPI_ISL_2716120, EPI_ISL_2716121, EPI_ISL_2716122, EPI_ISL_2716123, EPI_ISL_2716124, EPI_ISL_2716125, EPI_ISL_2716126, EPI_ISL_2716127, EPI_ISL_2716128, EPI_ISL_2716129, EPI_ISL_2716130, EPI_ISL_2716131, EPI_ISL_2716132, EPI_ISL_2716133, EPI_ISL_2716134, EPI_ISL_2716135, EPI_ISL_2716136, EPI_ISL_2716137, EPI_ISL_2716138, EPI_ISL_2716139, EPI_ISL_2716140, EPI_ISL_2716141, EPI_ISL_2716142, EPI_ISL_2716143, EPI_ISL_2716144, EPI_ISL_2716145, EPI_ISL_2716146, EPI_ISL_2716147, EPI_ISL_2716148, EPI_ISL_2716149, EPI_ISL_2716150, EPI_ISL_2716151, EPI_ISL_2716152, EPI_ISL_2716153, EPI_ISL_2716154, EPI_ISL_2716155, EPI_ISL_2716156, EPI_ISL_2716157, EPI_ISL_2716158, EPI_ISL_2716159, EPI_ISL_2716160, EPI_ISL_2716161, EPI_ISL_2716162, EPI_ISL_2716163, EPI_ISL_2716164, EPI_ISL_2716165, EPI_ISL_2716166, EPI_ISL_2716167, EPI_ISL_2716168, EPI_ISL_2716169, EPI_ISL_2716170, EPI_ISL_2716171, EPI_ISL_2716172, EPI_ISL_2716173, EPI_ISL_2716174, EPI_ISL_2716175, EPI_ISL_2716176, EPI_ISL_2716177, EPI_ISL_2716178, EPI_ISL_2716179, EPI_ISL_2716180, EPI_ISL_2716181, EPI_ISL_2716182, EPI_ISL_2716183, EPI_ISL_2716184, EPI_ISL_2716185, EPI_ISL_2716186, EPI_ISL_2716187, EPI_ISL_2716188, EPI_ISL_2716189, EPI_ISL_2716190, EPI_ISL_2716191, EPI_ISL_2716192, EPI_ISL_2716193, EPI_ISL_2716194, EPI_ISL_2716195, EPI_ISL_2716196, EPI_ISL_2716197, EPI_ISL_2716198, EPI_ISL_2716199, EPI_ISL_2716200, EPI_ISL_2716201, EPI_ISL_2716202, EPI_ISL_2716203, EPI_ISL_2716204, EPI_ISL_2716205, EPI_ISL_2716206, EPI_ISL_2716207

| | | | |
|---|---|---|---|
| see above | Pandemic Response Lab - NYC | Pandemic Response Lab, R&D | Henry Lee, Michael Hammerling, Melissa Hopkins, Cybill del Castillo, Shinyoung Clair Kang, William Ward, Pradeep Bugga, Sol Rey, Dylan Law, Katharine Nelson, Haiping Hao, Jon Laurent |
| EPI_ISL_2716237, EPI_ISL_2716240, EPI_ISL_2716245 | Yale Clinical Virology Lab | Grubaugh Lab - Yale School of Public Health | Joseph Fauver, Mallery Breban, Isabel Ott, Tara Alpert, Mary Petrone, Anderson Brito, Chantal Vogels, Annie Watkins, Chaney Kalinich, Jessica Rothman, Marie L. Landry, Nathan Grubaugh |
| EPI_ISL_2727102 | URMC LABS | Wadsworth Center, New York State Department of Health | Kirsten St. George, Daryl M. Lamson, Alexis Russell, Matthew Shudt, Melissa A Leisner, Jonathan Plitnick, Catharine Prussing, Navjot Singh, John Kelly, Erasmus Schneider, Erica Lasek-Nesselquist |
| EPI_ISL_2727103 | WESTCHESTER MEDICAL CENTER | Wadsworth Center, New York State Department of Health | Kirsten St. George, Daryl M. Lamson, Alexis Russell, Matthew Shudt, Melissa A Leisner, Jonathan Plitnick, Catharine Prussing, Navjot Singh, John Kelly, Erasmus Schneider, Erica Lasek-Nesselquist |
| EPI_ISL_2727104 | ALBANY MEDICAL CENTER | Wadsworth Center, New York State Department of Health | Kirsten St. George, Daryl M. Lamson, Alexis Russell, Matthew Shudt, Melissa A Leisner, Jonathan Plitnick, Catharine Prussing, Navjot Singh, John Kelly, Erasmus Schneider, Erica Lasek-Nesselquist |
| EPI_ISL_2727105 | NORTHWELL HEALTH LABORATORIES | Wadsworth Center, New York State Department of Health | Kirsten St. George, Daryl M. Lamson, Alexis Russell, Matthew Shudt, Melissa A Leisner, Jonathan Plitnick, Catharine Prussing, Navjot Singh, John Kelly, Erasmus Schneider, Erica Lasek-Nesselquist |
| EPI_ISL_2727106, EPI_ISL_2727107, EPI_ISL_2727108, EPI_ISL_2727109 | URMC LABS | Wadsworth Center, New York State Department of Health | Kirsten St. George, Daryl M. Lamson, Alexis Russell, Matthew Shudt, Melissa A Leisner, Jonathan Plitnick, Catharine Prussing, Navjot Singh, John Kelly, Erasmus Schneider, Erica Lasek-Nesselquist |
| EPI_ISL_2727110 | MONTEFIORE MEDICAL CENTER LABORATORIES | Wadsworth Center, New York State Department of Health | Kirsten St. George, Daryl M. Lamson, Alexis Russell, Matthew Shudt, Melissa A Leisner, Jonathan Plitnick, Catharine Prussing, Navjot Singh, John Kelly, Erasmus Schneider, Erica Lasek-Nesselquist |
| EPI_ISL_2727111 | ALBANY MEDICAL CENTER | Wadsworth Center, New York State Department of Health | Kirsten St. George, Daryl M. Lamson, Alexis Russell, Matthew Shudt, Melissa A Leisner, Jonathan Plitnick, Catharine Prussing, Navjot Singh, John Kelly, Erasmus Schneider, Erica Lasek-Nesselquist |
| EPI_ISL_2727112 | TEMPUS LABS INC | Wadsworth Center, New York State Department of Health | Kirsten St. George, Daryl M. Lamson, Alexis Russell, Matthew Shudt, Melissa A Leisner, Jonathan Plitnick, Catharine Prussing, Navjot Singh, John Kelly, Erasmus Schneider, Erica Lasek-Nesselquist |
| EPI_ISL_2727113, EPI_ISL_2727114 | ALBANY MEDICAL CENTER | Wadsworth Center, New York State Department of Health | Kirsten St. George, Daryl M. Lamson, Alexis Russell, Matthew Shudt, Melissa A Leisner, Jonathan Plitnick, Catharine Prussing, Navjot Singh, John Kelly, Erasmus Schneider, Erica Lasek-Nesselquist |

EPI_ISL_2753940, EPI_ISL_2753941, EPI_ISL_2753942, EPI_ISL_2753943, EPI_ISL_2753944, EPI_ISL_2753945, EPI_ISL_2753946, EPI_ISL_2753947, EPI_ISL_2753948, EPI_ISL_2753949, EPI_ISL_2753950, EPI_ISL_2753951, EPI_ISL_2753952

| | | | |
|---|---|---|---|
| see above | NYU Langone Health | Departments of Pathology and Medicine, New York University | Adriana Heguy, Dacia Dimartino, Emily Guzman, Christian Marier, Peter Meyn, Sitharam Ramaswami, Gael Westby, Paul Zappile, Yutong Zhang, Paolo |

| | | | |
|---|---|---|---|
| | | School of Medicine | Cotzia, Guiqing Wang |
| EPI_ISL_2759142, EPI_ISL_2759143, EPI_ISL_2759144, EPI_ISL_2759145, EPI_ISL_2759146, EPI_ISL_2759147, EPI_ISL_2759148, EPI_ISL_2759149, EPI_ISL_2759150, EPI_ISL_2759151, EPI_ISL_2759152, EPI_ISL_2759153, EPI_ISL_2759154, EPI_ISL_2759155, EPI_ISL_2759156, EPI_ISL_2759157, EPI_ISL_2759158, EPI_ISL_2759159, EPI_ISL_2759160, EPI_ISL_2759161, EPI_ISL_2759162, EPI_ISL_2759163, EPI_ISL_2759164, EPI_ISL_2759165, EPI_ISL_2759166, EPI_ISL_2759167, EPI_ISL_2759168, EPI_ISL_2759169, EPI_ISL_2759170, EPI_ISL_2759171, EPI_ISL_2759172, EPI_ISL_2759173, EPI_ISL_2759174, EPI_ISL_2759175, EPI_ISL_2759176, EPI_ISL_2759177, EPI_ISL_2759178, EPI_ISL_2759179, EPI_ISL_2759180, EPI_ISL_2759181, EPI_ISL_2759182, EPI_ISL_2759183, EPI_ISL_2759184, EPI_ISL_2759185, EPI_ISL_2759186, EPI_ISL_2759187, EPI_ISL_2759188, EPI_ISL_2759189, EPI_ISL_2759190, EPI_ISL_2759191, EPI_ISL_2759192, EPI_ISL_2759193, EPI_ISL_2759194, EPI_ISL_2759195, EPI_ISL_2759196, EPI_ISL_2759197, EPI_ISL_2759198, EPI_ISL_2759199, EPI_ISL_2759200, EPI_ISL_2759201, EPI_ISL_2759202, EPI_ISL_2759203, EPI_ISL_2759204, EPI_ISL_2759205, EPI_ISL_2759206, EPI_ISL_2759207, EPI_ISL_2759208, EPI_ISL_2759209, EPI_ISL_2759210, EPI_ISL_2759211, EPI_ISL_2759212, EPI_ISL_2759213, EPI_ISL_2759214, EPI_ISL_2759215, EPI_ISL_2759216, EPI_ISL_2759217, EPI_ISL_2759218, EPI_ISL_2759219, EPI_ISL_2759220, EPI_ISL_2759221, EPI_ISL_2759222, EPI_ISL_2759223, EPI_ISL_2759224, EPI_ISL_2759225, EPI_ISL_2759226, EPI_ISL_2759227, EPI_ISL_2759228, EPI_ISL_2759229, EPI_ISL_2759230, EPI_ISL_2759231, EPI_ISL_2759232, EPI_ISL_2759233, EPI_ISL_2759234, EPI_ISL_2759235, EPI_ISL_2759236, EPI_ISL_2759237, EPI_ISL_2759238, EPI_ISL_2759239, EPI_ISL_2759240, EPI_ISL_2759241, EPI_ISL_2759242, EPI_ISL_2759243, EPI_ISL_2759244, EPI_ISL_2759245, EPI_ISL_2759246, EPI_ISL_2759247, EPI_ISL_2759248, EPI_ISL_2759249, EPI_ISL_2759250, EPI_ISL_2759251, EPI_ISL_2759252, EPI_ISL_2759253, EPI_ISL_2759254, EPI_ISL_2759255, EPI_ISL_2759256, EPI_ISL_2759257, EPI_ISL_2759258, EPI_ISL_2759259, EPI_ISL_2759260, EPI_ISL_2759261, EPI_ISL_2759262, EPI_ISL_2759263, EPI_ISL_2759264, EPI_ISL_2759265, EPI_ISL_2759266, EPI_ISL_2759267, EPI_ISL_2759268, EPI_ISL_2759269, EPI_ISL_2759270, EPI_ISL_2759271, EPI_ISL_2759272, EPI_ISL_2759273, EPI_ISL_2759274, EPI_ISL_2759275, EPI_ISL_2759276, EPI_ISL_2759277, EPI_ISL_2759278, EPI_ISL_2759279, EPI_ISL_2759280, EPI_ISL_2759281, EPI_ISL_2759282, EPI_ISL_2759283, EPI_ISL_2759284, EPI_ISL_2759285, EPI_ISL_2759286, EPI_ISL_2759287, EPI_ISL_2759288, EPI_ISL_2759289, EPI_ISL_2759290, EPI_ISL_2759291, EPI_ISL_2759292, EPI_ISL_2759293, EPI_ISL_2759294, EPI_ISL_2759295, EPI_ISL_2759296, EPI_ISL_2759297, EPI_ISL_2759298, EPI_ISL_2759299, EPI_ISL_2759300, EPI_ISL_2759301, EPI_ISL_2759302, EPI_ISL_2759303, EPI_ISL_2759304, EPI_ISL_2759305, EPI_ISL_2759306, EPI_ISL_2759307, EPI_ISL_2759308, EPI_ISL_2759309, EPI_ISL_2759310, EPI_ISL_2759311, EPI_ISL_2759312, EPI_ISL_2759313, EPI_ISL_2759314, EPI_ISL_2759315, EPI_ISL_2759316, EPI_ISL_2759317, EPI_ISL_2759318, EPI_ISL_2759319, EPI_ISL_2759320, EPI_ISL_2759321, EPI_ISL_2759322, EPI_ISL_2759323, EPI_ISL_2759324, EPI_ISL_2759325, EPI_ISL_2759326, EPI_ISL_2759327, EPI_ISL_2759328, EPI_ISL_2759329, EPI_ISL_2759330, EPI_ISL_2759331, EPI_ISL_2759332, EPI_ISL_2759333, EPI_ISL_2759334, EPI_ISL_2759335, EPI_ISL_2759336, EPI_ISL_2759337, EPI_ISL_2759338, EPI_ISL_2759339, EPI_ISL_2759340, EPI_ISL_2759341, EPI_ISL_2759342, EPI_ISL_2759343, EPI_ISL_2759344, EPI_ISL_2759345, EPI_ISL_2759346, EPI_ISL_2759347, EPI_ISL_2759348, EPI_ISL_2759349, EPI_ISL_2759350, EPI_ISL_2759351, EPI_ISL_2759352, EPI_ISL_2759353, EPI_ISL_2759354, EPI_ISL_2759355, EPI_ISL_2759356, EPI_ISL_2759357, EPI_ISL_2759358, EPI_ISL_2759359, EPI_ISL_2759360, EPI_ISL_2759361, EPI_ISL_2759362, EPI_ISL_2759363, EPI_ISL_2759364, EPI_ISL_2759365, EPI_ISL_2759366, EPI_ISL_2759367, EPI_ISL_2759368, EPI_ISL_2759369, EPI_ISL_2759370, EPI_ISL_2759371, EPI_ISL_2759372, EPI_ISL_2759373, EPI_ISL_2759374, EPI_ISL_2759375, EPI_ISL_2759376, EPI_ISL_2759377, EPI_ISL_2759378, EPI_ISL_2759379, EPI_ISL_2759380, EPI_ISL_2759381, EPI_ISL_2759382, EPI_ISL_2759383, EPI_ISL_2759384, EPI_ISL_2759385, EPI_ISL_2759386, EPI_ISL_2759387, EPI_ISL_2759388, EPI_ISL_2759389, EPI_ISL_2759390, EPI_ISL_2759391, EPI_ISL_2759392, EPI_ISL_2759393, EPI_ISL_2759394, EPI_ISL_2759395, EPI_ISL_2759396, EPI_ISL_2759397, EPI_ISL_2759398, EPI_ISL_2759399, EPI_ISL_2759400, EPI_ISL_2759401, EPI_ISL_2759402, EPI_ISL_2759403, EPI_ISL_2759404, EPI_ISL_2759405, EPI_ISL_2759406, EPI_ISL_2759407, EPI_ISL_2759408, EPI_ISL_2759409, EPI_ISL_2759410, EPI_ISL_2759411, EPI_ISL_2759412, EPI_ISL_2759413, EPI_ISL_2759414, EPI_ISL_2759415, EPI_ISL_2759416, EPI_ISL_2759417, EPI_ISL_2759418, EPI_ISL_2759419, EPI_ISL_2759420, EPI_ISL_2759421, EPI_ISL_2759422, EPI_ISL_2759423, EPI_ISL_2759424, EPI_ISL_2759425, EPI_ISL_2759426, EPI_ISL_2759427, EPI_ISL_2759428, EPI_ISL_2759429, EPI_ISL_2759430, EPI_ISL_2759431, EPI_ISL_2759432, EPI_ISL_2759433, EPI_ISL_2759434, EPI_ISL_2759435, EPI_ISL_2759436, EPI_ISL_2759437, EPI_ISL_2759438, EPI_ISL_2759439, EPI_ISL_2759440, EPI_ISL_2759441, EPI_ISL_2759442, EPI_ISL_2759443, EPI_ISL_2759444, EPI_ISL_2759445, EPI_ISL_2759446, EPI_ISL_2759447, EPI_ISL_2759448, EPI_ISL_2759449, EPI_ISL_2759450, EPI_ISL_2759451, EPI_ISL_2759452, EPI_ISL_2759453, EPI_ISL_2759454, EPI_ISL_2759455, EPI_ISL_2759456, EPI_ISL_2759457, EPI_ISL_2759458, EPI_ISL_2759459, EPI_ISL_2759460, EPI_ISL_2759461, EPI_ISL_2759462, EPI_ISL_2759463, EPI_ISL_2759464, EPI_ISL_2759465, EPI_ISL_2759466, EPI_ISL_2759467, EPI_ISL_2759468, EPI_ISL_2759469, EPI_ISL_2759470, EPI_ISL_2759471, EPI_ISL_2759472, EPI_ISL_2759473, EPI_ISL_2759474, EPI_ISL_2759475, EPI_ISL_2759476, EPI_ISL_2759477, EPI_ISL_2759478, EPI_ISL_2759479, EPI_ISL_2759480, EPI_ISL_2759481, EPI_ISL_2759482, EPI_ISL_2759483, EPI_ISL_2759484, EPI_ISL_2759485, EPI_ISL_2759486, EPI_ISL_2759487, EPI_ISL_2759488, EPI_ISL_2759489, EPI_ISL_2759490, EPI_ISL_2759491, EPI_ISL_2759492, EPI_ISL_2759493, EPI_ISL_2759494, EPI_ISL_2759495, EPI_ISL_2759496, EPI_ISL_2759497, EPI_ISL_2759498, EPI_ISL_2759499, EPI_ISL_2759500, EPI_ISL_2759501, EPI_ISL_2759502, EPI_ISL_2759503, EPI_ISL_2759504, EPI_ISL_2759505, EPI_ISL_2759506, EPI_ISL_2759507, EPI_ISL_2759508, EPI_ISL_2759509, EPI_ISL_2759510, EPI_ISL_2759511, EPI_ISL_2759512, EPI_ISL_2759513, EPI_ISL_2759514, EPI_ISL_2759515, EPI_ISL_2759516, EPI_ISL_2759517, EPI_ISL_2759518, EPI_ISL_2759519, EPI_ISL_2759520, EPI_ISL_2759521, EPI_ISL_2759522, EPI_ISL_2759523, EPI_ISL_2759524, EPI_ISL_2759525, EPI_ISL_2759526, EPI_ISL_2759527, EPI_ISL_2759528, EPI_ISL_2759529, EPI_ISL_2759530, EPI_ISL_2759531, EPI_ISL_2759532, EPI_ISL_2759533, EPI_ISL_2759534, EPI_ISL_2759535, EPI_ISL_2759536, EPI_ISL_2759537, EPI_ISL_2759538, EPI_ISL_2759539, EPI_ISL_2759540, EPI_ISL_2759541, EPI_ISL_2759542, EPI_ISL_2759543, EPI_ISL_2759544, EPI_ISL_2759545, EPI_ISL_2759546, EPI_ISL_2759547, EPI_ISL_2759548, EPI_ISL_2759549, EPI_ISL_2759550, EPI_ISL_2759551, EPI_ISL_2759552, EPI_ISL_2759553, EPI_ISL_2759554, EPI_ISL_2759555, EPI_ISL_2759556, EPI_ISL_2759557, EPI_ISL_2759558, EPI_ISL_2759559, EPI_ISL_2759560, EPI_ISL_2759561, EPI_ISL_2759562, EPI_ISL_2759563, EPI_ISL_2759564, EPI_ISL_2759565, EPI_ISL_2759566, EPI_ISL_2759567, EPI_ISL_2759568, EPI_ISL_2759569, EPI_ISL_2759570, EPI_ISL_2759571, EPI_ISL_2759572, EPI_ISL_2759573, EPI_ISL_2759574, EPI_ISL_2759575, EPI_ISL_2759576, EPI_ISL_2759577, EPI_ISL_2759578, EPI_ISL_2759579, EPI_ISL_2759580, EPI_ISL_2759581, EPI_ISL_2759582, EPI_ISL_2759583, EPI_ISL_2759584, EPI_ISL_2759585, EPI_ISL_2759586, EPI_ISL_2759587, EPI_ISL_2759588, EPI_ISL_2759589, EPI_ISL_2759590, EPI_ISL_2759591, EPI_ISL_2759592, EPI_ISL_2759593, EPI_ISL_2759594, EPI_ISL_2759595, EPI_ISL_2759596, EPI_ISL_2759597, EPI_ISL_2759598, EPI_ISL_2759599, EPI_ISL_2759600, EPI_ISL_2759601, EPI_ISL_2759602, EPI_ISL_2759603, EPI_ISL_2759604, EPI_ISL_2759605, EPI_ISL_2759606, EPI_ISL_2759607, EPI_ISL_2759608, EPI_ISL_2759609, EPI_ISL_2759610, EPI_ISL_2759611, EPI_ISL_2759612, EPI_ISL_2759613, EPI_ISL_2759614, EPI_ISL_2759615, EPI_ISL_2759616, EPI_ISL_2759617, EPI_ISL_2759618, EPI_ISL_2759619, EPI_ISL_2759620, EPI_ISL_2759621, EPI_ISL_2759622, EPI_ISL_2759623, EPI_ISL_2759624, EPI_ISL_2759625, EPI_ISL_2759626, EPI_ISL_2759627, EPI_ISL_2759628, EPI_ISL_2759629, EPI_ISL_2759630, EPI_ISL_2759631, EPI_ISL_2759632, EPI_ISL_2759633, EPI_ISL_2759634, EPI_ISL_2759635, EPI_ISL_2759636, EPI_ISL_2759637, EPI_ISL_2759638, EPI_ISL_2759639, EPI_ISL_2759640, EPI_ISL_2759641, EPI_ISL_2759642, EPI_ISL_2759643, EPI_ISL_2759644 | | | |
| see above | Population Medicine and Diagnostic Sciences, Cornell University | Population Medicine and Diagnostic Sciences, Cornell University | Mitchell,P.K., Caserta,L.C., Cronk,B.D., Anderson,R.R., Laverack,M., Venugopalan,R., Plocharczyk,E., Diel,D.G. |
